# Development of Portable Electronic Health Record Based Algorithms to Identify Individuals with Diabetic Retinopathy

**DOI:** 10.1101/2023.11.10.23298311

**Authors:** Joseph H. Breeyear, Sabrina L. Mitchell, Cari L. Nealon, Jacklyn N. Hellwege, Brian Charest, Anjali Khakharia, Christopher W. Halladay, Janine Yang, Gustavo A. Garriga, Otis D. Wilson, Til B. Basnet, Adriana M. Hung, Peter D. Reaven, James B Meigs, Mary K Rhee, Yang Sun, Mary G. Lynch, Lucia Sobrin, Milam A. Brantley, Yan V. Sun, Peter W. Wilson, Sudha K. Iyengar, Neal S. Peachey, Lawrence S. Phillips, Todd L. Edwards, Ayush Giri

## Abstract

**Objectives:** To develop, validate and implement algorithms to identify diabetic retinopathy (DR) cases and controls from electronic health care records (EHR)s. **Methods**: We developed and validated EHR-based algorithms to identify DR cases and individuals with type I or II diabetes without DR (controls) in three independent EHR systems: Vanderbilt University Medical Center Synthetic Derivative (VUMC), the VA Northeast Ohio Healthcare System (VANEOHS), and Massachusetts General Brigham (MGB). Cases were required to meet one of three criteria: 1) two or more dates with any DR ICD-9/10 code documented in the EHR, or 2) at least one affirmative health-factor or EPIC code for DR along with an ICD9/10 code for DR on a different day, or 3) at least one ICD-9/10 code for any DR occurring within 24 hours of an ophthalmology exam. Criteria for controls included affirmative evidence for diabetes as well as an ophthalmology exam.

**Results:** The algorithms, developed and evaluated in VUMC through manual chart review, resulted in a positive predictive value (PPV) of 0.93 for cases and negative predictive value (NPV) of 0.97 for controls. Implementation of algorithms yielded similar metrics in VANEOHS (PPV=0.94; NPV=0.86) and lower in MGB (PPV=0.84; NPV=0.76). In comparison, use of DR definition as implemented in Phenome-wide association study (PheWAS) in VUMC, yielded similar PPV (0.92) but substantially reduced NPV (0.48). Implementation of the algorithms to the Million Veteran Program identified over 62,000 DR cases with genetic data including 14,549 African Americans and 6,209 Hispanics with DR.

**Conclusions/Discussion:** We demonstrate the robustness of the algorithms at three separate health-care centers, with a minimum PPV of 0.84 and substantially improved NPV than existing high-throughput methods. We strongly encourage independent validation and incorporation of features unique to each EHR to enhance algorithm performance for DR cases and controls.

## INTRODUCTION

Diabetic retinopathy (DR), is a leading cause of visual impairment and preventable blindness among working-age adults, affecting an estimated 103 million individuals worldwide.[1] As the global prevalence of diabetes mellitus (DM) rises, the global prevalence of DR will also rise, with estimates suggesting up to 160 million individuals with DR by 2045.[1]

The prevalence of DR varies between individuals with type 1 and type 2 diabetes, and is estimated to be between 37% and 94% for type 1 DM and 35% for type 2 DM.[2] Nearly one-third of those with DR progress to vision-threatening DR over years of disease.[2-4] Early-stage DR, termed non-proliferative DR, is characterized by blot hemorrhages, cotton wool spots, intraretinal microvascular abnormalities, retinal microaneurysms, and venous beading. Based on the extent of retinal findings, non-proliferative DR is classified as mild, moderate, or severe. The more advanced proliferative diabetic retinopathy (PDR) is defined by neovascularization of the retina or iris.

The strongest established risk factors for DR and for progression to vision-threatening DR are DM duration and poor glycemic control, while other reported risk factors include hypertension, diabetic neuropathy, diabetic nephropathy, and increased body mass index [BMI].[4-6] Compared with populations of European descent, the prevalence of DR and vision-threatening DR is higher in African American and Hispanic populations.[7-11] Conducting epidemiologic and genetic studies of DR based on manual reviews of retinal imaging and electronic health record (EHR) data, or prospective evaluation has been challenging, resulting in smaller sample sizes. Additionally, many prospective studies have been performed primarily in individuals of European ancestry. At present, only a small number of genetic loci have been associated with DR in studies primarily recruiting patients with face-to-face encounters.[12-19]

Large-scale EHR-linked biobanks provide important opportunities to pursue additional loci and to identify genetic and lifestyle-related risk factors for DR. To leverage these resources for understanding the etiology behind DR, an efficient and accurate means to identify individuals with diabetes and DR as well as individuals with diabetes without DR is required. High-throughput approaches such as Phenome-wide association study (PheWAS) have provided the opportunity to leverage EHR systems for research, however, the validity of the algorithms/definitions used in PheWAS, especially in the context of DR, has not been evaluated to our knowledge. Having validated DR algorithms for EHRs would increase analytical sample sizes, thereby improving statistical power and enabling research groups to ask previously intractable research questions to improve our understanding of the underlying pathophysiology of DR. To date, a single validated EHR-based algorithm for DR progression is presented in literature, one that was developed and validated in a single EHR system.[20] Here, we describe the development and portability of EHR-based algorithms to identify individuals with DR (cases) and individuals with DM but without DR (controls) across three healthcare systems in the US.

## RESEARCH DESIGN and METHODS

In this study, we utilized EHR data from three independent healthcare systems, the Vanderbilt University Medical Center (VUMC Synthetic Derivative (SD)), the VA Northeast Ohio Healthcare System (VANEOHS) Eye Clinic, and the Massachusetts General Brigham (MGB) biobank. This study was conducted upon approval of Institutional Review Boards of VUMC, VANEOHS, MGB, and Million Veteran Program (MVP) central and local offices.

### Algorithm Development

Algorithm conceptualization and initial development was performed in the VUMC SD, a database containing de-identified clinical information derived from VUMC’s electronic medical records, described elsewhere.[21] Briefly, the VUMC SD contains records for >2.2 million individuals and includes diagnostic and procedure codes, demographics (age, sex, race), clinical notes, laboratory values, and medication information. The development process is summarized as three hierarchical steps: 1) Algorithm Design provides the basic framework; 2) Algorithm Construct entails defining criteria to construct a given algorithm; and 3) Operationalizing criteria, which entails finding EHR-specific features that fulfil criteria for the algorithm construct (**Supplemental Table 1**; **Figure 1**). Algorithm development was an iterative process which required moving between steps 2 and 3 to generate multiple versions of algorithms. For example, cases defined under **Version 0** (**V0**) required that cases have at least one ICD-9/10 code for any form/severity of DR, while controls were required to have at least one ICD-9/10 code for DM and no codes from the DM exclusion list (**Supplemental Tables 1 – 3**) and at least one eye exam without codes for DR. In **V1**, we required cases to have at least two unique dates with an ICD-9/10 code for any form of DR, and controls to have at least three dates with an ICD-9/10 code for DM without any DR codes. This additional requirement for control was put in place to improve confidence that a given patient indeed had diabetes during chart review. In **V2**, we relaxed the algorithm for cases by adding an eligibility criterion allowing them to have a single ICD-9/10 code for any form of DR if accompanied within 24 hours of an ophthalmology exam. Along with the DR algorithm, we developed an algorithm for PDR based on restricting criteria in the “any DR” algorithm to only PDR ICD-9/10 codes.

**Figure 1.**
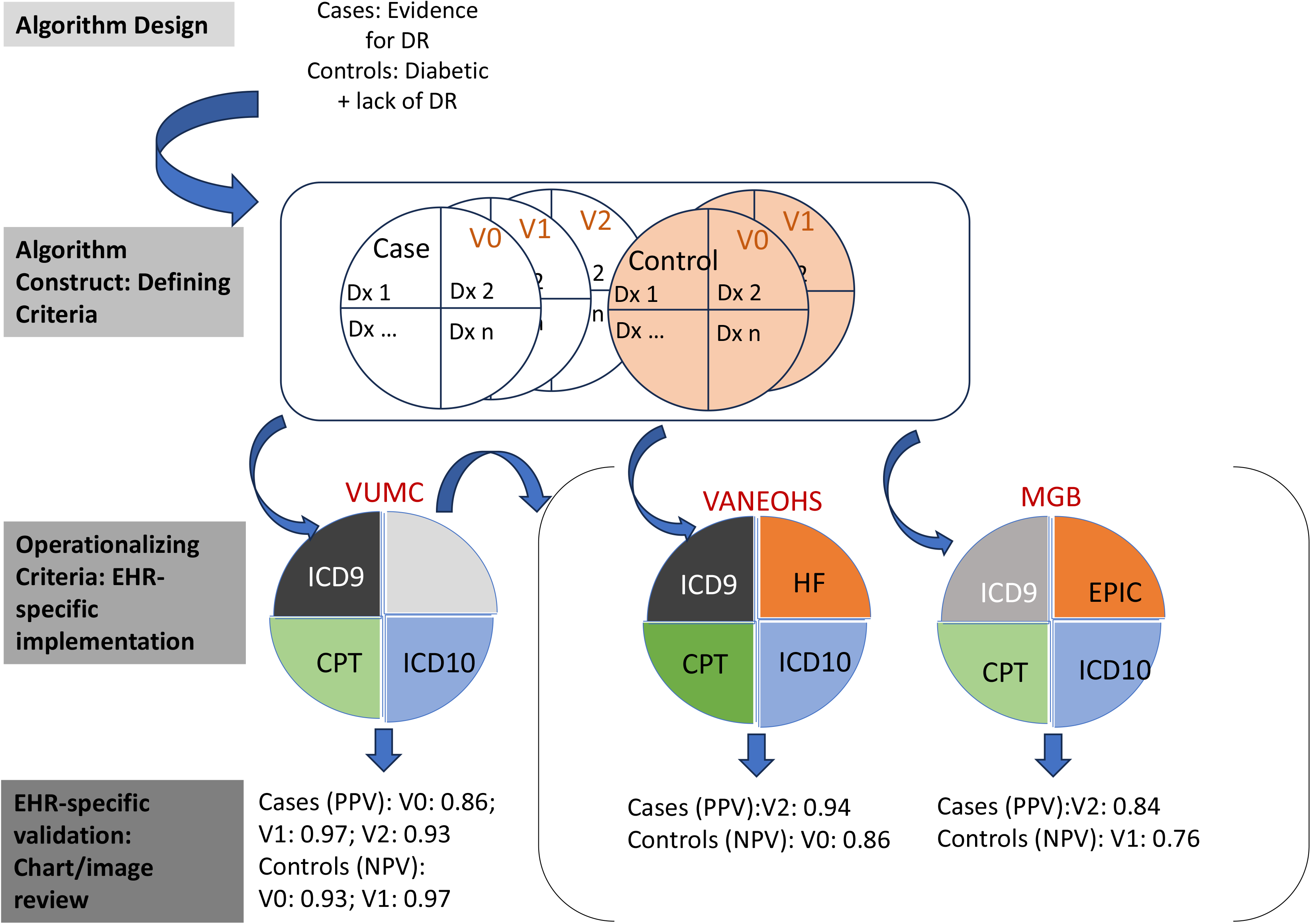
Flowchart describing stages of development and validation across three Electronic Health Record systems.

### Algorithm Portability: Operationalizing Criteria through EHR-specific implementation across institutions

#### VA Northeast Ohio Healthcare System

In the VANEOHS EHR, individuals with DM were first identified using a combination of diagnosis and procedure codes, clinic stop codes, and medications. Briefly, individuals with affirmative evidence of DM were identified based on presence of a diagnosis code at one or more face-to-face outpatient primary care physician visits or a DM diagnosis code at two or more outpatient visits of any kind, and at least one outpatient fill of a diabetes-indicated medication (**Supplemental Table 3**). In addition to ICD9/10 codes the VA also routinely utilizes Health Factor codes to annotate health records. To take advantage of this additional information source, the case-algorithm construct (**V2**) was operationalized in the VA to include Health Factor codes in addition ICD-9/10 diagnostic codes. The algorithm was subsequently applied in the U.S. Department of Veterans Affairs biorepository, the Million Veteran Program.

#### Mass General Brigham

In the MGB, individuals with DM were first identified with a previously curated DM phenotype.[22] To identify DR cases, we applied the DR case-algorithm construct (**V2**), whose criteria were operationalized to include an EPIC Provider code along with an ICD-9/10 code for any form of DR on unique days, to take advantage of features unique to MGB. We used the more stringent algorithm construct for controls in MGB (**V1**), which requires at least three unique days of diabetes codes to be included as controls, as applying only one ICD code for diabetes (**V0**) yielded a large proportion of records where diabetes status could not be validated with manual review.

### Algorithm Validation

In the VUMC SD, we conducted a blinded manual review of 495 de-identified patient EHRs by an ophthalmologist and 4 other trained reviewers, identifying DR status, DR severity, and control status, and compared the results to the algorithm definitions (**Supplemental Figure 1**). Of the 495 records manually reviewed, 45 were overlapped among reviewers to assess reviewer concordance. Performance of the algorithms was primarily assessed by computing positive predictive values (PPV) for cases and negative predictive values (NPV) for controls. Sensitivity and specificity were computed from resulting tabulations. If an algorithmically determined case or control could not be validated with manual review, they were classified as undetermined and were excluded from the numerator but included in the denominator for computing NPV. For the 45 overlapped records, if the reviewers disagreed, we used a conservative approach to include the manual review that did not bias our results towards a higher PPV or NPV. As we could not manually review retinal imaging data for individuals in the VUMC SD, we captured a subset of EHRs (n = 152) from the 495 that included EHR notes from the Vanderbilt Eye Institute (VEI). These VEI notes indicate that an individual was assessed in the clinic with a retinal exam with an ophthalmologist reporting on their findings. Of the 45 overlapped EHRs, 21 had VEI notes. Finally, in the VUMC SD, we compared the performance of our algorithm with the widely used PheCode definition of DR, by evaluating test characteristics of DR cases and controls identified using the PheCode definition.[23]

At the VANEOHS, we pulled 50 algorithm-defined cases and 50 algorithm-defined controls for chart review. Charts were reviewed by a single optometrist that was blinded to the algorithm assignments. Twenty-one additional charts were pulled for further review to improve precision of the PDR algorithm since there were too few PDR cases in the first 50 records with any-DR. Of the 50 any-DR cases, we identified 105 visits for 24 DR patients with at least one fundus imaging stored for manual review. We investigated the performance of the algorithm against these fundus images manually reviewed and staged by an optometrist.[24]

In the MGB biobank, we pulled 50 algorithm-defined cases (**V2**) and 50 algorithm-defined controls (**V1**) for chart review. Charts were reviewed by a fellowship-trained retinal specialist that was blinded to the algorithm assignments.

### Phenome-Wide Association Study

We performed a phenome-wide association study (PheWAS) of the (**V2**) DR algorithm in VUMC SD (n_max_ = 12,991) and the MVP (n_max_ = 165,322) leveraging the full catalog of ICD-9/10 codes.[25] The ∼1,800 PheCode traits were modeled with logistic regressions as a function of the (**V2**) algorithm designation, adjusted for age, sex, and the duration of DM, stratified by self-reported race/ethnicity (non-Hispanic White (NHW), non-Hispanic Black (NHB), and Hispanic (HIS)). Interpretation of results were limited to phenotypes with 20 or more cases. The results were meta-analyzed both within and across populations. Bonferroni-corrected statistical significance threshold was set to *p* - value ≤ 2.75 × 10^-5^. Analyses were conducted in R with the PheWAS package.[26]

## RESULTS

We compared the performance of the three DR algorithm versions developed in the VUMC SD dataset (**Table 1**). The initial algorithm (**V0**), aimed at identifying possible cases and controls, resulted in a PPV and NPV of 0.857 and 0.934, respectively. The second algorithm (**V1**), designed to increase algorithm precision, resulted in a substantially improved PPV (0.969) and provided minimal gains on NPV (0.966) compared to the V0 versions. The final algorithm for cases (**V2**), intended to maximize cases and maintain a high PPV by including ophthalmology exam data along with a single DR ICD code instead of two, resulted in a similar PPV (0.930) as **V1**, while increasing case-numbers. Compared to the PheCode definition (PPV = 0.922, NPV = 0.480), our algorithm classified cases (**V2**) with a similar PPV, but a considerably higher NPV (**V0** and **V1**). The same upward trend was seen for the PDR algorithm, with the PPV increasing from 0.802 to 0.914 (**Table 1**). In the subset of EHRs with VEI notes, we saw similar algorithm performance for PPV (0.938) and improved performance for NPV (1.0). The concordance of the five reviewers for DM, DR, and PDR designations was 96%, 91%, and 97%, respectively (**Supplemental Table 4**). When restricted to EHRs with notes from the Vanderbilt Eye Institute, the concordance for DM, DR, and PDR designations was 100%, 95%, and 100% of the individuals, respectively (**Supplemental Table 4**).

**Table 1.** Development and Evaluation of the Electronic Health Record-Based Diabetic Retinopathy Algorithm.

**Figure 1. Flowchart describing stages of development and validation across three Electronic Health Record systems**
|  |  |  |  |  |  |  |  |
| --- | --- | --- | --- | --- | --- | --- | --- |
| <u>Development</u> | Diabetic Retinopathy |  |  |  | Proliferative Diabetic Retinopathy |  |  |
|  | VUMC |  |  |  | VUMC |  |  |
|  | <u>Version 0</u> | <u>Version 1</u> | <u>Version 2</u> | <u>PheCode</u> | <u>Version 0</u> | <u>Version 1</u> | <u>Version 2</u> |
| <u>Manual Review</u> |  |  |  |  |  |  |  |
| Case (n) | 309 | 220 | 253 | 253 | 86 | 66 | 71 |
| Control (n) | 125 | 62 | 72 | 72 | 362* | 219* | 259* |
| Undetermined (n) | 14 | 3 | 5 | 5 | 0 | 0 | 0 |
| <u>Algorithm</u> |  |  |  |  |  |  |  |
| Case (n) | 357 | 226 | 271 | 205 | 101 | 59 | 70 |
| Control (n) | 91 | 59 | 59 | 125 | 347* | 226* | 260* |
| PPV | 0.857 | 0.969 | 0.930 | 0.922 | 0.802 | 0.932 | 0.914 |
| NPV | 0.934 | 0.966 | 0.966 | 0.480 | 0.986 | 0.951 | 0.973 |
| <u>Evaluation</u> | Diabetic Retinopathy |  |  |  | Proliferative Diabetic Retinopathy |  |  |
|  | VUMC<br>n = 330 | VANEOHS<br>n = 100 | MGB<br>n = 100 |  | VUMC<br>n = 330 | VANEOHS<br>n = 121 |  |
| PPV | 0.930 | 0.940 | 0.840 |  | 0.914 | 0.818 |  |
| NPV | 0.966 | 0.860 | 0.760 |  | 0.973 | 0.989 |  |
| * Includes individuals with less severe forms of diabetic retinopathy and those without diabetic retinopathy |  |  |  |  |  |  |  |

The algorithms performed well in VANEOHS, where the DR algorithm PPV was 0.940 and PDR algorithm PPV was 0.818 (**Table 1, Supplemental Table 5**). There was a slight drop in the NPV for the VANOEHS (NPV = 0.860), while the NPV for the PDR algorithm (NPV = 0.989) was comparable to the VUMC. Among the 50 algorithmically-defined any-DR cases, we found fundus photography images associated with 105 visits for 24 patients. On a visit-level, 99 of 105 (PPV = 0.94) algorithmically defined cases were verified to have DR by manual review of imaging. Six DR-determinations could not be verified directly via imaging due to poor image quality. Of the 99 visits with adequate image quality, staging of DR (background/mild non-proliferative diabetic retinopathy, moderate non-proliferative diabetic retinopathy, severe non-proliferative diabetic retinopathy, and proliferative diabetic retinopathy) agreed for 84 images between our manual review and algorithm-determined diagnosis (Supplemental Table 10). The algorithm tended to underestimate the staging of DR compared to manual review for the other remaining 15 images. On a patient-level 23 of the 24 patients (PPV = 0.96) were verified as DR cases through manual review of images, and 21 of 24 were verified for staging of DR. Application of the algorithm to MGB showed lower PPV (0.840) for **V2** cases and lower NPV (0.760) for **V1** controls compared to VUMC SD and VANEOHS.

We applied these algorithms to identify DR cases and DM controls in three separate EHR systems: 8,860 DR cases (**V2**) and 15,522 controls (**V0**) in the VUMC SD, 62,101 DR cases (**V2**) and 110,688 controls (**V0**) in the MVP, and 471 DR cases (**V2**) and 1,137 controls (**V1**) in the MGB (**Table 2**). The three cohorts differed substantially in their sex composition with VUMC SD and MGB having approximately 50% females, whereas MVP consisted of predominantly males (>95%) (**Table 2**). Despite these major differences in cohort compositions, seminal observations for DR cases and controls were consistent with those in epidemiologic literature, where individuals with DR on average were more likely to have diabetic macular edema, neovascular glaucoma, diabetic neuropathy, and diabetic nephropathy in both the VUMC SD and MVP EHR systems. By design, diabetics without DR were more likely to be older than DR cases across all three EHR systems. Average duration of diabetes, captured as time since first diagnosis of diabetes in EHR or first mention of antihypertensive medication in EHR, whichever came first, was similar between VUMC and MVP cases, while controls in the MVP had a longer duration than in VUMC. At VUMC SD, controls (**V1**) were more likely to have diagnostic codes for type II diabetes, more likely to have hypertension than DR cases and more likely to have ophthalmic visits in the last five years. Such phenomenon was not observed in the MVP EHR system. Cases and controls were similar on average with regard to BMI and had similar proportions of inherited retinal disease in VUMC SD and MVP.

**Table 2:** Characteristics of Algorithm-Identified Cohorts in VUMC, MVP, and MGB.

| Characteristic | VUMC |  |  | MVP |  | MGB |  |
| --- | --- | --- | --- | --- | --- | --- | --- |
|  | DR Cases<br>(N=8,860) | Controls (V0)<br>(N=15,522) | Controls (V1)<br>(N=11,802) | DR Cases<br>(N=62,101) | Controls<br>(N=110,688) | DR Cases<br>(N=471) | Controls<br>(N=1,137) |
| **Mean Age ± SD, years** | 58.9 ± 14.2 | 63.7 ± 14.8 | 63.1 ± 15.0 | 63.1 ± 9.8 | 68.5 ± 10.4 | 67.9 ± 12.4 | 69.1 ± 13.1 |
| **Mean BMI ± SD, kg/m<sup>2</sup>** | 32.0 ± 7.7 | 32.5 ± 7.8 | 32.7 ± 7.7 | 32.5 ± 6.2 | 32.1 ± 6.5 | — | — |
| **Mean Diabetes Duration ± SD, years** | 6.9 ± 5.3 | 4.5 ± 4.4 | 5.1 ± 4.5 | 6.2 ± 5.1 | 9.3 ± 5.4 | — | — |
| **Female, N (%)** | 4,441 (50%) | 8,209 (53%) | 6,201 (53%) | 2,648 (4.3%) | 7,229 (6.5%) | 219 (46%) | 512 (45%) |
| **Race/ethnicity** |  |  |  |  |  |  |  |
| **European American, N (%)** | 5,912 (67%) | 11,333 (72%) | 8,770 (74%) | 38,507 (62%) | 72,999 (66%) | 268 (57%) | 847 (74%) |
| **African American, N (%)** | 1,933 (22%) | 2,799 (18%) | 2,286 (19%) | 14,549 (23%) | 23,113 (21%) | 75 (16%) | 119 (10%) |
| **Hispanic, N (%)** | 204 (0.3%) | 259 (2%) | 208 (1.8%) | 6,209 (10%) | 9,596 (8.7%) | 81 (17%) | 32 (2.8%) |
| **Other, N (%)** | 811 (9.2%) | 1,131 (7%) | 556 (4.7%) | 2,836 (4.6%) | 4,980 (4.5%) | 47 (10%) | 139 (12%) |
| **Type 2 DM, N (%) \*** | 7,437 (84%) | 14,240 (92%) | 10,802 (92%) | 61,429 (99%) | 109,724 (99%) | — | — |
| **Genotyped, %** | 25% | 26% | 31% | 100% | 100% | 100% | 100% |
| **OPT exam in last 5 years, N (%)** | 2,243 (25%) | 6,597 (43%) | 4,851 (41%) | 44,726 (72%) | 78,495 (71%) | — | — |
| **Diabetic macular edema, N (%)** | 2,167 (25%) | 45 (0.3%) | 33 (0.3%) | 14,180 (22%) | 63 (0.05%) | — | — |
| **Neovascular glaucoma, N (%)** | 501 (5.7%) | 165 (1%) | 107 (0.9%) | 1,723 (2.7%) | 1,634 (1.5%) | — | — |
| **Hypertension, N (%)** | 4,927 (56%) | 9,737 (63%) | 8,288 (70%) | 36,342 (58%) | 66,114 (60%) | — | — |
| **Diabetic Neuropathy, N (%)** | 2,793 (32%) | 3,127 (20%) | 3,005 (25%) | 43,111 (69%) | 51,679 (47%) | — | — |
| **Diabetic Nephropathy, N (%)** | 2,157 (24%) | 2,035 (13%) | 1,976 (17%) | 27,783 (44%) | 24,348 (22%) | — | — |
| **Inherited retinal disease, N (%)** | 54 (0.6%) | 128 (0.8%) | 85 (0.7%) | 459 (0.7%) | 607 (0.5%) | — | — |
| **Age-related macular degeneration, N (%)** | 637 (7.3%) | 1,503 (10%) | 1,079 (9%) | 7,678 (12%) | 9,622 (6.7%) | — | — |
| *Type II DM was considered based on a 3:1 Type II to Type I ICD code ratio |  |  |  |  |  |  |  |

### Phenome-Wide Association Study

To systematically evaluate the relationship of algorithmically defined DR cases and controls with the phenome derived from EHR, we performed a Phenome-wide association study across study populations in the MVP and VUMC SD. The DR algorithm PheWAS identified a total of 959 unique PheCodes significantly associated with DR, with 99, 8, 8, and 7 significantly associated in the Multi, NHW, NHB, and HIS populations, respectively (**Table 3, Supplemental Tables 6 - 9**). In the Multi-ancestry meta-analysis, a total of 784 PheCodes were associated with increased DR risk while 151 were associated with decreased DR risk. The most significantly associated PheCodes included Diabetic Retinopathy, Type 2 DM with Ophthalmic Manifestations, and Type 1 DM with Ophthalmic Manifestations, all with *p* – values below 1.00 x 10^-305^. In addition, the DR algorithm designation was associated with other phenotype groups related to downstream consequences of DM in the multi-ancestry meta-analysis, including metabolic disorders (82), cardiovascular disorders (127), and renal disorders (70).

**Table 3.** Diabetes PheCodes Associated with the Diabetic Retinopathy Algorithm Designation.

|  |  |  |  |  |  |
| --- | --- | --- | --- | --- | --- |
| **Phenotype** | **PheCode** | **<u>Multi</u>**<br>**n<sub>max</sub> = 187,581**<br>**OR (95% CI)** | **<u>Non-Hispanic White</u>**<br>**n<sub>max</sub> = 128,777**<br>**OR (95% CI)** | **<u>Non-Hispanic Black</u>**<br>**n<sub>max</sub> = 43,183**<br>**OR (95% CI)** | **<u>Hispanic</u>**<br>**n<sub>max</sub> = 15,621**<br>**OR (95% CI)** |
| Diabetes Mellitus | 250 | 2.25 (2.16 – 2.35) | 2.17 (2.07 – 2.28) | 2.39 (2.21 – 2.58) | 68 (33 – 139) |
| Type I DM (T1D) | 250.1 | 3.48 (3.44 – 3.52) | 3.43 (3.38 – 3.48) | 3.60 (3.51 – 3.68) | 3.56 (3.42 – 3.71) |
| Type II DM (T2D) | 250.2 | 1.75 (1.66 – 1.85) | 1.62 (1.52 – 1.72) | 2.12 (1.88 – 2.39) | 46.4 (25.8 – 83.5) |
| T2D with Renal Manifestations | 250.22 | 3.25 (3.22 – 3.29) | 3.04 (2.99 – 3.08) | 3.63 (3.54 – 3.71) | 4.24 (4.08 – 4.41) |
| T2D with Neurological Manifestations | 250.24 | 2.91 (2.88 – 2.94) | 2.71 (2.68 – 2.75) | 3.23 (3.16 – 3.30) | 3.80 (3.67 – 3.94) |
| Polyneuropathy in DM | 250.6 | 3.05 (3.02 – 3.09) | 2.88 (2.84 – 2.93) | 3.41 (3.32 – 3.51) | 3.96 (3.78 – 4.14) |
| Diabetic Retinopathy | 250.7 | 419 (409 – 429) | 414 (402 – 426) | 383 (365 – 403) | 635 (580 – 696) |

## DISCUSSION

In this study, we repurposed diagnostic codes and administrative data routinely collected in clinical practice to develop algorithms that identify individuals with diabetes, with and without diabetic retinopathy, to be used for research purposes. We involved three separate healthcare systems to confirm the portability of the DR algorithms, along with a comparison of the algorithm with manually reviewed fundus imaging. While identification of individuals with DR typically relies on manual review or machine learning algorithms along with retinal imaging data, our algorithm focused on ICD diagnostic codes and is thus transferable to other sites, allows for modifications to capitalize on unique EHR features, and requires significantly fewer resources for identifying at-risk individuals. The DR algorithms tested accurately identified individuals with DR, with PPVs ranging from 0.840 – 0.944 and identified those with diabetes and without DR with NPVs ranging from 0.76 to 0.97. This is a vast improvement over existing algorithms implemented in high-throughput methods such as PheWAS (PPV = 0.93; NPV = 0.48) and represents a starting point for researchers looking to maximize use of EHR for DR research.

To date, most studies have used one of three methods to identify individuals with DR: 1) manual review of eye examination findings, (2) manual review of retinal imaging, or (3) machine learning algorithms to review retinal imaging data.[12 17 27 28] While these methods are considered the gold standard for phenotyping, they are labor intensive and can require retinal imaging data, which is not always available in research datasets. The largest DR genome-wide association study to date, including 19,372 cases and 19,615 controls, involved the manual review of retinal images for more than 38,987 individuals from multiple studies.[12] As studies of DR are often under-powered, a method to easily identify individuals with DR with relatively minimal effort while maintaining high accuracy through EHRs offers an attractive opportunity to scale up research related to DR. We first show through manual review of records that the algorithms accurately identify individuals coded as DR cases and controls. Following the premise that DR diagnoses of patients seen by eye-specialists would be most reliable, we stratified cases and controls in the VUMC SD chart review validation by whether they visited the Vanderbilt Eye Institute. We showed patients who had VEI notes had higher, yet comparable, accuracy as cases and controls who were not seen by eye-specialists suggesting the high PPVs and NPVs were not entirely driven by one stratum. Then in the VA, we verified high PPV of our any-DR case-algorithm by manually reviewing 115 fundus images for 24 of the 50 any-DR cases for whom images were stored in the VA system. Further evaluation of DR-staging between algorithm and manual review showed high accuracy with close matches for 84 of 99 images with good quality. Reasons for incomplete availability of images include: the VA does not systematically store fundus photography images; imaging could have been performed at a different VA location and the images would not be available at the local VA where the review was conducted, or the diagnosis may have come from outside of the VA system.

As with any study incorporating EHR data, there is a possibility of incomplete data, since individuals may receive care outside of a particular EHR system. Our experiences of developing and validating algorithms for DR cases and controls across three heterogeneous EHR systems has led to considerations and insights worth considering in the usage and interpretation of data derived from these sources, especially in the context of DR (**Table 4**). EHR systems, designed for healthcare utilization, often have center-specific methods and customizations, even within those using a single EHR platform (i.e. EPIC). As was the case in our study, data accessibility across EHR systems varies greatly either due to heterogenous data collection systems, provider practices, or due to data availability constraints imposed across each EHR system. For example, in the MGB we found a sizable proportion of individuals were missing adequate EHR notes to validate ICD diagnoses. At the expense of losing controls, we limited evaluation of records with 3 or more diabetes diagnosis codes (**V1** control algorithm). While this alleviated the problem and still, 13% of individuals were still missing adequate EHR notes needed for validation. Since these undetermined individuals were conservatively counted against the PPV and NPV to avoid inflation of predictive values, the resulting PPV and NPV are noticeably lower than the VUMC and VANEOHS.

**Table 4.**
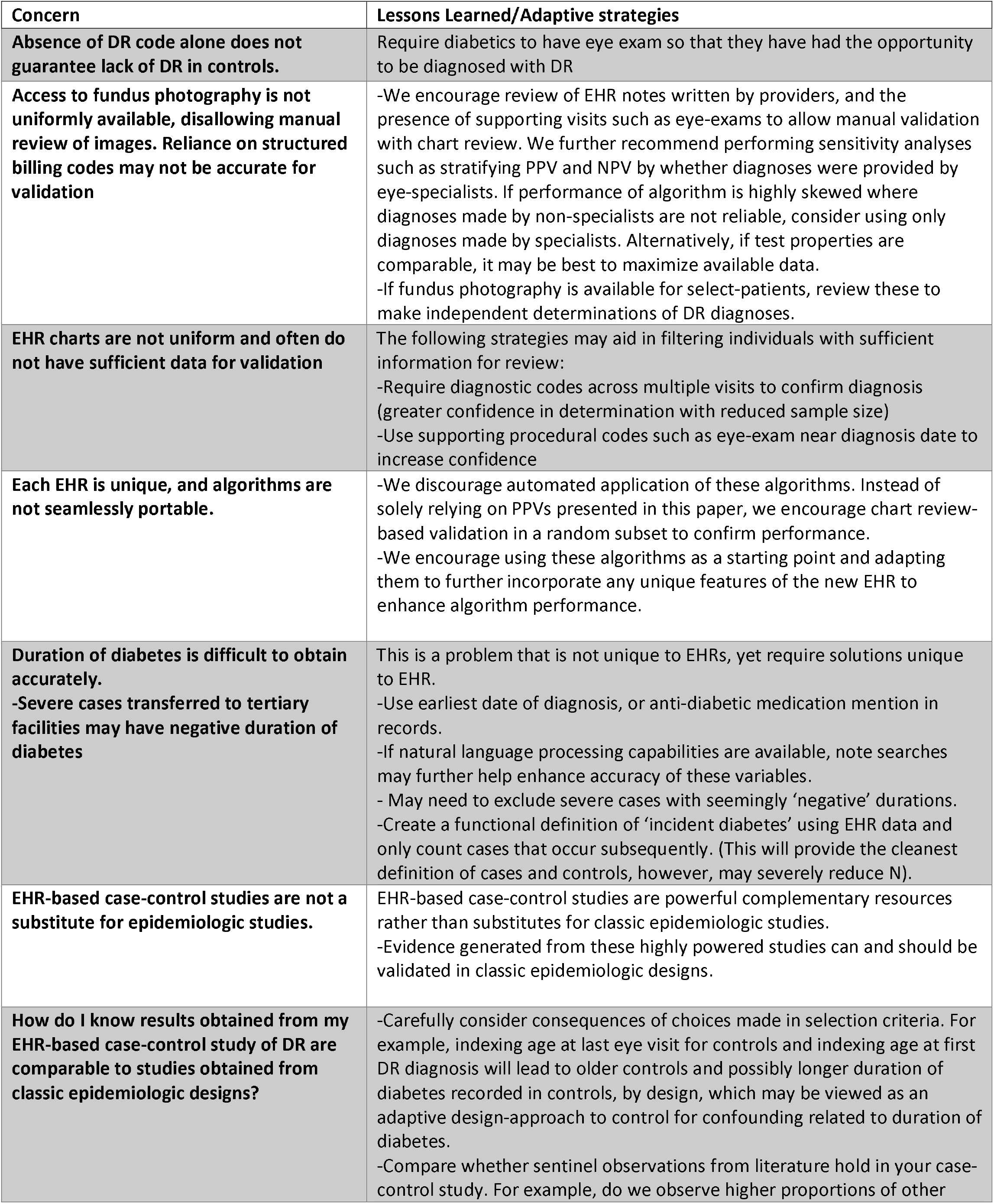
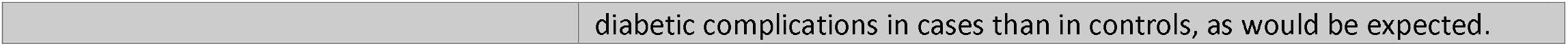
Lessons learned and adaptive strategies in developing case-control algorithms for DR in electronic health records (EHR).

Furthermore, we found that in addition to using common features across systems, embracing unique aspects of each system to retain as much detail as available greatly improves portability and validity of these algorithms. For example, in addition to the standard ICD9/10 diagnostic criteria, that formed the backbone for the primary algorithm at VUMC, the VANEOHS and the MGB systems each had additional coding systems that were unique to them, which we found more beneficial to incorporate rather than ignore. Limiting our algorithm to ICD9/10 codes and ignoring the Health Factor coding system in the VA would have led to more than 5,000 individuals misclassified as controls. In addition, we were able to capitalize on the richness of the VA EHR, which included detailed information on prescription, and the type of prescribing physician (primary care provider, specialist, inpatient, outpatient, etc.) to gain affirmative evidence of diabetes status, features that were not accessible for the VUMC SD or MGB.

We tested features of the algorithms which have consequences on predictive properties, on the number and the composition of individuals identified. Requiring two or more unique days with a diagnostic code for DR substantially improved PPV, but it also reduced the number of cases identified. Relaxing the criteria to one or more ICD and requiring evidence of eye-exam within 24 hours of the diagnostic code maintained previous gains in PPV from V1 and improved the number of cases. At VUMC, requiring at least three unique days of codes for diabetes for controls (compared to at least one), marginally improved NPV and substantially reduced sample size. On the other hand, in MGB, requiring at least three unique days of diagnostic codes specifically for diabetes was necessary for optimal performance of control selection. EHRs with limited availability of information on prescription (inpatient vs. outpatient) and details regarding prescriber (primary care provider, specialist or otherwise) or chart notes, other than structured ICD codes may benefit with these criteria.

Comparative analyses across versions of algorithms within and between institutions also led to substantive insights. In the VUMC SD, we observe a higher proportion of controls had T2D diagnostic codes in than DR cases, a phenomenon not observed in the VA. This is likely reflective of VUMC as a tertiary care facility, to which severe DR cases are referred for management. We note requiring at least three diabetes diagnostic codes for controls vs. one, further changes characteristics of controls, as it prioritizes individuals who have longer duration of diabetes, and are more likely to have other complications related to diabetes than controls with one ICD code. The VA’s role as a primary care facility for veterans is further evidenced in our selection of patients where both cases and controls are just as likely to have T2D diagnosis codes and just as likely to have had eye exams in the past five years. Duration of diabetes is a particularly difficult variable to quantify through EHR alone in the US as it is only as accurate as the first recorded ICD code/diabetes prescription in each EHR system. This was evident in both VUMC and MVP where for several individuals the first mention of T2D diagnosis came after mention of a DR code, suggesting individuals were either only identified as diabetics after they developed complications or that their diagnosis of diabetes from external institutions was not captured. To ensure controls with diabetes did not have DR but had the opportunity of being diagnosed with DR, we imposed requirements for controls including an ophthalmology/eye clinic visit and capturing age at last visit, whereas ages of cases were recorded at first mention of DR diagnosis. Consequently, we observe controls on average are older than cases across all three EHR systems. This strategy greatly reduced misclassification of outcome, which is highly desirable in the context of GWAS studies.

Overall, by developing and evaluating algorithms to identify diabetic individuals with and without DR, we show the potential of leveraging existing EHR systems to advance our understanding of the etiology of DR. We demonstrate these EHR-based algorithms to be portable and accurate across separate EHR systems and this represents a vast improvement over existing high-throughput method such as PheWAS. These algorithms provide advanced starting points for researchers looking to implement DR algorithms in EHR systems and rather than blind-automated application, we encourage researchers to investigate unique features in each EHR system to implement choices most appropriate to that system. As EHR systems increase in size, as well as the usage for research, a clear advantage to this approach is scalability as it offers an alternative to the manual review of large numbers of retinal images for research purposes while retaining validity.

## Data Availability

All summary-level data produced in the present work are contained in the manuscript or provided as Supplementary documents. Individual level data from electronic health records are not provided due to data-privacy concerns.

## ABBREVIATIONS

DR: Diabetic Retinopathy
PDR: Proliferative Diabetic Retinopathy
EHR: Electronic Health Record
DM: Diabetes Mellitus
MVP: Million Veteran Program
PPV: Positive Predictive Value
NPV: Negative Predictive Value

## ACKNOWLEDGEMENTS

This research is based on data from the Million Veteran Program, Office of Research and Development, and Veterans Health Administration. This publication does not represent the views of the Department of Veterans Affairs or the United States Government. Mass General Brigham Biobank for providing samples, genomic data, and health information data. We do not report any conflicts of interest. A.G. and T.L.E are the guarantors of this work and, as such, had full access to all the data in the study and take responsibility for the integrity of the data and the accuracy of the data analysis.

