## Supplemental Table 1 for "Development of Portable Electronic Health Record Based Algorithms to Identify Individuals with Diabetic Retinopathy"

**Supplemental Table 1a Algorithm Design, Construct, Criteria, and EHR-specific versions for cases.**

|  | **Case Algorithm** |  |  |
| --- | --- | --- | --- |
| **Algorithm Design** | Find individuals with verifiable evidence of diabetic retinopathy (DR) | | |
| **Algorithm versions** | **V0** | **V1** | **V2** |
| **Algorithm Construct: Defining Criteria** | 1 or more unique days of diagnostic code for DR | 2 or more unique dates of diagnostic code for DR | (2 or more unique dates of diagnostic code for DR) OR (ophthalmic exam AND 1 or more diagnostic code for DR within 24 hours of the ophthalmic exam) |
| **Operationalizing Criteria: EHR-specific implementation (VUMC)** | 1 or more unique days of ICD9 or ICD10 codes for DR of any severity (labeled as the following in supplementary tables: nsdr_icd, mnpdr_icd, snpdr_icd, pdr_icd) | 2 or more unique days of ICD9 or ICD10 codes for DR of any severity [labeled as the following in supplementary tables: nsdr_icd, mnpdr_icd, snpdr_icd, pdr_icd] | (2 or more unique dates of ICD9 or ICD10 codes for DR of any severity [nsdr_icd, mnpdr_icd, snpdr_icd, pdr_icd]) OR (ophthalmic exam CPT codes [opt_exam] AND 1 or more ICD9 or ICD10 codes for DR of any severity within 24 hours of the ophthalmic exam) |
| **Operationalizing Criteria: EHR-specific implementation (VANEOHS/MVP)** | 1 or more unique days of ICD9 or ICD10 codes for DR of any severity (nsdr_icd, mnpdr_icd, snpdr_icd, pdr_icd) OR 1 or more unique days of health factor codes for DR [any_dr_hf, nsdr_hf, mnpdr_hf, snpdr_hf, pdr_hf] | Any combination of 2 or more unique days of ICD9 or ICD10 codes for DR of any severity (nsdr_icd, mnpdr_icd, snpdr_icd, pdr_icd) OR health factor codes for DR [any_dr_hf, nsdr_hf, mnpdr_hf, snpdr_hf, pdr_hf] | (Any combination of 2 or more unique days of ICD9 or ICD10 codes for DR of any severity (nsdr_icd, mnpdr_icd, snpdr_icd, pdr_icd) OR health factor codes [any_dr_hf, nsdr_hf, mnpdr_hf, snpdr_hf, pdr_hf]) OR ((ophthalmic exam CPT codes specific to fundus photography [opt_exam] OR evidence of teleretinal visit through health factors [tri_evidence] OR general CPT cdoe with a 408 stop code to confirm visit to eye clinic [opt_code_use_with_stop_code]) AND (1 or more ICD9 or ICD10 codes for DR of any severity OR health factor codes for DR within 24 hours of the ophthalmic exam) |
| **Operationalizing Criteria-concepts: EHR-specific implementation (MGB)** | 1 or more unique days of ICD9 or ICD10 codes for DR of any severity (nsdr_icd, mnpdr_icd, snpdr_icd, pdr_icd) OR 1 or more unique days of EPIC Provider codes for DR specific to MGB | Any combination of 2 or more unique days of ICD9 or ICD10 codes for DR of any severity (nsdr_icd, mnpdr_icd, snpdr_icd, pdr_icd) OR EPIC Provider codes for DR specific to MGB | (Any combination of 2 or more unique days of ICD9 or ICD10 codes for DR of any severity (nsdr_icd, mnpdr_icd, snpdr_icd, pdr_icd) OR EPIC provider codes for DR specific to MGB) OR ((ophthalmic exam CPT codes [opt_exam]) AND (1 or more ICD9 or ICD10 codes or EPIC Provider codes specific for DR within 24 hours of the ophthalmic exam) |

**Supplemental Table 1b. Algorithm Design, Construct, Criteria, and EHR-specific versions for controls.**

|  | **Control Algorithm** |  |
| --- | --- | --- |
| **Algorithm Design** | Find individuals with verifiable evidence of diabetes mellitus in individuals and lack of evidence of diabetic retinopathy (DR) | |
| **Algorithm versions** | **V0** | **V1** |
| **Algorithm Construct: Defining Criteria** | Affirmative evidence of diabetes (through Diagnostic codes, medications or combination) AND no evidence for DR AND excludes unspecified/secondary diabetes AND at least one eye exam | 3 or more unique dates of diagnostic code for diabetes mellitus AND no evidence of DR and at least one eye exam |
| **Operationalizing Criteria: EHR-specific implementation (VUMC)** | (1 or more unique days of ICD9 or ICD10 codes for T1D [t1d_inclusions] OR T2D [dbm_inclusions]) AND no DR codes of any severity [nsdr_icd, mnpdr_icd, snpdr_icd, pdr_icd] AND no codes in unspecified/secondary diabetes list [dbm_exclusions] AND at least one ophthalmic exam code [opt_exam] after diabetes diagnosis date | (3 or more unique days of ICD9 or ICD10 codes for T1D [t1d_inclusions] OR T2D [dbm_inclusions]) AND no DR codes of any severity [nsdr_icd, mnpdr_icd, snpdr_icd, pdr_icd] AND no codes in unspecified/secondary diabetes list [dbm_exclusions] AND at least one ophthalmic exam code [opt_exam] after diabetes diagnosis date |
| **Operationalizing Criteria: EHR-specific implementation (VANEOHS/MVP)** | ((1 or more diagnostic code for diabetes mellitus through face-to-face outpatient primary care physician visit OR 2 or more unique days of diagnostic code for diabetes mellitus at outpatient visits of any kind) AND at least one outpatient fill of diabetes-indicated medications (Supplementary table 3)) AND no DR ICD9 or 10 diagnostic codes AND no health factor codes for DR AND (Evidence of eye exam: (ophthalmic exam CPT codes specific to fundus photography [opt_exam] OR general CPT code with a 408 stop code to confirm visit to eye clinic [opt_code_use_with_stop_code] OR evidence of teleretinal visit through health factors with affirmation for no DR [no_dr_evidence]*)) | Not implemented |
| **Operationalizng Criteria-concepts: EHR-specific implementation (MGB)** | (1 or more unique days of ICD9 or ICD10 codes for T1D [t1d_inclusions] OR T2D [dbm_inclusions]) AND (no DR codes of any severity [nsdr_icd, mnpdr_icd, snpdr_icd, pdr_icd] AND no EPIC provider codes specific to DR in MGB) AND no codes in unspecified/secondary diabetes list [dbm_exclusions] AND at least one ophthalmic exam code [opt_exam] after diabetes diagnosis date | (3 or more unique days of ICD9 or ICD10 codes for T1D [t1d_inclusions] OR T2D [dbm_inclusions]) AND (no DR codes of any severity [nsdr_icd, mnpdr_icd, snpdr_icd, pdr_icd] AND no EPIC provider codes specific to DR in MGB) AND no codes in unspecified/secondary diabetes list [dbm_exclusions] AND at least one ophthalmic exam code [opt_exam] after diabetes diagnosis date |

*For details on variable names, check supplemental tables 2 and 3.
