## Supplemental Table 2 for "Development of Portable Electronic Health Record Based Algorithms to Identify Individuals with Diabetic Retinopathy"

Supplemental Table 2a. List of ICD & CPT codes included in the DR algorithm at Vanderbilt University Medical Center

| Trait | Domain | Code | Concept Name |
| --- | --- | --- | --- |
| snpdr_icd | ICD9CM | 362.06 | Severe nonproliferative diabetic retinopathy |
| snpdr_icd | ICD10CM | E10.3411 | Type 1 diabetes mellitus with severe nonproliferative diabetic retinopathy with macular edema, right eye |
| snpdr_icd | ICD10CM | E10.3412 | Type 1 diabetes mellitus with severe nonproliferative diabetic retinopathy with macular edema, left eye |
| snpdr_icd | ICD10CM | E10.3413 | Type 1 diabetes mellitus with severe nonproliferative diabetic retinopathy with macular edema, bilateral |
| snpdr_icd | ICD10CM | E10.3419 | Type 1 diabetes mellitus with severe nonproliferative diabetic retinopathy with macular edema, unspecified eye |
| snpdr_icd | ICD10CM | E10.3491 | Type 1 diabetes mellitus with severe nonproliferative diabetic retinopathy without macular edema, right eye |
| snpdr_icd | ICD10CM | E10.3492 | Type 1 diabetes mellitus with severe nonproliferative diabetic retinopathy without macular edema, left eye |
| snpdr_icd | ICD10CM | E10.3493 | Type 1 diabetes mellitus with severe nonproliferative diabetic retinopathy without macular edema, bilateral |
| snpdr_icd | ICD10CM | E10.3499 | Type 1 diabetes mellitus with severe nonproliferative diabetic retinopathy without macular edema, unspecified eye |
| snpdr_icd | ICD10CM | E11.3411 | Type 2 diabetes mellitus with severe nonproliferative diabetic retinopathy with macular edema, right eye |
| snpdr_icd | ICD10CM | E11.3412 | Type 2 diabetes mellitus with severe nonproliferative diabetic retinopathy with macular edema, left eye |
| snpdr_icd | ICD10CM | E11.3413 | Type 2 diabetes mellitus with severe nonproliferative diabetic retinopathy with macular edema, bilateral |
| snpdr_icd | ICD10CM | E11.3419 | Type 2 diabetes mellitus with severe nonproliferative diabetic retinopathy with macular edema, unspecified eye |
| snpdr_icd | ICD10CM | E11.3491 | Type 2 diabetes mellitus with severe nonproliferative diabetic retinopathy without macular edema, right eye |
| snpdr_icd | ICD10CM | E11.3492 | Type 2 diabetes mellitus with severe nonproliferative diabetic retinopathy without macular edema, left eye |
| snpdr_icd | ICD10CM | E11.3493 | Type 2 diabetes mellitus with severe nonproliferative diabetic retinopathy without macular edema, bilateral |
| snpdr_icd | ICD10CM | E11.3499 | Type 2 diabetes mellitus with severe nonproliferative diabetic retinopathy without macular edema, unspecified eye |
| pdr_icd | ICD9CM | 362.02 | Proliferative diabetic retinopathy |
| pdr_icd | ICD10CM | E10.3511 | Type 1 diabetes mellitus with proliferative diabetic retinopathy with macular edema, right eye |
| pdr_icd | ICD10CM | E10.3512 | Type 1 diabetes mellitus with proliferative diabetic retinopathy with macular edema, left eye |
| pdr_icd | ICD10CM | E10.3513 | Type 1 diabetes mellitus with proliferative diabetic retinopathy with macular edema, bilateral |
| pdr_icd | ICD10CM | E10.3519 | Type 1 diabetes mellitus with proliferative diabetic retinopathy with macular edema, unspecified eye |
| pdr_icd | ICD10CM | E10.3521 | Type 1 diabetes mellitus with proliferative diabetic retinopathy with traction retinal detachment involving the macula, right eye |
| pdr_icd | ICD10CM | E10.3522 | Type 1 diabetes mellitus with proliferative diabetic retinopathy with traction retinal detachment involving the macula, left eye |
| pdr_icd | ICD10CM | E10.3523 | Type 1 diabetes mellitus with proliferative diabetic retinopathy with traction retinal detachment involving the macula, bilateral |
| pdr_icd | ICD10CM | E10.3529 | Type 1 diabetes mellitus with proliferative diabetic retinopathy with traction retinal detachment involving the macula, unspecified eye |
| pdr_icd | ICD10CM | E10.3531 | Type 1 diabetes mellitus with proliferative diabetic retinopathy with traction retinal detachment not involving the macula, right eye |
| pdr_icd | ICD10CM | E10.3532 | Type 1 diabetes mellitus with proliferative diabetic retinopathy with traction retinal detachment not involving the macula, left eye |
| pdr_icd | ICD10CM | E10.3533 | Type 1 diabetes mellitus with proliferative diabetic retinopathy with traction retinal detachment not involving the macula, bilateral |
| pdr_icd | ICD10CM | E10.3539 | Type 1 diabetes mellitus with proliferative diabetic retinopathy with traction retinal detachment not involving the macula, unspecified eye |
| pdr_icd | ICD10CM | E10.3541 | Type 1 diabetes mellitus with proliferative diabetic retinopathy with combined traction retinal detachment and rhegmatogenous retinal detachment, right eye |
| pdr_icd | ICD10CM | E10.3542 | Type 1 diabetes mellitus with proliferative diabetic retinopathy with combined traction retinal detachment and rhegmatogenous retinal detachment, left eye |
| pdr_icd | ICD10CM | E10.3543 | Type 1 diabetes mellitus with proliferative diabetic retinopathy with combined traction retinal detachment and rhegmatogenous retinal detachment, bilateral |
| pdr_icd | ICD10CM | E10.3549 | Type 1 diabetes mellitus with proliferative diabetic retinopathy with combined traction retinal detachment and rhegmatogenous retinal detachment, unspecified eye |
| pdr_icd | ICD10CM | E10.3551 | Type 1 diabetes mellitus with stable proliferative diabetic retinopathy, right eye |
| pdr_icd | ICD10CM | E10.3552 | Type 1 diabetes mellitus with stable proliferative diabetic retinopathy, left eye |
| pdr_icd | ICD10CM | E10.3553 | Type 1 diabetes mellitus with stable proliferative diabetic retinopathy, bilateral |
| pdr_icd | ICD10CM | E10.3559 | Type 1 diabetes mellitus with stable proliferative diabetic retinopathy, unspecified eye |
| pdr_icd | ICD10CM | E10.3591 | Type 1 diabetes mellitus with proliferative diabetic retinopathy without macular edema, right eye |
| pdr_icd | ICD10CM | E10.3592 | Type 1 diabetes mellitus with proliferative diabetic retinopathy without macular edema, left eye |
| pdr_icd | ICD10CM | E10.3593 | Type 1 diabetes mellitus with proliferative diabetic retinopathy without macular edema, bilateral |
| pdr_icd | ICD10CM | E10.3599 | Type 1 diabetes mellitus with proliferative diabetic retinopathy without macular edema, unspecified eye |
| pdr_icd | ICD10CM | E11.3511 | Type 2 diabetes mellitus with proliferative diabetic retinopathy with macular edema, right eye |
| pdr_icd | ICD10CM | E11.3512 | Type 2 diabetes mellitus with proliferative diabetic retinopathy with macular edema, left eye |
| pdr_icd | ICD10CM | E11.3513 | Type 2 diabetes mellitus with proliferative diabetic retinopathy with macular edema, bilateral |
| pdr_icd | ICD10CM | E11.3519 | Type 2 diabetes mellitus with proliferative diabetic retinopathy with macular edema, unspecified eye |
| pdr_icd | ICD10CM | E11.3521 | Type 2 diabetes mellitus with proliferative diabetic retinopathy with traction retinal detachment involving the macula, right eye |
| pdr_icd | ICD10CM | E11.3522 | Type 2 diabetes mellitus with proliferative diabetic retinopathy with traction retinal detachment involving the macula, left eye |
| pdr_icd | ICD10CM | E11.3523 | Type 2 diabetes mellitus with proliferative diabetic retinopathy with traction retinal detachment involving the macula, bilateral |
| pdr_icd | ICD10CM | E11.3529 | Type 2 diabetes mellitus with proliferative diabetic retinopathy with traction retinal detachment involving the macula, unspecified eye |
| pdr_icd | ICD10CM | E11.3531 | Type 2 diabetes mellitus with proliferative diabetic retinopathy with traction retinal detachment not involving the macula, right eye |
| pdr_icd | ICD10CM | E11.3532 | Type 2 diabetes mellitus with proliferative diabetic retinopathy with traction retinal detachment not involving the macula, left eye |
| pdr_icd | ICD10CM | E11.3533 | Type 2 diabetes mellitus with proliferative diabetic retinopathy with traction retinal detachment not involving the macula, bilateral |
| pdr_icd | ICD10CM | E11.3539 | Type 2 diabetes mellitus with proliferative diabetic retinopathy with traction retinal detachment not involving the macula, unspecified eye |
| pdr_icd | ICD10CM | E11.3541 | Type 2 diabetes mellitus with proliferative diabetic retinopathy with combined traction retinal detachment and rhegmatogenous retinal detachment, right eye |
| pdr_icd | ICD10CM | E11.3542 | Type 2 diabetes mellitus with proliferative diabetic retinopathy with combined traction retinal detachment and rhegmatogenous retinal detachment, left eye |
| pdr_icd | ICD10CM | E11.3543 | Type 2 diabetes mellitus with proliferative diabetic retinopathy with combined traction retinal detachment and rhegmatogenous retinal detachment, bilateral |
| pdr_icd | ICD10CM | E11.3549 | Type 2 diabetes mellitus with proliferative diabetic retinopathy with combined traction retinal detachment and rhegmatogenous retinal detachment, unspecified eye |
| pdr_icd | ICD10CM | E11.3551 | Type 2 diabetes mellitus with stable proliferative diabetic retinopathy, right eye |
| pdr_icd | ICD10CM | E11.3552 | Type 2 diabetes mellitus with stable proliferative diabetic retinopathy, left eye |
| pdr_icd | ICD10CM | E11.3553 | Type 2 diabetes mellitus with stable proliferative diabetic retinopathy, bilateral |
| pdr_icd | ICD10CM | E11.3559 | Type 2 diabetes mellitus with stable proliferative diabetic retinopathy, unspecified eye |
| pdr_icd | ICD10CM | E11.3591 | Type 2 diabetes mellitus with proliferative diabetic retinopathy without macular edema, right eye |
| pdr_icd | ICD10CM | E11.3592 | Type 2 diabetes mellitus with proliferative diabetic retinopathy without macular edema, left eye |
| pdr_icd | ICD10CM | E11.3593 | Type 2 diabetes mellitus with proliferative diabetic retinopathy without macular edema, bilateral |
| pdr_icd | ICD10CM | E11.3599 | Type 2 diabetes mellitus with proliferative diabetic retinopathy without macular edema, unspecified eye |
| opt_exam | CPT4 | 92004 | Ophthalmological services: medical examination and evaluation with initiation of diagnostic and treatment program; comprehensive, new patient, 1 or more visits |
| opt_exam | CPT4 | 92014 | Ophthalmological services: medical examination and evaluation, with initiation or continuation of diagnostic and treatment program; comprehensive, established patient, 1 or more visits |
| opt_exam | CPT4 | 92250 | Fundus photography with interpretation and report |
| opt_exam | CPT4 | 92227 | Ophthalmoscopy Procedures |
| opt_exam | CPT4 | 92228 | Ophthalmoscopy Procedures |
| opt_exam | CPT4 | 92002 | Ophthalmological services: medical examination and evaluation with initiation of diagnostic and treatment program. |
| opt_exam | CPT4 | 92012 | Ophthalmological services: medical examination and evaluation, with initiation or continuation of diagnostic and treatment program |
| nsdr_icd | ICD9CM | 362.01 | Background diabetic retinopathy |
| nsdr_icd | ICD9CM | 362.03 | Nonproliferative diabetic retinopathy NOS |
| nsdr_icd | ICD10CM | E10.311 | Type 1 diabetes mellitus with unspecified diabetic retinopathy with macular edema |
| nsdr_icd | ICD10CM | E10.319 | Type 1 diabetes mellitus with unspecified diabetic retinopathy without macular edema |
| nsdr_icd | ICD10CM | E10.321 | Type 1 diabetes mellitus with mild nonproliferative diabetic retinopathy with macular edema |
| nsdr_icd | ICD10CM | E10.329 | Type 1 diabetes mellitus with mild nonproliferative diabetic retinopathy without macular edema |
| nsdr_icd | ICD10CM | E11.311 | Type 2 diabetes mellitus with unspecified diabetic retinopathy with macular edema |
| nsdr_icd | ICD10CM | E11.319 | Type 2 diabetes mellitus with unspecified diabetic retinopathy without macular edema |
| nsdr_icd | ICD10CM | E11.321 | Type 2 diabetes mellitus with mild nonproliferative diabetic retinopathy with macular edema |
| nsdr_icd | ICD10CM | E11.329 | Type 2 diabetes mellitus with mild nonproliferative diabetic retinopathy without macular edema |
| mnpdr_icd | ICD9CM | 362.04 | Mild nonproliferative diabetic retinopathy |
| mnpdr_icd | ICD9CM | 362.05 | Moderate nonproliferative diabetic retinopathy |
| mnpdr_icd | ICD10CM | E10.3211 | Type 1 diabetes mellitus with mild nonproliferative diabetic retinopathy with macular edema, right eye |
| mnpdr_icd | ICD10CM | E10.3212 | Type 1 diabetes mellitus with mild nonproliferative diabetic retinopathy with macular edema, left eye |
| mnpdr_icd | ICD10CM | E10.3213 | Type 1 diabetes mellitus with mild nonproliferative diabetic retinopathy with macular edema, bilateral |
| mnpdr_icd | ICD10CM | E10.3219 | Type 1 diabetes mellitus with mild nonproliferative diabetic retinopathy with macular edema, unspecified eye |
| mnpdr_icd | ICD10CM | E10.3291 | Type 1 diabetes mellitus with mild nonproliferative diabetic retinopathy without macular edema, right eye |
| mnpdr_icd | ICD10CM | E10.3292 | Type 1 diabetes mellitus with mild nonproliferative diabetic retinopathy without macular edema, left eye |
| mnpdr_icd | ICD10CM | E10.3293 | Type 1 diabetes mellitus with mild nonproliferative diabetic retinopathy without macular edema, bilateral |
| mnpdr_icd | ICD10CM | E10.3299 | Type 1 diabetes mellitus with mild nonproliferative diabetic retinopathy without macular edema, unspecified eye |
| mnpdr_icd | ICD10CM | E10.3311 | Type 1 diabetes mellitus with moderate nonproliferative diabetic retinopathy with macular edema, right eye |
| mnpdr_icd | ICD10CM | E10.3312 | Type 1 diabetes mellitus with moderate nonproliferative diabetic retinopathy with macular edema, left eye |
| mnpdr_icd | ICD10CM | E10.3313 | Type 1 diabetes mellitus with moderate nonproliferative diabetic retinopathy with macular edema, bilateral |
| mnpdr_icd | ICD10CM | E10.3319 | Type 1 diabetes mellitus with moderate nonproliferative diabetic retinopathy with macular edema, unspecified eye |
| mnpdr_icd | ICD10CM | E10.3391 | Type 1 diabetes mellitus with moderate nonproliferative diabetic retinopathy without macular edema, right eye |
| mnpdr_icd | ICD10CM | E10.3392 | Type 1 diabetes mellitus with moderate nonproliferative diabetic retinopathy without macular edema, left eye |
| mnpdr_icd | ICD10CM | E10.3393 | Type 1 diabetes mellitus with moderate nonproliferative diabetic retinopathy without macular edema, bilateral |
| mnpdr_icd | ICD10CM | E10.3399 | Type 1 diabetes mellitus with moderate nonproliferative diabetic retinopathy without macular edema, unspecified eye |
| mnpdr_icd | ICD10CM | E11.3211 | Type 2 diabetes mellitus with mild nonproliferative diabetic retinopathy with macular edema, right eye |
| mnpdr_icd | ICD10CM | E11.3212 | Type 2 diabetes mellitus with mild nonproliferative diabetic retinopathy with macular edema, left eye |
| mnpdr_icd | ICD10CM | E11.3213 | Type 2 diabetes mellitus with mild nonproliferative diabetic retinopathy with macular edema, bilateral |
| mnpdr_icd | ICD10CM | E11.3219 | Type 2 diabetes mellitus with mild nonproliferative diabetic retinopathy with macular edema, unspecified eye |
| mnpdr_icd | ICD10CM | E11.3291 | Type 2 diabetes mellitus with mild nonproliferative diabetic retinopathy without macular edema, right eye |
| mnpdr_icd | ICD10CM | E11.3292 | Type 2 diabetes mellitus with mild nonproliferative diabetic retinopathy without macular edema, left eye |
| mnpdr_icd | ICD10CM | E11.3293 | Type 2 diabetes mellitus with mild nonproliferative diabetic retinopathy without macular edema, bilateral |
| mnpdr_icd | ICD10CM | E11.3299 | Type 2 diabetes mellitus with mild nonproliferative diabetic retinopathy without macular edema, unspecified eye |
| mnpdr_icd | ICD10CM | E11.3311 | Type 2 diabetes mellitus with moderate nonproliferative diabetic retinopathy with macular edema, right eye |
| mnpdr_icd | ICD10CM | E11.3312 | Type 2 diabetes mellitus with moderate nonproliferative diabetic retinopathy with macular edema, left eye |
| mnpdr_icd | ICD10CM | E11.3313 | Type 2 diabetes mellitus with moderate nonproliferative diabetic retinopathy with macular edema, bilateral |
| mnpdr_icd | ICD10CM | E11.3319 | Type 2 diabetes mellitus with moderate nonproliferative diabetic retinopathy with macular edema, unspecified eye |
| mnpdr_icd | ICD10CM | E11.3391 | Type 2 diabetes mellitus with moderate nonproliferative diabetic retinopathy without macular edema, right eye |
| mnpdr_icd | ICD10CM | E11.3392 | Type 2 diabetes mellitus with moderate nonproliferative diabetic retinopathy without macular edema, left eye |
| mnpdr_icd | ICD10CM | E11.3393 | Type 2 diabetes mellitus with moderate nonproliferative diabetic retinopathy without macular edema, bilateral |
| mnpdr_icd | ICD10CM | E11.3399 | Type 2 diabetes mellitus with moderate nonproliferative diabetic retinopathy without macular edema, unspecified eye |
| Codes additionally relevant for the VA | |  |  |
| opt_code_use_with_stop_code | CPT4 | 99202 | Use this general CPT code along with stop code 408nto count as an opthalmic visit |
| opt_code_use_with_stop_code | CPT4 | 99203 | Use this general CPT code along with stop code 408nto count as an opthalmic visit |
| opt_code_use_with_stop_code | CPT4 | 99204 | Use this general CPT code along with stop code 408nto count as an opthalmic visit |
| opt_code_use_with_stop_code | CPT4 | 99205 | Use this general CPT code along with stop code 408nto count as an opthalmic visit |
| opt_code_use_with_stop_code | CPT4 | 99212 | Use this general CPT code along with stop code 408nto count as an opthalmic visit |
| opt_code_use_with_stop_code | CPT4 | 99213 | Use this general CPT code along with stop code 408nto count as an opthalmic visit |
| opt_code_use_with_stop_code | CPT4 | 99214 | Use this general CPT code along with stop code 408nto count as an opthalmic visit |
| opt_code_use_with_stop_code | CPT4 | 99215 | Use this general CPT code along with stop code 408nto count as an opthalmic visit |
| opt_stop_code |  | 408 | Stop code to determine opthalmic visit in the VA |
| tri_evidence | HF_imaging_TRI_TRR | LTRI DM FINDING DR OTHER LT | Documentation of a teleretinal visit through health factors - DR present |
| tri_evidence | HF_imaging_TRI_TRR | LTRI DM FINDING DR OTHER RT | Documentation of a teleretinal visit through health factors - DR present |
| tri_evidence | HF_imaging_TRI_TRR | LTRI DM FINDING DR PREV RET LASER TX LT | Documentation of a teleretinal visit through health factors - DR present |
| tri_evidence | HF_imaging_TRI_TRR | LTRI DM FINDING DR PREV RET LASER TX RT | Documentation of a teleretinal visit through health factors - DR present |
| tri_evidence | HF_imaging_TRI_TRR | LTRI DM HX LASER THERAPY DIAB RETINOPATH | Documentation of a teleretinal visit through health factors - DR present |
| tri_evidence | HF_imaging_TRI_TRR | LTRI DM HX LASER TX DIAB RETINOPATHY | Documentation of a teleretinal visit through health factors - DR present |
| tri_evidence | HF_imaging_TRI_TRR | LTRI DM RETINOPATHY EXAM ABNORMAL | Documentation of a teleretinal visit through health factors - DR present |
| tri_evidence | HF_imaging_TRI_TRR | TRI DM FINDING DR OTHER LT | Documentation of a teleretinal visit through health factors - DR present |
| tri_evidence | HF_imaging_TRI_TRR | TRI DM FINDING DR OTHER RT | Documentation of a teleretinal visit through health factors - DR present |
| tri_evidence | HF_imaging_TRI_TRR | TRI DM FINDING DR PREV RET LASER TX LT | Documentation of a teleretinal visit through health factors - DR present |
| tri_evidence | HF_imaging_TRI_TRR | TRI DM FINDING DR PREV RET LASER TX RT | Documentation of a teleretinal visit through health factors - DR present |
| tri_evidence | HF_imaging_TRI_TRR | TRI DM HX LASER THERAPY DIAB RETINOPATHY | Documentation of a teleretinal visit through health factors - DR present |
| tri_evidence | HF_imaging_TRI_TRR | TRI DM RETINOPATHY EXAM ABNORMAL | Documentation of a teleretinal visit through health factors - DR present |
| tri_evidence | HF_imaging_TRI_TRR | TRI DM FINDING DR MACULAR EDEMA LT | Documentation of a teleretinal visit through health factors - DR present |
| tri_evidence | HF_imaging_TRI_TRR | TRI DM FINDING DR MACULAR EDEMA RT | Documentation of a teleretinal visit through health factors - DR present |
| tri_evidence | HF_imaging_TRI_TRR | DIABETES - RETINOPATHY | Documentation of a teleretinal visit through health factors - DR present |
| tri_evidence | HF_imaging_TRI_TRR | DIABETIC EYE RETINOPATHY FOUND | Documentation of a teleretinal visit through health factors - DR present |
| tri_evidence | HF_imaging_TRI_TRR | Diabetic Retina-Retinopathy | Documentation of a teleretinal visit through health factors - DR present |
| tri_evidence | HF_imaging_TRI_TRR | DIABETIC RETINOPATHY | Documentation of a teleretinal visit through health factors - DR present |
| tri_evidence | HF_imaging_TRI_TRR | DIABETIC RETINOPATHY PRESENT | Documentation of a teleretinal visit through health factors - DR present |
| tri_evidence | HF_imaging_TRI_TRR | HX DIABETIC RETINOPATHY | Documentation of a teleretinal visit through health factors - DR present |
| tri_evidence | HF_imaging_TRI_TRR | LTRI DM FINDING DR MACULAR EDEMA LT | Documentation of a teleretinal visit through health factors - DR present |
| tri_evidence | HF_imaging_TRI_TRR | LTRI DM FINDING DR MACULAR EDEMA RT | Documentation of a teleretinal visit through health factors - DR present |
| tri_evidence | HF_imaging_TRI_TRR | LTRI DM FINDING DR NO MAC EDEMA APP LT | Documentation of a teleretinal visit through health factors - DR present |
| tri_evidence | HF_imaging_TRI_TRR | LTRI DM FINDING DR NO MAC EDEMA APP RT | Documentation of a teleretinal visit through health factors - DR present |
| tri_evidence | HF_imaging_TRI_TRR | LTRI DM REFER RETINOPATHY | Documentation of a teleretinal visit through health factors - DR present |
| tri_evidence | HF_imaging_TRI_TRR | TRI DM REFER RETINOPATHY | Documentation of a teleretinal visit through health factors - DR present |
| tri_evidence | HF_imaging_TRI_TRR | VA-TRR DM RETINOPATHY EXAM ABNORMAL | Documentation of a teleretinal visit through health factors - DR present |
| tri_evidence | HF_imaging_TRI_TRR | VA-TRR DM2 FIND DR PREV RET LASER TX LT | Documentation of a teleretinal visit through health factors - DR present |
| tri_evidence | HF_imaging_TRI_TRR | VA-TRR DM2 FIND DR PREV RET LASER TX RT | Documentation of a teleretinal visit through health factors - DR present |
| tri_evidence | HF_imaging_TRI_TRR | VA-TRR DM2 FINDING DR OTHER LT | Documentation of a teleretinal visit through health factors - DR present |
| tri_evidence | HF_imaging_TRI_TRR | VA-TRR DM2 FINDING DR OTHER RT | Documentation of a teleretinal visit through health factors - DR present |
| tri_evidence | HF_imaging_TRI_TRR | LTRI DM FINDING DR MACULAR EDEMA LT | Documentation of a teleretinal visit through health factors - DR present |
| tri_evidence | HF_imaging_TRI_TRR | LTRI DM FINDING DR MACULAR EDEMA RT | Documentation of a teleretinal visit through health factors - DR present |
| tri_evidence | HF_imaging_TRI_TRR | LTRI DM FINDING DR MACULAR EDEMAL LT | Documentation of a teleretinal visit through health factors - DR present |
| tri_evidence | HF_imaging_TRI_TRR | TRI DM FINDING DR MACULAR EDEMA LT | Documentation of a teleretinal visit through health factors - DR present |
| tri_evidence | HF_imaging_TRI_TRR | TRI DM FINDING DR MACULAR EDEMA RT | Documentation of a teleretinal visit through health factors - DR present |
| tri_evidence | HF_imaging_TRI_TRR | DM MILD NONPROLIFERATIVE RETINOPATHY LT | Documentation of a teleretinal visit through health factors - DR present |
| tri_evidence | HF_imaging_TRI_TRR | DM MILD NONPROLIFERATIVE RETINOPATHY RT | Documentation of a teleretinal visit through health factors - DR present |
| tri_evidence | HF_imaging_TRI_TRR | DM MOD NONPROLIFERATIVE RETINOPATHY LT | Documentation of a teleretinal visit through health factors - DR present |
| tri_evidence | HF_imaging_TRI_TRR | DM MOD NONPROLIFERATIVE RETINOPATHY RT | Documentation of a teleretinal visit through health factors - DR present |
| tri_evidence | HF_imaging_TRI_TRR | LTRI DM DR MILD NONPROLIF LT | Documentation of a teleretinal visit through health factors - DR present |
| tri_evidence | HF_imaging_TRI_TRR | LTRI DM FINDING DR MILD NONPROLIF LT | Documentation of a teleretinal visit through health factors - DR present |
| tri_evidence | HF_imaging_TRI_TRR | LTRI DM FINDING DR MILD NONPROLIF RT | Documentation of a teleretinal visit through health factors - DR present |
| tri_evidence | HF_imaging_TRI_TRR | LTRI DM FINDING DR MOD NONPROLIF LT | Documentation of a teleretinal visit through health factors - DR present |
| tri_evidence | HF_imaging_TRI_TRR | LTRI DM FINDING DR MOD NONPROLIF RT | Documentation of a teleretinal visit through health factors - DR present |
| tri_evidence | HF_imaging_TRI_TRR | LTRI DM FINDING MILD NONPROLIF RT | Documentation of a teleretinal visit through health factors - DR present |
| tri_evidence | HF_imaging_TRI_TRR | LTRI DM MOD NONPROLIF LT | Documentation of a teleretinal visit through health factors - DR present |
| tri_evidence | HF_imaging_TRI_TRR | TRI DM FINDING DR MILD NONPROLIF LT | Documentation of a teleretinal visit through health factors - DR present |
| tri_evidence | HF_imaging_TRI_TRR | TRI DM FINDING DR MILD NONPROLIF RT | Documentation of a teleretinal visit through health factors - DR present |
| tri_evidence | HF_imaging_TRI_TRR | TRI DM FINDING DR MOD NONPROLIF LT | Documentation of a teleretinal visit through health factors - DR present |
| tri_evidence | HF_imaging_TRI_TRR | TRI DM FINDING DR MOD NONPROLIF RT | Documentation of a teleretinal visit through health factors - DR present |
| tri_evidence | HF_imaging_TRI_TRR | VA-TRR DM2 FIND DR MLD MAC NO EDEMA LT | Documentation of a teleretinal visit through health factors - DR present |
| tri_evidence | HF_imaging_TRI_TRR | VA-TRR DM2 FIND DR MLD MAC NO EDEMA RT | Documentation of a teleretinal visit through health factors - DR present |
| tri_evidence | HF_imaging_TRI_TRR | VA-TRR DM2 FIND DR MOD MAC NO EDEMA LT | Documentation of a teleretinal visit through health factors - DR present |
| tri_evidence | HF_imaging_TRI_TRR | VA-TRR DM2 FIND DR MOD MAC NO EDEMA RT | Documentation of a teleretinal visit through health factors - DR present |
| tri_evidence | HF_imaging_TRI_TRR | VA-TRR DM2 FINDING DR MLD MAC EDEMA LT | Documentation of a teleretinal visit through health factors - DR present |
| tri_evidence | HF_imaging_TRI_TRR | VA-TRR DM2 FINDING DR MLD MAC EDEMA RT | Documentation of a teleretinal visit through health factors - DR present |
| tri_evidence | HF_imaging_TRI_TRR | VA-TRR DM2 FINDING DR MOD MAC EDEMA LT | Documentation of a teleretinal visit through health factors - DR present |
| tri_evidence | HF_imaging_TRI_TRR | VA-TRR DM2 FINDING DR MOD MAC EDEMA RT | Documentation of a teleretinal visit through health factors - DR present |
| tri_evidence | HF_imaging_TRI_TRR | DIABETIC RETINA-MOD. BKGRD RETINOPATHY | Documentation of a teleretinal visit through health factors - DR present |
| tri_evidence | HF_imaging_TRI_TRR | DM PROLIFERATIVE RETINOPATHY LT | Documentation of a teleretinal visit through health factors - DR present |
| tri_evidence | HF_imaging_TRI_TRR | DM PROLIFERATIVE RETINOPATHY RT | Documentation of a teleretinal visit through health factors - DR present |
| tri_evidence | HF_imaging_TRI_TRR | LTRI DM FINDING DR PROLIFERATIVE LT | Documentation of a teleretinal visit through health factors - DR present |
| tri_evidence | HF_imaging_TRI_TRR | LTRI DM FINDING DR PROLIFERATIVE RT | Documentation of a teleretinal visit through health factors - DR present |
| tri_evidence | HF_imaging_TRI_TRR | TRI DM FINDING DR PROLIFERATIVE LT | Documentation of a teleretinal visit through health factors - DR present |
| tri_evidence | HF_imaging_TRI_TRR | TRI DM FINDING DR PROLIFERATIVE RT | Documentation of a teleretinal visit through health factors - DR present |
| tri_evidence | HF_imaging_TRI_TRR | VA-TRR DM2 FIND DR PDR MAC NO EDEMA LT | Documentation of a teleretinal visit through health factors - DR present |
| tri_evidence | HF_imaging_TRI_TRR | DM SEVERE NONPROLIFERATIVE RETINOPATHY L | Documentation of a teleretinal visit through health factors - DR present |
| tri_evidence | HF_imaging_TRI_TRR | DM SEVERE NONPROLIFERATIVE RETINOPATHY R | Documentation of a teleretinal visit through health factors - DR present |
| tri_evidence | HF_imaging_TRI_TRR | LTRI DM FINDING DR SEVERE NONPROLIF LT | Documentation of a teleretinal visit through health factors - DR present |
| tri_evidence | HF_imaging_TRI_TRR | LTRI DM FINDING DR SEVERE NONPROLIF RT | Documentation of a teleretinal visit through health factors - DR present |
| tri_evidence | HF_imaging_TRI_TRR | TRI DM FINDING DR SEVERE NONPROLIF LT | Documentation of a teleretinal visit through health factors - DR present |
| tri_evidence | HF_imaging_TRI_TRR | TRI DM FINDING DR SEVERE NONPROLIF RT | Documentation of a teleretinal visit through health factors - DR present |
| tri_evidence | HF_imaging_TRI_TRR | VA-TRR DM2 FIND DR SEV MAC NO EDEMA RT | Documentation of a teleretinal visit through health factors - DR present |
| tri_evidence | HF_imaging_TRI_TRR | LTRI DM DR MILD NONPROLIF RT | Documentation of a teleretinal visit through health factors - DR present |
| tri_evidence | HF_imaging_TRI_TRR | LTRI DM MOD NONPROLIF RT | Documentation of a teleretinal visit through health factors - DR present |
| tri_evidence | HF_imaging_TRI_TRR | VA-TRR DM2 FIND DR PDR MAC NO EDEMA RT | Documentation of a teleretinal visit through health factors - DR present |
| tri_evidence | HF_imaging_TRI_TRR | VA-TRR DM2 FIND DR SEV MAC NO EDEMA LT | Documentation of a teleretinal visit through health factors - DR present |
| tri_evidence | HF_imaging_TRI_TRR | VA-TRR DM2 FINDING DR NO RETINOPATHY LT | Documentation of a teleretinal visit through health factors - no DR present |
| tri_evidence | HF_imaging_TRI_TRR | VA-TRR DM2 FINDING DR NO RETINOPATHY RT | Documentation of a teleretinal visit through health factors - no DR present |
| tri_evidence | HF_imaging_TRI_TRR | VA-TRR DM RETINOPATHY EXAM NORMAL | Documentation of a teleretinal visit through health factors - no DR present |
| tri_evidence | HF_imaging_TRI_TRR | VA-TRR DM1 FINDING DR NO RETINOPATHY LT | Documentation of a teleretinal visit through health factors - no DR present |
| tri_evidence | HF_imaging_TRI_TRR | VA-TRR DM1 FINDING DR NO RETINOPATHY RT | Documentation of a teleretinal visit through health factors - no DR present |
| tri_evidence | HF_imaging_TRI_TRR | NO DIABETIC RETINOPATHY | Documentation of a teleretinal visit through health factors - no DR present |
| tri_evidence | HF_imaging_TRI_TRR | NORMAL RETINAL EXAM | Documentation of a teleretinal visit through health factors - no DR present |
| tri_evidence | HF_imaging_TRI_TRR | NO DIABETIC RETINOPATHY (OUTSIDE EXAM) | Documentation of a teleretinal visit through health factors - no DR present |
| tri_evidence | HF_imaging_TRI_TRR | DM NO DIABETIC RETINOPATHY LT | Documentation of a teleretinal visit through health factors - no DR present |
| tri_evidence | HF_imaging_TRI_TRR | DM NO DIABETIC RETINOPATHY RT | Documentation of a teleretinal visit through health factors - no DR present |
| tri_evidence | HF_imaging_TRI_TRR | LTRI DM FINDING DR NO RETINOPATHY RT | Documentation of a teleretinal visit through health factors - no DR present |
| tri_evidence | HF_imaging_TRI_TRR | LTRI DM FINDING DR NO RETINOPATHY LT | Documentation of a teleretinal visit through health factors - no DR present |
| tri_evidence | HF_imaging_TRI_TRR | LTRI DM RETINOPATHY EXAM NORMAL | Documentation of a teleretinal visit through health factors - no DR present |
| tri_evidence | HF_imaging_TRI_TRR | TRI DM FINDING DR NO RETINOPATHY RT | Documentation of a teleretinal visit through health factors - no DR present |
| tri_evidence | HF_imaging_TRI_TRR | TRI DM FINDING DR NO RETINOPATHY LT | Documentation of a teleretinal visit through health factors - no DR present |
| tri_evidence | HF_imaging_TRI_TRR | TRI DM RETINOPATHY EXAM NORMAL | Documentation of a teleretinal visit through health factors - no DR present |
| tri_evidence | HF_imaging_TRI_TRR | DIABETIC EYE NO RETINOPATHY FOUND | Documentation of a teleretinal visit through health factors - no DR present |
| tri_evidence | HF_imaging_TRI_TRR | DIABETIC RETINA-NO RETINOPATHY | Documentation of a teleretinal visit through health factors - no DR present |
| tri_evidence | HF_imaging_TRI_TRR | DIABETIC RETINOPATHY ABSENT | Documentation of a teleretinal visit through health factors - no DR present |
| tri_evidence | HF_imaging_TRI_TRR | DIABETES - NO RETINOPATHY | Documentation of a teleretinal visit through health factors - no DR present |
| tri_evidence | HF_imaging_TRI_TRR | DIABETIC RETINOPATHY - NO | Documentation of a teleretinal visit through health factors - no DR present |
| tri_evidence | HF_imaging_TRI_TRR | LTRI DM NO DIABETIC RETINOPATHY LT | Documentation of a teleretinal visit through health factors - no DR present |
| tri_evidence | HF_imaging_TRI_TRR | LTRI DM NO DIABETIC RETINOPATHY RT | Documentation of a teleretinal visit through health factors - no DR present |
| any_dr_hf | HF | LTRI DM FINDING DR OTHER LT | Diabetic retinopathy with no specific mention of severity |
| any_dr_hf | HF | LTRI DM FINDING DR OTHER RT | Diabetic retinopathy with no specific mention of severity |
| any_dr_hf | HF | LTRI DM FINDING DR PREV RET LASER TX LT | Diabetic retinopathy with no specific mention of severity + mention of previous treatment |
| any_dr_hf | HF | LTRI DM FINDING DR PREV RET LASER TX RT | Diabetic retinopathy with no specific mention of severity + mention of previous treatment |
| any_dr_hf | HF | LTRI DM HX LASER THERAPY DIAB RETINOPATH | Diabetic retinopathy with no specific mention of severity + mention of previous treatment |
| any_dr_hf | HF | LTRI DM HX LASER TX DIAB RETINOPATHY | Diabetic retinopathy with no specific mention of severity + mention of previous treatment |
| any_dr_hf | HF | LTRI DM RETINOPATHY EXAM ABNORMAL | Diabetic retinopathy with no specific mention of severity |
| any_dr_hf | HF | TRI DM FINDING DR OTHER LT | Diabetic retinopathy with no specific mention of severity |
| any_dr_hf | HF | TRI DM FINDING DR OTHER RT | Diabetic retinopathy with no specific mention of severity |
| any_dr_hf | HF | TRI DM FINDING DR PREV RET LASER TX LT | Diabetic retinopathy with no specific mention of severity + mention of previous treatment |
| any_dr_hf | HF | TRI DM FINDING DR PREV RET LASER TX RT | Diabetic retinopathy with no specific mention of severity + mention of previous treatment |
| any_dr_hf | HF | TRI DM HX LASER THERAPY DIAB RETINOPATHY | Diabetic retinopathy with no specific mention of severity + mention of previous treatment |
| any_dr_hf | HF | TRI DM RETINOPATHY EXAM ABNORMAL | Diabetic retinopathy with no specific mention of severity |
| any_dr_hf | HF | TRI DM FINDING DR MACULAR EDEMA LT | Diabetic retinopathy (with macular edema), left eye |
| any_dr_hf | HF | TRI DM FINDING DR MACULAR EDEMA RT | Diabetic retinopathy (with macular edema), right eye |
| any_dr_hf | HF | DIABETES - RETINOPATHY | Diabetic retinopathy with no specific mention of severity |
| any_dr_hf | HF | DIABETIC EYE RETINOPATHY FOUND | Diabetic retinopathy with no specific mention of severity |
| any_dr_hf | HF | Diabetic Retina-Retinopathy | Diabetic retinopathy with no specific mention of severity |
| any_dr_hf | HF | DIABETIC RETINOPATHY | Diabetic retinopathy with no specific mention of severity |
| any_dr_hf | HF | DIABETIC RETINOPATHY PRESENT | Diabetic retinopathy with no specific mention of severity |
| any_dr_hf | HF | HX DIABETIC RETINOPATHY | History of diabetic retinopathy with no specific mention of severity |
| any_dr_hf | HF | LTRI DM FINDING DR MACULAR EDEMA LT | Diabetic retinopathy with no specific mention of severity |
| any_dr_hf | HF | LTRI DM FINDING DR MACULAR EDEMA RT | Diabetic retinopathy with no specific mention of severity |
| any_dr_hf | HF | LTRI DM FINDING DR NO MAC EDEMA APP LT | Diabetic retinopathy with no specific mention of severity; no evidence of macular eduema |
| any_dr_hf | HF | LTRI DM FINDING DR NO MAC EDEMA APP RT | Diabetic retinopathy with no specific mention of severity; no evidence of macular eduema |
| any_dr_hf | HF | LTRI DM REFER RETINOPATHY | Diabetic retinopathy with no specific mention of severity |
| any_dr_hf | HF | TRI DM REFER RETINOPATHY | Diabetic retinopathy |
| any_dr_hf | HF | VA-TRR DM RETINOPATHY EXAM ABNORMAL | Diabetic retinopathy with no specific mention of severity |
| any_dr_hf | HF | VA-TRR DM2 FIND DR PREV RET LASER TX LT | Diabetic retinopathy with no specific mention of severity + mention of previous treatment |
| any_dr_hf | HF | VA-TRR DM2 FIND DR PREV RET LASER TX RT | Diabetic retinopathy with no specific mention of severity + mention of previous treatment |
| any_dr_hf | HF | VA-TRR DM2 FINDING DR OTHER LT | Diabetic retinopathy with no specific mention of severity |
| any_dr_hr | HF | VA-TRR DM2 FINDING DR OTHER RT | Diabetic retinopathy with no specific mention of severity |
| any_dr_hf | HF | LTRI DM FINDING DR MACULAR EDEMA LT | Diabetic macular edema |
| any_dr_hf | HF | LTRI DM FINDING DR MACULAR EDEMA RT | Diabetic macular edema |
| any_dr_hf | HF | LTRI DM FINDING DR MACULAR EDEMAL LT | Diabetic macular edema |
| any_dr_hf | HF | TRI DM FINDING DR MACULAR EDEMA LT | Diabetic macular edema |
| any_dr_hf | HF | TRI DM FINDING DR MACULAR EDEMA RT | Diabetic macular edema |
| mnpdr_hf | HF | DM MILD NONPROLIFERATIVE RETINOPATHY LT | Mild nonproliferative DR, left eye |
| mnpdr_hf | HF | DM MILD NONPROLIFERATIVE RETINOPATHY RT | Mild nonproliferative DR, right eye |
| mnpdr_hf | HF | DM MOD NONPROLIFERATIVE RETINOPATHY LT | Moderate nonproliferative DR, left eye |
| mnpdr_hf | HF | DM MOD NONPROLIFERATIVE RETINOPATHY RT | Moderate nonproliferative DR, right eye |
| mnpdr_hf | HF | LTRI DM DR MILD NONPROLIF LT | Mild nonproliferative DR, left eye |
| mnpdr_hf | HF | LTRI DM FINDING DR MILD NONPROLIF LT | Mild nonproliferative DR, left eye |
| mnpdr_hf | HF | LTRI DM FINDING DR MILD NONPROLIF RT | Mild nonproliferative DR, right eye |
| mnpdr_hf | HF | LTRI DM FINDING DR MOD NONPROLIF LT | Moderate nonproliferative DR, left eye |
| mnpdr_hf | HF | LTRI DM FINDING DR MOD NONPROLIF RT | Moderate nonproliferative DR, right eye |
| mnpdr_hf | HF | LTRI DM FINDING MILD NONPROLIF RT | Mild nonproliferative DR, right eye |
| mnpdr_hf | HF | LTRI DM MOD NONPROLIF LT | Moderate nonproliferative DR, left eye |
| mnpdr_hf | HF | TRI DM FINDING DR MILD NONPROLIF LT | Mild nonproliferative DR, left eye |
| mnpdr_hf | HF | TRI DM FINDING DR MILD NONPROLIF RT | Mild nonproliferative DR, right eye |
| mnpdr_hf | HF | TRI DM FINDING DR MOD NONPROLIF LT | Moderate nonproliferative DR, left eye |
| mnpdr_hf | HF | TRI DM FINDING DR MOD NONPROLIF RT | Moderate nonproliferative DR, right eye |
| mnpdr_hf | HF | VA-TRR DM2 FIND DR MLD MAC NO EDEMA LT | Mild nonproliferative DR with no macular edema, left eye |
| mnpdr_hf | HF | VA-TRR DM2 FIND DR MLD MAC NO EDEMA RT | Mild nonproliferative DR with no macular edema, right eye |
| mnpdr_hf | HF | VA-TRR DM2 FIND DR MOD MAC NO EDEMA LT | Moderate nonproliferative DR with no macular edema, left eye |
| mnpdr_hf | HF | VA-TRR DM2 FIND DR MOD MAC NO EDEMA RT | Moderate nonproliferative DR with no macular edema, right eye |
| mnpdr_hf | HF | VA-TRR DM2 FINDING DR MLD MAC EDEMA LT | Mild nonproliferative DR with macular edema, left eye |
| mnpdr_hf | HF | VA-TRR DM2 FINDING DR MLD MAC EDEMA RT | Mild nonproliferative DR with macular edema right eye |
| mnpdr_hf | HF | VA-TRR DM2 FINDING DR MOD MAC EDEMA LT | Moderate nonproliferative DR with macular edema, left eye |
| mnpdr_hf | HF | VA-TRR DM2 FINDING DR MOD MAC EDEMA RT | Moderate nonproliferative DR with macular edema, right eye |
| nsdr_hf | HF | DIABETIC RETINA-MOD. BKGRD RETINOPATHY | Nonspecific DR (moderate background retinopathy) |
| pdr_hf | HF | DM PROLIFERATIVE RETINOPATHY LT | Proliferative DR, left eye |
| pdr_hf | HF | DM PROLIFERATIVE RETINOPATHY RT | Proliferative DR, right eye |
| pdr_hf | HF | LTRI DM FINDING DR PROLIFERATIVE LT | Proliferative DR, left eye |
| pdr_hf | HF | LTRI DM FINDING DR PROLIFERATIVE RT | Proliferative DR, right eye |
| pdr_hf | HF | TRI DM FINDING DR PROLIFERATIVE LT | Proliferative DR, left eye |
| pdr_hf | HF | TRI DM FINDING DR PROLIFERATIVE RT | Proliferative DR, right eye |
| pdr_hf | HF | VA-TRR DM2 FIND DR PDR MAC NO EDEMA LT | Proliferative DR (no macular edema), left eye |
| snpdr_hf | HF | DM SEVERE NONPROLIFERATIVE RETINOPATHY L | Severe nonproliferative DR, left eye |
| snpdr_hf | HF | DM SEVERE NONPROLIFERATIVE RETINOPATHY R | Severe nonproliferative DR, right eye |
| snpdr_hf | HF | LTRI DM FINDING DR SEVERE NONPROLIF LT | Severe nonproliferative DR, left eye |
| snpdr_hf | HF | LTRI DM FINDING DR SEVERE NONPROLIF RT | Severe nonproliferative DR, right eye |
| snpdr_hf | HF | TRI DM FINDING DR SEVERE NONPROLIF LT | Severe nonproliferative DR, left eye |
| snpdr_hf | HF | TRI DM FINDING DR SEVERE NONPROLIF RT | Severe nonproliferative DR, right eye |
| snpdr_hf | HF | VA-TRR DM2 FIND DR SEV MAC NO EDEMA RT | Severe nonproliferative DR (no macular edema), right eye |
| mnpdr_hf | HF | LTRI DM DR MILD NONPROLIF RT | Mild nonproliferative DR right eye |
| mnpdr_hf | HF | LTRI DM MOD NONPROLIF RT | Moderate nonproliferative DR right eye |
| pdr_hf | HF | VA-TRR DM2 FIND DR PDR MAC NO EDEMA RT | Proliferative DR, with no macular edema, right eye |
| snpdr_hf | HF | VA-TRR DM2 FIND DR SEV MAC NO EDEMA LT | Severe non-proliferative DR, no macular edema, left eye |

Supplemental Table 2b. List of ICD codes included in the DM algorithm along with DM exclusions

| Trait | Domain | Code | Concept Name |
| --- | --- | --- | --- |
| t1d_inclusions | ICD9CM | 250.01 | Diabetes mellitus without mention of complication, type I [juvenile type], not stated as uncontrolled |
| t1d_inclusions | ICD9CM | 250.03 | Diabetes mellitus without mention of complication, type I [juvenile type], uncontrolled |
| t1d_inclusions | ICD9CM | 250.21 | Diabetes with hyperosmolarity, type I [juvenile type], not stated as uncontrolled |
| t1d_inclusions | ICD9CM | 250.23 | Diabetes with hyperosmolarity, type I [juvenile type], uncontrolled |
| t1d_inclusions | ICD9CM | 250.31 | Diabetes with other coma, type I [juvenile type], not stated as uncontrolled |
| t1d_inclusions | ICD9CM | 250.33 | Diabetes with other coma, type I [juvenile type], uncontrolled |
| t1d_inclusions | ICD9CM | 250.41 | Diabetes with renal manifestations, type I [juvenile type], not stated as uncontrolled |
| t1d_inclusions | ICD9CM | 250.43 | Diabetes with renal manifestations, type I [juvenile type], uncontrolled |
| t1d_inclusions | ICD9CM | 250.51 | Diabetes with ophthalmic manifestations, type I [juvenile type], not stated as uncontrolled |
| t1d_inclusions | ICD9CM | 250.53 | Diabetes with ophthalmic manifestations, type I [juvenile type], uncontrolled |
| t1d_inclusions | ICD9CM | 250.61 | Diabetes with neurological manifestations, type I [juvenile type], not stated as uncontrolled |
| t1d_inclusions | ICD9CM | 250.63 | Diabetes with neurological manifestations, type I [juvenile type], uncontrolled |
| t1d_inclusions | ICD9CM | 250.71 | Diabetes with peripheral circulatory disorders, type I [juvenile type], not stated as uncontrolled |
| t1d_inclusions | ICD9CM | 250.73 | Diabetes with peripheral circulatory disorders, type I [juvenile type], uncontrolled |
| t1d_inclusions | ICD9CM | 250.81 | Diabetes with other specified manifestations, type I [juvenile type], not stated as uncontrolled |
| t1d_inclusions | ICD9CM | 250.83 | Diabetes with other specified manifestations, type I [juvenile type], uncontrolled |
| t1d_inclusions | ICD9CM | 250.91 | Diabetes with unspecified complication, type I [juvenile type], not stated as uncontrolled |
| t1d_inclusions | ICD9CM | 250.93 | Diabetes with unspecified complication, type I [juvenile type], uncontrolled |
| t1d_inclusions | ICD10 | E10 | Type 1 diabetes mellitus |
| t1d_inclusions | ICD10CM | E10 | Type 1 diabetes mellitus |
| t1d_inclusions | ICD10 | E10.0 | Type 1 diabetes mellitus, With coma |
| t1d_inclusions | ICD10CM | E10.1 | Type 1 diabetes mellitus with ketoacidosis |
| t1d_inclusions | ICD10 | E10.1 | Type 1 diabetes mellitus, With ketoacidosis |
| t1d_inclusions | ICD10CM | E10.11 | Type 1 diabetes mellitus with ketoacidosis with coma |
| t1d_inclusions | ICD10CM | E10.2 | Type 1 diabetes mellitus with kidney complications |
| t1d_inclusions | ICD10 | E10.2 | Type 1 diabetes mellitus, With renal complications |
| t1d_inclusions | ICD10CM | E10.21 | Type 1 diabetes mellitus with diabetic nephropathy |
| t1d_inclusions | ICD10CM | E10.22 | Type 1 diabetes mellitus with diabetic chronic kidney disease |
| t1d_inclusions | ICD10CM | E10.29 | Type 1 diabetes mellitus with other diabetic kidney complication |
| t1d_inclusions | ICD10CM | E10.3 | Type 1 diabetes mellitus with ophthalmic complications |
| t1d_inclusions | ICD10 | E10.3 | Type 1 diabetes mellitus, With ophthalmic complications |
| t1d_inclusions | ICD10CM | E10.31 | Type 1 diabetes mellitus with unspecified diabetic retinopathy |
| t1d_inclusions | ICD10CM | E10.311 | Type 1 diabetes mellitus with unspecified diabetic retinopathy with macular edema |
| t1d_inclusions | ICD10CM | E10.319 | Type 1 diabetes mellitus with unspecified diabetic retinopathy without macular edema |
| t1d_inclusions | ICD10CM | E10.32 | Type 1 diabetes mellitus with mild nonproliferative diabetic retinopathy |
| t1d_inclusions | ICD10CM | E10.321 | Type 1 diabetes mellitus with mild nonproliferative diabetic retinopathy with macular edema |
| t1d_inclusions | ICD10CM | E10.3211 | Type 1 diabetes mellitus with mild nonproliferative diabetic retinopathy with macular edema, right eye |
| t1d_inclusions | ICD10CM | E10.3212 | Type 1 diabetes mellitus with mild nonproliferative diabetic retinopathy with macular edema, left eye |
| t1d_inclusions | ICD10CM | E10.3213 | Type 1 diabetes mellitus with mild nonproliferative diabetic retinopathy with macular edema, bilateral |
| t1d_inclusions | ICD10CM | E10.3219 | Type 1 diabetes mellitus with mild nonproliferative diabetic retinopathy with macular edema, unspecified eye |
| t1d_inclusions | ICD10CM | E10.329 | Type 1 diabetes mellitus with mild nonproliferative diabetic retinopathy without macular edema |
| t1d_inclusions | ICD10CM | E10.3291 | Type 1 diabetes mellitus with mild nonproliferative diabetic retinopathy without macular edema, right eye |
| t1d_inclusions | ICD10CM | E10.3292 | Type 1 diabetes mellitus with mild nonproliferative diabetic retinopathy without macular edema, left eye |
| t1d_inclusions | ICD10CM | E10.3293 | Type 1 diabetes mellitus with mild nonproliferative diabetic retinopathy without macular edema, bilateral |
| t1d_inclusions | ICD10CM | E10.3299 | Type 1 diabetes mellitus with mild nonproliferative diabetic retinopathy without macular edema, unspecified eye |
| t1d_inclusions | ICD10CM | E10.33 | Type 1 diabetes mellitus with moderate nonproliferative diabetic retinopathy |
| t1d_inclusions | ICD10CM | E10.331 | Type 1 diabetes mellitus with moderate nonproliferative diabetic retinopathy with macular edema |
| t1d_inclusions | ICD10CM | E10.3311 | Type 1 diabetes mellitus with moderate nonproliferative diabetic retinopathy with macular edema, right eye |
| t1d_inclusions | ICD10CM | E10.3312 | Type 1 diabetes mellitus with moderate nonproliferative diabetic retinopathy with macular edema, left eye |
| t1d_inclusions | ICD10CM | E10.3313 | Type 1 diabetes mellitus with moderate nonproliferative diabetic retinopathy with macular edema, bilateral |
| t1d_inclusions | ICD10CM | E10.3319 | Type 1 diabetes mellitus with moderate nonproliferative diabetic retinopathy with macular edema, unspecified eye |
| t1d_inclusions | ICD10CM | E10.339 | Type 1 diabetes mellitus with moderate nonproliferative diabetic retinopathy without macular edema |
| t1d_inclusions | ICD10CM | E10.3391 | Type 1 diabetes mellitus with moderate nonproliferative diabetic retinopathy without macular edema, right eye |
| t1d_inclusions | ICD10CM | E10.3392 | Type 1 diabetes mellitus with moderate nonproliferative diabetic retinopathy without macular edema, left eye |
| t1d_inclusions | ICD10CM | E10.3393 | Type 1 diabetes mellitus with moderate nonproliferative diabetic retinopathy without macular edema, bilateral |
| t1d_inclusions | ICD10CM | E10.3399 | Type 1 diabetes mellitus with moderate nonproliferative diabetic retinopathy without macular edema, unspecified eye |
| t1d_inclusions | ICD10CM | E10.34 | Type 1 diabetes mellitus with severe nonproliferative diabetic retinopathy |
| t1d_inclusions | ICD10CM | E10.341 | Type 1 diabetes mellitus with severe nonproliferative diabetic retinopathy with macular edema |
| t1d_inclusions | ICD10CM | E10.3411 | Type 1 diabetes mellitus with severe nonproliferative diabetic retinopathy with macular edema, right eye |
| t1d_inclusions | ICD10CM | E10.3412 | Type 1 diabetes mellitus with severe nonproliferative diabetic retinopathy with macular edema, left eye |
| t1d_inclusions | ICD10CM | E10.3413 | Type 1 diabetes mellitus with severe nonproliferative diabetic retinopathy with macular edema, bilateral |
| t1d_inclusions | ICD10CM | E10.3419 | Type 1 diabetes mellitus with severe nonproliferative diabetic retinopathy with macular edema, unspecified eye |
| t1d_inclusions | ICD10CM | E10.349 | Type 1 diabetes mellitus with severe nonproliferative diabetic retinopathy without macular edema |
| t1d_inclusions | ICD10CM | E10.3491 | Type 1 diabetes mellitus with severe nonproliferative diabetic retinopathy without macular edema, right eye |
| t1d_inclusions | ICD10CM | E10.3492 | Type 1 diabetes mellitus with severe nonproliferative diabetic retinopathy without macular edema, left eye |
| t1d_inclusions | ICD10CM | E10.3493 | Type 1 diabetes mellitus with severe nonproliferative diabetic retinopathy without macular edema, bilateral |
| t1d_inclusions | ICD10CM | E10.3499 | Type 1 diabetes mellitus with severe nonproliferative diabetic retinopathy without macular edema, unspecified eye |
| t1d_inclusions | ICD10CM | E10.35 | Type 1 diabetes mellitus with proliferative diabetic retinopathy |
| t1d_inclusions | ICD10CM | E10.351 | Type 1 diabetes mellitus with proliferative diabetic retinopathy with macular edema |
| t1d_inclusions | ICD10CM | E10.3511 | Type 1 diabetes mellitus with proliferative diabetic retinopathy with macular edema, right eye |
| t1d_inclusions | ICD10CM | E10.3512 | Type 1 diabetes mellitus with proliferative diabetic retinopathy with macular edema, left eye |
| t1d_inclusions | ICD10CM | E10.3513 | Type 1 diabetes mellitus with proliferative diabetic retinopathy with macular edema, bilateral |
| t1d_inclusions | ICD10CM | E10.3519 | Type 1 diabetes mellitus with proliferative diabetic retinopathy with macular edema, unspecified eye |
| t1d_inclusions | ICD10CM | E10.352 | Type 1 diabetes mellitus with proliferative diabetic retinopathy with traction retinal detachment involving the macula |
| t1d_inclusions | ICD10CM | E10.3521 | Type 1 diabetes mellitus with proliferative diabetic retinopathy with traction retinal detachment involving the macula, right eye |
| t1d_inclusions | ICD10CM | E10.3522 | Type 1 diabetes mellitus with proliferative diabetic retinopathy with traction retinal detachment involving the macula, left eye |
| t1d_inclusions | ICD10CM | E10.3523 | Type 1 diabetes mellitus with proliferative diabetic retinopathy with traction retinal detachment involving the macula, bilateral |
| t1d_inclusions | ICD10CM | E10.3529 | Type 1 diabetes mellitus with proliferative diabetic retinopathy with traction retinal detachment involving the macula, unspecified eye |
| t1d_inclusions | ICD10CM | E10.353 | Type 1 diabetes mellitus with proliferative diabetic retinopathy with traction retinal detachment not involving the macula |
| t1d_inclusions | ICD10CM | E10.3531 | Type 1 diabetes mellitus with proliferative diabetic retinopathy with traction retinal detachment not involving the macula, right eye |
| t1d_inclusions | ICD10CM | E10.3532 | Type 1 diabetes mellitus with proliferative diabetic retinopathy with traction retinal detachment not involving the macula, left eye |
| t1d_inclusions | ICD10CM | E10.3533 | Type 1 diabetes mellitus with proliferative diabetic retinopathy with traction retinal detachment not involving the macula, bilateral |
| t1d_inclusions | ICD10CM | E10.3539 | Type 1 diabetes mellitus with proliferative diabetic retinopathy with traction retinal detachment not involving the macula, unspecified eye |
| t1d_inclusions | ICD10CM | E10.354 | Type 1 diabetes mellitus with proliferative diabetic retinopathy with combined traction retinal detachment and rhegmatogenous retinal detachment |
| t1d_inclusions | ICD10CM | E10.3541 | Type 1 diabetes mellitus with proliferative diabetic retinopathy with combined traction retinal detachment and rhegmatogenous retinal detachment, right eye |
| t1d_inclusions | ICD10CM | E10.3542 | Type 1 diabetes mellitus with proliferative diabetic retinopathy with combined traction retinal detachment and rhegmatogenous retinal detachment, left eye |
| t1d_inclusions | ICD10CM | E10.3543 | Type 1 diabetes mellitus with proliferative diabetic retinopathy with combined traction retinal detachment and rhegmatogenous retinal detachment, bilateral |
| t1d_inclusions | ICD10CM | E10.3549 | Type 1 diabetes mellitus with proliferative diabetic retinopathy with combined traction retinal detachment and rhegmatogenous retinal detachment, unspecified eye |
| t1d_inclusions | ICD10CM | E10.355 | Type 1 diabetes mellitus with stable proliferative diabetic retinopathy |
| t1d_inclusions | ICD10CM | E10.3551 | Type 1 diabetes mellitus with stable proliferative diabetic retinopathy, right eye |
| t1d_inclusions | ICD10CM | E10.3552 | Type 1 diabetes mellitus with stable proliferative diabetic retinopathy, left eye |
| t1d_inclusions | ICD10CM | E10.3553 | Type 1 diabetes mellitus with stable proliferative diabetic retinopathy, bilateral |
| t1d_inclusions | ICD10CM | E10.3559 | Type 1 diabetes mellitus with stable proliferative diabetic retinopathy, unspecified eye |
| t1d_inclusions | ICD10CM | E10.359 | Type 1 diabetes mellitus with proliferative diabetic retinopathy without macular edema |
| t1d_inclusions | ICD10CM | E10.3591 | Type 1 diabetes mellitus with proliferative diabetic retinopathy without macular edema, right eye |
| t1d_inclusions | ICD10CM | E10.3592 | Type 1 diabetes mellitus with proliferative diabetic retinopathy without macular edema, left eye |
| t1d_inclusions | ICD10CM | E10.3593 | Type 1 diabetes mellitus with proliferative diabetic retinopathy without macular edema, bilateral |
| t1d_inclusions | ICD10CM | E10.3599 | Type 1 diabetes mellitus with proliferative diabetic retinopathy without macular edema, unspecified eye |
| t1d_inclusions | ICD10CM | E10.36 | Type 1 diabetes mellitus with diabetic cataract |
| t1d_inclusions | ICD10CM | E10.37 | Type 1 diabetes mellitus with diabetic macular edema, resolved following treatment |
| t1d_inclusions | ICD10CM | E10.37X1 | Type 1 diabetes mellitus with diabetic macular edema, resolved following treatment, right eye |
| t1d_inclusions | ICD10CM | E10.37X2 | Type 1 diabetes mellitus with diabetic macular edema, resolved following treatment, left eye |
| t1d_inclusions | ICD10CM | E10.37X3 | Type 1 diabetes mellitus with diabetic macular edema, resolved following treatment, bilateral |
| t1d_inclusions | ICD10CM | E10.37X9 | Type 1 diabetes mellitus with diabetic macular edema, resolved following treatment, unspecified eye |
| t1d_inclusions | ICD10CM | E10.39 | Type 1 diabetes mellitus with other diabetic ophthalmic complication |
| t1d_inclusions | ICD10 | E10.4 | Type 1 diabetes mellitus, With neurological complications |
| t1d_inclusions | ICD10CM | E10.4 | Type 1 diabetes mellitus with neurological complications |
| t1d_inclusions | ICD10CM | E10.40 | Type 1 diabetes mellitus with diabetic neuropathy, unspecified |
| t1d_inclusions | ICD10CM | E10.41 | Type 1 diabetes mellitus with diabetic mononeuropathy |
| t1d_inclusions | ICD10CM | E10.42 | Type 1 diabetes mellitus with diabetic polyneuropathy |
| t1d_inclusions | ICD10CM | E10.43 | Type 1 diabetes mellitus with diabetic autonomic (poly)neuropathy |
| t1d_inclusions | ICD10CM | E10.44 | Type 1 diabetes mellitus with diabetic amyotrophy |
| t1d_inclusions | ICD10CM | E10.49 | Type 1 diabetes mellitus with other diabetic neurological complication |
| t1d_inclusions | ICD10 | E10.5 | Type 1 diabetes mellitus, With peripheral circulatory complications |
| t1d_inclusions | ICD10CM | E10.5 | Type 1 diabetes mellitus with circulatory complications |
| t1d_inclusions | ICD10CM | E10.51 | Type 1 diabetes mellitus with diabetic peripheral angiopathy without gangrene |
| t1d_inclusions | ICD10CM | E10.52 | Type 1 diabetes mellitus with diabetic peripheral angiopathy with gangrene |
| t1d_inclusions | ICD10CM | E10.59 | Type 1 diabetes mellitus with other circulatory complications |
| t1d_inclusions | ICD10 | E10.6 | Type 1 diabetes mellitus, With other specified complications |
| t1d_inclusions | ICD10CM | E10.6 | Type 1 diabetes mellitus with other specified complications |
| t1d_inclusions | ICD10CM | E10.61 | Type 1 diabetes mellitus with diabetic arthropathy |
| t1d_inclusions | ICD10CM | E10.610 | Type 1 diabetes mellitus with diabetic neuropathic arthropathy |
| t1d_inclusions | ICD10CM | E10.618 | Type 1 diabetes mellitus with other diabetic arthropathy |
| t1d_inclusions | ICD10CM | E10.62 | Type 1 diabetes mellitus with skin complications |
| t1d_inclusions | ICD10CM | E10.620 | Type 1 diabetes mellitus with diabetic dermatitis |
| t1d_inclusions | ICD10CM | E10.621 | Type 1 diabetes mellitus with foot ulcer |
| t1d_inclusions | ICD10CM | E10.622 | Type 1 diabetes mellitus with other skin ulcer |
| t1d_inclusions | ICD10CM | E10.628 | Type 1 diabetes mellitus with other skin complications |
| t1d_inclusions | ICD10CM | E10.63 | Type 1 diabetes mellitus with oral complications |
| t1d_inclusions | ICD10CM | E10.630 | Type 1 diabetes mellitus with periodontal disease |
| t1d_inclusions | ICD10CM | E10.638 | Type 1 diabetes mellitus with other oral complications |
| t1d_inclusions | ICD10CM | E10.64 | Type 1 diabetes mellitus with hypoglycemia |
| t1d_inclusions | ICD10CM | E10.640 | Type 1 diabetes mellitus with hypoglycemia without coma |
| t1d_inclusions | ICD10CM | E10.641 | Type 1 diabetes mellitus with hypoglycemia with coma |
| t1d_inclusions | ICD10CM | E10.649 | Type 1 diabetes mellitus with hypoglycemia without coma |
| t1d_inclusions | ICD10CM | E10.65 | Type 1 diabetes mellitus with hyperglycemia |
| t1d_inclusions | ICD10CM | E10.69 | Type 1 diabetes mellitus with other specified complication |
| t1d_inclusions | ICD10 | E10.7 | Type 1 diabetes mellitus, With multiple complications |
| t1d_inclusions | ICD10CM | E10.8 | Type 1 diabetes mellitus with unspecified complications |
| t1d_inclusions | ICD10 | E10.8 | Type 1 diabetes mellitus, With unspecified complications |
| t1d_inclusions | ICD10CM | E10.9 | Type 1 diabetes mellitus without complications |
| t1d_inclusions | ICD10 | E10.9 | Type 1 diabetes mellitus, Without complications |
| t1d_inclusions | ICD9CM | 250.11 | Diabetes with ketoacidosis, type I [juvenile type], not stated as uncontrolled |
| t1d_inclusions | ICD9CM | 250.13 | Diabetes with ketoacidosis, type I [juvenile type], uncontrolled |
| t1d_inclusions | ICD10CM | E10.10 | Type 1 diabetes mellitus with ketoacidosis without coma |
| dbm_inclusions | ICD9CM | 250.00 | Diabetes mellitus without mention of complication, type II or unspecified type, not stated as uncontrolled |
| dbm_inclusions | ICD9CM | 250 | Diabetes mellitus without mention of complication |
| dbm_inclusions | ICD9CM | 250.02 | Diabetes mellitus without mention of complication, type II or unspecified type, uncontrolled |
| dbm_inclusions | ICD9CM | 250.2 | Diabetes mellitus with hyperosmolarity |
| dbm_inclusions | ICD9CM | 250.20 | Diabetes with hyperosmolarity, type II or unspecified type, not stated as uncontrolled |
| dbm_inclusions | ICD9CM | 250.22 | Diabetes with hyperosmolarity, type II or unspecified type, uncontrolled |
| dbm_inclusions | ICD9CM | 250.30 | Diabetes with other coma, type II or unspecified type, not stated as uncontrolled |
| dbm_inclusions | ICD9CM | 250.3 | Diabetes with other coma |
| dbm_inclusions | ICD9CM | 250.32 | Diabetes with other coma, type II or unspecified type, uncontrolled |
| dbm_inclusions | ICD9CM | 250.4 | Diabetes with renal manifestations |
| dbm_inclusions | ICD9CM | 250.40 | Diabetes with renal manifestations, type II or unspecified type, not stated as uncontrolled |
| dbm_inclusions | ICD9CM | 250.42 | Diabetes with renal manifestations, type II or unspecified type, uncontrolled |
| dbm_inclusions | ICD9CM | 250.50 | Diabetes with ophthalmic manifestations, type II or unspecified type, not stated as uncontrolled |
| dbm_inclusions | ICD9CM | 250.52 | Diabetes with ophthalmic manifestations, type II or unspecified type, uncontrolled |
| dbm_inclusions | ICD9CM | 250.60 | Diabetes with neurological manifestations, type II or unspecified type, not stated as uncontrolled |
| dbm_inclusions | ICD9CM | 250.62 | Diabetes with neurological manifestations, type II or unspecified type, uncontrolled |
| dbm_inclusions | ICD9CM | 250.7 | Diabetes with peripheral circulatory disorders, type II or unspecified type, not stated as uncontrolled |
| dbm_inclusions | ICD9CM | 250.72 | Diabetes with peripheral circulatory disorders, type II or unspecified type, uncontrolled |
| dbm_inclusions | ICD9CM | 250.80 | Diabetes with other specified manifestations, type II or unspecified type, not stated as uncontrolled |
| dbm_inclusions | ICD9CM | 250.82 | Diabetes with other specified manifestations, type II or unspecified type, uncontrolled |
| dbm_inclusions | ICD9CM | 250.90 | Diabetes with unspecified complication, type II or unspecified type, not stated as uncontrolled |
| dbm_inclusions | ICD9CM | 250.92 | Diabetes with unspecified complication, type II or unspecified type, uncontrolled |
| dbm_inclusions | ICD10CM | E11 | Type 2 diabetes mellitus |
| dbm_inclusions | ICD10CM | E11.0 | Type 2 diabetes mellitus with hyperosmolarity |
| dbm_inclusions | ICD10CM | E11.00 | Type 2 diabetes mellitus with hyperosmolarity without nonketotic hyperglycemic-hyperosmolar coma (NKHHC) |
| dbm_inclusions | ICD10CM | E11.01 | Type 2 diabetes mellitus with hyperosmolarity with coma |
| dbm_inclusions | ICD10CM | E11.2 | Type 2 diabetes mellitus with kidney complications |
| dbm_inclusions | ICD10CM | E11.21 | Type 2 diabetes mellitus with diabetic nephropathy |
| dbm_inclusions | ICD10CM | E11.22 | Type 2 diabetes mellitus with diabetic chronic kidney disease |
| dbm_inclusions | ICD10CM | E11.29 | Type 2 diabetes mellitus with other diabetic kidney complication |
| dbm_inclusions | ICD10CM | E11.3 | Type 2 diabetes mellitus with ophthalmic complications |
| dbm_inclusions | ICD10CM | E11.31 | Type 2 diabetes mellitus with unspecified diabetic retinopathy |
| dbm_inclusions | ICD10CM | E11.311 | Type 2 diabetes mellitus with unspecified diabetic retinopathy with macular edema |
| dbm_inclusions | ICD10CM | E11.319 | Type 2 diabetes mellitus with unspecified diabetic retinopathy without macular edema |
| dbm_inclusions | ICD10CM | E11.32 | Type 2 diabetes mellitus with mild nonproliferative diabetic retinopathy |
| dbm_inclusions | ICD10CM | E11.321 | Type 2 diabetes mellitus with mild nonproliferative diabetic retinopathy with macular edema |
| dbm_inclusions | ICD10CM | E11.3211 | Type 2 diabetes mellitus with mild nonproliferative diabetic retinopathy with macular edema, right eye |
| dbm_inclusions | ICD10CM | E11.3212 | Type 2 diabetes mellitus with mild nonproliferative diabetic retinopathy with macular edema, left eye |
| dbm_inclusions | ICD10CM | E11.3213 | Type 2 diabetes mellitus with mild nonproliferative diabetic retinopathy with macular edema, bilateral |
| dbm_inclusions | ICD10CM | E11.3219 | Type 2 diabetes mellitus with mild nonproliferative diabetic retinopathy with macular edema, unspecified eye |
| dbm_inclusions | ICD10CM | E11.329 | Type 2 diabetes mellitus with mild nonproliferative diabetic retinopathy without macular edema |
| dbm_inclusions | ICD10CM | E11.3291 | Type 2 diabetes mellitus with mild nonproliferative diabetic retinopathy without macular edema, right eye |
| dbm_inclusions | ICD10CM | E11.3292 | Type 2 diabetes mellitus with mild nonproliferative diabetic retinopathy without macular edema, left eye |
| dbm_inclusions | ICD10CM | E11.3293 | Type 2 diabetes mellitus with mild nonproliferative diabetic retinopathy without macular edema, bilateral |
| dbm_inclusions | ICD10CM | E11.3299 | Type 2 diabetes mellitus with mild nonproliferative diabetic retinopathy without macular edema, unspecified eye |
| dbm_inclusions | ICD10CM | E11.33 | Type 2 diabetes mellitus with moderate nonproliferative diabetic retinopathy |
| dbm_inclusions | ICD10CM | E11.331 | Type 2 diabetes mellitus with moderate nonproliferative diabetic retinopathy with macular edema |
| dbm_inclusions | ICD10CM | E11.3311 | Type 2 diabetes mellitus with moderate nonproliferative diabetic retinopathy with macular edema, right eye |
| dbm_inclusions | ICD10CM | E11.3312 | Type 2 diabetes mellitus with moderate nonproliferative diabetic retinopathy with macular edema, left eye |
| dbm_inclusions | ICD10CM | E11.3313 | Type 2 diabetes mellitus with moderate nonproliferative diabetic retinopathy with macular edema, bilateral |
| dbm_inclusions | ICD10CM | E11.3319 | Type 2 diabetes mellitus with moderate nonproliferative diabetic retinopathy with macular edema, unspecified eye |
| dbm_inclusions | ICD10CM | E11.339 | Type 2 diabetes mellitus with moderate nonproliferative diabetic retinopathy without macular edema |
| dbm_inclusions | ICD10CM | E11.3391 | Type 2 diabetes mellitus with moderate nonproliferative diabetic retinopathy without macular edema, right eye |
| dbm_inclusions | ICD10CM | E11.3392 | Type 2 diabetes mellitus with moderate nonproliferative diabetic retinopathy without macular edema, left eye |
| dbm_inclusions | ICD10CM | E11.3393 | Type 2 diabetes mellitus with moderate nonproliferative diabetic retinopathy without macular edema, bilateral |
| dbm_inclusions | ICD10CM | E11.3399 | Type 2 diabetes mellitus with moderate nonproliferative diabetic retinopathy without macular edema, unspecified eye |
| dbm_inclusions | ICD10CM | E11.34 | Type 2 diabetes mellitus with severe nonproliferative diabetic retinopathy |
| dbm_inclusions | ICD10CM | E11.341 | Type 2 diabetes mellitus with severe nonproliferative diabetic retinopathy with macular edema |
| dbm_inclusions | ICD10CM | E11.3411 | Type 2 diabetes mellitus with severe nonproliferative diabetic retinopathy with macular edema, right eye |
| dbm_inclusions | ICD10CM | E11.3412 | Type 2 diabetes mellitus with severe nonproliferative diabetic retinopathy with macular edema, left eye |
| dbm_inclusions | ICD10CM | E11.3413 | Type 2 diabetes mellitus with severe nonproliferative diabetic retinopathy with macular edema, bilateral |
| dbm_inclusions | ICD10CM | E11.3419 | Type 2 diabetes mellitus with severe nonproliferative diabetic retinopathy with macular edema, unspecified eye |
| dbm_inclusions | ICD10CM | E11.349 | Type 2 diabetes mellitus with severe nonproliferative diabetic retinopathy without macular edema |
| dbm_inclusions | ICD10CM | E11.3491 | Type 2 diabetes mellitus with severe nonproliferative diabetic retinopathy without macular edema, right eye |
| dbm_inclusions | ICD10CM | E11.3492 | Type 2 diabetes mellitus with severe nonproliferative diabetic retinopathy without macular edema, left eye |
| dbm_inclusions | ICD10CM | E11.3493 | Type 2 diabetes mellitus with severe nonproliferative diabetic retinopathy without macular edema, bilateral |
| dbm_inclusions | ICD10CM | E11.3499 | Type 2 diabetes mellitus with severe nonproliferative diabetic retinopathy without macular edema, unspecified eye |
| dbm_inclusions | ICD10CM | E11.35 | Type 2 diabetes mellitus with proliferative diabetic retinopathy |
| dbm_inclusions | ICD10CM | E11.351 | Type 2 diabetes mellitus with proliferative diabetic retinopathy with macular edema |
| dbm_inclusions | ICD10CM | E11.3511 | Type 2 diabetes mellitus with proliferative diabetic retinopathy with macular edema, right eye |
| dbm_inclusions | ICD10CM | E11.3512 | Type 2 diabetes mellitus with proliferative diabetic retinopathy with macular edema, left eye |
| dbm_inclusions | ICD10CM | E11.3513 | Type 2 diabetes mellitus with proliferative diabetic retinopathy with macular edema, bilateral |
| dbm_inclusions | ICD10CM | E11.3519 | Type 2 diabetes mellitus with proliferative diabetic retinopathy with macular edema, unspecified eye |
| dbm_inclusions | ICD10CM | E11.352 | Type 2 diabetes mellitus with proliferative diabetic retinopathy with traction retinal detachment involving the macula |
| dbm_inclusions | ICD10CM | E11.3521 | Type 2 diabetes mellitus with proliferative diabetic retinopathy with traction retinal detachment involving the macula, right eye |
| dbm_inclusions | ICD10CM | E11.3522 | Type 2 diabetes mellitus with proliferative diabetic retinopathy with traction retinal detachment involving the macula, left eye |
| dbm_inclusions | ICD10CM | E11.3523 | Type 2 diabetes mellitus with proliferative diabetic retinopathy with traction retinal detachment involving the macula, bilateral |
| dbm_inclusions | ICD10CM | E11.3529 | Type 2 diabetes mellitus with proliferative diabetic retinopathy with traction retinal detachment involving the macula, unspecified eye |
| dbm_inclusions | ICD10CM | E11.353 | Type 2 diabetes mellitus with proliferative diabetic retinopathy with traction retinal detachment not involving the macula |
| dbm_inclusions | ICD10CM | E11.3531 | Type 2 diabetes mellitus with proliferative diabetic retinopathy with traction retinal detachment not involving the macula, right eye |
| dbm_inclusions | ICD10CM | E11.3532 | Type 2 diabetes mellitus with proliferative diabetic retinopathy with traction retinal detachment not involving the macula, left eye |
| dbm_inclusions | ICD10CM | E11.3533 | Type 2 diabetes mellitus with proliferative diabetic retinopathy with traction retinal detachment not involving the macula, bilateral |
| dbm_inclusions | ICD10CM | E11.3539 | Type 2 diabetes mellitus with proliferative diabetic retinopathy with traction retinal detachment not involving the macula, unspecified eye |
| dbm_inclusions | ICD10CM | E11.354 | Type 2 diabetes mellitus with proliferative diabetic retinopathy with combined traction retinal detachment and rhegmatogenous retinal detachment |
| dbm_inclusions | ICD10CM | E11.3541 | Type 2 diabetes mellitus with proliferative diabetic retinopathy with combined traction retinal detachment and rhegmatogenous retinal detachment, right eye |
| dbm_inclusions | ICD10CM | E11.3542 | Type 2 diabetes mellitus with proliferative diabetic retinopathy with combined traction retinal detachment and rhegmatogenous retinal detachment, left eye |
| dbm_inclusions | ICD10CM | E11.3543 | Type 2 diabetes mellitus with proliferative diabetic retinopathy with combined traction retinal detachment and rhegmatogenous retinal detachment, bilateral |
| dbm_inclusions | ICD10CM | E11.3549 | Type 2 diabetes mellitus with proliferative diabetic retinopathy with combined traction retinal detachment and rhegmatogenous retinal detachment, unspecified eye |
| dbm_inclusions | ICD10CM | E11.355 | Type 2 diabetes mellitus with stable proliferative diabetic retinopathy |
| dbm_inclusions | ICD10CM | E11.3551 | Type 2 diabetes mellitus with stable proliferative diabetic retinopathy, right eye |
| dbm_inclusions | ICD10CM | E11.3552 | Type 2 diabetes mellitus with stable proliferative diabetic retinopathy, left eye |
| dbm_inclusions | ICD10CM | E11.3553 | Type 2 diabetes mellitus with stable proliferative diabetic retinopathy, bilateral |
| dbm_inclusions | ICD10CM | E11.3559 | Type 2 diabetes mellitus with stable proliferative diabetic retinopathy, unspecified eye |
| dbm_inclusions | ICD10CM | E11.359 | Type 2 diabetes mellitus with proliferative diabetic retinopathy without macular edema |
| dbm_inclusions | ICD10CM | E11.3591 | Type 2 diabetes mellitus with proliferative diabetic retinopathy without macular edema, right eye |
| dbm_inclusions | ICD10CM | E11.3592 | Type 2 diabetes mellitus with proliferative diabetic retinopathy without macular edema, left eye |
| dbm_inclusions | ICD10CM | E11.3593 | Type 2 diabetes mellitus with proliferative diabetic retinopathy without macular edema, bilateral |
| dbm_inclusions | ICD10CM | E11.3599 | Type 2 diabetes mellitus with proliferative diabetic retinopathy without macular edema, unspecified eye |
| dbm_inclusions | ICD10CM | E11.36 | Type 2 diabetes mellitus with diabetic cataract |
| dbm_inclusions | ICD10CM | E11.37 | Type 2 diabetes mellitus with diabetic macular edema, resolved following treatment |
| dbm_inclusions | ICD10CM | E11.37X1 | Type 2 diabetes mellitus with diabetic macular edema, resolved following treatment, right eye |
| dbm_inclusions | ICD10CM | E11.37X2 | Type 2 diabetes mellitus with diabetic macular edema, resolved following treatment, left eye |
| dbm_inclusions | ICD10CM | E11.37X3 | Type 2 diabetes mellitus with diabetic macular edema, resolved following treatment, bilateral |
| dbm_inclusions | ICD10CM | E11.37X9 | Type 2 diabetes mellitus with diabetic macular edema, resolved following treatment, unspecified eye |
| dbm_inclusions | ICD10CM | E11.39 | Type 2 diabetes mellitus with other diabetic ophthalmic complication |
| dbm_inclusions | ICD10CM | E11.4 | Type 2 diabetes mellitus with neurological complications |
| dbm_inclusions | ICD10CM | E11.40 | Type 2 diabetes mellitus with diabetic neuropathy, unspecified |
| dbm_inclusions | ICD10CM | E11.41 | Type 2 diabetes mellitus with diabetic mononeuropathy |
| dbm_inclusions | ICD10CM | E11.42 | Type 2 diabetes mellitus with diabetic polyneuropathy |
| dbm_inclusions | ICD10CM | E11.43 | Type 2 diabetes mellitus with diabetic autonomic (poly)neuropathy |
| dbm_inclusions | ICD10CM | E11.44 | Type 2 diabetes mellitus with diabetic amyotrophy |
| dbm_inclusions | ICD10CM | E11.49 | Type 2 diabetes mellitus with other diabetic neurological complication |
| dbm_inclusions | ICD10CM | E11.5 | Type 2 diabetes mellitus with circulatory complications |
| dbm_inclusions | ICD10CM | E11.51 | Type 2 diabetes mellitus with diabetic peripheral angiopathy without gangrene |
| dbm_inclusions | ICD10CM | E11.52 | Type 2 diabetes mellitus with diabetic peripheral angiopathy with gangrene |
| dbm_inclusions | ICD10CM | E11.59 | Type 2 diabetes mellitus with other circulatory complications |
| dbm_inclusions | ICD10CM | E11.6 | Type 2 diabetes mellitus with other specified complications |
| dbm_inclusions | ICD10CM | E11.61 | Type 2 diabetes mellitus with diabetic arthropathy |
| dbm_inclusions | ICD10CM | E11.610 | Type 2 diabetes mellitus with diabetic neuropathic arthropathy |
| dbm_inclusions | ICD10CM | E11.618 | Type 2 diabetes mellitus with other diabetic arthropathy |
| dbm_inclusions | ICD10CM | E11.62 | Type 2 diabetes mellitus with skin complications |
| dbm_inclusions | ICD10CM | E11.620 | Type 2 diabetes mellitus with diabetic dermatitis |
| dbm_inclusions | ICD10CM | E11.621 | Type 2 diabetes mellitus with foot ulcer |
| dbm_inclusions | ICD10CM | E11.622 | Type 2 diabetes mellitus with other skin ulcer |
| dbm_inclusions | ICD10CM | E11.628 | Type 2 diabetes mellitus with other skin complications |
| dbm_inclusions | ICD10CM | E11.63 | Type 2 diabetes mellitus with oral complications |
| dbm_inclusions | ICD10CM | E11.630 | Type 2 diabetes mellitus with periodontal disease |
| dbm_inclusions | ICD10CM | E11.638 | Type 2 diabetes mellitus with other oral complications |
| dbm_inclusions | ICD10CM | E11.64 | Type 2 diabetes mellitus with hypoglycemia |
| dbm_inclusions | ICD10CM | E11.640 | Type 2 diabetes mellitus with hypoglycemia without coma |
| dbm_inclusions | ICD10CM | E11.641 | Type 2 diabetes mellitus with hypoglycemia with coma |
| dbm_inclusions | ICD10CM | E11.649 | Type 2 diabetes mellitus with hypoglycemia without coma |
| dbm_inclusions | ICD10CM | E11.65 | Type 2 diabetes mellitus with hyperglycemia |
| dbm_inclusions | ICD10CM | E11.69 | Type 2 diabetes mellitus with other specified complication |
| dbm_inclusions | ICD10CM | E11.8 | Type 2 diabetes mellitus with unspecified complications |
| dbm_inclusions | ICD10CM | E11.9 | Type 2 diabetes mellitus without complications |
| dbm_inclusions | ICD9CM | 250.10 | Diabetes with ketoacidosis, type II or unspecified type, not stated as uncontrolled |
| dbm_inclusions | ICD9CM | 250.12 | Diabetes with ketoacidosis, type II or unspecified type, uncontrolled |
| dbm_inclusions | ICD10CM | E11.1 | Type 2 diabetes mellitus with ketoacidosis |
| dbm_inclusions | ICD10CM | E11.10 | Type 2 diabetes mellitus with ketoacidosis without coma |
| dbm_inclusions | ICD10CM | E11.11 | Type 2 diabetes mellitus with ketoacidosis with coma |
| dbm_exclusions | ICD10CM | E09 | Drug or chemical induced diabetes mellitus |
| dbm_exclusions | ICD10CM | E09.0 | Drug or chemical induced diabetes mellitus with hyperosmolarity |
| dbm_exclusions | ICD10CM | E09.00 | Drug or chemical induced diabetes mellitus with hyperosmolarity without nonketotic hyperglycemic-hyperosmolar coma (NKHHC) |
| dbm_exclusions | ICD10CM | E09.01 | Drug or chemical induced diabetes mellitus with hyperosmolarity with coma |
| dbm_exclusions | ICD10CM | E09.1 | Drug or chemical induced diabetes mellitus with ketoacidosis |
| dbm_exclusions | ICD10CM | E09.10 | Drug or chemical induced diabetes mellitus with ketoacidosis without coma |
| dbm_exclusions | ICD10CM | E09.11 | Drug or chemical induced diabetes mellitus with ketoacidosis with coma |
| dbm_exclusions | ICD10CM | E09.2 | Drug or chemical induced diabetes mellitus with kidney complications |
| dbm_exclusions | ICD10CM | E09.21 | Drug or chemical induced diabetes mellitus with diabetic nephropathy |
| dbm_exclusions | ICD10CM | E09.22 | Drug or chemical induced diabetes mellitus with diabetic chronic kidney disease |
| dbm_exclusions | ICD10CM | E09.29 | Drug or chemical induced diabetes mellitus with other diabetic kidney complication |
| dbm_exclusions | ICD10CM | E09.3 | Drug or chemical induced diabetes mellitus with ophthalmic complications |
| dbm_exclusions | ICD10CM | E09.31 | Drug or chemical induced diabetes mellitus with unspecified diabetic retinopathy |
| dbm_exclusions | ICD10CM | E09.311 | Drug or chemical induced diabetes mellitus with unspecified diabetic retinopathy with macular edema |
| dbm_exclusions | ICD10CM | E09.319 | Drug or chemical induced diabetes mellitus with unspecified diabetic retinopathy without macular edema |
| dbm_exclusions | ICD10CM | E09.32 | Drug or chemical induced diabetes mellitus with mild nonproliferative diabetic retinopathy |
| dbm_exclusions | ICD10CM | E09.321 | Drug or chemical induced diabetes mellitus with mild nonproliferative diabetic retinopathy with macular edema |
| dbm_exclusions | ICD10CM | E09.3211 | Drug or chemical induced diabetes mellitus with mild nonproliferative diabetic retinopathy with macular edema, right eye |
| dbm_exclusions | ICD10CM | E09.3212 | Drug or chemical induced diabetes mellitus with mild nonproliferative diabetic retinopathy with macular edema, left eye |
| dbm_exclusions | ICD10CM | E09.3213 | Drug or chemical induced diabetes mellitus with mild nonproliferative diabetic retinopathy with macular edema, bilateral |
| dbm_exclusions | ICD10CM | E09.3219 | Drug or chemical induced diabetes mellitus with mild nonproliferative diabetic retinopathy with macular edema, unspecified eye |
| dbm_exclusions | ICD10CM | E09.329 | Drug or chemical induced diabetes mellitus with mild nonproliferative diabetic retinopathy without macular edema |
| dbm_exclusions | ICD10CM | E09.3291 | Drug or chemical induced diabetes mellitus with mild nonproliferative diabetic retinopathy without macular edema, right eye |
| dbm_exclusions | ICD10CM | E09.3292 | Drug or chemical induced diabetes mellitus with mild nonproliferative diabetic retinopathy without macular edema, left eye |
| dbm_exclusions | ICD10CM | E09.3293 | Drug or chemical induced diabetes mellitus with mild nonproliferative diabetic retinopathy without macular edema, bilateral |
| dbm_exclusions | ICD10CM | E09.3299 | Drug or chemical induced diabetes mellitus with mild nonproliferative diabetic retinopathy without macular edema, unspecified eye |
| dbm_exclusions | ICD10CM | E09.33 | Drug or chemical induced diabetes mellitus with moderate nonproliferative diabetic retinopathy |
| dbm_exclusions | ICD10CM | E09.331 | Drug or chemical induced diabetes mellitus with moderate nonproliferative diabetic retinopathy with macular edema |
| dbm_exclusions | ICD10CM | E09.3311 | Drug or chemical induced diabetes mellitus with moderate nonproliferative diabetic retinopathy with macular edema, right eye |
| dbm_exclusions | ICD10CM | E09.3312 | Drug or chemical induced diabetes mellitus with moderate nonproliferative diabetic retinopathy with macular edema, left eye |
| dbm_exclusions | ICD10CM | E09.3313 | Drug or chemical induced diabetes mellitus with moderate nonproliferative diabetic retinopathy with macular edema, bilateral |
| dbm_exclusions | ICD10CM | E09.3319 | Drug or chemical induced diabetes mellitus with moderate nonproliferative diabetic retinopathy with macular edema, unspecified eye |
| dbm_exclusions | ICD10CM | E09.339 | Drug or chemical induced diabetes mellitus with moderate nonproliferative diabetic retinopathy without macular edema |
| dbm_exclusions | ICD10CM | E09.3391 | Drug or chemical induced diabetes mellitus with moderate nonproliferative diabetic retinopathy without macular edema, right eye |
| dbm_exclusions | ICD10CM | E09.3392 | Drug or chemical induced diabetes mellitus with moderate nonproliferative diabetic retinopathy without macular edema, left eye |
| dbm_exclusions | ICD10CM | E09.3393 | Drug or chemical induced diabetes mellitus with moderate nonproliferative diabetic retinopathy without macular edema, bilateral |
| dbm_exclusions | ICD10CM | E09.3399 | Drug or chemical induced diabetes mellitus with moderate nonproliferative diabetic retinopathy without macular edema, unspecified eye |
| dbm_exclusions | ICD10CM | E09.34 | Drug or chemical induced diabetes mellitus with severe nonproliferative diabetic retinopathy |
| dbm_exclusions | ICD10CM | E09.341 | Drug or chemical induced diabetes mellitus with severe nonproliferative diabetic retinopathy with macular edema |
| dbm_exclusions | ICD10CM | E09.3411 | Drug or chemical induced diabetes mellitus with severe nonproliferative diabetic retinopathy with macular edema, right eye |
| dbm_exclusions | ICD10CM | E09.3412 | Drug or chemical induced diabetes mellitus with severe nonproliferative diabetic retinopathy with macular edema, left eye |
| dbm_exclusions | ICD10CM | E09.3413 | Drug or chemical induced diabetes mellitus with severe nonproliferative diabetic retinopathy with macular edema, bilateral |
| dbm_exclusions | ICD10CM | E09.3419 | Drug or chemical induced diabetes mellitus with severe nonproliferative diabetic retinopathy with macular edema, unspecified eye |
| dbm_exclusions | ICD10CM | E09.349 | Drug or chemical induced diabetes mellitus with severe nonproliferative diabetic retinopathy without macular edema |
| dbm_exclusions | ICD10CM | E09.3491 | Drug or chemical induced diabetes mellitus with severe nonproliferative diabetic retinopathy without macular edema, right eye |
| dbm_exclusions | ICD10CM | E09.3492 | Drug or chemical induced diabetes mellitus with severe nonproliferative diabetic retinopathy without macular edema, left eye |
| dbm_exclusions | ICD10CM | E09.3493 | Drug or chemical induced diabetes mellitus with severe nonproliferative diabetic retinopathy without macular edema, bilateral |
| dbm_exclusions | ICD10CM | E09.3499 | Drug or chemical induced diabetes mellitus with severe nonproliferative diabetic retinopathy without macular edema, unspecified eye |
| dbm_exclusions | ICD10CM | E09.35 | Drug or chemical induced diabetes mellitus with proliferative diabetic retinopathy |
| dbm_exclusions | ICD10CM | E09.351 | Drug or chemical induced diabetes mellitus with proliferative diabetic retinopathy with macular edema |
| dbm_exclusions | ICD10CM | E09.3511 | Drug or chemical induced diabetes mellitus with proliferative diabetic retinopathy with macular edema, right eye |
| dbm_exclusions | ICD10CM | E09.3512 | Drug or chemical induced diabetes mellitus with proliferative diabetic retinopathy with macular edema, left eye |
| dbm_exclusions | ICD10CM | E09.3513 | Drug or chemical induced diabetes mellitus with proliferative diabetic retinopathy with macular edema, bilateral |
| dbm_exclusions | ICD10CM | E09.3519 | Drug or chemical induced diabetes mellitus with proliferative diabetic retinopathy with macular edema, unspecified eye |
| dbm_exclusions | ICD10CM | E09.352 | Drug or chemical induced diabetes mellitus with proliferative diabetic retinopathy with traction retinal detachment involving the macula |
| dbm_exclusions | ICD10CM | E09.3521 | Drug or chemical induced diabetes mellitus with proliferative diabetic retinopathy with traction retinal detachment involving the macula, right eye |
| dbm_exclusions | ICD10CM | E09.3522 | Drug or chemical induced diabetes mellitus with proliferative diabetic retinopathy with traction retinal detachment involving the macula, left eye |
| dbm_exclusions | ICD10CM | E09.3523 | Drug or chemical induced diabetes mellitus with proliferative diabetic retinopathy with traction retinal detachment involving the macula, bilateral |
| dbm_exclusions | ICD10CM | E09.3529 | Drug or chemical induced diabetes mellitus with proliferative diabetic retinopathy with traction retinal detachment involving the macula, unspecified eye |
| dbm_exclusions | ICD10CM | E09.353 | Drug or chemical induced diabetes mellitus with proliferative diabetic retinopathy with traction retinal detachment not involving the macula |
| dbm_exclusions | ICD10CM | E09.3531 | Drug or chemical induced diabetes mellitus with proliferative diabetic retinopathy with traction retinal detachment not involving the macula, right eye |
| dbm_exclusions | ICD10CM | E09.3532 | Drug or chemical induced diabetes mellitus with proliferative diabetic retinopathy with traction retinal detachment not involving the macula, left eye |
| dbm_exclusions | ICD10CM | E09.3533 | Drug or chemical induced diabetes mellitus with proliferative diabetic retinopathy with traction retinal detachment not involving the macula, bilateral |
| dbm_exclusions | ICD10CM | E09.3539 | Drug or chemical induced diabetes mellitus with proliferative diabetic retinopathy with traction retinal detachment not involving the macula, unspecified eye |
| dbm_exclusions | ICD10CM | E09.354 | Drug or chemical induced diabetes mellitus with proliferative diabetic retinopathy with combined traction retinal detachment and rhegmatogenous retinal detachment |
| dbm_exclusions | ICD10CM | E09.3541 | Drug or chemical induced diabetes mellitus with proliferative diabetic retinopathy with combined traction retinal detachment and rhegmatogenous retinal detachment, right eye |
| dbm_exclusions | ICD10CM | E09.3542 | Drug or chemical induced diabetes mellitus with proliferative diabetic retinopathy with combined traction retinal detachment and rhegmatogenous retinal detachment, left eye |
| dbm_exclusions | ICD10CM | E09.3543 | Drug or chemical induced diabetes mellitus with proliferative diabetic retinopathy with combined traction retinal detachment and rhegmatogenous retinal detachment, bilateral |
| dbm_exclusions | ICD10CM | E09.3549 | Drug or chemical induced diabetes mellitus with proliferative diabetic retinopathy with combined traction retinal detachment and rhegmatogenous retinal detachment, unspecified eye |
| dbm_exclusions | ICD10CM | E09.355 | Drug or chemical induced diabetes mellitus with stable proliferative diabetic retinopathy |
| dbm_exclusions | ICD10CM | E09.3551 | Drug or chemical induced diabetes mellitus with stable proliferative diabetic retinopathy, right eye |
| dbm_exclusions | ICD10CM | E09.3552 | Drug or chemical induced diabetes mellitus with stable proliferative diabetic retinopathy, left eye |
| dbm_exclusions | ICD10CM | E09.3553 | Drug or chemical induced diabetes mellitus with stable proliferative diabetic retinopathy, bilateral |
| dbm_exclusions | ICD10CM | E09.3559 | Drug or chemical induced diabetes mellitus with stable proliferative diabetic retinopathy, unspecified eye |
| dbm_exclusions | ICD10CM | E09.359 | Drug or chemical induced diabetes mellitus with proliferative diabetic retinopathy without macular edema |
| dbm_exclusions | ICD10CM | E09.3591 | Drug or chemical induced diabetes mellitus with proliferative diabetic retinopathy without macular edema, right eye |
| dbm_exclusions | ICD10CM | E09.3592 | Drug or chemical induced diabetes mellitus with proliferative diabetic retinopathy without macular edema, left eye |
| dbm_exclusions | ICD10CM | E09.3593 | Drug or chemical induced diabetes mellitus with proliferative diabetic retinopathy without macular edema, bilateral |
| dbm_exclusions | ICD10CM | E09.3599 | Drug or chemical induced diabetes mellitus with proliferative diabetic retinopathy without macular edema, unspecified eye |
| dbm_exclusions | ICD10CM | E09.36 | Drug or chemical induced diabetes mellitus with diabetic cataract |
| dbm_exclusions | ICD10CM | E09.37 | Drug or chemical induced diabetes mellitus with diabetic macular edema, resolved following treatment |
| dbm_exclusions | ICD10CM | E09.37X1 | Drug or chemical induced diabetes mellitus with diabetic macular edema, resolved following treatment, right eye |
| dbm_exclusions | ICD10CM | E09.37X2 | Drug or chemical induced diabetes mellitus with diabetic macular edema, resolved following treatment, left eye |
| dbm_exclusions | ICD10CM | E09.37X3 | Drug or chemical induced diabetes mellitus with diabetic macular edema, resolved following treatment, bilateral |
| dbm_exclusions | ICD10CM | E09.37X9 | Drug or chemical induced diabetes mellitus with diabetic macular edema, resolved following treatment, unspecified eye |
| dbm_exclusions | ICD10CM | E09.39 | Drug or chemical induced diabetes mellitus with other diabetic ophthalmic complication |
| dbm_exclusions | ICD10CM | E09.4 | Drug or chemical induced diabetes mellitus with neurological complications |
| dbm_exclusions | ICD10CM | E09.40 | Drug or chemical induced diabetes mellitus with neurological complications with diabetic neuropathy, unspecified |
| dbm_exclusions | ICD10CM | E09.41 | Drug or chemical induced diabetes mellitus with neurological complications with diabetic mononeuropathy |
| dbm_exclusions | ICD10CM | E09.42 | Drug or chemical induced diabetes mellitus with neurological complications with diabetic polyneuropathy |
| dbm_exclusions | ICD10CM | E09.43 | Drug or chemical induced diabetes mellitus with neurological complications with diabetic autonomic (poly)neuropathy |
| dbm_exclusions | ICD10CM | E09.44 | Drug or chemical induced diabetes mellitus with neurological complications with diabetic amyotrophy |
| dbm_exclusions | ICD10CM | E09.49 | Drug or chemical induced diabetes mellitus with neurological complications with other diabetic neurological complication |
| dbm_exclusions | ICD10CM | E09.5 | Drug or chemical induced diabetes mellitus with circulatory complications |
| dbm_exclusions | ICD10CM | E09.51 | Drug or chemical induced diabetes mellitus with diabetic peripheral angiopathy without gangrene |
| dbm_exclusions | ICD10CM | E09.52 | Drug or chemical induced diabetes mellitus with diabetic peripheral angiopathy with gangrene |
| dbm_exclusions | ICD10CM | E09.59 | Drug or chemical induced diabetes mellitus with other circulatory complications |
| dbm_exclusions | ICD10CM | E09.6 | Drug or chemical induced diabetes mellitus with other specified complications |
| dbm_exclusions | ICD10CM | E09.61 | Drug or chemical induced diabetes mellitus with diabetic arthropathy |
| dbm_exclusions | ICD10CM | E09.610 | Drug or chemical induced diabetes mellitus with diabetic neuropathic arthropathy |
| dbm_exclusions | ICD10CM | E09.618 | Drug or chemical induced diabetes mellitus with other diabetic arthropathy |
| dbm_exclusions | ICD10CM | E09.62 | Drug or chemical induced diabetes mellitus with skin complications |
| dbm_exclusions | ICD10CM | E09.620 | Drug or chemical induced diabetes mellitus with diabetic dermatitis |
| dbm_exclusions | ICD10CM | E09.621 | Drug or chemical induced diabetes mellitus with foot ulcer |
| dbm_exclusions | ICD10CM | E09.622 | Drug or chemical induced diabetes mellitus with other skin ulcer |
| dbm_exclusions | ICD10CM | E09.628 | Drug or chemical induced diabetes mellitus with other skin complications |
| dbm_exclusions | ICD10CM | E09.63 | Drug or chemical induced diabetes mellitus with oral complications |
| dbm_exclusions | ICD10CM | E09.630 | Drug or chemical induced diabetes mellitus with periodontal disease |
| dbm_exclusions | ICD10CM | E09.638 | Drug or chemical induced diabetes mellitus with other oral complications |
| dbm_exclusions | ICD10CM | E09.64 | Drug or chemical induced diabetes mellitus with hypoglycemia |
| dbm_exclusions | ICD10CM | E09.640 | Drug or chemical induced diabetes mellitus with hypoglycemia without coma |
| dbm_exclusions | ICD10CM | E09.641 | Drug or chemical induced diabetes mellitus with hypoglycemia with coma |
| dbm_exclusions | ICD10CM | E09.649 | Drug or chemical induced diabetes mellitus with hypoglycemia without coma |
| dbm_exclusions | ICD10CM | E09.65 | Drug or chemical induced diabetes mellitus with hyperglycemia |
| dbm_exclusions | ICD10CM | E09.69 | Drug or chemical induced diabetes mellitus with other specified complication |
| dbm_exclusions | ICD10CM | E09.8 | Drug or chemical induced diabetes mellitus with unspecified complications |
| dbm_exclusions | ICD10CM | E09.9 | Drug or chemical induced diabetes mellitus without complications |
| dbm_exclusions | ICD10CM | E13 | Other specified diabetes mellitus |
| dbm_exclusions | ICD10CM | E13.0 | Other specified diabetes mellitus with hyperosmolarity |
| dbm_exclusions | ICD10CM | E13.00 | Other specified diabetes mellitus with hyperosmolarity without nonketotic hyperglycemic-hyperosmolar coma (NKHHC) |
| dbm_exclusions | ICD10CM | E13.01 | Other specified diabetes mellitus with hyperosmolarity with coma |
| dbm_exclusions | ICD10CM | E13.1 | Other specified diabetes mellitus with ketoacidosis |
| dbm_exclusions | ICD10CM | E13.10 | Other specified diabetes mellitus with ketoacidosis without coma |
| dbm_exclusions | ICD10CM | E13.11 | Other specified diabetes mellitus with ketoacidosis with coma |
| dbm_exclusions | ICD10CM | E13.2 | Other specified diabetes mellitus with kidney complications |
| dbm_exclusions | ICD10CM | E13.21 | Other specified diabetes mellitus with diabetic nephropathy |
| dbm_exclusions | ICD10CM | E13.22 | Other specified diabetes mellitus with diabetic chronic kidney disease |
| dbm_exclusions | ICD10CM | E13.29 | Other specified diabetes mellitus with other diabetic kidney complication |
| dbm_exclusions | ICD10CM | E13.3 | Other specified diabetes mellitus with ophthalmic complications |
| dbm_exclusions | ICD10CM | E13.31 | Other specified diabetes mellitus with unspecified diabetic retinopathy |
| dbm_exclusions | ICD10CM | E13.311 | Other specified diabetes mellitus with unspecified diabetic retinopathy with macular edema |
| dbm_exclusions | ICD10CM | E13.319 | Other specified diabetes mellitus with unspecified diabetic retinopathy without macular edema |
| dbm_exclusions | ICD10CM | E13.32 | Other specified diabetes mellitus with mild nonproliferative diabetic retinopathy |
| dbm_exclusions | ICD10CM | E13.321 | Other specified diabetes mellitus with mild nonproliferative diabetic retinopathy with macular edema |
| dbm_exclusions | ICD10CM | E13.3211 | Other specified diabetes mellitus with mild nonproliferative diabetic retinopathy with macular edema, right eye |
| dbm_exclusions | ICD10CM | E13.3212 | Other specified diabetes mellitus with mild nonproliferative diabetic retinopathy with macular edema, left eye |
| dbm_exclusions | ICD10CM | E13.3213 | Other specified diabetes mellitus with mild nonproliferative diabetic retinopathy with macular edema, bilateral |
| dbm_exclusions | ICD10CM | E13.3219 | Other specified diabetes mellitus with mild nonproliferative diabetic retinopathy with macular edema, unspecified eye |
| dbm_exclusions | ICD10CM | E13.329 | Other specified diabetes mellitus with mild nonproliferative diabetic retinopathy without macular edema |
| dbm_exclusions | ICD10CM | E13.3291 | Other specified diabetes mellitus with mild nonproliferative diabetic retinopathy without macular edema, right eye |
| dbm_exclusions | ICD10CM | E13.3292 | Other specified diabetes mellitus with mild nonproliferative diabetic retinopathy without macular edema, left eye |
| dbm_exclusions | ICD10CM | E13.3293 | Other specified diabetes mellitus with mild nonproliferative diabetic retinopathy without macular edema, bilateral |
| dbm_exclusions | ICD10CM | E13.3299 | Other specified diabetes mellitus with mild nonproliferative diabetic retinopathy without macular edema, unspecified eye |
| dbm_exclusions | ICD10CM | E13.33 | Other specified diabetes mellitus with moderate nonproliferative diabetic retinopathy |
| dbm_exclusions | ICD10CM | E13.331 | Other specified diabetes mellitus with moderate nonproliferative diabetic retinopathy with macular edema |
| dbm_exclusions | ICD10CM | E13.3311 | Other specified diabetes mellitus with moderate nonproliferative diabetic retinopathy with macular edema, right eye |
| dbm_exclusions | ICD10CM | E13.3312 | Other specified diabetes mellitus with moderate nonproliferative diabetic retinopathy with macular edema, left eye |
| dbm_exclusions | ICD10CM | E13.3313 | Other specified diabetes mellitus with moderate nonproliferative diabetic retinopathy with macular edema, bilateral |
| dbm_exclusions | ICD10CM | E13.3319 | Other specified diabetes mellitus with moderate nonproliferative diabetic retinopathy with macular edema, unspecified eye |
| dbm_exclusions | ICD10CM | E13.339 | Other specified diabetes mellitus with moderate nonproliferative diabetic retinopathy without macular edema |
| dbm_exclusions | ICD10CM | E13.3391 | Other specified diabetes mellitus with moderate nonproliferative diabetic retinopathy without macular edema, right eye |
| dbm_exclusions | ICD10CM | E13.3392 | Other specified diabetes mellitus with moderate nonproliferative diabetic retinopathy without macular edema, left eye |
| dbm_exclusions | ICD10CM | E13.3393 | Other specified diabetes mellitus with moderate nonproliferative diabetic retinopathy without macular edema, bilateral |
| dbm_exclusions | ICD10CM | E13.3399 | Other specified diabetes mellitus with moderate nonproliferative diabetic retinopathy without macular edema, unspecified eye |
| dbm_exclusions | ICD10CM | E13.34 | Other specified diabetes mellitus with severe nonproliferative diabetic retinopathy |
| dbm_exclusions | ICD10CM | E13.341 | Other specified diabetes mellitus with severe nonproliferative diabetic retinopathy with macular edema |
| dbm_exclusions | ICD10CM | E13.3411 | Other specified diabetes mellitus with severe nonproliferative diabetic retinopathy with macular edema, right eye |
| dbm_exclusions | ICD10CM | E13.3412 | Other specified diabetes mellitus with severe nonproliferative diabetic retinopathy with macular edema, left eye |
| dbm_exclusions | ICD10CM | E13.3413 | Other specified diabetes mellitus with severe nonproliferative diabetic retinopathy with macular edema, bilateral |
| dbm_exclusions | ICD10CM | E13.3419 | Other specified diabetes mellitus with severe nonproliferative diabetic retinopathy with macular edema, unspecified eye |
| dbm_exclusions | ICD10CM | E13.349 | Other specified diabetes mellitus with severe nonproliferative diabetic retinopathy without macular edema |
| dbm_exclusions | ICD10CM | E13.3491 | Other specified diabetes mellitus with severe nonproliferative diabetic retinopathy without macular edema, right eye |
| dbm_exclusions | ICD10CM | E13.3492 | Other specified diabetes mellitus with severe nonproliferative diabetic retinopathy without macular edema, left eye |
| dbm_exclusions | ICD10CM | E13.3493 | Other specified diabetes mellitus with severe nonproliferative diabetic retinopathy without macular edema, bilateral |
| dbm_exclusions | ICD10CM | E13.3499 | Other specified diabetes mellitus with severe nonproliferative diabetic retinopathy without macular edema, unspecified eye |
| dbm_exclusions | ICD10CM | E13.35 | Other specified diabetes mellitus with proliferative diabetic retinopathy |
| dbm_exclusions | ICD10CM | E13.351 | Other specified diabetes mellitus with proliferative diabetic retinopathy with macular edema |
| dbm_exclusions | ICD10CM | E13.3511 | Other specified diabetes mellitus with proliferative diabetic retinopathy with macular edema, right eye |
| dbm_exclusions | ICD10CM | E13.3512 | Other specified diabetes mellitus with proliferative diabetic retinopathy with macular edema, left eye |
| dbm_exclusions | ICD10CM | E13.3513 | Other specified diabetes mellitus with proliferative diabetic retinopathy with macular edema, bilateral |
| dbm_exclusions | ICD10CM | E13.3519 | Other specified diabetes mellitus with proliferative diabetic retinopathy with macular edema, unspecified eye |
| dbm_exclusions | ICD10CM | E13.352 | Other specified diabetes mellitus with proliferative diabetic retinopathy with traction retinal detachment involving the macula |
| dbm_exclusions | ICD10CM | E13.3521 | Other specified diabetes mellitus with proliferative diabetic retinopathy with traction retinal detachment involving the macula, right eye |
| dbm_exclusions | ICD10CM | E13.3522 | Other specified diabetes mellitus with proliferative diabetic retinopathy with traction retinal detachment involving the macula, left eye |
| dbm_exclusions | ICD10CM | E13.3523 | Other specified diabetes mellitus with proliferative diabetic retinopathy with traction retinal detachment involving the macula, bilateral |
| dbm_exclusions | ICD10CM | E13.3529 | Other specified diabetes mellitus with proliferative diabetic retinopathy with traction retinal detachment involving the macula, unspecified eye |
| dbm_exclusions | ICD10CM | E13.353 | Other specified diabetes mellitus with proliferative diabetic retinopathy with traction retinal detachment not involving the macula |
| dbm_exclusions | ICD10CM | E13.3531 | Other specified diabetes mellitus with proliferative diabetic retinopathy with traction retinal detachment not involving the macula, right eye |
| dbm_exclusions | ICD10CM | E13.3532 | Other specified diabetes mellitus with proliferative diabetic retinopathy with traction retinal detachment not involving the macula, left eye |
| dbm_exclusions | ICD10CM | E13.3533 | Other specified diabetes mellitus with proliferative diabetic retinopathy with traction retinal detachment not involving the macula, bilateral |
| dbm_exclusions | ICD10CM | E13.3539 | Other specified diabetes mellitus with proliferative diabetic retinopathy with traction retinal detachment not involving the macula, unspecified eye |
| dbm_exclusions | ICD10CM | E13.354 | Other specified diabetes mellitus with proliferative diabetic retinopathy with combined traction retinal detachment and rhegmatogenous retinal detachment |
| dbm_exclusions | ICD10CM | E13.3541 | Other specified diabetes mellitus with proliferative diabetic retinopathy with combined traction retinal detachment and rhegmatogenous retinal detachment, right eye |
| dbm_exclusions | ICD10CM | E13.3542 | Other specified diabetes mellitus with proliferative diabetic retinopathy with combined traction retinal detachment and rhegmatogenous retinal detachment, left eye |
| dbm_exclusions | ICD10CM | E13.3543 | Other specified diabetes mellitus with proliferative diabetic retinopathy with combined traction retinal detachment and rhegmatogenous retinal detachment, bilateral |
| dbm_exclusions | ICD10CM | E13.3549 | Other specified diabetes mellitus with proliferative diabetic retinopathy with combined traction retinal detachment and rhegmatogenous retinal detachment, unspecified eye |
| dbm_exclusions | ICD10CM | E13.355 | Other specified diabetes mellitus with stable proliferative diabetic retinopathy |
| dbm_exclusions | ICD10CM | E13.3551 | Other specified diabetes mellitus with stable proliferative diabetic retinopathy, right eye |
| dbm_exclusions | ICD10CM | E13.3552 | Other specified diabetes mellitus with stable proliferative diabetic retinopathy, left eye |
| dbm_exclusions | ICD10CM | E13.3553 | Other specified diabetes mellitus with stable proliferative diabetic retinopathy, bilateral |
| dbm_exclusions | ICD10CM | E13.3559 | Other specified diabetes mellitus with stable proliferative diabetic retinopathy, unspecified eye |
| dbm_exclusions | ICD10CM | E13.359 | Other specified diabetes mellitus with proliferative diabetic retinopathy without macular edema |
| dbm_exclusions | ICD10CM | E13.3591 | Other specified diabetes mellitus with proliferative diabetic retinopathy without macular edema, right eye |
| dbm_exclusions | ICD10CM | E13.3592 | Other specified diabetes mellitus with proliferative diabetic retinopathy without macular edema, left eye |
| dbm_exclusions | ICD10CM | E13.3593 | Other specified diabetes mellitus with proliferative diabetic retinopathy without macular edema, bilateral |
| dbm_exclusions | ICD10CM | E13.3599 | Other specified diabetes mellitus with proliferative diabetic retinopathy without macular edema, unspecified eye |
| dbm_exclusions | ICD10CM | E13.36 | Other specified diabetes mellitus with diabetic cataract |
| dbm_exclusions | ICD10CM | E13.37 | Other specified diabetes mellitus with diabetic macular edema, resolved following treatment |
| dbm_exclusions | ICD10CM | E13.37X1 | Other specified diabetes mellitus with diabetic macular edema, resolved following treatment, right eye |
| dbm_exclusions | ICD10CM | E13.37X2 | Other specified diabetes mellitus with diabetic macular edema, resolved following treatment, left eye |
| dbm_exclusions | ICD10CM | E13.37X3 | Other specified diabetes mellitus with diabetic macular edema, resolved following treatment, bilateral |
| dbm_exclusions | ICD10CM | E13.37X9 | Other specified diabetes mellitus with diabetic macular edema, resolved following treatment, unspecified eye |
| dbm_exclusions | ICD10CM | E13.39 | Other specified diabetes mellitus with other diabetic ophthalmic complication |
| dbm_exclusions | ICD10CM | E13.4 | Other specified diabetes mellitus with neurological complications |
| dbm_exclusions | ICD10CM | E13.40 | Other specified diabetes mellitus with diabetic neuropathy, unspecified |
| dbm_exclusions | ICD10CM | E13.41 | Other specified diabetes mellitus with diabetic mononeuropathy |
| dbm_exclusions | ICD10CM | E13.42 | Other specified diabetes mellitus with diabetic polyneuropathy |
| dbm_exclusions | ICD10CM | E13.43 | Other specified diabetes mellitus with diabetic autonomic (poly)neuropathy |
| dbm_exclusions | ICD10CM | E13.44 | Other specified diabetes mellitus with diabetic amyotrophy |
| dbm_exclusions | ICD10CM | E13.49 | Other specified diabetes mellitus with other diabetic neurological complication |
| dbm_exclusions | ICD10CM | E13.5 | Other specified diabetes mellitus with circulatory complications |
| dbm_exclusions | ICD10CM | E13.51 | Other specified diabetes mellitus with diabetic peripheral angiopathy without gangrene |
| dbm_exclusions | ICD10CM | E13.52 | Other specified diabetes mellitus with diabetic peripheral angiopathy with gangrene |
| dbm_exclusions | ICD10CM | E13.59 | Other specified diabetes mellitus with other circulatory complications |
| dbm_exclusions | ICD10CM | E13.6 | Other specified diabetes mellitus with other specified complications |
| dbm_exclusions | ICD10CM | E13.61 | Other specified diabetes mellitus with diabetic arthropathy |
| dbm_exclusions | ICD10CM | E13.610 | Other specified diabetes mellitus with diabetic neuropathic arthropathy |
| dbm_exclusions | ICD10CM | E13.618 | Other specified diabetes mellitus with other diabetic arthropathy |
| dbm_exclusions | ICD10CM | E13.62 | Other specified diabetes mellitus with skin complications |
| dbm_exclusions | ICD10CM | E13.620 | Other specified diabetes mellitus with diabetic dermatitis |
| dbm_exclusions | ICD10CM | E13.621 | Other specified diabetes mellitus with foot ulcer |
| dbm_exclusions | ICD10CM | E13.622 | Other specified diabetes mellitus with other skin ulcer |
| dbm_exclusions | ICD10CM | E13.628 | Other specified diabetes mellitus with other skin complications |
| dbm_exclusions | ICD10CM | E13.63 | Other specified diabetes mellitus with oral complications |
| dbm_exclusions | ICD10CM | E13.630 | Other specified diabetes mellitus with periodontal disease |
| dbm_exclusions | ICD10CM | E13.638 | Other specified diabetes mellitus with other oral complications |
| dbm_exclusions | ICD10CM | E13.64 | Other specified diabetes mellitus with hypoglycemia |
| dbm_exclusions | ICD10CM | E13.640 | Other specified diabetes mellitus with hypoglycemia without coma |
| dbm_exclusions | ICD10CM | E13.641 | Other specified diabetes mellitus with hypoglycemia with coma |
| dbm_exclusions | ICD10CM | E13.649 | Other specified diabetes mellitus with hypoglycemia without coma |
| dbm_exclusions | ICD10CM | E13.65 | Other specified diabetes mellitus with hyperglycemia |
| dbm_exclusions | ICD10CM | E13.69 | Other specified diabetes mellitus with other specified complication |
| dbm_exclusions | ICD10CM | E13.8 | Other specified diabetes mellitus with unspecified complications |
| dbm_exclusions | ICD10CM | E13.9 | Other specified diabetes mellitus without complications |
| dbm_exclusions | ICD10CM | E08 | Diabetes mellitus due to underlying condition |
| dbm_exclusions | ICD10CM | E08.0 | Diabetes mellitus due to underlying condition with hyperosmolarity |
| dbm_exclusions | ICD10CM | E08.00 | Diabetes mellitus due to underlying condition with hyperosmolarity without nonketotic hyperglycemic-hyperosmolar coma (NKHHC) |
| dbm_exclusions | ICD10CM | E08.01 | Diabetes mellitus due to underlying condition with hyperosmolarity with coma |
| dbm_exclusions | ICD10CM | E08.1 | Diabetes mellitus due to underlying condition with ketoacidosis |
| dbm_exclusions | ICD10CM | E08.10 | Diabetes mellitus due to underlying condition with ketoacidosis without coma |
| dbm_exclusions | ICD10CM | E08.11 | Diabetes mellitus due to underlying condition with ketoacidosis with coma |
| dbm_exclusions | ICD10CM | E08.2 | Diabetes mellitus due to underlying condition with kidney complications |
| dbm_exclusions | ICD10CM | E08.21 | Diabetes mellitus due to underlying condition with diabetic nephropathy |
| dbm_exclusions | ICD10CM | E08.22 | Diabetes mellitus due to underlying condition with diabetic chronic kidney disease |
| dbm_exclusions | ICD10CM | E08.29 | Diabetes mellitus due to underlying condition with other diabetic kidney complication |
| dbm_exclusions | ICD10CM | E08.3 | Diabetes mellitus due to underlying condition with ophthalmic complications |
| dbm_exclusions | ICD10CM | E08.31 | Diabetes mellitus due to underlying condition with unspecified diabetic retinopathy |
| dbm_exclusions | ICD10CM | E08.311 | Diabetes mellitus due to underlying condition with unspecified diabetic retinopathy with macular edema |
| dbm_exclusions | ICD10CM | E08.319 | Diabetes mellitus due to underlying condition with unspecified diabetic retinopathy without macular edema |
| dbm_exclusions | ICD10CM | E08.32 | Diabetes mellitus due to underlying condition with mild nonproliferative diabetic retinopathy |
| dbm_exclusions | ICD10CM | E08.321 | Diabetes mellitus due to underlying condition with mild nonproliferative diabetic retinopathy with macular edema |
| dbm_exclusions | ICD10CM | E08.3211 | Diabetes mellitus due to underlying condition with mild nonproliferative diabetic retinopathy with macular edema, right eye |
| dbm_exclusions | ICD10CM | E08.3212 | Diabetes mellitus due to underlying condition with mild nonproliferative diabetic retinopathy with macular edema, left eye |
| dbm_exclusions | ICD10CM | E08.3213 | Diabetes mellitus due to underlying condition with mild nonproliferative diabetic retinopathy with macular edema, bilateral |
| dbm_exclusions | ICD10CM | E08.3219 | Diabetes mellitus due to underlying condition with mild nonproliferative diabetic retinopathy with macular edema, unspecified eye |
| dbm_exclusions | ICD10CM | E08.329 | Diabetes mellitus due to underlying condition with mild nonproliferative diabetic retinopathy without macular edema |
| dbm_exclusions | ICD10CM | E08.3291 | Diabetes mellitus due to underlying condition with mild nonproliferative diabetic retinopathy without macular edema, right eye |
| dbm_exclusions | ICD10CM | E08.3292 | Diabetes mellitus due to underlying condition with mild nonproliferative diabetic retinopathy without macular edema, left eye |
| dbm_exclusions | ICD10CM | E08.3293 | Diabetes mellitus due to underlying condition with mild nonproliferative diabetic retinopathy without macular edema, bilateral |
| dbm_exclusions | ICD10CM | E08.3299 | Diabetes mellitus due to underlying condition with mild nonproliferative diabetic retinopathy without macular edema, unspecified eye |
| dbm_exclusions | ICD10CM | E08.33 | Diabetes mellitus due to underlying condition with moderate nonproliferative diabetic retinopathy |
| dbm_exclusions | ICD10CM | E08.331 | Diabetes mellitus due to underlying condition with moderate nonproliferative diabetic retinopathy with macular edema |
| dbm_exclusions | ICD10CM | E08.3311 | Diabetes mellitus due to underlying condition with moderate nonproliferative diabetic retinopathy with macular edema, right eye |
| dbm_exclusions | ICD10CM | E08.3312 | Diabetes mellitus due to underlying condition with moderate nonproliferative diabetic retinopathy with macular edema, left eye |
| dbm_exclusions | ICD10CM | E08.3313 | Diabetes mellitus due to underlying condition with moderate nonproliferative diabetic retinopathy with macular edema, bilateral |
| dbm_exclusions | ICD10CM | E08.3319 | Diabetes mellitus due to underlying condition with moderate nonproliferative diabetic retinopathy with macular edema, unspecified eye |
| dbm_exclusions | ICD10CM | E08.339 | Diabetes mellitus due to underlying condition with moderate nonproliferative diabetic retinopathy without macular edema |
| dbm_exclusions | ICD10CM | E08.3391 | Diabetes mellitus due to underlying condition with moderate nonproliferative diabetic retinopathy without macular edema, right eye |
| dbm_exclusions | ICD10CM | E08.3392 | Diabetes mellitus due to underlying condition with moderate nonproliferative diabetic retinopathy without macular edema, left eye |
| dbm_exclusions | ICD10CM | E08.3393 | Diabetes mellitus due to underlying condition with moderate nonproliferative diabetic retinopathy without macular edema, bilateral |
| dbm_exclusions | ICD10CM | E08.3399 | Diabetes mellitus due to underlying condition with moderate nonproliferative diabetic retinopathy without macular edema, unspecified eye |
| dbm_exclusions | ICD10CM | E08.34 | Diabetes mellitus due to underlying condition with severe nonproliferative diabetic retinopathy |
| dbm_exclusions | ICD10CM | E08.341 | Diabetes mellitus due to underlying condition with severe nonproliferative diabetic retinopathy with macular edema |
| dbm_exclusions | ICD10CM | E08.3411 | Diabetes mellitus due to underlying condition with severe nonproliferative diabetic retinopathy with macular edema, right eye |
| dbm_exclusions | ICD10CM | E08.3412 | Diabetes mellitus due to underlying condition with severe nonproliferative diabetic retinopathy with macular edema, left eye |
| dbm_exclusions | ICD10CM | E08.3413 | Diabetes mellitus due to underlying condition with severe nonproliferative diabetic retinopathy with macular edema, bilateral |
| dbm_exclusions | ICD10CM | E08.3419 | Diabetes mellitus due to underlying condition with severe nonproliferative diabetic retinopathy with macular edema, unspecified eye |
| dbm_exclusions | ICD10CM | E08.349 | Diabetes mellitus due to underlying condition with severe nonproliferative diabetic retinopathy without macular edema |
| dbm_exclusions | ICD10CM | E08.3491 | Diabetes mellitus due to underlying condition with severe nonproliferative diabetic retinopathy without macular edema, right eye |
| dbm_exclusions | ICD10CM | E08.3492 | Diabetes mellitus due to underlying condition with severe nonproliferative diabetic retinopathy without macular edema, left eye |
| dbm_exclusions | ICD10CM | E08.3493 | Diabetes mellitus due to underlying condition with severe nonproliferative diabetic retinopathy without macular edema, bilateral |
| dbm_exclusions | ICD10CM | E08.3499 | Diabetes mellitus due to underlying condition with severe nonproliferative diabetic retinopathy without macular edema, unspecified eye |
| dbm_exclusions | ICD10CM | E08.35 | Diabetes mellitus due to underlying condition with proliferative diabetic retinopathy |
| dbm_exclusions | ICD10CM | E08.351 | Diabetes mellitus due to underlying condition with proliferative diabetic retinopathy with macular edema |
| dbm_exclusions | ICD10CM | E08.3511 | Diabetes mellitus due to underlying condition with proliferative diabetic retinopathy with macular edema, right eye |
| dbm_exclusions | ICD10CM | E08.3512 | Diabetes mellitus due to underlying condition with proliferative diabetic retinopathy with macular edema, left eye |
| dbm_exclusions | ICD10CM | E08.3513 | Diabetes mellitus due to underlying condition with proliferative diabetic retinopathy with macular edema, bilateral |
| dbm_exclusions | ICD10CM | E08.3519 | Diabetes mellitus due to underlying condition with proliferative diabetic retinopathy with macular edema, unspecified eye |
| dbm_exclusions | ICD10CM | E08.352 | Diabetes mellitus due to underlying condition with proliferative diabetic retinopathy with traction retinal detachment involving the macula |
| dbm_exclusions | ICD10CM | E08.3521 | Diabetes mellitus due to underlying condition with proliferative diabetic retinopathy with traction retinal detachment involving the macula, right eye |
| dbm_exclusions | ICD10CM | E08.3522 | Diabetes mellitus due to underlying condition with proliferative diabetic retinopathy with traction retinal detachment involving the macula, left eye |
| dbm_exclusions | ICD10CM | E08.3523 | Diabetes mellitus due to underlying condition with proliferative diabetic retinopathy with traction retinal detachment involving the macula, bilateral |
| dbm_exclusions | ICD10CM | E08.3529 | Diabetes mellitus due to underlying condition with proliferative diabetic retinopathy with traction retinal detachment involving the macula, unspecified eye |
| dbm_exclusions | ICD10CM | E08.353 | Diabetes mellitus due to underlying condition with proliferative diabetic retinopathy with traction retinal detachment not involving the macula |
| dbm_exclusions | ICD10CM | E08.3531 | Diabetes mellitus due to underlying condition with proliferative diabetic retinopathy with traction retinal detachment not involving the macula, right eye |
| dbm_exclusions | ICD10CM | E08.3532 | Diabetes mellitus due to underlying condition with proliferative diabetic retinopathy with traction retinal detachment not involving the macula, left eye |
| dbm_exclusions | ICD10CM | E08.3533 | Diabetes mellitus due to underlying condition with proliferative diabetic retinopathy with traction retinal detachment not involving the macula, bilateral |
| dbm_exclusions | ICD10CM | E08.3539 | Diabetes mellitus due to underlying condition with proliferative diabetic retinopathy with traction retinal detachment not involving the macula, unspecified eye |
| dbm_exclusions | ICD10CM | E08.354 | Diabetes mellitus due to underlying condition with proliferative diabetic retinopathy with combined traction retinal detachment and rhegmatogenous retinal detachment |
| dbm_exclusions | ICD10CM | E08.3541 | Diabetes mellitus due to underlying condition with proliferative diabetic retinopathy with combined traction retinal detachment and rhegmatogenous retinal detachment, right eye |
| dbm_exclusions | ICD10CM | E08.3542 | Diabetes mellitus due to underlying condition with proliferative diabetic retinopathy with combined traction retinal detachment and rhegmatogenous retinal detachment, left eye |
| dbm_exclusions | ICD10CM | E08.3543 | Diabetes mellitus due to underlying condition with proliferative diabetic retinopathy with combined traction retinal detachment and rhegmatogenous retinal detachment, bilateral |
| dbm_exclusions | ICD10CM | E08.3549 | Diabetes mellitus due to underlying condition with proliferative diabetic retinopathy with combined traction retinal detachment and rhegmatogenous retinal detachment, unspecified eye |
| dbm_exclusions | ICD10CM | E08.355 | Diabetes mellitus due to underlying condition with stable proliferative diabetic retinopathy |
| dbm_exclusions | ICD10CM | E08.3551 | Diabetes mellitus due to underlying condition with stable proliferative diabetic retinopathy, right eye |
| dbm_exclusions | ICD10CM | E08.3552 | Diabetes mellitus due to underlying condition with stable proliferative diabetic retinopathy, left eye |
| dbm_exclusions | ICD10CM | E08.3553 | Diabetes mellitus due to underlying condition with stable proliferative diabetic retinopathy, bilateral |
| dbm_exclusions | ICD10CM | E08.3559 | Diabetes mellitus due to underlying condition with stable proliferative diabetic retinopathy, unspecified eye |
| dbm_exclusions | ICD10CM | E08.359 | Diabetes mellitus due to underlying condition with proliferative diabetic retinopathy without macular edema |
| dbm_exclusions | ICD10CM | E08.3591 | Diabetes mellitus due to underlying condition with proliferative diabetic retinopathy without macular edema, right eye |
| dbm_exclusions | ICD10CM | E08.3592 | Diabetes mellitus due to underlying condition with proliferative diabetic retinopathy without macular edema, left eye |
| dbm_exclusions | ICD10CM | E08.3593 | Diabetes mellitus due to underlying condition with proliferative diabetic retinopathy without macular edema, bilateral |
| dbm_exclusions | ICD10CM | E08.3599 | Diabetes mellitus due to underlying condition with proliferative diabetic retinopathy without macular edema, unspecified eye |
| dbm_exclusions | ICD10CM | E08.36 | Diabetes mellitus due to underlying condition with diabetic cataract |
| dbm_exclusions | ICD10CM | E08.37 | Diabetes mellitus due to underlying condition with diabetic macular edema, resolved following treatment |
| dbm_exclusions | ICD10CM | E08.37X1 | Diabetes mellitus due to underlying condition with diabetic macular edema, resolved following treatment, right eye |
| dbm_exclusions | ICD10CM | E08.37X2 | Diabetes mellitus due to underlying condition with diabetic macular edema, resolved following treatment, left eye |
| dbm_exclusions | ICD10CM | E08.37X3 | Diabetes mellitus due to underlying condition with diabetic macular edema, resolved following treatment, bilateral |
| dbm_exclusions | ICD10CM | E08.37X9 | Diabetes mellitus due to underlying condition with diabetic macular edema, resolved following treatment, unspecified eye |
| dbm_exclusions | ICD10CM | E08.39 | Diabetes mellitus due to underlying condition with other diabetic ophthalmic complication |
| dbm_exclusions | ICD10CM | E08.4 | Diabetes mellitus due to underlying condition with neurological complications |
| dbm_exclusions | ICD10CM | E08.40 | Diabetes mellitus due to underlying condition with diabetic neuropathy, unspecified |
| dbm_exclusions | ICD10CM | E08.41 | Diabetes mellitus due to underlying condition with diabetic mononeuropathy |
| dbm_exclusions | ICD10CM | E08.42 | Diabetes mellitus due to underlying condition with diabetic polyneuropathy |
| dbm_exclusions | ICD10CM | E08.43 | Diabetes mellitus due to underlying condition with diabetic autonomic (poly)neuropathy |
| dbm_exclusions | ICD10CM | E08.44 | Diabetes mellitus due to underlying condition with diabetic amyotrophy |
| dbm_exclusions | ICD10CM | E08.49 | Diabetes mellitus due to underlying condition with other diabetic neurological complication |
| dbm_exclusions | ICD10CM | E08.5 | Diabetes mellitus due to underlying condition with circulatory complications |
| dbm_exclusions | ICD10CM | E08.51 | Diabetes mellitus due to underlying condition with diabetic peripheral angiopathy without gangrene |
| dbm_exclusions | ICD10CM | E08.52 | Diabetes mellitus due to underlying condition with diabetic peripheral angiopathy with gangrene |
| dbm_exclusions | ICD10CM | E08.59 | Diabetes mellitus due to underlying condition with other circulatory complications |
| dbm_exclusions | ICD10CM | E08.6 | Diabetes mellitus due to underlying condition with other specified complications |
| dbm_exclusions | ICD10CM | E08.61 | Diabetes mellitus due to underlying condition with diabetic arthropathy |
| dbm_exclusions | ICD10CM | E08.610 | Diabetes mellitus due to underlying condition with diabetic neuropathic arthropathy |
| dbm_exclusions | ICD10CM | E08.618 | Diabetes mellitus due to underlying condition with other diabetic arthropathy |
| dbm_exclusions | ICD10CM | E08.62 | Diabetes mellitus due to underlying condition with skin complications |
| dbm_exclusions | ICD10CM | E08.620 | Diabetes mellitus due to underlying condition with diabetic dermatitis |
| dbm_exclusions | ICD10CM | E08.621 | Diabetes mellitus due to underlying condition with foot ulcer |
| dbm_exclusions | ICD10CM | E08.622 | Diabetes mellitus due to underlying condition with other skin ulcer |
| dbm_exclusions | ICD10CM | E08.628 | Diabetes mellitus due to underlying condition with other skin complications |
| dbm_exclusions | ICD10CM | E08.63 | Diabetes mellitus due to underlying condition with oral complications |
| dbm_exclusions | ICD10CM | E08.630 | Diabetes mellitus due to underlying condition with periodontal disease |
| dbm_exclusions | ICD10CM | E08.638 | Diabetes mellitus due to underlying condition with other oral complications |
| dbm_exclusions | ICD10CM | E08.64 | Diabetes mellitus due to underlying condition with hypoglycemia |
| dbm_exclusions | ICD10CM | E08.640 | Diabetes mellitus due to underlying condition with hypoglycemia without coma |
| dbm_exclusions | ICD10CM | E08.641 | Diabetes mellitus due to underlying condition with hypoglycemia with coma |
| dbm_exclusions | ICD10CM | E08.649 | Diabetes mellitus due to underlying condition with hypoglycemia without coma |
| dbm_exclusions | ICD10CM | E08.65 | Diabetes mellitus due to underlying condition with hyperglycemia |
| dbm_exclusions | ICD10CM | E08.69 | Diabetes mellitus due to underlying condition with other specified complication |
| dbm_exclusions | ICD10CM | E08.8 | Diabetes mellitus due to underlying condition with unspecified complications |
| dbm_exclusions | ICD10CM | E08.9 | Diabetes mellitus due to underlying condition without complications |
| Codes additionally relevant for the VA | |  |  |
| no_dr_evidence | HF_imaging_TRI_TRR | VA-TRR DM2 FINDING DR NO RETINOPATHY LT | No evidence of DR, left eye |
| no_dr_evidence | HF_imaging_TRI_TRR | VA-TRR DM2 FINDING DR NO RETINOPATHY RT | No evidence of DR, right eye |
| no_dr_evidence | HF_imaging_TRI_TRR | VA-TRR DM RETINOPATHY EXAM NORMAL | No evidence of DR, both eyes |
| no_dr_evidence | HF_imaging_TRI_TRR | VA-TRR DM1 FINDING DR NO RETINOPATHY LT | No evidence of DR, left eye |
| no_dr_evidence | HF_imaging_TRI_TRR | VA-TRR DM1 FINDING DR NO RETINOPATHY RT | No evidence of DR, right eye |
| no_dr_evidence | HF_imaging_TRI_TRR | NO DIABETIC RETINOPATHY | No evidence of DR, both eyes |
| no_dr_evidence | HF_imaging_TRI_TRR | NORMAL RETINAL EXAM | No evidence of DR, both eyes |
| no_dr_evidence | HF_imaging_TRI_TRR | NO DIABETIC RETINOPATHY (OUTSIDE EXAM) | No evidence of DR, both eyes |
| no_dr_evidence | HF_imaging_TRI_TRR | DM NO DIABETIC RETINOPATHY LT | No evidence of DR, left eye |
| no_dr_evidence | HF_imaging_TRI_TRR | DM NO DIABETIC RETINOPATHY RT | No evidence of DR, right eye |
| no_dr_evidence | HF_imaging_TRI_TRR | LTRI DM FINDING DR NO RETINOPATHY RT | No evidence of DR, right eye |
| no_dr_evidence | HF_imaging_TRI_TRR | LTRI DM FINDING DR NO RETINOPATHY LT | No evidence of DR, left eye |
| no_dr_evidence | HF_imaging_TRI_TRR | LTRI DM RETINOPATHY EXAM NORMAL | No evidence of DR, both eyes |
| no_dr_evidence | HF_imaging_TRI_TRR | TRI DM FINDING DR NO RETINOPATHY RT | No evidence of DR, right eye |
| no_dr_evidence | HF_imaging_TRI_TRR | TRI DM FINDING DR NO RETINOPATHY LT | No evidence of DR, left eye |
| no_dr_evidence | HF_imaging_TRI_TRR | TRI DM RETINOPATHY EXAM NORMAL | No evidence of DR, both eyes |
| no_dr_evidence | HF_imaging_TRI_TRR | DIABETIC EYE NO RETINOPATHY FOUND | No evidence of DR, both eyes |
| no_dr_evidence | HF_imaging_TRI_TRR | DIABETIC RETINA-NO RETINOPATHY | No evidence of DR, both eyes |
| no_dr_evidence | HF_imaging_TRI_TRR | DIABETIC RETINOPATHY ABSENT | No evidence of DR, both eyes |
| no_dr_evidence | HF_imaging_TRI_TRR | DIABETES - NO RETINOPATHY | No evidence of DR, both eyes |
| no_dr_evidence | HF_imaging_TRI_TRR | DIABETIC RETINOPATHY - NO | No evidence of DR, both eyes |
| no_dr_evidence | HF_imaging_TRI_TRR | LTRI DM NO DIABETIC RETINOPATHY LT | No evidence of DR, left eye |
| no_dr_evidence | HF_imaging_TRI_TRR | LTRI DM NO DIABETIC RETINOPATHY RT | No evidence of DR, right eye |
