## Supplemental Table 3 for "Development of Portable Electronic Health Record Based Algorithms to Identify Individuals with Diabetic Retinopathy"

**Supplemental Table 3. List of drugs included in the DM algorithm for the VA**

| ID | Drug | OMOP ID | OMOP Name |
|---|---|---|---|
| INSULIN | INSULIN | 1596977 | Regular Insulin, Human |
| METFORMIN | METFORMIN | 1503328 | Metformin |
| INSULIN | INSULIN | 1596977 | Regular Insulin, Human |
| INSULIN | INSULIN | 1550023 | Insulin Lispro |
| INSULIN | INSULIN | 1567198 | Insulin, Aspart, Human |
| METFORMIN | METFORMIN | 40164929 | Metformin |
| METFORMIN | METFORMIN | 40164898 | Metformin |
| INSULIN | INSULIN | 1502905 | Insulin Glargine |
| METFORMIN | METFORMIN | 40164930 | Metformin |
| METFORMIN | METFORMIN | 1503327 | Metformin |
| INSULIN | INSULIN | 19071700 | Insulin Glargine |
| INSULIN | INSULIN | 1567198 | Insulin, Aspart, Human |
| INSULIN | INSULIN | 19071700 | Insulin Glargine |
| INSULIN | INSULIN | 46234045 | Insulin, Aspart, Human |
| METFORMIN | METFORMIN | 40164897 | Metformin |
| INSULIN | INSULIN | 1596977 | Regular Insulin, Human |
| METFORMIN | METFORMIN | 40063356 | Metformin |
| METFORMIN | METFORMIN | 40164929 | Metformin |
| INSULIN | INSULIN | 1567198 | Insulin, Aspart, Human |
| INSULIN | INSULIN | 1567232 | Insulin, Aspart, Human |
| INSULIN | INSULIN | 40051347 | Insulin Glargine |
| GLIMEPIRIDE | GLIMEPIRIDE | 1597760 | glimepiride |
| METFORMIN | METFORMIN | 40164932 | Metformin |
| INSULIN | INSULIN | 19120790 | Insulin Glargine |
| INSULIN | INSULIN | 36229086 | Insulin Glargine |
| SITAGLIPTIN | SITAGLIPTIN | 19125043 | sitagliptin |
| GLIPIZIDE | GLIPIZIDE | 19081295 | Glipizide |
| GLIPIZIDE | GLIPIZIDE | 1560233 | Glipizide |
| GLIMEPIRIDE | GLIMEPIRIDE | 1597757 | glimepiride |
| METFORMIN | METFORMIN | 1503297 | Metformin |
| GLYBURIDE | GLYBURIDE | 1559784 | Glyburide |
| INSULIN | INSULIN | 36229086 | Insulin Glargine |
| METFORMIN | METFORMIN | 40164897 | Metformin |
| INSULIN | INSULIN | 19071700 | Insulin Glargine |
| GLYBURIDE | GLYBURIDE | 19077682 | Glyburide |
| INSULIN | INSULIN | 19058398 | Insulin Lispro |
| METFORMIN | METFORMIN | 40117785 | Metformin |

| | | | |
|---|---|---|---|
| GLIMEPIRIDE | GLIMEPIRIDE | 1597775 | glimepiride |
| INSULIN | INSULIN | 1550023 | Insulin Lispro |
| METFORMIN | METFORMIN | 40164900 | Metformin |
| GLIPIZIDE | GLIPIZIDE | 19030443 | Glipizide |
| GLIPIZIDE | GLIPIZIDE | 19030445 | Glipizide |
| INSULIN | INSULIN | 40051347 | Insulin Glargine |
| INSULIN | INSULIN | 1502905 | Insulin Glargine |
| INSULIN | INSULIN | 1516981 | insulin detemir |
| INSULIN | INSULIN | 1596977 | Regular Insulin, Human |
| METFORMIN | METFORMIN | 40164929 | Metformin |
| METFORMIN | METFORMIN | 40063352 | Metformin |
| GLIPIZIDE | GLIPIZIDE | 19077638 | Glipizide |
| METFORMIN | METFORMIN | 40164947 | Metformin |
| GLIMEPIRIDE | GLIMEPIRIDE | 1597773 | glimepiride |
| SITAGLIPTIN | SITAGLIPTIN | 19125042 | sitagliptin |
| GLIPIZIDE | GLIPIZIDE | 19077636 | Glipizide |
| METFORMIN | METFORMIN | 40164930 | Metformin |
| GLIMEPIRIDE | GLIMEPIRIDE | 1597776 | glimepiride |
| INSULIN | INSULIN | 19078553 | insulin, isophane |
| GLYBURIDE | GLYBURIDE | 19077682 | Glyburide |
| INSULIN | INSULIN | 36229086 | Insulin Glargine |
| GLIMEPIRIDE | GLIMEPIRIDE | 1597759 | glimepiride |
| GLIMEPIRIDE | GLIMEPIRIDE | 19117200 | glimepiride |
| INSULIN | INSULIN | 1596977 | Regular Insulin, Human |
| METFORMIN | METFORMIN | 40164929 | Metformin |
| METFORMIN | METFORMIN | 40164938 | Metformin |
| GLYBURIDE | GLYBURIDE | 1559684 | Glyburide |
| PIOGLITAZONE | PIOGLITAZONE | 1525217 | pioglitazone |
| LIRAGLUTIDE | LIRAGLUTIDE | 40169223 | liraglutide |
| INSULIN | INSULIN | 19120315 | insulin, isophane |
| GLIMEPIRIDE | GLIMEPIRIDE | 1597772 | glimepiride |
| PIOGLITAZONE | PIOGLITAZONE | 1525218 | pioglitazone |
| INSULIN | INSULIN | 19071700 | Insulin Glargine |
| METFORMIN | METFORMIN | 40164929 | Metformin |
| METFORMIN | METFORMIN | 40164929 | Metformin |
| GLIPIZIDE | GLIPIZIDE | 19077636 | Glipizide |
| INSULIN | INSULIN | 40166274 | Insulin Lispro |
| METFORMIN | METFORMIN | 40164942 | Metformin |

| | | | |
|---|---|---|---|
| INSULIN | INSULIN | 1516979 | insulin detemir |
| GLIPIZIDE | GLIPIZIDE | 19085004 | Glipizide |
| GLYBURIDE | GLYBURIDE | 1559782 | Glyburide |
| GLIPIZIDE | GLIPIZIDE | 19077638 | Glipizide |
| METFORMIN | METFORMIN | 36235746 | Metformin |
| GLIPIZIDE | GLIPIZIDE | 19117945 | Glipizide |
| INSULIN | INSULIN | 19071700 | Insulin Glargine |
| PIOGLITAZONE | PIOGLITAZONE | 19099208 | pioglitazone |
| GLIPIZIDE | GLIPIZIDE | 19047638 | Glipizide |
| METFORMIN | METFORMIN | 40164929 | Metformin |
| INSULIN | INSULIN | 1502910 | Insulin Glargine |
| INSULIN | INSULIN | 1596977 | Regular Insulin, Human |
| INSULIN | INSULIN | 40052088 | Insulin Lispro |
| GLIMEPIRIDE | GLIMEPIRIDE | 19023585 | glimepiride |
| PIOGLITAZONE | PIOGLITAZONE | 19099209 | pioglitazone |
| METFORMIN | METFORMIN | 40164897 | Metformin |
| METFORMIN | METFORMIN | 40164946 | Metformin |
| GLIPIZIDE | GLIPIZIDE | 40044253 | Glipizide |
| METFORMIN | METFORMIN | 40164898 | Metformin |
| METFORMIN | METFORMIN | 40164923 | Metformin |
| SITAGLIPTIN | SITAGLIPTIN | 19125051 | sitagliptin |
| GLIMEPIRIDE | GLIMEPIRIDE | 1597760 | glimepiride |
| ROSIGLITAZONE | ROSIGLITAZONE | 19099245 | rosiglitazone |
| INSULIN | INSULIN | 19062999 | insulin, isophane |
| GLIMEPIRIDE | GLIMEPIRIDE | 1597774 | glimepiride |
| INSULIN | INSULIN | 19078557 | Regular Insulin, Human |
| INSULIN | INSULIN | 1596977 | Regular Insulin, Human |
| METFORMIN | METFORMIN | 40163926 | Metformin |
| GLYBURIDE | GLYBURIDE | 19030579 | Glyburide |
| METFORMIN | METFORMIN | 40164897 | Metformin |
| PIOGLITAZONE | PIOGLITAZONE | 1525217 | pioglitazone |
| METFORMIN | METFORMIN | 40164898 | Metformin |
| INSULIN | INSULIN | 1567232 | Insulin, Aspart, Human |
| INSULIN | INSULIN | 1567232 | Insulin, Aspart, Human |

| | | | |
|---|---|---|---|
| PIOGLITAZONE | PIOGLITAZONE | 1525216 | pioglitazone |
| INSULIN | INSULIN | 46234049 | Insulin, Aspart, Human |
| SITAGLIPTIN | SITAGLIPTIN | 19125043 | sitagliptin |
| ROSIGLITAZONE | ROSIGLITAZONE | 1547506 | rosiglitazone |
| NATEGLINIDE | NATEGLINIDE | 19071495 | nateglinide |
| METFORMIN | METFORMIN | 40163926 | Metformin |
| INSULIN | INSULIN | 36231450 | Insulin, Aspart, Human |
| GLIMEPIRIDE | GLIMEPIRIDE | 1597773 | glimepiride |
| GLYBURIDE | GLYBURIDE | 19077659 | Glyburide |
| EXENATIDE | EXENATIDE | 1583730 | exenatide |
| PIOGLITAZONE | PIOGLITAZONE | 1525218 | pioglitazone |
| INSULIN | INSULIN | 19078554 | Regular Insulin, Human |
| PIOGLITAZONE | PIOGLITAZONE | 19099207 | pioglitazone |
| GLIMEPIRIDE | GLIMEPIRIDE | 36214984 | glimepiride |
| GLIMEPIRIDE | GLIMEPIRIDE | 36234085 | glimepiride |
| METFORMIN | METFORMIN | 40164946 | Metformin |
| INSULIN | INSULIN | 19078557 | Regular Insulin, Human |
| METFORMIN | METFORMIN | 40164897 | Metformin |
| GLIPIZIDE | GLIPIZIDE | 1560171 | Glipizide |
| GLIMEPIRIDE | GLIMEPIRIDE | 1597761 | glimepiride |
| METFORMIN | METFORMIN | 40153151 | Metformin |
| METFORMIN | METFORMIN | 40164948 | Metformin |
| INSULIN | INSULIN | 40184294 | Insulin, Aspart Protamine, Human |
| INSULIN | INSULIN | 1567198 | Insulin, Aspart, Human |
| ROSIGLITAZONE | ROSIGLITAZONE | 1547506 | rosiglitazone |
| EXENATIDE | EXENATIDE | 19083253 | exenatide |
| METFORMIN | METFORMIN | 40164897 | Metformin |
| LINAGLIPTIN | LINAGLIPTIN | 40239219 | Linagliptin |
| GLYBURIDE | GLYBURIDE | 40046204 | Glyburide |
| GLIMEPIRIDE | GLIMEPIRIDE | 19117198 | glimepiride |
| INSULIN | INSULIN | 19078554 | Regular Insulin, Human |
| INSULIN | INSULIN | 40184293 | Insulin, Aspart Protamine, Human |
| METFORMIN | METFORMIN | 40063352 | Metformin |
| GLIMEPIRIDE | GLIMEPIRIDE | 1597757 | glimepiride |

| | | | |
|---|---|---|---|
| NATEGLINIDE | NATEGLINIDE | 19045606 | nateglinide |
| GLIMEPIRIDE | GLIMEPIRIDE | 1597756 | glimepiride |
| INSULIN | INSULIN | 36229086 | Insulin Glargine |
| INSULIN | INSULIN | 46233975 | Insulin Lispro |
| GLYBURIDE | GLYBURIDE | 40046201 | Glyburide |
| INSULIN | INSULIN | 1516981 | insulin detemir |
| INSULIN | INSULIN | 19078551 | insulin, isophane |
| INSULIN | INSULIN | 40166274 | Insulin Lispro |
| METFORMIN | METFORMIN | 40164930 | Metformin |
| SITAGLIPTIN | SITAGLIPTIN | 19125050 | sitagliptin |
| GLYBURIDE | GLYBURIDE | 19077659 | Glyburide |
| GLIMEPIRIDE | GLIMEPIRIDE | 1597772 | glimepiride |
| PIOGLITAZONE | PIOGLITAZONE | 1525216 | pioglitazone |
| INSULIN | INSULIN | 40184293 | Insulin, Aspart Protamine, Human |
| ROSIGLITAZONE | ROSIGLITAZONE | 19099246 | rosiglitazone |
| GLIPIZIDE | GLIPIZIDE | 36218767 | Glipizide |
| INSULIN | INSULIN | 40176981 | Insulin, Aspart Protamine, Human |
| INSULIN | INSULIN | 40051348 | Insulin Glargine |
| INSULIN | INSULIN | 46234045 | Insulin, Aspart, Human |
| INSULIN | INSULIN | 1596977 | Regular Insulin, Human |
| GLYBURIDE | GLYBURIDE | 19030575 | Glyburide |
| ROSIGLITAZONE | ROSIGLITAZONE | 1547507 | rosiglitazone |
| GLIPIZIDE | GLIPIZIDE | 19030445 | Glipizide |
| METFORMIN | METFORMIN | 40163928 | Metformin |
| GLYBURIDE | GLYBURIDE | 40168374 | Glyburide |
| INSULIN | INSULIN | 19078530 | insulin, isophane |
| METFORMIN | METFORMIN | 40164892 | Metformin |
| METFORMIN | METFORMIN | 40163926 | Metformin |
| METFORMIN | METFORMIN | 40164925 | Metformin |
| INSULIN | INSULIN | 36236032 | Insulin Lispro |
| GLYBURIDE | GLYBURIDE | 19077682 | Glyburide |
| INSULIN | INSULIN | 19078530 | insulin, isophane |

| | | | |
|---|---|---|---|
| LIRAGLUTIDE | LIRAGLUTIDE | 40170913 | liraglutide |
| INSULIN | INSULIN | 46233970 | Insulin Lispro |
| GLIPIZIDE | GLIPIZIDE | 36235748 | Glipizide |
| INSULIN | INSULIN | 19071700 | Insulin Glargine |
| GLIMEPIRIDE | GLIMEPIRIDE | 1597760 | glimepiride |
| INSULIN | INSULIN | 40184293 | Insulin, Aspart Protamine, Human |
| NATEGLINIDE | NATEGLINIDE | 19071495 | nateglinide |
| GLYBURIDE | GLYBURIDE | 40046201 | Glyburide |
| GLIPIZIDE | GLIPIZIDE | 19100559 | Glipizide |
| LIRAGLUTIDE | LIRAGLUTIDE | 36241903 | liraglutide |
| INSULIN | INSULIN | 1596999 | Regular Insulin, Human |
| INSULIN | INSULIN | 19009384 | insulin, isophane |
| GLIPIZIDE | GLIPIZIDE | 36218767 | Glipizide |
| METFORMIN | METFORMIN | 40164929 | Metformin |
| INSULIN | INSULIN | 46233975 | Insulin Lispro |
| SAXAGLIPTIN | SAXAGLIPTIN | 40166042 | saxagliptin |
| INSULIN | INSULIN | 19044971 | insulin detemir |
| EXENATIDE | EXENATIDE | 1583727 | exenatide |
| INSULIN | INSULIN | 36222190 | insulin detemir |
| GLIPIZIDE | GLIPIZIDE | 19030443 | Glipizide |
| INSULIN | INSULIN | 19009384 | insulin, isophane |
| INSULIN | INSULIN | 1513881 | Insulin Lispro |
| GLIPIZIDE | GLIPIZIDE | 19077638 | Glipizide |
| INSULIN | INSULIN | 1502910 | Insulin Glargine |
| INSULIN | INSULIN | 19018822 | Insulin Lispro |
| INSULIN | INSULIN | 1596916 | insulin, isophane |
| LINAGLIPTIN | LINAGLIPTIN | 40239220 | Linagliptin |
| INSULIN | INSULIN | 1513912 | Insulin Lispro |
| INSULIN | INSULIN | 19078554 | Regular Insulin, Human |
| CANAGLIFLOZIN | CANAGLIFLOZIN | 43526472 | canagliflozin |
| DAPAGLIFLOZIN | DAPAGLIFLOZIN | 44785473 | dapagliflozin |

| | | | |
|---|---|---|---|
| EXENATIDE | EXENATIDE | 19083254 | exenatide |
| METFORMIN | METFORMIN | 40163926 | Metformin |
| GLIMEPIRIDE | GLIMEPIRIDE | 1597760 | glimepiride |
| INSULIN | INSULIN | 19078551 | insulin, isophane |
| ROSIGLITAZONE | ROSIGLITAZONE | 1547507 | rosiglitazone |
| INSULIN | INSULIN | 1502905 | Insulin Glargine |
| GLIPIZIDE | GLIPIZIDE | 19077636 | Glipizide |
| INSULIN | INSULIN | 19071700 | Insulin Glargine |
| METFORMIN | METFORMIN | 40164898 | Metformin |
| INSULIN | INSULIN | 46221557 | Insulin Glargine |
| INSULIN | INSULIN | 1596916 | insulin, isophane |
| GLIMEPIRIDE | GLIMEPIRIDE | 1597773 | glimepiride |
| METFORMIN | METFORMIN | 40164947 | Metformin |
| GLYBURIDE | GLYBURIDE | 1559760 | Glyburide |
| INSULIN | INSULIN | 1596999 | Regular Insulin, Human |
| METFORMIN | METFORMIN | 40164893 | Metformin |
| METFORMIN | METFORMIN | 40135481 | Metformin |
| GLIPIZIDE | GLIPIZIDE | 19077638 | Glipizide |
| GLIPIZIDE | GLIPIZIDE | 19077636 | Glipizide |
| METFORMIN | METFORMIN | 40163924 | Metformin |
| NATEGLINIDE | NATEGLINIDE | 1502827 | nateglinide |
| LIRAGLUTIDE | LIRAGLUTIDE | 40170913 | liraglutide |
| GLYBURIDE | GLYBURIDE | 40165993 | Glyburide |
| METFORMIN | METFORMIN | 40164930 | Metformin |
| GLYBURIDE | GLYBURIDE | 19045630 | Glyburide |
| METFORMIN | METFORMIN | 40063352 | Metformin |
| INSULIN | INSULIN | 1567198 | Insulin, Aspart, Human |
| INSULIN | INSULIN | 19131582 | Insulin, Glulisine, Human |
| GLIMEPIRIDE | GLIMEPIRIDE | 1597757 | glimepiride |
| CANAGLIFLOZIN | CANAGLIFLOZIN | 43526468 | canagliflozin |
| GLIMEPIRIDE | GLIMEPIRIDE | 40059666 | glimepiride |
| INSULIN | INSULIN | 1550023 | Insulin Lispro |
| GLIPIZIDE | GLIPIZIDE | 19077638 | Glipizide |
| INSULIN | INSULIN | 1567198 | Insulin, Aspart, Human |

| | | | |
|---|---|---|---|
| CANAGLIFLOZIN | CANAGLIFLOZIN | 43526473 | canagliflozin |
| INSULIN | INSULIN | 1513912 | Insulin Lispro |
| INSULIN | INSULIN | 19078557 | Regular Insulin, Human |
| METFORMIN | METFORMIN | 36235746 | Metformin |
| EXENATIDE | EXENATIDE | 19083253 | exenatide |
| INSULIN | INSULIN | 19078530 | insulin, isophane |
| GLIPIZIDE | GLIPIZIDE | 40167636 | Glipizide |
| METFORMIN | METFORMIN | 36236322 | Metformin |
| INSULIN | INSULIN | 1513912 | Insulin Lispro |
| GLYBURIDE | GLYBURIDE | 19077682 | Glyburide |
| GLIPIZIDE | GLIPIZIDE | 19077638 | Glipizide |
| GLIPIZIDE | GLIPIZIDE | 19100559 | Glipizide |
| GLIMEPIRIDE | GLIMEPIRIDE | 1597759 | glimepiride |
| DAPAGLIFLOZIN | DAPAGLIFLOZIN | 44816049 | dapagliflozin |
| METFORMIN | METFORMIN | 40164929 | Metformin |
| INSULIN | INSULIN | 19131582 | Insulin, Glulisine, Human |
| GLYBURIDE | GLYBURIDE | 19077682 | Glyburide |
| INSULIN | INSULIN | 46233972 | Insulin Lispro |
| METFORMIN | METFORMIN | 40164924 | Metformin |
| INSULIN | INSULIN | 46233970 | Insulin Lispro |
| SITAGLIPTIN | SITAGLIPTIN | 19125043 | sitagliptin |
| INSULIN | INSULIN | 1567198 | Insulin, Aspart, Human |
| REPAGLINIDE | REPAGLINIDE | 19047663 | repaglinide |
| INSULIN | INSULIN | 19078530 | insulin, isophane |
| BROMOCRIPTINE | BROMOCRIPTINE | 19007101 | Bromocriptine |
| EXENATIDE | EXENATIDE | 42705842 | exenatide |
| GLYBURIDE | GLYBURIDE | 19077682 | Glyburide |
| INSULIN | INSULIN | 1502905 | Insulin Glargine |
| INSULIN | INSULIN | 1550027 | Insulin Lispro |
| GLIMEPIRIDE | GLIMEPIRIDE | 1597757 | glimepiride |
| METFORMIN | METFORMIN | 40135481 | Metformin |
| INSULIN | INSULIN | 36231450 | Insulin, Aspart, Human |
| GLIMEPIRIDE | GLIMEPIRIDE | 1597761 | glimepiride |

| | | | |
|---|---|---|---|
| INSULIN | INSULIN | 19078554 | Regular Insulin, Human |
| GLIMEPIRIDE | GLIMEPIRIDE | 1597772 | glimepiride |
| INSULIN | INSULIN | 19120315 | insulin, isophane |
| INSULIN | INSULIN | 1544873 | Insulin, Glulisine, Human |
| METFORMIN | METFORMIN | 40164904 | Metformin |
| INSULIN | INSULIN | 1567198 | Insulin, Aspart, Human |
| GLIPIZIDE | GLIPIZIDE | 19030445 | Glipizide |
| INSULIN | INSULIN | 19078555 | Regular Insulin, Human |
| SAXAGLIPTIN | SAXAGLIPTIN | 40166043 | saxagliptin |
| GLYBURIDE | GLYBURIDE | 40166003 | Glyburide |
| INSULIN | INSULIN | 1516981 | insulin detemir |
| SITAGLIPTIN | SITAGLIPTIN | 19125047 | sitagliptin |
| PIOGLITAZONE | PIOGLITAZONE | 1525218 | pioglitazone |
| GLYBURIDE | GLYBURIDE | 40166002 | Glyburide |
| EXENATIDE | EXENATIDE | 46234118 | exenatide |
| CANAGLIFLOZIN | CANAGLIFLOZIN | 43526469 | canagliflozin |
| INSULIN | INSULIN | 1596960 | insulin, isophane |
| INSULIN | INSULIN | 35602724 | insulin degludec |
| METFORMIN | METFORMIN | 40063354 | Metformin |
| METFORMIN | METFORMIN | 40163925 | Metformin |
| GLYBURIDE | GLYBURIDE | 40166003 | Glyburide |
| EXENATIDE | EXENATIDE | 19083253 | exenatide |
| SITAGLIPTIN | SITAGLIPTIN | 36221232 | sitagliptin |
| METFORMIN | METFORMIN | 40164880 | Metformin |
| METFORMIN | METFORMIN | 40063354 | Metformin |
| METFORMIN | METFORMIN | 19086484 | Metformin |
| PIOGLITAZONE | PIOGLITAZONE | 36236324 | pioglitazone |
| GLYBURIDE | GLYBURIDE | 1559785 | Glyburide |
| INSULIN | INSULIN | 1550028 | Insulin Lispro |
| METFORMIN | METFORMIN | 40164928 | Metformin |
| METFORMIN | METFORMIN | 40164923 | Metformin |

| | | | |
|---|---|---|---|
| INSULIN | INSULIN | 35602731 | insulin degludec |
| INSULIN | INSULIN | 19078557 | Regular Insulin, Human |
| INSULIN | INSULIN | 46233972 | Insulin Lispro |
| GLIMEPIRIDE | GLIMEPIRIDE | 36234085 | glimepiride |
| PIOGLITAZONE | PIOGLITAZONE | 1525242 | pioglitazone |
| EXENATIDE | EXENATIDE | 1583730 | exenatide |
| METFORMIN | METFORMIN | 40164941 | Metformin |
| GLIPIZIDE | GLIPIZIDE | 19077638 | Glipizide |
| EXENATIDE | EXENATIDE | 42708079 | exenatide |
| ROSIGLITAZONE | ROSIGLITAZONE | 1547506 | rosiglitazone |
| BROMOCRIPTINE | BROMOCRIPTINE | 19083957 | Bromocriptine |
| METFORMIN | METFORMIN | 40231396 | Metformin |
| INSULIN | INSULIN | 19078551 | insulin, isophane |
| ROSIGLITAZONE | ROSIGLITAZONE | 19120648 | rosiglitazone |
| REPAGLINIDE | REPAGLINIDE | 19078030 | repaglinide |
| INSULIN | INSULIN | 1596941 | insulin, isophane |
| GLYBURIDE | GLYBURIDE | 1559685 | Glyburide |
| INSULIN | INSULIN | 19120790 | Insulin Glargine |
| METFORMIN | METFORMIN | 40164946 | Metformin |
| METFORMIN | METFORMIN | 40164892 | Metformin |
| METFORMIN | METFORMIN | 40163928 | Metformin |
| GLYBURIDE | GLYBURIDE | 40166002 | Glyburide |
| GLIMEPIRIDE | GLIMEPIRIDE | 1597773 | glimepiride |
| NATEGLINIDE | NATEGLINIDE | 19045605 | nateglinide |
| INSULIN | INSULIN | 46234043 | Insulin, Aspart, Human |
| METFORMIN | METFORMIN | 40063352 | Metformin |
| SITAGLIPTIN | SITAGLIPTIN | 19125043 | sitagliptin |
| INSULIN | INSULIN | 19009384 | insulin, isophane |
| GLIPIZIDE | GLIPIZIDE | 19077636 | Glipizide |
| INSULIN | INSULIN | 40184293 | Insulin, Aspart Protamine, Human |
| GLIMEPIRIDE | GLIMEPIRIDE | 1597760 | glimepiride |
| PIOGLITAZONE | PIOGLITAZONE | 19079293 | pioglitazone |
| LIRAGLUTIDE | LIRAGLUTIDE | 40169223 | liraglutide |

| | | | |
|---|---|---|---|
| GLYBURIDE | GLYBURIDE | 40046201 | Glyburide |
| METFORMIN | METFORMIN | 40164946 | Metformin |
| INSULIN | INSULIN | 19078530 | insulin, isophane |
| INSULIN | INSULIN | 1567198 | Insulin, Aspart, Human |
| GLYBURIDE | GLYBURIDE | 19077659 | Glyburide |
| DAPAGLIFLOZIN | DAPAGLIFLOZIN | 44785477 | dapagliflozin |
| INSULIN | INSULIN | 19009384 | insulin, isophane |
| INSULIN | INSULIN | 19120315 | insulin, isophane |
| METFORMIN | METFORMIN | 40163926 | Metformin |
| INSULIN | INSULIN | 36235841 | Regular Insulin, Human |
| INSULIN | INSULIN | 19078555 | Regular Insulin, Human |
| GLYBURIDE | GLYBURIDE | 19077682 | Glyburide |
| DULAGLUTIDE | DULAGLUTIDE | 45774447 | dulaglutide |
| INSULIN | INSULIN | 1502905 | Insulin Glargine |
| PIOGLITAZONE | PIOGLITAZONE | 1525216 | pioglitazone |
| METFORMIN | METFORMIN | 40063352 | Metformin |
| INSULIN | INSULIN | 19095214 | insulin, isophane |
| REPAGLINIDE | REPAGLINIDE | 1516771 | repaglinide |
| INSULIN | INSULIN | 1550027 | Insulin Lispro |
| GLIMEPIRIDE | GLIMEPIRIDE | 1597757 | glimepiride |
| ROSIGLITAZONE | ROSIGLITAZONE | 1547505 | rosiglitazone |
| METFORMIN | METFORMIN | 40164897 | Metformin |
| METFORMIN | METFORMIN | 36223145 | Metformin |
| METFORMIN | METFORMIN | 40164898 | Metformin |
| PIOGLITAZONE | PIOGLITAZONE | 36236324 | pioglitazone |
| METFORMIN | METFORMIN | 40164946 | Metformin |
| SITAGLIPTIN | SITAGLIPTIN | 19125051 | sitagliptin |
| GLIMEPIRIDE | GLIMEPIRIDE | 1597759 | glimepiride |
| INSULIN | INSULIN | 46233948 | Insulin Lispro |
| INSULIN | INSULIN | 40052088 | Insulin Lispro |
| INSULIN | INSULIN | 19058398 | Insulin Lispro |
| PRAMLINTIDE | PRAMLINTIDE | 40165687 | Pramlintide |
| METFORMIN | METFORMIN | 40164918 | Metformin |
| METFORMIN | METFORMIN | 40163929 | Metformin |

| | | | |
|---|---|---|---|
| METFORMIN | METFORMIN | 40164944 | Metformin |
| GLIMEPIRIDE | GLIMEPIRIDE | 1597773 | glimepiride |
| INSULIN | INSULIN | 1596916 | insulin, isophane |
| INSULIN | INSULIN | 1596977 | Regular Insulin, Human |
| GLYBURIDE | GLYBURIDE | 19077684 | Glyburide |
| METFORMIN | METFORMIN | 40164930 | Metformin |
| PIOGLITAZONE | PIOGLITAZONE | 1525217 | pioglitazone |
| INSULIN | INSULIN | 1502908 | Insulin Glargine |
| GLYBURIDE | GLYBURIDE | 19077684 | Glyburide |
| INSULIN | INSULIN | 19120790 | Insulin Glargine |
| ROSIGLITAZONE | ROSIGLITAZONE | 1547505 | rosiglitazone |
| GLIPIZIDE | GLIPIZIDE | 19030445 | Glipizide |
| GLYBURIDE | GLYBURIDE | 19030572 | Glyburide |
| METFORMIN | METFORMIN | 40063350 | Metformin |
| EXENATIDE | EXENATIDE | 19083254 | exenatide |
| GLYBURIDE | GLYBURIDE | 40168370 | Glyburide |
| GLIPIZIDE | GLIPIZIDE | 36218767 | Glipizide |
| INSULIN | INSULIN | 19120315 | insulin, isophane |
| METFORMIN | METFORMIN | 40164946 | Metformin |
| GLIMEPIRIDE | GLIMEPIRIDE | 43011499 | glimepiride |
| GLYBURIDE | GLYBURIDE | 40166003 | Glyburide |
| INSULIN | INSULIN | 1516976 | insulin detemir |
| GLIMEPIRIDE | GLIMEPIRIDE | 1597772 | glimepiride |
| EXENATIDE | EXENATIDE | 19083254 | exenatide |
| PIOGLITAZONE | PIOGLITAZONE | 1525243 | pioglitazone |
| SITAGLIPTIN | SITAGLIPTIN | 19125043 | sitagliptin |
| METFORMIN | METFORMIN | 40164930 | Metformin |
| INSULIN | INSULIN | 19071700 | Insulin Glargine |
| METFORMIN | METFORMIN | 40164896 | Metformin |
| REPAGLINIDE | REPAGLINIDE | 1516770 | repaglinide |
| INSULIN | INSULIN | 1596999 | Regular Insulin, Human |
| INSULIN | INSULIN | 40051347 | Insulin Glargine |
| METFORMIN | METFORMIN | 40164926 | Metformin |

| | | | |
|---|---|---|---|
| GLIPIZIDE | GLIPIZIDE | 40166814 | Glipizide |
| INSULIN | INSULIN | 1567198 | Insulin, Aspart, Human |
| INSULIN | INSULIN | 40051347 | Insulin Glargine |
| INSULIN | INSULIN | 19078551 | insulin, isophane |
| GLYBURIDE | GLYBURIDE | 40166004 | Glyburide |
| METFORMIN | METFORMIN | 40164933 | Metformin |
| INSULIN | INSULIN | 36229086 | Insulin Glargine |
| PIOGLITAZONE | PIOGLITAZONE | 1525220 | pioglitazone |
| INSULIN | INSULIN | 1567198 | Insulin, Aspart, Human |
| METFORMIN | METFORMIN | 40164929 | Metformin |
| INSULIN | INSULIN | 1596916 | insulin, isophane |
| GLIPIZIDE | GLIPIZIDE | 40167638 | Glipizide |
| BROMOCRIPTINE | BROMOCRIPTINE | 730548 | Bromocriptine |
| DAPAGLIFLOZIN | DAPAGLIFLOZIN | 44815996 | dapagliflozin |
| METFORMIN | METFORMIN | 36221230 | Metformin |
| LIRAGLUTIDE | LIRAGLUTIDE | 36241903 | liraglutide |
| INSULIN | INSULIN | 46233975 | Insulin Lispro |
| METFORMIN | METFORMIN | 40163922 | Metformin |
| GLYBURIDE | GLYBURIDE | 19077682 | Glyburide |
| GLIPIZIDE | GLIPIZIDE | 19077636 | Glipizide |
| INSULIN | INSULIN | 36236032 | Insulin Lispro |
| INSULIN | INSULIN | 40184293 | Insulin, Aspart Protamine, Human |
| PIOGLITAZONE | PIOGLITAZONE | 1525244 | pioglitazone |
| INSULIN | INSULIN | 36222190 | insulin detemir |
| GLYBURIDE | GLYBURIDE | 40165998 | Glyburide |
| INSULIN | INSULIN | 46234049 | Insulin, Aspart, Human |
| METFORMIN | METFORMIN | 36235746 | Metformin |
| METFORMIN | METFORMIN | 40163926 | Metformin |
| DULAGLUTIDE | DULAGLUTIDE | 45774440 | dulaglutide |
| INSULIN | INSULIN | 40051347 | Insulin Glargine |
| GLIPIZIDE | GLIPIZIDE | 19077636 | Glipizide |
| METFORMIN | METFORMIN | 19113780 | Metformin |
| INSULIN | INSULIN | 36235840 | insulin, isophane |
| GLIPIZIDE | GLIPIZIDE | 40166816 | Glipizide |

| | | | |
|---|---|---|---|
| METFORMIN | METFORMIN | 40164881 | Metformin |
| ROSIGLITAZONE | ROSIGLITAZONE | 1547507 | rosiglitazone |
| INSULIN | INSULIN | 19071700 | Insulin Glargine |
| GLYBURIDE | GLYBURIDE | 40165998 | Glyburide |
| GLIMEPIRIDE | GLIMEPIRIDE | 1597773 | glimepiride |
| INSULIN | INSULIN | 1516981 | insulin detemir |
| INSULIN | INSULIN | 46234240 | insulin, isophane |
| INSULIN | INSULIN | 1550023 | Insulin Lispro |
| LIRAGLUTIDE | LIRAGLUTIDE | 40169223 | liraglutide |
| GLIMEPIRIDE | GLIMEPIRIDE | 1597760 | glimepiride |
| INSULIN | INSULIN | 1502905 | Insulin Glargine |
| SITAGLIPTIN | SITAGLIPTIN | 19125043 | sitagliptin |
| GLYBURIDE | GLYBURIDE | 1559685 | Glyburide |
| PIOGLITAZONE | PIOGLITAZONE | 40099897 | pioglitazone |
| GLYBURIDE | GLYBURIDE | 19081296 | Glyburide |
| GLIMEPIRIDE | GLIMEPIRIDE | 1597772 | glimepiride |
| EMPAGLIFLOZIN | EMPAGLIFLOZIN | 45774895 | empagliflozin |
| SITAGLIPTIN | SITAGLIPTIN | 19125046 | sitagliptin |
| INSULIN | INSULIN | 19078558 | Insulin, Aspart, Human |
| INSULIN | INSULIN | 19078551 | insulin, isophane |
| REPAGLINIDE | REPAGLINIDE | 19078029 | repaglinide |
| GLIPIZIDE | GLIPIZIDE | 19030443 | Glipizide |
| INSULIN | INSULIN | 46233975 | Insulin Lispro |
| METFORMIN | METFORMIN | 40164898 | Metformin |
| GLIPIZIDE | GLIPIZIDE | 19100559 | Glipizide |
| GLIMEPIRIDE | GLIMEPIRIDE | 1597761 | glimepiride |
| INSULIN | INSULIN | 1516979 | insulin detemir |
| INSULIN | INSULIN | 19078557 | Regular Insulin, Human |
| METFORMIN | METFORMIN | 40164923 | Metformin |
| INSULIN | INSULIN | 1567198 | Insulin, Aspart, Human |
| GLYBURIDE | GLYBURIDE | 36230035 | Glyburide |
| INSULIN | INSULIN | 1550023 | Insulin Lispro |
| INSULIN | INSULIN | 40051348 | Insulin Glargine |
| METFORMIN | METFORMIN | 40164946 | Metformin |
| METFORMIN | METFORMIN | 40063350 | Metformin |
| GLYBURIDE | GLYBURIDE | 19045629 | Glyburide |

| | | | |
|---|---|---|---|
| METFORMIN | METFORMIN | 40164898 | Metformin |
| EMPAGLIFLOZIN | EMPAGLIFLOZIN | 45774758 | empagliflozin |
| INSULIN | INSULIN | 1567232 | Insulin, Aspart, Human |
| NATEGLINIDE | NATEGLINIDE | 19085096 | nateglinide |
| METFORMIN | METFORMIN | 40164895 | Metformin |
| REPAGLINIDE | REPAGLINIDE | 19047663 | repaglinide |
| INSULIN | INSULIN | 1596955 | insulin, isophane |
| METFORMIN | METFORMIN | 40164895 | Metformin |
| REPAGLINIDE | REPAGLINIDE | 19078028 | repaglinide |
| INSULIN | INSULIN | 1550028 | Insulin Lispro |
| METFORMIN | METFORMIN | 42708167 | Metformin |
| INSULIN | INSULIN | 1550023 | Insulin Lispro |
| GLYBURIDE | GLYBURIDE | 40046201 | Glyburide |
| INSULIN | INSULIN | 40166275 | Insulin Lispro |
| INSULIN | INSULIN | 19120315 | insulin, isophane |
| EXENATIDE | EXENATIDE | 1583727 | exenatide |
| GLYBURIDE | GLYBURIDE | 19077659 | Glyburide |
| INSULIN | INSULIN | 19078553 | insulin, isophane |
| METFORMIN | METFORMIN | 36235787 | Metformin |
| INSULIN | INSULIN | 40166274 | Insulin Lispro |
| METFORMIN | METFORMIN | 40164899 | Metformin |
| INSULIN | INSULIN | 1550023 | Insulin Lispro |
| METFORMIN | METFORMIN | 40164931 | Metformin |
| PRAMLINTIDE | PRAMLINTIDE | 40163890 | Pramlintide |
| INSULIN | INSULIN | 19120315 | insulin, isophane |
| GLYBURIDE | GLYBURIDE | 19077659 | Glyburide |
| PIOGLITAZONE | PIOGLITAZONE | 1525221 | pioglitazone |
| INSULIN | INSULIN | 46234045 | Insulin, Aspart, Human |
| INSULIN | INSULIN | 1544873 | Insulin, Glulisine, Human |
| GLIPIZIDE | GLIPIZIDE | 19030445 | Glipizide |
| INSULIN | INSULIN | 1502905 | Insulin Glargine |
| GLIMEPIRIDE | GLIMEPIRIDE | 1597772 | glimepiride |

| | | | |
|---|---|---|---|
| METFORMIN | METFORMIN | 40164940 | Metformin |
| PIOGLITAZONE | PIOGLITAZONE | 1525217 | pioglitazone |
| INSULIN | INSULIN | 1567232 | Insulin, Aspart, Human |
| GLYBURIDE | GLYBURIDE | 19045626 | Glyburide |
| INSULIN | INSULIN | 46233975 | Insulin Lispro |
| GLYBURIDE | GLYBURIDE | 19077681 | Glyburide |
| METFORMIN | METFORMIN | 40163928 | Metformin |
| NATEGLINIDE | NATEGLINIDE | 1502829 | nateglinide |
| METFORMIN | METFORMIN | 42705990 | Metformin |
| INSULIN | INSULIN | 1596941 | insulin, isophane |
| SAXAGLIPTIN | SAXAGLIPTIN | 40166038 | saxagliptin |
| INSULIN | INSULIN | 19078559 | Insulin Glargine |
| GLYBURIDE | GLYBURIDE | 40168375 | Glyburide |
| METFORMIN | METFORMIN | 40164930 | Metformin |
| INSULIN | INSULIN | 36229086 | Insulin Glargine |
| METFORMIN | METFORMIN | 40164940 | Metformin |
| INSULIN | INSULIN | 1597001 | Regular Insulin, Human |
| METFORMIN | METFORMIN | 40164929 | Metformin |
| INSULIN | INSULIN | 19062999 | insulin, isophane |
| INSULIN | INSULIN | 19078559 | Insulin Glargine |
| INSULIN | INSULIN | 19062999 | insulin, isophane |
| GLYBURIDE | GLYBURIDE | 40046201 | Glyburide |
| METFORMIN | METFORMIN | 19106521 | Metformin |
| GLIPIZIDE | GLIPIZIDE | 19077638 | Glipizide |
| NATEGLINIDE | NATEGLINIDE | 19071495 | nateglinide |
| INSULIN | INSULIN | 19110357 | Regular Insulin, Human |
| BROMOCRIPTINE | BROMOCRIPTINE | 19116171 | Bromocriptine |
| SITAGLIPTIN | SITAGLIPTIN | 36221232 | sitagliptin |
| METFORMIN | METFORMIN | 40164947 | Metformin |
| DULAGLUTIDE | DULAGLUTIDE | 45774440 | dulaglutide |
| EMPAGLIFLOZIN | EMPAGLIFLOZIN | 45774894 | empagliflozin |
| INSULIN | INSULIN | 19062999 | insulin, isophane |

| | | | |
|---|---|---|---|
| GLYBURIDE | GLYBURIDE | 40165997 | Glyburide |
| METFORMIN | METFORMIN | 40163924 | Metformin |
| INSULIN | INSULIN | 1567198 | Insulin, Aspart, Human |
| INSULIN | INSULIN | 40184293 | Insulin, Aspart Protamine, Human |
| GLYBURIDE | GLYBURIDE | 40168374 | Glyburide |
| ACARBOSE | ACARBOSE | 19040969 | Acarbose |
| GLYBURIDE | GLYBURIDE | 19077681 | Glyburide |
| INSULIN | INSULIN | 1597001 | Regular Insulin, Human |
| INSULIN | INSULIN | 1502905 | Insulin Glargine |
| ROSIGLITAZONE | ROSIGLITAZONE | 36229365 | rosiglitazone |
| EXENATIDE | EXENATIDE | 19083253 | exenatide |
| NATEGLINIDE | NATEGLINIDE | 1502827 | nateglinide |
| GLIMEPIRIDE | GLIMEPIRIDE | 36214984 | glimepiride |
| INSULIN | INSULIN | 36231458 | insulin, isophane |
| INSULIN | INSULIN | 19078553 | insulin, isophane |
| GLIMEPIRIDE | GLIMEPIRIDE | 1597757 | glimepiride |
| INSULIN | INSULIN | 19078530 | insulin, isophane |
| INSULIN | INSULIN | 46233976 | Insulin Lispro |
| METFORMIN | METFORMIN | 40164898 | Metformin |
| GLIPIZIDE | GLIPIZIDE | 40166814 | Glipizide |
| EXENATIDE | EXENATIDE | 19083253 | exenatide |
| SITAGLIPTIN | SITAGLIPTIN | 19125041 | sitagliptin |
| INSULIN | INSULIN | 1513912 | Insulin Lispro |
| INSULIN | INSULIN | 19078552 | insulin, isophane |
| INSULIN | INSULIN | 1596977 | Regular Insulin, Human |
| EMPAGLIFLOZIN | EMPAGLIFLOZIN | 45774756 | empagliflozin |
| INSULIN | INSULIN | 19078555 | Regular Insulin, Human |
| NATEGLINIDE | NATEGLINIDE | 19085097 | nateglinide |
| EXENATIDE | EXENATIDE | 36234709 | exenatide |
| GLIMEPIRIDE | GLIMEPIRIDE | 1597759 | glimepiride |
| GLIMEPIRIDE | GLIMEPIRIDE | 43011831 | glimepiride |
| GLIPIZIDE | GLIPIZIDE | 19030445 | Glipizide |
| INSULIN | INSULIN | 46233975 | Insulin Lispro |
| INSULIN | INSULIN | 46233972 | Insulin Lispro |
| INSULIN | INSULIN | 46233970 | Insulin Lispro |

| | | | |
|---|---|---|---|
| GLIMEPIRIDE | GLIMEPIRIDE | 36234085 | glimepiride |
| EXENATIDE | EXENATIDE | 19083254 | exenatide |
| ACARBOSE | ACARBOSE | 19040970 | Acarbose |
| PIOGLITAZONE | PIOGLITAZONE | 1525218 | pioglitazone |
| INSULIN | INSULIN | 19078551 | insulin, isophane |
| GLYBURIDE | GLYBURIDE | 19077681 | Glyburide |
| EXENATIDE | EXENATIDE | 46234118 | exenatide |
| INSULIN | INSULIN | 19078553 | insulin, isophane |
| INSULIN | INSULIN | 19120790 | Insulin Glargine |
| METFORMIN | METFORMIN | 36235746 | Metformin |
| INSULIN | INSULIN | 1550023 | Insulin Lispro |
| INSULIN | INSULIN | 1513912 | Insulin Lispro |
| INSULIN | INSULIN | 46234045 | Insulin, Aspart, Human |
| GLIPIZIDE | GLIPIZIDE | 19077638 | Glipizide |
| METFORMIN | METFORMIN | 36235581 | Metformin |
| GLYBURIDE | GLYBURIDE | 36235750 | Glyburide |
| GLIMEPIRIDE | GLIMEPIRIDE | 19018043 | glimepiride |
| REPAGLINIDE | REPAGLINIDE | 1516770 | repaglinide |
| INSULIN | INSULIN | 40184293 | Insulin, Aspart Protamine, Human |
| METFORMIN | METFORMIN | 40164894 | Metformin |
| GLIMEPIRIDE | GLIMEPIRIDE | 1597760 | glimepiride |
| GLIPIZIDE | GLIPIZIDE | 40044251 | Glipizide |
| ACARBOSE | ACARBOSE | 19021786 | Acarbose |
| METFORMIN | METFORMIN | 40164890 | Metformin |
| METFORMIN | METFORMIN | 40163927 | Metformin |
| METFORMIN | METFORMIN | 40164940 | Metformin |
| GLYBURIDE | GLYBURIDE | 40165997 | Glyburide |
| GLIPIZIDE | GLIPIZIDE | 40166815 | Glipizide |
| INSULIN | INSULIN | 19009384 | insulin, isophane |
| METFORMIN | METFORMIN | 40231393 | Metformin |
| INSULIN | INSULIN | 36231459 | Regular Insulin, Human |

| | | | |
|---|---|---|---|
| METFORMIN | METFORMIN | 40164889 | Metformin |
| SAXAGLIPTIN | SAXAGLIPTIN | 40166042 | saxagliptin |
| GLIPIZIDE | GLIPIZIDE | 19079990 | Glipizide |
| GLIPIZIDE | GLIPIZIDE | 19030443 | Glipizide |
| INSULIN | INSULIN | 40052088 | Insulin Lispro |
| METFORMIN | METFORMIN | 40164881 | Metformin |
| CANAGLIFLOZIN | CANAGLIFLOZIN | 36246430 | canagliflozin |
| GLYBURIDE | GLYBURIDE | 40165998 | Glyburide |
| METFORMIN | METFORMIN | 36235581 | Metformin |
| GLIMEPIRIDE | GLIMEPIRIDE | 1597761 | glimepiride |
| INSULIN | INSULIN | 19120315 | insulin, isophane |
| PRAMLINTIDE | PRAMLINTIDE | 40165687 | Pramlintide |
| INSULIN | INSULIN | 1597001 | Regular Insulin, Human |
| INSULIN | INSULIN | 19078554 | Regular Insulin, Human |
| PIOGLITAZONE | PIOGLITAZONE | 19079293 | pioglitazone |
| METFORMIN | METFORMIN | 40153151 | Metformin |
| INSULIN | INSULIN | 19009384 | insulin, isophane |
| INSULIN | INSULIN | 1502905 | Insulin Glargine |
| PIOGLITAZONE | PIOGLITAZONE | 1525215 | pioglitazone |
| GLIPIZIDE | GLIPIZIDE | 19080793 | Glipizide |
| INSULIN | INSULIN | 1596977 | Regular Insulin, Human |
| GLIPIZIDE | GLIPIZIDE | 19077638 | Glipizide |
| INSULIN | INSULIN | 40176981 | Insulin, Aspart Protamine, Human |
| INSULIN | INSULIN | 35602724 | insulin degludec |
| METFORMIN | METFORMIN | 40164882 | Metformin |
| METFORMIN | METFORMIN | 40117785 | Metformin |
| INSULIN | INSULIN | 19062999 | insulin, isophane |
| GLYBURIDE | GLYBURIDE | 40168368 | Glyburide |
| GLYBURIDE | GLYBURIDE | 19077684 | Glyburide |
| GLIPIZIDE | GLIPIZIDE | 40166816 | Glipizide |
| ACARBOSE | ACARBOSE | 19047612 | Acarbose |
| INSULIN | INSULIN | 1567232 | Insulin, Aspart, Human |

| | | | |
|---|---|---|---|
| INSULIN | INSULIN | 1596959 | insulin, isophane |
| REPAGLINIDE | REPAGLINIDE | 1516771 | repaglinide |
| METFORMIN | METFORMIN | 40164930 | Metformin |
| INSULIN | INSULIN | 1596916 | insulin, isophane |
| GLIPIZIDE | GLIPIZIDE | 19077636 | Glipizide |
| PIOGLITAZONE | PIOGLITAZONE | 1525216 | pioglitazone |
| SITAGLIPTIN | SITAGLIPTIN | 19125040 | sitagliptin |
| GLIMEPIRIDE | GLIMEPIRIDE | 1597759 | glimepiride |
| GLIPIZIDE | GLIPIZIDE | 36218767 | Glipizide |
| INSULIN | INSULIN | 19078559 | Insulin Glargine |
| PIOGLITAZONE | PIOGLITAZONE | 1525217 | pioglitazone |
| INSULIN | INSULIN | 1502905 | Insulin Glargine |
| INSULIN | INSULIN | 40184293 | Insulin, Aspart Protamine, Human |
| BROMOCRIPTINE | BROMOCRIPTINE | 730551 | Bromocriptine |
| INSULIN | INSULIN | 1586344 | insulin, isophane |
| METFORMIN | METFORMIN | 19106521 | Metformin |
| INSULIN | INSULIN | 1596999 | Regular Insulin, Human |
| INSULIN | INSULIN | 1516981 | insulin detemir |
| METFORMIN | METFORMIN | 40164895 | Metformin |
| INSULIN | INSULIN | 19078558 | Insulin, Aspart, Human |
| INSULIN | INSULIN | 1596977 | Regular Insulin, Human |
| PIOGLITAZONE | PIOGLITAZONE | 1525220 | pioglitazone |
| ACARBOSE | ACARBOSE | 1529360 | Acarbose |
| GLIMEPIRIDE | GLIMEPIRIDE | 1597760 | glimepiride |
| METFORMIN | METFORMIN | 40063356 | Metformin |
| INSULIN | INSULIN | 19110219 | insulin, isophane |
| INSULIN | INSULIN | 19078553 | insulin, isophane |
| INSULIN | INSULIN | 19078557 | Regular Insulin, Human |
| GLIPIZIDE | GLIPIZIDE | 40166815 | Glipizide |
| GLIPIZIDE | GLIPIZIDE | 40166815 | Glipizide |
| PIOGLITAZONE | PIOGLITAZONE | 1525217 | pioglitazone |
| METFORMIN | METFORMIN | 40164898 | Metformin |
| INSULIN | INSULIN | 19078553 | insulin, isophane |

| | | | |
|---|---|---|---|
| INSULIN | INSULIN | 19071700 | Insulin Glargine |
| INSULIN | INSULIN | 46233946 | Insulin Lispro |
| GLIPIZIDE | GLIPIZIDE | 19030445 | Glipizide |
| METFORMIN | METFORMIN | 40164892 | Metformin |
| GLYBURIDE | GLYBURIDE | 40165994 | Glyburide |
| METFORMIN | METFORMIN | 40164929 | Metformin |
| GLYBURIDE | GLYBURIDE | 19077659 | Glyburide |
| SITAGLIPTIN | SITAGLIPTIN | 19125051 | sitagliptin |
| GLIPIZIDE | GLIPIZIDE | 19100559 | Glipizide |
| METFORMIN | METFORMIN | 40164901 | Metformin |
| INSULIN | INSULIN | 19110357 | Regular Insulin, Human |
| EXENATIDE | EXENATIDE | 19083253 | exenatide |
| METFORMIN | METFORMIN | 19113780 | Metformin |
| METFORMIN | METFORMIN | 40164889 | Metformin |
| GLYBURIDE | GLYBURIDE | 40166003 | Glyburide |
| INSULIN | INSULIN | 1502910 | Insulin Glargine |
| SITAGLIPTIN | SITAGLIPTIN | 19125051 | sitagliptin |
| METFORMIN | METFORMIN | 40164926 | Metformin |
| EXENATIDE | EXENATIDE | 1583727 | exenatide |
| DULAGLUTIDE | DULAGLUTIDE | 45774442 | dulaglutide |
| METFORMIN | METFORMIN | 36221230 | Metformin |
| EXENATIDE | EXENATIDE | 46234118 | exenatide |
| GLIMEPIRIDE | GLIMEPIRIDE | 36214984 | glimepiride |
| INSULIN | INSULIN | 35602731 | insulin degludec |
| INSULIN | INSULIN | 1550023 | Insulin Lispro |
| INSULIN | INSULIN | 40184293 | Insulin, Aspart Protamine, Human |
| METFORMIN | METFORMIN | 40164928 | Metformin |
| GLIMEPIRIDE | GLIMEPIRIDE | 43011831 | glimepiride |
| GLIPIZIDE | GLIPIZIDE | 19030445 | Glipizide |
| GLYBURIDE | GLYBURIDE | 19077659 | Glyburide |
| GLYBURIDE | GLYBURIDE | 40046201 | Glyburide |
| PIOGLITAZONE | PIOGLITAZONE | 36236324 | pioglitazone |
| GLYBURIDE | GLYBURIDE | 40166002 | Glyburide |

| | | | |
|---|---|---|---|
| SITAGLIPTIN | SITAGLIPTIN | 19125051 | sitagliptin |
| GLYBURIDE | GLYBURIDE | 40168371 | Glyburide |
| EXENATIDE | EXENATIDE | 19083253 | exenatide |
| INSULIN | INSULIN | 1567198 | Insulin, Aspart, Human |
| METFORMIN | METFORMIN | 1503297 | Metformin |
| SAXAGLIPTIN | SAXAGLIPTIN | 40166039 | saxagliptin |
| NATEGLINIDE | NATEGLINIDE | 1502830 | nateglinide |
| INSULIN | INSULIN | 19078610 | insulin, regular, beef-pork |
| INSULIN | INSULIN | 36236033 | Insulin Lispro |
| GLYBURIDE | GLYBURIDE | 40046201 | Glyburide |
| METFORMIN | METFORMIN | 1503297 | Metformin |
| INSULIN | INSULIN | 19078557 | Regular Insulin, Human |
| INSULIN | INSULIN | 19071700 | Insulin Glargine |
| GLIPIZIDE | GLIPIZIDE | 40165969 | Glipizide |
| GLYBURIDE | GLYBURIDE | 40168376 | Glyburide |
| BROMOCRIPTINE | BROMOCRIPTINE | 19083958 | Bromocriptine |
| METFORMIN | METFORMIN | 40164897 | Metformin |
| INSULIN | INSULIN | 1567198 | Insulin, Aspart, Human |
| NATEGLINIDE | NATEGLINIDE | 19071495 | nateglinide |
| INSULIN | INSULIN | 40051347 | Insulin Glargine |
| METFORMIN | METFORMIN | 40163924 | Metformin |
| GLYBURIDE | GLYBURIDE | 19106860 | Glyburide |
| GLIPIZIDE | GLIPIZIDE | 40165975 | Glipizide |
| METFORMIN | METFORMIN | 40163928 | Metformin |
| GLIPIZIDE | GLIPIZIDE | 40044251 | Glipizide |
| GLIPIZIDE | GLIPIZIDE | 36218767 | Glipizide |
| GLIPIZIDE | GLIPIZIDE | 36218767 | Glipizide |
| ACARBOSE | ACARBOSE | 1529352 | Acarbose |
| EXENATIDE | EXENATIDE | 19083253 | exenatide |
| GLYBURIDE | GLYBURIDE | 19077659 | Glyburide |
| INSULIN | INSULIN | 1516981 | insulin detemir |
| GLYBURIDE | GLYBURIDE | 40168370 | Glyburide |
| PIOGLITAZONE | PIOGLITAZONE | 1525218 | pioglitazone |
| INSULIN | INSULIN | 46233730 | Regular Insulin, Human |
| INSULIN | INSULIN | 40166274 | Insulin Lispro |
| INSULIN | INSULIN | 19120790 | Insulin Glargine |
| INSULIN | INSULIN | 36235841 | Regular Insulin, Human |

| | | | |
|---|---|---|---|
| METFORMIN | METFORMIN | 40164882 | Metformin |
| METFORMIN | METFORMIN | 40163928 | Metformin |
| INSULIN | INSULIN | 1513912 | Insulin Lispro |
| PIOGLITAZONE | PIOGLITAZONE | 1525221 | pioglitazone |
| METFORMIN | METFORMIN | 40163929 | Metformin |
| INSULIN | INSULIN | 19062999 | insulin, isophane |
| GLYBURIDE | GLYBURIDE | 19077684 | Glyburide |
| INSULIN | INSULIN | 1550023 | Insulin Lispro |
| METFORMIN | METFORMIN | 40063352 | Metformin |
| NATEGLINIDE | NATEGLINIDE | 36232423 | nateglinide |
| BROMOCRIPTINE | BROMOCRIPTINE | 730551 | Bromocriptine |
| INSULIN | INSULIN | 19095214 | insulin, isophane |
| INSULIN | INSULIN | 36229086 | Insulin Glargine |
| INSULIN | INSULIN | 1567198 | Insulin, Aspart, Human |
| INSULIN | INSULIN | 1596999 | Regular Insulin, Human |
| SITAGLIPTIN | SITAGLIPTIN | 19125051 | sitagliptin |
| INSULIN | INSULIN | 19110352 | insulin, isophane |
| ACARBOSE | ACARBOSE | 1529359 | Acarbose |
| INSULIN | INSULIN | 19078557 | Regular Insulin, Human |
| INSULIN | INSULIN | 1596955 | insulin, isophane |
| LIRAGLUTIDE | LIRAGLUTIDE | 40170913 | liraglutide |
| METFORMIN | METFORMIN | 40164939 | Metformin |
| INSULIN | INSULIN | 1550023 | Insulin Lispro |
| INSULIN | INSULIN | 35604094 | Insulin Glargine |
| INSULIN | INSULIN | 1596916 | insulin, isophane |
| GLYBURIDE | GLYBURIDE | 19110154 | Glyburide |
| INSULIN | INSULIN | 1567201 | Insulin, Aspart Protamine, Human |
| DAPAGLIFLOZIN | DAPAGLIFLOZIN | 44785473 | dapagliflozin |
| INSULIN | INSULIN | 1502910 | Insulin Glargine |
| METFORMIN | METFORMIN | 40164944 | Metformin |
| EXENATIDE | EXENATIDE | 1583730 | exenatide |

| | | | |
|---|---|---|---|
| GLIMEPIRIDE | GLIMEPIRIDE | 1597757 | glimepiride |
| GLIPIZIDE | GLIPIZIDE | 36235748 | Glipizide |
| INSULIN | INSULIN | 19131582 | Insulin, Glulisine, Human |
| METFORMIN | METFORMIN | 40063356 | Metformin |
| GLYBURIDE | GLYBURIDE | 36235750 | Glyburide |
| INSULIN | INSULIN | 40184294 | Insulin, Aspart Protamine, Human |
| PRAMLINTIDE | PRAMLINTIDE | 40165687 | Pramlintide |
| EMPAGLIFLOZIN | EMPAGLIFLOZIN | 45774895 | empagliflozin |
| METFORMIN | METFORMIN | 40164892 | Metformin |
| INSULIN | INSULIN | 1513912 | Insulin Lispro |
| GLYBURIDE | GLYBURIDE | 40166003 | Glyburide |
| INSULIN | INSULIN | 19120790 | Insulin Glargine |
| GLIPIZIDE | GLIPIZIDE | 19100559 | Glipizide |
| INSULIN | INSULIN | 1502905 | Insulin Glargine |
| GLYBURIDE | GLYBURIDE | 19030579 | Glyburide |
| INSULIN | INSULIN | 1596977 | Regular Insulin, Human |
| INSULIN | INSULIN | 1597000 | Regular Insulin, Human |
| INSULIN | INSULIN | 19100127 | insulin, isophane |
| INSULIN | INSULIN | 1513881 | Insulin Lispro |
| METFORMIN | METFORMIN | 40164891 | Metformin |
| INSULIN | INSULIN | 19131582 | Insulin, Glulisine, Human |
| GLIPIZIDE | GLIPIZIDE | 40165975 | Glipizide |
| INSULIN | INSULIN | 1550028 | Insulin Lispro |
| INSULIN | INSULIN | 19078557 | Regular Insulin, Human |
| INSULIN | INSULIN | 19120315 | insulin, isophane |
| GLIPIZIDE | GLIPIZIDE | 19030443 | Glipizide |
| INSULIN | INSULIN | 19078558 | Insulin, Aspart, Human |
| GLIMEPIRIDE | GLIMEPIRIDE | 1597757 | glimepiride |
| GLIPIZIDE | GLIPIZIDE | 40166815 | Glipizide |
| GLYBURIDE | GLYBURIDE | 1559684 | Glyburide |

| | | | |
|---|---|---|---|
| METFORMIN | METFORMIN | 40163924 | Metformin |
| SITAGLIPTIN | SITAGLIPTIN | 40243628 | sitagliptin |
| ACARBOSE | ACARBOSE | 19120257 | Acarbose |
| METFORMIN | METFORMIN | 40164920 | Metformin |
| PIOGLITAZONE | PIOGLITAZONE | 1525217 | pioglitazone |
| GLIMEPIRIDE | GLIMEPIRIDE | 1597759 | glimepiride |
| INSULIN | INSULIN | 1567232 | Insulin, Aspart, Human |
| NATEGLINIDE | NATEGLINIDE | 36232423 | nateglinide |
| INSULIN | INSULIN | 46234049 | Insulin, Aspart, Human |
| GLIPIZIDE | GLIPIZIDE | 19080793 | Glipizide |
| INSULIN | INSULIN | 1502910 | Insulin Glargine |
| GLIPIZIDE | GLIPIZIDE | 19030445 | Glipizide |
| INSULIN | INSULIN | 19110356 | Regular Insulin, Human |
| GLYBURIDE | GLYBURIDE | 36230035 | Glyburide |
| METFORMIN | METFORMIN | 40164930 | Metformin |
| GLIPIZIDE | GLIPIZIDE | 19030443 | Glipizide |
| INSULIN | INSULIN | 40051347 | Insulin Glargine |
| PRAMLINTIDE | PRAMLINTIDE | 36243018 | Pramlintide |
| INSULIN | INSULIN | 19120315 | insulin, isophane |
| GLIPIZIDE | GLIPIZIDE | 36235748 | Glipizide |
| METFORMIN | METFORMIN | 40163928 | Metformin |
| LINAGLIPTIN | LINAGLIPTIN | 42708087 | Linagliptin |
| INSULIN | INSULIN | 46221557 | Insulin Glargine |
| EXENATIDE | EXENATIDE | 19083253 | exenatide |
| LINAGLIPTIN | LINAGLIPTIN | 36229565 | Linagliptin |
| METFORMIN | METFORMIN | 19113780 | Metformin |
| INSULIN | INSULIN | 1513849 | Insulin, Zinc, Human |
| INSULIN | INSULIN | 40052088 | Insulin Lispro |
| METFORMIN | METFORMIN | 40164905 | Metformin |
| INSULIN | INSULIN | 1516979 | insulin detemir |
| INSULIN | INSULIN | 1597000 | Regular Insulin, Human |
| LIRAGLUTIDE | LIRAGLUTIDE | 40170911 | liraglutide |
| INSULIN | INSULIN | 1567198 | Insulin, Aspart, Human |

| | | | |
|---|---|---|---|
| INSULIN | INSULIN | 1567198 | Insulin, Aspart, Human |
| ACARBOSE | ACARBOSE | 19021787 | Acarbose |
| INSULIN | INSULIN | 19062999 | insulin, isophane |
| INSULIN | INSULIN | 1544873 | Insulin, Glulisine, Human |
| EXENATIDE | EXENATIDE | 19135404 | exenatide |
| METFORMIN | METFORMIN | 40229050 | Metformin |
| METFORMIN | METFORMIN | 40063352 | Metformin |
| INSULIN | INSULIN | 19014975 | insulin, isophane |
| INSULIN | INSULIN | 46233730 | Regular Insulin, Human |
| ROSIGLITAZONE | ROSIGLITAZONE | 19079465 | rosiglitazone |
| LIRAGLUTIDE | LIRAGLUTIDE | 40169223 | liraglutide |
| INSULIN | INSULIN | 1567198 | Insulin, Aspart, Human |
| GLYBURIDE | GLYBURIDE | 40165998 | Glyburide |
| METFORMIN | METFORMIN | 36236322 | Metformin |
| ROSIGLITAZONE | ROSIGLITAZONE | 1547506 | rosiglitazone |
| INSULIN | INSULIN | 19095218 | Regular Insulin, Human |
| INSULIN | INSULIN | 40176981 | Insulin, Aspart Protamine, Human |
| METFORMIN | METFORMIN | 40164944 | Metformin |
| GLYBURIDE | GLYBURIDE | 19030580 | Glyburide |
| GLYBURIDE | GLYBURIDE | 1559761 | Glyburide |
| INSULIN | INSULIN | 40052088 | Insulin Lispro |
| INSULIN | INSULIN | 40166274 | Insulin Lispro |
| GLIMEPIRIDE | GLIMEPIRIDE | 1597757 | glimepiride |
| ROSIGLITAZONE | ROSIGLITAZONE | 36229365 | rosiglitazone |
| INSULIN | INSULIN | 19058398 | Insulin Lispro |
| GLIMEPIRIDE | GLIMEPIRIDE | 1597760 | glimepiride |
| SITAGLIPTIN | SITAGLIPTIN | 19125041 | sitagliptin |
| GLIMEPIRIDE | GLIMEPIRIDE | 1597773 | glimepiride |
| INSULIN | INSULIN | 19078554 | Regular Insulin, Human |
| REPAGLINIDE | REPAGLINIDE | 36240429 | repaglinide |
| INSULIN | INSULIN | 1550023 | Insulin Lispro |
| DAPAGLIFLOZIN | DAPAGLIFLOZIN | 44785473 | dapagliflozin |
| INSULIN | INSULIN | 19062999 | insulin, isophane |

| | | | |
|---|---|---|---|
| LIRAGLUTIDE | LIRAGLUTIDE | 40169223 | liraglutide |
| GLIPIZIDE | GLIPIZIDE | 19134642 | Glipizide |
| INSULIN | INSULIN | 1502905 | Insulin Glargine |
| GLIPIZIDE | GLIPIZIDE | 40167636 | Glipizide |
| METFORMIN | METFORMIN | 40163929 | Metformin |
| MIGLITOL | MIGLITOL | 19067200 | miglitol |
| ACARBOSE | ACARBOSE | 19040969 | Acarbose |
| PRAMLINTIDE | PRAMLINTIDE | 40165687 | Pramlintide |
| INSULIN | INSULIN | 35604094 | Insulin Glargine |
| INSULIN | INSULIN | 40051347 | Insulin Glargine |
| INSULIN | INSULIN | 19078553 | insulin, isophane |
| INSULIN | INSULIN | 36233154 | Insulin, Glulisine, Human |
| METFORMIN | METFORMIN | 40164892 | Metformin |
| INSULIN | INSULIN | 1597012 | Regular Insulin, Human |
| INSULIN | INSULIN | 19111795 | insulin, isophane |
| INSULIN | INSULIN | 1513916 | Insulin Lispro |
| GLIMEPIRIDE | GLIMEPIRIDE | 1597760 | glimepiride |
| INSULIN | INSULIN | 19078553 | insulin, isophane |
| SITAGLIPTIN | SITAGLIPTIN | 42900896 | sitagliptin |
| BROMOCRIPTINE | BROMOCRIPTINE | 19007101 | Bromocriptine |
| INSULIN | INSULIN | 40184293 | Insulin, Aspart Protamine, Human |
| GLIPIZIDE | GLIPIZIDE | 19030445 | Glipizide |
| GLIMEPIRIDE | GLIMEPIRIDE | 1597761 | glimepiride |
| GLIMEPIRIDE | GLIMEPIRIDE | 1597761 | glimepiride |
| INSULIN | INSULIN | 1550023 | Insulin Lispro |
| GLIPIZIDE | GLIPIZIDE | 19134639 | Glipizide |
| INSULIN | INSULIN | 1502905 | Insulin Glargine |
| GLIPIZIDE | GLIPIZIDE | 40166814 | Glipizide |
| INSULIN | INSULIN | 19078557 | Regular Insulin, Human |
| METFORMIN | METFORMIN | 1503297 | Metformin |
| INSULIN | INSULIN | 1586370 | Ultralente Insulin, Human |
| PIOGLITAZONE | PIOGLITAZONE | 1525218 | pioglitazone |

| | | | |
|---|---|---|---|
| INSULIN | INSULIN | 1596999 | Regular Insulin, Human |
| INSULIN | INSULIN | 1513912 | Insulin Lispro |
| LINAGLIPTIN | LINAGLIPTIN | 40239219 | Linagliptin |
| METFORMIN | METFORMIN | 36235747 | Metformin |
| INSULIN | INSULIN | 1550027 | Insulin Lispro |
| INSULIN | INSULIN | 1513881 | Insulin Lispro |
| DULAGLUTIDE | DULAGLUTIDE | 45774447 | dulaglutide |
| EMPAGLIFLOZIN | EMPAGLIFLOZIN | 45774758 | empagliflozin |
| GLYBURIDE | GLYBURIDE | 19030580 | Glyburide |
| INSULIN | INSULIN | 19078554 | Regular Insulin, Human |
| INSULIN | INSULIN | 1516979 | insulin detemir |
| GLIPIZIDE | GLIPIZIDE | 19079990 | Glipizide |
| ROSIGLITAZONE | ROSIGLITAZONE | 19079465 | rosiglitazone |
| PIOGLITAZONE | PIOGLITAZONE | 19079293 | pioglitazone |
| INSULIN | INSULIN | 40176981 | Insulin, Aspart Protamine, Human |
| INSULIN | INSULIN | 19071700 | Insulin Glargine |
| GLYBURIDE | GLYBURIDE | 1559685 | Glyburide |
| PIOGLITAZONE | PIOGLITAZONE | 1525220 | pioglitazone |
| INSULIN | INSULIN | 1567198 | Insulin, Aspart, Human |
| ACARBOSE | ACARBOSE | 19023063 | Acarbose |
| NATEGLINIDE | NATEGLINIDE | 1502829 | nateglinide |
| DAPAGLIFLOZIN | DAPAGLIFLOZIN | 36247907 | dapagliflozin |
| DULAGLUTIDE | DULAGLUTIDE | 45774442 | dulaglutide |
| INSULIN | INSULIN | 1586344 | insulin, isophane |
| INSULIN | INSULIN | 1502910 | Insulin Glargine |
| INSULIN | INSULIN | 40184294 | Insulin, Aspart Protamine, Human |
| METFORMIN | METFORMIN | 36235752 | Metformin |
| INSULIN | INSULIN | 1550027 | Insulin Lispro |
| GLIPIZIDE | GLIPIZIDE | 19100559 | Glipizide |
| BROMOCRIPTINE | BROMOCRIPTINE | 19007101 | Bromocriptine |
| INSULIN | INSULIN | 1550023 | Insulin Lispro |
| INSULIN | INSULIN | 1596977 | Regular Insulin, Human |
| GLIPIZIDE | GLIPIZIDE | 19030443 | Glipizide |
| ACARBOSE | ACARBOSE | 19021312 | Acarbose |

| | | | |
|---|---|---|---|
| INSULIN | INSULIN | 1567198 | Insulin, Aspart, Human |
| INSULIN | INSULIN | 1597012 | Regular Insulin, Human |
| INSULIN | INSULIN | 40184293 | Insulin, Aspart Protamine, Human |
| EXENATIDE | EXENATIDE | 42708079 | exenatide |
| GLIMEPIRIDE | GLIMEPIRIDE | 36214984 | glimepiride |
| ROSIGLITAZONE | ROSIGLITAZONE | 1547506 | rosiglitazone |
| NATEGLINIDE | NATEGLINIDE | 1502829 | nateglinide |
| EXENATIDE | EXENATIDE | 45774533 | exenatide |
| INSULIN | INSULIN | 19078633 | Insulin, Regular, Pork |
| NATEGLINIDE | NATEGLINIDE | 19071495 | nateglinide |
| METFORMIN | METFORMIN | 40164882 | Metformin |
| LIRAGLUTIDE | LIRAGLUTIDE | 40170913 | liraglutide |
| INSULIN | INSULIN | 46234049 | Insulin, Aspart, Human |
| PIOGLITAZONE | PIOGLITAZONE | 40099898 | pioglitazone |
| INSULIN | INSULIN | 19058398 | Insulin Lispro |
| INSULIN | INSULIN | 19078554 | Regular Insulin, Human |
| METFORMIN | METFORMIN | 40164946 | Metformin |
| METFORMIN | METFORMIN | 36235746 | Metformin |
| LIRAGLUTIDE | LIRAGLUTIDE | 45892686 | liraglutide |
| INSULIN | INSULIN | 46234043 | Insulin, Aspart, Human |
| INSULIN | INSULIN | 36229086 | Insulin Glargine |
| INSULIN | INSULIN | 19078557 | Regular Insulin, Human |
| EXENATIDE | EXENATIDE | 36234709 | exenatide |
| INSULIN | INSULIN | 40166274 | Insulin Lispro |
| INSULIN | INSULIN | 19018822 | Insulin Lispro |
| ACARBOSE | ACARBOSE | 1529358 | Acarbose |
| ROSIGLITAZONE | ROSIGLITAZONE | 1547506 | rosiglitazone |
| METFORMIN | METFORMIN | 40063354 | Metformin |
| GLYBURIDE | GLYBURIDE | 19077681 | Glyburide |
| ROSIGLITAZONE | ROSIGLITAZONE | 19079465 | rosiglitazone |
| INSULIN | INSULIN | 46234241 | Regular Insulin, Human |
| SITAGLIPTIN | SITAGLIPTIN | 36221232 | sitagliptin |
| METFORMIN | METFORMIN | 40164926 | Metformin |
| PIOGLITAZONE | PIOGLITAZONE | 1525216 | pioglitazone |
| SAXAGLIPTIN | SAXAGLIPTIN | 40166042 | saxagliptin |

| | | | |
|---|---|---|---|
| INSULIN | INSULIN | 19018822 | Insulin Lispro |
| SAXAGLIPTIN | SAXAGLIPTIN | 36229291 | saxagliptin |
| INSULIN | INSULIN | 19078554 | Regular Insulin, Human |
| SITAGLIPTIN | SITAGLIPTIN | 19125049 | sitagliptin |
| CANAGLIFLOZIN | CANAGLIFLOZIN | 45774717 | canagliflozin |
| MIGLITOL | MIGLITOL | 1510207 | miglitol |
| GLYBURIDE | GLYBURIDE | 1559685 | Glyburide |
| INSULIN | INSULIN | 46234241 | Regular Insulin, Human |
| INSULIN | INSULIN | 1550028 | Insulin Lispro |
| METFORMIN | METFORMIN | 36235746 | Metformin |
| REPAGLINIDE | REPAGLINIDE | 19081914 | repaglinide |
| INSULIN | INSULIN | 1596999 | Regular Insulin, Human |
| PIOGLITAZONE | PIOGLITAZONE | 1525221 | pioglitazone |
| INSULIN | INSULIN | 19078610 | insulin, regular, beef-pork |
| INSULIN | INSULIN | 19078554 | Regular Insulin, Human |
| EXENATIDE | EXENATIDE | 1583727 | exenatide |
| SITAGLIPTIN | SITAGLIPTIN | 19125043 | sitagliptin |
| INSULIN | INSULIN | 19062999 | insulin, isophane |
| GLIMEPIRIDE | GLIMEPIRIDE | 19018043 | glimepiride |
| INSULIN | INSULIN | 1544873 | Insulin, Glulisine, Human |
| GLIMEPIRIDE | GLIMEPIRIDE | 19117199 | glimepiride |
| INSULIN | INSULIN | 19110356 | Regular Insulin, Human |
| EXENATIDE | EXENATIDE | 19083254 | exenatide |
| INSULIN | INSULIN | 19078557 | Regular Insulin, Human |
| INSULIN | INSULIN | 19120790 | Insulin Glargine |
| GLIPIZIDE | GLIPIZIDE | 19080793 | Glipizide |
| INSULIN | INSULIN | 19111797 | insulin, isophane |
| INSULIN | INSULIN | 1550028 | Insulin Lispro |
| INSULIN | INSULIN | 1596941 | insulin, isophane |
| ROSIGLITAZONE | ROSIGLITAZONE | 1547505 | rosiglitazone |
| NATEGLINIDE | NATEGLINIDE | 36223777 | nateglinide |
| METFORMIN | METFORMIN | 40164923 | Metformin |

| | | | |
|---|---|---|---|
| INSULIN | INSULIN | 19135263 | insulin, isophane |
| METFORMIN | METFORMIN | 42705990 | Metformin |
| INSULIN | INSULIN | 19062999 | insulin, isophane |
| EXENATIDE | EXENATIDE | 1583727 | exenatide |
| METFORMIN | METFORMIN | 40163923 | Metformin |
| METFORMIN | METFORMIN | 40164917 | Metformin |
| INSULIN | INSULIN | 36236032 | Insulin Lispro |
| GLIMEPIRIDE | GLIMEPIRIDE | 19100593 | glimepiride |
| GLIPIZIDE | GLIPIZIDE | 40167636 | Glipizide |
| ROSIGLITAZONE | ROSIGLITAZONE | 1547554 | rosiglitazone |
| GLYBURIDE | GLYBURIDE | 36230035 | Glyburide |
| INSULIN | INSULIN | 36233552 | insulin, isophane |
| INSULIN | INSULIN | 1502905 | Insulin Glargine |
| GLIPIZIDE | GLIPIZIDE | 19077638 | Glipizide |
| BROMOCRIPTINE | BROMOCRIPTINE | 19006288 | Bromocriptine |
| INSULIN | INSULIN | 1596999 | Regular Insulin, Human |
| METFORMIN | METFORMIN | 40164940 | Metformin |
| INSULIN | INSULIN | 19078557 | Regular Insulin, Human |
| GLIMEPIRIDE | GLIMEPIRIDE | 1597757 | glimepiride |
| INSULIN | INSULIN | 1596955 | insulin, isophane |
| INSULIN | INSULIN | 19120790 | Insulin Glargine |
| PIOGLITAZONE | PIOGLITAZONE | 1525218 | pioglitazone |
| INSULIN | INSULIN | 19071700 | Insulin Glargine |
| METFORMIN | METFORMIN | 40164898 | Metformin |
| GLYBURIDE | GLYBURIDE | 19045628 | Glyburide |
| BROMOCRIPTINE | BROMOCRIPTINE | 40016899 | Bromocriptine |
| SITAGLIPTIN | SITAGLIPTIN | 19125048 | sitagliptin |
| EXENATIDE | EXENATIDE | 19083254 | exenatide |
| GLIPIZIDE | GLIPIZIDE | 40165969 | Glipizide |
| GLYBURIDE | GLYBURIDE | 19077682 | Glyburide |
| GLIPIZIDE | GLIPIZIDE | 40139022 | Glipizide |
| NATEGLINIDE | NATEGLINIDE | 19071495 | nateglinide |

| | | | |
|---|---|---|---|
| INSULIN | INSULIN | 19078554 | Regular Insulin, Human |
| INSULIN | INSULIN | 1596972 | insulin, isophane |
| GLYBURIDE | GLYBURIDE | 19045625 | Glyburide |
| GLYBURIDE | GLYBURIDE | 40166001 | Glyburide |
| DULAGLUTIDE | DULAGLUTIDE | 45774442 | dulaglutide |
| METFORMIN | METFORMIN | 40164923 | Metformin |
| INSULIN | INSULIN | 19120315 | insulin, isophane |
| INSULIN | INSULIN | 19078552 | insulin, isophane |
| INSULIN | INSULIN | 19044971 | insulin detemir |
| INSULIN | INSULIN | 1596957 | insulin, isophane |
| GLIMEPIRIDE | GLIMEPIRIDE | 43011498 | glimepiride |
| INSULIN | INSULIN | 46234045 | Insulin, Aspart, Human |
| INSULIN | INSULIN | 35602731 | insulin degludec |
| EXENATIDE | EXENATIDE | 19083253 | exenatide |
| INSULIN | INSULIN | 46234043 | Insulin, Aspart, Human |
| INSULIN | INSULIN | 1550027 | Insulin Lispro |
| GLIPIZIDE | GLIPIZIDE | 1560171 | Glipizide |
| INSULIN | INSULIN | 1596960 | insulin, isophane |
| INSULIN | INSULIN | 19120790 | Insulin Glargine |
| EXENATIDE | EXENATIDE | 1583730 | exenatide |
| GLYBURIDE | GLYBURIDE | 19079986 | Glyburide |
| INSULIN | INSULIN | 46233975 | Insulin Lispro |
| METFORMIN | METFORMIN | 19106521 | Metformin |
| INSULIN | INSULIN | 40176981 | Insulin, Aspart Protamine, Human |
| GLIPIZIDE | GLIPIZIDE | 40160874 | Glipizide |
| LIRAGLUTIDE | LIRAGLUTIDE | 40169223 | liraglutide |
| EXENATIDE | EXENATIDE | 45774533 | exenatide |
| METFORMIN | METFORMIN | 40164891 | Metformin |
| INSULIN | INSULIN | 19062999 | insulin, isophane |
| INSULIN | INSULIN | 35604094 | Insulin Glargine |

| | | | |
|---|---|---|---|
| REPAGLINIDE | REPAGLINIDE | 19047663 | repaglinide |
| GLYBURIDE | GLYBURIDE | 1559685 | Glyburide |
| INSULIN | INSULIN | 1567198 | Insulin, Aspart, Human |
| ROSIGLITAZONE | ROSIGLITAZONE | 19079465 | rosiglitazone |
| EXENATIDE | EXENATIDE | 1583727 | exenatide |
| INSULIN | INSULIN | 1567232 | Insulin, Aspart, Human |
| GLIPIZIDE | GLIPIZIDE | 19134624 | Glipizide |
| INSULIN | INSULIN | 19078610 | insulin, regular, beef-pork |
| INSULIN | INSULIN | 36231459 | Regular Insulin, Human |
| GLYBURIDE | GLYBURIDE | 40165995 | Glyburide |
| EMPAGLIFLOZIN | EMPAGLIFLOZIN | 45774895 | empagliflozin |
| GLYBURIDE | GLYBURIDE | 19077684 | Glyburide |
| INSULIN | INSULIN | 1596941 | insulin, isophane |
| LINAGLIPTIN | LINAGLIPTIN | 40239218 | Linagliptin |
| INSULIN | INSULIN | 1596957 | insulin, isophane |
| ROSIGLITAZONE | ROSIGLITAZONE | 36229365 | rosiglitazone |
| PRAMLINTIDE | PRAMLINTIDE | 40165688 | Pramlintide |
| SITAGLIPTIN | SITAGLIPTIN | 19125043 | sitagliptin |
| SITAGLIPTIN | SITAGLIPTIN | 19125043 | sitagliptin |
| METFORMIN | METFORMIN | 40164947 | Metformin |
| INSULIN | INSULIN | 35602724 | insulin degludec |
| INSULIN | INSULIN | 19116559 | insulin, isophane |
| INSULIN | INSULIN | 1502905 | Insulin Glargine |
| CANAGLIFLOZIN | CANAGLIFLOZIN | 43526472 | canagliflozin |
| METFORMIN | METFORMIN | 1503297 | Metformin |
| GLIPIZIDE | GLIPIZIDE | 40166815 | Glipizide |
| GLIMEPIRIDE | GLIMEPIRIDE | 1597760 | glimepiride |
| INSULIN | INSULIN | 19078554 | Regular Insulin, Human |
| LINAGLIPTIN | LINAGLIPTIN | 40239219 | Linagliptin |
| INSULIN | INSULIN | 36231456 | insulin, isophane |
| INSULIN | INSULIN | 1586345 | insulin, isophane |
| EXENATIDE | EXENATIDE | 19083254 | exenatide |
| EXENATIDE | EXENATIDE | 19083254 | exenatide |
| GLIPIZIDE | GLIPIZIDE | 19079990 | Glipizide |

| | | | |
|---|---|---|---|
| SAXAGLIPTIN | SAXAGLIPTIN | 40166040 | saxagliptin |
| GLIPIZIDE | GLIPIZIDE | 19030445 | Glipizide |
| GLIPIZIDE | GLIPIZIDE | 19079990 | Glipizide |
| PRAMLINTIDE | PRAMLINTIDE | 40165687 | Pramlintide |
| GLIMEPIRIDE | GLIMEPIRIDE | 1597772 | glimepiride |
| METFORMIN | METFORMIN | 40164897 | Metformin |
| METFORMIN | METFORMIN | 40164927 | Metformin |
| CANAGLIFLOZIN | CANAGLIFLOZIN | 43526468 | canagliflozin |
| INSULIN | INSULIN | 36236032 | Insulin Lispro |
| INSULIN | INSULIN | 1596952 | insulin, isophane |
| GLYBURIDE | GLYBURIDE | 40165995 | Glyburide |
| EXENATIDE | EXENATIDE | 1583730 | exenatide |
| METFORMIN | METFORMIN | 40164944 | Metformin |
| INSULIN | INSULIN | 40176981 | Insulin, Aspart Protamine, Human |
| GLIPIZIDE | GLIPIZIDE | 19030443 | Glipizide |
| ACARBOSE | ACARBOSE | 19047612 | Acarbose |
| GLIMEPIRIDE | GLIMEPIRIDE | 36214984 | glimepiride |
| INSULIN | INSULIN | 19120790 | Insulin Glargine |
| INSULIN | INSULIN | 1596941 | insulin, isophane |
| INSULIN | INSULIN | 46233970 | Insulin Lispro |
| PIOGLITAZONE | PIOGLITAZONE | 1525217 | pioglitazone |
| EXENATIDE | EXENATIDE | 42708079 | exenatide |
| INSULIN | INSULIN | 1550027 | Insulin Lispro |
| ROSIGLITAZONE | ROSIGLITAZONE | 1547507 | rosiglitazone |
| EXENATIDE | EXENATIDE | 42708079 | exenatide |
| INSULIN | INSULIN | 19110219 | insulin, isophane |
| EXENATIDE | EXENATIDE | 1583728 | exenatide |
| GLIPIZIDE | GLIPIZIDE | 19134624 | Glipizide |
| METFORMIN | METFORMIN | 40164905 | Metformin |
| ALBIGLUTIDE | ALBIGLUTIDE | 44816086 | albiglutide |
| INSULIN | INSULIN | 36224228 | insulin detemir |
| MIGLITOL | MIGLITOL | 19067231 | miglitol |
| DULAGLUTIDE | DULAGLUTIDE | 45774448 | dulaglutide |
| METFORMIN | METFORMIN | 40164917 | Metformin |

| | | | |
|---|---|---|---|
| SITAGLIPTIN | SITAGLIPTIN | 42901620 | sitagliptin |
| INSULIN | INSULIN | 1550028 | Insulin Lispro |
| GLIMEPIRIDE | GLIMEPIRIDE | 36234085 | glimepiride |
| PIOGLITAZONE | PIOGLITAZONE | 36236324 | pioglitazone |
| INSULIN | INSULIN | 19071700 | Insulin Glargine |
| EXENATIDE | EXENATIDE | 45774533 | exenatide |
| GLYBURIDE | GLYBURIDE | 19106976 | Glyburide |
| ACARBOSE | ACARBOSE | 19040970 | Acarbose |
| INSULIN | INSULIN | 1596955 | insulin, isophane |
| GLYBURIDE | GLYBURIDE | 36213841 | Glyburide |
| INSULIN | INSULIN | 19135263 | insulin, isophane |
| GLIMEPIRIDE | GLIMEPIRIDE | 1597756 | glimepiride |
| LINAGLIPTIN | LINAGLIPTIN | 40239219 | Linagliptin |
| INSULIN | INSULIN | 19120315 | insulin, isophane |
| METFORMIN | METFORMIN | 40164947 | Metformin |
| INSULIN | INSULIN | 46234043 | Insulin, Aspart, Human |
| METFORMIN | METFORMIN | 40063352 | Metformin |
| METFORMIN | METFORMIN | 40164923 | Metformin |
| GLYBURIDE | GLYBURIDE | 19079986 | Glyburide |
| EXENATIDE | EXENATIDE | 19083254 | exenatide |
| GLYBURIDE | GLYBURIDE | 36231169 | Glyburide |
| GLIPIZIDE | GLIPIZIDE | 19134624 | Glipizide |
| METFORMIN | METFORMIN | 40164892 | Metformin |
| INSULIN | INSULIN | 1502905 | Insulin Glargine |
| INSULIN | INSULIN | 1502905 | Insulin Glargine |
| INSULIN | INSULIN | 1567198 | Insulin, Aspart, Human |
| GLIPIZIDE | GLIPIZIDE | 40166815 | Glipizide |
| ACARBOSE | ACARBOSE | 1529331 | Acarbose |
| SITAGLIPTIN | SITAGLIPTIN | 40136769 | sitagliptin |
| INSULIN | INSULIN | 36231450 | Insulin, Aspart, Human |
| INSULIN | INSULIN | 1550023 | Insulin Lispro |
| INSULIN | INSULIN | 1513912 | Insulin Lispro |
| INSULIN | INSULIN | 1550028 | Insulin Lispro |
| INSULIN | INSULIN | 19078527 | Insulin, Zinc, Human |

| | | | |
|---|---|---|---|
| INSULIN | INSULIN | 19111795 | insulin, isophane |
| GLYBURIDE | GLYBURIDE | 19060409 | Glyburide |
| INSULIN | INSULIN | 36235840 | insulin, isophane |
| INSULIN | INSULIN | 1513912 | Insulin Lispro |
| BROMOCRIPTINE | BROMOCRIPTINE | 730572 | Bromocriptine |
| INSULIN | INSULIN | 1567198 | Insulin, Aspart, Human |
| GLYBURIDE | GLYBURIDE | 19106860 | Glyburide |
| GLYBURIDE | GLYBURIDE | 19030575 | Glyburide |
| INSULIN | INSULIN | 40184294 | Insulin, Aspart Protamine, Human |
| EXENATIDE | EXENATIDE | 1583727 | exenatide |
| METFORMIN | METFORMIN | 40164895 | Metformin |
| GLYBURIDE | GLYBURIDE | 40165997 | Glyburide |
| PIOGLITAZONE | PIOGLITAZONE | 1525216 | pioglitazone |
| INSULIN | INSULIN | 36229086 | Insulin Glargine |
| MIGLITOL | MIGLITOL | 1510206 | miglitol |
| INSULIN | INSULIN | 1596960 | insulin, isophane |
| EXENATIDE | EXENATIDE | 46234118 | exenatide |
| INSULIN | INSULIN | 40184293 | Insulin, Aspart Protamine, Human |
| GLYBURIDE | GLYBURIDE | 36233495 | Glyburide |
| INSULIN | INSULIN | 1596977 | Regular Insulin, Human |
| GLIMEPIRIDE | GLIMEPIRIDE | 19017144 | glimepiride |
| SITAGLIPTIN | SITAGLIPTIN | 40243629 | sitagliptin |
| GLYBURIDE | GLYBURIDE | 40165995 | Glyburide |
| GLYBURIDE | GLYBURIDE | 19077684 | Glyburide |
| EXENATIDE | EXENATIDE | 19083253 | exenatide |
| REPAGLINIDE | REPAGLINIDE | 36240429 | repaglinide |
| DAPAGLIFLOZIN | DAPAGLIFLOZIN | 45775999 | dapagliflozin |
| INSULIN | INSULIN | 1596999 | Regular Insulin, Human |
| INSULIN | INSULIN | 19095211 | Insulin Lispro |
| METFORMIN | METFORMIN | 36235746 | Metformin |
| INSULIN | INSULIN | 1596977 | Regular Insulin, Human |

| | | | |
|---|---|---|---|
| METFORMIN | METFORMIN | 40163924 | Metformin |
| CHLORPROPAMIDE | CHLORPROPAMIDE | 1594975 | Chlorpropamide |
| INSULIN | INSULIN | 1596998 | Regular Insulin, Human |
| INSULIN | INSULIN | 19009384 | insulin, isophane |
| NATEGLINIDE | NATEGLINIDE | 1502827 | nateglinide |
| INSULIN | INSULIN | 19078530 | insulin, isophane |
| INSULIN | INSULIN | 19078555 | Regular Insulin, Human |
| ALBIGLUTIDE | ALBIGLUTIDE | 44816094 | albiglutide |
| NATEGLINIDE | NATEGLINIDE | 1502827 | nateglinide |
| INSULIN | INSULIN | 36227902 | Insulin Lispro |
| INSULIN | INSULIN | 36224228 | insulin detemir |
| METFORMIN | METFORMIN | 19088818 | Metformin |
| INSULIN | INSULIN | 19078553 | insulin, isophane |
| GLYBURIDE | GLYBURIDE | 19106860 | Glyburide |
| ROSIGLITAZONE | ROSIGLITAZONE | 1547507 | rosiglitazone |
| INSULIN | INSULIN | 1550023 | Insulin Lispro |
| EMPAGLIFLOZIN | EMPAGLIFLOZIN | 45892179 | empagliflozin |
| INSULIN | INSULIN | 40176981 | Insulin, Aspart Protamine, Human |
| LINAGLIPTIN | LINAGLIPTIN | 40239219 | Linagliptin |
| INSULIN | INSULIN | 46233975 | Insulin Lispro |
| EMPAGLIFLOZIN | EMPAGLIFLOZIN | 36246886 | empagliflozin |
| INSULIN | INSULIN | 19100127 | insulin, isophane |
| METFORMIN | METFORMIN | 40164904 | Metformin |
| METFORMIN | METFORMIN | 40063356 | Metformin |
| METFORMIN | METFORMIN | 40164905 | Metformin |
| DULAGLUTIDE | DULAGLUTIDE | 45774442 | dulaglutide |
| GLIPIZIDE | GLIPIZIDE | 19134624 | Glipizide |
| METFORMIN | METFORMIN | 1503297 | Metformin |
| INSULIN | INSULIN | 19078553 | insulin, isophane |
| SITAGLIPTIN | SITAGLIPTIN | 19125047 | sitagliptin |
| METFORMIN | METFORMIN | 40164881 | Metformin |
| INSULIN | INSULIN | 19110355 | insulin, isophane |

| | | | |
|---|---|---|---|
| INSULIN | INSULIN | 19120315 | insulin, isophane |
| NATEGLINIDE | NATEGLINIDE | 40058199 | nateglinide |
| METFORMIN | METFORMIN | 40231393 | Metformin |
| EXENATIDE | EXENATIDE | 1583727 | exenatide |
| INSULIN | INSULIN | 19110355 | insulin, isophane |
| GLIPIZIDE | GLIPIZIDE | 40165976 | Glipizide |
| METFORMIN | METFORMIN | 40164892 | Metformin |
| PIOGLITAZONE | PIOGLITAZONE | 1525216 | pioglitazone |
| ALBIGLUTIDE | ALBIGLUTIDE | 44816274 | albiglutide |
| INSULIN | INSULIN | 1550023 | Insulin Lispro |
| GLIPIZIDE | GLIPIZIDE | 36218768 | Glipizide |
| INSULIN | INSULIN | 19018822 | Insulin Lispro |
| INSULIN | INSULIN | 1550027 | Insulin Lispro |
| CANAGLIFLOZIN | CANAGLIFLOZIN | 43526472 | canagliflozin |
| INSULIN | INSULIN | 1550028 | Insulin Lispro |
| BROMOCRIPTINE | BROMOCRIPTINE | 19007070 | Bromocriptine |
| REPAGLINIDE | REPAGLINIDE | 19047663 | repaglinide |
| METFORMIN | METFORMIN | 19088818 | Metformin |
| GLYBURIDE | GLYBURIDE | 40168374 | Glyburide |
| GLYBURIDE | GLYBURIDE | 19063991 | Glyburide |
| GLYBURIDE | GLYBURIDE | 1559685 | Glyburide |
| GLYBURIDE | GLYBURIDE | 40165998 | Glyburide |
| INSULIN | INSULIN | 19016117 | Insulin Lispro |
| INSULIN | INSULIN | 19062999 | insulin, isophane |
| GLIMEPIRIDE | GLIMEPIRIDE | 19135386 | glimepiride |
| INSULIN | INSULIN | 19078552 | insulin, isophane |
| INSULIN | INSULIN | 19012679 | Regular Insulin, Human |
| EXENATIDE | EXENATIDE | 46234118 | exenatide |
| INSULIN | INSULIN | 46233975 | Insulin Lispro |
| SITAGLIPTIN | SITAGLIPTIN | 1580747 | sitagliptin |
| ROSIGLITAZONE | ROSIGLITAZONE | 1547510 | rosiglitazone |
| NATEGLINIDE | NATEGLINIDE | 1502826 | nateglinide |

| | | | |
|---|---|---|---|
| INSULIN | INSULIN | 1586345 | insulin, isophane |
| INSULIN | INSULIN | 46234240 | insulin, isophane |
| INSULIN | INSULIN | 40052088 | Insulin Lispro |
| INSULIN | INSULIN | 1596941 | insulin, isophane |
| DULAGLUTIDE | DULAGLUTIDE | 36247186 | dulaglutide |
| ACARBOSE | ACARBOSE | 40003412 | Acarbose |
| LIRAGLUTIDE | LIRAGLUTIDE | 40169223 | liraglutide |
| GLYBURIDE | GLYBURIDE | 40166003 | Glyburide |
| BROMOCRIPTINE | BROMOCRIPTINE | 19007101 | Bromocriptine |
| CHLORPROPAMIDE | CHLORPROPAMIDE | 1594979 | Chlorpropamide |
| EXENATIDE | EXENATIDE | 19083253 | exenatide |
| METFORMIN | METFORMIN | 40163924 | Metformin |
| GLYBURIDE | GLYBURIDE | 40166002 | Glyburide |
| INSULIN | INSULIN | 1596999 | Regular Insulin, Human |
| EXENATIDE | EXENATIDE | 19083253 | exenatide |
| INSULIN | INSULIN | 19120315 | insulin, isophane |
| GLIPIZIDE | GLIPIZIDE | 40166814 | Glipizide |
| INSULIN | INSULIN | 19120315 | insulin, isophane |
| METFORMIN | METFORMIN | 40164930 | Metformin |
| BROMOCRIPTINE | BROMOCRIPTINE | 19041784 | Bromocriptine |
| GLIPIZIDE | GLIPIZIDE | 40166816 | Glipizide |
| ROSIGLITAZONE | ROSIGLITAZONE | 1547510 | rosiglitazone |
| SITAGLIPTIN | SITAGLIPTIN | 19125043 | sitagliptin |
| METFORMIN | METFORMIN | 40164928 | Metformin |
| EXENATIDE | EXENATIDE | 19083254 | exenatide |
| INSULIN | INSULIN | 1516981 | insulin detemir |
| PIOGLITAZONE | PIOGLITAZONE | 1525216 | pioglitazone |
| SITAGLIPTIN | SITAGLIPTIN | 19125047 | sitagliptin |
| INSULIN | INSULIN | 1516979 | insulin detemir |
| EXENATIDE | EXENATIDE | 42705842 | exenatide |
| EXENATIDE | EXENATIDE | 19083254 | exenatide |
| INSULIN | INSULIN | 1596953 | insulin, isophane |
| INSULIN | INSULIN | 1567198 | Insulin, Aspart, Human |

| | | | |
|---|---|---|---|
| GLYBURIDE | GLYBURIDE | 19030576 | Glyburide |
| DAPAGLIFLOZIN | DAPAGLIFLOZIN | 44785477 | dapagliflozin |
| INSULIN | INSULIN | 46234045 | Insulin, Aspart, Human |
| NATEGLINIDE | NATEGLINIDE | 19071495 | nateglinide |
| BROMOCRIPTINE | BROMOCRIPTINE | 19007101 | Bromocriptine |
| LINAGLIPTIN | LINAGLIPTIN | 42708089 | Linagliptin |
| METFORMIN | METFORMIN | 40163924 | Metformin |
| GLIPIZIDE | GLIPIZIDE | 36235748 | Glipizide |
| GLIPIZIDE | GLIPIZIDE | 40166815 | Glipizide |
| INSULIN | INSULIN | 36227902 | Insulin Lispro |
| MIGLITOL | MIGLITOL | 1510207 | miglitol |
| METFORMIN | METFORMIN | 40164921 | Metformin |
| INSULIN | INSULIN | 19078554 | Regular Insulin, Human |
| INSULIN | INSULIN | 1513881 | Insulin Lispro |
| GLIMEPIRIDE | GLIMEPIRIDE | 1597756 | glimepiride |
| GLIPIZIDE | GLIPIZIDE | 40165977 | Glipizide |
| METFORMIN | METFORMIN | 40164947 | Metformin |
| MIGLITOL | MIGLITOL | 1510206 | miglitol |
| REPAGLINIDE | REPAGLINIDE | 1516794 | repaglinide |
| REPAGLINIDE | REPAGLINIDE | 1516771 | repaglinide |
| GLIMEPIRIDE | GLIMEPIRIDE | 36234085 | glimepiride |
| DAPAGLIFLOZIN | DAPAGLIFLOZIN | 44785477 | dapagliflozin |
| GLIPIZIDE | GLIPIZIDE | 40166815 | Glipizide |
| METFORMIN | METFORMIN | 40063354 | Metformin |
| INSULIN | INSULIN | 46233975 | Insulin Lispro |
| INSULIN | INSULIN | 19062999 | insulin, isophane |
| INSULIN | INSULIN | 1550023 | Insulin Lispro |
| INSULIN | INSULIN | 40051348 | Insulin Glargine |
| INSULIN | INSULIN | 1502910 | Insulin Glargine |
| INSULIN | INSULIN | 40176981 | Insulin, Aspart Protamine, Human |
| INSULIN | INSULIN | 19110352 | insulin, isophane |
| INSULIN | INSULIN | 46233975 | Insulin Lispro |
| EMPAGLIFLOZIN | EMPAGLIFLOZIN | 45774758 | empagliflozin |

| | | | |
|---|---|---|---|
| INSULIN | INSULIN | 19078553 | insulin, isophane |
| METFORMIN | METFORMIN | 40164887 | Metformin |
| INSULIN | INSULIN | 19078553 | insulin, isophane |
| LINAGLIPTIN | LINAGLIPTIN | 42708087 | Linagliptin |
| GLIMEPIRIDE | GLIMEPIRIDE | 1597759 | glimepiride |
| INSULIN | INSULIN | 1550023 | Insulin Lispro |
| INSULIN | INSULIN | 46233948 | Insulin Lispro |
| ACARBOSE | ACARBOSE | 19023063 | Acarbose |
| INSULIN | INSULIN | 19095218 | Regular Insulin, Human |
| GLIMEPIRIDE | GLIMEPIRIDE | 40059665 | glimepiride |
| PIOGLITAZONE | PIOGLITAZONE | 36236324 | pioglitazone |
| INSULIN | INSULIN | 19078603 | insulin, isophane |
| METFORMIN | METFORMIN | 1503297 | Metformin |
| EXENATIDE | EXENATIDE | 46234118 | exenatide |
| PIOGLITAZONE | PIOGLITAZONE | 1525221 | pioglitazone |
| GLIPIZIDE | GLIPIZIDE | 40167636 | Glipizide |
| EXENATIDE | EXENATIDE | 1583727 | exenatide |
| EXENATIDE | EXENATIDE | 19083253 | exenatide |
| GLIMEPIRIDE | GLIMEPIRIDE | 1597759 | glimepiride |
| METFORMIN | METFORMIN | 19106521 | Metformin |
| DULAGLUTIDE | DULAGLUTIDE | 36247186 | dulaglutide |
| INSULIN | INSULIN | 40051348 | Insulin Glargine |
| EXENATIDE | EXENATIDE | 36238252 | exenatide |
| GLIMEPIRIDE | GLIMEPIRIDE | 36234085 | glimepiride |
| ROSIGLITAZONE | ROSIGLITAZONE | 1547504 | rosiglitazone |
| EXENATIDE | EXENATIDE | 19083253 | exenatide |
| INSULIN | INSULIN | 46234045 | Insulin, Aspart, Human |
| INSULIN | INSULIN | 1596957 | insulin, isophane |
| INSULIN | INSULIN | 46234240 | insulin, isophane |
| GLIMEPIRIDE | GLIMEPIRIDE | 36234085 | glimepiride |
| INSULIN | INSULIN | 1596952 | insulin, isophane |
| METFORMIN | METFORMIN | 1503297 | Metformin |
| INSULIN | INSULIN | 1586345 | insulin, isophane |

| | | | |
|---|---|---|---|
| CANAGLIFLOZIN | CANAGLIFLOZIN | 43526468 | canagliflozin |
| NATEGLINIDE | NATEGLINIDE | 19071495 | nateglinide |
| INSULIN | INSULIN | 19120790 | Insulin Glargine |
| GLIPIZIDE | GLIPIZIDE | 19100559 | Glipizide |
| GLIPIZIDE | GLIPIZIDE | 40165970 | Glipizide |
| GLIMEPIRIDE | GLIMEPIRIDE | 1597796 | glimepiride |
| METFORMIN | METFORMIN | 40063356 | Metformin |
| SAXAGLIPTIN | SAXAGLIPTIN | 40166042 | saxagliptin |
| GLIMEPIRIDE | GLIMEPIRIDE | 1597801 | glimepiride |
| GLIMEPIRIDE | GLIMEPIRIDE | 40059666 | glimepiride |
| GLYBURIDE | GLYBURIDE | 40165994 | Glyburide |
| INSULIN | INSULIN | 19078553 | insulin, isophane |
| METFORMIN | METFORMIN | 40164929 | Metformin |
| METFORMIN | METFORMIN | 40063352 | Metformin |
| INSULIN | INSULIN | 19110219 | insulin, isophane |
| PIOGLITAZONE | PIOGLITAZONE | 1525221 | pioglitazone |
| SAXAGLIPTIN | SAXAGLIPTIN | 40166042 | saxagliptin |
| PIOGLITAZONE | PIOGLITAZONE | 36236324 | pioglitazone |
| BROMOCRIPTINE | BROMOCRIPTINE | 730551 | Bromocriptine |
| CANAGLIFLOZIN | CANAGLIFLOZIN | 43526468 | canagliflozin |
| METFORMIN | METFORMIN | 1503328 | Metformin |
| GLIMEPIRIDE | GLIMEPIRIDE | 1597759 | glimepiride |
| GLYBURIDE | GLYBURIDE | 1559685 | Glyburide |
| INSULIN | INSULIN | 19078554 | Regular Insulin, Human |
| INSULIN | INSULIN | 1502905 | Insulin Glargine |
| GLYBURIDE | GLYBURIDE | 19077682 | Glyburide |
| INSULIN | INSULIN | 19110352 | insulin, isophane |
| GLIMEPIRIDE | GLIMEPIRIDE | 43011830 | glimepiride |
| INSULIN | INSULIN | 19095214 | insulin, isophane |
| INSULIN | INSULIN | 36231450 | Insulin, Aspart, Human |
| INSULIN | INSULIN | 19078553 | insulin, isophane |
| INSULIN | INSULIN | 19095214 | insulin, isophane |
| METFORMIN | METFORMIN | 40164920 | Metformin |
| INSULIN | INSULIN | 1596941 | insulin, isophane |

| | | | |
|---|---|---|---|
| GLIMEPIRIDE | GLIMEPIRIDE | 1597798 | glimepiride |
| REPAGLINIDE | REPAGLINIDE | 1516770 | repaglinide |
| METFORMIN | METFORMIN | 40164926 | Metformin |
| NATEGLINIDE | NATEGLINIDE | 36232423 | nateglinide |
| ROSIGLITAZONE | ROSIGLITAZONE | 19099055 | rosiglitazone |
| PIOGLITAZONE | PIOGLITAZONE | 1525215 | pioglitazone |
| LIRAGLUTIDE | LIRAGLUTIDE | 40169223 | liraglutide |
| INSULIN | INSULIN | 40051348 | Insulin Glargine |
| GLIPIZIDE | GLIPIZIDE | 40139022 | Glipizide |
| GLIPIZIDE | GLIPIZIDE | 40165976 | Glipizide |
| INSULIN | INSULIN | 19078553 | insulin, isophane |
| SAXAGLIPTIN | SAXAGLIPTIN | 40166038 | saxagliptin |
| METFORMIN | METFORMIN | 1503297 | Metformin |
| INSULIN | INSULIN | 36233552 | insulin, isophane |
| EMPAGLIFLOZIN | EMPAGLIFLOZIN | 45892176 | empagliflozin |
| GLIMEPIRIDE | GLIMEPIRIDE | 40059665 | glimepiride |
| INSULIN | INSULIN | 1586344 | insulin, isophane |
| NATEGLINIDE | NATEGLINIDE | 1502830 | nateglinide |
| MIGLITOL | MIGLITOL | 1510206 | miglitol |
| GLIMEPIRIDE | GLIMEPIRIDE | 19135386 | glimepiride |
| GLYBURIDE | GLYBURIDE | 19030579 | Glyburide |
| METFORMIN | METFORMIN | 40164947 | Metformin |
| METFORMIN | METFORMIN | 40164929 | Metformin |
| INSULIN | INSULIN | 19044971 | insulin detemir |
| EXENATIDE | EXENATIDE | 19083253 | exenatide |
| METFORMIN | METFORMIN | 40164946 | Metformin |
| INSULIN | INSULIN | 1567198 | Insulin, Aspart, Human |
| GLIMEPIRIDE | GLIMEPIRIDE | 1597758 | glimepiride |
| ACARBOSE | ACARBOSE | 36242198 | Acarbose |
| CANAGLIFLOZIN | CANAGLIFLOZIN | 45774721 | canagliflozin |
| REPAGLINIDE | REPAGLINIDE | 1516766 | repaglinide |
| METFORMIN | METFORMIN | 40164947 | Metformin |
| INSULIN | INSULIN | 19111797 | insulin, isophane |
| ROSIGLITAZONE | ROSIGLITAZONE | 1547506 | rosiglitazone |
| METFORMIN | METFORMIN | 40164923 | Metformin |

| | | | |
|---|---|---|---|
| SITAGLIPTIN | SITAGLIPTIN | 36221232 | sitagliptin |
| CANAGLIFLOZIN | CANAGLIFLOZIN | 43526472 | canagliflozin |
| INSULIN | INSULIN | 46233975 | Insulin Lispro |
| CANAGLIFLOZIN | CANAGLIFLOZIN | 43526472 | canagliflozin |
| GLYBURIDE | GLYBURIDE | 40166002 | Glyburide |
| ALBIGLUTIDE | ALBIGLUTIDE | 44816283 | albiglutide |
| INSULIN | INSULIN | 19078553 | insulin, isophane |
| SITAGLIPTIN | SITAGLIPTIN | 19125041 | sitagliptin |
| INSULIN | INSULIN | 36233154 | Insulin, Glulisine, Human |
| LINAGLIPTIN | LINAGLIPTIN | 40239217 | Linagliptin |
| INSULIN | INSULIN | 1502905 | Insulin Glargine |
| METFORMIN | METFORMIN | 40164896 | Metformin |
| GLYBURIDE | GLYBURIDE | 19079986 | Glyburide |
| GLYBURIDE | GLYBURIDE | 19106976 | Glyburide |
| EXENATIDE | EXENATIDE | 1583730 | exenatide |
| METFORMIN | METFORMIN | 42708171 | Metformin |
| NATEGLINIDE | NATEGLINIDE | 19071495 | nateglinide |
| EXENATIDE | EXENATIDE | 1583730 | exenatide |
| BROMOCRIPTINE | BROMOCRIPTINE | 40164161 | Bromocriptine |
| INSULIN | INSULIN | 1586344 | insulin, isophane |
| SITAGLIPTIN | SITAGLIPTIN | 36221232 | sitagliptin |
| NATEGLINIDE | NATEGLINIDE | 1502830 | nateglinide |
| INSULIN | INSULIN | 46233948 | Insulin Lispro |
| INSULIN | INSULIN | 1586344 | insulin, isophane |
| INSULIN | INSULIN | 36222190 | insulin detemir |
| DAPAGLIFLOZIN | DAPAGLIFLOZIN | 44785473 | dapagliflozin |
| GLIPIZIDE | GLIPIZIDE | 36218767 | Glipizide |
| REPAGLINIDE | REPAGLINIDE | 19023426 | repaglinide |
| GLIPIZIDE | GLIPIZIDE | 40166816 | Glipizide |
| INSULIN | INSULIN | 19078553 | insulin, isophane |
| INSULIN | INSULIN | 19078553 | insulin, isophane |
| METFORMIN | METFORMIN | 40164886 | Metformin |
| INSULIN | INSULIN | 19094122 | Regular Insulin, Human |
| GLYBURIDE | GLYBURIDE | 36235750 | Glyburide |

| | | | |
|---|---|---|---|
| BROMOCRIPTINE | BROMOCRIPTINE | 730551 | Bromocriptine |
| INSULIN | INSULIN | 1596955 | insulin, isophane |
| INSULIN | INSULIN | 1513849 | Insulin, Zinc, Human |
| SITAGLIPTIN | SITAGLIPTIN | 19125049 | sitagliptin |
| ROSIGLITAZONE | ROSIGLITAZONE | 1547510 | rosiglitazone |
| BROMOCRIPTINE | BROMOCRIPTINE | 40016898 | Bromocriptine |
| METFORMIN | METFORMIN | 40173239 | Metformin |
| INSULIN | INSULIN | 19110219 | insulin, isophane |
| INSULIN | INSULIN | 40051348 | Insulin Glargine |
| INSULIN | INSULIN | 19110352 | insulin, isophane |
| INSULIN | INSULIN | 19111797 | insulin, isophane |
| INSULIN | INSULIN | 1586370 | Ultralente Insulin, Human |
| CHLORPROPAMIDE | CHLORPROPAMIDE | 1594976 | Chlorpropamide |
| GLYBURIDE | GLYBURIDE | 19030579 | Glyburide |
| INSULIN | INSULIN | 19062999 | insulin, isophane |
| INSULIN | INSULIN | 19100128 | Regular Insulin, Human |
| ACARBOSE | ACARBOSE | 1529352 | Acarbose |
| INSULIN | INSULIN | 19120315 | insulin, isophane |
| INSULIN | INSULIN | 1513881 | Insulin Lispro |
| REPAGLINIDE | REPAGLINIDE | 36213643 | repaglinide |
| INSULIN | INSULIN | 42902823 | insulin, isophane |
| SAXAGLIPTIN | SAXAGLIPTIN | 40166042 | saxagliptin |
| INSULIN | INSULIN | 46234045 | Insulin, Aspart, Human |
| METFORMIN | METFORMIN | 40163925 | Metformin |
| NATEGLINIDE | NATEGLINIDE | 19107111 | nateglinide |
| PRAMLINTIDE | PRAMLINTIDE | 40165687 | Pramlintide |
| INSULIN | INSULIN | 1550023 | Insulin Lispro |
| INSULIN | INSULIN | 1502905 | Insulin Glargine |
| INSULIN | INSULIN | 19078559 | Insulin Glargine |
| INSULIN | INSULIN | 1567198 | Insulin, Aspart, Human |
| PIOGLITAZONE | PIOGLITAZONE | 1525220 | pioglitazone |
| EMPAGLIFLOZIN | EMPAGLIFLOZIN | 45774759 | empagliflozin |
| INSULIN | INSULIN | 46233972 | Insulin Lispro |
| LINAGLIPTIN | LINAGLIPTIN | 42705859 | Linagliptin |

| | | | |
|---|---|---|---|
| INSULIN | INSULIN | 19100128 | Regular Insulin, Human |
| INSULIN | INSULIN | 40166274 | Insulin Lispro |
| METFORMIN | METFORMIN | 40163929 | Metformin |
| METFORMIN | METFORMIN | 40164945 | Metformin |
| INSULIN | INSULIN | 19135263 | insulin, isophane |
| CANAGLIFLOZIN | CANAGLIFLOZIN | 43526472 | canagliflozin |
| INSULIN | INSULIN | 1516981 | insulin detemir |
| LINAGLIPTIN | LINAGLIPTIN | 40239219 | Linagliptin |
| CANAGLIFLOZIN | CANAGLIFLOZIN | 45774717 | canagliflozin |
| SAXAGLIPTIN | SAXAGLIPTIN | 40166041 | saxagliptin |
| METFORMIN | METFORMIN | 42708171 | Metformin |
| INSULIN | INSULIN | 19095214 | insulin, isophane |
| GLYBURIDE | GLYBURIDE | 19077682 | Glyburide |
| GLIMEPIRIDE | GLIMEPIRIDE | 36214985 | glimepiride |
| INSULIN | INSULIN | 46233976 | Insulin Lispro |
| ROSIGLITAZONE | ROSIGLITAZONE | 1547506 | rosiglitazone |
| INSULIN | INSULIN | 1502905 | Insulin Glargine |
| METFORMIN | METFORMIN | 40164882 | Metformin |
| INSULIN | INSULIN | 40184294 | Insulin, Aspart Protamine, Human |
| EXENATIDE | EXENATIDE | 1583722 | exenatide |
| GLIPIZIDE | GLIPIZIDE | 19080793 | Glipizide |
| INSULIN | INSULIN | 40184293 | Insulin, Aspart Protamine, Human |
| INSULIN | INSULIN | 19009384 | insulin, isophane |
| METFORMIN | METFORMIN | 40229046 | Metformin |
| GLIPIZIDE | GLIPIZIDE | 40167638 | Glipizide |
| GLIMEPIRIDE | GLIMEPIRIDE | 1597757 | glimepiride |
| GLIMEPIRIDE | GLIMEPIRIDE | 19135386 | glimepiride |
| INSULIN | INSULIN | 46233972 | Insulin Lispro |
| GLYBURIDE | GLYBURIDE | 19077684 | Glyburide |
| INSULIN | INSULIN | 1550023 | Insulin Lispro |
| GLIMEPIRIDE | GLIMEPIRIDE | 1597759 | glimepiride |

| | | | |
|---|---|---|---|
| BROMOCRIPTINE | BROMOCRIPTINE | 730551 | Bromocriptine |
| INSULIN | INSULIN | 1531601 | Insulin, Aspart Protamine, Human |
| GLIMEPIRIDE | GLIMEPIRIDE | 1597798 | glimepiride |
| ALOGLIPTIN | ALOGLIPTIN | 43013925 | alogliptin |
| METFORMIN | METFORMIN | 1503328 | Metformin |
| INSULIN | INSULIN | 1544838 | Insulin, Glulisine, Human |
| INSULIN | INSULIN | 1596955 | insulin, isophane |
| INSULIN | INSULIN | 40051347 | Insulin Glargine |
| METFORMIN | METFORMIN | 40164892 | Metformin |
| METFORMIN | METFORMIN | 1503297 | Metformin |
| ROSIGLITAZONE | ROSIGLITAZONE | 1547507 | rosiglitazone |
| METFORMIN | METFORMIN | 40126689 | Metformin |
| EXENATIDE | EXENATIDE | 1583730 | exenatide |
| GLIPIZIDE | GLIPIZIDE | 40167636 | Glipizide |
| GLYBURIDE | GLYBURIDE | 19030580 | Glyburide |
| INSULIN | INSULIN | 19078558 | Insulin, Aspart, Human |
| INSULIN | INSULIN | 1567198 | Insulin, Aspart, Human |
| GLIPIZIDE | GLIPIZIDE | 19080793 | Glipizide |
| INSULIN | INSULIN | 40166275 | Insulin Lispro |
| EXENATIDE | EXENATIDE | 1583727 | exenatide |
| METFORMIN | METFORMIN | 40163926 | Metformin |
| GLYBURIDE | GLYBURIDE | 19030579 | Glyburide |
| INSULIN | INSULIN | 1550023 | Insulin Lispro |
| LIRAGLUTIDE | LIRAGLUTIDE | 40169223 | liraglutide |
| GLYBURIDE | GLYBURIDE | 19077684 | Glyburide |
| INSULIN | INSULIN | 1550023 | Insulin Lispro |
| METFORMIN | METFORMIN | 40164889 | Metformin |
| METFORMIN | METFORMIN | 36235787 | Metformin |
| INSULIN | INSULIN | 19062999 | insulin, isophane |
| INSULIN | INSULIN | 1596977 | Regular Insulin, Human |
| LINAGLIPTIN | LINAGLIPTIN | 36238286 | Linagliptin |
| INSULIN | INSULIN | 1567198 | Insulin, Aspart, Human |
| BROMOCRIPTINE | BROMOCRIPTINE | 36238866 | Bromocriptine |
| INSULIN | INSULIN | 46233975 | Insulin Lispro |
| GLYBURIDE | GLYBURIDE | 36221460 | Glyburide |
| ACARBOSE | ACARBOSE | 19040969 | Acarbose |

| | | | |
|---|---|---|---|
| LIRAGLUTIDE | LIRAGLUTIDE | 40170913 | liraglutide |
| METFORMIN | METFORMIN | 40164883 | Metformin |
| INSULIN | INSULIN | 46233976 | Insulin Lispro |
| INSULIN | INSULIN | 36231456 | insulin, isophane |
| INSULIN | INSULIN | 1502905 | Insulin Glargine |
| INSULIN | INSULIN | 1567232 | Insulin, Aspart, Human |
| INSULIN | INSULIN | 46234045 | Insulin, Aspart, Human |
| INSULIN | INSULIN | 1596960 | insulin, isophane |
| BROMOCRIPTINE | BROMOCRIPTINE | 40164162 | Bromocriptine |
| INSULIN | INSULIN | 1544873 | Insulin, Glulisine, Human |
| INSULIN | INSULIN | 40184294 | Insulin, Aspart Protamine, Human |
| EXENATIDE | EXENATIDE | 1583730 | exenatide |
| GLIMEPIRIDE | GLIMEPIRIDE | 1597772 | glimepiride |
| EMPAGLIFLOZIN | EMPAGLIFLOZIN | 45774895 | empagliflozin |
| METFORMIN | METFORMIN | 19109284 | Metformin |
| METFORMIN | METFORMIN | 40164947 | Metformin |
| INSULIN | INSULIN | 19120315 | insulin, isophane |
| INSULIN | INSULIN | 1550023 | Insulin Lispro |
| GLIMEPIRIDE | GLIMEPIRIDE | 19018043 | glimepiride |
| EXENATIDE | EXENATIDE | 1583727 | exenatide |
| METFORMIN | METFORMIN | 40164881 | Metformin |
| PIOGLITAZONE | PIOGLITAZONE | 1525217 | pioglitazone |
| EXENATIDE | EXENATIDE | 19135404 | exenatide |
| INSULIN | INSULIN | 1596977 | Regular Insulin, Human |
| GLIPIZIDE | GLIPIZIDE | 19077636 | Glipizide |
| GLIMEPIRIDE | GLIMEPIRIDE | 1597773 | glimepiride |
| REPAGLINIDE | REPAGLINIDE | 1516795 | repaglinide |
| GLYBURIDE | GLYBURIDE | 19077681 | Glyburide |
| LIRAGLUTIDE | LIRAGLUTIDE | 36241903 | liraglutide |
| EMPAGLIFLOZIN | EMPAGLIFLOZIN | 45774758 | empagliflozin |
| SITAGLIPTIN | SITAGLIPTIN | 19125047 | sitagliptin |
| GLIPIZIDE | GLIPIZIDE | 40044253 | Glipizide |
| INSULIN | INSULIN | 1596999 | Regular Insulin, Human |
| GLYBURIDE | GLYBURIDE | 36231169 | Glyburide |
| LIRAGLUTIDE | LIRAGLUTIDE | 40170911 | liraglutide |

| | | | |
|---|---|---|---|
| INSULIN | INSULIN | 19110357 | Regular Insulin, Human |
| INSULIN | INSULIN | 19078553 | insulin, isophane |
| METFORMIN | METFORMIN | 40164880 | Metformin |
| METFORMIN | METFORMIN | 40164905 | Metformin |
| INSULIN | INSULIN | 1513912 | Insulin Lispro |
| INSULIN | INSULIN | 19078553 | insulin, isophane |
| SITAGLIPTIN | SITAGLIPTIN | 19125045 | sitagliptin |
| INSULIN | INSULIN | 19078530 | insulin, isophane |
| DAPAGLIFLOZIN | DAPAGLIFLOZIN | 44785473 | dapagliflozin |
| BROMOCRIPTINE | BROMOCRIPTINE | 730572 | Bromocriptine |
| GLYBURIDE | GLYBURIDE | 19060409 | Glyburide |
| INSULIN | INSULIN | 1550023 | Insulin Lispro |
| GLYBURIDE | GLYBURIDE | 19077659 | Glyburide |
| INSULIN | INSULIN | 40166275 | Insulin Lispro |
| BROMOCRIPTINE | BROMOCRIPTINE | 730602 | Bromocriptine |
| GLIPIZIDE | GLIPIZIDE | 1560171 | Glipizide |
| INSULIN | INSULIN | 46234045 | Insulin, Aspart, Human |
| METFORMIN | METFORMIN | 36221230 | Metformin |
| INSULIN | INSULIN | 19100127 | insulin, isophane |
| METFORMIN | METFORMIN | 40163922 | Metformin |
| METFORMIN | METFORMIN | 36221230 | Metformin |
| GLIPIZIDE | GLIPIZIDE | 1560171 | Glipizide |
| INSULIN | INSULIN | 40184293 | Insulin, Aspart Protamine, Human |
| METFORMIN | METFORMIN | 19113780 | Metformin |
| BROMOCRIPTINE | BROMOCRIPTINE | 730579 | Bromocriptine |
| INSULIN | INSULIN | 1502908 | Insulin Glargine |
| INSULIN | INSULIN | 1550023 | Insulin Lispro |
| INSULIN | INSULIN | 46221557 | Insulin Glargine |
| INSULIN | INSULIN | 40184293 | Insulin, Aspart Protamine, Human |
| METFORMIN | METFORMIN | 40231396 | Metformin |
| PIOGLITAZONE | PIOGLITAZONE | 19079293 | pioglitazone |

| | | | |
|---|---|---|---|
| INSULIN | INSULIN | 19078530 | insulin, isophane |
| METFORMIN | METFORMIN | 1503297 | Metformin |
| INSULIN | INSULIN | 19116453 | insulin, isophane |
| ROSIGLITAZONE | ROSIGLITAZONE | 1547508 | rosiglitazone |
| INSULIN | INSULIN | 1596952 | insulin, isophane |
| INSULIN | INSULIN | 19123376 | Insulin Lispro |
| INSULIN | INSULIN | 19078607 | insulin, isophane |
| GLIPIZIDE | GLIPIZIDE | 40139022 | Glipizide |
| GLIMEPIRIDE | GLIMEPIRIDE | 43011830 | glimepiride |
| EXENATIDE | EXENATIDE | 1583727 | exenatide |
| INSULIN | INSULIN | 46234045 | Insulin, Aspart, Human |
| INSULIN | INSULIN | 19110219 | insulin, isophane |
| GLYBURIDE | GLYBURIDE | 19077682 | Glyburide |
| INSULIN | INSULIN | 19044971 | insulin detemir |
| INSULIN | INSULIN | 19110352 | insulin, isophane |
| LIRAGLUTIDE | LIRAGLUTIDE | 40170913 | liraglutide |
| EXENATIDE | EXENATIDE | 1583730 | exenatide |
| INSULIN | INSULIN | 1567232 | Insulin, Aspart, Human |
| DAPAGLIFLOZIN | DAPAGLIFLOZIN | 44785473 | dapagliflozin |
| METFORMIN | METFORMIN | 36235581 | Metformin |
| GLIPIZIDE | GLIPIZIDE | 40165976 | Glipizide |
| INSULIN | INSULIN | 46234045 | Insulin, Aspart, Human |
| INSULIN | INSULIN | 19078553 | insulin, isophane |
| INSULIN | INSULIN | 1596955 | insulin, isophane |
| INSULIN | INSULIN | 1502905 | Insulin Glargine |
| LIRAGLUTIDE | LIRAGLUTIDE | 40169223 | liraglutide |
| REPAGLINIDE | REPAGLINIDE | 19023425 | repaglinide |
| INSULIN | INSULIN | 1516981 | insulin detemir |
| METFORMIN | METFORMIN | 42708171 | Metformin |
| GLYBURIDE | GLYBURIDE | 40168369 | Glyburide |
| INSULIN | INSULIN | 1550023 | Insulin Lispro |
| INSULIN | INSULIN | 36231458 | insulin, isophane |
| INSULIN | INSULIN | 1513914 | Insulin Lispro |

| | | | |
|---|---|---|---|
| GLIPIZIDE | GLIPIZIDE | 19080793 | Glipizide |
| INSULIN | INSULIN | 1550023 | Insulin Lispro |
| GLIPIZIDE | GLIPIZIDE | 40166815 | Glipizide |
| INSULIN | INSULIN | 19078551 | insulin, isophane |
| INSULIN | INSULIN | 1516981 | insulin detemir |
| METFORMIN | METFORMIN | 40164917 | Metformin |
| EXENATIDE | EXENATIDE | 19083253 | exenatide |
| INSULIN | INSULIN | 1550023 | Insulin Lispro |
| INSULIN | INSULIN | 19044971 | insulin detemir |
| GLIPIZIDE | GLIPIZIDE | 40165976 | Glipizide |
| INSULIN | INSULIN | 36248870 | Insulin Glargine |
| EXENATIDE | EXENATIDE | 19083254 | exenatide |
| GLIMEPIRIDE | GLIMEPIRIDE | 43011831 | glimepiride |
| METFORMIN | METFORMIN | 40164943 | Metformin |
| INSULIN | INSULIN | 46234043 | Insulin, Aspart, Human |
| INSULIN | INSULIN | 19078603 | insulin, isophane |
| INSULIN | INSULIN | 40051348 | Insulin Glargine |
| INSULIN | INSULIN | 1596957 | insulin, isophane |
| EXENATIDE | EXENATIDE | 19083254 | exenatide |
| EXENATIDE | EXENATIDE | 19083253 | exenatide |
| SITAGLIPTIN | SITAGLIPTIN | 19125049 | sitagliptin |
| GLIPIZIDE | GLIPIZIDE | 40165975 | Glipizide |
| INSULIN | INSULIN | 46233948 | Insulin Lispro |
| INSULIN | INSULIN | 1586344 | insulin, isophane |
| INSULIN | INSULIN | 1596953 | insulin, isophane |
| INSULIN | INSULIN | 40184294 | Insulin, Aspart Protamine, Human |
| INSULIN | INSULIN | 1596977 | Regular Insulin, Human |
| INSULIN | INSULIN | 1550027 | Insulin Lispro |
| INSULIN | INSULIN | 19120790 | Insulin Glargine |
| ALOGLIPTIN | ALOGLIPTIN | 43013926 | alogliptin |
| INSULIN | INSULIN | 1586345 | insulin, isophane |
| INSULIN | INSULIN | 19014975 | insulin, isophane |

| | | | |
|---|---|---|---|
| BROMOCRIPTINE | BROMOCRIPTINE | 730548 | Bromocriptine |
| GLYBURIDE | GLYBURIDE | 19030580 | Glyburide |
| GLYBURIDE | GLYBURIDE | 40168370 | Glyburide |
| INSULIN | INSULIN | 46234045 | Insulin, Aspart, Human |
| INSULIN | INSULIN | 46233975 | Insulin Lispro |
| INSULIN | INSULIN | 1516981 | insulin detemir |
| METFORMIN | METFORMIN | 36235747 | Metformin |
| METFORMIN | METFORMIN | 19106521 | Metformin |
| INSULIN | INSULIN | 19112791 | Insulin, Glulisine, Human |
| METFORMIN | METFORMIN | 36223145 | Metformin |
| REPAGLINIDE | REPAGLINIDE | 19047663 | repaglinide |
| INSULIN | INSULIN | 1596955 | insulin, isophane |
| GLIPIZIDE | GLIPIZIDE | 19100559 | Glipizide |
| LIRAGLUTIDE | LIRAGLUTIDE | 40170912 | liraglutide |
| INSULIN | INSULIN | 19078553 | insulin, isophane |
| LIRAGLUTIDE | LIRAGLUTIDE | 45892686 | liraglutide |
| CANAGLIFLOZIN | CANAGLIFLOZIN | 43526468 | canagliflozin |
| METFORMIN | METFORMIN | 40063356 | Metformin |
| GLYBURIDE | GLYBURIDE | 19077681 | Glyburide |
| METFORMIN | METFORMIN | 40164918 | Metformin |
| INSULIN | INSULIN | 1516978 | insulin detemir |
| PIOGLITAZONE | PIOGLITAZONE | 1525218 | pioglitazone |
| PRAMLINTIDE | PRAMLINTIDE | 40165687 | Pramlintide |
| SITAGLIPTIN | SITAGLIPTIN | 36221233 | sitagliptin |
| GLYBURIDE | GLYBURIDE | 36230035 | Glyburide |
| INSULIN | INSULIN | 1516981 | insulin detemir |
| NATEGLINIDE | NATEGLINIDE | 1502829 | nateglinide |
| INSULIN | INSULIN | 1550023 | Insulin Lispro |
| PRAMLINTIDE | PRAMLINTIDE | 40163899 | Pramlintide |
| METFORMIN | METFORMIN | 40164932 | Metformin |
| INSULIN | INSULIN | 46234045 | Insulin, Aspart, Human |
| INSULIN | INSULIN | 1502905 | Insulin Glargine |
| CANAGLIFLOZIN | CANAGLIFLOZIN | 36246877 | canagliflozin |
| GLYBURIDE | GLYBURIDE | 19060491 | Glyburide |
| ROSIGLITAZONE | ROSIGLITAZONE | 36229365 | rosiglitazone |
| INSULIN | INSULIN | 1586345 | insulin, isophane |
| EMPAGLIFLOZIN | EMPAGLIFLOZIN | 45892178 | empagliflozin |

| | | | |
|---|---|---|---|
| GLYBURIDE | GLYBURIDE | 19030575 | Glyburide |
| GLYBURIDE | GLYBURIDE | 19030575 | Glyburide |
| INSULIN | INSULIN | 19095214 | insulin, isophane |
| SITAGLIPTIN | SITAGLIPTIN | 19125047 | sitagliptin |
| EXENATIDE | EXENATIDE | 1583722 | exenatide |
| EMPAGLIFLOZIN | EMPAGLIFLOZIN | 45892179 | empagliflozin |
| ACARBOSE | ACARBOSE | 40003413 | Acarbose |
| METFORMIN | METFORMIN | 40164930 | Metformin |
| INSULIN | INSULIN | 19116454 | insulin, isophane |
| GLIPIZIDE | GLIPIZIDE | 40165969 | Glipizide |
| NATEGLINIDE | NATEGLINIDE | 1502827 | nateglinide |
| DAPAGLIFLOZIN | DAPAGLIFLOZIN | 44785477 | dapagliflozin |
| GLIPIZIDE | GLIPIZIDE | 19077638 | Glipizide |
| SITAGLIPTIN | SITAGLIPTIN | 19125047 | sitagliptin |
| GLYBURIDE | GLYBURIDE | 40165994 | Glyburide |
| BROMOCRIPTINE | BROMOCRIPTINE | 730572 | Bromocriptine |
| INSULIN | INSULIN | 1567198 | Insulin, Aspart, Human |
| SITAGLIPTIN | SITAGLIPTIN | 40136770 | sitagliptin |
| METFORMIN | METFORMIN | 40063354 | Metformin |
| MIGLITOL | MIGLITOL | 1510208 | miglitol |
| GLYBURIDE | GLYBURIDE | 19030579 | Glyburide |
| DAPAGLIFLOZIN | DAPAGLIFLOZIN | 44785477 | dapagliflozin |
| INSULIN | INSULIN | 19078633 | Insulin, Regular, Pork |
| INSULIN | INSULIN | 36235843 | Ultralente Insulin, Human |
| METFORMIN | METFORMIN | 1503328 | Metformin |
| INSULIN | INSULIN | 1502905 | Insulin Glargine |
| INSULIN | INSULIN | 19078553 | insulin, isophane |
| INSULIN | INSULIN | 1513912 | Insulin Lispro |
| MIGLITOL | MIGLITOL | 1510208 | miglitol |
| GLYBURIDE | GLYBURIDE | 1559684 | Glyburide |
| INSULIN | INSULIN | 1513881 | Insulin Lispro |
| INSULIN | INSULIN | 1516981 | insulin detemir |
| CANAGLIFLOZIN | CANAGLIFLOZIN | 43526466 | canagliflozin |
| METFORMIN | METFORMIN | 1503297 | Metformin |
| INSULIN | INSULIN | 1597001 | Regular Insulin, Human |

| | | | |
|---|---|---|---|
| ROSIGLITAZONE | ROSIGLITAZONE | 1547505 | rosiglitazone |
| GLYBURIDE | GLYBURIDE | 19060491 | Glyburide |
| METFORMIN | METFORMIN | 36221230 | Metformin |
| INSULIN | INSULIN | 1513912 | Insulin Lispro |
| CHLORPROPAMIDE | CHLORPROPAMIDE | 1594980 | Chlorpropamide |
| INSULIN | INSULIN | 46233975 | Insulin Lispro |
| GLIPIZIDE | GLIPIZIDE | 40165973 | Glipizide |
| NATEGLINIDE | NATEGLINIDE | 1502827 | nateglinide |
| INSULIN | INSULIN | 19071700 | Insulin Glargine |
| INSULIN | INSULIN | 1596974 | insulin, isophane |
| BROMOCRIPTINE | BROMOCRIPTINE | 36238866 | Bromocriptine |
| GLYBURIDE | GLYBURIDE | 19063991 | Glyburide |
| ACARBOSE | ACARBOSE | 19047612 | Acarbose |
| METFORMIN | METFORMIN | 1503328 | Metformin |
| INSULIN | INSULIN | 1596977 | Regular Insulin, Human |
| INSULIN | INSULIN | 36236032 | Insulin Lispro |
| METFORMIN | METFORMIN | 40164903 | Metformin |
| INSULIN | INSULIN | 1567201 | Insulin, Aspart Protamine, Human |
| INSULIN | INSULIN | 46233975 | Insulin Lispro |
| GLIPIZIDE | GLIPIZIDE | 36235748 | Glipizide |
| METFORMIN | METFORMIN | 40229046 | Metformin |
| GLIMEPIRIDE | GLIMEPIRIDE | 1597798 | glimepiride |
| INSULIN | INSULIN | 19078553 | insulin, isophane |
| INSULIN | INSULIN | 40176981 | Insulin, Aspart Protamine, Human |
| EXENATIDE | EXENATIDE | 19083253 | exenatide |
| INSULIN | INSULIN | 19080091 | Ultralente Insulin, Human |
| GLYBURIDE | GLYBURIDE | 19106976 | Glyburide |
| INSULIN | INSULIN | 1550028 | Insulin Lispro |
| INSULIN | INSULIN | 40176981 | Insulin, Aspart Protamine, Human |
| INSULIN | INSULIN | 1567198 | Insulin, Aspart, Human |
| INSULIN | INSULIN | 1597001 | Regular Insulin, Human |
| INSULIN | INSULIN | 1513876 | Insulin, Protamine Lispro, Human |

| | | | |
|---|---|---|---|
| EXENATIDE | EXENATIDE | 1583727 | exenatide |
| ROSIGLITAZONE | ROSIGLITAZONE | 1547508 | rosiglitazone |
| INSULIN | INSULIN | 19094122 | Regular Insulin, Human |
| LIRAGLUTIDE | LIRAGLUTIDE | 40170912 | liraglutide |
| INSULIN | INSULIN | 19080091 | Ultralente Insulin, Human |
| SITAGLIPTIN | SITAGLIPTIN | 19125051 | sitagliptin |
| ACARBOSE | ACARBOSE | 19021312 | Acarbose |
| REPAGLINIDE | REPAGLINIDE | 1516771 | repaglinide |
| INSULIN | INSULIN | 19095218 | Regular Insulin, Human |
| INSULIN | INSULIN | 1596959 | insulin, isophane |
| INSULIN | INSULIN | 40130712 | insulin detemir |
| EXENATIDE | EXENATIDE | 42902866 | exenatide |
| INSULIN | INSULIN | 40184294 | Insulin, Aspart Protamine, Human |
| EXENATIDE | EXENATIDE | 19083254 | exenatide |
| GLYBURIDE | GLYBURIDE | 1559685 | Glyburide |
| CANAGLIFLOZIN | CANAGLIFLOZIN | 43526468 | canagliflozin |
| EXENATIDE | EXENATIDE | 19083254 | exenatide |
| REPAGLINIDE | REPAGLINIDE | 19023424 | repaglinide |
| GLIMEPIRIDE | GLIMEPIRIDE | 1597756 | glimepiride |
| METFORMIN | METFORMIN | 40164925 | Metformin |
| GLIPIZIDE | GLIPIZIDE | 40165970 | Glipizide |
| INSULIN | INSULIN | 19111795 | insulin, isophane |
| METFORMIN | METFORMIN | 40164897 | Metformin |
| INSULIN | INSULIN | 36236032 | Insulin Lispro |
| GLYBURIDE | GLYBURIDE | 19077682 | Glyburide |
| INSULIN | INSULIN | 19135263 | insulin, isophane |
| EXENATIDE | EXENATIDE | 19083253 | exenatide |
| REPAGLINIDE | REPAGLINIDE | 19023425 | repaglinide |
| LIRAGLUTIDE | LIRAGLUTIDE | 40169223 | liraglutide |
| INSULIN | INSULIN | 19078558 | Insulin, Aspart, Human |
| INSULIN | INSULIN | 1596974 | insulin, isophane |
| INSULIN | INSULIN | 1502905 | Insulin Glargine |
| SITAGLIPTIN | SITAGLIPTIN | 19125043 | sitagliptin |
| GLIMEPIRIDE | GLIMEPIRIDE | 1597761 | glimepiride |
| INSULIN | INSULIN | 1513916 | Insulin Lispro |

| | | | |
|---|---|---|---|
| INSULIN | INSULIN | 1516981 | insulin detemir |
| INSULIN | INSULIN | 1586370 | Ultralente Insulin, Human |
| SITAGLIPTIN | SITAGLIPTIN | 19125051 | sitagliptin |
| LIRAGLUTIDE | LIRAGLUTIDE | 36220119 | liraglutide |
| GLYBURIDE | GLYBURIDE | 40168374 | Glyburide |
| METFORMIN | METFORMIN | 42708167 | Metformin |
| GLYBURIDE | GLYBURIDE | 40168368 | Glyburide |
| INSULIN | INSULIN | 19120790 | Insulin Glargine |
| EMPAGLIFLOZIN | EMPAGLIFLOZIN | 45774752 | empagliflozin |
| GLIPIZIDE | GLIPIZIDE | 40166816 | Glipizide |
| TOLBUTAMIDE | TOLBUTAMIDE | 1502913 | Tolbutamide |
| BROMOCRIPTINE | BROMOCRIPTINE | 19006288 | Bromocriptine |
| REPAGLINIDE | REPAGLINIDE | 1516771 | repaglinide |
| LIRAGLUTIDE | LIRAGLUTIDE | 40169222 | liraglutide |
| GLYBURIDE | GLYBURIDE | 19030579 | Glyburide |
| INSULIN | INSULIN | 19078557 | Regular Insulin, Human |
| INSULIN | INSULIN | 36236032 | Insulin Lispro |
| INSULIN | INSULIN | 19110355 | insulin, isophane |
| INSULIN | INSULIN | 1544873 | Insulin, Glulisine, Human |
| REPAGLINIDE | REPAGLINIDE | 36240429 | repaglinide |
| GLYBURIDE | GLYBURIDE | 19106976 | Glyburide |
| INSULIN | INSULIN | 46234045 | Insulin, Aspart, Human |
| METFORMIN | METFORMIN | 40164895 | Metformin |
| GLIPIZIDE | GLIPIZIDE | 19030445 | Glipizide |
| GLYBURIDE | GLYBURIDE | 40165995 | Glyburide |
| PRAMLINTIDE | PRAMLINTIDE | 40165692 | Pramlintide |
| INSULIN | INSULIN | 19016117 | Insulin Lispro |
| INSULIN | INSULIN | 19078551 | insulin, isophane |
| REPAGLINIDE | REPAGLINIDE | 1516770 | repaglinide |
| ACARBOSE | ACARBOSE | 19040969 | Acarbose |
| ROSIGLITAZONE | ROSIGLITAZONE | 19099070 | rosiglitazone |
| GLYBURIDE | GLYBURIDE | 40165997 | Glyburide |
| EXENATIDE | EXENATIDE | 46234118 | exenatide |
| GLIMEPIRIDE | GLIMEPIRIDE | 1597758 | glimepiride |
| GLYBURIDE | GLYBURIDE | 19060409 | Glyburide |

| | | | |
|---|---|---|---|
| EXENATIDE | EXENATIDE | 19083253 | exenatide |
| METFORMIN | METFORMIN | 1503327 | Metformin |
| GLYBURIDE | GLYBURIDE | 19030580 | Glyburide |
| INSULIN | INSULIN | 19111795 | insulin, isophane |
| PRAMLINTIDE | PRAMLINTIDE | 40078056 | Pramlintide |
| LIRAGLUTIDE | LIRAGLUTIDE | 40169223 | liraglutide |
| ROSIGLITAZONE | ROSIGLITAZONE | 1547505 | rosiglitazone |
| INSULIN | INSULIN | 19120790 | Insulin Glargine |
| SITAGLIPTIN | SITAGLIPTIN | 42901620 | sitagliptin |
| GLIPIZIDE | GLIPIZIDE | 40044253 | Glipizide |
| METFORMIN | METFORMIN | 1503297 | Metformin |
| INSULIN | INSULIN | 19078554 | Regular Insulin, Human |
| ACARBOSE | ACARBOSE | 19047612 | Acarbose |
| DULAGLUTIDE | DULAGLUTIDE | 45774440 | dulaglutide |
| EXENATIDE | EXENATIDE | 46234116 | exenatide |
| GLIMEPIRIDE | GLIMEPIRIDE | 1597795 | glimepiride |
| INSULIN | INSULIN | 40052047 | insulin, isophane |
| METFORMIN | METFORMIN | 40164891 | Metformin |
| INSULIN | INSULIN | 19100127 | insulin, isophane |
| INSULIN | INSULIN | 19110357 | Regular Insulin, Human |
| INSULIN | INSULIN | 1550023 | Insulin Lispro |
| GLIPIZIDE | GLIPIZIDE | 40165969 | Glipizide |
| INSULIN | INSULIN | 19131582 | Insulin, Glulisine, Human |
| INSULIN | INSULIN | 19110352 | insulin, isophane |
| INSULIN | INSULIN | 1516976 | insulin detemir |
| INSULIN | INSULIN | 1596952 | insulin, isophane |
| ALOGLIPTIN | ALOGLIPTIN | 36246271 | alogliptin |
| GLYBURIDE | GLYBURIDE | 40046201 | Glyburide |
| INSULIN | INSULIN | 19014975 | insulin, isophane |
| GLIPIZIDE | GLIPIZIDE | 1560171 | Glipizide |
| GLYBURIDE | GLYBURIDE | 36235750 | Glyburide |
| METFORMIN | METFORMIN | 42708167 | Metformin |
| GLYBURIDE | GLYBURIDE | 40165996 | Glyburide |
| SITAGLIPTIN | SITAGLIPTIN | 19125049 | sitagliptin |

| | | | |
|---|---|---|---|
| INSULIN | INSULIN | 1513881 | Insulin Lispro |
| ACARBOSE | ACARBOSE | 1529331 | Acarbose |
| GLIMEPIRIDE | GLIMEPIRIDE | 19048032 | glimepiride |
| ACARBOSE | ACARBOSE | 36242198 | Acarbose |
| PIOGLITAZONE | PIOGLITAZONE | 19079293 | pioglitazone |
| METFORMIN | METFORMIN | 1503327 | Metformin |
| INSULIN | INSULIN | 1516976 | insulin detemir |
| INSULIN | INSULIN | 1596953 | insulin, isophane |
| INSULIN | INSULIN | 36231450 | Insulin, Aspart, Human |
| INSULIN | INSULIN | 19110357 | Regular Insulin, Human |
| INSULIN | INSULIN | 19061670 | Regular Insulin, Human |
| METFORMIN | METFORMIN | 40063354 | Metformin |
| INSULIN | INSULIN | 40166274 | Insulin Lispro |
| DULAGLUTIDE | DULAGLUTIDE | 45774440 | dulaglutide |
| NATEGLINIDE | NATEGLINIDE | 19071495 | nateglinide |
| INSULIN | INSULIN | 1596977 | Regular Insulin, Human |
| INSULIN | INSULIN | 19120315 | insulin, isophane |
| GLIMEPIRIDE | GLIMEPIRIDE | 1597758 | glimepiride |
| INSULIN | INSULIN | 19078551 | insulin, isophane |
| NATEGLINIDE | NATEGLINIDE | 1502830 | nateglinide |
| EXENATIDE | EXENATIDE | 19083254 | exenatide |
| GLIPIZIDE | GLIPIZIDE | 19134624 | Glipizide |
| GLIPIZIDE | GLIPIZIDE | 40167635 | Glipizide |
| SITAGLIPTIN | SITAGLIPTIN | 40243629 | sitagliptin |
| GLYBURIDE | GLYBURIDE | 40046204 | Glyburide |
| BROMOCRIPTINE | BROMOCRIPTINE | 36214531 | Bromocriptine |
| METFORMIN | METFORMIN | 40164905 | Metformin |
| EMPAGLIFLOZIN | EMPAGLIFLOZIN | 45892176 | empagliflozin |
| GLYBURIDE | GLYBURIDE | 1559761 | Glyburide |
| INSULIN | INSULIN | 1596977 | Regular Insulin, Human |
| GLIMEPIRIDE | GLIMEPIRIDE | 1597756 | glimepiride |
| CANAGLIFLOZIN | CANAGLIFLOZIN | 36246430 | canagliflozin |
| LINAGLIPTIN | LINAGLIPTIN | 36221103 | Linagliptin |
| INSULIN | INSULIN | 1550027 | Insulin Lispro |
| GLYBURIDE | GLYBURIDE | 40166002 | Glyburide |

| | | | |
|---|---|---|---|
| METFORMIN | METFORMIN | 40164920 | Metformin |
| INSULIN | INSULIN | 1516976 | insulin detemir |
| NATEGLINIDE | NATEGLINIDE | 1502827 | nateglinide |
| ROSIGLITAZONE | ROSIGLITAZONE | 1547507 | rosiglitazone |
| GLYBURIDE | GLYBURIDE | 19030580 | Glyburide |
| METFORMIN | METFORMIN | 40164928 | Metformin |
| ACARBOSE | ACARBOSE | 19021312 | Acarbose |
| METFORMIN | METFORMIN | 40164923 | Metformin |
| GLIPIZIDE | GLIPIZIDE | 40139022 | Glipizide |
| CANAGLIFLOZIN | CANAGLIFLOZIN | 45774712 | canagliflozin |
| INSULIN | INSULIN | 19018822 | Insulin Lispro |
| GLYBURIDE | GLYBURIDE | 36221460 | Glyburide |
| INSULIN | INSULIN | 19135320 | Regular Insulin, Human |
| ROSIGLITAZONE | ROSIGLITAZONE | 1547507 | rosiglitazone |
| INSULIN | INSULIN | 1597000 | Regular Insulin, Human |
| INSULIN | INSULIN | 36235841 | Regular Insulin, Human |
| INSULIN | INSULIN | 19080091 | Ultralente Insulin, Human |
| PIOGLITAZONE | PIOGLITAZONE | 1525216 | pioglitazone |
| ACARBOSE | ACARBOSE | 1529352 | Acarbose |
| DAPAGLIFLOZIN | DAPAGLIFLOZIN | 45775864 | dapagliflozin |
| METFORMIN | METFORMIN | 40164917 | Metformin |
| ACARBOSE | ACARBOSE | 19040970 | Acarbose |
| PIOGLITAZONE | PIOGLITAZONE | 1525218 | pioglitazone |
| REPAGLINIDE | REPAGLINIDE | 1516770 | repaglinide |
| INSULIN | INSULIN | 1502910 | Insulin Glargine |
| INSULIN | INSULIN | 1550028 | Insulin Lispro |
| LIRAGLUTIDE | LIRAGLUTIDE | 36241903 | liraglutide |
| INSULIN | INSULIN | 36236032 | Insulin Lispro |
| METFORMIN | METFORMIN | 1503297 | Metformin |
| METFORMIN | METFORMIN | 1503327 | Metformin |
| INSULIN | INSULIN | 1550023 | Insulin Lispro |
| INSULIN | INSULIN | 1596977 | Regular Insulin, Human |
| EXENATIDE | EXENATIDE | 1583727 | exenatide |

| | | | |
|---|---|---|---|
| INSULIN | INSULIN | 1502905 | Insulin Glargine |
| INSULIN | INSULIN | 19018822 | Insulin Lispro |
| GLIMEPIRIDE | GLIMEPIRIDE | 1597792 | glimepiride |
| GLIMEPIRIDE | GLIMEPIRIDE | 43011831 | glimepiride |
| GLYBURIDE | GLYBURIDE | 19030572 | Glyburide |
| ALBIGLUTIDE | ALBIGLUTIDE | 36246906 | albiglutide |
| LIRAGLUTIDE | LIRAGLUTIDE | 36241903 | liraglutide |
| INSULIN | INSULIN | 19078557 | Regular Insulin, Human |
| CANAGLIFLOZIN | CANAGLIFLOZIN | 43526468 | canagliflozin |
| INSULIN | INSULIN | 1502905 | Insulin Glargine |
| PIOGLITAZONE | PIOGLITAZONE | 40099897 | pioglitazone |
| INSULIN | INSULIN | 40184293 | Insulin, Aspart Protamine, Human |
| PRAMLINTIDE | PRAMLINTIDE | 40163890 | Pramlintide |
| METFORMIN | METFORMIN | 40164881 | Metformin |
| LIRAGLUTIDE | LIRAGLUTIDE | 40170913 | liraglutide |
| METFORMIN | METFORMIN | 40164904 | Metformin |
| REPAGLINIDE | REPAGLINIDE | 19023424 | repaglinide |
| METFORMIN | METFORMIN | 19088818 | Metformin |
| METFORMIN | METFORMIN | 40164898 | Metformin |
| INSULIN | INSULIN | 36235937 | insulin human, rDNA origin |
| INSULIN | INSULIN | 1596977 | Regular Insulin, Human |
| INSULIN | INSULIN | 46233975 | Insulin Lispro |
| GLIPIZIDE | GLIPIZIDE | 19077636 | Glipizide |
| SITAGLIPTIN | SITAGLIPTIN | 19125041 | sitagliptin |
| INSULIN | INSULIN | 46233730 | Regular Insulin, Human |
| METFORMIN | METFORMIN | 36235752 | Metformin |
| GLYBURIDE | GLYBURIDE | 40046201 | Glyburide |
| INSULIN | INSULIN | 46234043 | Insulin, Aspart, Human |
| LIRAGLUTIDE | LIRAGLUTIDE | 45892684 | liraglutide |
| LIRAGLUTIDE | LIRAGLUTIDE | 40169223 | liraglutide |
| INSULIN | INSULIN | 40051347 | Insulin Glargine |
| INSULIN | INSULIN | 46233947 | Insulin Lispro |
| GLYBURIDE | GLYBURIDE | 40165998 | Glyburide |
| INSULIN | INSULIN | 1544873 | Insulin, Glulisine, Human |
| INSULIN | INSULIN | 36231450 | Insulin, Aspart, Human |

| | | | |
|---|---|---|---|
| INSULIN | INSULIN | 1513881 | Insulin Lispro |
| INSULIN | INSULIN | 1513912 | Insulin Lispro |
| INSULIN | INSULIN | 19078554 | Regular Insulin, Human |
| METFORMIN | METFORMIN | 1503297 | Metformin |
| LIRAGLUTIDE | LIRAGLUTIDE | 45892686 | liraglutide |
| EXENATIDE | EXENATIDE | 1583727 | exenatide |
| LINAGLIPTIN | LINAGLIPTIN | 40239219 | Linagliptin |
| INSULIN | INSULIN | 19110357 | Regular Insulin, Human |
| BROMOCRIPTINE | BROMOCRIPTINE | 19007101 | Bromocriptine |
| METFORMIN | METFORMIN | 40164881 | Metformin |
| METFORMIN | METFORMIN | 40164923 | Metformin |
| INSULIN | INSULIN | 1567201 | Insulin, Aspart Protamine, Human |
| GLIPIZIDE | GLIPIZIDE | 40165973 | Glipizide |
| ROSIGLITAZONE | ROSIGLITAZONE | 1547510 | rosiglitazone |
| GLYBURIDE | GLYBURIDE | 40165993 | Glyburide |
| GLIMEPIRIDE | GLIMEPIRIDE | 1597795 | glimepiride |
| GLIPIZIDE | GLIPIZIDE | 19085004 | Glipizide |
| INSULIN | INSULIN | 19129469 | insulin human, rDNA origin |
| METFORMIN | METFORMIN | 40164919 | Metformin |
| INSULIN | INSULIN | 36229086 | Insulin Glargine |
| GLYBURIDE | GLYBURIDE | 19106860 | Glyburide |
| PRAMLINTIDE | PRAMLINTIDE | 1517998 | Pramlintide |
| INSULIN | INSULIN | 19078530 | insulin, isophane |
| DAPAGLIFLOZIN | DAPAGLIFLOZIN | 44785832 | dapagliflozin |
| INSULIN | INSULIN | 19120315 | insulin, isophane |
| GLIMEPIRIDE | GLIMEPIRIDE | 36234086 | glimepiride |
| INSULIN | INSULIN | 40184293 | Insulin, Aspart Protamine, Human |
| SITAGLIPTIN | SITAGLIPTIN | 40136769 | sitagliptin |

| | | | |
|---|---|---|---|
| INSULIN | INSULIN | 1596941 | insulin, isophane |
| INSULIN | INSULIN | 19078607 | insulin, isophane |
| GLIPIZIDE | GLIPIZIDE | 19134624 | Glipizide |
| INSULIN | INSULIN | 19078530 | insulin, isophane |
| METFORMIN | METFORMIN | 40164925 | Metformin |
| INSULIN | INSULIN | 1513849 | Insulin, Zinc, Human |
| INSULIN | INSULIN | 1550023 | Insulin Lispro |
| INSULIN | INSULIN | 19120315 | insulin, isophane |
| INSULIN | INSULIN | 46233972 | Insulin Lispro |
| PRAMLINTIDE | PRAMLINTIDE | 36243018 | Pramlintide |
| LIRAGLUTIDE | LIRAGLUTIDE | 40169223 | liraglutide |
| GLYBURIDE | GLYBURIDE | 40166003 | Glyburide |
| INSULIN | INSULIN | 35605622 | Regular Insulin, Human |
| INSULIN | INSULIN | 1513849 | Insulin, Zinc, Human |
| METFORMIN | METFORMIN | 40164926 | Metformin |
| ROSIGLITAZONE | ROSIGLITAZONE | 36229365 | rosiglitazone |
| INSULIN | INSULIN | 19129440 | insulin human, rDNA origin |
| METFORMIN | METFORMIN | 1503328 | Metformin |
| METFORMIN | METFORMIN | 40164944 | Metformin |
| ACARBOSE | ACARBOSE | 40003412 | Acarbose |
| GLIMEPIRIDE | GLIMEPIRIDE | 19135387 | glimepiride |
| METFORMIN | METFORMIN | 40164897 | Metformin |
| INSULIN | INSULIN | 1596941 | insulin, isophane |
| GLYBURIDE | GLYBURIDE | 36233495 | Glyburide |
| PIOGLITAZONE | PIOGLITAZONE | 40099897 | pioglitazone |
| ACARBOSE | ACARBOSE | 19040969 | Acarbose |
| METFORMIN | METFORMIN | 40164891 | Metformin |
| BROMOCRIPTINE | BROMOCRIPTINE | 19006288 | Bromocriptine |
| METFORMIN | METFORMIN | 36221231 | Metformin |
| INSULIN | INSULIN | 1550028 | Insulin Lispro |
| GLIMEPIRIDE | GLIMEPIRIDE | 1597796 | glimepiride |
| GLYBURIDE | GLYBURIDE | 40046201 | Glyburide |

| | | | |
|---|---|---|---|
| INSULIN | INSULIN | 19078557 | Regular Insulin, Human |
| INSULIN | INSULIN | 1550023 | Insulin Lispro |
| LINAGLIPTIN | LINAGLIPTIN | 42705862 | Linagliptin |
| GLIPIZIDE | GLIPIZIDE | 19085004 | Glipizide |
| INSULIN | INSULIN | 1516981 | insulin detemir |
| SAXAGLIPTIN | SAXAGLIPTIN | 36229291 | saxagliptin |
| EXENATIDE | EXENATIDE | 1583727 | exenatide |
| METFORMIN | METFORMIN | 40164946 | Metformin |
| ROSIGLITAZONE | ROSIGLITAZONE | 19079465 | rosiglitazone |
| SAXAGLIPTIN | SAXAGLIPTIN | 40166036 | saxagliptin |
| METFORMIN | METFORMIN | 40164906 | Metformin |
| INSULIN | INSULIN | 46233975 | Insulin Lispro |
| INSULIN | INSULIN | 1586370 | Ultralente Insulin, Human |
| INSULIN | INSULIN | 36235937 | insulin human, rDNA origin |
| EXENATIDE | EXENATIDE | 46234118 | exenatide |
| INSULIN | INSULIN | 1550023 | Insulin Lispro |
| GLYBURIDE | GLYBURIDE | 40046204 | Glyburide |
| INSULIN | INSULIN | 36235843 | Ultralente Insulin, Human |
| INSULIN | INSULIN | 1550027 | Insulin Lispro |
| SITAGLIPTIN | SITAGLIPTIN | 42901620 | sitagliptin |
| NATEGLINIDE | NATEGLINIDE | 36232423 | nateglinide |
| INSULIN | INSULIN | 1544838 | Insulin, Glulisine, Human |
| INSULIN | INSULIN | 19078557 | Regular Insulin, Human |
| INSULIN | INSULIN | 1516981 | insulin detemir |
| GLYBURIDE | GLYBURIDE | 36233495 | Glyburide |
| INSULIN | INSULIN | 1567198 | Insulin, Aspart, Human |
| INSULIN | INSULIN | 1550027 | Insulin Lispro |
| METFORMIN | METFORMIN | 40164917 | Metformin |
| INSULIN | INSULIN | 40176981 | Insulin, Aspart Protamine, Human |
| INSULIN | INSULIN | 19062999 | insulin, isophane |
| GLIMEPIRIDE | GLIMEPIRIDE | 1597758 | glimepiride |
| ROSIGLITAZONE | ROSIGLITAZONE | 1547508 | rosiglitazone |

| | | | |
|---|---|---|---|
| INSULIN | INSULIN | 19078554 | Regular Insulin, Human |
| LIRAGLUTIDE | LIRAGLUTIDE | 40170913 | liraglutide |
| INSULIN | INSULIN | 1502905 | Insulin Glargine |
| INSULIN | INSULIN | 1550023 | Insulin Lispro |
| PIOGLITAZONE | PIOGLITAZONE | 19079293 | pioglitazone |
| GLIMEPIRIDE | GLIMEPIRIDE | 1597775 | glimepiride |
| INSULIN | INSULIN | 1516976 | insulin detemir |
| INSULIN | INSULIN | 36229086 | Insulin Glargine |
| METFORMIN | METFORMIN | 40164897 | Metformin |
| GLYBURIDE | GLYBURIDE | 19030575 | Glyburide |
| INSULIN | INSULIN | 1596960 | insulin, isophane |
| INSULIN | INSULIN | 19110219 | insulin, isophane |
| INSULIN | INSULIN | 1513881 | Insulin Lispro |
| REPAGLINIDE | REPAGLINIDE | 1516770 | repaglinide |
| SITAGLIPTIN | SITAGLIPTIN | 19125047 | sitagliptin |
| INSULIN | INSULIN | 19062999 | insulin, isophane |
| INSULIN | INSULIN | 19078559 | Insulin Glargine |
| INSULIN | INSULIN | 40184293 | Insulin, Aspart Protamine, Human |
| GLYBURIDE | GLYBURIDE | 1559684 | Glyburide |
| GLIPIZIDE | GLIPIZIDE | 19077637 | Glipizide |
| REPAGLINIDE | REPAGLINIDE | 19047663 | repaglinide |
| INSULIN | INSULIN | 19135263 | insulin, isophane |
| SITAGLIPTIN | SITAGLIPTIN | 19125051 | sitagliptin |
| BROMOCRIPTINE | BROMOCRIPTINE | 730551 | Bromocriptine |
| LIRAGLUTIDE | LIRAGLUTIDE | 45892686 | liraglutide |
| INSULIN | INSULIN | 1516981 | insulin detemir |
| ACARBOSE | ACARBOSE | 1529352 | Acarbose |
| INSULIN | INSULIN | 19110354 | insulin, isophane |
| INSULIN | INSULIN | 40184293 | Insulin, Aspart Protamine, Human |
| INSULIN | INSULIN | 36235843 | Ultralente Insulin, Human |
| GLIPIZIDE | GLIPIZIDE | 36218767 | Glipizide |
| GLIMEPIRIDE | GLIMEPIRIDE | 1597761 | glimepiride |
| METFORMIN | METFORMIN | 1503297 | Metformin |

| | | | |
|---|---|---|---|
| CANAGLIFLOZIN | CANAGLIFLOZIN | 45774720 | canagliflozin |
| GLIMEPIRIDE | GLIMEPIRIDE | 36229116 | glimepiride |
| INSULIN | INSULIN | 1567201 | Insulin, Aspart Protamine, Human |
| METFORMIN | METFORMIN | 40164892 | Metformin |
| INSULIN | INSULIN | 46233972 | Insulin Lispro |
| INSULIN | INSULIN | 19110355 | insulin, isophane |
| CANAGLIFLOZIN | CANAGLIFLOZIN | 43526470 | canagliflozin |
| EXENATIDE | EXENATIDE | 1583728 | exenatide |
| INSULIN | INSULIN | 19062999 | insulin, isophane |
| INSULIN | INSULIN | 36231459 | Regular Insulin, Human |
| GLIPIZIDE | GLIPIZIDE | 40044251 | Glipizide |
| INSULIN | INSULIN | 1516978 | insulin detemir |
| INSULIN | INSULIN | 1596977 | Regular Insulin, Human |
| METFORMIN | METFORMIN | 40164882 | Metformin |
| GLYBURIDE | GLYBURIDE | 40165995 | Glyburide |
| GLIPIZIDE | GLIPIZIDE | 40167634 | Glipizide |
| INSULIN | INSULIN | 19111797 | insulin, isophane |
| GLYBURIDE | GLYBURIDE | 40168370 | Glyburide |
| SAXAGLIPTIN | SAXAGLIPTIN | 40166042 | saxagliptin |
| INSULIN | INSULIN | 19062999 | insulin, isophane |
| LIRAGLUTIDE | LIRAGLUTIDE | 36248956 | liraglutide |
| BROMOCRIPTINE | BROMOCRIPTINE | 730572 | Bromocriptine |
| METFORMIN | METFORMIN | 40164898 | Metformin |
| MIGLITOL | MIGLITOL | 19067232 | miglitol |
| METFORMIN | METFORMIN | 40146999 | Metformin |
| SITAGLIPTIN | SITAGLIPTIN | 19125045 | sitagliptin |
| NATEGLINIDE | NATEGLINIDE | 1502826 | nateglinide |
| EXENATIDE | EXENATIDE | 19083254 | exenatide |
| SAXAGLIPTIN | SAXAGLIPTIN | 40166035 | saxagliptin |
| GLIMEPIRIDE | GLIMEPIRIDE | 19017142 | glimepiride |
| DAPAGLIFLOZIN | DAPAGLIFLOZIN | 36247907 | dapagliflozin |
| INSULIN | INSULIN | 19095256 | insulin, isophane |
| GLYBURIDE | GLYBURIDE | 36231169 | Glyburide |
| EMPAGLIFLOZIN | EMPAGLIFLOZIN | 45774895 | empagliflozin |

| | | | |
|---|---|---|---|
| INSULIN | INSULIN | 1596916 | insulin, isophane |
| PRAMLINTIDE | PRAMLINTIDE | 36243018 | Pramlintide |
| INSULIN | INSULIN | 1550028 | Insulin Lispro |
| INSULIN | INSULIN | 1516978 | insulin detemir |
| INSULIN | INSULIN | 40184293 | Insulin, Aspart Protamine, Human |
| INSULIN | INSULIN | 1596941 | insulin, isophane |
| INSULIN | INSULIN | 1596977 | Regular Insulin, Human |
| METFORMIN | METFORMIN | 40163925 | Metformin |
| EXENATIDE | EXENATIDE | 19083253 | exenatide |
| CHLORPROPAMIDE | CHLORPROPAMIDE | 19117691 | Chlorpropamide |
| INSULIN | INSULIN | 1562586 | Insulin, Zinc, Pork |
| INSULIN | INSULIN | 19120315 | insulin, isophane |
| LIRAGLUTIDE | LIRAGLUTIDE | 40169222 | liraglutide |
| LINAGLIPTIN | LINAGLIPTIN | 36229565 | Linagliptin |
| REPAGLINIDE | REPAGLINIDE | 19047663 | repaglinide |
| GLYBURIDE | GLYBURIDE | 19030576 | Glyburide |
| GLIPIZIDE | GLIPIZIDE | 19079990 | Glipizide |
| ALBIGLUTIDE | ALBIGLUTIDE | 44816274 | albiglutide |
| INSULIN | INSULIN | 1550027 | Insulin Lispro |
| PIOGLITAZONE | PIOGLITAZONE | 1525217 | pioglitazone |
| GLIPIZIDE | GLIPIZIDE | 40165972 | Glipizide |
| GLIPIZIDE | GLIPIZIDE | 1560171 | Glipizide |
| INSULIN | INSULIN | 1502905 | Insulin Glargine |
| INSULIN | INSULIN | 19100127 | insulin, isophane |
| INSULIN | INSULIN | 19120315 | insulin, isophane |
| METFORMIN | METFORMIN | 40164882 | Metformin |
| INSULIN | INSULIN | 35602728 | insulin degludec |
| EXENATIDE | EXENATIDE | 36238252 | exenatide |
| NATEGLINIDE | NATEGLINIDE | 1502829 | nateglinide |
| LIRAGLUTIDE | LIRAGLUTIDE | 40169222 | liraglutide |
| INSULIN | INSULIN | 19078554 | Regular Insulin, Human |
| PIOGLITAZONE | PIOGLITAZONE | 36236325 | pioglitazone |
| GLIMEPIRIDE | GLIMEPIRIDE | 1597776 | glimepiride |
| INSULIN | INSULIN | 1596953 | insulin, isophane |

| | | | |
|---|---|---|---|
| INSULIN | INSULIN | 19016117 | Insulin Lispro |
| INSULIN | INSULIN | 19129441 | insulin human, rDNA origin |
| INSULIN | INSULIN | 36229086 | Insulin Glargine |
| INSULIN | INSULIN | 19120315 | insulin, isophane |
| METFORMIN | METFORMIN | 40171451 | Metformin |
| GLIPIZIDE | GLIPIZIDE | 1560171 | Glipizide |
| INSULIN | INSULIN | 1550027 | Insulin Lispro |
| INSULIN | INSULIN | 19078553 | insulin, isophane |
| ACARBOSE | ACARBOSE | 1529352 | Acarbose |
| INSULIN | INSULIN | 19080091 | Ultralente Insulin, Human |
| INSULIN | INSULIN | 40166274 | Insulin Lispro |
| INSULIN | INSULIN | 19078553 | insulin, isophane |
| ROSIGLITAZONE | ROSIGLITAZONE | 19079465 | rosiglitazone |
| EXENATIDE | EXENATIDE | 1583730 | exenatide |
| GLYBURIDE | GLYBURIDE | 19045624 | Glyburide |
| INSULIN | INSULIN | 19120790 | Insulin Glargine |
| SAXAGLIPTIN | SAXAGLIPTIN | 40166038 | saxagliptin |
| ALOGLIPTIN | ALOGLIPTIN | 43013916 | alogliptin |
| GLIPIZIDE | GLIPIZIDE | 19081295 | Glipizide |
| INSULIN | INSULIN | 1516981 | insulin detemir |
| METFORMIN | METFORMIN | 42705990 | Metformin |
| EMPAGLIFLOZIN | EMPAGLIFLOZIN | 36248789 | empagliflozin |
| INSULIN | INSULIN | 40184293 | Insulin, Aspart Protamine, Human |
| METFORMIN | METFORMIN | 40164946 | Metformin |
| INSULIN | INSULIN | 19012679 | Regular Insulin, Human |
| GLYBURIDE | GLYBURIDE | 40165998 | Glyburide |
| INSULIN | INSULIN | 19014975 | insulin, isophane |
| LIRAGLUTIDE | LIRAGLUTIDE | 40169223 | liraglutide |
| METFORMIN | METFORMIN | 40164889 | Metformin |

| | | | |
|---|---|---|---|
| INSULIN | INSULIN | 19111795 | insulin, isophane |
| PIOGLITAZONE | PIOGLITAZONE | 36236324 | pioglitazone |
| INSULIN | INSULIN | 46233972 | Insulin Lispro |
| GLIMEPIRIDE | GLIMEPIRIDE | 36214984 | glimepiride |
| INSULIN | INSULIN | 19071700 | Insulin Glargine |
| INSULIN | INSULIN | 1567198 | Insulin, Aspart, Human |
| INSULIN | INSULIN | 35605622 | Regular Insulin, Human |
| INSULIN | INSULIN | 19078551 | insulin, isophane |
| INSULIN | INSULIN | 40052773 | Regular Insulin, Human |
| GLIPIZIDE | GLIPIZIDE | 1560171 | Glipizide |
| ROSIGLITAZONE | ROSIGLITAZONE | 1547505 | rosiglitazone |
| METFORMIN | METFORMIN | 1503297 | Metformin |
| ACARBOSE | ACARBOSE | 19047612 | Acarbose |
| INSULIN | INSULIN | 1567232 | Insulin, Aspart, Human |
| ROSIGLITAZONE | ROSIGLITAZONE | 1547505 | rosiglitazone |
| GLIMEPIRIDE | GLIMEPIRIDE | 43011499 | glimepiride |
| LIRAGLUTIDE | LIRAGLUTIDE | 40170911 | liraglutide |
| INSULIN | INSULIN | 1502905 | Insulin Glargine |
| INSULIN | INSULIN | 19078553 | insulin, isophane |
| INSULIN | INSULIN | 1596941 | insulin, isophane |
| REPAGLINIDE | REPAGLINIDE | 40098707 | repaglinide |
| GLIPIZIDE | GLIPIZIDE | 19134642 | Glipizide |
| INSULIN | INSULIN | 46275390 | Regular Insulin, Human |
| METFORMIN | METFORMIN | 40164899 | Metformin |
| GLYBURIDE | GLYBURIDE | 19079986 | Glyburide |
| BROMOCRIPTINE | BROMOCRIPTINE | 19007101 | Bromocriptine |
| INSULIN | INSULIN | 19044971 | insulin detemir |
| METFORMIN | METFORMIN | 40171449 | Metformin |
| GLIPIZIDE | GLIPIZIDE | 19030443 | Glipizide |
| EXENATIDE | EXENATIDE | 46234118 | exenatide |
| EXENATIDE | EXENATIDE | 1583730 | exenatide |
| INSULIN | INSULIN | 36231450 | Insulin, Aspart, Human |
| GLYBURIDE | GLYBURIDE | 40046201 | Glyburide |
| GLIPIZIDE | GLIPIZIDE | 40044251 | Glipizide |

| | | | |
|---|---|---|---|
| PIOGLITAZONE | PIOGLITAZONE | 1525218 | pioglitazone |
| INSULIN | INSULIN | 46221552 | Insulin Glargine |
| DULAGLUTIDE | DULAGLUTIDE | 45774442 | dulaglutide |
| GLIMEPIRIDE | GLIMEPIRIDE | 1597801 | glimepiride |
| ROSIGLITAZONE | ROSIGLITAZONE | 1547504 | rosiglitazone |
| PIOGLITAZONE | PIOGLITAZONE | 1525221 | pioglitazone |
| INSULIN | INSULIN | 36233552 | insulin, isophane |
| INSULIN | INSULIN | 19009384 | insulin, isophane |
| METFORMIN | METFORMIN | 40063356 | Metformin |
| METFORMIN | METFORMIN | 40164917 | Metformin |
| GLIMEPIRIDE | GLIMEPIRIDE | 1597799 | glimepiride |
| METFORMIN | METFORMIN | 40164886 | Metformin |
| INSULIN | INSULIN | 35605622 | Regular Insulin, Human |
| INSULIN | INSULIN | 42902743 | Insulin Glargine |
| INSULIN | INSULIN | 1502905 | Insulin Glargine |
| METFORMIN | METFORMIN | 42708173 | Metformin |
| SITAGLIPTIN | SITAGLIPTIN | 19125051 | sitagliptin |
| METFORMIN | METFORMIN | 40163929 | Metformin |
| METFORMIN | METFORMIN | 36236322 | Metformin |
| GLIPIZIDE | GLIPIZIDE | 40044224 | Glipizide |
| METFORMIN | METFORMIN | 40164920 | Metformin |
| INSULIN | INSULIN | 46233730 | Regular Insulin, Human |
| EXENATIDE | EXENATIDE | 46234118 | exenatide |
| GLYBURIDE | GLYBURIDE | 19077659 | Glyburide |
| INSULIN | INSULIN | 19078554 | Regular Insulin, Human |
| INSULIN | INSULIN | 46233972 | Insulin Lispro |
| INSULIN | INSULIN | 1586344 | insulin, isophane |
| INSULIN | INSULIN | 46234045 | Insulin, Aspart, Human |
| INSULIN | INSULIN | 19078557 | Regular Insulin, Human |
| EXENATIDE | EXENATIDE | 1583727 | exenatide |
| GLYBURIDE | GLYBURIDE | 36230035 | Glyburide |

| | | | |
|---|---|---|---|
| GLYBURIDE | GLYBURIDE | 40046201 | Glyburide |
| PIOGLITAZONE | PIOGLITAZONE | 1525220 | pioglitazone |
| PIOGLITAZONE | PIOGLITAZONE | 1525220 | pioglitazone |
| INSULIN | INSULIN | 19078557 | Regular Insulin, Human |
| METFORMIN | METFORMIN | 40164930 | Metformin |
| INSULIN | INSULIN | 19111795 | insulin, isophane |
| INSULIN | INSULIN | 1596977 | Regular Insulin, Human |
| INSULIN | INSULIN | 1596972 | insulin, isophane |
| GLYBURIDE | GLYBURIDE | 36221460 | Glyburide |
| GLYBURIDE | GLYBURIDE | 40168374 | Glyburide |
| BROMOCRIPTINE | BROMOCRIPTINE | 730551 | Bromocriptine |
| PIOGLITAZONE | PIOGLITAZONE | 1525217 | pioglitazone |
| BROMOCRIPTINE | BROMOCRIPTINE | 40016898 | Bromocriptine |
| INSULIN | INSULIN | 40176981 | Insulin, Aspart Protamine, Human |
| INSULIN | INSULIN | 19131582 | Insulin, Glulisine, Human |
| INSULIN | INSULIN | 19009384 | insulin, isophane |
| EXENATIDE | EXENATIDE | 19083254 | exenatide |
| CANAGLIFLOZIN | CANAGLIFLOZIN | 45774721 | canagliflozin |
| INSULIN | INSULIN | 1513912 | Insulin Lispro |
| INSULIN | INSULIN | 1515107 | insulin, isophane |
| BROMOCRIPTINE | BROMOCRIPTINE | 36234233 | Bromocriptine |
| METFORMIN | METFORMIN | 40164905 | Metformin |
| INSULIN | INSULIN | 1544838 | Insulin, Glulisine, Human |
| SITAGLIPTIN | SITAGLIPTIN | 19125042 | sitagliptin |
| DAPAGLIFLOZIN | DAPAGLIFLOZIN | 44785473 | dapagliflozin |
| EXENATIDE | EXENATIDE | 46234118 | exenatide |
| INSULIN | INSULIN | 46234043 | Insulin, Aspart, Human |
| INSULIN | INSULIN | 46234098 | Insulin Glargine |
| EMPAGLIFLOZIN | EMPAGLIFLOZIN | 45892174 | empagliflozin |
| INSULIN | INSULIN | 1513881 | Insulin Lispro |

| | | | |
|---|---|---|---|
| GLYBURIDE | GLYBURIDE | 40168370 | Glyburide |
| DULAGLUTIDE | DULAGLUTIDE | 45774440 | dulaglutide |
| LINAGLIPTIN | LINAGLIPTIN | 40239218 | Linagliptin |
| GLIPIZIDE | GLIPIZIDE | 36235748 | Glipizide |
| REPAGLINIDE | REPAGLINIDE | 19023425 | repaglinide |
| INSULIN | INSULIN | 40184293 | Insulin, Aspart Protamine, Human |
| METFORMIN | METFORMIN | 40164944 | Metformin |
| SITAGLIPTIN | SITAGLIPTIN | 19125044 | sitagliptin |
| GLIPIZIDE | GLIPIZIDE | 19080793 | Glipizide |
| EXENATIDE | EXENATIDE | 46234118 | exenatide |
| INSULIN | INSULIN | 19009384 | insulin, isophane |
| INSULIN | INSULIN | 1513912 | Insulin Lispro |
| METFORMIN | METFORMIN | 40164914 | Metformin |
| INSULIN | INSULIN | 36222190 | insulin detemir |
| DULAGLUTIDE | DULAGLUTIDE | 45774440 | dulaglutide |
| INSULIN | INSULIN | 1502910 | Insulin Glargine |
| ALOGLIPTIN | ALOGLIPTIN | 36246267 | alogliptin |
| INSULIN | INSULIN | 1596956 | insulin, isophane |
| EXENATIDE | EXENATIDE | 1583727 | exenatide |
| DAPAGLIFLOZIN | DAPAGLIFLOZIN | 36247907 | dapagliflozin |
| INSULIN | INSULIN | 19111795 | insulin, isophane |
| INSULIN | INSULIN | 36229086 | Insulin Glargine |
| DAPAGLIFLOZIN | DAPAGLIFLOZIN | 45775999 | dapagliflozin |
| INSULIN | INSULIN | 19078551 | insulin, isophane |
| DULAGLUTIDE | DULAGLUTIDE | 45774448 | dulaglutide |
| LIRAGLUTIDE | LIRAGLUTIDE | 40169222 | liraglutide |
| METFORMIN | METFORMIN | 40164946 | Metformin |
| INSULIN | INSULIN | 36235937 | insulin human, rDNA origin |
| INSULIN | INSULIN | 1550023 | Insulin Lispro |
| GLYBURIDE | GLYBURIDE | 40046204 | Glyburide |
| SITAGLIPTIN | SITAGLIPTIN | 19125041 | sitagliptin |

| | | | |
|---|---|---|---|
| INSULIN | INSULIN | 1567232 | Insulin, Aspart, Human |
| ALOGLIPTIN | ALOGLIPTIN | 43013925 | alogliptin |
| INSULIN | INSULIN | 40051348 | Insulin Glargine |
| METFORMIN | METFORMIN | 40164904 | Metformin |
| INSULIN | INSULIN | 19078552 | insulin, isophane |
| INSULIN | INSULIN | 35602726 | insulin degludec |
| METFORMIN | METFORMIN | 1503297 | Metformin |
| SITAGLIPTIN | SITAGLIPTIN | 40243629 | sitagliptin |
| LIRAGLUTIDE | LIRAGLUTIDE | 36220119 | liraglutide |
| INSULIN | INSULIN | 1513912 | Insulin Lispro |
| INSULIN | INSULIN | 19078633 | Insulin, Regular, Pork |
| METFORMIN | METFORMIN | 40063350 | Metformin |
| INSULIN | INSULIN | 19078553 | insulin, isophane |
| GLIMEPIRIDE | GLIMEPIRIDE | 36234086 | glimepiride |
| INSULIN | INSULIN | 19078557 | Regular Insulin, Human |
| INSULIN | INSULIN | 19078553 | insulin, isophane |
| DAPAGLIFLOZIN | DAPAGLIFLOZIN | 44785830 | dapagliflozin |
| INSULIN | INSULIN | 19078526 | Insulin, Zinc, Human |
| PIOGLITAZONE | PIOGLITAZONE | 1525220 | pioglitazone |
| LIRAGLUTIDE | LIRAGLUTIDE | 40170913 | liraglutide |
| GLIMEPIRIDE | GLIMEPIRIDE | 36214985 | glimepiride |
| INSULIN | INSULIN | 1502905 | Insulin Glargine |
| INSULIN | INSULIN | 19116453 | insulin, isophane |
| INSULIN | INSULIN | 19078559 | Insulin Glargine |
| INSULIN | INSULIN | 1550028 | Insulin Lispro |
| INSULIN | INSULIN | 19110219 | insulin, isophane |
| INSULIN | INSULIN | 40051347 | Insulin Glargine |
| EXENATIDE | EXENATIDE | 46234118 | exenatide |
| GLIPIZIDE | GLIPIZIDE | 40167636 | Glipizide |
| METFORMIN | METFORMIN | 40164929 | Metformin |
| NATEGLINIDE | NATEGLINIDE | 36223777 | nateglinide |
| INSULIN | INSULIN | 36235840 | insulin, isophane |

| | | | |
|---|---|---|---|
| INSULIN | INSULIN | 36231450 | Insulin, Aspart, Human |
| METFORMIN | METFORMIN | 40164929 | Metformin |
| INSULIN | INSULIN | 19111797 | insulin, isophane |
| METFORMIN | METFORMIN | 1503297 | Metformin |
| GLIPIZIDE | GLIPIZIDE | 1560233 | Glipizide |
| METFORMIN | METFORMIN | 40063353 | Metformin |
| INSULIN | INSULIN | 45776625 | Regular Insulin, Human |
| GLIPIZIDE | GLIPIZIDE | 19030443 | Glipizide |
| METFORMIN | METFORMIN | 40164892 | Metformin |
| EMPAGLIFLOZIN | EMPAGLIFLOZIN | 45774758 | empagliflozin |
| INSULIN | INSULIN | 1586345 | insulin, isophane |
| INSULIN | INSULIN | 19078554 | Regular Insulin, Human |
| CANAGLIFLOZIN | CANAGLIFLOZIN | 36246430 | canagliflozin |
| SITAGLIPTIN | SITAGLIPTIN | 36221232 | sitagliptin |
| INSULIN | INSULIN | 19009384 | insulin, isophane |
| GLIMEPIRIDE | GLIMEPIRIDE | 1597798 | glimepiride |
| EXENATIDE | EXENATIDE | 1583727 | exenatide |
| PRAMLINTIDE | PRAMLINTIDE | 40165687 | Pramlintide |
| ALOGLIPTIN | ALOGLIPTIN | 43013919 | alogliptin |
| EMPAGLIFLOZIN | EMPAGLIFLOZIN | 46287410 | empagliflozin |
| GLYBURIDE | GLYBURIDE | 19079986 | Glyburide |
| INSULIN | INSULIN | 19078557 | Regular Insulin, Human |
| INSULIN | INSULIN | 19120315 | insulin, isophane |
| INSULIN | INSULIN | 1596999 | Regular Insulin, Human |
| BROMOCRIPTINE | BROMOCRIPTINE | 730581 | Bromocriptine |
| GLYBURIDE | GLYBURIDE | 40166003 | Glyburide |
| INSULIN | INSULIN | 46221559 | Insulin Glargine |
| BROMOCRIPTINE | BROMOCRIPTINE | 730548 | Bromocriptine |
| INSULIN | INSULIN | 40184294 | Insulin, Aspart Protamine, Human |
| INSULIN | INSULIN | 1516979 | insulin detemir |

| | | | |
|---|---|---|---|
| GLIMEPIRIDE | GLIMEPIRIDE | 36222784 | glimepiride |
| INSULIN | INSULIN | 1550023 | Insulin Lispro |
| MIGLITOL | MIGLITOL | 1510207 | miglitol |
| METFORMIN | METFORMIN | 40164882 | Metformin |
| GLIMEPIRIDE | GLIMEPIRIDE | 19048034 | glimepiride |
| EXENATIDE | EXENATIDE | 1583730 | exenatide |
| INSULIN | INSULIN | 19100127 | insulin, isophane |
| INSULIN | INSULIN | 46233976 | Insulin Lispro |
| LIRAGLUTIDE | LIRAGLUTIDE | 40169223 | liraglutide |
| METFORMIN | METFORMIN | 40163928 | Metformin |
| INSULIN | INSULIN | 1550028 | Insulin Lispro |
| LINAGLIPTIN | LINAGLIPTIN | 42708089 | Linagliptin |
| GLIMEPIRIDE | GLIMEPIRIDE | 40059665 | glimepiride |
| METFORMIN | METFORMIN | 36235787 | Metformin |
| METFORMIN | METFORMIN | 40164940 | Metformin |
| INSULIN | INSULIN | 19078553 | insulin, isophane |
| METFORMIN | METFORMIN | 36235746 | Metformin |
| BROMOCRIPTINE | BROMOCRIPTINE | 40016898 | Bromocriptine |
| GLIPIZIDE | GLIPIZIDE | 40167635 | Glipizide |
| INSULIN | INSULIN | 19112694 | insulin detemir |
| GLYBURIDE | GLYBURIDE | 19077681 | Glyburide |
| LIRAGLUTIDE | LIRAGLUTIDE | 40170913 | liraglutide |
| INSULIN | INSULIN | 1513914 | Insulin Lispro |
| INSULIN | INSULIN | 1513912 | Insulin Lispro |
| NATEGLINIDE | NATEGLINIDE | 1502827 | nateglinide |
| EMPAGLIFLOZIN | EMPAGLIFLOZIN | 45774758 | empagliflozin |
| INSULIN | INSULIN | 19135265 | Regular Insulin, Human |
| GLIMEPIRIDE | GLIMEPIRIDE | 1597772 | glimepiride |
| INSULIN | INSULIN | 19116610 | insulin, isophane |
| LIRAGLUTIDE | LIRAGLUTIDE | 40170912 | liraglutide |
| PIOGLITAZONE | PIOGLITAZONE | 1525215 | pioglitazone |
| LINAGLIPTIN | LINAGLIPTIN | 40239218 | Linagliptin |

| | | | |
|---|---|---|---|
| INSULIN | INSULIN | 19078557 | Regular Insulin, Human |
| INSULIN | INSULIN | 1596998 | Regular Insulin, Human |
| GLIMEPIRIDE | GLIMEPIRIDE | 19135385 | glimepiride |
| INSULIN | INSULIN | 1567198 | Insulin, Aspart, Human |
| NATEGLINIDE | NATEGLINIDE | 36232424 | nateglinide |
| INSULIN | INSULIN | 1596916 | insulin, isophane |
| INSULIN | INSULIN | 19110355 | insulin, isophane |
| LIRAGLUTIDE | LIRAGLUTIDE | 36241903 | liraglutide |
| INSULIN | INSULIN | 19111795 | insulin, isophane |
| GLYBURIDE | GLYBURIDE | 19077659 | Glyburide |
| GLIPIZIDE | GLIPIZIDE | 36235748 | Glipizide |
| SITAGLIPTIN | SITAGLIPTIN | 19125049 | sitagliptin |
| METFORMIN | METFORMIN | 42708167 | Metformin |
| INSULIN | INSULIN | 19100127 | insulin, isophane |
| INSULIN | INSULIN | 19009384 | insulin, isophane |
| METFORMIN | METFORMIN | 40163928 | Metformin |
| REPAGLINIDE | REPAGLINIDE | 1516770 | repaglinide |
| LIRAGLUTIDE | LIRAGLUTIDE | 36241903 | liraglutide |
| INSULIN | INSULIN | 1596998 | Regular Insulin, Human |
| BROMOCRIPTINE | BROMOCRIPTINE | 19007102 | Bromocriptine |
| INSULIN | INSULIN | 19078554 | Regular Insulin, Human |
| ROSIGLITAZONE | ROSIGLITAZONE | 40099633 | rosiglitazone |
| METFORMIN | METFORMIN | 1503328 | Metformin |
| INSULIN | INSULIN | 1502910 | Insulin Glargine |
| NATEGLINIDE | NATEGLINIDE | 1502827 | nateglinide |
| GLIMEPIRIDE | GLIMEPIRIDE | 1597760 | glimepiride |
| INSULIN | INSULIN | 36248196 | insulin degludec |
| INSULIN | INSULIN | 1597000 | Regular Insulin, Human |
| GLYBURIDE | GLYBURIDE | 19079986 | Glyburide |
| INSULIN | INSULIN | 1502905 | Insulin Glargine |
| METFORMIN | METFORMIN | 40163928 | Metformin |
| ROSIGLITAZONE | ROSIGLITAZONE | 1547505 | rosiglitazone |

| | | | |
|---|---|---|---|
| GLIPIZIDE | GLIPIZIDE | 40165976 | Glipizide |
| GLIMEPIRIDE | GLIMEPIRIDE | 36222784 | glimepiride |
| PIOGLITAZONE | PIOGLITAZONE | 40099897 | pioglitazone |
| INSULIN | INSULIN | 1597001 | Regular Insulin, Human |
| REPAGLINIDE | REPAGLINIDE | 19047663 | repaglinide |
| METFORMIN | METFORMIN | 1503297 | Metformin |
| METFORMIN | METFORMIN | 36242511 | Metformin |
| ALOGLIPTIN | ALOGLIPTIN | 43013920 | alogliptin |
| INSULIN | INSULIN | 1550023 | Insulin Lispro |
| METFORMIN | METFORMIN | 40164889 | Metformin |
| METFORMIN | METFORMIN | 40164896 | Metformin |
| METFORMIN | METFORMIN | 40164915 | Metformin |
| INSULIN | INSULIN | 19078553 | insulin, isophane |
| SAXAGLIPTIN | SAXAGLIPTIN | 40166038 | saxagliptin |
| INSULIN | INSULIN | 46234045 | Insulin, Aspart, Human |
| BROMOCRIPTINE | BROMOCRIPTINE | 19007101 | Bromocriptine |
| LIRAGLUTIDE | LIRAGLUTIDE | 40169223 | liraglutide |
| NATEGLINIDE | NATEGLINIDE | 1502829 | nateglinide |
| INSULIN | INSULIN | 19116609 | insulin, isophane |
| INSULIN | INSULIN | 1550027 | Insulin Lispro |
| ACARBOSE | ACARBOSE | 19040970 | Acarbose |
| METFORMIN | METFORMIN | 36240427 | Metformin |
| METFORMIN | METFORMIN | 40231404 | Metformin |
| METFORMIN | METFORMIN | 19106521 | Metformin |
| METFORMIN | METFORMIN | 40164943 | Metformin |
| INSULIN | INSULIN | 36229086 | Insulin Glargine |
| INSULIN | INSULIN | 19078554 | Regular Insulin, Human |
| INSULIN | INSULIN | 40176981 | Insulin, Aspart Protamine, Human |
| METFORMIN | METFORMIN | 40164930 | Metformin |

| | | | |
|---|---|---|---|
| METFORMIN | METFORMIN | 40164886 | Metformin |
| LINAGLIPTIN | LINAGLIPTIN | 40239216 | Linagliptin |
| INSULIN | INSULIN | 19120790 | Insulin Glargine |
| INSULIN | INSULIN | 19062456 | insulin, isophane |
| GLIPIZIDE | GLIPIZIDE | 40165976 | Glipizide |
| GLYBURIDE | GLYBURIDE | 36233495 | Glyburide |
| INSULIN | INSULIN | 46221557 | Insulin Glargine |
| INSULIN | INSULIN | 46233730 | Regular Insulin, Human |
| BROMOCRIPTINE | BROMOCRIPTINE | 40164161 | Bromocriptine |
| INSULIN | INSULIN | 36231451 | Insulin, Aspart Protamine, Human |
| METFORMIN | METFORMIN | 19113780 | Metformin |
| METFORMIN | METFORMIN | 40164943 | Metformin |
| ALOGLIPTIN | ALOGLIPTIN | 43013931 | alogliptin |
| LINAGLIPTIN | LINAGLIPTIN | 36229565 | Linagliptin |
| ALOGLIPTIN | ALOGLIPTIN | 43013917 | alogliptin |
| INSULIN | INSULIN | 1516978 | insulin detemir |
| INSULIN | INSULIN | 36229086 | Insulin Glargine |
| PIOGLITAZONE | PIOGLITAZONE | 19079293 | pioglitazone |
| INSULIN | INSULIN | 1596955 | insulin, isophane |
| GLIMEPIRIDE | GLIMEPIRIDE | 1597756 | glimepiride |
| PRAMLINTIDE | PRAMLINTIDE | 1517998 | Pramlintide |
| INSULIN | INSULIN | 1502910 | Insulin Glargine |
| LINAGLIPTIN | LINAGLIPTIN | 36229565 | Linagliptin |
| METFORMIN | METFORMIN | 40063352 | Metformin |
| LIRAGLUTIDE | LIRAGLUTIDE | 40169223 | liraglutide |
| ALOGLIPTIN | ALOGLIPTIN | 43013900 | alogliptin |
| INSULIN | INSULIN | 36231459 | Regular Insulin, Human |
| INSULIN | INSULIN | 1596955 | insulin, isophane |
| NATEGLINIDE | NATEGLINIDE | 36232424 | nateglinide |
| INSULIN | INSULIN | 36227902 | Insulin Lispro |
| PIOGLITAZONE | PIOGLITAZONE | 1525221 | pioglitazone |
| INSULIN | INSULIN | 19104092 | Regular Insulin, Human |
| REPAGLINIDE | REPAGLINIDE | 1516770 | repaglinide |
| METFORMIN | METFORMIN | 1503327 | Metformin |
| GLYBURIDE | GLYBURIDE | 19077659 | Glyburide |
| ROSIGLITAZONE | ROSIGLITAZONE | 1547505 | rosiglitazone |

| | | | |
|---|---|---|---|
| EMPAGLIFLOZIN | EMPAGLIFLOZIN | 45774758 | empagliflozin |
| INSULIN | INSULIN | 1596999 | Regular Insulin, Human |
| INSULIN | INSULIN | 46233975 | Insulin Lispro |
| METFORMIN | METFORMIN | 40164897 | Metformin |
| INSULIN | INSULIN | 19078553 | insulin, isophane |
| INSULIN | INSULIN | 1567198 | Insulin, Aspart, Human |
| INSULIN | INSULIN | 19112694 | insulin detemir |
| INSULIN | INSULIN | 1597001 | Regular Insulin, Human |
| INSULIN | INSULIN | 1550028 | Insulin Lispro |
| PIOGLITAZONE | PIOGLITAZONE | 40099897 | pioglitazone |
| INSULIN | INSULIN | 36231450 | Insulin, Aspart, Human |
| SITAGLIPTIN | SITAGLIPTIN | 19125043 | sitagliptin |
| INSULIN | INSULIN | 1513849 | Insulin, Zinc, Human |
| CHLORPROPAMIDE | CHLORPROPAMIDE | 1594973 | Chlorpropamide |
| GLIMEPIRIDE | GLIMEPIRIDE | 1597756 | glimepiride |
| INSULIN | INSULIN | 19078555 | Regular Insulin, Human |
| GLYBURIDE | GLYBURIDE | 19030572 | Glyburide |
| INSULIN | INSULIN | 46233975 | Insulin Lispro |
| INSULIN | INSULIN | 19061587 | insulin, isophane |
| INSULIN | INSULIN | 19120790 | Insulin Glargine |
| INSULIN | INSULIN | 19120790 | Insulin Glargine |
| INSULIN | INSULIN | 36236032 | Insulin Lispro |
| GLIPIZIDE | GLIPIZIDE | 1560171 | Glipizide |
| SAXAGLIPTIN | SAXAGLIPTIN | 40166037 | saxagliptin |
| INSULIN | INSULIN | 19095211 | Insulin Lispro |
| METFORMIN | METFORMIN | 40163924 | Metformin |
| ROSIGLITAZONE | ROSIGLITAZONE | 1547508 | rosiglitazone |
| INSULIN | INSULIN | 36231458 | insulin, isophane |
| INSULIN | INSULIN | 19018822 | Insulin Lispro |
| EMPAGLIFLOZIN | EMPAGLIFLOZIN | 36246886 | empagliflozin |
| NATEGLINIDE | NATEGLINIDE | 1502829 | nateglinide |
| GLYBURIDE | GLYBURIDE | 19077681 | Glyburide |
| CANAGLIFLOZIN | CANAGLIFLOZIN | 43526465 | canagliflozin |
| EMPAGLIFLOZIN | EMPAGLIFLOZIN | 45892179 | empagliflozin |
| BROMOCRIPTINE | BROMOCRIPTINE | 19006288 | Bromocriptine |
| METFORMIN | METFORMIN | 1503297 | Metformin |

| | | | |
|---|---|---|---|
| LIRAGLUTIDE | LIRAGLUTIDE | 36241903 | liraglutide |
| METFORMIN | METFORMIN | 40229046 | Metformin |
| EMPAGLIFLOZIN | EMPAGLIFLOZIN | 1592795 | empagliflozin |
| GLYBURIDE | GLYBURIDE | 19063991 | Glyburide |
| GLIPIZIDE | GLIPIZIDE | 40165973 | Glipizide |
| GLYBURIDE | GLYBURIDE | 40168368 | Glyburide |
| GLYBURIDE | GLYBURIDE | 40165993 | Glyburide |
| INSULIN | INSULIN | 19071700 | Insulin Glargine |
| ALBIGLUTIDE | ALBIGLUTIDE | 44816274 | albiglutide |
| EMPAGLIFLOZIN | EMPAGLIFLOZIN | 45892179 | empagliflozin |
| GLIPIZIDE | GLIPIZIDE | 19079990 | Glipizide |
| INSULIN | INSULIN | 36231459 | Regular Insulin, Human |
| INSULIN | INSULIN | 1567198 | Insulin, Aspart, Human |
| EXENATIDE | EXENATIDE | 19083254 | exenatide |
| INSULIN | INSULIN | 19111797 | insulin, isophane |
| CHLORPROPAMIDE | CHLORPROPAMIDE | 40030456 | Chlorpropamide |
| INSULIN | INSULIN | 1596977 | Regular Insulin, Human |
| DAPAGLIFLOZIN | DAPAGLIFLOZIN | 44785477 | dapagliflozin |
| METFORMIN | METFORMIN | 40163923 | Metformin |
| METFORMIN | METFORMIN | 40164897 | Metformin |
| INSULIN | INSULIN | 1596977 | Regular Insulin, Human |
| INSULIN | INSULIN | 19078556 | Regular Insulin, Human |
| ACARBOSE | ACARBOSE | 19040969 | Acarbose |
| PIOGLITAZONE | PIOGLITAZONE | 1525216 | pioglitazone |
| METFORMIN | METFORMIN | 40164895 | Metformin |
| INSULIN | INSULIN | 1567198 | Insulin, Aspart, Human |
| INSULIN | INSULIN | 1550027 | Insulin Lispro |
| METFORMIN | METFORMIN | 1503297 | Metformin |
| BROMOCRIPTINE | BROMOCRIPTINE | 40016898 | Bromocriptine |
| ALBIGLUTIDE | ALBIGLUTIDE | 44816273 | albiglutide |
| METFORMIN | METFORMIN | 40164932 | Metformin |
| GLYBURIDE | GLYBURIDE | 19077682 | Glyburide |
| METFORMIN | METFORMIN | 1503328 | Metformin |
| METFORMIN | METFORMIN | 1503328 | Metformin |

| | | | |
|---|---|---|---|
| INSULIN | INSULIN | 1597012 | Regular Insulin, Human |
| INSULIN | INSULIN | 46233975 | Insulin Lispro |
| GLYBURIDE | GLYBURIDE | 19063991 | Glyburide |
| METFORMIN | METFORMIN | 40164896 | Metformin |
| INSULIN | INSULIN | 19078554 | Regular Insulin, Human |
| INSULIN | INSULIN | 19044971 | insulin detemir |
| PIOGLITAZONE | PIOGLITAZONE | 1525217 | pioglitazone |
| INSULIN | INSULIN | 1544838 | Insulin, Glulisine, Human |
| METFORMIN | METFORMIN | 40164942 | Metformin |
| GLIPIZIDE | GLIPIZIDE | 40165976 | Glipizide |
| REPAGLINIDE | REPAGLINIDE | 36213643 | repaglinide |
| ROSIGLITAZONE | ROSIGLITAZONE | 36229365 | rosiglitazone |
| INSULIN | INSULIN | 19135320 | Regular Insulin, Human |
| INSULIN | INSULIN | 19044971 | insulin detemir |
| INSULIN | INSULIN | 1596972 | insulin, isophane |
| INSULIN | INSULIN | 1597001 | Regular Insulin, Human |
| CHLORPROPAMIDE | CHLORPROPAMIDE | 19025944 | Chlorpropamide |
| DULAGLUTIDE | DULAGLUTIDE | 45774448 | dulaglutide |
| INSULIN | INSULIN | 1596959 | insulin, isophane |
| CANAGLIFLOZIN | CANAGLIFLOZIN | 43526472 | canagliflozin |
| METFORMIN | METFORMIN | 40163929 | Metformin |
| METFORMIN | METFORMIN | 40164889 | Metformin |
| METFORMIN | METFORMIN | 1503297 | Metformin |
| BROMOCRIPTINE | BROMOCRIPTINE | 730548 | Bromocriptine |
| ACETOHEXAMIDE | ACETOHEXAMIDE | 1530020 | Acetohexamide |
| GLYBURIDE | GLYBURIDE | 40165997 | Glyburide |
| INSULIN | INSULIN | 1596918 | insulin, isophane |
| EMPAGLIFLOZIN | EMPAGLIFLOZIN | 36248376 | empagliflozin |
| INSULIN | INSULIN | 1597000 | Regular Insulin, Human |
| INSULIN | INSULIN | 19111795 | insulin, isophane |
| GLIMEPIRIDE | GLIMEPIRIDE | 1597760 | glimepiride |
| METFORMIN | METFORMIN | 40164923 | Metformin |

| | | | |
|---|---|---|---|
| INSULIN | INSULIN | 1597012 | Regular Insulin, Human |
| EXENATIDE | EXENATIDE | 46234118 | exenatide |
| METFORMIN | METFORMIN | 40163925 | Metformin |
| DAPAGLIFLOZIN | DAPAGLIFLOZIN | 44785834 | dapagliflozin |
| ALBIGLUTIDE | ALBIGLUTIDE | 44816283 | albiglutide |
| INSULIN | INSULIN | 1502905 | Insulin Glargine |
| LIRAGLUTIDE | LIRAGLUTIDE | 40170911 | liraglutide |
| SAXAGLIPTIN | SAXAGLIPTIN | 40166042 | saxagliptin |
| INSULIN | INSULIN | 1596955 | insulin, isophane |
| INSULIN | INSULIN | 1502910 | Insulin Glargine |
| GLYBURIDE | GLYBURIDE | 36230036 | Glyburide |
| CANAGLIFLOZIN | CANAGLIFLOZIN | 43526468 | canagliflozin |
| BROMOCRIPTINE | BROMOCRIPTINE | 730551 | Bromocriptine |
| MIGLITOL | MIGLITOL | 19029030 | miglitol |
| INSULIN | INSULIN | 1516979 | insulin detemir |
| INSULIN | INSULIN | 1597000 | Regular Insulin, Human |
| SITAGLIPTIN | SITAGLIPTIN | 40136769 | sitagliptin |
| INSULIN | INSULIN | 46233972 | Insulin Lispro |
| CANAGLIFLOZIN | CANAGLIFLOZIN | 36246877 | canagliflozin |
| INSULIN | INSULIN | 19135320 | Regular Insulin, Human |
| METFORMIN | METFORMIN | 1503297 | Metformin |
| DAPAGLIFLOZIN | DAPAGLIFLOZIN | 45775999 | dapagliflozin |
| INSULIN | INSULIN | 1567200 | Insulin, Aspart, Human |
| METFORMIN | METFORMIN | 36235581 | Metformin |
| LINAGLIPTIN | LINAGLIPTIN | 42708087 | Linagliptin |
| GLIMEPIRIDE | GLIMEPIRIDE | 1597756 | glimepiride |
| CHLORPROPAMIDE | CHLORPROPAMIDE | 19116145 | Chlorpropamide |
| INSULIN | INSULIN | 1567198 | Insulin, Aspart, Human |
| GLYBURIDE | GLYBURIDE | 36235750 | Glyburide |
| INSULIN | INSULIN | 1550027 | Insulin Lispro |
| INSULIN | INSULIN | 19078557 | Regular Insulin, Human |
| INSULIN | INSULIN | 1502905 | Insulin Glargine |
| MIGLITOL | MIGLITOL | 19029029 | miglitol |
| INSULIN | INSULIN | 19095214 | insulin, isophane |
| MIGLITOL | MIGLITOL | 36233498 | miglitol |

| | | | |
|---|---|---|---|
| METFORMIN | METFORMIN | 40164905 | Metformin |
| GLIMEPIRIDE | GLIMEPIRIDE | 36234085 | glimepiride |
| INSULIN | INSULIN | 40051347 | Insulin Glargine |
| ACARBOSE | ACARBOSE | 19023063 | Acarbose |
| INSULIN | INSULIN | 1596916 | insulin, isophane |
| INSULIN | INSULIN | 46234045 | Insulin, Aspart, Human |
| GLIMEPIRIDE | GLIMEPIRIDE | 1597761 | glimepiride |
| INSULIN | INSULIN | 1550023 | Insulin Lispro |
| DULAGLUTIDE | DULAGLUTIDE | 45774440 | dulaglutide |
| INSULIN | INSULIN | 19135265 | Regular Insulin, Human |
| INSULIN | INSULIN | 19111795 | insulin, isophane |
| EMPAGLIFLOZIN | EMPAGLIFLOZIN | 45774895 | empagliflozin |
| SAXAGLIPTIN | SAXAGLIPTIN | 40166044 | saxagliptin |
| INSULIN | INSULIN | 19078553 | insulin, isophane |
| INSULIN | INSULIN | 19058398 | Insulin Lispro |
| METFORMIN | METFORMIN | 40164931 | Metformin |
| INSULIN | INSULIN | 1596952 | insulin, isophane |
| METFORMIN | METFORMIN | 40164884 | Metformin |
| LINAGLIPTIN | LINAGLIPTIN | 42705859 | Linagliptin |
| INSULIN | INSULIN | 1550028 | Insulin Lispro |
| GLIPIZIDE | GLIPIZIDE | 36241808 | Glipizide |
| METFORMIN | METFORMIN | 1503297 | Metformin |
| LIRAGLUTIDE | LIRAGLUTIDE | 36248956 | liraglutide |
| LIRAGLUTIDE | LIRAGLUTIDE | 40170911 | liraglutide |
| EMPAGLIFLOZIN | EMPAGLIFLOZIN | 45774754 | empagliflozin |
| INSULIN | INSULIN | 36233154 | Insulin, Glulisine, Human |
| INSULIN | INSULIN | 19129440 | insulin human, rDNA origin |
| GLYBURIDE | GLYBURIDE | 40165994 | Glyburide |
| INSULIN | INSULIN | 19078555 | Regular Insulin, Human |
| METFORMIN | METFORMIN | 40164886 | Metformin |
| GLIMEPIRIDE | GLIMEPIRIDE | 1597757 | glimepiride |
| INSULIN | INSULIN | 40166275 | Insulin Lispro |

| | | | |
|---|---|---|---|
| METFORMIN | METFORMIN | 40229046 | Metformin |
| INSULIN | INSULIN | 19110355 | insulin, isophane |
| INSULIN | INSULIN | 19078557 | Regular Insulin, Human |
| DAPAGLIFLOZIN | DAPAGLIFLOZIN | 44785477 | dapagliflozin |
| GLYBURIDE | GLYBURIDE | 40165994 | Glyburide |
| INSULIN | INSULIN | 46221557 | Insulin Glargine |
| INSULIN | INSULIN | 19100128 | Regular Insulin, Human |
| ROSIGLITAZONE | ROSIGLITAZONE | 36227624 | rosiglitazone |
| SITAGLIPTIN | SITAGLIPTIN | 42901620 | sitagliptin |
| INSULIN | INSULIN | 19078530 | insulin, isophane |
| INSULIN | INSULIN | 1596977 | Regular Insulin, Human |
| REPAGLINIDE | REPAGLINIDE | 19023424 | repaglinide |
| DAPAGLIFLOZIN | DAPAGLIFLOZIN | 45775388 | dapagliflozin |
| INSULIN | INSULIN | 1502908 | Insulin Glargine |
| INSULIN | INSULIN | 19078557 | Regular Insulin, Human |
| INSULIN | INSULIN | 19129442 | insulin human, rDNA origin |
| INSULIN | INSULIN | 46233946 | Insulin Lispro |
| METFORMIN | METFORMIN | 40164881 | Metformin |
| INSULIN | INSULIN | 1513881 | Insulin Lispro |
| ROSIGLITAZONE | ROSIGLITAZONE | 1547506 | rosiglitazone |
| ALBIGLUTIDE | ALBIGLUTIDE | 44816094 | albiglutide |
| INSULIN | INSULIN | 1502905 | Insulin Glargine |
| GLYBURIDE | GLYBURIDE | 36235750 | Glyburide |
| SAXAGLIPTIN | SAXAGLIPTIN | 40166038 | saxagliptin |
| GLIMEPIRIDE | GLIMEPIRIDE | 40059666 | glimepiride |
| GLYBURIDE | GLYBURIDE | 19077659 | Glyburide |
| INSULIN | INSULIN | 40176981 | Insulin, Aspart Protamine, Human |
| GLIPIZIDE | GLIPIZIDE | 1560171 | Glipizide |
| METFORMIN | METFORMIN | 40163925 | Metformin |

| | | | |
|---|---|---|---|
| GLYBURIDE | GLYBURIDE | 40168374 | Glyburide |
| METFORMIN | METFORMIN | 40164929 | Metformin |
| METFORMIN | METFORMIN | 40163924 | Metformin |
| GLIPIZIDE | GLIPIZIDE | 36218768 | Glipizide |
| INSULIN | INSULIN | 1597012 | Regular Insulin, Human |
| ROSIGLITAZONE | ROSIGLITAZONE | 1547510 | rosiglitazone |
| METFORMIN | METFORMIN | 40164919 | Metformin |
| NATEGLINIDE | NATEGLINIDE | 1502830 | nateglinide |
| INSULIN | INSULIN | 46233976 | Insulin Lispro |
| METFORMIN | METFORMIN | 1503327 | Metformin |
| INSULIN | INSULIN | 19116559 | insulin, isophane |
| DAPAGLIFLOZIN | DAPAGLIFLOZIN | 45775860 | dapagliflozin |
| GLYBURIDE | GLYBURIDE | 19077659 | Glyburide |
| METFORMIN | METFORMIN | 1503297 | Metformin |
| INSULIN | INSULIN | 1596941 | insulin, isophane |
| INSULIN | INSULIN | 1516981 | insulin detemir |
| INSULIN | INSULIN | 1502908 | Insulin Glargine |
| INSULIN | INSULIN | 19110219 | insulin, isophane |
| GLIPIZIDE | GLIPIZIDE | 40165969 | Glipizide |
| CANAGLIFLOZIN | CANAGLIFLOZIN | 43526472 | canagliflozin |
| GLIMEPIRIDE | GLIMEPIRIDE | 40059666 | glimepiride |
| GLIMEPIRIDE | GLIMEPIRIDE | 19122367 | glimepiride |
| METFORMIN | METFORMIN | 40164896 | Metformin |
| INSULIN | INSULIN | 1502905 | Insulin Glargine |
| EMPAGLIFLOZIN | EMPAGLIFLOZIN | 46287685 | empagliflozin |
| INSULIN | INSULIN | 1516978 | insulin detemir |
| ALBIGLUTIDE | ALBIGLUTIDE | 44816086 | albiglutide |
| INSULIN | INSULIN | 1513881 | Insulin Lispro |
| INSULIN | INSULIN | 19078554 | Regular Insulin, Human |
| INSULIN | INSULIN | 1718711 | insulin degludec |
| GLYBURIDE | GLYBURIDE | 36235750 | Glyburide |
| INSULIN | INSULIN | 1567198 | Insulin, Aspart, Human |

| | | | |
|---|---|---|---|
| INSULIN | INSULIN | 19129441 | insulin human, rDNA origin |
| LINAGLIPTIN | LINAGLIPTIN | 40239219 | Linagliptin |
| GLYBURIDE | GLYBURIDE | 19079986 | Glyburide |
| GLYBURIDE | GLYBURIDE | 19060409 | Glyburide |
| INSULIN | INSULIN | 19062999 | insulin, isophane |
| DULAGLUTIDE | DULAGLUTIDE | 45774442 | dulaglutide |
| GLYBURIDE | GLYBURIDE | 19106860 | Glyburide |
| METFORMIN | METFORMIN | 40229048 | Metformin |
| PIOGLITAZONE | PIOGLITAZONE | 40099898 | pioglitazone |
| INSULIN | INSULIN | 40051348 | Insulin Glargine |
| GLYBURIDE | GLYBURIDE | 40165998 | Glyburide |
| INSULIN | INSULIN | 19111797 | insulin, isophane |
| EXENATIDE | EXENATIDE | 19083254 | exenatide |
| EXENATIDE | EXENATIDE | 1583727 | exenatide |
| INSULIN | INSULIN | 1596955 | insulin, isophane |
| INSULIN | INSULIN | 19071700 | Insulin Glargine |
| INSULIN | INSULIN | 1550027 | Insulin Lispro |
| METFORMIN | METFORMIN | 1503328 | Metformin |
| GLIMEPIRIDE | GLIMEPIRIDE | 1597773 | glimepiride |
| LIRAGLUTIDE | LIRAGLUTIDE | 36241903 | liraglutide |
| INSULIN | INSULIN | 19078553 | insulin, isophane |
| GLIMEPIRIDE | GLIMEPIRIDE | 36229116 | glimepiride |
| INSULIN | INSULIN | 46233948 | Insulin Lispro |
| BROMOCRIPTINE | BROMOCRIPTINE | 19116172 | Bromocriptine |
| INSULIN | INSULIN | 19110356 | Regular Insulin, Human |
| INSULIN | INSULIN | 1567201 | Insulin, Aspart Protamine, Human |
| PRAMLINTIDE | PRAMLINTIDE | 40165688 | Pramlintide |
| INSULIN | INSULIN | 40176981 | Insulin, Aspart Protamine, Human |
| INSULIN | INSULIN | 19120315 | insulin, isophane |
| EXENATIDE | EXENATIDE | 19083254 | exenatide |
| INSULIN | INSULIN | 45776625 | Regular Insulin, Human |
| PRAMLINTIDE | PRAMLINTIDE | 40165686 | Pramlintide |

| | | | |
|---|---|---|---|
| BROMOCRIPTINE | BROMOCRIPTINE | 36238866 | Bromocriptine |
| DULAGLUTIDE | DULAGLUTIDE | 45774445 | dulaglutide |
| LIRAGLUTIDE | LIRAGLUTIDE | 40170911 | liraglutide |
| BROMOCRIPTINE | BROMOCRIPTINE | 730603 | Bromocriptine |
| METFORMIN | METFORMIN | 1503297 | Metformin |
| CHLORPROPAMIDE | CHLORPROPAMIDE | 36231727 | Chlorpropamide |
| METFORMIN | METFORMIN | 36223145 | Metformin |
| INSULIN | INSULIN | 1586344 | insulin, isophane |
| INSULIN | INSULIN | 19062999 | insulin, isophane |
| ALBIGLUTIDE | ALBIGLUTIDE | 44816282 | albiglutide |
| METFORMIN | METFORMIN | 40231404 | Metformin |
| INSULIN | INSULIN | 46234241 | Regular Insulin, Human |
| GLYBURIDE | GLYBURIDE | 40046143 | Glyburide |
| ALBIGLUTIDE | ALBIGLUTIDE | 44816274 | albiglutide |
| GLYBURIDE | GLYBURIDE | 36213837 | Glyburide |
| GLIMEPIRIDE | GLIMEPIRIDE | 1597756 | glimepiride |
| INSULIN | INSULIN | 1502905 | Insulin Glargine |
| INSULIN | INSULIN | 19018822 | Insulin Lispro |
| SITAGLIPTIN | SITAGLIPTIN | 42901620 | sitagliptin |
| INSULIN | INSULIN | 19078553 | insulin, isophane |
| GLIMEPIRIDE | GLIMEPIRIDE | 1597775 | glimepiride |
| INSULIN | INSULIN | 19120315 | insulin, isophane |
| INSULIN | INSULIN | 19078554 | Regular Insulin, Human |
| METFORMIN | METFORMIN | 40164905 | Metformin |
| INSULIN | INSULIN | 36231450 | Insulin, Aspart, Human |
| NATEGLINIDE | NATEGLINIDE | 40058198 | nateglinide |
| ACARBOSE | ACARBOSE | 19040969 | Acarbose |
| GLIPIZIDE | GLIPIZIDE | 19081295 | Glipizide |
| DAPAGLIFLOZIN | DAPAGLIFLOZIN | 44785831 | dapagliflozin |
| INSULIN | INSULIN | 1596953 | insulin, isophane |
| GLYBURIDE | GLYBURIDE | 1559782 | Glyburide |
| MIGLITOL | MIGLITOL | 1510206 | miglitol |

| | | | |
|---|---|---|---|
| INSULIN | INSULIN | 19010204 | insulin, isophane |
| INSULIN | INSULIN | 46233946 | Insulin Lispro |
| GLIMEPIRIDE | GLIMEPIRIDE | 1597759 | glimepiride |
| NATEGLINIDE | NATEGLINIDE | 19107111 | nateglinide |
| INSULIN | INSULIN | 36236033 | Insulin Lispro |
| EXENATIDE | EXENATIDE | 1583730 | exenatide |
| INSULIN | INSULIN | 40051347 | Insulin Glargine |
| INSULIN | INSULIN | 40051348 | Insulin Glargine |
| INSULIN | INSULIN | 46233976 | Insulin Lispro |
| INSULIN | INSULIN | 19078553 | insulin, isophane |
| INSULIN | INSULIN | 1596999 | Regular Insulin, Human |
| METFORMIN | METFORMIN | 1503297 | Metformin |
| INSULIN | INSULIN | 1567198 | Insulin, Aspart, Human |
| GLYBURIDE | GLYBURIDE | 36235751 | Glyburide |
| NATEGLINIDE | NATEGLINIDE | 36232423 | nateglinide |
| INSULIN | INSULIN | 19135265 | Regular Insulin, Human |
| PRAMLINTIDE | PRAMLINTIDE | 40165688 | Pramlintide |
| INSULIN | INSULIN | 1502905 | Insulin Glargine |
| INSULIN | INSULIN | 1596959 | insulin, isophane |
| INSULIN | INSULIN | 19078558 | Insulin, Aspart, Human |
| INSULIN | INSULIN | 1513881 | Insulin Lispro |
| INSULIN | INSULIN | 19120790 | Insulin Glargine |
| METFORMIN | METFORMIN | 40229046 | Metformin |
| GLYBURIDE | GLYBURIDE | 40165994 | Glyburide |
| INSULIN | INSULIN | 1550023 | Insulin Lispro |
| EXENATIDE | EXENATIDE | 19083254 | exenatide |
| ACETOHEXAMIDE | ACETOHEXAMIDE | 19088427 | Acetohexamide |
| PRAMLINTIDE | PRAMLINTIDE | 40165687 | Pramlintide |
| METFORMIN | METFORMIN | 40229046 | Metformin |
| REPAGLINIDE | REPAGLINIDE | 19023426 | repaglinide |
| DAPAGLIFLOZIN | DAPAGLIFLOZIN | 44785473 | dapagliflozin |
| INSULIN | INSULIN | 46234043 | Insulin, Aspart, Human |

| | | | |
|---|---|---|---|
| INSULIN | INSULIN | 19078551 | insulin, isophane |
| LIRAGLUTIDE | LIRAGLUTIDE | 40169223 | liraglutide |
| INSULIN | INSULIN | 1567232 | Insulin, Aspart, Human |
| GLYBURIDE | GLYBURIDE | 19030572 | Glyburide |
| DULAGLUTIDE | DULAGLUTIDE | 45774435 | dulaglutide |
| METFORMIN | METFORMIN | 19106521 | Metformin |
| GLIPIZIDE | GLIPIZIDE | 40165974 | Glipizide |
| EMPAGLIFLOZIN | EMPAGLIFLOZIN | 46287409 | empagliflozin |
| INSULIN | INSULIN | 19078553 | insulin, isophane |
| CANAGLIFLOZIN | CANAGLIFLOZIN | 45774714 | canagliflozin |
| INSULIN | INSULIN | 1562586 | Insulin, Zinc, Pork |
| BROMOCRIPTINE | BROMOCRIPTINE | 19134681 | Bromocriptine |
| ACARBOSE | ACARBOSE | 19023063 | Acarbose |
| GLIMEPIRIDE | GLIMEPIRIDE | 1597757 | glimepiride |
| INSULIN | INSULIN | 19009384 | insulin, isophane |
| INSULIN | INSULIN | 1596998 | Regular Insulin, Human |
| INSULIN | INSULIN | 1596957 | insulin, isophane |
| INSULIN | INSULIN | 40166274 | Insulin Lispro |
| INSULIN | INSULIN | 36248870 | Insulin Glargine |
| METFORMIN | METFORMIN | 42708171 | Metformin |
| SAXAGLIPTIN | SAXAGLIPTIN | 40166041 | saxagliptin |
| INSULIN | INSULIN | 46233972 | Insulin Lispro |
| METFORMIN | METFORMIN | 40164886 | Metformin |
| GLIMEPIRIDE | GLIMEPIRIDE | 1597775 | glimepiride |
| SITAGLIPTIN | SITAGLIPTIN | 19125041 | sitagliptin |
| DULAGLUTIDE | DULAGLUTIDE | 45774447 | dulaglutide |
| INSULIN | INSULIN | 46233975 | Insulin Lispro |
| CHLORPROPAMIDE | CHLORPROPAMIDE | 1594976 | Chlorpropamide |
| GLIPIZIDE | GLIPIZIDE | 36241808 | Glipizide |
| METFORMIN | METFORMIN | 40164932 | Metformin |

| | | | |
|---|---|---|---|
| INSULIN | INSULIN | 19080091 | Ultralente Insulin, Human |
| SITAGLIPTIN | SITAGLIPTIN | 40136769 | sitagliptin |
| INSULIN | INSULIN | 46221552 | Insulin Glargine |
| INSULIN | INSULIN | 45777022 | Regular Insulin, Human |
| INSULIN | INSULIN | 46233975 | Insulin Lispro |
| EXENATIDE | EXENATIDE | 42902689 | exenatide |
| INSULIN | INSULIN | 19009384 | insulin, isophane |
| CANAGLIFLOZIN | CANAGLIFLOZIN | 43526472 | canagliflozin |
| INSULIN | INSULIN | 1596977 | Regular Insulin, Human |
| INSULIN | INSULIN | 1597001 | Regular Insulin, Human |
| INSULIN | INSULIN | 1567232 | Insulin, Aspart, Human |
| GLIMEPIRIDE | GLIMEPIRIDE | 19018043 | glimepiride |
| EXENATIDE | EXENATIDE | 19083253 | exenatide |
| METFORMIN | METFORMIN | 36235746 | Metformin |
| INSULIN | INSULIN | 19135320 | Regular Insulin, Human |
| INSULIN | INSULIN | 46221557 | Insulin Glargine |
| BROMOCRIPTINE | BROMOCRIPTINE | 19062900 | Bromocriptine |
| GLIMEPIRIDE | GLIMEPIRIDE | 36214984 | glimepiride |
| INSULIN | INSULIN | 19078558 | Insulin, Aspart, Human |
| PIOGLITAZONE | PIOGLITAZONE | 1525220 | pioglitazone |
| INSULIN | INSULIN | 1502910 | Insulin Glargine |
| INSULIN | INSULIN | 1550028 | Insulin Lispro |
| PRAMLINTIDE | PRAMLINTIDE | 40165688 | Pramlintide |
| INSULIN | INSULIN | 19120315 | insulin, isophane |
| ACARBOSE | ACARBOSE | 19047612 | Acarbose |
| INSULIN | INSULIN | 19010204 | insulin, isophane |
| INSULIN | INSULIN | 19078557 | Regular Insulin, Human |
| INSULIN | INSULIN | 19078554 | Regular Insulin, Human |
| CHLORPROPAMIDE | CHLORPROPAMIDE | 19006907 | Chlorpropamide |
| EXENATIDE | EXENATIDE | 1583727 | exenatide |
| METFORMIN | METFORMIN | 36236322 | Metformin |
| INSULIN | INSULIN | 46234045 | Insulin, Aspart, Human |

| | | | |
|---|---|---|---|
| GLYBURIDE | GLYBURIDE | 36221460 | Glyburide |
| INSULIN | INSULIN | 19135320 | Regular Insulin, Human |
| EXENATIDE | EXENATIDE | 1583727 | exenatide |
| INSULIN | INSULIN | 19078555 | Regular Insulin, Human |
| NATEGLINIDE | NATEGLINIDE | 19071495 | nateglinide |
| METFORMIN | METFORMIN | 1503328 | Metformin |
| INSULIN | INSULIN | 1596977 | Regular Insulin, Human |
| GLIMEPIRIDE | GLIMEPIRIDE | 36214984 | glimepiride |
| METFORMIN | METFORMIN | 1503297 | Metformin |
| INSULIN | INSULIN | 1567232 | Insulin, Aspart, Human |
| INSULIN | INSULIN | 46233945 | Insulin Lispro |
| METFORMIN | METFORMIN | 40063352 | Metformin |
| INSULIN | INSULIN | 19078530 | insulin, isophane |
| METFORMIN | METFORMIN | 40231404 | Metformin |
| EXENATIDE | EXENATIDE | 46234118 | exenatide |
| SITAGLIPTIN | SITAGLIPTIN | 19125041 | sitagliptin |
| INSULIN | INSULIN | 19078559 | Insulin Glargine |
| GLIMEPIRIDE | GLIMEPIRIDE | 40059666 | glimepiride |
| LIRAGLUTIDE | LIRAGLUTIDE | 40170913 | liraglutide |
| METFORMIN | METFORMIN | 1503297 | Metformin |
| DAPAGLIFLOZIN | DAPAGLIFLOZIN | 44816049 | dapagliflozin |
| INSULIN | INSULIN | 19110352 | insulin, isophane |
| INSULIN | INSULIN | 19009384 | insulin, isophane |
| INSULIN | INSULIN | 36231450 | Insulin, Aspart, Human |
| PIOGLITAZONE | PIOGLITAZONE | 1525221 | pioglitazone |
| INSULIN | INSULIN | 19071700 | Insulin Glargine |
| LINAGLIPTIN | LINAGLIPTIN | 42708087 | Linagliptin |
| INSULIN | INSULIN | 46234240 | insulin, isophane |
| INSULIN | INSULIN | 1567198 | Insulin, Aspart, Human |
| INSULIN | INSULIN | 1596960 | insulin, isophane |
| METFORMIN | METFORMIN | 1503297 | Metformin |
| GLIPIZIDE | GLIPIZIDE | 40167636 | Glipizide |
| METFORMIN | METFORMIN | 42708175 | Metformin |

| | | | |
|---|---|---|---|
| INSULIN | INSULIN | 1502905 | Insulin Glargine |
| METFORMIN | METFORMIN | 36235747 | Metformin |
| GLIPIZIDE | GLIPIZIDE | 40166815 | Glipizide |
| CANAGLIFLOZIN | CANAGLIFLOZIN | 45774714 | canagliflozin |
| INSULIN | INSULIN | 46233978 | Insulin Lispro |
| INSULIN | INSULIN | 40051348 | Insulin Glargine |
| GLIPIZIDE | GLIPIZIDE | 40044253 | Glipizide |
| LIRAGLUTIDE | LIRAGLUTIDE | 36241903 | liraglutide |
| INSULIN | INSULIN | 46233975 | Insulin Lispro |
| INSULIN | INSULIN | 1586344 | insulin, isophane |
| INSULIN | INSULIN | 1567198 | Insulin, Aspart, Human |
| GLIMEPIRIDE | GLIMEPIRIDE | 19135386 | glimepiride |
| METFORMIN | METFORMIN | 40164925 | Metformin |
| CANAGLIFLOZIN | CANAGLIFLOZIN | 43526474 | canagliflozin |
| INSULIN | INSULIN | 1596977 | Regular Insulin, Human |
| INSULIN | INSULIN | 36231458 | insulin, isophane |
| EXENATIDE | EXENATIDE | 42903341 | exenatide |
| ALOGLIPTIN | ALOGLIPTIN | 43013925 | alogliptin |
| GLYBURIDE | GLYBURIDE | 40166002 | Glyburide |
| INSULIN | INSULIN | 1596977 | Regular Insulin, Human |
| INSULIN | INSULIN | 19100128 | Regular Insulin, Human |
| INSULIN | INSULIN | 19120790 | Insulin Glargine |
| INSULIN | INSULIN | 1550027 | Insulin Lispro |
| INSULIN | INSULIN | 1550028 | Insulin Lispro |
| GLIPIZIDE | GLIPIZIDE | 19134624 | Glipizide |
| METFORMIN | METFORMIN | 36242511 | Metformin |
| INSULIN | INSULIN | 19080091 | Ultralente Insulin, Human |
| DULAGLUTIDE | DULAGLUTIDE | 45774448 | dulaglutide |
| CHLORPROPAMIDE | CHLORPROPAMIDE | 1594976 | Chlorpropamide |
| METFORMIN | METFORMIN | 1503297 | Metformin |
| INSULIN | INSULIN | 40176981 | Insulin, Aspart Protamine, Human |
| INSULIN | INSULIN | 1544873 | Insulin, Glulisine, Human |
| SITAGLIPTIN | SITAGLIPTIN | 19125045 | sitagliptin |
| GLIPIZIDE | GLIPIZIDE | 40165970 | Glipizide |
| ACARBOSE | ACARBOSE | 40003412 | Acarbose |
| DULAGLUTIDE | DULAGLUTIDE | 45774447 | dulaglutide |

| | | | |
|---|---|---|---|
| INSULIN | INSULIN | 1596977 | Regular Insulin, Human |
| LINAGLIPTIN | LINAGLIPTIN | 40239219 | Linagliptin |
| INSULIN | INSULIN | 19120315 | insulin, isophane |
| INSULIN | INSULIN | 1513849 | Insulin, Zinc, Human |
| METFORMIN | METFORMIN | 40229050 | Metformin |
| INSULIN | INSULIN | 19078551 | insulin, isophane |
| GLYBURIDE | GLYBURIDE | 40168374 | Glyburide |
| PRAMLINTIDE | PRAMLINTIDE | 40165687 | Pramlintide |
| INSULIN | INSULIN | 46234045 | Insulin, Aspart, Human |
| LINAGLIPTIN | LINAGLIPTIN | 36238284 | Linagliptin |
| METFORMIN | METFORMIN | 40163926 | Metformin |
| METFORMIN | METFORMIN | 1503327 | Metformin |
| METFORMIN | METFORMIN | 40164880 | Metformin |
| PIOGLITAZONE | PIOGLITAZONE | 19099208 | pioglitazone |
| INSULIN | INSULIN | 1596941 | insulin, isophane |
| GLIPIZIDE | GLIPIZIDE | 40165971 | Glipizide |
| GLIPIZIDE | GLIPIZIDE | 19077638 | Glipizide |
| LINAGLIPTIN | LINAGLIPTIN | 42708091 | Linagliptin |
| INSULIN | INSULIN | 19110219 | insulin, isophane |
| GLIPIZIDE | GLIPIZIDE | 40044224 | Glipizide |
| INSULIN | INSULIN | 1513914 | Insulin Lispro |
| INSULIN | INSULIN | 19078603 | insulin, isophane |
| EXENATIDE | EXENATIDE | 19135404 | exenatide |
| CANAGLIFLOZIN | CANAGLIFLOZIN | 43526467 | canagliflozin |
| NATEGLINIDE | NATEGLINIDE | 36232423 | nateglinide |
| LINAGLIPTIN | LINAGLIPTIN | 42708087 | Linagliptin |
| GLIPIZIDE | GLIPIZIDE | 40165972 | Glipizide |
| METFORMIN | METFORMIN | 36235752 | Metformin |
| ACARBOSE | ACARBOSE | 19023063 | Acarbose |
| BROMOCRIPTINE | BROMOCRIPTINE | 40164161 | Bromocriptine |
| METFORMIN | METFORMIN | 40164911 | Metformin |

| | | | |
|---|---|---|---|
| INSULIN | INSULIN | 1567198 | Insulin, Aspart, Human |
| GLIMEPIRIDE | GLIMEPIRIDE | 1597756 | glimepiride |
| INSULIN | INSULIN | 19120315 | insulin, isophane |
| METFORMIN | METFORMIN | 40229046 | Metformin |
| INSULIN | INSULIN | 1513916 | Insulin Lispro |
| INSULIN | INSULIN | 1516976 | insulin detemir |
| MIGLITOL | MIGLITOL | 19029029 | miglitol |
| GLYBURIDE | GLYBURIDE | 19030576 | Glyburide |
| DAPAGLIFLOZIN | DAPAGLIFLOZIN | 36247908 | dapagliflozin |
| EXENATIDE | EXENATIDE | 19083254 | exenatide |
| DAPAGLIFLOZIN | DAPAGLIFLOZIN | 44785833 | dapagliflozin |
| INSULIN | INSULIN | 1513916 | Insulin Lispro |
| INSULIN | INSULIN | 19078553 | insulin, isophane |
| ROSIGLITAZONE | ROSIGLITAZONE | 1547510 | rosiglitazone |
| GLYBURIDE | GLYBURIDE | 19030576 | Glyburide |
| INSULIN | INSULIN | 1596941 | insulin, isophane |
| INSULIN | INSULIN | 19116559 | insulin, isophane |
| CANAGLIFLOZIN | CANAGLIFLOZIN | 43526468 | canagliflozin |
| DAPAGLIFLOZIN | DAPAGLIFLOZIN | 45775860 | dapagliflozin |
| INSULIN | INSULIN | 35602730 | insulin degludec |
| INSULIN | INSULIN | 1567198 | Insulin, Aspart, Human |
| CANAGLIFLOZIN | CANAGLIFLOZIN | 45774721 | canagliflozin |
| EXENATIDE | EXENATIDE | 1583730 | exenatide |
| INSULIN | INSULIN | 19110220 | Regular Insulin, Human |
| EXENATIDE | EXENATIDE | 46234117 | exenatide |
| INSULIN | INSULIN | 1596959 | insulin, isophane |
| INSULIN | INSULIN | 19078552 | insulin, isophane |
| ACARBOSE | ACARBOSE | 19120257 | Acarbose |
| REPAGLINIDE | REPAGLINIDE | 19023425 | repaglinide |
| METFORMIN | METFORMIN | 40164891 | Metformin |

| | | | |
|---|---|---|---|
| LINAGLIPTIN | LINAGLIPTIN | 42708087 | Linagliptin |
| INSULIN | INSULIN | 19078559 | Insulin Glargine |
| INSULIN | INSULIN | 1596952 | insulin, isophane |
| ROSIGLITAZONE | ROSIGLITAZONE | 1547506 | rosiglitazone |
| INSULIN | INSULIN | 36231456 | insulin, isophane |
| GLYBURIDE | GLYBURIDE | 19077681 | Glyburide |
| INSULIN | INSULIN | 19078610 | insulin, regular, beef-pork |
| LIRAGLUTIDE | LIRAGLUTIDE | 36241903 | liraglutide |
| SITAGLIPTIN | SITAGLIPTIN | 36221233 | sitagliptin |
| METFORMIN | METFORMIN | 40164919 | Metformin |
| INSULIN | INSULIN | 19078551 | insulin, isophane |
| METFORMIN | METFORMIN | 36236323 | Metformin |
| INSULIN | INSULIN | 1596960 | insulin, isophane |
| INSULIN | INSULIN | 1550027 | Insulin Lispro |
| INSULIN | INSULIN | 1502908 | Insulin Glargine |
| ALBIGLUTIDE | ALBIGLUTIDE | 44816283 | albiglutide |
| CANAGLIFLOZIN | CANAGLIFLOZIN | 36246430 | canagliflozin |
| METFORMIN | METFORMIN | 40063352 | Metformin |
| GLIMEPIRIDE | GLIMEPIRIDE | 1597772 | glimepiride |
| INSULIN | INSULIN | 1596960 | insulin, isophane |
| REPAGLINIDE | REPAGLINIDE | 36240429 | repaglinide |
| SAXAGLIPTIN | SAXAGLIPTIN | 40166038 | saxagliptin |
| DAPAGLIFLOZIN | DAPAGLIFLOZIN | 44785473 | dapagliflozin |
| GLYBURIDE | GLYBURIDE | 1559786 | Glyburide |
| PRAMLINTIDE | PRAMLINTIDE | 1517998 | Pramlintide |
| INSULIN | INSULIN | 36229086 | Insulin Glargine |
| INSULIN | INSULIN | 1550023 | Insulin Lispro |
| INSULIN | INSULIN | 40166275 | Insulin Lispro |
| INSULIN | INSULIN | 1596959 | insulin, isophane |
| INSULIN | INSULIN | 1550023 | Insulin Lispro |
| INSULIN | INSULIN | 40176981 | Insulin, Aspart Protamine, Human |
| MIGLITOL | MIGLITOL | 1510202 | miglitol |
| PIOGLITAZONE | PIOGLITAZONE | 40099898 | pioglitazone |
| INSULIN | INSULIN | 40051347 | Insulin Glargine |

| | | | |
|---|---|---|---|
| EXENATIDE | EXENATIDE | 19083253 | exenatide |
| GLIMEPIRIDE | GLIMEPIRIDE | 1597796 | glimepiride |
| DULAGLUTIDE | DULAGLUTIDE | 45774436 | dulaglutide |
| REPAGLINIDE | REPAGLINIDE | 1516771 | repaglinide |
| METFORMIN | METFORMIN | 40164900 | Metformin |
| SITAGLIPTIN | SITAGLIPTIN | 19125051 | sitagliptin |
| CANAGLIFLOZIN | CANAGLIFLOZIN | 43526472 | canagliflozin |
| INSULIN | INSULIN | 19131582 | Insulin, Glulisine, Human |
| INSULIN | INSULIN | 46233945 | Insulin Lispro |
| SITAGLIPTIN | SITAGLIPTIN | 19125041 | sitagliptin |
| GLIMEPIRIDE | GLIMEPIRIDE | 19135386 | glimepiride |
| GLIPIZIDE | GLIPIZIDE | 40165973 | Glipizide |
| GLIMEPIRIDE | GLIMEPIRIDE | 19018043 | glimepiride |
| GLYBURIDE | GLYBURIDE | 19063991 | Glyburide |
| INSULIN | INSULIN | 19110352 | insulin, isophane |
| METFORMIN | METFORMIN | 40164923 | Metformin |
| INSULIN | INSULIN | 1596941 | insulin, isophane |
| GLIPIZIDE | GLIPIZIDE | 19077638 | Glipizide |
| INSULIN | INSULIN | 19078527 | Insulin, Zinc, Human |
| INSULIN | INSULIN | 40166274 | Insulin Lispro |
| INSULIN | INSULIN | 1550023 | Insulin Lispro |
| GLIMEPIRIDE | GLIMEPIRIDE | 1597792 | glimepiride |
| INSULIN | INSULIN | 40166274 | Insulin Lispro |
| REPAGLINIDE | REPAGLINIDE | 19023424 | repaglinide |
| INSULIN | INSULIN | 19100127 | insulin, isophane |
| PRAMLINTIDE | PRAMLINTIDE | 40127005 | Pramlintide |
| SAXAGLIPTIN | SAXAGLIPTIN | 40166041 | saxagliptin |
| METFORMIN | METFORMIN | 40164926 | Metformin |
| INSULIN | INSULIN | 19131582 | Insulin, Glulisine, Human |
| INSULIN | INSULIN | 19014975 | insulin, isophane |
| TOLAZAMIDE | TOLAZAMIDE | 1502818 | Tolazamide |
| EMPAGLIFLOZIN | EMPAGLIFLOZIN | 45774895 | empagliflozin |
| INSULIN | INSULIN | 1550023 | Insulin Lispro |
| INSULIN | INSULIN | 19120790 | Insulin Glargine |

| | | | |
|---|---|---|---|
| GLYBURIDE | GLYBURIDE | 40046211 | Glyburide |
| BROMOCRIPTINE | BROMOCRIPTINE | 40016898 | Bromocriptine |
| INSULIN | INSULIN | 19116559 | insulin, isophane |
| PIOGLITAZONE | PIOGLITAZONE | 40099898 | pioglitazone |
| GLIPIZIDE | GLIPIZIDE | 40166815 | Glipizide |
| INSULIN | INSULIN | 19131582 | Insulin, Glulisine, Human |
| GLIMEPIRIDE | GLIMEPIRIDE | 19135386 | glimepiride |
| INSULIN | INSULIN | 1597012 | Regular Insulin, Human |
| GLIPIZIDE | GLIPIZIDE | 19030443 | Glipizide |
| INSULIN | INSULIN | 19062999 | insulin, isophane |
| INSULIN | INSULIN | 1567198 | Insulin, Aspart, Human |
| CANAGLIFLOZIN | CANAGLIFLOZIN | 45774717 | canagliflozin |
| INSULIN | INSULIN | 36229086 | Insulin Glargine |
| LINAGLIPTIN | LINAGLIPTIN | 36229565 | Linagliptin |
| REPAGLINIDE | REPAGLINIDE | 40098707 | repaglinide |
| INSULIN | INSULIN | 1567198 | Insulin, Aspart, Human |
| INSULIN | INSULIN | 19078555 | Regular Insulin, Human |
| INSULIN | INSULIN | 1550028 | Insulin Lispro |
| METFORMIN | METFORMIN | 40164917 | Metformin |
| CANAGLIFLOZIN | CANAGLIFLOZIN | 43526475 | canagliflozin |
| PRAMLINTIDE | PRAMLINTIDE | 40165687 | Pramlintide |
| INSULIN | INSULIN | 1502905 | Insulin Glargine |
| INSULIN | INSULIN | 1596960 | insulin, isophane |
| INSULIN | INSULIN | 1596941 | insulin, isophane |
| GLIMEPIRIDE | GLIMEPIRIDE | 1597798 | glimepiride |
| INSULIN | INSULIN | 19120315 | insulin, isophane |
| INSULIN | INSULIN | 1596977 | Regular Insulin, Human |
| EMPAGLIFLOZIN | EMPAGLIFLOZIN | 45892176 | empagliflozin |
| INSULIN | INSULIN | 19078559 | Insulin Glargine |
| INSULIN | INSULIN | 40052088 | Insulin Lispro |
| NATEGLINIDE | NATEGLINIDE | 40058199 | nateglinide |
| METFORMIN | METFORMIN | 1503297 | Metformin |

| | | | |
|---|---|---|---|
| INSULIN | INSULIN | 19078551 | insulin, isophane |
| METFORMIN | METFORMIN | 40164929 | Metformin |
| INSULIN | INSULIN | 19078557 | Regular Insulin, Human |
| METFORMIN | METFORMIN | 1503297 | Metformin |
| INSULIN | INSULIN | 1597000 | Regular Insulin, Human |
| INSULIN | INSULIN | 40184293 | Insulin, Aspart Protamine, Human |
| INSULIN | INSULIN | 46233948 | Insulin Lispro |
| INSULIN | INSULIN | 19078552 | insulin, isophane |
| TOLBUTAMIDE | TOLBUTAMIDE | 1502860 | Tolbutamide |
| GLYBURIDE | GLYBURIDE | 19060491 | Glyburide |
| METFORMIN | METFORMIN | 40164899 | Metformin |
| GLYBURIDE | GLYBURIDE | 19030576 | Glyburide |
| METFORMIN | METFORMIN | 40063354 | Metformin |
| EMPAGLIFLOZIN | EMPAGLIFLOZIN | 46287410 | empagliflozin |
| INSULIN | INSULIN | 19110355 | insulin, isophane |
| LIRAGLUTIDE | LIRAGLUTIDE | 45892686 | liraglutide |
| INSULIN | INSULIN | 19116559 | insulin, isophane |
| INSULIN | INSULIN | 19018822 | Insulin Lispro |
| INSULIN | INSULIN | 1513912 | Insulin Lispro |
| GLYBURIDE | GLYBURIDE | 36213841 | Glyburide |
| INSULIN | INSULIN | 19062999 | insulin, isophane |
| METFORMIN | METFORMIN | 1503297 | Metformin |
| GLIMEPIRIDE | GLIMEPIRIDE | 1597799 | glimepiride |
| GLYBURIDE | GLYBURIDE | 36231169 | Glyburide |
| SAXAGLIPTIN | SAXAGLIPTIN | 40166042 | saxagliptin |
| METFORMIN | METFORMIN | 40164944 | Metformin |
| INSULIN | INSULIN | 1513881 | Insulin Lispro |
| INSULIN | INSULIN | 40051347 | Insulin Glargine |
| METFORMIN | METFORMIN | 40164927 | Metformin |

| | | | |
|---|---|---|---|
| INSULIN | INSULIN | 19110219 | insulin, isophane |
| INSULIN | INSULIN | 19078530 | insulin, isophane |
| INSULIN | INSULIN | 19095214 | insulin, isophane |
| GLIMEPIRIDE | GLIMEPIRIDE | 1597774 | glimepiride |
| INSULIN | INSULIN | 19078553 | insulin, isophane |
| INSULIN | INSULIN | 1502908 | Insulin Glargine |
| INSULIN | INSULIN | 46234241 | Regular Insulin, Human |
| DULAGLUTIDE | DULAGLUTIDE | 45774442 | dulaglutide |
| INSULIN | INSULIN | 19078553 | insulin, isophane |
| INSULIN | INSULIN | 46233730 | Regular Insulin, Human |
| MIGLITOL | MIGLITOL | 1510206 | miglitol |
| INSULIN | INSULIN | 19062999 | insulin, isophane |
| METFORMIN | METFORMIN | 1503297 | Metformin |
| SITAGLIPTIN | SITAGLIPTIN | 1580747 | sitagliptin |
| GLIPIZIDE | GLIPIZIDE | 36235749 | Glipizide |
| GLIMEPIRIDE | GLIMEPIRIDE | 1597757 | glimepiride |
| DAPAGLIFLOZIN | DAPAGLIFLOZIN | 45775388 | dapagliflozin |
| LINAGLIPTIN | LINAGLIPTIN | 40239218 | Linagliptin |
| INSULIN | INSULIN | 19080091 | Ultralente Insulin, Human |
| INSULIN | INSULIN | 1586328 | insulin, isophane |
| METFORMIN | METFORMIN | 36223145 | Metformin |
| CANAGLIFLOZIN | CANAGLIFLOZIN | 43526475 | canagliflozin |
| REPAGLINIDE | REPAGLINIDE | 36213643 | repaglinide |
| INSULIN | INSULIN | 19078558 | Insulin, Aspart, Human |
| METFORMIN | METFORMIN | 40164882 | Metformin |
| GLYBURIDE | GLYBURIDE | 36230035 | Glyburide |
| INSULIN | INSULIN | 1596952 | insulin, isophane |
| METFORMIN | METFORMIN | 36235581 | Metformin |
| EMPAGLIFLOZIN | EMPAGLIFLOZIN | 46287678 | empagliflozin |
| GLIMEPIRIDE | GLIMEPIRIDE | 1597760 | glimepiride |
| EMPAGLIFLOZIN | EMPAGLIFLOZIN | 45774753 | empagliflozin |
| NATEGLINIDE | NATEGLINIDE | 36223777 | nateglinide |
| INSULIN | INSULIN | 1502910 | Insulin Glargine |
| ROSIGLITAZONE | ROSIGLITAZONE | 19107077 | rosiglitazone |

| | | | |
|---|---|---|---|
| NATEGLINIDE | NATEGLINIDE | 40058198 | nateglinide |
| INSULIN | INSULIN | 1502905 | Insulin Glargine |
| METFORMIN | METFORMIN | 40164897 | Metformin |
| SITAGLIPTIN | SITAGLIPTIN | 40136769 | sitagliptin |
| INSULIN | INSULIN | 1550023 | Insulin Lispro |
| INSULIN | INSULIN | 19111797 | insulin, isophane |
| BROMOCRIPTINE | BROMOCRIPTINE | 40016897 | Bromocriptine |
| INSULIN | INSULIN | 1596957 | insulin, isophane |
| GLYBURIDE | GLYBURIDE | 19030573 | Glyburide |
| GLYBURIDE | GLYBURIDE | 36233495 | Glyburide |
| INSULIN | INSULIN | 1550023 | Insulin Lispro |
| METFORMIN | METFORMIN | 40163926 | Metformin |
| INSULIN | INSULIN | 1550027 | Insulin Lispro |
| INSULIN | INSULIN | 19058398 | Insulin Lispro |
| INSULIN | INSULIN | 1596972 | insulin, isophane |
| METFORMIN | METFORMIN | 1503297 | Metformin |
| BROMOCRIPTINE | BROMOCRIPTINE | 19083957 | Bromocriptine |
| GLYBURIDE | GLYBURIDE | 40168370 | Glyburide |
| EXENATIDE | EXENATIDE | 19135404 | exenatide |
| INSULIN | INSULIN | 1502905 | Insulin Glargine |
| INSULIN | INSULIN | 1567198 | Insulin, Aspart, Human |
| INSULIN | INSULIN | 19110355 | insulin, isophane |
| INSULIN | INSULIN | 1513881 | Insulin Lispro |
| GLYBURIDE | GLYBURIDE | 1559685 | Glyburide |
| INSULIN | INSULIN | 1502910 | Insulin Glargine |
| ALBIGLUTIDE | ALBIGLUTIDE | 44816283 | albiglutide |
| EXENATIDE | EXENATIDE | 19083254 | exenatide |
| SAXAGLIPTIN | SAXAGLIPTIN | 40166042 | saxagliptin |
| DULAGLUTIDE | DULAGLUTIDE | 45774448 | dulaglutide |
| LIRAGLUTIDE | LIRAGLUTIDE | 40170911 | liraglutide |
| INSULIN | INSULIN | 19078527 | Insulin, Zinc, Human |
| INSULIN | INSULIN | 1516980 | insulin detemir |
| INSULIN | INSULIN | 1596999 | Regular Insulin, Human |

| | | | |
|---|---|---|---|
| INSULIN | INSULIN | 1596999 | Regular Insulin, Human |
| INSULIN | INSULIN | 19110356 | Regular Insulin, Human |
| INSULIN | INSULIN | 1596999 | Regular Insulin, Human |
| INSULIN | INSULIN | 46221552 | Insulin Glargine |
| INSULIN | INSULIN | 46234045 | Insulin, Aspart, Human |
| NATEGLINIDE | NATEGLINIDE | 1502830 | nateglinide |
| INSULIN | INSULIN | 1597000 | Regular Insulin, Human |
| GLYBURIDE | GLYBURIDE | 40046143 | Glyburide |
| REPAGLINIDE | REPAGLINIDE | 1516766 | repaglinide |
| INSULIN | INSULIN | 19078553 | insulin, isophane |
| INSULIN | INSULIN | 46234240 | insulin, isophane |
| INSULIN | INSULIN | 40051347 | Insulin Glargine |
| INSULIN | INSULIN | 1502905 | Insulin Glargine |
| METFORMIN | METFORMIN | 42708167 | Metformin |
| METFORMIN | METFORMIN | 40164900 | Metformin |
| METFORMIN | METFORMIN | 40063352 | Metformin |
| GLIMEPIRIDE | GLIMEPIRIDE | 36222784 | glimepiride |
| METFORMIN | METFORMIN | 40063356 | Metformin |
| TOLAZAMIDE | TOLAZAMIDE | 1502809 | Tolazamide |
| REPAGLINIDE | REPAGLINIDE | 19023425 | repaglinide |
| INSULIN | INSULIN | 1502908 | Insulin Glargine |
| EMPAGLIFLOZIN | EMPAGLIFLOZIN | 46287684 | empagliflozin |
| GLIPIZIDE | GLIPIZIDE | 40167634 | Glipizide |
| INSULIN | INSULIN | 46233945 | Insulin Lispro |
| SITAGLIPTIN | SITAGLIPTIN | 40243629 | sitagliptin |
| INSULIN | INSULIN | 1567232 | Insulin, Aspart, Human |
| LINAGLIPTIN | LINAGLIPTIN | 42705865 | Linagliptin |
| INSULIN | INSULIN | 1550027 | Insulin Lispro |
| GLIPIZIDE | GLIPIZIDE | 19085004 | Glipizide |
| INSULIN | INSULIN | 36235841 | Regular Insulin, Human |
| PRAMLINTIDE | PRAMLINTIDE | 1517998 | Pramlintide |
| INSULIN | INSULIN | 19116455 | insulin, isophane |

| | | | |
|---|---|---|---|
| ALOGLIPTIN | ALOGLIPTIN | 43013925 | alogliptin |
| INSULIN | INSULIN | 19095218 | Regular Insulin, Human |
| ROSIGLITAZONE | ROSIGLITAZONE | 36229365 | rosiglitazone |
| INSULIN | INSULIN | 19095214 | insulin, isophane |
| METFORMIN | METFORMIN | 40164914 | Metformin |
| SAXAGLIPTIN | SAXAGLIPTIN | 40166041 | saxagliptin |
| INSULIN | INSULIN | 19078553 | insulin, isophane |
| DULAGLUTIDE | DULAGLUTIDE | 45774442 | dulaglutide |
| METFORMIN | METFORMIN | 40164928 | Metformin |
| INSULIN | INSULIN | 45774400 | Regular Insulin, Human |
| GLYBURIDE | GLYBURIDE | 19079986 | Glyburide |
| GLIPIZIDE | GLIPIZIDE | 40165976 | Glipizide |
| INSULIN | INSULIN | 19120790 | Insulin Glargine |
| GLIMEPIRIDE | GLIMEPIRIDE | 1597798 | glimepiride |
| ACARBOSE | ACARBOSE | 19040969 | Acarbose |
| INSULIN | INSULIN | 1567198 | Insulin, Aspart, Human |
| INSULIN | INSULIN | 1596977 | Regular Insulin, Human |
| INSULIN | INSULIN | 36248195 | insulin degludec |
| INSULIN | INSULIN | 19135263 | insulin, isophane |
| INSULIN | INSULIN | 1567198 | Insulin, Aspart, Human |
| METFORMIN | METFORMIN | 40164940 | Metformin |
| INSULIN | INSULIN | 19044971 | insulin detemir |
| METFORMIN | METFORMIN | 40164940 | Metformin |
| METFORMIN | METFORMIN | 40164943 | Metformin |
| INSULIN | INSULIN | 1516976 | insulin detemir |
| DULAGLUTIDE | DULAGLUTIDE | 45774442 | dulaglutide |
| INSULIN | INSULIN | 19058398 | Insulin Lispro |
| INSULIN | INSULIN | 1513912 | Insulin Lispro |
| EXENATIDE | EXENATIDE | 1583730 | exenatide |
| INSULIN | INSULIN | 36235843 | Ultralente Insulin, Human |

| | | | |
|---|---|---|---|
| GLIPIZIDE | GLIPIZIDE | 40165970 | Glipizide |
| METFORMIN | METFORMIN | 40164880 | Metformin |
| INSULIN | INSULIN | 1567198 | Insulin, Aspart, Human |
| INSULIN | INSULIN | 19100128 | Regular Insulin, Human |
| INSULIN | INSULIN | 46233730 | Regular Insulin, Human |
| PIOGLITAZONE | PIOGLITAZONE | 40099897 | pioglitazone |
| INSULIN | INSULIN | 1567198 | Insulin, Aspart, Human |
| INSULIN | INSULIN | 19094122 | Regular Insulin, Human |
| INSULIN | INSULIN | 19018822 | Insulin Lispro |
| INSULIN | INSULIN | 1596999 | Regular Insulin, Human |
| ROSIGLITAZONE | ROSIGLITAZONE | 1547507 | rosiglitazone |
| GLYBURIDE | GLYBURIDE | 19106860 | Glyburide |
| INSULIN | INSULIN | 19131582 | Insulin, Glulisine, Human |
| GLIPIZIDE | GLIPIZIDE | 19077636 | Glipizide |
| METFORMIN | METFORMIN | 40164917 | Metformin |
| SITAGLIPTIN | SITAGLIPTIN | 19125040 | sitagliptin |
| EMPAGLIFLOZIN | EMPAGLIFLOZIN | 45774751 | empagliflozin |
| INSULIN | INSULIN | 1596941 | insulin, isophane |
| GLIMEPIRIDE | GLIMEPIRIDE | 1597756 | glimepiride |
| GLIPIZIDE | GLIPIZIDE | 40165975 | Glipizide |
| INSULIN | INSULIN | 19078555 | Regular Insulin, Human |
| LIRAGLUTIDE | LIRAGLUTIDE | 36241903 | liraglutide |
| INSULIN | INSULIN | 36227902 | Insulin Lispro |
| METFORMIN | METFORMIN | 40164938 | Metformin |
| GLIPIZIDE | GLIPIZIDE | 19077636 | Glipizide |
| INSULIN | INSULIN | 1596916 | insulin, isophane |
| INSULIN | INSULIN | 19120790 | Insulin Glargine |
| INSULIN | INSULIN | 19095218 | Regular Insulin, Human |
| GLYBURIDE | GLYBURIDE | 19106860 | Glyburide |
| INSULIN | INSULIN | 19078607 | insulin, isophane |
| CANAGLIFLOZIN | CANAGLIFLOZIN | 36246431 | canagliflozin |
| METFORMIN | METFORMIN | 1503328 | Metformin |
| GLYBURIDE | GLYBURIDE | 1559684 | Glyburide |
| METFORMIN | METFORMIN | 40164922 | Metformin |

| | | | |
|---|---|---|---|
| INSULIN | INSULIN | 19100127 | insulin, isophane |
| INSULIN | INSULIN | 1586345 | insulin, isophane |
| DULAGLUTIDE | DULAGLUTIDE | 45774448 | dulaglutide |
| EMPAGLIFLOZIN | EMPAGLIFLOZIN | 36246886 | empagliflozin |
| INSULIN | INSULIN | 46234045 | Insulin, Aspart, Human |
| DAPAGLIFLOZIN | DAPAGLIFLOZIN | 44815995 | dapagliflozin |
| EMPAGLIFLOZIN | EMPAGLIFLOZIN | 36248789 | empagliflozin |
| INSULIN | INSULIN | 1567201 | Insulin, Aspart Protamine, Human |
| GLYBURIDE | GLYBURIDE | 40165997 | Glyburide |
| GLYBURIDE | GLYBURIDE | 1559684 | Glyburide |
| INSULIN | INSULIN | 1596957 | insulin, isophane |
| MIGLITOL | MIGLITOL | 19029029 | miglitol |
| GLYBURIDE | GLYBURIDE | 1559685 | Glyburide |
| INSULIN | INSULIN | 1567198 | Insulin, Aspart, Human |
| EXENATIDE | EXENATIDE | 19083254 | exenatide |
| INSULIN | INSULIN | 19078530 | insulin, isophane |
| GLYBURIDE | GLYBURIDE | 40165993 | Glyburide |
| METFORMIN | METFORMIN | 40164900 | Metformin |
| INSULIN | INSULIN | 46233946 | Insulin Lispro |
| INSULIN | INSULIN | 36222190 | insulin detemir |
| INSULIN | INSULIN | 19078530 | insulin, isophane |
| INSULIN | INSULIN | 46233730 | Regular Insulin, Human |
| INSULIN | INSULIN | 19110356 | Regular Insulin, Human |
| ACARBOSE | ACARBOSE | 40003412 | Acarbose |
| INSULIN | INSULIN | 19120315 | insulin, isophane |
| GLYBURIDE | GLYBURIDE | 1559784 | Glyburide |
| INSULIN | INSULIN | 19062999 | insulin, isophane |
| INSULIN | INSULIN | 19104458 | insulin, isophane |
| ROSIGLITAZONE | ROSIGLITAZONE | 19099245 | rosiglitazone |
| INSULIN | INSULIN | 36231450 | Insulin, Aspart, Human |
| INSULIN | INSULIN | 19062999 | insulin, isophane |

| | | | |
|---|---|---|---|
| GLIMEPIRIDE | GLIMEPIRIDE | 1597756 | glimepiride |
| METFORMIN | METFORMIN | 36235752 | Metformin |
| GLYBURIDE | GLYBURIDE | 19030575 | Glyburide |
| METFORMIN | METFORMIN | 42708169 | Metformin |
| NATEGLINIDE | NATEGLINIDE | 1502827 | nateglinide |
| INSULIN | INSULIN | 19078530 | insulin, isophane |
| INSULIN | INSULIN | 19078633 | Insulin, Regular, Pork |
| GLYBURIDE | GLYBURIDE | 40046211 | Glyburide |
| INSULIN | INSULIN | 1502905 | Insulin Glargine |
| INSULIN | INSULIN | 19120790 | Insulin Glargine |
| INSULIN | INSULIN | 35602731 | insulin degludec |
| METFORMIN | METFORMIN | 40163929 | Metformin |
| INSULIN | INSULIN | 19078557 | Regular Insulin, Human |
| GLIMEPIRIDE | GLIMEPIRIDE | 19117200 | glimepiride |
| INSULIN | INSULIN | 19095214 | insulin, isophane |
| ALOGLIPTIN | ALOGLIPTIN | 43013906 | alogliptin |
| INSULIN | INSULIN | 19009384 | insulin, isophane |
| INSULIN | INSULIN | 19078557 | Regular Insulin, Human |
| INSULIN | INSULIN | 36235841 | Regular Insulin, Human |
| INSULIN | INSULIN | 19110220 | Regular Insulin, Human |
| INSULIN | INSULIN | 19082658 | Insulin, Zinc, Pork |
| GLYBURIDE | GLYBURIDE | 19063991 | Glyburide |
| LIRAGLUTIDE | LIRAGLUTIDE | 40169223 | liraglutide |
| METFORMIN | METFORMIN | 42708175 | Metformin |
| INSULIN | INSULIN | 1562586 | Insulin, Zinc, Pork |
| INSULIN | INSULIN | 19110219 | insulin, isophane |
| INSULIN | INSULIN | 1567198 | Insulin, Aspart, Human |
| METFORMIN | METFORMIN | 40164896 | Metformin |
| METFORMIN | METFORMIN | 40164893 | Metformin |
| INSULIN | INSULIN | 19095214 | insulin, isophane |
| PRAMLINTIDE | PRAMLINTIDE | 40165686 | Pramlintide |
| EXENATIDE | EXENATIDE | 36234709 | exenatide |

| | | | |
|---|---|---|---|
| EMPAGLIFLOZIN | EMPAGLIFLOZIN | 1592795 | empagliflozin |
| METFORMIN | METFORMIN | 40153895 | Metformin |
| INSULIN | INSULIN | 19010204 | insulin, isophane |
| GLIMEPIRIDE | GLIMEPIRIDE | 1597781 | glimepiride |
| INSULIN | INSULIN | 36229086 | Insulin Glargine |
| GLIPIZIDE | GLIPIZIDE | 1560171 | Glipizide |
| LINAGLIPTIN | LINAGLIPTIN | 40239219 | Linagliptin |
| BROMOCRIPTINE | BROMOCRIPTINE | 19083958 | Bromocriptine |
| INSULIN | INSULIN | 19120315 | insulin, isophane |
| EXENATIDE | EXENATIDE | 1583727 | exenatide |
| NATEGLINIDE | NATEGLINIDE | 19071495 | nateglinide |
| INSULIN | INSULIN | 46233978 | Insulin Lispro |
| BROMOCRIPTINE | BROMOCRIPTINE | 730572 | Bromocriptine |
| INSULIN | INSULIN | 36233552 | insulin, isophane |
| INSULIN | INSULIN | 40184293 | Insulin, Aspart Protamine, Human |
| CHLORPROPAMIDE | CHLORPROPAMIDE | 19088819 | Chlorpropamide |
| METFORMIN | METFORMIN | 40063356 | Metformin |
| INSULIN | INSULIN | 1596955 | insulin, isophane |
| INSULIN | INSULIN | 1596977 | Regular Insulin, Human |
| INSULIN | INSULIN | 19078553 | insulin, isophane |
| REPAGLINIDE | REPAGLINIDE | 19023425 | repaglinide |
| NATEGLINIDE | NATEGLINIDE | 19085097 | nateglinide |
| INSULIN | INSULIN | 1550027 | Insulin Lispro |
| REPAGLINIDE | REPAGLINIDE | 36240430 | repaglinide |
| INSULIN | INSULIN | 19120315 | insulin, isophane |
| GLIMEPIRIDE | GLIMEPIRIDE | 1597756 | glimepiride |
| INSULIN | INSULIN | 1597000 | Regular Insulin, Human |
| INSULIN | INSULIN | 1513881 | Insulin Lispro |
| METFORMIN | METFORMIN | 40164924 | Metformin |
| METFORMIN | METFORMIN | 1503328 | Metformin |
| GLYBURIDE | GLYBURIDE | 1559684 | Glyburide |
| ACARBOSE | ACARBOSE | 19040970 | Acarbose |
| EXENATIDE | EXENATIDE | 19083253 | exenatide |

| | | | |
|---|---|---|---|
| INSULIN | INSULIN | 1596977 | Regular Insulin, Human |
| REPAGLINIDE | REPAGLINIDE | 19023425 | repaglinide |
| INSULIN | INSULIN | 19012679 | Regular Insulin, Human |
| INSULIN | INSULIN | 19112791 | Insulin, Glulisine, Human |
| METFORMIN | METFORMIN | 40164922 | Metformin |
| INSULIN | INSULIN | 19120315 | insulin, isophane |
| INSULIN | INSULIN | 1586344 | insulin, isophane |
| NATEGLINIDE | NATEGLINIDE | 1502830 | nateglinide |
| INSULIN | INSULIN | 19078557 | Regular Insulin, Human |
| INSULIN | INSULIN | 19111795 | insulin, isophane |
| EXENATIDE | EXENATIDE | 42708079 | exenatide |
| METFORMIN | METFORMIN | 1503297 | Metformin |
| GLIPIZIDE | GLIPIZIDE | 40166816 | Glipizide |
| GLYBURIDE | GLYBURIDE | 19030572 | Glyburide |
| METFORMIN | METFORMIN | 40164892 | Metformin |
| INSULIN | INSULIN | 1502905 | Insulin Glargine |
| INSULIN | INSULIN | 1586327 | insulin, isophane |
| INSULIN | INSULIN | 40051347 | Insulin Glargine |
| INSULIN | INSULIN | 19062999 | insulin, isophane |
| GLIMEPIRIDE | GLIMEPIRIDE | 1597756 | glimepiride |
| INSULIN | INSULIN | 46221552 | Insulin Glargine |
| INSULIN | INSULIN | 1596955 | insulin, isophane |
| INSULIN | INSULIN | 19078559 | Insulin Glargine |
| METFORMIN | METFORMIN | 36242511 | Metformin |
| METFORMIN | METFORMIN | 1503328 | Metformin |
| INSULIN | INSULIN | 1550023 | Insulin Lispro |
| INSULIN | INSULIN | 1544838 | Insulin, Glulisine, Human |
| DAPAGLIFLOZIN | DAPAGLIFLOZIN | 45775864 | dapagliflozin |
| GLYBURIDE | GLYBURIDE | 1559684 | Glyburide |
| INSULIN | INSULIN | 36231450 | Insulin, Aspart, Human |
| GLIPIZIDE | GLIPIZIDE | 40165973 | Glipizide |
| EXENATIDE | EXENATIDE | 1583730 | exenatide |
| REPAGLINIDE | REPAGLINIDE | 19023426 | repaglinide |
| INSULIN | INSULIN | 1502910 | Insulin Glargine |

| | | | |
|---|---|---|---|
| INSULIN | INSULIN | 19078554 | Regular Insulin, Human |
| INSULIN | INSULIN | 19078559 | Insulin Glargine |
| CHLORPROPAMIDE | CHLORPROPAMIDE | 19025944 | Chlorpropamide |
| INSULIN | INSULIN | 19116559 | insulin, isophane |
| ROSIGLITAZONE | ROSIGLITAZONE | 36229366 | rosiglitazone |
| INSULIN | INSULIN | 1596957 | insulin, isophane |
| INSULIN | INSULIN | 19078603 | insulin, isophane |
| INSULIN | INSULIN | 36229086 | Insulin Glargine |
| INSULIN | INSULIN | 35605622 | Regular Insulin, Human |
| GLYBURIDE | GLYBURIDE | 1559685 | Glyburide |
| LIRAGLUTIDE | LIRAGLUTIDE | 36241903 | liraglutide |
| METFORMIN | METFORMIN | 42708175 | Metformin |
| INSULIN | INSULIN | 1550028 | Insulin Lispro |
| SAXAGLIPTIN | SAXAGLIPTIN | 36229291 | saxagliptin |
| GLYBURIDE | GLYBURIDE | 19030573 | Glyburide |
| EXENATIDE | EXENATIDE | 1583727 | exenatide |
| EXENATIDE | EXENATIDE | 19083253 | exenatide |
| INSULIN | INSULIN | 1550027 | Insulin Lispro |
| REPAGLINIDE | REPAGLINIDE | 19023426 | repaglinide |
| INSULIN | INSULIN | 46234049 | Insulin, Aspart, Human |
| EXENATIDE | EXENATIDE | 1583727 | exenatide |
| INSULIN | INSULIN | 1567201 | Insulin, Aspart Protamine, Human |
| CANAGLIFLOZIN | CANAGLIFLOZIN | 45774723 | canagliflozin |
| GLYBURIDE | GLYBURIDE | 1559684 | Glyburide |
| INSULIN | INSULIN | 1596957 | insulin, isophane |
| INSULIN | INSULIN | 46221552 | Insulin Glargine |
| CANAGLIFLOZIN | CANAGLIFLOZIN | 43526472 | canagliflozin |
| METFORMIN | METFORMIN | 40063354 | Metformin |
| GLIMEPIRIDE | GLIMEPIRIDE | 19023585 | glimepiride |
| INSULIN | INSULIN | 46234043 | Insulin, Aspart, Human |
| INSULIN | INSULIN | 36222190 | insulin detemir |

| | | | |
|---|---|---|---|
| DAPAGLIFLOZIN | DAPAGLIFLOZIN | 44785477 | dapagliflozin |
| METFORMIN | METFORMIN | 40164914 | Metformin |
| INSULIN | INSULIN | 1718604 | Insulin Glargine |
| INSULIN | INSULIN | 19018822 | Insulin Lispro |
| LINAGLIPTIN | LINAGLIPTIN | 36238286 | Linagliptin |
| SITAGLIPTIN | SITAGLIPTIN | 40243628 | sitagliptin |
| ROSIGLITAZONE | ROSIGLITAZONE | 40099633 | rosiglitazone |
| PRAMLINTIDE | PRAMLINTIDE | 40165687 | Pramlintide |
| INSULIN | INSULIN | 19078610 | insulin, regular, beef-pork |
| INSULIN | INSULIN | 19135276 | Insulin Glargine |
| INSULIN | INSULIN | 19062999 | insulin, isophane |
| ALOGLIPTIN | ALOGLIPTIN | 36246271 | alogliptin |
| ROSIGLITAZONE | ROSIGLITAZONE | 1547508 | rosiglitazone |
| INSULIN | INSULIN | 1550028 | Insulin Lispro |
| INSULIN | INSULIN | 1502905 | Insulin Glargine |
| INSULIN | INSULIN | 1513912 | Insulin Lispro |
| INSULIN | INSULIN | 19078530 | insulin, isophane |
| INSULIN | INSULIN | 1596977 | Regular Insulin, Human |
| INSULIN | INSULIN | 1550027 | Insulin Lispro |
| LIXISENATIDE | LIXISENATIDE | 44506754 | Lixisenatide |
| INSULIN | INSULIN | 19094122 | Regular Insulin, Human |
| METFORMIN | METFORMIN | 40163929 | Metformin |
| METFORMIN | METFORMIN | 40164897 | Metformin |
| ACARBOSE | ACARBOSE | 36242198 | Acarbose |
| INSULIN | INSULIN | 19078551 | insulin, isophane |
| ALBIGLUTIDE | ALBIGLUTIDE | 44816086 | albiglutide |
| GLIMEPIRIDE | GLIMEPIRIDE | 40059665 | glimepiride |
| SAXAGLIPTIN | SAXAGLIPTIN | 40166042 | saxagliptin |
| INSULIN | INSULIN | 1550023 | Insulin Lispro |
| INSULIN | INSULIN | 46233975 | Insulin Lispro |
| INSULIN | INSULIN | 19135265 | Regular Insulin, Human |

| | | | |
|---|---|---|---|
| SITAGLIPTIN | SITAGLIPTIN | 19125042 | sitagliptin |
| GLYBURIDE | GLYBURIDE | 19063991 | Glyburide |
| BROMOCRIPTINE | BROMOCRIPTINE | 19007101 | Bromocriptine |
| EXENATIDE | EXENATIDE | 19135404 | exenatide |
| ALOGLIPTIN | ALOGLIPTIN | 43013914 | alogliptin |
| GLIPIZIDE | GLIPIZIDE | 19100559 | Glipizide |
| ACARBOSE | ACARBOSE | 19021312 | Acarbose |
| DAPAGLIFLOZIN | DAPAGLIFLOZIN | 36247907 | dapagliflozin |
| GLIPIZIDE | GLIPIZIDE | 36235749 | Glipizide |
| INSULIN | INSULIN | 19111795 | insulin, isophane |
| SITAGLIPTIN | SITAGLIPTIN | 40136770 | sitagliptin |
| EXENATIDE | EXENATIDE | 36234709 | exenatide |
| GLIPIZIDE | GLIPIZIDE | 19081295 | Glipizide |
| SITAGLIPTIN | SITAGLIPTIN | 42901620 | sitagliptin |
| EXENATIDE | EXENATIDE | 46234118 | exenatide |
| DULAGLUTIDE | DULAGLUTIDE | 45774435 | dulaglutide |
| CANAGLIFLOZIN | CANAGLIFLOZIN | 36246431 | canagliflozin |
| ACARBOSE | ACARBOSE | 1529331 | Acarbose |
| INSULIN | INSULIN | 19078530 | insulin, isophane |
| INSULIN | INSULIN | 19016117 | Insulin Lispro |
| INSULIN | INSULIN | 1513881 | Insulin Lispro |
| INSULIN | INSULIN | 19078559 | Insulin Glargine |
| INSULIN | INSULIN | 1502910 | Insulin Glargine |
| METFORMIN | METFORMIN | 40164926 | Metformin |
| DULAGLUTIDE | DULAGLUTIDE | 45774440 | dulaglutide |
| METFORMIN | METFORMIN | 36223145 | Metformin |
| METFORMIN | METFORMIN | 40164891 | Metformin |
| GLIPIZIDE | GLIPIZIDE | 40044253 | Glipizide |
| INSULIN | INSULIN | 19095218 | Regular Insulin, Human |
| GLYBURIDE | GLYBURIDE | 1559684 | Glyburide |
| GLYBURIDE | GLYBURIDE | 1559684 | Glyburide |
| GLIMEPIRIDE | GLIMEPIRIDE | 43011830 | glimepiride |
| REPAGLINIDE | REPAGLINIDE | 19023424 | repaglinide |
| METFORMIN | METFORMIN | 36235582 | Metformin |
| EXENATIDE | EXENATIDE | 1583730 | exenatide |
| INSULIN | INSULIN | 36229086 | Insulin Glargine |

| | | | |
|---|---|---|---|
| INSULIN | INSULIN | 1516979 | insulin detemir |
| INSULIN | INSULIN | 19131582 | Insulin, Glulisine, Human |
| INSULIN | INSULIN | 46234045 | Insulin, Aspart, Human |
| METFORMIN | METFORMIN | 1503297 | Metformin |
| LINAGLIPTIN | LINAGLIPTIN | 42708087 | Linagliptin |
| INSULIN | INSULIN | 40051347 | Insulin Glargine |
| METFORMIN | METFORMIN | 1503297 | Metformin |
| INSULIN | INSULIN | 36224228 | insulin detemir |
| INSULIN | INSULIN | 19078527 | Insulin, Zinc, Human |
| ALBIGLUTIDE | ALBIGLUTIDE | 44816274 | albiglutide |
| INSULIN | INSULIN | 1596957 | insulin, isophane |
| INSULIN | INSULIN | 19010204 | insulin, isophane |
| INSULIN | INSULIN | 19095211 | Insulin Lispro |
| INSULIN | INSULIN | 40051347 | Insulin Glargine |
| INSULIN | INSULIN | 1596941 | insulin, isophane |
| INSULIN | INSULIN | 1596941 | insulin, isophane |
| INSULIN | INSULIN | 1567232 | Insulin, Aspart, Human |
| INSULIN | INSULIN | 40166274 | Insulin Lispro |
| INSULIN | INSULIN | 1544838 | Insulin, Glulisine, Human |
| INSULIN | INSULIN | 1502905 | Insulin Glargine |
| GLYBURIDE | GLYBURIDE | 40165993 | Glyburide |
| INSULIN | INSULIN | 19078551 | insulin, isophane |
| CANAGLIFLOZIN | CANAGLIFLOZIN | 45774714 | canagliflozin |
| INSULIN | INSULIN | 19078610 | insulin, regular, beef-pork |
| INSULIN | INSULIN | 40176981 | Insulin, Aspart Protamine, Human |
| GLIPIZIDE | GLIPIZIDE | 40165972 | Glipizide |
| INSULIN | INSULIN | 1718711 | insulin degludec |
| GLYBURIDE | GLYBURIDE | 19106976 | Glyburide |
| EMPAGLIFLOZIN | EMPAGLIFLOZIN | 45892176 | empagliflozin |
| METFORMIN | METFORMIN | 1503328 | Metformin |

| | | | |
|---|---|---|---|
| GLIMEPIRIDE | GLIMEPIRIDE | 43011830 | glimepiride |
| INSULIN | INSULIN | 19078558 | Insulin, Aspart, Human |
| PIOGLITAZONE | PIOGLITAZONE | 1525218 | pioglitazone |
| INSULIN | INSULIN | 19116883 | insulin, isophane |
| LIRAGLUTIDE | LIRAGLUTIDE | 40170912 | liraglutide |
| PIOGLITAZONE | PIOGLITAZONE | 40099898 | pioglitazone |
| INSULIN | INSULIN | 19111797 | insulin, isophane |
| INSULIN | INSULIN | 1597001 | Regular Insulin, Human |
| LINAGLIPTIN | LINAGLIPTIN | 36238286 | Linagliptin |
| INSULIN | INSULIN | 1596972 | insulin, isophane |
| LIRAGLUTIDE | LIRAGLUTIDE | 40169223 | liraglutide |
| METFORMIN | METFORMIN | 40231404 | Metformin |
| INSULIN | INSULIN | 19095199 | Lente Insulin, Beef-Pork |
| EXENATIDE | EXENATIDE | 1583730 | exenatide |
| METFORMIN | METFORMIN | 40153151 | Metformin |
| GLYBURIDE | GLYBURIDE | 36235750 | Glyburide |
| INSULIN | INSULIN | 40184293 | Insulin, Aspart Protamine, Human |
| METFORMIN | METFORMIN | 40063354 | Metformin |
| CANAGLIFLOZIN | CANAGLIFLOZIN | 45774719 | canagliflozin |
| INSULIN | INSULIN | 19071700 | Insulin Glargine |
| BROMOCRIPTINE | BROMOCRIPTINE | 36238866 | Bromocriptine |
| GLIMEPIRIDE | GLIMEPIRIDE | 1597801 | glimepiride |
| INSULIN | INSULIN | 19120315 | insulin, isophane |
| INSULIN | INSULIN | 1596960 | insulin, isophane |
| INSULIN | INSULIN | 40184293 | Insulin, Aspart Protamine, Human |
| INSULIN | INSULIN | 1516976 | insulin detemir |
| METFORMIN | METFORMIN | 40164944 | Metformin |
| GLYBURIDE | GLYBURIDE | 40166003 | Glyburide |
| GLIPIZIDE | GLIPIZIDE | 19100559 | Glipizide |
| EXENATIDE | EXENATIDE | 1583728 | exenatide |

| | | | |
|---|---|---|---|
| GLIMEPIRIDE | GLIMEPIRIDE | 1597796 | glimepiride |
| BROMOCRIPTINE | BROMOCRIPTINE | 40016896 | Bromocriptine |
| EXENATIDE | EXENATIDE | 42708079 | exenatide |
| INSULIN | INSULIN | 46234045 | Insulin, Aspart, Human |
| METFORMIN | METFORMIN | 1503297 | Metformin |
| METFORMIN | METFORMIN | 40164897 | Metformin |
| METFORMIN | METFORMIN | 1503297 | Metformin |
| INSULIN | INSULIN | 1596955 | insulin, isophane |
| GLIPIZIDE | GLIPIZIDE | 1560233 | Glipizide |
| INSULIN | INSULIN | 1513912 | Insulin Lispro |
| INSULIN | INSULIN | 1550027 | Insulin Lispro |
| PIOGLITAZONE | PIOGLITAZONE | 1525217 | pioglitazone |
| INSULIN | INSULIN | 19110219 | insulin, isophane |
| METFORMIN | METFORMIN | 40164920 | Metformin |
| DULAGLUTIDE | DULAGLUTIDE | 45774436 | dulaglutide |
| SITAGLIPTIN | SITAGLIPTIN | 19125043 | sitagliptin |
| INSULIN | INSULIN | 1516976 | insulin detemir |
| GLIPIZIDE | GLIPIZIDE | 40139022 | Glipizide |
| INSULIN | INSULIN | 19078553 | insulin, isophane |
| METFORMIN | METFORMIN | 40164908 | Metformin |
| INSULIN | INSULIN | 46233975 | Insulin Lispro |
| PIOGLITAZONE | PIOGLITAZONE | 19079293 | pioglitazone |
| INSULIN | INSULIN | 46221557 | Insulin Glargine |
| INSULIN | INSULIN | 1596941 | insulin, isophane |
| EXENATIDE | EXENATIDE | 1583727 | exenatide |
| INSULIN | INSULIN | 46233950 | Insulin Lispro |
| INSULIN | INSULIN | 40051347 | Insulin Glargine |
| GLIMEPIRIDE | GLIMEPIRIDE | 19117200 | glimepiride |
| INSULIN | INSULIN | 1567198 | Insulin, Aspart, Human |
| INSULIN | INSULIN | 40184293 | Insulin, Aspart Protamine, Human |
| GLIPIZIDE | GLIPIZIDE | 40165970 | Glipizide |
| INSULIN | INSULIN | 36248196 | insulin degludec |
| INSULIN | INSULIN | 40051347 | Insulin Glargine |
| SITAGLIPTIN | SITAGLIPTIN | 19125047 | sitagliptin |

| | | | |
|---|---|---|---|
| GLYBURIDE | GLYBURIDE | 1559684 | Glyburide |
| INSULIN | INSULIN | 19078558 | Insulin, Aspart, Human |
| METFORMIN | METFORMIN | 36236322 | Metformin |
| ACARBOSE | ACARBOSE | 19040969 | Acarbose |
| INSULIN | INSULIN | 19078552 | insulin, isophane |
| METFORMIN | METFORMIN | 1503328 | Metformin |
| CANAGLIFLOZIN | CANAGLIFLOZIN | 45774721 | canagliflozin |
| SITAGLIPTIN | SITAGLIPTIN | 36240263 | sitagliptin |
| SITAGLIPTIN | SITAGLIPTIN | 40243629 | sitagliptin |
| METFORMIN | METFORMIN | 1503297 | Metformin |
| SITAGLIPTIN | SITAGLIPTIN | 40136770 | sitagliptin |
| INSULIN | INSULIN | 1567198 | Insulin, Aspart, Human |
| LINAGLIPTIN | LINAGLIPTIN | 40239219 | Linagliptin |
| GLYBURIDE | GLYBURIDE | 19065480 | Glyburide |
| INSULIN | INSULIN | 40184293 | Insulin, Aspart Protamine, Human |
| INSULIN | INSULIN | 36233154 | Insulin, Glulisine, Human |
| METFORMIN | METFORMIN | 36236481 | Metformin |
| METFORMIN | METFORMIN | 1503328 | Metformin |
| INSULIN | INSULIN | 1586370 | Ultralente Insulin, Human |
| GLYBURIDE | GLYBURIDE | 40165998 | Glyburide |
| METFORMIN | METFORMIN | 19088818 | Metformin |
| GLIPIZIDE | GLIPIZIDE | 36218767 | Glipizide |
| INSULIN | INSULIN | 1596955 | insulin, isophane |
| SAXAGLIPTIN | SAXAGLIPTIN | 40166045 | saxagliptin |
| ALBIGLUTIDE | ALBIGLUTIDE | 44816283 | albiglutide |
| GLYBURIDE | GLYBURIDE | 19077682 | Glyburide |
| INSULIN | INSULIN | 46233975 | Insulin Lispro |
| INSULIN | INSULIN | 19016117 | Insulin Lispro |
| REPAGLINIDE | REPAGLINIDE | 19023426 | repaglinide |
| EXENATIDE | EXENATIDE | 19135404 | exenatide |
| LIRAGLUTIDE | LIRAGLUTIDE | 40170913 | liraglutide |
| INSULIN | INSULIN | 19062999 | insulin, isophane |
| INSULIN | INSULIN | 19100131 | insulin, isophane |
| INSULIN | INSULIN | 1516979 | insulin detemir |
| EMPAGLIFLOZIN | EMPAGLIFLOZIN | 45892179 | empagliflozin |
| INSULIN | INSULIN | 1550028 | Insulin Lispro |

| | | | |
|---|---|---|---|
| SITAGLIPTIN | SITAGLIPTIN | 19125047 | sitagliptin |
| INSULIN | INSULIN | 40176981 | Insulin, Aspart Protamine, Human |
| CHLORPROPAMIDE | CHLORPROPAMIDE | 19006907 | Chlorpropamide |
| GLIPIZIDE | GLIPIZIDE | 40167634 | Glipizide |
| ALBIGLUTIDE | ALBIGLUTIDE | 44816283 | albiglutide |
| PIOGLITAZONE | PIOGLITAZONE | 1525216 | pioglitazone |
| INSULIN | INSULIN | 46234049 | Insulin, Aspart, Human |
| INSULIN | INSULIN | 19123376 | Insulin Lispro |
| INSULIN | INSULIN | 1550028 | Insulin Lispro |
| INSULIN | INSULIN | 35605621 | Regular Insulin, Human |
| LINAGLIPTIN | LINAGLIPTIN | 36229566 | Linagliptin |
| INSULIN | INSULIN | 1513912 | Insulin Lispro |
| METFORMIN | METFORMIN | 42705990 | Metformin |
| NATEGLINIDE | NATEGLINIDE | 36223777 | nateglinide |
| INSULIN | INSULIN | 1596941 | insulin, isophane |
| INSULIN | INSULIN | 1513850 | Insulin, Zinc, Human |
| EMPAGLIFLOZIN | EMPAGLIFLOZIN | 45892176 | empagliflozin |
| INSULIN | INSULIN | 1597001 | Regular Insulin, Human |
| CANAGLIFLOZIN | CANAGLIFLOZIN | 45774724 | canagliflozin |
| INSULIN | INSULIN | 36231457 | Insulin, Zinc, Human |
| EMPAGLIFLOZIN | EMPAGLIFLOZIN | 45774893 | empagliflozin |
| GLYBURIDE | GLYBURIDE | 19106976 | Glyburide |
| GLYBURIDE | GLYBURIDE | 1559685 | Glyburide |
| DULAGLUTIDE | DULAGLUTIDE | 45774442 | dulaglutide |
| INSULIN | INSULIN | 19110352 | insulin, isophane |
| INSULIN | INSULIN | 19058398 | Insulin Lispro |
| GLIPIZIDE | GLIPIZIDE | 19077637 | Glipizide |
| INSULIN | INSULIN | 1516981 | insulin detemir |
| GLYBURIDE | GLYBURIDE | 36230036 | Glyburide |
| REPAGLINIDE | REPAGLINIDE | 36240429 | repaglinide |
| DULAGLUTIDE | DULAGLUTIDE | 45774438 | dulaglutide |
| INSULIN | INSULIN | 36236032 | Insulin Lispro |
| INSULIN | INSULIN | 1516976 | insulin detemir |
| EXENATIDE | EXENATIDE | 46234117 | exenatide |
| GLYBURIDE | GLYBURIDE | 40168374 | Glyburide |
| NATEGLINIDE | NATEGLINIDE | 36232423 | nateglinide |

| | | | |
|---|---|---|---|
| GLIPIZIDE | GLIPIZIDE | 19077636 | Glipizide |
| METFORMIN | METFORMIN | 40163928 | Metformin |
| BROMOCRIPTINE | BROMOCRIPTINE | 19006288 | Bromocriptine |
| INSULIN | INSULIN | 40051347 | Insulin Glargine |
| GLIPIZIDE | GLIPIZIDE | 1560171 | Glipizide |
| INSULIN | INSULIN | 19116453 | insulin, isophane |
| INSULIN | INSULIN | 36229086 | Insulin Glargine |
| METFORMIN | METFORMIN | 1503297 | Metformin |
| REPAGLINIDE | REPAGLINIDE | 1516771 | repaglinide |
| INSULIN | INSULIN | 19120790 | Insulin Glargine |
| SITAGLIPTIN | SITAGLIPTIN | 40243629 | sitagliptin |
| INSULIN | INSULIN | 35602728 | insulin degludec |
| INSULIN | INSULIN | 1544873 | Insulin, Glulisine, Human |
| CHLORPROPAMIDE | CHLORPROPAMIDE | 19088817 | Chlorpropamide |
| INSULIN | INSULIN | 1516976 | insulin detemir |
| INSULIN | INSULIN | 46234045 | Insulin, Aspart, Human |
| INSULIN | INSULIN | 1516976 | insulin detemir |
| GLYBURIDE | GLYBURIDE | 1559684 | Glyburide |
| METFORMIN | METFORMIN | 1503328 | Metformin |
| GLIPIZIDE | GLIPIZIDE | 19100559 | Glipizide |
| METFORMIN | METFORMIN | 40164889 | Metformin |
| INSULIN | INSULIN | 1567201 | Insulin, Aspart Protamine, Human |
| EXENATIDE | EXENATIDE | 45774533 | exenatide |
| GLIMEPIRIDE | GLIMEPIRIDE | 36214985 | glimepiride |
| GLIMEPIRIDE | GLIMEPIRIDE | 43011831 | glimepiride |
| GLIMEPIRIDE | GLIMEPIRIDE | 1597799 | glimepiride |
| INSULIN | INSULIN | 1596999 | Regular Insulin, Human |
| INSULIN | INSULIN | 1596998 | Regular Insulin, Human |
| METFORMIN | METFORMIN | 40164900 | Metformin |
| ROSIGLITAZONE | ROSIGLITAZONE | 36227624 | rosiglitazone |
| METFORMIN | METFORMIN | 40164929 | Metformin |
| GLIPIZIDE | GLIPIZIDE | 40044224 | Glipizide |
| INSULIN | INSULIN | 1596955 | insulin, isophane |
| METFORMIN | METFORMIN | 40176046 | Metformin |
| INSULIN | INSULIN | 19078555 | Regular Insulin, Human |

| | | | |
|---|---|---|---|
| INSULIN | INSULIN | 1596952 | insulin, isophane |
| INSULIN | INSULIN | 19131582 | Insulin, Glulisine, Human |
| INSULIN | INSULIN | 40184293 | Insulin, Aspart Protamine, Human |
| INSULIN | INSULIN | 19100128 | Regular Insulin, Human |
| INSULIN | INSULIN | 19078557 | Regular Insulin, Human |
| DULAGLUTIDE | DULAGLUTIDE | 45774448 | dulaglutide |
| METFORMIN | METFORMIN | 42708173 | Metformin |
| GLIPIZIDE | GLIPIZIDE | 19030445 | Glipizide |
| METFORMIN | METFORMIN | 1503327 | Metformin |
| INSULIN | INSULIN | 40052088 | Insulin Lispro |
| INSULIN | INSULIN | 19111795 | insulin, isophane |
| INSULIN | INSULIN | 19078554 | Regular Insulin, Human |
| INSULIN | INSULIN | 19078555 | Regular Insulin, Human |
| INSULIN | INSULIN | 36247261 | Regular Insulin, Human |
| NATEGLINIDE | NATEGLINIDE | 1502826 | nateglinide |
| ACARBOSE | ACARBOSE | 19021312 | Acarbose |
| INSULIN | INSULIN | 36236033 | Insulin Lispro |
| METFORMIN | METFORMIN | 1503297 | Metformin |
| GLIPIZIDE | GLIPIZIDE | 19079990 | Glipizide |
| INSULIN | INSULIN | 19110219 | insulin, isophane |
| LINAGLIPTIN | LINAGLIPTIN | 42708087 | Linagliptin |
| BROMOCRIPTINE | BROMOCRIPTINE | 40016896 | Bromocriptine |
| INSULIN | INSULIN | 19095211 | Insulin Lispro |
| EMPAGLIFLOZIN | EMPAGLIFLOZIN | 45774895 | empagliflozin |
| REPAGLINIDE | REPAGLINIDE | 19023426 | repaglinide |
| ACARBOSE | ACARBOSE | 19023063 | Acarbose |
| INSULIN | INSULIN | 19116449 | insulin, isophane |
| INSULIN | INSULIN | 19116559 | insulin, isophane |
| DULAGLUTIDE | DULAGLUTIDE | 45774442 | dulaglutide |
| CANAGLIFLOZIN | CANAGLIFLOZIN | 43526468 | canagliflozin |
| METFORMIN | METFORMIN | 36235787 | Metformin |
| INSULIN | INSULIN | 1597001 | Regular Insulin, Human |

| | | | |
|---|---|---|---|
| METFORMIN | METFORMIN | 40164881 | Metformin |
| INSULIN | INSULIN | 1597000 | Regular Insulin, Human |
| INSULIN | INSULIN | 40051347 | Insulin Glargine |
| METFORMIN | METFORMIN | 40164901 | Metformin |
| INSULIN | INSULIN | 19062999 | insulin, isophane |
| INSULIN | INSULIN | 19078551 | insulin, isophane |
| INSULIN | INSULIN | 19131582 | Insulin, Glulisine, Human |
| METFORMIN | METFORMIN | 1503328 | Metformin |
| DAPAGLIFLOZIN | DAPAGLIFLOZIN | 44785473 | dapagliflozin |
| METFORMIN | METFORMIN | 36242511 | Metformin |
| ALOGLIPTIN | ALOGLIPTIN | 36246195 | alogliptin |
| INSULIN | INSULIN | 40166275 | Insulin Lispro |
| GLYBURIDE | GLYBURIDE | 1559684 | Glyburide |
| BROMOCRIPTINE | BROMOCRIPTINE | 730548 | Bromocriptine |
| INSULIN | INSULIN | 19110219 | insulin, isophane |
| PIOGLITAZONE | PIOGLITAZONE | 1525216 | pioglitazone |
| EXENATIDE | EXENATIDE | 1583722 | exenatide |
| PIOGLITAZONE | PIOGLITAZONE | 1525218 | pioglitazone |
| INSULIN | INSULIN | 19078530 | insulin, isophane |
| INSULIN | INSULIN | 19078553 | insulin, isophane |
| GLIPIZIDE | GLIPIZIDE | 40166814 | Glipizide |
| REPAGLINIDE | REPAGLINIDE | 19023424 | repaglinide |
| INSULIN | INSULIN | 1596918 | insulin, isophane |
| METFORMIN | METFORMIN | 1503328 | Metformin |
| INSULIN | INSULIN | 36235839 | Insulin, Zinc, Human |
| INSULIN | INSULIN | 46234237 | insulin, isophane |
| INSULIN | INSULIN | 19110355 | insulin, isophane |
| INSULIN | INSULIN | 36227902 | Insulin Lispro |
| INSULIN | INSULIN | 19120790 | Insulin Glargine |
| GLYBURIDE | GLYBURIDE | 19024508 | Glyburide |
| ROSIGLITAZONE | ROSIGLITAZONE | 36227624 | rosiglitazone |
| METFORMIN | METFORMIN | 36235746 | Metformin |
| TOLBUTAMIDE | TOLBUTAMIDE | 1502912 | Tolbutamide |
| INSULIN | INSULIN | 46233945 | Insulin Lispro |

| | | | |
|---|---|---|---|
| EXENATIDE | EXENATIDE | 36234709 | exenatide |
| INSULIN | INSULIN | 1596960 | insulin, isophane |
| ACARBOSE | ACARBOSE | 19047612 | Acarbose |
| INSULIN | INSULIN | 1567198 | Insulin, Aspart, Human |
| METFORMIN | METFORMIN | 1503297 | Metformin |
| METFORMIN | METFORMIN | 40164929 | Metformin |
| INSULIN | INSULIN | 19110356 | Regular Insulin, Human |
| BROMOCRIPTINE | BROMOCRIPTINE | 19006288 | Bromocriptine |
| EXENATIDE | EXENATIDE | 46234116 | exenatide |
| PIOGLITAZONE | PIOGLITAZONE | 40099898 | pioglitazone |
| INSULIN | INSULIN | 19078555 | Regular Insulin, Human |
| INSULIN | INSULIN | 1596977 | Regular Insulin, Human |
| NATEGLINIDE | NATEGLINIDE | 19107111 | nateglinide |
| GLIPIZIDE | GLIPIZIDE | 1560233 | Glipizide |
| INSULIN | INSULIN | 40051348 | Insulin Glargine |
| INSULIN | INSULIN | 1596959 | insulin, isophane |
| INSULIN | INSULIN | 19078555 | Regular Insulin, Human |
| INSULIN | INSULIN | 1550023 | Insulin Lispro |
| NATEGLINIDE | NATEGLINIDE | 36223778 | nateglinide |
| INSULIN | INSULIN | 46233976 | Insulin Lispro |
| GLYBURIDE | GLYBURIDE | 1559685 | Glyburide |
| INSULIN | INSULIN | 19062999 | insulin, isophane |
| INSULIN | INSULIN | 46233945 | Insulin Lispro |
| INSULIN | INSULIN | 19012679 | Regular Insulin, Human |
| METFORMIN | METFORMIN | 36235581 | Metformin |
| INSULIN | INSULIN | 1586344 | insulin, isophane |
| INSULIN | INSULIN | 19078551 | insulin, isophane |
| METFORMIN | METFORMIN | 40164928 | Metformin |
| INSULIN | INSULIN | 1596977 | Regular Insulin, Human |
| MIGLITOL | MIGLITOL | 1510206 | miglitol |
| METFORMIN | METFORMIN | 40164933 | Metformin |
| GLYBURIDE | GLYBURIDE | 1559684 | Glyburide |
| INSULIN | INSULIN | 40176981 | Insulin, Aspart Protamine, Human |
| INSULIN | INSULIN | 46234240 | insulin, isophane |

| | | | |
|---|---|---|---|
| INSULIN | INSULIN | 40184294 | Insulin, Aspart Protamine, Human |
| GLIMEPIRIDE | GLIMEPIRIDE | 43011831 | glimepiride |
| MIGLITOL | MIGLITOL | 36233498 | miglitol |
| METFORMIN | METFORMIN | 40231395 | Metformin |
| INSULIN | INSULIN | 1567201 | Insulin, Aspart Protamine, Human |
| INSULIN | INSULIN | 19078603 | insulin, isophane |
| INSULIN | INSULIN | 1567198 | Insulin, Aspart, Human |
| INSULIN | INSULIN | 19110356 | Regular Insulin, Human |
| METFORMIN | METFORMIN | 1503328 | Metformin |
| INSULIN | INSULIN | 1544873 | Insulin, Glulisine, Human |
| EXENATIDE | EXENATIDE | 1583728 | exenatide |
| INSULIN | INSULIN | 19078557 | Regular Insulin, Human |
| GLIPIZIDE | GLIPIZIDE | 1560171 | Glipizide |
| TOLBUTAMIDE | TOLBUTAMIDE | 1502855 | Tolbutamide |
| INSULIN | INSULIN | 1596941 | insulin, isophane |
| DULAGLUTIDE | DULAGLUTIDE | 36247186 | dulaglutide |
| INSULIN | INSULIN | 19010204 | insulin, isophane |
| INSULIN | INSULIN | 1596959 | insulin, isophane |
| TOLBUTAMIDE | TOLBUTAMIDE | 19047751 | Tolbutamide |
| METFORMIN | METFORMIN | 1503297 | Metformin |
| EXENATIDE | EXENATIDE | 1583730 | exenatide |
| ALBIGLUTIDE | ALBIGLUTIDE | 44816282 | albiglutide |
| INSULIN | INSULIN | 36236033 | Insulin Lispro |
| INSULIN | INSULIN | 1567198 | Insulin, Aspart, Human |
| METFORMIN | METFORMIN | 1503297 | Metformin |
| INSULIN | INSULIN | 19131582 | Insulin, Glulisine, Human |
| INSULIN | INSULIN | 40051347 | Insulin Glargine |
| INSULIN | INSULIN | 19078554 | Regular Insulin, Human |
| METFORMIN | METFORMIN | 36236323 | Metformin |
| INSULIN | INSULIN | 19110357 | Regular Insulin, Human |
| LINAGLIPTIN | LINAGLIPTIN | 40239218 | Linagliptin |
| INSULIN | INSULIN | 19078555 | Regular Insulin, Human |

| | | | |
|---|---|---|---|
| INSULIN | INSULIN | 19058398 | Insulin Lispro |
| MIGLITOL | MIGLITOL | 1510207 | miglitol |
| LIRAGLUTIDE | LIRAGLUTIDE | 40170911 | liraglutide |
| LIRAGLUTIDE | LIRAGLUTIDE | 40169222 | liraglutide |
| METFORMIN | METFORMIN | 1503297 | Metformin |
| INSULIN | INSULIN | 19018822 | Insulin Lispro |
| GLIMEPIRIDE | GLIMEPIRIDE | 19117200 | glimepiride |
| INSULIN | INSULIN | 1513849 | Insulin, Zinc, Human |
| ROSIGLITAZONE | ROSIGLITAZONE | 1547507 | rosiglitazone |
| METFORMIN | METFORMIN | 40229050 | Metformin |
| INSULIN | INSULIN | 1513912 | Insulin Lispro |
| INSULIN | INSULIN | 40184293 | Insulin, Aspart Protamine, Human |
| INSULIN | INSULIN | 19095214 | insulin, isophane |
| INSULIN | INSULIN | 19078556 | Regular Insulin, Human |
| INSULIN | INSULIN | 19014975 | insulin, isophane |
| METFORMIN | METFORMIN | 36223143 | Metformin |
| INSULIN | INSULIN | 19120315 | insulin, isophane |
| INSULIN | INSULIN | 1502910 | Insulin Glargine |
| CANAGLIFLOZIN | CANAGLIFLOZIN | 43526472 | canagliflozin |
| CANAGLIFLOZIN | CANAGLIFLOZIN | 43526468 | canagliflozin |
| ALOGLIPTIN | ALOGLIPTIN | 43013901 | alogliptin |
| METFORMIN | METFORMIN | 40164924 | Metformin |
| INSULIN | INSULIN | 19062999 | insulin, isophane |
| INSULIN | INSULIN | 1596955 | insulin, isophane |
| TOLBUTAMIDE | TOLBUTAMIDE | 1502856 | Tolbutamide |
| METFORMIN | METFORMIN | 40164941 | Metformin |
| INSULIN | INSULIN | 1513881 | Insulin Lispro |
| INSULIN | INSULIN | 19044971 | insulin detemir |
| EXENATIDE | EXENATIDE | 1583730 | exenatide |
| INSULIN | INSULIN | 1596977 | Regular Insulin, Human |
| INSULIN | INSULIN | 19110352 | insulin, isophane |
| METFORMIN | METFORMIN | 1503297 | Metformin |

| | | | |
|---|---|---|---|
| INSULIN | INSULIN | 36229086 | Insulin Glargine |
| EXENATIDE | EXENATIDE | 1583722 | exenatide |
| INSULIN | INSULIN | 19110357 | Regular Insulin, Human |
| LIRAGLUTIDE | LIRAGLUTIDE | 45892686 | liraglutide |
| INSULIN | INSULIN | 1502910 | Insulin Glargine |
| METFORMIN | METFORMIN | 1503328 | Metformin |
| INSULIN | INSULIN | 19078557 | Regular Insulin, Human |
| INSULIN | INSULIN | 19078558 | Insulin, Aspart, Human |
| GLIPIZIDE | GLIPIZIDE | 40167637 | Glipizide |
| METFORMIN | METFORMIN | 40164900 | Metformin |
| MIGLITOL | MIGLITOL | 19029030 | miglitol |
| INSULIN | INSULIN | 1596953 | insulin, isophane |
| INSULIN | INSULIN | 1516981 | insulin detemir |
| METFORMIN | METFORMIN | 40063350 | Metformin |
| INSULIN | INSULIN | 46234045 | Insulin, Aspart, Human |
| INSULIN | INSULIN | 1513912 | Insulin Lispro |
| GLIPIZIDE | GLIPIZIDE | 40044224 | Glipizide |
| METFORMIN | METFORMIN | 1503327 | Metformin |
| METFORMIN | METFORMIN | 40164931 | Metformin |
| GLYBURIDE | GLYBURIDE | 19065480 | Glyburide |
| INSULIN | INSULIN | 19095211 | Insulin Lispro |
| METFORMIN | METFORMIN | 40164938 | Metformin |
| GLYBURIDE | GLYBURIDE | 1559760 | Glyburide |
| EXENATIDE | EXENATIDE | 42708079 | exenatide |
| INSULIN | INSULIN | 1502905 | Insulin Glargine |
| DAPAGLIFLOZIN | DAPAGLIFLOZIN | 45776000 | dapagliflozin |
| INSULIN | INSULIN | 19135264 | Regular Insulin, Human |
| METFORMIN | METFORMIN | 40164928 | Metformin |
| REPAGLINIDE | REPAGLINIDE | 19047663 | repaglinide |
| INSULIN | INSULIN | 19110355 | insulin, isophane |
| INSULIN | INSULIN | 40176981 | Insulin, Aspart Protamine, Human |
| ACARBOSE | ACARBOSE | 19021312 | Acarbose |
| EXENATIDE | EXENATIDE | 42903341 | exenatide |
| BROMOCRIPTINE | BROMOCRIPTINE | 40016896 | Bromocriptine |

| | | | |
|---|---|---|---|
| CANAGLIFLOZIN | CANAGLIFLOZIN | 45774724 | canagliflozin |
| INSULIN | INSULIN | 1502905 | Insulin Glargine |
| MIGLITOL | MIGLITOL | 1510206 | miglitol |
| INSULIN | INSULIN | 19078558 | Insulin, Aspart, Human |
| NATEGLINIDE | NATEGLINIDE | 1502830 | nateglinide |
| INSULIN | INSULIN | 36235841 | Regular Insulin, Human |
| GLIPIZIDE | GLIPIZIDE | 1560171 | Glipizide |
| GLIPIZIDE | GLIPIZIDE | 1560233 | Glipizide |
| INSULIN | INSULIN | 35604094 | Insulin Glargine |
| METFORMIN | METFORMIN | 1503297 | Metformin |
| GLIMEPIRIDE | GLIMEPIRIDE | 43011499 | glimepiride |
| LIRAGLUTIDE | LIRAGLUTIDE | 40170913 | liraglutide |
| GLIMEPIRIDE | GLIMEPIRIDE | 1597756 | glimepiride |
| GLIMEPIRIDE | GLIMEPIRIDE | 1597775 | glimepiride |
| ALBIGLUTIDE | ALBIGLUTIDE | 36246906 | albiglutide |
| INSULIN | INSULIN | 36222190 | insulin detemir |
| GLIMEPIRIDE | GLIMEPIRIDE | 1597776 | glimepiride |
| INSULIN | INSULIN | 19078553 | insulin, isophane |
| GLYBURIDE | GLYBURIDE | 19106976 | Glyburide |
| INSULIN | INSULIN | 46233975 | Insulin Lispro |
| INSULIN | INSULIN | 46233975 | Insulin Lispro |
| METFORMIN | METFORMIN | 40164889 | Metformin |
| METFORMIN | METFORMIN | 40164929 | Metformin |
| METFORMIN | METFORMIN | 36235753 | Metformin |
| INSULIN | INSULIN | 1513849 | Insulin, Zinc, Human |
| INSULIN | INSULIN | 19058398 | Insulin Lispro |
| INSULIN | INSULIN | 1502905 | Insulin Glargine |
| METFORMIN | METFORMIN | 36223145 | Metformin |
| LINAGLIPTIN | LINAGLIPTIN | 42708091 | Linagliptin |
| EMPAGLIFLOZIN | EMPAGLIFLOZIN | 45774758 | empagliflozin |
| GLYBURIDE | GLYBURIDE | 19060491 | Glyburide |
| ALOGLIPTIN | ALOGLIPTIN | 43013925 | alogliptin |
| INSULIN | INSULIN | 1596977 | Regular Insulin, Human |
| INSULIN | INSULIN | 19095218 | Regular Insulin, Human |
| INSULIN | INSULIN | 19078530 | insulin, isophane |

| | | | |
|---|---|---|---|
| INSULIN | INSULIN | 1550023 | Insulin Lispro |
| GLYBURIDE | GLYBURIDE | 1559684 | Glyburide |
| METFORMIN | METFORMIN | 40164930 | Metformin |
| GLIMEPIRIDE | GLIMEPIRIDE | 19048036 | glimepiride |
| GLYBURIDE | GLYBURIDE | 1559684 | Glyburide |
| LIRAGLUTIDE | LIRAGLUTIDE | 40169223 | liraglutide |
| INSULIN | INSULIN | 46233975 | Insulin Lispro |
| GLIMEPIRIDE | GLIMEPIRIDE | 19135386 | glimepiride |
| DULAGLUTIDE | DULAGLUTIDE | 45774442 | dulaglutide |
| REPAGLINIDE | REPAGLINIDE | 1516771 | repaglinide |
| ALOGLIPTIN | ALOGLIPTIN | 43013915 | alogliptin |
| INSULIN | INSULIN | 19120790 | Insulin Glargine |
| GLYBURIDE | GLYBURIDE | 40166003 | Glyburide |
| GLIMEPIRIDE | GLIMEPIRIDE | 1597792 | glimepiride |
| METFORMIN | METFORMIN | 1503327 | Metformin |
| METFORMIN | METFORMIN | 1503297 | Metformin |
| INSULIN | INSULIN | 1550028 | Insulin Lispro |
| INSULIN | INSULIN | 19123376 | Insulin Lispro |
| METFORMIN | METFORMIN | 1503297 | Metformin |
| INSULIN | INSULIN | 1550027 | Insulin Lispro |
| INSULIN | INSULIN | 1567232 | Insulin, Aspart, Human |
| INSULIN | INSULIN | 46234098 | Insulin Glargine |
| REPAGLINIDE | REPAGLINIDE | 36240429 | repaglinide |
| METFORMIN | METFORMIN | 40164923 | Metformin |
| INSULIN | INSULIN | 35602724 | insulin degludec |
| ACARBOSE | ACARBOSE | 19040970 | Acarbose |
| INSULIN | INSULIN | 46233975 | Insulin Lispro |
| INSULIN | INSULIN | 19095215 | insulin, isophane |
| SITAGLIPTIN | SITAGLIPTIN | 19125042 | sitagliptin |
| METFORMIN | METFORMIN | 1503297 | Metformin |
| INSULIN | INSULIN | 1513881 | Insulin Lispro |
| INSULIN | INSULIN | 40184294 | Insulin, Aspart Protamine, Human |
| INSULIN | INSULIN | 1502905 | Insulin Glargine |
| GLIPIZIDE | GLIPIZIDE | 40139022 | Glipizide |
| GLIPIZIDE | GLIPIZIDE | 40044224 | Glipizide |

| | | | |
|---|---|---|---|
| NATEGLINIDE | NATEGLINIDE | 36223777 | nateglinide |
| INSULIN | INSULIN | 19078553 | insulin, isophane |
| METFORMIN | METFORMIN | 36235746 | Metformin |
| INSULIN | INSULIN | 46233976 | Insulin Lispro |
| INSULIN | INSULIN | 36229086 | Insulin Glargine |
| EMPAGLIFLOZIN | EMPAGLIFLOZIN | 45892179 | empagliflozin |
| SITAGLIPTIN | SITAGLIPTIN | 36221232 | sitagliptin |
| METFORMIN | METFORMIN | 40164883 | Metformin |
| INSULIN | INSULIN | 45777022 | Regular Insulin, Human |
| INSULIN | INSULIN | 19100127 | insulin, isophane |
| METFORMIN | METFORMIN | 40231393 | Metformin |
| INSULIN | INSULIN | 19078559 | Insulin Glargine |
| EXENATIDE | EXENATIDE | 45774533 | exenatide |
| PIOGLITAZONE | PIOGLITAZONE | 19099207 | pioglitazone |
| INSULIN | INSULIN | 19116559 | insulin, isophane |
| METFORMIN | METFORMIN | 40164914 | Metformin |
| GLIMEPIRIDE | GLIMEPIRIDE | 1597756 | glimepiride |
| INSULIN | INSULIN | 19110352 | insulin, isophane |
| INSULIN | INSULIN | 19120315 | insulin, isophane |
| METFORMIN | METFORMIN | 40164897 | Metformin |
| INSULIN | INSULIN | 19058398 | Insulin Lispro |
| ACARBOSE | ACARBOSE | 1529352 | Acarbose |
| TOLAZAMIDE | TOLAZAMIDE | 1502812 | Tolazamide |
| ALOGLIPTIN | ALOGLIPTIN | 43013922 | alogliptin |
| DULAGLUTIDE | DULAGLUTIDE | 45774447 | dulaglutide |
| EXENATIDE | EXENATIDE | 42708078 | exenatide |
| ACARBOSE | ACARBOSE | 1529352 | Acarbose |
| INSULIN | INSULIN | 1502905 | Insulin Glargine |
| LIRAGLUTIDE | LIRAGLUTIDE | 40169223 | liraglutide |
| GLYBURIDE | GLYBURIDE | 40046211 | Glyburide |
| INSULIN | INSULIN | 1516976 | insulin detemir |
| INSULIN | INSULIN | 1586345 | insulin, isophane |
| EXENATIDE | EXENATIDE | 19083254 | exenatide |

| | | | |
|---|---|---|---|
| INSULIN | INSULIN | 1502910 | Insulin Glargine |
| DAPAGLIFLOZIN | DAPAGLIFLOZIN | 45775999 | dapagliflozin |
| INSULIN | INSULIN | 1513881 | Insulin Lispro |
| METFORMIN | METFORMIN | 40164901 | Metformin |
| METFORMIN | METFORMIN | 42708167 | Metformin |
| INSULIN | INSULIN | 1596959 | insulin, isophane |
| LIRAGLUTIDE | LIRAGLUTIDE | 40170913 | liraglutide |
| INSULIN | INSULIN | 19078530 | insulin, isophane |
| GLIPIZIDE | GLIPIZIDE | 19085004 | Glipizide |
| INSULIN | INSULIN | 19110355 | insulin, isophane |
| INSULIN | INSULIN | 46233975 | Insulin Lispro |
| INSULIN | INSULIN | 19116453 | insulin, isophane |
| INSULIN | INSULIN | 19100128 | Regular Insulin, Human |
| ALBIGLUTIDE | ALBIGLUTIDE | 44816274 | albiglutide |
| INSULIN | INSULIN | 1567198 | Insulin, Aspart, Human |
| INSULIN | INSULIN | 1596953 | insulin, isophane |
| INSULIN | INSULIN | 1597001 | Regular Insulin, Human |
| INSULIN | INSULIN | 19010204 | insulin, isophane |
| INSULIN | INSULIN | 1596959 | insulin, isophane |
| INSULIN | INSULIN | 1513881 | Insulin Lispro |
| LINAGLIPTIN | LINAGLIPTIN | 40239216 | Linagliptin |
| INSULIN | INSULIN | 40166274 | Insulin Lispro |
| ALBIGLUTIDE | ALBIGLUTIDE | 44816274 | albiglutide |
| GLYBURIDE | GLYBURIDE | 40168368 | Glyburide |
| EXENATIDE | EXENATIDE | 1583728 | exenatide |
| GLYBURIDE | GLYBURIDE | 40165994 | Glyburide |
| INSULIN | INSULIN | 46234241 | Regular Insulin, Human |
| GLIMEPIRIDE | GLIMEPIRIDE | 1597775 | glimepiride |
| METFORMIN | METFORMIN | 1503327 | Metformin |
| INSULIN | INSULIN | 19062456 | insulin, isophane |
| NATEGLINIDE | NATEGLINIDE | 40058199 | nateglinide |
| NATEGLINIDE | NATEGLINIDE | 1502829 | nateglinide |

| | | | |
|---|---|---|---|
| INSULIN | INSULIN | 1596959 | insulin, isophane |
| BROMOCRIPTINE | BROMOCRIPTINE | 19062899 | Bromocriptine |
| INSULIN | INSULIN | 40184294 | Insulin, Aspart Protamine, Human |
| GLIPIZIDE | GLIPIZIDE | 36218768 | Glipizide |
| GLIMEPIRIDE | GLIMEPIRIDE | 19100593 | glimepiride |
| SITAGLIPTIN | SITAGLIPTIN | 40243629 | sitagliptin |
| MIGLITOL | MIGLITOL | 1510208 | miglitol |
| EXENATIDE | EXENATIDE | 19083254 | exenatide |
| METFORMIN | METFORMIN | 40164938 | Metformin |
| INSULIN | INSULIN | 19120315 | insulin, isophane |
| SITAGLIPTIN | SITAGLIPTIN | 36221232 | sitagliptin |
| INSULIN | INSULIN | 1502910 | Insulin Glargine |
| ALOGLIPTIN | ALOGLIPTIN | 43013897 | alogliptin |
| INSULIN | INSULIN | 19110357 | Regular Insulin, Human |
| INSULIN | INSULIN | 1597000 | Regular Insulin, Human |
| METFORMIN | METFORMIN | 40164932 | Metformin |
| METFORMIN | METFORMIN | 40164895 | Metformin |
| GLIPIZIDE | GLIPIZIDE | 40167637 | Glipizide |
| INSULIN | INSULIN | 46233975 | Insulin Lispro |
| PRAMLINTIDE | PRAMLINTIDE | 40163890 | Pramlintide |
| INSULIN | INSULIN | 40176981 | Insulin, Aspart Protamine, Human |
| GLIMEPIRIDE | GLIMEPIRIDE | 1597756 | glimepiride |
| INSULIN | INSULIN | 19078559 | Insulin Glargine |
| INSULIN | INSULIN | 1502910 | Insulin Glargine |
| INSULIN | INSULIN | 1596974 | insulin, isophane |
| METFORMIN | METFORMIN | 40164908 | Metformin |
| INSULIN | INSULIN | 1550023 | Insulin Lispro |
| METFORMIN | METFORMIN | 40164929 | Metformin |
| INSULIN | INSULIN | 19078553 | insulin, isophane |
| GLICLAZIDE | GLICLAZIDE | 19102035 | Gliclazide |
| METFORMIN | METFORMIN | 36239257 | Metformin |
| GLYBURIDE | GLYBURIDE | 36231169 | Glyburide |
| GLIMEPIRIDE | GLIMEPIRIDE | 1597796 | glimepiride |

| | | | |
|---|---|---|---|
| INSULIN | INSULIN | 1596960 | insulin, isophane |
| GLIPIZIDE | GLIPIZIDE | 19081295 | Glipizide |
| INSULIN | INSULIN | 1513849 | Insulin, Zinc, Human |
| METFORMIN | METFORMIN | 36221230 | Metformin |
| EXENATIDE | EXENATIDE | 19083253 | exenatide |
| METFORMIN | METFORMIN | 19088818 | Metformin |
| INSULIN | INSULIN | 1513881 | Insulin Lispro |
| LINAGLIPTIN | LINAGLIPTIN | 42708087 | Linagliptin |
| INSULIN | INSULIN | 19044971 | insulin detemir |
| INSULIN | INSULIN | 40051341 | Ultralente Insulin, Human |
| EMPAGLIFLOZIN | EMPAGLIFLOZIN | 46287684 | empagliflozin |
| EXENATIDE | EXENATIDE | 1583722 | exenatide |
| GLIPIZIDE | GLIPIZIDE | 40165972 | Glipizide |
| INSULIN | INSULIN | 40051347 | Insulin Glargine |
| INSULIN | INSULIN | 19014975 | insulin, isophane |
| GLYBURIDE | GLYBURIDE | 19065480 | Glyburide |
| GLIPIZIDE | GLIPIZIDE | 36218766 | Glipizide |
| INSULIN | INSULIN | 1502908 | Insulin Glargine |
| GLIMEPIRIDE | GLIMEPIRIDE | 19117198 | glimepiride |
| PIOGLITAZONE | PIOGLITAZONE | 1525243 | pioglitazone |
| METFORMIN | METFORMIN | 40063352 | Metformin |
| METFORMIN | METFORMIN | 1503327 | Metformin |
| INSULIN | INSULIN | 1596977 | Regular Insulin, Human |
| GLYBURIDE | GLYBURIDE | 19081296 | Glyburide |
| GLIPIZIDE | GLIPIZIDE | 1560233 | Glipizide |
| METFORMIN | METFORMIN | 1503327 | Metformin |
| GLIMEPIRIDE | GLIMEPIRIDE | 19023585 | glimepiride |
| BROMOCRIPTINE | BROMOCRIPTINE | 40016898 | Bromocriptine |
| METFORMIN | METFORMIN | 40164904 | Metformin |
| INSULIN | INSULIN | 1550023 | Insulin Lispro |
| INSULIN | INSULIN | 1516979 | insulin detemir |
| CANAGLIFLOZIN | CANAGLIFLOZIN | 45774708 | canagliflozin |
| METFORMIN | METFORMIN | 40229048 | Metformin |

| | | | |
|---|---|---|---|
| INSULIN | INSULIN | 19078557 | Regular Insulin, Human |
| SAXAGLIPTIN | SAXAGLIPTIN | 36229291 | saxagliptin |
| INSULIN | INSULIN | 19120790 | Insulin Glargine |
| CANAGLIFLOZIN | CANAGLIFLOZIN | 1718901 | canagliflozin |
| PIOGLITAZONE | PIOGLITAZONE | 1525215 | pioglitazone |
| INSULIN | INSULIN | 40176981 | Insulin, Aspart Protamine, Human |
| EXENATIDE | EXENATIDE | 46234118 | exenatide |
| GLIPIZIDE | GLIPIZIDE | 40044253 | Glipizide |
| INSULIN | INSULIN | 19095211 | Insulin Lispro |
| INSULIN | INSULIN | 19078553 | insulin, isophane |
| INSULIN | INSULIN | 40176981 | Insulin, Aspart Protamine, Human |
| INSULIN | INSULIN | 40176981 | Insulin, Aspart Protamine, Human |
| METFORMIN | METFORMIN | 40164894 | Metformin |
| REPAGLINIDE | REPAGLINIDE | 19078030 | repaglinide |
| INSULIN | INSULIN | 19061670 | Regular Insulin, Human |
| INSULIN | INSULIN | 1513912 | Insulin Lispro |
| INSULIN | INSULIN | 1502905 | Insulin Glargine |
| PRAMLINTIDE | PRAMLINTIDE | 40165687 | Pramlintide |
| GLIPIZIDE | GLIPIZIDE | 19077636 | Glipizide |
| GLYBURIDE | GLYBURIDE | 36213841 | Glyburide |
| NATEGLINIDE | NATEGLINIDE | 19110957 | nateglinide |
| METFORMIN | METFORMIN | 40063350 | Metformin |
| ACARBOSE | ACARBOSE | 19040970 | Acarbose |
| EXENATIDE | EXENATIDE | 42708079 | exenatide |
| PIOGLITAZONE | PIOGLITAZONE | 1525216 | pioglitazone |
| EXENATIDE | EXENATIDE | 19083254 | exenatide |
| LIRAGLUTIDE | LIRAGLUTIDE | 45892686 | liraglutide |
| INSULIN | INSULIN | 1550023 | Insulin Lispro |
| INSULIN | INSULIN | 19120315 | insulin, isophane |
| INSULIN | INSULIN | 40166274 | Insulin Lispro |
| INSULIN | INSULIN | 1550023 | Insulin Lispro |
| DAPAGLIFLOZIN | DAPAGLIFLOZIN | 44815995 | dapagliflozin |

| | | | |
|---|---|---|---|
| GLIPIZIDE | GLIPIZIDE | 40167634 | Glipizide |
| DAPAGLIFLOZIN | DAPAGLIFLOZIN | 45776000 | dapagliflozin |
| INSULIN | INSULIN | 19078607 | insulin, isophane |
| INSULIN | INSULIN | 19135263 | insulin, isophane |
| METFORMIN | METFORMIN | 40164897 | Metformin |
| DAPAGLIFLOZIN | DAPAGLIFLOZIN | 44816049 | dapagliflozin |
| PIOGLITAZONE | PIOGLITAZONE | 40099897 | pioglitazone |
| INSULIN | INSULIN | 35602724 | insulin degludec |
| INSULIN | INSULIN | 19095211 | Insulin Lispro |
| NATEGLINIDE | NATEGLINIDE | 36223777 | nateglinide |
| INSULIN | INSULIN | 19078552 | insulin, isophane |
| INSULIN | INSULIN | 1596953 | insulin, isophane |
| BROMOCRIPTINE | BROMOCRIPTINE | 730548 | Bromocriptine |
| INSULIN | INSULIN | 1596918 | insulin, isophane |
| GLYBURIDE | GLYBURIDE | 19030573 | Glyburide |
| INSULIN | INSULIN | 19078633 | Insulin, Regular, Pork |
| LINAGLIPTIN | LINAGLIPTIN | 36221103 | Linagliptin |
| GLYBURIDE | GLYBURIDE | 19110155 | Glyburide |
| INSULIN | INSULIN | 19044971 | insulin detemir |
| INSULIN | INSULIN | 19100128 | Regular Insulin, Human |
| METFORMIN | METFORMIN | 19113780 | Metformin |
| INSULIN | INSULIN | 40051348 | Insulin Glargine |
| INSULIN | INSULIN | 19078551 | insulin, isophane |
| GLIPIZIDE | GLIPIZIDE | 19081295 | Glipizide |
| GLIPIZIDE | GLIPIZIDE | 1560171 | Glipizide |
| LINAGLIPTIN | LINAGLIPTIN | 40239222 | Linagliptin |
| INSULIN | INSULIN | 19078553 | insulin, isophane |
| INSULIN | INSULIN | 40184293 | Insulin, Aspart Protamine, Human |
| INSULIN | INSULIN | 1513912 | Insulin Lispro |
| INSULIN | INSULIN | 46233975 | Insulin Lispro |
| PIOGLITAZONE | PIOGLITAZONE | 1525220 | pioglitazone |
| INSULIN | INSULIN | 1502905 | Insulin Glargine |
| INSULIN | INSULIN | 36236033 | Insulin Lispro |

| | | | |
|---|---|---|---|
| DULAGLUTIDE | DULAGLUTIDE | 45774440 | dulaglutide |
| ACARBOSE | ACARBOSE | 19047612 | Acarbose |
| INSULIN | INSULIN | 19116559 | insulin, isophane |
| INSULIN | INSULIN | 1596955 | insulin, isophane |
| INSULIN | INSULIN | 46233975 | Insulin Lispro |
| INSULIN | INSULIN | 1718604 | Insulin Glargine |
| SAXAGLIPTIN | SAXAGLIPTIN | 36229291 | saxagliptin |
| METFORMIN | METFORMIN | 40164897 | Metformin |
| EXENATIDE | EXENATIDE | 19083253 | exenatide |
| TOLBUTAMIDE | TOLBUTAMIDE | 1502856 | Tolbutamide |
| METFORMIN | METFORMIN | 1503327 | Metformin |
| INSULIN | INSULIN | 1596977 | Regular Insulin, Human |
| INSULIN | INSULIN | 1586344 | insulin, isophane |
| INSULIN | INSULIN | 1567198 | Insulin, Aspart, Human |
| METFORMIN | METFORMIN | 40164932 | Metformin |
| MIGLITOL | MIGLITOL | 1510207 | miglitol |
| EXENATIDE | EXENATIDE | 42705842 | exenatide |
| INSULIN | INSULIN | 19078633 | Insulin, Regular, Pork |
| DULAGLUTIDE | DULAGLUTIDE | 36247186 | dulaglutide |
| INSULIN | INSULIN | 1597012 | Regular Insulin, Human |
| MIGLITOL | MIGLITOL | 19029029 | miglitol |
| GLYBURIDE | GLYBURIDE | 19030580 | Glyburide |
| INSULIN | INSULIN | 1516976 | insulin detemir |
| INSULIN | INSULIN | 1550023 | Insulin Lispro |
| METFORMIN | METFORMIN | 1503328 | Metformin |
| EXENATIDE | EXENATIDE | 1583730 | exenatide |
| GLIPIZIDE | GLIPIZIDE | 19077638 | Glipizide |
| DULAGLUTIDE | DULAGLUTIDE | 45774440 | dulaglutide |
| METFORMIN | METFORMIN | 40164925 | Metformin |
| GLIMEPIRIDE | GLIMEPIRIDE | 1597756 | glimepiride |
| ACARBOSE | ACARBOSE | 1529358 | Acarbose |
| PIOGLITAZONE | PIOGLITAZONE | 36236324 | pioglitazone |
| DULAGLUTIDE | DULAGLUTIDE | 45774445 | dulaglutide |
| GLIMEPIRIDE | GLIMEPIRIDE | 1597798 | glimepiride |
| EMPAGLIFLOZIN | EMPAGLIFLOZIN | 36246886 | empagliflozin |
| GLIPIZIDE | GLIPIZIDE | 40165975 | Glipizide |
| CANAGLIFLOZIN | CANAGLIFLOZIN | 45774714 | canagliflozin |

| | | | |
|---|---|---|---|
| GLIPIZIDE | GLIPIZIDE | 1560171 | Glipizide |
| METFORMIN | METFORMIN | 40167192 | Metformin |
| DULAGLUTIDE | DULAGLUTIDE | 45774442 | dulaglutide |
| INSULIN | INSULIN | 46234241 | Regular Insulin, Human |
| ROSIGLITAZONE | ROSIGLITAZONE | 19079465 | rosiglitazone |
| INSULIN | INSULIN | 1597012 | Regular Insulin, Human |
| GLIPIZIDE | GLIPIZIDE | 19081295 | Glipizide |
| INSULIN | INSULIN | 1596960 | insulin, isophane |
| GLIPIZIDE | GLIPIZIDE | 40044224 | Glipizide |
| INSULIN | INSULIN | 19078558 | Insulin, Aspart, Human |
| INSULIN | INSULIN | 19110352 | insulin, isophane |
| GLYBURIDE | GLYBURIDE | 19030576 | Glyburide |
| METFORMIN | METFORMIN | 40164939 | Metformin |
| CHLORPROPAMIDE | CHLORPROPAMIDE | 1594976 | Chlorpropamide |
| GLIPIZIDE | GLIPIZIDE | 1560233 | Glipizide |
| EXENATIDE | EXENATIDE | 19083254 | exenatide |
| INSULIN | INSULIN | 19110356 | Regular Insulin, Human |
| DULAGLUTIDE | DULAGLUTIDE | 45774442 | dulaglutide |
| INSULIN | INSULIN | 1597000 | Regular Insulin, Human |
| INSULIN | INSULIN | 1596972 | insulin, isophane |
| PIOGLITAZONE | PIOGLITAZONE | 1525217 | pioglitazone |
| ROSIGLITAZONE | ROSIGLITAZONE | 1547506 | rosiglitazone |
| INSULIN | INSULIN | 46234241 | Regular Insulin, Human |
| INSULIN | INSULIN | 1567232 | Insulin, Aspart, Human |
| INSULIN | INSULIN | 19062999 | insulin, isophane |
| SITAGLIPTIN | SITAGLIPTIN | 19125045 | sitagliptin |
| INSULIN | INSULIN | 36236032 | Insulin Lispro |
| INSULIN | INSULIN | 19014975 | insulin, isophane |
| GLYBURIDE | GLYBURIDE | 1559785 | Glyburide |
| GLYBURIDE | GLYBURIDE | 1559684 | Glyburide |
| SAXAGLIPTIN | SAXAGLIPTIN | 36229291 | saxagliptin |
| INSULIN | INSULIN | 19078554 | Regular Insulin, Human |
| METFORMIN | METFORMIN | 40164930 | Metformin |
| INSULIN | INSULIN | 1567198 | Insulin, Aspart, Human |
| INSULIN | INSULIN | 19110352 | insulin, isophane |

| | | | |
|---|---|---|---|
| DULAGLUTIDE | DULAGLUTIDE | 45774442 | dulaglutide |
| MIGLITOL | MIGLITOL | 19081705 | miglitol |
| INSULIN | INSULIN | 1596955 | insulin, isophane |
| INSULIN | INSULIN | 19044971 | insulin detemir |
| INSULIN | INSULIN | 35604094 | Insulin Glargine |
| METFORMIN | METFORMIN | 36236481 | Metformin |
| METFORMIN | METFORMIN | 40164929 | Metformin |
| SITAGLIPTIN | SITAGLIPTIN | 40136769 | sitagliptin |
| GLYBURIDE | GLYBURIDE | 36230035 | Glyburide |
| GLIPIZIDE | GLIPIZIDE | 36235748 | Glipizide |
| GLIPIZIDE | GLIPIZIDE | 19081295 | Glipizide |
| METFORMIN | METFORMIN | 1503328 | Metformin |
| GLIPIZIDE | GLIPIZIDE | 40166814 | Glipizide |
| INSULIN | INSULIN | 19120790 | Insulin Glargine |
| BROMOCRIPTINE | BROMOCRIPTINE | 730572 | Bromocriptine |
| INSULIN | INSULIN | 36236032 | Insulin Lispro |
| BROMOCRIPTINE | BROMOCRIPTINE | 40164160 | Bromocriptine |
| INSULIN | INSULIN | 40051347 | Insulin Glargine |
| INSULIN | INSULIN | 36231450 | Insulin, Aspart, Human |
| GLYBURIDE | GLYBURIDE | 40046211 | Glyburide |
| METFORMIN | METFORMIN | 1503297 | Metformin |
| ALBIGLUTIDE | ALBIGLUTIDE | 44816283 | albiglutide |
| PRAMLINTIDE | PRAMLINTIDE | 40165688 | Pramlintide |
| CANAGLIFLOZIN | CANAGLIFLOZIN | 1718901 | canagliflozin |
| INSULIN | INSULIN | 1596999 | Regular Insulin, Human |
| INSULIN | INSULIN | 19078554 | Regular Insulin, Human |
| EXENATIDE | EXENATIDE | 36234709 | exenatide |
| INSULIN | INSULIN | 46233943 | Insulin Lispro |
| INSULIN | INSULIN | 46234045 | Insulin, Aspart, Human |
| INSULIN | INSULIN | 1513850 | Insulin, Zinc, Human |
| INSULIN | INSULIN | 46234241 | Regular Insulin, Human |
| SITAGLIPTIN | SITAGLIPTIN | 19125047 | sitagliptin |
| INSULIN | INSULIN | 19010204 | insulin, isophane |
| METFORMIN | METFORMIN | 40164920 | Metformin |
| GLYBURIDE | GLYBURIDE | 19106976 | Glyburide |

| | | | |
|---|---|---|---|
| GLIPIZIDE | GLIPIZIDE | 19030445 | Glipizide |
| ACARBOSE | ACARBOSE | 40003412 | Acarbose |
| ALBIGLUTIDE | ALBIGLUTIDE | 44816283 | albiglutide |
| INSULIN | INSULIN | 19120790 | Insulin Glargine |
| METFORMIN | METFORMIN | 40164932 | Metformin |
| INSULIN | INSULIN | 19078558 | Insulin, Aspart, Human |
| INSULIN | INSULIN | 36231458 | insulin, isophane |
| CANAGLIFLOZIN | CANAGLIFLOZIN | 45774721 | canagliflozin |
| GLYBURIDE | GLYBURIDE | 19106976 | Glyburide |
| LIRAGLUTIDE | LIRAGLUTIDE | 40170911 | liraglutide |
| INSULIN | INSULIN | 1567198 | Insulin, Aspart, Human |
| INSULIN | INSULIN | 1596977 | Regular Insulin, Human |
| DULAGLUTIDE | DULAGLUTIDE | 45774442 | dulaglutide |
| SAXAGLIPTIN | SAXAGLIPTIN | 40166037 | saxagliptin |
| METFORMIN | METFORMIN | 19106521 | Metformin |
| GLIPIZIDE | GLIPIZIDE | 19047638 | Glipizide |
| GLIPIZIDE | GLIPIZIDE | 19081295 | Glipizide |
| REPAGLINIDE | REPAGLINIDE | 36213644 | repaglinide |
| BROMOCRIPTINE | BROMOCRIPTINE | 19062900 | Bromocriptine |
| INSULIN | INSULIN | 1502910 | Insulin Glargine |
| INSULIN | INSULIN | 1596977 | Regular Insulin, Human |
| GLIPIZIDE | GLIPIZIDE | 19030443 | Glipizide |
| LIRAGLUTIDE | LIRAGLUTIDE | 36248956 | liraglutide |
| INSULIN | INSULIN | 19012269 | Insulin, Zinc, Human |
| GLIMEPIRIDE | GLIMEPIRIDE | 1597756 | glimepiride |
| GLYBURIDE | GLYBURIDE | 19030579 | Glyburide |
| METFORMIN | METFORMIN | 40164881 | Metformin |
| INSULIN | INSULIN | 1516981 | insulin detemir |
| LIXISENATIDE | LIXISENATIDE | 44506754 | Lixisenatide |
| INSULIN | INSULIN | 40051348 | Insulin Glargine |
| SAXAGLIPTIN | SAXAGLIPTIN | 40166038 | saxagliptin |
| GLIMEPIRIDE | GLIMEPIRIDE | 1597756 | glimepiride |
| LINAGLIPTIN | LINAGLIPTIN | 40239217 | Linagliptin |
| GLIMEPIRIDE | GLIMEPIRIDE | 19117200 | glimepiride |
| EXENATIDE | EXENATIDE | 19083253 | exenatide |
| METFORMIN | METFORMIN | 40153151 | Metformin |
| INSULIN | INSULIN | 19078557 | Regular Insulin, Human |

| | | | |
|---|---|---|---|
| INSULIN | INSULIN | 1502905 | Insulin Glargine |
| PIOGLITAZONE | PIOGLITAZONE | 1525215 | pioglitazone |
| GLYBURIDE | GLYBURIDE | 1559684 | Glyburide |
| INSULIN | INSULIN | 1596999 | Regular Insulin, Human |
| METFORMIN | METFORMIN | 40164891 | Metformin |
| INSULIN | INSULIN | 40184294 | Insulin, Aspart Protamine, Human |
| BROMOCRIPTINE | BROMOCRIPTINE | 19007101 | Bromocriptine |
| PIOGLITAZONE | PIOGLITAZONE | 1525215 | pioglitazone |
| INSULIN | INSULIN | 19110219 | insulin, isophane |
| GLYBURIDE | GLYBURIDE | 40165995 | Glyburide |
| EXENATIDE | EXENATIDE | 19135404 | exenatide |
| INSULIN | INSULIN | 19078607 | insulin, isophane |
| INSULIN | INSULIN | 1516981 | insulin detemir |
| INSULIN | INSULIN | 1596972 | insulin, isophane |
| INSULIN | INSULIN | 1550027 | Insulin Lispro |
| CHLORPROPAMIDE | CHLORPROPAMIDE | 40030456 | Chlorpropamide |
| DAPAGLIFLOZIN | DAPAGLIFLOZIN | 44815996 | dapagliflozin |
| BROMOCRIPTINE | BROMOCRIPTINE | 19006288 | Bromocriptine |
| INSULIN | INSULIN | 1516981 | insulin detemir |
| GLIPIZIDE | GLIPIZIDE | 19047638 | Glipizide |
| EXENATIDE | EXENATIDE | 45774533 | exenatide |
| GLIMEPIRIDE | GLIMEPIRIDE | 1597756 | glimepiride |
| DAPAGLIFLOZIN | DAPAGLIFLOZIN | 44785831 | dapagliflozin |
| INSULIN | INSULIN | 36231459 | Regular Insulin, Human |
| INSULIN | INSULIN | 46233972 | Insulin Lispro |
| METFORMIN | METFORMIN | 1503297 | Metformin |
| INSULIN | INSULIN | 36233552 | insulin, isophane |
| ACARBOSE | ACARBOSE | 40003413 | Acarbose |
| INSULIN | INSULIN | 46233945 | Insulin Lispro |
| INSULIN | INSULIN | 46234045 | Insulin, Aspart, Human |
| DULAGLUTIDE | DULAGLUTIDE | 45774440 | dulaglutide |
| LINAGLIPTIN | LINAGLIPTIN | 40239219 | Linagliptin |
| INSULIN | INSULIN | 1550023 | Insulin Lispro |
| INSULIN | INSULIN | 19078555 | Regular Insulin, Human |
| GLYBURIDE | GLYBURIDE | 19030579 | Glyburide |

| | | | |
|---|---|---|---|
| INSULIN | INSULIN | 1597012 | Regular Insulin, Human |
| SITAGLIPTIN | SITAGLIPTIN | 19125049 | sitagliptin |
| SITAGLIPTIN | SITAGLIPTIN | 19125051 | sitagliptin |
| SITAGLIPTIN | SITAGLIPTIN | 36221233 | sitagliptin |
| METFORMIN | METFORMIN | 40231396 | Metformin |
| INSULIN | INSULIN | 19104458 | insulin, isophane |
| INSULIN | INSULIN | 19078553 | insulin, isophane |
| MIGLITOL | MIGLITOL | 40056705 | miglitol |
| INSULIN | INSULIN | 36224228 | insulin detemir |
| INSULIN | INSULIN | 36222190 | insulin detemir |
| GLIPIZIDE | GLIPIZIDE | 19080793 | Glipizide |
| INSULIN | INSULIN | 1516976 | insulin detemir |
| METFORMIN | METFORMIN | 1503328 | Metformin |
| GLIPIZIDE | GLIPIZIDE | 1560171 | Glipizide |
| ROSIGLITAZONE | ROSIGLITAZONE | 1547508 | rosiglitazone |
| GLIPIZIDE | GLIPIZIDE | 1560171 | Glipizide |
| GLYBURIDE | GLYBURIDE | 19077682 | Glyburide |
| INSULIN | INSULIN | 19100127 | insulin, isophane |
| NATEGLINIDE | NATEGLINIDE | 19045605 | nateglinide |
| GLIPIZIDE | GLIPIZIDE | 1560233 | Glipizide |
| PRAMLINTIDE | PRAMLINTIDE | 1517998 | Pramlintide |
| INSULIN | INSULIN | 1567198 | Insulin, Aspart, Human |
| METFORMIN | METFORMIN | 40164941 | Metformin |
| GLICLAZIDE | GLICLAZIDE | 19059796 | Gliclazide |
| CHLORPROPAMIDE | CHLORPROPAMIDE | 19006907 | Chlorpropamide |
| INSULIN | INSULIN | 19110219 | insulin, isophane |
| INSULIN | INSULIN | 19078610 | insulin, regular, beef-pork |
| LIRAGLUTIDE | LIRAGLUTIDE | 40169223 | liraglutide |
| GLIMEPIRIDE | GLIMEPIRIDE | 1597776 | glimepiride |
| METFORMIN | METFORMIN | 40164905 | Metformin |
| GLYBURIDE | GLYBURIDE | 19030580 | Glyburide |
| ALOGLIPTIN | ALOGLIPTIN | 43013916 | alogliptin |
| METFORMIN | METFORMIN | 40164932 | Metformin |
| GLYBURIDE | GLYBURIDE | 1559684 | Glyburide |
| INSULIN | INSULIN | 19071700 | Insulin Glargine |

| | | | |
|---|---|---|---|
| METFORMIN | METFORMIN | 42708167 | Metformin |
| METFORMIN | METFORMIN | 40164919 | Metformin |
| GLIPIZIDE | GLIPIZIDE | 40165976 | Glipizide |
| ACETOHEXAMIDE | ACETOHEXAMIDE | 19018784 | Acetohexamide |
| INSULIN | INSULIN | 19062999 | insulin, isophane |
| ALBIGLUTIDE | ALBIGLUTIDE | 44816274 | albiglutide |
| INSULIN | INSULIN | 1502905 | Insulin Glargine |
| INSULIN | INSULIN | 19078633 | Insulin, Regular, Pork |
| GLYBURIDE | GLYBURIDE | 19106976 | Glyburide |
| CHLORPROPAMIDE | CHLORPROPAMIDE | 19025944 | Chlorpropamide |
| METFORMIN | METFORMIN | 19113780 | Metformin |
| METFORMIN | METFORMIN | 42708171 | Metformin |
| DAPAGLIFLOZIN | DAPAGLIFLOZIN | 45775999 | dapagliflozin |
| LINAGLIPTIN | LINAGLIPTIN | 42708088 | Linagliptin |
| INSULIN | INSULIN | 19116455 | insulin, isophane |
| GLIPIZIDE | GLIPIZIDE | 40165970 | Glipizide |
| METFORMIN | METFORMIN | 40164882 | Metformin |
| INSULIN | INSULIN | 19009384 | insulin, isophane |
| EMPAGLIFLOZIN | EMPAGLIFLOZIN | 36246887 | empagliflozin |
| INSULIN | INSULIN | 19078559 | Insulin Glargine |
| ACARBOSE | ACARBOSE | 19023063 | Acarbose |
| INSULIN | INSULIN | 1550023 | Insulin Lispro |
| GLIPIZIDE | GLIPIZIDE | 1560233 | Glipizide |
| INSULIN | INSULIN | 1513912 | Insulin Lispro |
| INSULIN | INSULIN | 1550023 | Insulin Lispro |
| GLYBURIDE | GLYBURIDE | 19001408 | Glyburide |
| DAPAGLIFLOZIN | DAPAGLIFLOZIN | 44816049 | dapagliflozin |
| INSULIN | INSULIN | 46234045 | Insulin, Aspart, Human |
| INSULIN | INSULIN | 1597012 | Regular Insulin, Human |

| | | | |
|---|---|---|---|
| GLYBURIDE | GLYBURIDE | 1559684 | Glyburide |
| INSULIN | INSULIN | 40176981 | Insulin, Aspart Protamine, Human |
| CANAGLIFLOZIN | CANAGLIFLOZIN | 43526471 | canagliflozin |
| INSULIN | INSULIN | 40176981 | Insulin, Aspart Protamine, Human |
| INSULIN | INSULIN | 1586345 | insulin, isophane |
| EXENATIDE | EXENATIDE | 42902527 | exenatide |
| INSULIN | INSULIN | 1596960 | insulin, isophane |
| ALOGLIPTIN | ALOGLIPTIN | 43013925 | alogliptin |
| INSULIN | INSULIN | 36231451 | Insulin, Aspart Protamine, Human |
| GLIPIZIDE | GLIPIZIDE | 1560171 | Glipizide |
| NATEGLINIDE | NATEGLINIDE | 19071495 | nateglinide |
| INSULIN | INSULIN | 36231450 | Insulin, Aspart, Human |
| EXENATIDE | EXENATIDE | 1583730 | exenatide |
| GLYBURIDE | GLYBURIDE | 36221460 | Glyburide |
| INSULIN | INSULIN | 19095218 | Regular Insulin, Human |
| GLIMEPIRIDE | GLIMEPIRIDE | 1597774 | glimepiride |
| GLYBURIDE | GLYBURIDE | 40046204 | Glyburide |
| INSULIN | INSULIN | 40176981 | Insulin, Aspart Protamine, Human |
| INSULIN | INSULIN | 35602718 | insulin degludec |
| GLIPIZIDE | GLIPIZIDE | 19077636 | Glipizide |
| GLIPIZIDE | GLIPIZIDE | 40165973 | Glipizide |
| LINAGLIPTIN | LINAGLIPTIN | 36229565 | Linagliptin |
| PIOGLITAZONE | PIOGLITAZONE | 19099208 | pioglitazone |
| METFORMIN | METFORMIN | 1503328 | Metformin |
| REPAGLINIDE | REPAGLINIDE | 19023424 | repaglinide |
| INSULIN | INSULIN | 46233947 | Insulin Lispro |
| INSULIN | INSULIN | 40052045 | insulin, isophane |
| INSULIN | INSULIN | 1567198 | Insulin, Aspart, Human |
| INSULIN | INSULIN | 1586370 | Ultralente Insulin, Human |
| INSULIN | INSULIN | 1516981 | insulin detemir |
| CANAGLIFLOZIN | CANAGLIFLOZIN | 45774714 | canagliflozin |
| INSULIN | INSULIN | 19016117 | Insulin Lispro |
| PIOGLITAZONE | PIOGLITAZONE | 1525242 | pioglitazone |

| | | | |
|---|---|---|---|
| METFORMIN | METFORMIN | 40164920 | Metformin |
| INSULIN | INSULIN | 19080091 | Ultralente Insulin, Human |
| METFORMIN | METFORMIN | 40164900 | Metformin |
| ACARBOSE | ACARBOSE | 36216898 | Acarbose |
| INSULIN | INSULIN | 19111797 | insulin, isophane |
| INSULIN | INSULIN | 1586345 | insulin, isophane |
| METFORMIN | METFORMIN | 1503328 | Metformin |
| METFORMIN | METFORMIN | 40231393 | Metformin |
| INSULIN | INSULIN | 19129441 | insulin human, rDNA origin |
| EXENATIDE | EXENATIDE | 46234118 | exenatide |
| REPAGLINIDE | REPAGLINIDE | 19023425 | repaglinide |
| DAPAGLIFLOZIN | DAPAGLIFLOZIN | 45775860 | dapagliflozin |
| METFORMIN | METFORMIN | 40164932 | Metformin |
| INSULIN | INSULIN | 19078557 | Regular Insulin, Human |
| TOLBUTAMIDE | TOLBUTAMIDE | 19037213 | Tolbutamide |
| LIRAGLUTIDE | LIRAGLUTIDE | 40169223 | liraglutide |
| METFORMIN | METFORMIN | 1503327 | Metformin |
| INSULIN | INSULIN | 1596977 | Regular Insulin, Human |
| INSULIN | INSULIN | 1596941 | insulin, isophane |
| INSULIN | INSULIN | 1596918 | insulin, isophane |
| GLYBURIDE | GLYBURIDE | 19110154 | Glyburide |
| CHLORPROPAMIDE | CHLORPROPAMIDE | 36231727 | Chlorpropamide |
| INSULIN | INSULIN | 1502908 | Insulin Glargine |
| INSULIN | INSULIN | 1516976 | insulin detemir |
| GLYBURIDE | GLYBURIDE | 19060491 | Glyburide |
| GLIPIZIDE | GLIPIZIDE | 1560233 | Glipizide |
| INSULIN | INSULIN | 1513849 | Insulin, Zinc, Human |
| INSULIN | INSULIN | 1550028 | Insulin Lispro |
| GLYBURIDE | GLYBURIDE | 19045630 | Glyburide |
| INSULIN | INSULIN | 42902823 | insulin, isophane |
| INSULIN | INSULIN | 46233945 | Insulin Lispro |

| | | | |
|---|---|---|---|
| INSULIN | INSULIN | 40176981 | Insulin, Aspart Protamine, Human |
| EXENATIDE | EXENATIDE | 42902527 | exenatide |
| METFORMIN | METFORMIN | 40164919 | Metformin |
| METFORMIN | METFORMIN | 36236322 | Metformin |
| ALOGLIPTIN | ALOGLIPTIN | 43013898 | alogliptin |
| METFORMIN | METFORMIN | 40164919 | Metformin |
| ACARBOSE | ACARBOSE | 19040970 | Acarbose |
| ALOGLIPTIN | ALOGLIPTIN | 43013897 | alogliptin |
| INSULIN | INSULIN | 40176981 | Insulin, Aspart Protamine, Human |
| INSULIN | INSULIN | 19061670 | Regular Insulin, Human |
| METFORMIN | METFORMIN | 36235752 | Metformin |
| EXENATIDE | EXENATIDE | 19135404 | exenatide |
| INSULIN | INSULIN | 1596959 | insulin, isophane |
| ROSIGLITAZONE | ROSIGLITAZONE | 1547504 | rosiglitazone |
| DAPAGLIFLOZIN | DAPAGLIFLOZIN | 45776000 | dapagliflozin |
| INSULIN | INSULIN | 35602728 | insulin degludec |
| SITAGLIPTIN | SITAGLIPTIN | 40243628 | sitagliptin |
| INSULIN | INSULIN | 1502910 | Insulin Glargine |
| EXENATIDE | EXENATIDE | 1583728 | exenatide |
| METFORMIN | METFORMIN | 40164886 | Metformin |
| EXENATIDE | EXENATIDE | 1583727 | exenatide |
| INSULIN | INSULIN | 1596956 | insulin, isophane |
| DAPAGLIFLOZIN | DAPAGLIFLOZIN | 36248559 | dapagliflozin |
| METFORMIN | METFORMIN | 40164947 | Metformin |
| ROSIGLITAZONE | ROSIGLITAZONE | 40099633 | rosiglitazone |
| INSULIN | INSULIN | 1550027 | Insulin Lispro |
| INSULIN | INSULIN | 19078553 | insulin, isophane |
| GLYBURIDE | GLYBURIDE | 1559760 | Glyburide |
| INSULIN | INSULIN | 1567232 | Insulin, Aspart, Human |
| EXENATIDE | EXENATIDE | 19083253 | exenatide |

| | | | |
|---|---|---|---|
| GLYBURIDE | GLYBURIDE | 40168370 | Glyburide |
| METFORMIN | METFORMIN | 36239257 | Metformin |
| INSULIN | INSULIN | 46234047 | Insulin, Aspart, Human |
| INSULIN | INSULIN | 1596977 | Regular Insulin, Human |
| GLIPIZIDE | GLIPIZIDE | 1560233 | Glipizide |
| GLIMEPIRIDE | GLIMEPIRIDE | 1597759 | glimepiride |
| LIRAGLUTIDE | LIRAGLUTIDE | 40169223 | liraglutide |
| EXENATIDE | EXENATIDE | 1583727 | exenatide |
| GLYBURIDE | GLYBURIDE | 19030572 | Glyburide |
| INSULIN | INSULIN | 1567232 | Insulin, Aspart, Human |
| PIOGLITAZONE | PIOGLITAZONE | 1525216 | pioglitazone |
| INSULIN | INSULIN | 1586370 | Ultralente Insulin, Human |
| INSULIN | INSULIN | 46233976 | Insulin Lispro |
| INSULIN | INSULIN | 19078552 | insulin, isophane |
| ALBIGLUTIDE | ALBIGLUTIDE | 44816274 | albiglutide |
| METFORMIN | METFORMIN | 40229046 | Metformin |
| ALBIGLUTIDE | ALBIGLUTIDE | 44816274 | albiglutide |
| GLYBURIDE | GLYBURIDE | 40046143 | Glyburide |
| GLIMEPIRIDE | GLIMEPIRIDE | 1597795 | glimepiride |
| MIGLITOL | MIGLITOL | 1510207 | miglitol |
| INSULIN | INSULIN | 36233552 | insulin, isophane |
| INSULIN | INSULIN | 19120790 | Insulin Glargine |
| INSULIN | INSULIN | 40166275 | Insulin Lispro |
| CANAGLIFLOZIN | CANAGLIFLOZIN | 43526467 | canagliflozin |
| METFORMIN | METFORMIN | 40164948 | Metformin |
| GLIPIZIDE | GLIPIZIDE | 1560171 | Glipizide |
| METFORMIN | METFORMIN | 40163926 | Metformin |
| GLIMEPIRIDE | GLIMEPIRIDE | 1597756 | glimepiride |
| GLYBURIDE | GLYBURIDE | 40168370 | Glyburide |
| INSULIN | INSULIN | 1516976 | insulin detemir |
| ALOGLIPTIN | ALOGLIPTIN | 43013899 | alogliptin |
| INSULIN | INSULIN | 40051347 | Insulin Glargine |
| INSULIN | INSULIN | 19111795 | insulin, isophane |
| METFORMIN | METFORMIN | 42705992 | Metformin |
| METFORMIN | METFORMIN | 40164932 | Metformin |

| | | | |
|---|---|---|---|
| EMPAGLIFLOZIN | EMPAGLIFLOZIN | 1592800 | empagliflozin |
| INSULIN | INSULIN | 1596977 | Regular Insulin, Human |
| INSULIN | INSULIN | 19009384 | insulin, isophane |
| EXENATIDE | EXENATIDE | 46234116 | exenatide |
| PIOGLITAZONE | PIOGLITAZONE | 19099208 | pioglitazone |
| GLYBURIDE | GLYBURIDE | 36233495 | Glyburide |
| METFORMIN | METFORMIN | 40164932 | Metformin |
| ROSIGLITAZONE | ROSIGLITAZONE | 36227624 | rosiglitazone |
| METFORMIN | METFORMIN | 42705992 | Metformin |
| GLYBURIDE | GLYBURIDE | 1559784 | Glyburide |
| INSULIN | INSULIN | 19110357 | Regular Insulin, Human |
| LIRAGLUTIDE | LIRAGLUTIDE | 40169223 | liraglutide |
| GLIPIZIDE | GLIPIZIDE | 1560171 | Glipizide |
| METFORMIN | METFORMIN | 40164891 | Metformin |
| INSULIN | INSULIN | 1562586 | Insulin, Zinc, Pork |
| INSULIN | INSULIN | 46234045 | Insulin, Aspart, Human |
| INSULIN | INSULIN | 1550023 | Insulin Lispro |
| EXENATIDE | EXENATIDE | 46234118 | exenatide |
| EXENATIDE | EXENATIDE | 36234709 | exenatide |
| METFORMIN | METFORMIN | 42708167 | Metformin |
| ALOGLIPTIN | ALOGLIPTIN | 36246267 | alogliptin |
| INSULIN | INSULIN | 1513881 | Insulin Lispro |
| INSULIN | INSULIN | 1513912 | Insulin Lispro |
| METFORMIN | METFORMIN | 40063356 | Metformin |
| INSULIN | INSULIN | 1550023 | Insulin Lispro |
| NATEGLINIDE | NATEGLINIDE | 1502827 | nateglinide |
| INSULIN | INSULIN | 40051348 | Insulin Glargine |
| INSULIN | INSULIN | 1550027 | Insulin Lispro |
| REPAGLINIDE | REPAGLINIDE | 36213643 | repaglinide |
| GLIPIZIDE | GLIPIZIDE | 1560233 | Glipizide |
| INSULIN | INSULIN | 19120315 | insulin, isophane |
| GLYBURIDE | GLYBURIDE | 19077682 | Glyburide |
| EXENATIDE | EXENATIDE | 1583727 | exenatide |

| | | | |
|---|---|---|---|
| INSULIN | INSULIN | 1596973 | insulin, isophane |
| GLYBURIDE | GLYBURIDE | 1559784 | Glyburide |
| METFORMIN | METFORMIN | 40164929 | Metformin |
| METFORMIN | METFORMIN | 40163927 | Metformin |
| GLIMEPIRIDE | GLIMEPIRIDE | 40059665 | glimepiride |
| METFORMIN | METFORMIN | 40164944 | Metformin |
| METFORMIN | METFORMIN | 1503328 | Metformin |
| INSULIN | INSULIN | 19116559 | insulin, isophane |
| EXENATIDE | EXENATIDE | 46234118 | exenatide |
| GLIPIZIDE | GLIPIZIDE | 19077638 | Glipizide |
| INSULIN | INSULIN | 19116454 | insulin, isophane |
| INSULIN | INSULIN | 19078553 | insulin, isophane |
| INSULIN | INSULIN | 19078553 | insulin, isophane |
| GLIPIZIDE | GLIPIZIDE | 40166816 | Glipizide |
| GLIMEPIRIDE | GLIMEPIRIDE | 1597801 | glimepiride |
| INSULIN | INSULIN | 19110352 | insulin, isophane |
| SITAGLIPTIN | SITAGLIPTIN | 19125049 | sitagliptin |
| INSULIN | INSULIN | 1516979 | insulin detemir |
| GLYBURIDE | GLYBURIDE | 36221460 | Glyburide |
| INSULIN | INSULIN | 40166274 | Insulin Lispro |
| ALOGLIPTIN | ALOGLIPTIN | 43013912 | alogliptin |
| INSULIN | INSULIN | 19120790 | Insulin Glargine |
| INSULIN | INSULIN | 36231450 | Insulin, Aspart, Human |
| METFORMIN | METFORMIN | 1503327 | Metformin |
| INSULIN | INSULIN | 19131582 | Insulin, Glulisine, Human |
| GLIMEPIRIDE | GLIMEPIRIDE | 1597758 | glimepiride |
| INSULIN | INSULIN | 36235840 | insulin, isophane |
| GLIPIZIDE | GLIPIZIDE | 1560171 | Glipizide |
| ACARBOSE | ACARBOSE | 40003412 | Acarbose |
| BROMOCRIPTINE | BROMOCRIPTINE | 19006288 | Bromocriptine |
| INSULIN | INSULIN | 46234241 | Regular Insulin, Human |

| | | | |
|---|---|---|---|
| INSULIN | INSULIN | 19078551 | insulin, isophane |
| INSULIN | INSULIN | 19094122 | Regular Insulin, Human |
| INSULIN | INSULIN | 19090210 | Lente Insulin, Beef |
| INSULIN | INSULIN | 19095214 | insulin, isophane |
| INSULIN | INSULIN | 19061670 | Regular Insulin, Human |
| METFORMIN | METFORMIN | 40231388 | Metformin |
| METFORMIN | METFORMIN | 1503327 | Metformin |
| INSULIN | INSULIN | 1567198 | Insulin, Aspart, Human |
| METFORMIN | METFORMIN | 42708175 | Metformin |
| GLIPIZIDE | GLIPIZIDE | 36241808 | Glipizide |
| GLIMEPIRIDE | GLIMEPIRIDE | 1597798 | glimepiride |
| LIRAGLUTIDE | LIRAGLUTIDE | 40170913 | liraglutide |
| INSULIN | INSULIN | 19062999 | insulin, isophane |
| INSULIN | INSULIN | 19135263 | insulin, isophane |
| INSULIN | INSULIN | 19010204 | insulin, isophane |
| INSULIN | INSULIN | 1513914 | Insulin Lispro |
| INSULIN | INSULIN | 1597012 | Regular Insulin, Human |
| INSULIN | INSULIN | 19116559 | insulin, isophane |
| INSULIN | INSULIN | 1516981 | insulin detemir |
| GLYBURIDE | GLYBURIDE | 40165995 | Glyburide |
| METFORMIN | METFORMIN | 1503328 | Metformin |
| METFORMIN | METFORMIN | 40171449 | Metformin |
| METFORMIN | METFORMIN | 40164924 | Metformin |
| GLIMEPIRIDE | GLIMEPIRIDE | 1597795 | glimepiride |
| INSULIN | INSULIN | 1550023 | Insulin Lispro |
| INSULIN | INSULIN | 1596919 | insulin, isophane |
| INSULIN | INSULIN | 1567198 | Insulin, Aspart, Human |
| INSULIN | INSULIN | 1550027 | Insulin Lispro |
| BROMOCRIPTINE | BROMOCRIPTINE | 19083957 | Bromocriptine |

| | | | |
|---|---|---|---|
| GLYBURIDE | GLYBURIDE | 40165993 | Glyburide |
| EMPAGLIFLOZIN | EMPAGLIFLOZIN | 45892179 | empagliflozin |
| SITAGLIPTIN | SITAGLIPTIN | 19125047 | sitagliptin |
| GLIPIZIDE | GLIPIZIDE | 40167638 | Glipizide |
| ACARBOSE | ACARBOSE | 36242198 | Acarbose |
| INSULIN | INSULIN | 19123376 | Insulin Lispro |
| INSULIN | INSULIN | 1550023 | Insulin Lispro |
| SITAGLIPTIN | SITAGLIPTIN | 40136770 | sitagliptin |
| INSULIN | INSULIN | 19062999 | insulin, isophane |
| INSULIN | INSULIN | 19095199 | Lente Insulin, Beef-Pork |
| INSULIN | INSULIN | 1550023 | Insulin Lispro |
| LIRAGLUTIDE | LIRAGLUTIDE | 40170911 | liraglutide |
| EXENATIDE | EXENATIDE | 19083254 | exenatide |
| METFORMIN | METFORMIN | 36221231 | Metformin |
| METFORMIN | METFORMIN | 40164938 | Metformin |
| SAXAGLIPTIN | SAXAGLIPTIN | 40166041 | saxagliptin |
| INSULIN | INSULIN | 36222190 | insulin detemir |
| INSULIN | INSULIN | 1596940 | insulin, isophane |
| INSULIN | INSULIN | 19078557 | Regular Insulin, Human |
| INSULIN | INSULIN | 46233975 | Insulin Lispro |
| INSULIN | INSULIN | 40184294 | Insulin, Aspart Protamine, Human |
| GLIPIZIDE | GLIPIZIDE | 19081295 | Glipizide |
| PIOGLITAZONE | PIOGLITAZONE | 1525215 | pioglitazone |
| DAPAGLIFLOZIN | DAPAGLIFLOZIN | 44785831 | dapagliflozin |
| GLIMEPIRIDE | GLIMEPIRIDE | 1597774 | glimepiride |
| REPAGLINIDE | REPAGLINIDE | 19023426 | repaglinide |
| GLYBURIDE | GLYBURIDE | 40046204 | Glyburide |
| INSULIN | INSULIN | 36229086 | Insulin Glargine |
| METFORMIN | METFORMIN | 40164928 | Metformin |
| SITAGLIPTIN | SITAGLIPTIN | 19125047 | sitagliptin |
| METFORMIN | METFORMIN | 40164882 | Metformin |
| INSULIN | INSULIN | 1513881 | Insulin Lispro |
| INSULIN | INSULIN | 46233976 | Insulin Lispro |
| INSULIN | INSULIN | 1516981 | insulin detemir |
| INSULIN | INSULIN | 19111795 | insulin, isophane |

| | | | |
|---|---|---|---|
| INSULIN | INSULIN | 46233975 | Insulin Lispro |
| METFORMIN | METFORMIN | 1503297 | Metformin |
| LIRAGLUTIDE | LIRAGLUTIDE | 45892686 | liraglutide |
| INSULIN | INSULIN | 19120315 | insulin, isophane |
| GLIPIZIDE | GLIPIZIDE | 19047638 | Glipizide |
| GLIPIZIDE | GLIPIZIDE | 1560171 | Glipizide |
| METFORMIN | METFORMIN | 36235787 | Metformin |
| GLIPIZIDE | GLIPIZIDE | 40044253 | Glipizide |
| INSULIN | INSULIN | 19078609 | insulin, regular, beef-pork |
| INSULIN | INSULIN | 1597012 | Regular Insulin, Human |
| CANAGLIFLOZIN | CANAGLIFLOZIN | 45774716 | canagliflozin |
| EXENATIDE | EXENATIDE | 1583730 | exenatide |
| NATEGLINIDE | NATEGLINIDE | 19071495 | nateglinide |
| INSULIN | INSULIN | 19044971 | insulin detemir |
| INSULIN | INSULIN | 46233970 | Insulin Lispro |
| INSULIN | INSULIN | 1596977 | Regular Insulin, Human |
| INSULIN | INSULIN | 46233945 | Insulin Lispro |
| CANAGLIFLOZIN | CANAGLIFLOZIN | 43526468 | canagliflozin |
| METFORMIN | METFORMIN | 40164941 | Metformin |
| INSULIN | INSULIN | 40184294 | Insulin, Aspart Protamine, Human |
| INSULIN | INSULIN | 36231459 | Regular Insulin, Human |
| METFORMIN | METFORMIN | 40063350 | Metformin |
| ALOGLIPTIN | ALOGLIPTIN | 43013884 | alogliptin |
| INSULIN | INSULIN | 19120315 | insulin, isophane |
| GLYBURIDE | GLYBURIDE | 40166002 | Glyburide |
| INSULIN | INSULIN | 1550028 | Insulin Lispro |
| DAPAGLIFLOZIN | DAPAGLIFLOZIN | 45775867 | dapagliflozin |
| EMPAGLIFLOZIN | EMPAGLIFLOZIN | 46287691 | empagliflozin |
| INSULIN | INSULIN | 40176981 | Insulin, Aspart Protamine, Human |
| METFORMIN | METFORMIN | 42708171 | Metformin |
| INSULIN | INSULIN | 1596960 | insulin, isophane |

| | | | |
|---|---|---|---|
| DAPAGLIFLOZIN | DAPAGLIFLOZIN | 36247907 | dapagliflozin |
| INSULIN | INSULIN | 36229086 | Insulin Glargine |
| MIGLITOL | MIGLITOL | 1510208 | miglitol |
| INSULIN | INSULIN | 46234045 | Insulin, Aspart, Human |
| BROMOCRIPTINE | BROMOCRIPTINE | 19007070 | Bromocriptine |
| GLIMEPIRIDE | GLIMEPIRIDE | 40059666 | glimepiride |
| DULAGLUTIDE | DULAGLUTIDE | 45774447 | dulaglutide |
| METFORMIN | METFORMIN | 40163926 | Metformin |
| LIRAGLUTIDE | LIRAGLUTIDE | 45892686 | liraglutide |
| GLIPIZIDE | GLIPIZIDE | 19085004 | Glipizide |
| INSULIN | INSULIN | 1516978 | insulin detemir |
| GLIPIZIDE | GLIPIZIDE | 19117945 | Glipizide |
| GLYBURIDE | GLYBURIDE | 19030580 | Glyburide |
| METFORMIN | METFORMIN | 1503297 | Metformin |
| GLIMEPIRIDE | GLIMEPIRIDE | 1597776 | glimepiride |
| INSULIN | INSULIN | 19110355 | insulin, isophane |
| INSULIN | INSULIN | 19120790 | Insulin Glargine |
| GLYBURIDE | GLYBURIDE | 19106976 | Glyburide |
| DAPAGLIFLOZIN | DAPAGLIFLOZIN | 44785477 | dapagliflozin |
| INSULIN | INSULIN | 19120315 | insulin, isophane |
| EMPAGLIFLOZIN | EMPAGLIFLOZIN | 45774893 | empagliflozin |
| GLYBURIDE | GLYBURIDE | 1559684 | Glyburide |
| GLIMEPIRIDE | GLIMEPIRIDE | 1597760 | glimepiride |
| INSULIN | INSULIN | 1516976 | insulin detemir |
| GLIMEPIRIDE | GLIMEPIRIDE | 19117200 | glimepiride |
| METFORMIN | METFORMIN | 40164942 | Metformin |
| INSULIN | INSULIN | 1502905 | Insulin Glargine |
| INSULIN | INSULIN | 19078610 | insulin, regular, beef-pork |
| GLIPIZIDE | GLIPIZIDE | 19077638 | Glipizide |
| EXENATIDE | EXENATIDE | 1583727 | exenatide |
| INSULIN | INSULIN | 19078557 | Regular Insulin, Human |
| INSULIN | INSULIN | 40166274 | Insulin Lispro |
| INSULIN | INSULIN | 19080091 | Ultralente Insulin, Human |
| INSULIN | INSULIN | 19078554 | Regular Insulin, Human |
| ACARBOSE | ACARBOSE | 1529358 | Acarbose |

| | | | |
|---|---|---|---|
| INSULIN | INSULIN | 46233970 | Insulin Lispro |
| TOLAZAMIDE | TOLAZAMIDE | 19082949 | Tolazamide |
| INSULIN | INSULIN | 19078558 | Insulin, Aspart, Human |
| INSULIN | INSULIN | 19110357 | Regular Insulin, Human |
| GLIMEPIRIDE | GLIMEPIRIDE | 36234085 | glimepiride |
| GLIPIZIDE | GLIPIZIDE | 40165969 | Glipizide |
| TOLAZAMIDE | TOLAZAMIDE | 19082950 | Tolazamide |
| INSULIN | INSULIN | 19111795 | insulin, isophane |
| GLIMEPIRIDE | GLIMEPIRIDE | 1597775 | glimepiride |
| INSULIN | INSULIN | 1596972 | insulin, isophane |
| INSULIN | INSULIN | 19095211 | Insulin Lispro |
| METFORMIN | METFORMIN | 40231391 | Metformin |
| GLIPIZIDE | GLIPIZIDE | 40167636 | Glipizide |
| EMPAGLIFLOZIN | EMPAGLIFLOZIN | 45892176 | empagliflozin |
| GLIMEPIRIDE | GLIMEPIRIDE | 1597792 | glimepiride |
| ALBIGLUTIDE | ALBIGLUTIDE | 44816274 | albiglutide |
| PIOGLITAZONE | PIOGLITAZONE | 40099898 | pioglitazone |
| INSULIN | INSULIN | 1586370 | Ultralente Insulin, Human |
| EXENATIDE | EXENATIDE | 19083254 | exenatide |
| ROSIGLITAZONE | ROSIGLITAZONE | 19099278 | rosiglitazone |
| DULAGLUTIDE | DULAGLUTIDE | 45774447 | dulaglutide |
| INSULIN | INSULIN | 1502905 | Insulin Glargine |
| INSULIN | INSULIN | 1502908 | Insulin Glargine |
| INSULIN | INSULIN | 1502905 | Insulin Glargine |
| LIRAGLUTIDE | LIRAGLUTIDE | 40169222 | liraglutide |
| EXENATIDE | EXENATIDE | 45774533 | exenatide |
| GLYBURIDE | GLYBURIDE | 19030575 | Glyburide |
| ACARBOSE | ACARBOSE | 19040969 | Acarbose |
| GLIMEPIRIDE | GLIMEPIRIDE | 43011831 | glimepiride |
| INSULIN | INSULIN | 1550023 | Insulin Lispro |
| EXENATIDE | EXENATIDE | 19083253 | exenatide |
| GLIPIZIDE | GLIPIZIDE | 19079990 | Glipizide |
| INSULIN | INSULIN | 1516981 | insulin detemir |
| INSULIN | INSULIN | 1596977 | Regular Insulin, Human |
| METFORMIN | METFORMIN | 40164930 | Metformin |
| SITAGLIPTIN | SITAGLIPTIN | 40136770 | sitagliptin |
| ROSIGLITAZONE | ROSIGLITAZONE | 1547554 | rosiglitazone |
| SITAGLIPTIN | SITAGLIPTIN | 42901620 | sitagliptin |

| | | | |
|---|---|---|---|
| INSULIN | INSULIN | 19110352 | insulin, isophane |
| METFORMIN | METFORMIN | 40164897 | Metformin |
| INSULIN | INSULIN | 19078552 | insulin, isophane |
| INSULIN | INSULIN | 1513876 | Insulin, Protamine Lispro, Human |
| ALOGLIPTIN | ALOGLIPTIN | 36246271 | alogliptin |
| GLIPIZIDE | GLIPIZIDE | 1560171 | Glipizide |
| INSULIN | INSULIN | 46233946 | Insulin Lispro |
| INSULIN | INSULIN | 40184293 | Insulin, Aspart Protamine, Human |
| SITAGLIPTIN | SITAGLIPTIN | 40243630 | sitagliptin |
| DAPAGLIFLOZIN | DAPAGLIFLOZIN | 45775866 | dapagliflozin |
| GLIPIZIDE | GLIPIZIDE | 40139022 | Glipizide |
| EMPAGLIFLOZIN | EMPAGLIFLOZIN | 45774895 | empagliflozin |
| PRAMLINTIDE | PRAMLINTIDE | 46276272 | Pramlintide |
| BROMOCRIPTINE | BROMOCRIPTINE | 36238866 | Bromocriptine |
| ALOGLIPTIN | ALOGLIPTIN | 43013925 | alogliptin |
| METFORMIN | METFORMIN | 40231396 | Metformin |
| EXENATIDE | EXENATIDE | 19083253 | exenatide |
| INSULIN | INSULIN | 1597001 | Regular Insulin, Human |
| MIGLITOL | MIGLITOL | 1510202 | miglitol |
| LIRAGLUTIDE | LIRAGLUTIDE | 40170912 | liraglutide |
| LINAGLIPTIN | LINAGLIPTIN | 42708089 | Linagliptin |
| METFORMIN | METFORMIN | 40231393 | Metformin |
| INSULIN | INSULIN | 45776625 | Regular Insulin, Human |
| LIRAGLUTIDE | LIRAGLUTIDE | 45892684 | liraglutide |
| DAPAGLIFLOZIN | DAPAGLIFLOZIN | 36248560 | dapagliflozin |
| GLIMEPIRIDE | GLIMEPIRIDE | 1597799 | glimepiride |
| METFORMIN | METFORMIN | 1503327 | Metformin |
| METFORMIN | METFORMIN | 40164900 | Metformin |
| ACARBOSE | ACARBOSE | 36242198 | Acarbose |
| LIRAGLUTIDE | LIRAGLUTIDE | 40170913 | liraglutide |
| INSULIN | INSULIN | 19120790 | Insulin Glargine |
| PIOGLITAZONE | PIOGLITAZONE | 19099208 | pioglitazone |
| ROSIGLITAZONE | ROSIGLITAZONE | 19107077 | rosiglitazone |
| EXENATIDE | EXENATIDE | 1583730 | exenatide |
| INSULIN | INSULIN | 40166275 | Insulin Lispro |
| DAPAGLIFLOZIN | DAPAGLIFLOZIN | 44785477 | dapagliflozin |

| | | | |
|---|---|---|---|
| DULAGLUTIDE | DULAGLUTIDE | 45774447 | dulaglutide |
| METFORMIN | METFORMIN | 1503328 | Metformin |
| INSULIN | INSULIN | 1544873 | Insulin, Glulisine, Human |
| GLYBURIDE | GLYBURIDE | 19077681 | Glyburide |
| METFORMIN | METFORMIN | 40164930 | Metformin |
| EMPAGLIFLOZIN | EMPAGLIFLOZIN | 46287687 | empagliflozin |
| DULAGLUTIDE | DULAGLUTIDE | 46234471 | dulaglutide |
| GLIPIZIDE | GLIPIZIDE | 19077636 | Glipizide |
| INSULIN | INSULIN | 1513849 | Insulin, Zinc, Human |
| EXENATIDE | EXENATIDE | 42708079 | exenatide |
| METFORMIN | METFORMIN | 42705990 | Metformin |
| CANAGLIFLOZIN | CANAGLIFLOZIN | 1718901 | canagliflozin |
| LIRAGLUTIDE | LIRAGLUTIDE | 40169223 | liraglutide |
| EXENATIDE | EXENATIDE | 1583727 | exenatide |
| INSULIN | INSULIN | 1516976 | insulin detemir |
| INSULIN | INSULIN | 45777022 | Regular Insulin, Human |
| LINAGLIPTIN | LINAGLIPTIN | 42708089 | Linagliptin |
| INSULIN | INSULIN | 36231459 | Regular Insulin, Human |
| INSULIN | INSULIN | 19095211 | Insulin Lispro |
| EXENATIDE | EXENATIDE | 1583730 | exenatide |
| GLIMEPIRIDE | GLIMEPIRIDE | 19117198 | glimepiride |
| INSULIN | INSULIN | 19110352 | insulin, isophane |
| INSULIN | INSULIN | 1502905 | Insulin Glargine |
| INSULIN | INSULIN | 1596977 | Regular Insulin, Human |
| INSULIN | INSULIN | 36229086 | Insulin Glargine |
| INSULIN | INSULIN | 1567198 | Insulin, Aspart, Human |
| INSULIN | INSULIN | 42899447 | Insulin, Pork |
| INSULIN | INSULIN | 40051347 | Insulin Glargine |
| INSULIN | INSULIN | 19078553 | insulin, isophane |
| INSULIN | INSULIN | 1596998 | Regular Insulin, Human |
| INSULIN | INSULIN | 19110352 | insulin, isophane |
| INSULIN | INSULIN | 1513849 | Insulin, Zinc, Human |
| ROSIGLITAZONE | ROSIGLITAZONE | 1547507 | rosiglitazone |
| SAXAGLIPTIN | SAXAGLIPTIN | 40166042 | saxagliptin |

| | | | |
|---|---|---|---|
| EXENATIDE | EXENATIDE | 1583730 | exenatide |
| ALOGLIPTIN | ALOGLIPTIN | 43013913 | alogliptin |
| BROMOCRIPTINE | BROMOCRIPTINE | 19063998 | Bromocriptine |
| ACARBOSE | ACARBOSE | 1529331 | Acarbose |
| INSULIN | INSULIN | 36222190 | insulin detemir |
| GLIPIZIDE | GLIPIZIDE | 40160874 | Glipizide |
| INSULIN | INSULIN | 36222190 | insulin detemir |
| INSULIN | INSULIN | 40176981 | Insulin, Aspart Protamine, Human |
| GLYBURIDE | GLYBURIDE | 40168374 | Glyburide |
| MIGLITOL | MIGLITOL | 19029030 | miglitol |
| INSULIN | INSULIN | 1596941 | insulin, isophane |
| CANAGLIFLOZIN | CANAGLIFLOZIN | 36246877 | canagliflozin |
| INSULIN | INSULIN | 1550023 | Insulin Lispro |
| INSULIN | INSULIN | 46221557 | Insulin Glargine |
| INSULIN | INSULIN | 19078557 | Regular Insulin, Human |
| GLYBURIDE | GLYBURIDE | 19060409 | Glyburide |
| INSULIN | INSULIN | 35602724 | insulin degludec |
| INSULIN | INSULIN | 1567198 | Insulin, Aspart, Human |
| DAPAGLIFLOZIN | DAPAGLIFLOZIN | 45776000 | dapagliflozin |
| INSULIN | INSULIN | 1718889 | Regular Insulin, Human |
| METFORMIN | METFORMIN | 1503297 | Metformin |
| CHLORPROPAMIDE | CHLORPROPAMIDE | 1594975 | Chlorpropamide |
| SAXAGLIPTIN | SAXAGLIPTIN | 36229292 | saxagliptin |
| METFORMIN | METFORMIN | 40164905 | Metformin |
| METFORMIN | METFORMIN | 40164909 | Metformin |
| INSULIN | INSULIN | 35602728 | insulin degludec |
| INSULIN | INSULIN | 19078554 | Regular Insulin, Human |
| METFORMIN | METFORMIN | 40164893 | Metformin |
| EXENATIDE | EXENATIDE | 19083254 | exenatide |
| GLIMEPIRIDE | GLIMEPIRIDE | 1597774 | glimepiride |

| | | | |
|---|---|---|---|
| TOLAZAMIDE | TOLAZAMIDE | 19036965 | Tolazamide |
| INSULIN | INSULIN | 1596977 | Regular Insulin, Human |
| INSULIN | INSULIN | 40184293 | Insulin, Aspart Protamine, Human |
| INSULIN | INSULIN | 1502905 | Insulin Glargine |
| PIOGLITAZONE | PIOGLITAZONE | 19099207 | pioglitazone |
| INSULIN | INSULIN | 19078554 | Regular Insulin, Human |
| INSULIN | INSULIN | 19078553 | insulin, isophane |
| REPAGLINIDE | REPAGLINIDE | 19023425 | repaglinide |
| INSULIN | INSULIN | 19078554 | Regular Insulin, Human |
| CHLORPROPAMIDE | CHLORPROPAMIDE | 19060470 | Chlorpropamide |
| INSULIN | INSULIN | 46233947 | Insulin Lispro |
| INSULIN | INSULIN | 1502910 | Insulin Glargine |
| METFORMIN | METFORMIN | 40164882 | Metformin |
| TOLAZAMIDE | TOLAZAMIDE | 1502817 | Tolazamide |
| GLIMEPIRIDE | GLIMEPIRIDE | 1597776 | glimepiride |
| INSULIN | INSULIN | 1586344 | insulin, isophane |
| CHLORPROPAMIDE | CHLORPROPAMIDE | 40030456 | Chlorpropamide |
| REPAGLINIDE | REPAGLINIDE | 36213643 | repaglinide |
| METFORMIN | METFORMIN | 40063352 | Metformin |
| LIRAGLUTIDE | LIRAGLUTIDE | 40169223 | liraglutide |
| INSULIN | INSULIN | 1516979 | insulin detemir |
| INSULIN | INSULIN | 1567198 | Insulin, Aspart, Human |
| INSULIN | INSULIN | 40184294 | Insulin, Aspart Protamine, Human |
| INSULIN | INSULIN | 19104092 | Regular Insulin, Human |
| INSULIN | INSULIN | 1596975 | insulin, isophane |
| GLIMEPIRIDE | GLIMEPIRIDE | 1597792 | glimepiride |
| INSULIN | INSULIN | 19094122 | Regular Insulin, Human |
| LINAGLIPTIN | LINAGLIPTIN | 42708089 | Linagliptin |
| DULAGLUTIDE | DULAGLUTIDE | 45774440 | dulaglutide |
| METFORMIN | METFORMIN | 40164932 | Metformin |
| METFORMIN | METFORMIN | 40164940 | Metformin |
| INSULIN | INSULIN | 1567198 | Insulin, Aspart, Human |

| | | | |
|---|---|---|---|
| INSULIN | INSULIN | 1513912 | Insulin Lispro |
| PRAMLINTIDE | PRAMLINTIDE | 40165687 | Pramlintide |
| GLIPIZIDE | GLIPIZIDE | 36241808 | Glipizide |
| INSULIN | INSULIN | 1502910 | Insulin Glargine |
| INSULIN | INSULIN | 1596972 | insulin, isophane |
| BROMOCRIPTINE | BROMOCRIPTINE | 36234233 | Bromocriptine |
| INSULIN | INSULIN | 19116453 | insulin, isophane |
| INSULIN | INSULIN | 35602731 | insulin degludec |
| INSULIN | INSULIN | 1567198 | Insulin, Aspart, Human |
| REPAGLINIDE | REPAGLINIDE | 1516771 | repaglinide |
| GLIMEPIRIDE | GLIMEPIRIDE | 1597757 | glimepiride |
| METFORMIN | METFORMIN | 36235747 | Metformin |
| INSULIN | INSULIN | 19062999 | insulin, isophane |
| INSULIN | INSULIN | 1544873 | Insulin, Glulisine, Human |
| INSULIN | INSULIN | 19100128 | Regular Insulin, Human |
| METFORMIN | METFORMIN | 40164897 | Metformin |
| INSULIN | INSULIN | 19120790 | Insulin Glargine |
| GLYBURIDE | GLYBURIDE | 1559684 | Glyburide |
| DAPAGLIFLOZIN | DAPAGLIFLOZIN | 44785834 | dapagliflozin |
| INSULIN | INSULIN | 1596956 | insulin, isophane |
| METFORMIN | METFORMIN | 40164932 | Metformin |
| PRAMLINTIDE | PRAMLINTIDE | 40165688 | Pramlintide |
| GLYBURIDE | GLYBURIDE | 36231170 | Glyburide |
| METFORMIN | METFORMIN | 40164898 | Metformin |
| INSULIN | INSULIN | 1502905 | Insulin Glargine |
| INSULIN | INSULIN | 19078607 | insulin, isophane |
| GLIMEPIRIDE | GLIMEPIRIDE | 36229116 | glimepiride |
| INSULIN | INSULIN | 1550023 | Insulin Lispro |
| MIGLITOL | MIGLITOL | 1510208 | miglitol |
| INSULIN | INSULIN | 19078553 | insulin, isophane |
| INSULIN | INSULIN | 1567232 | Insulin, Aspart, Human |
| GLYBURIDE | GLYBURIDE | 40168368 | Glyburide |
| METFORMIN | METFORMIN | 40063352 | Metformin |

| | | | |
|---|---|---|---|
| INSULIN | INSULIN | 19095211 | Insulin Lispro |
| METFORMIN | METFORMIN | 40171449 | Metformin |
| GLIPIZIDE | GLIPIZIDE | 40166815 | Glipizide |
| INSULIN | INSULIN | 19078553 | insulin, isophane |
| EXENATIDE | EXENATIDE | 1583722 | exenatide |
| METFORMIN | METFORMIN | 40164900 | Metformin |
| METFORMIN | METFORMIN | 40164929 | Metformin |
| INSULIN | INSULIN | 19104459 | insulin, isophane |
| INSULIN | INSULIN | 1516979 | insulin detemir |
| INSULIN | INSULIN | 1597000 | Regular Insulin, Human |
| GLYBURIDE | GLYBURIDE | 1559784 | Glyburide |
| INSULIN | INSULIN | 1502905 | Insulin Glargine |
| ALBIGLUTIDE | ALBIGLUTIDE | 44816283 | albiglutide |
| METFORMIN | METFORMIN | 40164944 | Metformin |
| METFORMIN | METFORMIN | 1503327 | Metformin |
| GLIMEPIRIDE | GLIMEPIRIDE | 1597756 | glimepiride |
| EXENATIDE | EXENATIDE | 36234709 | exenatide |
| METFORMIN | METFORMIN | 40164919 | Metformin |
| INSULIN | INSULIN | 46233946 | Insulin Lispro |
| GLYBURIDE | GLYBURIDE | 36233495 | Glyburide |
| GLIPIZIDE | GLIPIZIDE | 19134639 | Glipizide |
| EXENATIDE | EXENATIDE | 19083254 | exenatide |
| EXENATIDE | EXENATIDE | 36234709 | exenatide |
| INSULIN | INSULIN | 36231450 | Insulin, Aspart, Human |
| INSULIN | INSULIN | 1502910 | Insulin Glargine |
| CHLORPROPAMIDE | CHLORPROPAMIDE | 1594975 | Chlorpropamide |
| EXENATIDE | EXENATIDE | 46234118 | exenatide |
| LINAGLIPTIN | LINAGLIPTIN | 40239220 | Linagliptin |
| EXENATIDE | EXENATIDE | 19083253 | exenatide |
| METFORMIN | METFORMIN | 40164946 | Metformin |
| INSULIN | INSULIN | 1597012 | Regular Insulin, Human |
| INSULIN | INSULIN | 1596977 | Regular Insulin, Human |
| GLIPIZIDE | GLIPIZIDE | 19117945 | Glipizide |
| EXENATIDE | EXENATIDE | 19083253 | exenatide |

| | | | |
|---|---|---|---|
| METFORMIN | METFORMIN | 40164890 | Metformin |
| EXENATIDE | EXENATIDE | 46234118 | exenatide |
| INSULIN | INSULIN | 19078530 | insulin, isophane |
| INSULIN | INSULIN | 1597001 | Regular Insulin, Human |
| INSULIN | INSULIN | 19116453 | insulin, isophane |
| BROMOCRIPTINE | BROMOCRIPTINE | 40016898 | Bromocriptine |
| INSULIN | INSULIN | 19095218 | Regular Insulin, Human |
| INSULIN | INSULIN | 1550023 | Insulin Lispro |
| EMPAGLIFLOZIN | EMPAGLIFLOZIN | 45892176 | empagliflozin |
| LIRAGLUTIDE | LIRAGLUTIDE | 40169222 | liraglutide |
| LIRAGLUTIDE | LIRAGLUTIDE | 40169223 | liraglutide |
| DULAGLUTIDE | DULAGLUTIDE | 45774440 | dulaglutide |
| EXENATIDE | EXENATIDE | 19135404 | exenatide |
| INSULIN | INSULIN | 36233154 | Insulin, Glulisine, Human |
| GLIPIZIDE | GLIPIZIDE | 19081295 | Glipizide |
| SITAGLIPTIN | SITAGLIPTIN | 19125042 | sitagliptin |
| EXENATIDE | EXENATIDE | 42902527 | exenatide |
| GLIMEPIRIDE | GLIMEPIRIDE | 36234085 | glimepiride |
| GLYBURIDE | GLYBURIDE | 40168376 | Glyburide |
| GLIPIZIDE | GLIPIZIDE | 19081295 | Glipizide |
| GLIMEPIRIDE | GLIMEPIRIDE | 1597774 | glimepiride |
| INSULIN | INSULIN | 46234043 | Insulin, Aspart, Human |
| DAPAGLIFLOZIN | DAPAGLIFLOZIN | 44816049 | dapagliflozin |
| SAXAGLIPTIN | SAXAGLIPTIN | 40166044 | saxagliptin |
| INSULIN | INSULIN | 1596998 | Regular Insulin, Human |
| METFORMIN | METFORMIN | 1503328 | Metformin |
| METFORMIN | METFORMIN | 36221230 | Metformin |
| GLIPIZIDE | GLIPIZIDE | 1560171 | Glipizide |
| INSULIN | INSULIN | 1550027 | Insulin Lispro |
| METFORMIN | METFORMIN | 1503328 | Metformin |
| EXENATIDE | EXENATIDE | 36238252 | exenatide |
| BROMOCRIPTINE | BROMOCRIPTINE | 40164159 | Bromocriptine |
| INSULIN | INSULIN | 19100127 | insulin, isophane |
| GLICLAZIDE | GLICLAZIDE | 19102035 | Gliclazide |
| BROMOCRIPTINE | BROMOCRIPTINE | 19116170 | Bromocriptine |
| INSULIN | INSULIN | 19135276 | Insulin Glargine |
| INSULIN | INSULIN | 1596977 | Regular Insulin, Human |

| | | | |
|---|---|---|---|
| INSULIN | INSULIN | 46233975 | Insulin Lispro |
| INSULIN | INSULIN | 36224228 | insulin detemir |
| METFORMIN | METFORMIN | 40164929 | Metformin |
| INSULIN | INSULIN | 19112791 | Insulin, Glulisine, Human |
| INSULIN | INSULIN | 19110219 | insulin, isophane |
| INSULIN | INSULIN | 19078553 | insulin, isophane |
| LINAGLIPTIN | LINAGLIPTIN | 40239222 | Linagliptin |
| DULAGLUTIDE | DULAGLUTIDE | 45774440 | dulaglutide |
| INSULIN | INSULIN | 1550028 | Insulin Lispro |
| METFORMIN | METFORMIN | 40063352 | Metformin |
| INSULIN | INSULIN | 19078553 | insulin, isophane |
| METFORMIN | METFORMIN | 42708167 | Metformin |
| METFORMIN | METFORMIN | 1503297 | Metformin |
| DULAGLUTIDE | DULAGLUTIDE | 45774447 | dulaglutide |
| CANAGLIFLOZIN | CANAGLIFLOZIN | 45774717 | canagliflozin |
| LINAGLIPTIN | LINAGLIPTIN | 36249793 | Linagliptin |
| GLYBURIDE | GLYBURIDE | 40046211 | Glyburide |
| GLIPIZIDE | GLIPIZIDE | 19006910 | Glipizide |
| SITAGLIPTIN | SITAGLIPTIN | 19125042 | sitagliptin |
| GLIPIZIDE | GLIPIZIDE | 36235749 | Glipizide |
| INSULIN | INSULIN | 46233730 | Regular Insulin, Human |
| GLIPIZIDE | GLIPIZIDE | 1560171 | Glipizide |
| METFORMIN | METFORMIN | 40164923 | Metformin |
| GLYBURIDE | GLYBURIDE | 19077681 | Glyburide |
| EXENATIDE | EXENATIDE | 46234117 | exenatide |
| METFORMIN | METFORMIN | 36221230 | Metformin |
| INSULIN | INSULIN | 1502905 | Insulin Glargine |
| GLIMEPIRIDE | GLIMEPIRIDE | 43011830 | glimepiride |
| METFORMIN | METFORMIN | 40231396 | Metformin |
| METFORMIN | METFORMIN | 40164932 | Metformin |
| PRAMLINTIDE | PRAMLINTIDE | 40165692 | Pramlintide |
| GLIMEPIRIDE | GLIMEPIRIDE | 19018043 | glimepiride |
| INSULIN | INSULIN | 36231458 | insulin, isophane |

| | | | |
|---|---|---|---|
| INSULIN | INSULIN | 19071700 | Insulin Glargine |
| INSULIN | INSULIN | 19131582 | Insulin, Glulisine, Human |
| ALOGLIPTIN | ALOGLIPTIN | 43013924 | alogliptin |
| METFORMIN | METFORMIN | 40063354 | Metformin |
| GLYBURIDE | GLYBURIDE | 19106974 | Glyburide |
| DAPAGLIFLOZIN | DAPAGLIFLOZIN | 45775862 | dapagliflozin |
| BROMOCRIPTINE | BROMOCRIPTINE | 19083958 | Bromocriptine |
| DULAGLUTIDE | DULAGLUTIDE | 45774442 | dulaglutide |
| INSULIN | INSULIN | 19111797 | insulin, isophane |
| INSULIN | INSULIN | 1718604 | Insulin Glargine |
| GLIMEPIRIDE | GLIMEPIRIDE | 1597759 | glimepiride |
| INSULIN | INSULIN | 46233948 | Insulin Lispro |
| INSULIN | INSULIN | 46233945 | Insulin Lispro |
| ROSIGLITAZONE | ROSIGLITAZONE | 1547505 | rosiglitazone |
| GLIPIZIDE | GLIPIZIDE | 1560171 | Glipizide |
| INSULIN | INSULIN | 1567198 | Insulin, Aspart, Human |
| METFORMIN | METFORMIN | 1503297 | Metformin |
| METFORMIN | METFORMIN | 1503328 | Metformin |
| INSULIN | INSULIN | 1550028 | Insulin Lispro |
| METFORMIN | METFORMIN | 40164917 | Metformin |
| GLIMEPIRIDE | GLIMEPIRIDE | 36222784 | glimepiride |
| METFORMIN | METFORMIN | 40164894 | Metformin |
| PIOGLITAZONE | PIOGLITAZONE | 36236324 | pioglitazone |
| METFORMIN | METFORMIN | 1503297 | Metformin |
| INSULIN | INSULIN | 19110357 | Regular Insulin, Human |
| SAXAGLIPTIN | SAXAGLIPTIN | 40166044 | saxagliptin |
| INSULIN | INSULIN | 46234045 | Insulin, Aspart, Human |
| METFORMIN | METFORMIN | 40153151 | Metformin |
| METFORMIN | METFORMIN | 40164926 | Metformin |
| GLIPIZIDE | GLIPIZIDE | 1560233 | Glipizide |
| INSULIN | INSULIN | 36235937 | insulin human, rDNA origin |
| METFORMIN | METFORMIN | 40171451 | Metformin |
| EXENATIDE | EXENATIDE | 42708079 | exenatide |
| INSULIN | INSULIN | 1596918 | insulin, isophane |

| | | | |
|---|---|---|---|
| INSULIN | INSULIN | 19078553 | insulin, isophane |
| INSULIN | INSULIN | 46233730 | Regular Insulin, Human |
| METFORMIN | METFORMIN | 40164947 | Metformin |
| METFORMIN | METFORMIN | 40164914 | Metformin |
| INSULIN | INSULIN | 19078633 | Insulin, Regular, Pork |
| METFORMIN | METFORMIN | 40229047 | Metformin |
| INSULIN | INSULIN | 40176981 | Insulin, Aspart Protamine, Human |
| INSULIN | INSULIN | 46233972 | Insulin Lispro |
| GLYBURIDE | GLYBURIDE | 40166003 | Glyburide |
| INSULIN | INSULIN | 40166274 | Insulin Lispro |
| LINAGLIPTIN | LINAGLIPTIN | 42708089 | Linagliptin |
| GLYBURIDE | GLYBURIDE | 19077681 | Glyburide |
| REPAGLINIDE | REPAGLINIDE | 19054445 | repaglinide |
| METFORMIN | METFORMIN | 40164885 | Metformin |
| BROMOCRIPTINE | BROMOCRIPTINE | 19116170 | Bromocriptine |
| GLIMEPIRIDE | GLIMEPIRIDE | 1597799 | glimepiride |
| INSULIN | INSULIN | 1567200 | Insulin, Aspart, Human |
| DULAGLUTIDE | DULAGLUTIDE | 45774447 | dulaglutide |
| PRAMLINTIDE | PRAMLINTIDE | 40127005 | Pramlintide |
| SITAGLIPTIN | SITAGLIPTIN | 40243630 | sitagliptin |
| INSULIN | INSULIN | 1596977 | Regular Insulin, Human |
| EMPAGLIFLOZIN | EMPAGLIFLOZIN | 1592804 | empagliflozin |
| METFORMIN | METFORMIN | 42708167 | Metformin |
| INSULIN | INSULIN | 46234240 | insulin, isophane |
| INSULIN | INSULIN | 35602726 | insulin degludec |
| GLIMEPIRIDE | GLIMEPIRIDE | 19117198 | glimepiride |
| INSULIN | INSULIN | 1502910 | Insulin Glargine |
| METFORMIN | METFORMIN | 40063350 | Metformin |
| INSULIN | INSULIN | 19116455 | insulin, isophane |
| METFORMIN | METFORMIN | 40164948 | Metformin |

| | | | |
|---|---|---|---|
| EXENATIDE | EXENATIDE | 1583730 | exenatide |
| GLIMEPIRIDE | GLIMEPIRIDE | 43011498 | glimepiride |
| INSULIN | INSULIN | 36236032 | Insulin Lispro |
| INSULIN | INSULIN | 19078553 | insulin, isophane |
| INSULIN | INSULIN | 1596999 | Regular Insulin, Human |
| INSULIN | INSULIN | 40184294 | Insulin, Aspart Protamine, Human |
| GLYBURIDE | GLYBURIDE | 19065480 | Glyburide |
| INSULIN | INSULIN | 19078553 | insulin, isophane |
| GLYBURIDE | GLYBURIDE | 40166003 | Glyburide |
| CHLORPROPAMIDE | CHLORPROPAMIDE | 36231727 | Chlorpropamide |
| INSULIN | INSULIN | 1502910 | Insulin Glargine |
| GLYBURIDE | GLYBURIDE | 36233496 | Glyburide |
| INSULIN | INSULIN | 1513881 | Insulin Lispro |
| INSULIN | INSULIN | 19058398 | Insulin Lispro |
| INSULIN | INSULIN | 19078559 | Insulin Glargine |
| METFORMIN | METFORMIN | 1503297 | Metformin |
| METFORMIN | METFORMIN | 19113780 | Metformin |
| GLYBURIDE | GLYBURIDE | 1559782 | Glyburide |
| METFORMIN | METFORMIN | 40164900 | Metformin |
| INSULIN | INSULIN | 1502910 | Insulin Glargine |
| INSULIN | INSULIN | 35602724 | insulin degludec |
| LINAGLIPTIN | LINAGLIPTIN | 36249798 | Linagliptin |
| DAPAGLIFLOZIN | DAPAGLIFLOZIN | 44815995 | dapagliflozin |
| GLICLAZIDE | GLICLAZIDE | 19059797 | Gliclazide |
| METFORMIN | METFORMIN | 40164891 | Metformin |
| GLIMEPIRIDE | GLIMEPIRIDE | 19117198 | glimepiride |
| INSULIN | INSULIN | 19009384 | insulin, isophane |
| CANAGLIFLOZIN | CANAGLIFLOZIN | 36246430 | canagliflozin |
| INSULIN | INSULIN | 1596941 | insulin, isophane |
| DULAGLUTIDE | DULAGLUTIDE | 45774447 | dulaglutide |
| EXENATIDE | EXENATIDE | 36234709 | exenatide |

| | | | |
|---|---|---|---|
| DAPAGLIFLOZIN | DAPAGLIFLOZIN | 36247908 | dapagliflozin |
| EMPAGLIFLOZIN | EMPAGLIFLOZIN | 45892176 | empagliflozin |
| METFORMIN | METFORMIN | 40164928 | Metformin |
| NATEGLINIDE | NATEGLINIDE | 36232423 | nateglinide |
| EMPAGLIFLOZIN | EMPAGLIFLOZIN | 1592800 | empagliflozin |
| GLIPIZIDE | GLIPIZIDE | 19047638 | Glipizide |
| INSULIN | INSULIN | 19116559 | insulin, isophane |
| INSULIN | INSULIN | 19110219 | insulin, isophane |
| GLIMEPIRIDE | GLIMEPIRIDE | 19100593 | glimepiride |
| INSULIN | INSULIN | 19078553 | insulin, isophane |
| EXENATIDE | EXENATIDE | 19083253 | exenatide |
| DULAGLUTIDE | DULAGLUTIDE | 45774446 | dulaglutide |
| ACARBOSE | ACARBOSE | 1529360 | Acarbose |
| GLIMEPIRIDE | GLIMEPIRIDE | 1597772 | glimepiride |
| EXENATIDE | EXENATIDE | 19083254 | exenatide |
| METFORMIN | METFORMIN | 1503327 | Metformin |
| INSULIN | INSULIN | 42902743 | Insulin Glargine |
| METFORMIN | METFORMIN | 42705990 | Metformin |
| GLIMEPIRIDE | GLIMEPIRIDE | 19117198 | glimepiride |
| METFORMIN | METFORMIN | 40164896 | Metformin |
| GLIMEPIRIDE | GLIMEPIRIDE | 1597775 | glimepiride |
| GLYBURIDE | GLYBURIDE | 1559685 | Glyburide |
| GLIMEPIRIDE | GLIMEPIRIDE | 1597776 | glimepiride |
| SAXAGLIPTIN | SAXAGLIPTIN | 40166038 | saxagliptin |
| INSULIN | INSULIN | 19078555 | Regular Insulin, Human |
| INSULIN | INSULIN | 1513849 | Insulin, Zinc, Human |
| INSULIN | INSULIN | 19062999 | insulin, isophane |
| INSULIN | INSULIN | 40051347 | Insulin Glargine |
| GLIMEPIRIDE | GLIMEPIRIDE | 19122367 | glimepiride |
| METFORMIN | METFORMIN | 36236322 | Metformin |
| EXENATIDE | EXENATIDE | 46234118 | exenatide |
| INSULIN | INSULIN | 46233728 | Regular Insulin, Human |

| | | | |
|---|---|---|---|
| INSULIN | INSULIN | 19078551 | insulin, isophane |
| INSULIN | INSULIN | 1550023 | Insulin Lispro |
| INSULIN | INSULIN | 19135265 | Regular Insulin, Human |
| LIRAGLUTIDE | LIRAGLUTIDE | 40169223 | liraglutide |
| SITAGLIPTIN | SITAGLIPTIN | 19125046 | sitagliptin |
| BROMOCRIPTINE | BROMOCRIPTINE | 730579 | Bromocriptine |
| GLIMEPIRIDE | GLIMEPIRIDE | 1597775 | glimepiride |
| INSULIN | INSULIN | 1586370 | Ultralente Insulin, Human |
| INSULIN | INSULIN | 46234043 | Insulin, Aspart, Human |
| INSULIN | INSULIN | 19120790 | Insulin Glargine |
| INSULIN | INSULIN | 35602724 | insulin degludec |
| GLIPIZIDE | GLIPIZIDE | 19047638 | Glipizide |
| ROSIGLITAZONE | ROSIGLITAZONE | 40099633 | rosiglitazone |
| INSULIN | INSULIN | 19123376 | Insulin Lispro |
| INSULIN | INSULIN | 1544838 | Insulin, Glulisine, Human |
| INSULIN | INSULIN | 1597000 | Regular Insulin, Human |
| INSULIN | INSULIN | 1562586 | Insulin, Zinc, Pork |
| METFORMIN | METFORMIN | 1503297 | Metformin |
| GLYBURIDE | GLYBURIDE | 1559782 | Glyburide |
| METFORMIN | METFORMIN | 40229050 | Metformin |
| LIRAGLUTIDE | LIRAGLUTIDE | 45892684 | liraglutide |
| GLIMEPIRIDE | GLIMEPIRIDE | 40159908 | glimepiride |
| ACARBOSE | ACARBOSE | 1529331 | Acarbose |
| NATEGLINIDE | NATEGLINIDE | 40058199 | nateglinide |
| EXENATIDE | EXENATIDE | 19083254 | exenatide |
| INSULIN | INSULIN | 1567198 | Insulin, Aspart, Human |
| INSULIN | INSULIN | 19009384 | insulin, isophane |
| INSULIN | INSULIN | 19078633 | Insulin, Regular, Pork |
| NATEGLINIDE | NATEGLINIDE | 36232423 | nateglinide |
| METFORMIN | METFORMIN | 40171450 | Metformin |
| REPAGLINIDE | REPAGLINIDE | 19023425 | repaglinide |
| INSULIN | INSULIN | 19078555 | Regular Insulin, Human |
| INSULIN | INSULIN | 1502910 | Insulin Glargine |
| INSULIN | INSULIN | 19100127 | insulin, isophane |
| GLIPIZIDE | GLIPIZIDE | 1560171 | Glipizide |

| | | | |
|---|---|---|---|
| NATEGLINIDE | NATEGLINIDE | 1502826 | nateglinide |
| INSULIN | INSULIN | 40051348 | Insulin Glargine |
| INSULIN | INSULIN | 36229086 | Insulin Glargine |
| SITAGLIPTIN | SITAGLIPTIN | 19125042 | sitagliptin |
| GLIPIZIDE | GLIPIZIDE | 40167636 | Glipizide |
| LIRAGLUTIDE | LIRAGLUTIDE | 36241903 | liraglutide |
| METFORMIN | METFORMIN | 40164932 | Metformin |
| BROMOCRIPTINE | BROMOCRIPTINE | 40164161 | Bromocriptine |
| METFORMIN | METFORMIN | 40164929 | Metformin |
| PRAMLINTIDE | PRAMLINTIDE | 40165692 | Pramlintide |
| METFORMIN | METFORMIN | 40164944 | Metformin |
| METFORMIN | METFORMIN | 40164933 | Metformin |
| INSULIN | INSULIN | 19062999 | insulin, isophane |
| DAPAGLIFLOZIN | DAPAGLIFLOZIN | 44785473 | dapagliflozin |
| INSULIN | INSULIN | 19078553 | insulin, isophane |
| METFORMIN | METFORMIN | 40063354 | Metformin |
| INSULIN | INSULIN | 1502905 | Insulin Glargine |
| INSULIN | INSULIN | 40166274 | Insulin Lispro |
| INSULIN | INSULIN | 19135320 | Regular Insulin, Human |
| INSULIN | INSULIN | 1596977 | Regular Insulin, Human |
| METFORMIN | METFORMIN | 1503297 | Metformin |
| METFORMIN | METFORMIN | 40164894 | Metformin |
| DAPAGLIFLOZIN | DAPAGLIFLOZIN | 36247907 | dapagliflozin |
| GLIPIZIDE | GLIPIZIDE | 1560171 | Glipizide |
| GLYBURIDE | GLYBURIDE | 19077684 | Glyburide |
| INSULIN | INSULIN | 1550028 | Insulin Lispro |
| LIRAGLUTIDE | LIRAGLUTIDE | 40170911 | liraglutide |
| REPAGLINIDE | REPAGLINIDE | 19023426 | repaglinide |
| INSULIN | INSULIN | 1513881 | Insulin Lispro |
| INSULIN | INSULIN | 36248870 | Insulin Glargine |
| INSULIN | INSULIN | 1567232 | Insulin, Aspart, Human |
| ROSIGLITAZONE | ROSIGLITAZONE | 36229365 | rosiglitazone |
| INSULIN | INSULIN | 46234043 | Insulin, Aspart, Human |
| METFORMIN | METFORMIN | 1503327 | Metformin |
| TOLBUTAMIDE | TOLBUTAMIDE | 1502856 | Tolbutamide |

| | | | |
|---|---|---|---|
| EXENATIDE | EXENATIDE | 1583722 | exenatide |
| CANAGLIFLOZIN | CANAGLIFLOZIN | 43526469 | canagliflozin |
| INSULIN | INSULIN | 19061670 | Regular Insulin, Human |
| ALOGLIPTIN | ALOGLIPTIN | 43013912 | alogliptin |
| INSULIN | INSULIN | 1596916 | insulin, isophane |
| INSULIN | INSULIN | 1513916 | Insulin Lispro |
| SITAGLIPTIN | SITAGLIPTIN | 40136769 | sitagliptin |
| EMPAGLIFLOZIN | EMPAGLIFLOZIN | 45774751 | empagliflozin |
| INSULIN | INSULIN | 19062999 | insulin, isophane |
| SITAGLIPTIN | SITAGLIPTIN | 19125045 | sitagliptin |
| INSULIN | INSULIN | 19111797 | insulin, isophane |
| LINAGLIPTIN | LINAGLIPTIN | 36249798 | Linagliptin |
| ROSIGLITAZONE | ROSIGLITAZONE | 1547506 | rosiglitazone |
| INSULIN | INSULIN | 19110357 | Regular Insulin, Human |
| ACARBOSE | ACARBOSE | 19047612 | Acarbose |
| INSULIN | INSULIN | 1516978 | insulin detemir |
| METFORMIN | METFORMIN | 42705990 | Metformin |
| METFORMIN | METFORMIN | 40163928 | Metformin |
| METFORMIN | METFORMIN | 36223143 | Metformin |
| METFORMIN | METFORMIN | 40229050 | Metformin |
| REPAGLINIDE | REPAGLINIDE | 19120529 | repaglinide |
| GLYBURIDE | GLYBURIDE | 1559684 | Glyburide |
| INSULIN | INSULIN | 1567198 | Insulin, Aspart, Human |
| INSULIN | INSULIN | 46233976 | Insulin Lispro |
| CANAGLIFLOZIN | CANAGLIFLOZIN | 45774717 | canagliflozin |
| SITAGLIPTIN | SITAGLIPTIN | 19125049 | sitagliptin |
| CANAGLIFLOZIN | CANAGLIFLOZIN | 45774717 | canagliflozin |
| INSULIN | INSULIN | 1562586 | Insulin, Zinc, Pork |
| DULAGLUTIDE | DULAGLUTIDE | 45774442 | dulaglutide |
| TOLBUTAMIDE | TOLBUTAMIDE | 19037213 | Tolbutamide |

| | | | |
|---|---|---|---|
| INSULIN | INSULIN | 1596972 | insulin, isophane |
| REPAGLINIDE | REPAGLINIDE | 36213643 | repaglinide |
| METFORMIN | METFORMIN | 40164939 | Metformin |
| INSULIN | INSULIN | 35602731 | insulin degludec |
| INSULIN | INSULIN | 19111797 | insulin, isophane |
| BROMOCRIPTINE | BROMOCRIPTINE | 730572 | Bromocriptine |
| INSULIN | INSULIN | 19110357 | Regular Insulin, Human |
| INSULIN | INSULIN | 1596977 | Regular Insulin, Human |
| GLIMEPIRIDE | GLIMEPIRIDE | 19122367 | glimepiride |
| INSULIN | INSULIN | 19131582 | Insulin, Glulisine, Human |
| INSULIN | INSULIN | 1596916 | insulin, isophane |
| GLYBURIDE | GLYBURIDE | 1559684 | Glyburide |
| METFORMIN | METFORMIN | 1503328 | Metformin |
| ACARBOSE | ACARBOSE | 36242198 | Acarbose |
| EMPAGLIFLOZIN | EMPAGLIFLOZIN | 36248376 | empagliflozin |
| ALBIGLUTIDE | ALBIGLUTIDE | 44816083 | albiglutide |
| INSULIN | INSULIN | 46234241 | Regular Insulin, Human |
| REPAGLINIDE | REPAGLINIDE | 40098707 | repaglinide |
| CANAGLIFLOZIN | CANAGLIFLOZIN | 43526475 | canagliflozin |
| INSULIN | INSULIN | 40166274 | Insulin Lispro |
| INSULIN | INSULIN | 19078527 | Insulin, Zinc, Human |
| GLIMEPIRIDE | GLIMEPIRIDE | 1597776 | glimepiride |
| METFORMIN | METFORMIN | 1503327 | Metformin |
| GLIPIZIDE | GLIPIZIDE | 40165975 | Glipizide |
| INSULIN | INSULIN | 1550028 | Insulin Lispro |
| LIRAGLUTIDE | LIRAGLUTIDE | 40170912 | liraglutide |
| INSULIN | INSULIN | 36236032 | Insulin Lispro |
| GLIMEPIRIDE | GLIMEPIRIDE | 1597756 | glimepiride |
| DAPAGLIFLOZIN | DAPAGLIFLOZIN | 36248557 | dapagliflozin |
| INSULIN | INSULIN | 19078607 | insulin, isophane |
| ALOGLIPTIN | ALOGLIPTIN | 43013929 | alogliptin |
| INSULIN | INSULIN | 46233946 | Insulin Lispro |
| INSULIN | INSULIN | 36235841 | Regular Insulin, Human |
| INSULIN | INSULIN | 46221555 | Insulin Glargine |
| DULAGLUTIDE | DULAGLUTIDE | 45774440 | dulaglutide |
| GLIPIZIDE | GLIPIZIDE | 19085004 | Glipizide |

| | | | |
|---|---|---|---|
| INSULIN | INSULIN | 19044971 | insulin detemir |
| INSULIN | INSULIN | 1550027 | Insulin Lispro |
| INSULIN | INSULIN | 1550023 | Insulin Lispro |
| CANAGLIFLOZIN | CANAGLIFLOZIN | 43526468 | canagliflozin |
| INSULIN | INSULIN | 1586328 | insulin, isophane |
| ALBIGLUTIDE | ALBIGLUTIDE | 44816274 | albiglutide |
| METFORMIN | METFORMIN | 40063354 | Metformin |
| INSULIN | INSULIN | 1596977 | Regular Insulin, Human |
| DULAGLUTIDE | DULAGLUTIDE | 45774445 | dulaglutide |
| EXENATIDE | EXENATIDE | 19083254 | exenatide |
| INSULIN | INSULIN | 1513916 | Insulin Lispro |
| GLIPIZIDE | GLIPIZIDE | 1560171 | Glipizide |
| GLIPIZIDE | GLIPIZIDE | 36218767 | Glipizide |
| METFORMIN | METFORMIN | 40231392 | Metformin |
| EXENATIDE | EXENATIDE | 19083253 | exenatide |
| GLIMEPIRIDE | GLIMEPIRIDE | 1597760 | glimepiride |
| INSULIN | INSULIN | 35602728 | insulin degludec |
| METFORMIN | METFORMIN | 1503327 | Metformin |
| INSULIN | INSULIN | 19009384 | insulin, isophane |
| ALBIGLUTIDE | ALBIGLUTIDE | 44816283 | albiglutide |
| INSULIN | INSULIN | 1502905 | Insulin Glargine |
| INSULIN | INSULIN | 19078633 | Insulin, Regular, Pork |
| GLIMEPIRIDE | GLIMEPIRIDE | 1597775 | glimepiride |
| GLYBURIDE | GLYBURIDE | 1559684 | Glyburide |
| INSULIN | INSULIN | 19120315 | insulin, isophane |
| INSULIN | INSULIN | 36236032 | Insulin Lispro |
| INSULIN | INSULIN | 19062999 | insulin, isophane |
| LIRAGLUTIDE | LIRAGLUTIDE | 45892686 | liraglutide |
| BROMOCRIPTINE | BROMOCRIPTINE | 730548 | Bromocriptine |
| INSULIN | INSULIN | 19061670 | Regular Insulin, Human |
| INSULIN | INSULIN | 1502905 | Insulin Glargine |
| INSULIN | INSULIN | 1550023 | Insulin Lispro |
| INSULIN | INSULIN | 19010204 | insulin, isophane |
| INSULIN | INSULIN | 19110352 | insulin, isophane |

| | | | |
|---|---|---|---|
| INSULIN | INSULIN | 1513881 | Insulin Lispro |
| BROMOCRIPTINE | BROMOCRIPTINE | 730572 | Bromocriptine |
| METFORMIN | METFORMIN | 40163923 | Metformin |
| METFORMIN | METFORMIN | 40063352 | Metformin |
| GLIPIZIDE | GLIPIZIDE | 40165972 | Glipizide |
| METFORMIN | METFORMIN | 40164905 | Metformin |
| GLIPIZIDE | GLIPIZIDE | 19079990 | Glipizide |
| EXENATIDE | EXENATIDE | 1583730 | exenatide |
| INSULIN | INSULIN | 36231456 | insulin, isophane |
| METFORMIN | METFORMIN | 36235746 | Metformin |
| EXENATIDE | EXENATIDE | 1583728 | exenatide |
| INSULIN | INSULIN | 1567198 | Insulin, Aspart, Human |
| GLIPIZIDE | GLIPIZIDE | 1560171 | Glipizide |
| DULAGLUTIDE | DULAGLUTIDE | 45774447 | dulaglutide |
| TOLAZAMIDE | TOLAZAMIDE | 19082961 | Tolazamide |
| INSULIN | INSULIN | 19018822 | Insulin Lispro |
| INSULIN | INSULIN | 1550028 | Insulin Lispro |
| INSULIN | INSULIN | 1531601 | Insulin, Aspart Protamine, Human |
| METFORMIN | METFORMIN | 40171451 | Metformin |
| INSULIN | INSULIN | 36231450 | Insulin, Aspart, Human |
| INSULIN | INSULIN | 1516976 | insulin detemir |
| INSULIN | INSULIN | 1596941 | insulin, isophane |
| METFORMIN | METFORMIN | 40164942 | Metformin |
| GLIPIZIDE | GLIPIZIDE | 40167636 | Glipizide |
| PIOGLITAZONE | PIOGLITAZONE | 1525221 | pioglitazone |
| METFORMIN | METFORMIN | 40063350 | Metformin |
| CANAGLIFLOZIN | CANAGLIFLOZIN | 45774714 | canagliflozin |
| INSULIN | INSULIN | 1596999 | Regular Insulin, Human |
| DAPAGLIFLOZIN | DAPAGLIFLOZIN | 45775862 | dapagliflozin |
| INSULIN | INSULIN | 1567200 | Insulin, Aspart, Human |
| INSULIN | INSULIN | 19009384 | insulin, isophane |
| INSULIN | INSULIN | 1516978 | insulin detemir |

| | | | |
|---|---|---|---|
| INSULIN | INSULIN | 40051348 | Insulin Glargine |
| GLIMEPIRIDE | GLIMEPIRIDE | 1597756 | glimepiride |
| SAXAGLIPTIN | SAXAGLIPTIN | 40166042 | saxagliptin |
| INSULIN | INSULIN | 19120315 | insulin, isophane |
| GLIPIZIDE | GLIPIZIDE | 1560171 | Glipizide |
| INSULIN | INSULIN | 19120315 | insulin, isophane |
| INSULIN | INSULIN | 1596919 | insulin, isophane |
| GLYBURIDE | GLYBURIDE | 40165995 | Glyburide |
| BROMOCRIPTINE | BROMOCRIPTINE | 19062900 | Bromocriptine |
| INSULIN | INSULIN | 19071700 | Insulin Glargine |
| INSULIN | INSULIN | 19135265 | Regular Insulin, Human |
| INSULIN | INSULIN | 19078557 | Regular Insulin, Human |
| INSULIN | INSULIN | 36229086 | Insulin Glargine |
| INSULIN | INSULIN | 19080091 | Ultralente Insulin, Human |
| EXENATIDE | EXENATIDE | 19083253 | exenatide |
| INSULIN | INSULIN | 19131582 | Insulin, Glulisine, Human |
| LIRAGLUTIDE | LIRAGLUTIDE | 40170913 | liraglutide |
| METFORMIN | METFORMIN | 40164895 | Metformin |
| METFORMIN | METFORMIN | 40171449 | Metformin |
| EXENATIDE | EXENATIDE | 19135404 | exenatide |
| INSULIN | INSULIN | 1596955 | insulin, isophane |
| INSULIN | INSULIN | 19120315 | insulin, isophane |
| METFORMIN | METFORMIN | 36236481 | Metformin |
| PIOGLITAZONE | PIOGLITAZONE | 1525215 | pioglitazone |
| EMPAGLIFLOZIN | EMPAGLIFLOZIN | 45892177 | empagliflozin |
| BROMOCRIPTINE | BROMOCRIPTINE | 40164161 | Bromocriptine |
| METFORMIN | METFORMIN | 1503328 | Metformin |
| INSULIN | INSULIN | 19078603 | insulin, isophane |

| | | | |
|---|---|---|---|
| PIOGLITAZONE | PIOGLITAZONE | 1525215 | pioglitazone |
| INSULIN | INSULIN | 19094122 | Regular Insulin, Human |
| INSULIN | INSULIN | 1567198 | Insulin, Aspart, Human |
| SITAGLIPTIN | SITAGLIPTIN | 40243633 | sitagliptin |
| INSULIN | INSULIN | 19082658 | Insulin, Zinc, Pork |
| METFORMIN | METFORMIN | 40153151 | Metformin |
| ACARBOSE | ACARBOSE | 40003412 | Acarbose |
| INSULIN | INSULIN | 42899447 | Insulin, Pork |
| EXENATIDE | EXENATIDE | 46234117 | exenatide |
| INSULIN | INSULIN | 19120315 | insulin, isophane |
| EMPAGLIFLOZIN | EMPAGLIFLOZIN | 45774758 | empagliflozin |
| METFORMIN | METFORMIN | 40164880 | Metformin |
| INSULIN | INSULIN | 19110355 | insulin, isophane |
| INSULIN | INSULIN | 1718889 | Regular Insulin, Human |
| INSULIN | INSULIN | 36236032 | Insulin Lispro |
| PIOGLITAZONE | PIOGLITAZONE | 19099208 | pioglitazone |
| INSULIN | INSULIN | 1550023 | Insulin Lispro |
| INSULIN | INSULIN | 19110357 | Regular Insulin, Human |
| CANAGLIFLOZIN | CANAGLIFLOZIN | 43526465 | canagliflozin |
| DULAGLUTIDE | DULAGLUTIDE | 45774442 | dulaglutide |
| LIRAGLUTIDE | LIRAGLUTIDE | 40170913 | liraglutide |
| METFORMIN | METFORMIN | 40153151 | Metformin |
| METFORMIN | METFORMIN | 40163926 | Metformin |
| LINAGLIPTIN | LINAGLIPTIN | 42708087 | Linagliptin |
| CANAGLIFLOZIN | CANAGLIFLOZIN | 45774713 | canagliflozin |
| INSULIN | INSULIN | 19116559 | insulin, isophane |
| INSULIN | INSULIN | 46233730 | Regular Insulin, Human |
| INSULIN | INSULIN | 19110219 | insulin, isophane |
| BROMOCRIPTINE | BROMOCRIPTINE | 730580 | Bromocriptine |
| GLIMEPIRIDE | GLIMEPIRIDE | 43011831 | glimepiride |
| ROSIGLITAZONE | ROSIGLITAZONE | 19079465 | rosiglitazone |
| DAPAGLIFLOZIN | DAPAGLIFLOZIN | 44785833 | dapagliflozin |

| | | | |
|---|---|---|---|
| INSULIN | INSULIN | 40052771 | Regular Insulin, Human |
| PIOGLITAZONE | PIOGLITAZONE | 19099209 | pioglitazone |
| GLIPIZIDE | GLIPIZIDE | 19077638 | Glipizide |
| GLIMEPIRIDE | GLIMEPIRIDE | 19117198 | glimepiride |
| INSULIN | INSULIN | 1596999 | Regular Insulin, Human |
| INSULIN | INSULIN | 1550028 | Insulin Lispro |
| INSULIN | INSULIN | 1513849 | Insulin, Zinc, Human |
| INSULIN | INSULIN | 19012679 | Regular Insulin, Human |
| CHLORPROPAMIDE | CHLORPROPAMIDE | 19025944 | Chlorpropamide |
| INSULIN | INSULIN | 19120790 | Insulin Glargine |
| LIRAGLUTIDE | LIRAGLUTIDE | 40169223 | liraglutide |
| EMPAGLIFLOZIN | EMPAGLIFLOZIN | 45774754 | empagliflozin |
| INSULIN | INSULIN | 35602728 | insulin degludec |
| INSULIN | INSULIN | 1597012 | Regular Insulin, Human |
| GLYBURIDE | GLYBURIDE | 40168368 | Glyburide |
| GLIMEPIRIDE | GLIMEPIRIDE | 1597756 | glimepiride |
| INSULIN | INSULIN | 35602731 | insulin degludec |
| ALOGLIPTIN | ALOGLIPTIN | 43013910 | alogliptin |
| INSULIN | INSULIN | 1550028 | Insulin Lispro |
| INSULIN | INSULIN | 35602731 | insulin degludec |
| DULAGLUTIDE | DULAGLUTIDE | 36247186 | dulaglutide |
| DAPAGLIFLOZIN | DAPAGLIFLOZIN | 45775999 | dapagliflozin |
| INSULIN | INSULIN | 46233975 | Insulin Lispro |
| METFORMIN | METFORMIN | 1503297 | Metformin |
| GLYBURIDE | GLYBURIDE | 1559684 | Glyburide |
| INSULIN | INSULIN | 19104458 | insulin, isophane |
| INSULIN | INSULIN | 1596999 | Regular Insulin, Human |
| ROSIGLITAZONE | ROSIGLITAZONE | 1547508 | rosiglitazone |
| INSULIN | INSULIN | 40051347 | Insulin Glargine |
| EXENATIDE | EXENATIDE | 1583727 | exenatide |
| EXENATIDE | EXENATIDE | 1583722 | exenatide |
| GLIPIZIDE | GLIPIZIDE | 19081295 | Glipizide |
| INSULIN | INSULIN | 19078553 | insulin, isophane |

| | | | |
|---|---|---|---|
| INSULIN | INSULIN | 1596999 | Regular Insulin, Human |
| INSULIN | INSULIN | 46233943 | Insulin Lispro |
| INSULIN | INSULIN | 1567232 | Insulin, Aspart, Human |
| INSULIN | INSULIN | 46233730 | Regular Insulin, Human |
| INSULIN | INSULIN | 1562586 | Insulin, Zinc, Pork |
| INSULIN | INSULIN | 19061670 | Regular Insulin, Human |
| REPAGLINIDE | REPAGLINIDE | 1516771 | repaglinide |
| INSULIN | INSULIN | 1567200 | Insulin, Aspart, Human |
| CANAGLIFLOZIN | CANAGLIFLOZIN | 36246431 | canagliflozin |
| EXENATIDE | EXENATIDE | 19083254 | exenatide |
| EXENATIDE | EXENATIDE | 1583728 | exenatide |
| INSULIN | INSULIN | 19110352 | insulin, isophane |
| INSULIN | INSULIN | 1596977 | Regular Insulin, Human |
| INSULIN | INSULIN | 46233972 | Insulin Lispro |
| GLIPIZIDE | GLIPIZIDE | 1560171 | Glipizide |
| METFORMIN | METFORMIN | 40164902 | Metformin |
| INSULIN | INSULIN | 46234049 | Insulin, Aspart, Human |
| GLIMEPIRIDE | GLIMEPIRIDE | 1597756 | glimepiride |
| LIRAGLUTIDE | LIRAGLUTIDE | 40169223 | liraglutide |
| INSULIN | INSULIN | 19135264 | Regular Insulin, Human |
| GLIPIZIDE | GLIPIZIDE | 1560171 | Glipizide |
| INSULIN | INSULIN | 1586344 | insulin, isophane |
| INSULIN | INSULIN | 19061670 | Regular Insulin, Human |
| INSULIN | INSULIN | 19100127 | insulin, isophane |
| MIGLITOL | MIGLITOL | 19081706 | miglitol |
| INSULIN | INSULIN | 19120315 | insulin, isophane |
| METFORMIN | METFORMIN | 40164926 | Metformin |
| INSULIN | INSULIN | 46233972 | Insulin Lispro |
| INSULIN | INSULIN | 40166274 | Insulin Lispro |
| INSULIN | INSULIN | 19094122 | Regular Insulin, Human |
| METFORMIN | METFORMIN | 36239257 | Metformin |
| LIRAGLUTIDE | LIRAGLUTIDE | 40170911 | liraglutide |
| GLIPIZIDE | GLIPIZIDE | 19085004 | Glipizide |
| INSULIN | INSULIN | 1596955 | insulin, isophane |
| METFORMIN | METFORMIN | 1503297 | Metformin |
| INSULIN | INSULIN | 1597012 | Regular Insulin, Human |

| | | | |
|---|---|---|---|
| INSULIN | INSULIN | 46233975 | Insulin Lispro |
| METFORMIN | METFORMIN | 40164881 | Metformin |
| NATEGLINIDE | NATEGLINIDE | 36232423 | nateglinide |
| METFORMIN | METFORMIN | 40163928 | Metformin |
| SITAGLIPTIN | SITAGLIPTIN | 19125043 | sitagliptin |
| EMPAGLIFLOZIN | EMPAGLIFLOZIN | 45774757 | empagliflozin |
| INSULIN | INSULIN | 19062999 | insulin, isophane |
| GLIMEPIRIDE | GLIMEPIRIDE | 1597775 | glimepiride |
| GLIMEPIRIDE | GLIMEPIRIDE | 36214984 | glimepiride |
| SITAGLIPTIN | SITAGLIPTIN | 1580747 | sitagliptin |
| INSULIN | INSULIN | 1502910 | Insulin Glargine |
| INSULIN | INSULIN | 35602731 | insulin degludec |
| GLIPIZIDE | GLIPIZIDE | 1560233 | Glipizide |
| INSULIN | INSULIN | 35602731 | insulin degludec |
| LIRAGLUTIDE | LIRAGLUTIDE | 40169223 | liraglutide |
| PRAMLINTIDE | PRAMLINTIDE | 36243018 | Pramlintide |
| INSULIN | INSULIN | 40176981 | Insulin, Aspart Protamine, Human |
| GLYBURIDE | GLYBURIDE | 19110153 | Glyburide |
| METFORMIN | METFORMIN | 42708167 | Metformin |
| INSULIN | INSULIN | 19120790 | Insulin Glargine |
| GLYBURIDE | GLYBURIDE | 1559684 | Glyburide |
| LINAGLIPTIN | LINAGLIPTIN | 36249793 | Linagliptin |
| METFORMIN | METFORMIN | 40164938 | Metformin |
| INSULIN | INSULIN | 1550027 | Insulin Lispro |
| INSULIN | INSULIN | 35602724 | insulin degludec |
| LINAGLIPTIN | LINAGLIPTIN | 40239222 | Linagliptin |
| METFORMIN | METFORMIN | 1503327 | Metformin |
| METFORMIN | METFORMIN | 40171451 | Metformin |
| INSULIN | INSULIN | 1596977 | Regular Insulin, Human |
| BROMOCRIPTINE | BROMOCRIPTINE | 19103828 | Bromocriptine |
| BROMOCRIPTINE | BROMOCRIPTINE | 40016899 | Bromocriptine |
| INSULIN | INSULIN | 1550027 | Insulin Lispro |
| METFORMIN | METFORMIN | 40173227 | Metformin |

| | | | |
|---|---|---|---|
| INSULIN | INSULIN | 40051347 | Insulin Glargine |
| INSULIN | INSULIN | 1567232 | Insulin, Aspart, Human |
| METFORMIN | METFORMIN | 1503328 | Metformin |
| INSULIN | INSULIN | 1513914 | Insulin Lispro |
| INSULIN | INSULIN | 19095211 | Insulin Lispro |
| GLIMEPIRIDE | GLIMEPIRIDE | 19018043 | glimepiride |
| ACARBOSE | ACARBOSE | 19023063 | Acarbose |
| GLIMEPIRIDE | GLIMEPIRIDE | 1597781 | glimepiride |
| INSULIN | INSULIN | 1567198 | Insulin, Aspart, Human |
| ROSIGLITAZONE | ROSIGLITAZONE | 36227624 | rosiglitazone |
| METFORMIN | METFORMIN | 40164905 | Metformin |
| BROMOCRIPTINE | BROMOCRIPTINE | 19007101 | Bromocriptine |
| ALOGLIPTIN | ALOGLIPTIN | 43013897 | alogliptin |
| BROMOCRIPTINE | BROMOCRIPTINE | 19062900 | Bromocriptine |
| SITAGLIPTIN | SITAGLIPTIN | 19125042 | sitagliptin |
| EXENATIDE | EXENATIDE | 42708079 | exenatide |
| GLIPIZIDE | GLIPIZIDE | 40166814 | Glipizide |
| GLIPIZIDE | GLIPIZIDE | 19077638 | Glipizide |
| METFORMIN | METFORMIN | 1503328 | Metformin |
| GLIPIZIDE | GLIPIZIDE | 40044224 | Glipizide |
| INSULIN | INSULIN | 19123376 | Insulin Lispro |
| GLYBURIDE | GLYBURIDE | 19001408 | Glyburide |
| INSULIN | INSULIN | 19078553 | insulin, isophane |
| INSULIN | INSULIN | 1513916 | Insulin Lispro |
| INSULIN | INSULIN | 36249657 | Regular Insulin, Human |
| NATEGLINIDE | NATEGLINIDE | 1502826 | nateglinide |
| INSULIN | INSULIN | 19110357 | Regular Insulin, Human |
| ALBIGLUTIDE | ALBIGLUTIDE | 44816283 | albiglutide |
| INSULIN | INSULIN | 40051347 | Insulin Glargine |
| METFORMIN | METFORMIN | 40164895 | Metformin |

| | | | |
|---|---|---|---|
| METFORMIN | METFORMIN | 40163923 | Metformin |
| BROMOCRIPTINE | BROMOCRIPTINE | 730603 | Bromocriptine |
| GLICLAZIDE | GLICLAZIDE | 19059796 | Gliclazide |
| INSULIN | INSULIN | 1513912 | Insulin Lispro |
| INSULIN | INSULIN | 1597000 | Regular Insulin, Human |
| GLIPIZIDE | GLIPIZIDE | 36239236 | Glipizide |
| CANAGLIFLOZIN | CANAGLIFLOZIN | 43526474 | canagliflozin |
| GLIMEPIRIDE | GLIMEPIRIDE | 19023585 | glimepiride |
| EXENATIDE | EXENATIDE | 19083253 | exenatide |
| MIGLITOL | MIGLITOL | 36233498 | miglitol |
| INSULIN | INSULIN | 1567198 | Insulin, Aspart, Human |
| INSULIN | INSULIN | 1597012 | Regular Insulin, Human |
| INSULIN | INSULIN | 1516976 | insulin detemir |
| INSULIN | INSULIN | 36236032 | Insulin Lispro |
| INSULIN | INSULIN | 1596941 | insulin, isophane |
| INSULIN | INSULIN | 19009422 | insulin, isophane |
| GLIMEPIRIDE | GLIMEPIRIDE | 1597756 | glimepiride |
| INSULIN | INSULIN | 19131582 | Insulin, Glulisine, Human |
| SITAGLIPTIN | SITAGLIPTIN | 19125050 | sitagliptin |
| BROMOCRIPTINE | BROMOCRIPTINE | 19006288 | Bromocriptine |
| GLIMEPIRIDE | GLIMEPIRIDE | 1597756 | glimepiride |
| EXENATIDE | EXENATIDE | 42708078 | exenatide |
| GLYBURIDE | GLYBURIDE | 1559684 | Glyburide |
| ACARBOSE | ACARBOSE | 19040970 | Acarbose |
| INSULIN | INSULIN | 19078553 | insulin, isophane |
| INSULIN | INSULIN | 1550023 | Insulin Lispro |
| METFORMIN | METFORMIN | 40063352 | Metformin |
| BROMOCRIPTINE | BROMOCRIPTINE | 730548 | Bromocriptine |
| METFORMIN | METFORMIN | 40126689 | Metformin |
| GLIMEPIRIDE | GLIMEPIRIDE | 1597776 | glimepiride |
| INSULIN | INSULIN | 19078557 | Regular Insulin, Human |
| LIRAGLUTIDE | LIRAGLUTIDE | 36241903 | liraglutide |
| EXENATIDE | EXENATIDE | 1583730 | exenatide |
| INSULIN | INSULIN | 19062456 | insulin, isophane |

| | | | |
|---|---|---|---|
| INSULIN | INSULIN | 19078553 | insulin, isophane |
| METFORMIN | METFORMIN | 1503328 | Metformin |
| METFORMIN | METFORMIN | 40164918 | Metformin |
| BROMOCRIPTINE | BROMOCRIPTINE | 19007101 | Bromocriptine |
| GLIPIZIDE | GLIPIZIDE | 19134624 | Glipizide |
| GLIPIZIDE | GLIPIZIDE | 19081295 | Glipizide |
| ROSIGLITAZONE | ROSIGLITAZONE | 1547506 | rosiglitazone |
| ALBIGLUTIDE | ALBIGLUTIDE | 44816283 | albiglutide |
| INSULIN | INSULIN | 19129442 | insulin human, rDNA origin |
| INSULIN | INSULIN | 19078610 | insulin, regular, beef-pork |
| METFORMIN | METFORMIN | 42708169 | Metformin |
| ACARBOSE | ACARBOSE | 19021312 | Acarbose |
| INSULIN | INSULIN | 40176981 | Insulin, Aspart Protamine, Human |
| INSULIN | INSULIN | 1596941 | insulin, isophane |
| DULAGLUTIDE | DULAGLUTIDE | 45774440 | dulaglutide |
| INSULIN | INSULIN | 46233943 | Insulin Lispro |
| GLYBURIDE | GLYBURIDE | 19030573 | Glyburide |
| METFORMIN | METFORMIN | 1503297 | Metformin |
| INSULIN | INSULIN | 19110356 | Regular Insulin, Human |
| INSULIN | INSULIN | 46234240 | insulin, isophane |
| ROSIGLITAZONE | ROSIGLITAZONE | 1547507 | rosiglitazone |
| BROMOCRIPTINE | BROMOCRIPTINE | 730551 | Bromocriptine |
| INSULIN | INSULIN | 19095214 | insulin, isophane |
| ACARBOSE | ACARBOSE | 19040970 | Acarbose |
| BROMOCRIPTINE | BROMOCRIPTINE | 730548 | Bromocriptine |
| PIOGLITAZONE | PIOGLITAZONE | 1525215 | pioglitazone |
| INSULIN | INSULIN | 1567198 | Insulin, Aspart, Human |
| METFORMIN | METFORMIN | 40164886 | Metformin |
| INSULIN | INSULIN | 40052088 | Insulin Lispro |
| INSULIN | INSULIN | 19078603 | insulin, isophane |
| INSULIN | INSULIN | 40052088 | Insulin Lispro |
| INSULIN | INSULIN | 36221713 | Regular Insulin, Human |

| | | | |
|---|---|---|---|
| EXENATIDE | EXENATIDE | 42708078 | exenatide |
| INSULIN | INSULIN | 1502910 | Insulin Glargine |
| INSULIN | INSULIN | 1550023 | Insulin Lispro |
| GLIMEPIRIDE | GLIMEPIRIDE | 1597758 | glimepiride |
| INSULIN | INSULIN | 46233730 | Regular Insulin, Human |
| GLYBURIDE | GLYBURIDE | 1559684 | Glyburide |
| INSULIN | INSULIN | 1567198 | Insulin, Aspart, Human |
| EMPAGLIFLOZIN | EMPAGLIFLOZIN | 36248376 | empagliflozin |
| INSULIN | INSULIN | 19078553 | insulin, isophane |
| METFORMIN | METFORMIN | 40229048 | Metformin |
| GLIPIZIDE | GLIPIZIDE | 1560171 | Glipizide |
| METFORMIN | METFORMIN | 1503297 | Metformin |
| METFORMIN | METFORMIN | 36221230 | Metformin |
| GLYBURIDE | GLYBURIDE | 40166002 | Glyburide |
| INSULIN | INSULIN | 19110357 | Regular Insulin, Human |
| SAXAGLIPTIN | SAXAGLIPTIN | 40166037 | saxagliptin |
| PRAMLINTIDE | PRAMLINTIDE | 40165687 | Pramlintide |
| INSULIN | INSULIN | 46221559 | Insulin Glargine |
| METFORMIN | METFORMIN | 1503327 | Metformin |
| SITAGLIPTIN | SITAGLIPTIN | 1580747 | sitagliptin |
| LINAGLIPTIN | LINAGLIPTIN | 42708087 | Linagliptin |
| GLIMEPIRIDE | GLIMEPIRIDE | 1597756 | glimepiride |
| BROMOCRIPTINE | BROMOCRIPTINE | 36214531 | Bromocriptine |
| METFORMIN | METFORMIN | 1503297 | Metformin |
| PIOGLITAZONE | PIOGLITAZONE | 1525215 | pioglitazone |
| METFORMIN | METFORMIN | 1503297 | Metformin |
| EXENATIDE | EXENATIDE | 1583730 | exenatide |
| PIOGLITAZONE | PIOGLITAZONE | 19099208 | pioglitazone |
| METFORMIN | METFORMIN | 1503327 | Metformin |
| GLYBURIDE | GLYBURIDE | 36233495 | Glyburide |
| GLYBURIDE | GLYBURIDE | 19030579 | Glyburide |
| METFORMIN | METFORMIN | 40063354 | Metformin |
| INSULIN | INSULIN | 46233975 | Insulin Lispro |
| INSULIN | INSULIN | 1596916 | insulin, isophane |
| INSULIN | INSULIN | 1550023 | Insulin Lispro |
| METFORMIN | METFORMIN | 1503328 | Metformin |

| | | | |
|---|---|---|---|
| INSULIN | INSULIN | 1550023 | Insulin Lispro |
| INSULIN | INSULIN | 19078526 | Insulin, Zinc, Human |
| INSULIN | INSULIN | 19116609 | insulin, isophane |
| INSULIN | INSULIN | 46234045 | Insulin, Aspart, Human |
| DULAGLUTIDE | DULAGLUTIDE | 36247185 | dulaglutide |
| BROMOCRIPTINE | BROMOCRIPTINE | 36234234 | Bromocriptine |
| GLIMEPIRIDE | GLIMEPIRIDE | 1597781 | glimepiride |
| INSULIN | INSULIN | 46221552 | Insulin Glargine |
| INSULIN | INSULIN | 19095214 | insulin, isophane |
| METFORMIN | METFORMIN | 40231396 | Metformin |
| INSULIN | INSULIN | 19061670 | Regular Insulin, Human |
| INSULIN | INSULIN | 19104458 | insulin, isophane |
| SITAGLIPTIN | SITAGLIPTIN | 36224404 | sitagliptin |
| PIOGLITAZONE | PIOGLITAZONE | 1525242 | pioglitazone |
| METFORMIN | METFORMIN | 40164904 | Metformin |
| INSULIN | INSULIN | 46233948 | Insulin Lispro |
| REPAGLINIDE | REPAGLINIDE | 36240429 | repaglinide |
| INSULIN | INSULIN | 19078555 | Regular Insulin, Human |
| INSULIN | INSULIN | 1502905 | Insulin Glargine |
| METFORMIN | METFORMIN | 1503297 | Metformin |
| GLIMEPIRIDE | GLIMEPIRIDE | 1597759 | glimepiride |
| EXENATIDE | EXENATIDE | 1583728 | exenatide |
| EXENATIDE | EXENATIDE | 19135404 | exenatide |
| INSULIN | INSULIN | 40051347 | Insulin Glargine |
| LINAGLIPTIN | LINAGLIPTIN | 42708091 | Linagliptin |
| REPAGLINIDE | REPAGLINIDE | 19023425 | repaglinide |
| ALOGLIPTIN | ALOGLIPTIN | 43013897 | alogliptin |
| PIOGLITAZONE | PIOGLITAZONE | 36236324 | pioglitazone |
| INSULIN | INSULIN | 19009384 | insulin, isophane |
| METFORMIN | METFORMIN | 1503297 | Metformin |
| DAPAGLIFLOZIN | DAPAGLIFLOZIN | 45775999 | dapagliflozin |

| | | | |
|---|---|---|---|
| METFORMIN | METFORMIN | 40117785 | Metformin |
| ALOGLIPTIN | ALOGLIPTIN | 36246195 | alogliptin |
| PIOGLITAZONE | PIOGLITAZONE | 1525215 | pioglitazone |
| INSULIN | INSULIN | 19058398 | Insulin Lispro |
| TOLBUTAMIDE | TOLBUTAMIDE | 1502856 | Tolbutamide |
| ACARBOSE | ACARBOSE | 19047612 | Acarbose |
| GLIMEPIRIDE | GLIMEPIRIDE | 1597776 | glimepiride |
| DULAGLUTIDE | DULAGLUTIDE | 45774435 | dulaglutide |
| TOLAZAMIDE | TOLAZAMIDE | 36230840 | Tolazamide |
| INSULIN | INSULIN | 19078610 | insulin, regular, beef-pork |
| INSULIN | INSULIN | 36231450 | Insulin, Aspart, Human |
| METFORMIN | METFORMIN | 1503297 | Metformin |
| GLIMEPIRIDE | GLIMEPIRIDE | 1597760 | glimepiride |
| INSULIN | INSULIN | 19110356 | Regular Insulin, Human |
| METFORMIN | METFORMIN | 40164946 | Metformin |
| METFORMIN | METFORMIN | 36235752 | Metformin |
| ACARBOSE | ACARBOSE | 1529331 | Acarbose |
| INSULIN | INSULIN | 19078554 | Regular Insulin, Human |
| INSULIN | INSULIN | 36222190 | insulin detemir |
| INSULIN | INSULIN | 46234045 | Insulin, Aspart, Human |
| INSULIN | INSULIN | 19095199 | Lente Insulin, Beef-Pork |
| LINAGLIPTIN | LINAGLIPTIN | 40239221 | Linagliptin |
| INSULIN | INSULIN | 19078553 | insulin, isophane |
| PRAMLINTIDE | PRAMLINTIDE | 40127005 | Pramlintide |
| INSULIN | INSULIN | 1502910 | Insulin Glargine |
| INSULIN | INSULIN | 19100127 | insulin, isophane |
| INSULIN | INSULIN | 1516981 | insulin detemir |
| LIRAGLUTIDE | LIRAGLUTIDE | 36220119 | liraglutide |
| EMPAGLIFLOZIN | EMPAGLIFLOZIN | 46287410 | empagliflozin |
| GLIPIZIDE | GLIPIZIDE | 19085004 | Glipizide |
| GLIPIZIDE | GLIPIZIDE | 1560171 | Glipizide |
| LINAGLIPTIN | LINAGLIPTIN | 42708091 | Linagliptin |
| LINAGLIPTIN | LINAGLIPTIN | 40239217 | Linagliptin |
| GLYBURIDE | GLYBURIDE | 1559684 | Glyburide |
| DULAGLUTIDE | DULAGLUTIDE | 46234471 | dulaglutide |

| | | | |
|---|---|---|---|
| INSULIN | INSULIN | 40176981 | Insulin, Aspart Protamine, Human |
| ALOGLIPTIN | ALOGLIPTIN | 43013925 | alogliptin |
| INSULIN | INSULIN | 1516981 | insulin detemir |
| GLIPIZIDE | GLIPIZIDE | 1560171 | Glipizide |
| INSULIN | INSULIN | 19062999 | insulin, isophane |
| DAPAGLIFLOZIN | DAPAGLIFLOZIN | 44785477 | dapagliflozin |
| CANAGLIFLOZIN | CANAGLIFLOZIN | 36246430 | canagliflozin |
| INSULIN | INSULIN | 1567198 | Insulin, Aspart, Human |
| GLIPIZIDE | GLIPIZIDE | 40167635 | Glipizide |
| INSULIN | INSULIN | 19116609 | insulin, isophane |
| METFORMIN | METFORMIN | 40164897 | Metformin |
| METFORMIN | METFORMIN | 36235746 | Metformin |
| INSULIN | INSULIN | 19071700 | Insulin Glargine |
| ALBIGLUTIDE | ALBIGLUTIDE | 44816332 | albiglutide |
| INSULIN | INSULIN | 19071700 | Insulin Glargine |
| EXENATIDE | EXENATIDE | 42708079 | exenatide |
| INSULIN | INSULIN | 19078553 | insulin, isophane |
| GLYBURIDE | GLYBURIDE | 1559784 | Glyburide |
| METFORMIN | METFORMIN | 40176046 | Metformin |
| INSULIN | INSULIN | 19095218 | Regular Insulin, Human |
| REPAGLINIDE | REPAGLINIDE | 1516770 | repaglinide |
| INSULIN | INSULIN | 1550023 | Insulin Lispro |
| GLYBURIDE | GLYBURIDE | 36231169 | Glyburide |
| INSULIN | INSULIN | 40051348 | Insulin Glargine |
| INSULIN | INSULIN | 19010204 | insulin, isophane |
| METFORMIN | METFORMIN | 1503328 | Metformin |
| INSULIN | INSULIN | 1516979 | insulin detemir |
| METFORMIN | METFORMIN | 40164932 | Metformin |
| ALBIGLUTIDE | ALBIGLUTIDE | 44816283 | albiglutide |
| TOLBUTAMIDE | TOLBUTAMIDE | 36214386 | Tolbutamide |
| METFORMIN | METFORMIN | 40229046 | Metformin |
| INSULIN | INSULIN | 1596957 | insulin, isophane |
| CANAGLIFLOZIN | CANAGLIFLOZIN | 36246431 | canagliflozin |

| | | | |
|---|---|---|---|
| ALBIGLUTIDE | ALBIGLUTIDE | 44816274 | albiglutide |
| PIOGLITAZONE | PIOGLITAZONE | 36220121 | pioglitazone |
| ROSIGLITAZONE | ROSIGLITAZONE | 19079465 | rosiglitazone |
| EXENATIDE | EXENATIDE | 36234709 | exenatide |
| LIRAGLUTIDE | LIRAGLUTIDE | 45892686 | liraglutide |
| INSULIN | INSULIN | 19135263 | insulin, isophane |
| GLIPIZIDE | GLIPIZIDE | 40044251 | Glipizide |
| METFORMIN | METFORMIN | 1503297 | Metformin |
| METFORMIN | METFORMIN | 1503327 | Metformin |
| INSULIN | INSULIN | 19110220 | Regular Insulin, Human |
| INSULIN | INSULIN | 46221555 | Insulin Glargine |
| INSULIN | INSULIN | 46221558 | Insulin Glargine |
| INSULIN | INSULIN | 36231450 | Insulin, Aspart, Human |
| ROSIGLITAZONE | ROSIGLITAZONE | 1547506 | rosiglitazone |
| INSULIN | INSULIN | 36231450 | Insulin, Aspart, Human |
| ROSIGLITAZONE | ROSIGLITAZONE | 19099246 | rosiglitazone |
| SAXAGLIPTIN | SAXAGLIPTIN | 40166037 | saxagliptin |
| CANAGLIFLOZIN | CANAGLIFLOZIN | 43526471 | canagliflozin |
| INSULIN | INSULIN | 1597012 | Regular Insulin, Human |
| INSULIN | INSULIN | 1567198 | Insulin, Aspart, Human |
| INSULIN | INSULIN | 19104469 | insulin, isophane |
| GLIMEPIRIDE | GLIMEPIRIDE | 19023585 | glimepiride |
| GLIPIZIDE | GLIPIZIDE | 19100559 | Glipizide |
| INSULIN | INSULIN | 1596932 | insulin, isophane |
| EMPAGLIFLOZIN | EMPAGLIFLOZIN | 46287410 | empagliflozin |
| GLIPIZIDE | GLIPIZIDE | 19081295 | Glipizide |
| GLIMEPIRIDE | GLIMEPIRIDE | 1597773 | glimepiride |
| GLIPIZIDE | GLIPIZIDE | 19117945 | Glipizide |
| INSULIN | INSULIN | 40176981 | Insulin, Aspart Protamine, Human |
| INSULIN | INSULIN | 19120790 | Insulin Glargine |
| INSULIN | INSULIN | 1567198 | Insulin, Aspart, Human |
| GLIMEPIRIDE | GLIMEPIRIDE | 19117200 | glimepiride |
| INSULIN | INSULIN | 19062999 | insulin, isophane |
| INSULIN | INSULIN | 19016117 | Insulin Lispro |
| GLYBURIDE | GLYBURIDE | 1559684 | Glyburide |

| | | | |
|---|---|---|---|
| LINAGLIPTIN | LINAGLIPTIN | 36238287 | Linagliptin |
| METFORMIN | METFORMIN | 40231392 | Metformin |
| PIOGLITAZONE | PIOGLITAZONE | 1525244 | pioglitazone |
| INSULIN | INSULIN | 1567232 | Insulin, Aspart, Human |
| EXENATIDE | EXENATIDE | 1583728 | exenatide |
| METFORMIN | METFORMIN | 40164897 | Metformin |
| GLIPIZIDE | GLIPIZIDE | 19081295 | Glipizide |
| GLIPIZIDE | GLIPIZIDE | 40165972 | Glipizide |
| GLIMEPIRIDE | GLIMEPIRIDE | 1597774 | glimepiride |
| METFORMIN | METFORMIN | 40164929 | Metformin |
| INSULIN | INSULIN | 19062999 | insulin, isophane |
| INSULIN | INSULIN | 19078557 | Regular Insulin, Human |
| GLYBURIDE | GLYBURIDE | 1559784 | Glyburide |
| EXENATIDE | EXENATIDE | 1583727 | exenatide |
| REPAGLINIDE | REPAGLINIDE | 1516766 | repaglinide |
| GLIPIZIDE | GLIPIZIDE | 19077636 | Glipizide |
| GLYBURIDE | GLYBURIDE | 1559684 | Glyburide |
| DAPAGLIFLOZIN | DAPAGLIFLOZIN | 44785833 | dapagliflozin |
| PRAMLINTIDE | PRAMLINTIDE | 40163890 | Pramlintide |
| GLIPIZIDE | GLIPIZIDE | 36218767 | Glipizide |
| INSULIN | INSULIN | 36233154 | Insulin, Glulisine, Human |
| INSULIN | INSULIN | 19078526 | Insulin, Zinc, Human |
| PIOGLITAZONE | PIOGLITAZONE | 19099209 | pioglitazone |
| INSULIN | INSULIN | 36229086 | Insulin Glargine |
| INSULIN | INSULIN | 1550028 | Insulin Lispro |
| GLIPIZIDE | GLIPIZIDE | 19117945 | Glipizide |
| METFORMIN | METFORMIN | 1503297 | Metformin |
| GLIPIZIDE | GLIPIZIDE | 19077636 | Glipizide |
| INSULIN | INSULIN | 1502910 | Insulin Glargine |
| LINAGLIPTIN | LINAGLIPTIN | 42708090 | Linagliptin |
| TOLAZAMIDE | TOLAZAMIDE | 19036966 | Tolazamide |
| ROSIGLITAZONE | ROSIGLITAZONE | 19099246 | rosiglitazone |
| METFORMIN | METFORMIN | 40164882 | Metformin |
| GLIPIZIDE | GLIPIZIDE | 36218767 | Glipizide |

| | | | |
|---|---|---|---|
| METFORMIN | METFORMIN | 40063350 | Metformin |
| METFORMIN | METFORMIN | 40126695 | Metformin |
| GLYBURIDE | GLYBURIDE | 1559784 | Glyburide |
| CHLORPROPAMIDE | CHLORPROPAMIDE | 1594976 | Chlorpropamide |
| INSULIN | INSULIN | 19110352 | insulin, isophane |
| LIRAGLUTIDE | LIRAGLUTIDE | 40169223 | liraglutide |
| GLYBURIDE | GLYBURIDE | 1559684 | Glyburide |
| REPAGLINIDE | REPAGLINIDE | 1516771 | repaglinide |
| BROMOCRIPTINE | BROMOCRIPTINE | 730548 | Bromocriptine |
| INSULIN | INSULIN | 19012269 | Insulin, Zinc, Human |
| GLICLAZIDE | GLICLAZIDE | 19059797 | Gliclazide |
| METFORMIN | METFORMIN | 36239258 | Metformin |
| EMPAGLIFLOZIN | EMPAGLIFLOZIN | 45774895 | empagliflozin |
| GLIMEPIRIDE | GLIMEPIRIDE | 40059666 | glimepiride |
| INSULIN | INSULIN | 42899447 | Insulin, Pork |
| INSULIN | INSULIN | 1550028 | Insulin Lispro |
| BROMOCRIPTINE | BROMOCRIPTINE | 40016899 | Bromocriptine |
| ACARBOSE | ACARBOSE | 1529352 | Acarbose |
| EMPAGLIFLOZIN | EMPAGLIFLOZIN | 36248789 | empagliflozin |
| INSULIN | INSULIN | 19078555 | Regular Insulin, Human |
| GLIMEPIRIDE | GLIMEPIRIDE | 1597756 | glimepiride |
| INSULIN | INSULIN | 46234241 | Regular Insulin, Human |
| INSULIN | INSULIN | 1550023 | Insulin Lispro |
| INSULIN | INSULIN | 35602731 | insulin degludec |
| INSULIN | INSULIN | 19110355 | insulin, isophane |
| GLIPIZIDE | GLIPIZIDE | 19117945 | Glipizide |
| INSULIN | INSULIN | 19110219 | insulin, isophane |
| INSULIN | INSULIN | 19078603 | insulin, isophane |
| METFORMIN | METFORMIN | 1503328 | Metformin |
| INSULIN | INSULIN | 19014975 | insulin, isophane |
| GLIMEPIRIDE | GLIMEPIRIDE | 1597774 | glimepiride |
| GLIPIZIDE | GLIPIZIDE | 40165973 | Glipizide |
| INSULIN | INSULIN | 19078553 | insulin, isophane |
| MIGLITOL | MIGLITOL | 36233498 | miglitol |
| INSULIN | INSULIN | 19071700 | Insulin Glargine |

| | | | |
|---|---|---|---|
| INSULIN | INSULIN | 40184293 | Insulin, Aspart Protamine, Human |
| INSULIN | INSULIN | 19078553 | insulin, isophane |
| INSULIN | INSULIN | 40166274 | Insulin Lispro |
| INSULIN | INSULIN | 19100128 | Regular Insulin, Human |
| METFORMIN | METFORMIN | 40164893 | Metformin |
| GLIMEPIRIDE | GLIMEPIRIDE | 1597773 | glimepiride |
| GLYBURIDE | GLYBURIDE | 1559684 | Glyburide |
| INSULIN | INSULIN | 1596941 | insulin, isophane |
| CANAGLIFLOZIN | CANAGLIFLOZIN | 1718909 | canagliflozin |
| BROMOCRIPTINE | BROMOCRIPTINE | 730551 | Bromocriptine |
| MIGLITOL | MIGLITOL | 36215710 | miglitol |
| INSULIN | INSULIN | 45777025 | Regular Insulin, Human |
| INSULIN | INSULIN | 19116559 | insulin, isophane |
| INSULIN | INSULIN | 1596960 | insulin, isophane |
| INSULIN | INSULIN | 1550027 | Insulin Lispro |
| INSULIN | INSULIN | 19078554 | Regular Insulin, Human |
| ACARBOSE | ACARBOSE | 40003413 | Acarbose |
| EXENATIDE | EXENATIDE | 19083254 | exenatide |
| ACARBOSE | ACARBOSE | 1529331 | Acarbose |
| GLIMEPIRIDE | GLIMEPIRIDE | 1597756 | glimepiride |
| CANAGLIFLOZIN | CANAGLIFLOZIN | 45774714 | canagliflozin |
| METFORMIN | METFORMIN | 40164907 | Metformin |
| LIRAGLUTIDE | LIRAGLUTIDE | 40170913 | liraglutide |
| METFORMIN | METFORMIN | 40117785 | Metformin |
| GLIMEPIRIDE | GLIMEPIRIDE | 1597795 | glimepiride |
| INSULIN | INSULIN | 36236032 | Insulin Lispro |
| ACARBOSE | ACARBOSE | 19120257 | Acarbose |
| INSULIN | INSULIN | 19123376 | Insulin Lispro |
| INSULIN | INSULIN | 19044971 | insulin detemir |
| ALOGLIPTIN | ALOGLIPTIN | 43013918 | alogliptin |
| LIRAGLUTIDE | LIRAGLUTIDE | 36220119 | liraglutide |

| | | | |
|---|---|---|---|
| INSULIN | INSULIN | 1550023 | Insulin Lispro |
| INSULIN | INSULIN | 1567198 | Insulin, Aspart, Human |
| INSULIN | INSULIN | 19100127 | insulin, isophane |
| EMPAGLIFLOZIN | EMPAGLIFLOZIN | 46287410 | empagliflozin |
| METFORMIN | METFORMIN | 36235582 | Metformin |
| INSULIN | INSULIN | 36235843 | Ultralente Insulin, Human |
| GLIMEPIRIDE | GLIMEPIRIDE | 1597776 | glimepiride |
| INSULIN | INSULIN | 1513881 | Insulin Lispro |
| GLIPIZIDE | GLIPIZIDE | 36218767 | Glipizide |
| INSULIN | INSULIN | 1550027 | Insulin Lispro |
| INSULIN | INSULIN | 1567200 | Insulin, Aspart, Human |
| METFORMIN | METFORMIN | 40164929 | Metformin |
| INSULIN | INSULIN | 19094121 | Regular Insulin, Human |
| METFORMIN | METFORMIN | 40117785 | Metformin |
| INSULIN | INSULIN | 1596916 | insulin, isophane |
| METFORMIN | METFORMIN | 1503328 | Metformin |
| INSULIN | INSULIN | 1544838 | Insulin, Glulisine, Human |
| INSULIN | INSULIN | 1513912 | Insulin Lispro |
| INSULIN | INSULIN | 19010204 | insulin, isophane |
| INSULIN | INSULIN | 19078551 | insulin, isophane |
| REPAGLINIDE | REPAGLINIDE | 19078029 | repaglinide |
| GLYBURIDE | GLYBURIDE | 1559684 | Glyburide |
| INSULIN | INSULIN | 19135265 | Regular Insulin, Human |
| EXENATIDE | EXENATIDE | 1583730 | exenatide |
| REPAGLINIDE | REPAGLINIDE | 19078030 | repaglinide |
| INSULIN | INSULIN | 35602725 | insulin degludec |
| INSULIN | INSULIN | 35602724 | insulin degludec |
| INSULIN | INSULIN | 45774400 | Regular Insulin, Human |
| INSULIN | INSULIN | 1516976 | insulin detemir |
| INSULIN | INSULIN | 19095199 | Lente Insulin, Beef-Pork |
| INSULIN | INSULIN | 46234049 | Insulin, Aspart, Human |
| METFORMIN | METFORMIN | 40163924 | Metformin |

| | | | |
|---|---|---|---|
| METFORMIN | METFORMIN | 40164926 | Metformin |
| DAPAGLIFLOZIN | DAPAGLIFLOZIN | 44815995 | dapagliflozin |
| INSULIN | INSULIN | 1516981 | insulin detemir |
| LINAGLIPTIN | LINAGLIPTIN | 42708091 | Linagliptin |
| GLYBURIDE | GLYBURIDE | 1559782 | Glyburide |
| METFORMIN | METFORMIN | 40163926 | Metformin |
| INSULIN | INSULIN | 19078553 | insulin, isophane |
| INSULIN | INSULIN | 40184294 | Insulin, Aspart Protamine, Human |
| INSULIN | INSULIN | 1596999 | Regular Insulin, Human |
| PRAMLINTIDE | PRAMLINTIDE | 1517998 | Pramlintide |
| REPAGLINIDE | REPAGLINIDE | 1516766 | repaglinide |
| METFORMIN | METFORMIN | 36221231 | Metformin |
| INSULIN | INSULIN | 19078553 | insulin, isophane |
| MIGLITOL | MIGLITOL | 1510206 | miglitol |
| SITAGLIPTIN | SITAGLIPTIN | 36221233 | sitagliptin |
| GLIPIZIDE | GLIPIZIDE | 40165975 | Glipizide |
| PIOGLITAZONE | PIOGLITAZONE | 19099209 | pioglitazone |
| METFORMIN | METFORMIN | 40163927 | Metformin |
| INSULIN | INSULIN | 19094121 | Regular Insulin, Human |
| INSULIN | INSULIN | 19120315 | insulin, isophane |
| GLYBURIDE | GLYBURIDE | 1559782 | Glyburide |
| INSULIN | INSULIN | 19116450 | insulin, isophane |
| INSULIN | INSULIN | 46233969 | Insulin Lispro |
| LIRAGLUTIDE | LIRAGLUTIDE | 40170911 | liraglutide |
| INSULIN | INSULIN | 19095211 | Insulin Lispro |
| GLIPIZIDE | GLIPIZIDE | 1560171 | Glipizide |
| METFORMIN | METFORMIN | 40164882 | Metformin |
| GLIPIZIDE | GLIPIZIDE | 1560171 | Glipizide |
| GLIMEPIRIDE | GLIMEPIRIDE | 1597775 | glimepiride |
| INSULIN | INSULIN | 1516979 | insulin detemir |

| | | | |
|---|---|---|---|
| INSULIN | INSULIN | 36231450 | Insulin, Aspart, Human |
| INSULIN | INSULIN | 19078553 | insulin, isophane |
| INSULIN | INSULIN | 1596957 | insulin, isophane |
| INSULIN | INSULIN | 46275452 | Regular Insulin, Human |
| LINAGLIPTIN | LINAGLIPTIN | 42708088 | Linagliptin |
| DULAGLUTIDE | DULAGLUTIDE | 45774435 | dulaglutide |
| GLICLAZIDE | GLICLAZIDE | 40044222 | Gliclazide |
| INSULIN | INSULIN | 1596952 | insulin, isophane |
| INSULIN | INSULIN | 40184294 | Insulin, Aspart Protamine, Human |
| INSULIN | INSULIN | 19014975 | insulin, isophane |
| DAPAGLIFLOZIN | DAPAGLIFLOZIN | 44785831 | dapagliflozin |
| INSULIN | INSULIN | 19009384 | insulin, isophane |
| INSULIN | INSULIN | 40184293 | Insulin, Aspart Protamine, Human |
| GLYBURIDE | GLYBURIDE | 40165994 | Glyburide |
| INSULIN | INSULIN | 1596960 | insulin, isophane |
| GLYBURIDE | GLYBURIDE | 1559784 | Glyburide |
| GLIMEPIRIDE | GLIMEPIRIDE | 1597795 | glimepiride |
| INSULIN | INSULIN | 40051348 | Insulin Glargine |
| METFORMIN | METFORMIN | 36223145 | Metformin |
| DAPAGLIFLOZIN | DAPAGLIFLOZIN | 44816049 | dapagliflozin |
| EXENATIDE | EXENATIDE | 42705842 | exenatide |
| METFORMIN | METFORMIN | 36236481 | Metformin |
| METFORMIN | METFORMIN | 40164918 | Metformin |
| GLYBURIDE | GLYBURIDE | 1559782 | Glyburide |
| METFORMIN | METFORMIN | 36235788 | Metformin |
| METFORMIN | METFORMIN | 40231393 | Metformin |
| INSULIN | INSULIN | 1502905 | Insulin Glargine |
| METFORMIN | METFORMIN | 1503297 | Metformin |

| | | | |
|---|---|---|---|
| INSULIN | INSULIN | 19058398 | Insulin Lispro |
| REPAGLINIDE | REPAGLINIDE | 19023424 | repaglinide |
| GLYBURIDE | GLYBURIDE | 40165996 | Glyburide |
| METFORMIN | METFORMIN | 40231402 | Metformin |
| METFORMIN | METFORMIN | 1503327 | Metformin |
| METFORMIN | METFORMIN | 40164932 | Metformin |
| GLIMEPIRIDE | GLIMEPIRIDE | 1597796 | glimepiride |
| METFORMIN | METFORMIN | 40164912 | Metformin |
| INSULIN | INSULIN | 19078557 | Regular Insulin, Human |
| INSULIN | INSULIN | 19094122 | Regular Insulin, Human |
| METFORMIN | METFORMIN | 40164933 | Metformin |
| METFORMIN | METFORMIN | 1503297 | Metformin |
| INSULIN | INSULIN | 19120315 | insulin, isophane |
| GLIMEPIRIDE | GLIMEPIRIDE | 1597781 | glimepiride |
| INSULIN | INSULIN | 1596972 | insulin, isophane |
| INSULIN | INSULIN | 36235840 | insulin, isophane |
| EXENATIDE | EXENATIDE | 1583722 | exenatide |
| INSULIN | INSULIN | 19012269 | Insulin, Zinc, Human |
| INSULIN | INSULIN | 46233730 | Regular Insulin, Human |
| METFORMIN | METFORMIN | 1503297 | Metformin |
| GLYBURIDE | GLYBURIDE | 40166001 | Glyburide |
| ROSIGLITAZONE | ROSIGLITAZONE | 36229365 | rosiglitazone |
| INSULIN | INSULIN | 1567198 | Insulin, Aspart, Human |
| METFORMIN | METFORMIN | 36235787 | Metformin |
| CANAGLIFLOZIN | CANAGLIFLOZIN | 45774717 | canagliflozin |
| DULAGLUTIDE | DULAGLUTIDE | 45774447 | dulaglutide |
| GLIPIZIDE | GLIPIZIDE | 40166814 | Glipizide |
| METFORMIN | METFORMIN | 1503327 | Metformin |
| INSULIN | INSULIN | 19078554 | Regular Insulin, Human |

| | | | |
|---|---|---|---|
| INSULIN | INSULIN | 19120315 | insulin, isophane |
| EXENATIDE | EXENATIDE | 1583730 | exenatide |
| DAPAGLIFLOZIN | DAPAGLIFLOZIN | 36248559 | dapagliflozin |
| LINAGLIPTIN | LINAGLIPTIN | 36229565 | Linagliptin |
| METFORMIN | METFORMIN | 40063352 | Metformin |
| METFORMIN | METFORMIN | 40063352 | Metformin |
| INSULIN | INSULIN | 19120315 | insulin, isophane |
| INSULIN | INSULIN | 19116454 | insulin, isophane |
| INSULIN | INSULIN | 19095214 | insulin, isophane |
| GLIPIZIDE | GLIPIZIDE | 40044253 | Glipizide |
| BROMOCRIPTINE | BROMOCRIPTINE | 730548 | Bromocriptine |
| REPAGLINIDE | REPAGLINIDE | 19054444 | repaglinide |
| INSULIN | INSULIN | 40184293 | Insulin, Aspart Protamine, Human |
| GLIPIZIDE | GLIPIZIDE | 36235748 | Glipizide |
| GLIMEPIRIDE | GLIMEPIRIDE | 40059665 | glimepiride |
| INSULIN | INSULIN | 19112790 | Insulin, Glulisine, Human |
| GLIMEPIRIDE | GLIMEPIRIDE | 1597758 | glimepiride |
| LINAGLIPTIN | LINAGLIPTIN | 40239220 | Linagliptin |
| GLIMEPIRIDE | GLIMEPIRIDE | 19117199 | glimepiride |
| INSULIN | INSULIN | 36235840 | insulin, isophane |
| INSULIN | INSULIN | 19120315 | insulin, isophane |
| LIRAGLUTIDE | LIRAGLUTIDE | 40169223 | liraglutide |
| GLIPIZIDE | GLIPIZIDE | 40167637 | Glipizide |
| INSULIN | INSULIN | 1596959 | insulin, isophane |
| METFORMIN | METFORMIN | 1503297 | Metformin |
| GLYBURIDE | GLYBURIDE | 40165997 | Glyburide |
| METFORMIN | METFORMIN | 36223145 | Metformin |
| METFORMIN | METFORMIN | 36242511 | Metformin |
| SAXAGLIPTIN | SAXAGLIPTIN | 40166037 | saxagliptin |
| METFORMIN | METFORMIN | 40164928 | Metformin |
| METFORMIN | METFORMIN | 40164948 | Metformin |
| GLIPIZIDE | GLIPIZIDE | 19047638 | Glipizide |
| INSULIN | INSULIN | 1513914 | Insulin Lispro |

| | | | |
|---|---|---|---|
| PRAMLINTIDE | PRAMLINTIDE | 1517998 | Pramlintide |
| INSULIN | INSULIN | 42902743 | Insulin Glargine |
| METFORMIN | METFORMIN | 40164895 | Metformin |
| SAXAGLIPTIN | SAXAGLIPTIN | 36224321 | saxagliptin |
| REPAGLINIDE | REPAGLINIDE | 36213643 | repaglinide |
| INSULIN | INSULIN | 35602723 | insulin degludec |
| METFORMIN | METFORMIN | 40164942 | Metformin |
| GLYBURIDE | GLYBURIDE | 40046201 | Glyburide |
| METFORMIN | METFORMIN | 40164892 | Metformin |
| INSULIN | INSULIN | 19078610 | insulin, regular, beef-pork |
| PIOGLITAZONE | PIOGLITAZONE | 19099207 | pioglitazone |
| INSULIN | INSULIN | 19014975 | insulin, isophane |
| GLIPIZIDE | GLIPIZIDE | 1560233 | Glipizide |
| ALBIGLUTIDE | ALBIGLUTIDE | 44816274 | albiglutide |
| GLIPIZIDE | GLIPIZIDE | 19080793 | Glipizide |
| METFORMIN | METFORMIN | 1503297 | Metformin |
| INSULIN | INSULIN | 19016117 | Insulin Lispro |
| METFORMIN | METFORMIN | 40164942 | Metformin |
| METFORMIN | METFORMIN | 19106521 | Metformin |
| SITAGLIPTIN | SITAGLIPTIN | 19125042 | sitagliptin |
| INSULIN | INSULIN | 19062999 | insulin, isophane |
| METFORMIN | METFORMIN | 40229050 | Metformin |
| INSULIN | INSULIN | 19129469 | insulin human, rDNA origin |
| INSULIN | INSULIN | 19110357 | Regular Insulin, Human |
| METFORMIN | METFORMIN | 40164948 | Metformin |
| REPAGLINIDE | REPAGLINIDE | 19023426 | repaglinide |
| METFORMIN | METFORMIN | 40164920 | Metformin |
| ROSIGLITAZONE | ROSIGLITAZONE | 19099245 | rosiglitazone |
| METFORMIN | METFORMIN | 1503297 | Metformin |
| SAXAGLIPTIN | SAXAGLIPTIN | 40166041 | saxagliptin |
| INSULIN | INSULIN | 1516976 | insulin detemir |
| INSULIN | INSULIN | 40184294 | Insulin, Aspart Protamine, Human |

| | | | |
|---|---|---|---|
| GLIPIZIDE | GLIPIZIDE | 19077636 | Glipizide |
| GLIPIZIDE | GLIPIZIDE | 36218767 | Glipizide |
| INSULIN | INSULIN | 1502910 | Insulin Glargine |
| INSULIN | INSULIN | 19018822 | Insulin Lispro |
| INSULIN | INSULIN | 19116454 | insulin, isophane |
| GLYBURIDE | GLYBURIDE | 1559782 | Glyburide |
| LIRAGLUTIDE | LIRAGLUTIDE | 36241903 | liraglutide |
| LINAGLIPTIN | LINAGLIPTIN | 36221103 | Linagliptin |
| INSULIN | INSULIN | 1596955 | insulin, isophane |
| INSULIN | INSULIN | 1550023 | Insulin Lispro |
| METFORMIN | METFORMIN | 19086484 | Metformin |
| LINAGLIPTIN | LINAGLIPTIN | 36238286 | Linagliptin |
| INSULIN | INSULIN | 1596974 | insulin, isophane |
| METFORMIN | METFORMIN | 40164918 | Metformin |
| INSULIN | INSULIN | 19058398 | Insulin Lispro |
| PIOGLITAZONE | PIOGLITAZONE | 1525242 | pioglitazone |
| ALBIGLUTIDE | ALBIGLUTIDE | 44816283 | albiglutide |
| ACARBOSE | ACARBOSE | 1529331 | Acarbose |
| METFORMIN | METFORMIN | 1503297 | Metformin |
| SITAGLIPTIN | SITAGLIPTIN | 19125050 | sitagliptin |
| ACARBOSE | ACARBOSE | 19047612 | Acarbose |
| DAPAGLIFLOZIN | DAPAGLIFLOZIN | 45775867 | dapagliflozin |
| INSULIN | INSULIN | 36233552 | insulin, isophane |
| ALBIGLUTIDE | ALBIGLUTIDE | 44816274 | albiglutide |
| METFORMIN | METFORMIN | 1503297 | Metformin |
| SITAGLIPTIN | SITAGLIPTIN | 19125042 | sitagliptin |
| EXENATIDE | EXENATIDE | 19083254 | exenatide |
| CANAGLIFLOZIN | CANAGLIFLOZIN | 36246430 | canagliflozin |
| PIOGLITAZONE | PIOGLITAZONE | 1525216 | pioglitazone |
| INSULIN | INSULIN | 40166274 | Insulin Lispro |
| INSULIN | INSULIN | 1596999 | Regular Insulin, Human |
| GLIPIZIDE | GLIPIZIDE | 1560233 | Glipizide |
| INSULIN | INSULIN | 1596955 | insulin, isophane |
| PRAMLINTIDE | PRAMLINTIDE | 40078056 | Pramlintide |
| METFORMIN | METFORMIN | 40164900 | Metformin |

| | | | |
|---|---|---|---|
| GLIMEPIRIDE | GLIMEPIRIDE | 19117200 | glimepiride |
| DAPAGLIFLOZIN | DAPAGLIFLOZIN | 45775860 | dapagliflozin |
| ACARBOSE | ACARBOSE | 1529331 | Acarbose |
| GLIPIZIDE | GLIPIZIDE | 19081295 | Glipizide |
| INSULIN | INSULIN | 19062999 | insulin, isophane |
| METFORMIN | METFORMIN | 1503328 | Metformin |
| INSULIN | INSULIN | 1596977 | Regular Insulin, Human |
| GLYBURIDE | GLYBURIDE | 19030576 | Glyburide |
| INSULIN | INSULIN | 1586369 | Ultralente Insulin, Human |
| INSULIN | INSULIN | 19080091 | Ultralente Insulin, Human |
| INSULIN | INSULIN | 1516981 | insulin detemir |
| METFORMIN | METFORMIN | 19088818 | Metformin |
| GLIPIZIDE | GLIPIZIDE | 19047638 | Glipizide |
| GLIPIZIDE | GLIPIZIDE | 1560233 | Glipizide |
| BROMOCRIPTINE | BROMOCRIPTINE | 36238866 | Bromocriptine |
| INSULIN | INSULIN | 35602724 | insulin degludec |
| METFORMIN | METFORMIN | 40163925 | Metformin |
| INSULIN | INSULIN | 1502905 | Insulin Glargine |
| INSULIN | INSULIN | 1513849 | Insulin, Zinc, Human |
| INSULIN | INSULIN | 19078559 | Insulin Glargine |
| LIRAGLUTIDE | LIRAGLUTIDE | 40170911 | liraglutide |
| INSULIN | INSULIN | 1550023 | Insulin Lispro |
| LIRAGLUTIDE | LIRAGLUTIDE | 40170913 | liraglutide |
| METFORMIN | METFORMIN | 1503328 | Metformin |
| GLIPIZIDE | GLIPIZIDE | 19077638 | Glipizide |
| INSULIN | INSULIN | 1550023 | Insulin Lispro |
| BROMOCRIPTINE | BROMOCRIPTINE | 19083958 | Bromocriptine |
| GLYBURIDE | GLYBURIDE | 19110154 | Glyburide |
| METFORMIN | METFORMIN | 1503328 | Metformin |
| BROMOCRIPTINE | BROMOCRIPTINE | 19007101 | Bromocriptine |
| LIRAGLUTIDE | LIRAGLUTIDE | 36241903 | liraglutide |
| REPAGLINIDE | REPAGLINIDE | 36213643 | repaglinide |
| METFORMIN | METFORMIN | 40164918 | Metformin |
| METFORMIN | METFORMIN | 40171451 | Metformin |

| | | | |
|---|---|---|---|
| EXENATIDE | EXENATIDE | 19135404 | exenatide |
| INSULIN | INSULIN | 19078554 | Regular Insulin, Human |
| INSULIN | INSULIN | 40184293 | Insulin, Aspart Protamine, Human |
| INSULIN | INSULIN | 19120790 | Insulin Glargine |
| INSULIN | INSULIN | 1550028 | Insulin Lispro |
| INSULIN | INSULIN | 19078553 | insulin, isophane |
| LIRAGLUTIDE | LIRAGLUTIDE | 40170912 | liraglutide |
| INSULIN | INSULIN | 1544838 | Insulin, Glulisine, Human |
| INSULIN | INSULIN | 46233970 | Insulin Lispro |
| GLIPIZIDE | GLIPIZIDE | 19081295 | Glipizide |
| INSULIN | INSULIN | 46233948 | Insulin Lispro |
| INSULIN | INSULIN | 46234045 | Insulin, Aspart, Human |
| CANAGLIFLOZIN | CANAGLIFLOZIN | 45774724 | canagliflozin |
| INSULIN | INSULIN | 19016117 | Insulin Lispro |
| GLIPIZIDE | GLIPIZIDE | 1560233 | Glipizide |
| METFORMIN | METFORMIN | 40164929 | Metformin |
| METFORMIN | METFORMIN | 40164897 | Metformin |
| SITAGLIPTIN | SITAGLIPTIN | 19125042 | sitagliptin |
| DULAGLUTIDE | DULAGLUTIDE | 45774447 | dulaglutide |
| SAXAGLIPTIN | SAXAGLIPTIN | 40166043 | saxagliptin |
| INSULIN | INSULIN | 1550028 | Insulin Lispro |
| METFORMIN | METFORMIN | 36221230 | Metformin |
| INSULIN | INSULIN | 1502905 | Insulin Glargine |
| CANAGLIFLOZIN | CANAGLIFLOZIN | 36246430 | canagliflozin |
| INSULIN | INSULIN | 19078551 | insulin, isophane |
| INSULIN | INSULIN | 36227902 | Insulin Lispro |
| BROMOCRIPTINE | BROMOCRIPTINE | 40016897 | Bromocriptine |
| GLIPIZIDE | GLIPIZIDE | 1560233 | Glipizide |
| GLIPIZIDE | GLIPIZIDE | 1560171 | Glipizide |
| METFORMIN | METFORMIN | 1503327 | Metformin |
| INSULIN | INSULIN | 1502905 | Insulin Glargine |
| METFORMIN | METFORMIN | 19086484 | Metformin |
| INSULIN | INSULIN | 46233976 | Insulin Lispro |
| INSULIN | INSULIN | 19131582 | Insulin, Glulisine, Human |

| | | | |
|---|---|---|---|
| DAPAGLIFLOZIN | DAPAGLIFLOZIN | 36247907 | dapagliflozin |
| METFORMIN | METFORMIN | 40164900 | Metformin |
| GLYBURIDE | GLYBURIDE | 19106976 | Glyburide |
| GLIMEPIRIDE | GLIMEPIRIDE | 1597775 | glimepiride |
| INSULIN | INSULIN | 19110356 | Regular Insulin, Human |
| MIGLITOL | MIGLITOL | 19029029 | miglitol |
| CANAGLIFLOZIN | CANAGLIFLOZIN | 43526467 | canagliflozin |
| GLIMEPIRIDE | GLIMEPIRIDE | 1597776 | glimepiride |
| INSULIN | INSULIN | 46233945 | Insulin Lispro |
| INSULIN | INSULIN | 46221552 | Insulin Glargine |
| INSULIN | INSULIN | 19095214 | insulin, isophane |
| GLIMEPIRIDE | GLIMEPIRIDE | 1597756 | glimepiride |
| INSULIN | INSULIN | 40166275 | Insulin Lispro |
| INSULIN | INSULIN | 19078555 | Regular Insulin, Human |
| ALBIGLUTIDE | ALBIGLUTIDE | 44816086 | albiglutide |
| EXENATIDE | EXENATIDE | 1583722 | exenatide |
| INSULIN | INSULIN | 1596955 | insulin, isophane |
| EMPAGLIFLOZIN | EMPAGLIFLOZIN | 45774753 | empagliflozin |
| TOLBUTAMIDE | TOLBUTAMIDE | 1502914 | Tolbutamide |
| METFORMIN | METFORMIN | 40135481 | Metformin |
| METFORMIN | METFORMIN | 40146999 | Metformin |
| CHLORPROPAMIDE | CHLORPROPAMIDE | 19006907 | Chlorpropamide |
| INSULIN | INSULIN | 19131582 | Insulin, Glulisine, Human |
| DULAGLUTIDE | DULAGLUTIDE | 36247186 | dulaglutide |
| GLIPIZIDE | GLIPIZIDE | 40044251 | Glipizide |
| METFORMIN | METFORMIN | 1503328 | Metformin |
| INSULIN | INSULIN | 1596999 | Regular Insulin, Human |
| INSULIN | INSULIN | 40052088 | Insulin Lispro |
| REPAGLINIDE | REPAGLINIDE | 19023424 | repaglinide |
| INSULIN | INSULIN | 19120790 | Insulin Glargine |
| INSULIN | INSULIN | 36248870 | Insulin Glargine |
| INSULIN | INSULIN | 40052088 | Insulin Lispro |
| BROMOCRIPTINE | BROMOCRIPTINE | 40016899 | Bromocriptine |
| METFORMIN | METFORMIN | 40164943 | Metformin |
| TOLAZAMIDE | TOLAZAMIDE | 19036966 | Tolazamide |
| GLIPIZIDE | GLIPIZIDE | 19085004 | Glipizide |
| SITAGLIPTIN | SITAGLIPTIN | 1580747 | sitagliptin |

| | | | |
|---|---|---|---|
| INSULIN | INSULIN | 1550028 | Insulin Lispro |
| PRAMLINTIDE | PRAMLINTIDE | 40165687 | Pramlintide |
| INSULIN | INSULIN | 19095218 | Regular Insulin, Human |
| INSULIN | INSULIN | 1550027 | Insulin Lispro |
| GLIMEPIRIDE | GLIMEPIRIDE | 1597799 | glimepiride |
| INSULIN | INSULIN | 46233945 | Insulin Lispro |
| INSULIN | INSULIN | 1597001 | Regular Insulin, Human |
| INSULIN | INSULIN | 36229086 | Insulin Glargine |
| EMPAGLIFLOZIN | EMPAGLIFLOZIN | 45774895 | empagliflozin |
| GLIMEPIRIDE | GLIMEPIRIDE | 1597756 | glimepiride |
| METFORMIN | METFORMIN | 40163929 | Metformin |
| INSULIN | INSULIN | 46234045 | Insulin, Aspart, Human |
| DAPAGLIFLOZIN | DAPAGLIFLOZIN | 44785473 | dapagliflozin |
| INSULIN | INSULIN | 19135265 | Regular Insulin, Human |
| EMPAGLIFLOZIN | EMPAGLIFLOZIN | 36246885 | empagliflozin |
| PIOGLITAZONE | PIOGLITAZONE | 1525218 | pioglitazone |
| PIOGLITAZONE | PIOGLITAZONE | 19099207 | pioglitazone |
| BROMOCRIPTINE | BROMOCRIPTINE | 19134681 | Bromocriptine |
| METFORMIN | METFORMIN | 36223138 | Metformin |
| DAPAGLIFLOZIN | DAPAGLIFLOZIN | 45775866 | dapagliflozin |
| LINAGLIPTIN | LINAGLIPTIN | 36238284 | Linagliptin |
| METFORMIN | METFORMIN | 40164897 | Metformin |
| REPAGLINIDE | REPAGLINIDE | 36240429 | repaglinide |
| DAPAGLIFLOZIN | DAPAGLIFLOZIN | 44815996 | dapagliflozin |
| ROSIGLITAZONE | ROSIGLITAZONE | 40099633 | rosiglitazone |
| INSULIN | INSULIN | 1513914 | Insulin Lispro |
| INSULIN | INSULIN | 19095214 | insulin, isophane |
| LIRAGLUTIDE | LIRAGLUTIDE | 40170911 | liraglutide |
| PIOGLITAZONE | PIOGLITAZONE | 40099898 | pioglitazone |
| PRAMLINTIDE | PRAMLINTIDE | 40165687 | Pramlintide |
| INSULIN | INSULIN | 19078555 | Regular Insulin, Human |
| GLIMEPIRIDE | GLIMEPIRIDE | 1597781 | glimepiride |
| CANAGLIFLOZIN | CANAGLIFLOZIN | 43526472 | canagliflozin |
| EMPAGLIFLOZIN | EMPAGLIFLOZIN | 45774758 | empagliflozin |
| METFORMIN | METFORMIN | 40164897 | Metformin |
| SITAGLIPTIN | SITAGLIPTIN | 40136769 | sitagliptin |

| | | | |
|---|---|---|---|
| INSULIN | INSULIN | 40184294 | Insulin, Aspart Protamine, Human |
| INSULIN | INSULIN | 19120315 | insulin, isophane |
| INSULIN | INSULIN | 19100128 | Regular Insulin, Human |
| METFORMIN | METFORMIN | 40164885 | Metformin |
| ACARBOSE | ACARBOSE | 19023063 | Acarbose |
| INSULIN | INSULIN | 19090210 | Lente Insulin, Beef |
| ALBIGLUTIDE | ALBIGLUTIDE | 44816094 | albiglutide |
| INSULIN | INSULIN | 1513849 | Insulin, Zinc, Human |
| GLIMEPIRIDE | GLIMEPIRIDE | 1597776 | glimepiride |
| ALOGLIPTIN | ALOGLIPTIN | 43013919 | alogliptin |
| INSULIN | INSULIN | 19095214 | insulin, isophane |
| INSULIN | INSULIN | 19016117 | Insulin Lispro |
| GLIPIZIDE | GLIPIZIDE | 40165977 | Glipizide |
| ROSIGLITAZONE | ROSIGLITAZONE | 19107077 | rosiglitazone |
| INSULIN | INSULIN | 46233972 | Insulin Lispro |
| INSULIN | INSULIN | 19078552 | insulin, isophane |
| INSULIN | INSULIN | 19078553 | insulin, isophane |
| INSULIN | INSULIN | 40051347 | Insulin Glargine |
| METFORMIN | METFORMIN | 1503327 | Metformin |
| INSULIN | INSULIN | 1596941 | insulin, isophane |
| CANAGLIFLOZIN | CANAGLIFLOZIN | 45774718 | canagliflozin |
| INSULIN | INSULIN | 1596999 | Regular Insulin, Human |
| METFORMIN | METFORMIN | 36239257 | Metformin |
| INSULIN | INSULIN | 1550023 | Insulin Lispro |
| ALBIGLUTIDE | ALBIGLUTIDE | 44816283 | albiglutide |
| INSULIN | INSULIN | 19058398 | Insulin Lispro |
| REPAGLINIDE | REPAGLINIDE | 19023424 | repaglinide |
| GLIPIZIDE | GLIPIZIDE | 40044253 | Glipizide |
| NATEGLINIDE | NATEGLINIDE | 19045606 | nateglinide |
| INSULIN | INSULIN | 19094122 | Regular Insulin, Human |
| LIRAGLUTIDE | LIRAGLUTIDE | 40170911 | liraglutide |
| SAXAGLIPTIN | SAXAGLIPTIN | 40166043 | saxagliptin |
| LIRAGLUTIDE | LIRAGLUTIDE | 40169222 | liraglutide |
| INSULIN | INSULIN | 19078526 | Insulin, Zinc, Human |

| | | | |
|---|---|---|---|
| EMPAGLIFLOZIN | EMPAGLIFLOZIN | 1592656 | empagliflozin |
| INSULIN | INSULIN | 19120315 | insulin, isophane |
| INSULIN | INSULIN | 19012703 | insulin, isophane |
| GLIMEPIRIDE | GLIMEPIRIDE | 19018043 | glimepiride |
| GLYBURIDE | GLYBURIDE | 1559684 | Glyburide |
| INSULIN | INSULIN | 1562586 | Insulin, Zinc, Pork |
| METFORMIN | METFORMIN | 40164929 | Metformin |
| DAPAGLIFLOZIN | DAPAGLIFLOZIN | 44785834 | dapagliflozin |
| METFORMIN | METFORMIN | 40164886 | Metformin |
| GLIMEPIRIDE | GLIMEPIRIDE | 1597796 | glimepiride |
| INSULIN | INSULIN | 19062999 | insulin, isophane |
| INSULIN | INSULIN | 1586370 | Ultralente Insulin, Human |
| LINAGLIPTIN | LINAGLIPTIN | 36238286 | Linagliptin |
| GLYBURIDE | GLYBURIDE | 1559684 | Glyburide |
| INSULIN | INSULIN | 1513881 | Insulin Lispro |
| INSULIN | INSULIN | 40166274 | Insulin Lispro |
| METFORMIN | METFORMIN | 40063354 | Metformin |
| EXENATIDE | EXENATIDE | 36238252 | exenatide |
| INSULIN | INSULIN | 1502910 | Insulin Glargine |
| DULAGLUTIDE | DULAGLUTIDE | 45774442 | dulaglutide |
| METFORMIN | METFORMIN | 1503297 | Metformin |
| ALOGLIPTIN | ALOGLIPTIN | 43013927 | alogliptin |
| PIOGLITAZONE | PIOGLITAZONE | 19099209 | pioglitazone |
| INSULIN | INSULIN | 46234049 | Insulin, Aspart, Human |
| INSULIN | INSULIN | 46233975 | Insulin Lispro |
| INSULIN | INSULIN | 1596959 | insulin, isophane |
| INSULIN | INSULIN | 36231458 | insulin, isophane |
| INSULIN | INSULIN | 1567232 | Insulin, Aspart, Human |
| EMPAGLIFLOZIN | EMPAGLIFLOZIN | 46287691 | empagliflozin |
| INSULIN | INSULIN | 1596916 | insulin, isophane |

| | | | |
|---|---|---|---|
| EMPAGLIFLOZIN | EMPAGLIFLOZIN | 1592795 | empagliflozin |
| INSULIN | INSULIN | 36231459 | Regular Insulin, Human |
| INSULIN | INSULIN | 19044971 | insulin detemir |
| BROMOCRIPTINE | BROMOCRIPTINE | 36238867 | Bromocriptine |
| ROSIGLITAZONE | ROSIGLITAZONE | 40099632 | rosiglitazone |
| INSULIN | INSULIN | 1567232 | Insulin, Aspart, Human |
| INSULIN | INSULIN | 19078559 | Insulin Glargine |
| METFORMIN | METFORMIN | 40164897 | Metformin |
| LIRAGLUTIDE | LIRAGLUTIDE | 36241903 | liraglutide |
| LIRAGLUTIDE | LIRAGLUTIDE | 36241903 | liraglutide |
| METFORMIN | METFORMIN | 40231396 | Metformin |
| CANAGLIFLOZIN | CANAGLIFLOZIN | 43526465 | canagliflozin |
| PRAMLINTIDE | PRAMLINTIDE | 40163890 | Pramlintide |
| INSULIN | INSULIN | 1513881 | Insulin Lispro |
| LIXISENATIDE | LIXISENATIDE | 44506754 | Lixisenatide |
| INSULIN | INSULIN | 19016117 | Insulin Lispro |
| ACARBOSE | ACARBOSE | 1529331 | Acarbose |
| INSULIN | INSULIN | 1586369 | Ultralente Insulin, Human |
| METFORMIN | METFORMIN | 40164930 | Metformin |
| METFORMIN | METFORMIN | 40163928 | Metformin |
| GLIPIZIDE | GLIPIZIDE | 1560171 | Glipizide |
| METFORMIN | METFORMIN | 36236481 | Metformin |
| INSULIN | INSULIN | 19095211 | Insulin Lispro |
| LINAGLIPTIN | LINAGLIPTIN | 40239216 | Linagliptin |
| INSULIN | INSULIN | 1516979 | insulin detemir |
| SAXAGLIPTIN | SAXAGLIPTIN | 40166044 | saxagliptin |
| ACARBOSE | ACARBOSE | 1529331 | Acarbose |
| INSULIN | INSULIN | 19078607 | insulin, isophane |
| PIOGLITAZONE | PIOGLITAZONE | 36236324 | pioglitazone |
| GLIPIZIDE | GLIPIZIDE | 1560171 | Glipizide |
| METFORMIN | METFORMIN | 40164930 | Metformin |
| INSULIN | INSULIN | 40176981 | Insulin, Aspart Protamine, Human |
| METFORMIN | METFORMIN | 40063353 | Metformin |
| METFORMIN | METFORMIN | 19106521 | Metformin |

| | | | |
|---|---|---|---|
| INSULIN | INSULIN | 36236032 | Insulin Lispro |
| INSULIN | INSULIN | 19078607 | insulin, isophane |
| ACARBOSE | ACARBOSE | 1529331 | Acarbose |
| GLYBURIDE | GLYBURIDE | 1559785 | Glyburide |
| INSULIN | INSULIN | 1586370 | Ultralente Insulin, Human |
| METFORMIN | METFORMIN | 36235747 | Metformin |
| GLIPIZIDE | GLIPIZIDE | 19085004 | Glipizide |
| GLIMEPIRIDE | GLIMEPIRIDE | 1597756 | glimepiride |
| INSULIN | INSULIN | 46233946 | Insulin Lispro |
| INSULIN | INSULIN | 1597001 | Regular Insulin, Human |
| METFORMIN | METFORMIN | 19088818 | Metformin |
| INSULIN | INSULIN | 1596977 | Regular Insulin, Human |
| ACARBOSE | ACARBOSE | 1529359 | Acarbose |
| GLIPIZIDE | GLIPIZIDE | 40165975 | Glipizide |
| INSULIN | INSULIN | 1586370 | Ultralente Insulin, Human |
| METFORMIN | METFORMIN | 40063353 | Metformin |
| INSULIN | INSULIN | 19078553 | insulin, isophane |
| GLIMEPIRIDE | GLIMEPIRIDE | 1597775 | glimepiride |
| INSULIN | INSULIN | 1516981 | insulin detemir |
| EXENATIDE | EXENATIDE | 1583722 | exenatide |
| INSULIN | INSULIN | 40176981 | Insulin, Aspart Protamine, Human |
| GLIMEPIRIDE | GLIMEPIRIDE | 1597756 | glimepiride |
| METFORMIN | METFORMIN | 40063353 | Metformin |
| METFORMIN | METFORMIN | 1503328 | Metformin |
| DULAGLUTIDE | DULAGLUTIDE | 45774435 | dulaglutide |
| METFORMIN | METFORMIN | 1503327 | Metformin |
| GLIMEPIRIDE | GLIMEPIRIDE | 1597781 | glimepiride |
| NATEGLINIDE | NATEGLINIDE | 19045605 | nateglinide |
| INSULIN | INSULIN | 35605622 | Regular Insulin, Human |
| INSULIN | INSULIN | 46234045 | Insulin, Aspart, Human |
| METFORMIN | METFORMIN | 1503327 | Metformin |
| BROMOCRIPTINE | BROMOCRIPTINE | 19103828 | Bromocriptine |
| METFORMIN | METFORMIN | 1503327 | Metformin |
| INSULIN | INSULIN | 19078557 | Regular Insulin, Human |
| INSULIN | INSULIN | 19078553 | insulin, isophane |

| | | | |
|---|---|---|---|
| INSULIN | INSULIN | 19120790 | Insulin Glargine |
| INSULIN | INSULIN | 19120790 | Insulin Glargine |
| GLIPIZIDE | GLIPIZIDE | 1560171 | Glipizide |
| REPAGLINIDE | REPAGLINIDE | 1516770 | repaglinide |
| GLIPIZIDE | GLIPIZIDE | 19077636 | Glipizide |
| CHLORPROPAMIDE | CHLORPROPAMIDE | 1594976 | Chlorpropamide |
| INSULIN | INSULIN | 19078553 | insulin, isophane |
| DAPAGLIFLOZIN | DAPAGLIFLOZIN | 45775865 | dapagliflozin |
| SITAGLIPTIN | SITAGLIPTIN | 40136769 | sitagliptin |
| DULAGLUTIDE | DULAGLUTIDE | 45774438 | dulaglutide |
| INSULIN | INSULIN | 40052045 | insulin, isophane |
| INSULIN | INSULIN | 19078554 | Regular Insulin, Human |
| INSULIN | INSULIN | 1586344 | insulin, isophane |
| METFORMIN | METFORMIN | 1503297 | Metformin |
| INSULIN | INSULIN | 46234045 | Insulin, Aspart, Human |
| GLIPIZIDE | GLIPIZIDE | 19117945 | Glipizide |
| INSULIN | INSULIN | 40051363 | insulin, isophane |
| BROMOCRIPTINE | BROMOCRIPTINE | 730603 | Bromocriptine |
| GLICLAZIDE | GLICLAZIDE | 19101729 | Gliclazide |
| INSULIN | INSULIN | 1586369 | Ultralente Insulin, Human |
| REPAGLINIDE | REPAGLINIDE | 19078030 | repaglinide |
| INSULIN | INSULIN | 1502905 | Insulin Glargine |
| EXENATIDE | EXENATIDE | 1583722 | exenatide |
| GLYBURIDE | GLYBURIDE | 19106976 | Glyburide |
| GLYBURIDE | GLYBURIDE | 40046204 | Glyburide |
| GLIPIZIDE | GLIPIZIDE | 1560233 | Glipizide |
| INSULIN | INSULIN | 19120790 | Insulin Glargine |
| INSULIN | INSULIN | 19014976 | insulin, isophane |
| METFORMIN | METFORMIN | 40164918 | Metformin |
| INSULIN | INSULIN | 1550023 | Insulin Lispro |
| GLIMEPIRIDE | GLIMEPIRIDE | 40059666 | glimepiride |
| INSULIN | INSULIN | 46233947 | Insulin Lispro |
| INSULIN | INSULIN | 1567198 | Insulin, Aspart, Human |
| LIRAGLUTIDE | LIRAGLUTIDE | 40169222 | liraglutide |
| INSULIN | INSULIN | 19116559 | insulin, isophane |

| | | | |
|---|---|---|---|
| EXENATIDE | EXENATIDE | 46234116 | exenatide |
| INSULIN | INSULIN | 1596977 | Regular Insulin, Human |
| METFORMIN | METFORMIN | 40063350 | Metformin |
| INSULIN | INSULIN | 19078553 | insulin, isophane |
| LIRAGLUTIDE | LIRAGLUTIDE | 40170912 | liraglutide |
| GLIMEPIRIDE | GLIMEPIRIDE | 1597756 | glimepiride |
| ACETOHEXAMIDE | ACETOHEXAMIDE | 1530014 | Acetohexamide |
| ALOGLIPTIN | ALOGLIPTIN | 43013925 | alogliptin |
| METFORMIN | METFORMIN | 40164897 | Metformin |
| INSULIN | INSULIN | 19018822 | Insulin Lispro |
| INSULIN | INSULIN | 1596955 | insulin, isophane |
| INSULIN | INSULIN | 36231458 | insulin, isophane |
| INSULIN | INSULIN | 1513849 | Insulin, Zinc, Human |
| GLIPIZIDE | GLIPIZIDE | 1560233 | Glipizide |
| INSULIN | INSULIN | 1596916 | insulin, isophane |
| GLIMEPIRIDE | GLIMEPIRIDE | 36229116 | glimepiride |
| INSULIN | INSULIN | 19110354 | insulin, isophane |
| INSULIN | INSULIN | 19123376 | Insulin Lispro |
| INSULIN | INSULIN | 36231456 | insulin, isophane |
| INSULIN | INSULIN | 46234241 | Regular Insulin, Human |
| METFORMIN | METFORMIN | 40164894 | Metformin |
| DAPAGLIFLOZIN | DAPAGLIFLOZIN | 45775864 | dapagliflozin |
| INSULIN | INSULIN | 1567198 | Insulin, Aspart, Human |
| BROMOCRIPTINE | BROMOCRIPTINE | 730548 | Bromocriptine |
| CHLORPROPAMIDE | CHLORPROPAMIDE | 1594975 | Chlorpropamide |
| METFORMIN | METFORMIN | 40063352 | Metformin |
| GLIMEPIRIDE | GLIMEPIRIDE | 1597775 | glimepiride |
| CANAGLIFLOZIN | CANAGLIFLOZIN | 43526469 | canagliflozin |
| GLIPIZIDE | GLIPIZIDE | 36235749 | Glipizide |
| INSULIN | INSULIN | 1513881 | Insulin Lispro |
| INSULIN | INSULIN | 19123376 | Insulin Lispro |
| INSULIN | INSULIN | 1596972 | insulin, isophane |
| PIOGLITAZONE | PIOGLITAZONE | 40099897 | pioglitazone |
| LIRAGLUTIDE | LIRAGLUTIDE | 40169223 | liraglutide |
| INSULIN | INSULIN | 40166274 | Insulin Lispro |

| | | | |
|---|---|---|---|
| DAPAGLIFLOZIN | DAPAGLIFLOZIN | 44785831 | dapagliflozin |
| INSULIN | INSULIN | 1567198 | Insulin, Aspart, Human |
| CHLORPROPAMIDE | CHLORPROPAMIDE | 40030456 | Chlorpropamide |
| INSULIN | INSULIN | 1596977 | Regular Insulin, Human |
| INSULIN | INSULIN | 1502905 | Insulin Glargine |
| INSULIN | INSULIN | 19116559 | insulin, isophane |
| GLIMEPIRIDE | GLIMEPIRIDE | 40059666 | glimepiride |
| GLYBURIDE | GLYBURIDE | 19077659 | Glyburide |
| INSULIN | INSULIN | 1567232 | Insulin, Aspart, Human |
| GLIMEPIRIDE | GLIMEPIRIDE | 1597756 | glimepiride |
| BROMOCRIPTINE | BROMOCRIPTINE | 40016896 | Bromocriptine |
| LINAGLIPTIN | LINAGLIPTIN | 40239216 | Linagliptin |
| INSULIN | INSULIN | 1597000 | Regular Insulin, Human |
| LIRAGLUTIDE | LIRAGLUTIDE | 40170911 | liraglutide |
| GLYBURIDE | GLYBURIDE | 1559684 | Glyburide |
| INSULIN | INSULIN | 40176981 | Insulin, Aspart Protamine, Human |
| INSULIN | INSULIN | 46221558 | Insulin Glargine |
| BROMOCRIPTINE | BROMOCRIPTINE | 730551 | Bromocriptine |
| INSULIN | INSULIN | 1513849 | Insulin, Zinc, Human |
| SITAGLIPTIN | SITAGLIPTIN | 36221232 | sitagliptin |
| BROMOCRIPTINE | BROMOCRIPTINE | 19006288 | Bromocriptine |
| INSULIN | INSULIN | 19071700 | Insulin Glargine |
| INSULIN | INSULIN | 19078552 | insulin, isophane |
| GLYBURIDE | GLYBURIDE | 40166004 | Glyburide |
| GLIPIZIDE | GLIPIZIDE | 19077637 | Glipizide |
| GLICLAZIDE | GLICLAZIDE | 40044222 | Gliclazide |
| INSULIN | INSULIN | 19080091 | Ultralente Insulin, Human |
| GLYBURIDE | GLYBURIDE | 1559784 | Glyburide |
| METFORMIN | METFORMIN | 1503327 | Metformin |
| GLIPIZIDE | GLIPIZIDE | 1560171 | Glipizide |
| INSULIN | INSULIN | 1502910 | Insulin Glargine |
| INSULIN | INSULIN | 45777023 | Regular Insulin, Human |
| SITAGLIPTIN | SITAGLIPTIN | 19125051 | sitagliptin |
| METFORMIN | METFORMIN | 1503297 | Metformin |

| | | | |
|---|---|---|---|
| METFORMIN | METFORMIN | 40231396 | Metformin |
| MIGLITOL | MIGLITOL | 40056706 | miglitol |
| METFORMIN | METFORMIN | 40164893 | Metformin |
| SITAGLIPTIN | SITAGLIPTIN | 42900896 | sitagliptin |
| INSULIN | INSULIN | 19058398 | Insulin Lispro |
| INSULIN | INSULIN | 19010204 | insulin, isophane |
| INSULIN | INSULIN | 1502905 | Insulin Glargine |
| INSULIN | INSULIN | 19078557 | Regular Insulin, Human |
| INSULIN | INSULIN | 36229086 | Insulin Glargine |
| GLIMEPIRIDE | GLIMEPIRIDE | 36234086 | glimepiride |
| INSULIN | INSULIN | 1502905 | Insulin Glargine |
| INSULIN | INSULIN | 46234045 | Insulin, Aspart, Human |
| METFORMIN | METFORMIN | 40164930 | Metformin |
| LINAGLIPTIN | LINAGLIPTIN | 42708089 | Linagliptin |
| GLYBURIDE | GLYBURIDE | 40165998 | Glyburide |
| INSULIN | INSULIN | 1596977 | Regular Insulin, Human |
| INSULIN | INSULIN | 1596999 | Regular Insulin, Human |
| MIGLITOL | MIGLITOL | 1510207 | miglitol |
| LIRAGLUTIDE | LIRAGLUTIDE | 45892686 | liraglutide |
| INSULIN | INSULIN | 19110219 | insulin, isophane |
| INSULIN | INSULIN | 1550027 | Insulin Lispro |
| PRAMLINTIDE | PRAMLINTIDE | 40163899 | Pramlintide |
| INSULIN | INSULIN | 1596960 | insulin, isophane |
| ALOGLIPTIN | ALOGLIPTIN | 43013895 | alogliptin |
| INSULIN | INSULIN | 1562586 | Insulin, Zinc, Pork |
| GLYBURIDE | GLYBURIDE | 40046204 | Glyburide |
| SITAGLIPTIN | SITAGLIPTIN | 19125042 | sitagliptin |
| EMPAGLIFLOZIN | EMPAGLIFLOZIN | 36246886 | empagliflozin |
| EXENATIDE | EXENATIDE | 19083253 | exenatide |
| INSULIN | INSULIN | 46233972 | Insulin Lispro |
| METFORMIN | METFORMIN | 1503328 | Metformin |
| INSULIN | INSULIN | 19120790 | Insulin Glargine |
| GLIPIZIDE | GLIPIZIDE | 40166816 | Glipizide |

| | | | |
|---|---|---|---|
| INSULIN | INSULIN | 1596998 | Regular Insulin, Human |
| INSULIN | INSULIN | 1567232 | Insulin, Aspart, Human |
| BROMOCRIPTINE | BROMOCRIPTINE | 730572 | Bromocriptine |
| BROMOCRIPTINE | BROMOCRIPTINE | 40164163 | Bromocriptine |
| GLYBURIDE | GLYBURIDE | 1559784 | Glyburide |
| INSULIN | INSULIN | 19078633 | Insulin, Regular, Pork |
| INSULIN | INSULIN | 1550023 | Insulin Lispro |
| TOLAZAMIDE | TOLAZAMIDE | 1502810 | Tolazamide |
| INSULIN | INSULIN | 40052088 | Insulin Lispro |
| INSULIN | INSULIN | 46234045 | Insulin, Aspart, Human |
| METFORMIN | METFORMIN | 40164948 | Metformin |
| DAPAGLIFLOZIN | DAPAGLIFLOZIN | 44816049 | dapagliflozin |
| INSULIN | INSULIN | 19078554 | Regular Insulin, Human |
| NATEGLINIDE | NATEGLINIDE | 1502830 | nateglinide |
| METFORMIN | METFORMIN | 40164930 | Metformin |
| MIGLITOL | MIGLITOL | 19029030 | miglitol |
| CHLORPROPAMIDE | CHLORPROPAMIDE | 1594978 | Chlorpropamide |
| GLIMEPIRIDE | GLIMEPIRIDE | 1597756 | glimepiride |
| INSULIN | INSULIN | 36236032 | Insulin Lispro |
| GLIPIZIDE | GLIPIZIDE | 1560233 | Glipizide |
| DULAGLUTIDE | DULAGLUTIDE | 36247186 | dulaglutide |
| METFORMIN | METFORMIN | 40143322 | Metformin |
| LINAGLIPTIN | LINAGLIPTIN | 42705856 | Linagliptin |
| GLIMEPIRIDE | GLIMEPIRIDE | 1597775 | glimepiride |
| INSULIN | INSULIN | 40166274 | Insulin Lispro |
| GLIPIZIDE | GLIPIZIDE | 19081295 | Glipizide |
| INSULIN | INSULIN | 19078555 | Regular Insulin, Human |
| DULAGLUTIDE | DULAGLUTIDE | 45774440 | dulaglutide |
| INSULIN | INSULIN | 1596977 | Regular Insulin, Human |
| METFORMIN | METFORMIN | 40231393 | Metformin |
| INSULIN | INSULIN | 36235840 | insulin, isophane |
| GLIPIZIDE | GLIPIZIDE | 40167637 | Glipizide |
| INSULIN | INSULIN | 46234045 | Insulin, Aspart, Human |
| GLIMEPIRIDE | GLIMEPIRIDE | 1597756 | glimepiride |
| GLYBURIDE | GLYBURIDE | 19045629 | Glyburide |
| INSULIN | INSULIN | 19078552 | insulin, isophane |
| METFORMIN | METFORMIN | 40063354 | Metformin |

| | | | |
|---|---|---|---|
| METFORMIN | METFORMIN | 1503327 | Metformin |
| INSULIN | INSULIN | 1596959 | insulin, isophane |
| INSULIN | INSULIN | 19095218 | Regular Insulin, Human |
| ACARBOSE | ACARBOSE | 1529331 | Acarbose |
| GLIMEPIRIDE | GLIMEPIRIDE | 1597801 | glimepiride |
| INSULIN | INSULIN | 1596977 | Regular Insulin, Human |
| INSULIN | INSULIN | 1596977 | Regular Insulin, Human |
| GLIPIZIDE | GLIPIZIDE | 19081295 | Glipizide |
| INSULIN | INSULIN | 1586345 | insulin, isophane |
| INSULIN | INSULIN | 19135265 | Regular Insulin, Human |
| INSULIN | INSULIN | 1567198 | Insulin, Aspart, Human |
| INSULIN | INSULIN | 46233975 | Insulin Lispro |
| INSULIN | INSULIN | 19078552 | insulin, isophane |
| GLYBURIDE | GLYBURIDE | 19030579 | Glyburide |
| METFORMIN | METFORMIN | 36235787 | Metformin |
| INSULIN | INSULIN | 19078554 | Regular Insulin, Human |
| METFORMIN | METFORMIN | 40164882 | Metformin |
| INSULIN | INSULIN | 19062999 | insulin, isophane |
| INSULIN | INSULIN | 19062999 | insulin, isophane |
| INSULIN | INSULIN | 19014975 | insulin, isophane |
| ALBIGLUTIDE | ALBIGLUTIDE | 44816274 | albiglutide |
| GLYBURIDE | GLYBURIDE | 1559782 | Glyburide |
| METFORMIN | METFORMIN | 1503297 | Metformin |
| INSULIN | INSULIN | 19018822 | Insulin Lispro |
| ALOGLIPTIN | ALOGLIPTIN | 43013919 | alogliptin |
| LIRAGLUTIDE | LIRAGLUTIDE | 36248956 | liraglutide |
| INSULIN | INSULIN | 40166274 | Insulin Lispro |
| INSULIN | INSULIN | 19120315 | insulin, isophane |
| GLIPIZIDE | GLIPIZIDE | 19030445 | Glipizide |
| METFORMIN | METFORMIN | 40164924 | Metformin |
| DULAGLUTIDE | DULAGLUTIDE | 45774440 | dulaglutide |
| INSULIN | INSULIN | 40166275 | Insulin Lispro |
| INSULIN | INSULIN | 36235841 | Regular Insulin, Human |

| | | | |
|---|---|---|---|
| INSULIN | INSULIN | 19116559 | insulin, isophane |
| INSULIN | INSULIN | 46233975 | Insulin Lispro |
| INSULIN | INSULIN | 1586344 | insulin, isophane |
| INSULIN | INSULIN | 19100128 | Regular Insulin, Human |
| INSULIN | INSULIN | 19120790 | Insulin Glargine |
| INSULIN | INSULIN | 19078554 | Regular Insulin, Human |
| INSULIN | INSULIN | 35604094 | Insulin Glargine |
| INSULIN | INSULIN | 46234043 | Insulin, Aspart, Human |
| INSULIN | INSULIN | 1550027 | Insulin Lispro |
| PIOGLITAZONE | PIOGLITAZONE | 36236324 | pioglitazone |
| INSULIN | INSULIN | 1516981 | insulin detemir |
| INSULIN | INSULIN | 19078526 | Insulin, Zinc, Human |
| INSULIN | INSULIN | 19116454 | insulin, isophane |
| INSULIN | INSULIN | 40184293 | Insulin, Aspart Protamine, Human |
| LIRAGLUTIDE | LIRAGLUTIDE | 40169223 | liraglutide |
| INSULIN | INSULIN | 19100128 | Regular Insulin, Human |
| INSULIN | INSULIN | 1567198 | Insulin, Aspart, Human |
| METFORMIN | METFORMIN | 40164919 | Metformin |
| METFORMIN | METFORMIN | 1503297 | Metformin |
| INSULIN | INSULIN | 46233948 | Insulin Lispro |
| INSULIN | INSULIN | 19078560 | Lente Insulin, Beef-Pork |
| METFORMIN | METFORMIN | 40164897 | Metformin |
| INSULIN | INSULIN | 19110219 | insulin, isophane |
| INSULIN | INSULIN | 19078633 | Insulin, Regular, Pork |
| ACARBOSE | ACARBOSE | 19021312 | Acarbose |
| INSULIN | INSULIN | 40051347 | Insulin Glargine |
| INSULIN | INSULIN | 19078527 | Insulin, Zinc, Human |
| INSULIN | INSULIN | 36233154 | Insulin, Glulisine, Human |
| GLIPIZIDE | GLIPIZIDE | 40166814 | Glipizide |
| INSULIN | INSULIN | 1562586 | Insulin, Zinc, Pork |
| INSULIN | INSULIN | 19116454 | insulin, isophane |

| | | | |
|---|---|---|---|
| INSULIN | INSULIN | 19078557 | Regular Insulin, Human |
| ROSIGLITAZONE | ROSIGLITAZONE | 1547505 | rosiglitazone |
| GLYBURIDE | GLYBURIDE | 1559760 | Glyburide |
| GLIPIZIDE | GLIPIZIDE | 40166814 | Glipizide |
| SITAGLIPTIN | SITAGLIPTIN | 40243627 | sitagliptin |
| BROMOCRIPTINE | BROMOCRIPTINE | 19007101 | Bromocriptine |
| EMPAGLIFLOZIN | EMPAGLIFLOZIN | 45774753 | empagliflozin |
| NATEGLINIDE | NATEGLINIDE | 1502826 | nateglinide |
| GLYBURIDE | GLYBURIDE | 1559784 | Glyburide |
| EMPAGLIFLOZIN | EMPAGLIFLOZIN | 45892179 | empagliflozin |
| INSULIN | INSULIN | 1596977 | Regular Insulin, Human |
| INSULIN | INSULIN | 19078555 | Regular Insulin, Human |
| INSULIN | INSULIN | 19010204 | insulin, isophane |
| GLYBURIDE | GLYBURIDE | 19001408 | Glyburide |
| INSULIN | INSULIN | 19078558 | Insulin, Aspart, Human |
| LIRAGLUTIDE | LIRAGLUTIDE | 40170913 | liraglutide |
| SITAGLIPTIN | SITAGLIPTIN | 19125049 | sitagliptin |
| METFORMIN | METFORMIN | 40063350 | Metformin |
| INSULIN | INSULIN | 19078553 | insulin, isophane |
| INSULIN | INSULIN | 19062999 | insulin, isophane |
| LINAGLIPTIN | LINAGLIPTIN | 40239220 | Linagliptin |
| METFORMIN | METFORMIN | 40164932 | Metformin |
| METFORMIN | METFORMIN | 1503297 | Metformin |
| METFORMIN | METFORMIN | 40231393 | Metformin |
| ROSIGLITAZONE | ROSIGLITAZONE | 19099245 | rosiglitazone |
| INSULIN | INSULIN | 19100127 | insulin, isophane |
| METFORMIN | METFORMIN | 40231395 | Metformin |
| INSULIN | INSULIN | 40166274 | Insulin Lispro |
| INSULIN | INSULIN | 40051347 | Insulin Glargine |
| GLYBURIDE | GLYBURIDE | 40165993 | Glyburide |
| BROMOCRIPTINE | BROMOCRIPTINE | 40164161 | Bromocriptine |
| METFORMIN | METFORMIN | 19088818 | Metformin |
| EXENATIDE | EXENATIDE | 19083254 | exenatide |
| GLIMEPIRIDE | GLIMEPIRIDE | 36222784 | glimepiride |

| | | | |
|---|---|---|---|
| INSULIN | INSULIN | 19095218 | Regular Insulin, Human |
| INSULIN | INSULIN | 1567198 | Insulin, Aspart, Human |
| METFORMIN | METFORMIN | 40164900 | Metformin |
| INSULIN | INSULIN | 46234045 | Insulin, Aspart, Human |
| INSULIN | INSULIN | 46233730 | Regular Insulin, Human |
| INSULIN | INSULIN | 40184293 | Insulin, Aspart Protamine, Human |
| INSULIN | INSULIN | 19120315 | insulin, isophane |
| GLIPIZIDE | GLIPIZIDE | 1560171 | Glipizide |
| INSULIN | INSULIN | 36248870 | Insulin Glargine |
| METFORMIN | METFORMIN | 40063352 | Metformin |
| METFORMIN | METFORMIN | 40164939 | Metformin |
| LIXISENATIDE | LIXISENATIDE | 44506754 | Lixisenatide |
| METFORMIN | METFORMIN | 1503297 | Metformin |
| DULAGLUTIDE | DULAGLUTIDE | 45774440 | dulaglutide |
| INSULIN | INSULIN | 40176981 | Insulin, Aspart Protamine, Human |
| METFORMIN | METFORMIN | 40164925 | Metformin |
| METFORMIN | METFORMIN | 1503297 | Metformin |
| INSULIN | INSULIN | 1567232 | Insulin, Aspart, Human |
| GLIMEPIRIDE | GLIMEPIRIDE | 19117198 | glimepiride |
| INSULIN | INSULIN | 1596956 | insulin, isophane |
| INSULIN | INSULIN | 40051347 | Insulin Glargine |
| EXENATIDE | EXENATIDE | 45774535 | exenatide |
| GLIPIZIDE | GLIPIZIDE | 40167635 | Glipizide |
| METFORMIN | METFORMIN | 40063354 | Metformin |
| METFORMIN | METFORMIN | 40146999 | Metformin |
| INSULIN | INSULIN | 36247261 | Regular Insulin, Human |
| INSULIN | INSULIN | 40176981 | Insulin, Aspart Protamine, Human |
| METFORMIN | METFORMIN | 40164896 | Metformin |
| SAXAGLIPTIN | SAXAGLIPTIN | 40166038 | saxagliptin |
| METFORMIN | METFORMIN | 40164880 | Metformin |
| INSULIN | INSULIN | 1567198 | Insulin, Aspart, Human |
| METFORMIN | METFORMIN | 1503327 | Metformin |
| INSULIN | INSULIN | 46234244 | Regular Insulin, Human |

| | | | |
|---|---|---|---|
| LIRAGLUTIDE | LIRAGLUTIDE | 36241903 | liraglutide |
| PIOGLITAZONE | PIOGLITAZONE | 1525215 | pioglitazone |
| BROMOCRIPTINE | BROMOCRIPTINE | 40016899 | Bromocriptine |
| INSULIN | INSULIN | 46221558 | Insulin Glargine |
| SAXAGLIPTIN | SAXAGLIPTIN | 40166044 | saxagliptin |
| METFORMIN | METFORMIN | 40164938 | Metformin |
| TOLAZAMIDE | TOLAZAMIDE | 1502819 | Tolazamide |
| INSULIN | INSULIN | 19058398 | Insulin Lispro |
| GLYBURIDE | GLYBURIDE | 19045629 | Glyburide |
| INSULIN | INSULIN | 19078555 | Regular Insulin, Human |
| METFORMIN | METFORMIN | 1503328 | Metformin |
| INSULIN | INSULIN | 1550023 | Insulin Lispro |
| INSULIN | INSULIN | 19078555 | Regular Insulin, Human |
| GLIMEPIRIDE | GLIMEPIRIDE | 1597796 | glimepiride |
| METFORMIN | METFORMIN | 1503297 | Metformin |
| GLIMEPIRIDE | GLIMEPIRIDE | 1597756 | glimepiride |
| INSULIN | INSULIN | 46233975 | Insulin Lispro |
| EXENATIDE | EXENATIDE | 19083253 | exenatide |
| METFORMIN | METFORMIN | 40164947 | Metformin |
| INSULIN | INSULIN | 19078559 | Insulin Glargine |
| INSULIN | INSULIN | 1597012 | Regular Insulin, Human |
| INSULIN | INSULIN | 40051348 | Insulin Glargine |
| INSULIN | INSULIN | 1502910 | Insulin Glargine |
| GLIMEPIRIDE | GLIMEPIRIDE | 43011498 | glimepiride |
| METFORMIN | METFORMIN | 40164922 | Metformin |
| INSULIN | INSULIN | 40051347 | Insulin Glargine |
| SITAGLIPTIN | SITAGLIPTIN | 19125042 | sitagliptin |
| METFORMIN | METFORMIN | 40164918 | Metformin |
| GLYBURIDE | GLYBURIDE | 19030579 | Glyburide |
| INSULIN | INSULIN | 46221557 | Insulin Glargine |
| CANAGLIFLOZIN | CANAGLIFLOZIN | 43526471 | canagliflozin |
| GLIPIZIDE | GLIPIZIDE | 40044224 | Glipizide |
| INSULIN | INSULIN | 1567232 | Insulin, Aspart, Human |
| CHLORPROPAMIDE | CHLORPROPAMIDE | 1594975 | Chlorpropamide |
| INSULIN | INSULIN | 1596972 | insulin, isophane |
| GLYBURIDE | GLYBURIDE | 1559684 | Glyburide |
| INSULIN | INSULIN | 40051347 | Insulin Glargine |

| | | | |
|---|---|---|---|
| GLIMEPIRIDE | GLIMEPIRIDE | 1597776 | glimepiride |
| METFORMIN | METFORMIN | 40164892 | Metformin |
| ALOGLIPTIN | ALOGLIPTIN | 43013884 | alogliptin |
| METFORMIN | METFORMIN | 40164898 | Metformin |
| EXENATIDE | EXENATIDE | 19083253 | exenatide |
| METFORMIN | METFORMIN | 40171451 | Metformin |
| INSULIN | INSULIN | 19090210 | Lente Insulin, Beef |
| GLYBURIDE | GLYBURIDE | 19030575 | Glyburide |
| GLYBURIDE | GLYBURIDE | 1559785 | Glyburide |
| GLYBURIDE | GLYBURIDE | 40046204 | Glyburide |
| METFORMIN | METFORMIN | 40164932 | Metformin |
| METFORMIN | METFORMIN | 42708177 | Metformin |
| INSULIN | INSULIN | 19078633 | Insulin, Regular, Pork |
| INSULIN | INSULIN | 40184293 | Insulin, Aspart Protamine, Human |
| DULAGLUTIDE | DULAGLUTIDE | 45774440 | dulaglutide |
| GLIPIZIDE | GLIPIZIDE | 1560171 | Glipizide |
| EXENATIDE | EXENATIDE | 42708079 | exenatide |
| PIOGLITAZONE | PIOGLITAZONE | 1525215 | pioglitazone |
| INSULIN | INSULIN | 1597012 | Regular Insulin, Human |
| INSULIN | INSULIN | 19116453 | insulin, isophane |
| EXENATIDE | EXENATIDE | 19083254 | exenatide |
| INSULIN | INSULIN | 19044971 | insulin detemir |
| INSULIN | INSULIN | 19062999 | insulin, isophane |
| GLIPIZIDE | GLIPIZIDE | 19081295 | Glipizide |
| EXENATIDE | EXENATIDE | 46234118 | exenatide |
| INSULIN | INSULIN | 40052088 | Insulin Lispro |
| METFORMIN | METFORMIN | 40164929 | Metformin |
| INSULIN | INSULIN | 1597001 | Regular Insulin, Human |
| INSULIN | INSULIN | 19131582 | Insulin, Glulisine, Human |
| METFORMIN | METFORMIN | 36236322 | Metformin |
| TOLAZAMIDE | TOLAZAMIDE | 36230840 | Tolazamide |
| INSULIN | INSULIN | 46221555 | Insulin Glargine |

| | | | |
|---|---|---|---|
| INSULIN | INSULIN | 1567232 | Insulin, Aspart, Human |
| INSULIN | INSULIN | 36222190 | insulin detemir |
| DAPAGLIFLOZIN | DAPAGLIFLOZIN | 36248559 | dapagliflozin |
| INSULIN | INSULIN | 19078553 | insulin, isophane |
| INSULIN | INSULIN | 19078632 | Insulin, Regular, Pork |
| METFORMIN | METFORMIN | 40164929 | Metformin |
| INSULIN | INSULIN | 19120315 | insulin, isophane |
| INSULIN | INSULIN | 35602728 | insulin degludec |
| GLYBURIDE | GLYBURIDE | 1559784 | Glyburide |
| NATEGLINIDE | NATEGLINIDE | 36223777 | nateglinide |
| ROSIGLITAZONE | ROSIGLITAZONE | 19079465 | rosiglitazone |
| INSULIN | INSULIN | 1718711 | insulin degludec |
| INSULIN | INSULIN | 19078555 | Regular Insulin, Human |
| BROMOCRIPTINE | BROMOCRIPTINE | 730548 | Bromocriptine |
| INSULIN | INSULIN | 1567232 | Insulin, Aspart, Human |
| DAPAGLIFLOZIN | DAPAGLIFLOZIN | 45775867 | dapagliflozin |
| BROMOCRIPTINE | BROMOCRIPTINE | 730548 | Bromocriptine |
| LINAGLIPTIN | LINAGLIPTIN | 40239218 | Linagliptin |
| EXENATIDE | EXENATIDE | 45774533 | exenatide |
| DAPAGLIFLOZIN | DAPAGLIFLOZIN | 45776000 | dapagliflozin |
| GLIPIZIDE | GLIPIZIDE | 40165969 | Glipizide |
| INSULIN | INSULIN | 19135276 | Insulin Glargine |
| INSULIN | INSULIN | 36231459 | Regular Insulin, Human |
| METFORMIN | METFORMIN | 40063352 | Metformin |
| METFORMIN | METFORMIN | 40229047 | Metformin |
| SAXAGLIPTIN | SAXAGLIPTIN | 40166038 | saxagliptin |
| INSULIN | INSULIN | 46234045 | Insulin, Aspart, Human |
| INSULIN | INSULIN | 19012679 | Regular Insulin, Human |
| GLIPIZIDE | GLIPIZIDE | 1560171 | Glipizide |
| SITAGLIPTIN | SITAGLIPTIN | 19125042 | sitagliptin |
| METFORMIN | METFORMIN | 40164929 | Metformin |

| | | | |
|---|---|---|---|
| GLIMEPIRIDE | GLIMEPIRIDE | 19117200 | glimepiride |
| METFORMIN | METFORMIN | 40164900 | Metformin |
| METFORMIN | METFORMIN | 19106521 | Metformin |
| BROMOCRIPTINE | BROMOCRIPTINE | 40016896 | Bromocriptine |
| GLIMEPIRIDE | GLIMEPIRIDE | 1597773 | glimepiride |
| INSULIN | INSULIN | 19110219 | insulin, isophane |
| INSULIN | INSULIN | 1567198 | Insulin, Aspart, Human |
| ALBIGLUTIDE | ALBIGLUTIDE | 44816274 | albiglutide |
| GLYBURIDE | GLYBURIDE | 40046211 | Glyburide |
| INSULIN | INSULIN | 19129437 | insulin human, rDNA origin |
| GLYBURIDE | GLYBURIDE | 1559684 | Glyburide |
| METFORMIN | METFORMIN | 40164929 | Metformin |
| GLYBURIDE | GLYBURIDE | 1559782 | Glyburide |
| GLIPIZIDE | GLIPIZIDE | 19085004 | Glipizide |
| ACARBOSE | ACARBOSE | 1529360 | Acarbose |
| INSULIN | INSULIN | 19078557 | Regular Insulin, Human |
| INSULIN | INSULIN | 36235840 | insulin, isophane |
| METFORMIN | METFORMIN | 40164923 | Metformin |
| SITAGLIPTIN | SITAGLIPTIN | 19125042 | sitagliptin |
| METFORMIN | METFORMIN | 40164942 | Metformin |
| METFORMIN | METFORMIN | 1503328 | Metformin |
| GLYBURIDE | GLYBURIDE | 1559784 | Glyburide |
| METFORMIN | METFORMIN | 40164902 | Metformin |
| INSULIN | INSULIN | 1596977 | Regular Insulin, Human |
| INSULIN | INSULIN | 1550027 | Insulin Lispro |
| GLYBURIDE | GLYBURIDE | 40046201 | Glyburide |
| INSULIN | INSULIN | 40052088 | Insulin Lispro |
| TOLBUTAMIDE | TOLBUTAMIDE | 1502855 | Tolbutamide |
| GLIPIZIDE | GLIPIZIDE | 36241808 | Glipizide |
| METFORMIN | METFORMIN | 40231396 | Metformin |
| DAPAGLIFLOZIN | DAPAGLIFLOZIN | 44816049 | dapagliflozin |
| METFORMIN | METFORMIN | 1503328 | Metformin |
| INSULIN | INSULIN | 40166274 | Insulin Lispro |
| INSULIN | INSULIN | 19131582 | Insulin, Glulisine, Human |
| ACETOHEXAMIDE | ACETOHEXAMIDE | 1530014 | Acetohexamide |
| GLIPIZIDE | GLIPIZIDE | 40166816 | Glipizide |

| | | | |
|---|---|---|---|
| METFORMIN | METFORMIN | 40164883 | Metformin |
| INSULIN | INSULIN | 1544838 | Insulin, Glulisine, Human |
| INSULIN | INSULIN | 46234049 | Insulin, Aspart, Human |
| EXENATIDE | EXENATIDE | 19083254 | exenatide |
| INSULIN | INSULIN | 1516979 | insulin detemir |
| METFORMIN | METFORMIN | 40173237 | Metformin |
| LIRAGLUTIDE | LIRAGLUTIDE | 40170913 | liraglutide |
| INSULIN | INSULIN | 19116453 | insulin, isophane |
| GLYBURIDE | GLYBURIDE | 40166003 | Glyburide |
| METFORMIN | METFORMIN | 1503327 | Metformin |
| INSULIN | INSULIN | 36236032 | Insulin Lispro |
| INSULIN | INSULIN | 1513912 | Insulin Lispro |
| GLYBURIDE | GLYBURIDE | 1559684 | Glyburide |
| INSULIN | INSULIN | 1567198 | Insulin, Aspart, Human |
| INSULIN | INSULIN | 19078553 | insulin, isophane |
| GLIPIZIDE | GLIPIZIDE | 1560171 | Glipizide |
| INSULIN | INSULIN | 19110356 | Regular Insulin, Human |
| GLIPIZIDE | GLIPIZIDE | 40166816 | Glipizide |
| DAPAGLIFLOZIN | DAPAGLIFLOZIN | 36248558 | dapagliflozin |
| GLIPIZIDE | GLIPIZIDE | 40167636 | Glipizide |
| METFORMIN | METFORMIN | 40164939 | Metformin |
| INSULIN | INSULIN | 46221552 | Insulin Glargine |
| INSULIN | INSULIN | 1550023 | Insulin Lispro |
| INSULIN | INSULIN | 40176981 | Insulin, Aspart Protamine, Human |
| GLIMEPIRIDE | GLIMEPIRIDE | 1597757 | glimepiride |
| ACETOHEXAMIDE | ACETOHEXAMIDE | 36235641 | Acetohexamide |
| GLIMEPIRIDE | GLIMEPIRIDE | 1597776 | glimepiride |
| METFORMIN | METFORMIN | 40164924 | Metformin |
| PIOGLITAZONE | PIOGLITAZONE | 19099207 | pioglitazone |
| ALBIGLUTIDE | ALBIGLUTIDE | 36246906 | albiglutide |
| SAXAGLIPTIN | SAXAGLIPTIN | 36224321 | saxagliptin |
| INSULIN | INSULIN | 19120315 | insulin, isophane |
| GLIPIZIDE | GLIPIZIDE | 19100559 | Glipizide |

| | | | |
|---|---|---|---|
| LINAGLIPTIN | LINAGLIPTIN | 40239220 | Linagliptin |
| METFORMIN | METFORMIN | 40164928 | Metformin |
| INSULIN | INSULIN | 1567232 | Insulin, Aspart, Human |
| INSULIN | INSULIN | 19058398 | Insulin Lispro |
| INSULIN | INSULIN | 1596977 | Regular Insulin, Human |
| LIRAGLUTIDE | LIRAGLUTIDE | 36241903 | liraglutide |
| METFORMIN | METFORMIN | 40164938 | Metformin |
| ACARBOSE | ACARBOSE | 19040970 | Acarbose |
| GLYBURIDE | GLYBURIDE | 19060409 | Glyburide |
| GLIMEPIRIDE | GLIMEPIRIDE | 1597759 | glimepiride |
| INSULIN | INSULIN | 19078551 | insulin, isophane |
| SITAGLIPTIN | SITAGLIPTIN | 40136770 | sitagliptin |
| INSULIN | INSULIN | 19018822 | Insulin Lispro |
| METFORMIN | METFORMIN | 36242511 | Metformin |
| GLIPIZIDE | GLIPIZIDE | 19081295 | Glipizide |
| GLIPIZIDE | GLIPIZIDE | 19085004 | Glipizide |
| METFORMIN | METFORMIN | 40063352 | Metformin |
| SITAGLIPTIN | SITAGLIPTIN | 19125045 | sitagliptin |
| SITAGLIPTIN | SITAGLIPTIN | 19125042 | sitagliptin |
| INSULIN | INSULIN | 19111797 | insulin, isophane |
| ACARBOSE | ACARBOSE | 1529331 | Acarbose |
| INSULIN | INSULIN | 1596952 | insulin, isophane |
| EMPAGLIFLOZIN | EMPAGLIFLOZIN | 45774759 | empagliflozin |
| INSULIN | INSULIN | 1516981 | insulin detemir |
| GLIPIZIDE | GLIPIZIDE | 1560171 | Glipizide |
| ROSIGLITAZONE | ROSIGLITAZONE | 36229365 | rosiglitazone |
| PRAMLINTIDE | PRAMLINTIDE | 40163890 | Pramlintide |
| INSULIN | INSULIN | 35602724 | insulin degludec |
| INSULIN | INSULIN | 19018822 | Insulin Lispro |
| NATEGLINIDE | NATEGLINIDE | 1502827 | nateglinide |
| INSULIN | INSULIN | 19062999 | insulin, isophane |
| ROSIGLITAZONE | ROSIGLITAZONE | 19099246 | rosiglitazone |
| SITAGLIPTIN | SITAGLIPTIN | 19125045 | sitagliptin |
| INSULIN | INSULIN | 40051348 | Insulin Glargine |

| | | | |
|---|---|---|---|
| EXENATIDE | EXENATIDE | 1583727 | exenatide |
| ALBIGLUTIDE | ALBIGLUTIDE | 44816273 | albiglutide |
| INSULIN | INSULIN | 40166274 | Insulin Lispro |
| PRAMLINTIDE | PRAMLINTIDE | 40165685 | Pramlintide |
| INSULIN | INSULIN | 1550023 | Insulin Lispro |
| INSULIN | INSULIN | 19018822 | Insulin Lispro |
| INSULIN | INSULIN | 19061670 | Regular Insulin, Human |
| ALOGLIPTIN | ALOGLIPTIN | 43013917 | alogliptin |
| INSULIN | INSULIN | 1586370 | Ultralente Insulin, Human |
| REPAGLINIDE | REPAGLINIDE | 1516771 | repaglinide |
| INSULIN | INSULIN | 1513912 | Insulin Lispro |
| GLYBURIDE | GLYBURIDE | 19077684 | Glyburide |
| INSULIN | INSULIN | 36229086 | Insulin Glargine |
| INSULIN | INSULIN | 46233728 | Regular Insulin, Human |
| INSULIN | INSULIN | 1596977 | Regular Insulin, Human |
| EMPAGLIFLOZIN | EMPAGLIFLOZIN | 45892179 | empagliflozin |
| INSULIN | INSULIN | 19078555 | Regular Insulin, Human |
| INSULIN | INSULIN | 19078557 | Regular Insulin, Human |
| METFORMIN | METFORMIN | 40173237 | Metformin |
| INSULIN | INSULIN | 46233946 | Insulin Lispro |
| INSULIN | INSULIN | 19012679 | Regular Insulin, Human |
| SITAGLIPTIN | SITAGLIPTIN | 40243628 | sitagliptin |
| GLIPIZIDE | GLIPIZIDE | 19134642 | Glipizide |
| INSULIN | INSULIN | 1516981 | insulin detemir |
| INSULIN | INSULIN | 36231450 | Insulin, Aspart, Human |
| EXENATIDE | EXENATIDE | 19135404 | exenatide |
| BROMOCRIPTINE | BROMOCRIPTINE | 40016898 | Bromocriptine |
| GLIPIZIDE | GLIPIZIDE | 36218767 | Glipizide |
| INSULIN | INSULIN | 1596955 | insulin, isophane |
| METFORMIN | METFORMIN | 40164925 | Metformin |
| INSULIN | INSULIN | 46234045 | Insulin, Aspart, Human |
| ROSIGLITAZONE | ROSIGLITAZONE | 1547504 | rosiglitazone |
| INSULIN | INSULIN | 1516978 | insulin detemir |
| INSULIN | INSULIN | 46233970 | Insulin Lispro |
| GLIMEPIRIDE | GLIMEPIRIDE | 19023585 | glimepiride |

| | | | |
|---|---|---|---|
| INSULIN | INSULIN | 1567232 | Insulin, Aspart, Human |
| METFORMIN | METFORMIN | 36235746 | Metformin |
| GLIMEPIRIDE | GLIMEPIRIDE | 1597776 | glimepiride |
| PIOGLITAZONE | PIOGLITAZONE | 1525215 | pioglitazone |
| LIRAGLUTIDE | LIRAGLUTIDE | 40170911 | liraglutide |
| DULAGLUTIDE | DULAGLUTIDE | 45774438 | dulaglutide |
| EXENATIDE | EXENATIDE | 1583722 | exenatide |
| REPAGLINIDE | REPAGLINIDE | 19047663 | repaglinide |
| DAPAGLIFLOZIN | DAPAGLIFLOZIN | 44785477 | dapagliflozin |
| SITAGLIPTIN | SITAGLIPTIN | 19125050 | sitagliptin |
| LINAGLIPTIN | LINAGLIPTIN | 42708089 | Linagliptin |
| EXENATIDE | EXENATIDE | 1583727 | exenatide |
| INSULIN | INSULIN | 19078556 | Regular Insulin, Human |
| DULAGLUTIDE | DULAGLUTIDE | 45774438 | dulaglutide |
| METFORMIN | METFORMIN | 40164897 | Metformin |
| METFORMIN | METFORMIN | 40164893 | Metformin |
| REPAGLINIDE | REPAGLINIDE | 36240429 | repaglinide |
| INSULIN | INSULIN | 1567232 | Insulin, Aspart, Human |
| INSULIN | INSULIN | 19100127 | insulin, isophane |
| ALOGLIPTIN | ALOGLIPTIN | 43013929 | alogliptin |
| ACARBOSE | ACARBOSE | 1529331 | Acarbose |
| METFORMIN | METFORMIN | 19086484 | Metformin |
| INSULIN | INSULIN | 19112791 | Insulin, Glulisine, Human |
| GLYBURIDE | GLYBURIDE | 19115458 | Glyburide |
| INSULIN | INSULIN | 36222190 | insulin detemir |
| INSULIN | INSULIN | 1597000 | Regular Insulin, Human |
| INSULIN | INSULIN | 1502910 | Insulin Glargine |
| METFORMIN | METFORMIN | 40164893 | Metformin |
| INSULIN | INSULIN | 46233970 | Insulin Lispro |
| ROSIGLITAZONE | ROSIGLITAZONE | 1547554 | rosiglitazone |
| ALOGLIPTIN | ALOGLIPTIN | 43013924 | alogliptin |
| MIGLITOL | MIGLITOL | 1510208 | miglitol |
| LIRAGLUTIDE | LIRAGLUTIDE | 40170913 | liraglutide |
| EXENATIDE | EXENATIDE | 1583730 | exenatide |
| GLYBURIDE | GLYBURIDE | 1559782 | Glyburide |
| METFORMIN | METFORMIN | 40164897 | Metformin |

| | | | |
|---|---|---|---|
| METFORMIN | METFORMIN | 40153895 | Metformin |
| INSULIN | INSULIN | 19110357 | Regular Insulin, Human |
| INSULIN | INSULIN | 36229086 | Insulin Glargine |
| GLIMEPIRIDE | GLIMEPIRIDE | 1597756 | glimepiride |
| INSULIN | INSULIN | 1597000 | Regular Insulin, Human |
| DAPAGLIFLOZIN | DAPAGLIFLOZIN | 45775388 | dapagliflozin |
| GLYBURIDE | GLYBURIDE | 1559684 | Glyburide |
| INSULIN | INSULIN | 19116455 | insulin, isophane |
| LIRAGLUTIDE | LIRAGLUTIDE | 40170913 | liraglutide |
| INSULIN | INSULIN | 19110354 | insulin, isophane |
| METFORMIN | METFORMIN | 40164881 | Metformin |
| EXENATIDE | EXENATIDE | 19083254 | exenatide |
| GLYBURIDE | GLYBURIDE | 1559785 | Glyburide |
| INSULIN | INSULIN | 46221552 | Insulin Glargine |
| GLIPIZIDE | GLIPIZIDE | 1560171 | Glipizide |
| INSULIN | INSULIN | 1596959 | insulin, isophane |
| PIOGLITAZONE | PIOGLITAZONE | 19099208 | pioglitazone |
| INSULIN | INSULIN | 19078553 | insulin, isophane |
| GLIPIZIDE | GLIPIZIDE | 19081295 | Glipizide |
| EXENATIDE | EXENATIDE | 19083253 | exenatide |
| EXENATIDE | EXENATIDE | 46234118 | exenatide |
| GLIMEPIRIDE | GLIMEPIRIDE | 36234086 | glimepiride |
| METFORMIN | METFORMIN | 40164897 | Metformin |
| INSULIN | INSULIN | 40176981 | Insulin, Aspart Protamine, Human |
| MIGLITOL | MIGLITOL | 19029061 | miglitol |
| GLIPIZIDE | GLIPIZIDE | 19081295 | Glipizide |
| DAPAGLIFLOZIN | DAPAGLIFLOZIN | 45776000 | dapagliflozin |
| LIRAGLUTIDE | LIRAGLUTIDE | 40169223 | liraglutide |
| GLIMEPIRIDE | GLIMEPIRIDE | 1597757 | glimepiride |
| METFORMIN | METFORMIN | 40164901 | Metformin |
| INSULIN | INSULIN | 36227902 | Insulin Lispro |
| INSULIN | INSULIN | 19100127 | insulin, isophane |
| EXENATIDE | EXENATIDE | 36234709 | exenatide |
| INSULIN | INSULIN | 1502905 | Insulin Glargine |

| | | | |
|---|---|---|---|
| INSULIN | INSULIN | 19078553 | insulin, isophane |
| METFORMIN | METFORMIN | 40164938 | Metformin |
| INSULIN | INSULIN | 36229086 | Insulin Glargine |
| ALBIGLUTIDE | ALBIGLUTIDE | 44816093 | albiglutide |
| NATEGLINIDE | NATEGLINIDE | 19045605 | nateglinide |
| PRAMLINTIDE | PRAMLINTIDE | 40163899 | Pramlintide |
| INSULIN | INSULIN | 40051348 | Insulin Glargine |
| EXENATIDE | EXENATIDE | 19083254 | exenatide |
| INSULIN | INSULIN | 19078554 | Regular Insulin, Human |
| CANAGLIFLOZIN | CANAGLIFLOZIN | 45774717 | canagliflozin |
| EXENATIDE | EXENATIDE | 46234118 | exenatide |
| METFORMIN | METFORMIN | 40117785 | Metformin |
| GLYBURIDE | GLYBURIDE | 40166003 | Glyburide |
| METFORMIN | METFORMIN | 40164946 | Metformin |
| METFORMIN | METFORMIN | 40231393 | Metformin |
| METFORMIN | METFORMIN | 40063350 | Metformin |
| INSULIN | INSULIN | 19078554 | Regular Insulin, Human |
| INSULIN | INSULIN | 19100127 | insulin, isophane |
| INSULIN | INSULIN | 19009384 | insulin, isophane |
| METFORMIN | METFORMIN | 40164886 | Metformin |
| EMPAGLIFLOZIN | EMPAGLIFLOZIN | 45774754 | empagliflozin |
| LINAGLIPTIN | LINAGLIPTIN | 40239220 | Linagliptin |
| METFORMIN | METFORMIN | 40164893 | Metformin |
| GLIMEPIRIDE | GLIMEPIRIDE | 1597756 | glimepiride |
| GLICLAZIDE | GLICLAZIDE | 19087996 | Gliclazide |
| GLIMEPIRIDE | GLIMEPIRIDE | 19023585 | glimepiride |
| METFORMIN | METFORMIN | 40163928 | Metformin |
| INSULIN | INSULIN | 40166274 | Insulin Lispro |
| GLIMEPIRIDE | GLIMEPIRIDE | 36222785 | glimepiride |
| INSULIN | INSULIN | 1596959 | insulin, isophane |
| METFORMIN | METFORMIN | 40164897 | Metformin |

| | | | |
|---|---|---|---|
| INSULIN | INSULIN | 19131582 | Insulin, Glulisine, Human |
| BROMOCRIPTINE | BROMOCRIPTINE | 40016886 | Bromocriptine |
| INSULIN | INSULIN | 19078559 | Insulin Glargine |
| INSULIN | INSULIN | 35602728 | insulin degludec |
| INSULIN | INSULIN | 19078527 | Insulin, Zinc, Human |
| INSULIN | INSULIN | 1596999 | Regular Insulin, Human |
| NATEGLINIDE | NATEGLINIDE | 19107110 | nateglinide |
| METFORMIN | METFORMIN | 40164898 | Metformin |
| DULAGLUTIDE | DULAGLUTIDE | 36247185 | dulaglutide |
| MIGLITOL | MIGLITOL | 19067232 | miglitol |
| TOLAZAMIDE | TOLAZAMIDE | 1502811 | Tolazamide |
| GLIPIZIDE | GLIPIZIDE | 19030445 | Glipizide |
| INSULIN | INSULIN | 36222190 | insulin detemir |
| GLYBURIDE | GLYBURIDE | 1559782 | Glyburide |
| LINAGLIPTIN | LINAGLIPTIN | 42705862 | Linagliptin |
| METFORMIN | METFORMIN | 40164896 | Metformin |
| SITAGLIPTIN | SITAGLIPTIN | 19125042 | sitagliptin |
| GLYBURIDE | GLYBURIDE | 40165997 | Glyburide |
| METFORMIN | METFORMIN | 40063350 | Metformin |
| INSULIN | INSULIN | 40166275 | Insulin Lispro |
| METFORMIN | METFORMIN | 40164938 | Metformin |
| METFORMIN | METFORMIN | 1503328 | Metformin |
| GLIMEPIRIDE | GLIMEPIRIDE | 40059665 | glimepiride |
| GLYBURIDE | GLYBURIDE | 1559684 | Glyburide |
| GLIMEPIRIDE | GLIMEPIRIDE | 36214984 | glimepiride |
| METFORMIN | METFORMIN | 36235753 | Metformin |
| LIRAGLUTIDE | LIRAGLUTIDE | 45892686 | liraglutide |
| METFORMIN | METFORMIN | 1503328 | Metformin |
| ROSIGLITAZONE | ROSIGLITAZONE | 19099055 | rosiglitazone |
| SITAGLIPTIN | SITAGLIPTIN | 19125048 | sitagliptin |
| INSULIN | INSULIN | 46233950 | Insulin Lispro |
| NATEGLINIDE | NATEGLINIDE | 19045606 | nateglinide |
| GLIPIZIDE | GLIPIZIDE | 19030445 | Glipizide |
| INSULIN | INSULIN | 19112791 | Insulin, Glulisine, Human |
| METFORMIN | METFORMIN | 1503297 | Metformin |

| | | | |
|---|---|---|---|
| ACARBOSE | ACARBOSE | 1529331 | Acarbose |
| INSULIN | INSULIN | 35605622 | Regular Insulin, Human |
| BROMOCRIPTINE | BROMOCRIPTINE | 730548 | Bromocriptine |
| METFORMIN | METFORMIN | 40164930 | Metformin |
| METFORMIN | METFORMIN | 40063356 | Metformin |
| DULAGLUTIDE | DULAGLUTIDE | 45774442 | dulaglutide |
| METFORMIN | METFORMIN | 40163926 | Metformin |
| GLIMEPIRIDE | GLIMEPIRIDE | 1597756 | glimepiride |
| INSULIN | INSULIN | 40052088 | Insulin Lispro |
| PIOGLITAZONE | PIOGLITAZONE | 1525215 | pioglitazone |
| INSULIN | INSULIN | 46221557 | Insulin Glargine |
| INSULIN | INSULIN | 36235841 | Regular Insulin, Human |
| PIOGLITAZONE | PIOGLITAZONE | 1525243 | pioglitazone |
| GLYBURIDE | GLYBURIDE | 40166002 | Glyburide |
| EXENATIDE | EXENATIDE | 1583727 | exenatide |
| METFORMIN | METFORMIN | 36223145 | Metformin |
| INSULIN | INSULIN | 1550028 | Insulin Lispro |
| GLYBURIDE | GLYBURIDE | 19077682 | Glyburide |
| INSULIN | INSULIN | 1596977 | Regular Insulin, Human |
| INSULIN | INSULIN | 40166274 | Insulin Lispro |
| INSULIN | INSULIN | 19104173 | insulin, isophane |
| INSULIN | INSULIN | 19095211 | Insulin Lispro |
| ACARBOSE | ACARBOSE | 1529331 | Acarbose |
| METFORMIN | METFORMIN | 40164882 | Metformin |
| GLYBURIDE | GLYBURIDE | 1559684 | Glyburide |
| GLYBURIDE | GLYBURIDE | 19065480 | Glyburide |
| GLIMEPIRIDE | GLIMEPIRIDE | 19117200 | glimepiride |
| EXENATIDE | EXENATIDE | 42708079 | exenatide |
| METFORMIN | METFORMIN | 36236323 | Metformin |
| ROSIGLITAZONE | ROSIGLITAZONE | 1547505 | rosiglitazone |
| GLIPIZIDE | GLIPIZIDE | 1560171 | Glipizide |
| CHLORPROPAMIDE | CHLORPROPAMIDE | 1594976 | Chlorpropamide |
| DULAGLUTIDE | DULAGLUTIDE | 45774440 | dulaglutide |
| CANAGLIFLOZIN | CANAGLIFLOZIN | 43526465 | canagliflozin |
| METFORMIN | METFORMIN | 36235747 | Metformin |
| INSULIN | INSULIN | 1502905 | Insulin Glargine |

| | | | |
|---|---|---|---|
| INSULIN | INSULIN | 19078553 | insulin, isophane |
| GLYBURIDE | GLYBURIDE | 1559684 | Glyburide |
| INSULIN | INSULIN | 19110357 | Regular Insulin, Human |
| EXENATIDE | EXENATIDE | 19083253 | exenatide |
| INSULIN | INSULIN | 19078553 | insulin, isophane |
| INSULIN | INSULIN | 1562586 | Insulin, Zinc, Pork |
| METFORMIN | METFORMIN | 1503327 | Metformin |
| NATEGLINIDE | NATEGLINIDE | 19110958 | nateglinide |
| INSULIN | INSULIN | 40052047 | insulin, isophane |
| REPAGLINIDE | REPAGLINIDE | 40098705 | repaglinide |
| INSULIN | INSULIN | 19090205 | Lente Insulin, Beef |
| INSULIN | INSULIN | 19078553 | insulin, isophane |
| GLYBURIDE | GLYBURIDE | 36213841 | Glyburide |
| METFORMIN | METFORMIN | 40163922 | Metformin |
| PIOGLITAZONE | PIOGLITAZONE | 1525220 | pioglitazone |
| INSULIN | INSULIN | 1596953 | insulin, isophane |
| INSULIN | INSULIN | 1550028 | Insulin Lispro |
| EXENATIDE | EXENATIDE | 46234118 | exenatide |
| METFORMIN | METFORMIN | 40163926 | Metformin |
| INSULIN | INSULIN | 35602728 | insulin degludec |
| METFORMIN | METFORMIN | 40164932 | Metformin |
| INSULIN | INSULIN | 19010204 | insulin, isophane |
| NATEGLINIDE | NATEGLINIDE | 1502827 | nateglinide |
| INSULIN | INSULIN | 19095218 | Regular Insulin, Human |
| METFORMIN | METFORMIN | 1503327 | Metformin |
| GLYBURIDE | GLYBURIDE | 1559684 | Glyburide |
| GLIPIZIDE | GLIPIZIDE | 40167636 | Glipizide |
| INSULIN | INSULIN | 35602726 | insulin degludec |
| INSULIN | INSULIN | 19120790 | Insulin Glargine |
| GLIPIZIDE | GLIPIZIDE | 1560233 | Glipizide |
| INSULIN | INSULIN | 36235843 | Ultralente Insulin, Human |
| GLIPIZIDE | GLIPIZIDE | 40165970 | Glipizide |
| TOLAZAMIDE | TOLAZAMIDE | 40089088 | Tolazamide |

| | | | |
|---|---|---|---|
| INSULIN | INSULIN | 19062999 | insulin, isophane |
| GLYBURIDE | GLYBURIDE | 19067365 | Glyburide |
| SITAGLIPTIN | SITAGLIPTIN | 40243629 | sitagliptin |
| INSULIN | INSULIN | 1502905 | Insulin Glargine |
| INSULIN | INSULIN | 19120315 | insulin, isophane |
| EMPAGLIFLOZIN | EMPAGLIFLOZIN | 36248788 | empagliflozin |
| INSULIN | INSULIN | 19078554 | Regular Insulin, Human |
| INSULIN | INSULIN | 1502905 | Insulin Glargine |
| INSULIN | INSULIN | 1550023 | Insulin Lispro |
| DULAGLUTIDE | DULAGLUTIDE | 45774445 | dulaglutide |
| INSULIN | INSULIN | 1550028 | Insulin Lispro |
| METFORMIN | METFORMIN | 40164918 | Metformin |
| INSULIN | INSULIN | 1596977 | Regular Insulin, Human |
| PIOGLITAZONE | PIOGLITAZONE | 1525215 | pioglitazone |
| MIGLITOL | MIGLITOL | 1510207 | miglitol |
| SITAGLIPTIN | SITAGLIPTIN | 42900902 | sitagliptin |
| INSULIN | INSULIN | 19071700 | Insulin Glargine |
| INSULIN | INSULIN | 19078553 | insulin, isophane |
| DULAGLUTIDE | DULAGLUTIDE | 45774440 | dulaglutide |
| INSULIN | INSULIN | 1596977 | Regular Insulin, Human |
| METFORMIN | METFORMIN | 1503328 | Metformin |
| GLIMEPIRIDE | GLIMEPIRIDE | 1597792 | glimepiride |
| INSULIN | INSULIN | 46234235 | insulin, isophane |
| INSULIN | INSULIN | 1513881 | Insulin Lispro |
| GLIPIZIDE | GLIPIZIDE | 19117945 | Glipizide |
| GLYBURIDE | GLYBURIDE | 19045630 | Glyburide |
| INSULIN | INSULIN | 19100128 | Regular Insulin, Human |
| INSULIN | INSULIN | 1562586 | Insulin, Zinc, Pork |
| INSULIN | INSULIN | 40166274 | Insulin Lispro |
| INSULIN | INSULIN | 19078558 | Insulin, Aspart, Human |
| GLIPIZIDE | GLIPIZIDE | 19079990 | Glipizide |
| METFORMIN | METFORMIN | 40231392 | Metformin |

| | | | |
|---|---|---|---|
| INSULIN | INSULIN | 19078554 | Regular Insulin, Human |
| CHLORPROPAMIDE | CHLORPROPAMIDE | 1594976 | Chlorpropamide |
| INSULIN | INSULIN | 40184294 | Insulin, Aspart Protamine, Human |
| METFORMIN | METFORMIN | 40063350 | Metformin |
| GLYBURIDE | GLYBURIDE | 19060491 | Glyburide |
| REPAGLINIDE | REPAGLINIDE | 19078028 | repaglinide |
| INSULIN | INSULIN | 1596960 | insulin, isophane |
| INSULIN | INSULIN | 1502905 | Insulin Glargine |
| MIGLITOL | MIGLITOL | 19029029 | miglitol |
| METFORMIN | METFORMIN | 40063354 | Metformin |
| INSULIN | INSULIN | 1596941 | insulin, isophane |
| INSULIN | INSULIN | 1567232 | Insulin, Aspart, Human |
| GLIPIZIDE | GLIPIZIDE | 19081295 | Glipizide |
| INSULIN | INSULIN | 19044971 | insulin detemir |
| ALOGLIPTIN | ALOGLIPTIN | 43013912 | alogliptin |
| METFORMIN | METFORMIN | 1503328 | Metformin |
| INSULIN | INSULIN | 1550027 | Insulin Lispro |
| INSULIN | INSULIN | 1596977 | Regular Insulin, Human |
| DAPAGLIFLOZIN | DAPAGLIFLOZIN | 45775866 | dapagliflozin |
| INSULIN | INSULIN | 19135320 | Regular Insulin, Human |
| INSULIN | INSULIN | 1596955 | insulin, isophane |
| INSULIN | INSULIN | 19078527 | Insulin, Zinc, Human |
| INSULIN | INSULIN | 1513849 | Insulin, Zinc, Human |
| INSULIN | INSULIN | 36231458 | insulin, isophane |
| EMPAGLIFLOZIN | EMPAGLIFLOZIN | 45774894 | empagliflozin |
| NATEGLINIDE | NATEGLINIDE | 36223777 | nateglinide |
| INSULIN | INSULIN | 40166274 | Insulin Lispro |
| LINAGLIPTIN | LINAGLIPTIN | 42708089 | Linagliptin |
| NATEGLINIDE | NATEGLINIDE | 19045605 | nateglinide |
| METFORMIN | METFORMIN | 1503297 | Metformin |
| GLYBURIDE | GLYBURIDE | 1559784 | Glyburide |
| INSULIN | INSULIN | 1567198 | Insulin, Aspart, Human |
| DAPAGLIFLOZIN | DAPAGLIFLOZIN | 44785833 | dapagliflozin |

| | | | |
|---|---|---|---|
| METFORMIN | METFORMIN | 40164918 | Metformin |
| INSULIN | INSULIN | 1562586 | Insulin, Zinc, Pork |
| INSULIN | INSULIN | 36222190 | insulin detemir |
| GLIPIZIDE | GLIPIZIDE | 19134639 | Glipizide |
| INSULIN | INSULIN | 40166275 | Insulin Lispro |
| INSULIN | INSULIN | 46234043 | Insulin, Aspart, Human |
| INSULIN | INSULIN | 19095211 | Insulin Lispro |
| INSULIN | INSULIN | 1516976 | insulin detemir |
| METFORMIN | METFORMIN | 40164928 | Metformin |
| INSULIN | INSULIN | 19078526 | Insulin, Zinc, Human |
| INSULIN | INSULIN | 19078553 | insulin, isophane |
| METFORMIN | METFORMIN | 1503297 | Metformin |
| SITAGLIPTIN | SITAGLIPTIN | 19125050 | sitagliptin |
| INSULIN | INSULIN | 1550027 | Insulin Lispro |
| INSULIN | INSULIN | 1597001 | Regular Insulin, Human |
| METFORMIN | METFORMIN | 1503297 | Metformin |
| DULAGLUTIDE | DULAGLUTIDE | 45774436 | dulaglutide |
| INSULIN | INSULIN | 19078527 | Insulin, Zinc, Human |
| INSULIN | INSULIN | 46233975 | Insulin Lispro |
| INSULIN | INSULIN | 19078633 | Insulin, Regular, Pork |
| INSULIN | INSULIN | 1550023 | Insulin Lispro |
| INSULIN | INSULIN | 19078551 | insulin, isophane |
| GLYBURIDE | GLYBURIDE | 40046201 | Glyburide |
| METFORMIN | METFORMIN | 40164922 | Metformin |
| INSULIN | INSULIN | 19131582 | Insulin, Glulisine, Human |
| INSULIN | INSULIN | 1596977 | Regular Insulin, Human |
| GLIPIZIDE | GLIPIZIDE | 40166816 | Glipizide |
| INSULIN | INSULIN | 19078633 | Insulin, Regular, Pork |
| DAPAGLIFLOZIN | DAPAGLIFLOZIN | 44785832 | dapagliflozin |
| INSULIN | INSULIN | 1596999 | Regular Insulin, Human |
| INSULIN | INSULIN | 19061670 | Regular Insulin, Human |
| INSULIN | INSULIN | 19129437 | insulin human, rDNA origin |

| | | | |
|---|---|---|---|
| INSULIN | INSULIN | 1502905 | Insulin Glargine |
| INSULIN | INSULIN | 35602728 | insulin degludec |
| METFORMIN | METFORMIN | 40164883 | Metformin |
| INSULIN | INSULIN | 19111795 | insulin, isophane |
| INSULIN | INSULIN | 19110355 | insulin, isophane |
| GLYBURIDE | GLYBURIDE | 1559684 | Glyburide |
| INSULIN | INSULIN | 36248870 | Insulin Glargine |
| INSULIN | INSULIN | 19120315 | insulin, isophane |
| INSULIN | INSULIN | 1567232 | Insulin, Aspart, Human |
| METFORMIN | METFORMIN | 40164938 | Metformin |
| GLYBURIDE | GLYBURIDE | 19077659 | Glyburide |
| INSULIN | INSULIN | 1502905 | Insulin Glargine |
| GLIMEPIRIDE | GLIMEPIRIDE | 1597776 | glimepiride |
| INSULIN | INSULIN | 1567201 | Insulin, Aspart Protamine, Human |
| INSULIN | INSULIN | 19062999 | insulin, isophane |
| INSULIN | INSULIN | 19110356 | Regular Insulin, Human |
| LIRAGLUTIDE | LIRAGLUTIDE | 40169223 | liraglutide |
| GLYBURIDE | GLYBURIDE | 36213841 | Glyburide |
| INSULIN | INSULIN | 1544838 | Insulin, Glulisine, Human |
| METFORMIN | METFORMIN | 36235746 | Metformin |
| INSULIN | INSULIN | 19078632 | Insulin, Regular, Pork |
| INSULIN | INSULIN | 19012703 | insulin, isophane |
| EXENATIDE | EXENATIDE | 46234119 | exenatide |
| DAPAGLIFLOZIN | DAPAGLIFLOZIN | 36247907 | dapagliflozin |
| INSULIN | INSULIN | 1513916 | Insulin Lispro |
| INSULIN | INSULIN | 1516979 | insulin detemir |
| GLYBURIDE | GLYBURIDE | 1559782 | Glyburide |
| INSULIN | INSULIN | 19129469 | insulin human, rDNA origin |
| INSULIN | INSULIN | 1513912 | Insulin Lispro |
| METFORMIN | METFORMIN | 40126689 | Metformin |
| INSULIN | INSULIN | 1567198 | Insulin, Aspart, Human |

| | | | |
|---|---|---|---|
| INSULIN | INSULIN | 1586345 | insulin, isophane |
| INSULIN | INSULIN | 19078557 | Regular Insulin, Human |
| LIXISENATIDE | LIXISENATIDE | 44506754 | Lixisenatide |
| INSULIN | INSULIN | 1567232 | Insulin, Aspart, Human |
| INSULIN | INSULIN | 1550028 | Insulin Lispro |
| SAXAGLIPTIN | SAXAGLIPTIN | 40166044 | saxagliptin |
| METFORMIN | METFORMIN | 40164890 | Metformin |
| METFORMIN | METFORMIN | 40164930 | Metformin |
| ROSIGLITAZONE | ROSIGLITAZONE | 1547504 | rosiglitazone |
| EXENATIDE | EXENATIDE | 19083253 | exenatide |
| METFORMIN | METFORMIN | 40164893 | Metformin |
| INSULIN | INSULIN | 36235937 | insulin human, rDNA origin |
| INSULIN | INSULIN | 1502905 | Insulin Glargine |
| ACARBOSE | ACARBOSE | 1529331 | Acarbose |
| INSULIN | INSULIN | 19078553 | insulin, isophane |
| METFORMIN | METFORMIN | 40164929 | Metformin |
| INSULIN | INSULIN | 19095475 | Ultralente Insulin, Human |
| GLIPIZIDE | GLIPIZIDE | 40165969 | Glipizide |
| GLIPIZIDE | GLIPIZIDE | 40165976 | Glipizide |
| EXENATIDE | EXENATIDE | 42708079 | exenatide |
| PIOGLITAZONE | PIOGLITAZONE | 19099207 | pioglitazone |
| ACETOHEXAMIDE | ACETOHEXAMIDE | 1530019 | Acetohexamide |
| GLYBURIDE | GLYBURIDE | 19079986 | Glyburide |
| GLIPIZIDE | GLIPIZIDE | 19117945 | Glipizide |
| REPAGLINIDE | REPAGLINIDE | 19023426 | repaglinide |
| METFORMIN | METFORMIN | 40164938 | Metformin |
| METFORMIN | METFORMIN | 42708177 | Metformin |
| INSULIN | INSULIN | 35605622 | Regular Insulin, Human |
| DULAGLUTIDE | DULAGLUTIDE | 45774442 | dulaglutide |
| INSULIN | INSULIN | 19078559 | Insulin Glargine |
| INSULIN | INSULIN | 19078633 | Insulin, Regular, Pork |
| NATEGLINIDE | NATEGLINIDE | 1502827 | nateglinide |

| | | | |
|---|---|---|---|
| INSULIN | INSULIN | 46233975 | Insulin Lispro |
| INSULIN | INSULIN | 46233945 | Insulin Lispro |
| INSULIN | INSULIN | 40051347 | Insulin Glargine |
| GLIPIZIDE | GLIPIZIDE | 1560171 | Glipizide |
| GLYBURIDE | GLYBURIDE | 1559785 | Glyburide |
| INSULIN | INSULIN | 1531601 | Insulin, Aspart Protamine, Human |
| INSULIN | INSULIN | 1596916 | insulin, isophane |
| GLYBURIDE | GLYBURIDE | 40168376 | Glyburide |
| ROSIGLITAZONE | ROSIGLITAZONE | 19120648 | rosiglitazone |
| INSULIN | INSULIN | 1502910 | Insulin Glargine |
| GLIPIZIDE | GLIPIZIDE | 19077638 | Glipizide |
| EXENATIDE | EXENATIDE | 19083253 | exenatide |
| METFORMIN | METFORMIN | 36235787 | Metformin |
| METFORMIN | METFORMIN | 40164942 | Metformin |
| PIOGLITAZONE | PIOGLITAZONE | 19099209 | pioglitazone |
| INSULIN | INSULIN | 1513876 | Insulin, Protamine Lispro, Human |
| BROMOCRIPTINE | BROMOCRIPTINE | 730548 | Bromocriptine |
| BROMOCRIPTINE | BROMOCRIPTINE | 36234233 | Bromocriptine |
| INSULIN | INSULIN | 1596999 | Regular Insulin, Human |
| INSULIN | INSULIN | 1586345 | insulin, isophane |
| INSULIN | INSULIN | 19078552 | insulin, isophane |
| INSULIN | INSULIN | 1596952 | insulin, isophane |
| METFORMIN | METFORMIN | 36235746 | Metformin |
| LINAGLIPTIN | LINAGLIPTIN | 36221103 | Linagliptin |
| INSULIN | INSULIN | 1544873 | Insulin, Glulisine, Human |
| ALOGLIPTIN | ALOGLIPTIN | 43013922 | alogliptin |
| INSULIN | INSULIN | 36229086 | Insulin Glargine |
| INSULIN | INSULIN | 40176981 | Insulin, Aspart Protamine, Human |
| INSULIN | INSULIN | 1596974 | insulin, isophane |
| METFORMIN | METFORMIN | 40164948 | Metformin |
| CANAGLIFLOZIN | CANAGLIFLOZIN | 43526469 | canagliflozin |
| INSULIN | INSULIN | 40166274 | Insulin Lispro |
| CANAGLIFLOZIN | CANAGLIFLOZIN | 43526469 | canagliflozin |
| METFORMIN | METFORMIN | 40229046 | Metformin |
| GLYBURIDE | GLYBURIDE | 40046201 | Glyburide |

| | | | |
|---|---|---|---|
| INSULIN | INSULIN | 1596952 | insulin, isophane |
| INSULIN | INSULIN | 42903322 | Regular Insulin, Human |
| INSULIN | INSULIN | 19095472 | insulin, isophane |
| INSULIN | INSULIN | 1597000 | Regular Insulin, Human |
| ROSIGLITAZONE | ROSIGLITAZONE | 1547506 | rosiglitazone |
| INSULIN | INSULIN | 19095218 | Regular Insulin, Human |
| GLIPIZIDE | GLIPIZIDE | 19134624 | Glipizide |
| GLYBURIDE | GLYBURIDE | 1559784 | Glyburide |
| MIGLITOL | MIGLITOL | 19081704 | miglitol |
| INSULIN | INSULIN | 42903270 | Insulin, Aspart Protamine, Human |
| INSULIN | INSULIN | 1562586 | Insulin, Zinc, Pork |
| LINAGLIPTIN | LINAGLIPTIN | 40239222 | Linagliptin |
| INSULIN | INSULIN | 1516978 | insulin detemir |
| EMPAGLIFLOZIN | EMPAGLIFLOZIN | 36248376 | empagliflozin |
| DULAGLUTIDE | DULAGLUTIDE | 45774440 | dulaglutide |
| BROMOCRIPTINE | BROMOCRIPTINE | 40164161 | Bromocriptine |
| MIGLITOL | MIGLITOL | 19029030 | miglitol |
| EXENATIDE | EXENATIDE | 36234709 | exenatide |
| GLIPIZIDE | GLIPIZIDE | 36241808 | Glipizide |
| GLIMEPIRIDE | GLIMEPIRIDE | 1597756 | glimepiride |
| PIOGLITAZONE | PIOGLITAZONE | 36236324 | pioglitazone |
| INSULIN | INSULIN | 1513881 | Insulin Lispro |
| NATEGLINIDE | NATEGLINIDE | 19102037 | nateglinide |
| INSULIN | INSULIN | 36231459 | Regular Insulin, Human |
| INSULIN | INSULIN | 19116454 | insulin, isophane |
| GLIMEPIRIDE | GLIMEPIRIDE | 19023585 | glimepiride |
| ACARBOSE | ACARBOSE | 19040969 | Acarbose |
| INSULIN | INSULIN | 36231450 | Insulin, Aspart, Human |
| GLYBURIDE | GLYBURIDE | 1559684 | Glyburide |
| BROMOCRIPTINE | BROMOCRIPTINE | 36238867 | Bromocriptine |
| PIOGLITAZONE | PIOGLITAZONE | 1525243 | pioglitazone |
| INSULIN | INSULIN | 1596972 | insulin, isophane |
| DULAGLUTIDE | DULAGLUTIDE | 36247186 | dulaglutide |
| INSULIN | INSULIN | 36231456 | insulin, isophane |
| METFORMIN | METFORMIN | 40164929 | Metformin |
| INSULIN | INSULIN | 46233975 | Insulin Lispro |

| | | | |
|---|---|---|---|
| GLIPIZIDE | GLIPIZIDE | 40166816 | Glipizide |
| METFORMIN | METFORMIN | 40164893 | Metformin |
| INSULIN | INSULIN | 36233552 | insulin, isophane |
| METFORMIN | METFORMIN | 40164932 | Metformin |
| METFORMIN | METFORMIN | 1503327 | Metformin |
| CANAGLIFLOZIN | CANAGLIFLOZIN | 45774714 | canagliflozin |
| TOLAZAMIDE | TOLAZAMIDE | 1502810 | Tolazamide |
| INSULIN | INSULIN | 36224228 | insulin detemir |
| GLIMEPIRIDE | GLIMEPIRIDE | 19117200 | glimepiride |
| GLYBURIDE | GLYBURIDE | 19077681 | Glyburide |
| GLIPIZIDE | GLIPIZIDE | 19077636 | Glipizide |
| INSULIN | INSULIN | 46233943 | Insulin Lispro |
| GLIMEPIRIDE | GLIMEPIRIDE | 1597756 | glimepiride |
| INSULIN | INSULIN | 1550023 | Insulin Lispro |
| PIOGLITAZONE | PIOGLITAZONE | 19079293 | pioglitazone |
| GLYBURIDE | GLYBURIDE | 19045630 | Glyburide |
| EXENATIDE | EXENATIDE | 36234709 | exenatide |
| METFORMIN | METFORMIN | 1503297 | Metformin |
| GLYBURIDE | GLYBURIDE | 19030580 | Glyburide |
| GLIMEPIRIDE | GLIMEPIRIDE | 1597776 | glimepiride |
| CANAGLIFLOZIN | CANAGLIFLOZIN | 45774724 | canagliflozin |
| INSULIN | INSULIN | 19078556 | Regular Insulin, Human |
| EXENATIDE | EXENATIDE | 1583727 | exenatide |
| EMPAGLIFLOZIN | EMPAGLIFLOZIN | 45774756 | empagliflozin |
| ACARBOSE | ACARBOSE | 19021786 | Acarbose |
| INSULIN | INSULIN | 1550023 | Insulin Lispro |
| INSULIN | INSULIN | 35602728 | insulin degludec |
| INSULIN | INSULIN | 19062999 | insulin, isophane |
| EXENATIDE | EXENATIDE | 46234118 | exenatide |
| EMPAGLIFLOZIN | EMPAGLIFLOZIN | 45892172 | empagliflozin |
| INSULIN | INSULIN | 19062999 | insulin, isophane |
| INSULIN | INSULIN | 19062999 | insulin, isophane |
| METFORMIN | METFORMIN | 1503328 | Metformin |
| LIRAGLUTIDE | LIRAGLUTIDE | 45892686 | liraglutide |

| | | | |
|---|---|---|---|
| GLYBURIDE | GLYBURIDE | 40046201 | Glyburide |
| ALBIGLUTIDE | ALBIGLUTIDE | 44816283 | albiglutide |
| GLIMEPIRIDE | GLIMEPIRIDE | 36234086 | glimepiride |
| METFORMIN | METFORMIN | 40164880 | Metformin |
| DULAGLUTIDE | DULAGLUTIDE | 45774440 | dulaglutide |
| GLYBURIDE | GLYBURIDE | 36233495 | Glyburide |
| DAPAGLIFLOZIN | DAPAGLIFLOZIN | 36247907 | dapagliflozin |
| INSULIN | INSULIN | 1550028 | Insulin Lispro |
| METFORMIN | METFORMIN | 40164904 | Metformin |
| INSULIN | INSULIN | 1567198 | Insulin, Aspart, Human |
| GLIMEPIRIDE | GLIMEPIRIDE | 19018043 | glimepiride |
| ALOGLIPTIN | ALOGLIPTIN | 43013903 | alogliptin |
| GLIMEPIRIDE | GLIMEPIRIDE | 1597756 | glimepiride |
| INSULIN | INSULIN | 19111795 | insulin, isophane |
| INSULIN | INSULIN | 1597001 | Regular Insulin, Human |
| METFORMIN | METFORMIN | 40164914 | Metformin |
| GLIMEPIRIDE | GLIMEPIRIDE | 19023585 | glimepiride |
| ROSIGLITAZONE | ROSIGLITAZONE | 19099245 | rosiglitazone |
| METFORMIN | METFORMIN | 40164893 | Metformin |
| GLIPIZIDE | GLIPIZIDE | 19030445 | Glipizide |
| METFORMIN | METFORMIN | 40164880 | Metformin |
| METFORMIN | METFORMIN | 40164894 | Metformin |
| CANAGLIFLOZIN | CANAGLIFLOZIN | 45774714 | canagliflozin |
| PIOGLITAZONE | PIOGLITAZONE | 40099897 | pioglitazone |
| GLIPIZIDE | GLIPIZIDE | 40166814 | Glipizide |
| METFORMIN | METFORMIN | 19088818 | Metformin |
| INSULIN | INSULIN | 1550023 | Insulin Lispro |
| INSULIN | INSULIN | 19111795 | insulin, isophane |
| PIOGLITAZONE | PIOGLITAZONE | 19099209 | pioglitazone |
| GLYBURIDE | GLYBURIDE | 1559684 | Glyburide |
| INSULIN | INSULIN | 46233950 | Insulin Lispro |
| ALBIGLUTIDE | ALBIGLUTIDE | 44816332 | albiglutide |
| INSULIN | INSULIN | 1596916 | insulin, isophane |
| GLIPIZIDE | GLIPIZIDE | 1560171 | Glipizide |
| NATEGLINIDE | NATEGLINIDE | 19085097 | nateglinide |
| MIGLITOL | MIGLITOL | 1510206 | miglitol |

| | | | |
|---|---|---|---|
| BROMOCRIPTINE | BROMOCRIPTINE | 19041784 | Bromocriptine |
| INSULIN | INSULIN | 46221557 | Insulin Glargine |
| INSULIN | INSULIN | 1597001 | Regular Insulin, Human |
| SITAGLIPTIN | SITAGLIPTIN | 19125050 | sitagliptin |
| INSULIN | INSULIN | 1516981 | insulin detemir |
| METFORMIN | METFORMIN | 40171449 | Metformin |
| GLYBURIDE | GLYBURIDE | 40168368 | Glyburide |
| INSULIN | INSULIN | 19058398 | Insulin Lispro |
| SAXAGLIPTIN | SAXAGLIPTIN | 36224321 | saxagliptin |
| INSULIN | INSULIN | 1502905 | Insulin Glargine |
| CANAGLIFLOZIN | CANAGLIFLOZIN | 43526473 | canagliflozin |
| INSULIN | INSULIN | 1550027 | Insulin Lispro |
| PIOGLITAZONE | PIOGLITAZONE | 36236324 | pioglitazone |
| INSULIN | INSULIN | 19014975 | insulin, isophane |
| INSULIN | INSULIN | 36236032 | Insulin Lispro |
| GLYBURIDE | GLYBURIDE | 1559760 | Glyburide |
| GLYBURIDE | GLYBURIDE | 40165998 | Glyburide |
| BROMOCRIPTINE | BROMOCRIPTINE | 40164161 | Bromocriptine |
| EXENATIDE | EXENATIDE | 42708079 | exenatide |
| GLIPIZIDE | GLIPIZIDE | 36235748 | Glipizide |
| REPAGLINIDE | REPAGLINIDE | 19078028 | repaglinide |
| INSULIN | INSULIN | 1550028 | Insulin Lispro |
| INSULIN | INSULIN | 1596999 | Regular Insulin, Human |
| SITAGLIPTIN | SITAGLIPTIN | 19125042 | sitagliptin |
| INSULIN | INSULIN | 1513881 | Insulin Lispro |
| INSULIN | INSULIN | 19116559 | insulin, isophane |
| GLIPIZIDE | GLIPIZIDE | 19077636 | Glipizide |
| INSULIN | INSULIN | 19044971 | insulin detemir |
| GLYBURIDE | GLYBURIDE | 19077682 | Glyburide |
| INSULIN | INSULIN | 40176981 | Insulin, Aspart Protamine, Human |
| INSULIN | INSULIN | 1502905 | Insulin Glargine |
| DAPAGLIFLOZIN | DAPAGLIFLOZIN | 45775999 | dapagliflozin |
| GLIMEPIRIDE | GLIMEPIRIDE | 1597776 | glimepiride |

| | | | |
|---|---|---|---|
| INSULIN | INSULIN | 1567198 | Insulin, Aspart, Human |
| INSULIN | INSULIN | 46234049 | Insulin, Aspart, Human |
| INSULIN | INSULIN | 19110352 | insulin, isophane |
| REPAGLINIDE | REPAGLINIDE | 40098707 | repaglinide |
| GLIPIZIDE | GLIPIZIDE | 19030445 | Glipizide |
| LIRAGLUTIDE | LIRAGLUTIDE | 40169223 | liraglutide |
| GLYBURIDE | GLYBURIDE | 19106860 | Glyburide |
| PIOGLITAZONE | PIOGLITAZONE | 19099208 | pioglitazone |
| METFORMIN | METFORMIN | 40163922 | Metformin |
| METFORMIN | METFORMIN | 40164898 | Metformin |
| GLYBURIDE | GLYBURIDE | 1559782 | Glyburide |
| INSULIN | INSULIN | 46234043 | Insulin, Aspart, Human |
| INSULIN | INSULIN | 1596977 | Regular Insulin, Human |
| INSULIN | INSULIN | 1596959 | insulin, isophane |
| CANAGLIFLOZIN | CANAGLIFLOZIN | 36246878 | canagliflozin |
| INSULIN | INSULIN | 1596998 | Regular Insulin, Human |
| LINAGLIPTIN | LINAGLIPTIN | 42708087 | Linagliptin |
| INSULIN | INSULIN | 46221557 | Insulin Glargine |
| INSULIN | INSULIN | 46234241 | Regular Insulin, Human |
| EXENATIDE | EXENATIDE | 46234116 | exenatide |
| INSULIN | INSULIN | 19111795 | insulin, isophane |
| METFORMIN | METFORMIN | 40164904 | Metformin |
| GLIPIZIDE | GLIPIZIDE | 19030445 | Glipizide |
| INSULIN | INSULIN | 46234045 | Insulin, Aspart, Human |
| GLYBURIDE | GLYBURIDE | 1559782 | Glyburide |
| INSULIN | INSULIN | 1567201 | Insulin, Aspart Protamine, Human |
| GLIMEPIRIDE | GLIMEPIRIDE | 40059665 | glimepiride |
| TOLAZAMIDE | TOLAZAMIDE | 19036966 | Tolazamide |
| EMPAGLIFLOZIN | EMPAGLIFLOZIN | 45892179 | empagliflozin |
| INSULIN | INSULIN | 1596959 | insulin, isophane |
| GLIMEPIRIDE | GLIMEPIRIDE | 1597757 | glimepiride |
| GLYBURIDE | GLYBURIDE | 1559785 | Glyburide |
| INSULIN | INSULIN | 35602722 | insulin degludec |
| INSULIN | INSULIN | 19014976 | insulin, isophane |
| INSULIN | INSULIN | 1502905 | Insulin Glargine |
| INSULIN | INSULIN | 40166274 | Insulin Lispro |

| | | | |
|---|---|---|---|
| SITAGLIPTIN | SITAGLIPTIN | 19125042 | sitagliptin |
| CHLORPROPAMIDE | CHLORPROPAMIDE | 1594973 | Chlorpropamide |
| INSULIN | INSULIN | 46233975 | Insulin Lispro |
| ALBIGLUTIDE | ALBIGLUTIDE | 44816274 | albiglutide |
| EMPAGLIFLOZIN | EMPAGLIFLOZIN | 46287690 | empagliflozin |
| METFORMIN | METFORMIN | 40231393 | Metformin |
| INSULIN | INSULIN | 1544873 | Insulin, Glulisine, Human |
| METFORMIN | METFORMIN | 40164897 | Metformin |
| GLYBURIDE | GLYBURIDE | 1559784 | Glyburide |
| ROSIGLITAZONE | ROSIGLITAZONE | 19099246 | rosiglitazone |
| GLYBURIDE | GLYBURIDE | 36213841 | Glyburide |
| GLIPIZIDE | GLIPIZIDE | 1560233 | Glipizide |
| GLIMEPIRIDE | GLIMEPIRIDE | 1597796 | glimepiride |
| METFORMIN | METFORMIN | 40164930 | Metformin |
| INSULIN | INSULIN | 46221559 | Insulin Glargine |
| INSULIN | INSULIN | 1502905 | Insulin Glargine |
| INSULIN | INSULIN | 1513849 | Insulin, Zinc, Human |
| INSULIN | INSULIN | 19110220 | Regular Insulin, Human |
| INSULIN | INSULIN | 40166274 | Insulin Lispro |
| GLIMEPIRIDE | GLIMEPIRIDE | 1597776 | glimepiride |
| GLIPIZIDE | GLIPIZIDE | 19077636 | Glipizide |
| INSULIN | INSULIN | 19078555 | Regular Insulin, Human |
| LIRAGLUTIDE | LIRAGLUTIDE | 45892684 | liraglutide |
| ACETOHEXAMIDE | ACETOHEXAMIDE | 19088426 | Acetohexamide |
| PRAMLINTIDE | PRAMLINTIDE | 40165692 | Pramlintide |
| METFORMIN | METFORMIN | 1503327 | Metformin |
| INSULIN | INSULIN | 19078554 | Regular Insulin, Human |
| GLYBURIDE | GLYBURIDE | 40165998 | Glyburide |
| INSULIN | INSULIN | 1502905 | Insulin Glargine |
| METFORMIN | METFORMIN | 19088818 | Metformin |
| INSULIN | INSULIN | 46275452 | Regular Insulin, Human |
| INSULIN | INSULIN | 19095211 | Insulin Lispro |
| INSULIN | INSULIN | 19095214 | insulin, isophane |
| INSULIN | INSULIN | 46233975 | Insulin Lispro |

| | | | |
|---|---|---|---|
| INSULIN | INSULIN | 19099013 | Ultralente Insulin, Beef-Pork |
| METFORMIN | METFORMIN | 40164942 | Metformin |
| INSULIN | INSULIN | 1586344 | insulin, isophane |
| INSULIN | INSULIN | 19120315 | insulin, isophane |
| INSULIN | INSULIN | 19095214 | insulin, isophane |
| INSULIN | INSULIN | 1596973 | insulin, isophane |
| INSULIN | INSULIN | 36248870 | Insulin Glargine |
| ROSIGLITAZONE | ROSIGLITAZONE | 40099633 | rosiglitazone |
| INSULIN | INSULIN | 19080091 | Ultralente Insulin, Human |
| METFORMIN | METFORMIN | 40164935 | Metformin |
| GLYBURIDE | GLYBURIDE | 19077681 | Glyburide |
| GLIPIZIDE | GLIPIZIDE | 19134642 | Glipizide |
| METFORMIN | METFORMIN | 40229050 | Metformin |
| INSULIN | INSULIN | 19078633 | Insulin, Regular, Pork |
| PIOGLITAZONE | PIOGLITAZONE | 1525217 | pioglitazone |
| INSULIN | INSULIN | 1550028 | Insulin Lispro |
| INSULIN | INSULIN | 1597012 | Regular Insulin, Human |
| INSULIN | INSULIN | 1513912 | Insulin Lispro |
| EXENATIDE | EXENATIDE | 46234118 | exenatide |
| EXENATIDE | EXENATIDE | 46234118 | exenatide |
| GLIPIZIDE | GLIPIZIDE | 19134639 | Glipizide |
| INSULIN | INSULIN | 1586345 | insulin, isophane |
| REPAGLINIDE | REPAGLINIDE | 40098707 | repaglinide |
| ACARBOSE | ACARBOSE | 19021312 | Acarbose |
| GLYBURIDE | GLYBURIDE | 40046201 | Glyburide |
| GLIMEPIRIDE | GLIMEPIRIDE | 1597757 | glimepiride |
| INSULIN | INSULIN | 19078530 | insulin, isophane |
| INSULIN | INSULIN | 1567198 | Insulin, Aspart, Human |
| INSULIN | INSULIN | 19100127 | insulin, isophane |
| GLYBURIDE | GLYBURIDE | 19081296 | Glyburide |
| INSULIN | INSULIN | 40130712 | insulin detemir |
| ACARBOSE | ACARBOSE | 1529331 | Acarbose |

| | | | |
|---|---|---|---|
| INSULIN | INSULIN | 19104457 | insulin, isophane |
| METFORMIN | METFORMIN | 40164925 | Metformin |
| METFORMIN | METFORMIN | 40164939 | Metformin |
| GLYBURIDE | GLYBURIDE | 1559784 | Glyburide |
| ROSIGLITAZONE | ROSIGLITAZONE | 36229366 | rosiglitazone |
| GLYBURIDE | GLYBURIDE | 1559760 | Glyburide |
| INSULIN | INSULIN | 1513881 | Insulin Lispro |
| INSULIN | INSULIN | 36236032 | Insulin Lispro |
| GLIPIZIDE | GLIPIZIDE | 40166816 | Glipizide |
| INSULIN | INSULIN | 46221557 | Insulin Glargine |
| DAPAGLIFLOZIN | DAPAGLIFLOZIN | 44785477 | dapagliflozin |
| BROMOCRIPTINE | BROMOCRIPTINE | 730551 | Bromocriptine |
| METFORMIN | METFORMIN | 40117785 | Metformin |
| INSULIN | INSULIN | 19078553 | insulin, isophane |
| PIOGLITAZONE | PIOGLITAZONE | 19099208 | pioglitazone |
| INSULIN | INSULIN | 1513912 | Insulin Lispro |
| EXENATIDE | EXENATIDE | 1583728 | exenatide |
| INSULIN | INSULIN | 46233976 | Insulin Lispro |
| INSULIN | INSULIN | 35604094 | Insulin Glargine |
| NATEGLINIDE | NATEGLINIDE | 19045605 | nateglinide |
| SITAGLIPTIN | SITAGLIPTIN | 19125046 | sitagliptin |
| ALOGLIPTIN | ALOGLIPTIN | 36246195 | alogliptin |
| INSULIN | INSULIN | 1502905 | Insulin Glargine |
| GLIPIZIDE | GLIPIZIDE | 1560233 | Glipizide |
| GLIPIZIDE | GLIPIZIDE | 19100559 | Glipizide |
| EMPAGLIFLOZIN | EMPAGLIFLOZIN | 1592804 | empagliflozin |
| TOLBUTAMIDE | TOLBUTAMIDE | 36242646 | Tolbutamide |
| REPAGLINIDE | REPAGLINIDE | 19078030 | repaglinide |
| EXENATIDE | EXENATIDE | 36234709 | exenatide |
| INSULIN | INSULIN | 1567198 | Insulin, Aspart, Human |
| DULAGLUTIDE | DULAGLUTIDE | 45774438 | dulaglutide |
| ROSIGLITAZONE | ROSIGLITAZONE | 36227624 | rosiglitazone |
| GLIPIZIDE | GLIPIZIDE | 19117945 | Glipizide |
| BROMOCRIPTINE | BROMOCRIPTINE | 19007102 | Bromocriptine |
| GLIMEPIRIDE | GLIMEPIRIDE | 1597759 | glimepiride |

| | | | |
|---|---|---|---|
| INSULIN | INSULIN | 19078551 | insulin, isophane |
| CANAGLIFLOZIN | CANAGLIFLOZIN | 43526467 | canagliflozin |
| GLIPIZIDE | GLIPIZIDE | 19077638 | Glipizide |
| GLYBURIDE | GLYBURIDE | 40166003 | Glyburide |
| INSULIN | INSULIN | 19078553 | insulin, isophane |
| PIOGLITAZONE | PIOGLITAZONE | 19099208 | pioglitazone |
| METFORMIN | METFORMIN | 40164908 | Metformin |
| CHLORPROPAMIDE | CHLORPROPAMIDE | 19006907 | Chlorpropamide |
| LIRAGLUTIDE | LIRAGLUTIDE | 40170913 | liraglutide |
| METFORMIN | METFORMIN | 36236481 | Metformin |
| ACARBOSE | ACARBOSE | 19047612 | Acarbose |
| INSULIN | INSULIN | 19062999 | insulin, isophane |
| NATEGLINIDE | NATEGLINIDE | 1502826 | nateglinide |
| INSULIN | INSULIN | 1544838 | Insulin, Glulisine, Human |
| GLYBURIDE | GLYBURIDE | 19045630 | Glyburide |
| INSULIN | INSULIN | 19078530 | insulin, isophane |
| INSULIN | INSULIN | 19062999 | insulin, isophane |
| INSULIN | INSULIN | 46233975 | Insulin Lispro |
| METFORMIN | METFORMIN | 40164941 | Metformin |
| NATEGLINIDE | NATEGLINIDE | 40058199 | nateglinide |
| LINAGLIPTIN | LINAGLIPTIN | 40239221 | Linagliptin |
| METFORMIN | METFORMIN | 40164926 | Metformin |
| INSULIN | INSULIN | 36247261 | Regular Insulin, Human |
| LIRAGLUTIDE | LIRAGLUTIDE | 40170913 | liraglutide |
| PIOGLITAZONE | PIOGLITAZONE | 1525244 | pioglitazone |
| CANAGLIFLOZIN | CANAGLIFLOZIN | 43526473 | canagliflozin |
| METFORMIN | METFORMIN | 40164936 | Metformin |
| INSULIN | INSULIN | 46234049 | Insulin, Aspart, Human |
| GLYBURIDE | GLYBURIDE | 1559684 | Glyburide |
| INSULIN | INSULIN | 19058398 | Insulin Lispro |
| GLYBURIDE | GLYBURIDE | 1559684 | Glyburide |
| METFORMIN | METFORMIN | 40164946 | Metformin |

| | | | |
|---|---|---|---|
| ACARBOSE | ACARBOSE | 1529359 | Acarbose |
| METFORMIN | METFORMIN | 1503297 | Metformin |
| METFORMIN | METFORMIN | 40164883 | Metformin |
| GLYBURIDE | GLYBURIDE | 1559784 | Glyburide |
| METFORMIN | METFORMIN | 40164900 | Metformin |
| LINAGLIPTIN | LINAGLIPTIN | 40239220 | Linagliptin |
| GLIMEPIRIDE | GLIMEPIRIDE | 1597756 | glimepiride |
| INSULIN | INSULIN | 1502910 | Insulin Glargine |
| BROMOCRIPTINE | BROMOCRIPTINE | 40016896 | Bromocriptine |
| METFORMIN | METFORMIN | 19113780 | Metformin |
| INSULIN | INSULIN | 40051347 | Insulin Glargine |
| GLIPIZIDE | GLIPIZIDE | 40167638 | Glipizide |
| INSULIN | INSULIN | 40166275 | Insulin Lispro |
| INSULIN | INSULIN | 1502905 | Insulin Glargine |
| GLYBURIDE | GLYBURIDE | 19030572 | Glyburide |
| INSULIN | INSULIN | 19111795 | insulin, isophane |
| INSULIN | INSULIN | 1516981 | insulin detemir |
| GLIMEPIRIDE | GLIMEPIRIDE | 36229116 | glimepiride |
| INSULIN | INSULIN | 36227902 | Insulin Lispro |
| GLIPIZIDE | GLIPIZIDE | 19081295 | Glipizide |
| INSULIN | INSULIN | 46234045 | Insulin, Aspart, Human |
| INSULIN | INSULIN | 1550023 | Insulin Lispro |
| GLIMEPIRIDE | GLIMEPIRIDE | 19117200 | glimepiride |
| INSULIN | INSULIN | 19120790 | Insulin Glargine |
| ACARBOSE | ACARBOSE | 1529358 | Acarbose |
| EXENATIDE | EXENATIDE | 42705842 | exenatide |
| GLYBURIDE | GLYBURIDE | 1559784 | Glyburide |
| EMPAGLIFLOZIN | EMPAGLIFLOZIN | 46287684 | empagliflozin |
| INSULIN | INSULIN | 19078557 | Regular Insulin, Human |
| EXENATIDE | EXENATIDE | 1583722 | exenatide |
| PIOGLITAZONE | PIOGLITAZONE | 19099208 | pioglitazone |
| METFORMIN | METFORMIN | 40164880 | Metformin |
| GLIMEPIRIDE | GLIMEPIRIDE | 1597775 | glimepiride |
| METFORMIN | METFORMIN | 36235746 | Metformin |
| INSULIN | INSULIN | 46234045 | Insulin, Aspart, Human |
| METFORMIN | METFORMIN | 40135481 | Metformin |
| INSULIN | INSULIN | 40176981 | Insulin, Aspart Protamine, Human |

| | | | |
|---|---|---|---|
| BROMOCRIPTINE | BROMOCRIPTINE | 36234233 | Bromocriptine |
| METFORMIN | METFORMIN | 40164933 | Metformin |
| INSULIN | INSULIN | 19078554 | Regular Insulin, Human |
| GLIMEPIRIDE | GLIMEPIRIDE | 1597756 | glimepiride |
| BROMOCRIPTINE | BROMOCRIPTINE | 36238866 | Bromocriptine |
| LINAGLIPTIN | LINAGLIPTIN | 42708091 | Linagliptin |
| GLYBURIDE | GLYBURIDE | 40166002 | Glyburide |
| INSULIN | INSULIN | 40176981 | Insulin, Aspart Protamine, Human |
| NATEGLINIDE | NATEGLINIDE | 1502826 | nateglinide |
| INSULIN | INSULIN | 1513916 | Insulin Lispro |
| INSULIN | INSULIN | 19116559 | insulin, isophane |
| METFORMIN | METFORMIN | 40164943 | Metformin |
| INSULIN | INSULIN | 1596972 | insulin, isophane |
| ALBIGLUTIDE | ALBIGLUTIDE | 44816283 | albiglutide |
| INSULIN | INSULIN | 42899447 | Insulin, Pork |
| GLIPIZIDE | GLIPIZIDE | 1560171 | Glipizide |
| ALOGLIPTIN | ALOGLIPTIN | 43013916 | alogliptin |
| GLIPIZIDE | GLIPIZIDE | 19081295 | Glipizide |
| SAXAGLIPTIN | SAXAGLIPTIN | 36229292 | saxagliptin |
| PIOGLITAZONE | PIOGLITAZONE | 1525215 | pioglitazone |
| INSULIN | INSULIN | 19078551 | insulin, isophane |
| INSULIN | INSULIN | 19116454 | insulin, isophane |
| INSULIN | INSULIN | 36231459 | Regular Insulin, Human |
| INSULIN | INSULIN | 19095218 | Regular Insulin, Human |
| INSULIN | INSULIN | 19120315 | insulin, isophane |
| INSULIN | INSULIN | 19116454 | insulin, isophane |
| INSULIN | INSULIN | 46233975 | Insulin Lispro |
| INSULIN | INSULIN | 1596955 | insulin, isophane |
| INSULIN | INSULIN | 19061670 | Regular Insulin, Human |

| | | | |
|---|---|---|---|
| METFORMIN | METFORMIN | 1503297 | Metformin |
| GLIPIZIDE | GLIPIZIDE | 1560233 | Glipizide |
| METFORMIN | METFORMIN | 40231404 | Metformin |
| MIGLITOL | MIGLITOL | 19029061 | miglitol |
| INSULIN | INSULIN | 19116455 | insulin, isophane |
| METFORMIN | METFORMIN | 40164893 | Metformin |
| GLIPIZIDE | GLIPIZIDE | 1560233 | Glipizide |
| GLYBURIDE | GLYBURIDE | 19077682 | Glyburide |
| METFORMIN | METFORMIN | 40164929 | Metformin |
| BROMOCRIPTINE | BROMOCRIPTINE | 19007101 | Bromocriptine |
| ACARBOSE | ACARBOSE | 1529359 | Acarbose |
| METFORMIN | METFORMIN | 40117785 | Metformin |
| INSULIN | INSULIN | 1567198 | Insulin, Aspart, Human |
| INSULIN | INSULIN | 19044971 | insulin detemir |
| ALBIGLUTIDE | ALBIGLUTIDE | 44816282 | albiglutide |
| EXENATIDE | EXENATIDE | 45774533 | exenatide |
| INSULIN | INSULIN | 35602731 | insulin degludec |
| METFORMIN | METFORMIN | 40164929 | Metformin |
| SAXAGLIPTIN | SAXAGLIPTIN | 40166044 | saxagliptin |
| METFORMIN | METFORMIN | 40163926 | Metformin |
| INSULIN | INSULIN | 1516978 | insulin detemir |
| ROSIGLITAZONE | ROSIGLITAZONE | 36229365 | rosiglitazone |
| METFORMIN | METFORMIN | 36235746 | Metformin |
| INSULIN | INSULIN | 46233730 | Regular Insulin, Human |
| INSULIN | INSULIN | 19078553 | insulin, isophane |
| EXENATIDE | EXENATIDE | 42705842 | exenatide |
| METFORMIN | METFORMIN | 40164929 | Metformin |
| LINAGLIPTIN | LINAGLIPTIN | 40239220 | Linagliptin |
| INSULIN | INSULIN | 35602731 | insulin degludec |
| GLYBURIDE | GLYBURIDE | 40046204 | Glyburide |
| EXENATIDE | EXENATIDE | 1583722 | exenatide |
| METFORMIN | METFORMIN | 40231391 | Metformin |
| INSULIN | INSULIN | 1516981 | insulin detemir |
| EMPAGLIFLOZIN | EMPAGLIFLOZIN | 36246886 | empagliflozin |
| INSULIN | INSULIN | 19135264 | Regular Insulin, Human |

| | | | |
|---|---|---|---|
| GLIPIZIDE | GLIPIZIDE | 40167638 | Glipizide |
| METFORMIN | METFORMIN | 40164901 | Metformin |
| INSULIN | INSULIN | 1516981 | insulin detemir |
| GLIMEPIRIDE | GLIMEPIRIDE | 1597756 | glimepiride |
| METFORMIN | METFORMIN | 40063356 | Metformin |
| INSULIN | INSULIN | 40166274 | Insulin Lispro |
| INSULIN | INSULIN | 1502908 | Insulin Glargine |
| METFORMIN | METFORMIN | 40164935 | Metformin |
| INSULIN | INSULIN | 19078557 | Regular Insulin, Human |
| SITAGLIPTIN | SITAGLIPTIN | 19125042 | sitagliptin |
| CHLORPROPAMIDE | CHLORPROPAMIDE | 40030456 | Chlorpropamide |
| INSULIN | INSULIN | 19078559 | Insulin Glargine |
| INSULIN | INSULIN | 19120315 | insulin, isophane |
| LIRAGLUTIDE | LIRAGLUTIDE | 40170911 | liraglutide |
| SITAGLIPTIN | SITAGLIPTIN | 40243633 | sitagliptin |
| INSULIN | INSULIN | 19095214 | insulin, isophane |
| CANAGLIFLOZIN | CANAGLIFLOZIN | 43526473 | canagliflozin |
| GLIMEPIRIDE | GLIMEPIRIDE | 1597796 | glimepiride |
| INSULIN | INSULIN | 19078530 | insulin, isophane |
| METFORMIN | METFORMIN | 40164918 | Metformin |
| INSULIN | INSULIN | 19071700 | Insulin Glargine |
| BROMOCRIPTINE | BROMOCRIPTINE | 19116171 | Bromocriptine |
| PIOGLITAZONE | PIOGLITAZONE | 40099898 | pioglitazone |
| PIOGLITAZONE | PIOGLITAZONE | 19099208 | pioglitazone |
| INSULIN | INSULIN | 40051347 | Insulin Glargine |
| METFORMIN | METFORMIN | 40164880 | Metformin |
| DAPAGLIFLOZIN | DAPAGLIFLOZIN | 44815996 | dapagliflozin |
| INSULIN | INSULIN | 1502905 | Insulin Glargine |
| BROMOCRIPTINE | BROMOCRIPTINE | 19007070 | Bromocriptine |
| METFORMIN | METFORMIN | 19113780 | Metformin |
| GLIMEPIRIDE | GLIMEPIRIDE | 1597776 | glimepiride |
| INSULIN | INSULIN | 35604094 | Insulin Glargine |
| INSULIN | INSULIN | 1586344 | insulin, isophane |
| INSULIN | INSULIN | 19120790 | Insulin Glargine |
| INSULIN | INSULIN | 19062999 | insulin, isophane |

| | | | |
|---|---|---|---|
| INSULIN | INSULIN | 1513850 | Insulin, Zinc, Human |
| INSULIN | INSULIN | 46233950 | Insulin Lispro |
| INSULIN | INSULIN | 1516979 | insulin detemir |
| INSULIN | INSULIN | 19090210 | Lente Insulin, Beef |
| INSULIN | INSULIN | 19063035 | Regular Insulin, Human |
| GLIPIZIDE | GLIPIZIDE | 19134642 | Glipizide |
| INSULIN | INSULIN | 1596959 | insulin, isophane |
| METFORMIN | METFORMIN | 40164932 | Metformin |
| ALBIGLUTIDE | ALBIGLUTIDE | 44816283 | albiglutide |
| EXENATIDE | EXENATIDE | 1583728 | exenatide |
| GLIMEPIRIDE | GLIMEPIRIDE | 19117198 | glimepiride |
| SITAGLIPTIN | SITAGLIPTIN | 19125042 | sitagliptin |
| METFORMIN | METFORMIN | 1503297 | Metformin |
| EXENATIDE | EXENATIDE | 1583730 | exenatide |
| INSULIN | INSULIN | 1516979 | insulin detemir |
| GLIMEPIRIDE | GLIMEPIRIDE | 1597799 | glimepiride |
| INSULIN | INSULIN | 1502910 | Insulin Glargine |
| GLYBURIDE | GLYBURIDE | 19045628 | Glyburide |
| GLIMEPIRIDE | GLIMEPIRIDE | 1597757 | glimepiride |
| GLIMEPIRIDE | GLIMEPIRIDE | 1597795 | glimepiride |
| METFORMIN | METFORMIN | 1503297 | Metformin |
| DAPAGLIFLOZIN | DAPAGLIFLOZIN | 44785473 | dapagliflozin |
| REPAGLINIDE | REPAGLINIDE | 36240429 | repaglinide |
| INSULIN | INSULIN | 1596959 | insulin, isophane |
| INSULIN | INSULIN | 1567232 | Insulin, Aspart, Human |
| METFORMIN | METFORMIN | 40063352 | Metformin |
| INSULIN | INSULIN | 1516979 | insulin detemir |
| INSULIN | INSULIN | 1596999 | Regular Insulin, Human |
| GLIMEPIRIDE | GLIMEPIRIDE | 1597775 | glimepiride |
| METFORMIN | METFORMIN | 40063356 | Metformin |
| INSULIN | INSULIN | 19078633 | Insulin, Regular, Pork |
| PIOGLITAZONE | PIOGLITAZONE | 40099897 | pioglitazone |
| ROSIGLITAZONE | ROSIGLITAZONE | 1547504 | rosiglitazone |
| EXENATIDE | EXENATIDE | 46234116 | exenatide |
| INSULIN | INSULIN | 35602731 | insulin degludec |
| GLYBURIDE | GLYBURIDE | 1559760 | Glyburide |
| INSULIN | INSULIN | 1596974 | insulin, isophane |
| METFORMIN | METFORMIN | 1503328 | Metformin |

| | | | |
|---|---|---|---|
| INSULIN | INSULIN | 19014975 | insulin, isophane |
| INSULIN | INSULIN | 1567198 | Insulin, Aspart, Human |
| EXENATIDE | EXENATIDE | 1583722 | exenatide |
| METFORMIN | METFORMIN | 19086484 | Metformin |
| GLIMEPIRIDE | GLIMEPIRIDE | 1597756 | glimepiride |
| INSULIN | INSULIN | 19078555 | Regular Insulin, Human |
| INSULIN | INSULIN | 19078557 | Regular Insulin, Human |
| METFORMIN | METFORMIN | 40063354 | Metformin |
| INSULIN | INSULIN | 1567198 | Insulin, Aspart, Human |
| EXENATIDE | EXENATIDE | 46234116 | exenatide |
| ALOGLIPTIN | ALOGLIPTIN | 43013925 | alogliptin |
| INSULIN | INSULIN | 1562586 | Insulin, Zinc, Pork |
| EXENATIDE | EXENATIDE | 19135404 | exenatide |
| INSULIN | INSULIN | 36229086 | Insulin Glargine |
| GLYBURIDE | GLYBURIDE | 19077659 | Glyburide |
| ALBIGLUTIDE | ALBIGLUTIDE | 36246906 | albiglutide |
| GLYBURIDE | GLYBURIDE | 1559684 | Glyburide |
| GLIMEPIRIDE | GLIMEPIRIDE | 1597759 | glimepiride |
| INSULIN | INSULIN | 1567198 | Insulin, Aspart, Human |
| DAPAGLIFLOZIN | DAPAGLIFLOZIN | 44785833 | dapagliflozin |
| METFORMIN | METFORMIN | 40164895 | Metformin |
| NATEGLINIDE | NATEGLINIDE | 19045606 | nateglinide |
| LIRAGLUTIDE | LIRAGLUTIDE | 40169222 | liraglutide |
| NATEGLINIDE | NATEGLINIDE | 1502826 | nateglinide |
| METFORMIN | METFORMIN | 40164908 | Metformin |
| METFORMIN | METFORMIN | 40164891 | Metformin |
| METFORMIN | METFORMIN | 19086484 | Metformin |
| GLIMEPIRIDE | GLIMEPIRIDE | 36222784 | glimepiride |
| EMPAGLIFLOZIN | EMPAGLIFLOZIN | 46287682 | empagliflozin |
| INSULIN | INSULIN | 1586327 | insulin, isophane |
| SITAGLIPTIN | SITAGLIPTIN | 19125042 | sitagliptin |
| METFORMIN | METFORMIN | 19113780 | Metformin |
| INSULIN | INSULIN | 19078558 | Insulin, Aspart, Human |
| METFORMIN | METFORMIN | 40164942 | Metformin |
| SAXAGLIPTIN | SAXAGLIPTIN | 40166045 | saxagliptin |

| | | | |
|---|---|---|---|
| PIOGLITAZONE | PIOGLITAZONE | 19099209 | pioglitazone |
| SITAGLIPTIN | SITAGLIPTIN | 19125042 | sitagliptin |
| METFORMIN | METFORMIN | 40164880 | Metformin |
| METFORMIN | METFORMIN | 40164912 | Metformin |
| INSULIN | INSULIN | 46233972 | Insulin Lispro |
| EXENATIDE | EXENATIDE | 1583722 | exenatide |
| INSULIN | INSULIN | 19095199 | Lente Insulin, Beef-Pork |
| SITAGLIPTIN | SITAGLIPTIN | 40243630 | sitagliptin |
| GLYBURIDE | GLYBURIDE | 40046201 | Glyburide |
| INSULIN | INSULIN | 36222190 | insulin detemir |
| GLIMEPIRIDE | GLIMEPIRIDE | 1597775 | glimepiride |
| INSULIN | INSULIN | 40176981 | Insulin, Aspart Protamine, Human |
| INSULIN | INSULIN | 36227902 | Insulin Lispro |
| INSULIN | INSULIN | 36227902 | Insulin Lispro |
| METFORMIN | METFORMIN | 40164946 | Metformin |
| CANAGLIFLOZIN | CANAGLIFLOZIN | 45774717 | canagliflozin |
| METFORMIN | METFORMIN | 40164897 | Metformin |
| INSULIN | INSULIN | 19120315 | insulin, isophane |
| GLIPIZIDE | GLIPIZIDE | 19047638 | Glipizide |
| NATEGLINIDE | NATEGLINIDE | 19045605 | nateglinide |
| INSULIN | INSULIN | 19012679 | Regular Insulin, Human |
| METFORMIN | METFORMIN | 36236323 | Metformin |
| METFORMIN | METFORMIN | 40063350 | Metformin |
| SITAGLIPTIN | SITAGLIPTIN | 36221233 | sitagliptin |
| SITAGLIPTIN | SITAGLIPTIN | 42900896 | sitagliptin |
| SITAGLIPTIN | SITAGLIPTIN | 19125043 | sitagliptin |
| SITAGLIPTIN | SITAGLIPTIN | 19125042 | sitagliptin |
| LINAGLIPTIN | LINAGLIPTIN | 36238286 | Linagliptin |
| GLIPIZIDE | GLIPIZIDE | 1560233 | Glipizide |
| INSULIN | INSULIN | 1550023 | Insulin Lispro |
| METFORMIN | METFORMIN | 1503328 | Metformin |
| INSULIN | INSULIN | 1502910 | Insulin Glargine |
| MIGLITOL | MIGLITOL | 36215710 | miglitol |
| INSULIN | INSULIN | 19094123 | Regular Insulin, Human |

| | | | |
|---|---|---|---|
| INSULIN | INSULIN | 19009384 | insulin, isophane |
| SITAGLIPTIN | SITAGLIPTIN | 19125047 | sitagliptin |
| GLYBURIDE | GLYBURIDE | 36213841 | Glyburide |
| INSULIN | INSULIN | 1597012 | Regular Insulin, Human |
| GLIPIZIDE | GLIPIZIDE | 1560171 | Glipizide |
| GLYBURIDE | GLYBURIDE | 1559782 | Glyburide |
| SITAGLIPTIN | SITAGLIPTIN | 42900896 | sitagliptin |
| NATEGLINIDE | NATEGLINIDE | 19107110 | nateglinide |
| INSULIN | INSULIN | 40166274 | Insulin Lispro |
| INSULIN | INSULIN | 19078558 | Insulin, Aspart, Human |
| METFORMIN | METFORMIN | 40164904 | Metformin |
| INSULIN | INSULIN | 19104459 | insulin, isophane |
| METFORMIN | METFORMIN | 1503327 | Metformin |
| GLIPIZIDE | GLIPIZIDE | 40165975 | Glipizide |
| INSULIN | INSULIN | 40184293 | Insulin, Aspart Protamine, Human |
| INSULIN | INSULIN | 1596977 | Regular Insulin, Human |
| METFORMIN | METFORMIN | 1503327 | Metformin |
| INSULIN | INSULIN | 46234049 | Insulin, Aspart, Human |
| INSULIN | INSULIN | 1513849 | Insulin, Zinc, Human |
| INSULIN | INSULIN | 19123376 | Insulin Lispro |
| INSULIN | INSULIN | 1596921 | insulin, isophane |
| INSULIN | INSULIN | 1596959 | insulin, isophane |
| PIOGLITAZONE | PIOGLITAZONE | 19099208 | pioglitazone |
| INSULIN | INSULIN | 1550023 | Insulin Lispro |
| GLYBURIDE | GLYBURIDE | 1559784 | Glyburide |
| METFORMIN | METFORMIN | 40164932 | Metformin |
| NATEGLINIDE | NATEGLINIDE | 19085097 | nateglinide |
| METFORMIN | METFORMIN | 40164943 | Metformin |
| INSULIN | INSULIN | 19010204 | insulin, isophane |
| INSULIN | INSULIN | 40166274 | Insulin Lispro |
| DULAGLUTIDE | DULAGLUTIDE | 45774445 | dulaglutide |
| REPAGLINIDE | REPAGLINIDE | 1516770 | repaglinide |
| PIOGLITAZONE | PIOGLITAZONE | 19099208 | pioglitazone |

| | | | |
|---|---|---|---|
| LIRAGLUTIDE | LIRAGLUTIDE | 40169223 | liraglutide |
| BROMOCRIPTINE | BROMOCRIPTINE | 730548 | Bromocriptine |
| INSULIN | INSULIN | 1513849 | Insulin, Zinc, Human |
| GLIMEPIRIDE | GLIMEPIRIDE | 1597756 | glimepiride |
| INSULIN | INSULIN | 1718889 | Regular Insulin, Human |
| METFORMIN | METFORMIN | 40164938 | Metformin |
| INSULIN | INSULIN | 19123376 | Insulin Lispro |
| GLYBURIDE | GLYBURIDE | 19077659 | Glyburide |
| INSULIN | INSULIN | 19116455 | insulin, isophane |
| METFORMIN | METFORMIN | 40164942 | Metformin |
| INSULIN | INSULIN | 1502905 | Insulin Glargine |
| INSULIN | INSULIN | 1550028 | Insulin Lispro |
| GLIPIZIDE | GLIPIZIDE | 1560171 | Glipizide |
| INSULIN | INSULIN | 1513881 | Insulin Lispro |
| EMPAGLIFLOZIN | EMPAGLIFLOZIN | 36248789 | empagliflozin |
| INSULIN | INSULIN | 1596977 | Regular Insulin, Human |
| INSULIN | INSULIN | 19131582 | Insulin, Glulisine, Human |
| INSULIN | INSULIN | 1513881 | Insulin Lispro |
| EXENATIDE | EXENATIDE | 19083253 | exenatide |
| PIOGLITAZONE | PIOGLITAZONE | 40099897 | pioglitazone |
| INSULIN | INSULIN | 1596999 | Regular Insulin, Human |
| INSULIN | INSULIN | 19010204 | insulin, isophane |
| METFORMIN | METFORMIN | 40164940 | Metformin |
| INSULIN | INSULIN | 19100128 | Regular Insulin, Human |
| PIOGLITAZONE | PIOGLITAZONE | 36236324 | pioglitazone |
| METFORMIN | METFORMIN | 40164938 | Metformin |
| GLIPIZIDE | GLIPIZIDE | 40044251 | Glipizide |
| METFORMIN | METFORMIN | 40063354 | Metformin |
| GLYBURIDE | GLYBURIDE | 19077682 | Glyburide |

| | | | |
|---|---|---|---|
| GLIMEPIRIDE | GLIMEPIRIDE | 43011830 | glimepiride |
| INSULIN | INSULIN | 1550027 | Insulin Lispro |
| INSULIN | INSULIN | 1596940 | insulin, isophane |
| METFORMIN | METFORMIN | 40164919 | Metformin |
| SITAGLIPTIN | SITAGLIPTIN | 19125042 | sitagliptin |
| METFORMIN | METFORMIN | 40164893 | Metformin |
| CANAGLIFLOZIN | CANAGLIFLOZIN | 36246877 | canagliflozin |
| GLIMEPIRIDE | GLIMEPIRIDE | 36229117 | glimepiride |
| METFORMIN | METFORMIN | 36223145 | Metformin |
| INSULIN | INSULIN | 19116454 | insulin, isophane |
| EXENATIDE | EXENATIDE | 42902527 | exenatide |
| INSULIN | INSULIN | 19111797 | insulin, isophane |
| INSULIN | INSULIN | 19120790 | Insulin Glargine |
| BROMOCRIPTINE | BROMOCRIPTINE | 730548 | Bromocriptine |
| ALOGLIPTIN | ALOGLIPTIN | 36246272 | alogliptin |
| INSULIN | INSULIN | 19078553 | insulin, isophane |
| METFORMIN | METFORMIN | 40164942 | Metformin |
| LIXISENATIDE | LIXISENATIDE | 44506754 | Lixisenatide |
| INSULIN | INSULIN | 1502905 | Insulin Glargine |
| INSULIN | INSULIN | 1567198 | Insulin, Aspart, Human |
| INSULIN | INSULIN | 40052047 | insulin, isophane |
| GLYBURIDE | GLYBURIDE | 36230036 | Glyburide |
| PIOGLITAZONE | PIOGLITAZONE | 19099209 | pioglitazone |
| BROMOCRIPTINE | BROMOCRIPTINE | 19083958 | Bromocriptine |
| INSULIN | INSULIN | 19095211 | Insulin Lispro |
| GLIMEPIRIDE | GLIMEPIRIDE | 1597774 | glimepiride |
| BROMOCRIPTINE | BROMOCRIPTINE | 19116171 | Bromocriptine |
| SITAGLIPTIN | SITAGLIPTIN | 40243628 | sitagliptin |
| GLYBURIDE | GLYBURIDE | 19077659 | Glyburide |
| INSULIN | INSULIN | 19010204 | insulin, isophane |
| CHLORPROPAMIDE | CHLORPROPAMIDE | 19060470 | Chlorpropamide |
| SITAGLIPTIN | SITAGLIPTIN | 36221233 | sitagliptin |
| GLIPIZIDE | GLIPIZIDE | 1560171 | Glipizide |
| INSULIN | INSULIN | 19120315 | insulin, isophane |
| METFORMIN | METFORMIN | 40164898 | Metformin |

| | | | |
|---|---|---|---|
| CANAGLIFLOZIN | CANAGLIFLOZIN | 43526468 | canagliflozin |
| MIGLITOL | MIGLITOL | 36233498 | miglitol |
| METFORMIN | METFORMIN | 40171449 | Metformin |
| PIOGLITAZONE | PIOGLITAZONE | 1525215 | pioglitazone |
| EMPAGLIFLOZIN | EMPAGLIFLOZIN | 45774758 | empagliflozin |
| INSULIN | INSULIN | 19078553 | insulin, isophane |
| METFORMIN | METFORMIN | 40063356 | Metformin |
| INSULIN | INSULIN | 19078603 | insulin, isophane |
| INSULIN | INSULIN | 19078553 | insulin, isophane |
| INSULIN | INSULIN | 46233948 | Insulin Lispro |
| INSULIN | INSULIN | 46234045 | Insulin, Aspart, Human |
| METFORMIN | METFORMIN | 36223134 | Metformin |
| INSULIN | INSULIN | 19078553 | insulin, isophane |
| EXENATIDE | EXENATIDE | 46234118 | exenatide |
| ACARBOSE | ACARBOSE | 19021787 | Acarbose |
| EXENATIDE | EXENATIDE | 19135404 | exenatide |
| METFORMIN | METFORMIN | 40164932 | Metformin |
| INSULIN | INSULIN | 19095214 | insulin, isophane |
| INSULIN | INSULIN | 19120790 | Insulin Glargine |
| INSULIN | INSULIN | 36227902 | Insulin Lispro |
| ACARBOSE | ACARBOSE | 19047612 | Acarbose |
| INSULIN | INSULIN | 36231450 | Insulin, Aspart, Human |
| METFORMIN | METFORMIN | 40173237 | Metformin |
| INSULIN | INSULIN | 42902607 | insulin, isophane |
| INSULIN | INSULIN | 1513881 | Insulin Lispro |
| BROMOCRIPTINE | BROMOCRIPTINE | 730551 | Bromocriptine |
| INSULIN | INSULIN | 1513876 | Insulin, Protamine Lispro, Human |
| GLYBURIDE | GLYBURIDE | 19060491 | Glyburide |
| GLIPIZIDE | GLIPIZIDE | 19134642 | Glipizide |
| INSULIN | INSULIN | 46233975 | Insulin Lispro |
| GLIPIZIDE | GLIPIZIDE | 19085004 | Glipizide |
| INSULIN | INSULIN | 19100131 | insulin, isophane |

| | | | |
|---|---|---|---|
| METFORMIN | METFORMIN | 1503297 | Metformin |
| SITAGLIPTIN | SITAGLIPTIN | 1580747 | sitagliptin |
| INSULIN | INSULIN | 1596956 | insulin, isophane |
| INSULIN | INSULIN | 36236032 | Insulin Lispro |
| LIRAGLUTIDE | LIRAGLUTIDE | 40170913 | liraglutide |
| GLYBURIDE | GLYBURIDE | 1559728 | Glyburide |
| TOLAZAMIDE | TOLAZAMIDE | 19036966 | Tolazamide |
| EXENATIDE | EXENATIDE | 1583727 | exenatide |
| INSULIN | INSULIN | 1550023 | Insulin Lispro |
| GLIMEPIRIDE | GLIMEPIRIDE | 1597798 | glimepiride |
| CANAGLIFLOZIN | CANAGLIFLOZIN | 45774721 | canagliflozin |
| GLYBURIDE | GLYBURIDE | 1559784 | Glyburide |
| EXENATIDE | EXENATIDE | 46234116 | exenatide |
| ALOGLIPTIN | ALOGLIPTIN | 43013912 | alogliptin |
| GLIMEPIRIDE | GLIMEPIRIDE | 1597775 | glimepiride |
| INSULIN | INSULIN | 40052088 | Insulin Lispro |
| INSULIN | INSULIN | 1596977 | Regular Insulin, Human |
| ALBIGLUTIDE | ALBIGLUTIDE | 44816283 | albiglutide |
| INSULIN | INSULIN | 35602731 | insulin degludec |
| GLIPIZIDE | GLIPIZIDE | 1560233 | Glipizide |
| INSULIN | INSULIN | 19012679 | Regular Insulin, Human |
| INSULIN | INSULIN | 19078553 | insulin, isophane |
| INSULIN | INSULIN | 46234043 | Insulin, Aspart, Human |
| INSULIN | INSULIN | 19012703 | insulin, isophane |
| METFORMIN | METFORMIN | 40173239 | Metformin |
| METFORMIN | METFORMIN | 36235752 | Metformin |
| NATEGLINIDE | NATEGLINIDE | 19107111 | nateglinide |
| INSULIN | INSULIN | 1596974 | insulin, isophane |
| SITAGLIPTIN | SITAGLIPTIN | 19125050 | sitagliptin |
| GLIPIZIDE | GLIPIZIDE | 40044251 | Glipizide |
| EMPAGLIFLOZIN | EMPAGLIFLOZIN | 45774895 | empagliflozin |
| METFORMIN | METFORMIN | 40164918 | Metformin |
| INSULIN | INSULIN | 1550023 | Insulin Lispro |

| | | | |
|---|---|---|---|
| METFORMIN | METFORMIN | 36223145 | Metformin |
| GLIPIZIDE | GLIPIZIDE | 19077637 | Glipizide |
| INSULIN | INSULIN | 1567232 | Insulin, Aspart, Human |
| INSULIN | INSULIN | 19116455 | insulin, isophane |
| INSULIN | INSULIN | 19078530 | insulin, isophane |
| GLIPIZIDE | GLIPIZIDE | 1560233 | Glipizide |
| INSULIN | INSULIN | 46234049 | Insulin, Aspart, Human |
| GLYBURIDE | GLYBURIDE | 1559782 | Glyburide |
| GLIPIZIDE | GLIPIZIDE | 19085004 | Glipizide |
| METFORMIN | METFORMIN | 40163924 | Metformin |
| INSULIN | INSULIN | 36248870 | Insulin Glargine |
| INSULIN | INSULIN | 1544873 | Insulin, Glulisine, Human |
| INSULIN | INSULIN | 1596977 | Regular Insulin, Human |
| METFORMIN | METFORMIN | 40164897 | Metformin |
| EMPAGLIFLOZIN | EMPAGLIFLOZIN | 36248790 | empagliflozin |
| INSULIN | INSULIN | 40051347 | Insulin Glargine |
| GLIPIZIDE | GLIPIZIDE | 36218767 | Glipizide |
| METFORMIN | METFORMIN | 40063352 | Metformin |
| METFORMIN | METFORMIN | 40164946 | Metformin |
| INSULIN | INSULIN | 46233972 | Insulin Lispro |
| GLIPIZIDE | GLIPIZIDE | 19081295 | Glipizide |
| SITAGLIPTIN | SITAGLIPTIN | 19125041 | sitagliptin |
| SITAGLIPTIN | SITAGLIPTIN | 40136770 | sitagliptin |
| METFORMIN | METFORMIN | 40164942 | Metformin |
| INSULIN | INSULIN | 1516981 | insulin detemir |
| INSULIN | INSULIN | 40166274 | Insulin Lispro |
| INSULIN | INSULIN | 36227902 | Insulin Lispro |
| INSULIN | INSULIN | 19078557 | Regular Insulin, Human |
| INSULIN | INSULIN | 1567232 | Insulin, Aspart, Human |
| GLIMEPIRIDE | GLIMEPIRIDE | 1597756 | glimepiride |
| METFORMIN | METFORMIN | 1503297 | Metformin |
| INSULIN | INSULIN | 1567232 | Insulin, Aspart, Human |
| GLIPIZIDE | GLIPIZIDE | 19047638 | Glipizide |

| | | | |
|---|---|---|---|
| REPAGLINIDE | REPAGLINIDE | 1516766 | repaglinide |
| METFORMIN | METFORMIN | 40164929 | Metformin |
| INSULIN | INSULIN | 40051348 | Insulin Glargine |
| GLYBURIDE | GLYBURIDE | 1559785 | Glyburide |
| INSULIN | INSULIN | 19062999 | insulin, isophane |
| DULAGLUTIDE | DULAGLUTIDE | 45774435 | dulaglutide |
| INSULIN | INSULIN | 36231456 | insulin, isophane |
| INSULIN | INSULIN | 19094122 | Regular Insulin, Human |
| GLIMEPIRIDE | GLIMEPIRIDE | 36234085 | glimepiride |
| INSULIN | INSULIN | 19078558 | Insulin, Aspart, Human |
| EMPAGLIFLOZIN | EMPAGLIFLOZIN | 45774758 | empagliflozin |
| INSULIN | INSULIN | 40052088 | Insulin Lispro |
| INSULIN | INSULIN | 1596999 | Regular Insulin, Human |
| INSULIN | INSULIN | 1502905 | Insulin Glargine |
| INSULIN | INSULIN | 19016117 | Insulin Lispro |
| INSULIN | INSULIN | 19095211 | Insulin Lispro |
| INSULIN | INSULIN | 19071700 | Insulin Glargine |
| INSULIN | INSULIN | 19078527 | Insulin, Zinc, Human |
| INSULIN | INSULIN | 1550027 | Insulin Lispro |
| SAXAGLIPTIN | SAXAGLIPTIN | 40166035 | saxagliptin |
| INSULIN | INSULIN | 19078552 | insulin, isophane |
| INSULIN | INSULIN | 46233950 | Insulin Lispro |
| INSULIN | INSULIN | 46234045 | Insulin, Aspart, Human |
| INSULIN | INSULIN | 42902743 | Insulin Glargine |
| EXENATIDE | EXENATIDE | 1583730 | exenatide |
| INSULIN | INSULIN | 19044971 | insulin detemir |
| CANAGLIFLOZIN | CANAGLIFLOZIN | 43526471 | canagliflozin |
| METFORMIN | METFORMIN | 40164932 | Metformin |
| METFORMIN | METFORMIN | 40164940 | Metformin |
| INSULIN | INSULIN | 46234241 | Regular Insulin, Human |
| INSULIN | INSULIN | 19014975 | insulin, isophane |
| GLIMEPIRIDE | GLIMEPIRIDE | 36234086 | glimepiride |
| TOLBUTAMIDE | TOLBUTAMIDE | 1502860 | Tolbutamide |
| INSULIN | INSULIN | 19078554 | Regular Insulin, Human |
| EMPAGLIFLOZIN | EMPAGLIFLOZIN | 45774754 | empagliflozin |

| | | | |
|---|---|---|---|
| INSULIN | INSULIN | 1596977 | Regular Insulin, Human |
| LINAGLIPTIN | LINAGLIPTIN | 40239216 | Linagliptin |
| METFORMIN | METFORMIN | 1503327 | Metformin |
| CHLORPROPAMIDE | CHLORPROPAMIDE | 40030456 | Chlorpropamide |
| GLIPIZIDE | GLIPIZIDE | 40167637 | Glipizide |
| INSULIN | INSULIN | 19010204 | insulin, isophane |
| METFORMIN | METFORMIN | 40173239 | Metformin |
| GLYBURIDE | GLYBURIDE | 36231170 | Glyburide |
| GLIMEPIRIDE | GLIMEPIRIDE | 43011499 | glimepiride |
| METFORMIN | METFORMIN | 40164880 | Metformin |
| INSULIN | INSULIN | 19135263 | insulin, isophane |
| METFORMIN | METFORMIN | 40117785 | Metformin |
| INSULIN | INSULIN | 46234240 | insulin, isophane |
| INSULIN | INSULIN | 19078553 | insulin, isophane |
| EXENATIDE | EXENATIDE | 46234118 | exenatide |
| INSULIN | INSULIN | 46233948 | Insulin Lispro |
| INSULIN | INSULIN | 40051348 | Insulin Glargine |
| INSULIN | INSULIN | 19120790 | Insulin Glargine |
| INSULIN | INSULIN | 1596956 | insulin, isophane |
| GLIMEPIRIDE | GLIMEPIRIDE | 43011830 | glimepiride |
| BROMOCRIPTINE | BROMOCRIPTINE | 19022176 | Bromocriptine |
| INSULIN | INSULIN | 46234045 | Insulin, Aspart, Human |
| INSULIN | INSULIN | 19094122 | Regular Insulin, Human |
| LIRAGLUTIDE | LIRAGLUTIDE | 40169223 | liraglutide |
| EMPAGLIFLOZIN | EMPAGLIFLOZIN | 45774895 | empagliflozin |
| INSULIN | INSULIN | 35604094 | Insulin Glargine |
| SITAGLIPTIN | SITAGLIPTIN | 42900896 | sitagliptin |
| BROMOCRIPTINE | BROMOCRIPTINE | 730548 | Bromocriptine |
| GLIPIZIDE | GLIPIZIDE | 19085004 | Glipizide |
| GLIPIZIDE | GLIPIZIDE | 19047638 | Glipizide |
| INSULIN | INSULIN | 19120315 | insulin, isophane |
| ALOGLIPTIN | ALOGLIPTIN | 43013916 | alogliptin |
| METFORMIN | METFORMIN | 40164929 | Metformin |
| LIRAGLUTIDE | LIRAGLUTIDE | 45892686 | liraglutide |

| | | | |
|---|---|---|---|
| INSULIN | INSULIN | 1544838 | Insulin, Glulisine, Human |
| PIOGLITAZONE | PIOGLITAZONE | 1525215 | pioglitazone |
| PIOGLITAZONE | PIOGLITAZONE | 36236325 | pioglitazone |
| METFORMIN | METFORMIN | 40164923 | Metformin |
| INSULIN | INSULIN | 36248195 | insulin degludec |
| INSULIN | INSULIN | 1596916 | insulin, isophane |
| EXENATIDE | EXENATIDE | 1583727 | exenatide |
| SITAGLIPTIN | SITAGLIPTIN | 19125050 | sitagliptin |
| INSULIN | INSULIN | 35604094 | Insulin Glargine |
| INSULIN | INSULIN | 46234045 | Insulin, Aspart, Human |
| INSULIN | INSULIN | 1586344 | insulin, isophane |
| GLYBURIDE | GLYBURIDE | 1559784 | Glyburide |
| GLYBURIDE | GLYBURIDE | 1559684 | Glyburide |
| INSULIN | INSULIN | 46234049 | Insulin, Aspart, Human |
| METFORMIN | METFORMIN | 40164890 | Metformin |
| INSULIN | INSULIN | 19062999 | insulin, isophane |
| GLIPIZIDE | GLIPIZIDE | 40167637 | Glipizide |
| METFORMIN | METFORMIN | 40164946 | Metformin |
| BROMOCRIPTINE | BROMOCRIPTINE | 19007101 | Bromocriptine |
| EXENATIDE | EXENATIDE | 19135404 | exenatide |
| EXENATIDE | EXENATIDE | 46234116 | exenatide |
| INSULIN | INSULIN | 1544873 | Insulin, Glulisine, Human |
| INSULIN | INSULIN | 19110352 | insulin, isophane |
| METFORMIN | METFORMIN | 1503327 | Metformin |
| METFORMIN | METFORMIN | 42708175 | Metformin |
| GLYBURIDE | GLYBURIDE | 1559684 | Glyburide |
| GLIMEPIRIDE | GLIMEPIRIDE | 1597775 | glimepiride |
| BROMOCRIPTINE | BROMOCRIPTINE | 730548 | Bromocriptine |
| GLYBURIDE | GLYBURIDE | 40168374 | Glyburide |
| METFORMIN | METFORMIN | 40063356 | Metformin |
| GLIPIZIDE | GLIPIZIDE | 40167636 | Glipizide |
| INSULIN | INSULIN | 19095218 | Regular Insulin, Human |
| CANAGLIFLOZIN | CANAGLIFLOZIN | 43526475 | canagliflozin |
| EXENATIDE | EXENATIDE | 19083254 | exenatide |

| | | | |
|---|---|---|---|
| INSULIN | INSULIN | 35605622 | Regular Insulin, Human |
| INSULIN | INSULIN | 19062999 | insulin, isophane |
| DAPAGLIFLOZIN | DAPAGLIFLOZIN | 44816049 | dapagliflozin |
| INSULIN | INSULIN | 40176981 | Insulin, Aspart Protamine, Human |
| EXENATIDE | EXENATIDE | 19135404 | exenatide |
| INSULIN | INSULIN | 19120790 | Insulin Glargine |
| INSULIN | INSULIN | 1502908 | Insulin Glargine |
| GLIPIZIDE | GLIPIZIDE | 1560171 | Glipizide |
| INSULIN | INSULIN | 19078553 | insulin, isophane |
| LINAGLIPTIN | LINAGLIPTIN | 40239220 | Linagliptin |
| METFORMIN | METFORMIN | 40164929 | Metformin |
| ALOGLIPTIN | ALOGLIPTIN | 43013929 | alogliptin |
| GLYBURIDE | GLYBURIDE | 40168376 | Glyburide |
| PIOGLITAZONE | PIOGLITAZONE | 19099209 | pioglitazone |
| INSULIN | INSULIN | 35604094 | Insulin Glargine |
| SITAGLIPTIN | SITAGLIPTIN | 19125050 | sitagliptin |
| METFORMIN | METFORMIN | 1503297 | Metformin |
| MIGLITOL | MIGLITOL | 1510202 | miglitol |
| INSULIN | INSULIN | 1550027 | Insulin Lispro |
| INSULIN | INSULIN | 1596941 | insulin, isophane |
| GLIPIZIDE | GLIPIZIDE | 19077636 | Glipizide |
| INSULIN | INSULIN | 19110354 | insulin, isophane |
| SITAGLIPTIN | SITAGLIPTIN | 1580747 | sitagliptin |
| INSULIN | INSULIN | 1567198 | Insulin, Aspart, Human |
| BROMOCRIPTINE | BROMOCRIPTINE | 40164160 | Bromocriptine |
| GLIMEPIRIDE | GLIMEPIRIDE | 1597756 | glimepiride |
| GLIMEPIRIDE | GLIMEPIRIDE | 1597774 | glimepiride |
| GLIPIZIDE | GLIPIZIDE | 19081295 | Glipizide |
| GLIMEPIRIDE | GLIMEPIRIDE | 43011499 | glimepiride |
| METFORMIN | METFORMIN | 42708177 | Metformin |
| METFORMIN | METFORMIN | 40164932 | Metformin |
| METFORMIN | METFORMIN | 40164900 | Metformin |
| PIOGLITAZONE | PIOGLITAZONE | 19099209 | pioglitazone |
| INSULIN | INSULIN | 19120790 | Insulin Glargine |
| INSULIN | INSULIN | 19112791 | Insulin, Glulisine, Human |

| | | | |
|---|---|---|---|
| INSULIN | INSULIN | 19095211 | Insulin Lispro |
| EXENATIDE | EXENATIDE | 42903341 | exenatide |
| GLIPIZIDE | GLIPIZIDE | 19030443 | Glipizide |
| INSULIN | INSULIN | 19094122 | Regular Insulin, Human |
| BROMOCRIPTINE | BROMOCRIPTINE | 40016899 | Bromocriptine |
| INSULIN | INSULIN | 1596959 | insulin, isophane |
| INSULIN | INSULIN | 19078553 | insulin, isophane |
| EXENATIDE | EXENATIDE | 1583728 | exenatide |
| PIOGLITAZONE | PIOGLITAZONE | 19099207 | pioglitazone |
| INSULIN | INSULIN | 19078527 | Insulin, Zinc, Human |
| GLYBURIDE | GLYBURIDE | 36235751 | Glyburide |
| GLIMEPIRIDE | GLIMEPIRIDE | 36214984 | glimepiride |
| CANAGLIFLOZIN | CANAGLIFLOZIN | 45774717 | canagliflozin |
| METFORMIN | METFORMIN | 36236480 | Metformin |
| INSULIN | INSULIN | 19078557 | Regular Insulin, Human |
| GLIPIZIDE | GLIPIZIDE | 1560171 | Glipizide |
| PIOGLITAZONE | PIOGLITAZONE | 19099208 | pioglitazone |
| INSULIN | INSULIN | 19116454 | insulin, isophane |
| INSULIN | INSULIN | 1550028 | Insulin Lispro |
| PIOGLITAZONE | PIOGLITAZONE | 19099207 | pioglitazone |
| INSULIN | INSULIN | 19095214 | insulin, isophane |
| INSULIN | INSULIN | 46234049 | Insulin, Aspart, Human |
| METFORMIN | METFORMIN | 40164897 | Metformin |
| INSULIN | INSULIN | 36231450 | Insulin, Aspart, Human |
| INSULIN | INSULIN | 46233975 | Insulin Lispro |
| METFORMIN | METFORMIN | 1503297 | Metformin |
| INSULIN | INSULIN | 19078557 | Regular Insulin, Human |
| INSULIN | INSULIN | 40184294 | Insulin, Aspart Protamine, Human |
| PRAMLINTIDE | PRAMLINTIDE | 40165687 | Pramlintide |
| INSULIN | INSULIN | 46234045 | Insulin, Aspart, Human |
| GLIMEPIRIDE | GLIMEPIRIDE | 19017144 | glimepiride |
| NATEGLINIDE | NATEGLINIDE | 19045606 | nateglinide |
| GLIMEPIRIDE | GLIMEPIRIDE | 19023585 | glimepiride |

| | | | |
|---|---|---|---|
| GLIMEPIRIDE | GLIMEPIRIDE | 36234085 | glimepiride |
| MIGLITOL | MIGLITOL | 36215710 | miglitol |
| METFORMIN | METFORMIN | 40126695 | Metformin |
| ACARBOSE | ACARBOSE | 19047612 | Acarbose |
| DULAGLUTIDE | DULAGLUTIDE | 45774435 | dulaglutide |
| BROMOCRIPTINE | BROMOCRIPTINE | 19007102 | Bromocriptine |
| GLIPIZIDE | GLIPIZIDE | 19030445 | Glipizide |
| LIRAGLUTIDE | LIRAGLUTIDE | 40169222 | liraglutide |
| INSULIN | INSULIN | 19135320 | Regular Insulin, Human |
| INSULIN | INSULIN | 19135320 | Regular Insulin, Human |
| INSULIN | INSULIN | 36236032 | Insulin Lispro |
| INSULIN | INSULIN | 1596960 | insulin, isophane |
| INSULIN | INSULIN | 36235841 | Regular Insulin, Human |
| INSULIN | INSULIN | 19110356 | Regular Insulin, Human |
| LINAGLIPTIN | LINAGLIPTIN | 42708091 | Linagliptin |
| INSULIN | INSULIN | 19078530 | insulin, isophane |
| INSULIN | INSULIN | 40052088 | Insulin Lispro |
| INSULIN | INSULIN | 19078556 | Regular Insulin, Human |
| INSULIN | INSULIN | 19078633 | Insulin, Regular, Pork |
| INSULIN | INSULIN | 1502905 | Insulin Glargine |
| METFORMIN | METFORMIN | 40164930 | Metformin |
| INSULIN | INSULIN | 1502905 | Insulin Glargine |
| METFORMIN | METFORMIN | 40163922 | Metformin |
| METFORMIN | METFORMIN | 40063356 | Metformin |
| CHLORPROPAMIDE | CHLORPROPAMIDE | 19088819 | Chlorpropamide |
| LIRAGLUTIDE | LIRAGLUTIDE | 40170913 | liraglutide |
| METFORMIN | METFORMIN | 40117785 | Metformin |
| INSULIN | INSULIN | 46221557 | Insulin Glargine |
| ACARBOSE | ACARBOSE | 19047612 | Acarbose |
| INSULIN | INSULIN | 1567232 | Insulin, Aspart, Human |
| PIOGLITAZONE | PIOGLITAZONE | 19099207 | pioglitazone |
| INSULIN | INSULIN | 46221552 | Insulin Glargine |
| EXENATIDE | EXENATIDE | 36234709 | exenatide |

| | | | |
|---|---|---|---|
| INSULIN | INSULIN | 19116453 | insulin, isophane |
| GLIMEPIRIDE | GLIMEPIRIDE | 1597760 | glimepiride |
| INSULIN | INSULIN | 1502905 | Insulin Glargine |
| METFORMIN | METFORMIN | 40164896 | Metformin |
| METFORMIN | METFORMIN | 1503328 | Metformin |
| INSULIN | INSULIN | 1596977 | Regular Insulin, Human |
| INSULIN | INSULIN | 19078551 | insulin, isophane |
| INSULIN | INSULIN | 1513912 | Insulin Lispro |
| INSULIN | INSULIN | 19078553 | insulin, isophane |
| INSULIN | INSULIN | 36229086 | Insulin Glargine |
| INSULIN | INSULIN | 19071700 | Insulin Glargine |
| CANAGLIFLOZIN | CANAGLIFLOZIN | 45774721 | canagliflozin |
| PIOGLITAZONE | PIOGLITAZONE | 1525215 | pioglitazone |
| GLIPIZIDE | GLIPIZIDE | 19100559 | Glipizide |
| METFORMIN | METFORMIN | 40164901 | Metformin |
| GLYBURIDE | GLYBURIDE | 36213841 | Glyburide |
| ROSIGLITAZONE | ROSIGLITAZONE | 36229365 | rosiglitazone |
| METFORMIN | METFORMIN | 40164932 | Metformin |
| GLIPIZIDE | GLIPIZIDE | 40166816 | Glipizide |
| METFORMIN | METFORMIN | 40164898 | Metformin |
| NATEGLINIDE | NATEGLINIDE | 19045605 | nateglinide |
| METFORMIN | METFORMIN | 40173239 | Metformin |
| INSULIN | INSULIN | 46233946 | Insulin Lispro |
| METFORMIN | METFORMIN | 40164929 | Metformin |
| METFORMIN | METFORMIN | 40126689 | Metformin |
| GLIMEPIRIDE | GLIMEPIRIDE | 1597756 | glimepiride |
| GLIPIZIDE | GLIPIZIDE | 19117945 | Glipizide |
| INSULIN | INSULIN | 19078554 | Regular Insulin, Human |
| BROMOCRIPTINE | BROMOCRIPTINE | 19007101 | Bromocriptine |
| CANAGLIFLOZIN | CANAGLIFLOZIN | 36246430 | canagliflozin |
| INSULIN | INSULIN | 46233948 | Insulin Lispro |

| | | | |
|---|---|---|---|
| METFORMIN | METFORMIN | 40164904 | Metformin |
| INSULIN | INSULIN | 46233975 | Insulin Lispro |
| INSULIN | INSULIN | 40184293 | Insulin, Aspart Protamine, Human |
| INSULIN | INSULIN | 1567198 | Insulin, Aspart, Human |
| METFORMIN | METFORMIN | 40164894 | Metformin |
| INSULIN | INSULIN | 19135263 | insulin, isophane |
| GLIPIZIDE | GLIPIZIDE | 19085004 | Glipizide |
| INSULIN | INSULIN | 19111797 | insulin, isophane |
| INSULIN | INSULIN | 40051348 | Insulin Glargine |
| GLIPIZIDE | GLIPIZIDE | 40166815 | Glipizide |
| METFORMIN | METFORMIN | 40164938 | Metformin |
| INSULIN | INSULIN | 1502905 | Insulin Glargine |
| INSULIN | INSULIN | 19062999 | insulin, isophane |
| INSULIN | INSULIN | 45776625 | Regular Insulin, Human |
| GLYBURIDE | GLYBURIDE | 1559684 | Glyburide |
| INSULIN | INSULIN | 1516981 | insulin detemir |
| SITAGLIPTIN | SITAGLIPTIN | 1580747 | sitagliptin |
| INSULIN | INSULIN | 1596977 | Regular Insulin, Human |
| INSULIN | INSULIN | 19078530 | insulin, isophane |
| METFORMIN | METFORMIN | 40164929 | Metformin |
| EXENATIDE | EXENATIDE | 46234118 | exenatide |
| INSULIN | INSULIN | 1596952 | insulin, isophane |
| INSULIN | INSULIN | 40051347 | Insulin Glargine |
| EXENATIDE | EXENATIDE | 42903341 | exenatide |
| INSULIN | INSULIN | 1597001 | Regular Insulin, Human |
| INSULIN | INSULIN | 1567232 | Insulin, Aspart, Human |
| INSULIN | INSULIN | 40166274 | Insulin Lispro |
| METFORMIN | METFORMIN | 36235746 | Metformin |
| INSULIN | INSULIN | 19078633 | Insulin, Regular, Pork |
| GLIMEPIRIDE | GLIMEPIRIDE | 19100593 | glimepiride |
| GLIMEPIRIDE | GLIMEPIRIDE | 1597776 | glimepiride |
| GLIPIZIDE | GLIPIZIDE | 19030445 | Glipizide |
| GLIMEPIRIDE | GLIMEPIRIDE | 1597775 | glimepiride |

| | | | |
|---|---|---|---|
| INSULIN | INSULIN | 19131582 | Insulin, Glulisine, Human |
| GLYBURIDE | GLYBURIDE | 1559684 | Glyburide |
| ALBIGLUTIDE | ALBIGLUTIDE | 44816274 | albiglutide |
| INSULIN | INSULIN | 1567232 | Insulin, Aspart, Human |
| INSULIN | INSULIN | 19095213 | insulin, isophane |
| GLIPIZIDE | GLIPIZIDE | 1560171 | Glipizide |
| INSULIN | INSULIN | 19135320 | Regular Insulin, Human |
| METFORMIN | METFORMIN | 40146999 | Metformin |
| METFORMIN | METFORMIN | 40164880 | Metformin |
| GLYBURIDE | GLYBURIDE | 1559684 | Glyburide |
| SITAGLIPTIN | SITAGLIPTIN | 19125042 | sitagliptin |
| DULAGLUTIDE | DULAGLUTIDE | 45774435 | dulaglutide |
| METFORMIN | METFORMIN | 40229046 | Metformin |
| INSULIN | INSULIN | 1596977 | Regular Insulin, Human |
| SITAGLIPTIN | SITAGLIPTIN | 19125042 | sitagliptin |
| CANAGLIFLOZIN | CANAGLIFLOZIN | 36246430 | canagliflozin |
| INSULIN | INSULIN | 46233943 | Insulin Lispro |
| INSULIN | INSULIN | 19078633 | Insulin, Regular, Pork |
| PRAMLINTIDE | PRAMLINTIDE | 40163890 | Pramlintide |
| INSULIN | INSULIN | 19078527 | Insulin, Zinc, Human |
| EXENATIDE | EXENATIDE | 45774533 | exenatide |
| METFORMIN | METFORMIN | 1503328 | Metformin |
| NATEGLINIDE | NATEGLINIDE | 40058199 | nateglinide |
| INSULIN | INSULIN | 19044971 | insulin detemir |
| NATEGLINIDE | NATEGLINIDE | 40058198 | nateglinide |
| METFORMIN | METFORMIN | 40164925 | Metformin |
| LIRAGLUTIDE | LIRAGLUTIDE | 40170911 | liraglutide |
| DAPAGLIFLOZIN | DAPAGLIFLOZIN | 45775388 | dapagliflozin |
| METFORMIN | METFORMIN | 40164929 | Metformin |
| ROSIGLITAZONE | ROSIGLITAZONE | 36227624 | rosiglitazone |
| INSULIN | INSULIN | 19014975 | insulin, isophane |
| GLIMEPIRIDE | GLIMEPIRIDE | 1597773 | glimepiride |
| SITAGLIPTIN | SITAGLIPTIN | 40136770 | sitagliptin |
| GLIMEPIRIDE | GLIMEPIRIDE | 1597774 | glimepiride |
| INSULIN | INSULIN | 40166274 | Insulin Lispro |

| | | | |
|---|---|---|---|
| GLIPIZIDE | GLIPIZIDE | 40165976 | Glipizide |
| INSULIN | INSULIN | 19095218 | Regular Insulin, Human |
| GLIMEPIRIDE | GLIMEPIRIDE | 19017142 | glimepiride |
| METFORMIN | METFORMIN | 40164940 | Metformin |
| INSULIN | INSULIN | 19110355 | insulin, isophane |
| ALOGLIPTIN | ALOGLIPTIN | 43013884 | alogliptin |
| EXENATIDE | EXENATIDE | 1583722 | exenatide |
| INSULIN | INSULIN | 19061670 | Regular Insulin, Human |
| INSULIN | INSULIN | 19100127 | insulin, isophane |
| INSULIN | INSULIN | 35605622 | Regular Insulin, Human |
| METFORMIN | METFORMIN | 40164947 | Metformin |
| INSULIN | INSULIN | 19044971 | insulin detemir |
| GLYBURIDE | GLYBURIDE | 1559784 | Glyburide |
| GLYBURIDE | GLYBURIDE | 1559684 | Glyburide |
| GLYBURIDE | GLYBURIDE | 36233496 | Glyburide |
| TOLBUTAMIDE | TOLBUTAMIDE | 1502913 | Tolbutamide |
| INSULIN | INSULIN | 19116559 | insulin, isophane |
| METFORMIN | METFORMIN | 40164882 | Metformin |
| GLYBURIDE | GLYBURIDE | 1559785 | Glyburide |
| METFORMIN | METFORMIN | 40164925 | Metformin |
| INSULIN | INSULIN | 19071700 | Insulin Glargine |
| INSULIN | INSULIN | 36231450 | Insulin, Aspart, Human |
| GLYBURIDE | GLYBURIDE | 40166003 | Glyburide |
| INSULIN | INSULIN | 19110354 | insulin, isophane |
| INSULIN | INSULIN | 19071700 | Insulin Glargine |
| INSULIN | INSULIN | 46233730 | Regular Insulin, Human |
| NATEGLINIDE | NATEGLINIDE | 1502826 | nateglinide |
| INSULIN | INSULIN | 19078530 | insulin, isophane |
| ALOGLIPTIN | ALOGLIPTIN | 36246195 | alogliptin |
| INSULIN | INSULIN | 1550023 | Insulin Lispro |
| GLYBURIDE | GLYBURIDE | 1559784 | Glyburide |
| SAXAGLIPTIN | SAXAGLIPTIN | 40166043 | saxagliptin |
| INSULIN | INSULIN | 36229086 | Insulin Glargine |
| METFORMIN | METFORMIN | 40164929 | Metformin |

| | | | |
|---|---|---|---|
| GLIPIZIDE | GLIPIZIDE | 19047638 | Glipizide |
| INSULIN | INSULIN | 46234045 | Insulin, Aspart, Human |
| SITAGLIPTIN | SITAGLIPTIN | 19125042 | sitagliptin |
| INSULIN | INSULIN | 1596953 | insulin, isophane |
| INSULIN | INSULIN | 46233975 | Insulin Lispro |
| INSULIN | INSULIN | 40184293 | Insulin, Aspart Protamine, Human |
| INSULIN | INSULIN | 1513916 | Insulin Lispro |
| GLIMEPIRIDE | GLIMEPIRIDE | 1597775 | glimepiride |
| INSULIN | INSULIN | 46233975 | Insulin Lispro |
| CHLORPROPAMIDE | CHLORPROPAMIDE | 1594973 | Chlorpropamide |
| METFORMIN | METFORMIN | 40164932 | Metformin |
| EXENATIDE | EXENATIDE | 19083253 | exenatide |
| PIOGLITAZONE | PIOGLITAZONE | 19099209 | pioglitazone |
| INSULIN | INSULIN | 1596999 | Regular Insulin, Human |
| INSULIN | INSULIN | 46233975 | Insulin Lispro |
| GLYBURIDE | GLYBURIDE | 1559684 | Glyburide |
| INSULIN | INSULIN | 19071700 | Insulin Glargine |
| INSULIN | INSULIN | 19078555 | Regular Insulin, Human |
| ROSIGLITAZONE | ROSIGLITAZONE | 1547506 | rosiglitazone |
| INSULIN | INSULIN | 46233975 | Insulin Lispro |
| EMPAGLIFLOZIN | EMPAGLIFLOZIN | 45774756 | empagliflozin |
| CHLORPROPAMIDE | CHLORPROPAMIDE | 36231728 | Chlorpropamide |
| INSULIN | INSULIN | 1550023 | Insulin Lispro |
| METFORMIN | METFORMIN | 40164899 | Metformin |
| GLIMEPIRIDE | GLIMEPIRIDE | 19117199 | glimepiride |
| GLIPIZIDE | GLIPIZIDE | 1560171 | Glipizide |
| METFORMIN | METFORMIN | 1503297 | Metformin |
| METFORMIN | METFORMIN | 40164926 | Metformin |
| INSULIN | INSULIN | 1502910 | Insulin Glargine |
| GLYBURIDE | GLYBURIDE | 19030579 | Glyburide |
| GLIMEPIRIDE | GLIMEPIRIDE | 36229116 | glimepiride |
| METFORMIN | METFORMIN | 40164930 | Metformin |
| INSULIN | INSULIN | 1596975 | insulin, isophane |
| INSULIN | INSULIN | 1596977 | Regular Insulin, Human |
| LIRAGLUTIDE | LIRAGLUTIDE | 40170911 | liraglutide |

| | | | |
|---|---|---|---|
| SITAGLIPTIN | SITAGLIPTIN | 19125040 | sitagliptin |
| METFORMIN | METFORMIN | 42708175 | Metformin |
| LIRAGLUTIDE | LIRAGLUTIDE | 45892686 | liraglutide |
| INSULIN | INSULIN | 46233946 | Insulin Lispro |
| METFORMIN | METFORMIN | 19106521 | Metformin |
| INSULIN | INSULIN | 1550023 | Insulin Lispro |
| INSULIN | INSULIN | 19104092 | Regular Insulin, Human |
| INSULIN | INSULIN | 19110356 | Regular Insulin, Human |
| GLIPIZIDE | GLIPIZIDE | 19134624 | Glipizide |
| METFORMIN | METFORMIN | 40164929 | Metformin |
| INSULIN | INSULIN | 46275390 | Regular Insulin, Human |
| METFORMIN | METFORMIN | 36223145 | Metformin |
| GLIMEPIRIDE | GLIMEPIRIDE | 1597756 | glimepiride |
| INSULIN | INSULIN | 19116559 | insulin, isophane |
| METFORMIN | METFORMIN | 40164919 | Metformin |
| METFORMIN | METFORMIN | 40163929 | Metformin |
| GLYBURIDE | GLYBURIDE | 1559684 | Glyburide |
| DULAGLUTIDE | DULAGLUTIDE | 36247186 | dulaglutide |
| INSULIN | INSULIN | 46233975 | Insulin Lispro |
| GLYBURIDE | GLYBURIDE | 1559782 | Glyburide |
| INSULIN | INSULIN | 19062999 | insulin, isophane |
| METFORMIN | METFORMIN | 40164924 | Metformin |
| SAXAGLIPTIN | SAXAGLIPTIN | 36229291 | saxagliptin |
| INSULIN | INSULIN | 46233975 | Insulin Lispro |
| INSULIN | INSULIN | 19123376 | Insulin Lispro |
| ACARBOSE | ACARBOSE | 1529331 | Acarbose |
| BROMOCRIPTINE | BROMOCRIPTINE | 730548 | Bromocriptine |
| INSULIN | INSULIN | 19078530 | insulin, isophane |
| NATEGLINIDE | NATEGLINIDE | 19045606 | nateglinide |
| GLIPIZIDE | GLIPIZIDE | 19047638 | Glipizide |
| LIRAGLUTIDE | LIRAGLUTIDE | 45892684 | liraglutide |
| METFORMIN | METFORMIN | 40164930 | Metformin |
| REPAGLINIDE | REPAGLINIDE | 1516766 | repaglinide |

| | | | |
|---|---|---|---|
| CANAGLIFLOZIN | CANAGLIFLOZIN | 43526468 | canagliflozin |
| GLIPIZIDE | GLIPIZIDE | 1560233 | Glipizide |
| PIOGLITAZONE | PIOGLITAZONE | 19099209 | pioglitazone |
| METFORMIN | METFORMIN | 40164900 | Metformin |
| GLYBURIDE | GLYBURIDE | 1559760 | Glyburide |
| EXENATIDE | EXENATIDE | 1583730 | exenatide |
| NATEGLINIDE | NATEGLINIDE | 1502827 | nateglinide |
| METFORMIN | METFORMIN | 36242510 | Metformin |
| TOLAZAMIDE | TOLAZAMIDE | 36214131 | Tolazamide |
| DULAGLUTIDE | DULAGLUTIDE | 45774448 | dulaglutide |
| DULAGLUTIDE | DULAGLUTIDE | 45774440 | dulaglutide |
| GLIMEPIRIDE | GLIMEPIRIDE | 1597776 | glimepiride |
| INSULIN | INSULIN | 1596998 | Regular Insulin, Human |
| INSULIN | INSULIN | 35602723 | insulin degludec |
| METFORMIN | METFORMIN | 40164897 | Metformin |
| INSULIN | INSULIN | 1567198 | Insulin, Aspart, Human |
| SITAGLIPTIN | SITAGLIPTIN | 19125046 | sitagliptin |
| GLYBURIDE | GLYBURIDE | 1559784 | Glyburide |
| INSULIN | INSULIN | 19135263 | insulin, isophane |
| ALOGLIPTIN | ALOGLIPTIN | 43013903 | alogliptin |
| GLIMEPIRIDE | GLIMEPIRIDE | 1597801 | glimepiride |
| GLYBURIDE | GLYBURIDE | 19077682 | Glyburide |
| REPAGLINIDE | REPAGLINIDE | 1516794 | repaglinide |
| GLYBURIDE | GLYBURIDE | 36235750 | Glyburide |
| METFORMIN | METFORMIN | 1503297 | Metformin |
| INSULIN | INSULIN | 19014975 | insulin, isophane |
| INSULIN | INSULIN | 36248196 | insulin degludec |
| INSULIN | INSULIN | 1567198 | Insulin, Aspart, Human |
| INSULIN | INSULIN | 36227902 | Insulin Lispro |
| PIOGLITAZONE | PIOGLITAZONE | 19099209 | pioglitazone |
| INSULIN | INSULIN | 40184294 | Insulin, Aspart Protamine, Human |
| INSULIN | INSULIN | 1596941 | insulin, isophane |
| INSULIN | INSULIN | 46233728 | Regular Insulin, Human |
| INSULIN | INSULIN | 1513912 | Insulin Lispro |
| METFORMIN | METFORMIN | 40126695 | Metformin |
| TOLAZAMIDE | TOLAZAMIDE | 19036967 | Tolazamide |

| | | | |
|---|---|---|---|
| INSULIN | INSULIN | 1567232 | Insulin, Aspart, Human |
| BROMOCRIPTINE | BROMOCRIPTINE | 19116171 | Bromocriptine |
| METFORMIN | METFORMIN | 19113780 | Metformin |
| METFORMIN | METFORMIN | 40164932 | Metformin |
| METFORMIN | METFORMIN | 40164893 | Metformin |
| INSULIN | INSULIN | 1567232 | Insulin, Aspart, Human |
| GLYBURIDE | GLYBURIDE | 36235750 | Glyburide |
| INSULIN | INSULIN | 36235841 | Regular Insulin, Human |
| INSULIN | INSULIN | 1596999 | Regular Insulin, Human |
| INSULIN | INSULIN | 1567232 | Insulin, Aspart, Human |
| GLYBURIDE | GLYBURIDE | 40168376 | Glyburide |
| LIRAGLUTIDE | LIRAGLUTIDE | 36220119 | liraglutide |
| METFORMIN | METFORMIN | 1503297 | Metformin |
| SITAGLIPTIN | SITAGLIPTIN | 42900896 | sitagliptin |
| GLIPIZIDE | GLIPIZIDE | 1560171 | Glipizide |
| GLYBURIDE | GLYBURIDE | 40165994 | Glyburide |
| INSULIN | INSULIN | 1596972 | insulin, isophane |
| METFORMIN | METFORMIN | 40164882 | Metformin |
| PIOGLITAZONE | PIOGLITAZONE | 1525218 | pioglitazone |
| INSULIN | INSULIN | 1567232 | Insulin, Aspart, Human |
| DULAGLUTIDE | DULAGLUTIDE | 45774442 | dulaglutide |
| INSULIN | INSULIN | 19135263 | insulin, isophane |
| SITAGLIPTIN | SITAGLIPTIN | 19125047 | sitagliptin |
| DAPAGLIFLOZIN | DAPAGLIFLOZIN | 45775864 | dapagliflozin |
| INSULIN | INSULIN | 1596977 | Regular Insulin, Human |
| PRAMLINTIDE | PRAMLINTIDE | 40165688 | Pramlintide |
| INSULIN | INSULIN | 1596918 | insulin, isophane |
| INSULIN | INSULIN | 36222190 | insulin detemir |
| EXENATIDE | EXENATIDE | 19083254 | exenatide |
| ALOGLIPTIN | ALOGLIPTIN | 43013919 | alogliptin |
| CHLORPROPAMIDE | CHLORPROPAMIDE | 1594973 | Chlorpropamide |

| | | | |
|---|---|---|---|
| INSULIN | INSULIN | 46221552 | Insulin Glargine |
| INSULIN | INSULIN | 1513881 | Insulin Lispro |
| METFORMIN | METFORMIN | 1503297 | Metformin |
| METFORMIN | METFORMIN | 40063356 | Metformin |
| GLIPIZIDE | GLIPIZIDE | 19077638 | Glipizide |
| GLIPIZIDE | GLIPIZIDE | 19047638 | Glipizide |
| GLIPIZIDE | GLIPIZIDE | 40044251 | Glipizide |
| INSULIN | INSULIN | 1586370 | Ultralente Insulin, Human |
| GLYBURIDE | GLYBURIDE | 19077682 | Glyburide |
| METFORMIN | METFORMIN | 40229050 | Metformin |
| DULAGLUTIDE | DULAGLUTIDE | 45774442 | dulaglutide |
| INSULIN | INSULIN | 46233728 | Regular Insulin, Human |
| EMPAGLIFLOZIN | EMPAGLIFLOZIN | 45774753 | empagliflozin |
| GLIMEPIRIDE | GLIMEPIRIDE | 19117200 | glimepiride |
| EXENATIDE | EXENATIDE | 19083253 | exenatide |
| INSULIN | INSULIN | 35605622 | Regular Insulin, Human |
| ACETOHEXAMIDE | ACETOHEXAMIDE | 36235641 | Acetohexamide |
| ROSIGLITAZONE | ROSIGLITAZONE | 19099245 | rosiglitazone |
| INSULIN | INSULIN | 35602728 | insulin degludec |
| INSULIN | INSULIN | 1513849 | Insulin, Zinc, Human |
| GLYBURIDE | GLYBURIDE | 1559761 | Glyburide |
| GLIPIZIDE | GLIPIZIDE | 36218767 | Glipizide |
| INSULIN | INSULIN | 1597001 | Regular Insulin, Human |
| INSULIN | INSULIN | 1513849 | Insulin, Zinc, Human |
| BROMOCRIPTINE | BROMOCRIPTINE | 730572 | Bromocriptine |
| ROSIGLITAZONE | ROSIGLITAZONE | 19079465 | rosiglitazone |
| INSULIN | INSULIN | 36235937 | insulin human, rDNA origin |
| INSULIN | INSULIN | 1516981 | insulin detemir |
| TOLBUTAMIDE | TOLBUTAMIDE | 36214386 | Tolbutamide |
| INSULIN | INSULIN | 19111795 | insulin, isophane |
| INSULIN | INSULIN | 35602731 | insulin degludec |
| INSULIN | INSULIN | 1596972 | insulin, isophane |
| EXENATIDE | EXENATIDE | 19083254 | exenatide |
| INSULIN | INSULIN | 19078607 | insulin, isophane |
| GLYBURIDE | GLYBURIDE | 1559784 | Glyburide |
| ROSIGLITAZONE | ROSIGLITAZONE | 19099245 | rosiglitazone |

| | | | |
|---|---|---|---|
| INSULIN | INSULIN | 46234045 | Insulin, Aspart, Human |
| METFORMIN | METFORMIN | 1503297 | Metformin |
| INSULIN | INSULIN | 46234049 | Insulin, Aspart, Human |
| EXENATIDE | EXENATIDE | 46234116 | exenatide |
| BROMOCRIPTINE | BROMOCRIPTINE | 36238867 | Bromocriptine |
| INSULIN | INSULIN | 19120315 | insulin, isophane |
| METFORMIN | METFORMIN | 36242511 | Metformin |
| ACARBOSE | ACARBOSE | 36242199 | Acarbose |
| INSULIN | INSULIN | 19078527 | Insulin, Zinc, Human |
| INSULIN | INSULIN | 1502910 | Insulin Glargine |
| SITAGLIPTIN | SITAGLIPTIN | 19125050 | sitagliptin |
| INSULIN | INSULIN | 36229086 | Insulin Glargine |
| GLYBURIDE | GLYBURIDE | 40046201 | Glyburide |
| INSULIN | INSULIN | 1586344 | insulin, isophane |
| INSULIN | INSULIN | 1550023 | Insulin Lispro |
| INSULIN | INSULIN | 36224228 | insulin detemir |
| INSULIN | INSULIN | 1516981 | insulin detemir |
| INSULIN | INSULIN | 19078559 | Insulin Glargine |
| METFORMIN | METFORMIN | 40164897 | Metformin |
| GLIMEPIRIDE | GLIMEPIRIDE | 1597775 | glimepiride |
| LIRAGLUTIDE | LIRAGLUTIDE | 40170913 | liraglutide |
| METFORMIN | METFORMIN | 1503328 | Metformin |
| INSULIN | INSULIN | 19135263 | insulin, isophane |
| INSULIN | INSULIN | 40166275 | Insulin Lispro |
| INSULIN | INSULIN | 1567232 | Insulin, Aspart, Human |
| METFORMIN | METFORMIN | 40117785 | Metformin |
| GLYBURIDE | GLYBURIDE | 40046201 | Glyburide |
| INSULIN | INSULIN | 19078555 | Regular Insulin, Human |
| INSULIN | INSULIN | 1567200 | Insulin, Aspart, Human |
| INSULIN | INSULIN | 1567198 | Insulin, Aspart, Human |
| METFORMIN | METFORMIN | 40164897 | Metformin |
| INSULIN | INSULIN | 1596977 | Regular Insulin, Human |
| LIRAGLUTIDE | LIRAGLUTIDE | 45892686 | liraglutide |
| GLIPIZIDE | GLIPIZIDE | 40044251 | Glipizide |
| PIOGLITAZONE | PIOGLITAZONE | 19099208 | pioglitazone |
| INSULIN | INSULIN | 19094122 | Regular Insulin, Human |

| | | | |
|---|---|---|---|
| METFORMIN | METFORMIN | 40063350 | Metformin |
| NATEGLINIDE | NATEGLINIDE | 1502827 | nateglinide |
| METFORMIN | METFORMIN | 40164932 | Metformin |
| GLIPIZIDE | GLIPIZIDE | 19117945 | Glipizide |
| GLIPIZIDE | GLIPIZIDE | 1560233 | Glipizide |
| GLYBURIDE | GLYBURIDE | 40046201 | Glyburide |
| METFORMIN | METFORMIN | 40164940 | Metformin |
| LINAGLIPTIN | LINAGLIPTIN | 40239219 | Linagliptin |
| METFORMIN | METFORMIN | 40164880 | Metformin |
| GLIMEPIRIDE | GLIMEPIRIDE | 1597776 | glimepiride |
| METFORMIN | METFORMIN | 40164948 | Metformin |
| GLYBURIDE | GLYBURIDE | 19077659 | Glyburide |
| NATEGLINIDE | NATEGLINIDE | 1502826 | nateglinide |
| INSULIN | INSULIN | 19078558 | Insulin, Aspart, Human |
| GLIPIZIDE | GLIPIZIDE | 19085004 | Glipizide |
| INSULIN | INSULIN | 46234049 | Insulin, Aspart, Human |
| METFORMIN | METFORMIN | 19088818 | Metformin |
| INSULIN | INSULIN | 1502910 | Insulin Glargine |
| METFORMIN | METFORMIN | 40164939 | Metformin |
| INSULIN | INSULIN | 19078552 | insulin, isophane |
| INSULIN | INSULIN | 1596977 | Regular Insulin, Human |
| TOLAZAMIDE | TOLAZAMIDE | 1502816 | Tolazamide |
| INSULIN | INSULIN | 36231450 | Insulin, Aspart, Human |
| METFORMIN | METFORMIN | 40163928 | Metformin |
| METFORMIN | METFORMIN | 40164890 | Metformin |
| GLIPIZIDE | GLIPIZIDE | 1560171 | Glipizide |
| GLIPIZIDE | GLIPIZIDE | 40167636 | Glipizide |
| GLYBURIDE | GLYBURIDE | 19077659 | Glyburide |
| INSULIN | INSULIN | 1550027 | Insulin Lispro |
| LIRAGLUTIDE | LIRAGLUTIDE | 40170913 | liraglutide |
| INSULIN | INSULIN | 19135263 | insulin, isophane |
| INSULIN | INSULIN | 19104092 | Regular Insulin, Human |
| INSULIN | INSULIN | 1502910 | Insulin Glargine |
| INSULIN | INSULIN | 19044971 | insulin detemir |
| GLIMEPIRIDE | GLIMEPIRIDE | 1597756 | glimepiride |
| METFORMIN | METFORMIN | 36223137 | Metformin |

| | | | |
|---|---|---|---|
| INSULIN | INSULIN | 1515107 | insulin, isophane |
| GLYBURIDE | GLYBURIDE | 19077659 | Glyburide |
| METFORMIN | METFORMIN | 40163926 | Metformin |
| CANAGLIFLOZIN | CANAGLIFLOZIN | 45774717 | canagliflozin |
| INSULIN | INSULIN | 19078553 | insulin, isophane |
| GLICLAZIDE | GLICLAZIDE | 40044221 | Gliclazide |
| REPAGLINIDE | REPAGLINIDE | 1516766 | repaglinide |
| INSULIN | INSULIN | 40176981 | Insulin, Aspart Protamine, Human |
| METFORMIN | METFORMIN | 19086484 | Metformin |
| REPAGLINIDE | REPAGLINIDE | 40098707 | repaglinide |
| INSULIN | INSULIN | 46233975 | Insulin Lispro |
| SITAGLIPTIN | SITAGLIPTIN | 19125050 | sitagliptin |
| METFORMIN | METFORMIN | 40164947 | Metformin |
| INSULIN | INSULIN | 1567232 | Insulin, Aspart, Human |
| INSULIN | INSULIN | 46234049 | Insulin, Aspart, Human |
| TOLBUTAMIDE | TOLBUTAMIDE | 40089115 | Tolbutamide |
| INSULIN | INSULIN | 46233975 | Insulin Lispro |
| GLIPIZIDE | GLIPIZIDE | 1560171 | Glipizide |
| METFORMIN | METFORMIN | 19121940 | Metformin |
| GLIPIZIDE | GLIPIZIDE | 19134624 | Glipizide |
| EXENATIDE | EXENATIDE | 1583727 | exenatide |
| DAPAGLIFLOZIN | DAPAGLIFLOZIN | 44785473 | dapagliflozin |
| ACARBOSE | ACARBOSE | 19021787 | Acarbose |
| INSULIN | INSULIN | 1502905 | Insulin Glargine |
| GLYBURIDE | GLYBURIDE | 1559782 | Glyburide |
| SITAGLIPTIN | SITAGLIPTIN | 19125051 | sitagliptin |
| ACARBOSE | ACARBOSE | 40003413 | Acarbose |
| INSULIN | INSULIN | 19061587 | insulin, isophane |
| INSULIN | INSULIN | 1596972 | insulin, isophane |
| PIOGLITAZONE | PIOGLITAZONE | 19099209 | pioglitazone |
| METFORMIN | METFORMIN | 1503297 | Metformin |
| INSULIN | INSULIN | 46233970 | Insulin Lispro |

| | | | |
|---|---|---|---|
| METFORMIN | METFORMIN | 40173239 | Metformin |
| GLIMEPIRIDE | GLIMEPIRIDE | 40059666 | glimepiride |
| INSULIN | INSULIN | 19120315 | insulin, isophane |
| INSULIN | INSULIN | 1567198 | Insulin, Aspart, Human |
| INSULIN | INSULIN | 46233976 | Insulin Lispro |
| GLIPIZIDE | GLIPIZIDE | 1560171 | Glipizide |
| SITAGLIPTIN | SITAGLIPTIN | 19125048 | sitagliptin |
| INSULIN | INSULIN | 19123376 | Insulin Lispro |
| INSULIN | INSULIN | 1586370 | Ultralente Insulin, Human |
| METFORMIN | METFORMIN | 40171451 | Metformin |
| GLIPIZIDE | GLIPIZIDE | 40167634 | Glipizide |
| GLIPIZIDE | GLIPIZIDE | 40167635 | Glipizide |
| METFORMIN | METFORMIN | 40164925 | Metformin |
| INSULIN | INSULIN | 35602728 | insulin degludec |
| INSULIN | INSULIN | 19078553 | insulin, isophane |
| INSULIN | INSULIN | 19135320 | Regular Insulin, Human |
| GLIPIZIDE | GLIPIZIDE | 19081295 | Glipizide |
| GLYBURIDE | GLYBURIDE | 19079986 | Glyburide |
| INSULIN | INSULIN | 19044971 | insulin detemir |
| GLIPIZIDE | GLIPIZIDE | 19079990 | Glipizide |
| GLYBURIDE | GLYBURIDE | 40168375 | Glyburide |
| METFORMIN | METFORMIN | 40164882 | Metformin |
| INSULIN | INSULIN | 19078530 | insulin, isophane |
| METFORMIN | METFORMIN | 36242511 | Metformin |
| EXENATIDE | EXENATIDE | 19083253 | exenatide |
| INSULIN | INSULIN | 35602728 | insulin degludec |
| GLIPIZIDE | GLIPIZIDE | 1560171 | Glipizide |
| INSULIN | INSULIN | 35602724 | insulin degludec |
| METFORMIN | METFORMIN | 40164932 | Metformin |
| ALOGLIPTIN | ALOGLIPTIN | 43013897 | alogliptin |
| INSULIN | INSULIN | 19110352 | insulin, isophane |
| INSULIN | INSULIN | 36224228 | insulin detemir |
| METFORMIN | METFORMIN | 36235746 | Metformin |
| GLIPIZIDE | GLIPIZIDE | 19047638 | Glipizide |

| | | | |
|---|---|---|---|
| INSULIN | INSULIN | 40166274 | Insulin Lispro |
| INSULIN | INSULIN | 1596941 | insulin, isophane |
| EXENATIDE | EXENATIDE | 1583722 | exenatide |
| GLYBURIDE | GLYBURIDE | 40168375 | Glyburide |
| LIRAGLUTIDE | LIRAGLUTIDE | 36248956 | liraglutide |
| GLIPIZIDE | GLIPIZIDE | 19077638 | Glipizide |
| DAPAGLIFLOZIN | DAPAGLIFLOZIN | 44815996 | dapagliflozin |
| GLYBURIDE | GLYBURIDE | 19030575 | Glyburide |
| METFORMIN | METFORMIN | 36223145 | Metformin |
| ALBIGLUTIDE | ALBIGLUTIDE | 44816086 | albiglutide |
| METFORMIN | METFORMIN | 40164931 | Metformin |
| GLYBURIDE | GLYBURIDE | 36213839 | Glyburide |
| INSULIN | INSULIN | 19062999 | insulin, isophane |
| GLIMEPIRIDE | GLIMEPIRIDE | 1597774 | glimepiride |
| INSULIN | INSULIN | 19078556 | Regular Insulin, Human |
| METFORMIN | METFORMIN | 40164918 | Metformin |
| DULAGLUTIDE | DULAGLUTIDE | 45774442 | dulaglutide |
| GLIMEPIRIDE | GLIMEPIRIDE | 1597774 | glimepiride |
| INSULIN | INSULIN | 1596974 | insulin, isophane |
| GLIPIZIDE | GLIPIZIDE | 19081295 | Glipizide |
| METFORMIN | METFORMIN | 1503327 | Metformin |
| INSULIN | INSULIN | 19110220 | Regular Insulin, Human |
| INSULIN | INSULIN | 36233552 | insulin, isophane |
| METFORMIN | METFORMIN | 40164898 | Metformin |
| INSULIN | INSULIN | 46221558 | Insulin Glargine |
| ROSIGLITAZONE | ROSIGLITAZONE | 1547504 | rosiglitazone |
| EMPAGLIFLOZIN | EMPAGLIFLOZIN | 45892179 | empagliflozin |
| INSULIN | INSULIN | 46233945 | Insulin Lispro |
| METFORMIN | METFORMIN | 36235787 | Metformin |
| CANAGLIFLOZIN | CANAGLIFLOZIN | 45774714 | canagliflozin |
| EXENATIDE | EXENATIDE | 1583728 | exenatide |
| INSULIN | INSULIN | 1596953 | insulin, isophane |
| INSULIN | INSULIN | 46233975 | Insulin Lispro |
| INSULIN | INSULIN | 46234045 | Insulin, Aspart, Human |
| GLIMEPIRIDE | GLIMEPIRIDE | 1597759 | glimepiride |
| INSULIN | INSULIN | 1550023 | Insulin Lispro |

| | | | |
|---|---|---|---|
| GLICLAZIDE | GLICLAZIDE | 19059797 | Gliclazide |
| GLYBURIDE | GLYBURIDE | 1559684 | Glyburide |
| METFORMIN | METFORMIN | 40231393 | Metformin |
| INSULIN | INSULIN | 40051347 | Insulin Glargine |
| INSULIN | INSULIN | 1596916 | insulin, isophane |
| GLIMEPIRIDE | GLIMEPIRIDE | 19018043 | glimepiride |
| SAXAGLIPTIN | SAXAGLIPTIN | 40166044 | saxagliptin |
| BROMOCRIPTINE | BROMOCRIPTINE | 730580 | Bromocriptine |
| ACARBOSE | ACARBOSE | 40003412 | Acarbose |
| PIOGLITAZONE | PIOGLITAZONE | 1525215 | pioglitazone |
| INSULIN | INSULIN | 19135265 | Regular Insulin, Human |
| INSULIN | INSULIN | 1586370 | Ultralente Insulin, Human |
| METFORMIN | METFORMIN | 40164895 | Metformin |
| CANAGLIFLOZIN | CANAGLIFLOZIN | 36246877 | canagliflozin |
| LIRAGLUTIDE | LIRAGLUTIDE | 45892686 | liraglutide |
| MIGLITOL | MIGLITOL | 40056706 | miglitol |
| INSULIN | INSULIN | 40051347 | Insulin Glargine |
| DAPAGLIFLOZIN | DAPAGLIFLOZIN | 44785833 | dapagliflozin |
| GLIPIZIDE | GLIPIZIDE | 40167638 | Glipizide |
| GLYBURIDE | GLYBURIDE | 19116147 | Glyburide |
| INSULIN | INSULIN | 19080091 | Ultralente Insulin, Human |
| GLIPIZIDE | GLIPIZIDE | 19077638 | Glipizide |
| GLIMEPIRIDE | GLIMEPIRIDE | 1597774 | glimepiride |
| SITAGLIPTIN | SITAGLIPTIN | 19125042 | sitagliptin |
| GLIPIZIDE | GLIPIZIDE | 40044251 | Glipizide |
| GLIPIZIDE | GLIPIZIDE | 1560171 | Glipizide |
| METFORMIN | METFORMIN | 40063350 | Metformin |
| LINAGLIPTIN | LINAGLIPTIN | 40239220 | Linagliptin |
| METFORMIN | METFORMIN | 40164924 | Metformin |
| PRAMLINTIDE | PRAMLINTIDE | 36243018 | Pramlintide |
| GLYBURIDE | GLYBURIDE | 36230036 | Glyburide |
| METFORMIN | METFORMIN | 40164938 | Metformin |
| GLIMEPIRIDE | GLIMEPIRIDE | 1597772 | glimepiride |
| GLIMEPIRIDE | GLIMEPIRIDE | 1597775 | glimepiride |

| | | | |
|---|---|---|---|
| INSULIN | INSULIN | 19135263 | insulin, isophane |
| INSULIN | INSULIN | 46233730 | Regular Insulin, Human |
| METFORMIN | METFORMIN | 40164880 | Metformin |
| CANAGLIFLOZIN | CANAGLIFLOZIN | 36246430 | canagliflozin |
| CHLORPROPAMIDE | CHLORPROPAMIDE | 19006907 | Chlorpropamide |
| SITAGLIPTIN | SITAGLIPTIN | 40243628 | sitagliptin |
| INSULIN | INSULIN | 1596977 | Regular Insulin, Human |
| INSULIN | INSULIN | 1567198 | Insulin, Aspart, Human |
| GLIMEPIRIDE | GLIMEPIRIDE | 1597773 | glimepiride |
| INSULIN | INSULIN | 19120790 | Insulin Glargine |
| INSULIN | INSULIN | 1516981 | insulin detemir |
| METFORMIN | METFORMIN | 40164932 | Metformin |
| REPAGLINIDE | REPAGLINIDE | 40098705 | repaglinide |
| PIOGLITAZONE | PIOGLITAZONE | 19099208 | pioglitazone |
| GLIMEPIRIDE | GLIMEPIRIDE | 36229116 | glimepiride |
| METFORMIN | METFORMIN | 40164946 | Metformin |
| PIOGLITAZONE | PIOGLITAZONE | 19099207 | pioglitazone |
| GLIPIZIDE | GLIPIZIDE | 1560171 | Glipizide |
| CHLORPROPAMIDE | CHLORPROPAMIDE | 1594975 | Chlorpropamide |
| METFORMIN | METFORMIN | 40164938 | Metformin |
| GLIMEPIRIDE | GLIMEPIRIDE | 19100593 | glimepiride |
| INSULIN | INSULIN | 1596974 | insulin, isophane |
| ALOGLIPTIN | ALOGLIPTIN | 43013929 | alogliptin |
| INSULIN | INSULIN | 19120315 | insulin, isophane |
| INSULIN | INSULIN | 40052088 | Insulin Lispro |
| GLYBURIDE | GLYBURIDE | 40166004 | Glyburide |
| INSULIN | INSULIN | 1567198 | Insulin, Aspart, Human |
| INSULIN | INSULIN | 19078557 | Regular Insulin, Human |
| CANAGLIFLOZIN | CANAGLIFLOZIN | 43526473 | canagliflozin |
| BROMOCRIPTINE | BROMOCRIPTINE | 40016897 | Bromocriptine |
| SITAGLIPTIN | SITAGLIPTIN | 19125042 | sitagliptin |
| EXENATIDE | EXENATIDE | 1583730 | exenatide |
| INSULIN | INSULIN | 1550023 | Insulin Lispro |
| EXENATIDE | EXENATIDE | 46234118 | exenatide |
| INSULIN | INSULIN | 19120315 | insulin, isophane |
| GLIPIZIDE | GLIPIZIDE | 40044253 | Glipizide |

| | | | |
|---|---|---|---|
| INSULIN | INSULIN | 1567198 | Insulin, Aspart, Human |
| SAXAGLIPTIN | SAXAGLIPTIN | 40166039 | saxagliptin |
| GLIMEPIRIDE | GLIMEPIRIDE | 1597774 | glimepiride |
| EXENATIDE | EXENATIDE | 19083253 | exenatide |
| INSULIN | INSULIN | 46233976 | Insulin Lispro |
| GLYBURIDE | GLYBURIDE | 40166001 | Glyburide |
| METFORMIN | METFORMIN | 40164942 | Metformin |
| INSULIN | INSULIN | 40166274 | Insulin Lispro |
| DULAGLUTIDE | DULAGLUTIDE | 45774438 | dulaglutide |
| INSULIN | INSULIN | 36233552 | insulin, isophane |
| INSULIN | INSULIN | 19135265 | Regular Insulin, Human |
| METFORMIN | METFORMIN | 40164929 | Metformin |
| ROSIGLITAZONE | ROSIGLITAZONE | 19079465 | rosiglitazone |
| EXENATIDE | EXENATIDE | 36238252 | exenatide |
| EXENATIDE | EXENATIDE | 19083253 | exenatide |
| METFORMIN | METFORMIN | 40164918 | Metformin |
| METFORMIN | METFORMIN | 40164912 | Metformin |
| INSULIN | INSULIN | 35605621 | Regular Insulin, Human |
| METFORMIN | METFORMIN | 40063356 | Metformin |
| INSULIN | INSULIN | 35602728 | insulin degludec |
| INSULIN | INSULIN | 19078606 | insulin, isophane |
| INSULIN | INSULIN | 19123376 | Insulin Lispro |
| INSULIN | INSULIN | 1550027 | Insulin Lispro |
| GLYBURIDE | GLYBURIDE | 1559684 | Glyburide |
| GLYBURIDE | GLYBURIDE | 19030573 | Glyburide |
| CANAGLIFLOZIN | CANAGLIFLOZIN | 43526467 | canagliflozin |
| INSULIN | INSULIN | 36236033 | Insulin Lispro |
| INSULIN | INSULIN | 46234241 | Regular Insulin, Human |
| EMPAGLIFLOZIN | EMPAGLIFLOZIN | 45892177 | empagliflozin |
| INSULIN | INSULIN | 19120790 | Insulin Glargine |
| INSULIN | INSULIN | 36229086 | Insulin Glargine |
| METFORMIN | METFORMIN | 40164923 | Metformin |
| TOLAZAMIDE | TOLAZAMIDE | 1502812 | Tolazamide |
| METFORMIN | METFORMIN | 40163924 | Metformin |

| | | | |
|---|---|---|---|
| LINAGLIPTIN | LINAGLIPTIN | 40239220 | Linagliptin |
| INSULIN | INSULIN | 19078551 | insulin, isophane |
| GLIMEPIRIDE | GLIMEPIRIDE | 1597756 | glimepiride |
| INSULIN | INSULIN | 19094122 | Regular Insulin, Human |
| INSULIN | INSULIN | 19120790 | Insulin Glargine |
| INSULIN | INSULIN | 19078555 | Regular Insulin, Human |
| ALOGLIPTIN | ALOGLIPTIN | 43013911 | alogliptin |
| ROSIGLITAZONE | ROSIGLITAZONE | 1547504 | rosiglitazone |
| INSULIN | INSULIN | 46234241 | Regular Insulin, Human |
| METFORMIN | METFORMIN | 1503327 | Metformin |
| METFORMIN | METFORMIN | 1503297 | Metformin |
| INSULIN | INSULIN | 19062999 | insulin, isophane |
| METFORMIN | METFORMIN | 1503327 | Metformin |
| INSULIN | INSULIN | 1596941 | insulin, isophane |
| INSULIN | INSULIN | 1596977 | Regular Insulin, Human |
| ALOGLIPTIN | ALOGLIPTIN | 43013887 | alogliptin |
| EXENATIDE | EXENATIDE | 1583730 | exenatide |
| METFORMIN | METFORMIN | 40063354 | Metformin |
| INSULIN | INSULIN | 19094122 | Regular Insulin, Human |
| METFORMIN | METFORMIN | 36223145 | Metformin |
| CANAGLIFLOZIN | CANAGLIFLOZIN | 43526473 | canagliflozin |
| INSULIN | INSULIN | 1550023 | Insulin Lispro |
| NATEGLINIDE | NATEGLINIDE | 19102037 | nateglinide |
| METFORMIN | METFORMIN | 36235752 | Metformin |
| GLIPIZIDE | GLIPIZIDE | 1560233 | Glipizide |
| INSULIN | INSULIN | 1586370 | Ultralente Insulin, Human |
| INSULIN | INSULIN | 40184293 | Insulin, Aspart Protamine, Human |
| NATEGLINIDE | NATEGLINIDE | 19107111 | nateglinide |
| INSULIN | INSULIN | 35602726 | insulin degludec |
| INSULIN | INSULIN | 1567198 | Insulin, Aspart, Human |
| EXENATIDE | EXENATIDE | 45774533 | exenatide |
| GLIMEPIRIDE | GLIMEPIRIDE | 19117198 | glimepiride |
| GLIMEPIRIDE | GLIMEPIRIDE | 40059666 | glimepiride |
| BROMOCRIPTINE | BROMOCRIPTINE | 36214531 | Bromocriptine |
| INSULIN | INSULIN | 1596920 | insulin, isophane |
| GLIMEPIRIDE | GLIMEPIRIDE | 1597772 | glimepiride |

| | | | |
|---|---|---|---|
| NATEGLINIDE | NATEGLINIDE | 1502826 | nateglinide |
| INSULIN | INSULIN | 46233948 | Insulin Lispro |
| INSULIN | INSULIN | 19129437 | insulin human, rDNA origin |
| INSULIN | INSULIN | 1502910 | Insulin Glargine |
| INSULIN | INSULIN | 40166274 | Insulin Lispro |
| BROMOCRIPTINE | BROMOCRIPTINE | 19007102 | Bromocriptine |
| METFORMIN | METFORMIN | 1503297 | Metformin |
| GLIPIZIDE | GLIPIZIDE | 40165976 | Glipizide |
| INSULIN | INSULIN | 36235840 | insulin, isophane |
| INSULIN | INSULIN | 46234240 | insulin, isophane |
| NATEGLINIDE | NATEGLINIDE | 1502827 | nateglinide |
| ROSIGLITAZONE | ROSIGLITAZONE | 19120648 | rosiglitazone |
| PRAMLINTIDE | PRAMLINTIDE | 1517998 | Pramlintide |
| LINAGLIPTIN | LINAGLIPTIN | 40239220 | Linagliptin |
| METFORMIN | METFORMIN | 40164897 | Metformin |
| GLYBURIDE | GLYBURIDE | 1559784 | Glyburide |
| BROMOCRIPTINE | BROMOCRIPTINE | 730548 | Bromocriptine |
| SITAGLIPTIN | SITAGLIPTIN | 42901620 | sitagliptin |
| METFORMIN | METFORMIN | 40135481 | Metformin |
| LINAGLIPTIN | LINAGLIPTIN | 42708091 | Linagliptin |
| INSULIN | INSULIN | 1567232 | Insulin, Aspart, Human |
| INSULIN | INSULIN | 40184293 | Insulin, Aspart Protamine, Human |
| ACARBOSE | ACARBOSE | 40003413 | Acarbose |
| PRAMLINTIDE | PRAMLINTIDE | 1517998 | Pramlintide |
| INSULIN | INSULIN | 1567198 | Insulin, Aspart, Human |
| METFORMIN | METFORMIN | 40173239 | Metformin |
| INSULIN | INSULIN | 19016117 | Insulin Lispro |
| METFORMIN | METFORMIN | 40163926 | Metformin |
| SITAGLIPTIN | SITAGLIPTIN | 19125041 | sitagliptin |
| GLYBURIDE | GLYBURIDE | 1559684 | Glyburide |
| INSULIN | INSULIN | 19078558 | Insulin, Aspart, Human |
| METFORMIN | METFORMIN | 40164935 | Metformin |
| INSULIN | INSULIN | 36222190 | insulin detemir |
| GLIMEPIRIDE | GLIMEPIRIDE | 1597776 | glimepiride |

| | | | |
|---|---|---|---|
| INSULIN | INSULIN | 19120790 | Insulin Glargine |
| GLIPIZIDE | GLIPIZIDE | 40139022 | Glipizide |
| GLIPIZIDE | GLIPIZIDE | 36218768 | Glipizide |
| GLIMEPIRIDE | GLIMEPIRIDE | 1597759 | glimepiride |
| INSULIN | INSULIN | 1586370 | Ultralente Insulin, Human |
| ACARBOSE | ACARBOSE | 36242199 | Acarbose |
| INSULIN | INSULIN | 19110357 | Regular Insulin, Human |
| INSULIN | INSULIN | 19078633 | Insulin, Regular, Pork |
| INSULIN | INSULIN | 1502908 | Insulin Glargine |
| INSULIN | INSULIN | 46234240 | insulin, isophane |
| LINAGLIPTIN | LINAGLIPTIN | 40239216 | Linagliptin |
| INSULIN | INSULIN | 1513849 | Insulin, Zinc, Human |
| GLIPIZIDE | GLIPIZIDE | 40044251 | Glipizide |
| INSULIN | INSULIN | 19078554 | Regular Insulin, Human |
| METFORMIN | METFORMIN | 40231396 | Metformin |
| METFORMIN | METFORMIN | 40164893 | Metformin |
| GLYBURIDE | GLYBURIDE | 1559784 | Glyburide |
| METFORMIN | METFORMIN | 40164914 | Metformin |
| INSULIN | INSULIN | 1596959 | insulin, isophane |
| INSULIN | INSULIN | 19078633 | Insulin, Regular, Pork |
| INSULIN | INSULIN | 46233976 | Insulin Lispro |
| SAXAGLIPTIN | SAXAGLIPTIN | 36229291 | saxagliptin |
| INSULIN | INSULIN | 19100127 | insulin, isophane |
| INSULIN | INSULIN | 1596998 | Regular Insulin, Human |
| GLYBURIDE | GLYBURIDE | 1559684 | Glyburide |
| INSULIN | INSULIN | 19078530 | insulin, isophane |
| INSULIN | INSULIN | 1596960 | insulin, isophane |
| GLIMEPIRIDE | GLIMEPIRIDE | 1597760 | glimepiride |
| GLIMEPIRIDE | GLIMEPIRIDE | 1597801 | glimepiride |
| INSULIN | INSULIN | 19058398 | Insulin Lispro |
| INSULIN | INSULIN | 19078555 | Regular Insulin, Human |
| INSULIN | INSULIN | 19078530 | insulin, isophane |
| INSULIN | INSULIN | 36222190 | insulin detemir |
| INSULIN | INSULIN | 1502905 | Insulin Glargine |

| | | | |
|---|---|---|---|
| GLIMEPIRIDE | GLIMEPIRIDE | 1597756 | glimepiride |
| INSULIN | INSULIN | 1513881 | Insulin Lispro |
| METFORMIN | METFORMIN | 40164900 | Metformin |
| INSULIN | INSULIN | 36227902 | Insulin Lispro |
| INSULIN | INSULIN | 1567200 | Insulin, Aspart, Human |
| EXENATIDE | EXENATIDE | 42705842 | exenatide |
| INSULIN | INSULIN | 19116610 | insulin, isophane |
| GLYBURIDE | GLYBURIDE | 1559784 | Glyburide |
| INSULIN | INSULIN | 1550023 | Insulin Lispro |
| METFORMIN | METFORMIN | 40164893 | Metformin |
| ACARBOSE | ACARBOSE | 1529360 | Acarbose |
| GLYBURIDE | GLYBURIDE | 40166002 | Glyburide |
| INSULIN | INSULIN | 46221555 | Insulin Glargine |
| ACARBOSE | ACARBOSE | 36242198 | Acarbose |
| GLIMEPIRIDE | GLIMEPIRIDE | 19117199 | glimepiride |
| INSULIN | INSULIN | 1567198 | Insulin, Aspart, Human |
| INSULIN | INSULIN | 1567232 | Insulin, Aspart, Human |
| INSULIN | INSULIN | 19135263 | insulin, isophane |
| ACARBOSE | ACARBOSE | 1529331 | Acarbose |
| INSULIN | INSULIN | 1502905 | Insulin Glargine |
| INSULIN | INSULIN | 19094123 | Regular Insulin, Human |
| GLYBURIDE | GLYBURIDE | 1559782 | Glyburide |
| INSULIN | INSULIN | 19078610 | insulin, regular, beef-pork |
| INSULIN | INSULIN | 36233552 | insulin, isophane |
| GLIPIZIDE | GLIPIZIDE | 19085004 | Glipizide |
| GLYBURIDE | GLYBURIDE | 1559760 | Glyburide |
| INSULIN | INSULIN | 19062999 | insulin, isophane |
| INSULIN | INSULIN | 1502905 | Insulin Glargine |
| INSULIN | INSULIN | 1502908 | Insulin Glargine |
| INSULIN | INSULIN | 1550028 | Insulin Lispro |
| INSULIN | INSULIN | 40052047 | insulin, isophane |
| INSULIN | INSULIN | 46233975 | Insulin Lispro |
| INSULIN | INSULIN | 1531601 | Insulin, Aspart Protamine, Human |
| INSULIN | INSULIN | 1516981 | insulin detemir |
| GLYBURIDE | GLYBURIDE | 19030579 | Glyburide |

| | | | |
|---|---|---|---|
| PIOGLITAZONE | PIOGLITAZONE | 1525215 | pioglitazone |
| EXENATIDE | EXENATIDE | 42708079 | exenatide |
| INSULIN | INSULIN | 46234049 | Insulin, Aspart, Human |
| INSULIN | INSULIN | 19110357 | Regular Insulin, Human |
| INSULIN | INSULIN | 19120315 | insulin, isophane |
| GLICLAZIDE | GLICLAZIDE | 19059797 | Gliclazide |
| INSULIN | INSULIN | 1596921 | insulin, isophane |
| METFORMIN | METFORMIN | 36235788 | Metformin |
| GLYBURIDE | GLYBURIDE | 1559784 | Glyburide |
| INSULIN | INSULIN | 1596972 | insulin, isophane |
| METFORMIN | METFORMIN | 40164943 | Metformin |
| INSULIN | INSULIN | 40130712 | insulin detemir |
| GLIPIZIDE | GLIPIZIDE | 1560171 | Glipizide |
| BROMOCRIPTINE | BROMOCRIPTINE | 730548 | Bromocriptine |
| INSULIN | INSULIN | 19078558 | Insulin, Aspart, Human |
| INSULIN | INSULIN | 40184293 | Insulin, Aspart Protamine, Human |
| INSULIN | INSULIN | 1516979 | insulin detemir |
| INSULIN | INSULIN | 1502905 | Insulin Glargine |
| INSULIN | INSULIN | 40052045 | insulin, isophane |
| INSULIN | INSULIN | 1567198 | Insulin, Aspart, Human |
| PIOGLITAZONE | PIOGLITAZONE | 19099207 | pioglitazone |
| ROSIGLITAZONE | ROSIGLITAZONE | 36227624 | rosiglitazone |
| INSULIN | INSULIN | 1550028 | Insulin Lispro |
| EXENATIDE | EXENATIDE | 1583730 | exenatide |
| SITAGLIPTIN | SITAGLIPTIN | 19125040 | sitagliptin |
| INSULIN | INSULIN | 46233950 | Insulin Lispro |
| METFORMIN | METFORMIN | 40135481 | Metformin |
| INSULIN | INSULIN | 19095214 | insulin, isophane |
| SITAGLIPTIN | SITAGLIPTIN | 40243628 | sitagliptin |
| INSULIN | INSULIN | 19078553 | insulin, isophane |
| INSULIN | INSULIN | 1502905 | Insulin Glargine |
| METFORMIN | METFORMIN | 40146999 | Metformin |
| PIOGLITAZONE | PIOGLITAZONE | 1525218 | pioglitazone |
| METFORMIN | METFORMIN | 36235746 | Metformin |

| | | | |
|---|---|---|---|
| PRAMLINTIDE | PRAMLINTIDE | 40165686 | Pramlintide |
| PIOGLITAZONE | PIOGLITAZONE | 1525216 | pioglitazone |
| METFORMIN | METFORMIN | 40164900 | Metformin |
| INSULIN | INSULIN | 19116454 | insulin, isophane |
| METFORMIN | METFORMIN | 40164932 | Metformin |
| METFORMIN | METFORMIN | 40163926 | Metformin |
| GLYBURIDE | GLYBURIDE | 1559785 | Glyburide |
| EMPAGLIFLOZIN | EMPAGLIFLOZIN | 46287678 | empagliflozin |
| METFORMIN | METFORMIN | 40164897 | Metformin |
| GLIPIZIDE | GLIPIZIDE | 19081295 | Glipizide |
| INSULIN | INSULIN | 1513849 | Insulin, Zinc, Human |
| GLIPIZIDE | GLIPIZIDE | 19030443 | Glipizide |
| SITAGLIPTIN | SITAGLIPTIN | 42900896 | sitagliptin |
| DULAGLUTIDE | DULAGLUTIDE | 45774442 | dulaglutide |
| INSULIN | INSULIN | 19095214 | insulin, isophane |
| DAPAGLIFLOZIN | DAPAGLIFLOZIN | 44785834 | dapagliflozin |
| METFORMIN | METFORMIN | 42705990 | Metformin |
| INSULIN | INSULIN | 36227902 | Insulin Lispro |
| INSULIN | INSULIN | 1597001 | Regular Insulin, Human |
| BROMOCRIPTINE | BROMOCRIPTINE | 730548 | Bromocriptine |
| INSULIN | INSULIN | 19078553 | insulin, isophane |
| NATEGLINIDE | NATEGLINIDE | 40058198 | nateglinide |
| INSULIN | INSULIN | 1597000 | Regular Insulin, Human |
| INSULIN | INSULIN | 46234240 | insulin, isophane |
| INSULIN | INSULIN | 19061670 | Regular Insulin, Human |
| BROMOCRIPTINE | BROMOCRIPTINE | 19083958 | Bromocriptine |
| INSULIN | INSULIN | 19010204 | insulin, isophane |
| INSULIN | INSULIN | 19062999 | insulin, isophane |
| GLIPIZIDE | GLIPIZIDE | 40139022 | Glipizide |
| METFORMIN | METFORMIN | 40173239 | Metformin |
| CHLORPROPAMIDE | CHLORPROPAMIDE | 19107624 | Chlorpropamide |
| INSULIN | INSULIN | 19058398 | Insulin Lispro |
| INSULIN | INSULIN | 46233970 | Insulin Lispro |

| | | | |
|---|---|---|---|
| METFORMIN | METFORMIN | 40164923 | Metformin |
| INSULIN | INSULIN | 1596959 | insulin, isophane |
| INSULIN | INSULIN | 19100127 | insulin, isophane |
| ALOGLIPTIN | ALOGLIPTIN | 43013903 | alogliptin |
| INSULIN | INSULIN | 40184293 | Insulin, Aspart Protamine, Human |
| DAPAGLIFLOZIN | DAPAGLIFLOZIN | 36247907 | dapagliflozin |
| INSULIN | INSULIN | 36227902 | Insulin Lispro |
| INSULIN | INSULIN | 19120315 | insulin, isophane |
| INSULIN | INSULIN | 19078551 | insulin, isophane |
| EXENATIDE | EXENATIDE | 36234709 | exenatide |
| EXENATIDE | EXENATIDE | 46234122 | exenatide |
| METFORMIN | METFORMIN | 1503327 | Metformin |
| METFORMIN | METFORMIN | 40164932 | Metformin |
| INSULIN | INSULIN | 19120315 | insulin, isophane |
| METFORMIN | METFORMIN | 40117785 | Metformin |
| METFORMIN | METFORMIN | 40164898 | Metformin |
| MIGLITOL | MIGLITOL | 36233499 | miglitol |
| METFORMIN | METFORMIN | 1503327 | Metformin |
| INSULIN | INSULIN | 1596977 | Regular Insulin, Human |
| INSULIN | INSULIN | 1550023 | Insulin Lispro |
| GLIPIZIDE | GLIPIZIDE | 19077636 | Glipizide |
| INSULIN | INSULIN | 1597012 | Regular Insulin, Human |
| INSULIN | INSULIN | 1567198 | Insulin, Aspart, Human |
| INSULIN | INSULIN | 19095256 | insulin, isophane |
| INSULIN | INSULIN | 1586344 | insulin, isophane |
| METFORMIN | METFORMIN | 40163928 | Metformin |
| INSULIN | INSULIN | 46233975 | Insulin Lispro |
| METFORMIN | METFORMIN | 19086484 | Metformin |
| METFORMIN | METFORMIN | 40164942 | Metformin |
| BROMOCRIPTINE | BROMOCRIPTINE | 19006288 | Bromocriptine |
| PIOGLITAZONE | PIOGLITAZONE | 1525221 | pioglitazone |
| ROSIGLITAZONE | ROSIGLITAZONE | 19099246 | rosiglitazone |
| METFORMIN | METFORMIN | 40164928 | Metformin |

| | | | |
|---|---|---|---|
| TOLAZAMIDE | TOLAZAMIDE | 19036965 | Tolazamide |
| INSULIN | INSULIN | 19095214 | insulin, isophane |
| BROMOCRIPTINE | BROMOCRIPTINE | 19007101 | Bromocriptine |
| GLYBURIDE | GLYBURIDE | 19065480 | Glyburide |
| GLYBURIDE | GLYBURIDE | 19077682 | Glyburide |
| INSULIN | INSULIN | 35602724 | insulin degludec |
| DULAGLUTIDE | DULAGLUTIDE | 36247186 | dulaglutide |
| INSULIN | INSULIN | 1596932 | insulin, isophane |
| BROMOCRIPTINE | BROMOCRIPTINE | 19083958 | Bromocriptine |
| METFORMIN | METFORMIN | 40164880 | Metformin |
| EXENATIDE | EXENATIDE | 19083253 | exenatide |
| GLYBURIDE | GLYBURIDE | 40166003 | Glyburide |
| EXENATIDE | EXENATIDE | 1583730 | exenatide |
| METFORMIN | METFORMIN | 42708171 | Metformin |
| METFORMIN | METFORMIN | 40164897 | Metformin |
| ALBIGLUTIDE | ALBIGLUTIDE | 44816093 | albiglutide |
| GLIMEPIRIDE | GLIMEPIRIDE | 19023585 | glimepiride |
| INSULIN | INSULIN | 19116453 | insulin, isophane |
| ALBIGLUTIDE | ALBIGLUTIDE | 44816094 | albiglutide |
| INSULIN | INSULIN | 40052088 | Insulin Lispro |
| INSULIN | INSULIN | 1567198 | Insulin, Aspart, Human |
| INSULIN | INSULIN | 19116453 | insulin, isophane |
| GLIMEPIRIDE | GLIMEPIRIDE | 1597774 | glimepiride |
| INSULIN | INSULIN | 1567198 | Insulin, Aspart, Human |
| LINAGLIPTIN | LINAGLIPTIN | 40239220 | Linagliptin |
| GLIPIZIDE | GLIPIZIDE | 1560171 | Glipizide |
| METFORMIN | METFORMIN | 40231396 | Metformin |
| METFORMIN | METFORMIN | 40171449 | Metformin |
| EXENATIDE | EXENATIDE | 1583722 | exenatide |
| METFORMIN | METFORMIN | 19086484 | Metformin |
| INSULIN | INSULIN | 1597001 | Regular Insulin, Human |
| INSULIN | INSULIN | 19078557 | Regular Insulin, Human |
| INSULIN | INSULIN | 1502910 | Insulin Glargine |

| | | | |
|---|---|---|---|
| GLYBURIDE | GLYBURIDE | 1559784 | Glyburide |
| NATEGLINIDE | NATEGLINIDE | 1502826 | nateglinide |
| GLIPIZIDE | GLIPIZIDE | 19030445 | Glipizide |
| INSULIN | INSULIN | 40052088 | Insulin Lispro |
| CHLORPROPAMIDE | CHLORPROPAMIDE | 1594978 | Chlorpropamide |
| METFORMIN | METFORMIN | 42705992 | Metformin |
| NATEGLINIDE | NATEGLINIDE | 19045606 | nateglinide |
| INSULIN | INSULIN | 1550028 | Insulin Lispro |
| INSULIN | INSULIN | 19078530 | insulin, isophane |
| GLIPIZIDE | GLIPIZIDE | 19085004 | Glipizide |
| METFORMIN | METFORMIN | 40164948 | Metformin |
| METFORMIN | METFORMIN | 40164882 | Metformin |
| INSULIN | INSULIN | 19110356 | Regular Insulin, Human |
| GLIPIZIDE | GLIPIZIDE | 19047638 | Glipizide |
| GLIMEPIRIDE | GLIMEPIRIDE | 1597756 | glimepiride |
| GLIPIZIDE | GLIPIZIDE | 1560171 | Glipizide |
| ACARBOSE | ACARBOSE | 1529331 | Acarbose |
| ALBIGLUTIDE | ALBIGLUTIDE | 44816283 | albiglutide |
| INSULIN | INSULIN | 1586346 | Insulin, Regular, Pork |
| INSULIN | INSULIN | 19120315 | insulin, isophane |
| METFORMIN | METFORMIN | 40135481 | Metformin |
| LINAGLIPTIN | LINAGLIPTIN | 36221104 | Linagliptin |
| INSULIN | INSULIN | 19078633 | Insulin, Regular, Pork |
| LINAGLIPTIN | LINAGLIPTIN | 40239219 | Linagliptin |
| INSULIN | INSULIN | 46234049 | Insulin, Aspart, Human |
| INSULIN | INSULIN | 19078556 | Regular Insulin, Human |
| INSULIN | INSULIN | 19116453 | insulin, isophane |
| METFORMIN | METFORMIN | 1503297 | Metformin |
| INSULIN | INSULIN | 1596960 | insulin, isophane |
| PRAMLINTIDE | PRAMLINTIDE | 40078056 | Pramlintide |
| TOLBUTAMIDE | TOLBUTAMIDE | 19037213 | Tolbutamide |
| SITAGLIPTIN | SITAGLIPTIN | 19125042 | sitagliptin |
| METFORMIN | METFORMIN | 36235752 | Metformin |
| METFORMIN | METFORMIN | 40163926 | Metformin |

| | | | |
|---|---|---|---|
| SITAGLIPTIN | SITAGLIPTIN | 19125045 | sitagliptin |
| GLYBURIDE | GLYBURIDE | 19045630 | Glyburide |
| DULAGLUTIDE | DULAGLUTIDE | 45774442 | dulaglutide |
| INSULIN | INSULIN | 40166274 | Insulin Lispro |
| INSULIN | INSULIN | 1516976 | insulin detemir |
| INSULIN | INSULIN | 19120790 | Insulin Glargine |
| INSULIN | INSULIN | 19116454 | insulin, isophane |
| METFORMIN | METFORMIN | 40063354 | Metformin |
| EXENATIDE | EXENATIDE | 46234117 | exenatide |
| GLIPIZIDE | GLIPIZIDE | 40167636 | Glipizide |
| GLIPIZIDE | GLIPIZIDE | 19085004 | Glipizide |
| INSULIN | INSULIN | 19078527 | Insulin, Zinc, Human |
| METFORMIN | METFORMIN | 40164932 | Metformin |
| SITAGLIPTIN | SITAGLIPTIN | 42901620 | sitagliptin |
| INSULIN | INSULIN | 1596972 | insulin, isophane |
| GLIPIZIDE | GLIPIZIDE | 19030445 | Glipizide |
| GLYBURIDE | GLYBURIDE | 40166004 | Glyburide |
| LINAGLIPTIN | LINAGLIPTIN | 42708089 | Linagliptin |
| METFORMIN | METFORMIN | 40164948 | Metformin |
| EXENATIDE | EXENATIDE | 19083254 | exenatide |
| INSULIN | INSULIN | 19062999 | insulin, isophane |
| GLIPIZIDE | GLIPIZIDE | 40167637 | Glipizide |
| GLYBURIDE | GLYBURIDE | 19077682 | Glyburide |
| CHLORPROPAMIDE | CHLORPROPAMIDE | 40030456 | Chlorpropamide |
| INSULIN | INSULIN | 19078530 | insulin, isophane |
| METFORMIN | METFORMIN | 1503328 | Metformin |
| INSULIN | INSULIN | 40184293 | Insulin, Aspart Protamine, Human |
| INSULIN | INSULIN | 1531601 | Insulin, Aspart Protamine, Human |
| INSULIN | INSULIN | 1513881 | Insulin Lispro |
| NATEGLINIDE | NATEGLINIDE | 1502826 | nateglinide |
| GLYBURIDE | GLYBURIDE | 40046211 | Glyburide |

| | | | |
|---|---|---|---|
| INSULIN | INSULIN | 1718711 | insulin degludec |
| ACETOHEXAMIDE | ACETOHEXAMIDE | 19048692 | Acetohexamide |
| CANAGLIFLOZIN | CANAGLIFLOZIN | 36246431 | canagliflozin |
| INSULIN | INSULIN | 46233948 | Insulin Lispro |
| METFORMIN | METFORMIN | 1503297 | Metformin |
| INSULIN | INSULIN | 19123376 | Insulin Lispro |
| METFORMIN | METFORMIN | 40164942 | Metformin |
| INSULIN | INSULIN | 1567232 | Insulin, Aspart, Human |
| METFORMIN | METFORMIN | 1503297 | Metformin |
| SITAGLIPTIN | SITAGLIPTIN | 19125042 | sitagliptin |
| GLYBURIDE | GLYBURIDE | 19045629 | Glyburide |
| LINAGLIPTIN | LINAGLIPTIN | 40239220 | Linagliptin |
| INSULIN | INSULIN | 46234045 | Insulin, Aspart, Human |
| METFORMIN | METFORMIN | 40164880 | Metformin |
| INSULIN | INSULIN | 19116611 | insulin, isophane |
| SITAGLIPTIN | SITAGLIPTIN | 40243628 | sitagliptin |
| INSULIN | INSULIN | 1567198 | Insulin, Aspart, Human |
| INSULIN | INSULIN | 1502905 | Insulin Glargine |
| INSULIN | INSULIN | 19135265 | Regular Insulin, Human |
| INSULIN | INSULIN | 1596952 | insulin, isophane |
| INSULIN | INSULIN | 1567232 | Insulin, Aspart, Human |
| INSULIN | INSULIN | 36231459 | Regular Insulin, Human |
| GLYBURIDE | GLYBURIDE | 40046201 | Glyburide |
| ACARBOSE | ACARBOSE | 19021787 | Acarbose |
| LIRAGLUTIDE | LIRAGLUTIDE | 36241903 | liraglutide |
| GLIMEPIRIDE | GLIMEPIRIDE | 1597756 | glimepiride |
| METFORMIN | METFORMIN | 40231393 | Metformin |
| CANAGLIFLOZIN | CANAGLIFLOZIN | 45774714 | canagliflozin |
| INSULIN | INSULIN | 1567198 | Insulin, Aspart, Human |
| PIOGLITAZONE | PIOGLITAZONE | 19099208 | pioglitazone |
| GLYBURIDE | GLYBURIDE | 1559785 | Glyburide |
| INSULIN | INSULIN | 1596959 | insulin, isophane |
| INSULIN | INSULIN | 1596959 | insulin, isophane |

| | | | |
|---|---|---|---|
| LIRAGLUTIDE | LIRAGLUTIDE | 40170913 | liraglutide |
| GLIPIZIDE | GLIPIZIDE | 19077636 | Glipizide |
| INSULIN | INSULIN | 19116453 | insulin, isophane |
| METFORMIN | METFORMIN | 1503297 | Metformin |
| METFORMIN | METFORMIN | 40171451 | Metformin |
| METFORMIN | METFORMIN | 40231396 | Metformin |
| GLIMEPIRIDE | GLIMEPIRIDE | 1597756 | glimepiride |
| EMPAGLIFLOZIN | EMPAGLIFLOZIN | 1592800 | empagliflozin |
| GLIMEPIRIDE | GLIMEPIRIDE | 19117198 | glimepiride |
| ALOGLIPTIN | ALOGLIPTIN | 43013922 | alogliptin |
| METFORMIN | METFORMIN | 40164925 | Metformin |
| INSULIN | INSULIN | 40052047 | insulin, isophane |
| INSULIN | INSULIN | 19120315 | insulin, isophane |
| METFORMIN | METFORMIN | 40164929 | Metformin |
| GLYBURIDE | GLYBURIDE | 1559761 | Glyburide |
| INSULIN | INSULIN | 19078557 | Regular Insulin, Human |
| METFORMIN | METFORMIN | 1503297 | Metformin |
| GLIMEPIRIDE | GLIMEPIRIDE | 43011499 | glimepiride |
| METFORMIN | METFORMIN | 40164890 | Metformin |
| METFORMIN | METFORMIN | 42705990 | Metformin |
| INSULIN | INSULIN | 19078603 | insulin, isophane |
| INSULIN | INSULIN | 36231456 | insulin, isophane |
| INSULIN | INSULIN | 1597001 | Regular Insulin, Human |
| EXENATIDE | EXENATIDE | 46234118 | exenatide |
| METFORMIN | METFORMIN | 1503327 | Metformin |
| METFORMIN | METFORMIN | 40163922 | Metformin |
| INSULIN | INSULIN | 19078557 | Regular Insulin, Human |
| GLIPIZIDE | GLIPIZIDE | 40044253 | Glipizide |
| GLIMEPIRIDE | GLIMEPIRIDE | 1597774 | glimepiride |

| | | | |
|---|---|---|---|
| INSULIN | INSULIN | 36233154 | Insulin, Glulisine, Human |
| INSULIN | INSULIN | 1502908 | Insulin Glargine |
| GLIPIZIDE | GLIPIZIDE | 19085004 | Glipizide |
| INSULIN | INSULIN | 19082658 | Insulin, Zinc, Pork |
| METFORMIN | METFORMIN | 40164926 | Metformin |
| GLYBURIDE | GLYBURIDE | 1559782 | Glyburide |
| GLIMEPIRIDE | GLIMEPIRIDE | 1597776 | glimepiride |
| INSULIN | INSULIN | 1513881 | Insulin Lispro |
| INSULIN | INSULIN | 36214699 | Insulin Glargine |
| INSULIN | INSULIN | 19131582 | Insulin, Glulisine, Human |
| INSULIN | INSULIN | 1502910 | Insulin Glargine |
| INSULIN | INSULIN | 1567232 | Insulin, Aspart, Human |
| PIOGLITAZONE | PIOGLITAZONE | 19079293 | pioglitazone |
| LINAGLIPTIN | LINAGLIPTIN | 42705856 | Linagliptin |
| ACARBOSE | ACARBOSE | 1529331 | Acarbose |
| GLYBURIDE | GLYBURIDE | 19077682 | Glyburide |
| GLIPIZIDE | GLIPIZIDE | 40165970 | Glipizide |
| INSULIN | INSULIN | 40176981 | Insulin, Aspart Protamine, Human |
| INSULIN | INSULIN | 1597001 | Regular Insulin, Human |
| INSULIN | INSULIN | 19082658 | Insulin, Zinc, Pork |
| INSULIN | INSULIN | 46233970 | Insulin Lispro |
| METFORMIN | METFORMIN | 1503328 | Metformin |
| INSULIN | INSULIN | 1718604 | Insulin Glargine |
| INSULIN | INSULIN | 19078607 | insulin, isophane |
| PIOGLITAZONE | PIOGLITAZONE | 1525216 | pioglitazone |
| METFORMIN | METFORMIN | 40164898 | Metformin |
| GLYBURIDE | GLYBURIDE | 1559684 | Glyburide |
| BROMOCRIPTINE | BROMOCRIPTINE | 19083958 | Bromocriptine |
| INSULIN | INSULIN | 36236032 | Insulin Lispro |
| TOLAZAMIDE | TOLAZAMIDE | 1502810 | Tolazamide |
| INSULIN | INSULIN | 1567198 | Insulin, Aspart, Human |
| INSULIN | INSULIN | 35602730 | insulin degludec |

| | | | |
|---|---|---|---|
| METFORMIN | METFORMIN | 40164923 | Metformin |
| GLIMEPIRIDE | GLIMEPIRIDE | 1597757 | glimepiride |
| GLIMEPIRIDE | GLIMEPIRIDE | 1597760 | glimepiride |
| ACARBOSE | ACARBOSE | 36216897 | Acarbose |
| ACETOHEXAMIDE | ACETOHEXAMIDE | 19018783 | Acetohexamide |
| ACETOHEXAMIDE | ACETOHEXAMIDE | 19048693 | Acetohexamide |
| ACETOHEXAMIDE | ACETOHEXAMIDE | 36216774 | Acetohexamide |
| ACETOHEXAMIDE | ACETOHEXAMIDE | 40004923 | Acetohexamide |
| ACETOHEXAMIDE | ACETOHEXAMIDE | 40004924 | Acetohexamide |
| ACETOHEXAMIDE | ACETOHEXAMIDE | 36216775 | Acetohexamide |
| ACETOHEXAMIDE | ACETOHEXAMIDE | 36235642 | Acetohexamide |
| ALBIGLUTIDE | ALBIGLUTIDE | 44816284 | albiglutide |
| ALBIGLUTIDE | ALBIGLUTIDE | 44816285 | albiglutide |
| ALBIGLUTIDE | ALBIGLUTIDE | 44816275 | albiglutide |
| ALBIGLUTIDE | ALBIGLUTIDE | 44816276 | albiglutide |
| ALBIGLUTIDE | ALBIGLUTIDE | 36246905 | albiglutide |
| ALBIGLUTIDE | ALBIGLUTIDE | 46275422 | albiglutide |
| ALBIGLUTIDE | ALBIGLUTIDE | 46275423 | albiglutide |
| ALOGLIPTIN | ALOGLIPTIN | 36246265 | alogliptin |
| ALOGLIPTIN | ALOGLIPTIN | 43013885 | alogliptin |
| ALOGLIPTIN | ALOGLIPTIN | 36246266 | alogliptin |
| ALOGLIPTIN | ALOGLIPTIN | 36246269 | alogliptin |
| ALOGLIPTIN | ALOGLIPTIN | 43013888 | alogliptin |
| ALOGLIPTIN | ALOGLIPTIN | 43013893 | alogliptin |
| ALOGLIPTIN | ALOGLIPTIN | 36246270 | alogliptin |
| ALOGLIPTIN | ALOGLIPTIN | 43013896 | alogliptin |
| ALOGLIPTIN | ALOGLIPTIN | 43013904 | alogliptin |
| ALOGLIPTIN | ALOGLIPTIN | 43013902 | alogliptin |
| ALOGLIPTIN | ALOGLIPTIN | 43013907 | alogliptin |
| ALOGLIPTIN | ALOGLIPTIN | 43013905 | alogliptin |
| ALOGLIPTIN | ALOGLIPTIN | 43013908 | alogliptin |
| ALOGLIPTIN | ALOGLIPTIN | 43013909 | alogliptin |
| ALOGLIPTIN | ALOGLIPTIN | 43013923 | alogliptin |
| ALOGLIPTIN | ALOGLIPTIN | 43013921 | alogliptin |
| ALOGLIPTIN | ALOGLIPTIN | 43013930 | alogliptin |
| ALOGLIPTIN | ALOGLIPTIN | 43013928 | alogliptin |
| ALOGLIPTIN | ALOGLIPTIN | 36246193 | alogliptin |
| ALOGLIPTIN | ALOGLIPTIN | 43013932 | alogliptin |
| ALOGLIPTIN | ALOGLIPTIN | 36246194 | alogliptin |

| | | | |
|---|---|---|---|
| ALOGLIPTIN | ALOGLIPTIN | 36246268 | alogliptin |
| ALOGLIPTIN | ALOGLIPTIN | 36246196 | alogliptin |
| BROMOCRIPTINE | BROMOCRIPTINE | 19103829 | Bromocriptine |
| BROMOCRIPTINE | BROMOCRIPTINE | 19134682 | Bromocriptine |
| BROMOCRIPTINE | BROMOCRIPTINE | 19062603 | Bromocriptine |
| BROMOCRIPTINE | BROMOCRIPTINE | 36214529 | Bromocriptine |
| BROMOCRIPTINE | BROMOCRIPTINE | 40016894 | Bromocriptine |
| BROMOCRIPTINE | BROMOCRIPTINE | 36214530 | Bromocriptine |
| CANAGLIFLOZIN | CANAGLIFLOZIN | 1718899 | canagliflozin |
| CANAGLIFLOZIN | CANAGLIFLOZIN | 1718904 | canagliflozin |
| CANAGLIFLOZIN | CANAGLIFLOZIN | 1718905 | canagliflozin |
| CANAGLIFLOZIN | CANAGLIFLOZIN | 1718908 | canagliflozin |
| CANAGLIFLOZIN | CANAGLIFLOZIN | 1718912 | canagliflozin |
| CANAGLIFLOZIN | CANAGLIFLOZIN | 1718913 | canagliflozin |
| CANAGLIFLOZIN | CANAGLIFLOZIN | 1718898 | canagliflozin |
| CANAGLIFLOZIN | CANAGLIFLOZIN | 1718900 | canagliflozin |
| CANAGLIFLOZIN | CANAGLIFLOZIN | 36247438 | canagliflozin |
| CANAGLIFLOZIN | CANAGLIFLOZIN | 45774709 | canagliflozin |
| CANAGLIFLOZIN | CANAGLIFLOZIN | 36247439 | canagliflozin |
| CANAGLIFLOZIN | CANAGLIFLOZIN | 1718902 | canagliflozin |
| CANAGLIFLOZIN | CANAGLIFLOZIN | 1718903 | canagliflozin |
| CANAGLIFLOZIN | CANAGLIFLOZIN | 45774710 | canagliflozin |
| CANAGLIFLOZIN | CANAGLIFLOZIN | 1718906 | canagliflozin |
| CANAGLIFLOZIN | CANAGLIFLOZIN | 1718907 | canagliflozin |
| CANAGLIFLOZIN | CANAGLIFLOZIN | 45774715 | canagliflozin |
| CANAGLIFLOZIN | CANAGLIFLOZIN | 1718910 | canagliflozin |
| CANAGLIFLOZIN | CANAGLIFLOZIN | 1718911 | canagliflozin |
| CANAGLIFLOZIN | CANAGLIFLOZIN | 1718914 | canagliflozin |
| CANAGLIFLOZIN | CANAGLIFLOZIN | 1718915 | canagliflozin |
| CANAGLIFLOZIN | CANAGLIFLOZIN | 45774722 | canagliflozin |
| CANAGLIFLOZIN | CANAGLIFLOZIN | 36246400 | canagliflozin |
| CANAGLIFLOZIN | CANAGLIFLOZIN | 36246401 | canagliflozin |
| CHLORPROPAMIDE | CHLORPROPAMIDE | 36214817 | Chlorpropamide |
| CHLORPROPAMIDE | CHLORPROPAMIDE | 40030438 | Chlorpropamide |
| CHLORPROPAMIDE | CHLORPROPAMIDE | 36214818 | Chlorpropamide |
| CHLORPROPAMIDE | CHLORPROPAMIDE | 36214819 | Chlorpropamide |
| CHLORPROPAMIDE | CHLORPROPAMIDE | 40030439 | Chlorpropamide |
| CHLORPROPAMIDE | CHLORPROPAMIDE | 36214820 | Chlorpropamide |
| CHLORPROPAMIDE | CHLORPROPAMIDE | 19060469 | Chlorpropamide |
| CHLORPROPAMIDE | CHLORPROPAMIDE | 19107377 | Chlorpropamide |

| | | | |
|---|---|---|---|
| CHLORPROPAMIDE | CHLORPROPAMIDE | 19107661 | Chlorpropamide |
| CHLORPROPAMIDE | CHLORPROPAMIDE | 19113845 | Chlorpropamide |
| CHLORPROPAMIDE | CHLORPROPAMIDE | 19113846 | Chlorpropamide |
| CHLORPROPAMIDE | CHLORPROPAMIDE | 19109283 | Chlorpropamide |
| CHLORPROPAMIDE | CHLORPROPAMIDE | 19106977 | Chlorpropamide |
| CHLORPROPAMIDE | CHLORPROPAMIDE | 36214821 | Chlorpropamide |
| CHLORPROPAMIDE | CHLORPROPAMIDE | 40030457 | Chlorpropamide |
| CHLORPROPAMIDE | CHLORPROPAMIDE | 40108537 | Chlorpropamide |
| CHLORPROPAMIDE | CHLORPROPAMIDE | 36214822 | Chlorpropamide |
| CHLORPROPAMIDE | CHLORPROPAMIDE | 36233236 | Chlorpropamide |
| CHLORPROPAMIDE | CHLORPROPAMIDE | 36233237 | Chlorpropamide |
| DAPAGLIFLOZIN | DAPAGLIFLOZIN | 45775455 | dapagliflozin |
| DAPAGLIFLOZIN | DAPAGLIFLOZIN | 45775619 | dapagliflozin |
| DAPAGLIFLOZIN | DAPAGLIFLOZIN | 793826 | dapagliflozin |
| DAPAGLIFLOZIN | DAPAGLIFLOZIN | 793828 | dapagliflozin |
| DAPAGLIFLOZIN | DAPAGLIFLOZIN | 45775621 | dapagliflozin |
| DAPAGLIFLOZIN | DAPAGLIFLOZIN | 45775623 | dapagliflozin |
| DAPAGLIFLOZIN | DAPAGLIFLOZIN | 44785829 | dapagliflozin |
| DAPAGLIFLOZIN | DAPAGLIFLOZIN | 45775454 | dapagliflozin |
| DAPAGLIFLOZIN | DAPAGLIFLOZIN | 45775859 | dapagliflozin |
| DAPAGLIFLOZIN | DAPAGLIFLOZIN | 1592314 | dapagliflozin |
| DAPAGLIFLOZIN | DAPAGLIFLOZIN | 1592316 | dapagliflozin |
| DAPAGLIFLOZIN | DAPAGLIFLOZIN | 1592320 | dapagliflozin |
| DAPAGLIFLOZIN | DAPAGLIFLOZIN | 1592315 | dapagliflozin |
| DAPAGLIFLOZIN | DAPAGLIFLOZIN | 45775456 | dapagliflozin |
| DAPAGLIFLOZIN | DAPAGLIFLOZIN | 45775861 | dapagliflozin |
| DAPAGLIFLOZIN | DAPAGLIFLOZIN | 45775620 | dapagliflozin |
| DAPAGLIFLOZIN | DAPAGLIFLOZIN | 45775863 | dapagliflozin |
| DAPAGLIFLOZIN | DAPAGLIFLOZIN | 1592319 | dapagliflozin |
| DAPAGLIFLOZIN | DAPAGLIFLOZIN | 1592317 | dapagliflozin |
| DAPAGLIFLOZIN | DAPAGLIFLOZIN | 1592323 | dapagliflozin |
| DAPAGLIFLOZIN | DAPAGLIFLOZIN | 793825 | dapagliflozin |
| DAPAGLIFLOZIN | DAPAGLIFLOZIN | 793827 | dapagliflozin |
| DAPAGLIFLOZIN | DAPAGLIFLOZIN | 793829 | dapagliflozin |
| DAPAGLIFLOZIN | DAPAGLIFLOZIN | 793830 | dapagliflozin |
| DAPAGLIFLOZIN | DAPAGLIFLOZIN | 45775622 | dapagliflozin |
| DAPAGLIFLOZIN | DAPAGLIFLOZIN | 45775624 | dapagliflozin |
| DAPAGLIFLOZIN | DAPAGLIFLOZIN | 45775868 | dapagliflozin |
| DAPAGLIFLOZIN | DAPAGLIFLOZIN | 36247667 | dapagliflozin |
| DAPAGLIFLOZIN | DAPAGLIFLOZIN | 36247668 | dapagliflozin |

| | | | |
|---|---|---|---|
| DAPAGLIFLOZIN | DAPAGLIFLOZIN | 1592321 | dapagliflozin |
| DAPAGLIFLOZIN | DAPAGLIFLOZIN | 1592322 | dapagliflozin |
| DULAGLUTIDE | DULAGLUTIDE | 45774443 | dulaglutide |
| DULAGLUTIDE | DULAGLUTIDE | 45774444 | dulaglutide |
| DULAGLUTIDE | DULAGLUTIDE | 45774449 | dulaglutide |
| DULAGLUTIDE | DULAGLUTIDE | 45774450 | dulaglutide |
| DULAGLUTIDE | DULAGLUTIDE | 46234470 | dulaglutide |
| EMPAGLIFLOZIN | EMPAGLIFLOZIN | 1592653 | empagliflozin |
| EMPAGLIFLOZIN | EMPAGLIFLOZIN | 1592794 | empagliflozin |
| EMPAGLIFLOZIN | EMPAGLIFLOZIN | 1592798 | empagliflozin |
| EMPAGLIFLOZIN | EMPAGLIFLOZIN | 1592803 | empagliflozin |
| EMPAGLIFLOZIN | EMPAGLIFLOZIN | 36248787 | empagliflozin |
| EMPAGLIFLOZIN | EMPAGLIFLOZIN | 45892171 | empagliflozin |
| EMPAGLIFLOZIN | EMPAGLIFLOZIN | 45892175 | empagliflozin |
| EMPAGLIFLOZIN | EMPAGLIFLOZIN | 1592652 | empagliflozin |
| EMPAGLIFLOZIN | EMPAGLIFLOZIN | 1592655 | empagliflozin |
| EMPAGLIFLOZIN | EMPAGLIFLOZIN | 36248374 | empagliflozin |
| EMPAGLIFLOZIN | EMPAGLIFLOZIN | 46287679 | empagliflozin |
| EMPAGLIFLOZIN | EMPAGLIFLOZIN | 46287683 | empagliflozin |
| EMPAGLIFLOZIN | EMPAGLIFLOZIN | 36248375 | empagliflozin |
| EMPAGLIFLOZIN | EMPAGLIFLOZIN | 1592654 | empagliflozin |
| EMPAGLIFLOZIN | EMPAGLIFLOZIN | 1592657 | empagliflozin |
| EMPAGLIFLOZIN | EMPAGLIFLOZIN | 1592793 | empagliflozin |
| EMPAGLIFLOZIN | EMPAGLIFLOZIN | 1592796 | empagliflozin |
| EMPAGLIFLOZIN | EMPAGLIFLOZIN | 1592797 | empagliflozin |
| EMPAGLIFLOZIN | EMPAGLIFLOZIN | 46287408 | empagliflozin |
| EMPAGLIFLOZIN | EMPAGLIFLOZIN | 46287686 | empagliflozin |
| EMPAGLIFLOZIN | EMPAGLIFLOZIN | 46287688 | empagliflozin |
| EMPAGLIFLOZIN | EMPAGLIFLOZIN | 1592799 | empagliflozin |
| EMPAGLIFLOZIN | EMPAGLIFLOZIN | 1592801 | empagliflozin |
| EMPAGLIFLOZIN | EMPAGLIFLOZIN | 1592802 | empagliflozin |
| EMPAGLIFLOZIN | EMPAGLIFLOZIN | 1592805 | empagliflozin |
| EMPAGLIFLOZIN | EMPAGLIFLOZIN | 1592806 | empagliflozin |
| EMPAGLIFLOZIN | EMPAGLIFLOZIN | 46287689 | empagliflozin |
| EMPAGLIFLOZIN | EMPAGLIFLOZIN | 46287680 | empagliflozin |
| EMPAGLIFLOZIN | EMPAGLIFLOZIN | 36246884 | empagliflozin |
| EMPAGLIFLOZIN | EMPAGLIFLOZIN | 36248377 | empagliflozin |
| EXENATIDE | EXENATIDE | 45774531 | exenatide |
| EXENATIDE | EXENATIDE | 793111 | exenatide |
| EXENATIDE | EXENATIDE | 793114 | exenatide |

| | | | |
|---|---|---|---|
| EXENATIDE | EXENATIDE | 1583729 | exenatide |
| EXENATIDE | EXENATIDE | 19135405 | exenatide |
| EXENATIDE | EXENATIDE | 793109 | exenatide |
| EXENATIDE | EXENATIDE | 793112 | exenatide |
| EXENATIDE | EXENATIDE | 793115 | exenatide |
| EXENATIDE | EXENATIDE | 793116 | exenatide |
| EXENATIDE | EXENATIDE | 42705839 | exenatide |
| EXENATIDE | EXENATIDE | 45774534 | exenatide |
| EXENATIDE | EXENATIDE | 793110 | exenatide |
| EXENATIDE | EXENATIDE | 793113 | exenatide |
| EXENATIDE | EXENATIDE | 36222278 | exenatide |
| EXENATIDE | EXENATIDE | 46234120 | exenatide |
| EXENATIDE | EXENATIDE | 46234121 | exenatide |
| GLICLAZIDE | GLICLAZIDE | 19059800 | Gliclazide |
| GLICLAZIDE | GLICLAZIDE | 36218763 | Gliclazide |
| GLICLAZIDE | GLICLAZIDE | 36218764 | Gliclazide |
| GLIMEPIRIDE | GLIMEPIRIDE | 36214980 | glimepiride |
| GLIMEPIRIDE | GLIMEPIRIDE | 40135391 | glimepiride |
| GLIMEPIRIDE | GLIMEPIRIDE | 40143966 | glimepiride |
| GLIMEPIRIDE | GLIMEPIRIDE | 36214981 | glimepiride |
| GLIMEPIRIDE | GLIMEPIRIDE | 36214982 | glimepiride |
| GLIMEPIRIDE | GLIMEPIRIDE | 40129534 | glimepiride |
| GLIMEPIRIDE | GLIMEPIRIDE | 36214983 | glimepiride |
| GLIMEPIRIDE | GLIMEPIRIDE | 19122366 | glimepiride |
| GLIMEPIRIDE | GLIMEPIRIDE | 19126618 | glimepiride |
| GLIMEPIRIDE | GLIMEPIRIDE | 19135388 | glimepiride |
| GLIMEPIRIDE | GLIMEPIRIDE | 19122368 | glimepiride |
| GLIMEPIRIDE | GLIMEPIRIDE | 19126619 | glimepiride |
| GLIPIZIDE | GLIPIZIDE | 36235739 | Glipizide |
| GLIPIZIDE | GLIPIZIDE | 36235740 | Glipizide |
| GLIPIZIDE | GLIPIZIDE | 36218765 | Glipizide |
| GLIPIZIDE | GLIPIZIDE | 19117693 | Glipizide |
| GLIPIZIDE | GLIPIZIDE | 19025947 | Glipizide |
| GLIPIZIDE | GLIPIZIDE | 19117692 | Glipizide |
| GLIPIZIDE | GLIPIZIDE | 19116148 | Glipizide |
| GLIPIZIDE | GLIPIZIDE | 19025946 | Glipizide |
| GLIPIZIDE | GLIPIZIDE | 40044252 | Glipizide |
| GLIPIZIDE | GLIPIZIDE | 40044254 | Glipizide |
| GLIPIZIDE | GLIPIZIDE | 36229921 | Glipizide |
| GLIPIZIDE | GLIPIZIDE | 36229922 | Glipizide |

| | | | |
|---|---|---|---|
| GLIQUIDONE | GLIQUIDONE | 19097821 | gliquidone |
| GLIQUIDONE | GLIQUIDONE | 19092911 | gliquidone |
| GLIQUIDONE | GLIQUIDONE | 19006931 | gliquidone |
| GLIQUIDONE | GLIQUIDONE | 36214986 | gliquidone |
| GLIQUIDONE | GLIQUIDONE | 40059670 | gliquidone |
| GLIQUIDONE | GLIQUIDONE | 36214987 | gliquidone |
| GLYBURIDE | GLYBURIDE | 36234721 | Glyburide |
| GLYBURIDE | GLYBURIDE | 36234722 | Glyburide |
| GLYBURIDE | GLYBURIDE | 36220647 | Glyburide |
| GLYBURIDE | GLYBURIDE | 36220648 | Glyburide |
| GLYBURIDE | GLYBURIDE | 36230037 | Glyburide |
| GLYBURIDE | GLYBURIDE | 36230038 | Glyburide |
| GLYBURIDE | GLYBURIDE | 36235405 | Glyburide |
| GLYBURIDE | GLYBURIDE | 36235406 | Glyburide |
| GLYBURIDE | GLYBURIDE | 36233481 | Glyburide |
| GLYBURIDE | GLYBURIDE | 36233482 | Glyburide |
| GLYBURIDE | GLYBURIDE | 36213838 | Glyburide |
| GLYBURIDE | GLYBURIDE | 40046145 | Glyburide |
| GLYBURIDE | GLYBURIDE | 19125218 | Glyburide |
| GLYBURIDE | GLYBURIDE | 19125219 | Glyburide |
| GLYBURIDE | GLYBURIDE | 19120562 | Glyburide |
| GLYBURIDE | GLYBURIDE | 19106975 | Glyburide |
| GLYBURIDE | GLYBURIDE | 19116146 | Glyburide |
| GLYBURIDE | GLYBURIDE | 19117420 | Glyburide |
| GLYBURIDE | GLYBURIDE | 19117418 | Glyburide |
| GLYBURIDE | GLYBURIDE | 19125220 | Glyburide |
| GLYBURIDE | GLYBURIDE | 19117416 | Glyburide |
| GLYBURIDE | GLYBURIDE | 19117419 | Glyburide |
| GLYBURIDE | GLYBURIDE | 19117417 | Glyburide |
| GLYBURIDE | GLYBURIDE | 19006908 | Glyburide |
| GLYBURIDE | GLYBURIDE | 19024506 | Glyburide |
| GLYBURIDE | GLYBURIDE | 19024504 | Glyburide |
| GLYBURIDE | GLYBURIDE | 19125221 | Glyburide |
| GLYBURIDE | GLYBURIDE | 19024502 | Glyburide |
| GLYBURIDE | GLYBURIDE | 19024505 | Glyburide |
| GLYBURIDE | GLYBURIDE | 19024503 | Glyburide |
| GLYBURIDE | GLYBURIDE | 19099409 | Glyburide |
| GLYBURIDE | GLYBURIDE | 1559729 | Glyburide |
| GLYBURIDE | GLYBURIDE | 19099408 | Glyburide |
| GLYBURIDE | GLYBURIDE | 19117424 | Glyburide |

| | | | |
|---|---|---|---|
| GLYBURIDE | GLYBURIDE | 19117422 | Glyburide |
| GLYBURIDE | GLYBURIDE | 19125222 | Glyburide |
| GLYBURIDE | GLYBURIDE | 19117421 | Glyburide |
| GLYBURIDE | GLYBURIDE | 19117423 | Glyburide |
| GLYBURIDE | GLYBURIDE | 19006909 | Glyburide |
| GLYBURIDE | GLYBURIDE | 19024510 | Glyburide |
| GLYBURIDE | GLYBURIDE | 19125223 | Glyburide |
| GLYBURIDE | GLYBURIDE | 19024507 | Glyburide |
| GLYBURIDE | GLYBURIDE | 19024509 | Glyburide |
| GLYBURIDE | GLYBURIDE | 19120563 | Glyburide |
| GLYBURIDE | GLYBURIDE | 19067366 | Glyburide |
| GLYBURIDE | GLYBURIDE | 36213840 | Glyburide |
| GLYBURIDE | GLYBURIDE | 40046202 | Glyburide |
| GLYBURIDE | GLYBURIDE | 40046203 | Glyburide |
| GLYBURIDE | GLYBURIDE | 40046205 | Glyburide |
| GLYBURIDE | GLYBURIDE | 40046206 | Glyburide |
| GLYBURIDE | GLYBURIDE | 40136854 | Glyburide |
| GLYBURIDE | GLYBURIDE | 40046207 | Glyburide |
| GLYBURIDE | GLYBURIDE | 40046209 | Glyburide |
| GLYBURIDE | GLYBURIDE | 40046210 | Glyburide |
| GLYBURIDE | GLYBURIDE | 40046212 | Glyburide |
| GLYBURIDE | GLYBURIDE | 36233491 | Glyburide |
| GLYBURIDE | GLYBURIDE | 36233492 | Glyburide |
| GLYBURIDE | GLYBURIDE | 36240751 | Glyburide |
| GLYBURIDE | GLYBURIDE | 36240752 | Glyburide |
| GLYBURIDE | GLYBURIDE | 36231233 | Glyburide |
| GLYBURIDE | GLYBURIDE | 36231234 | Glyburide |
| GLYBURIDE | GLYBURIDE | 36232128 | Glyburide |
| GLYBURIDE | GLYBURIDE | 36232129 | Glyburide |
| INSULIN | INSULIN | 35605670 | insulin argine |
| INSULIN | INSULIN | 1718706 | insulin degludec |
| INSULIN | INSULIN | 35602720 | insulin degludec |
| INSULIN | INSULIN | 35602729 | insulin degludec |
| INSULIN | INSULIN | 793408 | insulin degludec |
| INSULIN | INSULIN | 793413 | insulin degludec |
| INSULIN | INSULIN | 35602717 | insulin degludec |
| INSULIN | INSULIN | 793406 | insulin degludec |
| INSULIN | INSULIN | 793407 | insulin degludec |
| INSULIN | INSULIN | 793411 | insulin degludec |
| INSULIN | INSULIN | 1718704 | insulin degludec |

| | | | |
|---|---|---|---|
| INSULIN | INSULIN | 1718705 | insulin degludec |
| INSULIN | INSULIN | 1718709 | insulin degludec |
| INSULIN | INSULIN | 1718708 | insulin degludec |
| INSULIN | INSULIN | 1718712 | insulin degludec |
| INSULIN | INSULIN | 1718713 | insulin degludec |
| INSULIN | INSULIN | 35602732 | insulin degludec |
| INSULIN | INSULIN | 35602733 | insulin degludec |
| INSULIN | INSULIN | 793405 | insulin degludec |
| INSULIN | INSULIN | 793410 | insulin degludec |
| INSULIN | INSULIN | 793498 | insulin degludec |
| INSULIN | INSULIN | 793499 | insulin degludec |
| INSULIN | INSULIN | 35602719 | insulin degludec |
| INSULIN | INSULIN | 793412 | insulin degludec |
| INSULIN | INSULIN | 1718710 | insulin degludec |
| INSULIN | INSULIN | 42902821 | insulin detemir |
| INSULIN | INSULIN | 42903168 | insulin detemir |
| INSULIN | INSULIN | 40056632 | insulin detemir |
| INSULIN | INSULIN | 46234190 | insulin detemir |
| INSULIN | INSULIN | 46234191 | insulin detemir |
| INSULIN | INSULIN | 46221553 | Insulin Glargine |
| INSULIN | INSULIN | 1718599 | Insulin Glargine |
| INSULIN | INSULIN | 42544059 | Insulin Glargine |
| INSULIN | INSULIN | 42544060 | Insulin Glargine |
| INSULIN | INSULIN | 36248425 | Insulin Glargine |
| INSULIN | INSULIN | 1718597 | Insulin Glargine |
| INSULIN | INSULIN | 1718598 | Insulin Glargine |
| INSULIN | INSULIN | 1718602 | Insulin Glargine |
| INSULIN | INSULIN | 1718601 | Insulin Glargine |
| INSULIN | INSULIN | 1718605 | Insulin Glargine |
| INSULIN | INSULIN | 1718606 | Insulin Glargine |
| INSULIN | INSULIN | 35604092 | Insulin Glargine |
| INSULIN | INSULIN | 42902742 | Insulin Glargine |
| INSULIN | INSULIN | 35604095 | Insulin Glargine |
| INSULIN | INSULIN | 46234097 | Insulin Glargine |
| INSULIN | INSULIN | 35604093 | Insulin Glargine |
| INSULIN | INSULIN | 46234099 | Insulin Glargine |
| INSULIN | INSULIN | 1718603 | Insulin Glargine |
| INSULIN | INSULIN | 19129443 | insulin human, rDNA origin |
| INSULIN | INSULIN | 19129439 | insulin human, rDNA origin |
| INSULIN | INSULIN | 1588986 | insulin human, rDNA origin |

| | | | |
|---|---|---|---|
| INSULIN | INSULIN | 19129428 | insulin human, rDNA origin |
| INSULIN | INSULIN | 19129431 | insulin human, rDNA origin |
| INSULIN | INSULIN | 19129433 | insulin human, rDNA origin |
| INSULIN | INSULIN | 19129435 | insulin human, rDNA origin |
| INSULIN | INSULIN | 42903044 | insulin human, rDNA origin |
| INSULIN | INSULIN | 19129438 | insulin human, rDNA origin |
| INSULIN | INSULIN | 40148823 | insulin human, rDNA origin |
| INSULIN | INSULIN | 40148825 | insulin human, rDNA origin |
| INSULIN | INSULIN | 40148826 | insulin human, rDNA origin |
| INSULIN | INSULIN | 36224229 | insulin human, rDNA origin |
| INSULIN | INSULIN | 1592474 | Insulin Lispro |
| INSULIN | INSULIN | 1592475 | Insulin Lispro |
| INSULIN | INSULIN | 46233974 | Insulin Lispro |
| INSULIN | INSULIN | 793225 | Insulin Lispro |
| INSULIN | INSULIN | 1513915 | Insulin Lispro |
| INSULIN | INSULIN | 1513913 | Insulin Lispro |
| INSULIN | INSULIN | 793222 | Insulin Lispro |
| INSULIN | INSULIN | 36214700 | Insulin Lispro |
| INSULIN | INSULIN | 40052077 | Insulin Lispro |
| INSULIN | INSULIN | 40143226 | Insulin Lispro |
| INSULIN | INSULIN | 46234016 | Insulin Lispro |
| INSULIN | INSULIN | 46234017 | Insulin Lispro |
| INSULIN | INSULIN | 793220 | Insulin Lispro |
| INSULIN | INSULIN | 19123375 | Insulin Lispro |
| INSULIN | INSULIN | 46233977 | Insulin Lispro |
| INSULIN | INSULIN | 793223 | Insulin Lispro |
| INSULIN | INSULIN | 46233971 | Insulin Lispro |
| INSULIN | INSULIN | 793226 | Insulin Lispro |
| INSULIN | INSULIN | 46233949 | Insulin Lispro |
| INSULIN | INSULIN | 1513877 | Insulin Lispro |
| INSULIN | INSULIN | 42902587 | Insulin Lispro |
| INSULIN | INSULIN | 42902923 | Insulin Lispro |
| INSULIN | INSULIN | 19033127 | Insulin Lispro |
| INSULIN | INSULIN | 19067324 | Insulin Lispro |
| INSULIN | INSULIN | 42902945 | Insulin Lispro |
| INSULIN | INSULIN | 42902599 | Insulin Lispro |
| INSULIN | INSULIN | 46233973 | Insulin Lispro |
| INSULIN | INSULIN | 36214701 | Insulin Lispro |
| INSULIN | INSULIN | 793221 | Insulin Lispro |
| INSULIN | INSULIN | 40130843 | Insulin Lispro |

| | | | |
|---|---|---|---|
| INSULIN | INSULIN | 46233944 | Insulin Lispro |
| INSULIN | INSULIN | 793224 | Insulin Lispro |
| INSULIN | INSULIN | 1567234 | Insulin, Aspart Protamine, Human |
| INSULIN | INSULIN | 36214702 | Insulin, Aspart Protamine, Human |
| INSULIN | INSULIN | 40051007 | Insulin, Aspart Protamine, Human |
| INSULIN | INSULIN | 40184291 | Insulin, Aspart Protamine, Human |
| INSULIN | INSULIN | 46234052 | Insulin, Aspart Protamine, Human |
| INSULIN | INSULIN | 46234053 | Insulin, Aspart Protamine, Human |
| INSULIN | INSULIN | 19133791 | Insulin, Aspart Protamine, Human |
| INSULIN | INSULIN | 19133793 | Insulin, Aspart Protamine, Human |
| INSULIN | INSULIN | 1531623 | Insulin, Aspart Protamine, Human |
| INSULIN | INSULIN | 42902356 | Insulin, Aspart Protamine, Human |
| INSULIN | INSULIN | 46234041 | Insulin, Aspart, Human |
| INSULIN | INSULIN | 792697 | Insulin, Aspart, Human |
| INSULIN | INSULIN | 792694 | Insulin, Aspart, Human |
| INSULIN | INSULIN | 792692 | Insulin, Aspart, Human |
| INSULIN | INSULIN | 19005729 | Insulin, Aspart, Human |
| INSULIN | INSULIN | 46234044 | Insulin, Aspart, Human |
| INSULIN | INSULIN | 792695 | Insulin, Aspart, Human |
| INSULIN | INSULIN | 46234050 | Insulin, Aspart, Human |
| INSULIN | INSULIN | 792698 | Insulin, Aspart, Human |
| INSULIN | INSULIN | 46234051 | Insulin, Aspart, Human |
| INSULIN | INSULIN | 19099834 | Insulin, Aspart, Human |
| INSULIN | INSULIN | 19133792 | Insulin, Aspart, Human |
| INSULIN | INSULIN | 46234040 | Insulin, Aspart, Human |
| INSULIN | INSULIN | 46234042 | Insulin, Aspart, Human |
| INSULIN | INSULIN | 36214703 | Insulin, Aspart, Human |
| INSULIN | INSULIN | 40051314 | Insulin, Aspart, Human |
| INSULIN | INSULIN | 792693 | Insulin, Aspart, Human |
| INSULIN | INSULIN | 40051316 | Insulin, Aspart, Human |
| INSULIN | INSULIN | 46234046 | Insulin, Aspart, Human |
| INSULIN | INSULIN | 792696 | Insulin, Aspart, Human |
| INSULIN | INSULIN | 46234048 | Insulin, Aspart, Human |
| INSULIN | INSULIN | 1544872 | Insulin, Glulisine, Human |
| INSULIN | INSULIN | 19131581 | Insulin, Glulisine, Human |
| INSULIN | INSULIN | 42902468 | Insulin, Glulisine, Human |
| INSULIN | INSULIN | 42902371 | Insulin, Glulisine, Human |
| INSULIN | INSULIN | 36214704 | Insulin, Glulisine, Human |
| INSULIN | INSULIN | 40051353 | Insulin, Glulisine, Human |
| INSULIN | INSULIN | 40153110 | Insulin, Glulisine, Human |

| | | | |
|---|---|---|---|
| INSULIN | INSULIN | 46233875 | Insulin, Glulisine, Human |
| INSULIN | INSULIN | 46233876 | Insulin, Glulisine, Human |
| INSULIN | INSULIN | 1596976 | insulin, isophane |
| INSULIN | INSULIN | 36235889 | insulin, isophane |
| INSULIN | INSULIN | 36236037 | insulin, isophane |
| INSULIN | INSULIN | 36236038 | insulin, isophane |
| INSULIN | INSULIN | 36235838 | insulin, isophane |
| INSULIN | INSULIN | 36232390 | insulin, isophane |
| INSULIN | INSULIN | 36232394 | insulin, isophane |
| INSULIN | INSULIN | 36232395 | insulin, isophane |
| INSULIN | INSULIN | 36221859 | insulin, isophane |
| INSULIN | INSULIN | 36236150 | insulin, isophane |
| INSULIN | INSULIN | 19095212 | insulin, isophane |
| INSULIN | INSULIN | 19117090 | insulin, isophane |
| INSULIN | INSULIN | 19017221 | insulin, isophane |
| INSULIN | INSULIN | 19014972 | insulin, isophane |
| INSULIN | INSULIN | 19095255 | insulin, isophane |
| INSULIN | INSULIN | 19120825 | insulin, isophane |
| INSULIN | INSULIN | 19069663 | insulin, isophane |
| INSULIN | INSULIN | 19078604 | insulin, isophane |
| INSULIN | INSULIN | 19078529 | insulin, isophane |
| INSULIN | INSULIN | 19078605 | insulin, isophane |
| INSULIN | INSULIN | 19116794 | insulin, isophane |
| INSULIN | INSULIN | 19116795 | insulin, isophane |
| INSULIN | INSULIN | 19063033 | insulin, isophane |
| INSULIN | INSULIN | 19116730 | insulin, isophane |
| INSULIN | INSULIN | 19012702 | insulin, isophane |
| INSULIN | INSULIN | 19012270 | insulin, isophane |
| INSULIN | INSULIN | 46234239 | insulin, isophane |
| INSULIN | INSULIN | 42902606 | insulin, isophane |
| INSULIN | INSULIN | 42902571 | insulin, isophane |
| INSULIN | INSULIN | 19104470 | insulin, isophane |
| INSULIN | INSULIN | 19117028 | insulin, isophane |
| INSULIN | INSULIN | 19116451 | insulin, isophane |
| INSULIN | INSULIN | 19010533 | insulin, isophane |
| INSULIN | INSULIN | 19009388 | insulin, isophane |
| INSULIN | INSULIN | 19116613 | insulin, isophane |
| INSULIN | INSULIN | 19010534 | insulin, isophane |
| INSULIN | INSULIN | 19010532 | insulin, isophane |
| INSULIN | INSULIN | 46234234 | insulin, isophane |

| | | | |
|---|---|---|---|
| INSULIN | INSULIN | 42903059 | insulin, isophane |
| INSULIN | INSULIN | 42903113 | insulin, isophane |
| INSULIN | INSULIN | 19101876 | insulin, isophane |
| INSULIN | INSULIN | 19009387 | insulin, isophane |
| INSULIN | INSULIN | 1596954 | insulin, isophane |
| INSULIN | INSULIN | 19010531 | insulin, isophane |
| INSULIN | INSULIN | 19009421 | insulin, isophane |
| INSULIN | INSULIN | 1596917 | insulin, isophane |
| INSULIN | INSULIN | 19009386 | insulin, isophane |
| INSULIN | INSULIN | 19010510 | insulin, isophane |
| INSULIN | INSULIN | 19009390 | insulin, isophane |
| INSULIN | INSULIN | 19117088 | insulin, isophane |
| INSULIN | INSULIN | 19062719 | insulin, isophane |
| INSULIN | INSULIN | 19116884 | insulin, isophane |
| INSULIN | INSULIN | 19017199 | insulin, isophane |
| INSULIN | INSULIN | 19016044 | insulin, isophane |
| INSULIN | INSULIN | 19116452 | insulin, isophane |
| INSULIN | INSULIN | 19115111 | insulin, isophane |
| INSULIN | INSULIN | 19009389 | insulin, isophane |
| INSULIN | INSULIN | 19095216 | insulin, isophane |
| INSULIN | INSULIN | 19117089 | insulin, isophane |
| INSULIN | INSULIN | 19017200 | insulin, isophane |
| INSULIN | INSULIN | 19016043 | insulin, isophane |
| INSULIN | INSULIN | 46221581 | insulin, isophane |
| INSULIN | INSULIN | 36249005 | insulin, isophane |
| INSULIN | INSULIN | 40052050 | insulin, isophane |
| INSULIN | INSULIN | 40052051 | insulin, isophane |
| INSULIN | INSULIN | 40050609 | insulin, isophane |
| INSULIN | INSULIN | 40052053 | insulin, isophane |
| INSULIN | INSULIN | 36249004 | insulin, isophane |
| INSULIN | INSULIN | 40051377 | insulin, isophane |
| INSULIN | INSULIN | 40051379 | insulin, isophane |
| INSULIN | INSULIN | 40051382 | insulin, isophane |
| INSULIN | INSULIN | 40051689 | insulin, isophane |
| INSULIN | INSULIN | 40051690 | insulin, isophane |
| INSULIN | INSULIN | 46234216 | insulin, isophane |
| INSULIN | INSULIN | 46234233 | insulin, isophane |
| INSULIN | INSULIN | 40159519 | insulin, isophane |
| INSULIN | INSULIN | 40159520 | insulin, isophane |
| INSULIN | INSULIN | 40159521 | insulin, isophane |

| | | | |
|---|---|---|---|
| INSULIN | INSULIN | 36249003 | insulin, isophane |
| INSULIN | INSULIN | 40051374 | insulin, isophane |
| INSULIN | INSULIN | 40052043 | insulin, isophane |
| INSULIN | INSULIN | 40052044 | insulin, isophane |
| INSULIN | INSULIN | 40051364 | insulin, isophane |
| INSULIN | INSULIN | 40052072 | insulin, isophane |
| INSULIN | INSULIN | 40051375 | insulin, isophane |
| INSULIN | INSULIN | 40052073 | insulin, isophane |
| INSULIN | INSULIN | 40052075 | insulin, isophane |
| INSULIN | INSULIN | 40052048 | insulin, isophane |
| INSULIN | INSULIN | 46234236 | insulin, isophane |
| INSULIN | INSULIN | 46234238 | insulin, isophane |
| INSULIN | INSULIN | 40159543 | insulin, isophane |
| INSULIN | INSULIN | 40159544 | insulin, isophane |
| INSULIN | INSULIN | 40159545 | insulin, isophane |
| INSULIN | INSULIN | 36228162 | insulin, isophane |
| INSULIN | INSULIN | 36242927 | insulin, isophane |
| INSULIN | INSULIN | 36242928 | insulin, isophane |
| INSULIN | INSULIN | 36241496 | insulin, isophane |
| INSULIN | INSULIN | 36241497 | insulin, isophane |
| INSULIN | INSULIN | 19090244 | Insulin, Prompt Zinc, Beef |
| INSULIN | INSULIN | 19090246 | Insulin, Prompt Zinc, Beef |
| INSULIN | INSULIN | 19090245 | Insulin, Prompt Zinc, Beef |
| INSULIN | INSULIN | 36214705 | Insulin, Prompt Zinc, Beef |
| INSULIN | INSULIN | 40052093 | Insulin, Prompt Zinc, Beef |
| INSULIN | INSULIN | 19090229 | Insulin, Prompt Zinc, Beef-Pork |
| INSULIN | INSULIN | 19090242 | Insulin, Prompt Zinc, Beef-Pork |
| INSULIN | INSULIN | 19078634 | Insulin, Prompt Zinc, Beef-Pork |
| INSULIN | INSULIN | 19090243 | Insulin, Prompt Zinc, Beef-Pork |
| INSULIN | INSULIN | 19078635 | Insulin, Prompt Zinc, Beef-Pork |
| INSULIN | INSULIN | 36214706 | Insulin, Prompt Zinc, Beef-Pork |
| INSULIN | INSULIN | 40052099 | Insulin, Prompt Zinc, Beef-Pork |
| INSULIN | INSULIN | 19090247 | Insulin, Prompt Zinc, Human |
| INSULIN | INSULIN | 36214707 | Insulin, Prompt Zinc, Human |
| INSULIN | INSULIN | 40052361 | Insulin, Prompt Zinc, Human |
| INSULIN | INSULIN | 36214708 | Insulin, Prompt Zinc, Human |
| INSULIN | INSULIN | 40051333 | Insulin, Prompt Zinc, Human |
| INSULIN | INSULIN | 19110359 | Insulin, Prompt Zinc, Human |
| INSULIN | INSULIN | 19104432 | Insulin, Prompt Zinc, Human |
| INSULIN | INSULIN | 19090248 | Insulin, Prompt Zinc, Human |

| | | | |
|---|---|---|---|
| INSULIN | INSULIN | 1586371 | Insulin, Prompt Zinc, Human |
| INSULIN | INSULIN | 19090249 | Insulin, Prompt Zinc, Pork |
| INSULIN | INSULIN | 19095477 | Insulin, Prompt Zinc, Pork |
| INSULIN | INSULIN | 19078636 | Insulin, Prompt Zinc, Pork |
| INSULIN | INSULIN | 19110361 | Insulin, Prompt Zinc, Pork |
| INSULIN | INSULIN | 19104442 | Insulin, Prompt Zinc, Pork |
| INSULIN | INSULIN | 36214709 | Insulin, Prompt Zinc, Pork |
| INSULIN | INSULIN | 40052370 | Insulin, Prompt Zinc, Pork |
| INSULIN | INSULIN | 1513879 | Insulin, Protamine Lispro, Human |
| INSULIN | INSULIN | 1513880 | Insulin, Protamine Lispro, Human |
| INSULIN | INSULIN | 36232393 | insulin, protamine zinc, beef |
| INSULIN | INSULIN | 19090180 | insulin, protamine zinc, beef |
| INSULIN | INSULIN | 19094096 | insulin, protamine zinc, beef |
| INSULIN | INSULIN | 19116446 | insulin, protamine zinc, beef |
| INSULIN | INSULIN | 19090181 | insulin, protamine zinc, beef |
| INSULIN | INSULIN | 19009383 | insulin, protamine zinc, beef |
| INSULIN | INSULIN | 19094097 | insulin, protamine zinc, beef |
| INSULIN | INSULIN | 19061541 | insulin, protamine zinc, beef |
| INSULIN | INSULIN | 19094098 | insulin, protamine zinc, beef |
| INSULIN | INSULIN | 19061627 | insulin, protamine zinc, beef |
| INSULIN | INSULIN | 36224230 | insulin, protamine zinc, beef |
| INSULIN | INSULIN | 40052382 | insulin, protamine zinc, beef |
| INSULIN | INSULIN | 40052383 | insulin, protamine zinc, beef |
| INSULIN | INSULIN | 19013926 | Insulin, Protamine Zinc, Beef-Pork |
| INSULIN | INSULIN | 19094042 | Insulin, Protamine Zinc, Beef-Pork |
| INSULIN | INSULIN | 19078608 | Insulin, Protamine Zinc, Beef-Pork |
| INSULIN | INSULIN | 19094043 | Insulin, Protamine Zinc, Beef-Pork |
| INSULIN | INSULIN | 19080064 | Insulin, Protamine Zinc, Beef-Pork |
| INSULIN | INSULIN | 36214710 | Insulin, Protamine Zinc, Beef-Pork |
| INSULIN | INSULIN | 40052389 | Insulin, Protamine Zinc, Beef-Pork |
| INSULIN | INSULIN | 19091621 | Insulin, Protamine Zinc, Human |
| INSULIN | INSULIN | 19091623 | Insulin, Protamine Zinc, Human |
| INSULIN | INSULIN | 19091622 | Insulin, Protamine Zinc, Human |
| INSULIN | INSULIN | 40174828 | Insulin, Protamine Zinc, Human |
| INSULIN | INSULIN | 40174831 | Insulin, Protamine Zinc, Human |
| INSULIN | INSULIN | 40174829 | Insulin, Protamine Zinc, Human |
| INSULIN | INSULIN | 40174830 | Insulin, Protamine Zinc, Human |
| INSULIN | INSULIN | 36214711 | Insulin, Protamine Zinc, Human |
| INSULIN | INSULIN | 40052393 | Insulin, Protamine Zinc, Human |
| INSULIN | INSULIN | 40174832 | Insulin, Protamine Zinc, Human |

| | | | |
|---|---|---|---|
| INSULIN | INSULIN | 36230590 | Insulin, Protamine Zinc, Human |
| INSULIN | INSULIN | 19090187 | Insulin, Protamine Zinc, Pork |
| INSULIN | INSULIN | 19094099 | Insulin, Protamine Zinc, Pork |
| INSULIN | INSULIN | 19090188 | Insulin, Protamine Zinc, Pork |
| INSULIN | INSULIN | 19094100 | Insulin, Protamine Zinc, Pork |
| INSULIN | INSULIN | 19061545 | Insulin, Protamine Zinc, Pork |
| INSULIN | INSULIN | 19094143 | Insulin, Protamine Zinc, Pork |
| INSULIN | INSULIN | 19061546 | Insulin, Protamine Zinc, Pork |
| INSULIN | INSULIN | 36214712 | Insulin, Protamine Zinc, Pork |
| INSULIN | INSULIN | 40052401 | Insulin, Protamine Zinc, Pork |
| INSULIN | INSULIN | 36232392 | Insulin, Regular, Beef |
| INSULIN | INSULIN | 19013951 | Insulin, Regular, Beef |
| INSULIN | INSULIN | 19095217 | Insulin, Regular, Beef |
| INSULIN | INSULIN | 19116444 | Insulin, Regular, Beef |
| INSULIN | INSULIN | 19078631 | Insulin, Regular, Beef |
| INSULIN | INSULIN | 19009381 | Insulin, Regular, Beef |
| INSULIN | INSULIN | 19013956 | Insulin, Regular, Beef |
| INSULIN | INSULIN | 19061543 | Insulin, Regular, Beef |
| INSULIN | INSULIN | 19013957 | Insulin, Regular, Beef |
| INSULIN | INSULIN | 19061624 | Insulin, Regular, Beef |
| INSULIN | INSULIN | 36214713 | Insulin, Regular, Beef |
| INSULIN | INSULIN | 40052410 | Insulin, Regular, Beef |
| INSULIN | INSULIN | 40052411 | Insulin, Regular, Beef |
| INSULIN | INSULIN | 36221860 | insulin, regular, beef-pork |
| INSULIN | INSULIN | 1590165 | insulin, regular, beef-pork |
| INSULIN | INSULIN | 19094016 | insulin, regular, beef-pork |
| INSULIN | INSULIN | 19069665 | insulin, regular, beef-pork |
| INSULIN | INSULIN | 1590170 | insulin, regular, beef-pork |
| INSULIN | INSULIN | 19094020 | insulin, regular, beef-pork |
| INSULIN | INSULIN | 19110358 | insulin, regular, beef-pork |
| INSULIN | INSULIN | 19104451 | insulin, regular, beef-pork |
| INSULIN | INSULIN | 36222968 | insulin, regular, beef-pork |
| INSULIN | INSULIN | 40052422 | insulin, regular, beef-pork |
| INSULIN | INSULIN | 36222969 | insulin, regular, beef-pork |
| INSULIN | INSULIN | 40052423 | insulin, regular, beef-pork |
| INSULIN | INSULIN | 40052425 | insulin, regular, beef-pork |
| INSULIN | INSULIN | 36232396 | Insulin, Regular, Pork |
| INSULIN | INSULIN | 36221858 | Insulin, Regular, Pork |
| INSULIN | INSULIN | 19110376 | Insulin, Regular, Pork |
| INSULIN | INSULIN | 19104091 | Insulin, Regular, Pork |

| | | | |
|---|---|---|---|
| INSULIN | INSULIN | 19094017 | Insulin, Regular, Pork |
| INSULIN | INSULIN | 19117087 | Insulin, Regular, Pork |
| INSULIN | INSULIN | 19069640 | Insulin, Regular, Pork |
| INSULIN | INSULIN | 19116886 | Insulin, Regular, Pork |
| INSULIN | INSULIN | 19017198 | Insulin, Regular, Pork |
| INSULIN | INSULIN | 1586367 | Insulin, Regular, Pork |
| INSULIN | INSULIN | 19016047 | Insulin, Regular, Pork |
| INSULIN | INSULIN | 19095219 | Insulin, Regular, Pork |
| INSULIN | INSULIN | 19094124 | Insulin, Regular, Pork |
| INSULIN | INSULIN | 19100132 | Insulin, Regular, Pork |
| INSULIN | INSULIN | 19094045 | Insulin, Regular, Pork |
| INSULIN | INSULIN | 19069661 | Insulin, Regular, Pork |
| INSULIN | INSULIN | 19028937 | Insulin, Regular, Pork |
| INSULIN | INSULIN | 19098280 | Insulin, Regular, Pork |
| INSULIN | INSULIN | 19094125 | Insulin, Regular, Pork |
| INSULIN | INSULIN | 19061544 | Insulin, Regular, Pork |
| INSULIN | INSULIN | 36222970 | Insulin, Regular, Pork |
| INSULIN | INSULIN | 40052794 | Insulin, Regular, Pork |
| INSULIN | INSULIN | 40052795 | Insulin, Regular, Pork |
| INSULIN | INSULIN | 40052796 | Insulin, Regular, Pork |
| INSULIN | INSULIN | 40052798 | Insulin, Regular, Pork |
| INSULIN | INSULIN | 36242926 | Insulin, Regular, Pork |
| INSULIN | INSULIN | 36236039 | Insulin, Zinc, Human |
| INSULIN | INSULIN | 36236040 | Insulin, Zinc, Human |
| INSULIN | INSULIN | 19095254 | Insulin, Zinc, Human |
| INSULIN | INSULIN | 19116792 | Insulin, Zinc, Human |
| INSULIN | INSULIN | 19116729 | Insulin, Zinc, Human |
| INSULIN | INSULIN | 19062454 | Insulin, Zinc, Human |
| INSULIN | INSULIN | 19063031 | Insulin, Zinc, Human |
| INSULIN | INSULIN | 19012680 | Insulin, Zinc, Human |
| INSULIN | INSULIN | 19110360 | Insulin, Zinc, Human |
| INSULIN | INSULIN | 19095473 | Insulin, Zinc, Human |
| INSULIN | INSULIN | 1513851 | Insulin, Zinc, Human |
| INSULIN | INSULIN | 36222971 | Insulin, Zinc, Human |
| INSULIN | INSULIN | 40053113 | Insulin, Zinc, Human |
| INSULIN | INSULIN | 40053114 | Insulin, Zinc, Human |
| INSULIN | INSULIN | 40053115 | Insulin, Zinc, Human |
| INSULIN | INSULIN | 40053116 | Insulin, Zinc, Human |
| INSULIN | INSULIN | 40053117 | Insulin, Zinc, Human |
| INSULIN | INSULIN | 36221857 | Insulin, Zinc, Pork |

| | | | |
|---|---|---|---|
| INSULIN | INSULIN | 19095200 | Insulin, Zinc, Pork |
| INSULIN | INSULIN | 19069638 | Insulin, Zinc, Pork |
| INSULIN | INSULIN | 19116728 | Insulin, Zinc, Pork |
| INSULIN | INSULIN | 19078528 | Insulin, Zinc, Pork |
| INSULIN | INSULIN | 19012268 | Insulin, Zinc, Pork |
| INSULIN | INSULIN | 1562623 | Insulin, Zinc, Pork |
| INSULIN | INSULIN | 19072747 | Insulin, Zinc, Pork |
| INSULIN | INSULIN | 19061547 | Insulin, Zinc, Pork |
| INSULIN | INSULIN | 43525653 | Insulin, Zinc, Pork |
| INSULIN | INSULIN | 19095474 | Insulin, Zinc, Pork |
| INSULIN | INSULIN | 19061548 | Insulin, Zinc, Pork |
| INSULIN | INSULIN | 36222972 | Insulin, Zinc, Pork |
| INSULIN | INSULIN | 40053128 | Insulin, Zinc, Pork |
| INSULIN | INSULIN | 40053129 | Insulin, Zinc, Pork |
| INSULIN | INSULIN | 40053131 | Insulin, Zinc, Pork |
| INSULIN | INSULIN | 43525383 | Insulin, Zinc, Pork |
| INSULIN | INSULIN | 36232130 | Insulin, Zinc, Pork |
| INSULIN | INSULIN | 36241982 | Insulin, Zinc, Pork |
| INSULIN | INSULIN | 35604829 | insulin-like growth factor 2 |
| INSULIN | INSULIN | 36232391 | Lente Insulin, Beef |
| INSULIN | INSULIN | 19090204 | Lente Insulin, Beef |
| INSULIN | INSULIN | 19090208 | Lente Insulin, Beef |
| INSULIN | INSULIN | 19116445 | Lente Insulin, Beef |
| INSULIN | INSULIN | 19078601 | Lente Insulin, Beef |
| INSULIN | INSULIN | 19009382 | Lente Insulin, Beef |
| INSULIN | INSULIN | 19090209 | Lente Insulin, Beef |
| INSULIN | INSULIN | 19090206 | Lente Insulin, Beef |
| INSULIN | INSULIN | 36218599 | Lente Insulin, Beef |
| INSULIN | INSULIN | 40052808 | Lente Insulin, Beef |
| INSULIN | INSULIN | 40052809 | Lente Insulin, Beef |
| INSULIN | INSULIN | 36221856 | Lente Insulin, Beef-Pork |
| INSULIN | INSULIN | 36222091 | Lente Insulin, Beef-Pork |
| INSULIN | INSULIN | 1513843 | Lente Insulin, Beef-Pork |
| INSULIN | INSULIN | 1513848 | Lente Insulin, Beef-Pork |
| INSULIN | INSULIN | 19062511 | Lente Insulin, Beef-Pork |
| INSULIN | INSULIN | 19116727 | Lente Insulin, Beef-Pork |
| INSULIN | INSULIN | 19112801 | Lente Insulin, Beef-Pork |
| INSULIN | INSULIN | 19021048 | Lente Insulin, Beef-Pork |
| INSULIN | INSULIN | 1513845 | Lente Insulin, Beef-Pork |
| INSULIN | INSULIN | 19012267 | Lente Insulin, Beef-Pork |

| | | | |
|---|---|---|---|
| INSULIN | INSULIN | 19112802 | Lente Insulin, Beef-Pork |
| INSULIN | INSULIN | 36218600 | Lente Insulin, Beef-Pork |
| INSULIN | INSULIN | 40052820 | Lente Insulin, Beef-Pork |
| INSULIN | INSULIN | 40052821 | Lente Insulin, Beef-Pork |
| INSULIN | INSULIN | 40052822 | Lente Insulin, Beef-Pork |
| INSULIN | INSULIN | 40052823 | Lente Insulin, Beef-Pork |
| INSULIN | INSULIN | 36229167 | Lente Insulin, Beef-Pork |
| INSULIN | INSULIN | 40220515 | long-(ARG3)insulin-like growth factor-I |
| INSULIN | INSULIN | 45777024 | Regular Insulin, Human |
| INSULIN | INSULIN | 1718876 | Regular Insulin, Human |
| INSULIN | INSULIN | 36249546 | Regular Insulin, Human |
| INSULIN | INSULIN | 35605620 | Regular Insulin, Human |
| INSULIN | INSULIN | 36236034 | Regular Insulin, Human |
| INSULIN | INSULIN | 36236035 | Regular Insulin, Human |
| INSULIN | INSULIN | 36235842 | Regular Insulin, Human |
| INSULIN | INSULIN | 36241498 | Regular Insulin, Human |
| INSULIN | INSULIN | 19116760 | Regular Insulin, Human |
| INSULIN | INSULIN | 19116790 | Regular Insulin, Human |
| INSULIN | INSULIN | 19062458 | Regular Insulin, Human |
| INSULIN | INSULIN | 19116791 | Regular Insulin, Human |
| INSULIN | INSULIN | 19116726 | Regular Insulin, Human |
| INSULIN | INSULIN | 19007871 | Regular Insulin, Human |
| INSULIN | INSULIN | 19012507 | Regular Insulin, Human |
| INSULIN | INSULIN | 19012678 | Regular Insulin, Human |
| INSULIN | INSULIN | 19012266 | Regular Insulin, Human |
| INSULIN | INSULIN | 42903369 | Regular Insulin, Human |
| INSULIN | INSULIN | 46234243 | Regular Insulin, Human |
| INSULIN | INSULIN | 46234242 | Regular Insulin, Human |
| INSULIN | INSULIN | 19110353 | Regular Insulin, Human |
| INSULIN | INSULIN | 46233729 | Regular Insulin, Human |
| INSULIN | INSULIN | 45776621 | Regular Insulin, Human |
| INSULIN | INSULIN | 19111798 | Regular Insulin, Human |
| INSULIN | INSULIN | 19061542 | Regular Insulin, Human |
| INSULIN | INSULIN | 19062459 | Regular Insulin, Human |
| INSULIN | INSULIN | 35605623 | Regular Insulin, Human |
| INSULIN | INSULIN | 35605624 | Regular Insulin, Human |
| INSULIN | INSULIN | 46233731 | Regular Insulin, Human |
| INSULIN | INSULIN | 45774398 | Regular Insulin, Human |
| INSULIN | INSULIN | 19111796 | Regular Insulin, Human |
| INSULIN | INSULIN | 45776620 | Regular Insulin, Human |

| | | | |
|---|---|---|---|
| INSULIN | INSULIN | 45776624 | Regular Insulin, Human |
| INSULIN | INSULIN | 36247260 | Regular Insulin, Human |
| INSULIN | INSULIN | 40052768 | Regular Insulin, Human |
| INSULIN | INSULIN | 40052769 | Regular Insulin, Human |
| INSULIN | INSULIN | 40052770 | Regular Insulin, Human |
| INSULIN | INSULIN | 40052772 | Regular Insulin, Human |
| INSULIN | INSULIN | 40052774 | Regular Insulin, Human |
| INSULIN | INSULIN | 40052775 | Regular Insulin, Human |
| INSULIN | INSULIN | 35605619 | Regular Insulin, Human |
| INSULIN | INSULIN | 40159688 | Regular Insulin, Human |
| INSULIN | INSULIN | 40159689 | Regular Insulin, Human |
| INSULIN | INSULIN | 36242327 | Regular Insulin, Human |
| INSULIN | INSULIN | 19090226 | Ultralente Insulin, Beef |
| INSULIN | INSULIN | 19095476 | Ultralente Insulin, Beef |
| INSULIN | INSULIN | 19078639 | Ultralente Insulin, Beef |
| INSULIN | INSULIN | 36223264 | Ultralente Insulin, Beef |
| INSULIN | INSULIN | 40051323 | Ultralente Insulin, Beef |
| INSULIN | INSULIN | 19090221 | Ultralente Insulin, Beef-Pork |
| INSULIN | INSULIN | 19099012 | Ultralente Insulin, Beef-Pork |
| INSULIN | INSULIN | 19078637 | Ultralente Insulin, Beef-Pork |
| INSULIN | INSULIN | 19078638 | Ultralente Insulin, Beef-Pork |
| INSULIN | INSULIN | 36223265 | Ultralente Insulin, Beef-Pork |
| INSULIN | INSULIN | 40051331 | Ultralente Insulin, Beef-Pork |
| INSULIN | INSULIN | 36235844 | Ultralente Insulin, Human |
| INSULIN | INSULIN | 1586383 | Ultralente Insulin, Human |
| INSULIN | INSULIN | 19120831 | Ultralente Insulin, Human |
| INSULIN | INSULIN | 19116793 | Ultralente Insulin, Human |
| INSULIN | INSULIN | 19012701 | Ultralente Insulin, Human |
| INSULIN | INSULIN | 36225587 | Ultralente Insulin, Human |
| INSULIN | INSULIN | 40051340 | Ultralente Insulin, Human |
| INSULIN | INSULIN | 40051342 | Ultralente Insulin, Human |
| LINAGLIPTIN | LINAGLIPTIN | 36249791 | Linagliptin |
| LINAGLIPTIN | LINAGLIPTIN | 36249796 | Linagliptin |
| LINAGLIPTIN | LINAGLIPTIN | 36249790 | Linagliptin |
| LINAGLIPTIN | LINAGLIPTIN | 36249792 | Linagliptin |
| LINAGLIPTIN | LINAGLIPTIN | 42705857 | Linagliptin |
| LINAGLIPTIN | LINAGLIPTIN | 42705866 | Linagliptin |
| LINAGLIPTIN | LINAGLIPTIN | 36238285 | Linagliptin |
| LINAGLIPTIN | LINAGLIPTIN | 36249794 | Linagliptin |
| LINAGLIPTIN | LINAGLIPTIN | 36249795 | Linagliptin |

| | | | |
|---|---|---|---|
| LINAGLIPTIN | LINAGLIPTIN | 42708086 | Linagliptin |
| LINAGLIPTIN | LINAGLIPTIN | 36249797 | Linagliptin |
| LINAGLIPTIN | LINAGLIPTIN | 36249799 | Linagliptin |
| LINAGLIPTIN | LINAGLIPTIN | 36249800 | Linagliptin |
| LIRAGLUTIDE | LIRAGLUTIDE | 1718703 | liraglutide |
| LIRAGLUTIDE | LIRAGLUTIDE | 42902992 | liraglutide |
| LIRAGLUTIDE | LIRAGLUTIDE | 45892687 | liraglutide |
| LIRAGLUTIDE | LIRAGLUTIDE | 42903382 | liraglutide |
| LIRAGLUTIDE | LIRAGLUTIDE | 46234110 | liraglutide |
| LIRAGLUTIDE | LIRAGLUTIDE | 46234112 | liraglutide |
| LIRAGLUTIDE | LIRAGLUTIDE | 46234111 | liraglutide |
| LIXISENATIDE | LIXISENATIDE | 40220770 | Lixisenatide |
| LIXISENATIDE | LIXISENATIDE | 40221118 | Lixisenatide |
| LIXISENATIDE | LIXISENATIDE | 40220858 | Lixisenatide |
| LIXISENATIDE | LIXISENATIDE | 40220859 | Lixisenatide |
| LIXISENATIDE | LIXISENATIDE | 40220860 | Lixisenatide |
| LIXISENATIDE | LIXISENATIDE | 40220862 | Lixisenatide |
| LIXISENATIDE | LIXISENATIDE | 40220856 | Lixisenatide |
| LIXISENATIDE | LIXISENATIDE | 1718596 | Lixisenatide |
| LIXISENATIDE | LIXISENATIDE | 44506520 | Lixisenatide |
| LIXISENATIDE | LIXISENATIDE | 40220854 | Lixisenatide |
| LIXISENATIDE | LIXISENATIDE | 40220852 | Lixisenatide |
| LIXISENATIDE | LIXISENATIDE | 40220857 | Lixisenatide |
| LIXISENATIDE | LIXISENATIDE | 44506518 | Lixisenatide |
| LIXISENATIDE | LIXISENATIDE | 40220861 | Lixisenatide |
| LIXISENATIDE | LIXISENATIDE | 40220863 | Lixisenatide |
| LIXISENATIDE | LIXISENATIDE | 40220864 | Lixisenatide |
| LIXISENATIDE | LIXISENATIDE | 36247618 | Lixisenatide |
| LIXISENATIDE | LIXISENATIDE | 40220851 | Lixisenatide |
| LIXISENATIDE | LIXISENATIDE | 40220855 | Lixisenatide |
| METFORMIN | METFORMIN | 19107329 | Metformin |
| METFORMIN | METFORMIN | 40171448 | Metformin |
| METFORMIN | METFORMIN | 40229045 | Metformin |
| METFORMIN | METFORMIN | 42708166 | Metformin |
| METFORMIN | METFORMIN | 42708170 | Metformin |
| METFORMIN | METFORMIN | 40229049 | Metformin |
| METFORMIN | METFORMIN | 42708174 | Metformin |
| METFORMIN | METFORMIN | 793297 | Metformin |
| METFORMIN | METFORMIN | 793299 | Metformin |
| METFORMIN | METFORMIN | 793303 | Metformin |

| | | | |
|---|---|---|---|
| METFORMIN | METFORMIN | 793298 | Metformin |
| METFORMIN | METFORMIN | 793302 | Metformin |
| METFORMIN | METFORMIN | 793300 | Metformin |
| METFORMIN | METFORMIN | 793306 | Metformin |
| METFORMIN | METFORMIN | 793308 | Metformin |
| METFORMIN | METFORMIN | 793307 | Metformin |
| METFORMIN | METFORMIN | 793309 | Metformin |
| METFORMIN | METFORMIN | 793312 | Metformin |
| METFORMIN | METFORMIN | 793311 | Metformin |
| METFORMIN | METFORMIN | 793313 | Metformin |
| METFORMIN | METFORMIN | 793315 | Metformin |
| METFORMIN | METFORMIN | 793314 | Metformin |
| METFORMIN | METFORMIN | 793316 | Metformin |
| METFORMIN | METFORMIN | 36235744 | Metformin |
| METFORMIN | METFORMIN | 36235745 | Metformin |
| METFORMIN | METFORMIN | 40173226 | Metformin |
| METFORMIN | METFORMIN | 36223131 | Metformin |
| METFORMIN | METFORMIN | 36223132 | Metformin |
| METFORMIN | METFORMIN | 36223133 | Metformin |
| METFORMIN | METFORMIN | 40153147 | Metformin |
| METFORMIN | METFORMIN | 40153892 | Metformin |
| METFORMIN | METFORMIN | 36223135 | Metformin |
| METFORMIN | METFORMIN | 40117725 | Metformin |
| METFORMIN | METFORMIN | 36223136 | Metformin |
| METFORMIN | METFORMIN | 40063306 | Metformin |
| METFORMIN | METFORMIN | 40231387 | Metformin |
| METFORMIN | METFORMIN | 36223139 | Metformin |
| METFORMIN | METFORMIN | 36223140 | Metformin |
| METFORMIN | METFORMIN | 42705988 | Metformin |
| METFORMIN | METFORMIN | 36223141 | Metformin |
| METFORMIN | METFORMIN | 40139098 | Metformin |
| METFORMIN | METFORMIN | 36223142 | Metformin |
| METFORMIN | METFORMIN | 40173236 | Metformin |
| METFORMIN | METFORMIN | 40173234 | Metformin |
| METFORMIN | METFORMIN | 40173235 | Metformin |
| METFORMIN | METFORMIN | 40173238 | Metformin |
| METFORMIN | METFORMIN | 40164888 | Metformin |
| METFORMIN | METFORMIN | 40231394 | Metformin |
| METFORMIN | METFORMIN | 42708168 | Metformin |
| METFORMIN | METFORMIN | 42708172 | Metformin |

| | | | |
|---|---|---|---|
| METFORMIN | METFORMIN | 40164910 | Metformin |
| METFORMIN | METFORMIN | 40164913 | Metformin |
| METFORMIN | METFORMIN | 40164916 | Metformin |
| METFORMIN | METFORMIN | 40231403 | Metformin |
| METFORMIN | METFORMIN | 42708176 | Metformin |
| METFORMIN | METFORMIN | 40167189 | Metformin |
| METFORMIN | METFORMIN | 40167190 | Metformin |
| METFORMIN | METFORMIN | 19006932 | Metformin |
| METFORMIN | METFORMIN | 19027257 | Metformin |
| METFORMIN | METFORMIN | 40164934 | Metformin |
| METFORMIN | METFORMIN | 40164937 | Metformin |
| METFORMIN | METFORMIN | 40167191 | Metformin |
| METFORMIN | METFORMIN | 40167193 | Metformin |
| METFORMIN | METFORMIN | 19006933 | Metformin |
| METFORMIN | METFORMIN | 19027259 | Metformin |
| METFORMIN | METFORMIN | 36223144 | Metformin |
| METFORMIN | METFORMIN | 40063355 | Metformin |
| METFORMIN | METFORMIN | 40063357 | Metformin |
| METFORMIN | METFORMIN | 40220371 | Metformin |
| METFORMIN | METFORMIN | 40220375 | Metformin |
| METFORMIN | METFORMIN | 36239753 | Metformin |
| METFORMIN | METFORMIN | 36239754 | Metformin |
| METFORMIN | METFORMIN | 40220373 | Metformin |
| METFORMIN | METFORMIN | 40220376 | Metformin |
| METFORMIN | METFORMIN | 36240428 | Metformin |
| METFORMIN | METFORMIN | 793304 | Metformin |
| METFORMIN | METFORMIN | 793305 | Metformin |
| MIGLITOL | MIGLITOL | 36215711 | miglitol |
| NATEGLINIDE | NATEGLINIDE | 1502873 | nateglinide |
| PIOGLITAZONE | PIOGLITAZONE | 36220120 | pioglitazone |
| PRAMLINTIDE | PRAMLINTIDE | 40163889 | Pramlintide |
| PRAMLINTIDE | PRAMLINTIDE | 40163898 | Pramlintide |
| PRAMLINTIDE | PRAMLINTIDE | 40165689 | Pramlintide |
| PRAMLINTIDE | PRAMLINTIDE | 42903071 | Pramlintide |
| PRAMLINTIDE | PRAMLINTIDE | 42902983 | Pramlintide |
| PRAMLINTIDE | PRAMLINTIDE | 36223440 | Pramlintide |
| PRAMLINTIDE | PRAMLINTIDE | 46276271 | Pramlintide |
| REPAGLINIDE | REPAGLINIDE | 36231452 | repaglinide |
| REPAGLINIDE | REPAGLINIDE | 36231453 | repaglinide |
| REPAGLINIDE | REPAGLINIDE | 19120528 | repaglinide |

| | | | |
|---|---|---|---|
| REPAGLINIDE | REPAGLINIDE | 19054443 | repaglinide |
| REPAGLINIDE | REPAGLINIDE | 19120530 | repaglinide |
| REPAGLINIDE | REPAGLINIDE | 40098706 | repaglinide |
| ROSIGLITAZONE | ROSIGLITAZONE | 36227625 | rosiglitazone |
| SAXAGLIPTIN | SAXAGLIPTIN | 36224108 | saxagliptin |
| SEMAGLUTIDE | SEMAGLUTIDE | 793147 | semaglutide |
| SEMAGLUTIDE | SEMAGLUTIDE | 793152 | semaglutide |
| SEMAGLUTIDE | SEMAGLUTIDE | 793153 | semaglutide |
| SEMAGLUTIDE | SEMAGLUTIDE | 793154 | semaglutide |
| SEMAGLUTIDE | SEMAGLUTIDE | 793151 | semaglutide |
| SEMAGLUTIDE | SEMAGLUTIDE | 793143 | semaglutide |
| SEMAGLUTIDE | SEMAGLUTIDE | 793144 | semaglutide |
| SEMAGLUTIDE | SEMAGLUTIDE | 793149 | semaglutide |
| SEMAGLUTIDE | SEMAGLUTIDE | 793145 | semaglutide |
| SEMAGLUTIDE | SEMAGLUTIDE | 793146 | semaglutide |
| SEMAGLUTIDE | SEMAGLUTIDE | 793150 | semaglutide |
| SITAGLIPTIN | SITAGLIPTIN | 793332 | sitagliptin |
| SITAGLIPTIN | SITAGLIPTIN | 793334 | sitagliptin |
| SITAGLIPTIN | SITAGLIPTIN | 793338 | sitagliptin |
| SITAGLIPTIN | SITAGLIPTIN | 793333 | sitagliptin |
| SITAGLIPTIN | SITAGLIPTIN | 793337 | sitagliptin |
| SITAGLIPTIN | SITAGLIPTIN | 793335 | sitagliptin |
| SITAGLIPTIN | SITAGLIPTIN | 793341 | sitagliptin |
| SITAGLIPTIN | SITAGLIPTIN | 793343 | sitagliptin |
| SITAGLIPTIN | SITAGLIPTIN | 793342 | sitagliptin |
| SITAGLIPTIN | SITAGLIPTIN | 793344 | sitagliptin |
| SITAGLIPTIN | SITAGLIPTIN | 36240264 | sitagliptin |
| SITAGLIPTIN | SITAGLIPTIN | 36240261 | sitagliptin |
| SITAGLIPTIN | SITAGLIPTIN | 40243622 | sitagliptin |
| SITAGLIPTIN | SITAGLIPTIN | 40243624 | sitagliptin |
| SITAGLIPTIN | SITAGLIPTIN | 36240262 | sitagliptin |
| SITAGLIPTIN | SITAGLIPTIN | 40243626 | sitagliptin |
| SITAGLIPTIN | SITAGLIPTIN | 42901618 | sitagliptin |
| SITAGLIPTIN | SITAGLIPTIN | 42901619 | sitagliptin |
| SITAGLIPTIN | SITAGLIPTIN | 40243631 | sitagliptin |
| SITAGLIPTIN | SITAGLIPTIN | 42900899 | sitagliptin |
| SITAGLIPTIN | SITAGLIPTIN | 42901621 | sitagliptin |
| SITAGLIPTIN | SITAGLIPTIN | 40243634 | sitagliptin |
| SITAGLIPTIN | SITAGLIPTIN | 40243632 | sitagliptin |
| SITAGLIPTIN | SITAGLIPTIN | 42901622 | sitagliptin |

| | | | |
|---|---|---|---|
| SITAGLIPTIN | SITAGLIPTIN | 42901623 | sitagliptin |
| SITAGLIPTIN | SITAGLIPTIN | 36224403 | sitagliptin |
| SITAGLIPTIN | SITAGLIPTIN | 793339 | sitagliptin |
| SITAGLIPTIN | SITAGLIPTIN | 793340 | sitagliptin |
| TOLAZAMIDE | TOLAZAMIDE | 40089089 | Tolazamide |
| TOLAZAMIDE | TOLAZAMIDE | 40089090 | Tolazamide |
| TOLAZAMIDE | TOLAZAMIDE | 36214132 | Tolazamide |
| TOLAZAMIDE | TOLAZAMIDE | 36230841 | Tolazamide |
| TOLBUTAMIDE | TOLBUTAMIDE | 36242644 | Tolbutamide |
| TOLBUTAMIDE | TOLBUTAMIDE | 36242650 | Tolbutamide |
| TOLBUTAMIDE | TOLBUTAMIDE | 1502859 | Tolbutamide |
| TOLBUTAMIDE | TOLBUTAMIDE | 1502915 | Tolbutamide |
| TOLBUTAMIDE | TOLBUTAMIDE | 19075975 | Tolbutamide |
| TOLBUTAMIDE | TOLBUTAMIDE | 1502861 | Tolbutamide |
| TOLBUTAMIDE | TOLBUTAMIDE | 19040829 | Tolbutamide |
| TOLBUTAMIDE | TOLBUTAMIDE | 19034783 | Tolbutamide |
| TOLBUTAMIDE | TOLBUTAMIDE | 1502916 | Tolbutamide |
| TOLBUTAMIDE | TOLBUTAMIDE | 36214385 | Tolbutamide |
| TOLBUTAMIDE | TOLBUTAMIDE | 40089113 | Tolbutamide |
| TOLBUTAMIDE | TOLBUTAMIDE | 40089114 | Tolbutamide |
| TOLBUTAMIDE | TOLBUTAMIDE | 40089116 | Tolbutamide |
| TOLBUTAMIDE | TOLBUTAMIDE | 40089117 | Tolbutamide |
| TOLBUTAMIDE | TOLBUTAMIDE | 36214387 | Tolbutamide |
| TOLBUTAMIDE | TOLBUTAMIDE | 36229273 | Tolbutamide |
| TOLBUTAMIDE | TOLBUTAMIDE | 36229274 | Tolbutamide |

| Drug Name | Drug Route |
| --- | --- |
| Regular Insulin, Human | SUBCUT |
| Metformin 500 MG | |
| Regular Insulin, Human | IV |
| Insulin Lispro | SUBCUT |
| Insulin, Aspart, Human | SUBCUT |
| Metformin hydrochloride 500 MG Oral Tablet | by mouth |
| Metformin hydrochloride 1000 MG Oral Tablet [Glucophage] | by mouth |
| Insulin Glargine | SUBCUT |
| Metformin hydrochloride 500 MG Oral Tablet [Glucophage] | by mouth |
| Metformin 1000 MG | |
| Insulin Glargine 100 UNT/ML Injectable Solution [Lantus] | subcutaneously |
| Insulin, Aspart, Human | |
| Insulin Glargine 100 UNT/ML Injectable Solution [Lantus] | subcutaneous |
| Insulin, Aspart, Human 100 UNT/ML Cartridge [NovoLog] | |
| Metformin hydrochloride 1000 MG Oral Tablet | by mouth |
| Regular Insulin, Human | subcut |
| Metformin Oral Tablet [Glucophage] | by mouth |
| Metformin hydrochloride 500 MG Oral Tablet | po |
| Insulin, Aspart, Human | subcut |
| Insulin, Aspart, Human 100 UNT/ML Injectable Solution [NovoLog] | subcutaneously |
| Insulin Glargine Injectable Solution | |
| glimepiride 4 MG Oral Tablet [Amaryl] | by mouth |
| Metformin hydrochloride 500 MG [Glucophage] | |
| Insulin Glargine 100 UNT/ML [Lantus] | |
| Lantus Injectable Product | subcut |
| sitagliptin 100 MG Oral Tablet [Januvia] | by mouth |
| Glipizide 5 MG | |
| Glipizide 10 MG | |
| glimepiride 2 MG Oral Tablet [Amaryl] | by mouth |
| Metformin | |
| Glyburide 5 MG | |
| Lantus Injectable Product | SUBCUT |
| Metformin hydrochloride 1000 MG Oral Tablet | po |
| Insulin Glargine 100 UNT/ML Injectable Solution [Lantus] | Subcutaneous |
| Glyburide 5 MG Oral Tablet | by mouth |
| Insulin Lispro 100 UNT/ML Injectable Solution | subcutaneous |
| Metformin Extended Release Oral Tablet [Fortamet] | |

| | |
|---|---|
| glimepiride 4 MG | |
| Insulin Lispro | |
| Metformin hydrochloride 1000 MG [Glucophage] | |
| Glipizide 5 MG Oral Tablet [Glucotrol] | by mouth |
| Glipizide 10 MG Oral Tablet [Glucotrol] | by mouth |
| Insulin Glargine Injectable Solution | SUBCUT |
| Insulin Glargine | |
| 3 ML insulin detemir 100 UNT/ML Pen Injector [Levemir] | subcutaneous |
| Regular Insulin, Human | INFUSION |
| Metformin hydrochloride 500 MG Oral Tablet | |
| Metformin Oral Solution | by mouth |
| Glipizide 5 MG Oral Tablet | by mouth |
| Metformin hydrochloride 850 MG Oral Tablet [Glucophage] | by mouth |
| glimepiride 4 MG Oral Tablet | by mouth |
| sitagliptin 100 MG [Januvia] | |
| Glipizide 10 MG Oral Tablet | by mouth |
| Metformin hydrochloride 500 MG Oral Tablet [Glucophage] | po |
| glimepiride 2 MG | |
| insulin pork, isophane 100 UNT/ML Injectable Suspension [NPH Iletin II] | |
| Glyburide 5 MG Oral Tablet | po |
| Lantus Injectable Product | subcutaneously |
| glimepiride 1 MG Oral Tablet [Amaryl] | by mouth |
| glimepiride 4 MG [Amaryl] | |
| Regular Insulin, Human | |
| Metformin hydrochloride 500 MG Oral Tablet | oral |
| Metformin hydrochloride 750 MG | |
| Glyburide | |
| pioglitazone 30 MG Oral Tablet [Actos] | by mouth |
| 3 ML liraglutide 6 MG/ML Pen Injector [Victoza] | subcutaneous |
| insulin human, isophane 70 UNT/ML / Regular Insulin, Human 30 UNT/ML [Humulin] | |
| glimepiride 2 MG Oral Tablet | by mouth |
| pioglitazone 45 MG Oral Tablet [Actos] | by mouth |
| Insulin Glargine 100 UNT/ML Injectable Solution [Lantus] | SUBCUT |
| Metformin hydrochloride 500 MG Oral Tablet | orally |
| Metformin hydrochloride 500 MG Oral Tablet | PO |
| Glipizide 10 MG Oral Tablet | po |
| Insulin Lispro 100 UNT/ML Injectable Solution [Humalog] | SUBCUT |
| Metformin hydrochloride 850 MG | |

| | |
|---|---|
| insulin detemir 100 UNT/ML Injectable Solution [Levemir] | subcutaneously |
| Glipizide 2.5 MG | |
| Glyburide 2.5 MG | |
| Glipizide 5 MG Oral Tablet | po |
| Glucophage Oral Product | by mouth |
| Glipizide 10 MG [Glucotrol] | |
| Insulin Glargine 100 UNT/ML Injectable Solution [Lantus] | SUBCUTANEOUS |
| pioglitazone 30 MG [Actos] | |
| Glipizide 5 MG [Glucotrol] | |
| Metformin hydrochloride 500 MG Oral Tablet | ORAL |
| 3 ML Insulin Glargine 100 UNT/ML Pen Injector [Lantus] | SUBCUTANEOUS |
| Regular Insulin, Human | iv |
| Insulin Lispro Injectable Solution | subcut |
| glimepiride 2 MG [Amaryl] | |
| pioglitazone 45 MG [Actos] | |
| Metformin hydrochloride 1000 MG Oral Tablet | oral |
| Metformin hydrochloride 850 MG Oral Tablet | by mouth |
| Glipizide Oral Tablet [Glucotrol] | by mouth |
| Metformin hydrochloride 1000 MG Oral Tablet [Glucophage] | po |
| Metformin hydrochloride 500 MG / sitagliptin 50 MG Oral Tablet [Janumet] | by mouth |
| sitagliptin 50 MG Oral Tablet [Januvia] | by mouth |
| glimepiride 4 MG Oral Tablet [Amaryl] | po |
| rosiglitazone 4 MG [Avandia] | |
| insulin human, isophane 70 UNT/ML / Regular Insulin, Human 30 UNT/ML [Novolin] | |
| glimepiride 1 MG | |
| Regular Insulin, Human 100 UNT/ML Injectable Solution [Humulin R] | |
| Regular Insulin, Human | SQ |
| 24 HR Metformin hydrochloride 500 MG Extended Release Oral Tablet [Glucophage] | ORAL |
| Glyburide 5 MG Oral Tablet [Diabeta] | by mouth |
| Metformin hydrochloride 1000 MG Oral Tablet | orally |
| pioglitazone 30 MG Oral Tablet [Actos] | po |
| Metformin hydrochloride 1000 MG Oral Tablet [Glucophage] | ORAL |
| Insulin, Aspart, Human 100 UNT/ML Injectable Solution [NovoLog] | SUBCUT |
| Insulin, Aspart, Human 100 UNT/ML Injectable Solution [NovoLog] | SUBCUTANEOUS |

| | |
|---|---|
| pioglitazone 15 MG Oral Tablet [Actos] | by mouth |
| 3 ML Insulin, Aspart, Human 100 UNT/ML Pen Injector [NovoLog] | SUBCUTANEOUS |
| sitagliptin 100 MG Oral Tablet [Januvia] | po |
| rosiglitazone 4 MG Oral Tablet [Avandia] | po |
| nateglinide 120 MG Oral Tablet [Starlix] | by mouth |
| 24 HR Metformin hydrochloride 500 MG Extended Release Oral Tablet [Glucophage] | |
| NovoLog Injectable Product | subcutaneously |
| glimepiride 4 MG Oral Tablet | po |
| Glyburide 2.5 MG Oral Tablet | by mouth |
| 60 ACTUAT exenatide 0.005 MG/ACTUAT Pen Injector [Byetta] | subcutaneously |
| pioglitazone 45 MG Oral Tablet [Actos] | po |
| Regular Insulin, Human 100 UNT/ML Injectable Solution [Novolin R] | |
| pioglitazone 15 MG [Actos] | |
| glimepiride Oral Product | by mouth |
| Amaryl Oral Product | by mouth |
| Metformin hydrochloride 850 MG Oral Tablet | po |
| Regular Insulin, Human 100 UNT/ML Injectable Solution [Humulin R] | subcutaneously |
| Metformin hydrochloride 1000 MG Oral Tablet | PO |
| Glipizide | |
| glimepiride 1 MG Oral Tablet | by mouth |
| Metformin Extended Release Oral Tablet [Glucophage] | |
| Metformin hydrochloride 850 MG [Glucophage] | |
| Insulin, Aspart Protamine, Human 70 UNT/ML / Insulin, Aspart, Human 30 UNT/ML [NovoLog Mix] | |
| Insulin, Aspart, Human | Subcutaneous |
| rosiglitazone 4 MG Oral Tablet [Avandia] | by mouth |
| exenatide 0.01 MG/ACTUAT [Byetta] | |
| Metformin hydrochloride 1000 MG Oral Tablet | |
| Linagliptin 5 MG Oral Tablet [Tradjenta] | by mouth |
| Glyburide Oral Tablet [Diabeta] | by mouth |
| glimepiride 1 MG [Amaryl] | |
| Regular Insulin, Human 100 UNT/ML Injectable Solution [Novolin R] | subcutaneously |
| Insulin, Aspart Protamine, Human 70 UNT/ML / Insulin, Aspart, Human 30 UNT/ML Injectable Suspension [NovoLog Mix] | subcutaneous |
| Metformin Oral Solution | po |
| glimepiride 2 MG Oral Tablet [Amaryl] | po |

| | |
|---|---|
| nateglinide 120 MG [Starlix] | |
| glimepiride | |
| Lantus Injectable Product | |
| Insulin Lispro Cartridge [Humalog] | |
| Glyburide Oral Tablet | po |
| 3 ML insulin detemir 100 UNT/ML Pen Injector [Levemir] | |
| insulin human, isophane 100 UNT/ML Injectable Suspension [Novolin N] | subcutaneous |
| Insulin Lispro 100 UNT/ML Injectable Solution [Humalog] | SUBCUTANEOUS |
| Metformin hydrochloride 500 MG Oral Tablet [Glucophage] | PO |
| sitagliptin 50 MG [Januvia] | |
| Glyburide 2.5 MG Oral Tablet | po |
| glimepiride 2 MG Oral Tablet | po |
| pioglitazone 15 MG Oral Tablet [Actos] | po |
| Insulin, Aspart Protamine, Human 70 UNT/ML / Insulin, Aspart, Human 30 UNT/ML Injectable Suspension [NovoLog Mix] | |
| rosiglitazone 8 MG [Avandia] | |
| Glipizide Oral Product | by mouth |
| 3 ML Insulin, Aspart Protamine, Human 70 UNT/ML / Insulin, Aspart, Human 30 UNT/ML Pen Injector [NovoLog Mix] | |
| Insulin Glargine Injectable Solution [Lantus] | subcutaneously |
| Insulin, Aspart, Human 100 UNT/ML Cartridge [NovoLog] | sq |
| Regular Insulin, Human | sq |
| Glyburide 2.5 MG Oral Tablet [Diabeta] | by mouth |
| rosiglitazone 8 MG Oral Tablet [Avandia] | po |
| Glipizide 10 MG Oral Tablet [Glucotrol] | po |
| 24 HR Metformin hydrochloride 750 MG Extended Release Oral Tablet | by mouth |
| Glyburide 6 MG Oral Tablet [Glynase] | by mouth |
| insulin human, isophane 100 UNT/ML Injectable Suspension [Humulin N] | subcutaneous |
| Metformin hydrochloride 1000 MG / sitagliptin 50 MG Oral Tablet [Janumet] | by mouth |
| 24 HR Metformin hydrochloride 500 MG Extended Release Oral Tablet [Glucophage] | oral |
| Metformin hydrochloride 500 MG Extended Release Oral Tablet | |
| Humalog Injectable Product | subcutaneously |
| Glyburide 5 MG Oral Tablet | PO |
| insulin human, isophane 100 UNT/ML Injectable Suspension [Humulin N] | subcutaneously |

| | |
|---|---|
| liraglutide 6 MG/ML [Victoza] | subcutaneously |
| 3 ML Insulin Lispro 100 UNT/ML Pen Injector [Humalog] | SUBCUTANEOUS |
| Glucotrol Oral Product | by mouth |
| Insulin Glargine 100 UNT/ML Injectable Solution [Lantus] | subq |
| glimepiride 4 MG Oral Tablet [Amaryl] | ORAL |
| Insulin, Aspart Protamine, Human 70 UNT/ML / Insulin, Aspart, Human 30 UNT/ML Injectable Suspension [NovoLog Mix] | subcutaneously |
| nateglinide 120 MG Oral Tablet [Starlix] | po |
| Glyburide Oral Tablet | by mouth |
| Glipizide 2.5 MG Oral Tablet | by mouth |
| Victoza Injectable Product | subcutaneously |
| Regular Insulin, Human 500 UNT/ML Injectable Solution [Humulin R] | subcutaneous |
| insulin human, isophane 70 UNT/ML / Regular Insulin, Human 30 UNT/ML Injectable Suspension [Humulin] | subcutaneously |
| Glipizide Oral Product | Oral |
| Metformin hydrochloride 500 MG Oral Tablet | |
| Insulin Lispro Cartridge [Humalog] | subcut |
| saxagliptin 5 MG Oral Tablet [Onglyza] | by mouth |
| insulin detemir 100 UNT/ML [Levemir] | |
| 60 ACTUAT exenatide 0.01 MG/ACTUAT Pen Injector [Byetta] | |
| Levemir Injectable Product | subcutaneously |
| Glipizide 5 MG Oral Tablet [Glucotrol] | po |
| insulin human, isophane 70 UNT/ML / Regular Insulin, Human 30 UNT/ML Injectable Suspension [Humulin] | subcutaneous |
| Insulin Lispro 50 UNT/ML / Insulin, Protamine Lispro, Human 50 UNT/ML Injectable Suspension [Humalog Mix] | |
| Glipizide 5 MG Oral Tablet | orally |
| 3 ML Insulin Glargine 100 UNT/ML Pen Injector [Lantus] | |
| Insulin Lispro 25 UNT/ML / Insulin, Protamine Lispro, Human 75 UNT/ML [Humalog Mix] | |
| insulin human, isophane 70 UNT/ML / Regular Insulin, Human 30 UNT/ML Injectable Suspension [Novolin] | subcutaneous |
| Linagliptin 5 MG [Tradjenta] | |
| Insulin Lispro 25 UNT/ML / Insulin, Protamine Lispro, Human 75 UNT/ML Injectable Suspension [Humalog Mix] | subcutaneous |
| Regular Insulin, Human 100 UNT/ML Injectable Solution [Novolin R] | SUBCUT |
| canagliflozin 300 MG Oral Tablet [Invokana] | by mouth |
| dapagliflozin 10 MG Oral Tablet [Farxiga] | by mouth |

| | |
|---|---|
| exenatide 0.005 MG/ACTUAT [Byetta] | |
| 24 HR Metformin hydrochloride 500 MG Extended Release Oral Tablet [Glucophage] | |
| glimepiride 4 MG Oral Tablet [Amaryl] | PO |
| insulin human, isophane 100 UNT/ML Injectable Suspension [Novolin N] | subcutaneously |
| rosiglitazone 8 MG Oral Tablet [Avandia] | by mouth |
| Insulin Glargine | subcutaneous |
| Glipizide 10 MG Oral Tablet | orally |
| Insulin Glargine 100 UNT/ML Injectable Solution [Lantus] | SubQ |
| Metformin hydrochloride 1000 MG Oral Tablet [Glucophage] | PO |
| 1.5 ML Insulin Glargine 300 UNT/ML Pen Injector [Toujeo] | subcutaneous |
| insulin human, isophane 70 UNT/ML / Regular Insulin, Human 30 UNT/ML Injectable Suspension [Novolin] | subcutaneously |
| glimepiride 4 MG Oral Tablet | orally |
| Metformin hydrochloride 850 MG Oral Tablet [Glucophage] | po |
| Glyburide 1.25 MG | |
| Regular Insulin, Human 500 UNT/ML Injectable Solution [Humulin R] | |
| Metformin hydrochloride 1000 MG / sitagliptin 50 MG [Janumet] | |
| Metformin Extended Release Oral Tablet [Glumetza] | |
| Glipizide 5 MG Oral Tablet | PO |
| Glipizide 10 MG Oral Tablet | PO |
| 24 HR Metformin hydrochloride 500 MG Extended Release Oral Tablet | by mouth |
| nateglinide 60 MG Oral Tablet [Starlix] | by mouth |
| liraglutide 6 MG/ML [Victoza] | |
| Glyburide / Metformin Oral Tablet [Glucovance] | by mouth |
| Metformin hydrochloride 500 MG Oral Tablet [Glucophage] | |
| Glyburide 5 MG [Diabeta] | |
| Metformin Oral Solution | oral |
| Insulin, Aspart, Human | npo |
| Insulin, Glulisine, Human 100 UNT/ML Injectable Solution [Apidra] | |
| glimepiride 2 MG Oral Tablet [Amaryl] | ORAL |
| canagliflozin 100 MG Oral Tablet [Invokana] | by mouth |
| glimepiride Oral Tablet [Amaryl] | by mouth |
| Insulin Lispro | subcut |
| Glipizide 5 MG Oral Tablet | ORAL |
| Insulin, Aspart, Human | subcutaneous |

| | |
|---|---|
| canagliflozin 300 MG [Invokana] | |
| Insulin Lispro 25 UNT/ML / Insulin, Protamine Lispro, Human 75 UNT/ML Injectable Suspension [Humalog Mix] | |
| Regular Insulin, Human 100 UNT/ML Injectable Solution [Humulin R] | IV |
| Glucophage Oral Product | po |
| exenatide 0.01 MG/ACTUAT [Byetta] | subcut |
| insulin human, isophane 100 UNT/ML Injectable Suspension [Humulin N] | |
| Glipizide 2.5 MG [Glucotrol] | |
| Actoplus Met Oral Product | by mouth |
| Insulin Lispro 25 UNT/ML / Insulin, Protamine Lispro, Human 75 UNT/ML Injectable Suspension [Humalog Mix] | subcutaneously |
| Glyburide 5 MG Oral Tablet | ORAL |
| Glipizide 5 MG Oral Tablet | oral |
| Glipizide 2.5 MG Oral Tablet | po |
| glimepiride 1 MG Oral Tablet [Amaryl] | po |
| dapagliflozin 10 MG [Farxiga] | |
| Metformin hydrochloride 500 MG Oral Tablet | p.o. |
| Insulin, Glulisine, Human 100 UNT/ML Injectable Solution [Apidra] | subcutaneously |
| Glyburide 5 MG Oral Tablet | orally |
| Insulin Lispro 100 UNT/ML Pen Injector [Humalog] | SubQ |
| Metformin hydrochloride 500 MG / sitagliptin 50 MG [Janumet] | |
| 3 ML Insulin Lispro 100 UNT/ML Pen Injector [Humalog] | |
| sitagliptin 100 MG Oral Tablet [Januvia] | ORAL |
| Insulin, Aspart, Human | SQ |
| repaglinide 2 MG Oral Tablet [Prandin] | by mouth |
| insulin human, isophane 100 UNT/ML Injectable Suspension [Humulin N] | SUBCUT |
| Bromocriptine 2.5 MG Oral Tablet [Parlodel] | by mouth |
| exenatide 3.08 MG/ML [Bydureon] | subcutaneous |
| Glyburide 5 MG Oral Tablet | |
| Insulin Glargine | SQ |
| Insulin Lispro 25 UNT/ML | |
| glimepiride 2 MG Oral Tablet [Amaryl] | PO |
| Metformin Extended Release Oral Tablet [Glumetza] | by mouth |
| NovoLog Injectable Product | |
| glimepiride 1 MG Oral Tablet | po |

| | |
|---|---|
| Regular Insulin, Human 100 UNT/ML Injectable Solution [Novolin R] | IV |
| glimepiride 2 MG Oral Tablet | orally |
| insulin human, isophane 70 UNT/ML / Regular Insulin, Human 30 UNT/ML [Humulin] | subcutaneous |
| 3 ML Insulin, Glulisine, Human 100 UNT/ML Pen Injector [Apidra] | subcutaneous |
| Metformin hydrochloride 500 MG / pioglitazone 15 MG Oral Tablet | |
| Insulin, Aspart, Human | NPO |
| Glipizide 10 MG Oral Tablet [Glucotrol] | ORAL |
| Regular Insulin, Human 100 UNT/ML Injectable Solution | subcutaneous |
| saxagliptin 5 MG [Onglyza] | |
| Glyburide 5 MG / Metformin hydrochloride 500 MG Oral Tablet [Glucovance] | |
| 3 ML insulin detemir 100 UNT/ML Pen Injector [Levemir] | SUBCUTANEOUS |
| sitagliptin 25 MG Oral Tablet [Januvia] | by mouth |
| pioglitazone 45 MG Oral Tablet [Actos] | PO |
| Glyburide 5 MG / Metformin hydrochloride 500 MG Oral Tablet | |
| exenatide 2 MG [Bydureon] | |
| canagliflozin 100 MG [Invokana] | |
| 3 ML insulin human, isophane 100 UNT/ML Pen Injector [Humulin N] | subcutaneous |
| 3 ML insulin degludec 100 UNT/ML Pen Injector [Tresiba] | subcutaneous |
| Metformin Oral Tablet | by mouth |
| Osmotic 24 HR Metformin hydrochloride 500 MG Extended Release Oral Tablet [Fortamet] | ORAL |
| Glyburide 5 MG / Metformin hydrochloride 500 MG Oral Tablet [Glucovance] | by mouth |
| exenatide 0.01 MG/ACTUAT [Byetta] | subcutaneous |
| Januvia Oral Product | by mouth |
| Metformin hydrochloride 100 MG/ML | |
| Metformin Oral Tablet | |
| Metformin 250 MG | |
| Actos Oral Product | po |
| Glyburide 6 MG | |
| Insulin Lispro 50 UNT/ML | |
| Metformin hydrochloride 500 MG Extended Release Oral Tablet [Glumetza] | by mouth |
| Metformin hydrochloride 500 MG / sitagliptin 50 MG Oral Tablet [Janumet] | |

| | |
|---|---|
| 3 ML insulin degludec 200 UNT/ML Pen Injector [Tresiba] | subcutaneous |
| Regular Insulin, Human 100 UNT/ML Injectable Solution [Humulin R] | SUBCUT |
| Insulin Lispro 100 UNT/ML Pen Injector [Humalog] | Subcutaneous |
| Amaryl Oral Product | po |
| pioglitazone 30 MG | |
| 60 ACTUAT exenatide 0.005 MG/ACTUAT Pen Injector [Byetta] | |
| Metformin hydrochloride 750 MG [Glucophage] | |
| Glipizide 5 MG Oral Tablet | |
| exenatide 2 MG Injection [Bydureon] | subcutaneous |
| rosiglitazone 4 MG Oral Tablet [Avandia] | PO |
| Bromocriptine 2.5 MG | |
| Metformin hydrochloride 1000 MG / saxagliptin 5 MG [Kombiglyze] | |
| insulin human, isophane 100 UNT/ML Injectable Suspension [Novolin N] | |
| rosiglitazone 2 MG [Avandia] | |
| repaglinide 2 MG [Prandin] | |
| insulin human, isophane 40 UNT/ML | |
| Glyburide 1.25 MG Oral Tablet | by mouth |
| Insulin Glargine 100 UNT/ML [Lantus] | SQ |
| Metformin hydrochloride 850 MG Oral Tablet | PO |
| Metformin hydrochloride 1000 MG / sitagliptin 50 MG Oral Tablet [Janumet] | po |
| 24 HR Metformin hydrochloride 750 MG Extended Release Oral Tablet | po |
| Glyburide 5 MG / Metformin hydrochloride 500 MG Oral Tablet | by mouth |
| glimepiride 4 MG Oral Tablet | oral |
| nateglinide 60 MG [Starlix] | |
| 3 ML Insulin, Aspart, Human 100 UNT/ML Cartridge [NovoLog] | SUBCUTANEOUS |
| Metformin Oral Solution | orally |
| sitagliptin 100 MG Oral Tablet [Januvia] | orally |
| insulin human, isophane 70 UNT/ML / Regular Insulin, Human 30 UNT/ML Injectable Suspension [Humulin] | |
| Glipizide 10 MG Oral Tablet | oral |
| Insulin, Aspart Protamine, Human 70 UNT/ML / Insulin, Aspart, Human 30 UNT/ML Injectable Suspension [NovoLog Mix] | Subcutaneous |
| glimepiride 4 MG Oral Tablet [Amaryl] | oral |
| pioglitazone 30 MG Oral Tablet | by mouth |
| 3 ML liraglutide 6 MG/ML Pen Injector [Victoza] | SUBCUTANEOUS |

| | |
|---|---|
| Glyburide Oral Tablet | PO |
| Metformin hydrochloride 850 MG Oral Tablet | ORAL |
| insulin human, isophane 100 UNT/ML Injectable Suspension [Humulin N] | Subcutaneous |
| Insulin, Aspart, Human | sq |
| Glyburide 2.5 MG Oral Tablet | PO |
| dapagliflozin 5 MG Oral Tablet [Farxiga] | by mouth |
| insulin human, isophane 70 UNT/ML / Regular Insulin, Human 30 UNT/ML Injectable Suspension [Humulin] | Subcutaneous |
| insulin human, isophane 70 UNT/ML / Regular Insulin, Human 30 UNT/ML [Humulin] | SubQ |
| 24 HR Metformin hydrochloride 500 MG Extended Release Oral Tablet [Glucophage] | by mouth |
| Humulin R Injectable Product | subcutaneously |
| Regular Insulin, Human 100 UNT/ML Injectable Solution | intravenous |
| Glyburide 5 MG Oral Tablet | oral |
| dulaglutide 3 MG/ML [Trulicity] | subcutaneous |
| Insulin Glargine | Subcutaneous |
| pioglitazone 15 MG Oral Tablet [Actos] | PO |
| Metformin Oral Solution | PO |
| insulin human, isophane 70 UNT/ML | |
| repaglinide 1 MG Oral Tablet [Prandin] | by mouth |
| Insulin Lispro 25 UNT/ML | subcut |
| glimepiride 2 MG Oral Tablet [Amaryl] | oral |
| rosiglitazone 2 MG Oral Tablet [Avandia] | po |
| Metformin hydrochloride 1000 MG Oral Tablet | p.o. |
| Metformin Pill | |
| Metformin hydrochloride 1000 MG Oral Tablet [Glucophage] | oral |
| Actos Oral Product | by mouth |
| Metformin hydrochloride 850 MG Oral Tablet | orally |
| sitagliptin 50 MG Oral Tablet [Januvia] | po |
| glimepiride 1 MG Oral Tablet [Amaryl] | ORAL |
| 3 ML Insulin Lispro 200 UNT/ML Pen Injector [Humalog] | subcutaneous |
| Insulin Lispro Injectable Solution | |
| Insulin Lispro 100 UNT/ML Injectable Solution | |
| pramlintide acetate 0.6 MG/ML Injectable Solution [Symlin] | Subcutaneous |
| Metformin hydrochloride 500 MG / rosiglitazone 2 MG [Avandamet] | |
| 24 HR Metformin hydrochloride 750 MG Extended Release Oral Tablet [Glucophage] | ORAL |

| | |
|---|---|
| Metformin hydrochloride 850 MG / pioglitazone 15 MG Oral Tablet [Actoplus Met] | by mouth |
| glimepiride 4 MG Oral Tablet | PO |
| insulin human, isophane 70 UNT/ML / Regular Insulin, Human 30 UNT/ML Injectable Suspension [Novolin] | Subcutaneous |
| Regular Insulin, Human | |
| Glyburide 6 MG Oral Tablet | po |
| Metformin hydrochloride 500 MG Oral Tablet [Glucophage] | orally |
| pioglitazone 30 MG Oral Tablet [Actos] | PO |
| Insulin Glargine 100 UNT/ML | |
| Glyburide 6 MG Oral Tablet | by mouth |
| Insulin Glargine 100 UNT/ML [Lantus] | subcut |
| rosiglitazone 2 MG Oral Tablet [Avandia] | by mouth |
| Glipizide 10 MG Oral Tablet [Glucotrol] | PO |
| Glyburide 1.25 MG Oral Tablet [Diabeta] | by mouth |
| Metformin Extended Release Oral Tablet | |
| exenatide 0.005 MG/ACTUAT [Byetta] | subcut |
| Glyburide 3 MG Oral Tablet [Glynase] | by mouth |
| Glipizide Oral Product | po |
| insulin human, isophane 70 UNT/ML / Regular Insulin, Human 30 UNT/ML [Humulin] | SQ |
| Metformin hydrochloride 850 MG Oral Tablet | oral |
| glimepiride 8 MG | |
| Glyburide 5 MG / Metformin hydrochloride 500 MG Oral Tablet [Glucovance] | po |
| insulin detemir | |
| glimepiride 2 MG Oral Tablet | PO |
| exenatide 0.005 MG/ACTUAT [Byetta] | SQ |
| pioglitazone 15 MG | |
| sitagliptin 100 MG Oral Tablet [Januvia] | PO |
| Metformin hydrochloride 500 MG Oral Tablet [Glucophage] | oral |
| Insulin Glargine 100 UNT/ML Injectable Solution [Lantus] | |
| Metformin hydrochloride 1000 MG Extended Release Oral Tablet [Glumetza] | by mouth |
| repaglinide 0.5 MG Oral Tablet [Prandin] | by mouth |
| Regular Insulin, Human 500 UNT/ML Injectable Solution [Humulin R] | subcutaneously |
| Insulin Glargine Injectable Solution | sq |
| Metformin hydrochloride 500 MG Extended Release Oral Tablet [Fortamet] | |

| | | |
|---|---|---|
| 24 HR Glipizide 10 MG Extended Release Oral Tablet [Glucotrol] | ORAL | |
| Insulin, Aspart, Human | po | |
| Insulin Glargine Injectable Solution | subcutaneously | |
| insulin human, isophane 100 UNT/ML Injectable Suspension [Novolin N] | Subcutaneous | |
| Glyburide 5 MG / Metformin hydrochloride 500 MG [Glucovance] | | |
| Metformin hydrochloride 500 MG [Glumetza] | | |
| Lantus Injectable Product | SQ | |
| pioglitazone 45 MG Oral Tablet | by mouth | |
| Insulin, Aspart, Human | subcutaneously | |
| Metformin hydrochloride 500 MG Oral Tablet | | 0 |
| insulin human, isophane 70 UNT/ML / Regular Insulin, Human 30 UNT/ML Injectable Suspension [Novolin] | SUBCUT | |
| Glipizide Extended Release Oral Tablet [Glucotrol] | | |
| Bromocriptine | | |
| dapagliflozin 5 MG [Farxiga] | | |
| Janumet Oral Product | by mouth | |
| Victoza Injectable Product | | |
| Insulin Lispro Cartridge [Humalog] | subcutaneous | |
| Osmotic 24 HR Metformin hydrochloride 1000 MG Extended Release Oral Tablet [Fortamet] | ORAL | |
| Glyburide 5 MG Oral Tablet | p.o. | |
| Glipizide 10 MG Oral Tablet | | |
| Humalog Injectable Product | | |
| Insulin, Aspart Protamine, Human 70 UNT/ML / Insulin, Aspart, Human 30 UNT/ML Injectable Suspension [NovoLog Mix] | subq | |
| pioglitazone 45 MG | | |
| Levemir Injectable Product | | |
| Glyburide 2.5 MG / Metformin hydrochloride 500 MG Oral Tablet [Glucovance] | by mouth | |
| 3 ML Insulin, Aspart, Human 100 UNT/ML Pen Injector [NovoLog] | | |
| Glucophage Oral Product | PO | |
| 24 HR Metformin hydrochloride 500 MG Extended Release Oral Tablet [Glucophage] | | 0 |
| dulaglutide 1.5 MG/ML [Trulicity] | subcutaneous | |
| Insulin Glargine Injectable Solution | subcutaneous | |
| Glipizide 10 MG Oral Tablet | Oral | |
| Metformin 500 MG / ropinirole 2 MG Oral Tablet | | |
| Humulin N Injectable Product | subcutaneously | |
| 24 HR Glipizide 5 MG Extended Release Oral Tablet [Glucotrol] | ORAL | |

| | | |
|---|---|---|
| Metformin hydrochloride 100 MG/ML Oral Solution | by mouth | |
| rosiglitazone 8 MG Oral Tablet [Avandia] | PO | |
| Insulin Glargine 100 UNT/ML Injectable Solution [Lantus] | subcu | |
| Glyburide 2.5 MG / Metformin hydrochloride 500 MG Oral Tablet [Glucovance] | | |
| glimepiride 4 MG Oral Tablet | | |
| 3 ML insulin detemir 100 UNT/ML Pen Injector [Levemir] | Subcutaneous | |
| insulin human, isophane 100 UNT/ML Pen Injector [Humulin N] | SubQ | |
| Insulin Lispro | NPO | |
| 3 ML liraglutide 6 MG/ML Pen Injector [Victoza] | subq | |
| glimepiride 4 MG Oral Tablet [Amaryl] | | |
| Insulin Glargine | subcut | |
| sitagliptin 100 MG Oral Tablet [Januvia] | oral | |
| Glyburide 1.25 MG Oral Tablet | po | |
| pioglitazone Oral Tablet | by mouth | |
| Glyburide 3 MG | | |
| glimepiride 2 MG Oral Tablet | oral | |
| empagliflozin 25 MG Oral Tablet [Jardiance] | by mouth | |
| sitagliptin 25 MG [Januvia] | | |
| Insulin, Aspart, Human 100 UNT/ML Injectable Solution | | |
| insulin human, isophane 100 UNT/ML Injectable Suspension [Novolin N] | SUBCUT | |
| repaglinide 1 MG [Prandin] | | |
| Glipizide 5 MG Oral Tablet [Glucotrol] | PO | |
| Insulin Lispro Cartridge [Humalog] | subq | |
| Metformin hydrochloride 1000 MG Oral Tablet [Glucophage] | | 0 |
| Glipizide 2.5 MG Oral Tablet | orally | |
| glimepiride 1 MG Oral Tablet | orally | |
| insulin detemir 100 UNT/ML Injectable Solution [Levemir] | SUBCUTANEOUS | |
| Regular Insulin, Human 100 UNT/ML Injectable Solution [Humulin R] | subcutaneous | |
| Metformin hydrochloride 500 MG / sitagliptin 50 MG Oral Tablet [Janumet] | po | |
| Insulin, Aspart, Human | SUBCUT. | |
| Diabeta Oral Product | by mouth | |
| Insulin Lispro | SQ | |
| Insulin Glargine Injectable Solution [Lantus] | | |
| Metformin hydrochloride 850 MG Oral Tablet | | |
| Metformin Extended Release Oral Tablet | by mouth | |
| Glyburide 2.5 MG [Diabeta] | | |

| | |
|---|---|
| Metformin hydrochloride 1000 MG Oral Tablet [Glucophage] | orally |
| empagliflozin 10 MG Oral Tablet [Jardiance] | by mouth |
| Insulin, Aspart, Human 100 UNT/ML Injectable Solution [NovoLog] | |
| nateglinide 120 MG | |
| Metformin hydrochloride 1000 MG Extended Release Oral Tablet [Fortamet] | by mouth |
| repaglinide 2 MG Oral Tablet [Prandin] | po |
| insulin human, isophane 60 UNT/ML | |
| Metformin hydrochloride 1000 MG Extended Release Oral Tablet [Fortamet] | |
| repaglinide 0.5 MG [Prandin] | |
| Insulin Lispro 50 UNT/ML | subcut |
| 24 HR Metformin hydrochloride 1000 MG / sitagliptin 100 MG Extended Release Oral Tablet [Janumet] | by mouth |
| Insulin Lispro | sq |
| Glyburide Oral Tablet | orally |
| Insulin Lispro Injectable Solution [Humalog] | subcutaneously |
| insulin human, isophane 70 UNT/ML / Regular Insulin, Human 30 UNT/ML [Humulin] | subcut |
| 60 ACTUAT exenatide 0.01 MG/ACTUAT Pen Injector [Byetta] | SUBCUTANEOUS |
| Glyburide 2.5 MG Oral Tablet | orally |
| insulin pork, isophane 100 UNT/ML Injectable Suspension [NPH Iletin II] | SQ |
| Fortamet Oral Product | by mouth |
| Insulin Lispro 100 UNT/ML Injectable Solution [Humalog] | |
| Metformin hydrochloride 1000 MG [Fortamet] | |
| Insulin Lispro | subcutaneous |
| Metformin hydrochloride 500 MG [Fortamet] | |
| 1.5 ML pramlintide acetate 1 MG/ML Pen Injector [Symlin] | subcutaneous |
| insulin human, isophane 70 UNT/ML / Regular Insulin, Human 30 UNT/ML [Humulin] | subq |
| Glyburide 2.5 MG Oral Tablet | ORAL |
| pioglitazone 15 MG Oral Tablet | by mouth |
| Insulin, Aspart, Human 100 UNT/ML Cartridge [NovoLog] | subcut |
| 3 ML Insulin, Glulisine, Human 100 UNT/ML Pen Injector [Apidra] | Subcutaneous |
| Glipizide 10 MG Oral Tablet [Glucotrol] | Oral |
| Insulin Glargine | by mouth |
| glimepiride 2 MG Oral Tablet | |

| | |
|---|---|
| Metformin hydrochloride 750 MG Extended Release Oral Tablet [Glucophage] | by mouth |
| pioglitazone 30 MG Oral Tablet [Actos] | ORAL |
| Insulin, Aspart, Human 100 UNT/ML Injectable Solution [NovoLog] | subcutaneous |
| Glyburide 5 MG [Micronase] | |
| Insulin Lispro Cartridge [Humalog] | SQ |
| Glyburide 3 MG Oral Tablet | po |
| 24 HR Metformin hydrochloride 750 MG Extended Release Oral Tablet | |
| nateglinide 120 MG Oral Tablet | by mouth |
| Metformin hydrochloride 1000 MG / sitagliptin 100 MG [Janumet] | |
| insulin human, isophane 40 UNT/ML | subcut |
| saxagliptin 2.5 MG Oral Tablet [Onglyza] | by mouth |
| Insulin Glargine 100 UNT/ML Injectable Solution | |
| Glyburide 6 MG [Glynase] | |
| Metformin hydrochloride 500 MG Oral Tablet [Glucophage] | p.o. |
| Lantus Injectable Product | sq |
| Metformin hydrochloride 750 MG Extended Release Oral Tablet [Glucophage] | |
| Regular Insulin, Human 10 UNT/ML | subcut |
| Metformin hydrochloride 500 MG Oral Tablet | Oral |
| insulin human, isophane 70 UNT/ML / Regular Insulin, Human 30 UNT/ML [Novolin] | SQ |
| Insulin Glargine 100 UNT/ML Injectable Solution | Subcutaneous |
| insulin human, isophane 70 UNT/ML / Regular Insulin, Human 30 UNT/ML [Novolin] | subcut |
| Glyburide Oral Tablet | p.o. |
| Metformin 250 MG Oral Tablet | by mouth |
| Glipizide 5 MG Oral Tablet | Oral |
| nateglinide 120 MG Oral Tablet [Starlix] | PO |
| Regular Insulin, Human 5 UNT/ML | subcut |
| Bromocriptine 2.5 MG [Parlodel] | |
| Januvia Oral Product | po |
| Metformin hydrochloride 850 MG Oral Tablet [Glucophage] | PO |
| dulaglutide 1.5 MG/ML [Trulicity] | |
| empagliflozin 25 MG [Jardiance] | |
| insulin human, isophane 70 UNT/ML / Regular Insulin, Human 30 UNT/ML [Novolin] | subcutaneous |

| | |
|---|---|
| Glyburide 2.5 MG / Metformin hydrochloride 500 MG Oral Tablet | |
| 24 HR Metformin hydrochloride 500 MG Extended Release Oral Tablet | oral |
| Insulin, Aspart, Human | subq |
| Insulin, Aspart Protamine, Human 70 UNT/ML / Insulin, Aspart, Human 30 UNT/ML Injectable Suspension [NovoLog Mix] | subQ |
| Glyburide 6 MG Oral Tablet [Glynase] | po |
| Acarbose 50 MG Oral Tablet [Precose] | by mouth |
| Glyburide 3 MG Oral Tablet | |
| Regular Insulin, Human 10 UNT/ML | iv |
| Insulin Glargine | orally |
| Avandia Oral Product | po |
| exenatide 0.01 MG/ACTUAT [Byetta] | SQ |
| nateglinide 60 MG Oral Tablet [Starlix] | po |
| glimepiride Oral Product | po |
| Novolin N Injectable Product | subcutaneously |
| insulin pork, isophane 100 UNT/ML Injectable Suspension [NPH Iletin II] | subcut |
| glimepiride 2 MG Oral Tablet [Amaryl] | |
| insulin human, isophane 100 UNT/ML Injectable Suspension [Humulin N] | SUBCUTANEOUS |
| 3 ML Insulin Lispro 100 UNT/ML Cartridge [Humalog] | SUBCUTANEOUS |
| Metformin hydrochloride 1000 MG Oral Tablet [Glucophage] | |
| 24 HR Glipizide 10 MG Extended Release Oral Tablet [Glucotrol] | |
| exenatide 0.01 MG/ACTUAT [Byetta] | subcut. |
| sitagliptin 100 MG Oral Tablet | orally |
| Insulin Lispro 25 UNT/ML / Insulin, Protamine Lispro, Human 75 UNT/ML Injectable Suspension [Humalog Mix] | subq |
| insulin human, isophane 100 UNT/ML Injectable Suspension | subcutaneous |
| Regular Insulin, Human | SUBCUT. |
| empagliflozin 10 MG [Jardiance] | |
| Regular Insulin, Human 100 UNT/ML Injectable Solution | |
| nateglinide 60 MG | |
| Byetta Injectable Product | subcutaneously |
| glimepiride 1 MG Oral Tablet [Amaryl] | PO |
| glimepiride 8 MG Oral Tablet | po |
| Glipizide 10 MG Oral Tablet [Glucotrol] | oral |
| Insulin Lispro Cartridge [Humalog] | subcut. |
| Insulin Lispro 100 UNT/ML Pen Injector [Humalog] | |
| 3 ML Insulin Lispro 100 UNT/ML Pen Injector [Humalog] | subcutaneous |

| | |
|---|---|
| Amaryl Oral Product | PO |
| exenatide 0.005 MG/ACTUAT [Byetta] | subcutaneous |
| Acarbose 100 MG Oral Tablet [Precose] | by mouth |
| pioglitazone 45 MG Oral Tablet [Actos] | ORAL |
| insulin human, isophane 100 UNT/ML Injectable Suspension [Novolin N] | SUBCUTANEOUS |
| Glyburide 3 MG Oral Tablet | by mouth |
| exenatide 2 MG [Bydureon] | SubQ |
| insulin pork, isophane 100 UNT/ML Injectable Suspension [NPH Iletin II] | subcutaneously |
| Insulin Glargine 100 UNT/ML [Lantus] | sq |
| Glucophage Oral Product | BY MOUTH |
| Insulin Lispro | npo |
| Insulin Lispro 25 UNT/ML / Insulin, Protamine Lispro, Human 75 UNT/ML Injectable Suspension [Humalog Mix] | Subcutaneous |
| Insulin, Aspart, Human 100 UNT/ML Cartridge [NovoLog] | SQ |
| Glipizide 5 MG Oral Tablet | |
| Avandamet Oral Product | po |
| Glucovance Oral Product | by mouth |
| glimepiride 3 MG Oral Tablet [Amaryl] | by mouth |
| repaglinide 0.5 MG Oral Tablet [Prandin] | po |
| Insulin, Aspart Protamine, Human 70 UNT/ML / Insulin, Aspart, Human 30 UNT/ML Injectable Suspension [NovoLog Mix] | SUBCUT |
| Metformin hydrochloride 1000 MG Extended Release Oral Tablet | |
| glimepiride 4 MG Oral Tablet [Amaryl] | orally |
| Glipizide Oral Tablet | by mouth |
| Acarbose 50 MG [Precose] | |
| Metformin hydrochloride 1000 MG / rosiglitazone 4 MG [Avandamet] | |
| Modified 24 HR Metformin hydrochloride 500 MG Extended Release Oral Tablet [Glumetza] | ORAL |
| Metformin hydrochloride 750 MG Extended Release Oral Tablet [Glucophage] | po |
| Glyburide 2.5 MG / Metformin hydrochloride 500 MG Oral Tablet | by mouth |
| 24 HR Glipizide 2.5 MG Extended Release Oral Tablet [Glucotrol] | ORAL |
| insulin human, isophane 70 UNT/ML / Regular Insulin, Human 30 UNT/ML Injectable Suspension [Humulin] | SUBCUTANEOUS |
| Metformin hydrochloride 1000 MG / saxagliptin 2.5 MG [Kombiglyze] | |
| Novolin R Injectable Product | subcutaneously |

| | |
|---|---|
| Metformin hydrochloride 1000 MG / rosiglitazone 4 MG Oral Tablet [Avandamet] | po |
| saxagliptin 5 MG Oral Tablet [Onglyza] | po |
| 24 HR Glipizide 5 MG Extended Release Oral Tablet | PO |
| Glipizide 5 MG Oral Tablet [Glucotrol] | Oral |
| Insulin Lispro Injectable Solution | sq |
| Metformin hydrochloride 100 MG/ML Oral Solution | po |
| Invokana Oral Product | by mouth |
| Glyburide 2.5 MG / Metformin hydrochloride 500 MG Oral Tablet [Glucovance] | po |
| Avandamet Oral Product | by mouth |
| glimepiride 1 MG Oral Tablet | oral |
| insulin human, isophane 70 UNT/ML / Regular Insulin, Human 30 UNT/ML [Humulin] | sq |
| pramlintide acetate 0.6 MG/ML Injectable Solution [Symlin] | subcutaneously |
| Regular Insulin, Human 10 UNT/ML | |
| Regular Insulin, Human 100 UNT/ML Injectable Solution [Novolin R] | subcutaneous |
| pioglitazone 30 MG Oral Tablet | orally |
| Metformin Extended Release Oral Tablet [Glucophage] | by mouth |
| insulin human, isophane 70 UNT/ML / Regular Insulin, Human 30 UNT/ML Injectable Suspension [Humulin] | SUBCUT |
| Insulin Glargine | sq |
| pioglitazone | |
| 24 HR Glipizide 10 MG Extended Release Oral Tablet | PO |
| Regular Insulin, Human | subcutaneous |
| Glipizide 5 MG Oral Tablet | p.o. |
| 3 ML Insulin, Aspart Protamine, Human 70 UNT/ML / Insulin, Aspart, Human 30 UNT/ML Pen Injector [NovoLog Mix] | SUBCUTANEOUS |
| 3 ML insulin degludec 100 UNT/ML Pen Injector [Tresiba] | |
| Metformin hydrochloride 100 MG/ML Oral Solution [Riomet] | by mouth |
| Metformin Extended Release Oral Tablet [Fortamet] | by mouth |
| insulin human, isophane 70 UNT/ML / Regular Insulin, Human 30 UNT/ML [Novolin] | subq |
| Glyburide 1.5 MG Oral Tablet [Glynase] | by mouth |
| Glyburide 6 MG Oral Tablet | |
| 24 HR Glipizide 5 MG Extended Release Oral Tablet [Glucotrol] | |
| Acarbose 25 MG Oral Tablet [Precose] | by mouth |
| Insulin, Aspart, Human 100 UNT/ML Injectable Solution [NovoLog] | |

| | |
|---|---|
| 3 ML insulin human, isophane 100 UNT/ML Prefilled Syringe | |
| repaglinide 1 MG Oral Tablet [Prandin] | po |
| Metformin hydrochloride 500 MG Oral Tablet [Glucophage] | ORAL |
| insulin human, isophane 70 UNT/ML / Regular Insulin, Human 30 UNT/ML Injectable Suspension [Novolin] | SUBCUTANEOUS |
| Glipizide 10 MG Oral Tablet | p.o. |
| pioglitazone 15 MG Oral Tablet [Actos] | ORAL |
| sitagliptin 100 MG | |
| glimepiride 1 MG Oral Tablet [Amaryl] | oral |
| Glipizide Oral Product | oral |
| Insulin Glargine 100 UNT/ML Injectable Solution | subcutaneous |
| pioglitazone 30 MG Oral Tablet [Actos] | orally |
| Insulin Glargine | subq |
| Insulin, Aspart Protamine, Human 70 UNT/ML / Insulin, Aspart, Human 30 UNT/ML Injectable Suspension [NovoLog Mix] | SQ |
| Bromocriptine 2.5 MG Oral Tablet | by mouth |
| insulin pork, isophane 80 UNT/ML | |
| Metformin 250 MG Oral Tablet | po |
| Regular Insulin, Human 500 UNT/ML Injectable Solution [Humulin R] | sq |
| 3 ML insulin detemir 100 UNT/ML Pen Injector [Levemir] | |
| Metformin hydrochloride 1000 MG Extended Release Oral Tablet [Fortamet] | po |
| Insulin, Aspart, Human 100 UNT/ML Injectable Solution | subcutaneous |
| Regular Insulin, Human | subq |
| pioglitazone 45 MG Oral Tablet | orally |
| Acarbose 50 MG | |
| glimepiride 4 MG Oral Tablet [Amaryl] | |
| Metformin Oral Tablet [Glucophage] | BY MOUTH |
| insulin human, isophane 16 UNT/ML | |
| insulin pork, isophane 100 UNT/ML Injectable Suspension [NPH Iletin II] | sq |
| Regular Insulin, Human 100 UNT/ML Injectable Solution [Humulin R] | intravenously |
| 24 HR Glipizide 2.5 MG Extended Release Oral Tablet [Glucotrol] | |
| 24 HR Glipizide 2.5 MG Extended Release Oral Tablet [Glucotrol] | by mouth |
| pioglitazone 30 MG Oral Tablet [Actos] | oral |
| Metformin hydrochloride 1000 MG Oral Tablet [Glucophage] | p.o. |
| insulin pork, isophane 100 UNT/ML Injectable Suspension [NPH Iletin II] | subcutaneous |

| | | |
|---|---|---|
| Insulin Glargine 100 UNT/ML Injectable Solution [Lantus] | subQ | |
| Insulin Lispro 200 UNT/ML [Humalog] | | |
| Glipizide 10 MG Oral Tablet [Glucotrol] | | |
| Metformin hydrochloride 1000 MG / sitagliptin 50 MG Oral Tablet [Janumet] | ORAL | |
| Glyburide 1.25 MG / Metformin hydrochloride 250 MG Oral Tablet | by mouth | |
| Metformin hydrochloride 500 MG Oral Tablet | BY MOUTH | |
| Glyburide 2.5 MG Oral Tablet | oral | |
| sitagliptin 50 MG Oral Tablet [Januvia] | PO | |
| Glipizide 2.5 MG Oral Tablet | PO | |
| Metformin hydrochloride 1000 MG [Glumetza] | | |
| Regular Insulin, Human 5 UNT/ML | | |
| exenatide 0.01 MG/ACTUAT [Byetta] | subq | |
| Metformin 500 MG / ropinirole 2 MG Oral Tablet | by mouth | |
| Metformin hydrochloride 1000 MG / rosiglitazone 4 MG Oral Tablet [Avandamet] | by mouth | |
| Glyburide 5 MG / Metformin hydrochloride 500 MG Oral Tablet [Glucovance] | PO | |
| 3 ML Insulin Glargine 100 UNT/ML Pen Injector [Lantus] | | 0 |
| sitagliptin 50 MG Oral Tablet [Januvia] | ORAL | |
| Metformin hydrochloride 500 MG Extended Release Oral Tablet [Fortamet] | by mouth | |
| 60 ACTUAT exenatide 0.01 MG/ACTUAT Pen Injector [Byetta] | po | |
| 0.5 ML dulaglutide 1.5 MG/ML Auto-Injector [Trulicity] | | |
| Janumet Oral Product | po | |
| exenatide 2 MG [Bydureon] | SQ | |
| glimepiride Oral Product | orally | |
| 3 ML insulin degludec 200 UNT/ML Pen Injector [Tresiba] | | |
| Insulin Lispro | SUB CONJ | |
| Insulin, Aspart Protamine, Human 70 UNT/ML / Insulin, Aspart, Human 30 UNT/ML Injectable Suspension [NovoLog Mix] | subcut | |
| Metformin hydrochloride 500 MG Extended Release Oral Tablet [Glumetza] | po | |
| glimepiride 8 MG Oral Tablet | by mouth | |
| Glipizide 10 MG Oral Tablet [Glucotrol] | | |
| Glyburide 2.5 MG Oral Tablet | | |
| Glyburide Oral Tablet | oral | |
| Actos Oral Product | PO | |
| Glyburide 5 MG / Metformin hydrochloride 500 MG Oral Tablet | po | |

| | | |
|---|---|---|
| sitagliptin 50 MG Oral Tablet [Januvia] | oral | |
| Glyburide 3 MG [Glynase] | | |
| exenatide 0.01 MG/ACTUAT [Byetta] | subcutaneously | |
| Insulin, Aspart, Human | npo. | |
| Metformin | PO | |
| saxagliptin 2.5 MG [Onglyza] | | |
| nateglinide 60 MG Oral Tablet | by mouth | |
| insulin, regular, beef-pork 40 UNT/ML Inhalant Solution | subcutaneously | |
| Humalog Mix Injectable Product | subcutaneously | |
| Glyburide Oral Tablet | | |
| Metformin | subcutaneously | |
| Regular Insulin, Human 100 UNT/ML Injectable Solution [Humulin R] | sq | |
| Insulin Glargine 100 UNT/ML Injectable Solution [Lantus] | | 0 |
| Glipizide 2.5 MG / Metformin hydrochloride 250 MG Oral Tablet | by mouth | |
| Glyburide Oral Tablet [Glynase] | by mouth | |
| Bromocriptine 5 MG | | |
| Metformin hydrochloride 1000 MG Oral Tablet | Oral | |
| Insulin, Aspart, Human | SC | |
| nateglinide 120 MG Oral Tablet [Starlix] | ORAL | |
| Insulin Glargine Injectable Solution | subcut | |
| 24 HR Metformin hydrochloride 500 MG Extended Release Oral Tablet | PO | |
| Glyburide 5 MG / Metformin 1000 MG Oral Tablet | po | |
| Glipizide 5 MG / Metformin hydrochloride 500 MG Oral Tablet | by mouth | |
| 24 HR Metformin hydrochloride 750 MG Extended Release Oral Tablet | oral | |
| Glipizide Oral Tablet | | |
| Glipizide Oral Product | orally | |
| Glipizide Oral Product | PO | |
| Acarbose 50 MG Oral Tablet | by mouth | |
| exenatide 0.01 MG/ACTUAT [Byetta] | po | |
| Glyburide 2.5 MG Oral Tablet | p.o. | |
| 3 ML insulin detemir 100 UNT/ML Pen Injector [Levemir] | subq | |
| Glyburide 3 MG Oral Tablet [Glynase] | po | |
| pioglitazone 45 MG Oral Tablet [Actos] | orally | |
| Regular Insulin, Human 8 UNT | subcut | |
| Insulin Lispro 100 UNT/ML Injectable Solution [Humalog] | subcutaneous | |
| Insulin Glargine 100 UNT/ML [Lantus] | Subcut | |
| Humulin R Injectable Product | | |

| | |
|---|---|
| Metformin hydrochloride 100 MG/ML Oral Solution [Riomet] | Oral |
| 24 HR Metformin hydrochloride 750 MG Extended Release Oral Tablet | orally |
| Insulin Lispro 25 UNT/ML / Insulin, Protamine Lispro, Human 75 UNT/ML Injectable Suspension [Humalog Mix] | subcut |
| pioglitazone 15 MG Oral Tablet | orally |
| 24 HR Metformin hydrochloride 750 MG Extended Release Oral Tablet [Glucophage] | oral |
| insulin human, isophane 70 UNT/ML / Regular Insulin, Human 30 UNT/ML [Novolin] | sq |
| Glyburide 6 MG Oral Tablet | PO |
| Insulin Lispro | subq |
| Metformin Oral Solution | |
| Starlix Oral Product | by mouth |
| Bromocriptine 2.5 MG Oral Tablet | po |
| insulin human, isophane 70 UNT/ML | subcut |
| Lantus Injectable Product | subcutaneous |
| Insulin, Aspart, Human | PO |
| Regular Insulin, Human 500 UNT/ML Injectable Solution [Humulin R] | SUBCUT |
| sitagliptin 50 MG Oral Tablet [Januvia] | orally |
| insulin human, isophane 28 UNT/ML | |
| Acarbose 25 MG | |
| Regular Insulin, Human 100 UNT/ML Injectable Solution [Humulin R] | SUBCUTANEOUS |
| insulin human, isophane 60 UNT/ML | subcut |
| liraglutide 6 MG/ML [Victoza] | SUBCUTANEOUS |
| Metformin hydrochloride 750 MG Extended Release Oral Tablet | |
| Insulin Lispro | by mouth |
| 3 ML Insulin Glargine 100 UNT/ML Pen Injector [Basaglar] | |
| insulin human, isophane 70 UNT/ML / Regular Insulin, Human 30 UNT/ML Injectable Suspension [Novolin] | |
| Glyburide 1 MG | |
| Insulin, Aspart Protamine, Human 70 UNT/ML / Insulin, Aspart, Human 30 UNT/ML Injectable Suspension | subcut |
| dapagliflozin 10 MG Oral Tablet [Farxiga] | oral |
| 3 ML Insulin Glargine 100 UNT/ML Pen Injector [Lantus] | SQ |
| Metformin hydrochloride 850 MG / pioglitazone 15 MG Oral Tablet [Actoplus Met] | po |
| 60 ACTUAT exenatide 0.005 MG/ACTUAT Pen Injector [Byetta] | SUBCUTANEOUS |

| | |
|---|---|
| glimepiride 2 MG Oral Tablet [Amaryl] | |
| Glucotrol Oral Product | po |
| Insulin, Glulisine, Human 100 UNT/ML Injectable Solution [Apidra] | SUBCUTANEOUS |
| Metformin Oral Tablet [Glucophage] | po |
| Glucovance Oral Product | po |
| Insulin, Aspart Protamine, Human 70 UNT/ML / Insulin, Aspart, Human 30 UNT/ML [NovoLog Mix] | SQ |
| pramlintide acetate 0.6 MG/ML Injectable Solution [Symlin] | subcutaneous |
| empagliflozin 25 MG Oral Tablet [Jardiance] | oral |
| Metformin hydrochloride 1000 MG / sitagliptin 50 MG Oral Tablet [Janumet] | orally |
| Insulin Lispro 25 UNT/ML / Insulin, Protamine Lispro, Human 75 UNT/ML Injectable Suspension [Humalog Mix] | SQ |
| Glyburide 5 MG / Metformin hydrochloride 500 MG Oral Tablet [Glucovance] | ORAL |
| Insulin Glargine 100 UNT/ML [Lantus] | subcut. |
| Glipizide 2.5 MG Oral Tablet | Oral |
| Insulin Glargine | subcu |
| Glyburide 5 MG Oral Tablet [Diabeta] | po |
| Regular Insulin, Human | SUB CONJ |
| Regular Insulin, Human 15 UNT/ML | subcut |
| insulin human, isophane 75 UNT/ML | |
| Insulin Lispro 50 UNT/ML / Insulin, Protamine Lispro, Human 50 UNT/ML Injectable Suspension [Humalog Mix] | subcutaneous |
| Metformin hydrochloride 1000 MG / sitagliptin 50 MG Oral Tablet | by mouth |
| Insulin, Glulisine, Human 100 UNT/ML Injectable Solution [Apidra] | sq |
| Glipizide 5 MG / Metformin hydrochloride 500 MG Oral Tablet | |
| Insulin Lispro 50 UNT/ML | subcutaneous |
| Regular Insulin, Human 100 UNT/ML Injectable Solution [Humulin R] | SQ |
| insulin human, isophane 70 UNT/ML / Regular Insulin, Human 30 UNT/ML [Humulin] | subcut. |
| Glipizide 5 MG Oral Tablet [Glucotrol] | |
| Insulin, Aspart, Human 100 UNT/ML Injectable Solution | Subcutaneous |
| glimepiride 2 MG Oral Tablet [Amaryl] | orally |
| 24 HR Glipizide 2.5 MG Extended Release Oral Tablet [Glucotrol] | po |
| Glyburide | PO |

| | | |
|---|---|---|
| 24 HR Metformin hydrochloride 500 MG Extended Release Oral Tablet | po | |
| Simvastatin 10 MG / sitagliptin 100 MG [Juvisync] | | |
| Acarbose 25 MG [Precose] | | |
| Metformin hydrochloride 500 MG / rosiglitazone 4 MG Oral Tablet [Avandamet] | po | |
| pioglitazone 30 MG Oral Tablet [Actos] | p.o. | |
| glimepiride 1 MG Oral Tablet [Amaryl] | | |
| Insulin, Aspart, Human 100 UNT/ML Injectable Solution [NovoLog] | | 0 |
| Starlix Oral Product | po | |
| 3 ML Insulin, Aspart, Human 100 UNT/ML Pen Injector [NovoLog] | | 0 |
| 24 HR Glipizide 10 MG Extended Release Oral Tablet | po | |
| 3 ML Insulin Glargine 100 UNT/ML Pen Injector [Lantus] | subcut | |
| Glipizide 10 MG Oral Tablet [Glucotrol] | orally | |
| Regular Insulin, Human 6 UNT/ML | subcut | |
| Diabeta Oral Product | PO | |
| Metformin hydrochloride 500 MG Oral Tablet [Glucophage] | Orally | |
| Glipizide 5 MG Oral Tablet [Glucotrol] | p.o. | |
| Insulin Glargine Injectable Solution | Subcutaneous | |
| Symlin Injectable Product | subcutaneously | |
| insulin human, isophane 70 UNT/ML / Regular Insulin, Human 30 UNT/ML [Humulin] | subcu | |
| Glucotrol Oral Product | PO | |
| 24 HR Metformin hydrochloride 750 MG Extended Release Oral Tablet | PO | |
| Linagliptin 2.5 MG / Metformin hydrochloride 1000 MG Oral Tablet [Jentadueto] | by mouth | |
| 1.5 ML Insulin Glargine 300 UNT/ML Pen Injector [Toujeo] | SUBCUTANEOUS | |
| exenatide 0.01 MG/ACTUAT [Byetta] | sq | |
| Tradjenta Oral Product | by mouth | |
| Metformin 500 MG / ropinirole 2 MG Oral Tablet | po | |
| Insulin, Zinc, Human | | |
| Insulin Lispro Injectable Solution | subcutaneously | |
| Metformin hydrochloride 500 MG / pioglitazone 15 MG Oral Tablet [Actoplus Met] | by mouth | |
| insulin detemir 100 UNT/ML Injectable Solution [Levemir] | subcutaneous | |
| Regular Insulin, Human 15 UNT/ML | | |
| liraglutide | | |
| Insulin, Aspart, Human | sc | |

| | | |
|---|---|---|
| Insulin, Aspart, Human | gt | |
| Acarbose 100 MG [Precose] | | |
| insulin human, isophane 70 UNT/ML / Regular Insulin, Human 30 UNT/ML [Novolin] | Subcutaneous | |
| 3 ML Insulin, Glulisine, Human 100 UNT/ML Pen Injector [Apidra] | SubQ | |
| exenatide 0.01 MG/ACTUAT | | |
| 24 HR Metformin hydrochloride 500 MG / saxagliptin 5 MG Extended Release Oral Tablet [Kombiglyze] | by mouth | |
| Metformin Oral Solution | p.o. | |
| insulin beef, isophane 40 UNT/ML | | |
| Regular Insulin, Human 8 UNT | | |
| rosiglitazone 4 MG Oral Tablet | PO | |
| 3 ML liraglutide 6 MG/ML Pen Injector [Victoza] | | |
| Insulin, Aspart, Human | oral | |
| Glyburide 2.5 MG / Metformin hydrochloride 500 MG Oral Tablet [Glucovance] | PO | |
| Actoplus Met Oral Product | po | |
| rosiglitazone 4 MG Oral Tablet [Avandia] | ORAL | |
| Regular Insulin, Human 30 UNT/ML | | |
| 3 ML Insulin, Aspart Protamine, Human 70 UNT/ML / Insulin, Aspart, Human 30 UNT/ML Pen Injector [NovoLog Mix] | subcut | |
| Metformin hydrochloride 850 MG / pioglitazone 15 MG Oral Tablet [Actoplus Met] | | |
| Glyburide 5 MG Oral Tablet [Micronase] | po | |
| Glyburide 1.5 MG | | |
| Insulin Lispro Injectable Solution | SQ | |
| Insulin Lispro 100 UNT/ML Injectable Solution [Humalog] | | |
| glimepiride 2 MG Oral Tablet [Amaryl] | p.o. | |
| Avandia Oral Product | PO | |
| Insulin Lispro 100 UNT/ML Injectable Solution | Subcutaneous | |
| glimepiride 4 MG Oral Tablet [Amaryl] | | 0 |
| sitagliptin 100 MG Oral Tablet | by mouth | |
| glimepiride 4 MG Oral Tablet | p.o. | |
| Regular Insulin, Human 100 UNT/ML Injectable Solution [Novolin R] | subq | |
| Prandin Oral Product | po | |
| Insulin Lispro | orally | |
| dapagliflozin 10 MG Oral Tablet [Farxiga] | ORAL | |
| insulin human, isophane 70 UNT/ML / Regular Insulin, Human 30 UNT/ML [Novolin] | subcut. | |

| | |
|---|---|
| 3 ML liraglutide 6 MG/ML Pen Injector [Victoza] | SQ |
| Glipizide 10 MG Extended Release Oral Tablet | |
| Insulin Glargine | po |
| Glipizide 2.5 MG [Glucotrol] | by mouth |
| 24 HR Metformin hydrochloride 750 MG Extended Release Oral Tablet [Glucophage] | |
| miglitol 25 MG [Glyset] | |
| Acarbose 50 MG Oral Tablet [Precose] | po |
| pramlintide acetate 0.6 MG/ML Injectable Solution [Symlin] | |
| 3 ML Insulin Glargine 100 UNT/ML Pen Injector [Basaglar] | subcutaneous |
| Insulin Glargine Injectable Solution | SQ |
| insulin pork, isophane 100 UNT/ML Injectable Suspension [NPH Iletin II] | subcu |
| Apidra Injectable Product | subcutaneously |
| Metformin hydrochloride 1000 MG / sitagliptin 50 MG Oral Tablet [Janumet] | oral |
| Regular Insulin, Human 20 UNT/ML | subcut |
| insulin human, isophane 32 UNT/ML | |
| 3 ML Insulin Lispro 25 UNT/ML / Insulin, Protamine Lispro, Human 75 UNT/ML Pen Injector [Humalog Mix] | SUBCUTANEOUS |
| glimepiride 4 MG Oral Tablet [Amaryl] | p.o. |
| insulin pork, isophane 100 UNT/ML Injectable Suspension [NPH Iletin II] | subq |
| Simvastatin 10 MG / sitagliptin 50 MG [Juvisync] | |
| Bromocriptine 2.5 MG Oral Tablet [Parlodel] | po |
| Insulin, Aspart Protamine, Human 70 UNT/ML / Insulin, Aspart, Human 30 UNT/ML Injectable Suspension [NovoLog Mix] | sq |
| Glipizide 10 MG Oral Tablet [Glucotrol] | p.o. |
| glimepiride 1 MG Oral Tablet | |
| glimepiride 1 MG Oral Tablet | PO |
| Insulin Lispro | subcu |
| Glipizide 5 MG Extended Release Oral Tablet | |
| Insulin Glargine | subcutaneously |
| 24 HR Glipizide 10 MG Extended Release Oral Tablet [Glucotrol] | by mouth |
| Regular Insulin, Human 100 UNT/ML Injectable Solution [Humulin R] | subq |
| Metformin | NPO |
| Ultralente Insulin, Human 100 UNT/ML Injectable Suspension | |
| pioglitazone 45 MG Oral Tablet [Actos] | oral |

| | |
|---|---|
| Regular Insulin, Human 500 UNT/ML Injectable Solution [Humulin R] | SUBCUTANEOUS |
| Insulin Lispro 25 UNT/ML / Insulin, Protamine Lispro, Human 75 UNT/ML Injectable Suspension [Humalog Mix] | SUBCUT |
| Linagliptin 5 MG Oral Tablet [Tradjenta] | po |
| Glucophage Pill | |
| Insulin Lispro 25 UNT/ML | subcutaneous |
| Insulin Lispro 50 UNT/ML / Insulin, Protamine Lispro, Human 50 UNT/ML Injectable Suspension [Humalog Mix] | subcut |
| dulaglutide 3 MG/ML [Trulicity] | |
| empagliflozin 10 MG Oral Tablet [Jardiance] | oral |
| Glyburide 5 MG Oral Tablet [Micronase] | by mouth |
| Regular Insulin, Human 100 UNT/ML Injectable Solution [Novolin R] | SUBCUTANEOUS |
| insulin detemir 100 UNT/ML Injectable Solution [Levemir] | |
| 24 HR Glipizide 5 MG Extended Release Oral Tablet | po |
| rosiglitazone 4 MG Oral Tablet | orally |
| pioglitazone 30 MG Oral Tablet | po |
| 3 ML Insulin, Aspart Protamine, Human 70 UNT/ML / Insulin, Aspart, Human 30 UNT/ML Pen Injector [NovoLog Mix] | subcutaneous |
| Insulin Glargine 100 UNT/ML Injectable Solution [Lantus] | SUBQ |
| Glyburide 1.25 MG Oral Tablet | orally |
| pioglitazone 45 MG Oral Tablet | po |
| Insulin, Aspart, Human | drip |
| Acarbose 25 MG Oral Tablet | by mouth |
| nateglinide 120 MG Oral Tablet | po |
| Farxiga Oral Product | by mouth |
| 0.5 ML dulaglutide 1.5 MG/ML Auto-Injector [Trulicity] | |
| insulin pork, isophane 80 UNT/ML | subcut |
| 3 ML Insulin Glargine 100 UNT/ML Pen Injector [Lantus] | subcutaneous |
| Insulin, Aspart Protamine, Human 70 UNT/ML / Insulin, Aspart, Human 30 UNT/ML [NovoLog Mix] | subcut |
| Glumetza Oral Product | by mouth |
| Insulin Lispro 25 UNT/ML | subcut. |
| Glipizide 2.5 MG Oral Tablet | oral |
| Bromocriptine 2.5 MG Oral Tablet [Parlodel] | ORAL |
| Insulin Lispro | SUBCUT. |
| Regular Insulin, Human | subcutaneously |
| Glipizide 5 MG Oral Tablet [Glucotrol] | orally |
| Acarbose 100 MG Oral Tablet | by mouth |

| | | |
|---|---|---|
| Insulin, Aspart, Human | by mouth | |
| Regular Insulin, Human 20 UNT/ML | | |
| Insulin, Aspart Protamine, Human 70 UNT/ML / Insulin, Aspart, Human 30 UNT/ML Injectable Suspension [NovoLog Mix] | SUBCUTANEOUS | |
| exenatide 2 MG Injection [Bydureon] | subcutaneously | |
| glimepiride Oral Product | oral | |
| rosiglitazone 4 MG Oral Tablet [Avandia] | p.o. | |
| nateglinide 120 MG Oral Tablet | orally | |
| 0.65 ML exenatide 3.08 MG/ML Pen Injector [Bydureon] | SUBCUTANEOUS | |
| Insulin, Regular, Pork 40 UNT/ML Injectable Solution | | |
| nateglinide 120 MG Oral Tablet [Starlix] | orally | |
| Metformin hydrochloride 100 MG/ML Oral Solution [Riomet] | | |
| liraglutide 6 MG/ML [Victoza] | sq | |
| 3 ML Insulin, Aspart, Human 100 UNT/ML Pen Injector [NovoLog] | subcutaneous | |
| pioglitazone Oral Tablet [Actos] | by mouth | |
| Insulin Lispro 100 UNT/ML Injectable Solution | | |
| Regular Insulin, Human 100 UNT/ML Injectable Solution [Novolin R] | INJECTION | |
| Metformin hydrochloride 850 MG Oral Tablet | p.o. | |
| Glucophage Oral Product | p.o. | |
| 3 ML liraglutide 6 MG/ML Pen Injector [Saxenda] | subcutaneous | |
| 3 ML Insulin, Aspart, Human 100 UNT/ML Cartridge [NovoLog] | | |
| Lantus Injectable Product | Subcutaneously | |
| Regular Insulin, Human 100 UNT/ML Injectable Solution [Humulin R] | INJECTION | |
| Byetta Injectable Product | | |
| Insulin Lispro 100 UNT/ML Injectable Solution [Humalog] | | 0 |
| Insulin Lispro 25 UNT/ML / Insulin, Protamine Lispro, Human 75 UNT/ML [Humalog Mix] | subcut | |
| Acarbose 100 MG | | |
| rosiglitazone 4 MG Oral Tablet [Avandia] | orally | |
| Metformin Oral Tablet | po | |
| Glyburide 3 MG Oral Tablet | PO | |
| rosiglitazone 4 MG Oral Tablet | by mouth | |
| Regular Insulin, Human 12 UNT | subcut | |
| Januvia Oral Product | oral | |
| Metformin hydrochloride 500 MG Extended Release Oral Tablet [Fortamet] | po | |
| pioglitazone 15 MG Oral Tablet [Actos] | orally | |
| saxagliptin 5 MG Oral Tablet [Onglyza] | orally | |

| | |
|---|---|
| Insulin Lispro 25 UNT/ML / Insulin, Protamine Lispro, Human 75 UNT/ML [Humalog Mix] | SQ |
| Onglyza Oral Product | by mouth |
| Regular Insulin, Human 100 UNT/ML Injectable Solution [Novolin R] | SQ |
| sitagliptin 50 MG Oral Tablet | orally |
| canagliflozin 150 MG / Metformin hydrochloride 500 MG Oral Tablet [Invokamet] | by mouth |
| miglitol 50 MG Oral Tablet [Glyset] | po |
| Glyburide 1.25 MG Oral Tablet | PO |
| Regular Insulin, Human 12 UNT | |
| Insulin Lispro 50 UNT/ML | SQ |
| Glucophage Oral Product | oral |
| repaglinide 1 MG | |
| Regular Insulin, Human 500 UNT/ML Injectable Solution [Humulin R] | SQ |
| pioglitazone 15 MG Oral Tablet | po |
| insulin, regular, beef-pork 40 UNT/ML Inhalant Solution | |
| Regular Insulin, Human 100 UNT/ML Injectable Solution [Novolin R] | sq |
| 60 ACTUAT exenatide 0.01 MG/ACTUAT Pen Injector [Byetta] | by mouth |
| sitagliptin 100 MG Oral Tablet [Januvia] | p.o. |
| insulin human, isophane 70 UNT/ML / Regular Insulin, Human 30 UNT/ML [Novolin] | SUBCUT |
| glimepiride 3 MG Oral Tablet [Amaryl] | po |
| 3 ML Insulin, Glulisine, Human 100 UNT/ML Pen Injector [Apidra] | SUBCUTANEOUS |
| glimepiride 3 MG [Amaryl] | |
| Regular Insulin, Human 6 UNT/ML | |
| exenatide 0.005 MG/ACTUAT [Byetta] | subq |
| Regular Insulin, Human 100 UNT/ML Injectable Solution [Humulin R] | subcut |
| Insulin Glargine 100 UNT/ML [Lantus] | sub q |
| 24 HR Glipizide 10 MG Extended Release Oral Tablet | orally |
| insulin human, isophane 36 UNT/ML | |
| Insulin Lispro 50 UNT/ML | subcut. |
| insulin human, isophane 40 UNT/ML | subcut. |
| rosiglitazone 2 MG Oral Tablet [Avandia] | PO |
| nateglinide Oral Product | by mouth |
| Metformin hydrochloride 500 MG / sitagliptin 50 MG Oral Tablet [Janumet] | orally |

| | | |
|---|---|---|
| 3 ML insulin human, isophane 100 UNT/ML Prefilled Syringe [Novolin N] | subcutaneous | |
| Metformin hydrochloride 1000 MG / sitagliptin 100 MG [Janumet] | by mouth | |
| insulin human, isophane 70 UNT/ML / Regular Insulin, Human 30 UNT/ML [Novolin] | SubQ | |
| 60 ACTUAT exenatide 0.01 MG/ACTUAT Pen Injector [Byetta] | subcutaneously | |
| Modified 24 HR Metformin hydrochloride 1000 MG Extended Release Oral Tablet [Glumetza] | ORAL | |
| Metformin hydrochloride 500 MG / rosiglitazone 2 MG Oral Tablet [Avandamet] | po | |
| Humalog Injectable Product | SUBCUT | |
| glimepiride 3 MG | | |
| Glipizide 2.5 MG [Glucotrol] | po | |
| rosiglitazone 4 MG | | |
| Diabeta Oral Product | po | |
| Humulin Injectable Product | subcutaneously | |
| Insulin Glargine | SC | |
| Glipizide 5 MG Oral Tablet | | 0 |
| Bromocriptine 5 MG Oral Capsule [Parlodel] | by mouth | |
| Regular Insulin, Human 500 UNT/ML Injectable Solution [Humulin R] | subq | |
| Metformin hydrochloride 750 MG Extended Release Oral Tablet [Glucophage] | PO | |
| Regular Insulin, Human 100 UNT/ML Injectable Solution [Humulin R] | INTRAVENOUS | |
| glimepiride 2 MG Oral Tablet [Amaryl] | | 0 |
| insulin human, isophane 60 UNT/ML | SQ | |
| Insulin Glargine 100 UNT/ML [Lantus] | sc | |
| pioglitazone 45 MG Oral Tablet [Actos] | p.o. | |
| Insulin Glargine 100 UNT/ML Injectable Solution [Lantus] | Subq | |
| Metformin hydrochloride 1000 MG Oral Tablet [Glucophage] | Orally | |
| Glyburide 1.25 MG [Diabeta] | | |
| Bromocriptine Oral Tablet [Parlodel] | by mouth | |
| sitagliptin 50 MG | | |
| exenatide 0.005 MG/ACTUAT [Byetta] | subcut. | |
| Glipizide 2.5 MG / Metformin hydrochloride 250 MG Oral Tablet | | |
| Glyburide 5 MG Oral Tablet | | |
| Glipizide Extended Release Oral Tablet | by mouth | |
| nateglinide 120 MG Oral Tablet [Starlix] | oral | |

| | |
|---|---|
| Regular Insulin, Human 100 UNT/ML Injectable Solution [Novolin R] | intravenously |
| 1.5 ML insulin human, isophane 70 UNT/ML / Regular Insulin, Human 30 UNT/ML Prefilled Syringe | |
| Glyburide 2.5 MG [Micronase] | |
| Glyburide 2.5 MG / Metformin hydrochloride 500 MG [Glucovance] | |
| 0.5 ML dulaglutide 1.5 MG/ML Auto-Injector [Trulicity] | SUBCUTANEOUS |
| Metformin hydrochloride 500 MG / sitagliptin 50 MG Oral Tablet [Janumet] | oral |
| insulin human, isophane 70 UNT/ML / Regular Insulin, Human 30 UNT/ML [Humulin] | sub q |
| insulin human, isophane 100 UNT/ML Injectable Suspension | Subcutaneous |
| insulin detemir 100 UNT/ML [Levemir] | SQ |
| 3 ML insulin human, isophane 70 UNT/ML / Regular Insulin, Human 30 UNT/ML Pen Injector [Humulin] | SUBCUTANEOUS |
| glimepiride 6 MG | |
| Insulin, Aspart, Human 100 UNT/ML Cartridge [NovoLog] | SC |
| 3 ML insulin degludec 200 UNT/ML Pen Injector [Tresiba] | SUBCUTANEOUS |
| exenatide 0.01 MG/ACTUAT [Byetta] | Subcut |
| 3 ML Insulin, Aspart, Human 100 UNT/ML Cartridge [NovoLog] | by mouth |
| Insulin Lispro 25 UNT/ML | SQ |
| Glipizide | PO |
| 3 ML insulin human, isophane 100 UNT/ML Pen Injector [Humulin N] | SUBCUTANEOUS |
| Insulin Glargine 100 UNT/ML [Lantus] | by mouth |
| 60 ACTUAT exenatide 0.005 MG/ACTUAT Pen Injector [Byetta] | po |
| Glyburide 1.5 MG Oral Tablet | po |
| Insulin Lispro Cartridge [Humalog] | sq |
| Metformin 250 MG Oral Tablet | orally |
| 3 ML Insulin, Aspart Protamine, Human 70 UNT/ML / Insulin, Aspart, Human 30 UNT/ML Pen Injector [NovoLog Mix] | subq |
| Glipizide / Metformin Oral Tablet [Metaglip] | by mouth |
| 3 ML liraglutide 6 MG/ML Pen Injector [Victoza] | by mouth |
| 0.65 ML exenatide 3.08 MG/ML Pen Injector [Bydureon] | |
| Metformin hydrochloride 1000 MG / sitagliptin 50 MG Oral Tablet | po |
| insulin human, isophane 70 UNT/ML / Regular Insulin, Human 30 UNT/ML [Novolin] | SC |
| 3 ML Insulin Glargine 100 UNT/ML Pen Injector [Basaglar] | SUBCUTANEOUS |

| | | |
|---|---|---|
| repaglinide 2 MG Oral Tablet [Prandin] | PO | |
| Glyburide 1.25 MG Oral Tablet | ORAL | |
| Insulin, Aspart, Human | NPO. | |
| rosiglitazone 4 MG Oral Tablet | po | |
| 60 ACTUAT exenatide 0.01 MG/ACTUAT Pen Injector [Byetta] | SQ | |
| Insulin, Aspart, Human 100 UNT/ML Injectable Solution [NovoLog] | Subcutaneously | |
| Glipizide 2.5 MG Extended Release Oral Tablet | PO | |
| insulin, regular, beef-pork 40 UNT/ML Inhalant Solution | subcut | |
| Novolin R Injectable Product | | |
| Glyburide 1.25 MG / Metformin hydrochloride 250 MG Oral Tablet [Glucovance] | | |
| empagliflozin 25 MG Oral Tablet [Jardiance] | | |
| Glyburide 6 MG Oral Tablet | orally | |
| insulin human, isophane 40 UNT/ML | subcutaneous | |
| Linagliptin 5 MG Oral Tablet | by mouth | |
| 3 ML insulin human, isophane 70 UNT/ML / Regular Insulin, Human 30 UNT/ML Pen Injector [Humulin] | Subcutaneous | |
| Avandia Oral Product | by mouth | |
| pramlintide acetate 0.6 MG/ML [Symlin] | | |
| sitagliptin 100 MG Oral Tablet [Januvia] | | |
| sitagliptin 100 MG Oral Tablet [Januvia] | | 0 |
| Metformin hydrochloride 850 MG Oral Tablet [Glucophage] | orally | |
| 3 ML insulin degludec 100 UNT/ML Pen Injector [Tresiba] | SUBCUTANEOUS | |
| insulin human, isophane 50 UNT/ML / Regular Insulin, Human 50 UNT/ML [Humulin] | | |
| Insulin Glargine | SUBCUT. | |
| canagliflozin 300 MG Oral Tablet [Invokana] | ORAL | |
| Metformin | topical | |
| 24 HR Glipizide 2.5 MG Extended Release Oral Tablet [Glucotrol] | Oral | |
| glimepiride 4 MG Oral Tablet [Amaryl] | Orally | |
| Regular Insulin, Human 100 UNT/ML Injectable Solution [Novolin R] | subcut | |
| Linagliptin 5 MG Oral Tablet [Tradjenta] | orally | |
| Novolin Injectable Product | subcutaneously | |
| insulin pork, isophane 70 UNT/ML / Insulin, Regular, Pork 30 UNT/ML Injectable Suspension | subcutaneously | |
| exenatide 0.005 MG/ACTUAT [Byetta] | subcutaneously | |
| exenatide 0.005 MG/ACTUAT [Byetta] | sq | |
| 24 HR Glipizide 5 MG Extended Release Oral Tablet | by mouth | |

| | | |
|---|---|---|
| saxagliptin 5 MG | | |
| Glipizide 10 MG Oral Tablet [Glucotrol] | | 0 |
| 24 HR Glipizide 5 MG Extended Release Oral Tablet | orally | |
| pramlintide acetate 0.6 MG/ML Injectable Solution [Symlin] | subq | |
| glimepiride 2 MG Oral Tablet | p.o. | |
| Metformin hydrochloride 1000 MG Oral Tablet | BY MOUTH | |
| Metformin hydrochloride 500 MG Extended Release Oral Tablet [Glucophage] | | |
| canagliflozin 100 MG Oral Tablet [Invokana] | ORAL | |
| Humalog Injectable Product | SQ | |
| insulin human, isophane 90 UNT/ML | | |
| Glyburide 1.25 MG / Metformin hydrochloride 250 MG Oral Tablet [Glucovance] | po | |
| 60 ACTUAT exenatide 0.005 MG/ACTUAT Pen Injector [Byetta] | subcut | |
| Metformin hydrochloride 850 MG / pioglitazone 15 MG Oral Tablet [Actoplus Met] | ORAL | |
| 3 ML Insulin, Aspart Protamine, Human 70 UNT/ML / Insulin, Aspart, Human 30 UNT/ML Pen Injector [NovoLog Mix] | subcut. | |
| Glipizide 5 MG Oral Tablet [Glucotrol] | oral | |
| Acarbose 25 MG Oral Tablet [Precose] | po | |
| glimepiride Oral Product | PO | |
| Insulin Glargine 100 UNT/ML [Lantus] | SC | |
| insulin human, isophane 40 UNT/ML | subq | |
| 3 ML Insulin Lispro 100 UNT/ML Pen Injector [Humalog] | | 0 |
| pioglitazone 30 MG Oral Tablet [Actos] | | |
| exenatide 2 MG Injection [Bydureon] | SUBCUTANEOUS | |
| Insulin Lispro 25 UNT/ML | subq | |
| rosiglitazone 8 MG Oral Tablet [Avandia] | p.o. | |
| exenatide 2 MG Injection [Bydureon] | | |
| insulin human, isophane 16 UNT/ML | subcut | |
| exenatide 0.005 MG/ACTUAT | | |
| Glipizide 2.5 MG Extended Release Oral Tablet | | |
| Metformin hydrochloride 500 MG / pioglitazone 15 MG Oral Tablet [Actoplus Met] | | |
| albiglutide 60 MG/ML [Tanzeum] | subcutaneous | |
| insulin detemir Injectable Product | | |
| miglitol 50 MG [Glyset] | | |
| 0.5 ML dulaglutide 3 MG/ML Auto-Injector [Trulicity] | SUBCUTANEOUS | |
| Metformin hydrochloride 500 MG / rosiglitazone 2 MG Oral Tablet [Avandamet] | by mouth | |

| | |
|---|---|
| Simvastatin 20 MG / sitagliptin 50 MG Oral Tablet | by mouth |
| Insulin Lispro 50 UNT/ML | sq |
| Amaryl Oral Product | BY MOUTH |
| Actos Oral Product | oral |
| Insulin Glargine 100 UNT/ML Injectable Solution [Lantus] | subcut |
| 0.65 ML exenatide 3.08 MG/ML Pen Injector [Bydureon] | subcutaneous |
| Glyburide 1 MG Oral Tablet | by mouth |
| Acarbose 100 MG Oral Tablet [Precose] | po |
| insulin human, isophane 60 UNT/ML | subcutaneous |
| Glyburide Pill | |
| 3 ML insulin human, isophane 100 UNT/ML Prefilled Syringe [Novolin N] | Subcutaneous |
| glimepiride | subcutaneously |
| Linagliptin 5 MG Oral Tablet [Tradjenta] | ORAL |
| insulin human, isophane 70 UNT/ML / Regular Insulin, Human 30 UNT/ML [Humulin] | Subcutaneous |
| Metformin hydrochloride 850 MG Oral Tablet [Glucophage] | |
| 3 ML Insulin, Aspart, Human 100 UNT/ML Cartridge [NovoLog] | |
| Metformin Oral Solution | Oral |
| Metformin hydrochloride 500 MG / sitagliptin 50 MG Oral Tablet [Janumet] | PO |
| Glyburide 1.5 MG Oral Tablet | by mouth |
| exenatide 0.005 MG/ACTUAT [Byetta] | po |
| Micronase Oral Product | po |
| Glipizide 2.5 MG Extended Release Oral Tablet | po |
| Metformin hydrochloride 1000 MG / sitagliptin 50 MG Oral Tablet [Janumet] | |
| Insulin Glargine | sc |
| Insulin Glargine | subQ |
| Insulin, Aspart, Human | subcu |
| 24 HR Glipizide 2.5 MG Extended Release Oral Tablet [Glucotrol] | oral |
| Acarbose | |
| sitagliptin Oral Tablet | by mouth |
| NovoLog Injectable Product | Subcutaneously |
| Insulin Lispro | po |
| Insulin Lispro 25 UNT/ML / Insulin, Protamine Lispro, Human 75 UNT/ML Injectable Suspension [Humalog Mix] | SUBCUTANEOUS |
| Insulin Lispro 50 UNT/ML | subq |
| Insulin, Zinc, Human 100 UNT/ML Injectable Suspension [Novolin L] | |

| | |
|---|---|
| insulin human, isophane 32 UNT/ML | subcut |
| Glyburide 2.5 MG / Phenformin 25 MG Oral Tablet | |
| Humulin N Injectable Product | |
| Insulin Lispro 25 UNT/ML / Insulin, Protamine Lispro, Human 75 UNT/ML Injectable Suspension [Humalog Mix] | sq |
| Bromocriptine 5 MG Oral Capsule | orally |
| Insulin, Aspart, Human | subcut. |
| Glyburide 5 MG / Metformin 1000 MG Oral Tablet | |
| Glyburide 2.5 MG Oral Tablet [Diabeta] | PO |
| Insulin, Aspart Protamine, Human 70 UNT/ML / Insulin, Aspart, Human 30 UNT/ML [NovoLog Mix] | sq |
| 60 ACTUAT exenatide 0.01 MG/ACTUAT Pen Injector [Byetta] | subq |
| Metformin hydrochloride 1000 MG Extended Release Oral Tablet [Fortamet] | Oral |
| Glyburide 2.5 MG / Metformin hydrochloride 500 MG Oral Tablet | po |
| pioglitazone 15 MG Oral Tablet [Actos] | oral |
| Lantus Injectable Product | SUBCUTANEOUSLY |
| miglitol 25 MG Oral Tablet [Glyset] | by mouth |
| 3 ML insulin human, isophane 100 UNT/ML Pen Injector [Humulin N] | |
| exenatide 2 MG [Bydureon] | subcutaneous |
| Insulin, Aspart Protamine, Human 70 UNT/ML / Insulin, Aspart, Human 30 UNT/ML Injectable Suspension [NovoLog Mix] | subcut. |
| Glynase Oral Product | by mouth |
| Regular Insulin, Human | VERIFY |
| glimepiride 2 MG / pioglitazone 30 MG [Duetact] | |
| Simvastatin 20 MG / sitagliptin 100 MG Oral Tablet | by mouth |
| Glyburide 1.25 MG / Metformin hydrochloride 250 MG Oral Tablet [Glucovance] | by mouth |
| Glyburide 6 MG Oral Tablet | ORAL |
| exenatide 0.01 MG/ACTUAT [Byetta] | SUBCUT |
| Prandin Oral Product | by mouth |
| 24 HR dapagliflozin 5 MG / Metformin hydrochloride 1000 MG Extended Release Oral Tablet [Xigduo] | by mouth |
| Regular Insulin, Human 500 UNT/ML Injectable Solution [Humulin R] | subcut |
| Insulin Lispro 100 UNT/ML | |
| Glucophage Oral Product | orally |
| Regular Insulin, Human | Subcutaneous |

| | |
|---|---|
| 24 HR Metformin hydrochloride 500 MG Extended Release Oral Tablet | BY MOUTH |
| Chlorpropamide 100 MG Oral Tablet | by mouth |
| Regular Insulin, Human 50 UNT/ML | |
| insulin human, isophane 70 UNT/ML / Regular Insulin, Human 30 UNT/ML Injectable Suspension [Humulin] | subq |
| nateglinide 60 MG Oral Tablet [Starlix] | PO |
| insulin human, isophane 100 UNT/ML Injectable Suspension [Humulin N] | sq |
| Regular Insulin, Human 100 UNT/ML Injectable Solution | subcutaneously |
| albiglutide 100 MG/ML [Tanzeum] | subcutaneous |
| nateglinide 60 MG Oral Tablet [Starlix] | ORAL |
| Lispro PRC Injectable Product | subcut |
| insulin detemir Injectable Product | subcutaneously |
| Metformin 400 MG | by mouth |
| insulin pork, isophane 100 UNT/ML Injectable Suspension [NPH Iletin II] | NPO |
| Glyburide 5 MG / Metformin 1000 MG Oral Tablet | by mouth |
| rosiglitazone 8 MG Oral Tablet [Avandia] | ORAL |
| Insulin Lispro | MISC |
| empagliflozin 25 MG / Linagliptin 5 MG Oral Tablet [Glyxambi] | by mouth |
| 3 ML Insulin, Aspart Protamine, Human 70 UNT/ML / Insulin, Aspart, Human 30 UNT/ML Pen Injector [NovoLog Mix] | |
| Linagliptin 5 MG Oral Tablet [Tradjenta] | PO |
| Insulin Lispro Cartridge [Humalog] | subQ |
| Jardiance Oral Product | by mouth |
| insulin human, isophane 75 UNT/ML | subcut |
| Metformin hydrochloride 500 MG / pioglitazone 15 MG Oral Tablet | by mouth |
| Metformin Oral Tablet [Glucophage] | |
| Metformin hydrochloride 500 MG / pioglitazone 15 MG Oral Tablet [Actoplus Met] | po |
| 0.5 ML dulaglutide 1.5 MG/ML Auto-Injector [Trulicity] | subcutaneous |
| Glipizide 2.5 MG Extended Release Oral Tablet | orally |
| Metformin | nostril |
| insulin pork, isophane 100 UNT/ML Injectable Suspension [NPH Iletin II] | drip |
| sitagliptin 25 MG Oral Tablet [Januvia] | po |
| Metformin hydrochloride 100 MG/ML Oral Solution | PO |
| insulin human, isophane 34 UNT/ML | subcut |

| | |
|---|---|
| insulin human, isophane 70 UNT/ML / Regular Insulin, Human 30 UNT/ML [Humulin] | SC |
| nateglinide Oral Tablet [Starlix] | by mouth |
| Metformin hydrochloride 1000 MG / saxagliptin 2.5 MG [Kombiglyze] | by mouth |
| 60 ACTUAT exenatide 0.01 MG/ACTUAT Pen Injector [Byetta] | subcut |
| insulin human, isophane 34 UNT/ML | |
| Glipizide 5 MG / Metformin hydrochloride 500 MG Oral Tablet [Metaglip] | by mouth |
| Metformin hydrochloride 1000 MG / sitagliptin 50 MG Oral Tablet [Janumet] | PO |
| pioglitazone 15 MG Oral Tablet [Actos] | p.o. |
| 0.5 ML albiglutide 60 MG/ML Pen Injector [Tanzeum] | |
| Insulin Lispro | Subcutaneous |
| Glipizide Pill | |
| Insulin Lispro 25 UNT/ML / Insulin, Protamine Lispro, Human 75 UNT/ML [Humalog Mix] | sq |
| Insulin Lispro 25 UNT/ML | sq |
| canagliflozin 300 MG Oral Tablet [Invokana] | po |
| Insulin Lispro 50 UNT/ML | iv |
| Bromocriptine 1 MG Oral Tablet [Parlodel] | by mouth |
| repaglinide 2 MG Oral Tablet [Prandin] | |
| Metformin 400 MG | |
| Glyburide 6 MG Oral Tablet [Glynase] | PO |
| Glyburide 2.5 MG / Metformin 400 MG Oral Tablet | po |
| Glyburide 1.25 MG Oral Tablet | oral |
| Glyburide 2.5 MG / Metformin hydrochloride 500 MG Oral Tablet [Glucovance] | ORAL |
| Insulin Lispro 100 UNT/ML [Humalog] | |
| insulin human, isophane 70 UNT/ML / Regular Insulin, Human 30 UNT/ML [Novolin] | subcu |
| glimepiride 4 MG / rosiglitazone 4 MG Oral Tablet [Avandaryl] | |
| insulin human, isophane 100 UNT/ML Injectable Suspension | |
| Regular Insulin, Human 100 UNT/ML Injectable Solution [Humulin S] | |
| exenatide 2 MG [Bydureon] | subq |
| Insulin Lispro Cartridge [Humalog] | SUBQ |
| sitagliptin | |
| rosiglitazone 8 MG Oral Tablet | po |
| nateglinide | |

| | | |
|---|---|---|
| insulin pork, isophane 70 UNT/ML / Insulin, Regular, Pork 30 UNT/ML Injectable Suspension | subcutaneous | |
| insulin human, isophane 100 UNT/ML Pen Injector [Humulin N] | subcutaneous | |
| Insulin Lispro Injectable Solution | SUBCUT | |
| insulin human, isophane 40 UNT/ML | SQ | |
| Trulicity Injectable Product | subcutaneously | |
| Acarbose Oral Tablet | by mouth | |
| 3 ML liraglutide 6 MG/ML Pen Injector [Victoza] | | 0 |
| Glyburide 5 MG / Metformin hydrochloride 500 MG Oral Tablet [Glucovance] | orally | |
| Bromocriptine 2.5 MG Oral Tablet [Parlodel] | PO | |
| Chlorpropamide 100 MG | | |
| exenatide 0.01 MG/ACTUAT [Byetta] | Subcutaneous | |
| 24 HR Metformin hydrochloride 500 MG Extended Release Oral Tablet | orally | |
| Glyburide 5 MG / Metformin hydrochloride 500 MG Oral Tablet | orally | |
| Regular Insulin, Human 500 UNT/ML Injectable Solution [Humulin R] | INJECTION | |
| exenatide 0.01 MG/ACTUAT [Byetta] | sc | |
| insulin human, isophane 70 UNT/ML / Regular Insulin, Human 30 UNT/ML [Humulin] | sc | |
| 24 HR Glipizide 10 MG Extended Release Oral Tablet [Glucotrol] | | 0 |
| insulin human, isophane 70 UNT/ML / Regular Insulin, Human 30 UNT/ML [Humulin] | by mouth | |
| Metformin hydrochloride 500 MG Oral Tablet [Glucophage] | Oral | |
| Bromocriptine 5 MG [Parlodel] | | |
| 24 HR Glipizide 5 MG Extended Release Oral Tablet [Glucotrol] | by mouth | |
| rosiglitazone 8 MG Oral Tablet | orally | |
| sitagliptin 100 MG Oral Tablet [Januvia] | BY MOUTH | |
| Metformin hydrochloride 500 MG Extended Release Oral Tablet [Glumetza] | | |
| exenatide 0.005 MG/ACTUAT [Byetta] | sc | |
| 3 ML insulin detemir 100 UNT/ML Pen Injector [Levemir] | SQ | |
| pioglitazone 15 MG Oral Tablet [Actos] | | |
| sitagliptin 25 MG Oral Tablet [Januvia] | orally | |
| insulin detemir 100 UNT/ML Injectable Solution [Levemir] | | |
| exenatide 3.08 MG/ML [Bydureon] | | |
| exenatide 0.005 MG/ACTUAT [Byetta] | Subcut | |
| insulin human, isophane 85 UNT/ML | | |
| Insulin, Aspart, Human | subQ | |

| | |
|---|---|
| Glyburide 2.5 MG Oral Tablet [Micronase] | po |
| dapagliflozin 5 MG Oral Tablet [Farxiga] | oral |
| Insulin, Aspart, Human 100 UNT/ML Cartridge [NovoLog] | sub q |
| nateglinide 120 MG Oral Tablet [Starlix] | |
| Bromocriptine 2.5 MG Oral Tablet [Parlodel] | oral |
| Linagliptin 2.5 MG / Metformin hydrochloride 500 MG Oral Tablet [Jentadueto] | by mouth |
| 24 HR Metformin hydrochloride 500 MG Extended Release Oral Tablet | |
| Glucotrol Oral Product | BY MOUTH |
| 24 HR Glipizide 2.5 MG Extended Release Oral Tablet [Glucotrol] | PO |
| Lispro PRC Injectable Product | subcutaneously |
| miglitol 50 MG Oral Tablet [Glyset] | by mouth |
| Metformin hydrochloride 500 MG / rosiglitazone 4 MG [Avandamet] | |
| Regular Insulin, Human 100 UNT/ML Injectable Solution [Novolin R] | INTRAVENOUS |
| Insulin Lispro 50 UNT/ML / Insulin, Protamine Lispro, Human 50 UNT/ML Injectable Suspension [Humalog Mix] | subq |
| glimepiride | PO |
| Glipizide 5 MG / Metformin hydrochloride 500 MG [Metaglip] | |
| Metformin hydrochloride 850 MG Oral Tablet [Glucophage] | ORAL |
| miglitol 25 MG Oral Tablet [Glyset] | po |
| repaglinide 2 MG | |
| repaglinide 1 MG Oral Tablet [Prandin] | PO |
| Amaryl Oral Product | orally |
| dapagliflozin 5 MG Oral Tablet [Farxiga] | ORAL |
| 24 HR Glipizide 2.5 MG Extended Release Oral Tablet [Glucotrol] | |
| Metformin Oral Tablet | oral |
| Insulin Lispro Cartridge [Humalog] | subcu |
| insulin human, isophane 70 UNT/ML / Regular Insulin, Human 30 UNT/ML [Novolin] | by mouth |
| Insulin Lispro | NPO. |
| Insulin Glargine Injectable Solution [Lantus] | subcut |
| 3 ML Insulin Glargine 100 UNT/ML Pen Injector [Lantus] | subq |
| 3 ML Insulin, Aspart Protamine, Human 70 UNT/ML / Insulin, Aspart, Human 30 UNT/ML Pen Injector [NovoLog Mix] | SQ |
| insulin human, isophane 28 UNT/ML | subcut |
| Insulin Lispro Cartridge [Humalog] | SC |
| empagliflozin 10 MG Oral Tablet [Jardiance] | |

| | |
|---|---|
| insulin pork, isophane 100 UNT/ML Injectable Suspension [NPH Iletin II] | subQ |
| Metformin hydrochloride 1000 MG / rosiglitazone 2 MG [Avandamet] | |
| insulin pork, isophane 100 UNT/ML Injectable Suspension [NPH Iletin II] | subcut. |
| Linagliptin 2.5 MG / Metformin hydrochloride 1000 MG Oral Tablet [Jentadueto] | |
| glimepiride 1 MG Oral Tablet [Amaryl] | |
| Insulin Lispro | sc |
| 3 ML Insulin Lispro 200 UNT/ML Pen Injector [Humalog] | |
| Acarbose 25 MG Oral Tablet | po |
| Regular Insulin, Human 30 UNT/ML | subcut |
| glimepiride Oral Tablet | by mouth |
| Actos Oral Product | orally |
| insulin human, isophane 70 UNT/ML / Regular Insulin, Human 30 UNT/ML Injectable Suspension | Subcutaneous |
| Metformin | subcutaneous |
| exenatide 2 MG [Bydureon] | by mouth |
| pioglitazone 15 MG Oral Tablet | |
| Glipizide 2.5 MG [Glucotrol] | PO |
| 60 ACTUAT exenatide 0.01 MG/ACTUAT Pen Injector [Byetta] | subcutaneous |
| exenatide 0.01 MG/ACTUAT [Byetta] | subcu |
| glimepiride 1 MG Oral Tablet [Amaryl] | p.o. |
| Metformin 250 MG Oral Tablet | PO |
| Trulicity Injectable Product | |
| Insulin Glargine Injectable Solution [Lantus] | subcutaneous |
| Bydureon Injectable Product | |
| Amaryl Oral Product | p.o. |
| rosiglitazone | PO |
| exenatide 0.01 MG/ACTUAT [Byetta] | by mouth |
| Insulin, Aspart, Human 100 UNT/ML Cartridge [NovoLog] | by mouth |
| 3 ML insulin human, isophane 70 UNT/ML / Regular Insulin, Human 30 UNT/ML Pen Injector [Humulin] | |
| insulin human, isophane 100 UNT/ML Pen Injector [Humulin N] | |
| Amaryl Oral Product | oral |
| insulin human, isophane 90 UNT/ML | subcut |
| Metformin | gt |
| insulin pork, isophane 70 UNT/ML / Insulin, Regular, Pork 30 UNT/ML Injectable Suspension | SubQ |

| | | |
|---|---|---|
| canagliflozin 100 MG Oral Tablet [Invokana] | oral | |
| nateglinide 120 MG Oral Tablet [Starlix] | Orally | |
| Insulin Glargine 100 UNT/ML [Lantus] | subc | |
| Glipizide 2.5 MG Oral Tablet | p.o. | |
| Glipizide 2.5 MG / Metformin hydrochloride 250 MG Oral Tablet [Metaglip] | by mouth | |
| glimepiride 2 MG / pioglitazone 30 MG Oral Tablet [Duetact] | by mouth | |
| Metformin Oral Tablet [Glucophage] | oral | |
| saxagliptin 5 MG Oral Tablet [Onglyza] | ORAL | |
| glimepiride 4 MG / rosiglitazone 8 MG Oral Tablet [Avandaryl] | by mouth | |
| glimepiride Oral Tablet [Amaryl] | BY MOUTH | |
| Glyburide 1.25 MG / Metformin hydrochloride 250 MG Oral Tablet | | |
| insulin pork, isophane 100 UNT/ML Injectable Suspension [NPH Iletin II] | sc | |
| Metformin hydrochloride 500 MG Oral Tablet | Orally | |
| Metformin Oral Solution | | |
| insulin human, isophane 16 UNT/ML | subcutaneous | |
| pioglitazone 15 MG Oral Tablet | PO | |
| saxagliptin 5 MG Oral Tablet [Onglyza] | oral | |
| Actos Oral Product | p.o. | |
| Bromocriptine 2.5 MG Oral Tablet | PO | |
| canagliflozin 100 MG Oral Tablet [Invokana] | po | |
| Metformin 500 MG | nostril | |
| glimepiride 1 MG Oral Tablet [Amaryl] | | 0 |
| Glyburide 1.25 MG Oral Tablet | p.o. | |
| Regular Insulin, Human 100 UNT/ML Injectable Solution [Novolin R] | DRIP | |
| Insulin Glargine | npo | |
| Glyburide 5 MG Oral Tablet | | 0 |
| insulin human, isophane 28 UNT/ML | subcut. | |
| glimepiride 6 MG Oral Tablet | by mouth | |
| insulin human, isophane 70 UNT/ML | SQ | |
| NovoLog Injectable Product | subcutaneous | |
| insulin pork, isophane 100 UNT/ML Injectable Suspension [NPH Iletin II] | npo | |
| insulin human, isophane 70 UNT/ML | subcut. | |
| Metformin hydrochloride 500 MG / rosiglitazone 4 MG Oral Tablet [Avandamet] | by mouth | |
| insulin human, isophane 40 UNT/ML | sq | |

| | | |
|---|---|---|
| glimepiride 2 MG / rosiglitazone 4 MG Oral Tablet [Avandaryl] | po | |
| repaglinide 0.5 MG Oral Tablet [Prandin] | PO | |
| Metformin hydrochloride 500 MG Extended Release Oral Tablet [Fortamet] | Oral | |
| Starlix Oral Product | PO | |
| rosiglitazone 8 MG | | |
| pioglitazone | PO | |
| 3 ML liraglutide 6 MG/ML Pen Injector [Victoza] | subQ | |
| Insulin Glargine Injectable Solution [Lantus] | Subcutaneously | |
| Glipizide Extended Release Oral Tablet | PO | |
| Glipizide 5 MG / Metformin hydrochloride 500 MG Oral Tablet [Metaglip] | | |
| insulin pork, isophane 100 UNT/ML Injectable Suspension [NPH Iletin II] | SC | |
| saxagliptin 2.5 MG Oral Tablet [Onglyza] | po | |
| Metformin | swallow | |
| Humulin Injectable Product | | |
| empagliflozin 10 MG / Linagliptin 5 MG Oral Tablet [Glyxambi] | by mouth | |
| glimepiride Oral Tablet | | |
| insulin pork, isophane 80 UNT/ML | subcut. | |
| nateglinide 60 MG Oral Tablet | po | |
| miglitol 25 MG Oral Tablet [Glyset] | PO | |
| glimepiride 4 MG / rosiglitazone 4 MG Oral Tablet [Avandaryl] | po | |
| Glyburide 5 MG Oral Tablet [Diabeta] | | |
| Metformin hydrochloride 850 MG Oral Tablet [Glucophage] | p.o. | |
| Metformin hydrochloride 500 MG Oral Tablet | Po | |
| insulin detemir 100 UNT/ML [Levemir] | subcut | |
| exenatide 0.01 MG/ACTUAT [Byetta] | sub Q | |
| Metformin hydrochloride 850 MG Oral Tablet | | 0 |
| Insulin, Aspart, Human | IV | |
| glimepiride 3 MG Oral Tablet | by mouth | |
| Precose Oral Product | by mouth | |
| canagliflozin 50 MG / Metformin hydrochloride 1000 MG Oral Tablet [Invokamet] | by mouth | |
| repaglinide | | |
| Metformin hydrochloride 850 MG Oral Tablet [Glucophage] | oral | |
| insulin human, isophane 36 UNT/ML | subcut | |
| rosiglitazone 4 MG Oral Tablet [Avandia] | oral | |
| Metformin hydrochloride 500 MG / sitagliptin 50 MG Oral Tablet [Janumet] | ORAL | |

| | | |
|---|---|---|
| Januvia Oral Product | PO | |
| canagliflozin 300 MG Oral Tablet [Invokana] | oral | |
| Insulin Lispro Cartridge [Humalog] | sc | |
| canagliflozin 300 MG Oral Tablet [Invokana] | | |
| Glyburide 5 MG / Metformin hydrochloride 500 MG Oral Tablet | PO | |
| 0.5 ML albiglutide 100 MG/ML Pen Injector [Tanzeum] | | |
| insulin pork, isophane 100 UNT/ML Injectable Suspension [NPH Iletin II] | Subcutaneous | |
| sitagliptin 100 MG Oral Tablet | po | |
| Apidra Injectable Product | | |
| Linagliptin 5 MG | | |
| Insulin Glargine | sub q | |
| Metformin hydrochloride 1000 MG Extended Release Oral Tablet [Glumetza] | po | |
| Glyburide 1.5 MG Oral Tablet | PO | |
| Glyburide 1 MG Oral Tablet | po | |
| 60 ACTUAT exenatide 0.005 MG/ACTUAT Pen Injector [Byetta] | by mouth | |
| 24 HR Metformin hydrochloride 1000 MG / sitagliptin 50 MG Extended Release Oral Tablet [Janumet] | oral | |
| nateglinide 120 MG Oral Tablet [Starlix] | p.o. | |
| 60 ACTUAT exenatide 0.005 MG/ACTUAT Pen Injector [Byetta] | SQ | |
| Bromocriptine 0.8 MG Oral Tablet [Cycloset] | by mouth | |
| insulin pork, isophane 80 UNT/ML | subq | |
| Januvia Oral Product | orally | |
| nateglinide 60 MG Oral Tablet | orally | |
| 3 ML Insulin Lispro 200 UNT/ML Pen Injector [Humalog] | SUBCUTANEOUS | |
| insulin pork, isophane 80 UNT/ML | subcutaneous | |
| Levemir Injectable Product | Subcutaneously | |
| dapagliflozin 10 MG Oral Tablet [Farxiga] | orally | |
| Glipizide Oral Product | p.o. | |
| repaglinide 2 MG Oral Tablet | by mouth | |
| 24 HR Glipizide 5 MG Extended Release Oral Tablet [Glucotrol] | | 0 |
| insulin pork, isophane 100 UNT/ML Injectable Suspension [NPH Iletin II] | oral | |
| insulin pork, isophane 100 UNT/ML Injectable Suspension [NPH Iletin II] | po | |
| Metformin hydrochloride 1000 MG / rosiglitazone 2 MG Oral Tablet [Avandamet] | po | |
| Regular Insulin, Human 40 UNT/ML | | |
| Glucovance Oral Product | PO | |

| | |
|---|---|
| Bromocriptine 2.5 MG Oral Tablet | orally |
| insulin human, isophane 60 UNT/ML | subcut. |
| Insulin, Zinc, Human | SQ |
| sitagliptin 50 MG Oral Tablet | |
| rosiglitazone 8 MG Oral Tablet | by mouth |
| Bromocriptine Oral Tablet | by mouth |
| Metformin hydrochloride 1000 MG / pioglitazone 30 MG [Actoplus Met] | |
| insulin human, isophane 16 UNT/ML | subq |
| Insulin Glargine Injectable Solution [Lantus] | SUBCUTANEOUSLY |
| insulin human, isophane 28 UNT/ML | subcutaneous |
| insulin human, isophane 36 UNT/ML | subcutaneous |
| Ultralente Insulin, Human 100 UNT/ML Injectable Suspension | sq |
| Chlorpropamide 250 MG Oral Tablet | by mouth |
| Glyburide 5 MG Oral Tablet [Diabeta] | p.o. |
| insulin human, isophane 70 UNT/ML / Regular Insulin, Human 30 UNT/ML [Novolin] | sub q |
| Regular Insulin, Human 25 UNT/ML | subcut |
| Acarbose 50 MG Oral Tablet | po |
| insulin human, isophane 70 UNT/ML / Regular Insulin, Human 30 UNT/ML [Humulin] | subQ |
| Insulin Lispro 50 UNT/ML / Insulin, Protamine Lispro, Human 50 UNT/ML Injectable Suspension [Humalog Mix] | SQ |
| repaglinide Oral Product | by mouth |
| insulin human, isophane 100 UNT/ML Prefilled Syringe | |
| saxagliptin 5 MG Oral Tablet [Onglyza] | PO |
| Insulin, Aspart, Human 100 UNT/ML Cartridge [NovoLog] | subcutaneously. |
| Osmotic 24 HR Metformin hydrochloride 500 MG Extended Release Oral Tablet [Fortamet] | |
| nateglinide 30 MG Oral Tablet | orally |
| pramlintide acetate 0.6 MG/ML Injectable Solution [Symlin] | subcut |
| Insulin Lispro | SC |
| Insulin Glargine | oral |
| Insulin Glargine 100 UNT/ML Injectable Solution | subcutaneously |
| Insulin, Aspart, Human | SUB CONJ |
| pioglitazone 45 MG Oral Tablet | PO |
| empagliflozin 25 MG | |
| Insulin Lispro 100 UNT/ML Pen Injector [Humalog] | SQ |
| Linagliptin 2.5 MG / Metformin hydrochloride 1000 MG [Jentadueto] | |

| | | |
|---|---|---|
| Regular Insulin, Human 25 UNT/ML | | |
| Insulin Lispro 100 UNT/ML Injectable Solution [Humalog] | subcutaneously | |
| 24 HR Metformin hydrochloride 750 MG Extended Release Oral Tablet [Glucophage] | | 0 |
| Metformin hydrochloride 850 MG / pioglitazone 15 MG [Actoplus Met] | | |
| 3 ML insulin human, isophane 100 UNT/ML Prefilled Syringe [Novolin N] | SubQ | |
| canagliflozin 300 MG Oral Tablet [Invokana] | orally | |
| 3 ML insulin detemir 100 UNT/ML Pen Injector [Levemir] | subQ | |
| Linagliptin 5 MG Oral Tablet [Tradjenta] | oral | |
| canagliflozin 150 MG / Metformin hydrochloride 500 MG Oral Tablet [Invokamet] | | |
| saxagliptin 5 MG Oral Tablet | by mouth | |
| 24 HR Metformin hydrochloride 1000 MG / sitagliptin 50 MG Extended Release Oral Tablet [Janumet] | | |
| insulin human, isophane 70 UNT/ML | sq | |
| Glyburide 5 MG Oral Tablet | Orally | |
| glimepiride Pill | | |
| 3 ML Insulin Lispro 100 UNT/ML Cartridge [Humalog] | subcut | |
| rosiglitazone 4 MG Oral Tablet [Avandia] | | |
| Insulin Glargine | iv | |
| Metformin hydrochloride 100 MG/ML Oral Solution [Riomet] | ORAL | |
| Insulin, Aspart Protamine, Human 70 UNT/ML / Insulin, Aspart, Human 30 UNT/ML [NovoLog Mix] | sc | |
| exenatide | | |
| 24 HR Glipizide 10 MG Extended Release Oral Tablet | by mouth | |
| Insulin, Aspart Protamine, Human 70 UNT/ML / Insulin, Aspart, Human 30 UNT/ML Injectable Suspension [NovoLog Mix] | SubQ | |
| insulin human, isophane 70 UNT/ML / Regular Insulin, Human 30 UNT/ML Injectable Suspension [Humulin] | subcut | |
| 24 HR Metformin hydrochloride 1000 MG / saxagliptin 2.5 MG Extended Release Oral Tablet [Kombiglyze] | by mouth | |
| Glipizide Extended Release Oral Tablet [Glucotrol] | by mouth | |
| glimepiride 2 MG Oral Tablet [Amaryl] | Orally | |
| glimepiride 4 MG / rosiglitazone 4 MG Oral Tablet [Avandaryl] | by mouth | |
| Insulin Lispro 100 UNT/ML Pen Injector [Humalog] | subcutaneous | |
| Glyburide 6 MG Oral Tablet | oral | |
| Insulin Lispro | drip | |
| glimepiride 1 MG Oral Tablet [Amaryl] | orally | |

| | |
|---|---|
| Bromocriptine 2.5 MG Oral Tablet | |
| Insulin, Aspart Protamine, Human | |
| glimepiride 2 MG / rosiglitazone 4 MG Oral Tablet [Avandaryl] | by mouth |
| alogliptin 25 MG Oral Tablet [Nesina] | by mouth |
| Metformin 500 MG | per tube |
| Insulin, Glulisine, Human | |
| insulin human, isophane 60 UNT/ML | subq |
| Insulin Glargine Injectable Solution | subq |
| Metformin hydrochloride 1000 MG / sitagliptin 50 MG Oral Tablet [Janumet] | |
| Metformin | IV |
| rosiglitazone 8 MG Oral Tablet [Avandia] | orally |
| Metformin / pioglitazone Oral Tablet | oral |
| 60 ACTUAT exenatide 0.005 MG/ACTUAT Pen Injector [Byetta] | subq |
| Glipizide 2.5 MG [Glucotrol] | orally |
| Glyburide 5 MG Oral Tablet [Micronase] | |
| Insulin, Aspart, Human 100 UNT/ML Injectable Solution | subcutaneously |
| Insulin, Aspart, Human | orally |
| 24 HR Glipizide 10 MG Extended Release Oral Tablet | BY MOUTH |
| Insulin Lispro Injectable Solution [Humalog] | Subcutaneously |
| 60 ACTUAT exenatide 0.01 MG/ACTUAT Pen Injector [Byetta] | sq |
| 24 HR Metformin hydrochloride 500 MG Extended Release Oral Tablet [Glucophage] | PO |
| Glyburide 5 MG Oral Tablet [Diabeta] | PO |
| Insulin Lispro | gt |
| 3 ML liraglutide 6 MG/ML Pen Injector [Victoza] | sub q |
| Glyburide 6 MG Oral Tablet | p.o. |
| Insulin Lispro | bedside |
| Metformin hydrochloride 1000 MG / rosiglitazone 4 MG Oral Tablet [Avandamet] | PO |
| Fortamet Oral Product | po |
| insulin human, isophane 70 UNT/ML / Regular Insulin, Human 30 UNT/ML [Novolin] | subQ |
| Regular Insulin, Human | by mouth |
| Jentadueto Oral Product | by mouth |
| Insulin, Aspart, Human | sub q |
| Parlodel Oral Product | by mouth |
| Insulin Lispro Cartridge [Humalog] | SUBCUT |
| Glyburide / Metformin Oral Product | po |
| Acarbose 50 MG Oral Tablet [Precose] | ORAL |

| | |
|---|---|
| liraglutide 6 MG/ML [Victoza] | SQ |
| Metformin hydrochloride 100 MG/ML [Riomet] | |
| 3 ML Insulin Lispro 100 UNT/ML Cartridge [Humalog] | |
| Novolin Injectable Product | |
| Insulin Glargine | subcut. |
| Insulin, Aspart, Human 100 UNT/ML Injectable Solution [NovoLog] | CONT SUBCUTAN INFUSI |
| Insulin, Aspart, Human 100 UNT/ML Cartridge [NovoLog] | SUBCUT |
| 3 ML insulin human, isophane 100 UNT/ML Pen Injector [Humulin N] | subq |
| Bromocriptine 0.8 MG [Cycloset] | |
| 3 ML Insulin, Glulisine, Human 100 UNT/ML Pen Injector [Apidra] | subq |
| Insulin, Aspart Protamine, Human 70 UNT/ML / Insulin, Aspart, Human 30 UNT/ML [NovoLog Mix] | sub q |
| 60 ACTUAT exenatide 0.005 MG/ACTUAT Pen Injector [Byetta] | subcutaneous |
| glimepiride 2 MG Oral Tablet | Oral |
| empagliflozin 25 MG Oral Tablet [Jardiance] | ORAL |
| Metformin 513 MG | nostril |
| Metformin hydrochloride 850 MG Oral Tablet [Glucophage] | Orally |
| insulin human, isophane 70 UNT/ML / Regular Insulin, Human 30 UNT/ML [Humulin] | sub Q |
| Insulin Lispro | PO |
| glimepiride 3 MG Oral Tablet [Amaryl] | PO |
| 60 ACTUAT exenatide 0.01 MG/ACTUAT Pen Injector [Byetta] | SC |
| Metformin hydrochloride 100 MG/ML Oral Solution | orally |
| pioglitazone 30 MG Oral Tablet [Actos] | |
| exenatide 0.01 MG/ACTUAT | subcut |
| Regular Insulin, Human | sub q |
| Glipizide 10 MG Oral Tablet | BY MOUTH |
| glimepiride 4 MG Oral Tablet | Oral |
| repaglinide 0.5 MG | |
| Glyburide 3 MG Oral Tablet | orally |
| Victoza Injectable Product | SUBCUTANEOUS |
| empagliflozin 10 MG Oral Tablet [Jardiance] | ORAL |
| sitagliptin 25 MG Oral Tablet [Januvia] | oral |
| Glipizide Oral Tablet [Glucotrol] | BY MOUTH |
| Regular Insulin, Human 500 UNT/ML Injectable Solution [Humulin R] | |
| Micronase Oral Product | PO |
| liraglutide | subcut |

| | | |
|---|---|---|
| Regular Insulin, Human 5 UNT/ML | iv | |
| insulin pork, isophane 100 UNT/ML Injectable Suspension [NPH Iletin II] | sub q | |
| Metformin hydrochloride 100 MG/ML | subcutaneous | |
| Metformin hydrochloride 500 MG / pioglitazone 15 MG Oral Tablet [Actoplus Met] | ORAL | |
| Insulin Lispro 25 UNT/ML / Insulin, Protamine Lispro, Human 75 UNT/ML Injectable Suspension [Humalog Mix] | subcu | |
| insulin pork, isophane 100 UNT/ML Injectable Suspension [NPH Iletin II] | SUBCUT | |
| sitagliptin 25 MG Oral Tablet | orally | |
| insulin human, isophane 100 UNT/ML Injectable Suspension [Humulin N] | subcut | |
| dapagliflozin 10 MG Oral Tablet [Farxiga] | po | |
| Bromocriptine 5 MG Oral Capsule | by mouth | |
| Glyburide 2.5 MG / Phenformin 25 MG Oral Tablet | po | |
| Insulin Lispro | subQ | |
| Glyburide 2.5 MG Oral Tablet | | |
| Insulin Lispro Injectable Solution [Humalog] | | |
| Bromocriptine 25 MG | | |
| Glipizide | nostril | |
| Insulin, Aspart, Human 100 UNT/ML Cartridge [NovoLog] | subcut. | |
| Janumet Oral Product | oral | |
| insulin human, isophane 75 UNT/ML | SQ | |
| Osmotic 24 HR Metformin hydrochloride 1000 MG Extended Release Oral Tablet [Fortamet] | | 0 |
| Janumet Oral Product | orally | |
| Glipizide | subcutaneously | |
| Insulin, Aspart Protamine, Human 70 UNT/ML / Insulin, Aspart, Human 30 UNT/ML Injectable Suspension [NovoLog Mix] | subcu | |
| Metformin 500 MG / ropinirole 2 MG Oral Tablet | PO | |
| Bromocriptine 10 MG | | |
| Insulin Glargine 100 UNT/ML | SUBCUT | |
| Insulin Lispro | subcutaneously | |
| 1.5 ML Insulin Glargine 300 UNT/ML Pen Injector [Toujeo] | | |
| Insulin, Aspart Protamine, Human 70 UNT/ML / Insulin, Aspart, Human 30 UNT/ML Injectable Suspension [NovoLog Mix] | sub q | |
| Metformin hydrochloride 1000 MG / saxagliptin 5 MG [Kombiglyze] | by mouth | |
| pioglitazone 30 MG Oral Tablet | PO | |

| | |
|---|---|
| insulin human, isophane 100 UNT/ML Injectable Suspension [Humulin N] | SQ |
| Metformin | IM |
| insulin human, isophane 90 UNT/ML / Regular Insulin, Human 10 UNT/ML [Humulin] | |
| rosiglitazone 2 MG Oral Tablet | orally |
| insulin human, isophane 90 UNT/ML | SQ |
| Insulin Lispro 100 UNT/ML Injectable Solution [Lispro PRC] | |
| insulin pork, isophane 100 UNT/ML Injectable Suspension | |
| Glipizide Extended Release Oral Tablet | orally |
| glimepiride 6 MG Oral Tablet | po |
| 60 ACTUAT exenatide 0.01 MG/ACTUAT Pen Injector [Byetta] | sc |
| Insulin, Aspart, Human 100 UNT/ML Cartridge [NovoLog] | sc |
| insulin human, isophane 16 UNT/ML | subcut. |
| Glyburide 5 MG Oral Tablet | BY MOUTH |
| insulin detemir 100 UNT/ML [Levemir] | sq |
| insulin human, isophane 28 UNT/ML | subq |
| liraglutide 6 MG/ML [Victoza] | subcut |
| 60 ACTUAT exenatide 0.005 MG/ACTUAT Pen Injector [Byetta] | sq |
| Insulin, Aspart, Human 100 UNT/ML Injectable Solution [NovoLog] | SUBCUTANEOUSLY |
| dapagliflozin 10 MG Oral Tablet [Farxiga] | |
| Avandamet Oral Product | PO |
| Glipizide 5 MG / Metformin hydrochloride 500 MG Oral Tablet [Metaglip] | ORAL |
| Insulin, Aspart, Human 100 UNT/ML Cartridge [NovoLog] | sub Q |
| insulin pork, isophane 100 UNT/ML Injectable Suspension [NPH Iletin II] | PO |
| insulin human, isophane 60 UNT/ML | sq |
| Insulin Glargine | IV |
| 3 ML liraglutide 6 MG/ML Pen Injector [Victoza] | subcut |
| repaglinide 0.5 MG Oral Tablet | by mouth |
| 3 ML insulin detemir 100 UNT/ML Pen Injector [Levemir] | SC |
| 24 HR Metformin hydrochloride 1000 MG / sitagliptin 50 MG Extended Release Oral Tablet [Janumet] | ORAL |
| Glyburide 1.5 MG [Glynase] | |
| Insulin Lispro | npo. |
| Novolin N Injectable Product | |
| 3 ML Insulin Lispro 50 UNT/ML / Insulin, Protamine Lispro, Human 50 UNT/ML Pen Injector [Humalog Mix] | subq |

| | | |
|---|---|---|
| 24 HR Glipizide 10 MG Extended Release Oral Tablet | oral | |
| Insulin Lispro | subcut. | |
| 24 HR Glipizide 2.5 MG Extended Release Oral Tablet [Glucotrol] | | 0 |
| insulin human, isophane 100 UNT/ML Injectable Suspension [Novolin N] | | |
| 3 ML insulin detemir 100 UNT/ML Pen Injector [Levemir] | sub q | |
| Metformin hydrochloride 500 MG / rosiglitazone 2 MG Oral Tablet [Avandamet] | PO | |
| exenatide 0.01 MG/ACTUAT [Byetta] | SubQ | |
| Insulin Lispro | oral | |
| insulin detemir 100 UNT/ML [Levemir] | subc | |
| Glipizide 5 MG / Metformin hydrochloride 500 MG Oral Tablet [Metaglip] | po | |
| Toujeo Injectable Product | subcutaneously | |
| exenatide 0.005 MG/ACTUAT [Byetta] | SC | |
| glimepiride 8 MG Oral Tablet | orally | |
| Metformin hydrochloride 850 MG / pioglitazone 15 MG Oral Tablet | by mouth | |
| 3 ML Insulin, Aspart, Human 100 UNT/ML Cartridge [NovoLog] | po | |
| insulin human, isophane 70 UNT/ML / Regular Insulin, Human 30 UNT/ML Injectable Suspension | subcutaneous | |
| Insulin Glargine Injectable Solution [Lantus] | SQ | |
| 3 ML insulin human, isophane 70 UNT/ML / Regular Insulin, Human 30 UNT/ML Pen Injector [Humulin] | subcutaneous | |
| exenatide 0.005 MG/ACTUAT [Byetta] | Subcutaneous | |
| exenatide 0.01 MG/ACTUAT [Byetta] | Subcut. | |
| sitagliptin 50 MG Oral Tablet | by mouth | |
| Glipizide 5 MG / Metformin hydrochloride 500 MG Oral Tablet | po | |
| 3 ML Insulin Lispro 200 UNT/ML Pen Injector [Humalog] | | |
| insulin pork, isophane 80 UNT/ML | SQ | |
| insulin human, isophane 85 UNT/ML | subcut | |
| Insulin, Aspart Protamine, Human 70 UNT/ML / Insulin, Aspart, Human 30 UNT/ML [NovoLog Mix] | SC | |
| Regular Insulin, Human | subQ | |
| Insulin Lispro 25 UNT/ML | subcu | |
| Insulin Glargine 100 UNT/ML [Lantus] | Sub Q | |
| alogliptin 25 MG [Nesina] | | |
| insulin pork, isophane 70 UNT/ML / Insulin, Regular, Pork 30 UNT/ML Injectable Suspension | subq | |
| insulin beef, isophane 40 UNT/ML | subcut | |

| | | |
|---|---|---|
| Bromocriptine | po | |
| Glyburide 5 MG Oral Tablet [Micronase] | PO | |
| Glyburide 3 MG Oral Tablet [Glynase] | PO | |
| Insulin, Aspart, Human 100 UNT/ML Cartridge [NovoLog] | Subcut | |
| Insulin Lispro Cartridge [Humalog] | Subcut | |
| 3 ML insulin detemir 100 UNT/ML Pen Injector [Levemir] | | 0 |
| Glucophage Pill | by mouth | |
| Metformin 250 MG Oral Tablet | oral | |
| Insulin, Glulisine, Human 100 UNT/ML Injectable Solution | | |
| Metformin Pill | by mouth | |
| repaglinide 2 MG Oral Tablet [Prandin] | ORAL | |
| insulin human, isophane 60 UNT/ML | sub q | |
| Glipizide 2.5 MG Oral Tablet | | |
| liraglutide 6 MG/ML | subcut | |
| insulin pork, isophane 100 UNT/ML Injectable Suspension [NPH Iletin II] | IV | |
| 3 ML liraglutide 6 MG/ML Pen Injector [Saxenda] | | |
| canagliflozin 100 MG Oral Tablet [Invokana] | | |
| Metformin Oral Tablet [Glucophage] | PO | |
| Glyburide 3 MG Oral Tablet | ORAL | |
| Metformin hydrochloride 500 MG / rosiglitazone 2 MG [Avandamet] | nostril | |
| insulin detemir 100 UNT/ML Injectable Solution | | |
| pioglitazone 45 MG Oral Tablet [Actos] | | |
| pramlintide acetate 0.6 MG/ML Injectable Solution [Symlin] | SUBCUTANEOUS | |
| Januvia Pill | | |
| Diabeta Oral Product | p.o. | |
| 3 ML insulin detemir 100 UNT/ML Pen Injector [Levemir] | subcut | |
| nateglinide 120 MG Oral Tablet | PO | |
| Insulin Lispro | iv | |
| 2.7 ML pramlintide acetate 1 MG/ML Pen Injector [Symlin] | SUBCUTANEOUS | |
| Metformin hydrochloride 500 MG [Glucophage] | per tube | |
| Insulin, Aspart, Human 100 UNT/ML Cartridge [NovoLog] | NPO | |
| Insulin Glargine | SubQ | |
| Invokamet Oral Product | by mouth | |
| Glyburide 5 MG / Phenformin 50 MG Oral Tablet | | |
| Avandia Oral Product | oral | |
| insulin pork, isophane 70 UNT/ML / Insulin, Regular, Pork 30 UNT/ML Injectable Suspension | | |
| empagliflozin 25 MG / Linagliptin 5 MG [Glyxambi] | | |

| | |
|---|---|
| Glyburide 2.5 MG Oral Tablet [Diabeta] | po |
| Glyburide 2.5 MG Oral Tablet [Diabeta] | oral |
| insulin human, isophane 70 UNT/ML | subcutaneously |
| sitagliptin 25 MG Oral Tablet [Januvia] | ORAL |
| exenatide | SUBCUT |
| empagliflozin 25 MG / Linagliptin 5 MG Oral Tablet [Glyxambi] | |
| Acarbose Oral Tablet [Precose] | by mouth |
| Metformin hydrochloride 500 MG Oral Tablet [Glucophage] | BY MOUTH |
| insulin human, isophane 80 UNT/ML / Regular Insulin, Human 20 UNT/ML [Humulin] | |
| Glipizide 2.5 MG / Metformin hydrochloride 250 MG Oral Tablet | oral |
| nateglinide 60 MG Oral Tablet [Starlix] | orally |
| dapagliflozin 5 MG Oral Tablet [Farxiga] | po |
| Glipizide 5 MG Oral Tablet | BY MOUTH |
| sitagliptin 25 MG Oral Tablet [Januvia] | |
| Glyburide 1.25 MG / Metformin hydrochloride 250 MG Oral Tablet | po |
| Bromocriptine 5 MG Oral Capsule | |
| Insulin, Aspart, Human | drip. |
| sitagliptin Oral Tablet [Januvia] | by mouth |
| Metformin Oral Tablet | PO |
| miglitol 100 MG Oral Tablet [Glyset] | by mouth |
| Glyburide 5 MG Oral Tablet [Diabeta] | orally |
| dapagliflozin 5 MG Oral Tablet [Farxiga] | orally |
| Insulin, Regular, Pork 40 UNT/ML Injectable Solution | subcut |
| Humulin U Ultralente Injectable Product | subcutaneously |
| Metformin 500 MG | by mouth. |
| Insulin Glargine | SUBQ |
| insulin pork, isophane 100 UNT/ML Injectable Suspension [NPH Iletin II] | IM |
| Insulin Lispro 25 UNT/ML / Insulin, Protamine Lispro, Human 75 UNT/ML Injectable Suspension [Humalog Mix] | sc |
| miglitol 100 MG Oral Tablet [Glyset] | po |
| Glyburide | NPO |
| Insulin Lispro 50 UNT/ML / Insulin, Protamine Lispro, Human 50 UNT/ML Injectable Suspension [Humalog Mix] | sq |
| 3 ML insulin detemir 100 UNT/ML Pen Injector [Levemir] | sq |
| canagliflozin 100 MG | |
| Metformin | npo |
| Regular Insulin, Human 10 UNT/ML | IV |

| | |
|---|---|
| rosiglitazone 2 MG Oral Tablet [Avandia] | ORAL |
| Glyburide 5 MG / Phenformin 50 MG Oral Tablet | po |
| Janumet Oral Product | PO |
| Insulin Lispro 25 UNT/ML / Insulin, Protamine Lispro, Human 75 UNT/ML Injectable Suspension [Humalog Mix] | subQ |
| Chlorpropamide 250 MG | |
| Insulin Lispro Cartridge [Humalog] | sub Q |
| Glipizide 2.5 MG / Metformin hydrochloride 500 MG Oral Tablet [Metaglip] | ORAL |
| nateglinide 60 MG Oral Tablet [Starlix] | oral |
| Insulin Glargine 100 UNT/ML Injectable Solution [Lantus] | Subcutaneously |
| 1.5 ML insulin human, isophane 100 UNT/ML Prefilled Syringe | Subcutaneous |
| Parlodel Oral Product | po |
| Glyburide 2.5 MG / Metformin 400 MG Oral Tablet | by mouth |
| Acarbose 25 MG Oral Tablet [Precose] | ORAL |
| Metformin 500 MG | subcutaneously |
| Regular Insulin, Human | subcu |
| Humalog Injectable Product | subcut. |
| Metformin hydrochloride 500 MG | |
| Insulin, Aspart Protamine, Human 70 UNT/ML / Insulin, Aspart, Human 30 UNT/ML Injectable Suspension | subq |
| Insulin Lispro Cartridge [Humalog] | Subq |
| Glucotrol Oral Product | p.o. |
| 24 HR Metformin hydrochloride 1000 MG / saxagliptin 2.5 MG Extended Release Oral Tablet [Kombiglyze] | po |
| glimepiride 2 MG / rosiglitazone 4 MG Oral Tablet [Avandaryl] | orally |
| insulin pork, isophane 100 UNT/ML Injectable Suspension [NPH Iletin II] | Subcutaneously |
| 3 ML Insulin, Aspart Protamine, Human 70 UNT/ML / Insulin, Aspart, Human 30 UNT/ML Pen Injector [NovoLog Mix] | Subcutaneous |
| exenatide 0.01 MG/ACTUAT [Byetta] | subQ |
| Ultralente Insulin, Human 100 UNT/ML Injectable Suspension [Humulin U Ultralente] | Subcutaneous |
| Glyburide 1 MG Oral Tablet | PO |
| Insulin Lispro 50 UNT/ML | subcu |
| 3 ML Insulin, Aspart Protamine, Human 70 UNT/ML / Insulin, Aspart, Human 30 UNT/ML Pen Injector [NovoLog Mix] | subcutaneously |
| Insulin, Aspart, Human | SEE COMMENT |
| Regular Insulin, Human 10 UNT/ML | sq |
| Insulin, Protamine Lispro, Human | |

| | |
|---|---|
| 60 ACTUAT exenatide 0.01 MG/ACTUAT Pen Injector [Byetta] | sub q |
| rosiglitazone 2 MG Oral Tablet | PO |
| Regular Insulin, Human 40 UNT/ML | subcut |
| liraglutide 6 MG/ML | |
| Ultralente Insulin, Human 100 UNT/ML Injectable Suspension [Humulin U Ultralente] | |
| sitagliptin 50 MG Oral Tablet [Januvia] | |
| Acarbose 100 MG Oral Tablet | orally |
| repaglinide 1 MG Oral Tablet [Prandin] | |
| Regular Insulin, Human 30 UNT/ML | IV |
| 3 ML insulin human, isophane 100 UNT/ML Prefilled Syringe | Subcutaneous |
| insulin detemir Injectable Solution [Levemir] | subcutaneously |
| exenatide 0.01 MG/ACTUAT Pen Injector [Byetta] | SUBCUT |
| Insulin, Aspart Protamine, Human 70 UNT/ML / Insulin, Aspart, Human 30 UNT/ML [NovoLog Mix] | subcut. |
| exenatide 0.005 MG/ACTUAT [Byetta] | sub q |
| Glyburide 1.25 MG Oral Tablet | |
| canagliflozin 100 MG Oral Tablet [Invokana] | orally |
| exenatide 0.005 MG/ACTUAT [Byetta] | by mouth |
| repaglinide 1 MG Oral Tablet | by mouth |
| glimepiride | nostril |
| Metformin hydrochloride 500 MG Extended Release Oral Tablet | by mouth |
| Glipizide 2.5 MG / Metformin hydrochloride 250 MG Oral Tablet [Metaglip] | |
| insulin human, isophane 32 UNT/ML | subcutaneous |
| Metformin hydrochloride 1000 MG Oral Tablet | Orally |
| Humalog Injectable Product | subcut |
| Glyburide 5 MG Oral Tablet | Oral |
| 3 ML insulin human, isophane 100 UNT/ML Prefilled Syringe [Novolin N] | subq |
| exenatide 0.01 MG/ACTUAT [Byetta] | sub q |
| repaglinide 0.5 MG Oral Tablet | orally |
| 3 ML liraglutide 6 MG/ML Pen Injector [Victoza] | sq |
| Insulin, Aspart, Human 100 UNT/ML Injectable Solution | SC |
| 1.5 ML insulin human, isophane 100 UNT/ML Prefilled Syringe | |
| Insulin Glargine | drip |
| sitagliptin 100 MG Oral Tablet [Januvia] | Oral |
| glimepiride 1 MG Oral Tablet | p.o. |
| 3 ML Insulin Lispro 25 UNT/ML / Insulin, Protamine Lispro, Human 75 UNT/ML Pen Injector [Humalog Mix] | |

| | |
|---|---|
| 3 ML insulin detemir 100 UNT/ML Pen Injector [Levemir] | subcu |
| Ultralente Insulin, Human 100 UNT/ML Injectable Suspension | subcut |
| sitagliptin 50 MG Oral Tablet [Januvia] | p.o. |
| liraglutide Injectable Product | |
| Glyburide 6 MG Oral Tablet [Glynase] | p.o. |
| 24 HR Metformin hydrochloride 1000 MG / sitagliptin 100 MG Extended Release Oral Tablet [Janumet] | oral |
| Glyburide 1.5 MG Oral Tablet [Glynase] | po |
| Insulin Glargine 100 UNT/ML [Lantus] | SUBCUT |
| empagliflozin 10 MG | |
| 24 HR Glipizide 5 MG Extended Release Oral Tablet [Glucotrol] | PO |
| Tolbutamide 500 MG | |
| Bromocriptine 5 MG Oral Capsule [Parlodel] | po |
| repaglinide 1 MG Oral Tablet [Prandin] | ORAL |
| 3 ML liraglutide 6 MG/ML Pen Injector | |
| Glyburide 5 MG Oral Tablet [Diabeta] | oral |
| Regular Insulin, Human 100 UNT/ML Injectable Solution [Humulin R] | subcut. |
| Humalog Injectable Product | sq |
| insulin human, isophane 34 UNT/ML | subcutaneous |
| 3 ML Insulin, Glulisine, Human 100 UNT/ML Pen Injector [Apidra] | |
| Prandin Oral Product | PO |
| Glyburide 1 MG Oral Tablet | orally |
| Insulin, Aspart, Human 100 UNT/ML Cartridge [NovoLog] | gt |
| Metformin hydrochloride 1000 MG Extended Release Oral Tablet [Fortamet] | PO |
| Glipizide 10 MG Oral Tablet [Glucotrol] | Orally |
| Glyburide 1.25 MG / Metformin hydrochloride 250 MG Oral Tablet [Glucovance] | PO |
| pramlintide acetate 1 MG/ML [Symlin] | |
| Insulin Lispro 100 UNT/ML [Humalog] | subcut |
| insulin human, isophane 100 UNT/ML Injectable Suspension [Novolin N] | sc |
| repaglinide 0.5 MG Oral Tablet [Prandin] | orally |
| Acarbose 50 MG Oral Tablet [Precose] | PO |
| rosiglitazone 2 MG | |
| Glyburide 2.5 MG / Metformin hydrochloride 500 MG Oral Tablet | orally |
| exenatide 2 MG [Bydureon] | SC |
| glimepiride 3 MG Oral Tablet | po |
| Glyburide 2.5 MG / Phenformin 25 MG Oral Tablet | by mouth |

| | |
|---|---|
| exenatide 0.01 MG/ACTUAT [Byetta] | SC |
| Metformin 1000 MG | nostril |
| Glyburide 5 MG Oral Tablet [Micronase] | orally |
| insulin human, isophane 32 UNT/ML | subcut. |
| Pramlintide Injectable Solution | |
| 3 ML liraglutide 6 MG/ML Pen Injector [Victoza] | SC |
| rosiglitazone 2 MG Oral Tablet [Avandia] | |
| Insulin Glargine 100 UNT/ML [Lantus] | sub Q |
| Simvastatin 20 MG / sitagliptin 50 MG Oral Tablet | po |
| Glipizide Oral Tablet [Glucotrol] | po |
| Metformin | drip |
| Regular Insulin, Human 100 UNT/ML Injectable Solution [Novolin R] | sc |
| Acarbose 25 MG Oral Tablet [Precose] | |
| dulaglutide 1.5 MG/ML [Trulicity] | SQ |
| exenatide 2 MG | |
| glimepiride 4 MG / pioglitazone 30 MG Oral Tablet [Duetact] | po |
| insulin, isophane Injectable Suspension [Novolin N] | subcutaneously |
| Metformin hydrochloride 1000 MG / sitagliptin 50 MG Oral Tablet | orally |
| insulin human, isophane 75 UNT/ML | subcutaneous |
| Regular Insulin, Human 5 UNT/ML | sq |
| Insulin Lispro | SQ INFUSION |
| Glipizide 2.5 MG / Metformin hydrochloride 250 MG Oral Tablet | po |
| Insulin, Glulisine, Human 100 UNT/ML Injectable Solution [Apidra] | Subcut |
| insulin human, isophane 28 UNT/ML | SQ |
| insulin detemir | subq |
| insulin human, isophane 90 UNT/ML | subcut. |
| Oseni Oral Product | by mouth |
| Glyburide Oral Tablet | |
| insulin beef, isophane 40 UNT/ML | SUBCUT |
| Glipizide | NPO |
| Glucovance Oral Product | orally |
| 24 HR Metformin hydrochloride 1000 MG / sitagliptin 100 MG Extended Release Oral Tablet [Janumet] | |
| Glyburide 1.25 MG / Metformin hydrochloride 250 MG [Glucovance] | |
| sitagliptin 50 MG Oral Tablet | po |

| | |
|---|---|
| Insulin Lispro 50 UNT/ML / Insulin, Protamine Lispro, Human 50 UNT/ML Injectable Suspension [Humalog Mix] | subcut. |
| Acarbose | PO |
| glimepiride 1 MG / rosiglitazone 4 MG [Avandaryl] | |
| Precose Oral Product | po |
| pioglitazone 30 MG Oral Tablet | |
| Metformin 1000 MG | by mouth. |
| insulin detemir | subcutaneous |
| insulin human, isophane 85 UNT/ML | subcut. |
| NovoLog Injectable Product | subQ |
| Regular Insulin, Human 5 UNT/ML | IV |
| Regular Insulin, Human 500 UNT/ML Injectable Solution | |
| Metformin Oral Tablet | orally |
| Insulin Lispro 100 UNT/ML Injectable Solution [Humalog] | INFUSI |
| dulaglutide 1.5 MG/ML [Trulicity] | subcutaneously |
| nateglinide 120 MG Oral Tablet [Starlix] | |
| Regular Insulin, Human | SC |
| insulin human, isophane 70 UNT/ML / Regular Insulin, Human 30 UNT/ML [Humulin] | Sub Q |
| glimepiride 3 MG Oral Tablet | orally |
| insulin human, isophane 100 UNT/ML Injectable Suspension [Novolin N] | SQ |
| nateglinide 60 MG Oral Tablet | PO |
| exenatide 0.005 MG/ACTUAT [Byetta] | subcu |
| Glipizide 2.5 MG Extended Release Oral Tablet | by mouth |
| Glipizide 2.5 MG Extended Release Oral Tablet [Glucotrol] | by mouth |
| Simvastatin 20 MG / sitagliptin 100 MG Oral Tablet | po |
| Glyburide Oral Tablet [Diabeta] | BY MOUTH |
| Bromocriptine Pill | |
| Metformin hydrochloride 500 MG / pioglitazone 15 MG Oral Tablet [Actoplus Met] | PO |
| empagliflozin 10 MG / Linagliptin 5 MG Oral Tablet [Glyxambi] | |
| Glyburide 1.5 MG | NPO |
| Regular Insulin, Human | Subcutaneously |
| glimepiride | subcutaneous |
| Invokana Oral Product | oral |
| Linagliptin Oral Product | by mouth |
| Insulin Lispro 25 UNT/ML | by mouth |
| Glyburide 5 MG / Metformin hydrochloride 500 MG Oral Tablet | oral |

| | | |
|---|---|---|
| Metformin hydrochloride 500 MG / rosiglitazone 4 MG Oral Tablet [Avandamet] | PO | |
| insulin detemir | subcut | |
| nateglinide 60 MG Oral Tablet [Starlix] | p.o. | |
| rosiglitazone 8 MG Oral Tablet [Avandia] | | |
| Glyburide 5 MG Oral Tablet [Micronase] | p.o. | |
| Metformin hydrochloride 500 MG Extended Release Oral Tablet [Glumetza] | PO | |
| Acarbose 100 MG Oral Tablet | po | |
| Metformin hydrochloride 500 MG / sitagliptin 50 MG Oral Tablet [Janumet] | | |
| Glipizide Extended Release Oral Tablet | po | |
| canagliflozin 150 MG / Metformin hydrochloride 1000 MG [Invokamet] | | |
| Insulin Lispro 25 UNT/ML / Insulin, Protamine Lispro, Human 75 UNT/ML [Humalog Mix] | sub q | |
| Glyburide / Metformin Oral Product | PO | |
| 1.5 ML Regular Insulin, Human 100 UNT/ML Prefilled Syringe | | |
| rosiglitazone 8 MG Oral Tablet [Avandia] | oral | |
| Regular Insulin, Human 15 UNT/ML | SQ | |
| Humulin R Injectable Product | sq | |
| Ultralente Insulin, Human 100 UNT/ML Injectable Suspension [Humulin U Ultralente] | subq | |
| pioglitazone 15 MG Oral Tablet [Actos] | Oral | |
| Acarbose 50 MG Oral Tablet | PO | |
| dapagliflozin 5 MG / Metformin hydrochloride 1000 MG [Xigduo] | by mouth | |
| Metformin hydrochloride 500 MG / rosiglitazone 2 MG Oral Tablet [Avandamet] | | |
| Acarbose 100 MG Oral Tablet [Precose] | ORAL | |
| pioglitazone 45 MG Oral Tablet [Actos] | | 0 |
| repaglinide 0.5 MG Oral Tablet [Prandin] | ORAL | |
| 3 ML Insulin Glargine 100 UNT/ML Pen Injector [Lantus] | by mouth | |
| Insulin Lispro 50 UNT/ML | sub q | |
| Victoza Injectable Product | SQ | |
| Humalog Injectable Product | subcutaneous | |
| Metformin | SC | |
| Metformin 1000 MG | per tube | |
| Insulin Lispro | sub Q | |
| Regular Insulin, Human | po | |
| 60 ACTUAT exenatide 0.01 MG/ACTUAT Pen Injector [Byetta] | subQ | |

| | | |
|---|---|---|
| Insulin Glargine | SUBCUTANEOUS | |
| Insulin Lispro 25 UNT/ML / Insulin, Protamine Lispro, Human 75 UNT/ML [Humalog Mix] | SC | |
| glimepiride 4 MG / pioglitazone 30 MG Oral Tablet | by mouth | |
| glimepiride 8 MG Oral Tablet | PO | |
| Glyburide 1.25 MG Oral Tablet [Diabeta] | po | |
| Tanzeum Injectable Product | subcutaneously | |
| Victoza Injectable Product | SUBCUTANEOUSLY | |
| Regular Insulin, Human 100 UNT/ML Injectable Solution [Humulin R] | subcu | |
| canagliflozin 100 MG Oral Tablet [Invokana] | PO | |
| Insulin Glargine | NPO | |
| pioglitazone Oral Tablet | orally | |
| Insulin, Aspart Protamine, Human 70 UNT/ML / Insulin, Aspart, Human 30 UNT/ML Injectable Suspension [NovoLog Mix] | Subq | |
| 1.5 ML pramlintide acetate 1 MG/ML Pen Injector [Symlin] | SUBCUTANEOUS | |
| Metformin hydrochloride 100 MG/ML Oral Solution | p.o. | |
| liraglutide 6 MG/ML [Victoza] | sub q | |
| Metformin hydrochloride 500 MG / pioglitazone 15 MG Oral Tablet | po | |
| repaglinide 1 MG Oral Tablet | orally | |
| Metformin 400 MG | po | |
| Metformin hydrochloride 1000 MG Oral Tablet [Glucophage] | BY MOUTH | |
| Exubera Inhalant Product | inhaled | |
| Regular Insulin, Human | orally | |
| Insulin Lispro Cartridge [Humalog] | sub q | |
| Glipizide 10 MG Oral Tablet | Po | |
| sitagliptin 100 MG Oral Tablet | PO | |
| Regular Insulin, Human 8 UNT | subcutaneously | |
| Glumetza Oral Product | po | |
| Glyburide Oral Tablet | Oral | |
| 3 ML Insulin, Aspart, Human 100 UNT/ML Cartridge [NovoLog] | | 0 |
| liraglutide 6 MG/ML [Saxenda] | | |
| 3 ML liraglutide 6 MG/ML Pen Injector [Victoza] | SubQ | |
| Insulin Glargine Injectable Solution | sc | |
| Insulin Lispro Pen Injector [Humalog] | | |
| Glyburide 2.5 MG / Metformin hydrochloride 500 MG Oral Tablet [Glucovance] | orally | |
| 3 ML Insulin, Glulisine, Human 100 UNT/ML Pen Injector [Apidra] | | |
| NovoLog Injectable Product | subcu | |

| | |
|---|---|
| Insulin Lispro 50 UNT/ML / Insulin, Protamine Lispro, Human 50 UNT/ML Injectable Suspension [Humalog Mix] | sub q |
| Insulin Lispro 25 UNT/ML / Insulin, Protamine Lispro, Human 75 UNT/ML Injectable Suspension [Humalog Mix] | subcut. |
| Regular Insulin, Human 100 UNT/ML Injectable Solution [Novolin R] | Subq |
| Metformin | SQ |
| 3 ML liraglutide 6 MG/ML Pen Injector [Saxenda] | SUBCUTANEOUS |
| 60 ACTUAT exenatide 0.01 MG/ACTUAT Pen Injector [Byetta] | sub Q |
| Linagliptin 5 MG Oral Tablet [Tradjenta] | |
| Regular Insulin, Human 5 UNT/ML | subcutaneous |
| Bromocriptine 2.5 MG Oral Tablet [Parlodel] | orally |
| Metformin hydrochloride 100 MG/ML Oral Solution | oral |
| Metformin hydrochloride 500 MG / sitagliptin 50 MG Oral Tablet [Janumet] | p.o. |
| Insulin, Aspart Protamine, Human 70 UNT/ML / Insulin, Aspart, Human 30 UNT/ML Injectable Suspension | Subcutaneous |
| Glipizide 2.5 MG / Metformin hydrochloride 500 MG Oral Tablet [Metaglip] | by mouth |
| rosiglitazone 8 MG Oral Tablet | PO |
| Glyburide / Metformin Oral Tablet [Glucovance] | |
| glimepiride 4 MG / pioglitazone 30 MG Oral Tablet [Duetact] | by mouth |
| Glipizide 2.5 MG | per tube |
| {180 (insulin human, rDNA origin 1 MG/ACTUAT Inhalant Powder [Exubera]) / 90 (insulin human, rDNA origin 3 MG/ACTUAT Inhalant Powder [Exubera]) } Pack [Exubera Kit] | Inhalation |
| Metformin hydrochloride 500 MG / rosiglitazone 4 MG Oral Tablet | by mouth |
| Lantus Injectable Product | subcut. |
| Glyburide 5 MG / Metformin 1000 MG Oral Tablet | PO |
| Pramlintide | |
| insulin human, isophane 100 UNT/ML Injectable Suspension [Humulin N] | SC |
| dapagliflozin 5 MG | |
| insulin human, isophane 70 UNT/ML / Regular Insulin, Human 30 UNT/ML [Humulin] | po |
| Amaryl Pill | |
| Insulin, Aspart Protamine, Human 70 UNT/ML / Insulin, Aspart, Human 30 UNT/ML Injectable Suspension [NovoLog Mix] | SUBQ |
| sitagliptin Oral Tablet | orally |

| | |
|---|---|
| insulin human, isophane 40 UNT/ML | subcu |
| insulin pork, isophane 100 UNT/ML Injectable Suspension | Subcutaneous |
| Glipizide 2.5 MG Extended Release Oral Tablet | oral |
| insulin human, isophane 100 UNT/ML Injectable Suspension [Humulin N] | subc. |
| Metformin hydrochloride 500 MG Extended Release Oral Tablet | by mouth. |
| Insulin, Zinc, Human | subcu |
| Insulin Lispro | SEE COMMENT |
| insulin human, isophane 70 UNT/ML / Regular Insulin, Human 30 UNT/ML [Humulin] | p.o. |
| Insulin Lispro 100 UNT/ML Pen Injector [Humalog] | po |
| Symlin Injectable Product | |
| 3 ML liraglutide 6 MG/ML Pen Injector [Victoza] | subq. |
| Glyburide 5 MG / Metformin hydrochloride 500 MG Oral Tablet [Glucovance] | p.o. |
| 3 ML Regular Insulin, Human 500 UNT/ML Pen Injector [Humulin R] | subcutaneous |
| Insulin, Zinc, Human | subcutaneously |
| Metformin hydrochloride 500 MG Extended Release Oral Tablet [Fortamet] | PO |
| Avandia Oral Product | p.o. |
| insulin human, rDNA origin 1 MG/ACTUAT Inhalant Powder [Exubera] | |
| Metformin 500 MG | per mouth |
| Metformin hydrochloride 850 MG / pioglitazone 15 MG Oral Tablet [Actoplus Met] | PO |
| Acarbose Oral Tablet | PO |
| glimepiride 2 MG / rosiglitazone 4 MG [Avandaryl] | |
| Metformin hydrochloride 1000 MG Oral Tablet | Po |
| insulin human, isophane 40 UNT/ML | sub q |
| Glynase Oral Product | po |
| pioglitazone Oral Tablet | po |
| Acarbose 50 MG Oral Tablet [Precose] | oral |
| Metformin hydrochloride 1000 MG / sitagliptin 50 MG Oral Tablet | PO |
| Bromocriptine 5 MG Oral Capsule [Parlodel] | PO |
| Janumet Pill | |
| Insulin Lispro 50 UNT/ML | sub Q |
| glimepiride 2 MG / pioglitazone 30 MG Oral Tablet [Duetact] | po |
| Glyburide Oral Tablet | Orally |

| | |
|---|---|
| Regular Insulin, Human 100 UNT/ML Injectable Solution [Humulin R] | Subcut |
| Insulin Lispro | SUBQ |
| Linagliptin 2.5 MG / Metformin hydrochloride 500 MG [Jentadueto] | |
| Glipizide 2.5 MG | nostril |
| 3 ML insulin detemir 100 UNT/ML Pen Injector [Levemir] | by mouth |
| Onglyza Oral Product | po |
| 60 ACTUAT exenatide 0.01 MG/ACTUAT Pen Injector [Byetta] | PO |
| Metformin hydrochloride 850 MG Oral Tablet | Oral |
| rosiglitazone 4 MG Oral Tablet | |
| saxagliptin 2.5 MG | |
| Metformin hydrochloride 500 MG / pioglitazone 15 MG [Actoplus Met] | |
| Insulin Lispro Cartridge [Humalog] | by mouth |
| Ultralente Insulin, Human 100 UNT/ML Injectable Suspension | SUBCUT |
| Exubera Inhalant Product | inhalation |
| exenatide 2 MG [Bydureon] | sub q |
| Insulin Lispro | IV |
| Glyburide Oral Tablet [Diabeta] | po |
| Humulin U Ultralente Injectable Product | subq |
| Insulin Lispro 25 UNT/ML | Subcut |
| Simvastatin 20 MG / sitagliptin 50 MG Oral Tablet | orally |
| Starlix Oral Product | oral |
| Insulin, Glulisine, Human | SQ |
| Regular Insulin, Human 100 UNT/ML Injectable Solution [Humulin R] | SC |
| 3 ML insulin detemir 100 UNT/ML Pen Injector [Levemir] | subcutaneously |
| Glynase Oral Product | PO |
| Insulin, Aspart, Human | Sub Q |
| Insulin Lispro 25 UNT/ML | sub q |
| Metformin hydrochloride 500 MG / rosiglitazone 2 MG Oral Tablet [Avandamet] | orally |
| 3 ML Insulin, Aspart Protamine, Human 70 UNT/ML / Insulin, Aspart, Human 30 UNT/ML Pen Injector [NovoLog Mix] | sq |
| insulin human, isophane 70 UNT/ML / Regular Insulin, Human 30 UNT/ML [Novolin] | sc |
| glimepiride 3 MG Oral Tablet | |
| rosiglitazone 2 MG Oral Tablet | po |

| | | |
|---|---|---|
| Regular Insulin, Human 100 UNT/ML Injectable Solution [Novolin R] | Sub q | |
| liraglutide 6 MG/ML [Victoza] | by mouth | |
| Insulin Glargine | sub Q | |
| Insulin Lispro | sub q | |
| pioglitazone 30 MG Oral Tablet | oral | |
| glimepiride 4 MG | per tube | |
| insulin detemir | by mouth | |
| Lantus Injectable Product | subq | |
| Metformin hydrochloride 1000 MG Oral Tablet | inhalation | |
| Glyburide 2.5 MG Oral Tablet [Diabeta] | p.o. | |
| 3 ML insulin human, isophane 100 UNT/ML Pen Injector [Humulin N] | Subcutaneous | |
| insulin human, isophane 16 UNT/ML | SQ | |
| Insulin Lispro 50 UNT/ML / Insulin, Protamine Lispro, Human 50 UNT/ML Injectable Suspension [Humalog Mix] | Subcut | |
| repaglinide 0.5 MG Oral Tablet [Prandin] | p.o. | |
| sitagliptin 25 MG Oral Tablet [Januvia] | PO | |
| insulin human, isophane 70 UNT/ML / Regular Insulin, Human 30 UNT/ML [Novolin] | sub Q | |
| Insulin Glargine 100 UNT/ML Injectable Solution | subq | |
| Insulin, Aspart Protamine, Human 70 UNT/ML / Insulin, Aspart, Human 30 UNT/ML Injectable Suspension [NovoLog Mix] | | |
| Glyburide | drip | |
| 24 HR Glipizide 2.5 MG Extended Release Oral Tablet | PO | |
| repaglinide 2 MG Oral Tablet [Prandin] | orally | |
| 3 ML insulin human, isophane 100 UNT/ML Prefilled Syringe [Novolin N] | | |
| sitagliptin 50 MG Oral Tablet [Januvia] | | 0 |
| Bromocriptine 2.5 MG Oral Tablet | oral | |
| 3 ML liraglutide 6 MG/ML Pen Injector [Saxenda] | | |
| 3 ML insulin detemir 100 UNT/ML Pen Injector [Levemir] | SubQ | |
| Acarbose 50 MG Oral Tablet | orally | |
| insulin human, isophane 30 UNT/ML | | |
| Insulin, Aspart Protamine, Human 70 UNT/ML / Insulin, Aspart, Human 30 UNT/ML Injectable Suspension [NovoLog Mix] | sub Q | |
| Humulin U Ultralente Injectable Product | subcutaneous | |
| Glipizide Oral Product | | |
| glimepiride 1 MG Oral Tablet | Oral | |
| Metformin | bedside | |

| | |
|---|---|
| canagliflozin 50 MG / Metformin hydrochloride 1000 MG [Invokamet] | |
| Avandaryl Oral Product | po |
| Insulin, Aspart Protamine, Human 70 UNT/ML / Insulin, Aspart, Human 30 UNT/ML Injectable Suspension | SQ |
| Metformin hydrochloride 1000 MG / sitagliptin 50 MG Oral Tablet [Janumet] | p.o. |
| Insulin Lispro 100 UNT/ML Pen Injector [Humalog] | Subcut |
| insulin human, isophane 34 UNT/ML | subq |
| canagliflozin 300 MG | |
| exenatide 0.005 MG/ACTUAT | subcut |
| insulin human, isophane 70 UNT/ML / Regular Insulin, Human 30 UNT/ML [Novolin] | IM |
| Novolin R Injectable Product | SC |
| Glipizide Oral Tablet | po |
| insulin detemir 100 UNT/ML Injectable Solution | SC |
| Regular Insulin, Human | iV |
| Metformin hydrochloride 100 MG/ML Oral Solution [Riomet] | po |
| Glyburide 1.25 MG / Metformin hydrochloride 250 MG Oral Tablet [Glucovance] | ORAL |
| Glipizide 10 MG Extended Release Oral Tablet [Glucotrol] | by mouth |
| insulin human, isophane 36 UNT/ML | subcut. |
| Glyburide 3 MG Oral Tablet [Glynase] | ORAL |
| saxagliptin 5 MG Oral Tablet [Onglyza] | |
| insulin human, isophane 70 UNT/ML / Regular Insulin, Human 30 UNT/ML [Novolin] | |
| Saxenda Injectable Product | subcutaneously |
| Bromocriptine 5 MG Oral Capsule | po |
| Metformin hydrochloride 1000 MG Oral Tablet [Glucophage] | Oral |
| miglitol 100 MG [Glyset] | |
| Metformin / sitagliptin Oral Tablet [Janumet] | by mouth |
| sitagliptin 25 MG Oral Tablet | PO |
| nateglinide | PO |
| exenatide 0.005 MG/ACTUAT [Byetta] | subQ |
| saxagliptin | |
| glimepiride 4 MG / pioglitazone 30 MG [Duetact] | |
| Farxiga Oral Product | oral |
| insulin human, isophane 100 UNT/ML | |
| Micronase Oral Product | p.o. |
| empagliflozin 25 MG Oral Tablet [Jardiance] | po |

| | | |
|---|---|---|
| insulin human, isophane 70 UNT/ML / Regular Insulin, Human 30 UNT/ML Injectable Suspension [Novolin] | SUBCUTANEOUSLY | |
| Symlin Injectable Product | subcut | |
| Insulin Lispro 50 UNT/ML | SUBCUT | |
| insulin detemir 100 UNT/ML Injectable Solution | subcu | |
| Insulin, Aspart Protamine, Human 70 UNT/ML / Insulin, Aspart, Human 30 UNT/ML Injectable Suspension [NovoLog Mix] | Subcut | |
| insulin human, isophane 40 UNT/ML | SUBQ | |
| Regular Insulin, Human | sc | |
| Osmotic 24 HR Metformin hydrochloride 500 MG Extended Release Oral Tablet [Fortamet] | oral | |
| exenatide 0.01 MG/ACTUAT [Byetta] | PO | |
| Chlorpropamide 250 MG [Diabinese] | | |
| Insulin, Zinc, Pork | | |
| insulin human, isophane 70 UNT/ML / Regular Insulin, Human 30 UNT/ML [Humulin] | SUBCUT | |
| 3 ML liraglutide 6 MG/ML Pen Injector | po | |
| Tradjenta Oral Product | oral | |
| repaglinide 2 MG Oral Tablet [Prandin] | oral | |
| Glyburide 2.5 MG Oral Tablet [Micronase] | by mouth | |
| 24 HR Glipizide 5 MG Extended Release Oral Tablet | oral | |
| 0.5 ML albiglutide 60 MG/ML Pen Injector [Tanzeum] | SUBCUTANEOUS | |
| Insulin Lispro 25 UNT/ML | SUBCUT | |
| pioglitazone 30 MG Oral Tablet [Actos] | | 0 |
| Glipizide 2.5 MG / Metformin hydrochloride 500 MG Oral Tablet | by mouth | |
| Glipizide | topical | |
| Insulin Glargine | npo. | |
| insulin human, isophane 75 UNT/ML | subcut. | |
| insulin human, isophane 70 UNT/ML / Regular Insulin, Human 30 UNT/ML [Humulin] | subq. | |
| Metformin hydrochloride 100 MG/ML Oral Solution [Riomet] | oral | |
| insulin degludec 200 UNT/ML | | |
| Bydureon Injectable Product | subcutaneously | |
| nateglinide 120 MG Oral Tablet | oral | |
| 3 ML liraglutide 6 MG/ML Pen Injector | by mouth | |
| Regular Insulin, Human 100 UNT/ML Injectable Solution [Novolin R] | subcut. | |
| Actos Pill | | |
| glimepiride 2 MG | nostril | |
| insulin human, isophane 85 UNT/ML | subcutaneous | |

| | |
|---|---|
| Insulin Lispro 100 UNT/ML [Humalog] | subq |
| insulin human, rDNA origin 3 MG/ACTUAT Inhalant Powder [Exubera] | |
| Lantus Injectable Product | SUBCUT. |
| insulin human, isophane 70 UNT/ML / Regular Insulin, Human 30 UNT/ML [Humulin] | SUB Q |
| 24 HR Metformin hydrochloride 1000 MG / pioglitazone 30 MG Extended Release Oral Tablet [Actoplus Met] | by mouth |
| Glipizide | subcutaneous |
| Insulin Lispro 25 UNT/ML | orally |
| insulin pork, isophane 100 UNT/ML Injectable Suspension [NPH Iletin II] | by mouth |
| Acarbose 50 MG Oral Tablet | |
| Ultralente Insulin, Human 100 UNT/ML Injectable Suspension [Humulin U Ultralente] | subcut. |
| Insulin Lispro 100 UNT/ML Injectable Solution [Humalog] | subcut |
| insulin pork, isophane 100 UNT/ML Injectable Suspension [NPH Iletin II] | SubQ |
| rosiglitazone 4 MG Oral Tablet | p.o. |
| 60 ACTUAT exenatide 0.005 MG/ACTUAT Pen Injector [Byetta] | subcut. |
| Glyburide 1.25 MG [Micronase] | |
| Insulin Glargine 100 UNT/ML [Lantus] | subq. |
| saxagliptin 2.5 MG Oral Tablet [Onglyza] | orally |
| alogliptin 25 MG / pioglitazone 15 MG Oral Tablet [Oseni] | by mouth |
| Glipizide 5 MG | nostril |
| 3 ML insulin detemir 100 UNT/ML Pen Injector [Levemir] | sc |
| Metformin hydrochloride 1000 MG / sitagliptin 100 MG [Janumet] | po |
| Glyxambi Oral Product | by mouth |
| Insulin, Aspart Protamine, Human 70 UNT/ML / Insulin, Aspart, Human 30 UNT/ML Injectable Suspension [NovoLog Mix] | SC |
| Metformin hydrochloride 850 MG Oral Tablet | Orally |
| Regular Insulin, Human 100 UNT/ML Injectable Solution [Humulin S] | subcutaneously |
| Glyburide 2.5 MG / Metformin hydrochloride 500 MG Oral Tablet [Glucovance] | oral |
| insulin beef, isophane 40 UNT/ML | subcutaneous |
| 3 ML liraglutide 6 MG/ML Pen Injector [Victoza] | subcu |
| Metformin hydrochloride 1000 MG / rosiglitazone 4 MG Oral Tablet [Avandamet] | ORAL |

| | |
|---|---|
| insulin human, isophane 32 UNT/ML | subq |
| Actos Oral Product | BY MOUTH |
| Insulin Lispro 100 UNT/ML Pen Injector [Humalog] | sq |
| glimepiride Oral Product | p.o. |
| Insulin Glargine 100 UNT/ML Injectable Solution [Lantus] | SUBCUTANEOUSLY |
| Insulin, Aspart, Human | PO. |
| 3 ML Regular Insulin, Human 500 UNT/ML Pen Injector [Humulin R] | |
| insulin human, isophane 100 UNT/ML Injectable Suspension [Novolin N] | subcut |
| Regular Insulin, Human Injectable Solution [Novolin R] | subcutaneously |
| Glipizide | drip |
| rosiglitazone 2 MG Oral Tablet [Avandia] | orally |
| Metformin | po. |
| Acarbose 25 MG Oral Tablet [Precose] | PO |
| Insulin, Aspart, Human 100 UNT/ML Injectable Solution [NovoLog] | SC |
| rosiglitazone 2 MG Oral Tablet [Avandia] | p.o. |
| glimepiride 8 MG | nostril |
| liraglutide | by mouth |
| Insulin Glargine | Subcut. |
| insulin pork, isophane 100 UNT/ML Injectable Suspension [NPH Iletin II] | p.o. |
| insulin human, isophane 40 UNT/ML | Subcut |
| repaglinide Oral Tablet [Prandin] | by mouth |
| Glipizide 10 MG Extended Release Oral Tablet | by mouth |
| {30 (Regular Insulin, Human 12 UNT Inhalant Powder) / 60 (Regular Insulin, Human 8 UNT Inhalant Powder) } Pack | |
| Metformin hydrochloride 1000 MG [Fortamet] | by mouth |
| Glyburide 1.5 MG Oral Tablet | orally |
| Bromocriptine 2.5 MG Oral Tablet [Parlodel] | |
| insulin detemir 100 UNT/ML [Levemir] | sub q |
| 24 HR Metformin hydrochloride 1000 MG / pioglitazone 15 MG Extended Release Oral Tablet [Actoplus Met] | |
| Glipizide 5 MG Oral Tablet [Glucotrol] | Po |
| exenatide 2 MG [Bydureon] | subcut |
| 60 ACTUAT exenatide 0.005 MG/ACTUAT Pen Injector [Byetta] | sc |
| NovoLog Injectable Product | subcut |
| Glyburide Oral Tablet | BY MOUTH |
| Glipizide Oral Tablet | oral |

| | | |
|---|---|---|
| pioglitazone 45 MG Oral Tablet [Actos] | BY MOUTH | |
| Insulin Glargine 300 UNT/ML | | |
| 0.5 ML dulaglutide 1.5 MG/ML Auto-Injector [Trulicity] | SQ | |
| glimepiride 4 MG / rosiglitazone 8 MG Oral Tablet [Avandaryl] | po | |
| rosiglitazone | | |
| pioglitazone 15 MG Oral Tablet | oral | |
| Humulin Injectable Product | Subq | |
| insulin human, isophane 70 UNT/ML / Regular Insulin, Human 30 UNT/ML Injectable Suspension [Humulin] | SUBQ | |
| Metformin Oral Tablet [Glucophage] | orally | |
| Metformin hydrochloride 500 MG / rosiglitazone 2 MG Oral Tablet [Avandamet] | ORAL | |
| glimepiride 1 MG / rosiglitazone 4 MG Oral Tablet [Avandaryl] | po | |
| Metformin hydrochloride 1000 MG / rosiglitazone 2 MG Oral Tablet [Avandamet] | PO | |
| 3 ML Regular Insulin, Human 500 UNT/ML Pen Injector [Humulin R] | SUBCUTANEOUS | |
| Insulin Glargine 100 UNT/ML Pen Injector [Lantus] | SC | |
| Insulin Glargine | Sub Q | |
| Metformin hydrochloride 1000 MG / sitagliptin 50 MG Extended Release Oral Tablet [Janumet] | | |
| sitagliptin 50 MG Oral Tablet [Januvia] | Oral | |
| 24 HR Metformin hydrochloride 750 MG Extended Release Oral Tablet [Glucophage] | PO | |
| Actoplus Met Oral Product | orally | |
| Glipizide / Metformin Oral Tablet | po | |
| Metformin hydrochloride 500 MG / rosiglitazone 4 MG Oral Tablet [Avandamet] | | |
| Regular Insulin, Human 8 UNT | subcutaneous | |
| exenatide 2 MG [Bydureon] | po | |
| Glyburide 2.5 MG Oral Tablet | | 0 |
| Regular Insulin, Human 100 UNT/ML Injectable Solution [Novolin R] | gt | |
| Insulin Lispro 100 UNT/ML Pen Injector [Humalog] | subcut | |
| insulin pork, isophane 80 UNT/ML | sq | |
| Insulin, Aspart, Human 100 UNT/ML Cartridge [NovoLog] | subcutaneous. | |
| Regular Insulin, Human 100 UNT/ML Injectable Solution [Humulin R] | subQ | |
| 60 ACTUAT exenatide 0.01 MG/ACTUAT Pen Injector [Byetta] | subcu | |
| Diabeta Oral Product | BY MOUTH | |

| | | |
|---|---|---|
| Glyburide Oral Tablet | Po | |
| pioglitazone 45 MG Oral Tablet | | |
| pioglitazone 45 MG Oral Tablet | oral | |
| Regular Insulin, Human 100 UNT/ML Injectable Solution [Humulin R] | gt | |
| Metformin hydrochloride 500 MG Oral Tablet [Glucophage] | Po | |
| insulin human, isophane 32 UNT/ML | SUBCUT | |
| Regular Insulin, Human | subcut. | |
| 1.5 ML insulin human, isophane 70 UNT/ML / Regular Insulin, Human 30 UNT/ML Prefilled Syringe | subcutaneous | |
| Glyburide / Metformin Oral Product | by mouth | |
| Glyburide 6 MG Oral Tablet [Glynase] | ORAL | |
| Bromocriptine 2.5 MG Oral Tablet | PER TUBE | |
| pioglitazone 30 MG Oral Tablet [Actos] | BY MOUTH | |
| Bromocriptine Oral Tablet | po | |
| 3 ML Insulin, Aspart Protamine, Human 70 UNT/ML / Insulin, Aspart, Human 30 UNT/ML Pen Injector [NovoLog Mix] | | 0 |
| Insulin, Glulisine, Human 100 UNT/ML Injectable Solution [Apidra] | subcut | |
| insulin human, isophane 70 UNT/ML / Regular Insulin, Human 30 UNT/ML Injectable Suspension [Humulin] | SC | |
| exenatide 0.005 MG/ACTUAT [Byetta] | SUBCUT | |
| canagliflozin 50 MG / Metformin hydrochloride 1000 MG Oral Tablet [Invokamet] | | |
| Insulin Lispro 25 UNT/ML / Insulin, Protamine Lispro, Human 75 UNT/ML Injectable Suspension [Humalog Mix] | | |
| insulin beef-pork, isophane 100 UNT/ML Injectable Suspension [Iletin NPH] | subcutaneously | |
| Cycloset Oral Product | by mouth | |
| Metformin hydrochloride 500 MG / pioglitazone 15 MG Oral Tablet [Actoplus Met] | orally | |
| Insulin, Glulisine, Human | SUBCUT | |
| sitagliptin 100 MG [Januvia] | by mouth. | |
| dapagliflozin 10 MG Oral Tablet [Farxiga] | PO | |
| exenatide 2 MG [Bydureon] | subcu | |
| 3 ML Insulin, Aspart, Human 100 UNT/ML Cartridge [NovoLog] | subcutaneous | |
| Insulin Glargine Pen Injector [Lantus] | SQ | |
| empagliflozin 10 MG / Linagliptin 5 MG [Glyxambi] | | |
| Insulin Lispro 50 UNT/ML / Insulin, Protamine Lispro, Human 50 UNT/ML Injectable Suspension [Humalog Mix] | SUBCUTANEOUS | |

| | |
|---|---|
| Glyburide 3 MG Oral Tablet [Glynase] | |
| dulaglutide 1.5 MG/ML [Trulicity] | by mouth |
| Linagliptin 5 MG Oral Tablet | PO |
| Glucotrol Oral Product | oral |
| repaglinide 0.5 MG Oral Tablet | PO |
| Insulin, Aspart Protamine, Human 70 UNT/ML / Insulin, Aspart, Human 30 UNT/ML Injectable Suspension [NovoLog Mix] | orally |
| Metformin hydrochloride 850 MG / pioglitazone 15 MG Oral Tablet [Actoplus Met] | orally |
| sitagliptin 25 MG | |
| 24 HR Glipizide 10 MG Extended Release Oral Tablet | p.o. |
| exenatide 2 MG [Bydureon] | sq |
| insulin human, isophane 70 UNT/ML / Regular Insulin, Human 30 UNT/ML Injectable Suspension [Humulin] | SQ |
| Insulin Lispro 25 UNT/ML / Insulin, Protamine Lispro, Human 75 UNT/ML Injectable Suspension [Humalog Mix] | Subq |
| Metformin hydrochloride 500 MG / rosiglitazone 1 MG Oral Tablet [Avandamet] | by mouth |
| Levemir Injectable Product | SQ |
| dulaglutide 1.5 MG/ML [Trulicity] | SC |
| 3 ML Insulin Glargine 100 UNT/ML Pen Injector [Lantus] | sq |
| Kazano Oral Product | by mouth |
| 3 ML insulin human, isophane 70 UNT/ML / Regular Insulin, Human 30 UNT/ML Pen Injector | |
| 60 ACTUAT exenatide 0.01 MG/ACTUAT Pen Injector [Byetta] | orally |
| Farxiga Oral Product | po |
| insulin human, isophane 32 UNT/ML | SQ |
| Lantus Injectable Product | Subcutaneous |
| 24 HR dapagliflozin 5 MG / Metformin hydrochloride 1000 MG Extended Release Oral Tablet [Xigduo] | oral |
| insulin human, isophane 100 UNT/ML Injectable Suspension [Novolin N] | sq |
| 0.5 ML dulaglutide 3 MG/ML Auto-Injector [Trulicity] | subcutaneous |
| 3 ML liraglutide 6 MG/ML Pen Injector | subcutaneous |
| Metformin hydrochloride 850 MG Oral Tablet | BY MOUTH |
| Exubera Inhalant Product | Inhalation |
| Insulin Lispro | p.o. |
| Glyburide Oral Tablet [Diabeta] | oral |
| sitagliptin 100 MG Oral Tablet | oral |

| | |
|---|---|
| Insulin, Aspart, Human 100 UNT/ML Injectable Solution [NovoLog] | VIA INSULIN PUMP |
| alogliptin 25 MG Oral Tablet [Nesina] | po |
| Insulin Glargine Injectable Solution [Lantus] | SubQ |
| Metformin hydrochloride 500 MG / pioglitazone 15 MG Oral Tablet | orally |
| insulin human, isophane 100 UNT/ML Injectable Suspension | |
| insulin degludec 100 UNT/ML Pen Injector [Tresiba] | |
| Metformin | PER TUBE |
| Simvastatin 20 MG / sitagliptin 100 MG Oral Tablet | orally |
| liraglutide Injectable Product | subcutaneously |
| Insulin Lispro 25 UNT/ML / Insulin, Protamine Lispro, Human 75 UNT/ML Injectable Suspension [Humalog Mix] | sub q |
| Insulin, Regular, Pork 40 UNT/ML Injectable Solution | subcut. |
| Metformin Extended Release Oral Tablet | oral |
| insulin pork, isophane 100 UNT/ML Injectable Suspension [NPH Iletin II] | sub Q |
| Amaryl Pill | by mouth |
| Regular Insulin, Human 100 UNT/ML Injectable Solution [Humulin R] | intravenous |
| insulin pork, isophane 100 UNT/ML Injectable Suspension [NPH Iletin II] | SUBq |
| dapagliflozin 10 MG | |
| Insulin, Zinc, Human 100 UNT/ML Injectable Suspension [Humulin L] | Subcutaneous |
| pioglitazone 45 MG Oral Tablet | p.o. |
| liraglutide 6 MG/ML [Victoza] | subcut. |
| glimepiride Pill | by mouth |
| Insulin Glargine | PO |
| insulin human, isophane 90 UNT/ML / Regular Insulin, Human 10 UNT/ML [Humulin] | subcut |
| Insulin Glargine 100 UNT/ML Injectable Solution | SQ |
| Insulin Lispro 50 UNT/ML | orally |
| insulin human, isophane 16 UNT/ML | subcu |
| Insulin Glargine Injectable Solution | subcu |
| exenatide 2 MG [Bydureon] | subQ |
| Glipizide 2.5 MG [Glucotrol] | p.o. |
| Metformin hydrochloride 500 MG Oral Tablet | PER TUBE |
| nateglinide Oral Product | orally |
| Humulin N Injectable Product | SUBCUT |

| | | |
|---|---|---|
| NovoLog Injectable Product | SQ | |
| Metformin hydrochloride 500 MG Oral Tablet | By Mouth | |
| insulin human, isophane 36 UNT/ML | subq | |
| Metformin | td | |
| Glipizide 10 MG | subcutaneously | |
| Metformin Oral Solution [Riomet] | | |
| Regular Insulin, Human 4 UNT Inhalant Powder [Afrezza] | inhalation | |
| Glipizide 5 MG Oral Tablet [Glucotrol] | ORAL | |
| Metformin hydrochloride 1000 MG / sitagliptin 50 MG Oral Tablet [Janumet] | | 0 |
| empagliflozin 10 MG Oral Tablet [Jardiance] | po | |
| insulin pork, isophane 70 UNT/ML / Insulin, Regular, Pork 30 UNT/ML Injectable Suspension | SUBCUTANEOUS | |
| Regular Insulin, Human 100 UNT/ML Injectable Solution [Novolin R] | subcutaneously. | |
| Invokana Oral Product | po | |
| Januvia Oral Product | p.o. | |
| insulin human, isophane 70 UNT/ML / Regular Insulin, Human 30 UNT/ML Injectable Suspension [Humulin] | sq | |
| glimepiride 2 MG / rosiglitazone 4 MG Oral Tablet [Avandaryl] | PO | |
| 60 ACTUAT exenatide 0.01 MG/ACTUAT Pen Injector [Byetta] | | 0 |
| pramlintide acetate 0.6 MG/ML Injectable Solution [Symlin] | SC | |
| alogliptin 25 MG / pioglitazone 30 MG Oral Tablet [Oseni] | by mouth | |
| empagliflozin 12.5 MG / Metformin hydrochloride 1000 MG Oral Tablet [Synjardy] | by mouth | |
| Glyburide 1.5 MG Oral Tablet | | |
| Regular Insulin, Human 100 UNT/ML Injectable Solution [Humulin R] | Subcutaneously | |
| insulin human, isophane 70 UNT/ML / Regular Insulin, Human 30 UNT/ML [Humulin] | Subq | |
| Regular Insulin, Human 500 UNT/ML Injectable Solution [Humulin R] | SC | |
| Bromocriptine 1 MG | | |
| Glyburide 5 MG / Metformin hydrochloride 500 MG Oral Tablet [Glucovance] | | 0 |
| Insulin Glargine 300 UNT/ML Pen Injector [Toujeo] | | |
| Bromocriptine | PO | |
| Insulin, Aspart Protamine, Human 70 UNT/ML / Insulin, Aspart, Human 30 UNT/ML [NovoLog Mix] | by mouth | |
| insulin detemir 100 UNT/ML Injectable Solution [Levemir] | | 0 |

| | | |
|---|---|---|
| Duetact Oral Product | po | |
| Insulin Lispro | subc | |
| miglitol 50 MG Oral Tablet [Glyset] | PO | |
| Metformin hydrochloride 100 MG/ML Oral Solution [Riomet] | | |
| glimepiride 2 MG / rosiglitazone 8 MG [Avandaryl] | | |
| 60 ACTUAT exenatide 0.005 MG/ACTUAT Pen Injector [Byetta] | | |
| insulin human, isophane 75 UNT/ML | subq | |
| 3 ML Insulin Lispro 100 UNT/ML Cartridge [Humalog] | | 0 |
| 3 ML liraglutide 6 MG/ML Pen Injector [Victoza] | Subcutaneous | |
| 24 HR Metformin hydrochloride 750 MG Extended Release Oral Tablet | p.o. | |
| Insulin Lispro 50 UNT/ML | SubQ | |
| Linagliptin 2.5 MG / Metformin hydrochloride 500 MG Oral Tablet [Jentadueto] | orally | |
| glimepiride Oral Tablet | oral | |
| Fortamet Oral Product | PO | |
| Metformin hydrochloride 750 MG Extended Release Oral Tablet [Glucophage] | orally | |
| insulin pork, isophane 100 UNT/ML Injectable Suspension [NPH Iletin II] | NPO. | |
| Glucophage Oral Product | by moutH | |
| Bromocriptine Oral Tablet | orally | |
| Glipizide 2.5 MG Extended Release Oral Tablet [Glucotrol] | | |
| insulin detemir 100 UNT/ML | SQ | |
| Glyburide 3 MG Oral Tablet | p.o. | |
| liraglutide 6 MG/ML [Victoza] | SC | |
| 3 ML Insulin Lispro 50 UNT/ML / Insulin, Protamine Lispro, Human 50 UNT/ML Pen Injector [Humalog Mix] | SUBCUTANEOUS | |
| Insulin Lispro 25 UNT/ML / Insulin, Protamine Lispro, Human 75 UNT/ML Injectable Suspension [Humalog Mix] | SubQ | |
| nateglinide 60 MG Oral Tablet [Starlix] | | |
| empagliflozin 10 MG Oral Tablet [Jardiance] | orally | |
| 3 ML Regular Insulin, Human 100 UNT/ML Prefilled Syringe [Novolin R] | | |
| glimepiride 2 MG Oral Tablet | BY MOUTH | |
| insulin human, isophane 80 UNT/ML / Regular Insulin, Human 20 UNT/ML [Human Mixtard] | subcut. | |
| liraglutide 6 MG/ML | sq | |
| pioglitazone | subcutaneously | |
| Linagliptin 5 MG Oral Tablet | po | |

| | | |
|---|---|---|
| Regular Insulin, Human 100 UNT/ML Injectable Solution [Humulin R] | by mouth | |
| Regular Insulin, Human 50 UNT/ML | IV | |
| glimepiride 4 MG / rosiglitazone 4 MG [Avandaryl] | | |
| Insulin, Aspart, Human | Subcutaneously | |
| Starlix Pill | | |
| insulin human, isophane 70 UNT/ML / Regular Insulin, Human 30 UNT/ML Injectable Suspension [Novolin] | | 0 |
| insulin human, isophane 34 UNT/ML | subcut. | |
| Victoza Injectable Product | MISCELLANEOUS | |
| insulin human, isophane 32 UNT/ML | Subcutaneous | |
| Glyburide 2.5 MG Oral Tablet | Po | |
| Glucotrol Oral Product | orally | |
| sitagliptin 50 MG Oral Tablet | Oral | |
| 24 HR Metformin hydrochloride 1000 MG / sitagliptin 100 MG Extended Release Oral Tablet [Janumet] | ORAL | |
| insulin human, isophane 75 UNT/ML | sq | |
| insulin human, isophane 70 UNT/ML / Regular Insulin, Human 30 UNT/ML Injectable Suspension [Humulin] | | 0 |
| 24 HR Metformin hydrochloride 750 MG Extended Release Oral Tablet | Oral | |
| repaglinide 0.5 MG Oral Tablet [Prandin] | oral | |
| Victoza Injectable Product | subcutaneous | |
| Regular Insulin, Human 50 UNT/ML | subcut | |
| Bromocriptine 10 MG Oral Capsule [Parlodel] | po | |
| Regular Insulin, Human 100 UNT/ML Injectable Solution [Novolin R] | | 0 |
| rosiglitazone Oral Tablet [Avandia] | by mouth | |
| Metformin 500 MG | per GT | |
| 3 ML Insulin Glargine 100 UNT/ML Pen Injector [Lantus] | SUBCUTANEOUSLY | |
| nateglinide 60 MG Oral Tablet [Starlix] | | |
| glimepiride 4 MG Oral Tablet [Amaryl] | Oral | |
| Tresiba Injectable Product | subcutaneously | |
| Regular Insulin, Human 15 UNT/ML | sq | |
| Glyburide 1.5 MG Oral Tablet | ORAL | |
| Insulin Glargine | Subcut | |
| 24 HR Metformin hydrochloride 750 MG Extended Release Oral Tablet | BY MOUTH | |
| rosiglitazone 2 MG Oral Tablet [Avandia] | Orally | |

| | |
|---|---|
| Glipizide 5 MG / Metformin hydrochloride 500 MG Oral Tablet [Metaglip] | p.o. |
| Duetact Oral Product | by mouth |
| pioglitazone Oral Tablet | PO |
| Regular Insulin, Human 10 UNT/ML | subq |
| repaglinide 2 MG Oral Tablet [Prandin] | p.o. |
| Metformin | NPO. |
| Riomet Oral Product | PO |
| alogliptin 25 MG / pioglitazone 30 MG [Oseni] | |
| Insulin Lispro | Subcut |
| Metformin hydrochloride 1000 MG / rosiglitazone 4 MG Oral Tablet [Avandamet] | p.o. |
| Metformin hydrochloride 1000 MG Extended Release Oral Tablet [Glumetza] | |
| Metformin hydrochloride 500 MG / rosiglitazone 1 MG [Avandamet] | |
| insulin pork, isophane 100 UNT/ML Injectable Suspension [NPH Iletin II] | G - tube |
| saxagliptin 2.5 MG Oral Tablet [Onglyza] | ORAL |
| Insulin, Aspart, Human 100 UNT/ML Cartridge [NovoLog] | subc |
| Bromocriptine 2.5 MG Oral Tablet [Parlodel] | |
| 3 ML liraglutide 6 MG/ML Pen Injector [Victoza] | Subq |
| nateglinide 120 MG Oral Tablet | p.o. |
| insulin human, isophane 90 UNT/ML / Regular Insulin, Human 10 UNT/ML [Human Mixtard] | SUBCUT |
| Insulin Lispro 25 UNT/ML | po |
| Acarbose 100 MG Oral Tablet [Precose] | PO |
| PrandiMet Oral Product | by mouth |
| Metformin hydrochloride 500 MG / saxagliptin 5 MG [Kombiglyze] | |
| Metformin 250 MG Oral Tablet | p.o. |
| Metformin hydrochloride 850 MG / pioglitazone 15 MG Oral Tablet | po |
| Lantus Injectable Product | sc |
| Regular Insulin, Human 100 UNT/ML Injectable Solution [Novolin R] | subcu |
| 3 ML Insulin, Aspart Protamine, Human 70 UNT/ML / Insulin, Aspart, Human 30 UNT/ML Pen Injector [NovoLog Mix] | sub q |
| Metformin hydrochloride 500 MG Oral Tablet [Glucophage] | P.O. |

| | |
|---|---|
| Metformin hydrochloride 1000 MG / rosiglitazone 2 MG Oral Tablet [Avandamet] | by mouth |
| Linagliptin | |
| Insulin Glargine 100 UNT/ML [Lantus] | npo |
| insulin human, isophane 100 UNT/ML [Humulin N] | subq |
| Glipizide 5 MG / Metformin hydrochloride 500 MG Oral Tablet [Metaglip] | |
| Glynase Oral Product | p.o. |
| 1.5 ML Insulin Glargine 300 UNT/ML Pen Injector [Toujeo] | subq |
| Regular Insulin, Human 8 UNT | sq |
| Bromocriptine 0.8 MG Oral Tablet [Cycloset] | po |
| NovoLog Mix Injectable Product | subcutaneously |
| Metformin 500 MG / ropinirole 2 MG Oral Tablet | orally |
| Metformin hydrochloride 850 MG / pioglitazone 15 MG Oral Tablet | |
| alogliptin Oral Tablet | by mouth |
| Tradjenta Oral Product | po |
| alogliptin 25 MG / pioglitazone 15 MG [Oseni] | |
| insulin detemir 100 UNT/ML Injectable Solution | subcutaneous |
| Lantus Injectable Product | subQ |
| pioglitazone 30 MG Oral Tablet | p.o. |
| insulin human, isophane 60 UNT/ML | SUBQ |
| glimepiride | NPO |
| Pramlintide | SUBCUT |
| 3 ML Insulin Glargine 100 UNT/ML Pen Injector [Lantus] | subcutaneously |
| Tradjenta Oral Product | orally |
| Metformin Oral Solution | PER TUBE |
| 3 ML liraglutide 6 MG/ML Pen Injector [Victoza] | po |
| alogliptin 12.5 MG / Metformin hydrochloride 500 MG Oral Tablet [Kazano] | by mouth |
| Novolin R Injectable Product | SUBCUTANEOUSLY |
| insulin human, isophane 60 UNT/ML | SC |
| Starlix Pill | by mouth |
| Lispro PRC Injectable Product | |
| pioglitazone 15 MG Oral Tablet | Oral |
| Regular Insulin, Human 5 UNT/ML Injectable Solution | subcutaneously |
| repaglinide 0.5 MG Oral Tablet [Prandin] | |
| Metformin 1000 MG | ng |
| Glyburide 2.5 MG Oral Tablet | BY MOUTH |
| rosiglitazone 2 MG Oral Tablet [Avandia] | oral |

| | |
|---|---|
| empagliflozin 10 MG Oral Tablet [Jardiance] | PO |
| Regular Insulin, Human 500 UNT/ML Injectable Solution [Humulin R] | subcutaneous. |
| Insulin Lispro Cartridge [Humalog] | Subcut. |
| Metformin hydrochloride 1000 MG Oral Tablet | By mouth |
| insulin pork, isophane 100 UNT/ML Injectable Suspension [NPH Iletin II] | gt |
| Insulin, Aspart, Human | Subcut |
| insulin detemir 100 UNT/ML | |
| Regular Insulin, Human 10 UNT/ML | subcutaneous |
| Insulin Lispro 50 UNT/ML | SC |
| pioglitazone Oral Tablet | oral |
| NovoLog Injectable Product | subq |
| sitagliptin 100 MG Oral Tablet [Januvia] | Po |
| Insulin, Zinc, Human | sq |
| Chlorpropamide | |
| glimepiride | by mouth |
| Regular Insulin, Human 100 UNT/ML Injectable Solution | Subcutaneous |
| Glyburide 1.25 MG Oral Tablet [Diabeta] | PO |
| Insulin Lispro Cartridge [Humalog] | Subcutaneous |
| insulin beef, isophane 80 UNT/ML Injectable Suspension | subcutaneously |
| Insulin Glargine 100 UNT/ML [Lantus] | po |
| Insulin Glargine 100 UNT/ML [Lantus] | Sub q |
| Humalog Injectable Product | Subcutaneously |
| Glipizide | IV |
| saxagliptin 2.5 MG Oral Tablet | by mouth |
| Insulin Lispro 100 UNT/ML | MISC |
| 24 HR Metformin hydrochloride 500 MG Extended Release Oral Tablet | p.o. |
| rosiglitazone 2 MG Oral Tablet | |
| Novolin N Injectable Product | Subcutaneously |
| Insulin Lispro 25 UNT/ML / Insulin, Protamine Lispro, Human 75 UNT/ML [Humalog Mix] | sc |
| Jardiance Oral Product | po |
| nateglinide 120 MG Oral Tablet | |
| Glyburide 3 MG Oral Tablet | oral |
| canagliflozin | |
| empagliflozin 25 MG / Linagliptin 5 MG Oral Tablet [Glyxambi] | |
| Bromocriptine 5 MG Oral Capsule [Parlodel] | ORAL |
| Metformin | OU |

| | | |
|---|---|---|
| Victoza Injectable Product | Subcutaneously | |
| 24 HR Metformin hydrochloride 1000 MG / saxagliptin 2.5 MG Extended Release Oral Tablet [Kombiglyze] | ORAL | |
| 24 HR empagliflozin 12.5 MG / Metformin hydrochloride 1000 MG Extended Release Oral Tablet [Synjardy] | oral | |
| Glyburide 2.5 MG / Metformin 400 MG Oral Tablet | oral | |
| Glipizide 2.5 MG / Metformin hydrochloride 500 MG Oral Tablet [Metaglip] | | |
| Glyburide 1.5 MG Oral Tablet [Glynase] | PO | |
| Glyburide / Metformin Oral Tablet [Glucovance] | oral | |
| Insulin Glargine 100 UNT/ML Injectable Solution [Lantus] | SQ | |
| 0.5 ML albiglutide 60 MG/ML Pen Injector [Tanzeum] | by mouth | |
| empagliflozin 25 MG / Linagliptin 5 MG Oral Tablet [Glyxambi] | oral | |
| 24 HR Glipizide 5 MG Extended Release Oral Tablet | p.o. | |
| Novolin R Injectable Product | Subcutaneously | |
| Insulin, Aspart, Human | Sq | |
| exenatide 0.005 MG/ACTUAT [Byetta] | SubQ | |
| insulin human, isophane 36 UNT/ML | subcu | |
| Chlorpropamide Oral Tablet | by mouth | |
| Regular Insulin, Human | SQ INFUSION | |
| dapagliflozin 5 MG Oral Tablet [Farxiga] | | |
| Modified 24 HR Metformin hydrochloride 1000 MG Extended Release Oral Tablet [Glumetza] | | |
| Metformin hydrochloride 1000 MG Oral Tablet | nostril | |
| Regular Insulin, Human | Sq | |
| Regular Insulin, Human 100 UNT/ML Injectable Solution [Velosulin] | | |
| Acarbose 50 MG Oral Tablet [Precose] | | |
| pioglitazone 15 MG Oral Tablet [Actos] | | 0 |
| Metformin hydrochloride 1000 MG Extended Release Oral Tablet [Fortamet] | oral | |
| Insulin, Aspart, Human | Subq | |
| Insulin Lispro 25 UNT/ML | sc | |
| Metformin | subQ | |
| Bromocriptine Oral Tablet | PO | |
| 0.5 ML albiglutide 60 MG/ML Pen Injector | | |
| Metformin hydrochloride 500 MG [Glucophage] | j - tube | |
| Glyburide 5 MG Oral Tablet | Po | |
| Metformin 500 MG | swallow | |
| Metformin 500 MG | IM | |

| | |
|---|---|
| Regular Insulin, Human 20 UNT/ML | iv |
| Insulin Lispro Cartridge [Humalog] | NPO |
| Glyburide 2.5 MG / Metformin 400 MG Oral Tablet | |
| Metformin hydrochloride 1000 MG Extended Release Oral Tablet [Glumetza] | oral |
| Regular Insulin, Human 100 UNT/ML Injectable Solution [Novolin R] | sub q |
| insulin detemir 100 UNT/ML [Levemir] | sc |
| pioglitazone 30 MG Oral Tablet [Actos] | Oral |
| Insulin, Glulisine, Human | subcutaneous |
| Metformin hydrochloride 850 MG | per tube |
| Glipizide 5 MG / Metformin hydrochloride 500 MG Oral Tablet [Metaglip] | oral |
| repaglinide Oral Product | orally |
| Avandia Oral Product | orally |
| 1.5 ML Regular Insulin, Human 100 UNT/ML Prefilled Syringe | iv |
| insulin detemir 100 UNT/ML [Levemir] | by mouth |
| 1.5 ML insulin human, isophane 70 UNT/ML / Regular Insulin, Human 30 UNT/ML Prefilled Syringe | subq |
| Regular Insulin, Human 10 UNT/ML | subcutaneously |
| Chlorpropamide 250 MG Oral Tablet [Diabinese] | by mouth |
| 0.5 ML dulaglutide 3 MG/ML Auto-Injector [Trulicity] | subq |
| 3 ML insulin human, isophane 100 UNT/ML Prefilled Syringe | SQ |
| canagliflozin 300 MG Oral Tablet [Invokana] | PO |
| 24 HR Metformin hydrochloride 750 MG Extended Release Oral Tablet [Glucophage] | |
| Metformin hydrochloride 1000 MG / rosiglitazone 4 MG Oral Tablet [Avandamet] | |
| Metformin | subq |
| Bromocriptine | by mouth |
| Acetohexamide 500 MG | |
| Glyburide 2.5 MG / Metformin hydrochloride 500 MG Oral Tablet | oral |
| insulin human, isophane 50 UNT/ML / Regular Insulin, Human 50 UNT/ML Injectable Suspension | |
| Synjardy Oral Product | by mouth |
| Regular Insulin, Human 15 UNT/ML | subcutaneously |
| insulin human, isophane 32 UNT/ML | SUBQ |
| glimepiride 4 MG Oral Tablet [Amaryl] | Po |
| Metformin hydrochloride 500 MG / sitagliptin 50 MG Oral Tablet [Janumet] | Po |

| | | |
|---|---|---|
| Regular Insulin, Human 20 UNT/ML | IV | |
| exenatide 2 MG [Bydureon] | IM | |
| Osmotic 24 HR Metformin hydrochloride 500 MG Extended Release Oral Tablet [Fortamet] | | |
| dapagliflozin Oral Tablet | by mouth | |
| 0.5 ML albiglutide 100 MG/ML Pen Injector [Tanzeum] | SUBCUTANEOUS | |
| Insulin Glargine | NPO. | |
| liraglutide | SC | |
| saxagliptin 5 MG Oral Tablet [Onglyza] | p.o. | |
| insulin human, isophane 60 UNT/ML | SUBCUT | |
| 3 ML Insulin Glargine 100 UNT/ML Pen Injector [Lantus] | ORAL | |
| Diabeta Pill | | |
| canagliflozin 100 MG Oral Tablet [Invokana] | | |
| Bromocriptine 2.5 MG Oral Tablet | p.o. | |
| miglitol 50 MG Oral Tablet | by mouth | |
| insulin detemir 100 UNT/ML Injectable Solution [Levemir] | SC | |
| Regular Insulin, Human 15 UNT/ML | IV | |
| sitagliptin Oral Tablet | | |
| Insulin Lispro 100 UNT/ML Pen Injector [Humalog] | subq | |
| Invokamet Oral Product | po | |
| 1.5 ML Regular Insulin, Human 100 UNT/ML Prefilled Syringe | subcutaneously | |
| Metformin | im | |
| 24 HR dapagliflozin 5 MG / Metformin hydrochloride 1000 MG Extended Release Oral Tablet [Xigduo] | ORAL | |
| Insulin, Aspart, Human 100 UNT/ML | | |
| Avandamet Oral Product | orally | |
| Linagliptin 2.5 MG / Metformin hydrochloride 1000 MG Oral Tablet [Jentadueto] | oral | |
| glimepiride | topical | |
| Chlorpropamide 100 MG [Diabinese] | | |
| Insulin, Aspart, Human | iv | |
| Glucovance Oral Product | p.o. | |
| Insulin Lispro 25 UNT/ML | subQ | |
| Regular Insulin, Human 100 UNT/ML Injectable Solution [Humulin R] | | 0 |
| Insulin Glargine | TOPICAL | |
| miglitol 25 MG Oral Tablet | po | |
| insulin human, isophane 70 UNT/ML | SC | |
| Glyset Oral Product | po | |

| | |
|---|---|
| Metformin hydrochloride 500 MG / pioglitazone 15 MG Oral Tablet [Actoplus Met] | |
| Amaryl Oral Product | Orally |
| Insulin Glargine Injectable Solution | by mouth |
| Acarbose 25 MG Oral Tablet | PO |
| insulin human, isophane 70 UNT/ML / Regular Insulin, Human 30 UNT/ML Injectable Suspension [Novolin] | Subcutaneously |
| Insulin, Aspart, Human 100 UNT/ML Cartridge [NovoLog] | Subc. |
| glimepiride 1 MG Oral Tablet | BY MOUTH |
| Insulin Lispro | subcutaneous. |
| dulaglutide 1.5 MG/ML [Trulicity] | IM |
| 3 ML Regular Insulin, Human 100 UNT/ML Prefilled Syringe [Novolin R] | subq |
| insulin human, isophane 32 UNT/ML | sq |
| empagliflozin 25 MG Oral Tablet [Jardiance] | orally |
| saxagliptin Oral Tablet | by mouth |
| insulin pork, isophane 100 UNT/ML Injectable Suspension [NPH Iletin II] | orally |
| Insulin Lispro 100 UNT/ML Injectable Solution | SUBCUTANEOUS |
| Metformin hydrochloride 500 MG [Fortamet] | po |
| insulin human, isophane 90 UNT/ML | sq |
| Metformin hydrochloride 1000 MG | |
| Linagliptin 2.5 MG / Metformin hydrochloride 1000 MG [Jentadueto] | by mouth. |
| Insulin Lispro 50 UNT/ML | by mouth |
| Metaglip Oral Product | by mouth |
| Metformin | Sl |
| Saxenda Injectable Product | |
| liraglutide | SQ |
| empagliflozin 10 MG Oral Tablet | by mouth |
| Apidra Injectable Product | subcutaneous |
| insulin human, rDNA origin 1 MG/ACTUAT Inhalant Powder [Exubera] | inhaled |
| Glyburide 1.25 MG / Metformin hydrochloride 250 MG Oral Tablet | oral |
| Regular Insulin, Human 100 UNT/ML Injectable Solution | INFUSION |
| Metformin hydrochloride 1000 MG / rosiglitazone 2 MG Oral Tablet [Avandamet] | ORAL |
| glimepiride 2 MG Oral Tablet [Amaryl] | BY MOUTH |
| Insulin Lispro Injectable Solution [Humalog] | subcut |

| | | |
|---|---|---|
| 24 HR Metformin hydrochloride 1000 MG / saxagliptin 2.5 MG Extended Release Oral Tablet [Kombiglyze] | PO | |
| insulin human, isophane 34 UNT/ML | sq | |
| Regular Insulin, Human 100 UNT/ML Injectable Solution [Humulin R] | Subcut. | |
| dapagliflozin 5 MG Oral Tablet [Farxiga] | | |
| Glyburide 1.25 MG / Metformin hydrochloride 250 MG Oral Tablet | orally | |
| 1.5 ML Insulin Glargine 300 UNT/ML Pen Injector [Toujeo] | | |
| Regular Insulin, Human 25 UNT/ML | sq | |
| rosiglitazone Oral Product | po | |
| Simvastatin 20 MG / sitagliptin 50 MG Oral Tablet | oral | |
| insulin human, isophane 100 UNT/ML Injectable Suspension [Humulin N] | Subcut | |
| Regular Insulin, Human | per tube | |
| repaglinide 1 MG Oral Tablet | oral | |
| dapagliflozin 10 MG / Metformin hydrochloride 1000 MG [Xigduo] | by mouth | |
| Insulin Glargine 100 UNT/ML | orally | |
| Regular Insulin, Human 100 UNT/ML Injectable Solution [Humulin R] | sub Q | |
| [Exubera]) / 90 (insulin human, rDNA origin 3 MG/ACTUAT Inhalant Powder [Exubera]) } Pack [Exubera Combination Pack 12] | Inhalation | |
| Insulin Lispro 200 UNT/ML [Humalog] | subcutaneously | |
| Metformin hydrochloride 100 MG/ML Oral Solution | | |
| Insulin Lispro 50 UNT/ML / Insulin, Protamine Lispro, Human 50 UNT/ML Injectable Suspension [Humalog Mix] | SubQ | |
| rosiglitazone 4 MG Oral Tablet [Avandia] | Po | |
| albiglutide 100 MG/ML [Tanzeum] | | |
| Insulin Glargine | swallow | |
| Glucovance Oral Product | oral | |
| saxagliptin 2.5 MG Oral Tablet [Onglyza] | oral | |
| glimepiride Oral Tablet [Amaryl] | orally | |
| Glyburide 2.5 MG Oral Tablet | Oral | |
| 3 ML Insulin, Aspart Protamine, Human 70 UNT/ML / Insulin, Aspart, Human 30 UNT/ML Pen Injector [NovoLog Mix] | Subcut | |
| Glipizide | gt | |
| Osmotic 24 HR Metformin hydrochloride 500 MG Extended Release Oral Tablet [Fortamet] | | 0 |

| | |
|---|---|
| Glyburide 6 MG Oral Tablet [Glynase] | |
| Metformin hydrochloride 500 MG Oral Tablet | ORALLY |
| 24 HR Metformin hydrochloride 500 MG Extended Release Oral Tablet | Oral |
| Glipizide Pill | by mouth |
| Regular Insulin, Human 20 UNT/ML | sq |
| rosiglitazone 8 MG Oral Tablet | |
| Metformin hydrochloride 500 MG / rosiglitazone 4 MG Oral Tablet | po |
| nateglinide 60 MG Oral Tablet | oral |
| 3 ML Insulin Lispro 100 UNT/ML Cartridge [Humalog] | subcutaneous |
| Metformin 1000 MG | per mouth |
| insulin human, isophane 50 UNT/ML / Regular Insulin, Human 50 UNT/ML [Humulin] | SQ |
| 24 HR dapagliflozin 10 MG / Metformin hydrochloride 1000 MG Extended Release Oral Tablet [Xigduo] | |
| Glyburide 2.5 MG Oral Tablet | Orally |
| Metformin | intramuscular |
| insulin human, isophane 40 UNT/ML | SC |
| 3 ML insulin detemir 100 UNT/ML Pen Injector [Levemir] | Sub Q |
| Insulin Glargine 100 UNT/ML | subcut |
| insulin human, isophane 16 UNT/ML | sq |
| Glipizide 2.5 MG / Metformin hydrochloride 250 MG Oral Tablet | PO |
| canagliflozin 300 MG Oral Tablet [Invokana] | |
| glimepiride Oral Tablet [Amaryl] | oral |
| glimepiride 2 MG / rosiglitazone 4 MG Oral Tablet | po |
| Metformin hydrochloride 1000 MG Extended Release Oral Tablet [Glumetza] | orally |
| Insulin Glargine | nostril |
| empagliflozin 12.5 MG | |
| insulin detemir 100 UNT/ML Injectable Solution | SUBCUTANEOUS |
| albiglutide 60 MG/ML [Tanzeum] | |
| Insulin Lispro 50 UNT/ML / Insulin, Protamine Lispro, Human 50 UNT/ML Injectable Suspension [Humalog Mix] | by mouth |
| Regular Insulin, Human 100 UNT/ML Injectable Solution [Novolin R] | by mouth |
| 3 ML insulin degludec 100 UNT/ML / liraglutide 3.6 MG/ML Pen Injector [Xultophy] | |
| Glucovance Oral Product | BY MOUTH |
| Insulin, Aspart, Human | po. |

| | | |
|---|---|---|
| insulin human, rDNA origin 3 MG/ACTUAT Inhalant Powder [Exubera] | inhaled | |
| Linagliptin 5 MG Oral Tablet [Tradjenta] | | 0 |
| Glyburide 1.5 MG Oral Tablet | oral | |
| Glyburide 2.5 MG / Phenformin 25 MG Oral Tablet | PO | |
| insulin human, isophane 70 UNT/ML / Regular Insulin, Human 30 UNT/ML [Novolin] | subcutaneously. | |
| 0.5 ML dulaglutide 1.5 MG/ML Auto-Injector [Trulicity] | SC | |
| Glyburide 5 MG / Metformin 1000 MG Oral Tablet | p.o. | |
| 24 HR Metformin hydrochloride 1000 MG / saxagliptin 5 MG Extended Release Oral Tablet [Kombiglyze] | ORAL | |
| pioglitazone Oral Tablet [Actos] | | |
| Insulin Glargine Injectable Solution [Lantus] | sc | |
| Glyburide 2.5 MG / Metformin hydrochloride 500 MG Oral Tablet [Glucovance] | p.o. | |
| insulin human, isophane 36 UNT/ML | SQ | |
| exenatide 0.005 MG/ACTUAT [Byetta] | IM | |
| 60 ACTUAT exenatide 0.01 MG/ACTUAT Pen Injector [Byetta] | Subcut | |
| insulin human, isophane 60 UNT/ML | subq. | |
| Insulin Glargine 100 UNT/ML Injectable Solution [Lantus] | by mouth | |
| Insulin Lispro 25 UNT/ML | SUBQ | |
| Metformin 500 MG | subcutaneous | |
| glimepiride 4 MG Oral Tablet | BY MOUTH | |
| Victoza Injectable Product | subq | |
| insulin pork, isophane 100 UNT/ML Injectable Suspension [NPH Iletin II] | subcu. | |
| Avandaryl Oral Product | PO | |
| 3 ML Insulin Lispro 200 UNT/ML Pen Injector [Humalog] | by mouth | |
| Bromocriptine 10 MG [Parlodel] | | |
| Regular Insulin, Human 6 UNT/ML | sq | |
| Insulin, Aspart Protamine, Human 70 UNT/ML / Insulin, Aspart, Human 30 UNT/ML Injectable Suspension | subcutaneous | |
| pramlintide acetate 0.6 MG/ML [Symlin] | Subcut | |
| 3 ML Insulin, Aspart Protamine, Human 70 UNT/ML / Insulin, Aspart, Human 30 UNT/ML Pen Injector [NovoLog Mix] | sc | |
| insulin human, isophane 70 UNT/ML / Regular Insulin, Human 30 UNT/ML [Humulin] | PO | |
| exenatide 0.005 MG/ACTUAT [Byetta] | Subq | |
| Regular Insulin, Human 4 UNT Inhalant Powder [Afrezza] | | |
| pramlintide acetate 0.6 MG/ML Injectable Solution | Subcutaneous | |

| | |
|---|---|
| Parlodel Oral Product | orally |
| dulaglutide 3 MG/ML | |
| liraglutide | oral |
| Bromocriptine 25 MG Oral Tablet | po |
| Metformin | PO. |
| Diabinese Oral Product | po |
| Metformin Pill | po |
| insulin pork, isophane 80 UNT/ML | subcu |
| insulin human, isophane 70 UNT/ML / Regular Insulin, Human 30 UNT/ML [Novolin] | po |
| 0.5 ML albiglutide 100 MG/ML Pen Injector | subq |
| Metformin hydrochloride 500 MG / saxagliptin 5 MG [Kombiglyze] | PO |
| Regular Insulin, Human 12 UNT | sq |
| Glyburide / Metformin Oral Tablet | |
| 0.5 ML albiglutide 60 MG/ML Pen Injector [Tanzeum] | subQ |
| Glyburide / Metformin Pill | |
| glimepiride | gt |
| Insulin Glargine | topical |
| Insulin Lispro 25 UNT/ML / Insulin, Protamine Lispro, Human 75 UNT/ML [Humalog Mix] | SUBCUT |
| Simvastatin 20 MG / sitagliptin 50 MG Oral Tablet | PO |
| insulin pork, isophane 100 UNT/ML Injectable Suspension [NPH Iletin II] | im |
| glimepiride 4 MG | subq |
| insulin human, isophane 70 UNT/ML / Regular Insulin, Human 30 UNT/ML [Humulin] | oral |
| Regular Insulin, Human 100 UNT/ML Injectable Solution [Novolin R] | SC |
| Metformin hydrochloride 500 MG / pioglitazone 15 MG Oral Tablet [Actoplus Met] | oral |
| NovoLog Injectable Product | SUBCUTANEOUSLY |
| nateglinide Oral Tablet | |
| Acarbose 50 MG Oral Tablet [Precose] | |
| Glipizide 5 MG | per mouth |
| dapagliflozin 10 MG Oral Tablet | |
| insulin human, isophane 85 UNT/ML | subcu |
| Glyburide 2.5 MG | per tube |
| miglitol 25 MG Oral Tablet [Glyset] | |

| | | |
|---|---|---|
| insulin human, isophane 50 UNT/ML / Regular Insulin, Human 50 UNT/ML Injectable Suspension [Humulin] | SQ | |
| Insulin Lispro 200 UNT/ML [Humalog] | subcut | |
| glimepiride 1 MG Oral Tablet [Amaryl] | Orally | |
| nateglinide 30 MG Oral Tablet | by mouth | |
| Humalog Mix Injectable Product | subcut | |
| 60 ACTUAT exenatide 0.005 MG/ACTUAT Pen Injector [Byetta] | subcu | |
| Insulin Glargine Injectable Solution | Subcutaneously | |
| Insulin Glargine Injectable Solution [Lantus] | sq | |
| 3 ML Insulin Lispro 100 UNT/ML Cartridge [Humalog] | sub Q | |
| insulin pork, isophane 100 UNT/ML Injectable Suspension [NPH Iletin II] | bedside | |
| Regular Insulin, Human 500 UNT/ML Injectable Solution [Humulin R] | by mouth | |
| Metformin | Im | |
| Insulin, Aspart, Human | IM | |
| Glucovance Pill | | |
| Starlix Oral Product | p.o. | |
| 3 ML Regular Insulin, Human 100 UNT/ML Prefilled Syringe [Novolin R] | Subcutaneous | |
| pramlintide acetate 0.6 MG/ML [Symlin] | subcut. | |
| Insulin Glargine | per tube | |
| 3 ML insulin human, isophane 100 UNT/ML Prefilled Syringe | by mouth | |
| Insulin, Aspart, Human 100 UNT/ML Injectable Solution | subcu | |
| Insulin Lispro 50 UNT/ML / Insulin, Protamine Lispro, Human 50 UNT/ML Injectable Suspension [Humalog Mix] | sub Q | |
| Insulin Glargine 100 UNT/ML [Lantus] | Subcutaneous | |
| 24 HR Metformin hydrochloride 1000 MG / saxagliptin 2.5 MG Extended Release Oral Tablet [Kombiglyze] | orally | |
| Glyburide 1.25 MG / Metformin hydrochloride 250 MG Oral Tablet | PO | |
| Insulin Lispro | drip. | |
| exenatide 0.005 MG/ACTUAT [Byetta] | SUBQ | |
| Acetohexamide 500 MG [Dymelor] | | |
| pramlintide acetate 0.6 MG/ML Injectable Solution [Symlin] | Subcu | |
| 24 HR Metformin hydrochloride 1000 MG / saxagliptin 2.5 MG Extended Release Oral Tablet [Kombiglyze] | | |
| repaglinide 2 MG Oral Tablet | orally | |
| dapagliflozin 10 MG Oral Tablet [Farxiga] | | 0 |
| 3 ML Insulin, Aspart, Human 100 UNT/ML Cartridge [NovoLog] | oral | |

| | |
|---|---|
| insulin human, isophane 100 UNT/ML Injectable Suspension [Novolin N] | subcut. |
| 3 ML liraglutide 6 MG/ML Pen Injector [Victoza] | IM |
| Insulin, Aspart, Human 100 UNT/ML Injectable Solution [NovoLog] | IV |
| Glyburide 1.25 MG Oral Tablet [Diabeta] | oral |
| dulaglutide | |
| Metformin 250 MG Oral Tablet | |
| Glipizide 2.5 MG / Metformin hydrochloride 500 MG [Metaglip] | |
| empagliflozin 12.5 MG / Metformin hydrochloride 1000 MG [Synjardy] | |
| insulin pork, isophane 100 UNT/ML Injectable Suspension [NPH Iletin II] | drip. |
| canagliflozin 150 MG / Metformin hydrochloride 1000 MG Oral Tablet [Invokamet] | by mouth |
| Insulin, Zinc, Pork | SQ |
| Bromocriptine 2.5 MG Extended Release Oral Capsule | |
| Acarbose 25 MG Oral Tablet | orally |
| glimepiride 2 MG Oral Tablet [Amaryl] | Oral |
| insulin human, isophane 70 UNT/ML / Regular Insulin, Human 30 UNT/ML Injectable Suspension [Humulin] | subcu |
| Regular Insulin, Human 50 UNT/ML | INFUSION |
| 3 ML insulin human, isophane 70 UNT/ML / Regular Insulin, Human 30 UNT/ML Pen Injector [Humulin] | subq |
| Insulin Lispro 100 UNT/ML Injectable Solution [Humalog] | VIA INSULIN PUMP |
| Toujeo Injectable Product | |
| 24 HR Metformin hydrochloride 1000 MG / sitagliptin 50 MG Extended Release Oral Tablet [Janumet] | by mouth |
| saxagliptin 5 MG Oral Tablet | po |
| Insulin Lispro 100 UNT/ML Pen Injector [Humalog] | SC |
| Metformin hydrochloride 1000 MG / rosiglitazone 2 MG Oral Tablet [Avandamet] | orally |
| glimepiride 4 MG | by mouth. |
| sitagliptin 100 MG Oral Tablet | |
| dulaglutide 3 MG/ML [Trulicity] | SQ |
| Insulin Lispro Cartridge [Humalog] | SUBCU |
| Chlorpropamide 250 MG Oral Tablet | PO |
| Metaglip Oral Product | po |
| Metformin hydrochloride 500 MG [Glucophage] | by mouth. |

| | | |
|---|---|---|
| Ultralente Insulin, Human 100 UNT/ML Injectable Suspension [Humulin U Ultralente] | SUBCUT | |
| sitagliptin Oral Tablet | oral | |
| Insulin Glargine 300 UNT/ML | subcutaneously | |
| {60 (Regular Insulin, Human 4 UNT Inhalant Powder) / 30 (Regular Insulin, Human 8 UNT Inhalant Powder) } Pack | | |
| Insulin Lispro Cartridge [Humalog] | SubQ | |
| exenatide 0.005 MG/ACTUAT Pen Injector [Byetta] | SUBCUT | |
| insulin human, isophane 70 UNT/ML / Regular Insulin, Human 30 UNT/ML Injectable Suspension [Humulin] | SubQ | |
| canagliflozin 300 MG Oral Tablet [Invokana] | | 0 |
| Regular Insulin, Human | NPO | |
| Regular Insulin, Human 10 UNT/ML | SUBCUT | |
| Insulin, Aspart, Human 100 UNT/ML Injectable Solution [NovoLog] | CONTINUOUS INFUSION | |
| glimepiride 3 MG Oral Tablet [Amaryl] | p.o. | |
| exenatide 0.01 MG/ACTUAT [Byetta] | IM | |
| Glucophage Oral Product | Orally | |
| 1.5 ML Regular Insulin, Human 100 UNT/ML Prefilled Syringe | SQ | |
| 1.5 ML Insulin Glargine 300 UNT/ML Pen Injector [Toujeo] | SQ | |
| Bromocriptine 10 MG Oral Capsule | orally | |
| glimepiride Oral Product | Oral | |
| Insulin, Aspart, Human 100 UNT/ML Injectable Solution | subc. | |
| pioglitazone 45 MG Oral Tablet | BY MOUTH | |
| 3 ML Insulin Glargine 100 UNT/ML Pen Injector [Lantus] | subQ | |
| Insulin Lispro 50 UNT/ML | sc | |
| pramlintide acetate 0.6 MG/ML [Symlin] | subcut | |
| insulin human, isophane 70 UNT/ML / Regular Insulin, Human 30 UNT/ML [Humulin] | SUBCUTANEOUS | |
| Acarbose 25 MG Oral Tablet [Precose] | oral | |
| insulin human, isophane 50 UNT/ML / Regular Insulin, Human 50 UNT/ML Injectable Suspension [Humulin] | subcutaneously | |
| Regular Insulin, Human 100 UNT/ML Injectable Solution [Humulin R] | SubQ | |
| Regular Insulin, Human 100 UNT/ML Injectable Solution [Novolin R] | Subcutaneously | |
| Chlorpropamide 100 MG Oral Tablet [Diabinese] | po | |
| 60 ACTUAT exenatide 0.01 MG/ACTUAT Pen Injector [Byetta] | Subq | |
| Actoplus Met Oral Product | oral | |
| Insulin, Aspart, Human 100 UNT/ML Cartridge [NovoLog] | Sub Q | |

| | | |
|---|---|---|
| Glyburide / Metformin Oral Product | orally | |
| 1.5 ML Regular Insulin, Human 100 UNT/ML Prefilled Syringe | subcut | |
| 60 ACTUAT exenatide 0.01 MG/ACTUAT Pen Injector [Byetta] | IM | |
| Regular Insulin, Human 100 UNT/ML Injectable Solution | iv | |
| nateglinide 120 MG Oral Tablet [Starlix] | | 0 |
| Metformin 500 MG | po. | |
| Regular Insulin, Human | SubQ | |
| glimepiride Oral Product | | |
| Metformin | drip. | |
| Insulin, Aspart, Human 100 UNT/ML Injectable Solution [NovoLog] | SQ | |
| 3 ML Insulin Lispro 200 UNT/ML Pen Injector | | |
| Metformin Oral Solution | BY MOUTH | |
| insulin human, isophane 100 UNT/ML Injectable Suspension [Humulin N] | Sub Q | |
| Metformin hydrochloride 500 MG / saxagliptin 5 MG [Kombiglyze] | by mouth | |
| exenatide 2 MG [Bydureon] | sc | |
| sitagliptin 100 MG Oral Tablet | p.o. | |
| Insulin Glargine 100 UNT/ML Injectable Solution | SC | |
| glimepiride Oral Tablet [Amaryl] | po | |
| liraglutide 6 MG/ML [Victoza] | subcutaneous. | |
| Metformin | inhalation | |
| dapagliflozin 10 MG [Farxiga] | per tube | |
| insulin human, isophane 28 UNT/ML | Subcut | |
| insulin human, isophane 70 UNT/ML / Regular Insulin, Human 30 UNT/ML Injectable Suspension [Humulin] | subQ | |
| NovoLog Injectable Product | sq | |
| pioglitazone 15 MG Oral Tablet | p.o. | |
| Insulin Glargine 100 UNT/ML Injectable Solution [Lantus] | sc | |
| Linagliptin 2.5 MG / Metformin hydrochloride 1000 MG Oral Tablet [Jentadueto] | | |
| insulin human, isophane 100 UNT/ML Pen Injector [Humulin N] | Subcutaneous | |
| Insulin, Aspart, Human | | 0 |
| 3 ML insulin human, isophane 100 UNT/ML Pen Injector [Humulin N] | SubQ | |
| Metformin | sublingual | |
| Glipizide 2.5 MG [Glucotrol] | oral | |
| 24 HR Metformin hydrochloride 500 MG / sitagliptin 50 MG Extended Release Oral Tablet [Janumet] | oral | |

| | |
|---|---|
| Insulin Glargine | inhalation |
| Glucophage Pill | po |
| 24 HR Glipizide 2.5 MG Extended Release Oral Tablet [Glucotrol] | orally |
| canagliflozin 150 MG / Metformin hydrochloride 1000 MG Oral Tablet [Invokamet] | ORAL |
| Insulin Lispro 100 UNT/ML Cartridge [Humalog] | |
| Insulin Glargine Injectable Solution [Lantus] | subcut. |
| Glipizide Oral Tablet [Glucotrol] | oral |
| Victoza Injectable Product | sub q |
| Insulin Lispro Cartridge [Humalog] | oral |
| insulin pork, isophane 80 UNT/ML | SUBCUT |
| Insulin, Aspart, Human | SubQ |
| glimepiride 4 MG / rosiglitazone 4 MG Oral Tablet [Avandaryl] | orally |
| Metformin hydrochloride 500 MG Extended Release Oral Tablet | gt |
| canagliflozin Oral Tablet | by mouth |
| Regular Insulin, Human | Intravenous |
| Novolin N Injectable Product | SC |
| exenatide 0.01 MG/ACTUAT Pen Injector | |
| alogliptin 25 MG Oral Tablet [Nesina] | orally |
| Glyburide 5 MG / Metformin hydrochloride 500 MG Oral Tablet | p.o. |
| Regular Insulin, Human | IM |
| Regular Insulin, Human 25 UNT/ML | sub q |
| Insulin Glargine 100 UNT/ML [Lantus] | orally |
| Insulin Lispro 25 UNT/ML | subcutaneously |
| Insulin Lispro 50 UNT/ML | SUBQ |
| Glipizide 2.5 MG Extended Release Oral Tablet | p.o. |
| Riomet Oral Product | by mouth |
| Ultralente Insulin, Human 100 UNT/ML Injectable Suspension [Humulin U Ultralente] | subcutaneously |
| 0.5 ML dulaglutide 3 MG/ML Auto-Injector [Trulicity] | |
| Chlorpropamide 250 MG Oral Tablet | po |
| Metformin | subcut |
| 3 ML Insulin, Aspart Protamine, Human 70 UNT/ML / Insulin, Aspart, Human 30 UNT/ML Pen Injector [NovoLog Mix] | by mouth |
| 3 ML Insulin, Glulisine, Human 100 UNT/ML Pen Injector [Apidra] | SC |
| sitagliptin 25 MG Oral Tablet | by mouth |
| Glipizide 2.5 MG / Metformin hydrochloride 250 MG Oral Tablet [Metaglip] | po |
| Acarbose Oral Tablet | po |
| dulaglutide 3 MG/ML [Trulicity] | subq |

| | |
|---|---|
| Regular Insulin, Human | oral |
| Linagliptin 5 MG Oral Tablet [Tradjenta] | p.o. |
| insulin human, isophane 70 UNT/ML / Regular Insulin, Human 30 UNT/ML [Humulin] | |
| Insulin, Zinc, Human | subcut |
| 24 HR Metformin hydrochloride 500 MG / saxagliptin 5 MG Extended Release Oral Tablet [Kombiglyze] | |
| insulin human, isophane 100 UNT/ML Injectable Suspension [Novolin N] | SC |
| Glyburide 6 MG Oral Tablet [Glynase] | oral |
| pramlintide acetate 0.6 MG/ML Injectable Solution [Symlin] | SUBCUT |
| Insulin, Aspart, Human 100 UNT/ML Cartridge [NovoLog] | sub Q. |
| Linagliptin / Metformin Oral Product | by mouth |
| 24 HR Metformin hydrochloride 500 MG Extended Release Oral Tablet [Glucophage] | po |
| Metformin 1000 MG | subcutaneously |
| Metformin hydrochloride 100 MG/ML | subcutaneously |
| pioglitazone 30 MG [Actos] | per tube |
| insulin human, isophane 40 UNT/ML | sub Q |
| Glipizide 2.5 MG / Metformin hydrochloride 250 MG [Metaglip] | |
| Glipizide 5 MG Oral Tablet | Orally |
| Linagliptin 2.5 MG / Metformin hydrochloride 850 MG Oral Tablet [Jentadueto] | by mouth |
| insulin human, isophane 16 UNT/ML | subcutaneous. |
| Glipizide / Metformin Oral Tablet | by mouth |
| 3 ML Insulin Lispro 50 UNT/ML / Insulin, Protamine Lispro, Human 50 UNT/ML Pen Injector [Humalog Mix] | subcutaneous |
| insulin human, isophane 70 UNT/ML / Regular Insulin, Human 30 UNT/ML Injectable Suspension | |
| exenatide 0.01 MG/ACTUAT | by mouth |
| canagliflozin 100 MG Oral Tablet | by mouth |
| Starlix Oral Product | orally |
| Linagliptin 2.5 MG / Metformin hydrochloride 1000 MG Oral Tablet [Jentadueto] | ng |
| Glipizide 2.5 MG / Metformin hydrochloride 500 MG Oral Tablet | po |
| Glumetza Oral Product | |
| Acarbose 25 MG Oral Tablet | oral |
| Bromocriptine 0.8 MG Oral Tablet [Cycloset] | |
| Metformin hydrochloride 500 MG / repaglinide 2 MG Oral Tablet [PrandiMet] | by mouth |

| | | |
|---|---|---|
| Insulin, Aspart, Human | bedside | |
| glimepiride | subq | |
| insulin human, isophane 70 UNT/ML / Regular Insulin, Human 30 UNT/ML [Humulin] | sQ | |
| 24 HR Metformin hydrochloride 1000 MG / saxagliptin 2.5 MG Extended Release Oral Tablet [Kombiglyze] | oral | |
| 3 ML Insulin Lispro 25 UNT/ML / Insulin, Protamine Lispro, Human 75 UNT/ML Pen Injector [Humalog Mix] | | 0 |
| insulin detemir | sq | |
| miglitol 25 MG Oral Tablet | | |
| Glyburide 2.5 MG Oral Tablet [Micronase] | p.o. | |
| Farxiga Pill | | |
| exenatide 0.005 MG/ACTUAT [Byetta] | PO | |
| dapagliflozin 5 MG Oral Tablet | by mouth | |
| 3 ML Insulin Lispro 25 UNT/ML / Insulin, Protamine Lispro, Human 75 UNT/ML Pen Injector [Humalog Mix] | subcutaneous | |
| insulin pork, isophane 100 UNT/ML Injectable Suspension [NPH Iletin II] | Subcut | |
| rosiglitazone 8 MG Oral Tablet | p.o. | |
| Glyburide 2.5 MG Oral Tablet [Micronase] | PO | |
| insulin human, isophane 40 UNT/ML | sc | |
| insulin human, isophane 50 UNT/ML / Regular Insulin, Human 50 UNT/ML [Humulin] | subcut | |
| canagliflozin 100 MG Oral Tablet [Invokana] | | 0 |
| 24 HR dapagliflozin 10 MG / Metformin hydrochloride 1000 MG Extended Release Oral Tablet [Xigduo] | oral | |
| insulin degludec 200 UNT/ML [Tresiba] | | |
| Insulin, Aspart, Human | SQ INFUSION | |
| canagliflozin 50 MG / Metformin hydrochloride 1000 MG Oral Tablet [Invokamet] | oral | |
| 60 ACTUAT exenatide 0.005 MG/ACTUAT Pen Injector [Byetta] | sub q | |
| Regular Insulin, Human 24 UNT/ML | subcut | |
| exenatide Injection | subcutaneously | |
| 3 ML insulin human, isophane 100 UNT/ML Prefilled Syringe | COMBO ROUTE) | |
| insulin human, isophane 100 UNT/ML Injectable Suspension | subcut | |
| Acarbose 25 MG [Precose] | G - tube | |
| repaglinide 0.5 MG Oral Tablet | oral | |
| Metformin hydrochloride 1000 MG / sitagliptin 50 MG Oral Tablet | | |

| | | |
|---|---|---|
| Linagliptin 2.5 MG / Metformin hydrochloride 1000 MG Oral Tablet [Jentadueto] | orally | |
| Insulin Glargine 100 UNT/ML Injectable Solution | SUBCUT | |
| insulin human, isophane 90 UNT/ML | subcutaneous | |
| rosiglitazone 4 MG Oral Tablet [Avandia] | Orally | |
| Novolin Injectable Product | SUBCUT | |
| Glyburide 3 MG Oral Tablet | BY MOUTH | |
| insulin, regular, beef-pork 40 UNT/ML Inhalant Solution | SUBCUT | |
| Victoza Injectable Product | COMBO ROUTE) | |
| Januvia Pill | by mouth | |
| Metformin hydrochloride 500 MG / rosiglitazone 4 MG Oral Tablet | | |
| insulin human, isophane 100 UNT/ML Injectable Suspension [Novolin N] | | 0 |
| Actoplus Met Pill | | |
| 3 ML insulin human, isophane 100 UNT/ML Pen Injector [Humulin N] | by mouth | |
| Insulin Lispro 25 UNT/ML | sub Q | |
| Insulin Glargine 100 UNT/ML | SQ | |
| 0.5 ML albiglutide 100 MG/ML Pen Injector [Tanzeum] | by mouth | |
| Invokana Oral Product | orally | |
| Metformin Oral Solution | Orally | |
| glimepiride 2 MG Oral Tablet | Po | |
| 3 ML insulin human, isophane 100 UNT/ML Pen Injector [Humulin N] | subQ | |
| Prandin Oral Product | p.o. | |
| saxagliptin 2.5 MG Oral Tablet [Onglyza] | | |
| dapagliflozin 10 MG Oral Tablet [Farxiga] | | |
| Glyburide 4.5 MG | | |
| Pramlintide | subcut | |
| Lantus Injectable Product | sub Q | |
| Insulin Lispro | IM | |
| Insulin Lispro Injectable Solution [Humalog] | subcu | |
| 3 ML insulin human, isophane 100 UNT/ML Prefilled Syringe | SubQ | |
| Insulin Lispro | TOPICAL | |
| 3 ML Insulin, Aspart Protamine, Human 70 UNT/ML / Insulin, Aspart, Human 30 UNT/ML Pen Injector [NovoLog Mix] | subcu | |
| miglitol | | |
| pioglitazone Oral Tablet [Actos] | po | |
| Insulin Glargine Injectable Solution | sub q | |

| | |
|---|---|
| exenatide 0.01 MG/ACTUAT [Byetta] | orally |
| glimepiride 2 MG / pioglitazone 30 MG Oral Tablet [Duetact] | orally |
| dulaglutide 1.5 MG/ML | |
| repaglinide 1 MG Oral Tablet [Prandin] | p.o. |
| Metformin hydrochloride 1000 MG [Glucophage] | po. |
| sitagliptin 50 MG Oral Tablet [Januvia] | BY MOUTH |
| canagliflozin 300 MG Oral Tablet [Invokana] | BY MOUTH |
| Insulin, Glulisine, Human 100 UNT/ML Injectable Solution [Apidra] | subcut. |
| 3 ML Insulin Lispro 200 UNT/ML Pen Injector | NPO |
| sitagliptin 100 MG Oral Tablet | Oral |
| glimepiride 4 MG / rosiglitazone 4 MG Oral Tablet [Avandaryl] | ORAL |
| Glipizide 2.5 MG / Metformin hydrochloride 500 MG Oral Tablet [Metaglip] | oral |
| glimepiride 3 MG Oral Tablet [Amaryl] | |
| Glyburide 2.5 MG / Metformin 400 MG Oral Tablet | PO |
| insulin human, isophane 28 UNT/ML | SC |
| Metformin hydrochloride 500 MG / sitagliptin 50 MG Oral Tablet [Janumet] | BY MOUTH |
| insulin human, isophane 40 UNT/ML | SUBCUTANEOUS |
| Glipizide 5 MG Oral Tablet | Po |
| Insulin, Zinc, Human 100 UNT/ML Injectable Suspension [Novolin L] | SQ |
| Insulin Lispro 100 UNT/ML Injectable Solution [Humalog] | subq. |
| Insulin Lispro | |
| glimepiride 4 MG / pioglitazone 30 MG Oral Tablet | |
| Insulin Lispro 100 UNT/ML Injectable Solution [Humalog] | Subcut |
| repaglinide 1 MG Oral Tablet | |
| insulin human, isophane 75 UNT/ML | sub q |
| Pramlintide Injectable Solution [Symlin] | |
| saxagliptin 5 MG Oral Tablet | PO |
| Metformin hydrochloride 500 MG Extended Release Oral Tablet [Fortamet] | orally |
| Insulin, Glulisine, Human 100 UNT/ML Injectable Solution [Apidra] | SQ |
| insulin beef, isophane 40 UNT/ML | subq |
| Tolazamide 250 MG | |
| empagliflozin 25 MG Oral Tablet [Jardiance] | PO |
| Insulin Lispro | PO. |
| Insulin Glargine 100 UNT/ML [Lantus] | SQ. |

| | | |
|---|---|---|
| Glyburide Oral Tablet [Micronase] | by mouth | |
| Bromocriptine Oral Tablet | | |
| insulin human, isophane 50 UNT/ML / Regular Insulin, Human 50 UNT/ML [Humulin] | sq | |
| pioglitazone Oral Tablet [Actos] | BY MOUTH | |
| 24 HR Glipizide 2.5 MG Extended Release Oral Tablet [Glucotrol] | p.o. | |
| Insulin, Glulisine, Human 100 UNT/ML Injectable Solution [Apidra] | | 0 |
| glimepiride 4 MG / rosiglitazone 4 MG Oral Tablet [Avandaryl] | p.o. | |
| Regular Insulin, Human 20 UNT/ML | SQ | |
| Glipizide 5 MG Oral Tablet [Glucotrol] | Orally | |
| insulin human, isophane 70 UNT/ML / Regular Insulin, Human 30 UNT/ML [Novolin] | oral | |
| Insulin, Aspart, Human | subcutaneous. | |
| canagliflozin 150 MG / Metformin hydrochloride 500 MG Oral Tablet [Invokamet] | po | |
| Lantus Injectable Product | IM | |
| Tradjenta Oral Product | PO | |
| repaglinide Oral Tablet [Prandin] | | |
| Insulin, Aspart, Human | swallow | |
| Regular Insulin, Human 100 UNT/ML Injectable Solution | intravenously | |
| Insulin Lispro 50 UNT/ML | subq. | |
| Metformin hydrochloride 500 MG / rosiglitazone 2 MG Oral Tablet [Avandamet] | Oral | |
| canagliflozin Oral Tablet [Invokana] | | |
| pramlintide acetate 0.6 MG/ML Injectable Solution [Symlin] | sub q | |
| Insulin Glargine | bedside | |
| 3 ML insulin human, isophane 100 UNT/ML Pen Injector [Humulin N] | subcutaneously | |
| insulin human, isophane 40 UNT/ML | subQ | |
| glimepiride 2 MG / rosiglitazone 4 MG Oral Tablet [Avandaryl] | oral | |
| insulin human, isophane 70 UNT/ML / Regular Insulin, Human 30 UNT/ML [Humulin] | Subcut | |
| Regular Insulin, Human | drip | |
| empagliflozin 10 MG / Linagliptin 5 MG Oral Tablet [Glyxambi] | oral | |
| Insulin Glargine 100 UNT/ML Injectable Solution | subQ | |
| Insulin Lispro Injectable Solution | Subcutaneous | |
| nateglinide Oral Tablet [Starlix] | | |
| Metformin | swallow. | |

| | |
|---|---|
| insulin human, isophane 100 UNT/ML Injectable Suspension [Novolin N] | subq |
| Metformin hydrochloride 500 MG Oral Tablet | p.o |
| Regular Insulin, Human 100 UNT/ML Injectable Solution [Humulin R] | npo |
| Metformin | ng |
| Regular Insulin, Human 15 UNT/ML | subcutaneous |
| Insulin, Aspart Protamine, Human 70 UNT/ML / Insulin, Aspart, Human 30 UNT/ML Injectable Suspension [NovoLog Mix] | by mouth |
| 3 ML Insulin Lispro 200 UNT/ML Pen Injector [Humalog] | subq |
| insulin human, isophane 100 UNT/ML Injectable Suspension | SC |
| Tolbutamide 250 MG Oral Tablet | |
| Glyburide 5 MG / Phenformin 50 MG Oral Tablet | by mouth |
| Metformin hydrochloride 1000 MG [Fortamet] | po |
| Glyburide 2.5 MG Oral Tablet [Micronase] | orally |
| Metformin Oral Tablet | BY MOUTH |
| empagliflozin 12.5 MG / Metformin hydrochloride 1000 MG Oral Tablet [Synjardy] | |
| insulin human, isophane 34 UNT/ML | Subcut |
| 3 ML liraglutide 6 MG/ML Pen Injector [Saxenda] | subq |
| insulin human, isophane 50 UNT/ML / Regular Insulin, Human 50 UNT/ML [Humulin] | subQ |
| Insulin Lispro 25 UNT/ML / Insulin, Protamine Lispro, Human 75 UNT/ML [Humalog Mix] | IM |
| Insulin Lispro 25 UNT/ML / Insulin, Protamine Lispro, Human 75 UNT/ML Injectable Suspension [Humalog Mix] | SUB Q |
| Glyburide Pill | by mouth |
| insulin human, isophane 70 UNT/ML / Regular Insulin, Human 30 UNT/ML [Novolin] | Sq |
| Metformin | |
| glimepiride 1 MG / rosiglitazone 4 MG Oral Tablet [Avandaryl] | orally |
| Micronase Oral Product | by mouth |
| saxagliptin 5 MG Oral Tablet [Onglyza] | BY MOUTH |
| Metformin hydrochloride 850 MG / pioglitazone 15 MG Oral Tablet [Actoplus Met] | oral |
| Insulin Lispro 50 UNT/ML / Insulin, Protamine Lispro, Human 50 UNT/ML Injectable Suspension [Humalog Mix] | SC |
| Insulin Glargine Injectable Solution | SUBCUT. |
| Metformin hydrochloride 500 MG Extended Release Oral Tablet [Glucophage] | by mouth |

| | | |
|---|---|---|
| insulin human, isophane 16 UNT/ML | SC | |
| insulin human, isophane 100 UNT/ML Injectable Suspension [Humulin N] | sc | |
| insulin human, isophane 70 UNT/ML | sc | |
| glimepiride 1 MG | nostril | |
| insulin pork, isophane 100 UNT/ML Injectable Suspension [NPH Iletin II] | po. | |
| Insulin Glargine 100 UNT/ML | sq | |
| Regular Insulin, Human 12 UNT | Sq | |
| 0.5 ML dulaglutide 1.5 MG/ML Auto-Injector [Trulicity] | sc | |
| insulin pork, isophane 100 UNT/ML Injectable Suspension [NPH Iletin II] | | 0 |
| Regular Insulin, Human 8 UNT | SQ | |
| miglitol 25 MG Oral Tablet [Glyset] | orally | |
| insulin human, isophane 70 UNT/ML / Regular Insulin, Human 30 UNT/ML [Novolin] | Subq | |
| Metformin | sq | |
| sitagliptin | PO | |
| Glucotrol Pill | | |
| glimepiride 2 MG Oral Tablet [Amaryl] | Po | |
| dapagliflozin 10 MG / Metformin hydrochloride 1000 MG [Xigduo] | | |
| Linagliptin 5 MG Oral Tablet | orally | |
| Ultralente Insulin, Human 100 UNT/ML Injectable Suspension [Humulin U Ultralente] | SQ | |
| insulin pork, isophane 80 UNT/ML Injectable Suspension | subcutaneously | |
| Metformin Pill | PO | |
| canagliflozin Oral Tablet [Invokana] | by mouth | |
| repaglinide Oral Product | oral | |
| Insulin, Aspart, Human 100 UNT/ML Injectable Solution | SUBCUTANEOUS | |
| Metformin hydrochloride 100 MG/ML Oral Solution [Riomet] | PO | |
| Diabeta Oral Product | oral | |
| insulin human, isophane 90 UNT/ML | subq | |
| Avandamet Oral Product | p.o. | |
| empagliflozin 5 MG | | |
| glimepiride 4 MG Oral Tablet [Amaryl] | BY MOUTH | |
| empagliflozin Oral Tablet | by mouth | |
| nateglinide Oral Product | oral | |
| 3 ML Insulin Glargine 100 UNT/ML Pen Injector [Lantus] | drip | |
| rosiglitazone 1 MG Oral Tablet | | |

| | | |
|---|---|---|
| nateglinide Oral Tablet | by mouth | |
| Insulin Glargine | topically | |
| Metformin hydrochloride 1000 MG Oral Tablet | ORAL | |
| sitagliptin Oral Tablet | po | |
| Insulin Lispro | SubQ | |
| insulin human, isophane 36 UNT/ML | Subcut | |
| Bromocriptine Oral Capsule [Parlodel] | by mouth | |
| 3 ML insulin human, isophane 70 UNT/ML / Regular Insulin, Human 30 UNT/ML Pen Injector [Humulin] | subQ | |
| Glyburide 1.25 MG Oral Tablet [Micronase] | PO | |
| Glynase Oral Product | orally | |
| Insulin Lispro | bedside. | |
| 24 HR Metformin hydrochloride 500 MG Extended Release Oral Tablet [Glucophage] | orally | |
| Insulin Lispro 25 UNT/ML | SUB Q | |
| Insulin Lispro 100 UNT/ML Injectable Solution | SC | |
| 1.5 ML insulin human, isophane 70 UNT/ML / Regular Insulin, Human 30 UNT/ML Prefilled Syringe | subcut. | |
| Metformin | sl | |
| Bromocriptine 2.5 MG | per tube | |
| Glyburide 3 MG Oral Tablet [Glynase] | p.o. | |
| exenatide 0.01 MG/ACTUAT | subcutaneous | |
| Insulin Glargine | p.o. | |
| Insulin, Aspart, Human | nostril | |
| insulin human, isophane 34 UNT/ML | SQ | |
| Insulin Lispro 50 UNT/ML / Insulin, Protamine Lispro, Human 50 UNT/ML Injectable Suspension [Humalog Mix] | subcutaneously | |
| Glyburide 1.25 MG Oral Tablet | Oral | |
| 3 ML Insulin Glargine 100 UNT/ML Pen Injector [Lantus] | sub q | |
| 0.5 ML albiglutide 100 MG/ML Pen Injector [Tanzeum] | subcutaneous | |
| exenatide 0.005 MG/ACTUAT [Byetta] | subcu. | |
| saxagliptin 5 MG Oral Tablet [Onglyza] | | |
| 0.5 ML dulaglutide 3 MG/ML Auto-Injector [Trulicity] | | 0 |
| liraglutide | subcutaneous | |
| Insulin, Zinc, Human 100 UNT/ML Injectable Suspension [Novolin L] | sub q | |
| 3 ML insulin detemir 100 UNT/ML Pen Injector | subcutaneously | |
| Regular Insulin, Human 500 UNT/ML Injectable Solution [Humulin R] | intramuscular | |

| | |
|---|---|
| Regular Insulin, Human 500 UNT/ML Injectable Solution [Humulin R] | Subcutaneous |
| Regular Insulin, Human 6 UNT/ML | subcutaneous |
| Regular Insulin, Human 500 UNT/ML Injectable Solution [Humulin R] | sc |
| Insulin Glargine 300 UNT/ML | orally |
| Insulin, Aspart, Human 100 UNT/ML Cartridge [NovoLog] | subq. |
| nateglinide 60 MG Oral Tablet | Oral |
| Regular Insulin, Human 15 UNT/ML | iv |
| Glyburide / Metformin Oral Tablet | by mouth |
| repaglinide | PO |
| insulin pork, isophane 100 UNT/ML Injectable Suspension [NPH Iletin II] | GT |
| insulin human, isophane 100 UNT/ML Pen Injector [Humulin N] | SQ |
| Insulin Glargine Injectable Solution | subcut. |
| Insulin Glargine | subq. |
| 24 HR Metformin hydrochloride 1000 MG / sitagliptin 100 MG Extended Release Oral Tablet [Janumet] | po |
| Metformin hydrochloride 1000 MG [Glucophage] | intravenously |
| Metformin Oral Solution | Po |
| Duetact Oral Product | BY MOUTH |
| Metformin Oral Tablet [Glucophage] | intravenously |
| Tolazamide | |
| repaglinide 0.5 MG Oral Tablet | ORAL |
| Insulin Glargine 100 UNT/ML | by mouth |
| empagliflozin 5 MG / Metformin hydrochloride 500 MG Oral Tablet [Synjardy] | by mouth |
| Glipizide 10 MG Extended Release Oral Tablet [Glucotrol] | |
| 3 ML Insulin Lispro 200 UNT/ML Pen Injector | orally |
| Simvastatin 20 MG / sitagliptin 100 MG Oral Tablet | PO |
| Insulin, Aspart, Human 100 UNT/ML Injectable Solution [NovoLog] | PER INSULIN PUMP |
| Linagliptin 2.5 MG / Metformin hydrochloride 850 MG [Jentadueto] | |
| Insulin Lispro 25 UNT/ML | subq. |
| Glipizide 2.5 MG | SQ |
| Humulin R Injectable Product | subq |
| Pramlintide | SQ |
| insulin human, isophane 60 UNT/ML / Regular Insulin, Human 40 UNT/ML [Humulin] | |

| | | |
|---|---|---|
| alogliptin 25 MG Oral Tablet [Nesina] | PO | |
| Regular Insulin, Human 30 UNT/ML | subcutaneous | |
| Avandia Oral Product | Oral | |
| insulin human, isophane 70 UNT/ML | SUBCUT | |
| Metformin hydrochloride 500 MG / rosiglitazone 1 MG Oral Tablet [Avandamet] | po | |
| saxagliptin 5 MG Oral Tablet | orally | |
| insulin pork, isophane 100 UNT/ML Injectable Suspension [NPH Iletin II] | npo. | |
| 0.5 ML dulaglutide 1.5 MG/ML Auto-Injector [Trulicity] | sub q | |
| Metformin hydrochloride 500 MG Extended Release Oral Tablet [Glumetza] | orally | |
| Regular Insulin, Human 8 UNT Inhalant Powder [Afrezza] | | |
| Glyburide 1.5 MG Oral Tablet | p.o. | |
| Glipizide 5 MG / Metformin hydrochloride 500 MG Oral Tablet [Metaglip] | | 0 |
| Insulin Glargine 100 UNT/ML [Lantus] | oral | |
| glimepiride 2 MG / rosiglitazone 4 MG Oral Tablet [Avandaryl] | ORAL | |
| Acarbose 50 MG Oral Tablet [Precose] | p.o. | |
| Insulin, Aspart, Human | p.o. | |
| Regular Insulin, Human | topical | |
| insulin degludec Injectable Product | subcutaneously | |
| 3 ML insulin human, isophane 100 UNT/ML Prefilled Syringe [Novolin N] | SUBCUTANEOUS | |
| Insulin, Aspart, Human | | |
| Metformin hydrochloride 750 MG Extended Release Oral Tablet [Glucophage] | p.o. | |
| insulin detemir 100 UNT/ML [Levemir] | sub Q | |
| Metformin hydrochloride 750 MG Extended Release Oral Tablet [Glucophage] | oral | |
| Metformin hydrochloride 850 MG / pioglitazone 15 MG Oral Tablet | PO. | |
| insulin detemir | SQ | |
| 0.5 ML dulaglutide 1.5 MG/ML Auto-Injector [Trulicity] | by mouth | |
| Insulin Lispro 100 UNT/ML Injectable Solution | pump | |
| Insulin Lispro 25 UNT/ML / Insulin, Protamine Lispro, Human 75 UNT/ML Injectable Suspension [Humalog Mix] | by mouth | |
| 60 ACTUAT exenatide 0.005 MG/ACTUAT Pen Injector [Byetta] | PO | |
| Humulin U Ultralente Injectable Product | | |

| | | |
|---|---|---|
| Glipizide 2.5 MG / Metformin hydrochloride 250 MG Oral Tablet [Metaglip] | oral | |
| Metformin hydrochloride 100 MG/ML | Neb | |
| Insulin, Aspart, Human | subc | |
| Regular Insulin, Human 25 UNT/ML | IV | |
| Regular Insulin, Human 8 UNT | subQ | |
| pioglitazone Oral Tablet | | |
| Insulin, Aspart, Human | Bedside | |
| Regular Insulin, Human 40 UNT/ML | IV | |
| Insulin Lispro 25 UNT/ML / Insulin, Protamine Lispro, Human 75 UNT/ML [Humalog Mix] | by mouth | |
| Regular Insulin, Human 500 UNT/ML Injectable Solution [Humulin R] | | 0 |
| rosiglitazone 8 MG Oral Tablet [Avandia] | Orally | |
| Glyburide 5 MG / Metformin 1000 MG Oral Tablet | oral | |
| Insulin, Glulisine, Human 100 UNT/ML Injectable Solution [Apidra] | subcutaneous | |
| Glipizide 10 MG Oral Tablet | ORAL | |
| Metformin hydrochloride 500 MG / rosiglitazone 2 MG Oral Tablet [Avandamet] | p.o. | |
| sitagliptin 100 MG | IM | |
| empagliflozin | | |
| insulin human, isophane 40 UNT/ML | SubQ | |
| glimepiride | intramuscular | |
| Glipizide 5 MG / Metformin hydrochloride 500 MG Oral Tablet | oral | |
| Regular Insulin, Human 100 UNT/ML Injectable Solution | SQ | |
| Victoza Injectable Product | IM injection | |
| Lispro PRC Injectable Product | sq | |
| Metformin hydrochloride 750 MG | nostril | |
| Glipizide 10 MG Oral Tablet | inhalation | |
| insulin human, isophane 70 UNT/ML / Regular Insulin, Human 30 UNT/ML Injectable Suspension [Novolin] | subcut | |
| Insulin Glargine 100 UNT/ML [Lantus] | topical | |
| Regular Insulin, Human 30 UNT/ML | iv | |
| Glyburide 5 MG / Metformin 1000 MG Oral Tablet | orally | |
| insulin pork, isophane 100 UNT/ML Injectable Suspension | subcut | |
| Invokana Pill | | |
| Metformin 500 MG | ou | |
| Glyburide | nostril | |
| Metformin hydrochloride 500 MG / sitagliptin 50 MG Oral Tablet | po | |

| | | |
|---|---|---|
| insulin human, isophane 75 UNT/ML | SC | |
| insulin pork, isophane 70 UNT/ML / Insulin, Regular, Pork 30 UNT/ML Injectable Suspension | subcu | |
| 0.5 ML dulaglutide 3 MG/ML Auto-Injector [Trulicity] | subQ | |
| Jardiance Oral Product | orally | |
| Insulin, Aspart, Human 100 UNT/ML Cartridge [NovoLog] | subc. | |
| dapagliflozin Oral Tablet [Farxiga] | by mouth | |
| Glyxambi Oral Product | po | |
| Insulin, Aspart Protamine, Human 70 UNT/ML / Insulin, Aspart, Human 30 UNT/ML Injectable Suspension | sq | |
| Glyburide 2.5 MG / Metformin hydrochloride 500 MG Oral Tablet | PO | |
| Glyburide | bedside | |
| 3 ML insulin human, isophane 70 UNT/ML / Regular Insulin, Human 30 UNT/ML Pen Injector [Humulin] | subcutaneously | |
| miglitol 25 MG Oral Tablet | by mouth | |
| Glyburide 1.25 MG Oral Tablet | Orally | |
| Insulin, Aspart, Human | sub Q | |
| exenatide 0.005 MG/ACTUAT [Byetta] | subc. | |
| insulin human, isophane 100 UNT/ML Injectable Suspension [Humulin N] | G - Tube | |
| Glyburide / Metformin Oral Tablet [Glucovance] | po | |
| Metformin hydrochloride 1000 MG [Glucophage] | subcutaneously | |
| Insulin Lispro 200 UNT/ML [Humalog] | SQ | |
| Levemir Injectable Product | subcut | |
| insulin human, isophane 100 UNT/ML Injectable Suspension [Humulin N] | | 0 |
| Regular Insulin, Human 8 UNT | iv | |
| Regular Insulin, Human 6 UNT/ML | iv | |
| Acarbose Oral Tablet | orally | |
| insulin human, isophane 70 UNT/ML / Regular Insulin, Human 30 UNT/ML [Humulin] | orally | |
| Glyburide 5 MG | per tube | |
| insulin human, isophane 70 UNT/ML / Regular Insulin, Human 30 UNT/ML [Novolin] | SUBQ | |
| insulin human, isophane 30 UNT/ML / Regular Insulin, Human 10 UNT/ML Injectable Suspension | | |
| rosiglitazone 4 MG [Avandia] | NPO | |
| NovoLog Injectable Product | SUBCUT | |
| insulin human, isophane 70 UNT/ML / Regular Insulin, Human 30 UNT/ML [Novolin] | Subcut | |

| | | |
|---|---|---|
| glimepiride | Sublingual | |
| Glumetza Oral Product | oral | |
| Glyburide 2.5 MG Oral Tablet [Diabeta] | | |
| Metformin hydrochloride 1000 MG / sitagliptin 100 MG Extended Release Oral Tablet [Janumet] | | |
| nateglinide 60 MG Oral Tablet [Starlix] | | 0 |
| insulin human, isophane 100 UNT/ML Injectable Suspension [Humulin N] | sub q | |
| Insulin, Regular, Pork 40 UNT/ML Injectable Solution | subcutaneous | |
| Glyburide Oral Tablet [Micronase] | po | |
| Insulin Glargine | | 0 |
| Insulin Glargine 100 UNT/ML [Lantus] | SUB Q | |
| 3 ML insulin degludec 200 UNT/ML Pen Injector [Tresiba] | injection | |
| 24 HR Metformin hydrochloride 750 MG Extended Release Oral Tablet [Glucophage] | by mouth | |
| Regular Insulin, Human 100 UNT/ML Injectable Solution [Humulin R] | sc | |
| glimepiride 4 MG [Amaryl] | per tube | |
| insulin human, isophane 70 UNT/ML | sub q | |
| alogliptin 12.5 MG / pioglitazone 30 MG Oral Tablet [Oseni] | by mouth | |
| insulin human, isophane 70 UNT/ML / Regular Insulin, Human 30 UNT/ML Injectable Suspension [Humulin] | sub q | |
| Regular Insulin, Human 100 UNT/ML Injectable Solution [Humulin R] | sub q | |
| Humulin R Injectable Product | subcut | |
| Regular Insulin, Human 24 UNT/ML | | |
| Insulin, Zinc, Pork 100 UNT/ML Injectable Suspension | | |
| Glyburide 2.5 MG / Metformin 400 MG Oral Tablet | p.o. | |
| 3 ML liraglutide 6 MG/ML Pen Injector [Victoza] | orally | |
| 24 HR Metformin hydrochloride 500 MG / sitagliptin 50 MG Extended Release Oral Tablet [Janumet] | by mouth | |
| Insulin, Zinc, Pork | sq | |
| insulin human, isophane 16 UNT/ML | sub Q | |
| Insulin, Aspart, Human | TPN | |
| Metformin hydrochloride 1000 MG Extended Release Oral Tablet [Glumetza] | PO | |
| Metformin hydrochloride 1000 MG / sitagliptin 50 MG [Janumet] | per tube | |
| insulin human, isophane 70 UNT/ML | PO | |
| pramlintide acetate 0.6 MG/ML Injectable Solution | | |
| Byetta Injectable Product | subcutaneous | |

| | |
|---|---|
| 24 HR empagliflozin 12.5 MG / Metformin hydrochloride 1000 MG Extended Release Oral Tablet [Synjardy] | |
| Metformin / rosiglitazone Oral Tablet [Avandamet] | |
| insulin human, isophane 50 UNT/ML / Regular Insulin, Human 50 UNT/ML Injectable Suspension [Humulin] | |
| glimepiride 2 MG / pioglitazone 30 MG Oral Tablet | PO |
| Lantus Injectable Product | sub q |
| Glipizide | npo |
| Linagliptin 5 MG Oral Tablet [Tradjenta] | Oral |
| Bromocriptine 5 MG | per tube |
| insulin human, isophane 70 UNT/ML / Regular Insulin, Human 30 UNT/ML [Humulin] | topical |
| 60 ACTUAT exenatide 0.01 MG/ACTUAT Pen Injector [Byetta] | Subcutaneous |
| nateglinide 120 MG Oral Tablet [Starlix] | Po |
| Insulin Lispro 100 UNT/ML Cartridge [Humalog] | subcutaneous |
| Bromocriptine 5 MG Oral Capsule | PO |
| Humulin Injectable Product | subcut |
| Insulin, Aspart Protamine, Human 70 UNT/ML / Insulin, Aspart, Human 30 UNT/ML Injectable Suspension [NovoLog Mix] | Sub Q |
| Chlorpropamide 200 MG | |
| Metformin Oral Tablet [Glucophage] | p.o. |
| insulin human, isophane 60 UNT/ML | SUBCUTANEOUS |
| Regular Insulin, Human | Sub Q |
| insulin pork, isophane 100 UNT/ML Injectable Suspension [NPH Iletin II] | SUB Q |
| repaglinide 0.5 MG Oral Tablet | |
| nateglinide 60 MG | per mouth |
| Insulin Lispro 25 UNT/ML | npo |
| Prandin Pill | |
| insulin human, isophane 70 UNT/ML / Regular Insulin, Human 30 UNT/ML [Humulin] | subcu. |
| glimepiride | OU |
| Regular Insulin, Human 15 UNT/ML | SUBCUT |
| Insulin Lispro 50 UNT/ML / Insulin, Protamine Lispro, Human 50 UNT/ML Injectable Suspension [Humalog Mix] | gt |
| Metformin hydrochloride 500 MG / sitagliptin 50 MG [Janumet] | subcutaneously |
| Metformin 500 MG | IV |
| Glyburide | IV |
| Acarbose 100 MG Oral Tablet [Precose] | oral |
| exenatide 0.01 MG/ACTUAT [Byetta] | sub q. |

| | | |
|---|---|---|
| Regular Insulin, Human | SEE COMMENT | |
| repaglinide 0.5 MG Oral Tablet | po | |
| Regular Insulin, Human 100 UNT/ML Injectable Solution [Humulin S] | npo | |
| Insulin, Glulisine, Human 100 UNT/ML Injectable Solution | subcutaneous | |
| Metformin hydrochloride 500 MG / sitagliptin 50 MG Oral Tablet | by mouth | |
| insulin human, isophane 70 UNT/ML / Regular Insulin, Human 30 UNT/ML [Humulin] | SUBQ | |
| insulin pork, isophane 80 UNT/ML | sub q | |
| nateglinide 60 MG Oral Tablet | p.o. | |
| Regular Insulin, Human 100 UNT/ML Injectable Solution [Humulin R] | SQ. | |
| insulin human, isophane 32 UNT/ML | SubQ | |
| exenatide 2 MG Injection [Bydureon] | | |
| Metformin | subcu | |
| 24 HR Glipizide 5 MG Extended Release Oral Tablet [Glucotrol] | oral | |
| Glyburide 1.25 MG Oral Tablet [Diabeta] | p.o. | |
| Metformin hydrochloride 1000 MG / sitagliptin 50 MG Oral Tablet [Janumet] | Oral | |
| Insulin Glargine | gt | |
| insulin pork, isophane 40 UNT/ML Injectable Suspension | | |
| Insulin Glargine Injectable Solution | Subcut | |
| insulin human, isophane 70 UNT/ML / Regular Insulin, Human 30 UNT/ML [Novolin] | SUB Q | |
| glimepiride | po | |
| Insulin Glargine 300 UNT/ML | by mouth | |
| insulin human, isophane 60 UNT/ML | sc | |
| Insulin Glargine 100 UNT/ML Injectable Solution | sub q | |
| Riomet Oral Product | po | |
| Metformin 500 MG | IM injection | |
| Insulin Lispro | | 0 |
| Insulin, Glulisine, Human | subcutaneously | |
| dapagliflozin 5 MG / Metformin hydrochloride 1000 MG [Xigduo] | | |
| Glyburide | npo | |
| NovoLog Injectable Product | IM injection | |
| Glipizide 2.5 MG / Metformin hydrochloride 500 MG Oral Tablet [Metaglip] | | |
| 60 ACTUAT exenatide 0.005 MG/ACTUAT Pen Injector [Byetta] | | 0 |
| repaglinide 2 MG Oral Tablet | | |
| 3 ML Insulin Glargine 100 UNT/ML Pen Injector [Lantus] | npo | |

| | | |
|---|---|---|
| Regular Insulin, Human 100 UNT/ML Injectable Solution [Novolin R] | Subcutaneous | |
| Insulin Glargine 100 UNT/ML Injectable Solution | oral | |
| Chlorpropamide 250 MG Oral Tablet [Diabinese] | po | |
| insulin human, isophane 50 UNT/ML / Regular Insulin, Human 50 UNT/ML [Humulin] | subq. | |
| Avandia Pill | | |
| 3 ML insulin human, isophane 70 UNT/ML / Regular Insulin, Human 30 UNT/ML Pen Injector [Humulin] | | 0 |
| insulin human, isophane 70 UNT/ML / Regular Insulin, Human 30 UNT/ML Injectable Suspension | subcutaneously | |
| Lantus Injectable Product | MISCELLANEOUS | |
| 3 ML Regular Insulin, Human 500 UNT/ML Pen Injector [Humulin R] | | |
| Glyburide 1.25 MG Oral Tablet | | |
| Victoza Injectable Product | by mouth | |
| 24 HR Metformin hydrochloride 500 MG / sitagliptin 50 MG Extended Release Oral Tablet [Janumet] | | |
| Insulin Lispro 50 UNT/ML | po | |
| Onglyza Oral Product | oral | |
| Glyburide 1.25 MG Oral Tablet [Micronase] | by mouth | |
| 60 ACTUAT exenatide 0.01 MG/ACTUAT Pen Injector [Byetta] | subcut. | |
| exenatide 0.01 MG/ACTUAT [Byetta] | Subq | |
| Insulin Lispro 25 UNT/ML | iv | |
| repaglinide 2 MG Oral Tablet | ORAL | |
| 3 ML Insulin, Aspart, Human 100 UNT/ML Pen Injector [NovoLog] | SQ | |
| 60 ACTUAT exenatide 0.01 MG/ACTUAT Pen Injector [Byetta] | | |
| Insulin, Aspart Protamine, Human 70 UNT/ML / Insulin, Aspart, Human 30 UNT/ML Injectable Suspension | SUBCUT | |
| canagliflozin 50 MG / Metformin hydrochloride 500 MG [Invokamet] | | |
| Glyburide | subcutaneously | |
| 3 ML insulin human, isophane 70 UNT/ML / Regular Insulin, Human 30 UNT/ML Pen Injector [Humulin] | by mouth | |
| Insulin Glargine 300 UNT/ML | po | |
| canagliflozin 300 MG Oral Tablet [Invokana] | p.o. | |
| Metformin Oral Tablet | p.o. | |
| glimepiride 2 MG [Amaryl] | per tube | |
| 3 ML Insulin, Aspart, Human 100 UNT/ML Cartridge [NovoLog] | PO | |
| Levemir Injectable Product | subq | |

| | | |
|---|---|---|
| dapagliflozin 5 MG Oral Tablet [Farxiga] | PO | |
| Metformin hydrochloride 500 MG / rosiglitazone 1 MG Oral Tablet [Avandamet] | PO | |
| 3 ML Insulin Glargine 100 UNT/ML / Lixisenatide 0.033 MG/ML Pen Injector [Soliqua] | subcutaneous | |
| Insulin Lispro 25 UNT/ML / Insulin, Protamine Lispro, Human 75 UNT/ML [Humalog Mix] | subcut. | |
| Jentadueto Oral Product | orally | |
| Simvastatin 10 MG / sitagliptin 100 MG [Juvisync] | subcutaneous | |
| rosiglitazone Oral Tablet [Avandia] | | |
| pramlintide acetate 0.6 MG/ML Injectable Solution [Symlin] | SQ | |
| insulin, regular, beef-pork 40 UNT/ML Inhalant Solution | subc. | |
| 3 ML Insulin Glargine 100 UNT/ML Pen Injector | subcutaneous | |
| insulin human, isophane 70 UNT/ML / Regular Insulin, Human 30 UNT/ML [Novolin] | sq. | |
| Oseni Oral Product | oral | |
| rosiglitazone 2 MG Oral Tablet | by mouth | |
| Insulin Lispro 50 UNT/ML | subQ | |
| Insulin Glargine | SUB Q | |
| Insulin Lispro 25 UNT/ML / Insulin, Protamine Lispro, Human 75 UNT/ML Injectable Suspension [Humalog Mix] | SUBCUTANEOUSLY | |
| insulin human, isophane 100 UNT/ML Injectable Suspension [Humulin N] | subcut. | |
| Regular Insulin, Human | SQ. | |
| Insulin Lispro 25 UNT/ML | Subcutaneous | |
| Lixisenatide | | |
| Regular Insulin, Human 40 UNT/ML | subq | |
| 24 HR Metformin hydrochloride 750 MG Extended Release Oral Tablet [Glucophage] | po | |
| Metformin hydrochloride 1000 MG Oral Tablet | | 0 |
| Precose Oral Product | PO | |
| insulin human, isophane 100 UNT/ML Injectable Suspension [Novolin N] | Subcut | |
| albiglutide 60 MG/ML [Tanzeum] | subq | |
| glimepiride Oral Tablet | orally | |
| saxagliptin 5 MG Oral Tablet [Onglyza] | | 0 |
| Insulin Lispro | nostril | |
| Insulin Lispro Cartridge [Humalog] | p.o. | |
| 3 ML Regular Insulin, Human 100 UNT/ML Prefilled Syringe [Novolin R] | SUBCUTANEOUS | |

| | | |
|---|---|---|
| sitagliptin 100 MG [Januvia] | subcutaneous | |
| Glyburide 2.5 MG / Metformin 400 MG Oral Tablet | orally | |
| Bromocriptine 2.5 MG Oral Tablet [Parlodel] | p.o. | |
| exenatide 0.01 MG/ACTUAT | sc | |
| alogliptin 25 MG | | |
| Glipizide 2.5 MG Oral Tablet | BY MOUTH | |
| Acarbose 100 MG Oral Tablet | p.o. | |
| Farxiga Oral Product | orally | |
| Glucotrol Pill | by mouth | |
| insulin human, isophane 32 UNT/ML | Subcut. | |
| sitagliptin Oral Tablet [Januvia] | orally | |
| Byetta Injectable Product | COMBO ROUTE) | |
| Glipizide 5 MG | OU | |
| Simvastatin 20 MG / sitagliptin 50 MG Oral Tablet | p.o. | |
| exenatide 2 MG [Bydureon] | sub Q | |
| dulaglutide | by mouth | |
| Invokana Pill | by mouth | |
| Acarbose | G - tube | |
| insulin human, isophane 100 UNT/ML Injectable Suspension [Humulin N] | Subcutaneously | |
| Insulin Lispro 100 UNT/ML [Humalog] | sq | |
| Insulin Lispro 50 UNT/ML / Insulin, Protamine Lispro, Human 50 UNT/ML Injectable Suspension [Humalog Mix] | SUBCUT | |
| Insulin Glargine 100 UNT/ML Injectable Solution | subcut | |
| 3 ML Insulin Glargine 100 UNT/ML Pen Injector [Lantus] | sc | |
| Metformin hydrochloride 500 MG Extended Release Oral Tablet [Fortamet] | oral | |
| dulaglutide 1.5 MG/ML [Trulicity] | sub q | |
| Metformin Pill | swallow | |
| Metformin hydrochloride 1000 MG / sitagliptin 50 MG Oral Tablet | p.o. | |
| Glipizide Oral Tablet [Glucotrol] | orally | |
| Regular Insulin, Human 30 UNT/ML | subcutaneously | |
| Glyburide | SQ | |
| Glyburide | gt | |
| glimepiride 6 MG Oral Tablet | | |
| repaglinide 1 MG Oral Tablet | ORAL | |
| Avandamet Pill | | |
| 60 ACTUAT exenatide 0.005 MG/ACTUAT Pen Injector [Byetta] | SubQ | |
| Lantus Injectable Product | | 0 |

| | | |
|---|---|---|
| insulin detemir 100 UNT/ML Injectable Solution [Levemir] | Subcutaneously | |
| Insulin, Glulisine, Human 100 UNT/ML Injectable Solution [Apidra] | by mouth | |
| Insulin, Aspart, Human 100 UNT/ML Cartridge [NovoLog] | | 0 |
| Metformin | Bedside | |
| Linagliptin 2.5 MG / Metformin hydrochloride 1000 MG Oral Tablet [Jentadueto] | ORAL | |
| Insulin Glargine Injectable Solution | Subq | |
| Metformin | SL | |
| insulin detemir Injectable Product | subcutaneous | |
| Insulin, Zinc, Human 100 UNT/ML Injectable Suspension [Novolin L] | subcutaneously | |
| 0.5 ML albiglutide 60 MG/ML Pen Injector [Tanzeum] | subcutaneous | |
| 3 ML insulin human, isophane 70 UNT/ML / Regular Insulin, Human 30 UNT/ML Pen Injector [Humulin] | subcut | |
| insulin human, isophane 50 UNT/ML / Regular Insulin, Human 50 UNT/ML Injectable Suspension [Humulin] | SUBCUT | |
| Insulin Lispro 100 UNT/ML | SUBCUT | |
| Insulin Glargine Injectable Solution | iv | |
| insulin human, isophane 40 UNT/ML | im | |
| insulin human, isophane 40 UNT/ML | subc. | |
| Insulin, Aspart, Human 100 UNT/ML Injectable Solution [NovoLog] | subq | |
| Insulin Lispro 100 UNT/ML Injectable Solution [Humalog] | IV | |
| Insulin, Glulisine, Human | subcut | |
| Insulin Glargine | Sub q | |
| Glyburide / Metformin Oral Tablet [Glucovance] | BY MOUTH | |
| insulin human, isophane 100 UNT/ML Injectable Suspension [Novolin N] | by mouth | |
| canagliflozin 150 MG / Metformin hydrochloride 1000 MG Oral Tablet [Invokamet] | orally | |
| insulin, regular, beef-pork 40 UNT/ML Inhalant Solution | sc | |
| 3 ML Insulin, Aspart Protamine, Human 70 UNT/ML / Insulin, Aspart, Human 30 UNT/ML Pen Injector [NovoLog Mix] | gt | |
| Glipizide 2.5 MG / Metformin hydrochloride 500 MG Oral Tablet | PO | |
| 3 ML insulin degludec 100 UNT/ML / liraglutide 3.6 MG/ML Pen Injector [Xultophy] | subcutaneous | |
| Glyburide 1 MG Oral Tablet | Oral | |
| empagliflozin 10 MG / Linagliptin 5 MG Oral Tablet [Glyxambi] | | |
| Metformin 500 MG | sl | |

| | | |
|---|---|---|
| glimepiride 6 MG Oral Tablet | orally | |
| Insulin, Aspart, Human 100 UNT/ML Injectable Solution | subq | |
| pioglitazone 45 MG Oral Tablet [Actos] | Po | |
| insulin pork, isophane 70 UNT/ML / Insulin, Regular, Pork 30 UNT/ML [Pork Mixtard 30] | | |
| liraglutide 6 MG/ML | SQ | |
| pioglitazone Oral Tablet [Actos] | orally | |
| insulin human, isophane 36 UNT/ML | sub q | |
| Regular Insulin, Human 10 UNT/ML | Subcutaneous | |
| Jentadueto Oral Product | po | |
| 1.5 ML insulin human, isophane 70 UNT/ML / Regular Insulin, Human 30 UNT/ML Prefilled Syringe | subcut | |
| 3 ML liraglutide 6 MG/ML Pen Injector [Victoza] | Subcutaneously | |
| Metformin hydrochloride 500 MG / saxagliptin 5 MG [Kombiglyze] | po | |
| Lente Insulin, Beef-Pork 40 UNT/ML | SQ | |
| 60 ACTUAT exenatide 0.005 MG/ACTUAT Pen Injector [Byetta] | Subcutaneously | |
| Metformin Extended Release Oral Tablet [Glucophage] | po | |
| Glucovance Oral Product | Po | |
| Insulin, Aspart Protamine, Human 70 UNT/ML / Insulin, Aspart, Human 30 UNT/ML Injectable Suspension [NovoLog Mix] | | 0 |
| Metformin Oral Tablet | swallow | |
| canagliflozin 50 MG / Metformin hydrochloride 1000 MG Oral Tablet | by mouth | |
| Insulin Glargine 100 UNT/ML Injectable Solution [Lantus] | | |
| Parlodel Oral Product | PO | |
| glimepiride 4 MG / rosiglitazone 8 MG Oral Tablet [Avandaryl] | orally | |
| insulin human, isophane 70 UNT/ML / Regular Insulin, Human 30 UNT/ML [Humulin] | Sq | |
| 3 ML insulin human, isophane 100 UNT/ML Pen Injector [Humulin N] | SQ | |
| Insulin, Aspart Protamine, Human 70 UNT/ML / Insulin, Aspart, Human 30 UNT/ML Injectable Suspension [NovoLog Mix] | sc | |
| insulin detemir | PO | |
| Metformin hydrochloride 850 MG / pioglitazone 15 MG Oral Tablet [Actoplus Met] | | 0 |
| Glyburide 5 MG / Metformin hydrochloride 500 MG Oral Tablet [Glucovance] | oral | |
| Glipizide 2.5 MG Oral Tablet | ORAL | |
| exenatide 0.005 MG/ACTUAT | subq | |

| | | |
|---|---|---|
| glimepiride 2 MG / pioglitazone 30 MG Oral Tablet [Duetact] | ORAL | |
| Bromocriptine Oral Capsule | | |
| exenatide 2 MG Injection [Bydureon] | | 0 |
| Insulin, Aspart, Human 100 UNT/ML Cartridge [NovoLog] | TD | |
| Metformin | sc | |
| Metformin hydrochloride 1000 MG Oral Tablet | subcutaneously | |
| Metformin | oral | |
| insulin human, isophane 60 UNT/ML | subcutaneously | |
| Glipizide 10 MG | ng | |
| Insulin Lispro 25 UNT/ML / Insulin, Protamine Lispro, Human 75 UNT/ML Injectable Suspension [Humalog Mix] | SUBQ | |
| Insulin Lispro 25 UNT/ML | PO | |
| pioglitazone 30 MG Oral Tablet [Actos] | Po | |
| insulin human, isophane 16 UNT/ML | Subcut | |
| Metformin hydrochloride 500 MG / rosiglitazone 4 MG Oral Tablet [Avandamet] | ORAL | |
| dulaglutide 1.5 MG/ML | SQ | |
| sitagliptin 100 MG Oral Tablet [Januvia] | p.o | |
| insulin detemir | sub q | |
| Glipizide Extended Release Oral Tablet | oral | |
| insulin pork, isophane 100 UNT/ML Injectable Suspension [NPH Iletin II] | swallow | |
| Metformin hydrochloride 500 MG / repaglinide 1 MG Oral Tablet [PrandiMet] | po | |
| Insulin Lispro Cartridge [Humalog] | orally | |
| pioglitazone 30 MG Oral Tablet | Oral | |
| 1.5 ML Insulin Glargine 300 UNT/ML Pen Injector [Toujeo] | | 0 |
| insulin human, isophane 40 UNT/ML | SUBCUT | |
| 60 ACTUAT exenatide 0.01 MG/ACTUAT Pen Injector [Byetta] | SubQ | |
| Insulin Lispro 200 UNT/ML Pen Injector [Humalog] | | |
| Insulin Glargine Injectable Solution | orally | |
| glimepiride 4 MG [Amaryl] | subcutaneously | |
| Insulin, Aspart, Human | Oral | |
| Insulin, Aspart Protamine, Human 70 UNT/ML / Insulin, Aspart, Human 30 UNT/ML Injectable Suspension [NovoLog Mix] | po | |
| Glipizide 2.5 MG / Metformin hydrochloride 250 MG Oral Tablet [Metaglip] | ORAL | |
| Tresiba Injectable Product | | |
| Insulin Glargine Injectable Solution | oral | |
| sitagliptin 25 MG Oral Tablet [Januvia] | | 0 |

| | | |
|---|---|---|
| Glyburide | subcutaneous | |
| Insulin, Aspart, Human 100 UNT/ML Injectable Solution | subcut | |
| Actoplus Met Oral Product | PO | |
| Acarbose 50 MG Oral Tablet [Precose] | orally | |
| insulin human, isophane 100 UNT/ML Injectable Suspension | subcutaneously | |
| Metformin 500 MG | SQ | |
| canagliflozin 50 MG / Metformin hydrochloride 1000 MG Oral Tablet [Invokamet] | po | |
| Juvisync Oral Product | by mouth | |
| Simvastatin 20 MG / sitagliptin 100 MG Oral Tablet | | |
| Metformin | inh | |
| sitagliptin Oral Tablet [Januvia] | po | |
| Insulin, Aspart, Human | SUBCUTANEOUS | |
| Linagliptin 5 MG Oral Tablet [Tradjenta] | BY MOUTH | |
| Glyburide 4.5 MG Oral Tablet | po | |
| Insulin, Aspart Protamine, Human 70 UNT/ML / Insulin, Aspart, Human 30 UNT/ML Injectable Suspension [NovoLog Mix] | Subcutaneously | |
| Apidra Injectable Product | Subcutaneously | |
| Kombiglyze Oral Product | by mouth | |
| Metformin 500 MG | sublingual | |
| Ultralente Insulin, Human 100 UNT/ML Injectable Suspension | SC | |
| Glyburide 2.5 MG / Metformin hydrochloride 500 MG Oral Tablet [Glucovance] | | 0 |
| Metformin 400 MG | orally | |
| Glipizide Oral Product | Orally | |
| insulin human, isophane 60 UNT/ML | subcu. | |
| saxagliptin Oral Tablet [Onglyza] | by mouth | |
| 0.5 ML albiglutide 100 MG/ML Pen Injector [Tanzeum] | sub q | |
| Glyburide 5 MG Oral Tablet | P.O. | |
| Insulin Lispro Cartridge [Humalog] | gt | |
| Insulin Lispro 100 UNT/ML [Humalog] | SubQ | |
| repaglinide 2 MG Oral Tablet | oral | |
| exenatide 0.01 MG/ACTUAT | Subcutaneous | |
| liraglutide 6 MG/ML [Victoza] | Sub Q | |
| insulin human, isophane 70 UNT/ML / Regular Insulin, Human 30 UNT/ML [Novolin] | NPO. | |
| insulin pork, isophane 50 UNT/ML | subcut | |
| insulin detemir 100 UNT/ML Injectable Solution [Levemir] | subc | |
| empagliflozin 25 MG / Linagliptin 5 MG Oral Tablet [Glyxambi] | orally | |
| Insulin Lispro 50 UNT/ML | Subcut | |

| | | |
|---|---|---|
| sitagliptin 25 MG Oral Tablet [Januvia] | p.o. | |
| 3 ML Insulin, Aspart Protamine, Human 70 UNT/ML / Insulin, Aspart, Human 30 UNT/ML Pen Injector [NovoLog Mix] | NPO | |
| Chlorpropamide 100 MG Oral Tablet [Diabinese] | ORAL | |
| Glipizide 10 MG Extended Release Oral Tablet [Glucotrol] | po | |
| 0.5 ML albiglutide 100 MG/ML Pen Injector [Tanzeum] | SQ | |
| pioglitazone 15 MG Oral Tablet [Actos] | PER TUBE | |
| 3 ML Insulin, Aspart, Human 100 UNT/ML Pen Injector [NovoLog] | injection | |
| Insulin Lispro 100 UNT/ML Injectable Solution [Lispro PRC] | orally | |
| Insulin Lispro 50 UNT/ML | subcu. | |
| Regular Insulin, Human Pen Injector [Humulin R] | subQ | |
| Tradjenta Pill | | |
| Insulin Lispro 25 UNT/ML / Insulin, Protamine Lispro, Human 75 UNT/ML Injectable Suspension [Humalog Mix] | SC | |
| Metformin hydrochloride 1000 MG / sitagliptin 100 MG [Janumet] | orally | |
| nateglinide Oral Product | po | |
| insulin human, isophane 40 UNT/ML | Subcutaneous | |
| Insulin, Zinc, Human 100 UNT/ML Injectable Suspension | Subcutaneous | |
| empagliflozin 10 MG / Linagliptin 5 MG Oral Tablet [Glyxambi] | po | |
| Regular Insulin, Human 10 UNT/ML | subQ | |
| canagliflozin 50 MG / Metformin hydrochloride 500 MG Oral Tablet [Invokamet] | by mouth | |
| Novolin L Injectable Product | subcutaneously | |
| empagliflozin 25 MG Oral Tablet | by mouth | |
| Glyburide 1 MG Oral Tablet | | |
| Glyburide 1.25 MG Oral Tablet | | 0 |
| 0.5 ML dulaglutide 1.5 MG/ML Auto-Injector [Trulicity] | | 0 |
| insulin human, isophane 28 UNT/ML | subQ | |
| Insulin Lispro 100 UNT/ML Injectable Solution | subcutaneously | |
| 24 HR Glipizide 2.5 MG Extended Release Oral Tablet | | |
| 3 ML insulin detemir 100 UNT/ML Pen Injector [Levemir] | injection | |
| Diabeta Pill | by mouth | |
| Prandin Oral Product | oral | |
| 0.5 ML dulaglutide 1.5 MG/ML Auto-Injector | | |
| Humalog Injectable Product | subq | |
| insulin detemir | orally | |
| exenatide Injection | | |
| Glyburide 6 MG Oral Tablet [Glynase] | orally | |
| Starlix Oral Product | BY MOUTH | |

| | | |
|---|---|---|
| Glipizide 10 MG Oral Tablet | nostril | |
| 24 HR Metformin hydrochloride 750 MG Extended Release Oral Tablet | Orally | |
| Bromocriptine 5 MG Oral Capsule [Parlodel] | oral | |
| Insulin Glargine Injectable Solution | po | |
| Glipizide | inhalation | |
| insulin human, isophane 90 UNT/ML / Regular Insulin, Human 10 UNT/ML [Humulin] | subq | |
| Lantus Injectable Product | IM injection | |
| Metformin | sub q | |
| repaglinide 1 MG Oral Tablet [Prandin] | oral | |
| Insulin Glargine 100 UNT/ML [Lantus] | drip | |
| Simvastatin 20 MG / sitagliptin 100 MG Oral Tablet | oral | |
| insulin degludec 200 UNT/ML | sub q | |
| 3 ML Insulin, Glulisine, Human 100 UNT/ML Pen Injector [Apidra] | subcut | |
| Chlorpropamide 125 MG | | |
| insulin detemir | subc | |
| Insulin, Aspart, Human 100 UNT/ML Cartridge [NovoLog] | po | |
| insulin detemir | sc | |
| Glyburide | by mouth | |
| Metformin 500 MG | topically | |
| Glipizide 2.5 MG Oral Tablet | Po | |
| Metformin hydrochloride 1000 MG / rosiglitazone 4 MG Oral Tablet [Avandamet] | orally | |
| Insulin, Aspart Protamine, Human 70 UNT/ML / Insulin, Aspart, Human 30 UNT/ML Injectable Suspension | | |
| 0.65 ML exenatide 3.08 MG/ML Pen Injector [Bydureon] | | 0 |
| glimepiride Pill | po | |
| glimepiride 8 MG Oral Tablet | | |
| glimepiride 1 MG / rosiglitazone 4 MG Oral Tablet [Avandaryl] | by mouth | |
| Regular Insulin, Human 500 UNT/ML Injectable Solution [Humulin R] | subQ | |
| Regular Insulin, Human 50 UNT/ML | iv | |
| Metformin hydrochloride 1000 MG [Glucophage] | per tube | |
| rosiglitazone Oral Product | orally | |
| Metformin hydrochloride 500 MG Oral Tablet | P.O. | |
| Glipizide / Metformin Oral Tablet | orally | |
| insulin human, isophane 60 UNT/ML | Sub Q | |
| Metformin hydrochloride 850 MG Extended Release Oral Tablet | | |
| Regular Insulin, Human 100 UNT/ML Injectable Solution | SC | |

| | |
|---|---|
| insulin human, isophane 90 UNT/ML | SUBCUTANEOUS |
| Insulin, Glulisine, Human 100 UNT/ML Injectable Solution [Apidra] | sub q |
| Insulin, Aspart Protamine, Human 70 UNT/ML / Insulin, Aspart, Human 30 UNT/ML Injectable Suspension [NovoLog Mix] | NPO |
| Regular Insulin, Human 25 UNT/ML | subcutaneously |
| Regular Insulin, Human 100 UNT/ML Injectable Solution [Humulin R] | Subcutaneous |
| 0.5 ML dulaglutide 3 MG/ML Auto-Injector [Trulicity] | SubQ |
| Metformin hydrochloride 1000 MG / sitagliptin 50 MG Extended Release Oral Tablet [Janumet] | by mouth |
| Glipizide 10 MG Oral Tablet [Glucotrol] | P.O. |
| Metformin 1000 MG | PO. |
| Insulin Lispro Injectable Solution | subcutaneous |
| insulin human, isophane 32 UNT/ML | subcu |
| Regular Insulin, Human 100 UNT/ML Injectable Solution [Novolin R] | npo |
| Regular Insulin, Human 100 UNT/ML Injectable Solution | orally |
| Afrezza Inhalant Product | inhaled |
| nateglinide | Bedside |
| Acarbose 100 MG Oral Tablet | PO |
| Humalog Mix Injectable Product | subq |
| Metformin | npo. |
| 24 HR Glipizide 5 MG Extended Release Oral Tablet | Oral |
| insulin human, isophane 16 UNT/ML | SUBCUTANEOUS |
| Linagliptin 2.5 MG / Metformin hydrochloride 1000 MG Oral Tablet [Jentadueto] | PO |
| Bromocriptine Oral Capsule | by mouth |
| Insulin Lispro 100 UNT/ML | INSUPUMP |
| empagliflozin 25 MG Oral Tablet [Jardiance] | |
| repaglinide 2 MG Oral Tablet | po |
| Acarbose 25 MG Oral Tablet | |
| insulin human, isophane 85 UNT/ML / Regular Insulin, Human 15 UNT/ML [Pur-In Mix] | |
| insulin human, isophane 50 UNT/ML / Regular Insulin, Human 50 UNT/ML [Humulin] | SUB Q |
| 0.5 ML dulaglutide 1.5 MG/ML Auto-Injector [Trulicity] | injection |
| canagliflozin 100 MG Oral Tablet [Invokana] | p.o. |
| Fortamet Oral Product | oral |
| Regular Insulin, Human 10 UNT/ML | SQ |

| | |
|---|---|
| Metformin hydrochloride 100 MG/ML Oral Solution | Po |
| Regular Insulin, Human 15 UNT/ML | Subcutaneous |
| Insulin Glargine Injectable Solution | SUBCUTANEOUS |
| Metformin hydrochloride 1000 MG [Glumetza] | by mouth |
| insulin human, isophane 70 UNT/ML / Regular Insulin, Human 30 UNT/ML [Novolin] | orally |
| insulin human, isophane 100 UNT/ML Injectable Suspension [Novolin N] | Subc. |
| Insulin, Glulisine, Human 100 UNT/ML Injectable Solution [Apidra] | SC |
| Metformin 500 MG | gt |
| dapagliflozin 10 MG Oral Tablet [Farxiga] | p.o. |
| Riomet Oral Product | oral |
| Nesina Oral Product | by mouth |
| Insulin Lispro Injectable Solution [Humalog] | SUBCUTANEOUSLY |
| Glyburide | SEE COMMENT |
| Bromocriptine | orally |
| insulin human, isophane 16 UNT/ML | SUBCUT |
| pioglitazone 15 MG Oral Tablet [Actos] | Po |
| exenatide | by mouth |
| pioglitazone 45 MG Oral Tablet [Actos] | Oral |
| insulin human, isophane 100 UNT/ML Injectable Suspension [Humulin N] | by mouth |
| insulin pork, isophane 100 UNT/ML Injectable Suspension [NPH Iletin II] | Sq |
| 24 HR Glipizide 10 MG Extended Release Oral Tablet [Glucotrol] | PO |
| repaglinide 1 MG Oral Tablet | po |
| insulin human, isophane 50 UNT/ML / Regular Insulin, Human 50 UNT/ML Injectable Suspension | SUBCUTANEOUS |
| Metformin 500 MG | topical |
| Humulin L Injectable Product | subcutaneously |
| 3 ML insulin human, isophane 100 UNT/ML Pen Injector | |
| insulin human, isophane 34 UNT/ML | SubQ |
| Lispro PRC Injectable Product | SQ |
| Insulin Glargine 100 UNT/ML [Lantus] | subcutaneously. |
| Glyburide 5 MG Oral Tablet [Euglucon] | po |
| rosiglitazone Oral Product | PO |
| Glucophage Oral Product | |
| Tolbutamide 250 MG | |
| 3 ML Insulin Lispro 200 UNT/ML Pen Injector | subcut |

| | |
|---|---|
| Byetta Injectable Product | subq |
| 3 ML insulin human, isophane 100 UNT/ML Pen Injector [Humulin N] | SUBQ |
| Acarbose 25 MG Oral Tablet [Precose] | g-tube |
| Insulin, Aspart, Human | MISC |
| Metformin | by mouth. |
| Metformin hydrochloride 500 MG Oral Tablet | by mouth. |
| Regular Insulin, Human 6 UNT/ML | subQ |
| Bromocriptine 5 MG Oral Capsule [Parlodel] | orally |
| exenatide 2 MG | subcutaneous |
| pioglitazone Oral Tablet [Actos] | oral |
| Regular Insulin, Human 100 UNT/ML Injectable Solution | IV |
| Regular Insulin, Human | PO |
| nateglinide 30 MG Oral Tablet | po |
| Glipizide 10 MG | NPO |
| Insulin Glargine Injectable Solution [Lantus] | by mouth |
| 3 ML insulin human, isophane 100 UNT/ML Prefilled Syringe | sq |
| Regular Insulin, Human 100 UNT/ML Injectable Solution | subcut |
| Insulin Lispro | INSUPUMP |
| nateglinide Pill | |
| 3 ML Insulin Lispro 100 UNT/ML Cartridge [Humalog] | subq |
| Glyburide 1.25 MG Oral Tablet | BY MOUTH |
| insulin human, isophane 70 UNT/ML / Regular Insulin, Human 30 UNT/ML [Novolin] | NPO |
| 3 ML Insulin Lispro 200 UNT/ML Pen Injector | sq |
| Regular Insulin, Human 100 UNT/ML Injectable Solution [Humulin S] | sq |
| Avandamet Oral Product | oral |
| insulin pork, isophane 80 UNT/ML | SUBQ |
| insulin human, isophane 100 UNT/ML Injectable Suspension [Novolin N] | Sub Q |
| Metformin hydrochloride 500 MG Extended Release Oral Tablet [Glumetza] | oral |
| Regular Insulin, Human | MISC |
| miglitol 25 MG Oral Tablet [Glyset] | p.o. |
| Metformin hydrochloride 500 MG [Glumetza] | by mouth |
| Glyburide | PO. |
| 3 ML Insulin, Aspart Protamine, Human 70 UNT/ML / Insulin, Aspart, Human 30 UNT/ML Pen Injector [NovoLog Mix] | subQ |
| insulin human, isophane 100 UNT/ML Pen Injector [Humulin N] | Sq |

| | | |
|---|---|---|
| Insulin, Aspart Protamine, Human 70 UNT/ML / Insulin, Aspart, Human 30 UNT/ML [NovoLog Mix] | subcutaneous. | |
| glimepiride 8 MG Oral Tablet | p.o. | |
| Glyset Oral Product | PO | |
| Metformin hydrochloride 1000 MG / saxagliptin 5 MG Extended Release Oral Tablet [Kombiglyze] | by mouth | |
| Insulin, Aspart Protamine, Human 70 UNT/ML / Insulin, Aspart, Human 30 UNT/ML Injectable Suspension | npo | |
| insulin human, isophane 70 UNT/ML / Regular Insulin, Human 30 UNT/ML Injectable Suspension | SUBCUTANEOUS | |
| Insulin, Aspart, Human | bedside. | |
| Regular Insulin, Human 6 UNT/ML | IV | |
| Metformin 500 MG | OU | |
| 3 ML Insulin, Glulisine, Human 100 UNT/ML Pen Injector [Apidra] | | 0 |
| exenatide 0.005 MG/ACTUAT | subcutaneous | |
| Regular Insulin, Human 100 UNT/ML Injectable Solution [Humulin R] | SUBCUTANEOUSLY | |
| Glipizide | NPO. | |
| Tolbutamide | | |
| insulin human, isophane 40 UNT/ML | subcu. | |
| Trulicity Injectable Product | IM injection | |
| insulin human, isophane 50 UNT/ML / Regular Insulin, Human 50 UNT/ML Injectable Suspension [Humulin] | subcutaneous | |
| 3 ML insulin human, isophane 100 UNT/ML Prefilled Syringe | NPO | |
| Tolbutamide 500 MG [Orinase] | | |
| Metformin | TOPICAL | |
| 60 ACTUAT exenatide 0.005 MG/ACTUAT Pen Injector [Byetta] | p.o. | |
| 0.5 ML albiglutide 100 MG/ML Pen Injector | | |
| Humalog Mix Injectable Product | | |
| Insulin, Aspart, Human | td | |
| Metformin | iv | |
| Insulin, Glulisine, Human 100 UNT/ML Injectable Solution [Apidra] | Subcutaneously | |
| Insulin Glargine Injectable Solution | Sub Q | |
| Regular Insulin, Human 100 UNT/ML Injectable Solution [Novolin R] | intravenous | |
| Actoplus Met Pill | by mouth | |
| Regular Insulin, Human 5 UNT/ML | INFUSION | |
| Linagliptin 5 MG Oral Tablet | p.o. | |
| Regular Insulin, Human 100 UNT/ML Injectable Solution | drip | |

| | |
|---|---|
| Insulin Lispro 100 UNT/ML Injectable Solution | subQ |
| miglitol 50 MG Oral Tablet [Glyset] | ORAL |
| liraglutide | orally |
| 3 ML liraglutide 6 MG/ML Pen Injector | subcut |
| Metformin | inhaled |
| Insulin Lispro 25 UNT/ML / Insulin, Protamine Lispro, Human 75 UNT/ML [Humalog Mix] | subq. |
| glimepiride 4 MG [Amaryl] | gt |
| Insulin, Zinc, Human | p.o. |
| rosiglitazone 8 MG Oral Tablet [Avandia] | Po |
| 24 HR Metformin hydrochloride 500 MG / saxagliptin 5 MG Extended Release Oral Tablet [Kombiglyze] | oral |
| Insulin Lispro 25 UNT/ML / Insulin, Protamine Lispro, Human 75 UNT/ML Injectable Suspension [Humalog Mix] | po |
| Insulin, Aspart Protamine, Human 70 UNT/ML / Insulin, Aspart, Human 30 UNT/ML Injectable Suspension [NovoLog Mix] | gt |
| insulin human, isophane 70 UNT/ML | Subcutaneously |
| Regular Insulin, Human 100 UNT/ML Injectable Solution [Velosulin] | subcutaneous |
| insulin beef, isophane 40 UNT/ML | SQ |
| Metformin Oral Liquid Product | by mouth |
| insulin human, isophane 70 UNT/ML / Regular Insulin, Human 30 UNT/ML [Humulin] | MISCELLANEOUS |
| 3 ML Insulin Glargine 100 UNT/ML Pen Injector [Lantus] | Subcutaneous |
| canagliflozin 300 MG Oral Tablet [Invokana] | Oral |
| canagliflozin 100 MG Oral Tablet [Invokana] | BY MOUTH |
| alogliptin 12.5 MG / Metformin hydrochloride 500 MG [Kazano] | |
| Metformin hydrochloride 500 MG / sitagliptin 50 MG [Janumet] | per tube |
| insulin human, isophane 70 UNT/ML / Regular Insulin, Human 30 UNT/ML [Novolin] | p.o. |
| insulin human, isophane 60 UNT/ML | subcu |
| Tolbutamide 500 MG Oral Tablet | po |
| Metformin hydrochloride 750 MG [Glucophage] | by mouth |
| Insulin Lispro 50 UNT/ML / Insulin, Protamine Lispro, Human 50 UNT/ML Injectable Suspension [Humalog Mix] | subQ |
| insulin detemir 100 UNT/ML [Levemir] | SC |
| 60 ACTUAT exenatide 0.005 MG/ACTUAT Pen Injector [Byetta] | SC |
| Regular Insulin, Human | gt |
| insulin human, isophane 28 UNT/ML | SUBQ |
| Metformin | topically |

| | | |
|---|---|---|
| Lantus Injectable Product | subcu | |
| exenatide | oral | |
| Regular Insulin, Human 5 UNT/ML | Subcutaneous | |
| 3 ML liraglutide 6 MG/ML Pen Injector [Saxenda] | by mouth | |
| 3 ML Insulin Glargine 100 UNT/ML Pen Injector [Lantus] | subcu | |
| Metformin 500 MG | drip | |
| Regular Insulin, Human 100 UNT/ML Injectable Solution [Humulin R] | IM | |
| Insulin, Aspart, Human 100 UNT/ML Injectable Solution | by mouth | |
| Glipizide 5 MG Extended Release Oral Tablet [Glucotrol] | by mouth | |
| Metformin hydrochloride 1000 MG [Glucophage] | gt | |
| miglitol 50 MG Oral Tablet | orally | |
| insulin human, isophane 85 UNT/ML | subq | |
| 3 ML insulin detemir 100 UNT/ML Pen Injector [Levemir] | subc. | |
| Metformin Extended Release Oral Tablet | by mouth. | |
| Insulin, Aspart, Human 100 UNT/ML Cartridge [NovoLog] | oral | |
| Insulin Lispro 25 UNT/ML / Insulin, Protamine Lispro, Human 75 UNT/ML Injectable Suspension [Humalog Mix] | | 0 |
| Glipizide / Metformin Oral Tablet | p.o. | |
| Metformin 1000 MG | IM | |
| Metformin hydrochloride 500 MG [Fortamet] | by mouth | |
| Glyburide 4.5 MG Oral Tablet | by mouth | |
| Insulin Lispro 100 UNT/ML | by mouth | |
| Metformin hydrochloride 750 MG | by mouth. | |
| Glyburide 1.25 MG | by mouth. | |
| exenatide 2 MG Injection [Bydureon] | Subcutaneously | |
| Insulin Glargine | SUB CONJ | |
| 24 HR dapagliflozin 10 MG / Metformin hydrochloride 500 MG Extended Release Oral Tablet [Xigduo] | by mouth | |
| 3 ML Regular Insulin, Human 100 UNT/ML Prefilled Syringe | | |
| Metformin hydrochloride 500 MG Extended Release Oral Tablet [Glumetza] | p.o. | |
| repaglinide 2 MG Oral Tablet [Prandin] | Oral | |
| insulin human, isophane 34 UNT/ML | subcu | |
| 3 ML Insulin, Aspart Protamine, Human 70 UNT/ML / Insulin, Aspart, Human 30 UNT/ML Pen Injector [NovoLog Mix] | Subcutaneously | |
| Acarbose 100 MG Oral Tablet | oral | |
| exenatide 0.01 MG/ACTUAT Pen Injector | sc | |
| Bromocriptine Oral Capsule | PO | |

| | |
|---|---|
| canagliflozin 50 MG / Metformin hydrochloride 500 MG Oral Tablet [Invokamet] | ORAL |
| Insulin Glargine | inhalatn |
| miglitol 25 MG Oral Tablet [Glyset] | ORAL |
| Insulin, Aspart, Human 100 UNT/ML Injectable Solution | subQ |
| nateglinide 60 MG Oral Tablet | |
| Humulin R Injectable Product | subcutaneous |
| Glipizide | swallow |
| Glipizide 10 MG | by mouth. |
| 3 ML Insulin Glargine 100 UNT/ML Pen Injector [Basaglar] | injection |
| Metformin | orally. |
| glimepiride 8 MG | NPO |
| liraglutide 6 MG/ML [Victoza] | orally |
| glimepiride | SC |
| glimepiride 4 MG | NPO |
| Tanzeum Injectable Product | |
| Levemir Injectable Product | SUBCUTANEOUSLY |
| glimepiride 2 MG | inhaled |
| insulin pork, isophane 100 UNT/ML Injectable Suspension [NPH Iletin II] | subcutaneously. |
| Glyburide 1 MG Oral Tablet | |
| Insulin Lispro Cartridge [Humalog] | Sub Q |
| Insulin Lispro Cartridge [Humalog] | subcutaneous. |
| Metformin hydrochloride 1000 MG / rosiglitazone 4 MG Oral Tablet [Avandamet] | oral |
| Metformin hydrochloride 500 MG Oral Tablet | g-tube |
| Glumetza Pill | |
| Insulin, Zinc, Human | subcutaneous |
| Insulin Lispro 100 UNT/ML Injectable Solution | subq |
| Insulin Glargine | subcutaneously. |
| Metformin Pill | oral |
| Linagliptin 2.5 MG / Metformin hydrochloride 850 MG Oral Tablet [Jentadueto] | oral |
| empagliflozin 10 MG Oral Tablet [Jardiance] | |
| Glyburide 5 MG / Phenformin 50 MG Oral Tablet | p.o. |
| alogliptin 25 MG Oral Tablet [Nesina] | oral |
| Regular Insulin, Human | PERIDIAL |
| Regular Insulin, Human 30 UNT/ML | sq |
| insulin human, isophane 100 UNT/ML Injectable Suspension [Humulin N] | SUBQ |

| | | |
|---|---|---|
| Insulin Lispro | topically | |
| Glyburide | NPO. | |
| Metformin hydrochloride 500 MG Oral Tablet [Glucophage] | By mouth | |
| glimepiride 4 MG / rosiglitazone 8 MG [Avandaryl] | | |
| Glyburide | po. | |
| 3 ML liraglutide 6 MG/ML Pen Injector [Victoza] | sub Q | |
| Insulin Lispro Cartridge [Humalog] | po | |
| glimepiride 4 MG / rosiglitazone 4 MG Oral Tablet [Avandaryl] | PO | |
| 0.5 ML dulaglutide 1.5 MG/ML Auto-Injector [Trulicity] | sq | |
| repaglinide 1 MG Oral Tablet [Prandin] | orally | |
| alogliptin 25 MG / pioglitazone 15 MG Oral Tablet | by mouth | |
| Insulin Glargine 100 UNT/ML [Lantus] | | 0 |
| Glyburide 5 MG / Metformin hydrochloride 500 MG Oral Tablet [Glucovance] | BY MOUTH | |
| glimepiride 4 MG / pioglitazone 30 MG Oral Tablet | po | |
| Metformin 1000 MG | subcut | |
| Metformin | intravenous | |
| Insulin Lispro 50 UNT/ML | Sub Q | |
| Insulin Lispro 100 UNT/ML Injectable Solution [Lispro PRC] | by mouth | |
| Metformin | INH | |
| Insulin Lispro 25 UNT/ML | NPO | |
| Insulin, Aspart, Human 100 UNT/ML Injectable Solution [NovoLog] | SUBCUTANEOUS BY INSULIN PUMP | |
| Insulin Glargine Pen Injector [Lantus] | | |
| Prandin Oral Product | orally | |
| Metformin hydrochloride 500 MG / sitagliptin 50 MG Oral Tablet [Janumet] | | 0 |
| 3 ML insulin degludec 100 UNT/ML Pen Injector [Tresiba] | | |
| Acarbose 100 MG Oral Tablet [Precose] | | |
| Insulin Lispro Cartridge [Humalog] | SUB Q | |
| insulin pork, isophane 100 UNT/ML | | |
| sitagliptin 100 MG [Januvia] | sc | |
| Metformin | per tube | |
| Insulin Lispro 50 UNT/ML / Insulin, Protamine Lispro, Human 50 UNT/ML Injectable Suspension [Humalog Mix] | SUBQ | |
| Insulin, Aspart Protamine, Human 70 UNT/ML / Insulin, Aspart, Human 30 UNT/ML [NovoLog Mix] | SUBCUT | |
| Insulin Glargine | | |
| Glipizide Extended Release Oral Tablet | p.o. | |
| Glipizide / Metformin Oral Tablet | Orally | |

| | |
|---|---|
| nateglinide Oral Product | |
| insulin pork, isophane 100 UNT/ML Injectable Suspension [NPH Iletin II] | SUBQ |
| Glucophage Oral Product | Oral |
| 3 ML Insulin Lispro 100 UNT/ML Cartridge [Humalog] | SubQ |
| Lantus Injectable Product | by mouth |
| empagliflozin 25 MG / Linagliptin 5 MG Oral Tablet [Glyxambi] | ORAL |
| Januvia Oral Product | |
| Metformin hydrochloride 100 MG/ML [Riomet] | g - tube |
| {60 (Regular Insulin, Human 4 UNT Inhalant Powder) / 30 (Regular Insulin, Human 8 UNT Inhalant Powder) } Pack | SQ |
| insulin human, isophane 75 UNT/ML | SUBCUT |
| Metformin hydrochloride 1000 MG / saxagliptin 2.5 MG [Kombiglyze] | orally |
| Insulin Glargine 100 UNT/ML Injectable Solution | sq |
| 0.65 ML exenatide 3.08 MG/ML Pen Injector [Bydureon] | injection |
| pioglitazone 15 MG [Actos] | PO. |
| insulin human, isophane 50 UNT/ML / Regular Insulin, Human 50 UNT/ML [Humulin] | subcutaneous |
| Metformin hydrochloride 500 MG / rosiglitazone 1 MG Oral Tablet [Avandamet] | |
| glimepiride | inhalation |
| insulin human, isophane 28 UNT/ML | SUBCUTANEOUS |
| insulin human, isophane 70 UNT/ML / Regular Insulin, Human 30 UNT/ML [Humulin] | IM |
| Metformin hydrochloride 1000 MG Oral Tablet | IV |
| Insulin Lispro 100 UNT/ML Injectable Solution | subcut |
| Acarbose 50 MG Oral Tablet | oral |
| Tolazamide 500 MG Oral Tablet | po |
| alogliptin 25 MG / pioglitazone 45 MG Oral Tablet [Oseni] | by mouth |
| dulaglutide 3 MG/ML [Trulicity] | subcut |
| exenatide 2 MG Injection | subcutaneously |
| Acarbose 50 MG Oral Tablet | p.o. |
| Insulin Glargine | SEE COMMENT |
| 3 ML liraglutide 6 MG/ML Pen Injector [Victoza] | SUBQ |
| Glyburide Oral Tablet [Micronase] | |
| insulin detemir | subcut. |
| insulin pork, isophane 70 UNT/ML / Insulin, Regular, Pork 30 UNT/ML Injectable Suspension | subQ |
| exenatide 0.005 MG/ACTUAT [Byetta] | subq. |

| | |
|---|---|
| 3 ML Insulin Glargine 100 UNT/ML Pen Injector [Lantus] | Subcut |
| 24 HR dapagliflozin 5 MG / Metformin hydrochloride 1000 MG Extended Release Oral Tablet [Xigduo] | |
| Insulin Lispro 50 UNT/ML / Insulin, Protamine Lispro, Human 50 UNT/ML Injectable Suspension [Humalog Mix] | subcu |
| Metformin hydrochloride 1000 MG [Glumetza] | orally |
| 24 HR Metformin hydrochloride 1000 MG / sitagliptin 100 MG Extended Release Oral Tablet [Janumet] | |
| 3 ML insulin human, isophane 100 UNT/ML Prefilled Syringe | subcutaneously |
| liraglutide 6 MG/ML [Victoza] | sub Q |
| insulin human, isophane 100 UNT/ML Injectable Suspension [Humulin N] | subcutaneously. |
| Glipizide 2.5 MG | subcutaneously |
| insulin human, isophane 34 UNT/ML | SUBCUT |
| Insulin Lispro Cartridge [Humalog] | IV |
| insulin human, isophane 90 UNT/ML / Regular Insulin, Human 10 UNT/ML [Humulin] | sub q |
| Regular Insulin, Human 25 UNT/ML | INFUSION |
| 0.5 ML albiglutide 60 MG/ML Pen Injector [Tanzeum] | po |
| Insulin, Aspart, Human | im |
| insulin human, isophane 85 UNT/ML | SQ |
| Regular Insulin, Human 10 UNT/ML | subcut. |
| insulin human, isophane 50 UNT/ML / Regular Insulin, Human 50 UNT/ML Injectable Suspension [Humulin] | subcut |
| 3 ML insulin human, isophane 100 UNT/ML Prefilled Syringe | MISCELLANEOUS |
| Insulin Lispro 50 UNT/ML / Insulin, Protamine Lispro, Human 50 UNT/ML Injectable Suspension [Humalog Mix] | Sub Q |
| Linagliptin | subcutaneous |
| Insulin Lispro 100 UNT/ML Injectable Solution [Humalog] | Subcutaneous |
| 0.5 ML albiglutide 60 MG/ML Pen Injector [Tanzeum] | SQ |
| Glyburide 1.5 MG Oral Tablet [Glynase] | ORAL |
| exenatide 0.005 MG/ACTUAT | Subq |
| Glyburide 1.25 MG / Metformin hydrochloride 250 MG Oral Tablet | BY MOUTH |
| Regular Insulin, Human 12 UNT | iv |
| glimepiride 4 MG | subcutaneously |
| Metformin 1000 MG | topical |
| insulin human, isophane 100 UNT/ML [Humulin N] | |
| nateglinide Oral Tablet [Starlix] | BY MOUTH |
| nateglinide 120 MG Oral Tablet | Oral |

| | |
|---|---|
| 3 ML insulin human, isophane 100 UNT/ML Prefilled Syringe | inhaled |
| Bromocriptine 1 MG Oral Tablet | po |
| Insulin, Aspart Protamine, Human 70 UNT/ML / Insulin, Aspart, Human 30 UNT/ML [NovoLog Mix] | sc. |
| Glipizide Pill | po |
| glimepiride 3 MG | subcutaneous |
| Simvastatin 20 MG / sitagliptin 100 MG Oral Tablet | p.o. |
| miglitol 100 MG Oral Tablet [Glyset] | PO |
| exenatide 0.005 MG/ACTUAT [Byetta] | Sq |
| Metformin hydrochloride 750 MG | subcutaneously |
| insulin human, isophane 70 UNT/ML / Regular Insulin, Human 30 UNT/ML [Humulin] | intravenously |
| Januvia Oral Product | Oral |
| 3 ML Insulin Glargine 100 UNT/ML Pen Injector [Lantus] | PO |
| alogliptin 12.5 MG / Metformin hydrochloride 1000 MG Oral Tablet [Kazano] | oral |
| Regular Insulin, Human 5 UNT/ML | SUBCUT |
| Regular Insulin, Human 15 UNT/ML | subcu |
| Metformin hydrochloride 500 MG [Glucophage] | nostril |
| Metformin hydrochloride 1000 MG Extended Release Oral Tablet [Fortamet] | orally |
| Glipizide 5 MG Extended Release Oral Tablet [Glucotrol] | |
| Insulin Lispro Cartridge [Humalog] | SUBCUTANEOUS |
| 1.5 ML pramlintide acetate 1 MG/ML Pen Injector [Symlin] | subq |
| 3 ML Insulin, Aspart Protamine, Human 70 UNT/ML / Insulin, Aspart, Human 30 UNT/ML Pen Injector [NovoLog Mix] | Sq |
| glimepiride | SQ |
| Insulin Glargine 100 UNT/ML Injectable Solution | by mouth |
| 3 ML Insulin Glargine 100 UNT/ML Pen Injector [Lantus] | po |
| 1.5 ML insulin human, isophane 100 UNT/ML Prefilled Syringe | IV |
| Metformin hydrochloride 500 MG / repaglinide 1 MG Oral Tablet [PrandiMet] | by mouth |
| Insulin Lispro | inhalatn |
| Metformin hydrochloride 500 MG Oral Tablet | ORAL |
| insulin pork, isophane 100 UNT/ML Injectable Suspension [NPH Iletin II] | subq. |
| Gliclazide 30 MG | po |
| Metforming Oral Product | PO |
| Micronase Oral Product | orally |
| glimepiride 2 MG / pioglitazone 30 MG Oral Tablet [Duetact] | p.o. |

| | |
|---|---|
| 3 ML insulin human, isophane 100 UNT/ML Pen Injector [Humulin N] | subcu |
| Glipizide 5 MG | npo |
| Insulin, Zinc, Human | subcut. |
| Janumet Oral Product | p.o. |
| exenatide 0.01 MG/ACTUAT [Byetta] | subcu. |
| Metformin 400 MG | PO |
| Insulin Lispro 50 UNT/ML / Insulin, Protamine Lispro, Human 50 UNT/ML Injectable Suspension [Humalog Mix] | Subcutaneous |
| Linagliptin 2.5 MG / Metformin hydrochloride 1000 MG Oral Tablet [Jentadueto] | po |
| insulin detemir 100 UNT/ML [Levemir] | Sub Q |
| Ultralente Insulin, Human Injectable Suspension [Humulin U Ultralente] | subcutaneously |
| empagliflozin 5 MG / Metformin hydrochloride 500 MG Oral Tablet [Synjardy] | |
| exenatide | Subq |
| Glipizide 2.5 MG / Metformin hydrochloride 500 MG Oral Tablet | orally |
| Insulin Glargine Injectable Solution | npo |
| insulin beef, isophane 40 UNT/ML | subcut. |
| Glyburide 4.5 MG Oral Tablet | |
| Glipizide / Metformin Pill | |
| Insulin Glargine 100 UNT/ML | iv |
| glimepiride 1 MG [Amaryl] | per tube |
| pioglitazone 15 MG | per tube |
| Metformin Oral Solution | ORAL |
| Metformin 1000 MG | subcutaneous |
| Regular Insulin, Human | INHALATION |
| Glyburide 3 MG | per tube |
| Glipizide 10 MG | drip |
| Metformin 1000 MG | NPO |
| glimepiride 2 MG [Amaryl] | nostril |
| Bromocriptine Oral Tablet | oral |
| Metformin hydrochloride 500 MG / pioglitazone 15 MG Oral Tablet | PO |
| Insulin Lispro | topical |
| insulin detemir 100 UNT/ML Injectable Solution [Levemir] | injection |
| canagliflozin 150 MG | |
| 24 HR Metformin hydrochloride 1000 MG / saxagliptin 5 MG Extended Release Oral Tablet [Kombiglyze] | by mouth |

| | |
|---|---|
| Regular Insulin, Human 100 UNT/ML Injectable Solution [Humulin R] | subc |
| Onglyza Oral Product | PO |
| Insulin Glargine 100 UNT/ML [Lantus] | NPO |
| 24 HR canagliflozin 150 MG / Metformin hydrochloride 1000 MG Extended Release Oral Tablet [Invokamet] | oral |
| pioglitazone | nostril |
| 3 ML Insulin, Aspart Protamine, Human 70 UNT/ML / Insulin, Aspart, Human 30 UNT/ML Pen Injector [NovoLog Mix] | SubQ |
| exenatide 2 MG [Bydureon] | Sc |
| Glipizide Oral Tablet [Glucotrol] | |
| Insulin Lispro 100 UNT/ML | SQ INFUSION |
| insulin pork, isophane 100 UNT/ML Injectable Suspension [NPH Iletin II] | g - tube |
| 3 ML Insulin, Aspart Protamine, Human 70 UNT/ML / Insulin, Aspart, Human 30 UNT/ML Pen Injector [NovoLog Mix] | SUBQ |
| 3 ML Insulin, Aspart Protamine, Human 70 UNT/ML / Insulin, Aspart, Human 30 UNT/ML Pen Injector [NovoLog Mix] | subcutaneous. |
| Metformin hydrochloride 1000 MG Extended Release Oral Tablet | by mouth. |
| repaglinide 2 MG [Prandin] | neb |
| Regular Insulin, Human 500 UNT/ML Injectable Solution | iv |
| Insulin Lispro 25 UNT/ML / Insulin, Protamine Lispro, Human 75 UNT/ML Injectable Suspension [Humalog Mix] | gt |
| Insulin Glargine | subcutaneous. |
| pramlintide acetate 0.6 MG/ML Injectable Solution [Symlin] | SUBCUTANEOUSLY |
| Glipizide 10 MG Oral Tablet | ORALLY |
| Glyburide Pill | po |
| nateglinide 30 MG | |
| Metformin Extended Release Oral Tablet | PO |
| Acarbose 100 MG Oral Tablet [Precose] | p.o. |
| exenatide 2 MG Injection [Bydureon] | by mouth |
| pioglitazone 15 MG Oral Tablet [Actos] | g tube |
| exenatide 0.005 MG/ACTUAT [Byetta] | SubCutaneous |
| 3 ML liraglutide 6 MG/ML Pen Injector [Saxenda] | injection |
| Insulin Lispro | Oral |
| insulin human, isophane 70 UNT/ML / Regular Insulin, Human 30 UNT/ML [Humulin] | drip |
| Insulin Lispro 100 UNT/ML Injectable Solution [Humalog] | INFUSION |
| Insulin Lispro | subcu. |
| dapagliflozin Oral Tablet [Farxiga] | PO |

| | |
|---|---|
| Glipizide 10 MG Extended Release Oral Tablet [Glucotrol] | PO |
| 24 HR dapagliflozin 10 MG / Metformin hydrochloride 500 MG Extended Release Oral Tablet [Xigduo] | |
| insulin pork, isophane 100 UNT/ML Injectable Suspension | subcutaneous |
| 3 ML insulin human, isophane 100 UNT/ML Prefilled Syringe [Novolin N] | Subq |
| Metformin hydrochloride 1000 MG Oral Tablet | IM |
| dapagliflozin 10 MG [Farxiga] | Sub Q |
| pioglitazone Oral Tablet | p.o. |
| 3 ML insulin degludec 100 UNT/ML Pen Injector [Tresiba] | injection |
| Insulin Lispro 100 UNT/ML | orally |
| nateglinide Oral Product | PO |
| insulin human, isophane 100 UNT/ML Injectable Suspension | SUBCUTANEOUS |
| insulin human, isophane 85 UNT/ML | sq |
| Bromocriptine | IM |
| insulin human, isophane 50 UNT/ML / Regular Insulin, Human 50 UNT/ML Injectable Suspension | subcutaneous |
| Glyburide 1.25 MG Oral Tablet [Micronase] | po |
| Insulin, Regular, Pork 40 UNT/ML Injectable Solution | subcu |
| Linagliptin Oral Product | po |
| Glyburide 3.5 MG | |
| insulin detemir 100 UNT/ML [Levemir] | orally |
| Regular Insulin, Human 25 UNT/ML | iv |
| Metformin 500 MG / ropinirole 2 MG Oral Tablet | oral |
| Insulin Glargine Injectable Solution [Lantus] | MISCELLANEOUS |
| insulin human, isophane 100 UNT/ML Injectable Suspension [Novolin N] | sub Q |
| Glipizide 5 MG | gt |
| Glipizide | SQ |
| Linagliptin Oral Tablet [Tradjenta] | by mouth |
| insulin pork, isophane 100 UNT/ML Injectable Suspension [NPH Iletin II] | nG |
| Insulin, Aspart Protamine, Human 70 UNT/ML / Insulin, Aspart, Human 30 UNT/ML Injectable Suspension [NovoLog Mix] | nostril |
| Insulin Lispro 25 UNT/ML / Insulin, Protamine Lispro, Human 75 UNT/ML Injectable Suspension [Humalog Mix] | SUBq |
| Insulin Lispro Cartridge [Humalog] | PO |
| pioglitazone 45 MG Oral Tablet | Oral |
| Insulin Glargine | subcu. |
| Humalog Mix Injectable Product | SUBCUT |

| | | |
|---|---|---|
| dulaglutide 1.5 MG/ML [Trulicity] | orally | |
| Acarbose 25 MG Oral Tablet [Precose] | p.o. | |
| insulin human, isophane 50 UNT/ML / Regular Insulin, Human 50 UNT/ML [Humulin] | sc | |
| insulin human, isophane 60 UNT/ML | subQ | |
| Insulin Lispro Cartridge [Humalog] | MISCELLANEOUS | |
| 3 ML Insulin Glargine 100 UNT/ML / Lixisenatide 0.033 MG/ML Pen Injector [Soliqua] | | |
| Onglyza Oral Product | p.o. | |
| Metformin hydrochloride 1000 MG Oral Tablet | P.O. | |
| exenatide 0.01 MG/ACTUAT [Byetta] | SUBQ | |
| Tolbutamide 500 MG Oral Tablet | PO | |
| Metformin 1000 MG | gt | |
| Regular Insulin, Human | p.o. | |
| insulin pork, isophane 80 UNT/ML | SUBCUTANEOUS | |
| Insulin, Aspart, Human | SUBCUTANEOUSLY | |
| Metformin hydrochloride 500 MG [Glucophage] | subcutaneously | |
| miglitol 50 MG Oral Tablet [Glyset] | oral | |
| exenatide 3.08 MG/ML [Bydureon] | SC | |
| Insulin, Regular, Pork 40 UNT/ML Injectable Solution | PO | |
| Trulicity Injectable Product | sub q | |
| Regular Insulin, Human 20 UNT/ML | per tube | |
| miglitol 25 MG Oral Tablet | orally | |
| Glyburide 5 MG Oral Tablet [Micronase] | Orally | |
| insulin detemir | SC | |
| Insulin Lispro | Subq | |
| Metformin 500 MG | | 0 |
| 60 ACTUAT exenatide 0.005 MG/ACTUAT Pen Injector [Byetta] | SUBQ | |
| Glipizide 5 MG Oral Tablet | P.O. | |
| dulaglutide 1.5 MG/ML [Trulicity] | Subcutaneously | |
| Metformin hydrochloride 500 MG Extended Release Oral Tablet | po. | |
| glimepiride | sublingual | |
| Acarbose 100 MG | SEE COMMENT | |
| Actos Oral Product | Oral | |
| dulaglutide 3 MG/ML | subcutaneously | |
| glimepiride 2 MG / rosiglitazone 4 MG Oral Tablet [Avandaryl] | p.o. | |
| Jardiance Oral Product | oral | |
| Glipizide 5 MG / Metformin hydrochloride 500 MG Oral Tablet | orally | |
| canagliflozin 150 MG / Metformin hydrochloride 1000 MG Oral Tablet [Invokamet] | | |

| | | |
|---|---|---|
| Glipizide | sublingual | |
| Metformin hydrochloride 850 MG [Metforming] | | |
| 0.5 ML dulaglutide 1.5 MG/ML Auto-Injector [Trulicity] | subq | |
| Regular Insulin, Human 12 UNT | subcutaneously | |
| rosiglitazone 4 MG Oral Tablet | ORAL | |
| Regular Insulin, Human 20 UNT/ML | subcutaneous | |
| Glipizide 5 MG | PO. | |
| 3 ML insulin human, isophane 100 UNT/ML Pen Injector [Humulin N] | | 0 |
| Glipizide / Metformin Oral Tablet | | |
| Insulin, Aspart, Human 100 UNT/ML Injectable Solution | Subcut | |
| insulin human, isophane 28 UNT/ML | sub Q | |
| Glyburide 2.5 MG Oral Tablet [Micronase] | | |
| Metformin hydrochloride 750 MG Extended Release Oral Tablet | by mouth. | |
| Chlorpropamide 250 MG Oral Tablet | p.o. | |
| Glipizide 10 MG | per mouth | |
| exenatide 0.005 MG/ACTUAT [Byetta] | Subcut. | |
| Regular Insulin, Human 6 UNT/ML | npo | |
| 0.5 ML dulaglutide 1.5 MG/ML Auto-Injector [Trulicity] | IM | |
| Regular Insulin, Human 15 UNT/ML | NPO | |
| 1.5 ML insulin human, isophane 70 UNT/ML / Regular Insulin, Human 30 UNT/ML Prefilled Syringe | sub q | |
| pioglitazone 30 MG Oral Tablet [Actos] | Orally | |
| rosiglitazone 4 MG Oral Tablet [Avandia] | | |
| Regular Insulin, Human 12 UNT | SQ | |
| Insulin, Aspart, Human 100 UNT/ML Injectable Solution [NovoLog] | by mouth | |
| insulin human, isophane 70 UNT/ML / Regular Insulin, Human 30 UNT/ML [Novolin] | im | |
| sitagliptin 25 MG Oral Tablet | po | |
| Humalog Injectable Product | by mouth | |
| insulin beef, isophane 40 UNT/ML | sc | |
| Glyburide 6 MG | per tube | |
| Glyburide | drip. | |
| Onglyza Oral Product | orally | |
| Regular Insulin, Human 100 UNT/ML Injectable Solution [Novolin R] | IM | |
| Metformin hydrochloride 500 MG Oral Tablet [Glucophage] | | 0 |
| Insulin, Aspart, Human | OU | |
| insulin human, isophane 28 UNT/ML | subcu | |

| | | |
|---|---|---|
| 0.5 ML dulaglutide 1.5 MG/ML Auto-Injector [Trulicity] | po | |
| miglitol 25 MG | | |
| insulin human, isophane 60 UNT/ML | sub Q | |
| insulin detemir 100 UNT/ML [Levemir] | subcut. | |
| 3 ML Insulin Glargine 100 UNT/ML Pen Injector [Basaglar] | | 0 |
| Kombiglyze Oral Product | po | |
| Metformin hydrochloride 500 MG Oral Tablet | By mouth | |
| sitagliptin Oral Tablet | PO | |
| Diabeta Oral Product | orally | |
| Glucotrol Oral Product | | |
| Glipizide 5 MG | SL | |
| Metformin 500 MG | sq | |
| 24 HR Glipizide 10 MG Extended Release Oral Tablet [Glucotrol] | oral | |
| Insulin Glargine 100 UNT/ML [Lantus] | p.o. | |
| Bromocriptine 5 MG Oral Capsule | oral | |
| Humalog Injectable Product | IM injection | |
| Bromocriptine 0.8 MG Oral Tablet | PO | |
| Insulin Glargine Injectable Solution | SC | |
| NovoLog Injectable Product | npo | |
| Glyburide Oral Tablet [Micronase] | orally | |
| Metformin | Topical | |
| 0.5 ML albiglutide 100 MG/ML Pen Injector [Tanzeum] | subq | |
| pramlintide acetate 0.6 MG/ML [Symlin] | sub q | |
| 24 HR canagliflozin 150 MG / Metformin hydrochloride 1000 MG Extended Release Oral Tablet [Invokamet] | ORAL | |
| Regular Insulin, Human 500 UNT/ML Injectable Solution [Humulin R] | SQ. | |
| Regular Insulin, Human 100 UNT/ML Injectable Solution [Novolin R] | SUBCUTANEOUSLY | |
| Byetta Injectable Product | MISCELLANEOUS | |
| Insulin Lispro 200 UNT/ML | | |
| Insulin, Aspart, Human 100 UNT/ML Cartridge [NovoLog] | GT | |
| Insulin, Zinc, Human 100 UNT/ML Injectable Suspension | SUBCUT | |
| Regular Insulin, Human 12 UNT | subq | |
| sitagliptin 25 MG Oral Tablet [Januvia] | By Mouth | |
| insulin human, isophane 50 UNT/ML / Regular Insulin, Human 50 UNT/ML Injectable Suspension [Humulin] | sq | |
| Metformin hydrochloride 500 MG / rosiglitazone 4 MG Oral Tablet [Avandamet] | orally | |
| Glyburide 1 MG Oral Tablet | | 0 |

| | |
|---|---|
| Glipizide 10 MG Oral Tablet [Glucotrol] | BY MOUTH |
| Acarbose Oral Tablet | |
| 0.5 ML albiglutide 100 MG/ML Pen Injector [Tanzeum] | sq |
| Insulin Glargine 100 UNT/ML [Lantus] | subcu. |
| Metformin hydrochloride 500 MG [Glucophage] | IV |
| Insulin, Aspart, Human 100 UNT/ML Injectable Solution | |
| Novolin N Injectable Product | SQ |
| canagliflozin 50 MG / Metformin hydrochloride 1000 MG Oral Tablet [Invokamet] | ORAL |
| Glyburide 1 MG Oral Tablet | ORAL |
| liraglutide | sq |
| Insulin, Aspart, Human | SUB Q |
| Regular Insulin, Human | nostril |
| 0.5 ML dulaglutide 1.5 MG/ML Auto-Injector [Trulicity] | subQ |
| saxagliptin 2.5 MG Oral Tablet | po |
| Metformin 250 MG Oral Tablet | Oral |
| Glipizide 5 MG [Glucotrol] | IM |
| Glipizide 5 MG | per tube |
| repaglinide Pill | |
| Bromocriptine 10 MG Oral Capsule | po |
| 3 ML Insulin Glargine 100 UNT/ML Pen Injector [Lantus] | NPO |
| Regular Insulin, Human | Subcut |
| Glipizide 5 MG Oral Tablet [Glucotrol] | BY MOUTH |
| Saxenda Injectable Product | SUBCUTANEOUS |
| Insulin, Zinc, Human 100 UNT/ML Injectable Suspension [Human Ultratard] | iv |
| glimepiride | topically |
| Glyburide 5 MG Oral Tablet [Diabeta] | Orally |
| Metformin hydrochloride 100 MG/ML Oral Solution | Oral |
| 3 ML insulin detemir 100 UNT/ML Pen Injector [Levemir] | nostril |
| Lixisenatide | SQ |
| Insulin Glargine Injectable Solution [Lantus] | subc. |
| saxagliptin 2.5 MG Oral Tablet [Onglyza] | PO |
| glimepiride | swallow |
| Linagliptin 5 MG | subcutaneously |
| glimepiride 4 MG [Amaryl] | subcut |
| exenatide 0.01 MG/ACTUAT [Byetta] | subc. |
| Metformin Extended Release Oral Tablet [Glucophage] | orally |
| Regular Insulin, Human 100 UNT/ML Injectable Solution [Humulin R] | by mouth. |

| | | |
|---|---|---|
| Insulin Glargine | drip. | |
| pioglitazone | subcutaneous | |
| Glyburide | po | |
| Regular Insulin, Human 500 UNT/ML Injectable Solution [Humulin R] | NPO | |
| Metformin hydrochloride 1000 MG / sitagliptin 50 MG Oral Tablet | oral | |
| Insulin, Aspart Protamine, Human 70 UNT/ML / Insulin, Aspart, Human 30 UNT/ML [NovoLog Mix] | subq. | |
| Bromocriptine 2.5 MG Oral Tablet [Parlodel] | | 0 |
| pioglitazone | subcut | |
| insulin human, isophane 16 UNT/ML | Subcutaneous | |
| Glyburide 1.25 MG / Metformin hydrochloride 250 MG Oral Tablet [Glucovance] | p.o. | |
| exenatide 0.01 MG/ACTUAT | subcutaneously | |
| insulin pork, isophane 100 UNT/ML Injectable Suspension | SQ | |
| 3 ML insulin detemir 100 UNT/ML Pen Injector [Levemir] | Subcu | |
| 1.5 ML insulin human, isophane 70 UNT/ML / Regular Insulin, Human 30 UNT/ML Prefilled Syringe | SQ | |
| Insulin Lispro 25 UNT/ML | subcu. | |
| Chlorpropamide Oral Tablet | oral | |
| dapagliflozin 5 MG [Farxiga] | ng | |
| Bromocriptine 5 MG Oral Capsule [Parlodel] | | |
| 3 ML insulin detemir 100 UNT/ML Pen Injector [Levemir] | ou | |
| Glipizide 5 MG [Glucotrol] | per mouth | |
| 0.65 ML exenatide 3.08 MG/ML Pen Injector [Bydureon] | | |
| glimepiride | IV | |
| dapagliflozin 10 MG Oral Tablet | po | |
| Novolin R Injectable Product | subcutaneous | |
| Insulin Lispro 100 UNT/ML Pen Injector [Humalog] | subcut. | |
| Metformin | ou | |
| Humulin Injectable Product | Subcutaneously | |
| Acarbose Oral Tablet [Precose] | | |
| 3 ML Insulin Lispro 200 UNT/ML Pen Injector | subcutaneous | |
| Insulin, Aspart, Human 100 UNT/ML Cartridge [NovoLog] | NPO. | |
| dulaglutide 1.5 MG/ML [Trulicity] | subq | |
| Linagliptin 5 MG Oral Tablet [Tradjenta] | PO. | |
| Insulin Lispro | po. | |
| Regular Insulin, Human 100 UNT/ML Injectable Solution | SUBCUTANEOUS | |
| Glyburide 5 MG Oral Tablet [Diabeta] | ORAL | |

| | | |
|---|---|---|
| Regular Insulin, Human 20 UNT/ML | sub Q | |
| sitagliptin 50 MG Oral Tablet | p.o. | |
| sitagliptin 50 MG Oral Tablet [Januvia] | g-tube | |
| Januvia Pill | po | |
| Metformin hydrochloride 1000 MG / saxagliptin 5 MG [Kombiglyze] | po | |
| insulin human, isophane 30 UNT/ML / Regular Insulin, Human 10 UNT/ML Injectable Suspension | subcut | |
| insulin pork, isophane 100 UNT/ML Injectable Suspension [NPH Iletin II] | Sub Q | |
| miglitol Oral Tablet | by mouth | |
| insulin detemir Injectable Product | SUBCUT | |
| Levemir Injectable Product | by mouth | |
| 24 HR Glipizide 10 MG Extended Release Oral Tablet | Oral | |
| insulin detemir | oral | |
| Metformin 500 MG | drip. | |
| Glipizide | intramuscular | |
| rosiglitazone 2 MG Oral Tablet | p.o. | |
| Glipizide | sl | |
| Glyburide 5 MG Oral Tablet | p.o | |
| insulin human, isophane 75 UNT/ML | subq. | |
| nateglinide 60 MG [Starlix] | per mouth | |
| Glipizide 10 MG | nostril | |
| Pramlintide | sub q | |
| Insulin, Aspart, Human | Drip | |
| Metformin hydrochloride 750 MG [Glucophage] | orally | |
| Gliclazide | | |
| Chlorpropamide 100 MG Oral Tablet [Diabinese] | by mouth | |
| insulin human, isophane 16 UNT/ML | subQ | |
| insulin, regular, beef-pork 40 UNT/ML Inhalant Solution | po | |
| 3 ML liraglutide 6 MG/ML Pen Injector [Victoza] | subc | |
| glimepiride 2 MG | subcutaneously | |
| Metformin hydrochloride 500 MG / pioglitazone 15 MG Oral Tablet [Actoplus Met] | | 0 |
| Glyburide 5 MG Oral Tablet [Micronase] | ORAL | |
| alogliptin 25 MG / pioglitazone 15 MG Oral Tablet [Oseni] | | |
| Metformin hydrochloride 500 MG [Glucophage] | po. | |
| Glyburide | ng | |
| Insulin Glargine 100 UNT/ML Injectable Solution [Lantus] | SUBCU | |

| | | |
|---|---|---|
| 24 HR Metformin hydrochloride 1000 MG / sitagliptin 100 MG Extended Release Oral Tablet [Janumet] | by mouth. | |
| Metformin hydrochloride 500 MG / rosiglitazone 4 MG Oral Tablet | oral | |
| Glipizide 5 MG / Metformin hydrochloride 500 MG Oral Tablet [Metaglip] | PO | |
| Acetohexamide 500 MG Oral Tablet | po | |
| insulin human, isophane 70 UNT/ML / Regular Insulin, Human 30 UNT/ML [Novolin] | gt | |
| 0.5 ML albiglutide 60 MG/ML Pen Injector [Tanzeum] | SubQ | |
| Insulin Glargine | Oral | |
| Insulin, Regular, Pork 40 UNT/ML Injectable Solution | SQ | |
| Glyburide 1 MG Oral Tablet | oral | |
| Chlorpropamide 250 MG Oral Tablet [Diabinese] | p.o. | |
| Metformin 500 MG / ropinirole 2 MG Oral Tablet | p.o. | |
| 24 HR Metformin hydrochloride 1000 MG / sitagliptin 50 MG Extended Release Oral Tablet [Janumet] | | 0 |
| 24 HR dapagliflozin 5 MG / Metformin hydrochloride 1000 MG Extended Release Oral Tablet [Xigduo] | | |
| Linagliptin 2.5 MG / Metformin hydrochloride 500 MG Oral Tablet | by mouth | |
| insulin human, isophane 60 UNT/ML / Regular Insulin, Human 40 UNT/ML [Humulin] | SQ | |
| Glipizide 2.5 MG / Metformin hydrochloride 250 MG Oral Tablet [Metaglip] | | |
| Metformin hydrochloride 100 MG/ML Oral Solution [Riomet] | | 0 |
| insulin human, isophane 70 UNT/ML / Regular Insulin, Human 30 UNT/ML Injectable Suspension [Humulin] | SUBCUTANEOUSLY | |
| Jardiance Pill | | |
| Insulin Glargine 100 UNT/ML Injectable Solution | orally | |
| Acarbose 25 MG Oral Tablet | p.o. | |
| Insulin Lispro | per tube | |
| Glipizide 10 MG | npo | |
| Insulin Lispro 25 UNT/ML / Insulin, Protamine Lispro, Human 75 UNT/ML Injectable Suspension [Humalog Mix] | nostril | |
| Insulin Lispro | FLUSH | |
| Glyburide 5 MG Oral Tablet [Daonil] | po | |
| dapagliflozin 10 MG [Farxiga] | subcutaneous | |
| Insulin, Aspart, Human 100 UNT/ML Cartridge [NovoLog] | Sq | |
| Regular Insulin, Human 20 UNT/ML | Subcutaneous | |

| | | |
|---|---|---|
| Glyburide | subcu | |
| 3 ML Insulin, Aspart Protamine, Human 70 UNT/ML / Insulin, Aspart, Human 30 UNT/ML Pen Injector [NovoLog Mix] | oral | |
| canagliflozin 300 MG Oral Tablet | by mouth | |
| 3 ML Insulin, Aspart Protamine, Human 70 UNT/ML / Insulin, Aspart, Human 30 UNT/ML Pen Injector [NovoLog Mix] | SC | |
| insulin pork, isophane 70 UNT/ML / Insulin, Regular, Pork 30 UNT/ML Injectable Suspension | SUBQ | |
| exenatide 0.005 MG/ACTUAT Pen Injector | by mouth | |
| 3 ML insulin human, isophane 100 UNT/ML Pen Injector [Humulin N] | sq | |
| alogliptin 25 MG Oral Tablet [Nesina] | | |
| NovoLog Mix Injectable Product | | |
| Glipizide | npo. | |
| nateglinide 120 MG Oral Tablet [Starlix] | Oral | |
| NovoLog Injectable Product | SUBCUTANEOUSLY | |
| 60 ACTUAT exenatide 0.005 MG/ACTUAT Pen Injector [Byetta] | IM | |
| Glyburide / Metformin Oral Product | oral | |
| Regular Insulin, Human 30 UNT/ML | SQ | |
| glimepiride 1 MG | per tube | |
| Glyburide Oral Tablet [Diabeta] | | |
| 3 ML Insulin, Aspart Protamine, Human 70 UNT/ML / Insulin, Aspart, Human 30 UNT/ML Pen Injector [NovoLog Mix] | MISC.(NON-DRUG; COMBO ROUTE) | |
| insulin degludec 100 UNT/ML | | |
| Glipizide 10 MG Oral Tablet | sublingual | |
| Glipizide 2.5 MG / Metformin hydrochloride 500 MG Oral Tablet [Metaglip] | | 0 |
| Tradjenta Oral Product | inhalation | |
| pioglitazone 30 MG [Actos] | per mouth | |
| Metformin 500 MG | j - tube | |
| repaglinide 1 MG Oral Tablet | | 0 |
| Insulin Lispro Pen Injector [Humalog] | SUBCUT | |
| insulin, isophane Injectable Suspension [Humulin N] | subcutaneously | |
| Insulin, Aspart, Human | sl | |
| Ultralente Insulin, Human 100 UNT/ML Injectable Suspension | subcutaneous | |
| 3 ML insulin detemir 100 UNT/ML Pen Injector [Levemir] | SUBCUT | |
| canagliflozin 150 MG / Metformin hydrochloride 1000 MG Oral Tablet [Invokamet] | oral | |
| Insulin Lispro 100 UNT/ML [Humalog] | SC | |
| pioglitazone 30 MG | per tube | |

| | | |
|---|---|---|
| Metformin hydrochloride 500 MG / rosiglitazone 4 MG Oral Tablet [Avandamet] | p.o. | |
| Ultralente Insulin, Human 100 UNT/ML Injectable Suspension [Humulin U Ultralente] | sq | |
| Metformin hydrochloride 1000 MG [Glucophage] | nostril | |
| Acarbose Pill | | |
| insulin human, isophane 36 UNT/ML | SubQ | |
| insulin pork, isophane 70 UNT/ML / Insulin, Regular, Pork 30 UNT/ML Injectable Suspension | Subcutaneously | |
| Metformin 500 MG | SC | |
| Metformin hydrochloride 1000 MG / saxagliptin 2.5 MG [Kombiglyze] | topical | |
| insulin human, rDNA origin 3 MG/ACTUAT Inhalant Powder [Exubera] | inhaled. | |
| exenatide 2 MG [Bydureon] | Subcutaneous | |
| repaglinide 0.5 MG Oral Tablet | | |
| 24 HR dapagliflozin 10 MG / Metformin hydrochloride 1000 MG Extended Release Oral Tablet [Xigduo] | by mouth | |
| Metformin hydrochloride 500 MG [Glucophage] | gt | |
| Regular Insulin, Human 100 UNT/ML Injectable Solution [Humulin R] | subcutaneously. | |
| Tolbutamide 500 MG Oral Tablet [Orinase] | PO | |
| 3 ML liraglutide 6 MG/ML Pen Injector [Victoza] | sub Q. | |
| Metformin 1000 MG | SQ | |
| Regular Insulin, Human | Subq | |
| insulin human, isophane 40 UNT/ML | IM | |
| insulin human, isophane 50 UNT/ML / Regular Insulin, Human 50 UNT/ML Injectable Suspension | subcu | |
| Glyburide 1 MG | | 0 |
| Diabinese Oral Product | p.o. | |
| Insulin Glargine 100 UNT/ML | SC | |
| insulin detemir | SubQ | |
| Glyburide 5 MG / Phenformin 50 MG Oral Tablet | PO | |
| Glipizide 10 MG | per tube | |
| Insulin, Zinc, Human | subq | |
| Insulin Lispro 50 UNT/ML | gt | |
| Glyburide 5 MG [Diabeta] | per tube | |
| insulin human, isophane 100 UNT/ML Prefilled Syringe | SC | |
| 3 ML Insulin Lispro 200 UNT/ML Pen Injector | by mouth | |

| | |
|---|---|
| 3 ML Insulin, Aspart Protamine, Human 70 UNT/ML / Insulin, Aspart, Human 30 UNT/ML Pen Injector [NovoLog Mix] | MISCELLANEOUS |
| exenatide 0.005 MG/ACTUAT Pen Injector | |
| Metformin hydrochloride 500 MG / rosiglitazone 4 MG Oral Tablet | orally |
| Actoplus Met Oral Product | BY MOUTH |
| alogliptin 12.5 MG / Metformin hydrochloride 1000 MG [Kazano] | |
| Metformin hydrochloride 500 MG / rosiglitazone 4 MG Oral Tablet | PO |
| Acarbose 100 MG Oral Tablet [Precose] | orally |
| alogliptin 12.5 MG / Metformin hydrochloride 1000 MG Oral Tablet [Kazano] | by mouth |
| 3 ML Insulin, Aspart Protamine, Human 70 UNT/ML / Insulin, Aspart, Human 30 UNT/ML Pen Injector [NovoLog Mix] | NPO. |
| Regular Insulin, Human 500 UNT/ML Injectable Solution | subcutaneous |
| Glumetza Oral Product | PO |
| exenatide 0.01 MG/ACTUAT | subq |
| 3 ML insulin human, isophane 100 UNT/ML Prefilled Syringe | G - tube |
| rosiglitazone | PER TUBE |
| 24 HR dapagliflozin 10 MG / Metformin hydrochloride 500 MG Extended Release Oral Tablet [Xigduo] | |
| insulin degludec 200 UNT/ML | by mouth |
| Simvastatin 10 MG / sitagliptin 100 MG [Juvisync] | subcutaneously |
| 3 ML Insulin Glargine 100 UNT/ML Pen Injector [Lantus] | IV |
| exenatide 0.005 MG/ACTUAT | SC |
| Metformin hydrochloride 1000 MG / rosiglitazone 2 MG Oral Tablet [Avandamet] | p.o. |
| 60 ACTUAT exenatide 0.01 MG/ACTUAT Pen Injector [Byetta] | SUBQ |
| 3 ML insulin human, isophane 70 UNT/ML / Regular Insulin, Human 30 UNT/ML Pen Injector | IM |
| Xigduo Oral Product | by mouth |
| Metformin hydrochloride 850 MG Oral Tablet [Glucophage] | BY MOUTH |
| rosiglitazone Oral Tablet [Avandia] | PO |
| Insulin Lispro 25 UNT/ML | Sub Q |
| insulin pork, isophane 100 UNT/ML Injectable Suspension [NPH Iletin II] | bedside. |
| Glyburide 1.25 MG | per tube |
| Insulin, Aspart, Human 100 UNT/ML Injectable Solution [NovoLog] | SUBCUTANEOUS PER INSULIN PUMP |
| exenatide 0.01 MG/ACTUAT [Byetta] | p.o. |

| | | |
|---|---|---|
| Glyburide 3 MG Oral Tablet [Glynase] | orally | |
| Metforming Oral Product | po | |
| 3 ML Insulin, Aspart, Human 100 UNT/ML Pen Injector | SUBCUTANEOUS | |
| Regular Insulin, Human | TPN | |
| Glipizide 10 MG | gt | |
| glimepiride 1 MG Oral Tablet [Amaryl] | Oral | |
| 3 ML liraglutide 6 MG/ML Pen Injector [Victoza] | sUBQ | |
| 60 ACTUAT exenatide 0.01 MG/ACTUAT Pen Injector [Byetta] | Sub Q | |
| Glyburide 1.25 MG Oral Tablet [Diabeta] | | |
| Insulin, Aspart, Human 100 UNT/ML Injectable Solution [NovoLog] | pump | |
| pioglitazone 15 MG Oral Tablet [Actos] | p.o | |
| Ultralente Insulin, Human 100 UNT/ML Injectable Suspension | SQ | |
| 3 ML Insulin Lispro 100 UNT/ML Cartridge [Humalog] | SUBQ | |
| insulin human, isophane 100 UNT/ML Injectable Suspension | SQ | |
| 0.5 ML albiglutide 60 MG/ML Pen Injector [Tanzeum] | | |
| 24 HR Metformin hydrochloride 1000 MG / saxagliptin 2.5 MG Extended Release Oral Tablet [Kombiglyze] | | 0 |
| 0.5 ML albiglutide 60 MG/ML Pen Injector [Tanzeum] | subq | |
| Glyburide / Metformin Oral Tablet | po | |
| glimepiride 4 MG / pioglitazone 30 MG Oral Tablet [Duetact] | ORAL | |
| miglitol 50 MG Oral Tablet [Glyset] | p.o. | |
| Humulin Injectable Product | SQ | |
| Insulin Glargine 100 UNT/ML [Lantus] | subc. | |
| Insulin Lispro Injectable Solution [Humalog] | subcutaneous | |
| canagliflozin 100 MG Oral Tablet | orally | |
| Metformin hydrochloride 850 MG [Glucophage] | per tube | |
| Glipizide | bedside | |
| 24 HR Metformin hydrochloride 500 MG Extended Release Oral Tablet [Glucophage] | p.o. | |
| glimepiride | drip | |
| Glyburide 3 MG Oral Tablet [Glynase] | oral | |
| insulin detemir | subQ | |
| alogliptin 12.5 MG / Metformin hydrochloride 500 MG Oral Tablet | by mouth | |
| Insulin Glargine Injectable Solution | SUBCUTANEOUSLY | |
| insulin human, isophane 32 UNT/ML | subq. | |
| Metformin / sitagliptin Extended Release Oral Tablet [Janumet] | | |
| Metformin hydrochloride 500 MG [Glucophage] | ng | |

| | | |
|---|---|---|
| 24 HR empagliflozin 25 MG / Metformin hydrochloride 1000 MG Extended Release Oral Tablet [Synjardy] | | |
| Regular Insulin, Human | | 0 |
| insulin human, isophane 70 UNT/ML / Regular Insulin, Human 30 UNT/ML Injectable Suspension [Humulin] | by mouth | |
| exenatide 2 MG | subQ | |
| pioglitazone 30 MG [Actos] | bedside | |
| Glynase Oral Product | Orally | |
| Metformin hydrochloride 500 MG [Glucophage] | SQ | |
| rosiglitazone Oral Product | p.o. | |
| Metformin / sitagliptin Extended Release Oral Tablet [Janumet] | by mouth | |
| Glyburide 5 MG | per mouth | |
| Regular Insulin, Human 5 UNT/ML | subq | |
| 3 ML liraglutide 6 MG/ML Pen Injector [Victoza] | subcut. | |
| Glipizide | im | |
| Metformin hydrochloride 1000 MG / sitagliptin 50 MG Oral Tablet | BY MOUTH | |
| Insulin, Zinc, Pork | subcu | |
| Insulin, Aspart, Human 100 UNT/ML Cartridge [NovoLog] | SQ. | |
| Insulin Lispro | Subcutaneously | |
| exenatide 2 MG [Bydureon] | orally | |
| Byetta Injectable Product | | 0 |
| 24 HR Metformin hydrochloride 1000 MG / sitagliptin 100 MG Extended Release Oral Tablet [Janumet] | orally | |
| Kazano Oral Product | po | |
| Insulin Lispro 50 UNT/ML / Insulin, Protamine Lispro, Human 50 UNT/ML Injectable Suspension [Humalog Mix] | PO | |
| Insulin Lispro 25 UNT/ML / Insulin, Protamine Lispro, Human 75 UNT/ML Injectable Suspension [Humalog Mix] | SEE COMMENT | |
| Metformin Oral Tablet [Glucophage] | trans | |
| Insulin Lispro | subq. | |
| nateglinide 60 MG Oral Tablet [Starlix] | Oral | |
| Insulin Glargine Injectable Solution [Lantus] | subq | |
| Insulin Lispro 25 UNT/ML | SC | |
| repaglinide Oral Product | po | |
| Glipizide 10 MG | po. | |
| insulin human, isophane 70 UNT/ML / Regular Insulin, Human 30 UNT/ML [Humulin] | IV | |
| Glyburide 5 MG Oral Tablet | PER TUBE | |
| 60 ACTUAT exenatide 0.01 MG/ACTUAT Pen Injector [Byetta] | p.o. | |

| | | |
|---|---|---|
| 1.5 ML insulin human, isophane 70 UNT/ML / Regular Insulin, Human 30 UNT/ML Prefilled Syringe [Novolin] | | |
| Glyburide 5 MG | gt | |
| Metformin hydrochloride 500 MG Oral Tablet | per tube | |
| Modified 24 HR Metformin hydrochloride 500 MG Extended Release Oral Tablet [Glumetza] | | 0 |
| glimepiride Oral Tablet | po | |
| Metformin hydrochloride 850 MG / pioglitazone 15 MG Oral Tablet [Actoplus Met] | BY MOUTH | |
| Metformin 500 MG | subcut | |
| insulin human, isophane 50 UNT/ML / Regular Insulin, Human 50 UNT/ML [Humulin] | subcut. | |
| exenatide 2 MG [Bydureon] | PO | |
| Glipizide 5 MG Oral Tablet | PER TUBE | |
| insulin human, isophane 80 UNT/ML / Regular Insulin, Human 20 UNT/ML [Humulin] | SQ | |
| insulin pork, isophane 100 UNT/ML Injectable Suspension [NPH Iletin II] | iv | |
| insulin pork, isophane 100 UNT/ML Injectable Suspension [NPH Iletin II] | inhalation | |
| 24 HR Glipizide 5 MG Extended Release Oral Tablet [Glucotrol] | | |
| glimepiride 4 MG / rosiglitazone 8 MG Oral Tablet [Avandaryl] | ORAL | |
| insulin human, isophane 28 UNT/ML | Subcut. | |
| sitagliptin 50 MG Oral Tablet | PO | |
| insulin detemir 100 UNT/ML Injectable Solution [Levemir] | SUBCUTANEOUSLY | |
| Glyburide / Metformin Oral Product | p.o. | |
| Insulin Lispro 100 UNT/ML Injectable Solution [Humalog] | injection | |
| alogliptin 12.5 MG Oral Tablet [Nesina] | by mouth | |
| Insulin Glargine 100 UNT/ML [Lantus] | Subcut. | |
| NovoLog Injectable Product | by mouth | |
| Metformin 1000 MG | G - TUBE | |
| Insulin, Glulisine, Human 100 UNT/ML Injectable Solution [Apidra] | sc | |
| glimepiride 3 MG Oral Tablet | PO | |
| Humulin N Injectable Product | Subq | |
| Glipizide | subq | |
| Acarbose Oral Tablet | oral | |
| Bromocriptine 5 MG Oral Capsule [Parlodel] | | |
| Regular Insulin, Human 12 UNT | NPO | |

| | |
|---|---|
| insulin human, isophane 100 UNT/ML Injectable Suspension [Novolin N] | sub q |
| Regular Insulin, Human 40 UNT/ML | subcutaneous |
| Lente Insulin, Beef 80 UNT/ML | subcu |
| insulin human, isophane 70 UNT/ML | Subq. |
| Regular Insulin, Human 500 UNT/ML Injectable Solution | SC |
| Metformin / saxagliptin Extended Release Oral Tablet [Kombiglyze] | |
| Metformin 1000 MG | IV |
| Insulin, Aspart, Human | subcutaneously. |
| 24 HR Metformin hydrochloride 500 MG / sitagliptin 50 MG Extended Release Oral Tablet [Janumet] | ORAL |
| Metaglip Oral Product | oral |
| glimepiride 2 MG / rosiglitazone 4 MG Oral Tablet [Avandaryl] | |
| liraglutide 6 MG/ML [Victoza] | po |
| insulin human, isophane 70 UNT/ML / Regular Insulin, Human 30 UNT/ML [Novolin] | SUBCUTANEOUS |
| 3 ML insulin human, isophane 100 UNT/ML Prefilled Syringe [Novolin N] | by mouth |
| insulin human, isophane 50 UNT/ML / Regular Insulin, Human 50 UNT/ML Injectable Suspension [Humulin] | Subcutaneous |
| 3 ML Insulin Lispro 50 UNT/ML / Insulin, Protamine Lispro, Human 50 UNT/ML Pen Injector [Humalog Mix] | |
| Regular Insulin, Human 20 UNT/ML | subcutaneously |
| insulin human, isophane 50 UNT/ML / Regular Insulin, Human 50 UNT/ML [Humulin] | subq |
| 3 ML insulin detemir 100 UNT/ML Pen Injector [Levemir] | SUBQ |
| Glyburide 1.25 MG / Metformin hydrochloride 250 MG Oral Tablet [Glucovance] | orally |
| Metformin 500 MG | PO. |
| 24 HR Metformin hydrochloride 1000 MG / pioglitazone 15 MG Extended Release Oral Tablet [Actoplus Met] | ORAL |
| Metformin hydrochloride 500 MG / sitagliptin 50 MG [Janumet] | gt |
| glimepiride 4 MG / pioglitazone 30 MG Oral Tablet [Duetact] | |
| Insulin Lispro | iv. |
| insulin human, isophane 90 UNT/ML / Regular Insulin, Human 10 UNT/ML Injectable Suspension | |
| Insulin, Aspart, Human | inhalation |
| Insulin Lispro 25 UNT/ML | sub q. |
| Bromocriptine 2.5 MG | gtube |

| | | |
|---|---|---|
| Glyburide / Metformin Oral Tablet [Glucovance] | orally | |
| empagliflozin 25 MG / Linagliptin 5 MG Oral Tablet [Glyxambi] | po | |
| sitagliptin 25 MG Oral Tablet [Januvia] | Oral | |
| Glipizide Extended Release Oral Tablet [Glucotrol] | BY MOUTH | |
| Precose Oral Product | p.o. | |
| Insulin Lispro 100 UNT/ML Injectable Solution [Lispro PRC] | bedside | |
| Insulin Lispro | inhalation | |
| sitagliptin Oral Tablet [Januvia] | oral | |
| insulin human, isophane 70 UNT/ML / Regular Insulin, Human 30 UNT/ML [Novolin] | | 0 |
| Lente Insulin, Beef-Pork 40 UNT/ML | | |
| Insulin Lispro | *TOPICAL | |
| liraglutide | | |
| exenatide 0.005 MG/ACTUAT [Byetta] | p.o. | |
| Janumet Pill | by mouth | |
| Metformin hydrochloride 750 MG | ou | |
| saxagliptin 5 MG Oral Tablet | p.o. | |
| Levemir Injectable Product | SC | |
| insulin human, isophane 50 UNT/ML | | |
| Regular Insulin, Human 100 UNT/ML Injectable Solution [Humulin R] | npo. | |
| Insulin Lispro Cartridge [Humalog] | Sq | |
| Insulin, Aspart Protamine, Human 70 UNT/ML / Insulin, Aspart, Human 30 UNT/ML [NovoLog Mix] | subcutaneously. | |
| Glipizide 5 MG | IM | |
| pioglitazone | swallow | |
| dapagliflozin 10 MG Oral Tablet | by mouth | |
| glimepiride 1 MG | npo | |
| repaglinide 2 MG Oral Tablet | p.o. | |
| Glyburide Oral Tablet [Diabeta] | orally | |
| Lantus Injectable Product | SUBCUTANEOUSLY | |
| Metformin hydrochloride 500 MG Extended Release Oral Tablet [Glumetza] | Po | |
| sitagliptin 25 MG Oral Tablet [Januvia] | BY MOUTH | |
| Metformin hydrochloride 100 MG/ML Oral Solution [Riomet] | swallow | |
| Insulin Lispro 50 UNT/ML / Insulin, Protamine Lispro, Human 50 UNT/ML Injectable Suspension [Humalog Mix] | sc | |
| 3 ML Insulin Lispro 100 UNT/ML Cartridge [Humalog] | SQ | |
| 3 ML insulin detemir 100 UNT/ML Pen Injector [Levemir] | subcutaneously. | |
| insulin human, isophane 32 UNT/ML | sub q | |

| | | |
|---|---|---|
| Insulin Lispro Cartridge [Humalog] | bedside | |
| Metformin | | 0 |
| 3 ML liraglutide 6 MG/ML Pen Injector [Saxenda] | SQ | |
| insulin human, isophane 70 UNT/ML / Regular Insulin, Human 30 UNT/ML [Humulin] | SUBQ. | |
| Glipizide 5 MG [Glucotrol] | IV | |
| Glipizide | SC | |
| Fortamet Oral Product | Oral | |
| Glipizide Oral Tablet [Glucotrol] | PO | |
| insulin, regular, beef-pork 100 UNT/ML Injectable Solution | subcutaneously | |
| Regular Insulin, Human 20 UNT/ML | SUBCUT | |
| canagliflozin 150 MG / Metformin hydrochloride 500 MG [Invokamet] | | |
| 60 ACTUAT exenatide 0.005 MG/ACTUAT Pen Injector [Byetta] | Subcut | |
| nateglinide 120 MG Oral Tablet [Starlix] | BY MOUTH | |
| insulin detemir 100 UNT/ML [Levemir] | po | |
| 3 ML Insulin Lispro 100 UNT/ML Pen Injector [Humalog] | miscellaneous | |
| Regular Insulin, Human | inhalatn | |
| 3 ML Insulin Lispro 200 UNT/ML Pen Injector | IM | |
| canagliflozin 100 MG Oral Tablet [Invokana] | Oral | |
| Metformin hydrochloride 750 MG [Glucophage] | po | |
| Insulin, Aspart Protamine, Human 70 UNT/ML / Insulin, Aspart, Human 30 UNT/ML [NovoLog Mix] | subc | |
| Novolin R Injectable Product | subq | |
| Metformin Extended Release Oral Tablet | po | |
| alogliptin | | |
| insulin human, isophane 70 UNT/ML / Regular Insulin, Human 30 UNT/ML [Humulin] | | 0 |
| Glyburide 5 MG / Metformin hydrochloride 500 MG Oral Tablet | BY MOUTH | |
| Insulin Lispro 50 UNT/ML | p.o. | |
| 24 HR dapagliflozin 5 MG / Metformin hydrochloride 500 MG Extended Release Oral Tablet [Xigduo] | oral | |
| empagliflozin 5 MG / Metformin hydrochloride 1000 MG Oral Tablet [Synjardy] | by mouth | |
| 3 ML Insulin, Aspart Protamine, Human 70 UNT/ML / Insulin, Aspart, Human 30 UNT/ML Pen Injector [NovoLog Mix] | sub Q | |
| 24 HR Metformin hydrochloride 1000 MG / sitagliptin 50 MG Extended Release Oral Tablet [Janumet] | | |
| 3 ML insulin human, isophane 100 UNT/ML Pen Injector [Humulin N] | subcut. | |

| | |
|---|---|
| Farxiga Oral Product | PO |
| Lantus Injectable Product | subcutaneous. |
| miglitol 100 MG Oral Tablet [Glyset] | |
| Insulin, Aspart, Human 100 UNT/ML Cartridge [NovoLog] | orally |
| Bromocriptine 1 MG Oral Tablet [Parlodel] | po |
| glimepiride Oral Tablet [Amaryl] | |
| dulaglutide 3 MG/ML [Trulicity] | Subcutaneous |
| 24 HR Metformin hydrochloride 500 MG Extended Release Oral Tablet [Glucophage] | gt |
| 3 ML liraglutide 6 MG/ML Pen Injector [Saxenda] | Sub Q |
| Glipizide 2.5 MG | subq |
| insulin detemir 100 UNT/ML Injectable Solution | subcutaneously |
| Glipizide 10 MG [Glucotrol] | subcutaneously |
| Glyburide 5 MG Oral Tablet [Micronase] | p.o |
| Metformin | neb |
| glimepiride 2 MG | npo |
| insulin human, isophane 34 UNT/ML | SUB Q |
| Insulin Glargine 100 UNT/ML [Lantus] | gt |
| Glyburide 1 MG Oral Tablet | p.o. |
| dapagliflozin 5 MG Oral Tablet [Farxiga] | p.o. |
| insulin human, isophane 70 UNT/ML / Regular Insulin, Human 30 UNT/ML [Humulin] | im |
| empagliflozin 25 MG Oral Tablet | orally |
| Glyburide | sq |
| glimepiride 4 MG Oral Tablet [Amaryl] | p.o |
| insulin detemir | sub Q |
| glimepiride 4 MG [Amaryl] | nostril |
| Metformin hydrochloride 850 MG | by mouth. |
| Insulin Glargine | inhaled |
| insulin, regular, beef-pork 40 UNT/ML Inhalant Solution | npo |
| Glipizide 5 MG Oral Tablet | pO |
| 60 ACTUAT exenatide 0.01 MG/ACTUAT Pen Injector [Byetta] | oral |
| Regular Insulin, Human 100 UNT/ML Injectable Solution [Humulin R] | subq. |
| Insulin Lispro 100 UNT/ML Injectable Solution [Humalog] | subq |
| Ultralente Insulin, Human 100 UNT/ML Injectable Suspension [Humulin U Ultralente] | SUBCUTANEOUS |
| Regular Insulin, Human 100 UNT/ML Injectable Solution [Novolin R] | SUBQ |
| Acarbose 100 MG | G - tube |

| | | |
|---|---|---|
| 3 ML Insulin Lispro 100 UNT/ML Pen Injector [Humalog] | injection | |
| Tolazamide 100 MG [Tolinase] | | |
| Insulin, Aspart, Human 100 UNT/ML Injectable Solution | sq | |
| Regular Insulin, Human 5 UNT/ML | SQ | |
| Amaryl Oral Product | Oral | |
| Glipizide 2.5 MG / Metformin hydrochloride 250 MG Oral Tablet | orally | |
| Tolazamide 250 MG [Tolinase] | | |
| insulin human, isophane 32 UNT/ML | SC | |
| glimepiride 4 MG | gt | |
| 1.5 ML insulin human, isophane 70 UNT/ML / Regular Insulin, Human 30 UNT/ML Prefilled Syringe | SC | |
| Insulin Lispro 100 UNT/ML | SQ | |
| Metformin hydrochloride 1000 MG / saxagliptin 2.5 MG Extended Release Oral Tablet | by mouth | |
| Glipizide 2.5 MG [Glucotrol] | npo | |
| empagliflozin 10 MG / Linagliptin 5 MG Oral Tablet [Glyxambi] | ORAL | |
| glimepiride 4 MG / pioglitazone 30 MG Oral Tablet | orally | |
| 0.5 ML albiglutide 60 MG/ML Pen Injector [Tanzeum] | | 0 |
| pioglitazone Oral Tablet [Actos] | Po | |
| Ultralente Insulin, Human 100 UNT/ML Injectable Suspension | subcu | |
| exenatide 0.005 MG/ACTUAT [Byetta] | | 0 |
| rosiglitazone 1 MG | | |
| dulaglutide 3 MG/ML [Trulicity] | subQ | |
| Insulin Glargine | IM | |
| Insulin Glargine 100 UNT/ML | po | |
| Insulin Glargine | sQ | |
| 3 ML liraglutide 6 MG/ML Pen Injector | orally | |
| 0.65 ML exenatide 3.08 MG/ML Pen Injector [Bydureon] | subcutaneously | |
| Glyburide 2.5 MG Oral Tablet [Diabeta] | orally | |
| Acarbose 50 MG Oral Tablet [Precose] | | 0 |
| glimepiride 8 MG Oral Tablet | oral | |
| Insulin Lispro | im | |
| exenatide 0.01 MG/ACTUAT [Byetta] | subc | |
| 24 HR Glipizide 5 MG Extended Release Oral Tablet | | |
| 3 ML insulin detemir 100 UNT/ML Pen Injector [Levemir] | oral | |
| Regular Insulin, Human | npo | |
| Metformin hydrochloride 500 MG Oral Tablet [Glucophage] | ORALLY | |
| sitagliptin Oral Tablet [Januvia] | | |
| rosiglitazone 4 MG | per tube | |
| Simvastatin 20 MG / sitagliptin 50 MG Oral Tablet | | |

| | | |
|---|---|---|
| insulin human, isophane 28 UNT/ML | Subcutaneous | |
| Metformin hydrochloride 1000 MG Oral Tablet | p.o | |
| insulin human, isophane 100 UNT/ML Injectable Suspension | orally | |
| Insulin, Protamine Lispro, Human | subcutaneously | |
| Oseni Oral Product | orally | |
| Glipizide | subcu | |
| Insulin Lispro 200 UNT/ML [Humalog] | by mouth | |
| Insulin, Aspart Protamine, Human 70 UNT/ML / Insulin, Aspart, Human 30 UNT/ML Injectable Suspension [NovoLog Mix] | oral | |
| Simvastatin 20 MG / sitagliptin 100 MG Oral Tablet [Juvisync] | oral | |
| dapagliflozin 5 MG / Metformin hydrochloride 500 MG [Xigduo] | | |
| Glipizide Extended Release Oral Tablet | | |
| empagliflozin 25 MG Oral Tablet [Jardiance] | | 0 |
| Pramlintide Pen Injector [Symlin] | | |
| Parlodel Oral Product | p.o. | |
| alogliptin 25 MG Oral Tablet [Nesina] | ORAL | |
| Metformin hydrochloride 1000 MG / saxagliptin 5 MG [Kombiglyze] | topical | |
| exenatide 0.01 MG/ACTUAT [Byetta] | Sub q | |
| Regular Insulin, Human 10 UNT/ML | per tube | |
| miglitol | PO | |
| liraglutide 6 MG/ML | subcutaneously | |
| Linagliptin 2.5 MG / Metformin hydrochloride 500 MG Oral Tablet [Jentadueto] | ORAL | |
| Metformin hydrochloride 1000 MG / saxagliptin 2.5 MG [Kombiglyze] | po | |
| Regular Insulin, Human 4 UNT Inhalant Powder [Afrezza] | INHALATION | |
| liraglutide 6 MG/ML [Saxenda] | SQ | |
| Xigduo Pill | | |
| glimepiride 1 MG / rosiglitazone 4 MG Oral Tablet [Avandaryl] | PO | |
| Metformin 1000 MG | npo. | |
| Metformin hydrochloride 1000 MG [Glucophage] | PO. | |
| Precose Oral Product | BY MOUTH | |
| liraglutide 6 MG/ML [Victoza] | Subcutaneously | |
| Insulin Glargine 100 UNT/ML [Lantus] | Sq | |
| pioglitazone 30 MG [Actos] | ng | |
| rosiglitazone 1 MG Oral Tablet | po | |
| 60 ACTUAT exenatide 0.005 MG/ACTUAT Pen Injector [Byetta] | subq. | |
| Insulin Lispro Injectable Solution [Humalog] | SUBCUT | |
| dapagliflozin 5 MG Oral Tablet [Farxiga] | | 0 |

| | |
|---|---|
| dulaglutide 3 MG/ML [Trulicity] | sc |
| Metformin 500 MG | inhalation |
| 3 ML Insulin, Glulisine, Human 100 UNT/ML Pen Injector [Apidra] | subQ |
| Glyburide 3 MG Oral Tablet | |
| Metformin hydrochloride 500 MG Oral Tablet [Glucophage] | per tube |
| empagliflozin 12.5 MG / Metformin hydrochloride 500 MG [Synjardy] | |
| dulaglutide Auto-Injector [Trulicity] | |
| Glipizide 10 MG Oral Tablet | BY mouth |
| Insulin, Zinc, Human | SUBCUT |
| exenatide 2 MG Injection [Bydureon] | IM Injection |
| Metformin hydrochloride 1000 MG / sitagliptin 100 MG [Janumet] | PO |
| 24 HR canagliflozin 150 MG / Metformin hydrochloride 1000 MG Extended Release Oral Tablet [Invokamet] | |
| 3 ML liraglutide 6 MG/ML Pen Injector [Victoza] | SUBCU |
| 60 ACTUAT exenatide 0.01 MG/ACTUAT Pen Injector [Byetta] | SUB Q |
| insulin detemir | SEE COMMENT |
| {60 (Regular Insulin, Human 4 UNT Inhalant Powder) / 30 (Regular Insulin, Human 8 UNT Inhalant Powder) } Pack | subcut |
| Linagliptin 2.5 MG / Metformin hydrochloride 500 MG Oral Tablet [Jentadueto] | oral |
| Novolin R Injectable Product | Subcut |
| Insulin Lispro 100 UNT/ML | subcutaneously. |
| 60 ACTUAT exenatide 0.005 MG/ACTUAT Pen Injector [Byetta] | SUB Q |
| glimepiride 1 MG [Amaryl] | by mouth. |
| insulin human, isophane 28 UNT/ML | po |
| Insulin Glargine | MISC |
| Regular Insulin, Human | drip. |
| Lantus Injectable Product | COMBO ROUTE) |
| Insulin, Aspart, Human | per tube |
| Insulin, Pork | IV |
| Insulin Glargine Injectable Solution | subQ |
| insulin pork, isophane 100 UNT/ML Injectable Suspension [NPH Iletin II] | sq. |
| Regular Insulin, Human 50 UNT/ML | SEE COMMENT |
| insulin human, isophane 28 UNT/ML | SUBCUT |
| Insulin, Zinc, Human | IM |
| rosiglitazone 8 MG Oral Tablet [Avandia] | Oral |
| saxagliptin 5 MG Oral Tablet [Onglyza] | BY mouth |

| | |
|---|---|
| 60 ACTUAT exenatide 0.005 MG/ACTUAT Pen Injector [Byetta] | subQ |
| alogliptin 12.5 MG [Nesina] | |
| Bromocriptine 5 MG Oral Tablet | |
| Acarbose | PER TUBE |
| Levemir Injectable Product | sub q |
| Glipizide / Metformin Oral Tablet [Metaglip] | oral |
| Levemir Injectable Product | subcutaneous |
| 3 ML Insulin, Aspart Protamine, Human 70 UNT/ML / Insulin, Aspart, Human 30 UNT/ML Pen Injector [NovoLog Mix] | po |
| Glyburide 6 MG Oral Tablet [Glynase] | Po |
| miglitol 50 MG Oral Tablet | po |
| insulin human, isophane 40 UNT/ML | by mouth |
| Invokamet Oral Product | orally |
| Insulin Lispro | subcutaneously. |
| 1.5 ML Insulin Glargine 300 UNT/ML Pen Injector [Toujeo] | Subcutaneous |
| Regular Insulin, Human 100 UNT/ML Injectable Solution [Humulin R] | Subq |
| Glyburide 2.5 MG / Phenformin 25 MG Oral Tablet | p.o. |
| 3 ML insulin degludec 100 UNT/ML Pen Injector [Tresiba] | oral |
| Insulin, Aspart, Human | NG |
| 24 HR dapagliflozin 10 MG / Metformin hydrochloride 500 MG Extended Release Oral Tablet [Xigduo] | oral |
| {60 (Regular Insulin, Human 12 UNT Inhalant Powder [Afrezza]) / 60 (Regular Insulin, Human 4 UNT Inhalant Powder [Afrezza]) / 60 (Regular Insulin, Human 8 UNT Inhalant Powder [Afrezza]) } Pack [Afrezza 180 Titration Pack- 60 (4 UNT), 60 (8 UNT), 60 (12 UN | |
| Metformin | bedside. |
| Chlorpropamide 100 MG Oral Tablet | po |
| Onglyza Pill | |
| Metformin hydrochloride 500 MG / pioglitazone 15 MG Oral Tablet [Actoplus Met] | p.o. |
| Metformin hydrochloride 500 MG / repaglinide 1 MG [PrandiMet] | |
| insulin degludec 200 UNT/ML | orally |
| Regular Insulin, Human 100 UNT/ML Injectable Solution [Novolin R] | PERIDIAL |
| Metformin hydrochloride 1000 MG / sitagliptin 50 MG [Janumet] | swallow |
| exenatide 0.005 MG/ACTUAT [Byetta] | subc |
| glimepiride 1 MG | ng |

| | | |
|---|---|---|
| Tolazamide 100 MG Oral Tablet [Tolinase] | PO | |
| Regular Insulin, Human | inhalation | |
| Insulin, Aspart Protamine, Human 70 UNT/ML / Insulin, Aspart, Human 30 UNT/ML Injectable Suspension [NovoLog Mix] | PO | |
| Insulin Glargine | Subcutaneously | |
| pioglitazone 15 MG [Actos] | per tube | |
| Regular Insulin, Human 100 UNT/ML Injectable Solution [Novolin R] | Subcut | |
| insulin pork, isophane 100 UNT/ML Injectable Suspension [NPH Iletin II] | Drip | |
| repaglinide 0.5 MG Oral Tablet | | 0 |
| Regular Insulin, Human 100 UNT/ML Injectable Solution [Novolin R] | npo. | |
| Chlorpropamide 200 MG / Phenformin 25 MG Oral Tablet | | |
| Insulin Lispro Pen Injector [Humalog] | SUBQ | |
| 3 ML Insulin Glargine 100 UNT/ML Pen Injector [Lantus] | subcut. | |
| Metformin hydrochloride 100 MG/ML Oral Solution [Riomet] | G tube | |
| Tolazamide 100 MG | | |
| glimepiride 2 MG | gt | |
| insulin pork, isophane 80 UNT/ML | subq. | |
| Chlorpropamide Oral Tablet | p.o. | |
| repaglinide Oral Product | Oral | |
| Metformin Oral Solution | topical | |
| 3 ML liraglutide 6 MG/ML Pen Injector [Victoza] | sc | |
| insulin detemir 100 UNT/ML Injectable Solution [Levemir] | by mouth | |
| Insulin, Aspart, Human | TOPICAL | |
| Insulin, Aspart Protamine, Human 70 UNT/ML / Insulin, Aspart, Human 30 UNT/ML [NovoLog Mix] | SQ. | |
| Regular Insulin, Human 5 UNT/ML Injectable Solution | | |
| 1.5 ML insulin human, isophane 100 UNT/ML Prefilled Syringe [Humulin N] | | |
| glimepiride 4 MG / pioglitazone 30 MG Oral Tablet | PO | |
| Regular Insulin, Human 40 UNT/ML | iv | |
| Linagliptin 2.5 MG / Metformin hydrochloride 500 MG Oral Tablet [Jentadueto] | po | |
| dulaglutide 1.5 MG/ML [Trulicity] | subcut | |
| Metformin hydrochloride 500 MG [Glucophage] | NG | |
| Metformin hydrochloride 750 MG Extended Release Oral Tablet [Glucophage] | Oral | |
| Insulin, Aspart, Human | MISCELLANEOUS | |

| | |
|---|---|
| Insulin Lispro 25 UNT/ML / Insulin, Protamine Lispro, Human 75 UNT/ML Injectable Suspension [Humalog Mix] | subq. |
| pramlintide acetate 0.6 MG/ML Injectable Solution [Symlin] | subcut. |
| Metaglip Oral Product | PO |
| 3 ML Insulin Glargine 100 UNT/ML Pen Injector [Lantus] | oral |
| 1.5 ML insulin human, isophane 70 UNT/ML / Regular Insulin, Human 30 UNT/ML Prefilled Syringe | subcutaneously |
| Cycloset Oral Product | PO |
| insulin human, isophane 90 UNT/ML / Regular Insulin, Human 10 UNT/ML [Humulin] | SQ |
| 3 ML insulin degludec 200 UNT/ML Pen Injector [Tresiba] | miscellaneous |
| Insulin, Aspart, Human | ??? |
| repaglinide 1 MG Oral Tablet [Prandin] | Oral |
| glimepiride 2 MG Oral Tablet [Amaryl] | By Mouth |
| Glucophage Pill | PO |
| insulin human, isophane 70 UNT/ML / Regular Insulin, Human 30 UNT/ML [Novolin] | topical |
| 3 ML Insulin, Glulisine, Human 100 UNT/ML Pen Injector [Apidra] | SQ |
| Regular Insulin, Human 25 UNT/ML | SQ |
| Metformin hydrochloride 1000 MG Oral Tablet | ORAL |
| Insulin Glargine 100 UNT/ML [Lantus] | SubCut |
| Glyburide | subq |
| dapagliflozin Oral Tablet | orally |
| 3 ML insulin human, isophane 70 UNT/ML / Regular Insulin, Human 30 UNT/ML Pen Injector | subcut |
| Metformin hydrochloride 500 MG [Glucophage] | SL |
| pramlintide acetate 0.6 MG/ML [Symlin] | Subcutaneous |
| Micronase Pill | po |
| Metformin hydrochloride 1000 MG Oral Tablet [Glucophage] | By mouth |
| Insulin Glargine | SUBCU |
| insulin pork, isophane 100 UNT/ML Injectable Suspension | po |
| Avandaryl Oral Product | by mouth |
| Insulin Lispro | SUBCUTANEOUS |
| miglitol 100 MG Oral Tablet [Glyset] | ORAL |
| insulin pork, isophane 100 UNT/ML Injectable Suspension [NPH Iletin II] | NG |
| Insulin, Aspart, Human 100 UNT/ML Injectable Solution [NovoLog] | injection |
| Glyburide 1.5 MG Oral Tablet [Glynase] | p.o. |
| Metformin Oral Solution | ORAL |

| | |
|---|---|
| Insulin Lispro 100 UNT/ML | SEE COMMENT |
| 24 HR Metformin hydrochloride 1000 MG / pioglitazone 15 MG Extended Release Oral Tablet [Actoplus Met] | oral |
| 24 HR Glipizide 2.5 MG Extended Release Oral Tablet [Glucotrol] | Orally |
| insulin pork, isophane 100 UNT/ML Injectable Suspension [NPH Iletin II] | PO. |
| exenatide | IV |
| Metformin hydrochloride 1000 MG [Glucophage] | by mouth. |
| Metformin hydrochloride 500 MG Oral Tablet | im |
| insulin human, isophane 34 UNT/ML / Regular Insulin, Human 6 UNT/ML Injectable Suspension | subcut |
| insulin detemir 100 UNT/ML Injectable Solution [Levemir] | SUBCUTANEOUSLY |
| Regular Insulin, Human 15 UNT/ML | npo |
| Glyburide 5 MG | by mouth. |
| Insulin Glargine | subc |
| 0.5 ML albiglutide 100 MG/ML Pen Injector [Tanzeum] | sub Q |
| Metformin hydrochloride 850 MG / pioglitazone 15 MG Oral Tablet [Actoplus Met] | p.o. |
| Metformin 1000 MG | NOSTRIL |
| glimepiride | IM |
| Byetta Injectable Product | oral |
| Metformin hydrochloride 500 MG / rosiglitazone 4 MG Oral Tablet | sub q |
| Insulin Lispro 200 UNT/ML [Humalog] | orally |
| Glynase Oral Product | oral |
| Glipizide 5 MG Extended Release Oral Tablet | by mouth |
| exenatide 0.005 MG/ACTUAT [Byetta] | subcutaneously. |
| Byetta Injectable Product | SUBCUTANEOUSLY |
| NovoLog Injectable Product | 0 |
| 3 ML Insulin Glargine 100 UNT/ML Pen Injector [Lantus] | SUBCUT |
| Chlorpropamide 100 MG Oral Tablet | PO |
| exenatide 2 MG [Bydureon] | Im |
| Linagliptin 5 MG [Tradjenta] | per tube |
| exenatide 0.01 MG/ACTUAT [Byetta] | SUBCUTANEOUS |
| Metformin hydrochloride 850 MG Oral Tablet | PER TUBE |
| Regular Insulin, Human 20 UNT/ML | po |
| Regular Insulin, Human | ng |
| Glipizide 10 MG [Glucotrol] | per mouth |
| exenatide 0.01 MG/ACTUAT [Byetta] | subq. |

| | |
|---|---|
| Metformin hydrochloride 1000 MG / rosiglitazone 4 MG [Avandamet] | NPO |
| exenatide 2 MG [Bydureon] | Subq |
| insulin human, isophane 100 UNT/ML Injectable Suspension [Humulin N] | NPO |
| Regular Insulin, Human 10 UNT/ML | orally |
| insulin human, isophane 90 UNT/ML / Regular Insulin, Human 10 UNT/ML [Humulin] | sq |
| Bromocriptine Oral Tablet | p.o. |
| Regular Insulin, Human 30 UNT/ML | orally |
| Insulin Lispro | Subcut. |
| empagliflozin 10 MG / Linagliptin 5 MG Oral Tablet [Glyxambi] | orally |
| 3 ML liraglutide 6 MG/ML Pen Injector | SC |
| 3 ML liraglutide 6 MG/ML Pen Injector [Victoza] | subcu. |
| dulaglutide 1.5 MG/ML [Trulicity] | sq |
| exenatide 0.01 MG/ACTUAT | SUBCUT |
| Apidra Injectable Product | SQ |
| Glipizide 5 MG | NPO |
| sitagliptin 100 MG [Januvia] | nostril |
| exenatide 0.005 MG/ACTUAT Pen Injector | subcut |
| Amaryl Oral Product | |
| Glyburide Oral Tablet [Glynase] | |
| Glipizide 5 MG | subcutaneous |
| glimepiride 1 MG | drip |
| 3 ML Insulin, Aspart, Human 100 UNT/ML Cartridge [NovoLog] | BY MOUTH |
| dapagliflozin 10 MG [Farxiga] | SC |
| saxagliptin Oral Tablet | PO |
| Regular Insulin, Human 50 UNT/ML | by mouth |
| Metformin 500 MG | bedside |
| Janumet Oral Product | BY MOUTH |
| Glipizide | sub Q |
| Insulin Lispro 25 UNT/ML | SubQ |
| Metformin 500 MG | subq |
| Bydureon Injectable Product | IM injection |
| Bromocriptine 0.8 MG | |
| insulin human, isophane 75 UNT/ML | sc |
| Gliclazide 30 MG | |
| Bromocriptine 1 MG [Parlodel] | g - tube |
| 3 ML Insulin Glargine 100 UNT/ML Pen Injector | orally |
| Regular Insulin, Human | subcutaneous. |

| | |
|---|---|
| Insulin Lispro Cartridge [Humalog] | drip |
| insulin detemir Injectable Product | SQ |
| Metformin hydrochloride 500 MG Oral Tablet | td |
| Insulin, Glulisine, Human 100 UNT/ML Injectable Solution | sc |
| insulin human, isophane 16 UNT/ML | sc |
| insulin pork, isophane 100 UNT/ML Injectable Suspension [NPH Iletin II] | MISCELLANEOUS |
| Linagliptin Oral Tablet [Tradjenta] | po |
| dulaglutide 1.5 MG/ML [Trulicity] | oral |
| Insulin Lispro 50 UNT/ML | PO |
| Metformin Oral Solution | inhalation |
| insulin pork, isophane 100 UNT/ML Injectable Suspension [NPH Iletin II] | SUBCUTANEOUS |
| 24 HR Metformin hydrochloride 1000 MG / sitagliptin 100 MG Extended Release Oral Tablet [Janumet] | PO |
| Metformin | NG |
| dulaglutide 3 MG/ML [Trulicity] | by mouth |
| canagliflozin 150 MG / Metformin hydrochloride 500 MG Oral Tablet [Invokamet] | oral |
| 24 HR Linagliptin 2.5 MG / Metformin hydrochloride 1000 MG Extended Release Oral Tablet [Jentadueto] | |
| Glyburide Oral Tablet [Micronase] | PO |
| Glipizide 5 MG Oral Tablet [Minidiab] | |
| sitagliptin 100 MG [Januvia] | subcutaneously |
| Glucotrol Pill | po |
| Regular Insulin, Human 8 UNT | SUBCUT |
| Glipizide | IM |
| Metformin hydrochloride 500 MG / sitagliptin 50 MG Oral Tablet [Janumet] | Oral |
| Glyburide 3 MG Oral Tablet | By Mouth |
| exenatide Injection | Subcutaneous |
| Janumet Oral Product | |
| Insulin Glargine | SQ. |
| glimepiride 6 MG Oral Tablet | Oral |
| Metformin hydrochloride 1000 MG / saxagliptin 5 MG [Kombiglyze] | IV |
| Metformin hydrochloride 500 MG [Glucophage] | drip. |
| pramlintide acetate 1 MG/ML [Symlin] | sc |
| glimepiride 3 MG Oral Tablet [Amaryl] | Po |
| Novolin N Injectable Product | subq |

| | |
|---|---|
| Insulin Glargine 100 UNT/ML Injectable Solution [Lantus] | SUBCUTANEOUSLY |
| Insulin, Glulisine, Human 100 UNT/ML Injectable Solution [Apidra] | Sub Q |
| alogliptin 25 MG Oral Tablet | by mouth |
| Metformin Oral Tablet | sublingual |
| Glyburide 3.5 MG Oral Tablet | by mouth |
| dapagliflozin 10 MG / Metformin hydrochloride 500 MG [Xigduo] | |
| Bromocriptine 5 MG | g - tube |
| 0.5 ML dulaglutide 1.5 MG/ML Auto-Injector [Trulicity] | intramuscular |
| insulin human, isophane 36 UNT/ML | SUBCUT |
| 3 ML Insulin Glargine 100 UNT/ML / Lixisenatide 0.033 MG/ML Pen Injector [Soliqua] | SUBCUTANEOUS |
| glimepiride 1 MG Oral Tablet [Amaryl] | BY MOUTH |
| 3 ML Insulin Lispro 200 UNT/ML Pen Injector [Humalog] | subcutaneously |
| 3 ML Insulin Lispro 200 UNT/ML Pen Injector | SUBQ |
| rosiglitazone 2 MG Oral Tablet [Avandia] | Po |
| Glipizide | subcut |
| Insulin, Aspart, Human | J tube |
| Metformin | MISC |
| Metformin 500 MG | SL |
| Insulin Lispro 50 UNT/ML | IV |
| Metformin hydrochloride 500 MG / rosiglitazone 2 MG Oral Tablet [Avandamet] | oral |
| Duetact Oral Product | PO |
| Metformin hydrochloride 1000 MG Extended Release Oral Tablet | by mouth |
| Actos Oral Product | |
| Metformin | iv. |
| Regular Insulin, Human 5 UNT/ML | po |
| saxagliptin Oral Tablet | orally |
| Insulin, Aspart, Human 100 UNT/ML Cartridge [NovoLog] | PO |
| Metformin Extended Release Oral Tablet [Glucophage] | subcutaneous |
| Metformin hydrochloride 500 MG Extended Release Oral Tablet [Fortamet] | gt |
| Glipizide 10 MG | SQ |
| Exubera Inhalant Product | Inhaled |
| 24 HR Metformin hydrochloride 1000 MG / pioglitazone 30 MG Extended Release Oral Tablet [Actoplus Met] | PO |
| exenatide 2 MG Injection [Bydureon] | subq |
| insulin human, isophane 50 UNT/ML / Regular Insulin, Human 50 UNT/ML Injectable Suspension | Subcutaneous |

| | | |
|---|---|---|
| insulin pork, isophane 100 UNT/ML Injectable Suspension [NPH Iletin II] | SQ. | |
| Regular Insulin, Human 8 UNT | Subcutaneous | |
| Metformin hydrochloride 850 MG Oral Tablet [Glucophage] | per tube | |
| Metformin hydrochloride 500 MG / rosiglitazone 1 MG Oral Tablet [Avandamet] | oral | |
| Insulin, Regular, Pork 40 UNT/ML Injectable Solution | npo | |
| 24 HR Metformin hydrochloride 1000 MG / saxagliptin 5 MG Extended Release Oral Tablet | by mouth | |
| 3 ML Insulin, Aspart Protamine, Human 70 UNT/ML / Insulin, Aspart, Human 30 UNT/ML Pen Injector [NovoLog Mix] | Subq | |
| Insulin Lispro 100 UNT/ML Pen Injector [Humalog] | sub q | |
| Glyburide 5 MG / Metformin hydrochloride 500 MG Oral Tablet [Glucovance] | g - tube | |
| Insulin Lispro 100 UNT/ML Injectable Solution [Humalog] | PER INSULIN PUMP | |
| Linagliptin 2.5 MG / Metformin hydrochloride 500 MG Oral Tablet [Jentadueto] | | |
| Glyburide 3 MG Oral Tablet | | 0 |
| repaglinide 2 MG Oral Tablet [NovoNorm] | by mouth | |
| Metformin hydrochloride 1000 MG / rosiglitazone 2 MG Oral Tablet | oral | |
| Bromocriptine 1 MG [Parlodel] | | |
| glimepiride 1 MG / rosiglitazone 4 MG Oral Tablet [Avandaryl] | | |
| Insulin, Aspart, Human 100 UNT/ML | SC | |
| dulaglutide 3 MG/ML [Trulicity] | subcutaneously | |
| Pramlintide Injectable Solution [Symlin] | subcutaneously | |
| Simvastatin 20 MG / sitagliptin 100 MG Oral Tablet [Juvisync] | orally | |
| Regular Insulin, Human | sub Q | |
| 24 HR empagliflozin 5 MG / Metformin hydrochloride 1000 MG Extended Release Oral Tablet [Synjardy] | oral | |
| 24 HR Metformin hydrochloride 1000 MG / sitagliptin 100 MG Extended Release Oral Tablet [Janumet] | | 0 |
| insulin human, isophane 100 UNT/ML Pen Injector [Humulin N] | subq | |
| insulin degludec 100 UNT/ML Pen Injector [Tresiba] | subcutaneously | |
| glimepiride 1 MG [Amaryl] | gt | |
| 3 ML Insulin Glargine 100 UNT/ML Pen Injector [Lantus] | Subcu | |
| Metformin Extended Release Oral Tablet | p.o. | |
| insulin human, isophane 60 UNT/ML / Regular Insulin, Human 40 UNT/ML [Humulin] | IM | |
| Metformin hydrochloride 850 MG [Glucophage] | ng | |

| | |
|---|---|
| 60 ACTUAT exenatide 0.005 MG/ACTUAT Pen Injector [Byetta] | Sub Q |
| glimepiride 6 MG | subcutaneously |
| Humalog Injectable Product | SUBCUTANEOUSLY |
| insulin pork, isophane 100 UNT/ML Injectable Suspension [NPH Iletin II] | SUBCUTANEOUSLY |
| Regular Insulin, Human 500 UNT/ML Injectable Solution [Humulin R] | orally |
| Insulin, Aspart Protamine, Human 70 UNT/ML / Insulin, Aspart, Human 30 UNT/ML [NovoLog Mix] | Sub Q |
| Glyburide 4.5 MG Oral Tablet | PO |
| insulin pork, isophane 100 UNT/ML Injectable Suspension [NPH Iletin II] | Gtube |
| Glyburide 5 MG / Metformin hydrochloride 500 MG Oral Tablet [Glucovance] | Po |
| Diabinese Oral Product | PO |
| 3 ML Insulin Glargine 100 UNT/ML Pen Injector [Lantus] | SC |
| Glynase Pill | by mouth |
| Insulin Lispro 50 UNT/ML / Insulin, Protamine Lispro, Human 50 UNT/ML Injectable Suspension [Humalog Mix] | orally |
| Insulin Lispro 100 UNT/ML Injectable Solution | subcu |
| Insulin Glargine 100 UNT/ML Injectable Solution | SUBCUTANEOUS |
| Metformin | G - tube |
| Metformin 500 MG / ropinirole 2 MG Oral Tablet | by mouth. |
| Glyburide 2.5 MG | ou |
| Metformin hydrochloride 1000 MG [Glucophage] | IM |
| 3 ML Insulin Glargine 100 UNT/ML Pen Injector [Lantus] | orally |
| 3 ML insulin degludec 100 UNT/ML Pen Injector [Tresiba] | subq |
| 24 HR Linagliptin 5 MG / Metformin hydrochloride 1000 MG Extended Release Oral Tablet [Jentadueto] | |
| dapagliflozin Oral Tablet [Farxiga] | |
| Gliclazide 80 MG Oral Tablet | |
| Metformin hydrochloride 1000 MG / sitagliptin 50 MG Oral Tablet | Po |
| glimepiride 1 MG [Amaryl] | po. |
| insulin human, isophane 70 UNT/ML / Regular Insulin, Human 30 UNT/ML Injectable Suspension [Humulin] | subc |
| Invokana Oral Product | PO |
| insulin human, isophane 40 UNT/ML | subq. |
| dulaglutide 3 MG/ML [Trulicity] | sub q |
| Byetta Injectable Product | SQ |

| | | |
|---|---|---|
| Farxiga Pill | by mouth | |
| empagliflozin 10 MG / Linagliptin 5 MG Oral Tablet [Glyxambi] | PO | |
| Metformin hydrochloride 500 MG Extended Release Oral Tablet [Glumetza] | Oral | |
| Starlix Oral Product | inhalation | |
| 24 HR empagliflozin 25 MG / Metformin hydrochloride 1000 MG Extended Release Oral Tablet [Synjardy] | oral | |
| Glipizide 5 MG [Glucotrol] | g - tube | |
| insulin human, isophane 50 UNT/ML / Regular Insulin, Human 50 UNT/ML [Humulin] | SUBCUT | |
| insulin human, isophane 16 UNT/ML | SUBQ | |
| glimepiride 3 MG | subcutaneously | |
| insulin pork, isophane 100 UNT/ML Injectable Suspension [NPH Iletin II] | p.o | |
| exenatide 0.01 MG/ACTUAT [Byetta] | oral | |
| 0.5 ML dulaglutide 3 MG/ML Auto-Injector | subcutaneous | |
| Acarbose 50 MG | PO | |
| glimepiride 2 MG Oral Tablet | PER TUBE | |
| exenatide 0.005 MG/ACTUAT [Byetta] | SUBCUTANEOUS | |
| Metformin 1000 MG | nares | |
| Insulin Glargine 100 UNT/ML Pen Injector [Lantus] | | |
| Metformin hydrochloride 1000 MG / sitagliptin 100 MG [Janumet] | oral | |
| glimepiride 1 MG [Amaryl] | subcutaneously | |
| Metformin hydrochloride 1000 MG Extended Release Oral Tablet [Glumetza] | p.o. | |
| glimepiride 4 MG | npo | |
| Glyburide 1.25 MG Oral Tablet | P.O. | |
| glimepiride 2 MG | per tube | |
| saxagliptin 2.5 MG Oral Tablet [Onglyza] | | |
| Regular Insulin, Human 100 UNT/ML Injectable Solution | SubQ | |
| Insulin, Zinc, Human | sub q | |
| insulin human, isophane 70 UNT/ML / Regular Insulin, Human 30 UNT/ML [Novolin] | Sub Q | |
| Insulin Glargine Injectable Solution | | 0 |
| glimepiride 2 MG / rosiglitazone 4 MG Oral Tablet | by mouth | |
| Actoplus Met Oral Product | Oral | |
| exenatide 2 MG [Bydureon] | SUBCUTANEOUSLY | |
| Regular Insulin, Human 4 UNT | | |

| | |
|---|---|
| insulin human, isophane 100 UNT/ML Injectable Suspension [Novolin N] | Sub q |
| Insulin Lispro | SQ. |
| 3 ML Regular Insulin, Human 100 UNT/ML Prefilled Syringe [Novolin R] | INTRAVENOUS |
| 3 ML liraglutide 6 MG/ML Pen Injector [Victoza] | Sq |
| sitagliptin 25 MG [Januvia] | SQ. |
| Bromocriptine 10 MG | per tube |
| glimepiride 4 MG | SC |
| Ultralente Insulin, Human 100 UNT/ML Injectable Suspension | Subcutaneous |
| 3 ML Insulin, Aspart, Human 100 UNT/ML Cartridge [NovoLog] | orally |
| Insulin Glargine 100 UNT/ML [Lantus] | PO |
| 3 ML insulin degludec 100 UNT/ML Pen Injector [Tresiba] | miscellaneous |
| Glipizide 5 MG [Glucotrol] | nostril |
| rosiglitazone Oral Tablet [Avandia] | oral |
| Insulin Lispro 100 UNT/ML Injectable Solution [Lispro PRC] | sq |
| Insulin, Glulisine, Human | by mouth |
| Regular Insulin, Human 15 UNT/ML | Sub Q |
| Insulin, Zinc, Pork | SUBCUT |
| Metformin | nares |
| Glyburide 2.5 MG | NPO |
| 24 HR Metformin hydrochloride 500 MG / saxagliptin 5 MG Extended Release Oral Tablet [Kombiglyze] | BY MOUTH |
| liraglutide 6 MG/ML [Saxenda] | subcutaneously |
| glimepiride / rosiglitazone Oral Tablet [Avandaryl] | po |
| Acarbose | G tube. |
| nateglinide Oral Tablet [Starlix] | oral |
| exenatide 0.005 MG/ACTUAT [Byetta] | IM injection |
| Insulin, Aspart, Human | nPO |
| insulin human, isophane 70 UNT/ML / Regular Insulin, Human 30 UNT/ML Injectable Suspension [Humulin] | subcut. |
| Insulin, Regular, Pork 40 UNT/ML Injectable Solution | sc |
| Starlix Oral Product | Oral |
| 24 HR Metformin hydrochloride 1000 MG / pioglitazone 30 MG Extended Release Oral Tablet | |
| repaglinide 0.5 MG Oral Tablet | Oral |
| Regular Insulin, Human 100 UNT/ML Injectable Solution | by mouth |
| 3 ML Insulin Glargine 100 UNT/ML Pen Injector [Lantus] | gt |
| insulin human, isophane 75 UNT/ML | SUBCU |
| Glipizide | drip. |

| | | |
|---|---|---|
| nateglinide | nostril | |
| Insulin Glargine Injectable Solution [Lantus] | SUBCUTANEOUSLY | |
| Lantus Injectable Product | SUBCUTANEOUS | |
| sitagliptin 100 MG [Januvia] | po. | |
| Glipizide 2.5 MG [Glucotrol] | per tube | |
| Victoza Injectable Product | | 0 |
| Metformin hydrochloride 500 MG [Glucophage] | subcutaneous | |
| Bromocriptine 0.8 MG Oral Tablet [Cycloset] | orally | |
| Metformin hydrochloride 500 MG Oral Tablet | swallow | |
| pramlintide acetate 1 MG/ML [Symlin] | subq | |
| Metformin hydrochloride 850 MG / pioglitazone 15 MG Oral Tablet [Actoplus Met] | Oral | |
| Metformin hydrochloride 500 MG [Glumetza] | PO | |
| insulin human, isophane 70 UNT/ML / Regular Insulin, Human 30 UNT/ML [Novolin] | subcutaneous. | |
| dapagliflozin 10 MG Oral Tablet [Farxiga] | BY MOUTH | |
| insulin pork, isophane 100 UNT/ML Injectable Suspension [NPH Iletin II] | subc | |
| Metformin Oral Tablet | NPO | |
| Insulin Glargine | Bedside | |
| Insulin Lispro 100 UNT/ML Injectable Solution [Humalog] | INSULIN PUMP | |
| 1.5 ML Regular Insulin, Human 100 UNT/ML Prefilled Syringe | sq | |
| Regular Insulin, Human | INJECT | |
| Metformin | subq. | |
| Metformin hydrochloride 1000 MG Extended Release Oral Tablet | gt | |
| Farxiga Oral Product | | |
| Glipizide | orally. | |
| Glyburide 6 MG Oral Tablet | | 0 |
| Insulin Lispro 50 UNT/ML | Subcutaneous | |
| liraglutide | SUBCUT | |
| repaglinide 2 MG Oral Tablet | | 0 |
| Insulin Lispro 50 UNT/ML / Insulin, Protamine Lispro, Human 50 UNT/ML Injectable Suspension [Humalog Mix] | oral | |
| Toujeo Injectable Product | oral | |
| Insulin, Aspart, Human 100 UNT/ML Injectable Solution [NovoLog] | intramuscular | |
| Avandia Oral Product | | |
| 3 ML Insulin, Aspart, Human 100 UNT/ML Cartridge [NovoLog] | SubQ | |
| Metformin 1000 MG | swallow | |
| Tolbutamide 500 MG Oral Tablet | p.o. | |

| | |
|---|---|
| exenatide | po |
| canagliflozin 100 MG [Invokana] | subcutaneous |
| Regular Insulin, Human 500 UNT/ML Injectable Solution | po |
| alogliptin 12.5 MG Oral Tablet [Nesina] | oral |
| insulin human, isophane 70 UNT/ML / Regular Insulin, Human 30 UNT/ML Injectable Suspension [Novolin] | sub Q |
| 3 ML Insulin Lispro 25 UNT/ML / Insulin, Protamine Lispro, Human 75 UNT/ML Pen Injector [Humalog Mix] | subq |
| sitagliptin Oral Tablet | p.o. |
| empagliflozin | subcut |
| insulin human, isophane 70 UNT/ML / Regular Insulin, Human 30 UNT/ML [Novolin] | subcu. |
| sitagliptin 25 MG Oral Tablet | Oral |
| insulin human, isophane 36 UNT/ML | sc |
| 24 HR Linagliptin 5 MG / Metformin hydrochloride 1000 MG Extended Release Oral Tablet [Jentadueto] | oral |
| rosiglitazone 4 MG Oral Tablet [Avandia] | Oral |
| Regular Insulin, Human 5 UNT/ML | sub q |
| Acarbose 25 MG Oral Tablet [Precose] | orally |
| insulin detemir 100 UNT/ML Injectable Solution | sq |
| Metformin hydrochloride 1000 MG / sitagliptin 100 MG [Janumet] | subcutaneous |
| 24 HR Metformin hydrochloride 750 MG Extended Release Oral Tablet | p.o |
| Metformin Oral Liquid Product | |
| 24 HR Metformin hydrochloride 500 MG / saxagliptin 5 MG Extended Release Oral Tablet [Kombiglyze] | ORAL |
| repaglinide 1 MG [NovoNorm] | |
| Glyburide | sl |
| Insulin, Aspart, Human | SC. |
| 3 ML Insulin Lispro 100 UNT/ML Cartridge [Humalog] | subcut. |
| canagliflozin 150 MG / Metformin hydrochloride 500 MG Oral Tablet [Invokamet] | orally |
| sitagliptin 50 MG Oral Tablet | Po |
| canagliflozin 150 MG / Metformin hydrochloride 500 MG Oral Tablet [Invokamet] | ORAL |
| Insulin, Zinc, Pork | SC |
| 0.5 ML dulaglutide 1.5 MG/ML Auto-Injector [Trulicity] | subcutaneously |
| Tolbutamide 500 MG Oral Tablet [Orinase] | p.o. |

| | |
|---|---|
| 1.5 ML insulin human, isophane 70 UNT/ML / Regular Insulin, Human 30 UNT/ML Prefilled Syringe | subcu |
| repaglinide Oral Product | |
| Metformin hydrochloride 750 MG Extended Release Oral Tablet | gt |
| 3 ML insulin degludec 200 UNT/ML Pen Injector [Tresiba] | oral |
| insulin human, isophane 36 UNT/ML | SUBCUT. |
| Bromocriptine 5 MG Oral Capsule | PER TUBE |
| Regular Insulin, Human 5 UNT/ML | sc |
| Regular Insulin, Human | Subc |
| glimepiride 2 MG / rosiglitazone 4 MG Oral Tablet | orally |
| Insulin, Glulisine, Human 100 UNT/ML Injectable Solution [Apidra] | CONT SUBCUTAN INFUSI |
| insulin human, isophane 70 UNT/ML / Regular Insulin, Human 30 UNT/ML Injectable Suspension [Novolin] | IM injection |
| Glyburide | subcut |
| Metformin 500 MG | Subq |
| Precose Oral Product | oral |
| Synjardy Oral Product | |
| albiglutide 60 MG/ML | |
| Regular Insulin, Human 12 UNT | Subcutaneous |
| repaglinide Oral Tablet [Prandin] | BY MOUTH |
| canagliflozin Oral Tablet [Invokana] | oral |
| Insulin Lispro 100 UNT/ML Injectable Solution [Humalog] | by mouth |
| Insulin, Zinc, Human 100 UNT/ML Injectable Suspension [Novolin L] | subcutaneous |
| glimepiride 2 MG | IM |
| Metformin 1000 MG | subq |
| Glipizide 5 MG / Metformin hydrochloride 500 MG Oral Tablet | PO |
| Insulin Lispro 50 UNT/ML | drip |
| liraglutide 6 MG/ML | by mouth |
| Humalog Injectable Product | im injection |
| glimepiride | Subq |
| dapagliflozin / Metformin Oral Product | by mouth |
| insulin pork, isophane 100 UNT/ML Injectable Suspension | subcutaneously |
| alogliptin 6.25 MG Oral Tablet [Nesina] | by mouth |
| Insulin Lispro 200 UNT/ML [Humalog] | gt |
| Humulin R Injectable Product | Subcutaneously |
| Insulin Glargine 300 UNT/ML [Toujeo] | SQ |
| dulaglutide 1.5 MG/ML [Trulicity] | intramuscular |
| Glipizide 2.5 MG | IV |

| | | |
|---|---|---|
| insulin detemir 100 UNT/ML [Levemir] | subcutaneous. | |
| Insulin Lispro 25 UNT/ML | IM | |
| Insulin Lispro | G tube | |
| canagliflozin 100 MG Oral Tablet [Invokana] | ORAL | |
| insulin pork, isophane 80 UNT/ML Injectable Suspension | | |
| 0.5 ML albiglutide 60 MG/ML Pen Injector [Tanzeum] | sq | |
| Metformin Oral Tablet | subcutaneously | |
| Regular Insulin, Human | im | |
| dulaglutide 3 MG/ML | subcut | |
| exenatide 0.005 MG/ACTUAT [Byetta] | SUB Q | |
| 3 ML Insulin Lispro 25 UNT/ML / Insulin, Protamine Lispro, Human 75 UNT/ML Pen Injector [Humalog Mix] | | |
| Glipizide | po. | |
| Glipizide Oral Product | Po | |
| Metformin hydrochloride 1000 MG / saxagliptin 2.5 MG Extended Release Oral Tablet [Kombiglyze] | by mouth | |
| exenatide 0.01 MG/ACTUAT [Byetta] | SUBCUTANEOUSLY | |
| glimepiride 4 MG Oral Tablet [Amaryl] | ORALLY | |
| insulin degludec 200 UNT/ML | PO | |
| Metformin 1000 MG | drip | |
| insulin human, isophane 70 UNT/ML / Regular Insulin, Human 30 UNT/ML Injectable Suspension [Humulin] | sc | |
| 0.5 ML albiglutide 100 MG/ML Pen Injector [Tanzeum] | | 0 |
| Insulin Glargine | MISCELLANEOUS | |
| Insulin, Regular, Pork 40 UNT/ML Injectable Solution | NPO | |
| glimepiride 4 MG | nostril | |
| Glyburide | sublingual | |
| insulin human, isophane 70 UNT/ML / Regular Insulin, Human 30 UNT/ML [Humulin] | po. | |
| Humalog Injectable Product | sc | |
| insulin human, isophane 70 UNT/ML / Regular Insulin, Human 30 UNT/ML [Novolin] | IV | |
| 3 ML liraglutide 6 MG/ML Pen Injector [Saxenda] | subcutaneously | |
| Bromocriptine | PER TUBE | |
| Regular Insulin, Human 500 UNT/ML Injectable Solution | Subcutaneous | |
| Insulin Glargine | SUBCUTANEOUSLY | |
| Insulin Lispro | nPO | |
| insulin human, isophane 50 UNT/ML / Regular Insulin, Human 50 UNT/ML Injectable Suspension [Humulin] | sc | |
| insulin human, isophane 28 UNT/ML | sub q | |

| | | |
|---|---|---|
| Insulin Lispro 50 UNT/ML / Insulin, Protamine Lispro, Human 50 UNT/ML Injectable Suspension [Humalog Mix] | po | |
| Bromocriptine 5 MG Oral Capsule | p.o. | |
| Modified 24 HR Metformin hydrochloride 1000 MG Extended Release Oral Tablet [Glumetza] | | 0 |
| Metformin Oral Solution | swallow | |
| Glipizide 2.5 MG / Metformin hydrochloride 500 MG Oral Tablet | | |
| Metformin hydrochloride 500 MG / pioglitazone 15 MG Oral Tablet [Actoplus Met] | Oral | |
| 24 HR Glipizide 5 MG Extended Release Oral Tablet | BY MOUTH | |
| 60 ACTUAT exenatide 0.005 MG/ACTUAT Pen Injector [Byetta] | iv | |
| Novolin Injectable Product | sq | |
| Glucophage Oral Product | Po | |
| exenatide 0.005 MG/ACTUAT | Subcutaneous | |
| Insulin, Aspart, Human | Subcu | |
| Glipizide | td | |
| dulaglutide 3 MG/ML [Trulicity] | SC | |
| Tolazamide 500 MG [Tolinase] | | |
| Insulin Lispro 25 UNT/ML / Insulin, Protamine Lispro, Human 75 UNT/ML [Humalog Mix] | sub Q. | |
| Insulin Lispro 50 UNT/ML | SUBCUTANEOUS | |
| Insulin, Aspart Protamine, Human | orally | |
| 24 HR Metformin hydrochloride 1000 MG / pioglitazone 30 MG Extended Release Oral Tablet [Actoplus Met] | | |
| NovoLog Injectable Product | sub q | |
| insulin detemir | Subcutaneous | |
| insulin human, isophane 40 UNT/ML | Sc | |
| Metformin hydrochloride 850 MG | subcutaneously | |
| Glipizide 2.5 MG [Glucotrol] | nostril | |
| pioglitazone 15 MG Oral Tablet | BY MOUTH | |
| Metformin Extended Release Oral Tablet | orally | |
| canagliflozin 150 MG / Metformin hydrochloride 1000 MG Oral Tablet [Invokamet] | | |
| Regular Insulin, Human 500 UNT/ML Injectable Solution [Humulin R] | oral | |
| dapagliflozin 10 MG / Metformin hydrochloride 500 MG [Xigduo] | by mouth | |
| Insulin, Aspart, Human 100 UNT/ML | subcut | |
| insulin human, isophane 70 UNT/ML / Regular Insulin, Human 30 UNT/ML Injectable Suspension [Humulin] | Sub Q | |
| insulin detemir 100 UNT/ML Injectable Solution | SQ | |

| | | |
|---|---|---|
| Insulin Glargine Injectable Solution [Lantus] | COMBO ROUTE) | |
| glimepiride | td | |
| saxagliptin 5 MG Oral Tablet [Onglyza] | Oral | |
| insulin human, isophane 70 UNT/ML / Regular Insulin, Human 30 UNT/ML [Humulin] | inhalation | |
| Glipizide | PER TUBE | |
| insulin human, isophane 70 UNT/ML / Regular Insulin, Human 30 UNT/ML [Humulin] | MISC.(NON-DRUG; COMBO ROUTE) | |
| insulin human, isophane 90 UNT/ML / Regular Insulin, Human 10 UNT/ML Injectable Suspension | subcutaneously | |
| Glyburide 1.25 MG / Metformin hydrochloride 250 MG Oral Tablet [Glucovance] | | 0 |
| Bromocriptine 10 MG Oral Capsule | by mouth | |
| Insulin Glargine 100 UNT/ML Injectable Solution [Lantus] | sq | |
| 3 ML Regular Insulin, Human 100 UNT/ML Prefilled Syringe [Novolin R] | subcut | |
| Regular Insulin, Human 100 UNT/ML Injectable Solution [Humulin R] | po | |
| Lantus Injectable Product | subc | |
| Ultralente Insulin, Human 100 UNT/ML Injectable Suspension [Humulin U Ultralente] | SC | |
| exenatide 0.01 MG/ACTUAT [Byetta] | SUBCU | |
| Insulin, Glulisine, Human 100 UNT/ML Injectable Solution [Apidra] | VIA INSULIN PUMP | |
| liraglutide 6 MG/ML [Victoza] | sc | |
| Metformin hydrochloride 1000 MG Extended Release Oral Tablet [Fortamet] | BY MOUTH | |
| 24 HR Metformin hydrochloride 1000 MG / pioglitazone 15 MG Extended Release Oral Tablet [Actoplus Met] | by mouth | |
| exenatide 0.01 MG/ACTUAT | PO | |
| insulin human, isophane 60 UNT/ML | Subq | |
| insulin human, isophane 70 UNT/ML / Regular Insulin, Human 30 UNT/ML [Humulin] | sub q. | |
| Kombiglyze Oral Product | orally | |
| pioglitazone | gt | |
| empagliflozin 25 MG / Linagliptin 5 MG Oral Tablet | by mouth | |
| Bromocriptine 0.8 MG Oral Tablet [Cycloset] | | |
| Metformin 500 MG | subcu | |
| insulin human, isophane 70 UNT/ML / Regular Insulin, Human 30 UNT/ML Injectable Suspension | subcut | |

| | | |
|---|---|---|
| pioglitazone | topical | |
| Regular Insulin, Human 40 UNT/ML | orally | |
| Insulin, Aspart, Human | neb | |
| Simvastatin 40 MG / sitagliptin 100 MG Oral Tablet [Juvisync] | by mouth | |
| Insulin, Zinc, Pork 100 UNT/ML Injectable Suspension | SQ | |
| Metformin Extended Release Oral Tablet [Glucophage] | oral | |
| Acarbose Oral Tablet | p.o. | |
| Insulin, Pork | iv | |
| exenatide Injection | oral | |
| insulin human, isophane 70 UNT/ML / Regular Insulin, Human 30 UNT/ML [Humulin] | gt | |
| empagliflozin 10 MG Oral Tablet [Jardiance] | | 0 |
| Metformin hydrochloride 100 MG/ML | IV | |
| insulin human, isophane 34 UNT/ML | SUBQ | |
| {60 (Regular Insulin, Human 12 UNT Inhalant Powder [Afrezza]) / 60 (Regular Insulin, Human 4 UNT Inhalant Powder [Afrezza]) / 60 (Regular Insulin, Human 8 UNT Inhalant Powder [Afrezza]) } Pack [Afrezza 180 Titration Pack- 60 (4 UNT), 60 (8 UNT), 60 (12 UN | inhalation | |
| Humalog Injectable Product | Sub Q | |
| pioglitazone 30 MG [Actos] | by mouth. | |
| Insulin Lispro | Sub Q | |
| Regular Insulin, Human 5 UNT/ML | NPO | |
| canagliflozin | subcutaneously | |
| 0.5 ML dulaglutide 1.5 MG/ML Auto-Injector [Trulicity] | oral | |
| liraglutide 6 MG/ML [Victoza] | | 0 |
| Metformin Extended Release Oral Tablet [Glucophage] | PO | |
| 24 HR Metformin hydrochloride 500 MG Extended Release Oral Tablet [Glucophage] | subcutaneously | |
| Linagliptin 2.5 MG / Metformin hydrochloride 1000 MG Oral Tablet [Jentadueto] | | 0 |
| canagliflozin / Metformin Oral Tablet [Invokamet] | by mouth | |
| insulin human, isophane 50 UNT/ML / Regular Insulin, Human 50 UNT/ML [Humulin] | subcu | |
| Regular Insulin, Human 8 UNT | sub Q | |
| insulin human, isophane 16 UNT/ML | by mouth | |
| Bromocriptine 50 MG | po | |
| glimepiride 8 MG Oral Tablet | Oral | |
| rosiglitazone 4 MG Oral Tablet | BY MOUTH | |
| dapagliflozin 5 MG Oral Tablet | oral | |

| | | |
|---|---|---|
| Regular Insulin, Human Injectable Solution [Humulin R] | subcutaneously | |
| pioglitazone 45 MG [Actos] | per tube | |
| Glipizide 5 MG Oral Tablet | p.o | |
| glimepiride 1 MG [Amaryl] | nostril | |
| Regular Insulin, Human 500 UNT/ML Injectable Solution [Humulin R] | po | |
| Insulin Lispro 50 UNT/ML | NPO | |
| Insulin, Zinc, Human | subq. | |
| Regular Insulin, Human 100 UNT/ML Injectable Solution [Humulin S] | by mouth | |
| Chlorpropamide 250 MG Oral Tablet [Diabinese] | PO | |
| Insulin Glargine 100 UNT/ML [Lantus] | drip. | |
| 3 ML liraglutide 6 MG/ML Pen Injector [Victoza] | PO | |
| empagliflozin 10 MG Oral Tablet | po | |
| insulin degludec 200 UNT/ML | po | |
| Regular Insulin, Human 20 UNT/ML | orally | |
| Glyburide 1.5 MG Oral Tablet [Glynase] | | |
| glimepiride | NPO. | |
| 3 ML insulin degludec 200 UNT/ML Pen Injector [Tresiba] | subq | |
| alogliptin 12.5 MG / pioglitazone 45 MG [Oseni] | | |
| Insulin Lispro 50 UNT/ML | Subq | |
| 3 ML insulin degludec 200 UNT/ML Pen Injector [Tresiba] | | 0 |
| Trulicity Injectable Product | by mouth | |
| 24 HR dapagliflozin 5 MG / Metformin hydrochloride 1000 MG Extended Release Oral Tablet [Xigduo] | po | |
| Insulin Lispro Cartridge [Humalog] | subq. | |
| Metformin | subcut. | |
| Glyburide | SubQ | |
| insulin human, isophane 30 UNT/ML / Regular Insulin, Human 10 UNT/ML Injectable Suspension | subcutaneously | |
| Regular Insulin, Human 500 UNT/ML Injectable Solution [Humulin R] | subcu | |
| rosiglitazone 2 MG Oral Tablet | oral | |
| Insulin Glargine Injectable Solution | IV | |
| 60 ACTUAT exenatide 0.01 MG/ACTUAT Pen Injector [Byetta] | subc. | |
| exenatide | sub q | |
| Glipizide 5 MG | TOPICAL | |
| insulin pork, isophane 100 UNT/ML Injectable Suspension [NPH Iletin II] | trans | |

| | | |
|---|---|---|
| Regular Insulin, Human 500 UNT/ML Injectable Solution [Humulin R] | gt | |
| Insulin Lispro 200 UNT/ML | po | |
| Insulin, Aspart, Human 100 UNT/ML Injectable Solution [NovoLog] | oral | |
| Regular Insulin, Human 8 UNT | by mouth | |
| Insulin, Zinc, Pork | Subcutaneous | |
| Regular Insulin, Human 500 UNT/ML Injectable Solution | orally | |
| repaglinide 1 MG Oral Tablet [Prandin] | | 0 |
| Insulin, Aspart, Human 100 UNT/ML | sq | |
| Invokana Pill | oral | |
| exenatide 0.005 MG/ACTUAT [Byetta] | oral | |
| exenatide 0.005 MG/ACTUAT | subcutaneously | |
| insulin human, isophane 28 UNT/ML | subq. | |
| Regular Insulin, Human | IV. | |
| Insulin Lispro 100 UNT/ML Pen Injector [Humalog] | subQ | |
| Glipizide | SL | |
| Metformin hydrochloride 250 MG | po | |
| 3 ML Insulin, Aspart, Human 100 UNT/ML Pen Injector [NovoLog] | SUBCUTANEOUSLY | |
| glimepiride | sub Q | |
| 3 ML liraglutide 6 MG/ML Pen Injector [Victoza] | SEE COMMENT | |
| 3 ML Regular Insulin, Human 100 UNT/ML Prefilled Syringe | IV | |
| Glipizide | nare | |
| insulin pork, isophane 80 UNT/ML | p.o. | |
| Regular Insulin, Human 500 UNT/ML Injectable Solution | subq | |
| insulin human, isophane 75 UNT/ML | sl | |
| miglitol 50 MG | | |
| insulin human, isophane 70 UNT/ML / Regular Insulin, Human 30 UNT/ML [Humulin] | nostril | |
| Metformin hydrochloride 500 MG Extended Release Oral Tablet [Fortamet] | p.o. | |
| Insulin Lispro 100 UNT/ML Pen Injector [Humalog] | SUBQ | |
| Insulin Lispro 100 UNT/ML Injectable Solution [Humalog] | SC | |
| Regular Insulin, Human 40 UNT/ML | sq | |
| Metforming Oral Product | oral | |
| liraglutide | Subcut | |
| Glipizide 2.5 MG | IM | |
| insulin human, isophane 60 UNT/ML | po | |
| Metformin | IV. | |
| Regular Insulin, Human 20 UNT/ML | sub q | |

| | | |
|---|---|---|
| Insulin Lispro Cartridge [Humalog] | IM | |
| Metformin hydrochloride 100 MG/ML Oral Solution | BY MOUTH | |
| Starlix Oral Product | | |
| 24 HR Metformin hydrochloride 750 MG Extended Release Oral Tablet | ORAL | |
| sitagliptin 100 MG Oral Tablet [Januvia] | P.O. | |
| empagliflozin Oral Tablet [Jardiance] | by mouth | |
| insulin human, isophane 70 UNT/ML / Regular Insulin, Human 30 UNT/ML [Novolin] | sQ | |
| glimepiride 4 MG | subcutaneous | |
| glimepiride Oral Product | BY MOUTH | |
| sitagliptin | IV | |
| 3 ML Insulin Glargine 100 UNT/ML Pen Injector [Lantus] | per tube | |
| 3 ML insulin degludec 200 UNT/ML Pen Injector [Tresiba] | intramuscular | |
| Glipizide 10 MG | bedside | |
| 3 ML insulin degludec 200 UNT/ML Pen Injector [Tresiba] | subconjunctival | |
| 3 ML liraglutide 6 MG/ML Pen Injector [Victoza] | subcutaneous. | |
| Symlin Injectable Product | COMBO ROUTE) | |
| 3 ML Insulin, Aspart Protamine, Human 70 UNT/ML / Insulin, Aspart, Human 30 UNT/ML Pen Injector [NovoLog Mix] | drip | |
| Glyburide 1.75 MG | | |
| 24 HR Metformin hydrochloride 1000 MG / sitagliptin 100 MG Extended Release Oral Tablet [Janumet] | BY MOUTH | |
| Insulin Glargine 100 UNT/ML [Lantus] | inhalation | |
| Glyburide | td | |
| 24 HR Linagliptin 2.5 MG / Metformin hydrochloride 1000 MG Extended Release Oral Tablet [Jentadueto] | oral | |
| Metformin hydrochloride 750 MG | | 0 |
| Insulin Lispro 25 UNT/ML | SUBCUTANEOUS | |
| 3 ML insulin degludec 100 UNT/ML Pen Injector [Tresiba] | subcutaneous | |
| Linagliptin Oral Tablet [Tradjenta] | | |
| Metformin 1000 MG | OU | |
| 24 HR Metformin hydrochloride 1000 MG / pioglitazone 30 MG Extended Release Oral Tablet [Actoplus Met] | po | |
| Regular Insulin, Human | SUBCUTANEOUS | |
| 24 HR Bromocriptine 2.5 MG Extended Release Oral Capsule | by mouth | |
| Bromocriptine Oral Tablet [Parlodel] | | |
| Insulin Lispro 25 UNT/ML | Oral | |
| Metformin / pioglitazone Extended Release Oral Tablet [Actoplus Met] | by mouth | |

| | | |
|---|---|---|
| Insulin Glargine Injectable Solution | NPO | |
| Insulin, Aspart, Human 100 UNT/ML Injectable Solution [NovoLog] | subc | |
| Metformin 500 MG | PER TUBE | |
| 3 ML Insulin Lispro 50 UNT/ML / Insulin, Protamine Lispro, Human 50 UNT/ML Pen Injector [Humalog Mix] | | 0 |
| Insulin Lispro 100 UNT/ML | subcut | |
| glimepiride 3 MG Oral Tablet [Amaryl] | BY MOUTH | |
| Acarbose 25 MG Oral Tablet | BY MOUTH | |
| glimepiride 2 MG / pioglitazone 30 MG Oral Tablet | | |
| Insulin, Aspart, Human | inhalatn | |
| rosiglitazone Oral Product | oral | |
| Metformin hydrochloride 500 MG / pioglitazone 15 MG Oral Tablet [Actoplus Met] | per tube | |
| Bromocriptine 2.5 MG Oral Tablet [Parlodel] | BY MOUTH | |
| alogliptin 12.5 MG / Metformin hydrochloride 1000 MG Oral Tablet [Kazano] | | |
| Bromocriptine 10 MG Oral Capsule | p.o. | |
| sitagliptin 100 MG [Januvia] | per tube | |
| exenatide 2 MG Injection [Bydureon] | SQ | |
| 24 HR Glipizide 10 MG Extended Release Oral Tablet [Glucotrol] | orally | |
| Glipizide 5 MG Oral Tablet | By Mouth | |
| Metformin 500 MG | NPO | |
| Glipizide / Metformin Oral Tablet | PO | |
| Insulin Lispro 100 UNT/ML Injectable Solution [Lispro PRC] | subcutaneously | |
| Glyburide 5 MG Oral Tablet [Daonil] | oral | |
| insulin pork, isophane 100 UNT/ML Injectable Suspension [NPH Iletin II] | Inh. | |
| 3 ML Insulin Lispro 25 UNT/ML / Insulin, Protamine Lispro, Human 75 UNT/ML Pen Injector [Humalog Mix] | injection | |
| {90 (Regular Insulin, Human 4 UNT Inhalant Powder [Afrezza]) / 90 (Regular Insulin, Human 8 UNT Inhalant Powder [Afrezza]) } Pack [Afrezza Titration Pack] | | |
| nateglinide | iv | |
| Regular Insulin, Human 5 UNT/ML | by mouth | |
| 0.5 ML albiglutide 100 MG/ML Pen Injector [Tanzeum] | SC | |
| Insulin Glargine Injectable Solution | subc | |
| Metformin hydrochloride 1000 MG Extended Release Oral Tablet [Fortamet] | subcutaneously | |

| | |
|---|---|
| Modified 24 HR Metformin hydrochloride 1000 MG Extended Release Oral Tablet [Glumetza] | by mouth |
| Bromocriptine 25 MG Oral Tablet | by mouth |
| Gliclazide | po |
| Insulin Lispro 25 UNT/ML / Insulin, Protamine Lispro, Human 75 UNT/ML Injectable Suspension [Humalog Mix] | oral |
| Regular Insulin, Human 15 UNT/ML | by mouth |
| Metaglip Pill | |
| canagliflozin Oral Tablet | oral |
| glimepiride 2 MG [Amaryl] | gt |
| exenatide 0.01 MG/ACTUAT [Byetta] | SEE COMMENT |
| Glyset Oral Product | p.o. |
| Insulin, Aspart, Human | COMBO ROUTE) |
| Regular Insulin, Human 20 UNT/ML | by mouth |
| insulin detemir | SUBCUT |
| Humalog Injectable Product | MISCELLANEOUS |
| insulin human, isophane 40 UNT/ML | NPO |
| insulin human, isophane 60 UNT/ML / Regular Insulin, Human 40 UNT/ML Injectable Suspension [Humulin] | |
| glimepiride | subcut |
| Insulin, Glulisine, Human 100 UNT/ML Injectable Solution [Apidra] | SUBQ BY INSULIN PUMP |
| sitagliptin 50 MG [Januvia] | nostril |
| Bromocriptine 5 MG Oral Capsule [Parlodel] | p.o. |
| glimepiride | Im |
| exenatide 2 MG Injection | |
| Glyburide | IM |
| Acarbose 100 MG Oral Tablet [Precose] | |
| insulin pork, isophane 100 UNT/ML Injectable Suspension [NPH Iletin II] | Oral |
| Insulin Lispro | NPo |
| Metformin Oral Solution | nostril |
| Bromocriptine | IV |
| Metformin / pioglitazone Oral Tablet | Oral |
| glimepiride 2 MG | NPO |
| Regular Insulin, Human 100 UNT/ML Injectable Solution [Humulin R] | Sub q |
| Victoza Injectable Product | sq |
| 60 ACTUAT exenatide 0.005 MG/ACTUAT Pen Injector [Byetta] | Subcut. |
| insulin human, isophane 100 UNT/ML [Humulin N] | subcut. |

| | | |
|---|---|---|
| insulin pork, isophane 100 UNT/ML Injectable Suspension [NPH Iletin II] | Subc | |
| Metformin 500 MG | nares | |
| Metformin hydrochloride 500 MG / rosiglitazone 2 MG [Avandamet] | inhaled | |
| Bromocriptine 2.5 MG Oral Tablet [Parlodel] | INTRAVAGINAL | |
| Glipizide 2.5 MG Extended Release Oral Tablet | BY MOUTH | |
| Glipizide 5 MG | inhalation | |
| rosiglitazone 4 MG Oral Tablet [Avandia] | BY MOUTH | |
| 0.5 ML albiglutide 100 MG/ML Pen Injector [Tanzeum] | po | |
| [Exubera]) / 90 (insulin human, rDNA origin 3 MG/ACTUAT Inhalant Powder [Exubera]) } Pack [Exubera Combination Pack 12] | | |
| insulin, regular, beef-pork 40 UNT/ML Inhalant Solution | Subcutaneously | |
| Metformin hydrochloride 1000 MG / sitagliptin 100 MG Extended Release Oral Tablet [Janumet] | by mouth | |
| Acarbose 100 MG Oral Tablet | PO. | |
| 3 ML Insulin, Aspart Protamine, Human 70 UNT/ML / Insulin, Aspart, Human 30 UNT/ML Pen Injector [NovoLog Mix] | Sub q | |
| insulin human, isophane 40 UNT/ML | po | |
| dulaglutide 1.5 MG/ML [Trulicity] | Sub q | |
| Insulin Lispro 200 UNT/ML | by mouth | |
| Glyburide 1.25 MG Oral Tablet [Micronase] | p.o. | |
| Metformin | SubQ | |
| Regular Insulin, Human 6 UNT/ML | Subcutaneous | |
| insulin human, isophane 100 UNT/ML Pen Injector [Humulin N] | by mouth | |
| rosiglitazone 8 MG Oral Tablet [Avandia] | P.o | |
| Bromocriptine 2.5 MG Oral Tablet | BY MOUTH | |
| insulin human, isophane 70 UNT/ML | subcutaneous. | |
| Acarbose 100 MG Oral Tablet [Precose] | | 0 |
| Bromocriptine | topical | |
| pioglitazone | sublingual | |
| Insulin, Aspart, Human | SUBQ | |
| Metformin hydrochloride 1000 MG / rosiglitazone 2 MG Oral Tablet [Avandamet] | | |
| Insulin Lispro Injectable Solution | npo | |
| insulin human, isophane 70 UNT/ML / Regular Insulin, Human 30 UNT/ML Injectable Suspension | by mouth | |
| Insulin Lispro Injectable Solution | iv | |
| Regular Insulin, Human Injectable Product | subcutaneously | |

| | | |
|---|---|---|
| exenatide 2 MG Injection | subcut | |
| 3 ML Insulin Glargine 100 UNT/ML Pen Injector [Lantus] | IM | |
| Insulin Lispro | nare | |
| glimepiride 3 MG Oral Tablet | Oral | |
| Regular Insulin, Human 8 UNT | IV | |
| Glyburide | inhalation | |
| Insulin, Aspart, Human | Po | |
| Synjardy Oral Product | orally | |
| insulin pork, isophane 100 UNT/ML Injectable Suspension [NPH Iletin II] | Im | |
| 24 HR Metformin hydrochloride 1000 MG / saxagliptin 5 MG Extended Release Oral Tablet [Kombiglyze] | | 0 |
| Glipizide | PO. | |
| Metformin | nare | |
| Janumet Oral Product | Oral | |
| Glyburide 5 MG / Metformin hydrochloride 500 MG Oral Tablet | Oral | |
| Regular Insulin, Human 5 UNT/ML | per tube | |
| saxagliptin 2.5 MG Oral Tablet | p.o. | |
| pramlintide acetate 0.6 MG/ML Injectable Solution [Symlin] | by mouth | |
| Insulin Glargine 300 UNT/ML Pen Injector [Toujeo] | po | |
| Metformin 1000 MG | inhaled | |
| sitagliptin | inh | |
| Linagliptin 2.5 MG / Metformin hydrochloride 1000 MG Oral Tablet [Jentadueto] | BY MOUTH | |
| glimepiride | npo | |
| Bromocriptine Pill | by mouth | |
| Metformin | IM. | |
| pioglitazone | PER TUBE | |
| Metformin | oral. | |
| 60 ACTUAT exenatide 0.005 MG/ACTUAT Pen Injector [Byetta] | oral | |
| pioglitazone 30 MG [Actos] | SUBCUT | |
| Metformin 1000 MG | | 0 |
| Glynase Oral Product | BY MOUTH | |
| Glyburide 5 MG Oral Tablet [Diabeta] | BY MOUTH | |
| Metformin Oral Tablet | swallow. | |
| Insulin Lispro Cartridge [Humalog] | subCUTANEOUS | |
| insulin human, isophane 70 UNT/ML / Regular Insulin, Human 30 UNT/ML Injectable Suspension [Novolin] | subq | |
| Insulin Lispro | swallow | |
| Metformin 500 MG | im | |

| | |
|---|---|
| Insulin Lispro | GTube |
| Insulin, Zinc, Human 100 UNT/ML Injectable Suspension [Humulin L] | |
| insulin human, isophane 90 UNT/ML / Regular Insulin, Human 10 UNT/ML [Human Mixtard] | sl |
| Insulin, Aspart, Human 100 UNT/ML Cartridge [NovoLog] | nostril |
| dulaglutide Injectable Product | subcutaneously |
| Cycloset Pill | |
| glimepiride 2 MG / pioglitazone 30 MG Oral Tablet | po |
| Insulin Glargine 300 UNT/ML | p.o. |
| insulin human, isophane 70 UNT/ML | by mouth |
| Metformin hydrochloride 1000 MG / saxagliptin 5 MG [Kombiglyze] | TOPICALLY |
| Regular Insulin, Human 500 UNT/ML Injectable Solution | per tube |
| insulin human, isophane 30 UNT/ML / Regular Insulin, Human 10 UNT/ML Injectable Suspension | subcut. |
| sitagliptin Pill | |
| pioglitazone 30 MG | nostril |
| Metformin hydrochloride 500 MG / pioglitazone 15 MG Oral Tablet | by mouth. |
| 3 ML Insulin Lispro 200 UNT/ML Pen Injector [Humalog] | miscellaneous |
| Prandin Oral Product | BY MOUTH |
| Regular Insulin, Human 100 UNT/ML Injectable Solution | INJECTION |
| Insulin Glargine | nares |
| Metformin | trans |
| glimepiride 1 MG Oral Tablet [Amaryl] | ORALLY |
| exenatide 0.005 MG/ACTUAT | subcu |
| exenatide 0.01 MG/ACTUAT | SubQ |
| Insulin Glargine Injectable Solution | PO |
| Linagliptin 2.5 MG / Metformin hydrochloride 850 MG Oral Tablet [Jentadueto] | ORALLY |
| repaglinide 0.5 MG Oral Tablet | p.o. |
| alogliptin 12.5 MG / Metformin hydrochloride 1000 MG Oral Tablet [Kazano] | ORAL |
| Actos Oral Product | ORAL |
| insulin human, isophane 70 UNT/ML / Regular Insulin, Human 30 UNT/ML Injectable Suspension [Humulin] | Subcutaneously |
| Metformin | per rectum |
| 24 HR dapagliflozin 5 MG / Metformin hydrochloride 1000 MG Extended Release Oral Tablet [Xigduo] | orally |

| | |
|---|---|
| Metformin Extended Release Oral Tablet [Fortamet] | by mouth. |
| Nesina Oral Product | orally |
| pioglitazone | npo |
| Insulin Lispro 100 UNT/ML Injectable Solution | SQ |
| Tolbutamide 500 MG Oral Tablet | by mouth |
| Acarbose 25 MG Oral Tablet [Precose] | PER G-TUBE |
| glimepiride 2 MG | SC |
| dulaglutide | subQ |
| Tolinase Oral Product | p.o. |
| insulin, regular, beef-pork 40 UNT/ML Inhalant Solution | SUBCUTANEOUSLY |
| NovoLog Injectable Product | COMBO ROUTE) |
| Metformin | G tube |
| glimepiride 4 MG Oral Tablet [Amaryl] | P.o. |
| Regular Insulin, Human 6 UNT/ML | inhalatn |
| Metformin hydrochloride 850 MG Oral Tablet | Po |
| Glumetza Oral Product | orally |
| Acarbose | SEE COMMENT |
| Regular Insulin, Human 100 UNT/ML Injectable Solution [Novolin R] | orally |
| Levemir Injectable Product | MISCELLANEOUS |
| Insulin, Aspart, Human 100 UNT/ML Cartridge [NovoLog] | SEE COMMENT |
| Lente Insulin, Beef-Pork 40 UNT/ML | subcu |
| Linagliptin Oral Tablet | by mouth |
| insulin pork, isophane 100 UNT/ML Injectable Suspension [NPH Iletin II] | G tube |
| Pramlintide Injectable Solution [Symlin] | SQ |
| 3 ML Insulin Glargine 100 UNT/ML Pen Injector [Lantus] | npo. |
| insulin human, isophane 75 UNT/ML | drip |
| 3 ML insulin detemir 100 UNT/ML Pen Injector [Levemir] | orally |
| liraglutide Injectable Product | SQ |
| empagliflozin 12.5 MG / Metformin hydrochloride 1000 MG Oral Tablet [Synjardy] | po |
| Glipizide 2.5 MG | po. |
| Glipizide | Im |
| Linagliptin 2.5 MG / Metformin hydrochloride 850 MG Oral Tablet [Jentadueto] | ORAL |
| Linagliptin 5 MG | per tube |
| Glyburide | ou |
| dulaglutide Auto-Injector [Trulicity] | by mouth |

| | |
|---|---|
| 3 ML Insulin, Aspart Protamine, Human 70 UNT/ML / Insulin, Aspart, Human 30 UNT/ML Pen Injector [NovoLog Mix] | PO |
| alogliptin 25 MG Oral Tablet [Nesina] | |
| 3 ML insulin detemir 100 UNT/ML Pen Injector [Levemir] | intramuscular |
| Glipizide | OU |
| insulin human, isophane 70 UNT/ML / Regular Insulin, Human 30 UNT/ML [Novolin] | PO |
| dapagliflozin 5 MG Oral Tablet [Farxiga] | BY MOUTH |
| Invokana Oral Product | PO. |
| Insulin, Aspart, Human | IV. |
| Glipizide 2.5 MG Extended Release Oral Tablet [Glucotrol] | po |
| insulin human, isophane 90 UNT/ML / Regular Insulin, Human 10 UNT/ML [Human Mixtard] | subcut |
| Metformin hydrochloride 1000 MG Oral Tablet | subcutaneous |
| Glucophage Oral Product | inhalation |
| Insulin Glargine 100 UNT/ML Injectable Solution [Lantus] | subCUTANEOUS |
| albiglutide | |
| Insulin Glargine 100 UNT/ML Injectable Solution [Lantus] | Subcu |
| exenatide 2 MG Injection [Bydureon] | SUBCUT |
| insulin pork, isophane 100 UNT/ML Injectable Suspension [NPH Iletin II] | gtube |
| Glyburide 5 MG | OU |
| Metformin hydrochloride 850 MG Extended Release Oral Tablet | by mouth |
| Regular Insulin, Human 30 UNT/ML | SUBCUT |
| repaglinide 0.5 MG Oral Tablet [Prandin] | Oral |
| Insulin Lispro | sl |
| Micronase Oral Product | oral |
| Insulin Glargine Injectable Solution [Lantus] | SC |
| insulin human, isophane 50 UNT/ML / Regular Insulin, Human 50 UNT/ML Injectable Suspension [Humulin] | p.o. |
| Metformin 500 MG | per gt |
| insulin detemir 100 UNT/ML Injectable Solution [Levemir] | Subcut. |
| Metformin hydrochloride 500 MG [Glucophage] | topical |
| 0.5 ML albiglutide 100 MG/ML Pen Injector [Tanzeum] | |
| Tolbutamide Oral Product | p.o. |
| 24 HR Metformin hydrochloride 1000 MG / saxagliptin 2.5 MG Extended Release Oral Tablet [Kombiglyze] | |
| 3 ML insulin human, isophane 70 UNT/ML / Regular Insulin, Human 30 UNT/ML Pen Injector [Humulin] | SubQ |
| Invokana Pill | swallow |

| | | |
|---|---|---|
| 0.5 ML albiglutide 60 MG/ML Pen Injector [Tanzeum] | subcut | |
| pioglitazone Pill | | |
| rosiglitazone 4 MG Oral Tablet | PER TUBE | |
| Byetta Injectable Product | sub q | |
| 3 ML liraglutide 6 MG/ML Pen Injector [Saxenda] | IM | |
| 3 ML insulin human, isophane 100 UNT/ML Prefilled Syringe [Novolin N] | Subcut | |
| Glipizide Oral Tablet | p.o. | |
| Metformin | ng. | |
| Metformin 1000 MG | po. | |
| Regular Insulin, Human 24 UNT/ML | sq | |
| Insulin Glargine 300 UNT/ML [Toujeo] | | |
| Insulin Glargine 300 UNT/ML Pen Injector | | |
| NovoLog Injectable Product | subcut. | |
| rosiglitazone 4 MG Oral Tablet [Avandia] | | 0 |
| NovoLog Injectable Product | SubQ | |
| rosiglitazone 8 MG [Avandia] | per tube | |
| saxagliptin 2.5 MG Oral Tablet | orally | |
| canagliflozin 300 MG Oral Tablet | PO | |
| Regular Insulin, Human 20 UNT/ML | oral | |
| Insulin, Aspart, Human | SQ. | |
| insulin human, isophane 32 UNT/ML / Regular Insulin, Human 8 UNT/ML Injectable Suspension | | |
| glimepiride 2 MG [Amaryl] | subcutaneously | |
| Glipizide 2.5 MG Oral Tablet | ORAL | |
| insulin human, isophane 40 UNT/ML Injectable Suspension | subcutaneously | |
| empagliflozin 12.5 MG / Metformin hydrochloride 1000 MG Oral Tablet [Synjardy] | PO | |
| Glipizide 5 MG | by mouth. | |
| glimepiride 4 MG Oral Tablet | Po | |
| Glipizide 10 MG [Glucotrol] | nostril | |
| 3 ML Insulin, Aspart Protamine, Human 70 UNT/ML / Insulin, Aspart, Human 30 UNT/ML Pen Injector [NovoLog Mix] | orally | |
| Insulin Glargine 100 UNT/ML [Lantus] | IV | |
| Insulin, Aspart, Human | IM injection | |
| glimepiride 4 MG [Amaryl] | subcutaneous | |
| insulin human, isophane 70 UNT/ML / Regular Insulin, Human 30 UNT/ML [Novolin] | Oral | |
| Insulin Lispro 100 UNT/ML [Humalog] | Im | |
| Glyburide | COMBO ROUTE) | |

| | |
|---|---|
| Jentadueto Pill | |
| Metformin hydrochloride 1000 MG / saxagliptin 2.5 MG Extended Release Oral Tablet [Kombiglyze] | |
| pioglitazone 45 MG | per tube |
| Insulin, Aspart, Human 100 UNT/ML Injectable Solution [NovoLog] | SUBCUTANEOUS VIA INSULIN PUMP |
| exenatide 0.005 MG/ACTUAT | SUBCUT |
| Metformin hydrochloride 1000 MG Oral Tablet | per tube |
| Glipizide 5 MG | topically |
| Glipizide 2.5 MG / Metformin hydrochloride 500 MG Oral Tablet | p.o. |
| glimepiride 1 MG | subcutaneously |
| Metformin hydrochloride 500 MG Oral Tablet | PEG |
| insulin human, isophane 70 UNT/ML / Regular Insulin, Human 30 UNT/ML [Novolin] | drip. |
| Regular Insulin, Human 100 UNT/ML Injectable Solution [Humulin R] | PERIDIAL |
| Glyburide 5 MG | IV |
| 60 ACTUAT exenatide 0.01 MG/ACTUAT Pen Injector [Byetta] | subcu. |
| repaglinide | NPO |
| Glipizide 10 MG Oral Tablet | PER TUBE |
| Glyburide | subcu. |
| dapagliflozin 5 MG Oral Tablet | PO |
| 1.5 ML pramlintide acetate 1 MG/ML Pen Injector [Symlin] | |
| Glipizide Oral Product | p.o |
| Apidra Injectable Product | by mouth |
| Insulin, Zinc, Human 100 UNT/ML Injectable Suspension [Humulin L] | SUBCUTANEOUS |
| pioglitazone 45 MG [Actos] | per mouth |
| Lantus Injectable Product | SUB CONJ |
| Insulin Lispro 50 UNT/ML | subcutaneously |
| Glipizide 10 MG [Glucotrol] | SL |
| Metformin | Npo |
| Glipizide 10 MG Oral Tablet | ORAL |
| 3 ML Insulin Glargine 100 UNT/ML Pen Injector [Lantus] | drip. |
| Linagliptin 2.5 MG / Metformin hydrochloride 850 MG Oral Tablet | by mouth |
| Tolazamide 250 MG Oral Tablet [Tolinase] | PO |
| rosiglitazone 8 MG [Avandia] | per mouth |
| Metformin hydrochloride 100 MG/ML Oral Solution [Riomet] | tube |
| Glipizide Oral Product | ORALLY |

| | | |
|---|---|---|
| Metformin Extended Release Oral Tablet | swallow | |
| Metformin Oral Tablet [Metforming] | | |
| Glyburide 5 MG | subcutaneously | |
| Chlorpropamide 250 MG Oral Tablet | ORAL | |
| insulin human, isophane 28 UNT/ML | IV | |
| 3 ML liraglutide 6 MG/ML Pen Injector [Victoza] | oral | |
| Glyburide | | 0 |
| repaglinide 1 MG Oral Tablet [Prandin] | BY MOUTH | |
| Bromocriptine | po. | |
| Insulin, Zinc, Human 100 UNT/ML Injectable Suspension [Human Ultratard] | po | |
| Gliclazide 80 MG Oral Tablet | by mouth | |
| Metforming Pill | | |
| empagliflozin 25 MG Oral Tablet [Jardiance] | p.o. | |
| glimepiride Oral Tablet [Amaryl] | p.o. | |
| Insulin, Pork | | |
| Insulin Lispro 50 UNT/ML | per tube | |
| Bromocriptine Oral Tablet [Parlodel] | BY MOUTH | |
| Acarbose 50 MG Oral Tablet | Oral | |
| Glyxambi Oral Product | PO | |
| Regular Insulin, Human 100 UNT/ML Injectable Solution | INTRAVENOUS | |
| glimepiride | ng | |
| Regular Insulin, Human 12 UNT | sub Q | |
| Insulin Lispro | MISCELLANEOUS | |
| 3 ML insulin degludec 200 UNT/ML Pen Injector [Tresiba] | subcutaneous | |
| insulin human, isophane 34 UNT/ML | sc | |
| Glipizide 10 MG [Glucotrol] | gt | |
| insulin human, isophane 16 UNT/ML | subq. | |
| insulin human, isophane 70 UNT/ML / Regular Insulin, Human 30 UNT/ML Injectable Suspension | | |
| Metformin 500 MG | ng | |
| insulin beef, isophane 40 UNT/ML | by mouth | |
| glimepiride 1 MG | gt | |
| Glipizide 2.5 MG / Metformin hydrochloride 500 MG Oral Tablet [Metaglip] | po | |
| insulin pork, isophane 100 UNT/ML Injectable Suspension [NPH Iletin II] | Subq | |
| Glyset Oral Product | by mouth | |
| Insulin Glargine 100 UNT/ML Injectable Solution [Lantus] | MISCELLANEOUS | |

| | | |
|---|---|---|
| Insulin, Aspart Protamine, Human 70 UNT/ML / Insulin, Aspart, Human 30 UNT/ML Injectable Suspension [NovoLog Mix] | sq. | |
| insulin pork, isophane 100 UNT/ML Injectable Suspension [NPH Iletin II] | J tube | |
| Insulin Lispro 100 UNT/ML Injectable Solution [Humalog] | pump | |
| Regular Insulin, Human 25 UNT/ML | subcutaneous | |
| Metformin hydrochloride 1000 MG / sitagliptin 50 MG [Janumet] | subcutaneously | |
| glimepiride 4 MG Oral Tablet | p.o | |
| Glyburide | PER TUBE | |
| insulin human, isophane 40 UNT/ML | Subcut. | |
| 24 HR canagliflozin 50 MG / Metformin hydrochloride 1000 MG Extended Release Oral Tablet [Invokamet] | ORAL | |
| Bromocriptine 2.5 MG Oral Tablet | Oral | |
| miglitol Oral Product | p.o. | |
| {30 (Regular Insulin, Human 4 UNT Inhalant Powder [Afrezza]) / 60 (Regular Insulin, Human 8 UNT Inhalant Powder [Afrezza]) } Pack [Afrezza 90 - 30 (4 UNT), 60 (8 UNT)] | INHALATION | |
| insulin human, isophane 50 UNT/ML / Regular Insulin, Human 50 UNT/ML [Humulin] | by mouth | |
| 3 ML insulin human, isophane 100 UNT/ML Pen Injector [Humulin N] | Subq | |
| Insulin Lispro 25 UNT/ML | p.o. | |
| Regular Insulin, Human 100 UNT/ML Injectable Solution [Novolin R] | SubQ | |
| Acarbose Oral Tablet [Precose] | oral | |
| exenatide 0.005 MG/ACTUAT [Byetta] | orally | |
| Acarbose | g - tube | |
| glimepiride | drip. | |
| canagliflozin 150 MG / Metformin hydrochloride 1000 MG Oral Tablet [Invokamet] | po | |
| Metformin hydrochloride 500 MG / repaglinide 1 MG Oral Tablet | by mouth | |
| liraglutide 6 MG/ML [Victoza] | oral | |
| Metformin Extended Release Oral Tablet [Fortamet] | subcutaneous | |
| glimepiride 4 MG / pioglitazone 30 MG Oral Tablet [Duetact] | orally | |
| Humalog Injectable Product | | 0 |
| Acarbose 25 MG [Precose] | G tube | |
| Insulin Lispro 100 UNT/ML Injectable Solution [Lispro PRC] | SQ | |
| insulin detemir 100 UNT/ML [Levemir] | oral | |
| alogliptin 25 MG / pioglitazone 30 MG Oral Tablet | by mouth | |
| liraglutide Injectable Product | SEE COMMENT | |

| | | |
|---|---|---|
| Insulin Lispro | subC | |
| Insulin, Aspart, Human | trans | |
| insulin human, isophane 75 UNT/ML | subcu | |
| empagliflozin 12.5 MG / Metformin hydrochloride 1000 MG Oral Tablet [Synjardy] | ORAL | |
| Avandamet Pill | PO | |
| Humulin U Ultralente Injectable Product | subcut. | |
| glimepiride 2 MG | inh | |
| Insulin Lispro 50 UNT/ML / Insulin, Protamine Lispro, Human 50 UNT/ML Injectable Suspension [Humalog Mix] | | |
| Glipizide Oral Product | BY MOUTH | |
| Insulin Lispro 25 UNT/ML | per tube | |
| Insulin, Aspart, Human 100 UNT/ML | subcu | |
| Metformin hydrochloride 500 MG Oral Tablet | nare | |
| Regular Insulin, Human 100 UNT/ML | iv | |
| Metformin Extended Release Oral Tablet [Fortamet] | BY MOUTH | |
| insulin human, isophane 70 UNT/ML / Regular Insulin, Human 30 UNT/ML Injectable Suspension [Novolin] | SQ | |
| Metformin 500 MG | td | |
| Insulin, Glulisine, Human | SC | |
| Insulin Lispro 25 UNT/ML / Insulin, Protamine Lispro, Human 75 UNT/ML Injectable Suspension [Humalog Mix] | PO | |
| insulin human, isophane 50 UNT/ML / Regular Insulin, Human 50 UNT/ML Injectable Suspension [Humulin] | sublingual | |
| insulin human, isophane 100 UNT/ML Injectable Suspension [Novolin N] | SUBCUTANEOUSLY | |
| repaglinide 1 MG [Prandin] | gt | |
| Glyburide | oral. | |
| 3 ML Regular Insulin, Human 100 UNT/ML Prefilled Syringe [Novolin R] | INJECTION | |
| 60 ACTUAT exenatide 0.005 MG/ACTUAT Pen Injector [Byetta] | subcutaneously. | |
| repaglinide 2 MG [Prandin] | per tube | |
| insulin degludec 100 UNT/ML Pen Injector | SQ | |
| 3 ML insulin degludec 100 UNT/ML Pen Injector [Tresiba] | | 0 |
| Regular Insulin, Human 8 UNT Inhalant Powder [Afrezza] | inhalation | |
| insulin detemir | SUB CONJ | |
| Lente Insulin, Beef-Pork 40 UNT/ML | subcutaneous | |
| 3 ML Insulin, Aspart, Human 100 UNT/ML Pen Injector [NovoLog] | SUBCUT | |
| 24 HR Metformin hydrochloride 500 MG Extended Release Oral Tablet | | 0 |

| | |
|---|---|
| Metformin hydrochloride 500 MG Extended Release Oral Tablet [Fortamet] | subcutaneously |
| dapagliflozin Oral Tablet [Farxiga] | nostril |
| 3 ML insulin detemir 100 UNT/ML Pen Injector [Levemir] | Subcut |
| Linagliptin 2.5 MG / Metformin hydrochloride 850 MG Oral Tablet [Jentadueto] | |
| Glyburide 2.5 MG | drip |
| 24 HR Metformin hydrochloride 500 MG Extended Release Oral Tablet [Glucophage] | BY MOUTH |
| insulin pork, isophane 100 UNT/ML Injectable Suspension [NPH Iletin II] | OU |
| Insulin, Aspart Protamine, Human 70 UNT/ML / Insulin, Aspart, Human 30 UNT/ML [NovoLog Mix] | Subcut |
| Regular Insulin, Human 500 UNT/ML Injectable Solution [Humulin R] | sub q |
| Pramlintide | sc |
| repaglinide | PER TUBE |
| Janumet Pill | po |
| insulin pork, isophane 100 UNT/ML Injectable Suspension [NPH Iletin II] | INH |
| miglitol 25 MG Oral Tablet [Glyset] | BY MOUTH |
| Januvia Pill | Im |
| Glipizide 5 MG / Metformin hydrochloride 500 MG Oral Tablet | Po |
| pioglitazone 45 MG [Actos] | drip |
| Modified 24 HR Metformin hydrochloride 500 MG Extended Release Oral Tablet [Glumetza] | PO |
| Regular Insulin, Human 100 UNT/ML | |
| insulin human, isophane 70 UNT/ML / Regular Insulin, Human 30 UNT/ML [Humulin] | subcutaneous. |
| Glyburide 2.5 MG | gt |
| insulin human, isophane 75 UNT/ML / Regular Insulin, Human 25 UNT/ML [Pur-In Mix] | |
| 3 ML Insulin Lispro 100 UNT/ML Pen Injector | SUBCUTANEOUS |
| liraglutide | inhaled |
| Insulin Lispro 100 UNT/ML | SC |
| Glipizide | topically |
| Metformin hydrochloride 100 MG/ML Oral Solution [Riomet] | BY MOUTH |
| Glipizide | iv. |
| glimepiride 4 MG | ng |
| insulin detemir 100 UNT/ML Injectable Solution [Levemir] | oral |

| | |
|---|---|
| NovoLog Injectable Product | MISCELLANEOUS |
| insulin pork, isophane 100 UNT/ML Injectable Suspension [NPH Iletin II] | SUBCUTANEOUSLY |
| 3 ML insulin human, isophane 70 UNT/ML / Regular Insulin, Human 30 UNT/ML Pen Injector [Humulin] | drip |
| {30 (Regular Insulin, Human 12 UNT Inhalant Powder [Afrezza]) / 60 (Regular Insulin, Human 8 UNT Inhalant Powder [Afrezza]) } Pack [Afrezza 90 - 60 (8 UNT), 30 (12 UNT)] | inhalation |
| Linagliptin 2.5 MG / Metformin hydrochloride 500 MG Oral Tablet | PO |
| dulaglutide | IM |
| Gliclazide Oral Tablet | by mouth |
| insulin human, isophane 90 UNT/ML | PO |
| Insulin, Aspart Protamine, Human 70 UNT/ML / Insulin, Aspart, Human 30 UNT/ML [NovoLog Mix] | Sub q |
| insulin beef, isophane 40 UNT/ML | oral |
| dapagliflozin 10 MG Oral Tablet | orally |
| insulin human, isophane 70 UNT/ML / Regular Insulin, Human 30 UNT/ML Injectable Suspension [Humulin] | nostril |
| Insulin, Aspart Protamine, Human 70 UNT/ML / Insulin, Aspart, Human 30 UNT/ML Injectable Suspension [NovoLog Mix] | IV |
| Glyburide 1.25 MG / Metformin hydrochloride 250 MG Oral Tablet | p.o. |
| 3 ML insulin human, isophane 100 UNT/ML Pen Injector [Humulin N] | sub q |
| Glyburide 5 MG | subcutaneous |
| glimepiride 4 MG / pioglitazone 30 MG Oral Tablet [Duetact] | p.o. |
| Insulin Glargine Injectable Solution [Lantus] | SUBCUTANEOUS |
| Metformin Pill | swallow. |
| dapagliflozin 10 MG [Farxiga] | subcutaneously |
| exenatide 3.08 MG/ML [Bydureon] | by mouth |
| Kombiglyze Oral Product | p.o. |
| Metformin hydrochloride 500 MG / rosiglitazone 2 MG [Avandamet] | nare |
| Glyburide 2.5 MG | per mouth |
| Fortamet Pill | |
| Metformin hydrochloride 1000 MG / saxagliptin 2.5 MG [Kombiglyze] | oral |
| Insulin Glargine | subling |
| Metformin | subcu. |

| | | |
|---|---|---|
| Insulin Lispro 100 UNT/ML Injectable Solution | injection | |
| repaglinide 1 MG Oral Tablet | | |
| Glyburide 1.25 MG / Metformin hydrochloride 250 MG [Glucovance] | PO | |
| Metformin hydrochloride 500 MG / saxagliptin 5 MG Extended Release Oral Tablet | | |
| Metformin 1000 MG | drip. | |
| Metformin hydrochloride 500 MG [Glucophage] | IM | |
| glimepiride 2 MG / pioglitazone 30 MG Oral Tablet [Duetact] | oral | |
| Metformin hydrochloride 500 MG / repaglinide 2 MG [PrandiMet] | | |
| Regular Insulin, Human 100 UNT/ML Injectable Solution [Humulin R] | Sc | |
| Regular Insulin, Human 40 UNT/ML | SUBCUT | |
| Metformin hydrochloride 500 MG [Glumetza] | po | |
| Metformin | J - tube | |
| insulin human, isophane 70 UNT/ML / Regular Insulin, Human 30 UNT/ML [Humulin] | npo | |
| glimepiride 2 MG / pioglitazone 30 MG Oral Tablet | nostril | |
| 1.5 ML insulin human, isophane 70 UNT/ML / Regular Insulin, Human 30 UNT/ML Prefilled Syringe | sq | |
| Humulin N Injectable Product | | 0 |
| exenatide | subcutaneous | |
| Insulin, Zinc, Human 100 UNT/ML Injectable Suspension [Human Ultratard] | | |
| Regular Insulin, Human 8 UNT | subcu | |
| Metformin | NOSTRIL | |
| Glyburide 2.5 MG / Metformin hydrochloride 500 MG [Glucovance] | PO | |
| Avandia Oral Product | Orally | |
| Insulin, Aspart, Human | Sub q | |
| Fortamet Oral Product | p.o. | |
| canagliflozin 150 MG / Metformin hydrochloride 500 MG Oral Tablet [Invokamet] | p.o. | |
| dulaglutide 3 MG/ML [Trulicity] | SubQ | |
| 24 HR Glipizide 10 MG Extended Release Oral Tablet [Glucotrol] | po | |
| Metformin 1000 MG | im | |
| Regular Insulin, Human 100 UNT/ML Injectable Solution [Novolin R] | subQ | |

| | |
|---|---|
| insulin human, isophane 70 UNT/ML / Regular Insulin, Human 30 UNT/ML [Humulin] | NPO |
| 60 ACTUAT exenatide 0.005 MG/ACTUAT Pen Injector [Byetta] | Subc. |
| Xigduo Oral Product | oral |
| Tradjenta Oral Product | ORAL |
| Metformin Oral Solution | inhaled |
| Metformin Oral Solution | p.o |
| insulin human, isophane 70 UNT/ML / Regular Insulin, Human 30 UNT/ML [Humulin] | subcutaneously. |
| insulin human, isophane 80 UNT/ML / Regular Insulin, Human 20 UNT/ML [Humulin] | p.o. |
| insulin human, isophane 70 UNT/ML | sub Q |
| Glipizide Oral Tablet [Glucotrol] | Oral |
| Bromocriptine | oral |
| repaglinide 1 MG Oral Tablet [NovoNorm] | po |
| Insulin, Aspart Protamine, Human 70 UNT/ML / Insulin, Aspart, Human 30 UNT/ML Injectable Suspension [NovoLog Mix] | IM |
| Glucotrol Oral Product | Oral |
| glimepiride Oral Tablet | PO |
| Insulin, Glulisine, Human 100 UNT/ML | |
| glimepiride 3 MG Oral Tablet | p.o. |
| Linagliptin 5 MG [Tradjenta] | sq |
| glimepiride 3 MG [Amaryl] | subcutaneous |
| Humulin N Injectable Product | SUBCUTANEOUSLY |
| insulin human, isophane 70 UNT/ML / Regular Insulin, Human 30 UNT/ML [Humulin] | G - tube |
| 3 ML liraglutide 6 MG/ML Pen Injector [Victoza] | Subcut. |
| Glipizide 5 MG Extended Release Oral Tablet [Glucotrol] | oral |
| 3 ML insulin human, isophane 100 UNT/ML Prefilled Syringe | subcu |
| Metformin | Trans |
| Glyburide 2.5 MG / Metformin hydrochloride 500 MG Oral Tablet | Oral |
| Metformin Pill | orally |
| Riomet Oral Product | orally |
| saxagliptin 2.5 MG Oral Tablet | PO |
| Metformin hydrochloride 500 MG Extended Release Oral Tablet [Glumetza] | gt |
| Metformin hydrochloride 850 MG [Glucophage] | per mouth |
| Glipizide 5 MG [Glucotrol] | subcutaneously |
| 3 ML Insulin Lispro 50 UNT/ML / Insulin, Protamine Lispro, Human 50 UNT/ML Pen Injector [Humalog Mix] | subcu |

| | |
|---|---|
| Pramlintide | SC |
| Insulin Glargine 100 UNT/ML Pen Injector [Lantus] | SQ |
| Metformin hydrochloride 1000 MG Extended Release Oral Tablet [Fortamet] | p.o. |
| saxagliptin Pill | by mouth |
| repaglinide Oral Product | p.o. |
| insulin degludec Pen Injector [Tresiba] | |
| Metformin hydrochloride 850 MG | SC |
| Glyburide Oral Tablet | ORAL |
| Metformin hydrochloride 1000 MG / sitagliptin 50 MG Oral Tablet [Janumet] | BY MOUTH |
| insulin, regular, beef-pork 40 UNT/ML Inhalant Solution | Subcutaneous |
| pioglitazone 15 MG [Actos] | by mouth. |
| insulin beef, isophane 40 UNT/ML | sq |
| Glipizide 10 MG | sq |
| 0.5 ML albiglutide 60 MG/ML Pen Injector [Tanzeum] | orally |
| 24 HR Glipizide 10 MG Extended Release Oral Tablet | |
| Metformin | Iv |
| Insulin Lispro 100 UNT/ML [Humalog] | subcu |
| Metformin hydrochloride 850 MG | subcutaneous |
| Metformin 250 MG Oral Tablet | ORAL |
| sitagliptin 100 MG [Januvia] | per mouth |
| insulin human, isophane 70 UNT/ML / Regular Insulin, Human 30 UNT/ML [Novolin] | MISCELLANEOUS |
| 24 HR Metformin hydrochloride 500 MG / saxagliptin 5 MG Extended Release Oral Tablet [Kombiglyze] | po |
| {180 (insulin human, rDNA origin 1 MG/ACTUAT Inhalant Powder [Exubera]) / 90 (insulin human, rDNA origin 3 MG/ACTUAT Inhalant Powder [Exubera]) } Pack [Exubera Kit] | |
| Regular Insulin, Human 5 UNT/ML | drip |
| Metformin hydrochloride 850 MG [Glucophage] | subcutaneously |
| repaglinide 2 MG Oral Tablet | PO |
| Metformin hydrochloride 500 MG / rosiglitazone 4 MG Oral Tablet [Avandamet] | oral |
| rosiglitazone 4 MG [Avandia] | per mouth |
| Metformin | intravenously |
| saxagliptin 5 MG Oral Tablet | |
| insulin detemir | MISC |
| Insulin, Aspart Protamine, Human 70 UNT/ML / Insulin, Aspart, Human 30 UNT/ML [NovoLog Mix] | MISCELLANEOUS |

| | | |
|---|---|---|
| Glipizide 10 MG Oral Tablet | P.O. | |
| Glipizide Oral Product | pO | |
| 3 ML Insulin Glargine 100 UNT/ML Pen Injector [Lantus] | SubQ | |
| Insulin Lispro 25 UNT/ML / Insulin, Protamine Lispro, Human 75 UNT/ML [Humalog Mix] | Sub Q | |
| insulin human, isophane 80 UNT/ML / Regular Insulin, Human 20 UNT/ML [Humulin] | subcut | |
| Glyburide 2.5 MG | bedside. | |
| Victoza Injectable Product | SUBCU | |
| Linagliptin Oral Product | orally | |
| insulin human, isophane 60 UNT/ML | SubQ | |
| Insulin Lispro | SUB Q | |
| Metformin 250 MG | IM | |
| Jentadueto Oral Product | oral | |
| 1.5 ML insulin human, isophane 100 UNT/ML Prefilled Syringe | subcutaneous | |
| Metformin hydrochloride 500 MG / rosiglitazone 2 MG [Avandamet] | IV | |
| Insulin Lispro 100 UNT/ML Injectable Solution | SubQ | |
| pioglitazone 30 MG | topically | |
| 0.5 ML albiglutide 100 MG/ML Pen Injector [Tanzeum] | sc | |
| Acarbose | IV | |
| Metformin | inh. | |
| sitagliptin 50 MG [Januvia] | td | |
| Acarbose 25 MG Oral Tablet [Precose] | | 0 |
| 24 HR dapagliflozin 5 MG / Metformin hydrochloride 500 MG Extended Release Oral Tablet [Xigduo] | | |
| Humulin Injectable Product | sq | |
| 0.5 ML albiglutide 60 MG/ML Pen Injector [Tanzeum] | sub q | |
| Metformin | NPo | |
| sitagliptin 100 MG [Januvia] | subcut | |
| exenatide 0.005 MG/ACTUAT [Byetta] | SEE COMMENT | |
| Invokana Oral Product | Oral | |
| pioglitazone 15 MG Oral Tablet [Actos] | sublingual | |
| Insulin Lispro 100 UNT/ML Injectable Solution [Humalog] | PUMP | |
| Regular Insulin, Human 500 UNT/ML Injectable Solution [Humulin R] | sub Q | |
| Glipizide 10 MG | IM | |
| insulin human, isophane 60 UNT/ML | by mouth | |
| Pramlintide Injectable Solution | subcutaneously | |
| Metformin hydrochloride 1000 MG [Glucophage] | subcutaneous | |

| | | |
|---|---|---|
| glimepiride 4 MG [Amaryl] | po. | |
| 24 HR dapagliflozin 10 MG / Metformin hydrochloride 1000 MG Extended Release Oral Tablet [Xigduo] | ORAL | |
| Acarbose | MISC | |
| Glipizide 5 MG | po. | |
| insulin human, isophane 70 UNT/ML / Regular Insulin, Human 30 UNT/ML [Novolin] | subq. | |
| Metformin 500 MG | swallow. | |
| Regular Insulin, Human | Intra_Muscular | |
| Glyburide 2.5 MG Oral Tablet [Micronase] | ORAL | |
| Ultralente Insulin, Human | sub q | |
| Ultralente Insulin, Human 100 UNT/ML Injectable Suspension [Humulin U Ultralente] | per tube | |
| 3 ML insulin detemir 100 UNT/ML Pen Injector [Levemir] | subcut. | |
| Metformin 400 MG | p.o. | |
| Glipizide 5 MG [Glucotrol] | gt | |
| Glipizide 10 MG | swallow | |
| Parlodel Oral Product | BY MOUTH | |
| 3 ML insulin degludec 100 UNT/ML Pen Injector [Tresiba] | intramuscular | |
| Osmotic 24 HR Metformin hydrochloride 500 MG Extended Release Oral Tablet [Fortamet] | PO | |
| Insulin Glargine | OU | |
| Insulin, Zinc, Human | sub Q | |
| Insulin Glargine 100 UNT/ML Injectable Solution | | 0 |
| liraglutide | SUBCUTANEOUS | |
| Insulin Lispro | SUBCUTANEOUSLY | |
| liraglutide 6 MG/ML [Victoza] | SUBCUTANEOUSLY | |
| Metformin 500 MG | Im | |
| Glipizide 5 MG Oral Tablet | g-tube | |
| Insulin Lispro | nares | |
| Bromocriptine 5 MG | swallow | |
| Glyburide 1 MG | nostril | |
| Metformin 500 MG | npo | |
| Bromocriptine 2.5 MG Oral Tablet [Parlodel] | INTRAVAGINALLY | |
| Victoza Injectable Product | IM INJECTION | |
| repaglinide Oral Product | PO | |
| Metformin hydrochloride 500 MG / rosiglitazone 2 MG [Avandamet] | subcutaneously | |
| 24 HR Metformin hydrochloride 1000 MG / pioglitazone 30 MG Extended Release Oral Tablet [Actoplus Met] | oral | |

| | | |
|---|---|---|
| exenatide 0.01 MG/ACTUAT | SC | |
| Regular Insulin, Human 100 UNT/ML Injectable Solution [Novolin R] | subcutaneous. | |
| Insulin, Aspart Protamine, Human 70 UNT/ML / Insulin, Aspart, Human 30 UNT/ML Injectable Suspension [NovoLog Mix] | p.o. | |
| Insulin Glargine 100 UNT/ML [Lantus] | sQ | |
| Insulin Lispro 50 UNT/ML | IM | |
| insulin pork, isophane 100 UNT/ML Injectable Suspension [NPH Iletin II] | td | |
| liraglutide 6 MG/ML | SC | |
| Insulin, Glulisine, Human | sc | |
| 3 ML Insulin Lispro 100 UNT/ML Pen Injector [Humalog] | subcutaneous | |
| Glipizide 5 MG | IV | |
| 3 ML Insulin Lispro 200 UNT/ML Pen Injector [Humalog] | | 0 |
| Insulin, Aspart, Human 100 UNT/ML Cartridge [NovoLog] | po. | |
| canagliflozin 50 MG / Metformin hydrochloride 500 MG Oral Tablet [Invokamet] | | 0 |
| Insulin Lispro 100 UNT/ML [Humalog] | Subcutaneous | |
| Glipizide 10 MG | intravenous | |
| Metformin hydrochloride 500 MG Oral Tablet | P.O | |
| Metformin hydrochloride 1000 MG Oral Tablet | topical | |
| sitagliptin 100 MG [Januvia] | ng | |
| dulaglutide 3 MG/ML [Trulicity] | SUBCU | |
| saxagliptin 5 MG [Onglyza] | by mouth. | |
| Insulin Lispro 50 UNT/ML | bedside | |
| Janumet Oral Product | Po | |
| Insulin Glargine | subC | |
| Invokana Oral Product | p.o. | |
| insulin human, isophane 100 UNT/ML Injectable Suspension [Novolin N] | SUBQ | |
| Lispro PRC Injectable Product | NPO | |
| Bromocriptine Oral Capsule [Parlodel] | | |
| Glipizide 10 MG | bedside. | |
| Glipizide | sq | |
| Metformin 1000 MG | Im | |
| Insulin Glargine | sublingual | |
| Metformin 250 MG | IV | |
| 3 ML Insulin Lispro 100 UNT/ML Cartridge [Humalog] | MISCELLANEOUS | |
| Insulin, Glulisine, Human 100 UNT/ML Injectable Solution [Apidra] | MISC | |

| | |
|---|---|
| Farxiga Oral Product | BY MOuth |
| Metformin hydrochloride 1000 MG [Glucophage] | per mouth |
| Glyburide 1 MG Oral Tablet | inhalation |
| glimepiride 4 MG | NPO. |
| Regular Insulin, Human 6 UNT/ML | subcutaneously |
| miglitol 25 MG Oral Tablet | PO |
| canagliflozin 100 MG Oral Tablet | PO |
| glimepiride 2 MG | inhalation |
| 3 ML Insulin Lispro 200 UNT/ML Pen Injector | SQ |
| Insulin Glargine 300 UNT/ML | PO |
| insulin human, isophane 70 UNT/ML | p.o. |
| glimepiride | sub q |
| Insulin Lispro Injectable Solution [Humalog] | by mouth |
| Regular Insulin, Human 100 UNT/ML Injectable Solution | subq |
| albiglutide 60 MG/ML [Tanzeum] | SQ |
| exenatide | drip |
| insulin human, isophane 60 UNT/ML | orally |
| empagliflozin Oral Tablet | orally |
| Tolbutamide 1000 MG | |
| Metformin Extended Release Oral Tablet [Glumetza] | subcutaneous |
| Metformin / sitagliptin Oral Tablet [Janumet] | |
| Chlorpropamide 100 MG Oral Tablet [Diabinese] | p.o. |
| Insulin, Glulisine, Human 100 UNT/ML Injectable Solution [Apidra] | gt |
| Trulicity Injectable Product | SUBCU |
| Glipizide Oral Tablet | PO |
| Metformin 500 MG | nare |
| Regular Insulin, Human 500 UNT/ML Injectable Solution [Humulin R] | CONT SUBCUTAN INFUSI |
| Insulin Lispro Injectable Solution | IV |
| repaglinide 1 MG Oral Tablet | PO |
| Insulin Glargine 100 UNT/ML [Lantus] | im |
| Toujeo Injectable Product | SUBCUTANEOUSLY |
| Insulin Lispro Injectable Solution | 0 |
| Bromocriptine Oral Tablet [Parlodel] | oral |
| Metformin hydrochloride 850 MG / pioglitazone 15 MG Oral Tablet | orally |
| Tolazamide 250 MG Oral Tablet [Tolinase] | p.o. |
| Glipizide 2.5 MG | NPO |
| sitagliptin | swallow |

| | | |
|---|---|---|
| Insulin Lispro 50 UNT/ML | swallow | |
| pramlintide acetate 0.6 MG/ML Injectable Solution [Symlin] | subcu | |
| Regular Insulin, Human 30 UNT/ML | PO | |
| Insulin Lispro 25 UNT/ML | | 0 |
| glimepiride 1 MG / rosiglitazone 4 MG Oral Tablet [Avandaryl] | ORAL | |
| 3 ML Insulin Lispro 200 UNT/ML Pen Injector | subq | |
| Regular Insulin, Human 10 UNT/ML | sc | |
| Lantus Injectable Product | SubQ | |
| empagliflozin 25 MG Oral Tablet [Jardiance] | BY mouth | |
| glimepiride | Topically | |
| 24 HR Metformin hydrochloride 750 MG Extended Release Oral Tablet [Glucophage] | orally | |
| Insulin, Aspart, Human 100 UNT/ML Cartridge [NovoLog] | bedside | |
| dapagliflozin 10 MG Oral Tablet [Farxiga] | Oral | |
| 3 ML Regular Insulin, Human 100 UNT/ML Prefilled Syringe [Novolin R] | subcutaneously | |
| empagliflozin Pill | | |
| pioglitazone 45 MG Oral Tablet [Actos] | ORAL | |
| pioglitazone 15 MG [Actos] | gt | |
| Bromocriptine 2.5 MG Extended Release Oral Capsule | by mouth | |
| Metformin / rosiglitazone Pill | | |
| dapagliflozin 5 MG / Metformin hydrochloride 500 MG [Xigduo] | BY MOUTH | |
| Linagliptin / Metformin Oral Product | p.o. | |
| Metformin hydrochloride 1000 MG Oral Tablet | G - tube | |
| Prandin Oral Product | Oral | |
| dapagliflozin 5 MG [Farxiga] | PO | |
| rosiglitazone Oral Tablet [Avandia] | p.o. | |
| 3 ML Insulin Lispro 50 UNT/ML / Insulin, Protamine Lispro, Human 50 UNT/ML Pen Injector [Humalog Mix] | SUBQ | |
| insulin human, isophane 70 UNT/ML | subcu. | |
| liraglutide | | 0 |
| pioglitazone Oral Tablet [Actos] | PO | |
| pramlintide acetate 0.6 MG/ML Injectable Solution [Symlin] | subQ | |
| Regular Insulin, Human 100 UNT/ML Injectable Solution | SEE COMMENT | |
| glimepiride 2 MG / pioglitazone 30 MG Oral Tablet | orally | |
| canagliflozin 300 MG Oral Tablet [Invokana] | Po | |
| empagliflozin 10 MG Oral Tablet [Jardiance] | p.o. | |
| Metformin hydrochloride 1000 MG Oral Tablet | Inhalation | |
| sitagliptin Oral Tablet | | |

| | |
|---|---|
| Insulin, Aspart Protamine, Human 70 UNT/ML / Insulin, Aspart, Human 30 UNT/ML [NovoLog Mix] | po |
| insulin human, isophane 70 UNT/ML / Regular Insulin, Human 30 UNT/ML [Humulin] | sublingual |
| Regular Insulin, Human 25 UNT/ML | per tube |
| Metformin hydrochloride 1000 MG / rosiglitazone 2 MG Oral Tablet | by mouth |
| Acarbose 25 MG Oral Tablet | Oral |
| Lente Insulin, Beef 80 UNT/ML | subcut |
| albiglutide 100 MG/ML [Tanzeum] | SQ |
| Insulin, Zinc, Human | PO |
| glimepiride 2 MG | NPO. |
| alogliptin 25 MG / pioglitazone 30 MG Oral Tablet [Oseni] | ORAL |
| insulin human, isophane 70 UNT/ML | oral |
| Insulin Lispro 100 UNT/ML [Humalog] | subcutaneous |
| Glipizide 5 MG / Metformin hydrochloride 500 MG [Metaglip] | drip |
| rosiglitazone 1 MG Oral Tablet | by mouth |
| Insulin Lispro 100 UNT/ML Pen Injector [Humalog] | by mouth |
| insulin human, isophane 100 UNT/ML Injectable Suspension | by mouth |
| insulin pork, isophane 100 UNT/ML Injectable Suspension [NPH Iletin II] | Sub q |
| Insulin Glargine Injectable Solution | bedside |
| Metformin 1000 MG | SL |
| insulin human, isophane 40 UNT/ML | oral |
| canagliflozin 50 MG | |
| Regular Insulin, Human 500 UNT/ML Injectable Solution [Humulin R] | ORAL |
| Metforming Oral Product | by mouth |
| Insulin Lispro | Bedside |
| 0.5 ML albiglutide 100 MG/ML Pen Injector [Tanzeum] | subcut |
| Insulin Lispro 100 UNT/ML Injectable Solution | infusion |
| repaglinide 1 MG Oral Tablet | p.o. |
| Glipizide Oral Tablet [Glucotrol] | p.o. |
| nateglinide 120 MG [Starlix] | subcutaneously |
| Regular Insulin, Human 40 UNT/ML | po |
| liraglutide | MISC |
| saxagliptin 5 MG [Onglyza] | SEE COMMENT |
| 3 ML liraglutide 6 MG/ML Pen Injector | oral |
| Insulin, Zinc, Human 100 UNT/ML Injectable Suspension [Humulin L] | subcut. |

| | | |
|---|---|---|
| 24 HR empagliflozin 10 MG / Metformin hydrochloride 1000 MG Extended Release Oral Tablet [Synjardy] | oral | |
| insulin human, isophane 70 UNT/ML / Regular Insulin, Human 30 UNT/ML [Humulin] | subQ. | |
| insulin human, isophane 100 UNT/ML Injectable Suspension [Humulin I] | by mouth | |
| glimepiride 3 MG Oral Tablet [Amaryl] | oral | |
| Glyburide | Im | |
| Insulin, Zinc, Pork | subcutaneously | |
| Metformin hydrochloride 500 MG Oral Tablet | Im | |
| dapagliflozin Oral Tablet | oral | |
| Metformin hydrochloride 1000 MG / rosiglitazone 2 MG Oral Tablet [Avandamet] | oral | |
| glimepiride 2 MG / pioglitazone 30 MG Oral Tablet [Duetact] | | |
| insulin human, isophane 70 UNT/ML / Regular Insulin, Human 30 UNT/ML [Novolin] | td | |
| Ultralente Insulin, Human 100 UNT/ML Injectable Suspension | npo | |
| Jentadueto Oral Product | PO | |
| Glyburide | topical | |
| Insulin Lispro 50 UNT/ML / Insulin, Protamine Lispro, Human 50 UNT/ML Injectable Suspension [Humalog Mix] | Sub q | |
| Insulin Lispro 100 UNT/ML Injectable Solution [Humalog] | Subq | |
| Metformin Oral Tablet | ORAL | |
| Bydureon Injectable Product | Subcutaneously | |
| 3 ML Insulin Glargine 100 UNT/ML Pen Injector [Lantus] | MISCELLANEOUS | |
| 0.5 ML dulaglutide 1.5 MG/ML Auto-Injector [Trulicity] | IM injection | |
| Metformin | sQ | |
| alogliptin 6.25 MG | | |
| pioglitazone 45 MG [Actos] | subcu | |
| 3 ML Insulin, Aspart, Human 100 UNT/ML Pen Injector [NovoLog] | subconjunctival | |
| Insulin Lispro Cartridge [Humalog] | | 0 |
| 3 ML insulin human, isophane 100 UNT/ML Prefilled Syringe | npo. | |
| Novolin N Injectable Product | | 0 |
| Insulin, Aspart, Human 100 UNT/ML Injectable Solution [NovoLog] | CONT SUBCUTAN INFUSION | |
| empagliflozin 5 MG / Metformin hydrochloride 1000 MG Oral Tablet [Synjardy] | ORAL | |
| insulin human, isophane 70 UNT/ML / Regular Insulin, Human 30 UNT/ML Injectable Suspension [Novolin] | sub q | |

| | | |
|---|---|---|
| 24 HR empagliflozin 12.5 MG / Metformin hydrochloride 1000 MG Extended Release Oral Tablet [Synjardy] | ORAL | |
| Novolin R Injectable Product | SUBCUTANEOUSLY | |
| insulin detemir 100 UNT/ML [Levemir] | SUB Q | |
| Parlodel Pill | | |
| rosiglitazone Oral Tablet | | |
| Insulin, Aspart, Human 100 UNT/ML Injectable Solution [NovoLog] | Subcutaneous | |
| Insulin Glargine 100 UNT/ML Injectable Solution | SubQ | |
| Metformin hydrochloride 1000 MG Oral Tablet | po. | |
| Victoza Injectable Product | SEE COMMENT | |
| Victoza Injectable Product | po | |
| Metformin hydrochloride 1000 MG / saxagliptin 5 MG [Kombiglyze] | NPO | |
| canagliflozin | PO | |
| 1.5 ML pramlintide acetate 1 MG/ML Pen Injector [Symlin] | | |
| Insulin Lispro 50 UNT/ML / Insulin, Protamine Lispro, Human 50 UNT/ML Injectable Suspension [Humalog Mix] | NPO | |
| Lixisenatide | | 0 |
| Insulin Lispro 100 UNT/ML [Humalog] | SQ | |
| Acarbose | PO / G - tube | |
| Ultralente Insulin, Human | by mouth | |
| Metformin hydrochloride 500 MG Oral Tablet [Glucophage] | swallow | |
| 24 HR Metformin hydrochloride 750 MG Extended Release Oral Tablet | subcutaneously | |
| Glipizide | intravenous | |
| Kombiglyze Oral Product | oral | |
| Insulin Lispro 100 UNT/ML | po | |
| Linagliptin | IV | |
| insulin detemir 100 UNT/ML Injectable Solution [Levemir] | subcut. | |
| saxagliptin Oral Tablet | oral | |
| Acarbose | per tube | |
| insulin pork, isophane 100 UNT/ML Injectable Suspension | subcu | |
| Actos Oral Product | Orally | |
| Glipizide | | 0 |
| Metformin hydrochloride 500 MG Oral Tablet [Glucophage] | . | |
| 3 ML Insulin, Aspart Protamine, Human 70 UNT/ML / Insulin, Aspart, Human 30 UNT/ML Pen Injector [NovoLog Mix] | injection | |
| Metformin Oral Solution [Riomet] | swallow | |
| Metformin 250 MG Oral Tablet | BY MOUTH | |

| | |
|---|---|
| Humalog Injectable Product | SUBCUTANEOUSLY |
| insulin pork, isophane 100 UNT/ML Injectable Suspension | by mouth |
| Acarbose | topical |
| Glyburide 6 MG | subcutaneous |
| Ultralente Insulin, Human 100 UNT/ML Injectable Suspension | by mouth |
| Glucophage Pill | oral |
| Glipizide 2.5 MG | gt |
| glimepiride | Bedside |
| Insulin Lispro 200 UNT/ML [Humalog] | po |
| Regular Insulin, Human 10 UNT/ML | im |
| Metformin 400 MG | GT |
| Regular Insulin, Human | subcutaneously. |
| Acarbose 25 MG | G tube |
| Glipizide 5 MG / Metformin hydrochloride 500 MG Oral Tablet | p.o. |
| Ultralente Insulin, Human 100 UNT/ML Injectable Suspension | subcu. |
| Metformin Oral Solution [Riomet] | po |
| insulin pork, isophane 100 UNT/ML Injectable Suspension [NPH Iletin II] | sc. |
| glimepiride 4 MG | subcu |
| 3 ML insulin detemir 100 UNT/ML Pen Injector [Levemir] | INJECTION |
| exenatide | SEE COMMENT |
| 3 ML Insulin, Aspart Protamine, Human 70 UNT/ML / Insulin, Aspart, Human 30 UNT/ML Pen Injector [NovoLog Mix] | IV |
| glimepiride | per tube |
| Metformin Oral Solution [Riomet] | oral |
| Metformin 500 MG | sub q |
| dulaglutide | SQ |
| Metformin 1000 MG | PER TUBE |
| glimepiride 2 MG / pioglitazone 30 MG Oral Tablet | by mouth |
| nateglinide 60 MG [Starlix] | inhaled |
| 3 ML Regular Insulin, Human 500 UNT/ML Pen Injector [Humulin R] | by mouth |
| Insulin, Aspart, Human 100 UNT/ML Cartridge [NovoLog] | SUB Q |
| Metformin 1000 MG | swallow. |
| 24 HR Bromocriptine 2.5 MG Extended Release Oral Capsule | oral |
| Metformin 1000 MG | topically |
| Regular Insulin, Human 100 UNT/ML Injectable Solution [Humulin R] | SUB Q |
| insulin pork, isophane 100 UNT/ML Injectable Suspension [NPH Iletin II] | Bedside |

| | |
|---|---|
| Insulin Glargine 100 UNT/ML [Lantus] | G - tube |
| Insulin Glargine 100 UNT/ML [Lantus] | subcutaneous |
| Glipizide | bedside. |
| repaglinide 0.5 MG Oral Tablet [Prandin] | BY MOUTH |
| Glipizide 10 MG Oral Tablet | IM |
| Chlorpropamide 250 MG Oral Tablet | oral |
| insulin pork, isophane 100 UNT/ML Injectable Suspension [NPH Iletin II] | subcutaneous. |
| dapagliflozin 5 MG / Metformin hydrochloride 1000 MG Extended Release Oral Tablet [Xigduo] | by mouth |
| sitagliptin Oral Tablet | Oral |
| 0.5 ML dulaglutide 1.5 MG/ML Auto-Injector | orally |
| insulin, isophane Injectable Suspension [Humulin N] | by mouth |
| Regular Insulin, Human 100 UNT/ML Injectable Solution [Novolin R] | SUBCUTANEOUSLY |
| insulin pork, isophane 80 UNT/ML | orally |
| Metformin | ORAL |
| Insulin, Aspart, Human 100 UNT/ML Cartridge [NovoLog] | sublingual |
| Glipizide 10 MG [Glucotrol] | topical |
| insulin, isophane Injectable Suspension | |
| Bromocriptine 25 MG Oral Tablet | PO |
| Gliclazide 30 MG Extended Release Oral Tablet | by mouth |
| Ultralente Insulin, Human | oral |
| repaglinide 2 MG [Prandin] | gt |
| Insulin Glargine | im |
| exenatide | sq |
| Glyburide 1 MG Oral Tablet | ORALLY |
| Glyburide Oral Tablet [Diabeta] | Oral |
| Glipizide 10 MG | IV |
| Insulin Glargine 100 UNT/ML [Lantus] | NG |
| insulin beef, isophane 80 UNT/ML | |
| Metformin hydrochloride 500 MG / rosiglitazone 2 MG [Avandamet] | swallow |
| Insulin Lispro | sublingual |
| glimepiride Oral Tablet [Amaryl] | TD |
| Insulin Lispro Pen Injector [Humalog] | by mouth |
| Insulin, Aspart, Human | Npo |
| 3 ML liraglutide 6 MG/ML Pen Injector | sq |
| insulin human, isophane 50 UNT/ML / Regular Insulin, Human 50 UNT/ML [Humulin] | sub q |

| | | |
|---|---|---|
| exenatide 2 MG | sub q | |
| Regular Insulin, Human | td | |
| Metformin Extended Release Oral Tablet | subcutaneously | |
| insulin pork, isophane 100 UNT/ML Injectable Suspension [NPH Iletin II] | MISC.(NON-DRUG; COMBO ROUTE) | |
| liraglutide 6 MG/ML | sub q | |
| glimepiride | SQ. | |
| Acetohexamide | | |
| alogliptin 25 MG Oral Tablet [Nesina] | | 0 |
| Metformin hydrochloride 1000 MG Oral Tablet | topically | |
| Insulin Lispro 25 UNT/ML / Insulin, Protamine Lispro, Human 75 UNT/ML [Humalog Mix] | gt | |
| insulin human, isophane 60 UNT/ML | subcutaneous. | |
| Novolin N Injectable Product | SUBCUTANEOUSLY | |
| Insulin, Zinc, Human | po | |
| Glipizide 10 MG | ou | |
| insulin human, isophane 70 UNT/ML / Regular Insulin, Human 30 UNT/ML Injectable Suspension [Novolin] | subcu | |
| Avandaryl Oral Product | | |
| insulin human, isophane 30 UNT/ML | subcu | |
| Insulin Lispro 100 UNT/ML Injectable Solution [Lispro PRC] | NPO. | |
| Novolin Injectable Product | SQ | |
| Regular Insulin, Human 12 UNT | SUBCUT | |
| Metformin hydrochloride 1000 MG Extended Release Oral Tablet | topically | |
| dapagliflozin 5 MG / Metformin hydrochloride 1000 MG [Xigduo] | PO | |
| Insulin, Aspart, Human | subcu. | |
| Bromocriptine | nostril | |
| Chlorpropamide 100 MG Oral Tablet | p.o. | |
| Metformin Oral Solution | P.o. | |
| glimepiride 4 MG | sc | |
| canagliflozin 100 MG [Invokana] | gt | |
| Glucotrol Pill | oral | |
| Insulin Lispro 50 UNT/ML / Insulin, Protamine Lispro, Human 50 UNT/ML Injectable Suspension [Humalog Mix] | subcu. | |
| Insulin Lispro 100 UNT/ML Injectable Solution [Lispro PRC] | NPO | |
| 1.5 ML insulin human, isophane 70 UNT/ML / Regular Insulin, Human 30 UNT/ML Prefilled Syringe | po | |
| pioglitazone Oral Tablet | Po | |
| 3 ML liraglutide 6 MG/ML Pen Injector [Victoza] | im | |
| Insulin Lispro 100 UNT/ML Injectable Solution [Humalog] | INSULIN PUMP | |

| | |
|---|---|
| dapagliflozin 10 MG Oral Tablet | PO |
| Insulin, Aspart, Human | subq. |
| Chlorpropamide Oral Tablet | po |
| Regular Insulin, Human | IV_Push |
| Insulin Glargine | PO. |
| insulin human, isophane 50 UNT/ML / Regular Insulin, Human 50 UNT/ML [Humulin] | po |
| glimepiride Oral Tablet [Amaryl] | PO |
| Glyburide 2.5 MG Oral Tablet | P.O. |
| Insulin, Aspart, Human 100 UNT/ML Injectable Solution [NovoLog] | SUBCUTANEOUS/PUMP |
| glimepiride | ou |
| Bromocriptine Oral Capsule | po |
| Linagliptin | nostril |
| Regular Insulin, Human 15 UNT/ML | per tube |
| liraglutide | SEE COMMENT |
| Glyburide | swallow |
| 3 ML Insulin, Aspart Protamine, Human 70 UNT/ML / Insulin, Aspart, Human 30 UNT/ML Pen Injector [NovoLog Mix] | intramuscular |
| Insulin Glargine 300 UNT/ML Pen Injector | subcutaneous |
| Bromocriptine 2.5 MG Oral Tablet | NG |
| Insulin, Zinc, Human | SUB Q |
| Januvia Oral Product | ORAL |
| Bromocriptine 5 MG Oral Capsule [Parlodel] | VAGINALLY |
| Insulin Glargine 100 UNT/ML Injectable Solution [Lantus] | sub q |
| insulin human, isophane 100 UNT/ML Injectable Suspension | injection |
| Glyburide 5 MG / Metformin hydrochloride 500 MG [Glucovance] | PO |
| 24 HR Glipizide 2.5 MG Extended Release Oral Tablet | inhalation |
| Gliclazide Oral Tablet | |
| Ultralente Insulin, Human 100 UNT/ML Injectable Suspension [Humulin U Ultralente] | subcutaneous |
| Glyburide 5 MG | ng |
| Metformin 1000 MG | bedside |
| Glipizide | Neb |
| 3 ML Insulin Glargine 100 UNT/ML Pen Injector [Lantus] | Drip |
| {60 (Regular Insulin, Human 4 UNT Inhalant Powder [Afrezza]) / 30 (Regular Insulin, Human 8 UNT Inhalant Powder [Afrezza]) } Pack [Afrezza 90 - 60 (4 UNT), 30 (8 UNT)] | INHALATION |
| sitagliptin 50 MG Oral Tablet [Januvia] | p.o |
| Metformin | g tube |

| | | |
|---|---|---|
| Metformin hydrochloride 1000 MG / saxagliptin 5 MG [Kombiglyze] | SEE COMMENT | |
| miglitol Oral Tablet [Glyset] | by mouth | |
| Metformin hydrochloride 1000 MG / sitagliptin 50 MG [Janumet] | IV | |
| Simvastatin 10 MG / sitagliptin 50 MG [Juvisync] | subcutaneously | |
| Insulin Lispro 100 UNT/ML Injectable Solution | IV | |
| insulin human, isophane 50 UNT/ML / Regular Insulin, Human 50 UNT/ML Injectable Suspension [Humulin] | subcu. | |
| Insulin Glargine | ADDITIVE | |
| Regular Insulin, Human 100 UNT/ML Injectable Solution [Humulin R] | Intravenous | |
| Lantus Injectable Product | drip. | |
| Amaryl Pill | nostril | |
| Insulin Glargine | sl | |
| Insulin, Aspart, Human 100 UNT/ML Cartridge [NovoLog] | SL | |
| Metformin hydrochloride 500 MG Oral Tablet [Glucophage] | by mouth. | |
| Linagliptin 2.5 MG / Metformin hydrochloride 500 MG Oral Tablet [Jentadueto] | | 0 |
| Glyburide 2.5 MG / Metformin hydrochloride 500 MG Oral Tablet [Glucovance] | SEE COMMENT | |
| Regular Insulin, Human | Subcut. | |
| Regular Insulin, Human 500 UNT/ML Injectable Solution [Humulin R] | VIA INSULIN PUMP | |
| miglitol 50 MG Oral Tablet [Glyset] | | |
| 3 ML liraglutide 6 MG/ML Pen Injector [Saxenda] | intramuscular | |
| insulin human, isophane 16 UNT/ML | subcu. | |
| Insulin Lispro 25 UNT/ML | Sub q | |
| 2.7 ML pramlintide acetate 1 MG/ML Pen Injector [Symlin] | | |
| 3 ML insulin human, isophane 100 UNT/ML Pen Injector [Humulin N] | SC | |
| alogliptin 12.5 MG | | |
| Insulin, Zinc, Pork | p.o. | |
| Glyburide Oral Tablet [Diabeta] | p.o. | |
| sitagliptin 100 MG [Januvia] | gt | |
| Jardiance Oral Product | | |
| exenatide 0.01 MG/ACTUAT [Byetta] | Subcu | |
| Insulin Lispro 100 UNT/ML Pen Injector [Humalog] | | 0 |
| Metformin 500 MG | Subcutaneous | |
| Insulin Glargine 100 UNT/ML [Lantus] | td | |
| 24 HR Glipizide 5 MG Extended Release Oral Tablet [Glucotrol] | gt | |

| | |
|---|---|
| Regular Insulin, Human 50 UNT/ML | per tube |
| Insulin, Aspart, Human 100 UNT/ML Injectable Solution [NovoLog] | sq |
| Bromocriptine 5 MG Oral Capsule | pO |
| Bromocriptine Oral Tablet [Cycloset] | PO |
| Glyburide 5 MG | nostril |
| Insulin, Regular, Pork 40 UNT/ML Injectable Solution | sub q |
| Insulin Lispro | inhaled |
| Tolazamide 100 MG Oral Tablet | p.o. |
| Insulin Lispro Injectable Solution | subcu |
| Insulin, Aspart, Human 100 UNT/ML Cartridge [NovoLog] | subcu. |
| Metformin hydrochloride 850 MG [Glucophage] | TOPICAL |
| dapagliflozin 10 MG [Farxiga] | nostril |
| Regular Insulin, Human 100 UNT/ML Injectable Solution [Novolin R] | po |
| nateglinide 60 MG Oral Tablet | BY MOUTH |
| Metformin hydrochloride 500 MG Oral Tablet [Glucophage] | gt |
| miglitol 50 MG Oral Tablet | PO |
| Chlorpropamide 125 MG Oral Tablet | p.o. |
| glimepiride | bedside. |
| Humalog Injectable Product | subcUTANEOUSLY |
| Glipizide 10 MG | npo. |
| Trulicity Injectable Product | SQ |
| Metformin / pioglitazone Oral Tablet [Actoplus Met] | orally |
| Linagliptin 2.5 MG | by mouth |
| glimepiride 4 MG | SQ |
| Insulin Lispro 100 UNT/ML Injectable Solution [Humalog] | THROUGH PUMP |
| Glipizide 5 MG | npo. |
| Regular Insulin, Human 100 UNT/ML Injectable Solution | po |
| dulaglutide 1.5 MG/ML [Trulicity] | SUBCUTANEOUS |
| Regular Insulin, Human | NG |
| Metformin hydrochloride 1000 MG / saxagliptin 2.5 MG [Kombiglyze] | PO |
| Humulin N Injectable Product | SUBCUTANEOUSLY |
| Glipizide 5 MG Extended Release Oral Tablet [Glucotrol] | PO |
| Insulin, Aspart, Human 100 UNT/ML Cartridge [NovoLog] | sl |
| glimepiride | im |
| Glyburide 2.5 MG [Diabeta] | per tube |
| insulin human, isophane 100 UNT/ML Injectable Suspension | PO |
| Metformin Oral Tablet | ORAL |

| | | |
|---|---|---|
| Metformin 1000 MG | ou | |
| 3 ML insulin human, isophane 100 UNT/ML Prefilled Syringe | subcutaneous | |
| Regular Insulin, Human 30 UNT/ML | subcu | |
| Acarbose | Gtube | |
| glimepiride 4 MG / rosiglitazone 8 MG Oral Tablet [Avandaryl] | | |
| Regular Insulin, Human | subcu. | |
| Regular Insulin, Human | UMBILICAL | |
| Glipizide 5 MG | subcutaneously | |
| insulin pork, isophane 70 UNT/ML / Insulin, Regular, Pork 30 UNT/ML Injectable Suspension | Subcutaneous | |
| 3 ML Regular Insulin, Human 100 UNT/ML Prefilled Syringe [Novolin R] | by mouth | |
| Insulin, Aspart, Human | Swallow | |
| Insulin Lispro Cartridge [Humalog] | subcutaneously. | |
| insulin human, isophane 100 UNT/ML Injectable Suspension | p.o. | |
| Glyburide 5 MG Oral Tablet [Diabeta] | Oral | |
| Fortamet Oral Product | | |
| Regular Insulin, Human 100 UNT/ML Injectable Solution [Novolin R] | CONT SUBCUTAN INFUSI | |
| Metformin hydrochloride 100 MG/ML Oral Solution [Riomet] | p.o. | |
| insulin human, isophane 70 UNT/ML / Regular Insulin, Human 30 UNT/ML [Novolin] | sublingual | |
| insulin human, isophane 70 UNT/ML / Regular Insulin, Human 30 UNT/ML [Novolin] | SUBCU | |
| insulin beef, isophane 40 UNT/ML | subcu | |
| 0.5 ML albiglutide 60 MG/ML Pen Injector [Tanzeum] | SC | |
| Glyburide 2.5 MG | by mouth. | |
| Metformin | Sublingual | |
| Insulin Lispro 25 UNT/ML / Insulin, Protamine Lispro, Human 75 UNT/ML [Humalog Mix] | | 0 |
| alogliptin 25 MG / pioglitazone 30 MG Oral Tablet [Oseni] | | |
| Saxenda Injectable Product | subQ | |
| Insulin Lispro 100 UNT/ML Injectable Solution [Humalog] | INFUSION PUMP | |
| insulin human, isophane 70 UNT/ML / Regular Insulin, Human 30 UNT/ML [Humulin] | npo. | |
| Glipizide 10 MG Oral Tablet [Glucotrol] | im | |
| Metformin hydrochloride 500 MG / sitagliptin 50 MG [Janumet] | PER TUBE | |
| dulaglutide 1.5 MG/ML [Trulicity] | po | |
| Insulin Lispro Injectable Solution [Humalog] | SUBCUTANEOUSLY | |
| Humulin R Injectable Product | nostril | |

| | |
|---|---|
| insulin human, isophane 50 UNT/ML / Regular Insulin, Human 50 UNT/ML [Humulin] | Subcutaneous |
| Insulin Lispro Cartridge [Humalog] | topical |
| insulin pork, isophane 80 UNT/ML | subcu. |
| Regular Insulin, Human 25 UNT/ML | po |
| Insulin Glargine 100 UNT/ML [Lantus] | OU |
| Regular Insulin, Human 100 UNT/ML Injectable Solution [Novolin R] | oral |
| 3 ML Insulin Glargine 100 UNT/ML Pen Injector [Basaglar] | oral |
| 3 ML Insulin, Aspart, Human 100 UNT/ML Cartridge [NovoLog] | pump |
| Insulin Lispro 25 UNT/ML | IV |
| Actos Oral Product | subcutaneous |
| 3 ML insulin detemir 100 UNT/ML Pen Injector [Levemir] | SUBCU |
| Insulin, Zinc, Human 100 UNT/ML Injectable Suspension [Humulin L] | subcu |
| insulin human, isophane 80 UNT/ML / Regular Insulin, Human 20 UNT/ML [Humulin] | sc. |
| Insulin, Aspart Protamine, Human 70 UNT/ML / Insulin, Aspart, Human 30 UNT/ML Injectable Suspension [NovoLog Mix] | injection |
| 3 ML liraglutide 6 MG/ML Pen Injector [Victoza] | per mouth |
| Regular Insulin, Human 25 UNT/ML | Subcutaneous |
| Insulin, Aspart, Human | J - tube |
| Metformin hydrochloride 500 MG / rosiglitazone 4 MG Oral Tablet | p.o. |
| Metformin | subcutaneously. |
| 3 ML Insulin Lispro 200 UNT/ML Pen Injector [Humalog] | oral |
| Lente Insulin, Beef-Pork 40 UNT/ML Injectable Suspension | |
| Metformin hydrochloride 1000 MG Oral Tablet | swallow |
| insulin human, isophane 16 UNT/ML | sub q |
| Insulin, Regular, Pork 40 UNT/ML Injectable Solution | subq |
| Acarbose 100 MG Oral Tablet | |
| Insulin Glargine Injectable Solution | sub Q |
| Insulin, Zinc, Human 100 UNT/ML Injectable Suspension [Novolin L] | po |
| Apidra Injectable Product | subq |
| 24 HR Glipizide 10 MG Extended Release Oral Tablet [Glucotrol] | subcutaneously |
| Insulin, Zinc, Pork | subcutaneous |
| insulin human, isophane 80 UNT/ML / Regular Insulin, Human 20 UNT/ML [Humulin] | drip. |

| | |
|---|---|
| Regular Insulin, Human 100 UNT/ML Injectable Solution [Humulin R] | SUBQ |
| rosiglitazone 2 MG Oral Tablet [Avandia] | Oral |
| Glyburide 1.25 MG | IV |
| 24 HR Glipizide 10 MG Extended Release Oral Tablet [Glucotrol] | gt |
| Simvastatin 10 MG / sitagliptin 100 MG Oral Tablet [Juvisync] | by mouth |
| Bromocriptine 2.5 MG Oral Tablet [Parlodel] | Po |
| empagliflozin Oral Tablet | oral |
| nateglinide | subcutaneously |
| Glyburide 5 MG | subcu |
| empagliflozin 25 MG / Linagliptin 5 MG Oral Tablet [Glyxambi] | PO |
| Regular Insulin, Human | ADDITIVE |
| Regular Insulin, Human 100 UNT/ML Injectable Solution | |
| insulin human, isophane 50 UNT/ML / Regular Insulin, Human 50 UNT/ML Injectable Suspension [Humulin] | SC |
| Glyburide 5 MG Oral Tablet [Daonil] | orally |
| Insulin, Aspart, Human 100 UNT/ML Injectable Solution | sc |
| liraglutide 6 MG/ML [Victoza] | Subcutaneous |
| sitagliptin 50 MG Oral Tablet | oral |
| Metformin Extended Release Oral Tablet | BY MOUTH |
| insulin pork, isophane 100 UNT/ML Injectable Suspension [NPH Iletin II] | IV. |
| insulin human, isophane 70 UNT/ML / Regular Insulin, Human 30 UNT/ML [Novolin] | drip |
| Linagliptin 5 MG [Tradjenta] | SUBQ |
| Metformin hydrochloride 500 MG [Glucophage] | ou |
| Metformin | sl. |
| Metformin hydrochloride 1000 MG / saxagliptin 2.5 MG [Kombiglyze] | Oral |
| rosiglitazone 4 MG [Avandia] | ng |
| insulin human, isophane 75 UNT/ML | oral |
| Metformin hydrochloride 1000 MG / saxagliptin 5 MG Extended Release Oral Tablet [Kombiglyze] | |
| Insulin Lispro 100 UNT/ML Injectable Solution [Humalog] | PER PUMP |
| Insulin Glargine Injectable Solution | IM |
| Glyburide / Metformin Oral Tablet [Glucovance] | p.o. |
| Bromocriptine 0.8 MG Oral Tablet [Cycloset] | ORAL |
| Metformin 400 MG | oral |
| exenatide 0.005 MG/ACTUAT [Byetta] | BY MOUTH |
| Duetact Oral Product | p.o. |

| | | |
|---|---|---|
| Regular Insulin, Human 30 UNT/ML | sub q | |
| Insulin, Aspart, Human | DRIP | |
| Metformin hydrochloride 1000 MG [Glucophage] | topical | |
| Insulin, Aspart, Human 100 UNT/ML Cartridge [NovoLog] | ORAL | |
| Regular Insulin, Human 8 UNT | orally | |
| Insulin, Aspart Protamine, Human 70 UNT/ML / Insulin, Aspart, Human 30 UNT/ML Injectable Suspension [NovoLog Mix] | MISCELLANEOUS | |
| insulin human, isophane 70 UNT/ML / Regular Insulin, Human 30 UNT/ML [Humulin] | SEE COMMENT | |
| Glipizide | iv | |
| Toujeo Injectable Product | SEE COMMENT | |
| Metformin Oral Solution | By mouth | |
| Metformin hydrochloride 750 MG Extended Release Oral Tablet | nostril | |
| Lixisenatide | subq | |
| Metformin | Inhalation | |
| dulaglutide 1.5 MG/ML [Trulicity] | SUBCUTANEOUSLY | |
| 3 ML Insulin, Aspart Protamine, Human 70 UNT/ML / Insulin, Aspart, Human 30 UNT/ML Pen Injector [NovoLog Mix] | SUBCUT | |
| Metformin hydrochloride 500 MG Extended Release Oral Tablet | swallow | |
| Metformin | MISCELLANEOUS | |
| Insulin, Aspart, Human 100 UNT/ML Injectable Solution [NovoLog] | SUBCUTANEOUS PUMP | |
| glimepiride 1 MG [Amaryl] | NPO | |
| 3 ML insulin human, isophane 70 UNT/ML / Regular Insulin, Human 30 UNT/ML Pen Injector | SQ | |
| Insulin Glargine Injectable Solution | per tube | |
| exenatide 3.08 MG/ML Pen Injector [Bydureon] | SQ | |
| Glipizide 2.5 MG Extended Release Oral Tablet [Glucotrol] | p.o. | |
| Metformin Oral Tablet | gt | |
| Metformin / sitagliptin Oral Tablet [Janumet] | po | |
| Afrezza Inhalant Product | inhalation | |
| 3 ML Insulin, Aspart Protamine, Human 70 UNT/ML / Insulin, Aspart, Human 30 UNT/ML Pen Injector [NovoLog Mix] | im | |
| Metformin hydrochloride 1000 MG Extended Release Oral Tablet [Glumetza] | Po | |
| saxagliptin 2.5 MG Oral Tablet [Onglyza] | | 0 |
| Metformin hydrochloride 100 MG/ML | nostril | |
| Insulin, Aspart, Human | topical | |
| Metformin 1000 MG | sub q | |
| Regular Insulin, Human 12 UNT Inhalant Powder [Afrezza] | | |

| | | |
|---|---|---|
| Victoza Injectable Product | oral | |
| pioglitazone | inh | |
| Bromocriptine Oral Tablet [Parlodel] | po | |
| Insulin Glargine 300 UNT/ML Pen Injector | subcut | |
| saxagliptin Oral Tablet | | |
| Metformin hydrochloride 750 MG | IM | |
| Tolazamide 500 MG | | |
| Insulin Lispro 100 UNT/ML Injectable Solution | by mouth | |
| Glyburide 2.5 MG [Diabeta] | nostril | |
| Regular Insulin, Human 100 UNT/ML Injectable Solution | SUBQ | |
| Metformin 500 MG | Sq | |
| Insulin Lispro | NG | |
| Regular Insulin, Human 100 UNT/ML Injectable Solution | oral | |
| glimepiride 2 MG / pioglitazone 30 MG Oral Tablet [Duetact] | | |
| Metformin | TD | |
| glimepiride | TD | |
| Insulin Lispro Cartridge [Humalog] | npo | |
| exenatide 0.01 MG/ACTUAT [Byetta] | BY MOUTH | |
| Metformin hydrochloride 850 MG Oral Tablet [Glucophage] | | 0 |
| Insulin Glargine 100 UNT/ML Injectable Solution | subcu | |
| Regular Insulin, Human 20 UNT/ML | subcu | |
| Insulin Glargine Injectable Solution [Lantus] | SUBCUT | |
| 3 ML Insulin Glargine 100 UNT/ML Pen Injector [Lantus] | sub Q | |
| glimepiride 6 MG | subcutaneous | |
| Metformin hydrochloride 500 MG / sitagliptin 50 MG Oral Tablet | PER TUBE | |
| Insulin Glargine Injectable Solution | TD | |
| sitagliptin 100 MG [Januvia] | OU | |
| Metformin hydrochloride 500 MG / rosiglitazone 2 MG [Avandamet] | inhalation | |
| Glyburide 5 MG Oral Tablet [Diabeta] | gt | |
| 1.5 ML Insulin Glargine 300 UNT/ML Pen Injector [Toujeo] | by mouth | |
| canagliflozin 300 MG Oral Tablet | po | |
| Glipizide / Metformin Oral Tablet | oral | |
| Insulin, Aspart, Human 100 UNT/ML Injectable Solution [NovoLog] | VIA PUMP | |
| Chlorpropamide 100 MG Oral Tablet | orally | |
| 1.5 ML insulin human, isophane 70 UNT/ML / Regular Insulin, Human 30 UNT/ML Prefilled Syringe | Oral | |
| Glyburide | iv | |
| Insulin Glargine Injectable Solution | SUBCUTANEOUSLY | |

| | | |
|---|---|---|
| glimepiride 2 MG | | 0 |
| Metformin hydrochloride 1000 MG / sitagliptin 50 MG Oral Tablet [Janumet] | PO. | |
| alogliptin | gt | |
| Metformin hydrochloride 1000 MG Oral Tablet [Glucophage] | PER TUBE | |
| exenatide 0.01 MG/ACTUAT [Byetta] | Subcutaneously | |
| 24 HR Metformin hydrochloride 1000 MG / pioglitazone 30 MG Extended Release Oral Tablet [Actoplus Met] | ORAL | |
| Lente Insulin, Beef 80 UNT/ML | subcut. | |
| Glyburide 2.5 MG Oral Tablet [Diabeta] | ORAL | |
| Glyburide 6 MG | nostril | |
| Glyburide Oral Tablet [Diabeta] | PO | |
| Metformin hydrochloride 500 MG [Glucophage] | per mouth | |
| Metformin hydrochloride 500 MG / sitagliptin 50 MG Extended Release Oral Tablet [Janumet] | | |
| Insulin, Regular, Pork 40 UNT/ML Injectable Solution | po | |
| Insulin, Aspart Protamine, Human 70 UNT/ML / Insulin, Aspart, Human 30 UNT/ML Injectable Suspension [NovoLog Mix] | subcu. | |
| dulaglutide 1.5 MG/ML [Trulicity] | SUBCU | |
| Glipizide | Subcutaneous | |
| exenatide 2 MG Injection [Bydureon] | sub q | |
| pioglitazone | TOPICAL | |
| Regular Insulin, Human 20 UNT/ML | subq | |
| insulin human, isophane 90 UNT/ML / Regular Insulin, Human 10 UNT/ML [Humulin] | p.o. | |
| exenatide 0.005 MG/ACTUAT [Byetta] | ou | |
| insulin detemir 100 UNT/ML [Levemir] | | 0 |
| insulin human, isophane 70 UNT/ML / Regular Insulin, Human 30 UNT/ML [Novolin] | Sublingual | |
| Glipizide 5 MG | NPO. | |
| exenatide 2 MG [Bydureon] | subcu. | |
| Insulin Lispro Injectable Solution | by mouth | |
| Metformin hydrochloride 500 MG Oral Tablet | po. | |
| Regular Insulin, Human 10 UNT/ML | sub q | |
| Insulin, Glulisine, Human 100 UNT/ML Injectable Solution [Apidra] | INTRAVENOUS | |
| Actoplus Met Oral Product | p.o. | |
| Tolinase Oral Product | ORAL | |
| Insulin Glargine 300 UNT/ML [Toujeo] | subcutaneous | |

| | | |
|---|---|---|
| Insulin, Aspart, Human 100 UNT/ML Injectable Solution [NovoLog] | IM | |
| Levemir Injectable Product | | 0 |
| Xigduo Oral Product | p.o. | |
| insulin pork, isophane 100 UNT/ML Injectable Suspension [NPH Iletin II] | intravenous | |
| Insulin, Regular, Pork 100 UNT/ML Injectable Solution | subcutaneous | |
| Metformin hydrochloride 500 MG Oral Tablet | bY mouth | |
| insulin human, isophane 70 UNT/ML / Regular Insulin, Human 30 UNT/ML [Humulin] | Oral | |
| insulin degludec 200 UNT/ML | per tube | |
| Glyburide 5 MG | Im | |
| nateglinide Oral Product | p.o. | |
| rosiglitazone 4 MG Oral Tablet | P.O. | |
| 3 ML insulin degludec 100 UNT/ML / liraglutide 3.6 MG/ML Pen Injector [Xultophy] | injection | |
| Regular Insulin, Human 100 UNT/ML Injectable Solution | subcu | |
| Bromocriptine | gt | |
| Insulin, Aspart, Human 100 UNT/ML Injectable Solution [NovoLog] | PER INSULIN PUMP DAILY | |
| 24 HR dapagliflozin 5 MG / Metformin hydrochloride 500 MG Extended Release Oral Tablet [Xigduo] | ORAL | |
| Bromocriptine | subcutaneous | |
| Linagliptin 5 MG Oral Tablet | Oral | |
| 0.65 ML exenatide 3.08 MG/ML Pen Injector [Bydureon] | intramuscular | |
| 24 HR dapagliflozin 10 MG / Metformin hydrochloride 500 MG Extended Release Oral Tablet [Xigduo] | orally | |
| Glipizide 2.5 MG / Metformin hydrochloride 250 MG Oral Tablet | p.o. | |
| 3 ML Insulin Glargine 100 UNT/ML Pen Injector | | |
| Novolin R Injectable Product | sq | |
| Metformin Oral Solution | | 0 |
| 24 HR Metformin hydrochloride 1000 MG / saxagliptin 5 MG Extended Release Oral Tablet | | |
| saxagliptin 2.5 MG Oral Tablet [Onglyza] | p.o. | |
| Insulin, Aspart, Human 100 UNT/ML Cartridge [NovoLog] | MISCELLANEOUS | |
| Regular Insulin, Human 100 UNT/ML Injectable Solution [Humulin S] | subcut | |
| Glipizide | inhaled | |
| sitagliptin 100 MG [Januvia] | SQ | |
| Metformin hydrochloride 500 MG Oral Tablet | NG | |

| | |
|---|---|
| glimepiride 4 MG [Amaryl] | by mouth. |
| Metformin hydrochloride 1000 MG [Glucophage] | intramuscular |
| Metformin 250 MG Oral Tablet | ORAL |
| Bromocriptine Oral Capsule | swallow |
| glimepiride 4 MG Oral Tablet | PER TUBE |
| insulin human, isophane 16 UNT/ML | p.o. |
| Insulin, Aspart, Human | ng |
| 0.5 ML albiglutide 60 MG/ML Pen Injector [Tanzeum] | Sq |
| Glyburide Oral Tablet [Micronase] | p.o. |
| insulin human, rDNA origin 1 MG/ACTUAT Inhalant Powder | inhalatn |
| Glyburide | orally |
| Metformin hydrochloride 500 MG Oral Tablet | Gtube. |
| Glyburide 2.5 MG | drip. |
| Glipizide 2.5 MG | orally. |
| Acarbose 50 MG | subcutaneously |
| Regular Insulin, Human 100 UNT/ML Injectable Solution [Humulin R] | INTRAMUSCULAR |
| Humulin N Injectable Product | subcut |
| Metformin hydrochloride 500 MG / sitagliptin 50 MG Oral Tablet [Janumet] | gt |
| sitagliptin 100 MG [Januvia] | subq |
| Metformin hydrochloride 850 MG | sl |
| Metformin 500 MG | ORAL |
| Glyburide 5 MG | orally. |
| Metformin hydrochloride 250 MG | |
| Regular Insulin, Human | sub q. |
| Insulin Lispro 25 UNT/ML | gt |
| Glyburide Oral Tablet | ORAL |
| Insulin Lispro Injectable Solution | subq |
| Tolbutamide | IV |
| Metaglip Oral Product | p.o. |
| Metformin hydrochloride 1000 MG / saxagliptin 5 MG [Kombiglyze] | gt |
| dapagliflozin 10 MG [Farxiga] | sq |
| Metformin 500 MG | Sublingual |
| Insulin Lispro 100 UNT/ML Injectable Solution [Humalog] | SQ |
| Insulin, Glulisine, Human 100 UNT/ML Injectable Solution [Apidra] | SEE COMMENT |
| Acetohexamide | PO |
| 24 HR Glipizide 5 MG Extended Release Oral Tablet [Glucotrol] | po |

| | |
|---|---|
| Metformin hydrochloride 100 MG/ML [Riomet] | by mouth. |
| Insulin, Glulisine, Human | subcu |
| 3 ML Insulin, Aspart, Human 100 UNT/ML Pen Injector [NovoLog] | intramuscular |
| exenatide 0.005 MG/ACTUAT [Byetta] | subcutaneous. |
| insulin detemir 100 UNT/ML Injectable Solution [Levemir] | Oral |
| Metformin hydrochloride 1000 MG / pioglitazone 30 MG Extended Release Oral Tablet | oral |
| liraglutide 6 MG/ML [Victoza] | IM |
| insulin human, isophane 90 UNT/ML / Regular Insulin, Human 10 UNT/ML [Humulin] | subcu |
| Glyburide 5 MG / Metformin hydrochloride 500 MG Oral Tablet [Glucovance] | P.O. |
| Metformin 1000 MG | subcu |
| Humalog Injectable Product | SUBCUTANEOUS |
| Insulin Lispro 25 UNT/ML / Insulin, Protamine Lispro, Human 75 UNT/ML Injectable Suspension [Humalog Mix] | sub Q |
| Glyburide | Swallow. |
| Insulin, Aspart, Human | topically |
| insulin pork, isophane 100 UNT/ML Injectable Suspension [NPH Iletin II] | iv. |
| Glipizide | INH |
| Regular Insulin, Human 6 UNT/ML | SUBCUT |
| 24 HR Glipizide 5 MG Extended Release Oral Tablet [Glucotrol] | BY MOUTH |
| dapagliflozin / Metformin Pill | |
| Glipizide 2.5 MG [Glucotrol] | ORAL |
| Metformin hydrochloride 750 MG Extended Release Oral Tablet | by mouth |
| Insulin Glargine 300 UNT/ML | gt |
| Insulin Lispro | Gtube |
| 3 ML Insulin, Aspart Protamine, Human 70 UNT/ML / Insulin, Aspart, Human 30 UNT/ML Pen Injector [NovoLog Mix] | abdominal subcutaneous |
| glimepiride 2 MG Oral Tablet [Amaryl] | per tube |
| Dymelor Oral Product | PO |
| glimepiride 2 MG | topical |
| Metformin hydrochloride 500 MG / sitagliptin 50 MG [Janumet] | PO |
| pioglitazone 15 MG [Actos] | IV |
| Tanzeum Injectable Product | SEE COMMENT |
| saxagliptin Pill | |
| insulin human, isophane 70 UNT/ML / Regular Insulin, Human 30 UNT/ML [Humulin] | Topical |
| Glipizide 2.5 MG Oral Tablet | P.O. |

| | |
|---|---|
| Linagliptin 5 MG [Tradjenta] | IV |
| Metformin hydrochloride 500 MG Extended Release Oral Tablet [Glumetza] | subcutaneously |
| Insulin, Aspart, Human 100 UNT/ML Injectable Solution [NovoLog] | MISC |
| Insulin Lispro 100 UNT/ML Injectable Solution | sq |
| Regular Insulin, Human | topically |
| Victoza Injectable Product | subcutaneously. |
| Metformin hydrochloride 750 MG | NPO |
| Acarbose 100 MG Oral Tablet [Precose] | Oral |
| Glyburide 2.5 MG / Phenformin 25 MG Oral Tablet | oral |
| glimepiride 1 MG Oral Tablet [Amaryl] | Po |
| insulin human, isophane 100 UNT/ML Injectable Suspension [Novolin N] | nostril |
| sitagliptin Oral Tablet [Januvia] | p.o. |
| Insulin Lispro 25 UNT/ML / Insulin, Protamine Lispro, Human 75 UNT/ML [Humalog Mix] | Sq |
| Riomet Oral Product | Oral |
| Glipizide 5 MG | td |
| Glipizide 2.5 MG | npo |
| Metformin Oral Solution | By Mouth |
| sitagliptin 25 MG Oral Tablet | p.o. |
| sitagliptin 100 MG [Januvia] | IM |
| insulin human, isophane 36 UNT/ML | sq |
| Acarbose | gt |
| insulin human, isophane 90 UNT/ML | SUB Q |
| empagliflozin 25 MG | gt |
| 3 ML insulin detemir 100 UNT/ML Pen Injector [Levemir] | subconjunctival |
| Glipizide | Swish and spit |
| Avandia Oral Product | Po |
| 1.5 ML pramlintide acetate 1 MG/ML Pen Injector [Symlin] | subcu |
| 3 ML insulin degludec 100 UNT/ML Pen Injector [Tresiba] | subdermal |
| Insulin Lispro 25 UNT/ML / Insulin, Protamine Lispro, Human 75 UNT/ML [Humalog Mix] | subQ. |
| nateglinide 60 MG Oral Tablet [Starlix] | P.O. |
| insulin human, isophane 70 UNT/ML / Regular Insulin, Human 30 UNT/ML [Novolin] | NG |
| rosiglitazone 8 MG [Avandia] | by mouth. |
| sitagliptin 25 MG Oral Tablet | oral |
| Insulin Glargine Injectable Solution [Lantus] | oral |

| | |
|---|---|
| 60 ACTUAT exenatide 0.01 MG/ACTUAT Pen Injector [Byetta] | subcutaneously. |
| 0.5 ML albiglutide 60 MG/ML Pen Injector | subcutaneous |
| Insulin Lispro 100 UNT/ML Injectable Solution [Humalog] | oral |
| pramlintide acetate 0.6 MG/ML | |
| Insulin Lispro | SUBcut |
| Insulin Lispro 25 UNT/ML / Insulin, Protamine Lispro, Human 75 UNT/ML [Humalog Mix] | po |
| Regular Insulin, Human 500 UNT/ML Injectable Solution | by mouth |
| alogliptin 25 MG / pioglitazone 15 MG [Oseni] | nostril |
| Ultralente Insulin, Human 100 UNT/ML Injectable Suspension | orally |
| repaglinide 1 MG Oral Tablet [Prandin] | po. |
| Insulin Lispro 25 UNT/ML / Insulin, Protamine Lispro, Human 75 UNT/ML Injectable Suspension [Humalog Mix] | im |
| Glyburide 6 MG Oral Tablet | Oral |
| Lantus Injectable Product | Subcut |
| Regular Insulin, Human 4 UNT | iv |
| Regular Insulin, Human | As_Directed |
| empagliflozin 25 MG / Linagliptin 5 MG Oral Tablet [Glyxambi] | p.o. |
| Regular Insulin, Human 100 UNT/ML Injectable Solution | per tube |
| Regular Insulin, Human 100 UNT/ML Injectable Solution [Humulin R] | per tube |
| Metformin hydrochloride 1000 MG / pioglitazone 30 MG Extended Release Oral Tablet | by mouth |
| Insulin Lispro 200 UNT/ML [Humalog] | subcutaneously. |
| Regular Insulin, Human 100 UNT/ML Injectable Solution [Humulin S] | sub q |
| Simvastatin 10 MG / sitagliptin 100 MG [Juvisync] | gt |
| Glipizide 10 MG Extended Release Oral Tablet | nostril |
| 3 ML insulin detemir 100 UNT/ML Pen Injector [Levemir] | po |
| NovoLog Injectable Product | subcutaneousLy |
| exenatide 0.01 MG/ACTUAT | NPO |
| Bromocriptine Oral Tablet | |
| Glipizide Oral Product | ORAL |
| insulin human, isophane 60 UNT/ML | PO |
| Metformin hydrochloride 500 MG Extended Release Oral Tablet | subcutaneously |
| Insulin, Aspart, Human 100 UNT/ML Cartridge [NovoLog] | p.o. |
| rosiglitazone | NG |
| insulin detemir 100 UNT/ML Injectable Solution | subQ |
| 3 ML Insulin Lispro 100 UNT/ML Pen Injector [Humalog] | intramuscular |
| glimepiride 2 MG [Amaryl] | subcutaneous |

| | |
|---|---|
| Insulin, Aspart, Human 100 UNT/ML Injectable Solution [NovoLog] | VIA INSULIN PUMP AS DIRECTED |
| Glucophage Oral Product | P.O. |
| glimepiride 2 MG | sublingual |
| pioglitazone | SQ |
| liraglutide | MISCELLANEOUS |
| 0.5 ML dulaglutide 1.5 MG/ML Auto-Injector | subcutaneous |
| exenatide | p.o. |
| repaglinide 2 MG Oral Tablet [Prandin] | BY MOUTH |
| dapagliflozin 5 MG Oral Tablet [Farxiga] | Oral |
| sitagliptin 50 MG [Januvia] | SEE COMMENT |
| Linagliptin 2.5 MG / Metformin hydrochloride 500 MG Oral Tablet [Jentadueto] | PO |
| 60 ACTUAT exenatide 0.01 MG/ACTUAT Pen Injector [Byetta] | SUBCU |
| Regular Insulin, Human 100 UNT/ML Injectable Solution [Velosulin] | po |
| 0.5 ML dulaglutide 1.5 MG/ML Auto-Injector | SQ |
| Metformin hydrochloride 1000 MG Oral Tablet | per tube. |
| Metformin hydrochloride 1000 MG / sitagliptin 50 MG [Janumet] | PER TUBE |
| Prandin Oral Product | Po |
| Insulin, Aspart, Human 100 UNT/ML Injectable Solution [NovoLog] | SUBCUTANEOUS PER INSULIN PUMP |
| insulin human, isophane 75 UNT/ML | subcu. |
| alogliptin 6.25 MG Oral Tablet [Nesina] | ORAL |
| Acarbose | by mouth. |
| Metformin 250 MG | Im |
| Insulin, Glulisine, Human 100 UNT/ML Injectable Solution | Subcutaneously |
| Glyburide 5 MG [Daonil] | |
| Levemir Injectable Product | po |
| Regular Insulin, Human 15 UNT/ML | po |
| 3 ML Insulin Glargine 100 UNT/ML Pen Injector [Lantus] | p.o. |
| Metformin hydrochloride 1000 MG / sitagliptin 50 MG [Janumet] | OU |
| 3 ML Insulin Lispro 100 UNT/ML Pen Injector [Humalog] | oral |
| rosiglitazone 4 MG | PER TUBE |
| alogliptin 25 MG Oral Tablet | |
| miglitol 100 MG Oral Tablet [Glyset] | p.o. |
| liraglutide 6 MG/ML [Victoza] | SEE COMMENT |
| 60 ACTUAT exenatide 0.005 MG/ACTUAT Pen Injector [Byetta] | subcutaneous. |
| Glyburide 2.5 MG | Per tube |
| Metformin hydrochloride 1000 MG Oral Tablet | swallow. |

| | |
|---|---|
| Metformin / rosiglitazone Oral Tablet [Avandamet] | orally |
| Regular Insulin, Human 5 UNT/ML | subcu |
| Lantus Injectable Product | SEE COMMENT |
| glimepiride | oral |
| Regular Insulin, Human 15 UNT/ML | subq |
| dapagliflozin 10 MG / Metformin hydrochloride 1000 MG [Xigduo] | orally |
| Glyburide | bedside. |
| insulin human, isophane 60 UNT/ML / Regular Insulin, Human 40 UNT/ML [Humulin] | sc |
| liraglutide 6 MG/ML [Victoza] | gt |
| insulin human, isophane 30 UNT/ML | subcut |
| Metformin hydrochloride 100 MG/ML Oral Solution | per tube |
| exenatide 0.005 MG/ACTUAT [Byetta] | sc. |
| Glyburide 6 MG | swallow |
| Insulin Glargine 300 UNT/ML | per tube |
| Glipizide | sub q |
| 3 ML insulin human, isophane 100 UNT/ML Prefilled Syringe | oral |
| pioglitazone 30 MG [Actos] | gt |
| insulin pork, isophane 100 UNT/ML Injectable Suspension [NPH Iletin II] | nostril |
| Glipizide 5 MG | oral. |
| exenatide 0.01 MG/ACTUAT [Byetta] | subcutaneously. |
| exenatide 2 MG [Bydureon] | SUBCU |
| Amaryl Pill | oral |
| Metformin hydrochloride 1000 MG Oral Tablet | per mouth |
| 3 ML Insulin, Aspart Protamine, Human 70 UNT/ML / Insulin, Aspart, Human 30 UNT/ML Pen Injector [NovoLog Mix] | Sub Q |
| miglitol 100 MG Oral Tablet | PO |
| Glipizide 5 MG | Im |
| 24 HR dapagliflozin 10 MG / Metformin hydrochloride 500 MG Extended Release Oral Tablet [Xigduo] | PO |
| 3 ML liraglutide 6 MG/ML Pen Injector [Victoza] | inhalation |
| glimepiride 2 MG Oral Tablet [Amaryl] | P.o. |
| Metformin hydrochloride 1000 MG [Glumetza] | PO |
| Lispro PRC Injectable Product | SUBCUT |
| insulin human, isophane 75 UNT/ML | sub Q |
| Byetta Injectable Product | PO |
| Insulin Glargine | ng |

| | | |
|---|---|---|
| insulin pork, isophane 100 UNT/ML Injectable Suspension [NPH Iletin II] | orally. | |
| Metformin hydrochloride 750 MG | subcutaneous | |
| Lantus Injectable Product | po | |
| albiglutide 100 MG/ML | | |
| nateglinide 60 MG [Starlix] | IV | |
| 2.7 ML pramlintide acetate 1 MG/ML Pen Injector [Symlin] | | 0 |
| Insulin Glargine Injectable Solution [Lantus] | Gtube | |
| exenatide 0.005 MG/ACTUAT [Byetta] | SQ. | |
| Regular Insulin, Human 100 UNT/ML Injectable Solution [Novolin R] | INFUSION | |
| canagliflozin 150 MG / Metformin hydrochloride 500 MG Oral Tablet [Invokamet] | | 0 |
| exenatide 2 MG [Bydureon] | SUBQ | |
| Metformin Extended Release Oral Tablet [Fortamet] | orally | |
| Glyburide 5 MG / Metformin hydrochloride 500 MG Oral Tablet [Glucovance] | Oral | |
| Metformin hydrochloride 850 MG Oral Tablet | p.o | |
| Metformin hydrochloride 1000 MG / saxagliptin 2.5 MG [Kombiglyze] | gt | |
| Metformin Extended Release Oral Tablet | NPO. | |
| Regular Insulin, Human 100 UNT/ML Injectable Solution [Novolin R] | CONTINUOUS INFUSION | |
| insulin human, isophane 75 UNT/ML | p.o. | |
| insulin human, isophane 70 UNT/ML / Regular Insulin, Human 30 UNT/ML Injectable Suspension [Humulin] | SUBCUTANEOUSLY | |
| Metformin hydrochloride 1000 MG / rosiglitazone 2 MG Oral Tablet [Avandamet] | Oral | |
| empagliflozin 10 MG Oral Tablet | oral | |
| Linagliptin 5 MG [Tradjenta] | nostril | |
| Metformin hydrochloride 1000 MG / sitagliptin 50 MG [Janumet] | subq | |
| glimepiride | | 0 |
| Gliclazide 80 MG | | |
| glimepiride 2 MG [Amaryl] | per mouth | |
| 24 HR Metformin hydrochloride 750 MG Extended Release Oral Tablet | | 0 |
| Insulin Lispro 100 UNT/ML Injectable Solution [Humalog] | MP | |
| Duetact Pill | | |
| 3 ML insulin human, isophane 100 UNT/ML Prefilled Syringe | npo | |
| Metformin hydrochloride 1000 MG Oral Tablet | sublingual | |

| | |
|---|---|
| Insulin, Glulisine, Human 100 UNT/ML Injectable Solution [Apidra] | SUBCUTANEOUS PER INSULIN PUMP |
| Bromocriptine Extended Release Oral Capsule | by mouth |
| Insulin Glargine 100 UNT/ML Injectable Solution | subc. |
| insulin degludec 200 UNT/ML | p.o. |
| Insulin, Zinc, Human 100 UNT/ML Injectable Suspension [Novolin L] | sc |
| Regular Insulin, Human 500 UNT/ML Injectable Solution [Humulin R] | p.o. |
| nateglinide 90 MG Oral Tablet | by mouth |
| Metformin hydrochloride 1000 MG Oral Tablet [Glucophage] | p.o |
| dulaglutide Injectable Product | |
| miglitol 100 MG [Glyset] | per tube |
| Tolazamide 250 MG Oral Tablet | |
| Glipizide 10 MG Oral Tablet [Glucotrol] | Po |
| Levemir Injectable Product | COMBO ROUTE) |
| Glyburide 2.5 MG | bedside |
| Linagliptin 2.5 MG / Metformin hydrochloride 500 MG [Jentadueto] | PO |
| Metformin hydrochloride 1000 MG Extended Release Oral Tablet [Glumetza] | BY MOUTH |
| sitagliptin 100 MG [Januvia] | orally. |
| Glyburide 2.5 MG / Metformin hydrochloride 500 MG Oral Tablet | p.o. |
| Metformin Extended Release Oral Tablet | ORAL |
| Insulin Lispro Injectable Solution [Humalog] | MISCELLANEOUS |
| Metformin hydrochloride 750 MG | IV |
| Metformin 500 MG | G - Tube |
| glimepiride Oral Tablet | p.o. |
| Glyburide | MISCELLANEOUS |
| glimepiride Oral Product | Po |
| Glumetza Pill | by mouth |
| 3 ML liraglutide 6 MG/ML Pen Injector [Saxenda] | Subcutaneous |
| Metformin 500 MG | orally. |
| rosiglitazone 8 MG | per tube |
| sitagliptin 50 MG | per tube |
| Insulin Lispro 200 UNT/ML Pen Injector [Humalog] | SubQ |
| nateglinide 120 MG [Starlix] | by mouth. |
| Glipizide 10 MG Oral Tablet [Glucotrol] | p.o |
| Insulin, Glulisine, Human 100 UNT/ML Injectable Solution | subcutaneously |
| Metformin | INH. |

| | | |
|---|---|---|
| Acarbose | Ng | |
| 3 ML Regular Insulin, Human 500 UNT/ML Pen Injector [Humulin R] | po | |
| Bromocriptine | NG | |
| Metformin hydrochloride 500 MG Oral Tablet [Glucophage] | NG | |
| Metformin Oral Tablet [Glucophage] | | 0 |
| 0.5 ML dulaglutide 1.5 MG/ML Auto-Injector [Trulicity] | orally | |
| 24 HR Metformin hydrochloride 500 MG Extended Release Oral Tablet [Glucophage] | inhalation | |
| glimepiride | sc | |
| Insulin Lispro Injectable Solution | sub q | |
| pioglitazone | td | |
| 1.5 ML Insulin Glargine 300 UNT/ML Pen Injector [Toujeo] | subconjunctival | |
| Humulin R Injectable Product | SUBCUTANEOUSLY | |
| pioglitazone 15 MG | sublingual | |
| Glyburide 5 MG / Metformin hydrochloride 500 MG Oral Tablet | SEE COMMENT | |
| 60 ACTUAT exenatide 0.01 MG/ACTUAT Pen Injector [Byetta] | injection | |
| Metformin Pill | gt | |
| Insulin Lispro 50 UNT/ML | sl | |
| Glyburide 5 MG Oral Tablet | by mouth. | |
| Regular Insulin, Human | bedside | |
| Insulin Lispro 100 UNT/ML Injectable Solution [Humalog] | SUBCUTANEOUSLY | |
| insulin human, isophane 16 UNT/ML / Regular Insulin, Human 24 UNT/ML Injectable Suspension | | |
| Insulin Lispro 100 UNT/ML | iv | |
| Acarbose | subcut | |
| Metformin hydrochloride 100 MG/ML Oral Solution [Riomet] | g-tube | |
| Glyburide | sub q | |
| Glyburide 4.5 MG Oral Tablet | orally | |
| glimepiride 4 MG [Amaryl] | drip | |
| exenatide 2 MG Injection [Bydureon] | SUBCUTANEOUSLY | |
| Actoplus Met Pill | orally | |
| rosiglitazone 2 MG Oral Tablet [Avandia] | PER TUBE | |
| Glipizide | sc | |
| Chlorpropamide 250 MG Oral Tablet | | |
| dulaglutide 1.5 MG/ML [Trulicity] | Sq | |
| canagliflozin | intramuscular | |
| Glucophage Pill | p.o. | |
| Insulin Glargine | Nares | |

| | |
|---|---|
| insulin pork, isophane 100 UNT/ML Injectable Suspension [NPH Iletin II] | SC. |
| Glyburide | MISC |
| Regular Insulin, Human 5 UNT/ML | gt |
| exenatide 0.01 MG/ACTUAT [Byetta] | im |
| insulin pork, isophane 100 UNT/ML Injectable Suspension [NPH Iletin II] | subQ. |
| Insulin, Zinc, Pork | subQ |
| Metformin 1000 MG | NPO. |
| nateglinide 90 MG | |
| insulin, isophane Injectable Suspension [Novolin N] | SQ |
| repaglinide Oral Tablet | |
| Lente Insulin, Beef 80 UNT/ML Injectable Suspension | subcutaneously |
| insulin pork, isophane 100 UNT/ML Injectable Suspension [NPH Iletin II] | Sc |
| Glyburide Pill | inhalation |
| Osmotic 24 HR Metformin hydrochloride 1000 MG Extended Release Oral Tablet [Fortamet] | BY MOUTH |
| pioglitazone 45 MG Oral Tablet | Po |
| insulin human, isophane 85 UNT/ML | SC |
| Insulin Lispro 50 UNT/ML | sub q. |
| exenatide 2 MG [Bydureon] | subcutaneous. |
| 24 HR Metformin hydrochloride 500 MG Extended Release Oral Tablet [Glucophage] | subcutaneous |
| insulin degludec 200 UNT/ML | IV |
| Metformin hydrochloride 500 MG [Glucophage] | swallow |
| insulin human, isophane 50 UNT/ML / Regular Insulin, Human 50 UNT/ML Injectable Suspension [Humulin] | by mouth |
| nateglinide 60 MG Oral Tablet [Starlix] | PER TUBE |
| Regular Insulin, Human 30 UNT/ML | subq |
| Metformin 1000 MG | orally. |
| Glyburide | OU |
| Glipizide 2.5 MG [Glucotrol] | PER TUBE |
| insulin degludec 100 UNT/ML Pen Injector [Tresiba] | by mouth |
| Insulin Glargine 100 UNT/ML [Lantus] | nostril |
| Glipizide 10 MG | subcu |
| Humulin U Ultralente Injectable Product | subQ |
| Glipizide 2.5 MG / Metformin hydrochloride 250 MG Oral Tablet [Metaglip] | orally |
| Tolazamide Oral Capsule | p.o. |

| | |
|---|---|
| insulin human, isophane 70 UNT/ML / Regular Insulin, Human 30 UNT/ML [Novolin] | IM. |
| Glyburide 1.5 MG Oral Tablet [Glycron] | po |
| Simvastatin 20 MG / sitagliptin 100 MG Oral Tablet | Oral |
| Insulin Glargine | swish and spit |
| insulin human, isophane 70 UNT/ML / Regular Insulin, Human 30 UNT/ML [Humulin] | drip. |
| empagliflozin / Linagliptin Pill | |
| Regular Insulin, Human 100 UNT/ML Injectable Solution [Novolin R] | Intravenous |
| Insulin Glargine | g - tube |
| Insulin Lispro | By mouth |
| dulaglutide 3 MG/ML | by mouth |
| Insulin Lispro 50 UNT/ML | subcutaneous. |
| Metformin hydrochloride 500 MG / rosiglitazone 2 MG [Avandamet] | subcutaneous |
| Regular Insulin, Human | SUBCUTANEOUSLY |
| pioglitazone | Im |
| miglitol 50 MG Oral Tablet [Glyset] | orally |
| Simvastatin 40 MG / sitagliptin 50 MG [Juvisync] | |
| Insulin Glargine 100 UNT/ML Injectable Solution [Lantus] | COMBO ROUTE) |
| insulin pork, isophane 100 UNT/ML Injectable Suspension [NPH Iletin II] | SL |
| dulaglutide 1.5 MG/ML [Trulicity] | pO |
| Regular Insulin, Human | TD |
| Metformin 500 MG | BUCCAL |
| glimepiride 4 MG / pioglitazone 30 MG Oral Tablet | oral |
| insulin human, isophane 70 UNT/ML / Regular Insulin, Human 30 UNT/ML Pen Injector [Humulin] | G - tube |
| Insulin Lispro 50 UNT/ML / Insulin, Protamine Lispro, Human 50 UNT/ML Injectable Suspension [Humalog Mix] | neb |
| Glipizide 10 MG [Glucotrol] | per tube |
| Glyburide 5 MG [Diabeta] | subcutaneously |
| Regular Insulin, Human 25 UNT/ML | orally |
| Insulin, Zinc, Pork | subq |
| Insulin Lispro 100 UNT/ML Injectable Solution [Humalog] | VIA PUMP |
| Insulin, Aspart, Human 100 UNT/ML Injectable Solution | SQ |
| 24 HR Glipizide 5 MG Extended Release Oral Tablet | ORAL |
| Metformin hydrochloride 1000 MG / saxagliptin 2.5 MG Extended Release Oral Tablet | p.o. |

| | |
|---|---|
| Regular Insulin, Human 100 UNT/ML Injectable Solution [Novolin R] | PER INSULIN PUMP |
| Chlorpropamide 250 MG Oral Tablet | orally |
| Insulin, Aspart Protamine, Human 70 UNT/ML / Insulin, Aspart, Human 30 UNT/ML [NovoLog Mix] | orally |
| Metformin Extended Release Oral Tablet | swallow. |
| Glyburide 5 MG / Phenformin 50 MG Oral Tablet | oral |
| repaglinide 0.5 MG [Prandin] | gt |
| 3 ML insulin human, isophane 100 UNT/ML Pen Injector [Humulin N] | MISCELLANEOUS |
| Insulin Glargine | BY MOUTH |
| miglitol 25 MG Oral Tablet | p.o. |
| Metformin Oral Tablet | Oral |
| insulin human, isophane 40 UNT/ML | orally |
| Insulin, Aspart, Human 100 UNT/ML Injectable Solution [NovoLog] | BY INSULIN PUMP |
| Glipizide 5 MG | swallow |
| insulin detemir 100 UNT/ML [Levemir] | SEE COMMENT |
| alogliptin 12.5 MG Oral Tablet [Nesina] | ORAL |
| Metformin 500 MG | per rectum. |
| Insulin Lispro 25 UNT/ML | Subq |
| Regular Insulin, Human | INTRAVENOUS |
| dapagliflozin 5 MG / Metformin hydrochloride 500 MG [Xigduo] | by mouth |
| 1.5 ML Regular Insulin, Human 100 UNT/ML Prefilled Syringe | IV |
| insulin human, isophane 60 UNT/ML | Subcut. |
| Insulin, Zinc, Human 100 UNT/ML Injectable Suspension [Novolin L] | by mouth |
| Insulin, Zinc, Human | subcu. |
| Novolin N Injectable Product | SUBCUTANEOUS |
| empagliflozin 25 MG [Jardiance] | SC |
| nateglinide Oral Product | Oral |
| Insulin Lispro 100 UNT/ML Injectable Solution [Humalog] | SUBCUTANEOUS |
| Linagliptin 2.5 MG / Metformin hydrochloride 500 MG Oral Tablet [Jentadueto] | p.o. |
| nateglinide 60 MG [Starlix] | NPO |
| Metformin | ORAL |
| Glyburide 5 MG | IM |
| Insulin, Aspart, Human | injection |
| dapagliflozin 5 MG Oral Tablet | |

| | | |
|---|---|---|
| Metformin hydrochloride 500 MG / rosiglitazone 2 MG [Avandamet] | SubQ | |
| Insulin, Zinc, Pork | subcut | |
| Levemir Injectable Product | SUBCUTANEOUSLY | |
| Glipizide 5 MG Extended Release Oral Tablet | gt | |
| Insulin Lispro Injectable Solution [Humalog] | | 0 |
| 3 ML Insulin, Aspart, Human 100 UNT/ML Cartridge [NovoLog] | inhalation | |
| Insulin Lispro 100 UNT/ML | p.o. | |
| insulin detemir | p.o. | |
| Metformin hydrochloride 500 MG Extended Release Oral Tablet [Glumetza] | BY MOUTH | |
| Insulin, Zinc, Human 100 UNT/ML Injectable Suspension [Humulin L] | subcutaneous | |
| insulin pork, isophane 100 UNT/ML Injectable Suspension [NPH Iletin II] | sub q. | |
| Metformin | intravenously. | |
| sitagliptin 50 MG [Januvia] | per tube | |
| Insulin Lispro 25 UNT/ML | subcutaneously. | |
| Regular Insulin, Human 10 UNT/ML | subcutaneous. | |
| Metformin | GT | |
| dulaglutide 1.5 MG/ML | subcutaneously | |
| Insulin, Zinc, Human 100 UNT/ML Injectable Suspension [Novolin L] | p.o | |
| Insulin Lispro Cartridge [Humalog] | SEE COMMENT | |
| Insulin, Regular, Pork 40 UNT/ML Injectable Solution | Subcutaneously | |
| Insulin Lispro | NEB | |
| insulin human, isophane 100 UNT/ML Injectable Suspension [Novolin N] | SUBCUTANEOUSLY | |
| Glyburide Oral Tablet | | 0 |
| Metformin hydrochloride 500 MG / sitagliptin 50 MG Oral Tablet | PO | |
| Insulin, Glulisine, Human 100 UNT/ML Injectable Solution [Apidra] | CONTINUOUS INFUSION | |
| Regular Insulin, Human | INFUSION. | |
| 24 HR Glipizide 5 MG Extended Release Oral Tablet [Glucotrol] | nostril | |
| Insulin, Regular, Pork 40 UNT/ML Injectable Solution | subQ | |
| dapagliflozin 5 MG | iv | |
| Regular Insulin, Human 500 UNT/ML Injectable Solution [Humulin R] | SUBCUTANEOUS PER INSULIN PUMP | |
| Regular Insulin, Human 500 UNT/ML Injectable Solution | SQ | |
| insulin human, rDNA origin 1 MG/ACTUAT Inhalant Powder | inhalation | |

| | | |
|---|---|---|
| Insulin Glargine | subc. | |
| insulin degludec 200 UNT/ML | subcut | |
| Metformin hydrochloride 100 MG/ML [Riomet] | G - Tube | |
| insulin human, isophane 32 UNT/ML | sub Q | |
| insulin human, isophane 34 UNT/ML | Subcut. | |
| Glyburide | npo. | |
| Toujeo Injectable Product | | 0 |
| insulin human, isophane 70 UNT/ML / Regular Insulin, Human 30 UNT/ML [Humulin] | subc. | |
| Insulin, Aspart, Human 100 UNT/ML Injectable Solution [NovoLog] | SUBCUTANEOUS INFUSION VIA | |
| Metformin hydrochloride 750 MG | orally. | |
| Glyburide 2.5 MG Oral Tablet | PER TUBE | |
| Insulin Glargine | G - tube | |
| glimepiride 2 MG | ng | |
| Insulin, Aspart Protamine, Human 70 UNT/ML / Insulin, Aspart, Human 30 UNT/ML Injectable Suspension | SUBCUTANEOUS | |
| insulin human, isophane 70 UNT/ML / Regular Insulin, Human 30 UNT/ML [Novolin] | iv | |
| Regular Insulin, Human 6 UNT/ML | SQ | |
| 3 ML liraglutide 6 MG/ML Pen Injector [Victoza] | Sub Q | |
| Glyburide Pill | swallow | |
| Insulin, Glulisine, Human | sq | |
| Glucophage Oral Product | ORALLY | |
| Insulin, Regular, Pork 100 UNT/ML Injectable Solution | subq | |
| insulin human, isophane 100 UNT/ML Injectable Suspension [Humulin I] | p.o. | |
| exenatide Injection [Bydureon] | | |
| Farxiga Oral Product | Oral | |
| 3 ML Insulin Lispro 25 UNT/ML / Insulin, Protamine Lispro, Human 75 UNT/ML Pen Injector [Humalog Mix] | subcutaneously | |
| insulin detemir 100 UNT/ML Injectable Solution [Levemir] | Subcutaneous | |
| Glyburide 2.5 MG | subcu | |
| {180 (insulin human, rDNA origin 1 MG/ACTUAT Inhalant Powder [Exubera]) / 90 (insulin human, rDNA origin 3 MG/ACTUAT Inhalant Powder [Exubera]) } Pack [Exubera Kit] | inhalation | |
| Insulin Lispro 25 UNT/ML / Insulin, Protamine Lispro, Human 75 UNT/ML Injectable Suspension [Humalog Mix] | Sub Q | |
| Metformin / pioglitazone Oral Tablet | by mouth | |
| Insulin, Aspart, Human | by mouth. | |

| | | |
|---|---|---|
| insulin pork, isophane 70 UNT/ML / Insulin, Regular, Pork 30 UNT/ML Injectable Suspension | | 0 |
| Regular Insulin, Human 100 UNT/ML Injectable Solution [Humulin R] | SUBCUTANEOUS BEFORE | |
| Lixisenatide | by mouth | |
| Insulin, Aspart, Human 100 UNT/ML Injectable Solution [NovoLog] | SUBCUTANEOUS,PER INSULIN PUMP | |
| Insulin Lispro 50 UNT/ML | MISC | |
| saxagliptin Oral Tablet | | |
| Metformin hydrochloride 1000 MG / rosiglitazone 4 MG [Avandamet] | SEE COMMENT | |
| Metformin hydrochloride 500 MG Oral Tablet [Glucophage] | by moutH | |
| rosiglitazone | NPO | |
| exenatide 0.01 MG/ACTUAT [Byetta] | MISC | |
| Metformin hydrochloride 1000 MG / sitagliptin 50 MG [Janumet] | PO | |
| Exubera Inhalant Product | | |
| Insulin Glargine | Sq | |
| Acarbose | Im | |
| insulin pork, isophane 100 UNT/ML Injectable Suspension [NPH Iletin II] | sublingual | |
| Metformin hydrochloride 500 MG Oral Tablet | G - TUBE | |
| Ultralente Insulin, Human 70 UNT/ML | sq | |
| Glipizide 2.5 MG / Metformin hydrochloride 250 MG Oral Tablet | Oral | |
| Glipizide 5 MG / Metformin hydrochloride 500 MG Oral Tablet [Metaglip] | orally | |
| exenatide 2 MG Injection [Bydureon] | oral | |
| pioglitazone 15 MG [Actos] | IM | |
| Acetohexamide 250 MG | | |
| Glyburide 1.5 MG Oral Tablet | | |
| Glipizide 10 MG [Glucotrol] | Sublingual | |
| repaglinide 2 MG Oral Tablet | | |
| Metformin hydrochloride 750 MG | gt | |
| Metformin hydrochloride 500 MG / sitagliptin 50 MG Extended Release Oral Tablet [Janumet] | oral | |
| 3 ML Regular Insulin, Human 500 UNT/ML Pen Injector [Humulin R] | | 0 |
| 0.5 ML dulaglutide 1.5 MG/ML Auto-Injector [Trulicity] | subcutaneous | |
| Insulin Glargine 100 UNT/ML Injectable Solution | subc | |
| Insulin, Regular, Pork 40 UNT/ML Injectable Solution | p.o. | |
| nateglinide 60 MG Oral Tablet [Starlix] | BY MOUTH | |

| | |
|---|---|
| Insulin Lispro Cartridge [Humalog] | subcu. |
| 3 ML Insulin Lispro 200 UNT/ML Pen Injector | subcu |
| Insulin Glargine Injectable Solution | SubQ |
| Glipizide | ng |
| Glyburide 6 MG | ng |
| Insulin, Aspart Protamine, Human | SUBCUT |
| insulin human, isophane 70 UNT/ML / Regular Insulin, Human 30 UNT/ML Injectable Suspension [Novolin] | Sub Q |
| Glyburide Oral Tablet [Glynase] | po |
| rosiglitazone 2 MG [Avandia] | inhalation |
| 3 ML Insulin Glargine 100 UNT/ML Pen Injector [Lantus] | SUB Q |
| Glipizide 5 MG Oral Tablet | P.O |
| exenatide 0.01 MG/ACTUAT [Byetta] | nostril |
| Fortamet Oral Product | orally |
| Metformin hydrochloride 850 MG | drip |
| pioglitazone 45 MG [Actos] | topical |
| Insulin, Protamine Lispro, Human | by mouth |
| Bromocriptine | ng |
| Cycloset Oral Product | oral |
| Regular Insulin, Human 500 UNT/ML Injectable Solution [Humulin R] | CONTINUOUS INFUSION |
| insulin pork, isophane 70 UNT/ML / Insulin, Regular, Pork 30 UNT/ML Injectable Suspension | SUBCUTANEOUSLY |
| insulin human, isophane 100 UNT/ML Injectable Suspension | po |
| insulin human, isophane 90 UNT/ML | subcu |
| Glucophage Oral Product | ORAL |
| Linagliptin Oral Product | p.o. |
| 3 ML Insulin, Glulisine, Human 100 UNT/ML Pen Injector [Apidra] | gt |
| alogliptin 25 MG / pioglitazone 45 MG Oral Tablet [Oseni] | |
| Lantus Injectable Product | IV |
| 3 ML Insulin, Aspart Protamine, Human 70 UNT/ML / Insulin, Aspart, Human 30 UNT/ML Pen Injector [NovoLog Mix] | subq. |
| 1.5 ML insulin human, isophane 100 UNT/ML Prefilled Syringe | po |
| Metformin hydrochloride 850 MG [Glucophage] | TD |
| canagliflozin 100 MG [Invokana] | by mouth. |
| Insulin Lispro 100 UNT/ML Injectable Solution [Humalog] | intravenously |
| canagliflozin 100 MG [Invokana] | subcutaneously |
| 24 HR Metformin hydrochloride 1000 MG / saxagliptin 2.5 MG Extended Release Oral Tablet [Kombiglyze] | p.o. |
| Glyburide Oral Tablet | inhalation |

| | | |
|---|---|---|
| insulin human, isophane 90 UNT/ML<br>R] | SUBQ | |
| insulin pork, isophane 40 UNT/ML | subq | |
| Regular Insulin, Human 15 UNT/ML | orally | |
| rosiglitazone 4 MG Oral Tablet [Avandia] | ORALLY | |
| Regular Insulin, Human 30 UNT/ML | per tube | |
| Glipizide 2.5 MG Extended Release Oral Tablet | gt | |
| Glyburide 5 MG | npo | |
| miglitol 100 MG | | |
| Insulin, Aspart Protamine, Human 70 UNT/ML / Insulin, Aspart, Human 30 UNT/ML Pen Injector [NovoLog Mix] | | |
| Insulin, Zinc, Pork | PO | |
| Linagliptin Oral Tablet [Tradjenta] | oral | |
| insulin detemir 100 UNT/ML Injectable Solution | Subcutaneous | |
| Synjardy Oral Product | po | |
| dulaglutide 1.5 MG/ML [Trulicity] | SEE COMMENT | |
| Bromocriptine 0.8 MG Oral Tablet [Cycloset] | Oral | |
| miglitol 50 MG Oral Tablet | p.o. | |
| Byetta Injectable Product | Subcutaneously | |
| Metaglip Oral Product | Po | |
| glimepiride | bedside | |
| Actos Oral Product | P.o | |
| Insulin Lispro 50 UNT/ML / Insulin, Protamine Lispro, Human 50 UNT/ML Injectable Suspension [Humalog Mix] | | 0 |
| nateglinide 180 MG | | |
| Novolin R Injectable Product | by mouth | |
| insulin human, isophane 80 UNT/ML / Regular Insulin, Human 20 UNT/ML [Humulin] | sub q | |
| glimepiride 2 MG [Amaryl] | po. | |
| Acarbose 50 MG Oral Tablet [Precose] | BY MOUTH | |
| NovoLog Injectable Product | SUBCUTANEOUS | |
| Glyburide | Per tube | |
| Parlodel Pill | by mouth | |
| pioglitazone 15 MG | Nares | |
| 1.5 ML insulin human, isophane 70 UNT/ML / Regular Insulin, Human 30 UNT/ML Prefilled Syringe | subcu. | |
| Trulicity Injectable Product | SUBCUTANEOUSLY | |
| Novolin Injectable Product | subcutaneous | |
| Metformin hydrochloride 500 MG Oral Tablet | swallow. | |
| Insulin Lispro Cartridge [Humalog] | COMBO ROUTE) | |

| | |
|---|---|
| 24 HR Glipizide 5 MG Extended Release Oral Tablet [Glucotrol] | orally |
| Metformin hydrochloride 1000 MG / sitagliptin 50 MG [Janumet] | MISC |
| Humulin Injectable Product | subcutaneous |
| Metformin hydrochloride 500 MG [Glucophage] | G - Tube |
| Metformin 1000 MG | td |
| canagliflozin 150 MG / Metformin hydrochloride 1000 MG Oral Tablet [Invokamet] | Oral |
| Tolazamide 100 MG Oral Tablet | po |
| insulin detemir Injectable Product | Subcutaneously |
| glimepiride 4 MG [Amaryl] | SC |
| Glyburide 3 MG Oral Tablet | SUBLING |
| Glipizide 10 MG Oral Tablet | p.o |
| Insulin Lispro 200 UNT/ML | sq |
| glimepiride | Subcut |
| Insulin Lispro | g - tube |
| pioglitazone 30 MG Oral Tablet | BY MOUTH |
| Glyburide 5 MG [Diabeta] | subcutaneous |
| Byetta Injectable Product | subcut |
| Metformin | Sq |
| Glyburide 5 MG Oral Tablet [Micronase] | oral |
| glimepiride 2 MG | bedside |
| canagliflozin 50 MG / Metformin hydrochloride 500 MG Oral Tablet [Invokamet] | |
| Regular Insulin, Human 100 UNT/ML Injectable Solution [Velosulin] | per tube |
| 60 ACTUAT exenatide 0.01 MG/ACTUAT Pen Injector [Byetta] | orally. |
| empagliflozin 10 MG [Jardiance] | inhalation |
| Acarbose 50 MG [Precose] | OU |
| Insulin Lispro | sq. |
| insulin degludec 200 UNT/ML | subcu. |
| insulin human, isophane 70 UNT/ML / Regular Insulin, Human 30 UNT/ML [Novolin] | bedside. |
| exenatide 2 MG [Bydureon] | gt |
| empagliflozin 10 MG / Linagliptin 5 MG Oral Tablet | by mouth |
| insulin human, isophane 70 UNT/ML / Regular Insulin, Human 30 UNT/ML [Novolin] | MISC |
| insulin human, isophane 70 UNT/ML / Regular Insulin, Human 30 UNT/ML [Novolin] | npo |
| Metformin 500 MG | sublingual. |
| 3 ML liraglutide 6 MG/ML Pen Injector [Saxenda] | oral |

| | | |
|---|---|---|
| Glyburide Oral Tablet | SEE COMMENT | |
| 0.5 ML albiglutide 100 MG/ML Pen Injector [Tanzeum] | PO | |
| Amaryl Pill | swallow | |
| Metformin hydrochloride 100 MG/ML | subq | |
| dulaglutide 1.5 MG/ML [Trulicity] | PO | |
| Glynase Oral Product | Oral | |
| Farxiga Oral Product | p.o. | |
| Insulin Lispro 50 UNT/ML | npo | |
| Metformin hydrochloride 500 MG / pioglitazone 15 MG Oral Tablet | oral | |
| Insulin, Aspart, Human | sublingual | |
| glimepiride 3 MG Oral Tablet [Amaryl] | orally | |
| alogliptin 12.5 MG / pioglitazone 15 MG Oral Tablet [Oseni] | ORAL | |
| glimepiride | inh | |
| insulin human, isophane 32 UNT/ML | sc | |
| Regular Insulin, Human 10 UNT/ML | subcu | |
| Metformin hydrochloride 500 MG / rosiglitazone 1 MG Oral Tablet [Avandamet] | p.o. | |
| glimepiride 2 MG [Amaryl] | bedside | |
| rosiglitazone 4 MG [Avandia] | per tube | |
| Metformin hydrochloride 1000 MG / sitagliptin 50 MG [Janumet] | subcut | |
| Glipizide 10 MG Oral Tablet [Glucotrol] | PER TUBE | |
| Metformin hydrochloride 100 MG/ML | per tube | |
| Metformin hydrochloride 1000 MG Extended Release Oral Tablet | subcutaneously | |
| canagliflozin 150 MG / Metformin hydrochloride 1000 MG Oral Tablet [Invokamet] | | 0 |
| pioglitazone Oral Tablet | Oral | |
| 24 HR Glipizide 10 MG Extended Release Oral Tablet [Glucotrol] | nostril | |
| Metformin 400 MG | by mouth. | |
| Insulin Lispro | intravenously. | |
| insulin human, isophane 32 UNT/ML | subcu. | |
| pioglitazone 45 MG [Actos] | drip. | |
| Glyburide | SC | |
| Insulin Lispro 200 UNT/ML Pen Injector [Humalog] | subq. | |
| albiglutide | subcutaneous | |
| insulin human, isophane 70 UNT/ML / Regular Insulin, Human 30 UNT/ML Injectable Suspension [Novolin] | SUBCUTANEOUSLY | |
| Glipizide | subQ | |
| nateglinide 60 MG | NPO | |
| miglitol 25 MG Oral Tablet [Glyset] | oral | |

| | | |
|---|---|---|
| Bromocriptine 5 MG [Parlodel] | per tube | |
| 1.5 ML Insulin Glargine 300 UNT/ML Pen Injector [Toujeo] | SubQ | |
| Regular Insulin, Human 10 UNT/ML | by mouth | |
| sitagliptin 50 MG [Januvia] | SQ | |
| 3 ML insulin detemir 100 UNT/ML Pen Injector [Levemir] | PO | |
| 24 HR Metformin hydrochloride 1000 MG / pioglitazone 15 MG Extended Release Oral Tablet [Actoplus Met] | po | |
| Glyburide 1.5 MG Oral Tablet [Glynase] | oral | |
| Insulin Lispro 100 UNT/ML Injectable Solution | po | |
| saxagliptin Pill | oral | |
| Insulin Glargine | SubCut. | |
| canagliflozin 300 MG [Invokana] | nostril | |
| Insulin Lispro 25 UNT/ML | oral | |
| Actos Oral Product | swallow | |
| insulin beef, isophane 40 UNT/ML | subcu. | |
| Humalog Injectable Product | IV | |
| Glyburide 1.25 MG | sublingual | |
| Glyburide 2.5 MG / Metformin hydrochloride 500 MG Oral Tablet [Glucovance] | bedside | |
| Bromocriptine 0.8 MG Oral Tablet [Cycloset] | oral | |
| exenatide 2 MG Injection [Bydureon] | Subcutaneous | |
| Glucotrol Oral Product | Po | |
| repaglinide 0.5 MG [Prandin] | inhaled | |
| Insulin Lispro 50 UNT/ML | oral | |
| Regular Insulin, Human 500 UNT/ML Injectable Solution [Humulin R] | Oral | |
| sitagliptin 100 MG [Januvia] | drip | |
| Insulin Lispro 50 UNT/ML / Insulin, Protamine Lispro, Human 50 UNT/ML Injectable Suspension [Humalog Mix] | Oral | |
| insulin human, isophane 50 UNT/ML / Regular Insulin, Human 50 UNT/ML [Humulin] | Sub Q | |
| Glipizide 10 MG Oral Tablet | topically | |
| insulin detemir 100 UNT/ML [Levemir] | gt | |
| Glyburide 5 MG Oral Tablet | By mouth | |
| 3 ML Insulin, Aspart Protamine, Human 70 UNT/ML / Insulin, Aspart, Human 30 UNT/ML Pen Injector [NovoLog Mix] | p.o. | |
| Insulin Glargine | SUBC | |
| 24 HR dapagliflozin 5 MG / Metformin hydrochloride 1000 MG Extended Release Oral Tablet [Xigduo] | | 0 |
| glimepiride 2 MG | subcutaneous | |

| | | |
|---|---|---|
| Insulin, Aspart, Human | BY MOUTH | |
| 3 ML Insulin, Aspart, Human 100 UNT/ML Pen Injector [NovoLog] | subcutaneous | |
| insulin human, isophane 28 UNT/ML | SUB Q | |
| repaglinide Oral Tablet [Prandin] | po | |
| Glipizide 10 MG Oral Tablet [Glucotrol] | gt | |
| 3 ML liraglutide 6 MG/ML Pen Injector [Victoza] | nostril | |
| Glyburide 5 MG / Metformin 1000 MG Oral Tablet | IV | |
| pioglitazone 30 MG [Actos] | drip | |
| Osmotic 24 HR Metformin hydrochloride 1000 MG Extended Release Oral Tablet [Fortamet] | | |
| Metformin hydrochloride 1000 MG Oral Tablet [Glucophage] | Po | |
| Glyburide 2.5 MG | subcutaneous. | |
| 3 ML Insulin, Aspart, Human 100 UNT/ML Cartridge [NovoLog] | subcutaneously | |
| Regular Insulin, Human | NPO. | |
| 3 ML insulin human, isophane 100 UNT/ML Prefilled Syringe | per rectum | |
| Invokamet Pill | | |
| Regular Insulin, Human 50 UNT/ML | po | |
| Linagliptin 2.5 MG / Metformin hydrochloride 1000 MG Oral Tablet [Jentadueto] | p.o. | |
| 1.5 ML Insulin Glargine 300 UNT/ML Pen Injector [Toujeo] | injection | |
| Regular Insulin, Human 12 UNT | subcutaneous | |
| exenatide 2 MG | SQ | |
| insulin human, isophane 32 UNT/ML | Subcut | |
| Metformin hydrochloride 500 MG / pioglitazone 15 MG Oral Tablet | swallow | |
| Glipizide 10 MG Oral Tablet [Glucotrol] | sublingual | |
| Insulin, Aspart, Human 100 UNT/ML Cartridge [NovoLog] | npo | |
| Glyburide 2.5 MG | swallow | |
| Insulin, Aspart Protamine, Human 70 UNT/ML / Insulin, Aspart, Human 30 UNT/ML Injectable Suspension | subcu | |
| glimepiride Oral Tablet | BY MOUTH | |
| Tolazamide 250 MG Oral Tablet [Tolinase] | oral | |
| empagliflozin 25 MG / Linagliptin 5 MG Oral Tablet [Glyxambi] | Oral | |
| 3 ML insulin human, isophane 100 UNT/ML Prefilled Syringe | | 0 |
| glimepiride 2 MG Oral Tablet [Amaryl] | g-tube | |
| Glyburide 6 MG | per mouth | |
| insulin degludec 100 UNT/ML [Tresiba] | | |
| insulin beef, isophane 80 UNT/ML | by mouth | |
| Insulin Glargine | Swallow | |
| Insulin Lispro 100 UNT/ML Injectable Solution [Humalog] | INSULIN PUMP | |

| | |
|---|---|
| sitagliptin 100 MG [Januvia] | td |
| Chlorpropamide | PO |
| Insulin Lispro Cartridge [Humalog] | subc |
| 0.5 ML albiglutide 60 MG/ML Pen Injector [Tanzeum] | Sc |
| empagliflozin 5 MG / Metformin hydrochloride 1000 MG [Synjardy] | |
| Metformin hydrochloride 1000 MG / saxagliptin 2.5 MG [Kombiglyze] | p.o. |
| 3 ML Insulin, Glulisine, Human 100 UNT/ML Pen Injector [Apidra] | subcu |
| Metformin hydrochloride 1000 MG Oral Tablet | subq |
| Glyburide 5 MG | TD |
| rosiglitazone 8 MG [Avandia] | po. |
| Glyburide Pill | bedside |
| Glipizide 10 MG | OU |
| glimepiride 2 MG / pioglitazone 30 MG Oral Tablet [Duetact] | PO |
| Metformin hydrochloride 500 MG Oral Tablet [Glucophage] | ORAL |
| Insulin Glargine 300 UNT/ML Pen Injector [Toujeo] | SEE COMMENT |
| Insulin Glargine | intravenous |
| Insulin, Zinc, Human | oral |
| Regular Insulin, Human 24 UNT/ML | SUBCUT |
| Insulin Lispro 100 UNT/ML Injectable Solution [Humalog] | PUMP |
| glimepiride 2 MG | by mouth. |
| Glipizide 10 MG Oral Tablet | Orally |
| Regular Insulin, Human 100 UNT/ML Injectable Solution | SUBCUTANEOUSLY |
| liraglutide 6 MG/ML [Saxenda] | sub q |
| Acetohexamide 250 MG [Dymelor] | |
| pramlintide acetate 1 MG/ML [Symlin] | SC |
| Metformin 1000 MG | TOPICAL |
| Regular Insulin, Human 100 UNT/ML Injectable Solution [Novolin R] | subcu. |
| Glyburide 2.5 MG / Metformin hydrochloride 500 MG Oral Tablet [Glucovance] | BY MOUTH |
| Insulin Glargine | po. |
| Metformin 400 MG | per mouth |
| {30 (Regular Insulin, Human 12 UNT Inhalant Powder [Afrezza]) / 60 (Regular Insulin, Human 8 UNT Inhalant Powder [Afrezza]) } Pack [Afrezza 90 - 60 (8 UNT), 30 (12 UNT)] | INHALATION |
| Insulin Lispro 100 UNT/ML | |
| insulin human, isophane 70 UNT/ML | subcutaneous |
| Insulin Lispro Cartridge [Humalog] | ou |

| | | |
|---|---|---|
| Ultralente Insulin, Beef-Pork 40 UNT/ML | | |
| Metformin hydrochloride 850 MG | per mouth | |
| insulin pork, isophane 80 UNT/ML | PO | |
| insulin human, isophane 70 UNT/ML / Regular Insulin, Human 30 UNT/ML [Humulin] | PO. | |
| insulin human, isophane 70 UNT/ML | | 0 |
| 1.5 ML insulin human, isophane 70 UNT/ML / Regular Insulin, Human 30 UNT/ML Prefilled Syringe [Novolin] | by mouth | |
| Toujeo Injectable Product | SUBCUTANEOUS | |
| rosiglitazone Oral Tablet [Avandia] | po | |
| Ultralente Insulin, Human 100 UNT/ML Injectable Suspension [Humulin U Ultralente] | | 0 |
| Metformin hydrochloride 625 MG Oral Tablet | by mouth | |
| Glyburide 3 MG Oral Tablet | Po | |
| Glipizide 10 MG Extended Release Oral Tablet | per tube | |
| 24 HR Metformin hydrochloride 500 MG / saxagliptin 5 MG Extended Release Oral Tablet [Kombiglyze] | orally | |
| Insulin, Regular, Pork 40 UNT/ML Injectable Solution | IM | |
| pioglitazone 30 MG Oral Tablet [Actos] | P.O. | |
| Insulin Lispro 50 UNT/ML | INH | |
| Regular Insulin, Human 20 UNT/ML | intravenous | |
| Insulin Lispro 25 UNT/ML / Insulin, Protamine Lispro, Human 75 UNT/ML Injectable Suspension [Humalog Mix] | IV | |
| exenatide 2 MG [Bydureon] | MISC | |
| exenatide 2 MG [Bydureon] | IM / SQ | |
| Glipizide 5 MG Extended Release Oral Tablet | p.o. | |
| insulin pork, isophane 70 UNT/ML / Insulin, Regular, Pork 30 UNT/ML Injectable Suspension | SUBCUTANEOUSLY | |
| repaglinide Oral Tablet [Prandin] | PO | |
| Acarbose 100 MG Oral Tablet | Oral | |
| Glyburide Oral Tablet | p.o | |
| glimepiride 2 MG Oral Tablet [Amaryl] | PER TUBE | |
| insulin human, isophane 100 UNT/ML Injectable Suspension [Humulin N] | subq. | |
| Insulin, Aspart, Human | VERIFY | |
| insulin human, isophane 75 UNT/ML | SUBCUTANEOUS | |
| Glyburide 3 MG | subcutaneous | |
| insulin detemir Injectable Solution [Levemir] | | |
| Acarbose | subcutaneously | |

| | |
|---|---|
| insulin human, isophane 28 UNT/ML / Regular Insulin, Human 12 UNT/ML Injectable Suspension | |
| Metformin hydrochloride 500 MG Extended Release Oral Tablet | nostril |
| Metformin hydrochloride 750 MG Extended Release Oral Tablet | subcutaneously |
| Glyburide 5 MG | intravenous |
| Avandia Pill | po |
| Glyburide 1.25 MG | po. |
| Insulin Lispro 50 UNT/ML / Insulin, Protamine Lispro, Human 50 UNT/ML Injectable Suspension [Humalog Mix] | im |
| Humalog Injectable Product | SUBCUTANEOUS |
| 24 HR Glipizide 5 MG Extended Release Oral Tablet [Glucotrol] | intramuscular |
| 1.5 ML Insulin Glargine 300 UNT/ML Pen Injector [Toujeo] | subcutaneously |
| dapagliflozin 5 MG Oral Tablet [Farxiga] | subcutaneously |
| Bromocriptine 2.5 MG Oral Tablet | ORAL |
| Metformin Extended Release Oral Tablet [Fortamet] | po |
| insulin pork, isophane 100 UNT/ML Injectable Suspension [NPH Iletin II] | J tube. |
| pioglitazone 30 MG [Actos] | nostril |
| Insulin Lispro 25 UNT/ML / Insulin, Protamine Lispro, Human 75 UNT/ML Injectable Suspension [Humalog Mix] | injection |
| exenatide 0.005 MG/ACTUAT | SQ |
| 3 ML Insulin Lispro 100 UNT/ML Cartridge [Humalog] | PUMP |
| 3 ML Insulin Glargine 100 UNT/ML Pen Injector [Basaglar] | miscellaneous |
| nateglinide 60 MG [Starlix] | po. |
| sitagliptin 25 MG [Januvia] | drip. |
| Nesina Oral Product | po |
| Insulin Glargine | J tube |
| Glipizide 10 MG | NPO. |
| Glipizide 2.5 MG Oral Tablet | gt |
| 24 HR empagliflozin 5 MG / Metformin hydrochloride 1000 MG Extended Release Oral Tablet [Synjardy] | |
| Orinase Oral Product | PO |
| repaglinide 2 MG [Prandin] | nostril |
| Byetta Injectable Product | IM injection |
| Insulin, Aspart, Human | Eye_Right |
| 0.5 ML dulaglutide 1.5 MG/ML Auto-Injector | RT EYE |
| rosiglitazone Oral Product | by mouth |
| Glipizide 10 MG [Glucotrol] | subcutaneous |
| Bromocriptine 10 MG Oral Capsule [Parlodel] | |
| glimepiride 1 MG Oral Tablet [Amaryl] | PER PEG |

| | |
|---|---|
| insulin human, isophane 100 UNT/ML Injectable Suspension [Novolin N] | SQ. |
| canagliflozin 100 MG Oral Tablet | po |
| Glipizide 5 MG Oral Tablet | ORAL |
| Glyburide 5 MG / Metformin hydrochloride 500 MG Oral Tablet [Glucovance] | PER TUBE |
| insulin pork, isophane 100 UNT/ML Injectable Suspension [NPH Iletin II] | ORAL |
| pioglitazone 30 MG [Actos] | subcutaneously |
| Metformin hydrochloride 500 MG / repaglinide 1 MG Oral Tablet [PrandiMet] | |
| Chlorpropamide 100 MG Oral Tablet [Diabinese] | PO |
| liraglutide 6 MG/ML [Victoza] | PO |
| Kombiglyze Oral Product | PO |
| Acarbose 25 MG Oral Tablet [Precose] | Oral |
| insulin human, isophane 70 UNT/ML / Regular Insulin, Human 30 UNT/ML [Novolin] | SL |
| nateglinide | inh. |
| Insulin, Glulisine, Human | subq |
| Glyburide 5 MG [Diabeta] | per tube. |
| insulin human, isophane 100 UNT/ML Injectable Suspension [Humulin N] | intravenously |
| insulin human, isophane 70 UNT/ML / Regular Insulin, Human 30 UNT/ML [Novolin] | sub q. |
| Insulin Lispro Cartridge [Humalog] | TOPICAL |
| Metformin hydrochloride 750 MG [Glucophage] | subcut |
| nateglinide Oral Tablet [Starlix] | PO |
| Linagliptin Oral Tablet | PO |
| Metformin hydrochloride 500 MG Extended Release Oral Tablet [Fortamet] | subcutaneous |
| Afrezza Inhalant Product | Inhaled |
| liraglutide 6 MG/ML [Victoza] | nostril |
| pioglitazone 45 MG | PER TUBE |
| canagliflozin 300 MG [Invokana] | subcutaneously |
| Metformin hydrochloride 625 MG Oral Tablet [Glucophage] | by mouth |
| 3 ML Insulin, Aspart, Human 100 UNT/ML Pen Injector [NovoLog] | pump |
| Glyburide | Sublingual |
| Insulin Lispro 100 UNT/ML Injectable Solution | other |
| Glyburide | inh. |
| Metformin hydrochloride 850 MG Oral Tablet | ORAL |

| | | |
|---|---|---|
| Acarbose 25 MG | gt | |
| Metformin | BY MOUTH | |
| Metformin hydrochloride 100 MG/ML [Riomet] | | 0 |
| Glyburide 5 MG | po. | |
| Metformin hydrochloride 1000 MG [Glucophage] | Inhalation | |
| Linagliptin 5 MG [Tradjenta] | subcutaneous | |
| glimepiride | po. | |
| 3 ML Insulin Glargine 100 UNT/ML Pen Injector [Lantus] | Oral | |
| Bromocriptine Oral Capsule | p.o. | |
| Metformin 500 MG / ropinirole 2 MG Oral Tablet | subq | |
| Insulin Glargine Injectable Solution | topical | |
| Glipizide Extended Release Oral Tablet [Glucotrol] | subcutaneous | |
| Insulin Lispro Injectable Solution [Humalog] | subq | |
| Insulin Glargine | IM / SQ | |
| Glyburide 1.25 MG Oral Tablet [Diabeta] | ORAL | |
| insulin human, isophane 32 UNT/ML | Subq | |
| 3 ML insulin detemir 100 UNT/ML Pen Injector [Levemir] | sub Q | |
| Avandaryl Oral Product | orally | |
| Lispro PRC Injectable Product | NPO. | |
| Glipizide 5 MG | trans | |
| Insulin, Aspart, Human 100 UNT/ML Cartridge [NovoLog] | ng | |
| Insulin Lispro | SUBCUTANEOUSLY | |
| glimepiride 4 MG [Amaryl] | Td | |
| Insulin Glargine 100 UNT/ML [Lantus] | COMBO ROUTE) | |
| Acarbose 100 MG | VERIFY ROUTE | |
| exenatide 3.08 MG/ML [Bydureon] | orally | |
| Glyburide 5 MG | drip. | |
| empagliflozin 5 MG / Metformin hydrochloride 500 MG Oral Tablet [Synjardy] | ORAL | |
| Regular Insulin, Human 100 UNT/ML Injectable Solution [Humulin R] | CONT SUBCUTAN INFUSI | |
| exenatide | subq | |
| pioglitazone 30 MG [Actos] | npo | |
| Metformin hydrochloride 100 MG/ML | | 0 |
| glimepiride 4 MG | IV | |
| Glucophage Oral Product | by MOUTH | |
| Insulin, Aspart, Human 100 UNT/ML Cartridge [NovoLog] | sQ | |
| Metformin Extended Release Oral Tablet [Glumetza] | gt | |
| 3 ML Insulin, Aspart Protamine, Human 70 UNT/ML / Insulin, Aspart, Human 30 UNT/ML Pen Injector [NovoLog Mix] | subcutaneously. | |

| | |
|---|---|
| Cycloset Oral Product | po |
| Metformin hydrochloride 500 MG [Glumetza] | subcutaneously |
| Regular Insulin, Human 100 UNT/ML Injectable Solution [Novolin R] | ORAL |
| glimepiride | PER TUBE |
| Parlodel Oral Product | g - tube |
| Linagliptin 2.5 MG / Metformin hydrochloride 850 MG Oral Tablet [Jentadueto] | |
| Glyburide 5 MG / Metformin hydrochloride 500 MG Oral Tablet | by mouth. |
| 3 ML Insulin, Aspart Protamine, Human 70 UNT/ML / Insulin, Aspart, Human 30 UNT/ML Pen Injector [NovoLog Mix] | other |
| nateglinide | subq |
| 3 ML Insulin Lispro 25 UNT/ML / Insulin, Protamine Lispro, Human 75 UNT/ML Pen Injector [Humalog Mix] | intramuscular |
| insulin human, isophane 50 UNT/ML / Regular Insulin, Human 50 UNT/ML [Humulin] | PO |
| Metformin hydrochloride 850 MG / pioglitazone 15 MG Oral Tablet | p.o. |
| 1.5 ML insulin human, isophane 70 UNT/ML / Regular Insulin, Human 30 UNT/ML Prefilled Syringe | NPO. |
| 0.5 ML albiglutide 100 MG/ML Pen Injector [Tanzeum] | oral |
| Insulin, Pork | p.o. |
| Glipizide | j - tube |
| alogliptin 25 MG / pioglitazone 15 MG Oral Tablet [Oseni] | |
| Glipizide 5 MG | Inhaled |
| Onglyza Pill | by mouth |
| pioglitazone | topically |
| insulin human, isophane 100 UNT/ML Injectable Suspension [Novolin N] | injection |
| insulin human, isophane 80 UNT/ML / Regular Insulin, Human 20 UNT/ML [Humulin] | drip |
| Novolin R Injectable Product | IV |
| Regular Insulin, Human 30 UNT/ML | Subcutaneous |
| insulin human, isophane 70 UNT/ML / Regular Insulin, Human 30 UNT/ML [Humulin] | Sq. |
| insulin human, isophane 80 UNT/ML / Regular Insulin, Human 20 UNT/ML [Humulin] | subcutaneous |
| Insulin Lispro Cartridge [Humalog] | sub q. |
| insulin human, isophane 60 UNT/ML | p.o. |
| Regular Insulin, Human 500 UNT/ML Injectable Solution | SUBCUTANEOUS |

| | | |
|---|---|---|
| Metformin | IM injection | |
| Glipizide 10 MG | inhaled | |
| Metformin hydrochloride 500 MG / saxagliptin 5 MG [Kombiglyze] | SQ | |
| miglitol 100 MG Oral Tablet | po | |
| insulin human, isophane 60 UNT/ML / Regular Insulin, Human 40 UNT/ML [Humulin] | subcutaneous | |
| Metformin hydrochloride 1000 MG / sitagliptin 50 MG [Janumet] | drip | |
| Glipizide 10 MG | | 0 |
| Glyburide 5 MG Oral Tablet | gt | |
| Metformin hydrochloride 500 MG Oral Tablet | feeding tube | |
| Bromocriptine 2.5 MG Oral Tablet [Parlodel] | VAGINALLY | |
| Acarbose 25 MG | topically | |
| Metformin Extended Release Oral Tablet [Fortamet] | inhalation | |
| Insulin, Aspart, Human | G - tube | |
| insulin detemir 100 UNT/ML [Levemir] | SUBQ | |
| 0.5 ML albiglutide 100 MG/ML Pen Injector | subcutaneously | |
| 0.65 ML exenatide 3.08 MG/ML Pen Injector [Bydureon] | oral | |
| 3 ML insulin degludec 200 UNT/ML Pen Injector [Tresiba] | subdermal | |
| Metformin hydrochloride 500 MG Oral Tablet | pressure line | |
| saxagliptin Oral Tablet | po | |
| 24 HR Metformin hydrochloride 500 MG Extended Release Oral Tablet [Glucophage] | nostril | |
| insulin detemir 100 UNT/ML Injectable Solution | subq | |
| Avandia Oral Product | ORAL | |
| Glucophage Oral Product | | 0 |
| Regular Insulin, Human 8 UNT | Sq | |
| insulin pork, isophane 100 UNT/ML Injectable Suspension [NPH Iletin II] | ORAL | |
| exenatide 3.08 MG/ML [Bydureon] | gt | |
| Metformin hydrochloride 500 MG Oral Tablet | MISC | |
| Linagliptin 5 MG [Tradjenta] | by mouth. | |
| 3 ML insulin degludec 200 UNT/ML Pen Injector [Tresiba] | wearable injector) | |
| Glyburide Oral Tablet [Diabeta] | gt | |
| exenatide | subQ | |
| Metformin hydrochloride 1000 MG / saxagliptin 2.5 MG Extended Release Oral Tablet | po | |
| 3 ML insulin detemir 100 UNT/ML Pen Injector [Levemir] | subq. | |
| Jardiance Oral Product | PO | |
| 3 ML Regular Insulin, Human 100 UNT/ML Prefilled Syringe | iv | |

| | | |
|---|---|---|
| Glipizide Extended Release Oral Tablet [Glucotrol] | orally | |
| Metformin hydrochloride 1000 MG [Glumetza] | nostril | |
| 3 ML insulin detemir 100 UNT/ML Pen Injector [Levemir] | Sq | |
| glimepiride | iv | |
| Metformin Oral Tablet [Glucophage] | P.O. | |
| Insulin Lispro 100 UNT/ML Injectable Solution [Humalog] | PUMP | |
| Insulin Glargine 100 UNT/ML | subcut. | |
| Metformin hydrochloride 625 MG Oral Tablet | p.o. | |
| Regular Insulin, Human 100 UNT/ML Injectable Solution [Humulin R] | MISCELLANEOUS | |
| sitagliptin 100 MG [Januvia] | Nares | |
| Chlorpropamide Oral Tablet | | |
| Insulin Glargine 100 UNT/ML Injectable Solution | po | |
| insulin human, isophane 70 UNT/ML / Regular Insulin, Human 30 UNT/ML [Humulin] | intravenous | |
| liraglutide | PO | |
| Simvastatin 40 MG / sitagliptin 100 MG Oral Tablet [Juvisync] | ORAL | |
| insulin human, isophane 70 UNT/ML | po | |
| canagliflozin 300 MG [Invokana] | topical | |
| glimepiride 2 MG / pioglitazone 30 MG Oral Tablet [Duetact] | Oral | |
| insulin human, isophane 100 UNT/ML Injectable Suspension [Humulin N] | oral | |
| Metformin hydrochloride 500 MG / rosiglitazone 2 MG [Avandamet] | inhalatn | |
| Insulin Glargine 100 UNT/ML Injectable Solution [Lantus] | INTRADERMAL | |
| Bromocriptine 2.5 MG [Parlodel] | | 0 |
| pioglitazone Oral Tablet [Actos] | im | |
| pioglitazone 30 MG [Actos] | IM | |
| Insulin Glargine Injectable Solution | npo. | |
| Metformin hydrochloride 100 MG/ML | SQ | |
| dapagliflozin 5 MG [Farxiga] | subcutaneous | |
| Insulin Glargine | Subq | |
| Bromocriptine 1 MG Oral Tablet [Parlodel] | ORAL | |
| Metformin 500 MG / ropinirole 2 MG Oral Tablet | per tube | |
| glimepiride 2 MG | sub q | |
| 3 ML Insulin Glargine 100 UNT/ML Pen Injector [Basaglar] | other | |
| insulin pork, isophane 80 UNT/ML | subcutaneously. | |
| Insulin Glargine 100 UNT/ML [Lantus] | BY MOUTH | |
| insulin human, isophane 70 UNT/ML / Regular Insulin, Human 30 UNT/ML [Novolin] | subling | |

| | | |
|---|---|---|
| Insulin, Zinc, Human 100 UNT/ML Injectable Suspension | p.o. | |
| Insulin Lispro 200 UNT/ML Pen Injector [Humalog] | by mouth | |
| insulin detemir 100 UNT/ML Injectable Solution [Levemir] | intramuscular | |
| Lente Insulin, Beef 80 UNT/ML | p.o. | |
| Regular Insulin, Human 100 UNT/ML [Novolin R] | | |
| Glipizide 10 MG Extended Release Oral Tablet | per mouth | |
| 3 ML insulin human, isophane 100 UNT/ML Prefilled Syringe | subq | |
| Metformin hydrochloride 500 MG [Glucophage] | Inhalation | |
| 0.5 ML albiglutide 100 MG/ML Pen Injector [Tanzeum] | IM | |
| exenatide 0.005 MG/ACTUAT | sc | |
| glimepiride 1 MG [Amaryl] | TOPICAL | |
| sitagliptin 100 MG [Januvia] | IV | |
| Metformin | Subq | |
| 60 ACTUAT exenatide 0.005 MG/ACTUAT Pen Injector [Byetta] | sub Q | |
| insulin detemir 100 UNT/ML Injectable Solution [Levemir] | miscellaneous | |
| glimepiride 1 MG / rosiglitazone 4 MG Oral Tablet [Avandaryl] | p.o. | |
| 3 ML Insulin Glargine 100 UNT/ML Pen Injector [Lantus] | sl | |
| Glyburide 1.25 MG [Diabeta] | per tube | |
| glimepiride 2 MG Oral Tablet [Amaryl] | gt | |
| glimepiride 4 MG / pioglitazone 30 MG Oral Tablet [Duetact] | oral | |
| Metformin | Subcut | |
| dapagliflozin 10 MG Oral Tablet [Farxiga] | combination | |
| Prandin Oral Product | | |
| 3 ML insulin human, isophane 100 UNT/ML Prefilled Syringe | subQ | |
| Insulin, Aspart, Human 100 UNT/ML Injectable Solution [NovoLog] | CONT SUBCUTAN | |
| Metformin Oral Solution | P.O. | |
| insulin detemir 100 UNT/ML Injectable Solution [Levemir] | subQ | |
| Regular Insulin, Human 500 UNT/ML Injectable Solution [Humulin R] | DRAWN WITH U-100 SYRINGE | |
| glimepiride 4 MG | subcut | |
| Metformin Oral Tablet [Glucophage] | ORAL | |
| Insulin, Regular, Pork 40 UNT/ML Injectable Solution | by mouth | |
| pioglitazone Oral Tablet | BY MOUTH | |
| rosiglitazone | subcu | |
| exenatide 2 MG | subq | |
| 3 ML insulin degludec 200 UNT/ML Pen Injector [Tresiba] | | |
| Glyburide 1.25 MG | | 0 |
| 1.5 ML insulin human, isophane 100 UNT/ML Prefilled Syringe | p.o. | |
| Metformin 500 MG | NG | |

| | |
|---|---|
| insulin beef, isophane 40 UNT/ML | PO |
| Insulin, Aspart, Human | sQ |
| exenatide | subcut |
| Metformin 250 MG | subq |
| glimepiride | TOPICAL |
| Regular Insulin, Human 100 UNT/ML Injectable Solution | im |
| Regular Insulin, Human 100 UNT/ML Injectable Solution [Humulin R] | oral |
| Metformin Oral Tablet | per tube |
| Insulin, Aspart, Human | NPo |
| exenatide 2 MG | IM |
| alogliptin 25 MG Oral Tablet [Nesina] | p.o. |
| Insulin, Zinc, Pork | subcu. |
| exenatide 0.01 MG/ACTUAT | SQ |
| Lantus Injectable Product | TO DELIVER |
| Glyburide 2.5 MG Oral Tablet | NG |
| Tanzeum Injectable Product | SUBCUTANEOUSLY |
| Glyburide | trans |
| glimepiride 1 MG Oral Tablet [Amaryl] | by mouth. |
| Insulin, Aspart, Human | INH |
| dapagliflozin 5 MG Oral Tablet | p.o. |
| Metformin hydrochloride 1000 MG Extended Release Oral Tablet [Fortamet] | subcut |
| nateglinide 120 MG [Starlix] | SC |
| 3 ML liraglutide 6 MG/ML Pen Injector | SL |
| nateglinide | NG |
| Metformin hydrochloride 500 MG / repaglinide 1 MG Oral Tablet [PrandiMet] | PO |
| Metformin hydrochloride 1000 MG / sitagliptin 50 MG Oral Tablet | Oral |
| Metformin 250 MG | pGT |
| Duetact Oral Product | oral |
| empagliflozin 5 MG / Metformin hydrochloride 500 MG [Synjardy] | |
| insulin pork, isophane 40 UNT/ML Injectable Suspension | per tube |
| sitagliptin 100 MG [Januvia] | subQ |
| Metformin 500 MG / ropinirole 2 MG Oral Tablet | im |
| Insulin, Aspart, Human 100 UNT/ML Injectable Solution | Intravenous |
| Metformin hydrochloride 850 MG | IM |
| saxagliptin Oral Tablet [Onglyza] | oral |

| | |
|---|---|
| pioglitazone 45 MG [Actos] | po. |
| sitagliptin 100 MG [Januvia] | ORAL |
| Metformin hydrochloride 100 MG/ML | swallow |
| Metformin hydrochloride 500 MG / repaglinide 2 MG [PrandiMet] | subq |
| Insulin Lispro 100 UNT/ML Pen Injector [Humalog] | Sq |
| exenatide | Subcutaneous |
| Lente Insulin, Beef-Pork 40 UNT/ML | p.o. |
| Simvastatin 20 MG / sitagliptin 100 MG Oral Tablet [Juvisync] | po |
| Glyburide Oral Tablet | inh |
| Levemir Injectable Product | sq |
| glimepiride 4 MG | td |
| 3 ML Insulin, Aspart Protamine, Human 70 UNT/ML / Insulin, Aspart, Human 30 UNT/ML Pen Injector [NovoLog Mix] | npo |
| Lispro PRC Injectable Product | gt |
| Lispro PRC Injectable Product | orally |
| Metformin hydrochloride 850 MG Oral Tablet | subcu |
| canagliflozin 150 MG / Metformin hydrochloride 500 MG Oral Tablet [Invokamet] | |
| Metformin hydrochloride 1000 MG Oral Tablet | intravenous |
| insulin human, isophane 70 UNT/ML / Regular Insulin, Human 30 UNT/ML [Humulin] | ORAL |
| Glipizide 5 MG [Glucotrol] | topical |
| nateglinide 60 MG [Starlix] | per tube |
| Regular Insulin, Human 100 UNT/ML Injectable Solution [Humulin S] | subcu. |
| Actoplus Met Pill | po |
| Metformin Extended Release Oral Tablet | Oral |
| Januvia Pill | PO |
| Simvastatin 10 MG / sitagliptin 50 MG [Juvisync] | topical |
| sitagliptin 100 MG Oral Tablet [Januvia] | ORAL |
| sitagliptin 100 MG [Januvia] | inhalation |
| Jentadueto Oral Product | p.o. |
| Glipizide 10 MG | NG |
| Insulin Lispro | Sq |
| Metformin 500 MG | g - tube |
| 3 ML Insulin Glargine 100 UNT/ML Pen Injector [Lantus] | COMBO ROUTE) |
| miglitol Oral Product | PO |
| Regular Insulin, Human 500 UNT/ML | |

| | | |
|---|---|---|
| insulin human, isophane 70 UNT/ML / Regular Insulin, Human 30 UNT/ML Injectable Suspension [Humulin] | ORAL | |
| sitagliptin 25 MG Oral Tablet [Januvia] | subcutaneously | |
| Glyburide Pill | orally | |
| Regular Insulin, Human 20 UNT/ML | im | |
| Glipizide | Bedside | |
| Glyburide 2.5 MG | PER TUBE | |
| Simvastatin 10 MG / sitagliptin 50 MG [Juvisync] | IM | |
| nateglinide 90 MG Oral Tablet | | |
| Insulin Lispro 100 UNT/ML Injectable Solution [Humalog] | WITH INSULIN PUMP | |
| Insulin, Aspart, Human 100 UNT/ML Injectable Solution | po | |
| Metformin hydrochloride 500 MG / pioglitazone 15 MG Oral Tablet | Oral | |
| insulin human, isophane 34 UNT/ML / Regular Insulin, Human 6 UNT/ML Injectable Suspension | drip | |
| Metformin 1000 MG | sl | |
| Glipizide 5 MG / Metformin hydrochloride 500 MG Oral Tablet | Oral | |
| Insulin, Aspart Protamine, Human 70 UNT/ML / Insulin, Aspart, Human 30 UNT/ML Injectable Suspension [NovoLog Mix] | by mouth. | |
| Regular Insulin, Human | im / iv | |
| Metformin 1000 MG | sublingual | |
| 3 ML Insulin, Aspart, Human 100 UNT/ML Pen Injector [NovoLog] | intravenous | |
| Insulin, Zinc, Human | by mouth | |
| Insulin Lispro 100 UNT/ML Injectable Solution [Lispro PRC] | TOPICAL | |
| insulin human, isophane 60 UNT/ML / Regular Insulin, Human 40 UNT/ML Injectable Suspension | iv | |
| 3 ML insulin human, isophane 100 UNT/ML Prefilled Syringe | orally | |
| pioglitazone 30 MG [Actos] | subcu | |
| Insulin Lispro | Intracavernosal | |
| Glyburide 5 MG | sl | |
| Metformin hydrochloride 500 MG [Glucophage] | subq | |
| nateglinide 60 MG | Bedside | |
| Metformin hydrochloride 850 MG / pioglitazone 15 MG Oral Tablet | SQ | |
| insulin human, isophane 50 UNT/ML / Regular Insulin, Human 50 UNT/ML Injectable Suspension [Humulin] | SUBCUTANEOUS | |
| Insulin Lispro 100 UNT/ML Injectable Solution [Humalog] | INSULIN PUMP | |
| dulaglutide 3 MG/ML | subcutaneous | |
| repaglinide 0.5 MG Oral Tablet [Prandin] | | 0 |
| pioglitazone 30 MG [Actos] | sq | |

| | |
|---|---|
| 3 ML liraglutide 6 MG/ML Pen Injector [Victoza] | intravenously |
| Bromocriptine | MISC |
| Insulin, Zinc, Human | drip. |
| glimepiride | inhaled. |
| {60 (Regular Insulin, Human 12 UNT Inhalant Powder [Afrezza]) / 60 (Regular Insulin, Human 4 UNT Inhalant Powder [Afrezza]) / 60 (Regular Insulin, Human 8 UNT Inhalant Powder [Afrezza]) } Pack [Afrezza 180 Titration Pack- 60 (4 UNT), 60 (8 UNT), 60 (12 UN | INHALATION |
| Metformin hydrochloride 750 MG | nare |
| Insulin Lispro 100 UNT/ML Injectable Solution [Lispro PRC] | subcut |
| Glyburide 2.5 MG Oral Tablet | g-tube |
| insulin human, isophane 60 UNT/ML / Regular Insulin, Human 40 UNT/ML [Humulin] | subcu |
| Metformin hydrochloride 850 MG | bedside |
| Insulin Glargine | NG |
| Insulin Lispro 50 UNT/ML | SUBCUTANEOUSLY |
| Glipizide | NOSTRIL |
| Insulin Lispro 50 UNT/ML / Insulin, Protamine Lispro, Human 50 UNT/ML Injectable Suspension [Humalog Mix] | inhalation |
| Glyxambi Oral Product | orally |
| Regular Insulin, Human | inhaled |
| Insulin, Glulisine, Human 100 UNT/ML Injectable Solution [Apidra] | oral |
| Insulin Lispro 50 UNT/ML / Insulin, Protamine Lispro, Human 50 UNT/ML Injectable Suspension [Humalog Mix] | per tube |
| exenatide 0.01 MG/ACTUAT [Byetta] | SQ. |
| pioglitazone Oral Tablet | P.o. |
| Regular Insulin, Human 500 UNT/ML Injectable Solution [Humulin R] | SUBCUTANEOUSLY |
| insulin human, isophane 50 UNT/ML / Regular Insulin, Human 50 UNT/ML Injectable Suspension [Humulin] | subcu |
| Metformin hydrochloride 750 MG Extended Release Oral Tablet [Glucophage] | subcutaneously |
| Regular Insulin, Human 25 UNT/ML | by mouth |
| Actos Oral Product | ORAL |
| Metformin hydrochloride 750 MG | SC |
| Glipizide Oral Tablet | swallow. |
| Metformin Oral Tablet | PO. |
| Glyburide 5 MG Oral Tablet | topical |

| | |
|---|---|
| glimepiride 6 MG Oral Tablet | PO |
| Insulin Lispro 25 UNT/ML | ORAL |
| insulin human, isophane 50 UNT/ML | subcutaneous |
| Metformin hydrochloride 500 MG / rosiglitazone 4 MG Oral Tablet | Im |
| sitagliptin 100 MG [Januvia] | topically |
| Metformin hydrochloride 1000 MG / sitagliptin 50 MG [Janumet] | by mouth. |
| Invokamet Oral Product | PO |
| Avandaryl Pill | |
| Metformin Pill | g - tube |
| insulin human, isophane 80 UNT/ML / Regular Insulin, Human 20 UNT/ML [Humulin] | per tube |
| exenatide 0.005 MG/ACTUAT Pen Injector | SQ |
| insulin human, isophane 36 UNT/ML | SC |
| Insulin Glargine 100 UNT/ML [Lantus] | ORAL |
| Bromocriptine | td |
| Oseni Pill | |
| insulin pork, isophane 100 UNT/ML Injectable Suspension [NPH Iletin II] | SWALLOW |
| Metformin hydrochloride 850 MG | iv |
| Lixisenatide | subcutaneous |
| Insulin Glargine | td |
| Insulin, Aspart, Human | Sublingual |
| insulin, isophane Injectable Suspension [Novolin N] | nostril |
| Diabeta Pill | oral |
| pioglitazone 45 MG [Actos] | SQ |
| Bromocriptine 5 MG | ng |
| Insulin Lispro 100 UNT/ML | drip |
| glimepiride 1 MG | intravenously |
| Bromocriptine 2.5 MG [Parlodel] | gtube |
| Simvastatin 10 MG / sitagliptin 100 MG [Juvisync] | sq |
| Glyburide 2.5 MG Oral Tablet | ORAL |
| insulin human, isophane 50 UNT/ML / Regular Insulin, Human 50 UNT/ML Injectable Suspension [Humulin] | subQ |
| Chlorpropamide 200 MG / Phenformin 25 MG Oral Tablet | by mouth |
| Januvia Pill | oral |
| Glipizide | Sq. |
| insulin human, isophane 70 UNT/ML / Regular Insulin, Human 30 UNT/ML [Humulin] | MISC |
| Metformin hydrochloride 1000 MG Oral Tablet [Glucophage] | BY MOUTH |

| | | |
|---|---|---|
| canagliflozin 100 MG Oral Tablet [Invokana] | By mouth | |
| Glyset Oral Product | oral | |
| 24 HR Metformin hydrochloride 1000 MG / pioglitazone 15 MG Extended Release Oral Tablet [Actoplus Met] | | 0 |
| pioglitazone | IV | |
| empagliflozin 10 MG Oral Tablet [Jardiance] | Oral | |
| insulin pork, isophane 100 UNT/ML Injectable Suspension [NPH Iletin II] | Sl | |
| Metformin Oral Tablet [Glucophage] | NPO | |
| insulin human, isophane 70 UNT/ML / Regular Insulin, Human 30 UNT/ML Injectable Suspension | | 0 |
| insulin pork, isophane 100 UNT/ML Injectable Suspension [NPH Iletin II] | P.O. | |
| 3 ML Insulin Lispro 200 UNT/ML Pen Injector [Humalog] | subcu | |
| Insulin, Aspart, Human 100 UNT/ML Cartridge [NovoLog] | NOSTRIL | |
| Metformin / repaglinide Pill | | |
| insulin pork, isophane 100 UNT/ML Injectable Suspension [NPH Iletin II] | IM. | |
| exenatide 2 MG [Bydureon] | SL | |
| Acarbose 100 MG [Precose] | sq | |
| exenatide 0.01 MG/ACTUAT | subcu | |
| Metformin hydrochloride 500 MG [Glucophage] | topically | |
| insulin human, isophane 70 UNT/ML | drip. | |
| Insulin Glargine 100 UNT/ML [Lantus] | sub Q. | |
| Lispro PRC Injectable Product | | 0 |
| Acarbose 25 MG Oral Tablet [Precose] | G tube | |
| NovoLog Injectable Product | Subcutaneous. | |
| Metformin hydrochloride 1000 MG / pioglitazone 30 MG Extended Release Oral Tablet | PO | |
| insulin human, isophane 70 UNT/ML / Regular Insulin, Human 30 UNT/ML Prefilled Syringe [Humulin] | | |
| Insulin Lispro 50 UNT/ML / Insulin, Protamine Lispro, Human 50 UNT/ML Injectable Suspension [Humalog Mix] | P.O. | |
| Bromocriptine 2.5 MG Oral Tablet | Po | |
| Insulin, Protamine Lispro, Human | PO | |
| Glyburide 5 MG / Phenformin 50 MG Oral Tablet | subcutaneously | |
| Glipizide 10 MG Extended Release Oral Tablet | gt | |
| Insulin Lispro Cartridge [Humalog] | intramuscular | |
| Glipizide 2.5 MG | PER TUBE | |
| insulin pork, isophane 50 UNT/ML | Subcutaneous | |

| | |
|---|---|
| Metformin | SWALLOW |
| sitagliptin | sublingual |
| 3 ML insulin human, isophane 70 UNT/ML / Regular Insulin, Human 30 UNT/ML Pen Injector | subcutaneous |
| Humalog Injectable Product | Oral |
| liraglutide 6 MG/ML [Victoza] | subcutaneously. |
| Glyburide 4.5 MG Oral Tablet [Glycron] | ORAL |
| Tolazamide 250 MG Oral Tablet [Tolinase] | po |
| 60 ACTUAT exenatide 0.01 MG/ACTUAT Pen Injector [Byetta] | im |
| Insulin Lispro | td |
| glimepiride 2 MG / rosiglitazone 4 MG Oral Tablet [Avandaryl] | |
| canagliflozin 50 MG / Metformin hydrochloride 1000 MG Oral Tablet [Invokamet] | BY MOUTH |
| Glyburide 5 MG | inhaled |
| exenatide 2 MG | SC |
| alogliptin 12.5 MG Oral Tablet [Nesina] | |
| glimepiride 4 MG | topical |
| Insulin Lispro Injectable Solution | npo. |
| Regular Insulin, Human | Intra_Dermal |
| 0.5 ML albiglutide 100 MG/ML Pen Injector [Tanzeum] | subcu |
| 3 ML insulin degludec 200 UNT/ML Pen Injector [Tresiba] | Sub Q |
| Glipizide 10 MG | SL. |
| Regular Insulin, Human 100 UNT/ML Injectable Solution [Humulin S] | SUBCUTANEOUSLY |
| insulin pork, isophane 100 UNT/ML Injectable Suspension [NPH Iletin II] | inhaled |
| 3 ML Insulin, Aspart, Human 100 UNT/ML Cartridge [NovoLog] | topical |
| insulin human, isophane 100 UNT/ML Injectable Suspension [Humulin I] | orally |
| Metformin hydrochloride 1000 MG / pioglitazone 30 MG [Actoplus Met] | subq |
| Glumetza Oral Product | p.o. |
| nateglinide 30 MG Oral Tablet | PO |
| 1.5 ML insulin human, isophane 100 UNT/ML Prefilled Syringe | subcu |
| sitagliptin 50 MG [Januvia] | subq |
| Glipizide Oral Tablet | subcutaneously |
| empagliflozin 25 MG Oral Tablet [Jardiance] | mouth/throat |
| Metformin hydrochloride 500 MG / rosiglitazone 2 MG [Avandamet] | ou |
| Insulin Lispro | INH |

| | |
|---|---|
| Metformin Pill | BY MOUTH |
| 24 HR Glipizide 2.5 MG Extended Release Oral Tablet | Inhalation |
| Insulin, Aspart, Human 100 UNT/ML Injectable Solution [NovoLog] | contin. subcutaneous infusion |
| insulin human, isophane 60 UNT/ML / Regular Insulin, Human 40 UNT/ML [Humulin] | sq |
| insulin human, isophane 100 UNT/ML Injectable Suspension [Humulin N] | PO |
| Glipizide 10 MG | subcutaneous |
| 3 ML Insulin, Aspart, Human 100 UNT/ML Pen Injector [NovoLog] | oral |
| Glyburide 2.5 MG | TD |
| Glipizide 2.5 MG | subcutaneous |
| 24 HR Metformin hydrochloride 500 MG Extended Release Oral Tablet | ORAL |
| Toujeo Injectable Product | SQ |
| 3 ML Insulin, Glulisine, Human 100 UNT/ML Pen Injector [Apidra] | po |
| Regular Insulin, Human | SL |
| Metformin hydrochloride 1000 MG Oral Tablet | By Mouth |
| Glyxambi Pill | |
| Insulin Glargine Injectable Solution | ng |
| Glipizide Oral Product | ORAL |
| Metformin Oral Solution | pO |
| Metformin hydrochloride 850 MG Oral Tablet | NG |
| Insulin Lispro 100 UNT/ML Pen Injector [Humalog] | SubQ. |
| Glipizide 5 MG | im |
| sitagliptin 100 MG Oral Tablet | SEE COMMENT |
| sitagliptin Oral Tablet [Januvia] | IV |
| Metformin hydrochloride 850 MG | gt |
| 3 ML insulin detemir 100 UNT/ML Pen Injector [Levemir] | inhalation |
| Insulin Lispro 100 UNT/ML Injectable Solution [Humalog] | intramuscular |
| Lispro PRC Injectable Product | po |
| Regular Insulin, Human 100 UNT/ML Injectable Solution [Humulin R] | SUBCUTANOUS |
| Insulin, Aspart, Human 100 UNT/ML Injectable Solution [NovoLog] | SUBCUTANEOUS INFUSION VIA |
| glimepiride | by mouth. |
| Metformin | BY MOUTH |
| Insulin, Aspart, Human 100 UNT/ML Injectable Solution [NovoLog] | PER PUMP |
| Glipizide 5 MG [Glucotrol] | per tube |

| | | |
|---|---|---|
| repaglinide | sublingual | |
| Metformin hydrochloride 500 MG Oral Tablet | TUBE | |
| Insulin Glargine Injectable Solution [Lantus] | | 0 |
| Glyburide 6 MG | by mouth. | |
| insulin human, isophane 70 UNT/ML / Regular Insulin, Human 30 UNT/ML [Novolin] | inhalation | |
| dulaglutide | subcutaneous | |
| Novolin Injectable Product | Subcutaneously | |
| Regular Insulin, Human 40 UNT/ML | per tube | |
| Amaryl Oral Product | P.O. | |
| Insulin, Aspart, Human 100 UNT/ML Injectable Solution | orally | |
| empagliflozin 10 MG Oral Tablet [Jardiance] | subcutaneously | |
| Insulin Lispro Injectable Solution | Sq | |
| Regular Insulin, Human 500 UNT/ML Injectable Solution [Humulin R] | iv | |
| Insulin Glargine | ORAL | |
| Insulin Lispro 100 UNT/ML [Humalog] | by mouth | |
| Insulin Lispro 100 UNT/ML | gt | |
| Insulin Glargine 100 UNT/ML Injectable Solution [Lantus] | oral | |
| Insulin, Zinc, Human 100 UNT/ML Injectable Suspension [Novolin L] | Subcut | |
| Insulin Lispro 25 UNT/ML | drip. | |
| saxagliptin | IV | |
| insulin human, isophane 100 UNT/ML Injectable Suspension | subdermal | |
| Insulin Lispro 200 UNT/ML Pen Injector [Humalog] | po | |
| Insulin, Aspart, Human 100 UNT/ML Cartridge [NovoLog] | im | |
| Insulin Glargine 100 UNT/ML Pen Injector [Lantus] | sub q | |
| 60 ACTUAT exenatide 0.005 MG/ACTUAT Pen Injector [Byetta] | SUBCUTANEOUS | |
| insulin detemir 100 UNT/ML [Levemir] | ORAL | |
| canagliflozin 300 MG Oral Tablet | p.o. | |
| Metformin hydrochloride 500 MG [Glucophage] | NPO | |
| Metformin hydrochloride 750 MG Extended Release Oral Tablet [Glucophage] | ORAL | |
| Regular Insulin, Human 12 UNT | by mouth | |
| insulin beef, isophane 40 UNT/ML | p.o. | |
| Amaryl Pill | po | |
| Tolbutamide 250 MG Oral Tablet | p.o. | |
| Regular Insulin, Human 100 UNT/ML Injectable Solution [Novolin R] | SUB Q | |
| empagliflozin 10 MG Oral Tablet | BY MOUTH | |

| | |
|---|---|
| Regular Insulin, Human | p.o. / IM |
| Linagliptin | sc |
| Metformin 1000 MG | iv |
| Chlorpropamide Oral Tablet | Oral |
| Glipizide 5 MG Extended Release Oral Tablet [Glucotrol] | po |
| insulin human, isophane 50 UNT/ML / Regular Insulin, Human 50 UNT/ML Injectable Suspension [Humulin] | po |
| Metformin hydrochloride 1000 MG / pioglitazone 30 MG [Actoplus Met] | IV |
| Micronase Pill | |
| glimepiride 8 MG | subcutaneously |
| Metformin hydrochloride 100 MG/ML | NG |
| 3 ML insulin human, isophane 100 UNT/ML Prefilled Syringe [Novolin N] | oral |
| Metformin Extended Release Oral Tablet [Fortamet] | subcutaneously |
| insulin human, isophane 100 UNT/ML Pen Injector [Humulin N] | SUBCUTANEOUSLY |
| insulin pork, isophane 100 UNT/ML Injectable Suspension [NPH Iletin II] | nare |
| exenatide 2 MG [Bydureon] | SC. |
| 3 ML Insulin Lispro 200 UNT/ML Pen Injector [Humalog] | SubQ |
| Insulin Glargine Injectable Solution [Lantus] | PO |
| Insulin Glargine 100 UNT/ML [Lantus] | Im |
| 3 ML insulin human, isophane 70 UNT/ML / Regular Insulin, Human 30 UNT/ML Pen Injector | sq |
| glimepiride 6 MG Oral Tablet | p.o. |
| Bromocriptine 10 MG Oral Tablet | by mouth |
| Insulin, Aspart, Human 100 UNT/ML Cartridge [NovoLog] | sc. |
| Regular Insulin, Human 40 UNT/ML | by mouth |
| 3 ML liraglutide 6 MG/ML Pen Injector [Victoza] | nares |
| empagliflozin 25 MG Oral Tablet [Jardiance] | subcutaneous |
| 3 ML Insulin Glargine 100 UNT/ML Pen Injector [Basaglar] | subconjunctival |
| Simvastatin 10 MG / sitagliptin 50 MG [Juvisync] | per tube |
| Bromocriptine | subq |
| Glipizide 2.5 MG | sq |
| Glipizide 5 MG [Glucotrol] | sl |
| insulin human, isophane 70 UNT/ML / Regular Insulin, Human 30 UNT/ML [Humulin] | INTRAVENOUS |
| alogliptin 25 MG / pioglitazone 15 MG Oral Tablet [Oseni] | p.o. |
| Metformin hydrochloride 500 MG Oral Tablet | TUBE |
| 3 ML liraglutide 6 MG/ML Pen Injector [Saxenda] | 0 |

| | | |
|---|---|---|
| Insulin, Glulisine, Human | p.o. | |
| pioglitazone | IM | |
| Actos Pill | by mouth | |
| Metformin hydrochloride 500 MG / sitagliptin 50 MG Oral Tablet [Janumet] | P.O. | |
| insulin degludec Injectable Product | Subcutaneously | |
| insulin human, isophane 70 UNT/ML / Regular Insulin, Human 30 UNT/ML Injectable Suspension [Novolin] | SUBCUTANEOUS WITH MEAL | |
| 60 ACTUAT exenatide 0.01 MG/ACTUAT Pen Injector [Byetta] | intramuscular | |
| sitagliptin 50 MG [Januvia] | OU | |
| 3 ML Insulin Glargine 100 UNT/ML Pen Injector [Basaglar] | intramuscular | |
| Insulin, Aspart, Human 100 UNT/ML Cartridge [NovoLog] | swallow | |
| insulin pork, isophane 80 UNT/ML | Sub q | |
| Glyburide 5 MG | | 0 |
| Glyburide | inhaled | |
| 3 ML Insulin, Aspart, Human 100 UNT/ML Pen Injector [NovoLog] | intradermal | |
| Metformin hydrochloride 1000 MG / rosiglitazone 4 MG [Avandamet] | SQ | |
| insulin human, isophane 70 UNT/ML / Regular Insulin, Human 30 UNT/ML [Novolin] | Inhalation | |
| Glipizide 5 MG Extended Release Oral Tablet [Glucotrol] | orally | |
| Metformin hydrochloride 850 MG Oral Tablet | by mouth. | |
| Bromocriptine 2.5 MG Oral Tablet [Parlodel] | PER VAGINA | |
| exenatide 0.01 MG/ACTUAT | | 0 |
| exenatide 2 MG | subcu | |
| 3 ML Insulin, Glulisine, Human 100 UNT/ML Pen Injector [Apidra] | SUBCUTANEOUSLY | |
| insulin human, isophane 28 UNT/ML | | 0 |
| Metformin 1000 MG | G - tube | |
| 24 HR Metformin hydrochloride 500 MG / sitagliptin 50 MG Extended Release Oral Tablet [Janumet] | po | |
| Glyburide | topically | |
| glimepiride 4 MG | OU | |
| Bromocriptine | trans | |
| Glyburide 6 MG Oral Tablet [Glynase] | | 0 |
| Metformin Oral Tablet [Glucophage] | swallow | |
| Glipizide 2.5 MG [Glucotrol] | Po | |
| Regular Insulin, Human 30 UNT/ML | po | |
| canagliflozin Oral Tablet [Invokana] | PO | |
| exenatide 0.005 MG/ACTUAT [Byetta] | Subcutaneously. | |

| | |
|---|---|
| 3 ML Regular Insulin, Human 500 UNT/ML Pen Injector [Humulin R] | injection |
| insulin human, isophane 70 UNT/ML / Regular Insulin, Human 30 UNT/ML [Novolin] | bedside |
| dapagliflozin 10 MG [Farxiga] | topically |
| 3 ML Insulin, Aspart Protamine, Human 70 UNT/ML / Insulin, Aspart, Human 30 UNT/ML Pen Injector [NovoLog Mix] | OU |
| exenatide 0.01 MG/ACTUAT | sq |
| Insulin Glargine 100 UNT/ML [Lantus] | sc. |
| Insulin Glargine 100 UNT/ML | drip |
| Glipizide | Sublingual |
| insulin pork, isophane 100 UNT/ML Injectable Suspension [NPH Iletin II] | GTube |
| Linagliptin 5 MG [Tradjenta] | G - Tube |
| Metformin hydrochloride 500 MG Oral Tablet | pO |
| alogliptin 6.25 MG Oral Tablet [Nesina] | |
| Glyburide Oral Tablet [Glynase] | BY MOUTH |
| pioglitazone 45 MG [Actos] | G - Tube |
| 3 ML Insulin Glargine 100 UNT/ML Pen Injector [Basaglar] | subcutaneous |
| sitagliptin 50 MG [Januvia] | ng |
| Metformin | SQ. |
| miglitol | PER TUBE |
| Insulin Lispro 25 UNT/ML | npo. |
| insulin human, isophane 40 UNT/ML | Sub q |
| Glipizide 10 MG Oral Tablet | TOPICAL |
| insulin human, isophane 30 UNT/ML | subcutaneously |
| sitagliptin | SEE COMMENT |
| Insulin, Aspart, Human | Intravenous |
| Bromocriptine 0.8 MG Oral Tablet | by mouth |
| glimepiride | ORAL |
| glimepiride 1 MG | by mouth. |
| Glipizide 5 MG | NG |
| glimepiride 8 MG | gt |
| Metformin hydrochloride 500 MG / sitagliptin 50 MG Extended Release Oral Tablet [Janumet] | by mouth |
| Metformin hydrochloride 500 MG [Glucophage] | INH |
| Metformin hydrochloride 1000 MG [Glucophage] | sublingual |
| pioglitazone 45 MG [Actos] | npo |
| Insulin Glargine 100 UNT/ML [Lantus] | SEE COMMENT |
| Insulin, Glulisine, Human 100 UNT/ML Injectable Solution | SQ |

| | |
|---|---|
| Insulin Lispro 100 UNT/ML | subcutaneous. |
| exenatide 0.01 MG/ACTUAT Pen Injector | SUBCUT |
| Glipizide 5 MG Oral Tablet [Glucotrol] | p.o |
| Regular Insulin, Human 40 UNT/ML | oral |
| Bromocriptine Oral Tablet [Parlodel] | p.o. |
| 3 ML insulin human, isophane 100 UNT/ML Prefilled Syringe | subcu. |
| insulin pork, isophane 100 UNT/ML Injectable Suspension [NPH Iletin II] | naris |
| exenatide 0.005 MG/ACTUAT | SEE COMMENT |
| pioglitazone 15 MG [Actos] | SQ |
| Insulin, Zinc, Human 100 UNT/ML Injectable Suspension [Novolin L] | gt |
| Glucovance Pill | by mouth |
| glimepiride Oral Product | inhalation |
| canagliflozin 150 MG / Metformin hydrochloride 500 MG Oral Tablet [Invokamet] | BY MOUTH |
| Kombiglyze Pill | |
| Regular Insulin, Human 100 UNT/ML Injectable Solution [Humulin R] | ORAL |
| Glipizide | Sq |
| pioglitazone 30 MG [Actos] | subq |
| insulin human, isophane 80 UNT/ML / Regular Insulin, Human 20 UNT/ML [Humulin] | NG |
| Insulin Lispro 50 UNT/ML | Oral |
| pioglitazone 15 MG [Actos] | td |
| insulin human, isophane 70 UNT/ML | subcu |
| 3 ML Insulin, Aspart, Human 100 UNT/ML Pen Injector [NovoLog] | SUB Q |
| Metformin hydrochloride 1000 MG Oral Tablet | Trans |
| NovoLog Injectable Product | SC |
| Insulin Lispro Cartridge [Humalog] | SQ. |
| Metformin | Intravenous |
| Regular Insulin, Human 100 UNT/ML Injectable Solution [Humulin R] | SUBCUTANOIUS |
| Insulin, Aspart Protamine, Human 70 UNT/ML / Insulin, Aspart, Human 30 UNT/ML [NovoLog Mix] | topically |
| pramlintide acetate 0.6 MG/ML Injectable Solution [Symlin] | subcutaneous. |
| Insulin, Aspart, Human 100 UNT/ML Cartridge [NovoLog] | TOPICAL |
| glimepiride 2 MG / pioglitazone 30 MG [Duetact] | subcutaneous |
| nateglinide 120 MG [Starlix] | po. |
| glimepiride 2 MG [Amaryl] | topical |

| | |
|---|---|
| Amaryl Oral Product | 0 |
| miglitol Oral Product | by mouth |
| Metformin Oral Tablet [Metforming] | po |
| Acarbose 25 MG Oral Tablet [Precose] | G TUBE |
| dulaglutide | |
| Bromocriptine 10 MG Oral Capsule [Parlodel] | PO |
| Glipizide 10 MG Oral Tablet [Glucotrol] | P.O |
| 3 ML liraglutide 6 MG/ML Pen Injector | per tube |
| 1.5 ML Regular Insulin, Human 100 UNT/ML Prefilled Syringe | oral |
| 1.5 ML Regular Insulin, Human 100 UNT/ML Prefilled Syringe | po |
| Humalog Injectable Product | iv |
| 3 ML insulin human, isophane 100 UNT/ML Pen Injector [Humulin N] | SUBCUTANEOUSLY |
| Humulin R Injectable Product | SQ |
| Regular Insulin, Human 6 UNT/ML | subcu |
| Linagliptin 2.5 MG / Metformin hydrochloride 850 MG Oral Tablet [Jentadueto] | po |
| insulin human, isophane 100 UNT/ML Injectable Suspension [Humulin N] | po |
| Insulin Lispro Injectable Solution | SUBCUTANEOUSLY |
| Regular Insulin, Human 100 UNT/ML Injectable Solution [Velosulin] | gt |
| Insulin, Regular, Pork 40 UNT/ML Injectable Solution | SUBCUTANEOUS |
| Insulin Glargine | COMBO ROUTE) |
| Metformin hydrochloride 500 MG Oral Tablet [Glucophage] | IV. |
| Insulin Glargine | SC. |
| Osmotic 24 HR Metformin hydrochloride 1000 MG Extended Release Oral Tablet [Fortamet] | PO |
| Metformin Oral Tablet [Glucophage] | npo |
| Chlorpropamide 200 MG | po |
| liraglutide 6 MG/ML [Victoza] | SUBCUT |
| Metformin Extended Release Oral Tablet [Fortamet] | gt |
| 1.5 ML Insulin Glargine 300 UNT/ML Pen Injector [Toujeo] | Subq |
| Acarbose 25 MG Oral Tablet [Precose] | G - tube |
| Insulin, Aspart, Human 100 UNT/ML Injectable Solution [NovoLog] | Infusn (via wearable injector) |
| pioglitazone 15 MG [Actos] | subcutaneously |
| Insulin Glargine 300 UNT/ML | subcut |
| Byetta Injectable Product | npo |

| | |
|---|---|
| insulin human, isophane 90 UNT/ML / Regular Insulin, Human 10 UNT/ML [Humulin] | sc |
| glimepiride 4 MG Oral Tablet [Amaryl] | PER TUBE |
| Insulin Glargine | Intra_Muscular |
| Metformin hydrochloride 1000 MG Extended Release Oral Tablet [Glumetza] | gt |
| Metformin 500 MG | TD |
| Regular Insulin, Human | IPPB |
| insulin human, isophane 100 UNT/ML Injectable Suspension [Novolin N] | IV |
| Insulin Lispro 25 UNT/ML / Insulin, Protamine Lispro, Human 75 UNT/ML Injectable Suspension [Humalog Mix] | orally |
| insulin pork, isophane 100 UNT/ML Injectable Suspension [NPH Iletin II] | Intravenous |
| Lantus Injectable Product | npo |
| Insulin Glargine 100 UNT/ML Injectable Solution [Lantus] | po |
| canagliflozin 50 MG / Metformin hydrochloride 1000 MG Oral Tablet [Invokamet] | subcutaneous |
| pioglitazone | inhaled |
| Glipizide 2.5 MG Oral Tablet | p.o |
| Metformin hydrochloride 1000 MG [Glumetza] | ng |
| Glyburide Pill | PO |
| Avandia Oral Product | drip |
| Metformin hydrochloride 500 MG [Glucophage] | Sublingual |
| 24 HR Glipizide 5 MG Extended Release Oral Tablet [Glucotrol] | p.o. |
| Metformin hydrochloride 1000 MG Oral Tablet [Glucophage] | . |
| nateglinide 60 MG [Starlix] | subcu |
| Metformin hydrochloride 1000 MG / pioglitazone 30 MG [Actoplus Met] | TOPICAL |
| Insulin Lispro 200 UNT/ML [Humalog] | sq |
| Metformin hydrochloride 500 MG Oral Tablet | subcutaneously |
| Metformin / pioglitazone Oral Tablet | PO |
| glimepiride | orally |
| Glipizide 10 MG [Glucotrol] | inhalation |
| Regular Insulin, Human 100 UNT/ML Injectable Solution [Novolin R] | SUBCUTANEOUS PER PUMP |
| Bromocriptine 2.5 MG Oral Tablet [Parlodel] | GTUBE |
| Invokana Oral Product | |
| 3 ML Insulin Lispro 200 UNT/ML Pen Injector [Humalog] | p.o. |

| | | |
|---|---|---|
| Metformin hydrochloride 500 MG / pioglitazone 15 MG Oral Tablet | subcutaneous | |
| Insulin Lispro Cartridge [Humalog] | sq. | |
| Insulin, Aspart Protamine, Human 70 UNT/ML / Insulin, Aspart, Human 30 UNT/ML Injectable Suspension [NovoLog Mix] | NG | |
| Insulin, Aspart, Human | orally. | |
| Metformin hydrochloride 1000 MG Extended Release Oral Tablet | | 0 |
| 3 ML insulin human, isophane 100 UNT/ML Prefilled Syringe [Novolin N] | orally | |
| Glipizide 2.5 MG | per mouth | |
| insulin human, isophane 36 UNT/ML | subcu. | |
| Insulin Glargine Injectable Solution [Lantus] | IM injection | |
| 24 HR Glipizide 2.5 MG Extended Release Oral Tablet [Glucotrol] | PEG | |
| Metformin hydrochloride 750 MG | ORAL | |
| Insulin Glargine | J - tube | |
| insulin human, isophane 70 UNT/ML / Regular Insulin, Human 30 UNT/ML [Novolin] | MISC.(NON-DRUG; COMBO ROUTE) | |
| Regular Insulin, Human 4 UNT Inhalant Powder [Afrezza] | | 0 |
| Glyburide | SUBLING | |
| 3 ML insulin detemir 100 UNT/ML Pen Injector [Levemir] | Oral | |
| sitagliptin | subcutaneously | |
| Regular Insulin, Human | npo. | |
| insulin human, isophane 100 UNT/ML Injectable Suspension [Humulin N] | ORAL | |
| Metformin hydrochloride 500 MG Oral Tablet | QAM | |
| exenatide 2 MG [Bydureon] | subcutaneously. | |
| insulin human, isophane 90 UNT/ML | IM | |
| Insulin Glargine Injectable Solution | DRIP | |
| exenatide 0.01 MG/ACTUAT Pen Injector | by mouth | |
| Regular Insulin, Human 10 UNT/ML | bedside | |
| Insulin, Aspart, Human 100 UNT/ML Injectable Solution [NovoLog] | SubQ | |
| Insulin Lispro 100 UNT/ML Injectable Solution [Humalog] | ORAL | |
| Glucophage Oral Product | subcutaneous | |
| Insulin, Regular, Pork 40 UNT/ML Injectable Solution | subcutaneously | |
| glimepiride 3 MG | po. | |
| glimepiride 2 MG | TOPICAL | |
| Glipizide 10 MG Oral Tablet [Glucotrol] | g-tube | |
| glimepiride 4 MG | ou | |

| | | |
|---|---|---|
| Insulin, Glulisine, Human 100 UNT/ML Injectable Solution [Apidra] | Sq | |
| Glyburide | SUBCUTANEOUS | |
| 0.5 ML albiglutide 60 MG/ML Pen Injector [Tanzeum] | subcu | |
| Insulin, Aspart, Human 100 UNT/ML Injectable Solution [NovoLog] | SUBCUTANEOUS INFUSION PER | |
| insulin beef-pork, isophane 40 UNT/ML | | |
| Glipizide | ORAL | |
| 1.5 ML Regular Insulin, Human 100 UNT/ML Prefilled Syringe | po. | |
| Metformin / sitagliptin Oral Tablet [Janumet] | oral | |
| Metformin hydrochloride 100 MG/ML | by mouth. | |
| Glyburide | NG. | |
| sitagliptin 100 MG [Januvia] | npo | |
| dulaglutide | orally | |
| 24 HR Metformin hydrochloride 1000 MG / saxagliptin 2.5 MG Extended Release Oral Tablet [Kombiglyze] | Oral | |
| Regular Insulin, Human | Misc_Route | |
| sitagliptin 100 MG [Januvia] | ou | |
| Invokana Oral Product | pO | |
| Insulin Lispro 200 UNT/ML | bedside | |
| Insulin, Regular, Pork 40 UNT/ML Injectable Solution | drip | |
| 1.5 ML pramlintide acetate 1 MG/ML Pen Injector [Symlin] | | 0 |
| Insulin, Zinc, Human 100 UNT/ML Injectable Suspension [Novolin L] | sq | |
| 0.65 ML exenatide 3.08 MG/ML Pen Injector [Bydureon] | miscellaneous | |
| Metformin 500 MG | with spacer | |
| nateglinide Oral Tablet [Starlix] | orally | |
| insulin detemir 100 UNT/ML [Levemir] | inhalation | |
| nateglinide Oral Tablet | orally | |
| Metformin hydrochloride 500 MG Extended Release Oral Tablet | subcutaneous | |
| liraglutide | sc | |
| dapagliflozin 10 MG / Metformin hydrochloride 1000 MG [Xigduo] | po | |
| Metformin hydrochloride 500 MG Oral Tablet | SC | |
| rosiglitazone Oral Product | Oral | |
| insulin beef, isophane 40 UNT/ML | Subcutaneous | |
| glimepiride 4 MG Oral Tablet | by mouth. | |
| sitagliptin Oral Tablet [Januvia] | PO | |
| glimepiride 1 MG | td | |
| Insulin Lispro 100 UNT/ML Injectable Solution [Humalog] | INFUSION | |

| | | |
|---|---|---|
| Glipizide 5 MG / Metformin hydrochloride 500 MG Oral Tablet [Metaglip] | po. | |
| Regular Insulin, Human 30 UNT/ML | drip | |
| glimepiride 4 MG / pioglitazone 30 MG [Duetact] | MISC | |
| Metformin hydrochloride 750 MG Extended Release Oral Tablet [Glucophage] | BY MOUTH | |
| insulin human, isophane 34 UNT/ML | subcutaneously. | |
| alogliptin | nostril | |
| exenatide | IM | |
| Regular Insulin, Human 500 UNT/ML Injectable Solution | INFUSION | |
| insulin human, isophane 75 UNT/ML | po | |
| 3 ML Regular Insulin, Human 500 UNT/ML Pen Injector [Humulin R] | swallow | |
| Metformin hydrochloride 850 MG Oral Tablet [Glucophage] | Oral | |
| insulin detemir 100 UNT/ML [Levemir] | Sub q. | |
| Glyburide 5 MG | PER TUBE | |
| Glyburide | subQ | |
| Glynase Pill | p.o. | |
| Tolbutamide 500 MG | per tube | |
| insulin human, isophane 50 UNT/ML / Regular Insulin, Human 50 UNT/ML [Humulin] | | 0 |
| Metformin hydrochloride 100 MG/ML Oral Solution [Riomet] | orally | |
| Glyburide 6 MG | SEE COMMENT | |
| Metformin hydrochloride 500 MG Extended Release Oral Tablet | OU | |
| Insulin Glargine 100 UNT/ML Injectable Solution [Lantus] | INJECTION | |
| NovoLog Injectable Product | subcutaneous. | |
| Glyburide 5 MG / Metformin hydrochloride 500 MG Oral Tablet [Glucovance] | by mouth. | |
| insulin human, isophane 30 UNT/ML | sq | |
| Insulin Glargine 100 UNT/ML Injectable Solution [Lantus] | nostril | |
| Regular Insulin, Human 8 UNT | subq | |
| nateglinide | sublingual | |
| insulin human, isophane 100 UNT/ML Injectable Suspension [Humulin N] | orally | |
| Nesina Oral Product | | |
| Insulin Lispro | Im | |
| Glyburide 5 MG | bedside | |
| saxagliptin 5 MG [Onglyza] | subcutaneously | |
| Lantus Injectable Product | im | |
| Metformin hydrochloride 500 MG Oral Tablet | NPO | |

| | |
|---|---|
| Glipizide 5 MG [Glucotrol] | nare |
| Insulin, Aspart, Human 100 UNT/ML Cartridge [NovoLog] | drip |
| sitagliptin 100 MG [Januvia] | NPO |
| insulin human, isophane 85 UNT/ML | Sub Q |
| Insulin Lispro Cartridge [Humalog] | Intravenous |
| Insulin, Aspart Protamine, Human 70 UNT/ML / Insulin, Aspart, Human 30 UNT/ML Injectable Suspension [NovoLog Mix] | INFUSION |
| 3 ML Insulin Lispro 25 UNT/ML / Insulin, Protamine Lispro, Human 75 UNT/ML Pen Injector [Humalog Mix] | miscellaneous |
| glimepiride 4 MG | G - Tube |
| Insulin Lispro Cartridge [Humalog] | neb |
| Chlorpropamide | NPO. |
| Metformin hydrochloride 500 MG [Glucophage] | intramuscular |
| exenatide 0.01 MG/ACTUAT [Byetta] | sc. |
| pioglitazone 45 MG [Actos] | subcutaneous |
| Regular Insulin, Human 500 UNT/ML Injectable Solution [Humulin R] | SUBCUTANEOUSLY |
| Insulin Lispro Cartridge [Humalog] | topically |
| Glyburide | IV. |
| Insulin Glargine 100 UNT/ML Injectable Solution [Lantus] | oral / sublingual |
| Regular Insulin, Human 100 UNT/ML Injectable Solution | p.o. |
| rosiglitazone 4 MG Oral Tablet [Avandia] | . |
| Insulin Lispro Cartridge [Humalog] | nasogastric tube |
| empagliflozin 10 MG [Jardiance] | SC |
| Diabinese Pill | |
| Insulin Lispro | AS DIR |
| Metformin hydrochloride 1000 MG [Fortamet] | gt |
| glimepiride 3 MG [Amaryl] | po. |
| Glipizide | Intravenous |
| Metformin | Td |
| Metformin hydrochloride 500 MG Extended Release Oral Tablet [Fortamet] | nostril |
| 3 ML Insulin Glargine 100 UNT/ML Pen Injector [Lantus] | intramuscular |
| Glyburide 5 MG Oral Tablet [Diabeta] | by mouth. |
| Avandaryl Oral Product | p.o. |
| Metformin hydrochloride 500 MG Oral Tablet [Glucophage] | topically |
| 1.5 ML insulin human, isophane 100 UNT/ML Prefilled Syringe [Humulin N] | p.o. |
| Regular Insulin, Human | bedside. |
| liraglutide | po |

| | | |
|---|---|---|
| sitagliptin 100 MG | SEE COMMENT | |
| 24 HR Metformin hydrochloride 500 MG / sitagliptin 50 MG Extended Release Oral Tablet [Janumet] | orally | |
| 3 ML liraglutide 6 MG/ML Pen Injector [Saxenda] | subQ | |
| Insulin Lispro 200 UNT/ML [Humalog] | sc | |
| Metformin 250 MG Oral Tablet | | 0 |
| Insulin Lispro | BEDSIDE | |
| Regular Insulin, Human 5 UNT/ML Injectable Solution | subcut | |
| Regular Insulin, Human 6 UNT/ML | by mouth | |
| Glipizide 2.5 MG Extended Release Oral Tablet | ORAL | |
| Metformin hydrochloride 500 MG Oral Tablet | PO/PT | |
| {30 (Regular Insulin, Human 12 UNT Inhalant Powder) / 60 (Regular Insulin, Human 8 UNT Inhalant Powder) } Pack | SQ | |
| Metformin Pill | inhalation | |
| glimepiride | subcu | |
| insulin human, isophane 50 UNT/ML / Regular Insulin, Human 50 UNT/ML [Humulin] | p.o. | |
| Metformin hydrochloride 500 MG / rosiglitazone 4 MG Oral Tablet | BY MOUTH | |
| 24 HR Metformin hydrochloride 750 MG Extended Release Oral Tablet [Glucophage] | gt | |
| Glyburide | Sq | |
| Trulicity Injectable Product | po | |
| Insulin Lispro Cartridge [Humalog] | SUBCUTANEOUSLY | |
| Glyburide 2.5 MG | SC | |
| insulin human, isophane 70 UNT/ML / Regular Insulin, Human 30 UNT/ML [Novolin] | nostril | |
| Metformin hydrochloride 500 MG / sitagliptin 50 MG [Janumet] | nostril | |
| Onglyza Oral Product | | |
| Insulin Lispro Cartridge [Humalog] | sl | |
| Insulin Lispro 100 UNT/ML Injectable Solution [Lispro PRC] | | 0 |
| Acarbose | G tube | |
| Bromocriptine | im | |
| insulin human, isophane 100 UNT/ML Injectable Suspension [Humulin N] | Intravenous | |
| nateglinide 120 MG [Starlix] | SQ | |
| Glipizide 5 MG [Glucotrol] | inhalation | |
| liraglutide 6 MG/ML [Saxenda] | by mouth | |
| Metformin hydrochloride 500 MG Oral Tablet [Glucophage] | td | |
| repaglinide | gt | |

| | | |
|---|---|---|
| canagliflozin 100 MG Oral Tablet [Invokana] | gt | |
| Glipizide 10 MG | subQ | |
| pioglitazone 45 MG [Actos] | td | |
| Metformin hydrochloride 1000 MG [Glucophage] | Topical | |
| Glyburide 1.25 MG | per mouth | |
| 60 ACTUAT exenatide 0.005 MG/ACTUAT Pen Injector [Byetta] | COMBO ROUTE) | |
| nateglinide 60 MG Oral Tablet [Starlix] | NG | |
| Riomet Oral Liquid Product | | |
| Tolazamide Oral Product | p.o. | |
| 0.5 ML dulaglutide 3 MG/ML Auto-Injector [Trulicity] | SUBCU | |
| dulaglutide 1.5 MG/ML [Trulicity] | Subq | |
| glimepiride 2 MG | topically | |
| Regular Insulin, Human 50 UNT/ML | p.o. | |
| insulin degludec Pen Injector [Tresiba] | SUBCUT | |
| Metformin hydrochloride 1000 MG Oral Tablet | ORLLAY | |
| Insulin, Aspart, Human | G - TUBE | |
| sitagliptin 25 MG [Januvia] | topical | |
| Glyburide 5 MG | BY MOUTH | |
| 3 ML insulin human, isophane 100 UNT/ML Prefilled Syringe [Novolin N] | sc | |
| alogliptin 12.5 MG / pioglitazone 15 MG Oral Tablet [Oseni] | | |
| glimepiride 4 MG / rosiglitazone 8 MG Oral Tablet [Avandaryl] | PO | |
| Glyburide 5 MG Oral Tablet | subcutaneous | |
| repaglinide 2 MG | topical | |
| Glucovance Oral Product | Oral | |
| Metformin | topical. | |
| insulin beef, isophane 40 UNT/ML | SubQ | |
| Tresiba Injectable Product | by mouth | |
| Insulin, Aspart, Human | Subcut. | |
| Lispro PRC Injectable Product | drip | |
| pioglitazone 45 MG [Actos] | sq | |
| Insulin, Aspart Protamine, Human 70 UNT/ML / Insulin, Aspart, Human 30 UNT/ML [NovoLog Mix] | | 0 |
| insulin human, isophane 40 UNT/ML | Sub Q | |
| Regular Insulin, Human 4 UNT | oral | |
| Insulin Lispro 25 UNT/ML / Insulin, Protamine Lispro, Human 75 UNT/ML Injectable Suspension [Humalog Mix] | p.o. | |
| Metformin Oral Tablet [Metforming] | by mouth | |
| Tolazamide 500 MG Oral Tablet [Tolinase] | p.o. | |

| | |
|---|---|
| Insulin, Aspart, Human 100 UNT/ML Injectable Solution [NovoLog] | FOR EVERY 8 G CARBS |
| Bromocriptine 2.5 MG [Parlodel] | PER TUBE |
| Metformin 500 MG / ropinirole 2 MG Oral Tablet | swallow |
| Metformin hydrochloride 500 MG [Glucophage] | sl |
| Metformin hydrochloride 1000 MG / sitagliptin 50 MG [Janumet] | gt |
| Insulin, Aspart, Human 100 UNT/ML Injectable Solution [NovoLog] | gt |
| Glucovance Oral Product | |
| Humulin R Injectable Product | sub q |
| Regular Insulin, Human 500 UNT/ML Injectable Solution [Humulin R] | td |
| Insulin, Aspart, Human 100 UNT/ML Injectable Solution [NovoLog] | miscellaneous |
| Glyburide Oral Tablet [Glynase] | oral |
| liraglutide Injectable Product | oral |
| Metformin | Subcu |
| Simvastatin 10 MG / sitagliptin 50 MG [Juvisync] | sq |
| Glipizide | nares |
| Glyburide 1.25 MG / Metformin hydrochloride 250 MG Oral Tablet | Oral |
| 1.5 ML insulin human, isophane 70 UNT/ML / Regular Insulin, Human 30 UNT/ML Prefilled Syringe | subcutaneous. |
| Metformin hydrochloride 100 MG/ML Oral Solution [Riomet] | VIS G TUBE |
| pioglitazone 45 MG Oral Tablet [Actos] | PER TUBE |
| Insulin, Aspart, Human 100 UNT/ML Injectable Solution [NovoLog] | INFUSION PER INSULIN PUMP |
| 0.5 ML dulaglutide 1.5 MG/ML Auto-Injector [Trulicity] | subcu |
| 3 ML insulin human, isophane 100 UNT/ML Prefilled Syringe [Novolin N] | SQ |
| sitagliptin 25 MG Oral Tablet [Januvia] | inhalation |
| dapagliflozin 5 MG / Metformin hydrochloride 1000 MG [Xigduo] | Oral |
| Regular Insulin, Human | MISCELLANEOUS |
| pramlintide acetate 0.6 MG/ML [Symlin] | sq |
| insulin human, isophane 50 UNT/ML / Regular Insulin, Human 50 UNT/ML Injectable Suspension | subq |
| Levemir Injectable Product | Subcutaneous |
| exenatide 0.005 MG/ACTUAT [Byetta] | td |
| alogliptin 25 MG / pioglitazone 30 MG Oral Tablet [Oseni] | oral |
| Chlorpropamide | sublingual |

| | | |
|---|---|---|
| Insulin Glargine 300 UNT/ML | Oral | |
| Insulin Lispro 50 UNT/ML / Insulin, Protamine Lispro, Human 50 UNT/ML Injectable Suspension [Humalog Mix] | iv | |
| Metformin | pGT | |
| Metformin Oral Tablet [Glucophage] | TRANS | |
| Glipizide 5 MG Oral Tablet | rectal | |
| Glipizide 5 MG [Glucotrol] | ORAL | |
| Glipizide Oral Tablet | sublingual | |
| Ultralente Insulin, Human 100 UNT/ML Injectable Suspension | oral | |
| Glyburide 5 MG Oral Tablet | td | |
| 24 HR Metformin hydrochloride 500 MG / saxagliptin 5 MG Extended Release Oral Tablet [Kombiglyze] | PO | |
| 0.5 ML dulaglutide 1.5 MG/ML Auto-Injector [Trulicity] | gt | |
| Regular Insulin, Human 4 UNT | po | |
| empagliflozin Oral Tablet | | |
| glimepiride 4 MG [Amaryl] | sc | |
| exenatide 0.01 MG/ACTUAT [Byetta] | Sc | |
| 3 ML Regular Insulin, Human 500 UNT/ML Pen Injector [Humulin R] | PO | |
| Dymelor Oral Product | p.o. | |
| rosiglitazone 4 MG [Avandia] | po. | |
| insulin degludec 200 UNT/ML | subcutaneously | |
| Insulin, Zinc, Human | | 0 |
| Glyburide 1.5 MG | npo | |
| Glipizide Oral Product | By Mouth | |
| Regular Insulin, Human 10 UNT/ML | npo. | |
| Insulin, Zinc, Human | sc. | |
| Bromocriptine 5 MG Oral Capsule | Oral | |
| rosiglitazone 4 MG Oral Tablet | oral | |
| Exubera Inhalant Product | INHALATION | |
| 3 ML insulin detemir 100 UNT/ML Pen Injector [Levemir] | intravenously | |
| Tolbutamide Oral Product | po | |
| insulin human, isophane 32 UNT/ML | subcutaneously | |
| 3 ML insulin degludec 200 UNT/ML Pen Injector [Tresiba] | one nostril | |
| 1.5 ML insulin human, isophane 70 UNT/ML / Regular Insulin, Human 30 UNT/ML Prefilled Syringe | NPO | |
| exenatide 0.005 MG/ACTUAT [Byetta] | sub Q | |
| insulin pork, isophane 100 UNT/ML Injectable Suspension | IM | |
| Glyburide 5 MG | Sl | |
| rosiglitazone 4 MG [Avandia] | topically | |

| | |
|---|---|
| Insulin, Aspart, Human 100 UNT/ML Cartridge [NovoLog] | sq. |
| Metformin | BY MOUTH |
| 3 ML Insulin, Aspart, Human 100 UNT/ML Pen Injector [NovoLog] | CORRECTION SCALE |
| exenatide 2 MG | subcutaneous. |
| Parlodel Pill | po |
| insulin human, isophane 70 UNT/ML / Regular Insulin, Human 30 UNT/ML [Humulin] | iv |
| Riomet Oral Product | |
| Precose Pill | by mouth |
| Insulin, Zinc, Human 100 UNT/ML Injectable Suspension [Novolin L] | SEE COMMENT |
| 3 ML Insulin Glargine 100 UNT/ML Pen Injector [Lantus] | SUBQ |
| sitagliptin 50 MG [Januvia] | by mouth. |
| Lantus Injectable Product | HEPLOCK |
| Glyburide Oral Tablet | po. |
| insulin pork, isophane 80 UNT/ML | by mouth |
| Insulin Lispro | _ |
| insulin detemir Injectable Product | SC |
| 3 ML insulin detemir 100 UNT/ML Pen Injector [Levemir] | subcutaneous |
| Insulin Glargine 100 UNT/ML Injectable Solution | COMBO ROUTE) |
| Metformin hydrochloride 1000 MG Oral Tablet | ORALLY |
| glimepiride 4 MG | swallow |
| liraglutide 6 MG/ML [Victoza] | Neb |
| Metformin 500 MG | SC. |
| 3 ML insulin human, isophane 100 UNT/ML Prefilled Syringe [Novolin N] | po |
| Insulin Lispro Injectable Solution [Humalog] | Subcut |
| Insulin, Aspart, Human 100 UNT/ML Injectable Solution [NovoLog] | subcutaneous (via wearable injector) |
| Metformin Extended Release Oral Tablet [Fortamet] | swallow |
| Glyburide Oral Tablet | drip. |
| Regular Insulin, Human 100 UNT/ML Injectable Solution | PO |
| Insulin, Aspart, Human 100 UNT/ML | subQ |
| Insulin, Aspart, Human | SL |
| Metformin hydrochloride 1000 MG Oral Tablet | iv |
| Regular Insulin, Human | intravenous |
| 3 ML liraglutide 6 MG/ML Pen Injector [Saxenda] | subdermal |
| Glipizide Oral Tablet | orally |
| pioglitazone 30 MG [Actos] | po. |
| Regular Insulin, Human 40 UNT/ML | p.o. |

| | |
|---|---|
| Metformin Extended Release Oral Tablet | ORAL |
| nateglinide 60 MG Oral Tablet [Starlix] | p.o |
| Metformin hydrochloride 500 MG [Glucophage] | OU |
| Glipizide 10 MG [Glucotrol] | po. |
| Glipizide 10 MG | Sq |
| Glyburide Oral Tablet | gt |
| Metformin hydrochloride 750 MG Extended Release Oral Tablet [Glucophage] | by mouth. |
| Linagliptin 5 MG Oral Tablet [Tradjenta] | ORAL (PER PEG) |
| Metformin hydrochloride 100 MG/ML | sub q |
| glimepiride 2 MG | SQ |
| Metformin hydrochloride 850 MG [Glucophage] | topically |
| Glyburide 2.5 MG Oral Tablet | p.o |
| nateglinide | gt |
| Insulin, Aspart, Human 100 UNT/ML Injectable Solution | Swish and spit |
| Glipizide 2.5 MG | subcu |
| 3 ML Insulin, Aspart, Human 100 UNT/ML Pen Injector [NovoLog] | subdermal |
| Metformin 400 MG | BY MOUTH |
| 3 ML Insulin Glargine 100 UNT/ML Pen Injector [Lantus] | SUBCUTANEUS |
| Metformin hydrochloride 750 MG Extended Release Oral Tablet | sl |
| insulin human, isophane 100 UNT/ML Injectable Suspension | sq |
| Regular Insulin, Human | INJ |
| Tolazamide 250 MG Oral Capsule | p.o. |
| NovoLog Injectable Product | Subcutaneous |
| 24 HR Metformin hydrochloride 750 MG Extended Release Oral Tablet | ORALLY |
| Metformin hydrochloride 1000 MG / rosiglitazone 4 MG [Avandamet] | sl |
| Glipizide | SubCut |
| Glipizide 2.5 MG [Glucotrol] | per mouth |
| Glyburide 2.5 MG Oral Tablet | PO/PT |
| Insulin Lispro 25 UNT/ML | SUBCUTANEOUSLY |
| liraglutide 6 MG/ML [Victoza] | subc |
| 3 ML insulin human, isophane 100 UNT/ML Prefilled Syringe [Novolin N] | subQ |
| Regular Insulin, Human 5 UNT/ML Injectable Solution | SQ |
| 3 ML Insulin Glargine 100 UNT/ML Pen Injector [Lantus] | IV. |
| insulin detemir 100 UNT/ML [Levemir] | p.o. |
| glimepiride | neb |
| Metformin / rosiglitazone Oral Product | po |

| | |
|---|---|
| insulin beef-pork, isophane 100 UNT/ML Injectable Suspension [Iletin NPH] | SUBCUTANEOUSLY |
| Glyburide 2.5 MG Oral Tablet | NPO |
| 24 HR Metformin hydrochloride 500 MG Extended Release Oral Tablet [Glucophage] | ORAL WITH FOOD |
| canagliflozin 150 MG / Metformin hydrochloride 500 MG Oral Tablet [Invokamet] | Oral |
| insulin pork, isophane 100 UNT/ML Injectable Suspension [NPH Iletin II] | TOPICAL |
| Gliclazide Extended Release Oral Tablet | |
| repaglinide | subcutaneous |
| 3 ML Insulin, Aspart Protamine, Human 70 UNT/ML / Insulin, Aspart, Human 30 UNT/ML Pen Injector [NovoLog Mix] | iv |
| Metformin 250 MG | ORAL |
| repaglinide Oral Tablet [Prandin] | p.o. |
| Insulin Lispro Cartridge [Humalog] | IV. |
| sitagliptin 50 MG [Januvia] | gt |
| Metformin hydrochloride 850 MG Oral Tablet [Glucophage] | p.o |
| Insulin, Aspart, Human 100 UNT/ML Injectable Solution [NovoLog] | SUBCUTANEOUS,INS ULIN PUMP |
| 3 ML Insulin, Aspart, Human 100 UNT/ML Pen Injector [NovoLog] | SUB |
| Tolbutamide Oral Tablet | |
| Insulin Lispro Cartridge [Humalog] | Npo |
| Glipizide | per tube. |
| Metformin hydrochloride 850 MG Oral Tablet [Metforming] | |
| Glipizide 2.5 MG Extended Release Oral Tablet | SQ |
| 60 ACTUAT exenatide 0.01 MG/ACTUAT Pen Injector [Byetta] | OU |
| dapagliflozin 10 MG Oral Tablet [Farxiga] | subcutaneous |
| Acarbose 100 MG [Precose] | subcutaneously |
| Insulin Glargine | NOSTRIL |
| Glyburide 2.5 MG | subcutaneous |
| sitagliptin 50 MG Oral Tablet [Januvia] | PER TUBE |
| Acarbose Oral Tablet [Precose] | po |
| insulin beef, isophane 80 UNT/ML Injectable Suspension | SUBCUTANEOUS |
| 1.5 ML insulin human, isophane 70 UNT/ML / Regular Insulin, Human 30 UNT/ML Prefilled Syringe | PO |
| pioglitazone 45 MG [Actos] | by mouth. |
| Metformin | Subcutaneously |
| 3 ML Insulin Lispro 100 UNT/ML Pen Injector [Humalog] | INSULIN PUMP |

| | | |
|---|---|---|
| Metformin hydrochloride 1000 MG / pioglitazone 30 MG [Actoplus Met] | subcutaneous | |
| glimepiride Oral Tablet [Amaryl] | subcutaneously | |
| insulin human, isophane 70 UNT/ML / Regular Insulin, Human 30 UNT/ML [Humulin] | td | |
| Insulin, Aspart, Human | IM. | |
| 3 ML Insulin Lispro 100 UNT/ML Cartridge [Humalog] | INFUSI | |
| Glipizide | TOPICAL | |
| sitagliptin 50 MG | td | |
| Insulin Lispro 100 UNT/ML Injectable Solution [Lispro PRC] | IV | |
| Ultralente Insulin, Human 100 UNT/ML Injectable Suspension | p.o. | |
| 24 HR Metformin hydrochloride 1000 MG / pioglitazone 30 MG Extended Release Oral Tablet [Actoplus Met] | orally | |
| Glipizide 10 MG Extended Release Oral Tablet [Glucotrol] | orally | |
| Glipizide 2.5 MG Extended Release Oral Tablet [Glucotrol] | PO | |
| Metformin hydrochloride 500 MG Extended Release Oral Tablet | oral | |
| insulin degludec 200 UNT/ML | gt | |
| insulin pork, isophane 100 UNT/ML Injectable Suspension [NPH Iletin II] | Drip. | |
| 1.5 ML Regular Insulin, Human 100 UNT/ML Prefilled Syringe | subc | |
| Glipizide 5 MG | drip | |
| Glyburide 1.5 MG Oral Tablet | PER TUBE | |
| insulin detemir 100 UNT/ML [Levemir] | Subcut | |
| 24 HR Glipizide 5 MG Extended Release Oral Tablet | | 0 |
| Glyburide 6 MG [Glynase] | sq | |
| Metformin hydrochloride 100 MG/ML Oral Solution [Riomet] | other | |
| insulin human, isophane 100 UNT/ML Injectable Suspension [Humulin N] | nostril | |
| Riomet Oral Product | p.o. | |
| exenatide 0.01 MG/ACTUAT [Byetta] | COMBO ROUTE) | |
| insulin degludec 200 UNT/ML | SQ | |
| Glipizide | INH. | |
| 3 ML insulin degludec 100 UNT/ML Pen Injector [Tresiba] | topical | |
| Metformin hydrochloride 500 MG [Glucophage] | td | |
| alogliptin 12.5 MG / Metformin hydrochloride 1000 MG Oral Tablet [Kazano] | Oral | |
| insulin human, isophane 28 UNT/ML | subcu. | |
| insulin detemir Injectable Product | by mouth | |
| Glucophage Oral Product | SC | |
| Glipizide 5 MG [Glucotrol] | subcutaneous | |

| | | |
|---|---|---|
| Insulin Lispro 100 UNT/ML Injectable Solution [Humalog] | SUBCUTANSOUS | |
| insulin human, isophane 40 UNT/ML | IV | |
| exenatide | gt | |
| Glyburide 6 MG [Glynase] | subcutaneous | |
| Saxenda Injectable Product | oral | |
| Glipizide 5 MG Oral Tablet | feeding tube | |
| dapagliflozin 5 MG [Farxiga] | subcutaneously | |
| Glyburide 2.5 MG Oral Tablet [Diabeta] | BY MOUTH | |
| Metformin Pill | p.o. | |
| albiglutide 60 MG/ML [Tanzeum] | by mouth | |
| Metformin hydrochloride 500 MG [Fortamet] | PO | |
| Glyburide / Phenformin Pill | | |
| insulin human, isophane 70 UNT/ML / Regular Insulin, Human 30 UNT/ML [Novolin] | Td | |
| glimepiride 1 MG | SC | |
| Regular Insulin, Human 100 UNT/ML Injectable Solution [Velosulin] | iv | |
| Metformin hydrochloride 500 MG / rosiglitazone 2 MG [Avandamet] | IV. | |
| 0.5 ML dulaglutide 1.5 MG/ML Auto-Injector [Trulicity] | p.o. | |
| glimepiride 1 MG | NPO. | |
| 1.5 ML insulin human, isophane 100 UNT/ML Prefilled Syringe | per tube | |
| Glipizide 5 MG | Sublingual | |
| Metformin 1000 MG | sc | |
| Regular Insulin, Human 24 UNT/ML | po | |
| Humulin Injectable Product | | 0 |
| Metformin hydrochloride 1000 MG Oral Tablet [Glucophage] | BY MOUTH | |
| Insulin Glargine 300 UNT/ML Pen Injector | SC | |
| rosiglitazone | iv | |
| empagliflozin 25 MG / Linagliptin 5 MG Oral Tablet [Glyxambi] | | 0 |
| 3 ML Insulin Lispro 200 UNT/ML Pen Injector | Subcutaneous | |
| Fortamet Oral Product | BY MOUTH | |
| canagliflozin 150 MG / Metformin hydrochloride 1000 MG Oral Tablet [Invokamet] | PO | |
| exenatide 0.005 MG/ACTUAT | sq | |
| insulin human, isophane 85 UNT/ML | sub q | |
| Insulin Lispro Cartridge [Humalog] | intravenously | |
| Insulin, Aspart, Human 100 UNT/ML Cartridge [NovoLog] | BY MOUTH | |
| glimepiride 1 MG Oral Tablet [Amaryl] | PEG | |
| Insulin Lispro | Drip | |

| | |
|---|---|
| Gliclazide 80 MG Oral Tablet | po |
| Glyburide | SL |
| Metformin hydrochloride 1000 MG / saxagliptin 2.5 MG [Kombiglyze] | subcutaneous |
| Insulin Glargine Injectable Solution | MISCELLANEOUS |
| insulin human, isophane 70 UNT/ML / Regular Insulin, Human 30 UNT/ML Injectable Suspension [Novolin] | by mouth |
| glimepiride 3 MG Oral Tablet [Amaryl] | ORAL |
| saxagliptin Oral Tablet | BY MOUTH |
| Bromocriptine 50 MG | p.o. |
| Acarbose Oral Tablet | By mouth |
| pioglitazone | sq |
| 3 ML Regular Insulin, Human 100 UNT/ML Prefilled Syringe [Novolin R] | SUBQ |
| Ultralente Insulin, Human 100 UNT/ML Injectable Suspension | subQ |
| Metformin hydrochloride 1000 MG Extended Release Oral Tablet [Fortamet] | nostril |
| Invokamet Oral Product | oral |
| 3 ML liraglutide 6 MG/ML Pen Injector [Saxenda] | SUBCU |
| miglitol Oral Tablet [Glyset] | po |
| Insulin Glargine Injectable Solution | nostril |
| dapagliflozin 5 MG Oral Tablet | po |
| Glipizide Extended Release Oral Tablet [Glucotrol] | PO |
| Glyburide 5 MG [Calabren] | |
| Ultralente Insulin, Human 100 UNT/ML Injectable Suspension [Humulin U Ultralente] | po |
| Glipizide 5 MG Oral Tablet | Im |
| glimepiride 1 MG | Im |
| sitagliptin 100 MG [Januvia] | topical |
| Glipizide Oral Tablet | ORAL |
| Glipizide | Topical |
| Metformin Extended Release Oral Tablet | IV |
| Linagliptin 5 MG [Tradjenta] | ng |
| Metformin hydrochloride 500 MG / sitagliptin 50 MG [Janumet] | NG |
| Symlin Injectable Product | MISCELLANEOUS |
| Diabeta Pill | gt |
| Metformin hydrochloride 750 MG | td |
| glimepiride 2 MG Oral Tablet | P.O. |
| glimepiride 4 MG | Im |

| | | |
|---|---|---|
| 3 ML insulin human, isophane 100 UNT/ML Prefilled Syringe [Novolin N] | subcu | |
| Regular Insulin, Human 8 UNT | per tube | |
| Metformin hydrochloride 100 MG/ML | sq | |
| Invokana Oral Product | BY MOUTH | |
| Chlorpropamide 100 MG Oral Tablet [Diabinese] | | 0 |
| Simvastatin 10 MG / sitagliptin 100 MG [Juvisync] | SubQ | |
| Regular Insulin, Human | INHALATN | |
| Insulin, Aspart, Human | Inhalatn | |
| glimepiride 4 MG Oral Tablet | ORAL | |
| Insulin Glargine 100 UNT/ML [Lantus] | SUBCUTANEOUS | |
| 3 ML insulin detemir 100 UNT/ML Pen Injector [Levemir] | Topical | |
| Metformin hydrochloride 500 MG [Glucophage] | iv | |
| repaglinide Oral Tablet | by mouth | |
| pioglitazone 30 MG [Actos] | SC | |
| Avandaryl Oral Product | oral | |
| Metformin hydrochloride 850 MG Oral Tablet | P.O | |
| pioglitazone 15 MG [Actos] | subcutaneous | |
| Glipizide | sQ | |
| Chlorpropamide 100 MG Oral Tablet | BY MOUTH | |
| Metformin hydrochloride 750 MG | topically | |
| glimepiride 3 MG | SubQ | |
| 1.5 ML insulin human, isophane 100 UNT/ML Prefilled Syringe | SQ | |
| alogliptin 6.25 MG Oral Tablet [Nesina] | p.o. | |
| insulin human, isophane 70 UNT/ML / Regular Insulin, Human 30 UNT/ML [Humulin] | Bedside | |
| Insulin Lispro Injectable Solution | SEE COMMENT | |
| Glyburide 5 MG / Metformin hydrochloride 500 MG [Glucovance] | per tube | |
| Insulin, Aspart, Human | sq. | |
| Regular Insulin, Human 100 UNT/ML Injectable Solution [Humulin R] | PO | |
| canagliflozin 300 MG [Invokana] | SC | |
| Bromocriptine Oral Capsule [Parlodel] | g - tube | |
| sitagliptin 100 MG [Januvia] | inh | |
| 60 ACTUAT exenatide 0.005 MG/ACTUAT Pen Injector [Byetta] | subcu. | |
| Insulin Lispro | ou | |
| exenatide 2 MG [Bydureon] | subcut. | |
| insulin human, isophane 70 UNT/ML / Regular Insulin, Human 30 UNT/ML [Humulin] | injection | |
| Glipizide Oral Tablet [Glucotrol] | by MOUTH | |

| | |
|---|---|
| Insulin, Aspart, Human | SUBCUTANEOUSLY |
| saxagliptin 2.5 MG [Onglyza] | bedside. |
| glimepiride 1 MG | po. |
| exenatide 0.01 MG/ACTUAT [Byetta] | gt |
| 3 ML Insulin Lispro 100 UNT/ML Cartridge [Humalog] | IM |
| Glyburide 2.5 MG / Metformin hydrochloride 500 MG [Glucovance] | PER TUBE |
| Metformin hydrochloride 850 MG | IV |
| Insulin Lispro 100 UNT/ML Injectable Solution [Humalog] | PUMP |
| 0.5 ML dulaglutide 1.5 MG/ML Auto-Injector | subcutaneously |
| Humulin Injectable Product | SUBCUTANEOUSLY |
| 3 ML Regular Insulin, Human 100 UNT/ML Prefilled Syringe [Novolin R] | sub Q |
| Metformin hydrochloride 500 MG Oral Tablet | mouth/throat |
| rosiglitazone 4 MG Oral Tablet | Oral |
| Bydureon Injectable Product | by mouth |
| exenatide 0.01 MG/ACTUAT [Byetta] | td |
| Metformin hydrochloride 500 MG / rosiglitazone 2 MG [Avandamet] | OU |
| Metformin hydrochloride 500 MG / repaglinide 2 MG [PrandiMet] | subcutaneous |
| Regular Insulin, Human Pen Injector [Humulin R] | subcut. |
| Metformin Oral Tablet [Glucophage] | ORALLY |
| insulin degludec 200 UNT/ML | iv |
| insulin beef, isophane 100 UNT/ML Injectable Suspension | subcutaneous |
| Insulin Lispro 100 UNT/ML Injectable Solution [Lispro PRC] | gt |
| Insulin Lispro 25 UNT/ML | intravenous |
| Glyburide | per tube |
| Glyburide 1.25 MG Oral Tablet [Micronase] | ORAL |
| canagliflozin 100 MG Oral Tablet | oral |
| Humalog Mix Injectable Product | SUBCUTANEOUSLY |
| Regular Insulin, Human 12 UNT | po |
| empagliflozin 25 MG / Linagliptin 5 MG Oral Tablet | p.o. |
| Insulin Glargine 100 UNT/ML [Lantus] | Sub Q. |
| Lantus Injectable Product | ADDITIVE |
| Metformin hydrochloride 500 MG / sitagliptin 50 MG Oral Tablet [Janumet] | ORAL |
| Tolazamide 500 MG Oral Tablet | PO |
| 24 HR Metformin hydrochloride 500 MG Extended Release Oral Tablet | NPO |

| | | |
|---|---|---|
| Linagliptin 5 MG [Tradjenta] | orally. | |
| insulin human, isophane 100 UNT/ML Injectable Suspension [Novolin N] | oral | |
| glimepiride | SubQ | |
| Regular Insulin, Human 40 UNT/ML | SC | |
| Insulin Glargine 100 UNT/ML [Lantus] | PO. | |
| Regular Insulin, Human 100 UNT/ML Injectable Solution | intravenous push | |
| alogliptin 12.5 MG Oral Tablet | by mouth | |
| rosiglitazone | IV | |
| Regular Insulin, Human 12 UNT | subcu | |
| Metformin 1000 MG | oral. | |
| Metformin | per tube. | |
| insulin human, isophane 70 UNT/ML / Regular Insulin, Human 30 UNT/ML [Novolin] | G - tube | |
| Metformin 1000 MG | inhalation | |
| insulin human, isophane 40 UNT/ML | subc | |
| Regular Insulin, Human | NOTE | |
| alogliptin / Metformin Oral Tablet [Kazano] | by mouth | |
| 60 ACTUAT exenatide 0.005 MG/ACTUAT Pen Injector [Byetta] | injection | |
| Metformin Oral Tablet | | 0 |
| Regular Insulin, Human 40 UNT/ML | sub Q | |
| Metformin Pill | Im | |
| canagliflozin 300 MG [Invokana] | iv | |
| Insulin Lispro | Npo | |
| nateglinide 180 MG | nostril | |
| Glumetza Oral Product | Oral | |
| Glipizide 10 MG | sc | |
| Ultralente Insulin, Human 100 UNT/ML Injectable Suspension | SEE COMMENT | |
| Insulin, Aspart Protamine, Human 70 UNT/ML / Insulin, Aspart, Human 30 UNT/ML Injectable Suspension [NovoLog Mix] | SEE COMMENT | |
| nateglinide 30 MG Oral Tablet | Oral | |
| insulin degludec 100 UNT/ML Pen Injector [Tresiba] | subcut | |
| Insulin, Aspart, Human | INSUPUMP | |
| 0.65 ML exenatide 3.08 MG/ML Pen Injector [Bydureon] | subcutaneous | |
| glimepiride 1 MG [Amaryl] | inhalation | |
| glimepiride Oral Tablet [Amaryl] | TOPICAL | |
| Bromocriptine Pill | PO | |
| insulin human, isophane 80 UNT/ML / Regular Insulin, Human 20 UNT/ML Injectable Suspension | per tube | |
| glimepiride 2 MG Oral Tablet | nostril | |

| | | |
|---|---|---|
| nateglinide | NPO | |
| 3 ML Insulin Lispro 200 UNT/ML Pen Injector [Humalog] | po | |
| insulin human, rDNA origin 1 MG/ACTUAT Inhalant Powder | | |
| 3 ML Insulin Glargine 100 UNT/ML Pen Injector [Lantus] | DAILY | |
| Insulin Lispro 100 UNT/ML Injectable Solution [Humalog] | INJECT | |
| Bromocriptine 10 MG Oral Capsule [Parlodel] | by mouth | |
| Metformin | inhalatn | |
| Glipizide 5 MG / Metformin hydrochloride 500 MG Oral Tablet [Metaglip] | DOBHOFF | |
| Humulin N Injectable Product | SQ | |
| insulin human, isophane 100 UNT/ML Pen Injector [Humulin N] | nostril | |
| nateglinide 60 MG Oral Tablet [Starlix] | ORAL | |
| rosiglitazone 2 MG [Avandia] | per tube | |
| Pramlintide | orally | |
| Linagliptin 5 MG [Tradjenta] | sublingual | |
| Metformin hydrochloride 1000 MG Oral Tablet | gt | |
| Glyburide 5 MG | subcu. | |
| Bromocriptine | p.o. | |
| Simvastatin 20 MG / sitagliptin 50 MG Oral Tablet | Oral | |
| Metformin Extended Release Oral Tablet [Glumetza] | po | |
| Linagliptin 2.5 MG / Metformin hydrochloride 850 MG Oral Tablet [Jentadueto] | Oral | |
| Insulin, Aspart, Human 100 UNT/ML Injectable Solution [NovoLog] | SUBCUTANEOUS INFUSION | |
| Insulin, Aspart Protamine, Human 70 UNT/ML / Insulin, Aspart, Human 30 UNT/ML Injectable Suspension [NovoLog Mix] | abdominal subcutaneous | |
| Acarbose Oral Tablet [Precose] | PO | |
| Pramlintide | oral | |
| Insulin, Aspart, Human | nares | |
| Metformin hydrochloride 1000 MG / pioglitazone 30 MG [Actoplus Met] | td | |
| Insulin Lispro 100 UNT/ML [Humalog] | | 0 |
| 24 HR Metformin hydrochloride 500 MG Extended Release Oral Tablet [Glucophage] | pressure line | |
| sitagliptin 100 MG Oral Tablet | BY MOUTH | |
| Glyburide | ORAL | |
| Insulin, Aspart, Human 100 UNT/ML Injectable Solution | SUBCUTANEOUSLY | |
| Metformin hydrochloride 625 MG Oral Tablet | oral | |
| Levemir Injectable Product | SUBCUT | |
| glimepiride 2 MG | sc | |

| | | |
|---|---|---|
| Insulin Glargine 100 UNT/ML [Lantus] | INHALATION | |
| Glipizide Extended Release Oral Tablet | ORAL | |
| Glipizide Pill | PO | |
| glimepiride 1 MG Oral Tablet [Amaryl] | PER TUBE | |
| Ultralente Insulin, Human 100 UNT/ML Injectable Suspension | subcutaneously | |
| Precose Pill | | |
| Regular Insulin, Human 5 UNT/ML | topical | |
| Insulin, Regular, Pork 40 UNT/ML Injectable Solution | oral | |
| Insulin Glargine 100 UNT/ML | oral | |
| insulin human, isophane 100 UNT/ML Pen Injector [Humulin N] | sq | |
| Linagliptin | im | |
| Insulin, Zinc, Human | subQ. | |
| Glipizide Oral Tablet | NPO | |
| Regular Insulin, Human 100 UNT/ML Injectable Solution [Novolin R] | Sub Q | |
| Metformin hydrochloride 1000 MG / saxagliptin 5 MG [Kombiglyze] | swallow | |
| Metformin hydrochloride 1000 MG / sitagliptin 50 MG [Janumet] | nostril | |
| Glyburide 5 MG | drip | |
| Metformin hydrochloride 500 MG / rosiglitazone 1 MG Oral Tablet [Avandamet] | ORAL | |
| 3 ML insulin human, isophane 100 UNT/ML Prefilled Syringe | per tube. | |
| Insulin, Regular, Pork 40 UNT/ML Injectable Solution | sq | |
| 3 ML Insulin Lispro 100 UNT/ML Cartridge [Humalog] | per tube | |
| Onglyza Oral Product | Oral | |
| insulin human, isophane 75 UNT/ML | subQ | |
| Regular Insulin, Human 50 UNT/ML | sq | |
| Glyburide | intravenous | |
| insulin human, isophane 100 UNT/ML Injectable Suspension [Humulin N] | IM | |
| 3 ML insulin human, isophane 100 UNT/ML Pen Injector [Humulin N] | topical | |
| glimepiride 4 MG Oral Tablet [Amaryl] | P.O. | |
| glimepiride 4 MG / rosiglitazone 8 MG Oral Tablet [Avandaryl] | oral | |
| Insulin Lispro 100 UNT/ML Injectable Solution | | 0 |
| Regular Insulin, Human 100 UNT/ML Injectable Solution | sq | |
| insulin human, isophane 100 UNT/ML Injectable Suspension [Humulin N] | UNDER THE SKIN | |
| Levemir Injectable Product | oral | |
| Insulin Glargine | Subcutan_Pump | |

| | | |
|---|---|---|
| glimepiride | trans | |
| Insulin Lispro 50 UNT/ML / Insulin, Protamine Lispro, Human 50 UNT/ML Injectable Suspension [Humalog Mix] | p.o. | |
| Metformin hydrochloride 1000 MG [Glucophage] | subq | |
| Lispro PRC Injectable Product | by mouth | |
| Insulin, Aspart, Human 100 UNT/ML | SQ | |
| exenatide 3.08 MG/ML [Bydureon] | sq | |
| insulin human, isophane 80 UNT/ML / Regular Insulin, Human 20 UNT/ML [Human Mixtard] | sub q | |
| Glyburide 5 MG | SL | |
| Insulin Lispro | Intravenous | |
| Metformin hydrochloride 1000 MG / sitagliptin 50 MG [Janumet] | subcutaneous | |
| Acarbose 50 MG | po. | |
| Glyburide 5 MG / Metformin hydrochloride 500 MG Oral Tablet | Po | |
| Insulin Glargine 300 UNT/ML [Toujeo] | SC | |
| Precose Oral Product | Oral | |
| glimepiride 3 MG [Amaryl] | gt | |
| Insulin, Aspart, Human | P.o. | |
| Insulin, Aspart, Human 100 UNT/ML Injectable Solution [NovoLog] | INFUSION PER INSULIN PUMP | |
| 3 ML insulin human, isophane 100 UNT/ML Prefilled Syringe [Novolin N] | SUBCU | |
| Acarbose | J - tube | |
| Insulin Glargine | intravenously | |
| Regular Insulin, Human 500 UNT/ML | iv | |
| Glyburide 2.5 MG | NPO. | |
| insulin, regular, beef-pork 40 UNT/ML Inhalant Solution | | 0 |
| Humulin Injectable Product | COMBO ROUTE) | |
| Glipizide 2.5 MG | npo. | |
| Glyburide 1.25 MG | td | |
| insulin human, isophane 70 UNT/ML / Regular Insulin, Human 30 UNT/ML [Novolin] | subc | |
| Insulin Glargine | intramuscular | |
| Insulin Glargine 100 UNT/ML | per tube | |
| Insulin Lispro 50 UNT/ML | drip. | |
| insulin, isophane Injectable Suspension [Novolin N] | per tube | |
| Insulin Lispro Cartridge [Humalog] | iv | |
| Insulin, Aspart Protamine, Human | Subcutaneous | |
| 3 ML insulin detemir 100 UNT/ML Pen Injector [Levemir] | intravenous | |
| Glyburide 5 MG Oral Tablet [Diabeta] | inhalation | |

| | | |
|---|---|---|
| pioglitazone | NARES | |
| exenatide 2 MG Injection [Bydureon] | SUBCUTANEOUSLY | |
| 3 ML Insulin, Aspart, Human 100 UNT/ML Pen Injector [NovoLog] | other | |
| Regular Insulin, Human 5 UNT/ML | subq. | |
| insulin human, isophane 70 UNT/ML / Regular Insulin, Human 30 UNT/ML [Humulin] | Sublingual | |
| Gliclazide 80 MG Oral Tablet | orally | |
| insulin human, isophane 60 UNT/ML / Regular Insulin, Human 40 UNT/ML Injectable Suspension | per tube | |
| Fortamet Pill | by mouth | |
| Glyburide 5 MG | ORAL | |
| 1.5 ML insulin human, isophane 70 UNT/ML / Regular Insulin, Human 30 UNT/ML Prefilled Syringe | drip | |
| Metformin hydrochloride 850 MG / pioglitazone 15 MG Oral Tablet | subcutaneous | |
| insulin detemir Injectable Solution [Levemir] | by mouth | |
| Glipizide | per tube | |
| Bromocriptine | Im | |
| Insulin, Aspart, Human 100 UNT/ML Injectable Solution | Oral | |
| Insulin, Aspart Protamine, Human 70 UNT/ML / Insulin, Aspart, Human 30 UNT/ML Injectable Suspension [NovoLog Mix] | SUBCUTANEOUSLY | |
| insulin detemir 100 UNT/ML Injectable Solution [Levemir] | SQ | |
| Insulin Glargine | HEPLOCK | |
| insulin, isophane Injectable Suspension [Humulin N] | nostril | |
| Insulin, Aspart, Human | Insulin_Pump | |
| pioglitazone 15 MG [Actos] | subcut | |
| rosiglitazone Oral Product | | |
| Insulin Lispro 50 UNT/ML | NPO. | |
| 60 ACTUAT exenatide 0.005 MG/ACTUAT Pen Injector [Byetta] | drip | |
| sitagliptin 100 MG | NPO | |
| Insulin Lispro 200 UNT/ML Pen Injector [Humalog] | subq | |
| Metformin Extended Release Oral Tablet [Glumetza] | | 0 |
| insulin human, isophane 70 UNT/ML | NPO | |
| Simvastatin 10 MG / sitagliptin 100 MG [Juvisync] | Subcutaneous | |
| insulin pork, isophane 100 UNT/ML Injectable Suspension [NPH Iletin II] | Po | |
| Insulin Glargine | SUBCUTANEOUS. | |
| Metformin / sitagliptin Oral Tablet [Janumet] | BY MOUTH | |
| pioglitazone 45 MG Oral Tablet [Actos] | p.o | |
| Glucophage Oral Product | ORAL | |

| | | |
|---|---|---|
| pramlintide acetate 0.6 MG/ML Injectable Solution | subcutaneous | |
| pioglitazone 15 MG Oral Tablet [Actos] | ORAL | |
| Metformin hydrochloride 1000 MG [Glucophage] | subQ | |
| insulin human, isophane 80 UNT/ML / Regular Insulin, Human 20 UNT/ML [Humulin] | oral | |
| Metformin hydrochloride 500 MG [Glucophage] | bedside | |
| 24 HR Metformin hydrochloride 500 MG Extended Release Oral Tablet [Glucophage] | THIS | |
| Glyburide 6 MG | inhalation | |
| empagliflozin 5 MG | orally | |
| Metformin hydrochloride 1000 MG Oral Tablet | inhaled | |
| Glipizide 5 MG | topical | |
| Insulin, Zinc, Human | drip | |
| Glipizide 5 MG Oral Tablet [Glucotrol] | | 0 |
| Simvastatin 10 MG / sitagliptin 50 MG [Juvisync] | subcutaneous | |
| 0.5 ML dulaglutide 1.5 MG/ML Auto-Injector [Trulicity] | Sc | |
| insulin human, isophane 70 UNT/ML | subq. | |
| dapagliflozin Oral Tablet | po | |
| Metformin hydrochloride 1000 MG / sitagliptin 100 MG [Janumet] | Oral | |
| Lispro PRC Injectable Product | iv | |
| Regular Insulin, Human 10 UNT/ML | p.o. | |
| Bromocriptine | per tube | |
| insulin pork, isophane 100 UNT/ML Injectable Suspension [NPH Iletin II] | inhalation. | |
| nateglinide Oral Tablet | sublingual | |
| Regular Insulin, Human 15 UNT/ML | ng | |
| insulin human, isophane 100 UNT/ML Pen Injector [Humulin N] | subcutaneous. | |
| Regular Insulin, Human 500 UNT/ML Injectable Solution | subQ | |
| Bromocriptine 5 MG | ORAL | |
| insulin human, isophane 50 UNT/ML / Regular Insulin, Human 50 UNT/ML Injectable Suspension [Humulin] | subq | |
| insulin human, isophane 70 UNT/ML / Regular Insulin, Human 30 UNT/ML [Novolin] | by mouth. | |
| Glipizide Extended Release Oral Tablet | BY MOUTH | |
| Metformin hydrochloride 1000 MG / pioglitazone 30 MG [Actoplus Met] | subcutaneously | |
| Chlorpropamide 125 MG / Metformin 500 MG Oral Tablet | p.o. | |
| Insulin Lispro 100 UNT/ML Injectable Solution | miscellaneous | |
| 3 ML Insulin Lispro 100 UNT/ML Pen Injector [Humalog] | topical | |

| | | |
|---|---|---|
| Metformin hydrochloride 500 MG / sitagliptin 50 MG Oral Tablet [Janumet] | inhaled | |
| 3 ML insulin human, isophane 100 UNT/ML Prefilled Syringe | po | |
| insulin human, isophane 75 UNT/ML | by mouth | |
| alogliptin 12.5 MG / pioglitazone 15 MG Oral Tablet [Oseni] | oral | |
| Insulin, Aspart Protamine, Human 70 UNT/ML / Insulin, Aspart, Human 30 UNT/ML Injectable Suspension [NovoLog Mix] | npo | |
| Farxiga Oral Product | per tube | |
| Lispro PRC Injectable Product | oral | |
| insulin human, isophane 70 UNT/ML / Regular Insulin, Human 30 UNT/ML [Humulin] | NPO. | |
| insulin human, isophane 100 UNT/ML Injectable Suspension [Novolin N] | p.o. | |
| Byetta Injectable Product | SUBCUT | |
| exenatide Pen Injector [Byetta] | | |
| Metformin 1000 MG | sub Q | |
| Metformin hydrochloride 500 MG [Glucophage] | inhalation | |
| insulin human, isophane 70 UNT/ML / Regular Insulin, Human 30 UNT/ML [Humulin] | bedside | |
| Metformin Extended Release Oral Tablet [Fortamet] | subq | |
| Metformin hydrochloride 1000 MG Oral Tablet [Glucophage] | DOBHOFF TUBE | |
| Glyset Pill | | |
| Metformin 1000 MG | IM. | |
| Regular Insulin, Human | IV PUSH | |
| Insulin Lispro | BY MOUTH | |
| Glipizide 10 MG Oral Tablet | | 0 |
| Regular Insulin, Human 20 UNT/ML | Sq | |
| Insulin, Aspart, Human | As_Directed | |
| insulin human, isophane 100 UNT/ML | IV | |
| insulin pork, isophane 80 UNT/ML | sc | |
| 24 HR Metformin hydrochloride 750 MG Extended Release Oral Tablet | Po | |
| Insulin Lispro Cartridge [Humalog] | Iv | |
| Metformin 250 MG | subcutaneously | |
| Metformin hydrochloride 850 MG | nostril | |
| Bromocriptine 5 MG Oral Capsule [Parlodel] | G-TUBE | |
| pioglitazone 15 MG Oral Tablet | PER TUBE | |
| rosiglitazone 8 MG [Avandia] | ng | |
| Metformin hydrochloride 500 MG Extended Release Oral Tablet [Glumetza] | SL | |

| | |
|---|---|
| Tolazamide 100 MG Oral Tablet [Tolinase] | p.o. |
| insulin human, isophane 70 UNT/ML | Subcutaneous |
| Bromocriptine 2.5 MG Oral Tablet [Parlodel] | Oral |
| Glyburide 4.5 MG Oral Tablet | p.o. |
| Glyburide 5 MG Oral Tablet | . |
| 3 ML insulin degludec 100 UNT/ML Pen Injector [Tresiba] | by mouth |
| Trulicity Injectable Product | SUBCUTANEOUSLY |
| insulin human, isophane 40 UNT/ML Injectable Suspension | SUBCUTANEOUSLY |
| Bromocriptine 5 MG | by mouth. |
| Metformin hydrochloride 100 MG/ML | bedside |
| exenatide 0.01 MG/ACTUAT [Byetta] | subcutaneous. |
| Glyburide 5 MG / Metformin hydrochloride 500 MG Oral Tablet [Glucovance] | IM |
| 60 ACTUAT exenatide 0.005 MG/ACTUAT Pen Injector [Byetta] | Subcutaneous |
| 24 HR Metformin hydrochloride 1000 MG / sitagliptin 50 MG Extended Release Oral Tablet [Janumet] | sublingual |
| Metformin hydrochloride 1000 MG Oral Tablet | IM. |
| albiglutide 100 MG/ML | subcutaneous |
| glimepiride 2 MG [Amaryl] | by mouth. |
| insulin human, isophane 90 UNT/ML / Regular Insulin, Human 10 UNT/ML [Humulin] | sub q. |
| albiglutide 100 MG/ML [Tanzeum] | by mouth |
| Insulin Lispro Injectable Solution | sc |
| Insulin, Aspart, Human | ou |
| insulin human, isophane 90 UNT/ML / Regular Insulin, Human 10 UNT/ML [Humulin] | subq. |
| glimepiride 1 MG | SQ. |
| Insulin, Aspart, Human | intravenous |
| Linagliptin 5 MG [Tradjenta] | subcutaneously |
| Glipizide | SubQ |
| Metformin hydrochloride 1000 MG / saxagliptin 5 MG [Kombiglyze] | p.o. |
| 24 HR Metformin hydrochloride 1000 MG / pioglitazone 15 MG Extended Release Oral Tablet [Actoplus Met] | |
| exenatide | td |
| Metformin 250 MG | subcutaneous |
| Regular Insulin, Human 10 UNT/ML | subcu. |
| Regular Insulin, Human 100 UNT/ML Injectable Solution [Humulin R] | SUBCU |
| 3 ML Insulin Glargine 100 UNT/ML Pen Injector [Lantus] | GT |

| | | |
|---|---|---|
| Glyburide 5 MG | NPO. | |
| nateglinide | IV | |
| Glipizide 10 MG Oral Tablet [Glucotrol] | PO/PT | |
| Insulin Lispro Injectable Solution | INSUPUMP | |
| Chlorpropamide 125 MG Oral Tablet | orally | |
| Metformin / sitagliptin Extended Release Oral Tablet [Janumet] | p.o. | |
| nateglinide 120 MG [Starlix] | gt | |
| Insulin Lispro 50 UNT/ML | | 0 |
| insulin human, isophane 100 UNT/ML Injectable Suspension [Humulin N] | sq. | |
| Glipizide 2.5 MG | Sublingual | |
| Metformin hydrochloride 850 MG [Glucophage] | subcutaneous | |
| Metformin hydrochloride 100 MG/ML Oral Solution [Riomet] | NPO | |
| Regular Insulin, Human 6 UNT/ML | po | |
| Glipizide 5 MG [Glucotrol] | po. | |
| glimepiride | orally. | |
| Glipizide | Inhalation | |
| Acarbose | sublingual | |
| 0.5 ML albiglutide 100 MG/ML Pen Injector [Tanzeum] | subQ | |
| Insulin, Regular, Pork | | |
| insulin human, isophane 70 UNT/ML / Regular Insulin, Human 30 UNT/ML [Humulin] | SQ. | |
| Metformin Extended Release Oral Tablet [Glumetza] | BY MOUTH | |
| Linagliptin Pill | by mouth | |
| Insulin, Regular, Pork 40 UNT/ML Injectable Solution | IV. | |
| Linagliptin 5 MG Oral Tablet [Tradjenta] | DOBHOFF | |
| 3 ML Insulin, Aspart, Human 100 UNT/ML Pen Injector [NovoLog] | SC | |
| Regular Insulin, Human 100 UNT/ML Injectable Solution [Velosulin] | IV | |
| insulin human, isophane 90 UNT/ML / Regular Insulin, Human 10 UNT/ML [Humulin] | orally | |
| Metformin | sub Q | |
| 3 ML insulin human, isophane 100 UNT/ML Pen Injector [Humulin N] | oral | |
| Pramlintide Injectable Solution | sub Q | |
| Tolbutamide 500 MG Oral Tablet [Orinase] | by mouth | |
| sitagliptin 100 MG [Januvia] | drip. | |
| Glumetza Oral Product | ORAL | |
| 24 HR Metformin hydrochloride 500 MG Extended Release Oral Tablet [Glucophage] | Oral | |

| | |
|---|---|
| sitagliptin 25 MG Oral Tablet | |
| Glyburide 5 MG [Diabeta] | SUBLING |
| 0.5 ML dulaglutide 1.5 MG/ML Auto-Injector [Trulicity] | subcut |
| Insulin Lispro 100 UNT/ML Injectable Solution [Humalog] | PUM |
| insulin detemir | Subq |
| Insulin Glargine 100 UNT/ML [Lantus] | SUBQ. |
| insulin human, isophane 80 UNT/ML / Regular Insulin, Human 20 UNT/ML [Humulin] | by mouth |
| Metformin Oral Tablet | by mouth. |
| exenatide Injection | MISCELLANEOUS |
| Glipizide 2.5 MG [Glucotrol] | gt |
| Glipizide 2.5 MG | by mouth. |
| Insulin, Zinc, Human 100 UNT/ML Injectable Suspension [Novolin L] | subcutaneously. |
| Metformin hydrochloride 500 MG [Glucophage] | Iv |
| Simvastatin 20 MG / sitagliptin 50 MG Oral Tablet | BY MOUTH |
| 1.5 ML insulin human, isophane 70 UNT/ML / Regular Insulin, Human 30 UNT/ML Prefilled Syringe | subq. |
| Glipizide 10 MG Oral Tablet [Glucotrol] | DOBHOFF TUBE |
| Glyburide 5 MG / Metformin hydrochloride 500 MG [Glucovance] | g - tube |
| Linagliptin 2.5 MG / Metformin hydrochloride 500 MG Oral Tablet [Jentadueto] | Oral |
| Metformin hydrochloride 850 MG [Glucophage] | gt |
| exenatide 0.005 MG/ACTUAT [Byetta] | SUBQ. |
| insulin human, isophane 70 UNT/ML / Regular Insulin, Human 30 UNT/ML [Novolin] | intramuscular |
| Glipizide 5 MG Extended Release Oral Tablet [Glucotrol] | p.o. |
| Glyburide 5 MG Oral Tablet | nostril |
| Chlorpropamide Oral Tablet | per tube |
| insulin human, isophane 100 UNT/ML Injectable Suspension [Humulin N] | IV |
| Metformin 500 MG | iv |
| Insulin, Aspart Protamine, Human 70 UNT/ML / Insulin, Aspart, Human 30 UNT/ML Injectable Suspension [NovoLog Mix] | SUB Q |
| Insulin, Aspart Protamine, Human | by mouth |
| Insulin Lispro 50 UNT/ML / Insulin, Protamine Lispro, Human 50 UNT/ML Injectable Suspension [Humalog Mix] | Subq |
| nateglinide | MISC |
| Glyburide Oral Tablet [Micronase] | oral |

| | |
|---|---|
| 3 ML insulin degludec 100 UNT/ML / liraglutide 3.6 MG/ML Pen Injector [Xultophy] | SUBCUTANEOUS |
| Acetohexamide 250 MG Oral Tablet [Dymelor] | p.o. |
| Invokana Pill | PO |
| 3 ML Insulin Lispro 200 UNT/ML Pen Injector [Humalog] | subQ |
| Metformin | IM injection. |
| Insulin Lispro 100 UNT/ML Injectable Solution [Lispro PRC] | oral |
| Metformin hydrochloride 850 MG | ou |
| Insulin, Aspart, Human 100 UNT/ML Injectable Solution [NovoLog] | SUBCUTANEOUSLY PER INSULIN PUMP |
| Metformin | per rectum. |
| sitagliptin 100 MG [Januvia] | sublingual |
| Glyburide 2.5 MG [Diabeta] | PER TUBE |
| Linagliptin 5 MG [Tradjenta] | SQ |
| Insulin, Aspart, Human 100 UNT/ML Cartridge [NovoLog] | OU |
| Metformin hydrochloride 100 MG/ML | Subcutaneous |
| insulin human, isophane 70 UNT/ML / Regular Insulin, Human 30 UNT/ML [Human Mixtard] | subcu |
| Simvastatin 10 MG / sitagliptin 100 MG [Juvisync] | subcu |
| Insulin, Aspart, Human | SPINAL |
| Insulin Glargine | SUBCUTANEOUSLY |
| 3 ML Regular Insulin, Human 100 UNT/ML Prefilled Syringe [Novolin R] | orally |
| insulin human, isophane 90 UNT/ML | sub q |
| Insulin, Aspart, Human 100 UNT/ML Injectable Solution [NovoLog] | TO 6 TIMES DAILY WITH MEALS AND |
| Novolin R Injectable Product | 0 |
| Glyburide Oral Tablet | P.O. |
| Acarbose 100 MG [Precose] | ng |
| Victoza Injectable Product | SUBCUTANEOUSLY |
| glimepiride | PO. |
| Metformin hydrochloride 1000 MG / saxagliptin 2.5 MG [Kombiglyze] | Bedside |
| canagliflozin 150 MG / Metformin hydrochloride 1000 MG Oral Tablet [Invokamet] | nostril |
| Insulin, Aspart, Human | subQ. |
| pioglitazone 30 MG [Actos] | swallow |
| Glyburide 6 MG | iv |
| 3 ML insulin human, isophane 100 UNT/ML Prefilled Syringe | Bedside |
| 3 ML insulin human, isophane 100 UNT/ML Prefilled Syringe | p.o. |

| | |
|---|---|
| liraglutide 6 MG/ML [Victoza] | td |
| Glipizide 10 MG Oral Tablet | subcutaneous |
| insulin human, isophane 90 UNT/ML / Regular Insulin, Human 10 UNT/ML [Humulin] | by mouth |
| Metformin | Subcutaneous |
| 24 HR Metformin hydrochloride 1000 MG / pioglitazone 30 MG Extended Release Oral Tablet [Actoplus Met] | |
| Metformin hydrochloride 1000 MG / saxagliptin 5 MG [Kombiglyze] | iv |
| glimepiride | Inhalation |
| 24 HR empagliflozin 25 MG / Metformin hydrochloride 1000 MG Extended Release Oral Tablet [Synjardy] | ORAL |
| glimepiride 1 MG [Amaryl] | IV |
| alogliptin 25 MG / pioglitazone 45 MG Oral Tablet [Oseni] | |
| Metformin hydrochloride 500 MG Extended Release Oral Tablet | sublingual |
| insulin, isophane Injectable Suspension [Novolin N] | SUBCUTANEOUSLY |
| insulin human, isophane 70 UNT/ML / Regular Insulin, Human 30 UNT/ML [Humulin] | OU |
| Metformin hydrochloride 500 MG Oral Tablet | Trans |
| Glyburide 1.5 MG | by mouth. |
| Regular Insulin, Human 100 UNT/ML Injectable Solution [Humulin R] | p.o. |
| Metformin | G tube. |
| glimepiride 8 MG | subq |
| Metformin hydrochloride 1000 MG / rosiglitazone 4 MG [Avandamet] | inhalation |
| Metformin hydrochloride 1000 MG / sitagliptin 100 MG [Janumet] | IV |
| insulin human, isophane 70 UNT/ML / Regular Insulin, Human 30 UNT/ML Injectable Suspension | SUBCUTANEOUSLY |
| Novolin Injectable Product | 0 |
| Regular Insulin, Human 10 UNT/ML | po |
| exenatide 2 MG [Bydureon] | 0 |
| Metformin 1000 MG | subQ |
| Osmotic 24 HR Metformin hydrochloride 1000 MG Extended Release Oral Tablet [Fortamet] | ORAL WITH FOOD |
| Regular Insulin, Human 100 UNT/ML Injectable Solution [Humulin R] | SUBCUTANEOUSLY |
| Glipizide Oral Tablet [Glucotrol] | SL |
| glimepiride 1 MG | subcutaneous |

| | | |
|---|---|---|
| Apidra Injectable Product | subcut. | |
| Insulin Glargine 100 UNT/ML | SUBCUT. | |
| Glipizide 2.5 MG | | 0 |
| Insulin, Zinc, Pork 100 UNT/ML Injectable Suspension | subcu | |
| Metformin hydrochloride 500 MG Extended Release Oral Tablet [Fortamet] | ORAL | |
| Glyburide 2.5 MG | BY MOUTH | |
| glimepiride 2 MG | po. | |
| Insulin Lispro 50 UNT/ML / Insulin, Protamine Lispro, Human 50 UNT/ML Injectable Suspension [Humalog Mix] | topical | |
| Insulin Glargine Injectable Product | | |
| Insulin, Glulisine, Human 100 UNT/ML Injectable Solution [Apidra] | PER PUMP CONT SUBCUTAN INFUSI | |
| 3 ML Insulin Glargine 100 UNT/ML Pen Injector [Lantus] | J - tube | |
| Insulin, Aspart, Human 100 UNT/ML Injectable Solution [NovoLog] | SUBCUTANEOUSLY | |
| pioglitazone 30 MG Oral Tablet | P.o. | |
| Linagliptin 2.5 MG | | |
| Acarbose | swallow. | |
| Glyburide 5 MG Oral Tablet | NG | |
| Glipizide 2.5 MG / Metformin hydrochloride 250 MG Oral Tablet [Metaglip] | | 0 |
| 3 ML Insulin, Aspart Protamine, Human 70 UNT/ML / Insulin, Aspart, Human 30 UNT/ML Pen Injector [NovoLog Mix] | miscellaneous | |
| Regular Insulin, Human 10 UNT/ML | NPO | |
| Insulin, Zinc, Pork 100 UNT/ML Injectable Suspension | SUBCUTANEOUS | |
| 3 ML Insulin Lispro 100 UNT/ML Pen Injector [Humalog] | SQ | |
| Metformin 500 MG | IV. | |
| 3 ML Insulin Glargine 100 UNT/ML / Lixisenatide 0.033 MG/ML Pen Injector [Soliqua] | injection | |
| insulin pork, isophane 100 UNT/ML Injectable Suspension | SUBCUT | |
| pioglitazone 15 MG Oral Tablet [Actos] | BY MOUTH | |
| Metformin hydrochloride 1000 MG Oral Tablet [Glucophage] | ORAL | |
| Glyburide | Bedside | |
| Bromocriptine 5 MG | Im | |
| Humalog Injectable Product | gt | |
| Tolazamide 100 MG Oral Tablet | PO | |
| Insulin, Aspart, Human | Sub_Q_Infusion | |
| insulin degludec 200 UNT/ML [Tresiba] | by mouth | |

| | |
|---|---|
| Metformin hydrochloride 500 MG / sitagliptin 50 MG Oral Tablet [Janumet] | IV |
| glimepiride 2 MG Oral Tablet [Amaryl] | mouth/throat |
| glimepiride 4 MG Oral Tablet [Amaryl] | Im |
| . | . |
| . | . |
| . | . |
| . | . |
| . | . |
| . | . |
| . | . |
| . | . |
| . | . |
| . | . |
| . | . |
| . | . |
| . | . |
| . | . |
| . | . |
| . | . |
| . | . |
| . | . |
| . | . |
| . | . |
| . | . |
| . | . |
| . | . |
| . | . |
| . | . |
| . | . |
| . | . |
| . | . |
| . | . |
| . | . |
| . | . |
| . | . |
| . | . |
| . | . |
| . | . |
