## Supplemental Table 4 for "Development of Portable Electronic Health Record Based Algorithms to Identify Individuals with Diabetic Retinopathy"

**Supplemental Table 4. Reviewer Agreement on Electronic Health Record-Based Diabetic Retinopathy Algorithm**

|  | **All Charts** | | | | **Charts with Vanderbilt Eye Institute Notes** | | | |
| --- | --- | --- | --- | --- | --- | --- | --- | --- |
| **Disease** | **Agree Disease** | **Disagree Disease** | **Agree No Disease** | **Agreement** | **Agree Disease** | **Disagree Disease** | **Agree No Disease** | **Agreement** |
| **DM** | 41 | 2 | 2 | 96% | 21 | 0 | 0 | 100% |
| **DR** | 29 | 4 | 12 | 91% | 16 | 1 | 4 | 95% |
| **PDR** | 9 | 1 | 35 | 98% | 6 | 0 | 15 | 100% |
