## Supplemental Table 5 for "Development of Portable Electronic Health Record Based Algorithms to Identify Individuals with Diabetic Retinopathy"

**Supplemental Table 5: Manual and Algorithm Designations in VANEOHS and MGB**

|  | **Chart review Disease** | | | **Chart review - No Disease** | | | **Chart review - Undetermined** | | |
| --- | --- | --- | --- | --- | --- | --- | --- | --- | --- |
|  | **VANEOHS** | | **MGB** | **VANEOHS** | | **MGB** | **VANEOHS** | | **MGB** |
|  | **DR** | **PDR** | **DR** | **DR** | **PDR** | **DR** | **DR** | **PDR** | **DR** |
| **Disease** | 47 | 27 | 42 | 3 | 6 | 6 | 0 | 0 | 2 |
| **No Disease** | 7 | 1 | 1 | 43 | 87 | 38 | 0 | 0 | 11 |
