## Supplemental Table 7 for "Development of Portable Electronic Health Record Based Algorithms to Identify Individuals with Diabetic Retinopathy"

Supplemental Table 7. European-Ancestry Phenome-Wide Association Study Meta-analysis Results for Diabetic Retinopathy Algorithm Designation adjusted for age, sex, and duration of diabetes

| PheCode | Phenotype | Phenotype Goup | Predictor | OR | LCI | UCI | p | n_total | k_studies |
|---|---|---|---|---|---|---|---|---|---|
| 250.7 | Diabetic retinopathy | endocrine/metabolic | DR | 414.40 | 402.23 | 426.93 | <1E-300 | 128777 | 2 |
| 250.23 | Type 2 diabetes with ophthalmic manifestations | endocrine/metabolic | DR | 75.89 | 74.37 | 77.44 | <1E-300 | 128777 | 2 |
| 250.13 | Type 1 diabetes with ophthalmic manifestations | endocrine/metabolic | DR | 19.32 | 18.41 | 20.28 | <1E-300 | 128777 | 2 |
| 362.9 | Retinal edema | sense organs | DR | 7.49 | 7.26 | 7.73 | <1E-300 | 128777 | 2 |
| 362.8 | Retinal hemorrhage/ischemia | sense organs | DR | 4.37 | 4.25 | 4.50 | <1E-300 | 128777 | 2 |
| 580.31 | Nephritis and nephropathy in diseases classified elsewhere | genitourinary | DR | 4.32 | 4.18 | 4.46 | <1E-300 | 128777 | 2 |
| 585.32 | End stage renal disease | genitourinary | DR | 4.06 | 3.93 | 4.19 | <1E-300 | 128777 | 2 |
| 362.23 | Cystoid macular degeneration of retina | sense organs | DR | 3.77 | 3.66 | 3.88 | <1E-300 | 128777 | 2 |
| 585.34 | Chronic Kidney Disease, Stage IV | genitourinary | DR | 3.70 | 3.61 | 3.79 | <1E-300 | 128777 | 2 |
| 250.3 | Insulin pump user | endocrine/metabolic | DR | 3.63 | 3.57 | 3.70 | <1E-300 | 128777 | 2 |
| 250.14 | Type 1 diabetes with neurological manifestations | endocrine/metabolic | DR | 3.59 | 3.49 | 3.68 | <1E-300 | 128777 | 2 |
| 285.21 | Anemia in chronic kidney disease | hematopoietic | DR | 3.56 | 3.48 | 3.65 | <1E-300 | 128777 | 2 |
| 250.1 | Type 1 diabetes | endocrine/metabolic | DR | 3.43 | 3.38 | 3.48 | <1E-300 | 128777 | 2 |
| 588.2 | Secondary hyperparathyroidism (of renal origin) | genitourinary | DR | 3.40 | 3.30 | 3.52 | <1E-300 | 128777 | 2 |
| 580.3 | Nephritis and nephropathy without mention of glomerulonephritis | genitourinary | DR | 3.32 | 3.22 | 3.42 | <1E-300 | 111475 | 1 |
| 362 | Other retinal disorders | sense organs | DR | 3.29 | 3.24 | 3.34 | <1E-300 | 128777 | 2 |
| 366 | Cataract | sense organs | DR | 3.25 | 3.18 | 3.32 | <1E-300 | 128777 | 2 |
| 588 | Disorders resulting from impaired renal function | genitourinary | DR | 3.04 | 2.95 | 3.13 | <1E-300 | 128777 | 2 |
| 250.22 | Type 2 diabetes with renal manifestations | endocrine/metabolic | DR | 3.04 | 2.99 | 3.08 | <1E-300 | 128777 | 2 |
| 362.4 | Retinal vascular changes and abnomalities | sense organs | DR | 2.99 | 2.92 | 3.06 | <1E-300 | 128777 | 2 |
| 580 | | | DR | 2.95 | 2.87 | 3.03 | <1E-300 | 111475 | 1 |
| 250.6 | Polyneuropathy in diabetes | endocrine/metabolic | DR | 2.88 | 2.84 | 2.93 | <1E-300 | 128777 | 2 |
| 710.19 | Unspecified osteomyelitis | musculoskeletal | DR | 2.87 | 2.79 | 2.95 | <1E-300 | 128777 | 2 |
| 362.2 | | | DR | 2.72 | 2.68 | 2.76 | <1E-300 | 111475 | 1 |
| 250.24 | Type 2 diabetes with neurological manifestations | endocrine/metabolic | DR | 2.71 | 2.68 | 2.75 | <1E-300 | 128777 | 2 |
| 707.2 | Chronic ulcer of leg or foot | dermatologic | DR | 2.70 | 2.66 | 2.75 | <1E-300 | 128777 | 2 |
| 401.22 | Hypertensive chronic kidney disease | circulatory system | DR | 2.70 | 2.66 | 2.74 | <1E-300 | 128777 | 2 |
| 585.3 | Chronic renal failure [CKD] | genitourinary | DR | 2.67 | 2.63 | 2.70 | <1E-300 | 128777 | 2 |
| 362.3 | Other nondiabetic retinopathy | sense organs | DR | 2.64 | 2.59 | 2.69 | <1E-300 | 128777 | 2 |
| 251.1 | Hypoglycemia | endocrine/metabolic | DR | 2.61 | 2.55 | 2.67 | <1E-300 | 128777 | 2 |
| 585.33 | Chronic Kidney Disease, Stage III | genitourinary | DR | 2.58 | 2.54 | 2.62 | <1E-300 | 128777 | 2 |
| 285.2 | Anemia of chronic disease | hematopoietic | DR | 2.56 | 2.51 | 2.62 | <1E-300 | 128777 | 2 |
| 707.1 | Decubitus ulcer | dermatologic | DR | 2.48 | 2.42 | 2.53 | <1E-300 | 128777 | 2 |
| 250.25 | Diabetes type 2 with peripheral circulatory disorders | endocrine/metabolic | DR | 2.46 | 2.42 | 2.51 | <1E-300 | 128777 | 2 |
| 249 | Secondary diabetes mellitus | endocrine/metabolic | DR | 2.42 | 2.38 | 2.47 | <1E-300 | 128777 | 2 |
| 362.26 | Macular puckering of retina | sense organs | DR | 2.42 | 2.37 | 2.46 | <1E-300 | 128777 | 2 |
| 276.13 | Hyperpotassemia | endocrine/metabolic | DR | 2.40 | 2.36 | 2.44 | <1E-300 | 128777 | 2 |
| 707 | Chronic ulcer of skin | dermatologic | DR | 2.37 | 2.33 | 2.41 | <1E-300 | 111475 | 1 |
| 585 | | | DR | 2.34 | 2.30 | 2.37 | <1E-300 | 111475 | 1 |
| 401.2 | Hypertensive heart and/or renal disease | circulatory system | DR | 2.33 | 2.30 | 2.37 | <1E-300 | 128777 | 2 |
| 269 | Proteinuria | endocrine/metabolic | DR | 2.30 | 2.26 | 2.35 | <1E-300 | 128777 | 2 |
| 428.4 | Heart failure with preserved EF [Diastolic heart failure] | circulatory system | DR | 2.12 | 2.08 | 2.16 | <1E-300 | 128777 | 2 |
| 379 | Other disorders of eye | sense organs | DR | 2.09 | 2.06 | 2.11 | <1E-300 | 128777 | 2 |
| 428.1 | Congestive heart failure (CHF) NOS | circulatory system | DR | 2.07 | 2.05 | 2.10 | <1E-300 | 128777 | 2 |
| 428 | | | DR | 2.06 | 2.03 | 2.09 | <1E-300 | 128777 | 2 |
| 443.9 | Peripheral vascular disease, unspecified | circulatory system | DR | 2.05 | 2.02 | 2.08 | <1E-300 | 128777 | 2 |
| 428.3 | Heart failure with reduced EF [Systolic or combined heart failure] | circulatory system | DR | 2.03 | 2.00 | 2.06 | <1E-300 | 128777 | 2 |
| 411.4 | Coronary atherosclerosis | circulatory system | DR | 2.02 | 2.00 | 2.05 | <1E-300 | 128777 | 2 |
| 443 | | | DR | 2.01 | 1.98 | 2.04 | <1E-300 | 111475 | 1 |
| 285 | Other anemias | hematopoietic | DR | 2.01 | 1.98 | 2.03 | <1E-300 | 128777 | 2 |
| 411 | | | DR | 2.00 | 1.97 | 2.03 | <1E-300 | 111475 | 1 |
| 585.1 | Acute renal failure | genitourinary | DR | 2.00 | 1.97 | 2.03 | <1E-300 | 128777 | 2 |
| 110.11 | Dermatophytosis of nail | infectious diseases | DR | 2.00 | 1.97 | 2.02 | <1E-300 | 128777 | 2 |
| 703 | Diseases of nail, NOS | dermatologic | DR | 1.91 | 1.88 | 1.94 | <1E-300 | 128777 | 2 |
| 440 | Atherosclerosis | circulatory system | DR | 1.89 | 1.86 | 1.92 | <1E-300 | 128777 | 2 |
| 433.1 | Occlusion and stenosis of precerebral arteries | circulatory system | DR | 1.88 | 1.85 | 1.91 | <1E-300 | 128777 | 2 |
| 433 | Cerebrovascular disease | circulatory system | DR | 1.87 | 1.84 | 1.89 | <1E-300 | 128777 | 2 |
| 366.2 | Senile cataract | sense organs | DR | 1.86 | 1.83 | 1.89 | <1E-300 | 128777 | 2 |
| 411.8 | Other chronic ischemic heart disease, unspecified | circulatory system | DR | 1.86 | 1.83 | 1.88 | <1E-300 | 128777 | 2 |
| 110.1 | Dermatophytosis | infectious diseases | DR | 1.81 | 1.79 | 1.84 | <1E-300 | 128777 | 2 |
| 110 | | | DR | 1.80 | 1.77 | 1.82 | <1E-300 | 111475 | 1 |
| 379.2 | Disorders of vitreous body | sense organs | DR | 1.78 | 1.75 | 1.80 | <1E-300 | 128777 | 2 |
| 411.2 | Myocardial infarction | circulatory system | DR | 1.74 | 1.71 | 1.76 | <1E-300 | 128777 | 2 |
| 276 | | | DR | 1.68 | 1.66 | 1.71 | <1E-300 | 111475 | 1 |
| 782.3 | Edema | symptoms | DR | 1.68 | 1.66 | 1.71 | <1E-300 | 128777 | 2 |
| 276.1 | Electrolyte imbalance | endocrine/metabolic | DR | 1.67 | 1.65 | 1.69 | <1E-300 | 128777 | 2 |
| 250.4 | Abnormal glucose | endocrine/metabolic | DR | 0.56 | 0.55 | 0.57 | <1E-300 | 111475 | 1 |
| 250.41 | Impaired fasting glucose | endocrine/metabolic | DR | 0.29 | 0.28 | 0.30 | <1E-300 | 128777 | 2 |
| 427.2 | Atrial fibrillation and flutter | circulatory system | DR | 1.74 | 1.71 | 1.76 | <1E-300 | 111475 | 1 |
| 710 | Osteomyelitis, periostitis, and other infections involving bone | musculoskeletal | DR | 2.49 | 2.43 | 2.56 | 2.64E-295 | 128777 | 2 |
| 710.1 | Osteomyelitis | musculoskeletal | DR | 2.56 | 2.49 | 2.63 | 7.77E-295 | 111475 | 1 |
| 350.2 | Abnormality of gait | neurological | DR | 1.63 | 1.60 | 1.65 | 4.92E-291 | 128777 | 2 |
| 440.2 | Atherosclerosis of the extremities | circulatory system | DR | 2.00 | 1.96 | 2.04 | 9.80E-284 | 128777 | 2 |
| 427.21 | Atrial fibrillation | circulatory system | DR | 1.67 | 1.65 | 1.70 | 1.40E-283 | 128777 | 2 |
| 585.2 | Renal failure NOS | genitourinary | DR | 2.49 | 2.43 | 2.55 | 2.09E-273 | 128777 | 2 |
| 710.11 | Acute osteomyelitis | musculoskeletal | DR | 3.01 | 2.92 | 3.11 | 2.12E-270 | 128777 | 2 |
| 700 | Corns and callosities | dermatologic | DR | 1.72 | 1.69 | 1.74 | 3.48E-267 | 128777 | 2 |
| 367 | Disorders of refraction and accommodation; blindness and low vision | sense organs | DR | 2.42 | 2.36 | 2.48 | 1.32E-264 | 128777 | 2 |
| 585.31 | Renal dialysis | genitourinary | DR | 4.31 | 4.13 | 4.49 | 3.16E-263 | 128777 | 2 |
| 710.12 | Chronic osteomyelitis | musculoskeletal | DR | 2.97 | 2.88 | 3.06 | 1.03E-261 | 128777 | 2 |
| 250.12 | Type 1 diabetes with renal manifestations | endocrine/metabolic | DR | 6.07 | 5.76 | 6.39 | 2.55E-260 | 128777 | 2 |
| 280.1 | Iron deficiency anemias, unspecified or not due to blood loss | hematopoietic | DR | 1.73 | 1.70 | 1.76 | 1.49E-258 | 128777 | 2 |
| 379.3 | Aphakia and other disorders of lens | sense organs | DR | 2.47 | 2.41 | 2.54 | 1.79E-258 | 128777 | 2 |
| 426 | Cardiac conduction disorders | circulatory system | DR | 1.58 | 1.56 | 1.60 | 3.77E-255 | 128777 | 2 |
| 1089 | Acquired absence of limb | | DR | 3.11 | 3.01 | 3.22 | 5.20E-254 | 128777 | 2 |
| 276.6 | Fluid overload | endocrine/metabolic | DR | 2.32 | 2.26 | 2.38 | 8.35E-254 | 128777 | 2 |
| 440.21 | Atherosclerosis of native arteries of the extremities with ulceration or gangrene | circulatory system | DR | 3.51 | 3.38 | 3.64 | 5.41E-253 | 128777 | 2 |
| 280 | | | DR | 1.73 | 1.70 | 1.76 | 3.58E-252 | 111475 | 1 |
| 367.9 | Blindness and low vision | sense organs | DR | 1.88 | 1.84 | 1.91 | 1.20E-246 | 128777 | 2 |
| 401.1 | Essential hypertension | circulatory system | DR | 2.60 | 2.52 | 2.67 | 6.36E-242 | 128777 | 2 |
| 772.3 | Muscle weakness | symptoms | DR | 1.64 | 1.61 | 1.66 | 4.87E-239 | 128777 | 2 |
| 350 | | | DR | 1.55 | 1.53 | 1.57 | 2.34E-234 | 111475 | 1 |
| 401 | Hypertension | circulatory system | DR | 3.86 | 3.70 | 4.02 | 7.32E-233 | 128777 | 2 |

| | | | | | | | | | |
|---|---|---|---|---|---|---|---|---|---|
| 586 | Other disorders of the kidney and ureters | genitourinary | DR | 1.65 | 1.62 | 1.67 | 1.87E-231 | 128777 | 2 |
| 395 | Heart valve disorders | circulatory system | DR | 1.72 | 1.69 | 1.75 | 1.45E-224 | 128777 | 2 |
| 433.3 | Cerebral ischemia | circulatory system | DR | 1.72 | 1.69 | 1.74 | 3.49E-224 | 128777 | 2 |
| 362.21 | Macular degeneration, dry | sense organs | DR | 1.87 | 1.84 | 1.91 | 7.41E-221 | 128777 | 2 |
| 433.31 | Transient cerebral ischemia | circulatory system | DR | 1.70 | 1.67 | 1.73 | 4.64E-220 | 128777 | 2 |
| 426.2 | Atrioventricular [AV] block | circulatory system | DR | 1.98 | 1.94 | 2.03 | 5.54E-219 | 128777 | 2 |
| 681.5 | Cellulitis and abscess of leg, except foot | dermatologic | DR | 1.70 | 1.67 | 1.73 | 8.11E-219 | 128777 | 2 |
| 681.6 | Cellulitis and abscess of foot, toe | dermatologic | DR | 2.53 | 2.45 | 2.60 | 1.01E-217 | 128777 | 2 |
| 456 | Chronic venous insufficiency [CVI] | circulatory system | DR | 1.70 | 1.67 | 1.73 | 2.73E-217 | 128777 | 2 |
| 735.2 | Acquired toe deformities | musculoskeletal | DR | 1.64 | 1.62 | 1.67 | 4.79E-217 | 128777 | 2 |
| 871.3 | Open wound of foot except toe(s) alone | injuries & poisonings | DR | 2.28 | 2.22 | 2.34 | 8.52E-215 | 128777 | 2 |
| 277 | Other disorders of metabolism | endocrine/metabolic | DR | 2.16 | 2.11 | 2.22 | 1.09E-211 | 128777 | 2 |
| 290 | Delirium dementia and amnestic and other cognitive disorders | mental disorders | DR | 1.72 | 1.69 | 1.75 | 2.71E-211 | 128777 | 2 |
| 367.4 | Presbyopia | sense organs | DR | 1.71 | 1.68 | 1.74 | 6.06E-210 | 128777 | 2 |
| 362.22 | Macular degeneration, wet | sense organs | DR | 2.91 | 2.81 | 3.02 | 1.05E-206 | 128777 | 2 |
| 791 | Gangrene | symptoms | DR | 3.77 | 3.61 | 3.94 | 9.10E-203 | 128777 | 2 |
| 681.1 | Cellulitis and abscess of fingers/toes | dermatologic | DR | 1.75 | 1.72 | 1.79 | 2.32E-200 | 128777 | 2 |
| 433.21 | Cerebral artery occlusion, with cerebral infarction | circulatory system | DR | 1.79 | 1.76 | 1.83 | 3.64E-200 | 128777 | 2 |
| 458 | Hypotension | circulatory system | DR | 1.55 | 1.53 | 1.57 | 1.86E-198 | 128777 | 2 |
| 371 | Inflammation of the eye | sense organs | DR | 1.52 | 1.50 | 1.55 | 7.82E-197 | 128777 | 2 |
| 427 | | | DR | 1.48 | 1.46 | 1.50 | 5.02E-191 | 128777 | 2 |
| 707.3 | Chronic ulcer of unspecified site | dermatologic | DR | 2.16 | 2.11 | 2.22 | 1.95E-190 | 128777 | 2 |
| 401.21 | Hypertensive heart disease | circulatory system | DR | 1.58 | 1.55 | 1.60 | 6.29E-190 | 128777 | 2 |
| 41 | Bacterial infection NOS | infectious diseases | DR | 1.58 | 1.56 | 1.61 | 5.01E-189 | 128777 | 2 |
| 317 | Alcohol-related disorders | mental disorders | DR | 0.61 | 0.60 | 0.62 | 2.42E-187 | 128777 | 2 |
| 735.21 | Hammer toe (acquired) | musculoskeletal | DR | 1.71 | 1.68 | 1.74 | 6.31E-186 | 128777 | 2 |
| 395.2 | Nonrheumatic aortic valve disorders | circulatory system | DR | 1.79 | 1.75 | 1.82 | 6.90E-185 | 128777 | 2 |
| 276.5 | Hypovolemia | endocrine/metabolic | DR | 1.55 | 1.53 | 1.58 | 2.23E-184 | 128777 | 2 |
| 371.3 | Inflammation of eyelids | sense organs | DR | 1.52 | 1.50 | 1.55 | 1.26E-183 | 128777 | 2 |
| 871 | Open wounds of extremities | injuries & poisonings | DR | 1.57 | 1.54 | 1.59 | 1.63E-183 | 128777 | 2 |
| 681 | Superficial cellulitis and abscess | dermatologic | DR | 1.46 | 1.44 | 1.47 | 4.93E-183 | 128777 | 2 |
| 433.2 | Occlusion of cerebral arteries | circulatory system | DR | 1.78 | 1.74 | 1.81 | 3.13E-182 | 128777 | 2 |
| 507 | Pleurisy; pleural effusion | respiratory | DR | 1.70 | 1.66 | 1.73 | 1.27E-181 | 128777 | 2 |
| 600 | Hyperplasia of prostate | genitourinary | DR | 1.47 | 1.45 | 1.49 | 2.75E-180 | 128777 | 2 |
| 362.29 | Macular degeneration (senile) of retina NOS | sense organs | DR | 2.11 | 2.05 | 2.16 | 3.13E-180 | 128777 | 2 |
| 271.3 | Intestinal disaccharidase deficiencies and disaccharide malabsorption | endocrine/metabolic | DR | 0.34 | 0.33 | 0.35 | 5.93E-180 | 128777 | 2 |
| 585.4 | Chronic kidney disease, Stage I or II | genitourinary | DR | 1.88 | 1.84 | 1.92 | 9.94E-180 | 128777 | 2 |
| 276.41 | Acidosis | endocrine/metabolic | DR | 1.78 | 1.74 | 1.81 | 2.50E-179 | 128777 | 2 |
| 316 | Substance addiction and disorders | mental disorders | DR | 0.59 | 0.58 | 0.60 | 5.59E-179 | 128777 | 2 |
| 735 | Acquired foot deformities | musculoskeletal | DR | 1.48 | 1.46 | 1.50 | 3.26E-177 | 128777 | 2 |
| 911 | Blister | injuries & poisonings | DR | 2.34 | 2.27 | 2.41 | 7.28E-171 | 128777 | 2 |
| 375.1 | Dry eyes | sense organs | DR | 1.42 | 1.41 | 1.44 | 8.78E-169 | 128777 | 2 |
| 458.9 | Hypotension NOS | circulatory system | DR | 1.54 | 1.51 | 1.56 | 6.48E-168 | 128777 | 2 |
| 588.1 | Renal osteodystrophy | genitourinary | DR | 3.41 | 3.26 | 3.57 | 3.43E-167 | 128777 | 2 |
| 459 | Other disorders of circulatory system | circulatory system | DR | 1.64 | 1.61 | 1.67 | 2.89E-166 | 128777 | 2 |
| 416 | Cardiomegaly | circulatory system | DR | 1.66 | 1.63 | 1.69 | 6.21E-164 | 128777 | 2 |
| 374 | Other disorders of eyelids | sense organs | DR | 1.57 | 1.54 | 1.59 | 2.23E-162 | 128777 | 2 |
| 426.9 | Cardiac pacemaker/device in situ | circulatory system | DR | 1.77 | 1.73 | 1.81 | 1.10E-161 | 128777 | 2 |
| 290.1 | Dementias | mental disorders | DR | 1.85 | 1.81 | 1.89 | 3.14E-161 | 128777 | 2 |
| 317.1 | Alcoholism | mental disorders | DR | 0.60 | 0.59 | 0.62 | 9.59E-161 | 128777 | 2 |
| 365 | Glaucoma | sense organs | DR | 1.43 | 1.42 | 1.45 | 8.82E-160 | 128777 | 2 |
| 367.2 | Astigmatism | sense organs | DR | 1.42 | 1.40 | 1.44 | 4.39E-159 | 128777 | 2 |
| 427.3 | Other specified cardiac dysrhythmias | circulatory system | DR | 1.48 | 1.46 | 1.50 | 2.11E-155 | 128777 | 2 |
| 702.1 | Actinic keratosis | dermatologic | DR | 1.44 | 1.42 | 1.46 | 4.67E-154 | 128777 | 2 |
| 271 | | | DR | 0.39 | 0.37 | 0.40 | 5.88E-154 | 111475 | 1 |
| 411.3 | Angina pectoris | circulatory system | DR | 1.46 | 1.44 | 1.48 | 8.15E-154 | 128777 | 2 |
| 38 | Septicemia | infectious diseases | DR | 1.58 | 1.55 | 1.61 | 4.12E-152 | 128777 | 2 |
| 367.8 | Hypermetropia | sense organs | DR | 1.42 | 1.40 | 1.44 | 1.73E-151 | 128777 | 2 |
| 433.8 | Late effects of cerebrovascular disease | circulatory system | DR | 1.77 | 1.73 | 1.81 | 4.08E-151 | 128777 | 2 |
| 798 | Malaise and fatigue | symptoms | DR | 1.39 | 1.37 | 1.40 | 2.85E-149 | 128777 | 2 |
| 459.9 | Circulatory disease NEC | circulatory system | DR | 1.64 | 1.61 | 1.68 | 3.83E-149 | 128777 | 2 |
| 426.21 | First degree AV block | circulatory system | DR | 2.02 | 1.97 | 2.08 | 1.79E-148 | 128777 | 2 |
| 172 | | | DR | 1.48 | 1.46 | 1.50 | 3.50E-148 | 111475 | 1 |
| 871.4 | Open wound of toe(s) | injuries & poisonings | DR | 2.31 | 2.23 | 2.38 | 7.79E-147 | 128777 | 2 |
| 300 | Anxiety disorders | mental disorders | DR | 0.71 | 0.70 | 0.72 | 6.93E-146 | 128777 | 2 |
| 426.91 | Cardiac pacemaker in situ | circulatory system | DR | 1.79 | 1.75 | 1.84 | 2.05E-143 | 128777 | 2 |
| 591 | Urinary tract infection | genitourinary | DR | 1.44 | 1.41 | 1.46 | 2.97E-141 | 128777 | 2 |
| 286.2 | | | DR | 1.52 | 1.49 | 1.54 | 3.46E-141 | 128777 | 2 |
| 411.1 | Unstable angina (intermediate coronary syndrome) | circulatory system | DR | 1.55 | 1.52 | 1.58 | 2.27E-138 | 128777 | 2 |
| 428.2 | Heart failure NOS | circulatory system | DR | 1.93 | 1.88 | 1.98 | 1.47E-137 | 128777 | 2 |
| 415.2 | Chronic pulmonary heart disease | circulatory system | DR | 1.70 | 1.66 | 1.73 | 7.16E-136 | 128777 | 2 |
| 872 | Traumatic amputation | injuries & poisonings | DR | 2.46 | 2.37 | 2.55 | 1.53E-135 | 128777 | 2 |
| 458.1 | Orthostatic hypotension | circulatory system | DR | 1.61 | 1.58 | 1.64 | 2.29E-135 | 128777 | 2 |
| 702 | Degenerative skin conditions and other dermatoses | dermatologic | DR | 1.40 | 1.38 | 1.42 | 2.37E-135 | 128777 | 2 |
| 172.2 | Other non-epithelial cancer of skin | neoplasms | DR | 1.47 | 1.44 | 1.49 | 2.39E-135 | 128777 | 2 |
| 701.1 | Keratoderma, acquired | dermatologic | DR | 1.61 | 1.58 | 1.65 | 4.95E-135 | 128777 | 2 |
| 356 | Hereditary and idiopathic peripheral neuropathy | neurological | DR | 1.45 | 1.43 | 1.47 | 5.18E-135 | 128777 | 2 |
| 599.2 | Retention of urine | genitourinary | DR | 1.53 | 1.50 | 1.56 | 5.98E-133 | 128777 | 2 |
| 713.5 | Arthropathy associated with neurological disorders | musculoskeletal | DR | 3.36 | 3.20 | 3.53 | 2.20E-132 | 128777 | 2 |
| 275 | Disorders of mineral metabolism | endocrine/metabolic | DR | 1.51 | 1.48 | 1.53 | 2.33E-132 | 128777 | 2 |
| 854 | Complications of cardiac/vascular device, implant, and graft | injuries & poisonings | DR | 1.82 | 1.77 | 1.86 | 2.73E-128 | 128777 | 2 |
| 300.1 | Anxiety disorder | mental disorders | DR | 0.72 | 0.71 | 0.73 | 7.63E-127 | 128777 | 2 |
| 250.15 | Diabetes type 1 with peripheral circulatory disorders | endocrine/metabolic | DR | 4.98 | 4.65 | 5.32 | 4.79E-126 | 128777 | 2 |
| 427.22 | Atrial flutter | circulatory system | DR | 1.59 | 1.56 | 1.62 | 1.03E-125 | 128777 | 2 |
| 440.22 | Atherosclerosis of native arteries of the extremities with intermittent claudication | circulatory system | DR | 1.74 | 1.70 | 1.78 | 8.07E-125 | 128777 | 2 |
| 994.2 | Sepsis | injuries & poisonings | DR | 1.56 | 1.54 | 1.59 | 2.41E-123 | 128777 | 2 |
| 426.3 | Bundle branch block | circulatory system | DR | 1.66 | 1.63 | 1.70 | 2.76E-123 | 128777 | 2 |
| 41.2 | Streptococcus infection | infectious diseases | DR | 1.99 | 1.93 | 2.05 | 5.04E-123 | 128777 | 2 |
| 706.8 | Other specified diseases of sebaceous glands | dermatologic | DR | 1.49 | 1.46 | 1.51 | 5.65E-123 | 128777 | 2 |
| 401.3 | Other hypertensive complications | circulatory system | DR | 1.83 | 1.78 | 1.88 | 1.94E-122 | 128777 | 2 |
| 276.4 | Acid-base balance disorder | endocrine/metabolic | DR | 1.61 | 1.58 | 1.64 | 2.00E-122 | 128777 | 2 |
| 503 | Pulmonary congestion and hypostasis | respiratory | DR | 1.99 | 1.93 | 2.05 | 1.44E-121 | 128777 | 2 |
| 414 | Other forms of chronic heart disease | circulatory system | DR | 1.56 | 1.53 | 1.59 | 7.06E-121 | 128777 | 2 |
| 292.4 | Altered mental status | mental disorders | DR | 1.56 | 1.53 | 1.59 | 7.20E-121 | 128777 | 2 |
| 340 | Migraine | neurological | DR | 0.57 | 0.56 | 0.59 | 9.83E-121 | 128777 | 2 |
| 41.1 | Staphylococcus infections | infectious diseases | DR | 1.67 | 1.64 | 1.71 | 3.09E-118 | 128777 | 2 |

| | | | | | | | | | |
|---|---|---|---|---|---|---|---|---|---|
| 361 | Retinal detachments and defects | sense organs | DR | 1.80 | 1.76 | 1.85 | 8.00E-118 | 128777 | 2 |
| 300.9 | Posttraumatic stress disorder | mental disorders | DR | 0.71 | 0.70 | 0.72 | 1.67E-117 | 128777 | 2 |
| 741.3 | Difficulty in walking | musculoskeletal | DR | 1.46 | 1.44 | 1.48 | 4.25E-114 | 128777 | 2 |
| 272.1 | Hyperlipidemia | endocrine/metabolic | DR | 1.79 | 1.74 | 1.83 | 7.60E-112 | 128777 | 2 |
| 509.1 | Respiratory failure | respiratory | DR | 1.47 | 1.45 | 1.50 | 1.44E-111 | 128777 | 2 |
| 296.1 | Bipolar | mental disorders | DR | 0.58 | 0.56 | 0.59 | 5.82E-111 | 128777 | 2 |
| 389.1 | Sensorineural hearing loss | sense organs | DR | 1.35 | 1.33 | 1.37 | 8.75E-111 | 128777 | 2 |
| 425.1 | Primary/intrinsic cardiomyopathies | circulatory system | DR | 1.53 | 1.50 | 1.56 | 3.88E-109 | 128777 | 2 |
| 512.7 | Shortness of breath | respiratory | DR | 1.31 | 1.30 | 1.33 | 3.20E-108 | 128777 | 2 |
| 731 | Osteitis deformans and osteopathies associated with other disorders classified elsewhere | musculoskeletal | DR | 3.00 | 2.85 | 3.16 | 1.27E-106 | 128777 | 2 |
| 380.4 | Impacted cerumen | sense organs | DR | 1.38 | 1.36 | 1.40 | 2.96E-106 | 128777 | 2 |
| 425 | Cardiomyopathy | circulatory system | DR | 1.53 | 1.50 | 1.56 | 3.85E-106 | 128777 | 2 |
| 260 | Protein-calorie malnutrition | endocrine/metabolic | DR | 1.63 | 1.59 | 1.67 | 4.46E-106 | 128777 | 2 |
| 396 | Abnormal heart sounds | circulatory system | DR | 1.58 | 1.55 | 1.61 | 1.00E-104 | 128777 | 2 |
| 480 | Pneumonia | respiratory | DR | 1.37 | 1.35 | 1.39 | 4.44E-104 | 128777 | 2 |
| 276.12 | Hyposmolality and/or hyponatremia | endocrine/metabolic | DR | 1.49 | 1.46 | 1.51 | 2.99E-103 | 128777 | 2 |
| 272 | | | DR | 2.12 | 2.04 | 2.19 | 5.13E-103 | 128777 | 2 |
| 874 | Complication of amputation stump | injuries & poisonings | DR | 2.92 | 2.77 | 3.07 | 8.31E-102 | 128777 | 2 |
| 703.1 | Ingrowing nail | dermatologic | DR | 1.42 | 1.40 | 1.44 | 2.41E-101 | 128777 | 2 |
| 433.6 | Acute, but ill-defined cerebrovascular disease | circulatory system | DR | 1.70 | 1.66 | 1.74 | 5.12E-101 | 128777 | 2 |
| 38.3 | Bacteremia | infectious diseases | DR | 1.73 | 1.69 | 1.78 | 6.83E-101 | 128777 | 2 |
| 1015 | Effects of other external causes | NULL | DR | 1.40 | 1.38 | 1.42 | 8.40E-101 | 128777 | 2 |
| 368 | Visual disturbances | sense organs | DR | 1.34 | 1.32 | 1.36 | 1.80E-100 | 128777 | 2 |
| 536.3 | Gastroparesis | digestive | DR | 2.22 | 2.14 | 2.31 | 1.80E-100 | 128777 | 2 |
| 366.1 | Nonsenile Cataract | sense organs | DR | 1.94 | 1.88 | 2.00 | 6.13E-99 | 128777 | 2 |
| 365.1 | Open-angle glaucoma | sense organs | DR | 1.37 | 1.35 | 1.39 | 7.79E-99 | 128777 | 2 |
| 509 | Respiratory failure, insufficiency, arrest | respiratory | DR | 1.41 | 1.39 | 1.44 | 1.46E-98 | 111475 | 1 |
| 915 | Superficial injury without mention of infection | injuries & poisonings | DR | 1.41 | 1.39 | 1.44 | 8.88E-98 | 128777 | 2 |
| 429 | | | DR | 1.45 | 1.42 | 1.48 | 1.72E-97 | 111475 | 1 |
| 365.11 | Primary open angle glaucoma | sense organs | DR | 1.56 | 1.53 | 1.60 | 6.53E-97 | 128777 | 2 |
| 573.7 | Abnormal results of function study of liver | digestive | DR | 0.68 | 0.67 | 0.69 | 3.83E-95 | 128777 | 2 |
| 395.1 | Nonrheumatic mitral valve disorders | circulatory system | DR | 1.57 | 1.53 | 1.60 | 1.02E-94 | 128777 | 2 |
| 250.11 | Type 1 diabetes with ketoacidosis | endocrine/metabolic | DR | 4.46 | 4.15 | 4.80 | 6.52E-94 | 128777 | 2 |
| 443.7 | Peripheral angiopathy in diseases classified elsewhere | circulatory system | DR | 3.73 | 3.50 | 3.97 | 1.31E-93 | 128777 | 2 |
| 994 | | | DR | 1.48 | 1.45 | 1.51 | 1.16E-90 | 111475 | 1 |
| 290.16 | Vascular dementia | mental disorders | DR | 2.13 | 2.05 | 2.21 | 3.82E-90 | 128777 | 2 |
| 292 | Neurological disorders | mental disorders | DR | 1.34 | 1.32 | 1.36 | 5.69E-90 | 128777 | 2 |
| 275.3 | Disorders of magnesium metabolism | endocrine/metabolic | DR | 1.52 | 1.49 | 1.55 | 5.03E-89 | 128777 | 2 |
| 348.8 | Encephalopathy, not elsewhere classified | neurological | DR | 1.66 | 1.62 | 1.70 | 7.88E-89 | 128777 | 2 |
| 799 | | | DR | 1.57 | 1.53 | 1.60 | 1.15E-88 | 128777 | 2 |
| 563 | Constipation | digestive | DR | 1.33 | 1.31 | 1.34 | 7.42E-88 | 128777 | 2 |
| 386.9 | Dizziness and giddiness (Light-headedness and vertigo) | sense organs | DR | 1.30 | 1.28 | 1.32 | 7.56E-87 | 128777 | 2 |
| 250.21 | Type 2 diabetes with ketoacidosis | endocrine/metabolic | DR | 2.33 | 2.23 | 2.43 | 1.27E-86 | 128777 | 2 |
| 372 | Disorders of conjunctiva | sense organs | DR | 1.55 | 1.51 | 1.58 | 2.33E-85 | 128777 | 2 |
| 272.11 | Hypercholesterolemia | endocrine/metabolic | DR | 1.30 | 1.28 | 1.31 | 4.64E-85 | 128777 | 2 |
| 771.1 | Swelling of limb | symptoms | DR | 1.42 | 1.40 | 1.45 | 5.75E-85 | 128777 | 2 |
| 415 | Pulmonary heart disease | circulatory system | DR | 1.45 | 1.42 | 1.48 | 1.40E-83 | 128777 | 2 |
| 457.3 | | | DR | 1.60 | 1.56 | 1.64 | 3.97E-83 | 128777 | 2 |
| 285.1 | Acute posthemorrhagic anemia | hematopoietic | DR | 1.56 | 1.53 | 1.60 | 8.40E-83 | 128777 | 2 |
| 394 | Rheumatic disease of the heart valves | circulatory system | DR | 1.67 | 1.62 | 1.71 | 2.47E-82 | 128777 | 2 |
| 508 | Pulmonary collapse; interstitial and compensatory emphysema | respiratory | DR | 1.46 | 1.43 | 1.48 | 1.33E-81 | 128777 | 2 |
| 1011 | Complications of surgical and medical procedures | NULL | DR | 1.47 | 1.44 | 1.50 | 3.43E-80 | 128777 | 2 |
| 426.7 | Abnormal electrocardiogram [ECG] [EKG] | circulatory system | DR | 1.33 | 1.31 | 1.35 | 7.96E-80 | 128777 | 2 |
| 172.22 | Squamous cell carcinoma | neoplasms | DR | 1.53 | 1.49 | 1.56 | 1.67E-78 | 128777 | 2 |
| 454.11 | Varicose veins of lower extremity, symptomtic | circulatory system | DR | 1.58 | 1.54 | 1.62 | 3.53E-78 | 128777 | 2 |
| 532 | Dysphagia | digestive | DR | 1.31 | 1.29 | 1.33 | 3.82E-78 | 128777 | 2 |
| 443.8 | Other specified peripheral vascular diseases | circulatory system | DR | 2.00 | 1.93 | 2.08 | 4.59E-78 | 128777 | 2 |
| 275.53 | Disorders of phosphorus metabolism | endocrine/metabolic | DR | 1.90 | 1.84 | 1.97 | 1.48E-77 | 128777 | 2 |
| 296 | Mood disorders | mental disorders | DR | 0.78 | 0.77 | 0.79 | 1.68E-77 | 128777 | 2 |
| 457 | | | DR | 1.54 | 1.51 | 1.58 | 2.37E-77 | 128777 | 2 |
| 300.11 | Generalized anxiety disorder | mental disorders | DR | 0.68 | 0.66 | 0.69 | 2.37E-77 | 128777 | 2 |
| 599 | Other symptoms/disorders or the urinary system | genitourinary | DR | 1.28 | 1.27 | 1.30 | 1.92E-76 | 128777 | 2 |
| 580.2 | Nephrotic syndrome without mention of glomerulonephritis | genitourinary | DR | 3.01 | 2.84 | 3.20 | 2.20E-76 | 128777 | 2 |
| 426.23 | Second degree AV block | circulatory system | DR | 2.45 | 2.34 | 2.58 | 6.94E-76 | 128777 | 2 |
| 743 | | | DR | 1.42 | 1.39 | 1.44 | 3.58E-75 | 128777 | 2 |
| 427.5 | Arrhythmia (cardiac) NOS | circulatory system | DR | 1.32 | 1.30 | 1.34 | 6.87E-75 | 128777 | 2 |
| 41.11 | Methicillin sensitive Staphylococcus aureus | infectious diseases | DR | 1.73 | 1.68 | 1.78 | 2.30E-73 | 128777 | 2 |
| 362.27 | Drusen (degenerative) of retina | sense organs | DR | 1.57 | 1.53 | 1.61 | 1.79E-72 | 128777 | 2 |
| 426.31 | Right bundle branch block | circulatory system | DR | 1.62 | 1.58 | 1.66 | 1.89E-71 | 128777 | 2 |
| 337.1 | Peripheral autonomic neuropathy | neurological | DR | 1.47 | 1.44 | 1.50 | 2.41E-71 | 128777 | 2 |
| 275.5 | Disorders of calcium/phosphorus metabolism | endocrine/metabolic | DR | 1.61 | 1.57 | 1.66 | 3.21E-71 | 128777 | 2 |
| 713 | Arthropathy associated with other disorders classified elsewhere | musculoskeletal | DR | 2.29 | 2.18 | 2.39 | 5.38E-71 | 111475 | 1 |
| 41.12 | Methicillin resistant Staphylococcus aureus | infectious diseases | DR | 1.73 | 1.68 | 1.79 | 9.67E-71 | 128777 | 2 |
| 689 | Disorder of skin and subcutaneous tissue NOS | dermatologic | DR | 1.29 | 1.27 | 1.30 | 1.67E-70 | 128777 | 2 |
| 260.3 | Adult failure to thrive | endocrine/metabolic | DR | 1.96 | 1.89 | 2.04 | 2.06E-70 | 128777 | 2 |
| 363 | Chorioretinal inflammations, scars, and other disorders of choroid | sense organs | DR | 1.57 | 1.53 | 1.61 | 7.15E-70 | 128777 | 2 |
| 296.22 | Major depressive disorder | mental disorders | DR | 0.79 | 0.78 | 0.80 | 1.89E-69 | 128777 | 2 |
| 587 | Kidney replaced by transpant | genitourinary | DR | 3.27 | 3.05 | 3.49 | 4.14E-69 | 128777 | 2 |
| 1013 | Asphyxia and hypoxemia | NULL | DR | 1.40 | 1.37 | 1.43 | 1.34E-68 | 128777 | 2 |
| 250.42 | Other abnormal glucose | endocrine/metabolic | DR | 0.77 | 0.76 | 0.78 | 1.36E-68 | 128777 | 2 |
| 368.4 | Visual field defects | sense organs | DR | 1.53 | 1.49 | 1.57 | 1.44E-68 | 128777 | 2 |
| 337 | Disorders of the autonomic nervous system | neurological | DR | 1.44 | 1.41 | 1.47 | 4.30E-68 | 128777 | 2 |
| 916 | Contusion | injuries & poisonings | DR | 1.30 | 1.28 | 1.32 | 1.34E-67 | 128777 | 2 |
| 110.12 | Althete's foot | infectious diseases | DR | 1.35 | 1.32 | 1.37 | 5.66E-67 | 128777 | 2 |
| 377 | Disorders of optic nerve and visual pathways | sense organs | DR | 1.52 | 1.49 | 1.56 | 1.78E-66 | 128777 | 2 |
| 300.12 | Agorophobia, social phobia, and panic disorder | mental disorders | DR | 0.61 | 0.60 | 0.63 | 1.87E-66 | 128777 | 2 |
| 447 | Other disorders of arteries and arterioles | circulatory system | DR | 1.66 | 1.61 | 1.71 | 2.95E-66 | 128777 | 2 |
| 348 | Other conditions of brain | neurological | DR | 1.53 | 1.49 | 1.57 | 2.86E-65 | 128777 | 2 |
| 297.1 | Suicidal ideation | mental disorders | DR | 0.63 | 0.61 | 0.65 | 8.92E-65 | 128777 | 2 |
| 296.2 | | | DR | 0.80 | 0.79 | 0.81 | 1.10E-64 | 128777 | 2 |
| 429.3 | Symptoms involving cardiovascular system | circulatory system | DR | 1.48 | 1.45 | 1.52 | 2.04E-64 | 128777 | 2 |
| 395.6 | Heart valve replaced | circulatory system | DR | 1.74 | 1.69 | 1.80 | 2.89E-64 | 128777 | 2 |
| 741 | Symptoms and disorders of the joints | musculoskeletal | DR | 1.27 | 1.25 | 1.29 | 4.02E-64 | 128777 | 2 |
| 851 | Complications of transplants and reattached limbs | injuries & poisonings | DR | 1.65 | 1.60 | 1.70 | 4.52E-64 | 128777 | 2 |
| 599.4 | Urinary incontinence | genitourinary | DR | 1.36 | 1.33 | 1.38 | 6.27E-64 | 128777 | 2 |

| | | | | | | | | | |
|---|---|---|---|---|---|---|---|---|---|
| 781 | Symptoms involving nervous and musculoskeletal systems | symptoms | DR | 1.48 | 1.45 | 1.51 | 8.26E-64 | 128777 | 2 |
| 426.24 | Atrioventricular block, complete | circulatory system | DR | 1.81 | 1.75 | 1.88 | 1.28E-63 | 128777 | 2 |
| 686 | Other local infections of skin and subcutaneous tissue | dermatologic | DR | 1.45 | 1.42 | 1.49 | 3.28E-63 | 128777 | 2 |
| 172.1 | Melanomas of skin, dx or hx | neoplasms | DR | 1.34 | 1.32 | 1.37 | 5.97E-63 | 128777 | 2 |
| 389 | Hearing loss | sense organs | DR | 1.26 | 1.24 | 1.28 | 9.86E-63 | 128777 | 2 |
| 379.5 | Disorders of iris and ciliary body | sense organs | DR | 1.70 | 1.65 | 1.76 | 1.65E-62 | 128777 | 2 |
| 454.1 | Varicose veins of lower extremity | circulatory system | DR | 1.43 | 1.40 | 1.46 | 5.03E-62 | 128777 | 2 |
| 414.2 | | | DR | 1.77 | 1.71 | 1.84 | 6.48E-62 | 128777 | 2 |
| 596 | Other disorders of bladder | genitourinary | DR | 1.36 | 1.33 | 1.38 | 3.27E-61 | 128777 | 2 |
| 364 | Corneal opacity and other disorders of cornea | sense organs | DR | 1.52 | 1.48 | 1.56 | 6.75E-61 | 128777 | 2 |
| 172.21 | Basal cell carcinoma | neoplasms | DR | 1.38 | 1.36 | 1.41 | 2.14E-60 | 128777 | 2 |
| 250 | Diabetes mellitus | endocrine/metabolic | DR | 2.17 | 2.07 | 2.28 | 3.47E-60 | 128777 | 2 |
| 252.1 | Hyperparathyroidism | endocrine/metabolic | DR | 1.78 | 1.71 | 1.84 | 4.78E-60 | 128777 | 2 |
| 433.11 | Occlusion of cerebral arteries, with cerebral infarction | circulatory system | DR | 1.94 | 1.86 | 2.02 | 9.14E-60 | 128777 | 2 |
| 505 | Other pulmonary inflamation or edema | respiratory | DR | 1.95 | 1.87 | 2.03 | 1.09E-59 | 128777 | 2 |
| 571 | | | DR | 0.75 | 0.74 | 0.77 | 1.80E-59 | 111475 | 1 |
| 426.32 | Left bundle branch block | circulatory system | DR | 1.63 | 1.58 | 1.68 | 4.87E-59 | 128777 | 2 |
| 788 | Syncope and collapse | symptoms | DR | 1.28 | 1.26 | 1.30 | 9.17E-58 | 128777 | 2 |
| 706 | | | DR | 1.25 | 1.23 | 1.27 | 1.09E-57 | 128777 | 2 |
| 374.3 | Ptosis of eyelid | sense organs | DR | 1.48 | 1.44 | 1.52 | 1.64E-57 | 128777 | 2 |
| 427.8 | Sinoatrial node dysfunction (Bradycardia) | circulatory system | DR | 1.65 | 1.59 | 1.70 | 1.99E-57 | 128777 | 2 |
| 252 | Disorders of parathyroid gland | endocrine/metabolic | DR | 1.76 | 1.70 | 1.82 | 6.71E-57 | 128777 | 2 |
| 313.1 | Attention deficit hyperactivity disorder | mental disorders | DR | 0.46 | 0.44 | 0.48 | 7.93E-57 | 128777 | 2 |
| 411.9 | Other acute and subacute forms of ischemic heart disease | circulatory system | DR | 1.80 | 1.74 | 1.87 | 1.87E-56 | 128777 | 2 |
| 370 | Keratitis | sense organs | DR | 1.44 | 1.41 | 1.48 | 4.70E-56 | 128777 | 2 |
| 453 | Chronic venous hypertension | circulatory system | DR | 1.91 | 1.83 | 1.99 | 1.17E-55 | 128777 | 2 |
| 701 | Other hypertrophic and atrophic conditions of skin | dermatologic | DR | 1.25 | 1.23 | 1.26 | 2.14E-55 | 128777 | 2 |
| 297 | Suicidal ideation or attempt | mental disorders | DR | 0.69 | 0.67 | 0.71 | 1.13E-54 | 111475 | 1 |
| 420 | Carditis | circulatory system | DR | 1.55 | 1.50 | 1.59 | 1.22E-54 | 111475 | 1 |
| 426.25 | Other heart block | circulatory system | DR | 2.19 | 2.08 | 2.30 | 6.83E-54 | 128777 | 2 |
| 1010.2 | screening for malignant neoplasms | | DR | 0.81 | 0.80 | 0.82 | 9.29E-54 | 128777 | 2 |
| 301 | Personality disorders | mental disorders | DR | 0.67 | 0.65 | 0.68 | 1.05E-53 | 128777 | 2 |
| 313 | Pervasive developmental disorders | mental disorders | DR | 0.53 | 0.50 | 0.55 | 1.92E-53 | 128777 | 2 |
| 292.2 | Mild cognitive impairment | mental disorders | DR | 1.41 | 1.38 | 1.44 | 1.97E-53 | 128777 | 2 |
| 286.9 | Abnormal coagulation profile | hematopoietic | DR | 1.60 | 1.55 | 1.65 | 9.53E-53 | 128777 | 2 |
| 571.5 | Other chronic nonalcoholic liver disease | digestive | DR | 0.77 | 0.76 | 0.78 | 1.84E-52 | 128777 | 2 |
| 224 | Benign neoplasm of eye | neoplasms | DR | 1.39 | 1.36 | 1.42 | 2.48E-52 | 128777 | 2 |
| 290.11 | Alzheimer's disease | mental disorders | DR | 1.95 | 1.87 | 2.04 | 4.22E-52 | 128777 | 2 |
| 440.9 | Atherosclerosis of aorta | circulatory system | DR | 1.60 | 1.55 | 1.65 | 4.69E-52 | 128777 | 2 |
| 480.1 | Bacterial pneumonia | respiratory | DR | 1.40 | 1.37 | 1.43 | 6.06E-52 | 128777 | 2 |
| 702.2 | Seborrheic keratosis | dermatologic | DR | 1.22 | 1.21 | 1.24 | 6.36E-52 | 128777 | 2 |
| 426.92 | Cardiac defibrillator in situ | circulatory system | DR | 1.53 | 1.49 | 1.57 | 6.99E-52 | 128777 | 2 |
| 374.1 | Ectropion or entropion | sense organs | DR | 1.60 | 1.55 | 1.65 | 7.04E-52 | 128777 | 2 |
| 270.2 | Disorders of amino-acid metabolism | endocrine/metabolic | DR | 0.38 | 0.35 | 0.40 | 3.18E-51 | 111475 | 1 |
| 870 | Open wounds of head; neck; and trunk | injuries & poisonings | DR | 1.35 | 1.32 | 1.37 | 3.59E-51 | 128777 | 2 |
| 281 | Other deficiency anemia | hematopoietic | DR | 1.39 | 1.36 | 1.42 | 4.67E-51 | 128777 | 2 |
| 454 | Varicose veins | circulatory system | DR | 1.36 | 1.33 | 1.38 | 5.64E-51 | 128777 | 2 |
| 38.2 | Gram positive septicemia | infectious diseases | DR | 1.90 | 1.82 | 1.98 | 6.88E-51 | 128777 | 2 |
| 374.6 | Dermatochalasis | sense organs | DR | 1.36 | 1.33 | 1.39 | 8.72E-51 | 128777 | 2 |
| 286 | | | DR | 1.42 | 1.39 | 1.46 | 1.91E-50 | 128777 | 2 |
| 363.3 | Chorioretinal scars | sense organs | DR | 1.50 | 1.46 | 1.54 | 2.34E-50 | 128777 | 2 |
| 261.2 | Vitamin B-complex deficiencies | endocrine/metabolic | DR | 1.34 | 1.32 | 1.37 | 6.13E-50 | 128777 | 2 |
| 287 | Purpura and other hemorrhagic conditions | hematopoietic | DR | 1.35 | 1.32 | 1.37 | 7.30E-50 | 128777 | 2 |
| 743.1 | | | DR | 1.51 | 1.46 | 1.55 | 1.51E-49 | 111475 | 1 |
| 512.9 | Other dyspnea | respiratory | DR | 1.20 | 1.19 | 1.22 | 5.44E-49 | 128777 | 2 |
| 580.32 | Nephritis and nephropathy with pathological lesion | genitourinary | DR | 2.11 | 2.01 | 2.22 | 1.03E-48 | 128777 | 2 |
| 402 | Elevated blood pressure reading without diagnosis of hypertension | circulatory system | DR | 0.74 | 0.73 | 0.76 | 1.30E-48 | 128777 | 2 |
| 41.4 | E. coli | infectious diseases | DR | 1.63 | 1.58 | 1.69 | 2.39E-48 | 128777 | 2 |
| 835 | Internal derangement of knee | injuries & poisonings | DR | 0.73 | 0.71 | 0.74 | 2.41E-47 | 128777 | 2 |
| 743.11 | Osteoporosis NOS | musculoskeletal | DR | 1.45 | 1.41 | 1.48 | 4.94E-47 | 128777 | 2 |
| 962.2 | | | DR | 2.94 | 2.73 | 3.17 | 7.91E-47 | 128777 | 2 |
| 224.1 | Benign neoplasm of eye, uveal | neoplasms | DR | 1.37 | 1.34 | 1.40 | 8.91E-47 | 128777 | 2 |
| 287.3 | Thrombocytopenia | hematopoietic | DR | 1.34 | 1.31 | 1.37 | 1.96E-46 | 128777 | 2 |
| 427.6 | Premature beats | circulatory system | DR | 1.39 | 1.36 | 1.43 | 5.27E-46 | 128777 | 2 |
| 351 | Other peripheral nerve disorders | neurological | DR | 1.20 | 1.19 | 1.22 | 7.37E-46 | 128777 | 2 |
| 280.2 | Iron deficiency anemia secondary to blood loss (chronic) | hematopoietic | DR | 1.49 | 1.45 | 1.53 | 2.51E-45 | 128777 | 2 |
| 292.1 | Aphasia/speech disturbance | mental disorders | DR | 1.40 | 1.37 | 1.43 | 8.63E-45 | 128777 | 2 |
| 690 | Erythematosquamous dermatosis | dermatologic | DR | 1.27 | 1.25 | 1.30 | 1.01E-44 | 111475 | 1 |
| 38.1 | Gram negative septicemia | infectious diseases | DR | 1.66 | 1.60 | 1.72 | 1.06E-44 | 128777 | 2 |
| 301.2 | Antisocial/borderline personality disorder | mental disorders | DR | 0.53 | 0.51 | 0.56 | 3.50E-44 | 128777 | 2 |
| 357 | Inflammatory and toxic neuropathy | neurological | DR | 1.33 | 1.30 | 1.36 | 6.54E-44 | 128777 | 2 |
| 429.2 | Abnormal function study of cardiovascular system | circulatory system | DR | 1.38 | 1.35 | 1.41 | 2.01E-43 | 128777 | 2 |
| 304 | Adjustment reaction | mental disorders | DR | 0.82 | 0.81 | 0.83 | 1.94E-42 | 128777 | 2 |
| 690.1 | Seborrheic dermatitis | dermatologic | DR | 1.26 | 1.24 | 1.28 | 2.49E-42 | 128777 | 2 |
| 420.3 | Endocarditis | circulatory system | DR | 1.67 | 1.60 | 1.73 | 9.22E-42 | 128777 | 2 |
| 377.1 | Optic atrophy | sense organs | DR | 1.56 | 1.51 | 1.61 | 9.44E-42 | 128777 | 2 |
| 297.2 | Suicide or self-inflicted injury | mental disorders | DR | 0.54 | 0.52 | 0.57 | 1.14E-41 | 128777 | 2 |
| 355.1 | Chronic pain syndrome | neurological | DR | 0.76 | 0.75 | 0.78 | 1.67E-41 | 128777 | 2 |
| 370.3 | Keratoconjunctivitis | sense organs | DR | 1.48 | 1.44 | 1.53 | 5.10E-41 | 128777 | 2 |
| 350.3 | Lack of coordination | neurological | DR | 1.51 | 1.46 | 1.56 | 6.94E-41 | 128777 | 2 |
| 394.1 | Mitral valve stenosis and aortic valve stenosis | circulatory system | DR | 1.80 | 1.72 | 1.88 | 8.70E-41 | 128777 | 2 |
| 459.7 | Blood vessel replaced | circulatory system | DR | 1.71 | 1.64 | 1.78 | 1.01E-40 | 128777 | 2 |
| 361.1 | Retinal detachment with retinal defect | sense organs | DR | 1.72 | 1.65 | 1.80 | 1.87E-40 | 128777 | 2 |
| 418 | Nonspecific chest pain | circulatory system | DR | 1.18 | 1.16 | 1.19 | 2.34E-40 | 128777 | 2 |
| 295.1 | Schizophrenia | mental disorders | DR | 0.62 | 0.59 | 0.64 | 2.37E-40 | 128777 | 2 |
| 695 | Erythematous conditions | dermatologic | DR | 1.25 | 1.23 | 1.27 | 6.98E-40 | 111475 | 1 |
| 789 | Nausea and vomiting | symptoms | DR | 1.22 | 1.20 | 1.24 | 8.66E-40 | 128777 | 2 |
| 783 | Fever of unknown origin | symptoms | DR | 1.26 | 1.23 | 1.28 | 2.67E-39 | 128777 | 2 |
| 270.3 | | | DR | 1.62 | 1.56 | 1.68 | 2.83E-39 | 128777 | 2 |
| 980 | | | DR | 1.66 | 1.59 | 1.72 | 3.99E-39 | 128777 | 2 |
| 458.2 | Iatrogenic hypotension | circulatory system | DR | 1.50 | 1.46 | 1.55 | 7.38E-39 | 128777 | 2 |
| 743.2 | Pathologic fracture | musculoskeletal | DR | 1.57 | 1.51 | 1.62 | 1.44E-38 | 128777 | 2 |
| 875 | | | DR | 1.61 | 1.55 | 1.67 | 1.54E-38 | 128777 | 2 |
| 512 | Other symptoms of respiratory system | respiratory | DR | 1.20 | 1.19 | 1.22 | 3.19E-38 | 111475 | 1 |
| 261 | Vitamin deficiency | endocrine/metabolic | DR | 1.19 | 1.17 | 1.21 | 4.55E-38 | 128777 | 2 |
| 427.1 | Paroxysmal tachycardia, unspecified | circulatory system | DR | 1.33 | 1.30 | 1.36 | 4.92E-38 | 128777 | 2 |
| 447.1 | Stricture of artery | circulatory system | DR | 1.82 | 1.74 | 1.91 | 1.06E-36 | 128777 | 2 |
| 327.3 | Sleep apnea | neurological | DR | 0.85 | 0.84 | 0.86 | 3.00E-36 | 128777 | 2 |

| | | | | | | | | | |
|---|---|---|---|---|---|---|---|---|---|
| 429.1 | Heart transplant/surgery | circulatory system | DR | 1.57 | 1.52 | 1.63 | 3.68E-36 | 128777 | 2 |
| 427.12 | Paroxysmal ventricular tachycardia | circulatory system | DR | 1.40 | 1.36 | 1.43 | 5.08E-36 | 128777 | 2 |
| 514 | Abnormal findings examination of lungs | respiratory | DR | 1.21 | 1.19 | 1.23 | 6.46E-36 | 128777 | 2 |
| 365.2 | Primary angle-closure glaucoma | sense organs | DR | 1.48 | 1.43 | 1.52 | 9.76E-36 | 128777 | 2 |
| 172.3 | Carcinoma in situ of skin | neoplasms | DR | 1.50 | 1.45 | 1.55 | 1.45E-35 | 128777 | 2 |
| 318 | Tobacco use disorder | mental disorders | DR | 0.86 | 0.85 | 0.87 | 2.13E-35 | 128777 | 2 |
| 394.7 | Disease of tricuspid valve | circulatory system | DR | 1.60 | 1.54 | 1.67 | 5.93E-35 | 128777 | 2 |
| 281.9 | Deficiency anemias | hematopoietic | DR | 1.59 | 1.53 | 1.65 | 6.69E-35 | 128777 | 2 |
| 331.9 | Cerebral degeneration, unspecified | neurological | DR | 1.68 | 1.61 | 1.76 | 1.43E-34 | 128777 | 2 |
| 327.4 | Insomnia | neurological | DR | 0.84 | 0.83 | 0.85 | 1.48E-34 | 128777 | 2 |
| 339 | Other headache syndromes | neurological | DR | 0.84 | 0.83 | 0.85 | 3.44E-34 | 128777 | 2 |
| 173 | Neoplasm of uncertain behavior of skin | neoplasms | DR | 1.19 | 1.17 | 1.20 | 7.13E-34 | 128777 | 2 |
| 338.2 | Chronic pain | neurological | DR | 0.84 | 0.83 | 0.85 | 1.42E-33 | 128777 | 2 |
| 395.3 | Nonrheumatic tricuspid valve disorders | circulatory system | DR | 1.56 | 1.50 | 1.62 | 1.57E-33 | 128777 | 2 |
| 800 | Fracture of lower limb | injuries & poisonings | DR | 1.37 | 1.34 | 1.41 | 2.50E-33 | 128777 | 2 |
| 274 | | | DR | 1.22 | 1.20 | 1.24 | 3.47E-33 | 111475 | 1 |
| 604.1 | Redundant prepuce and phimosis/BXO | genitourinary | DR | 1.67 | 1.60 | 1.74 | 3.81E-33 | 128777 | 2 |
| 362.6 | Peripheral retinal degenerations | sense organs | DR | 1.40 | 1.37 | 1.45 | 8.97E-33 | 128777 | 2 |
| 440.1 | Atherosclerosis of renal artery | circulatory system | DR | 2.08 | 1.96 | 2.22 | 1.26E-32 | 128777 | 2 |
| 452 | Other venous embolism and thrombosis | circulatory system | DR | 1.26 | 1.23 | 1.28 | 1.29E-32 | 128777 | 2 |
| 251 | Other disorders of pancreatic internal secretion | endocrine/metabolic | DR | 2.22 | 2.08 | 2.38 | 1.74E-32 | 128777 | 2 |
| 420.2 | Pericarditis | circulatory system | DR | 1.52 | 1.47 | 1.57 | 1.79E-32 | 128777 | 2 |
| 429.9 | Cardiac complications, not elsewhere classified | circulatory system | DR | 1.61 | 1.55 | 1.68 | 1.88E-32 | 128777 | 2 |
| 513 | Respiratory abnormalities | respiratory | DR | 1.28 | 1.25 | 1.30 | 1.94E-32 | 128777 | 2 |
| 729 | Other disorders of soft tissues | musculoskeletal | DR | 1.31 | 1.28 | 1.34 | 2.06E-32 | 128777 | 2 |
| 362.7 | Hereditary retinal dystrophies | sense organs | DR | 1.76 | 1.67 | 1.84 | 2.46E-32 | 128777 | 2 |
| 599.5 | Frequency of urination and polyuria | genitourinary | DR | 1.21 | 1.19 | 1.23 | 2.53E-32 | 128777 | 2 |
| 728.71 | Contracture of palmar fascia [Dupuytren's disease] | musculoskeletal | DR | 1.50 | 1.45 | 1.55 | 4.08E-32 | 128777 | 2 |
| 797 | Shock | symptoms | DR | 1.58 | 1.52 | 1.64 | 4.75E-32 | 128777 | 2 |
| 763 | Thoracic or lumbosacral neuritis or radiculitis, unspecified | symptoms | DR | 0.83 | 0.81 | 0.84 | 1.64E-31 | 128777 | 2 |
| 444 | Arterial embolism and thrombosis | circulatory system | DR | 1.51 | 1.46 | 1.56 | 1.95E-31 | 128777 | 2 |
| 800.1 | Fracture of neck of femur | injuries & poisonings | DR | 1.75 | 1.67 | 1.84 | 2.26E-31 | 128777 | 2 |
| 433.12 | Cerebral atherosclerosis | circulatory system | DR | 1.74 | 1.66 | 1.82 | 3.05E-31 | 128777 | 2 |
| 427.61 | Supraventricular premature beats | circulatory system | DR | 1.54 | 1.48 | 1.59 | 4.81E-31 | 128777 | 2 |
| 444.1 | Arterial embolism and thrombosis of lower extremity artery | circulatory system | DR | 1.69 | 1.62 | 1.77 | 5.23E-31 | 128777 | 2 |
| 561.1 | Diarrhea | digestive | DR | 1.17 | 1.16 | 1.19 | 6.34E-31 | 128777 | 2 |
| 281.1 | Megaloblastic anemia | hematopoietic | DR | 1.34 | 1.30 | 1.37 | 1.08E-30 | 111475 | 1 |
| 427.42 | Cardiac arrest | circulatory system | DR | 1.66 | 1.59 | 1.74 | 1.20E-30 | 128777 | 2 |
| 80 | Postoperative infection | infectious diseases | DR | 1.37 | 1.33 | 1.41 | 1.83E-30 | 128777 | 2 |
| 1010.6 | Persons encountering health services in circumstances related to reproduction | | DR | 0.37 | 0.34 | 0.41 | 1.89E-30 | 128777 | 2 |
| 244.4 | Hypothyroidism NOS | endocrine/metabolic | DR | 1.20 | 1.18 | 1.22 | 1.97E-30 | 128777 | 2 |
| 605 | Erectile dysfunction [ED] | genitourinary | DR | 1.16 | 1.15 | 1.18 | 2.00E-30 | 128777 | 2 |
| 1002 | Symptoms concerning nutrition, metabolism, and development | NULL | DR | 1.25 | 1.22 | 1.27 | 2.18E-30 | 128777 | 2 |
| 53 | Herpes zoster | infectious diseases | DR | 1.30 | 1.27 | 1.33 | 2.64E-30 | 128777 | 2 |
| 592.1 | Cystitis | genitourinary | DR | 1.39 | 1.35 | 1.43 | 3.00E-30 | 128777 | 2 |
| 8.52 | Intestinal infection due to C. difficile | infectious diseases | DR | 1.52 | 1.47 | 1.58 | 4.22E-30 | 128777 | 2 |
| 378.5 | Paralytic strabismus | sense organs | DR | 1.71 | 1.63 | 1.79 | 4.30E-30 | 128777 | 2 |
| 801 | Fracture of ankle and foot | injuries & poisonings | DR | 1.29 | 1.27 | 1.32 | 5.42E-30 | 128777 | 2 |
| 292.11 | Aphasia | mental disorders | DR | 1.65 | 1.58 | 1.73 | 9.08E-30 | 128777 | 2 |
| 368.2 | Diplopia and disorders of binocular vision | sense organs | DR | 1.34 | 1.30 | 1.37 | 3.66E-29 | 128777 | 2 |
| 340.1 | Migrain with aura | neurological | DR | 0.57 | 0.55 | 0.60 | 5.37E-29 | 128777 | 2 |
| 964 | Poisoning by agents primarily affecting blood constituents | injuries & poisonings | DR | 1.73 | 1.65 | 1.81 | 6.66E-29 | 111475 | 1 |
| 857 | Mechanical complication of unspecified genitourinary device, implant, and graft | injuries & poisonings | DR | 1.56 | 1.50 | 1.63 | 9.02E-29 | 128777 | 2 |
| 819 | Skull and face fracture and other intercranial injury | injuries & poisonings | DR | 0.73 | 0.71 | 0.75 | 9.91E-29 | 128777 | 2 |
| 327.32 | Obstructive sleep apnea | neurological | DR | 0.87 | 0.86 | 0.88 | 1.20E-28 | 128777 | 2 |
| 513.32 | Orthopnea | respiratory | DR | 1.80 | 1.70 | 1.89 | 1.65E-28 | 128777 | 2 |
| 378 | Strabismus and other disorders of binocular eye movements | sense organs | DR | 1.29 | 1.26 | 1.32 | 4.86E-28 | 128777 | 2 |
| 327 | Sleep disorders | neurological | DR | 0.87 | 0.86 | 0.88 | 5.25E-28 | 128777 | 2 |
| 578.9 | Hemorrhage of gastrointestinal tract | digestive | DR | 1.26 | 1.23 | 1.29 | 1.42E-27 | 128777 | 2 |
| 743.9 | Osteopenia or other disorder of bone and cartilage | musculoskeletal | DR | 1.28 | 1.25 | 1.31 | 1.86E-27 | 128777 | 2 |
| 561 | Symptoms involving digestive system | digestive | DR | 1.16 | 1.14 | 1.18 | 1.98E-27 | 128777 | 2 |
| 274.1 | Gout | endocrine/metabolic | DR | 1.20 | 1.18 | 1.22 | 2.47E-27 | 128777 | 2 |
| 564.1 | Irritable Bowel Syndrome | digestive | DR | 0.73 | 0.70 | 0.75 | 2.59E-27 | 128777 | 2 |
| 501 | Pneumonitis due to inhalation of food or vomitus | respiratory | DR | 1.47 | 1.42 | 1.53 | 2.69E-27 | 128777 | 2 |
| 450 | Noninfectious disorders of lymphatic channels | circulatory system | DR | 1.41 | 1.37 | 1.46 | 4.58E-27 | 128777 | 2 |
| 362.31 | Separation of retinal layers | sense organs | DR | 1.84 | 1.74 | 1.95 | 4.75E-27 | 128777 | 2 |
| 972 | Poisoning by agents primarily affecting the cardiovascular system | injuries & poisonings | DR | 1.53 | 1.47 | 1.60 | 5.04E-27 | 128777 | 2 |
| 509.8 | Dependence on respirator [Ventilator] or supplemental oxygen | respiratory | DR | 1.30 | 1.27 | 1.34 | 5.10E-27 | 128777 | 2 |
| 331 | Other cerebral degenerations | neurological | DR | 1.52 | 1.46 | 1.58 | 6.28E-27 | 128777 | 2 |
| 596.1 | Bladder neck obstruction | genitourinary | DR | 1.44 | 1.39 | 1.49 | 9.07E-27 | 128777 | 2 |
| 589 | Abnormal results of function study of kidney | genitourinary | DR | 1.48 | 1.42 | 1.53 | 1.06E-26 | 128777 | 2 |
| 574.1 | Cholelithiasis | digestive | DR | 1.25 | 1.22 | 1.27 | 1.53E-26 | 128777 | 2 |
| 377.3 | Optic neuritis/neuropathy | sense organs | DR | 1.46 | 1.41 | 1.51 | 1.75E-26 | 128777 | 2 |
| 797.1 | Cardiogenic shock | symptoms | DR | 1.79 | 1.70 | 1.89 | 1.80E-26 | 128777 | 2 |
| 8.5 | Bacterial enteritis | infectious diseases | DR | 1.44 | 1.39 | 1.48 | 2.38E-26 | 128777 | 2 |
| 800.2 | Fracture of unspecified part of femur | injuries & poisonings | DR | 1.76 | 1.67 | 1.86 | 2.80E-26 | 128777 | 2 |
| 495 | Asthma | respiratory | DR | 0.82 | 0.81 | 0.84 | 3.23E-26 | 128777 | 2 |
| 286.7 | Other and unspecified coagulation defects | hematopoietic | DR | 1.40 | 1.35 | 1.44 | 3.87E-26 | 128777 | 2 |
| 477 | Epistaxis or throat hemorrhage | respiratory | DR | 1.31 | 1.28 | 1.35 | 3.95E-26 | 128777 | 2 |
| 276.11 | Hyperosmolality and/or hypernatremia | endocrine/metabolic | DR | 1.56 | 1.50 | 1.63 | 5.29E-26 | 128777 | 2 |
| 112 | Candidiasis | infectious diseases | DR | 1.27 | 1.24 | 1.30 | 7.32E-26 | 128777 | 2 |
| 276.42 | Alkalosis | endocrine/metabolic | DR | 1.59 | 1.52 | 1.66 | 7.83E-26 | 128777 | 2 |
| 574 | | | DR | 1.24 | 1.21 | 1.26 | 9.40E-26 | 111475 | 1 |
| 364.5 | Corneal dystrophy | sense organs | DR | 1.47 | 1.41 | 1.52 | 1.17E-25 | 128777 | 2 |
| 850 | Hemorrhage or hematoma complicating a procedure | injuries & poisonings | DR | 1.37 | 1.33 | 1.42 | 1.40E-25 | 128777 | 2 |
| 964.1 | Anticoagulants causing adverse effects | injuries & poisonings | DR | 1.68 | 1.60 | 1.77 | 2.67E-25 | 128777 | 2 |
| 244 | | | DR | 1.19 | 1.17 | 1.21 | 4.66E-25 | 128777 | 2 |
| 740 | | | DR | 1.15 | 1.13 | 1.16 | 3.05E-24 | 128777 | 2 |
| 596.5 | Functional disorders of bladder | genitourinary | DR | 1.27 | 1.24 | 1.30 | 3.94E-24 | 128777 | 2 |
| 470 | Septal Deviations/Turbinate Hypertrophy | respiratory | DR | 0.77 | 0.75 | 0.79 | 5.13E-24 | 128777 | 2 |
| 415.21 | Primary pulmonary hypertension | circulatory system | DR | 1.59 | 1.52 | 1.66 | 5.33E-24 | 128777 | 2 |
| 457.2 | | | DR | 1.52 | 1.46 | 1.59 | 8.02E-24 | 128777 | 2 |
| 687 | Symptoms affecting skin | dermatologic | DR | 1.20 | 1.18 | 1.23 | 1.06E-23 | 128777 | 2 |
| 371.1 | Uveitis, noninfectious or NOS | sense organs | DR | 1.49 | 1.43 | 1.55 | 1.08E-23 | 128777 | 2 |

| | | | | | | | | | |
|---|---|---|---|---|---|---|---|---|---|
| 288.2 | Elevated white blood cell count | hematopoietic | DR | 1.21 | 1.19 | 1.23 | 1.12E-23 | 128777 | 2 |
| 367.1 | Myopia | sense organs | DR | 1.14 | 1.13 | 1.16 | 1.29E-23 | 128777 | 2 |
| 364.4 | Corneal degenerations | sense organs | DR | 1.51 | 1.45 | 1.57 | 1.67E-23 | 128777 | 2 |
| 765 | Cervical radiculitis | symptoms | DR | 0.82 | 0.80 | 0.83 | 1.84E-23 | 128777 | 2 |
| 368.3 | Anisometropia | sense organs | DR | 1.41 | 1.36 | 1.46 | 1.87E-23 | 128777 | 2 |
| 994.21 | Septic shock | injuries & poisonings | DR | 1.45 | 1.40 | 1.51 | 2.37E-23 | 128777 | 2 |
| 512.8 | Cough | respiratory | DR | 1.13 | 1.12 | 1.15 | 4.49E-23 | 128777 | 2 |
| 761 | Cervicalgia | symptoms | DR | 0.87 | 0.86 | 0.88 | 5.59E-23 | 128777 | 2 |
| 281.12 | Other vitamin B12 deficiency anemia | hematopoietic | DR | 1.29 | 1.26 | 1.33 | 5.90E-23 | 128777 | 2 |
| 41.9 | Infection with drug-resistant microorganisms | infectious diseases | DR | 1.55 | 1.48 | 1.62 | 6.28E-23 | 128777 | 2 |
| 626 | Disorders of menstruation and other abnormal bleeding from female genital tract | genitourinary | DR | 0.53 | 0.49 | 0.56 | 7.47E-23 | 128777 | 2 |
| 260.2 | severe protein-calorie malnutrition | endocrine/metabolic | DR | 1.57 | 1.50 | 1.64 | 7.58E-23 | 128777 | 2 |
| 342 | Hemiplegia | neurological | DR | 1.49 | 1.43 | 1.56 | 9.04E-23 | 128777 | 2 |
| 70 | Viral hepatitis | infectious diseases | DR | 0.76 | 0.74 | 0.78 | 9.48E-23 | 128777 | 2 |
| 740.9 | Osteoarthrosis NOS | musculoskeletal | DR | 1.13 | 1.12 | 1.15 | 9.88E-23 | 128777 | 2 |
| 807 | Fracture of ribs | injuries & poisonings | DR | 1.31 | 1.28 | 1.35 | 1.16E-22 | 128777 | 2 |
| 270.38 | Other specified disorders of plasma protein metabolism | endocrine/metabolic | DR | 1.73 | 1.64 | 1.83 | 1.29E-22 | 128777 | 2 |
| 593 | Hematuria | genitourinary | DR | 1.17 | 1.15 | 1.19 | 1.31E-22 | 128777 | 2 |
| 272.13 | Mixed hyperlipidemia | endocrine/metabolic | DR | 1.13 | 1.11 | 1.14 | 1.87E-22 | 128777 | 2 |
| 1010 | Other tests | | DR | 1.37 | 1.33 | 1.41 | 1.88E-22 | 128777 | 2 |
| 962 | Poisoning by hormones and synthetic substitutes | injuries & poisonings | DR | 1.44 | 1.39 | 1.49 | 2.07E-22 | 128777 | 2 |
| 626.1 | Irregular menstrual cycle/bleeding | genitourinary | DR | 0.50 | 0.47 | 0.54 | 4.85E-22 | 128777 | 2 |
| 735.3 | Hallux valgus (Bunion) | musculoskeletal | DR | 1.24 | 1.22 | 1.27 | 5.01E-22 | 128777 | 2 |
| 8 | Intestinal infection | infectious diseases | DR | 1.25 | 1.22 | 1.28 | 5.32E-22 | 128777 | 2 |
| 284 | Aplastic anemia | hematopoietic | DR | 1.40 | 1.35 | 1.45 | 1.14E-21 | 128777 | 2 |
| 796 | Elevated prostate specific antigen [PSA] | genitourinary | DR | 1.18 | 1.16 | 1.20 | 1.34E-21 | 128777 | 2 |
| 364.2 | Corneal edema | sense organs | DR | 1.68 | 1.59 | 1.77 | 1.41E-21 | 128777 | 2 |
| 598.9 | Other nonspecific findings on examination of urine | genitourinary | DR | 1.28 | 1.25 | 1.32 | 1.56E-21 | 128777 | 2 |
| 876 | | | DR | 1.57 | 1.49 | 1.64 | 2.15E-21 | 128777 | 2 |
| 70.3 | Viral hepatitis C | infectious diseases | DR | 0.76 | 0.74 | 0.78 | 2.99E-21 | 128777 | 2 |
| 817 | Concussion | injuries & poisonings | DR | 0.65 | 0.63 | 0.68 | 3.07E-21 | 128777 | 2 |
| 375 | Disorders of lacrimal system | sense organs | DR | 1.56 | 1.49 | 1.63 | 3.68E-21 | 128777 | 2 |
| 604 | Disorders of penis | genitourinary | DR | 1.32 | 1.29 | 1.36 | 3.98E-21 | 128777 | 2 |
| 427.4 | | | DR | 1.45 | 1.39 | 1.50 | 5.03E-21 | 111475 | 1 |
| 727.1 | Synovitis and tenosynovitis | musculoskeletal | DR | 1.19 | 1.17 | 1.21 | 6.02E-21 | 128777 | 2 |
| 189.2 | Cancer of bladder | neoplasms | DR | 1.35 | 1.31 | 1.40 | 6.07E-21 | 128777 | 2 |
| 474 | Acute and chronic tonsillitis | respiratory | DR | 0.60 | 0.57 | 0.64 | 1.08E-20 | 111475 | 1 |
| 270.32 | Paraproteinemia | endocrine/metabolic | DR | 1.55 | 1.48 | 1.63 | 1.78E-20 | 128777 | 2 |
| 695.9 | Unspecified erythematous condition | dermatologic | DR | 1.47 | 1.41 | 1.53 | 1.95E-20 | 128777 | 2 |
| 275.51 | Hypocalcemia | endocrine/metabolic | DR | 1.50 | 1.44 | 1.57 | 2.60E-20 | 128777 | 2 |
| 216.1 | | | DR | 1.23 | 1.20 | 1.25 | 3.79E-20 | 111475 | 1 |
| 769 | Nonallopathic lesions NEC | symptoms | DR | 0.75 | 0.72 | 0.77 | 3.81E-20 | 128777 | 2 |
| 601.4 | Balanoposthitis | genitourinary | DR | 1.46 | 1.40 | 1.52 | 4.17E-20 | 128777 | 2 |
| 572 | Ascites (non malignant) | digestive | DR | 1.36 | 1.32 | 1.41 | 4.30E-20 | 128777 | 2 |
| 722.1 | Displacement of intervertebral disc | musculoskeletal | DR | 0.83 | 0.81 | 0.84 | 4.75E-20 | 128777 | 2 |
| 368.9 | Subjective visual disturbances | sense organs | DR | 1.22 | 1.19 | 1.24 | 6.25E-20 | 128777 | 2 |
| 81 | Infection/inflammation of internal prosthetic device; implant; and graft | infectious diseases | DR | 1.43 | 1.37 | 1.49 | 6.26E-20 | 128777 | 2 |
| 510 | Other diseases of lung | respiratory | DR | 1.18 | 1.16 | 1.20 | 6.95E-20 | 128777 | 2 |
| 747 | Cardiac and circulatory congenital anomalies | congenital anomalies | DR | 1.36 | 1.31 | 1.41 | 7.00E-20 | 128777 | 2 |
| 418.1 | Precordial pain | circulatory system | DR | 1.30 | 1.27 | 1.34 | 7.14E-20 | 128777 | 2 |
| 257 | Testicular dysfunction | endocrine/metabolic | DR | 0.83 | 0.81 | 0.85 | 7.21E-20 | 128777 | 2 |
| 598 | Abnormal findings on examination of urine | genitourinary | DR | 1.28 | 1.25 | 1.31 | 7.23E-20 | 128777 | 2 |
| 972.6 | Antihypertensive agents causing adverse effects | injuries & poisonings | DR | 1.56 | 1.48 | 1.63 | 9.84E-20 | 128777 | 2 |
| 790 | Nonspecific findings on examination of blood | symptoms | DR | 1.44 | 1.38 | 1.50 | 1.13E-19 | 128777 | 2 |
| 590 | Pyelonephritis | genitourinary | DR | 1.39 | 1.34 | 1.44 | 1.85E-19 | 128777 | 2 |
| 370.2 | Superficial keratitis | sense organs | DR | 1.46 | 1.40 | 1.52 | 1.89E-19 | 128777 | 2 |
| 592 | | | DR | 1.29 | 1.25 | 1.33 | 2.03E-19 | 111475 | 1 |
| 185 | Cancer of prostate | neoplasms | DR | 1.21 | 1.18 | 1.23 | 2.14E-19 | 128777 | 2 |
| 189 | Cancer of urinary organs (incl. kidney and bladder) | neoplasms | DR | 1.27 | 1.24 | 1.31 | 3.14E-19 | 111475 | 1 |
| 261.4 | Vitamin D deficiency | endocrine/metabolic | DR | 1.13 | 1.11 | 1.14 | 4.09E-19 | 128777 | 2 |
| 153 | Colorectal cancer | neoplasms | DR | 1.33 | 1.29 | 1.38 | 4.36E-19 | 111475 | 1 |
| 284.1 | Pancytopenia | hematopoietic | DR | 1.38 | 1.33 | 1.43 | 5.09E-19 | 128777 | 2 |
| 760 | Back pain | symptoms | DR | 0.89 | 0.88 | 0.90 | 6.44E-19 | 128777 | 2 |
| 579.8 | Nonspecific abnormal findings in stool contents | digestive | DR | 1.21 | 1.19 | 1.24 | 7.83E-19 | 128777 | 2 |
| 272.9 | Unspecified disorder of lipoid metabolism | endocrine/metabolic | DR | 1.37 | 1.33 | 1.42 | 7.89E-19 | 128777 | 2 |
| 773 | Pain in limb | symptoms | DR | 1.12 | 1.10 | 1.13 | 7.94E-19 | 128777 | 2 |
| 586.2 | Cyst of kidney, acquired | genitourinary | DR | 1.23 | 1.20 | 1.26 | 8.41E-19 | 128777 | 2 |
| 599.9 | Other abnormality of urination | genitourinary | DR | 1.19 | 1.17 | 1.21 | 8.64E-19 | 128777 | 2 |
| 153.2 | Colon cancer | neoplasms | DR | 1.34 | 1.29 | 1.38 | 8.73E-19 | 128777 | 2 |
| 509.2 | Respiratory insufficiency | respiratory | DR | 1.41 | 1.36 | 1.47 | 1.55E-18 | 128777 | 2 |
| 626.2 | Dysmenorrhea | genitourinary | DR | 0.30 | 0.26 | 0.35 | 2.59E-18 | 128777 | 2 |
| 771.2 | | | DR | 1.30 | 1.26 | 1.34 | 2.81E-18 | 128777 | 2 |
| 257.1 | Testicular hypofunction | endocrine/metabolic | DR | 0.84 | 0.82 | 0.85 | 2.91E-18 | 128777 | 2 |
| 303.4 | Somatoform disorder | mental disorders | DR | 0.75 | 0.73 | 0.78 | 2.91E-18 | 128777 | 2 |
| 695.8 | Other specified erythematous conditions | dermatologic | DR | 1.27 | 1.23 | 1.30 | 3.65E-18 | 128777 | 2 |
| 332 | Parkinson's disease | neurological | DR | 1.36 | 1.31 | 1.41 | 3.74E-18 | 128777 | 2 |
| 960.1 | Adverse effects of antibacterials (not penicillins) | injuries & poisonings | DR | 1.57 | 1.49 | 1.65 | 3.81E-18 | 128777 | 2 |
| 279.1 | Immunity deficiency | endocrine/metabolic | DR | 1.48 | 1.41 | 1.54 | 4.20E-18 | 128777 | 2 |
| 360 | Disorders of the globe | sense organs | DR | 1.64 | 1.55 | 1.74 | 4.22E-18 | 128777 | 2 |
| 599.1 | Urinary obstruction | genitourinary | DR | 1.38 | 1.33 | 1.43 | 4.34E-18 | 128777 | 2 |
| 464 | Acute sinusitis | respiratory | DR | 0.87 | 0.86 | 0.88 | 4.38E-18 | 128777 | 2 |
| 711.1 | Pyogenic arthritis | musculoskeletal | DR | 1.52 | 1.45 | 1.60 | 4.76E-18 | 128777 | 2 |
| 480.11 | Pneumococcal pneumonia | respiratory | DR | 1.39 | 1.34 | 1.45 | 6.04E-18 | 128777 | 2 |
| 379.4 | Anomalies of pupillary function | sense organs | DR | 1.53 | 1.46 | 1.61 | 6.16E-18 | 128777 | 2 |
| 290.2 | Delirium due to conditions classified elsewhere | mental disorders | DR | 1.31 | 1.27 | 1.35 | 7.50E-18 | 128777 | 2 |
| 112.3 | Candidiasis of skin and nails | infectious diseases | DR | 1.47 | 1.41 | 1.54 | 7.71E-18 | 128777 | 2 |
| 302.1 | Decreased libido | mental disorders | DR | 0.63 | 0.60 | 0.67 | 7.98E-18 | 128777 | 2 |
| 189.21 | Malignant neoplasm of bladder | neoplasms | DR | 1.33 | 1.29 | 1.38 | 1.20E-17 | 128777 | 2 |
| 369 | Infection of the eye | sense organs | DR | 1.18 | 1.16 | 1.21 | 1.51E-17 | 128777 | 2 |
| 681.7 | Cellulitis and abscess of trunk | dermatologic | DR | 1.23 | 1.20 | 1.26 | 1.55E-17 | 128777 | 2 |
| 626.12 | Excessive or frequent menstruation | genitourinary | DR | 0.45 | 0.41 | 0.50 | 2.89E-17 | 128777 | 2 |
| 303 | Psychogenic and somatoform disorders | mental disorders | DR | 0.78 | 0.75 | 0.80 | 2.98E-17 | 111475 | 1 |
| 276.14 | Hypopotassemia | endocrine/metabolic | DR | 1.16 | 1.14 | 1.18 | 3.13E-17 | 128777 | 2 |
| 375.2 | Epiphora | sense organs | DR | 1.55 | 1.47 | 1.63 | 4.69E-17 | 128777 | 2 |
| 394.2 | Mitral valve disease | circulatory system | DR | 1.76 | 1.65 | 1.88 | 5.31E-17 | 128777 | 2 |
| 757 | Congenital anomalies of the integument | congenital anomalies | DR | 1.61 | 1.52 | 1.70 | 6.22E-17 | 111475 | 1 |
| 747.1 | Cardiac congenital anomalies | congenital anomalies | DR | 1.36 | 1.31 | 1.41 | 6.61E-17 | 128777 | 2 |

| | | | | | | | | | |
|---|---|---|---|---|---|---|---|---|---|
| 609.1 | Infertility, male | genitourinary | DR | 0.35 | 0.31 | 0.40 | 6.84E-17 | 111475 | 1 |
| 338 | | | DR | 0.89 | 0.88 | 0.90 | 7.17E-17 | 111475 | 1 |
| 536 | Disorders of function of stomach | digestive | DR | 1.20 | 1.18 | 1.23 | 7.66E-17 | 111475 | 1 |
| 979 | Adverse drug events and drug allergies | injuries & poisonings | DR | 1.18 | 1.16 | 1.21 | 8.18E-17 | 128777 | 2 |
| 288 | Diseases of white blood cells | hematopoietic | DR | 1.16 | 1.14 | 1.18 | 8.90E-17 | 128777 | 2 |
| 599.8 | Other symptoms involving urinary system | genitourinary | DR | 1.22 | 1.20 | 1.26 | 1.07E-16 | 128777 | 2 |
| 260.6 | Anorexia | endocrine/metabolic | DR | 1.43 | 1.37 | 1.49 | 1.51E-16 | 128777 | 2 |
| 452.2 | Deep vein thrombosis [DVT] | circulatory system | DR | 1.23 | 1.20 | 1.26 | 1.60E-16 | 128777 | 2 |
| 509.3 | Pulmonary insufficiency or respiratory failure following trauma and surgery | respiratory | DR | 1.38 | 1.33 | 1.43 | 1.84E-16 | 128777 | 2 |
| 295 | Schizophrenia and other psychotic disorders | mental disorders | DR | 0.81 | 0.79 | 0.83 | 1.87E-16 | 128777 | 2 |
| 317.11 | Alcoholic liver damage | mental disorders | DR | 0.75 | 0.72 | 0.78 | 1.92E-16 | 128777 | 2 |
| 859 | Complication due to other implant and internal device | injuries & poisonings | DR | 1.37 | 1.32 | 1.42 | 2.51E-16 | 128777 | 2 |
| 474.2 | Chronic tonsillitis and adenoiditis | respiratory | DR | 0.56 | 0.53 | 0.60 | 2.65E-16 | 128777 | 2 |
| 495.2 | Asthma with exacerbation | respiratory | DR | 0.72 | 0.69 | 0.75 | 4.12E-16 | 128777 | 2 |
| 698 | Pruritus and related conditions | dermatologic | DR | 1.19 | 1.17 | 1.22 | 4.13E-16 | 128777 | 2 |
| 425.2 | Secondary/extrinsic cardiomyopathies | circulatory system | DR | 1.41 | 1.35 | 1.47 | 6.13E-16 | 128777 | 2 |
| 289.8 | Polycythemia, secondary | hematopoietic | DR | 0.67 | 0.64 | 0.70 | 7.98E-16 | 128777 | 2 |
| 525.1 | Loss of teeth or edentulism | digestive | DR | 1.13 | 1.12 | 1.15 | 9.14E-16 | 128777 | 2 |
| 712 | Infective connective tissue disorders | musculoskeletal | DR | 1.85 | 1.71 | 2.00 | 9.56E-16 | 128777 | 2 |
| 597.2 | Urinary complications NEC | genitourinary | DR | 1.50 | 1.42 | 1.57 | 1.33E-15 | 128777 | 2 |
| 274.11 | Gouty arthropathy | endocrine/metabolic | DR | 1.24 | 1.20 | 1.27 | 1.55E-15 | 128777 | 2 |
| 250.2 | Type 2 diabetes | endocrine/metabolic | DR | 1.62 | 1.52 | 1.72 | 1.61E-15 | 128777 | 2 |
| 426.22 | | | DR | 2.33 | 2.10 | 2.59 | 1.67E-15 | 128777 | 2 |
| 599.6 | Oliguria and anuria | genitourinary | DR | 1.81 | 1.68 | 1.95 | 1.69E-15 | 128777 | 2 |
| 430 | Intracranial hemorrhage | circulatory system | DR | 1.42 | 1.36 | 1.48 | 1.91E-15 | 128777 | 2 |
| 71.1 | HIV infection, symptomatic | infectious diseases | DR | 0.52 | 0.48 | 0.56 | 2.24E-15 | 128777 | 2 |
| 531 | Peptic ulcer (excl. esophageal) | digestive | DR | 1.20 | 1.17 | 1.23 | 2.39E-15 | 128777 | 2 |
| 724 | | | DR | 0.83 | 0.82 | 0.85 | 3.48E-15 | 128777 | 2 |
| 519 | Other diseases of respiratory system, not elsewhere classified | respiratory | DR | 1.16 | 1.14 | 1.18 | 3.52E-15 | 128777 | 2 |
| 949.1 | Diaper or napkin rash | injuries & poisonings | DR | 2.08 | 1.90 | 2.29 | 3.52E-15 | 111475 | 1 |
| 687.3 | Changes in skin texture | dermatologic | DR | 1.57 | 1.48 | 1.66 | 3.64E-15 | 128777 | 2 |
| 722.6 | Degeneration of intervertebral disc | musculoskeletal | DR | 0.89 | 0.88 | 0.91 | 4.90E-15 | 128777 | 2 |
| 370.31 | Keratoconjunctivitis sicca | sense organs | DR | 1.27 | 1.24 | 1.31 | 5.78E-15 | 128777 | 2 |
| 939 | Atopic/contact dermatitis due to other or unspecified | dermatologic | DR | 1.11 | 1.10 | 1.13 | 6.09E-15 | 128777 | 2 |
| 724.8 | Other symptoms referable to back | musculoskeletal | DR | 0.78 | 0.75 | 0.80 | 6.21E-15 | 128777 | 2 |
| 974 | Poisoning by water, mineral, and uric acid metabolism drugs | injuries & poisonings | DR | 1.87 | 1.72 | 2.02 | 6.32E-15 | 128777 | 2 |
| 302 | Sexual and gender identity disorders | mental disorders | DR | 0.75 | 0.72 | 0.78 | 6.61E-15 | 128777 | 2 |
| 1010.1 | screening for infectious and parasitic diseases | | DR | 0.86 | 0.84 | 0.88 | 6.82E-15 | 128777 | 2 |
| 711 | Arthropathy associated with infections | musculoskeletal | DR | 1.38 | 1.32 | 1.43 | 7.29E-15 | 128777 | 2 |
| 722 | | | DR | 0.90 | 0.88 | 0.91 | 8.39E-15 | 128777 | 2 |
| 960 | Poisoning by antibiotics | injuries & poisonings | DR | 1.31 | 1.27 | 1.36 | 1.17E-14 | 128777 | 2 |
| 969 | Poisoning by psychotropic agents | injuries & poisonings | DR | 0.68 | 0.65 | 0.72 | 1.18E-14 | 128777 | 2 |
| 550.4 | Umbilical hernia | digestive | DR | 0.82 | 0.80 | 0.84 | 1.43E-14 | 128777 | 2 |
| 840.3 | Joint/ligament sprain | injuries & poisonings | DR | 0.83 | 0.81 | 0.85 | 1.44E-14 | 128777 | 2 |
| 290.3 | Other persistent mental disorders due to conditions classified elsewhere | mental disorders | DR | 1.20 | 1.17 | 1.23 | 1.79E-14 | 128777 | 2 |
| 306 | Other mental disorder | mental disorders | DR | 0.88 | 0.87 | 0.90 | 2.56E-14 | 128777 | 2 |
| 290.12 | Dementia with cerebral degenerations | mental disorders | DR | 1.62 | 1.52 | 1.73 | 3.30E-14 | 128777 | 2 |
| 735.22 | Claw toe (acquired) | musculoskeletal | DR | 1.88 | 1.73 | 2.04 | 4.49E-14 | 128777 | 2 |
| 592.11 | Acute cystitis | genitourinary | DR | 1.32 | 1.27 | 1.37 | 4.55E-14 | 128777 | 2 |
| 513.3 | Hypoventilation | respiratory | DR | 1.23 | 1.20 | 1.26 | 5.41E-14 | 128777 | 2 |
| 275.6 | Hypercalcemia | endocrine/metabolic | DR | 1.27 | 1.23 | 1.31 | 5.58E-14 | 128777 | 2 |
| 430.2 | Intracerebral hemorrhage | circulatory system | DR | 1.56 | 1.47 | 1.66 | 8.94E-14 | 128777 | 2 |
| 962.3 | Hormones and synthetic substitutes causing adverse effects in therapeutic use | injuries & poisonings | DR | 1.45 | 1.38 | 1.52 | 1.06E-13 | 128777 | 2 |
| 496 | Chronic airway obstruction | respiratory | DR | 1.10 | 1.09 | 1.12 | 1.26E-13 | 128777 | 2 |
| 910 | | | DR | 1.49 | 1.41 | 1.58 | 1.27E-13 | 128777 | 2 |
| 208 | Benign neoplasm of colon | neoplasms | DR | 1.10 | 1.08 | 1.11 | 1.58E-13 | 128777 | 2 |
| 256.4 | Polycystic ovaries | endocrine/metabolic | DR | 0.34 | 0.30 | 0.40 | 1.62E-13 | 128777 | 2 |
| 772.6 | Facial weakness | symptoms | DR | 1.43 | 1.37 | 1.51 | 1.67E-13 | 128777 | 2 |
| 722.8 | Postlaminectomy syndrome | musculoskeletal | DR | 0.74 | 0.71 | 0.77 | 1.81E-13 | 128777 | 2 |
| 580.4 | Renal sclerosis, NOS | genitourinary | DR | 1.76 | 1.63 | 1.90 | 1.95E-13 | 128777 | 2 |
| 378.1 | Strabismus (not specified as paralytic) | sense organs | DR | 1.22 | 1.19 | 1.26 | 2.45E-13 | 128777 | 2 |
| 364.51 | Fuchs' dystrophy | sense organs | DR | 1.39 | 1.33 | 1.45 | 2.51E-13 | 128777 | 2 |
| 537.1 | Lesions of stomach and duodenum | digestive | DR | 1.61 | 1.51 | 1.72 | 2.71E-13 | 128777 | 2 |
| 117 | Mycoses | infectious diseases | DR | 1.34 | 1.28 | 1.39 | 3.96E-13 | 128777 | 2 |
| 500.2 | Pneumoconiosis | respiratory | DR | 1.57 | 1.48 | 1.67 | 5.01E-13 | 111475 | 1 |
| 770 | Myalgia and myositis unspecified | symptoms | DR | 0.88 | 0.86 | 0.89 | 6.03E-13 | 128777 | 2 |
| 771 | Musculoskeletal symptoms referable to limbs | symptoms | DR | 1.33 | 1.28 | 1.39 | 6.58E-13 | 128777 | 2 |
| 136 | Other infectious and parasitic diseases | infectious diseases | DR | 1.23 | 1.19 | 1.27 | 1.54E-12 | 128777 | 2 |
| 727 | Other disorders of synovium, tendon, and bursa | musculoskeletal | DR | 1.11 | 1.09 | 1.12 | 2.42E-12 | 128777 | 2 |
| 253.4 | | | DR | 0.72 | 0.68 | 0.75 | 3.19E-12 | 128777 | 2 |
| 580.1 | | | DR | 1.62 | 1.51 | 1.74 | 4.22E-12 | 111475 | 1 |
| 513.8 | Disorders of diaphragm | respiratory | DR | 1.42 | 1.35 | 1.50 | 5.19E-12 | 128777 | 2 |
| 476 | Allergic rhinitis | respiratory | DR | 0.91 | 0.90 | 0.92 | 5.55E-12 | 128777 | 2 |
| 694 | | | DR | 1.16 | 1.13 | 1.18 | 5.77E-12 | 111475 | 1 |
| 300.4 | Dysthymic disorder | mental disorders | DR | 0.88 | 0.86 | 0.90 | 6.72E-12 | 128777 | 2 |
| 379.9 | Pain, swelling or discharge of eye | sense organs | DR | 1.20 | 1.17 | 1.23 | 6.96E-12 | 128777 | 2 |
| 71 | Human immunodeficiency virus [HIV] disease | infectious diseases | DR | 0.58 | 0.54 | 0.63 | 9.34E-12 | 128777 | 2 |
| 805 | Fracture of vertebral column without mention of spinal cord injury | injuries & poisonings | DR | 1.24 | 1.21 | 1.29 | 9.95E-12 | 128777 | 2 |
| 363.4 | Choroidal degenerations | sense organs | DR | 1.99 | 1.80 | 2.20 | 1.02E-11 | 111475 | 1 |
| 395.4 | Nonrheumatic pulmonary valve disorders | circulatory system | DR | 1.60 | 1.49 | 1.71 | 1.15E-11 | 128777 | 2 |
| 262 | Mineral deficiency NEC | endocrine/metabolic | DR | 1.31 | 1.26 | 1.36 | 1.33E-11 | 128777 | 2 |
| 274.2 | Crystal arthropathies | endocrine/metabolic | DR | 1.36 | 1.30 | 1.42 | 1.37E-11 | 128777 | 2 |
| 593.1 | Gross hematuria | genitourinary | DR | 1.17 | 1.14 | 1.20 | 1.39E-11 | 128777 | 2 |
| 349 | Other and unspecified disorders of the nervous system | neurological | DR | 1.26 | 1.22 | 1.31 | 1.46E-11 | 128777 | 2 |
| 426.8 | Other cardiac conduction disorders | circulatory system | DR | 1.38 | 1.32 | 1.45 | 1.47E-11 | 128777 | 2 |
| 803.1 | Fracture of humerus | injuries & poisonings | DR | 1.34 | 1.29 | 1.40 | 1.56E-11 | 128777 | 2 |
| 531.1 | Hemorrhage from gastrointestinal ulcer | digestive | DR | 1.38 | 1.31 | 1.45 | 1.82E-11 | 128777 | 2 |
| 609 | | | DR | 0.68 | 0.64 | 0.72 | 2.08E-11 | 111475 | 1 |
| 386 | Vertiginous syndromes and other disorders of vestibular system | sense organs | DR | 1.16 | 1.13 | 1.18 | 2.37E-11 | 128777 | 2 |
| 172.11 | Melanomas of skin | neoplasms | DR | 1.24 | 1.20 | 1.29 | 2.84E-11 | 128777 | 2 |
| 374.2 | Lagophthalmos | sense organs | DR | 1.57 | 1.47 | 1.69 | 3.11E-11 | 128777 | 2 |
| 626.13 | Irregular menstrual cycle | genitourinary | DR | 0.55 | 0.50 | 0.60 | 3.16E-11 | 128777 | 2 |
| 1000 | Burns | NULL | DR | 1.23 | 1.20 | 1.27 | 3.66E-11 | 128777 | 2 |
| 626.11 | Absent or infrequent menstruation | genitourinary | DR | 0.45 | 0.40 | 0.51 | 4.14E-11 | 128777 | 2 |

| | | | | | | | | | |
|---|---|---|---|---|---|---|---|---|---|
| 312 | Conduct disorders | mental disorders | DR | 0.76 | 0.73 | 0.79 | 5.60E-11 | 128777 | 2 |
| 361.2 | Retinoschisis and retinal cysts | sense organs | DR | 1.54 | 1.44 | 1.64 | 6.45E-11 | 128777 | 2 |
| 90 | Sexually transmitted infections (not HIV or hepatitis) | infectious diseases | DR | 1.34 | 1.28 | 1.40 | 6.53E-11 | 128777 | 2 |
| 8.6 | Viral Enteritis | infectious diseases | DR | 1.27 | 1.23 | 1.32 | 7.19E-11 | 128777 | 2 |
| 830 | Dislocation | injuries & poisonings | DR | 0.85 | 0.83 | 0.87 | 7.64E-11 | 128777 | 2 |
| 291.8 | Alteration of consciousness | mental disorders | DR | 1.21 | 1.18 | 1.25 | 8.02E-11 | 128777 | 2 |
| 556.11 | Angiodysplasia of intestine (without mention of hemorrhage) | digestive | DR | 1.50 | 1.41 | 1.59 | 1.07E-10 | 128777 | 2 |
| 938 | Dermatitis due to solar radiation | dermatologic | DR | 1.16 | 1.14 | 1.19 | 1.27E-10 | 128777 | 2 |
| 561.2 | Flatulence | digestive | DR | 1.18 | 1.15 | 1.22 | 1.42E-10 | 128777 | 2 |
| 681.3 | Cellulitis and abscess of arm/hand | dermatologic | DR | 1.18 | 1.15 | 1.22 | 1.43E-10 | 128777 | 2 |
| 199 | Neoplasm of uncertain behavior | neoplasms | DR | 1.12 | 1.10 | 1.14 | 2.42E-10 | 128777 | 2 |
| 592.12 | Chronic cystitis | genitourinary | DR | 1.50 | 1.41 | 1.61 | 2.62E-10 | 128777 | 2 |
| 790.6 | Other abnormal blood chemistry | symptoms | DR | 1.10 | 1.08 | 1.11 | 2.65E-10 | 128777 | 2 |
| 312.3 | Impulse control disorder | mental disorders | DR | 0.73 | 0.70 | 0.77 | 2.83E-10 | 128777 | 2 |
| 474.1 | Acute tonsillitis | respiratory | DR | 0.62 | 0.57 | 0.67 | 3.36E-10 | 128777 | 2 |
| 190 | Cancer of eye | neoplasms | DR | 1.63 | 1.50 | 1.76 | 3.49E-10 | 128777 | 2 |
| 394.3 | Aortic valve disease | circulatory system | DR | 1.92 | 1.73 | 2.13 | 3.59E-10 | 128777 | 2 |
| 1001 | Foreign body injury | NULL | DR | 1.17 | 1.14 | 1.20 | 3.76E-10 | 128777 | 2 |
| 228 | Hemangioma and lymphangioma, any site | neoplasms | DR | 1.16 | 1.13 | 1.19 | 3.79E-10 | 128777 | 2 |
| 256 | Ovarian dysfunction | endocrine/metabolic | DR | 0.40 | 0.34 | 0.46 | 3.84E-10 | 128777 | 2 |
| 960.2 | Allergy/adverse effect of penicillin | injuries & poisonings | DR | 1.29 | 1.24 | 1.35 | 3.93E-10 | 128777 | 2 |
| 1010.3 | screening for other diseases and disorders | | DR | 0.92 | 0.90 | 0.93 | 4.46E-10 | 128777 | 2 |
| 278 | Overweight, obesity and other hyperalimentation | endocrine/metabolic | DR | 0.91 | 0.90 | 0.92 | 4.51E-10 | 128777 | 2 |
| 856 | Vascular complications of surgery and medical procedures | injuries & poisonings | DR | 1.65 | 1.53 | 1.79 | 5.04E-10 | 128777 | 2 |
| 716.9 | Arthropathy NOS | musculoskeletal | DR | 1.10 | 1.09 | 1.12 | 5.24E-10 | 128777 | 2 |
| 480.2 | Viral pneumonia | respiratory | DR | 1.40 | 1.33 | 1.48 | 5.35E-10 | 128777 | 2 |
| 560 | | | DR | 1.18 | 1.15 | 1.21 | 5.70E-10 | 111475 | 1 |
| 573.6 | Nonspecific elevation of levels of transaminase or lactic acid dehydrogenase [LDH] | digestive | DR | 0.84 | 0.81 | 0.86 | 5.72E-10 | 128777 | 2 |
| 625 | Pain and other symptoms associated with female genital organs | genitourinary | DR | 0.62 | 0.58 | 0.67 | 6.16E-10 | 128777 | 2 |
| 578.8 | Hemorrhage of rectum and anus | digestive | DR | 0.88 | 0.86 | 0.89 | 7.23E-10 | 128777 | 2 |
| 695.7 | Prurigo and Lichen | dermatologic | DR | 1.18 | 1.15 | 1.21 | 7.66E-10 | 128777 | 2 |
| 706.1 | Acne | dermatologic | DR | 0.78 | 0.75 | 0.81 | 8.35E-10 | 128777 | 2 |
| 327.1 | Hypersomnia | neurological | DR | 0.82 | 0.80 | 0.85 | 8.88E-10 | 128777 | 2 |
| 772.1 | Muscular wasting and disuse atrophy | symptoms | DR | 1.39 | 1.32 | 1.47 | 9.57E-10 | 128777 | 2 |
| 702.4 | Degenerative skin disorders | dermatologic | DR | 1.89 | 1.70 | 2.09 | 9.81E-10 | 111475 | 1 |
| 1005 | Other symptoms | NULL | DR | 1.08 | 1.07 | 1.10 | 1.09E-09 | 128777 | 2 |
| 733 | Other disorders of bone and cartilage | musculoskeletal | DR | 1.15 | 1.13 | 1.18 | 1.12E-09 | 128777 | 2 |
| 742 | Derangement of joint, non-traumatic | musculoskeletal | DR | 0.86 | 0.83 | 0.88 | 1.45E-09 | 111475 | 1 |
| 389.4 | Tinnitus | sense organs | DR | 0.92 | 0.91 | 0.93 | 1.52E-09 | 128777 | 2 |
| 574.3 | Cholecystitis without cholelithiasis | digestive | DR | 1.21 | 1.17 | 1.25 | 1.55E-09 | 128777 | 2 |
| 519.8 | Other diseases of respiratory system, NEC | respiratory | DR | 1.18 | 1.15 | 1.22 | 1.64E-09 | 128777 | 2 |
| 333.1 | Essential tremor | neurological | DR | 1.16 | 1.13 | 1.19 | 1.82E-09 | 128777 | 2 |
| 446.5 | Giant cell arteritis | circulatory system | DR | 1.66 | 1.53 | 1.81 | 2.07E-09 | 128777 | 2 |
| 994.1 | Systemic inflammatory response syndrome (SIRS) | injuries & poisonings | DR | 1.30 | 1.25 | 1.36 | 2.13E-09 | 128777 | 2 |
| 580.14 | Chronic glomerulonephritis, NOS | genitourinary | DR | 1.84 | 1.66 | 2.04 | 2.64E-09 | 128777 | 2 |
| 195.3 | Malignant neoplasm of head, face, and neck | neoplasms | DR | 1.29 | 1.24 | 1.35 | 3.48E-09 | 128777 | 2 |
| 204 | Leukemia | neoplasms | DR | 1.29 | 1.23 | 1.35 | 3.50E-09 | 128777 | 2 |
| 459.1 | Hemorrhage NOS | circulatory system | DR | 1.37 | 1.30 | 1.44 | 3.74E-09 | 128777 | 2 |
| 427.11 | Paroxysmal supraventricular tachycardia | circulatory system | DR | 1.19 | 1.16 | 1.23 | 4.82E-09 | 128777 | 2 |
| 580.13 | Acute glomerulonephritis, NOS | genitourinary | DR | 2.03 | 1.80 | 2.28 | 4.92E-09 | 128777 | 2 |
| 241 | Nontoxic nodular goiter | endocrine/metabolic | DR | 1.17 | 1.14 | 1.21 | 5.79E-09 | 128777 | 2 |
| 211 | Benign neoplasm of other parts of digestive system | neoplasms | DR | 1.19 | 1.16 | 1.23 | 6.33E-09 | 128777 | 2 |
| 556 | Ulceration of the lower GI tract | digestive | DR | 1.30 | 1.25 | 1.37 | 6.95E-09 | 128777 | 2 |
| 780 | Hypothermia/Chills | symptoms | DR | 1.37 | 1.30 | 1.45 | 8.18E-09 | 128777 | 2 |
| 790.1 | Elevated sedimentation rate | symptoms | DR | 1.51 | 1.41 | 1.63 | 1.24E-08 | 128777 | 2 |
| 295.2 | Paranoid disorders | mental disorders | DR | 0.71 | 0.67 | 0.75 | 1.33E-08 | 128777 | 2 |
| 818 | Intracranial hemorrhage (injury) | injuries & poisonings | DR | 1.36 | 1.29 | 1.43 | 1.54E-08 | 128777 | 2 |
| 223 | Benign neoplasm of kidney and other urinary organs | neoplasms | DR | 1.35 | 1.28 | 1.42 | 1.70E-08 | 128777 | 2 |
| 717 | Polymyalgia Rheumatica | musculoskeletal | DR | 1.45 | 1.36 | 1.55 | 1.79E-08 | 128777 | 2 |
| 803 | Fracture of upper limb | injuries & poisonings | DR | 1.18 | 1.14 | 1.21 | 1.79E-08 | 128777 | 2 |
| 441 | Vascular insufficiency of intestine | circulatory system | DR | 1.39 | 1.31 | 1.47 | 1.83E-08 | 128777 | 2 |
| 512.3 | | | DR | 1.63 | 1.49 | 1.78 | 2.03E-08 | 128777 | 2 |
| 359.2 | Myopathy | neurological | DR | 1.32 | 1.26 | 1.39 | 2.53E-08 | 128777 | 2 |
| 281.11 | Pernicious anemia | hematopoietic | DR | 1.47 | 1.37 | 1.58 | 2.73E-08 | 128777 | 2 |
| 981 | Toxic effect of (non-ethyl) alcohol and petroleum and other solvents | injuries & poisonings | DR | 0.35 | 0.29 | 0.43 | 3.05E-08 | 111475 | 1 |
| 386.2 | Peripheral or central vertigo | sense organs | DR | 1.13 | 1.11 | 1.16 | 3.21E-08 | 128777 | 2 |
| 695.2 | Bullous dermatoses | dermatologic | DR | 1.51 | 1.40 | 1.63 | 3.47E-08 | 111475 | 1 |
| 53.1 | Herpes zoster with nervous system complications | infectious diseases | DR | 1.37 | 1.29 | 1.45 | 3.61E-08 | 128777 | 2 |
| 274.21 | Chondrocalcinosis | endocrine/metabolic | DR | 1.30 | 1.24 | 1.37 | 3.64E-08 | 128777 | 2 |
| 292.3 | Memory loss | mental disorders | DR | 1.13 | 1.11 | 1.16 | 3.71E-08 | 128777 | 2 |
| 738 | Other acquired musculoskeletal deformity | musculoskeletal | DR | 0.86 | 0.84 | 0.88 | 3.94E-08 | 128777 | 2 |
| 740.12 | Osteoarthrosis, localized, secondary | musculoskeletal | DR | 0.86 | 0.83 | 0.88 | 3.95E-08 | 128777 | 2 |
| 742.9 | Other derangement of joint | musculoskeletal | DR | 0.86 | 0.84 | 0.89 | 4.54E-08 | 128777 | 2 |
| 586.4 | Stricture/obstruction of ureter | genitourinary | DR | 1.23 | 1.18 | 1.27 | 4.66E-08 | 128777 | 2 |
| 241.2 | Nontoxic multinodular goiter | endocrine/metabolic | DR | 1.22 | 1.18 | 1.27 | 4.91E-08 | 128777 | 2 |
| 513.4 | Hyperventilation | respiratory | DR | 1.41 | 1.33 | 1.50 | 5.24E-08 | 128777 | 2 |
| 724.9 | Other unspecified back disorders | musculoskeletal | DR | 0.82 | 0.79 | 0.85 | 5.39E-08 | 128777 | 2 |
| 840 | Sprains and strains | injuries & poisonings | DR | 0.93 | 0.91 | 0.94 | 6.13E-08 | 128777 | 2 |
| 800.3 | Fracture of tibia and fibula | injuries & poisonings | DR | 1.23 | 1.18 | 1.27 | 6.81E-08 | 128777 | 2 |
| 793 | Nonspecific abnormal findings on radiological and other examination of musculoskeletal system | symptoms | DR | 1.21 | 1.17 | 1.25 | 7.50E-08 | 128777 | 2 |
| 733.9 | | | DR | 0.67 | 0.62 | 0.72 | 7.70E-08 | 111475 | 1 |
| 306.9 | Tension headache | mental disorders | DR | 0.76 | 0.73 | 0.80 | 7.74E-08 | 128777 | 2 |
| 723 | Other disorders of cervical region | musculoskeletal | DR | 0.79 | 0.75 | 0.82 | 7.86E-08 | 128777 | 2 |
| 726.4 | Calcaneal spur; Exostosis NOS | musculoskeletal | DR | 1.12 | 1.10 | 1.14 | 8.75E-08 | 128777 | 2 |
| 442 | Other aneurysm | circulatory system | DR | 1.13 | 1.10 | 1.15 | 9.08E-08 | 128777 | 2 |
| 531.4 | Peptic ulcer, site unspecified | digestive | DR | 1.17 | 1.14 | 1.21 | 9.43E-08 | 128777 | 2 |
| 560.1 | Paralytic ileus | digestive | DR | 1.22 | 1.18 | 1.27 | 9.99E-08 | 128777 | 2 |
| 556.1 | Ulceration of intestine | digestive | DR | 1.29 | 1.23 | 1.36 | 1.06E-07 | 128777 | 2 |
| 327.41 | Organic or persistent insomnia | neurological | DR | 0.83 | 0.80 | 0.86 | 1.11E-07 | 128777 | 2 |
| 502 | Postinflammatory pulmonary fibrosis | respiratory | DR | 1.19 | 1.15 | 1.23 | 1.17E-07 | 128777 | 2 |
| 740.3 | Osteoarthrosis involving more than one site, but not specified as generalized | musculoskeletal | DR | 1.20 | 1.16 | 1.24 | 1.24E-07 | 128777 | 2 |
| 446 | Polyarteritis nodosa and allied conditions | circulatory system | DR | 1.34 | 1.27 | 1.42 | 1.24E-07 | 111475 | 1 |
| 721.1 | Spondylosis without myelopathy | musculoskeletal | DR | 0.93 | 0.92 | 0.94 | 1.35E-07 | 128777 | 2 |
| 537 | Other disorders of stomach and duodenum | digestive | DR | 1.13 | 1.11 | 1.16 | 1.56E-07 | 128777 | 2 |

| | | | | | | | | | |
|---|---|---|---|---|---|---|---|---|---|
| 949 | Allergies, other | injuries & poisonings | DR | 0.88 | 0.86 | 0.91 | 1.64E-07 | 128777 | 2 |
| 578.2 | Blood in stool | digestive | DR | 1.10 | 1.08 | 1.12 | 1.66E-07 | 128777 | 2 |
| 931 | Contact dermatitis and other eczema due to plants [except food] | dermatologic | DR | 0.78 | 0.75 | 0.82 | 1.69E-07 | 128777 | 2 |
| 841 | | | DR | 0.88 | 0.86 | 0.90 | 1.90E-07 | 128777 | 2 |
| 497 | Bronchitis | respiratory | DR | 1.09 | 1.07 | 1.11 | 2.24E-07 | 128777 | 2 |
| 594.3 | Calculus of ureter | genitourinary | DR | 0.86 | 0.83 | 0.89 | 2.32E-07 | 128777 | 2 |
| 687.2 | Localized superficial swelling, mass, or lump | dermatologic | DR | 1.11 | 1.09 | 1.13 | 2.33E-07 | 128777 | 2 |
| 701.5 | Abnormal granulation tissue | dermatologic | DR | 1.35 | 1.27 | 1.43 | 2.79E-07 | 128777 | 2 |
| 687.1 | Rash and other nonspecific skin eruption | dermatologic | DR | 1.07 | 1.06 | 1.09 | 2.99E-07 | 128777 | 2 |
| 244.2 | Acquired hypothyroidism | endocrine/metabolic | DR | 1.20 | 1.16 | 1.24 | 3.47E-07 | 128777 | 2 |
| 275.11 | Hereditary hemochromatosis | hematopoietic | DR | 0.57 | 0.51 | 0.64 | 3.59E-07 | 128777 | 2 |
| 344 | Other paralytic syndromes | neurological | DR | 1.21 | 1.16 | 1.25 | 3.94E-07 | 128777 | 2 |
| 411.41 | Aneurysm and dissection of heart | circulatory system | DR | 1.48 | 1.37 | 1.60 | 4.09E-07 | 128777 | 2 |
| 870.3 | Other open wound of head and face | injuries & poisonings | DR | 1.20 | 1.16 | 1.25 | 4.48E-07 | 128777 | 2 |
| 348.7 | Coma | neurological | DR | 1.50 | 1.39 | 1.63 | 4.81E-07 | 128777 | 2 |
| 756 | Other congenital musculoskeletal anomalies | congenital anomalies | DR | 1.30 | 1.23 | 1.37 | 4.89E-07 | 111475 | 1 |
| 1010.7 | Persons with potential health hazards related to socioeconomic, psychosocial, and other circumstances | | DR | 1.11 | 1.09 | 1.14 | 4.93E-07 | 128777 | 2 |
| 500 | Lung disease due to external agents | respiratory | DR | 1.26 | 1.20 | 1.32 | 5.68E-07 | 128777 | 2 |
| 727.2 | Bursitis disorders | musculoskeletal | DR | 1.15 | 1.12 | 1.18 | 5.70E-07 | 128777 | 2 |
| 701.2 | Scar conditions and fibrosis of skin | dermatologic | DR | 1.14 | 1.11 | 1.17 | 5.80E-07 | 128777 | 2 |
| 753 | Congenital anomalies of the eye | congenital anomalies | DR | 1.29 | 1.22 | 1.36 | 5.91E-07 | 128777 | 2 |
| 599.3 | Dysuria | genitourinary | DR | 1.11 | 1.09 | 1.14 | 6.29E-07 | 128777 | 2 |
| 291.4 | Specific nonpsychotic mental disorders due to brain damage | mental disorders | DR | 0.79 | 0.76 | 0.83 | 6.56E-07 | 128777 | 2 |
| 574.2 | Calculus of bile duct | digestive | DR | 1.23 | 1.18 | 1.29 | 7.42E-07 | 128777 | 2 |
| 253.3 | Diabetes insipidus | endocrine/metabolic | DR | 1.52 | 1.39 | 1.65 | 7.99E-07 | 128777 | 2 |
| 747.12 | Valvular heart disease/ heart chambers | congenital anomalies | DR | 1.43 | 1.33 | 1.54 | 8.70E-07 | 128777 | 2 |
| 694.2 | Other dyschromia | dermatologic | DR | 1.11 | 1.09 | 1.14 | 8.77E-07 | 128777 | 2 |
| 54 | Herpes simplex | infectious diseases | DR | 0.85 | 0.82 | 0.87 | 9.19E-07 | 128777 | 2 |
| 626.8 | Infertility, female | genitourinary | DR | 0.31 | 0.25 | 0.40 | 9.38E-07 | 128777 | 2 |
| 286.5 | Hemorrhagic disorder due to intrinsic circulating anticoagulants | hematopoietic | DR | 1.46 | 1.35 | 1.58 | 9.64E-07 | 128777 | 2 |
| 747.13 | Congenital anomalies of great vessels | congenital anomalies | DR | 1.62 | 1.46 | 1.78 | 1.08E-06 | 128777 | 2 |
| 772.2 | Spasm of muscle | symptoms | DR | 0.90 | 0.88 | 0.92 | 1.08E-06 | 128777 | 2 |
| 597 | Other disorders of urethra and urinary tract | genitourinary | DR | 1.20 | 1.16 | 1.25 | 1.22E-06 | 128777 | 2 |
| 270.33 | Amyloidosis | endocrine/metabolic | DR | 1.84 | 1.62 | 2.08 | 1.31E-06 | 128777 | 2 |
| 716 | Other arthropathies | musculoskeletal | DR | 1.08 | 1.06 | 1.09 | 1.34E-06 | 128777 | 2 |
| 195.1 | Malignant neoplasm, other | neoplasms | DR | 1.16 | 1.13 | 1.20 | 1.45E-06 | 128777 | 2 |
| 368.7 | Disorders of accommodation | sense organs | DR | 0.49 | 0.42 | 0.57 | 1.48E-06 | 111475 | 1 |
| 526.41 | Temporomandibular joint disorder, unspecified | digestive | DR | 0.82 | 0.79 | 0.85 | 1.49E-06 | 128777 | 2 |
| 526.4 | Temporomandibular joint disorders | digestive | DR | 0.84 | 0.81 | 0.87 | 1.49E-06 | 128777 | 2 |
| 947 | Urticaria | dermatologic | DR | 0.85 | 0.83 | 0.88 | 1.52E-06 | 128777 | 2 |
| 715 | Other inflammatory spondylopathies | musculoskeletal | DR | 0.85 | 0.83 | 0.88 | 1.63E-06 | 128777 | 2 |
| 277.7 | Dysmetabolic syndrome X | endocrine/metabolic | DR | 0.85 | 0.83 | 0.88 | 1.69E-06 | 128777 | 2 |
| 442.8 | Aneurysm of other specified artery | circulatory system | DR | 1.40 | 1.31 | 1.51 | 1.71E-06 | 128777 | 2 |
| 1008 | Crushing or internal injury to organs | NULL | DR | 1.22 | 1.17 | 1.27 | 1.75E-06 | 128777 | 2 |
| 478 | Throat pain | respiratory | DR | 0.84 | 0.81 | 0.87 | 1.85E-06 | 128777 | 2 |
| 661 | Fetal distress and abnormal forces of labor | pregnancy complications | DR | 1.64 | 1.48 | 1.82 | 2.11E-06 | 128777 | 2 |
| 287.32 | Secondary thrombocytopenia | hematopoietic | DR | 1.23 | 1.18 | 1.28 | 2.12E-06 | 128777 | 2 |
| 228.1 | Hemangioma of skin and subcutaneous tissue | neoplasms | DR | 1.14 | 1.11 | 1.17 | 2.12E-06 | 128777 | 2 |
| 691 | Congenital anomalies of skin | dermatologic | DR | 1.24 | 1.19 | 1.30 | 2.16E-06 | 128777 | 2 |
| 295.3 | Psychosis | mental disorders | DR | 0.87 | 0.85 | 0.90 | 2.24E-06 | 128777 | 2 |
| 628 | Ovarian cyst | genitourinary | DR | 0.69 | 0.64 | 0.75 | 2.27E-06 | 128777 | 2 |
| 535.2 | Atrophic gastritis | digestive | DR | 1.16 | 1.13 | 1.20 | 2.29E-06 | 128777 | 2 |
| 722.9 | Other and unspecified disc disorder | musculoskeletal | DR | 0.87 | 0.84 | 0.89 | 2.32E-06 | 128777 | 2 |
| 369.2 | Eye infection, viral | sense organs | DR | 1.22 | 1.17 | 1.28 | 2.60E-06 | 128777 | 2 |
| 573.3 | Hepatomegaly | digestive | DR | 0.85 | 0.82 | 0.88 | 2.61E-06 | 128777 | 2 |
| 285.22 | Anemia in neoplastic disease | hematopoietic | DR | 1.27 | 1.20 | 1.33 | 2.79E-06 | 128777 | 2 |
| 727.8 | Plica syndrome | musculoskeletal | DR | 0.53 | 0.46 | 0.61 | 2.90E-06 | 128777 | 2 |
| 346.1 | Nonspecific abnormal findings on radiological and other examination of skull and head | neurological | DR | 1.22 | 1.17 | 1.28 | 3.04E-06 | 128777 | 2 |
| 535 | Gastritis and duodenitis | digestive | DR | 1.08 | 1.07 | 1.10 | 3.23E-06 | 111475 | 1 |
| 598.4 | | | DR | 1.70 | 1.51 | 1.90 | 3.69E-06 | 111475 | 1 |
| 253 | Disorders of the pituitary gland and its hypothalamic control | endocrine/metabolic | DR | 0.85 | 0.83 | 0.88 | 3.72E-06 | 128777 | 2 |
| 427.7 | Tachycardia NOS | circulatory system | DR | 1.09 | 1.07 | 1.11 | 4.28E-06 | 128777 | 2 |
| 743.21 | Pathologic fracture of vertebrae | musculoskeletal | DR | 1.31 | 1.24 | 1.39 | 4.31E-06 | 128777 | 2 |
| 961.1 | Poisoning/allergy of sulfonamides | injuries & poisonings | DR | 1.34 | 1.26 | 1.42 | 4.43E-06 | 128777 | 2 |
| 802 | Fracture of pelvis | injuries & poisonings | DR | 1.40 | 1.30 | 1.50 | 4.50E-06 | 128777 | 2 |
| 427.41 | Ventricular fibrillation and flutter | circulatory system | DR | 1.30 | 1.23 | 1.38 | 4.57E-06 | 128777 | 2 |
| 560.2 | Impaction of intestine | digestive | DR | 1.29 | 1.22 | 1.36 | 4.61E-06 | 128777 | 2 |
| 195 | Cancer, suspected or other | neoplasms | DR | 1.12 | 1.09 | 1.15 | 4.80E-06 | 128777 | 2 |
| 526 | Diseases of the jaws | digestive | DR | 0.89 | 0.87 | 0.91 | 4.85E-06 | 128777 | 2 |
| 562 | | | DR | 1.07 | 1.05 | 1.08 | 4.92E-06 | 111475 | 1 |
| 110.13 | Dermatophytosis of the body | infectious diseases | DR | 1.12 | 1.09 | 1.14 | 5.46E-06 | 128777 | 2 |
| 792 | Abnormal Papanicolaou smear of cervix and cervical HPV | genitourinary | DR | 0.69 | 0.63 | 0.74 | 5.64E-06 | 128777 | 2 |
| 81.12 | Chronic graft-versus-host disease | infectious diseases | DR | 0.25 | 0.18 | 0.34 | 6.57E-06 | 128777 | 2 |
| 573.1 | Chronic passive congestion of liver | digestive | DR | 1.67 | 1.49 | 1.87 | 6.71E-06 | 128777 | 2 |
| 117.1 | Histoplasmosis | infectious diseases | DR | 1.39 | 1.29 | 1.50 | 6.95E-06 | 128777 | 2 |
| 601 | | | DR | 1.09 | 1.07 | 1.11 | 6.97E-06 | 111475 | 1 |
| 240 | Simple and unspecified goiter | endocrine/metabolic | DR | 1.29 | 1.22 | 1.37 | 7.07E-06 | 128777 | 2 |
| 504 | Other alveolar and parietoalveolar pneumonopathy | respiratory | DR | 1.20 | 1.15 | 1.25 | 7.14E-06 | 128777 | 2 |
| 597.1 | Urethral stricture (not specified as infectious) | genitourinary | DR | 1.21 | 1.16 | 1.26 | 7.18E-06 | 128777 | 2 |
| 288.3 | Eosinophilia | hematopoietic | DR | 1.37 | 1.27 | 1.47 | 7.58E-06 | 128777 | 2 |
| 531.3 | Duodenal ulcer | digestive | DR | 1.22 | 1.17 | 1.27 | 7.71E-06 | 128777 | 2 |
| 465.2 | Acute pharyngitis | respiratory | DR | 0.92 | 0.90 | 0.94 | 8.16E-06 | 128777 | 2 |
| 253.1 | Pituitary hyperfunction | endocrine/metabolic | DR | 0.64 | 0.58 | 0.71 | 8.64E-06 | 128777 | 2 |
| 475 | Chronic sinusitis | respiratory | DR | 0.93 | 0.92 | 0.95 | 8.93E-06 | 128777 | 2 |
| 358 | Myoneural disorders | neurological | DR | 1.43 | 1.32 | 1.55 | 9.83E-06 | 128777 | 2 |
| 742.1 | Loose body in joint | musculoskeletal | DR | 0.61 | 0.55 | 0.68 | 1.02E-05 | 128777 | 2 |
| 264.2 | Failure to thrive (childhood) | endocrine/metabolic | DR | 1.72 | 1.52 | 1.94 | 1.04E-05 | 111475 | 1 |
| 283 | Acquired hemolytic anemias | hematopoietic | DR | 1.47 | 1.34 | 1.60 | 1.15E-05 | 111475 | 1 |
| 715.1 | Sacroiliitis NEC | musculoskeletal | DR | 0.84 | 0.81 | 0.87 | 1.15E-05 | 128777 | 2 |
| 938.2 | Chronic dermatitis due to solar radiation | dermatologic | DR | 1.13 | 1.10 | 1.17 | 1.33E-05 | 128777 | 2 |
| 801.1 | Fracture of foot | injuries & poisonings | DR | 1.18 | 1.14 | 1.23 | 1.38E-05 | 128777 | 2 |
| 241.1 | Nontoxic uninodular goiter | endocrine/metabolic | DR | 1.13 | 1.10 | 1.17 | 1.40E-05 | 128777 | 2 |
| 300.8 | Acute reaction to stress | mental disorders | DR | 0.85 | 0.81 | 0.88 | 1.47E-05 | 128777 | 2 |
| 615 | Endometriosis | genitourinary | DR | 0.58 | 0.51 | 0.65 | 1.58E-05 | 128777 | 2 |
| 530.9 | Heartburn | digestive | DR | 0.87 | 0.85 | 0.90 | 1.68E-05 | 128777 | 2 |

| | | | | | | | | | |
|---|---|---|---|---|---|---|---|---|---|
| 550 | Abdominal hernia | digestive | DR | 0.94 | 0.92 | 0.95 | 1.69E-05 | 128777 | 2 |
| 706.2 | Sebaceous cyst | dermatologic | DR | 1.07 | 1.06 | 1.09 | 1.75E-05 | 128777 | 2 |
| 290.13 | Senile dementia | mental disorders | DR | 1.49 | 1.36 | 1.63 | 1.79E-05 | 128777 | 2 |
| 350.1 | Abnormal involuntary movements | neurological | DR | 1.09 | 1.07 | 1.11 | 1.83E-05 | 128777 | 2 |
| 81.1 | Graft-versus-host disease | infectious diseases | DR | 0.22 | 0.15 | 0.31 | 1.86E-05 | 128777 | 2 |
| 264 | | | DR | 1.67 | 1.48 | 1.88 | 1.93E-05 | 111475 | 1 |
| 276.8 | Polydipsia | endocrine/metabolic | DR | 0.66 | 0.60 | 0.73 | 2.02E-05 | 128777 | 2 |
| 327.5 | Parasomnia | neurological | DR | 0.89 | 0.86 | 0.91 | 2.08E-05 | 128777 | 2 |
| 480.3 | Pneumonia due to fungus (mycoses) | respiratory | DR | 1.24 | 1.18 | 1.31 | 2.21E-05 | 128777 | 2 |
| 222 | Benign neoplasm of male genital organs | neoplasms | DR | 1.17 | 1.13 | 1.21 | 2.25E-05 | 111475 | 1 |
| 747.2 | Congenital anomalies of peripheral vascular system | congenital anomalies | DR | 1.29 | 1.21 | 1.37 | 2.31E-05 | 128777 | 2 |
| 204.4 | Multiple myeloma | neoplasms | DR | 1.36 | 1.27 | 1.47 | 2.43E-05 | 128777 | 2 |
| 1019 | Other ill-defined and unknown causes of morbidity and mortality | NULL | DR | 1.08 | 1.06 | 1.09 | 2.47E-05 | 128777 | 2 |
| 571.8 | Liver abscess and sequelae of chronic liver disease | digestive | DR | 1.16 | 1.12 | 1.20 | 2.55E-05 | 128777 | 2 |
| 726.1 | Enthesopathy | musculoskeletal | DR | 1.06 | 1.05 | 1.08 | 2.65E-05 | 128777 | 2 |
| 809 | Fracture of unspecified bones | injuries & poisonings | DR | 1.13 | 1.10 | 1.16 | 2.80E-05 | 128777 | 2 |
| 348.9 | Other conditions of brain, NOS | neurological | DR | 1.16 | 1.12 | 1.21 | 3.07E-05 | 128777 | 2 |
| 1004 | Other signs and symptoms involving emotional state | NULL | DR | 0.85 | 0.81 | 0.88 | 3.09E-05 | 128777 | 2 |
| 840.2 | | | DR | 1.12 | 1.09 | 1.15 | 3.20E-05 | 128777 | 2 |
| 743.4 | Stress fracture | musculoskeletal | DR | 1.28 | 1.21 | 1.36 | 3.33E-05 | 128777 | 2 |
| 649.1 | Diabetes or abnormal glucose tolerance complicating pregnancy | pregnancy complications | DR | 1.61 | 1.43 | 1.80 | 3.38E-05 | 128777 | 2 |
| 756.3 | Congenital anomalies of muscle, tendon, fascia, and connective tissue | congenital anomalies | DR | 1.51 | 1.37 | 1.67 | 3.42E-05 | 111475 | 1 |
| 473 | Diseases of the larynx and vocal cords | respiratory | DR | 1.10 | 1.08 | 1.13 | 3.43E-05 | 128777 | 2 |
| 550.6 | Incisional hernia | digestive | DR | 0.85 | 0.81 | 0.88 | 3.59E-05 | 128777 | 2 |
| 451 | Phlebitis and thrombophlebitis | circulatory system | DR | 1.16 | 1.12 | 1.20 | 3.62E-05 | 128777 | 2 |
| 333 | Extrapyramidal disease and abnormal movement disorders | neurological | DR | 1.09 | 1.07 | 1.11 | 3.79E-05 | 128777 | 2 |
| 977 | | | DR | 1.29 | 1.21 | 1.37 | 4.09E-05 | 128777 | 2 |
| 512.2 | Painful respiration | respiratory | DR | 1.09 | 1.07 | 1.11 | 4.29E-05 | 128777 | 2 |
| 289 | Other diseases of blood and blood-forming organs | hematopoietic | DR | 1.16 | 1.12 | 1.20 | 4.52E-05 | 128777 | 2 |
| 571.81 | Portal hypertension | digestive | DR | 1.18 | 1.14 | 1.23 | 4.54E-05 | 128777 | 2 |
| 574.11 | Cholelithiasis with acute cholecystitis | digestive | DR | 1.23 | 1.17 | 1.29 | 4.59E-05 | 128777 | 2 |
| 578 | | | DR | 1.06 | 1.05 | 1.08 | 4.69E-05 | 111475 | 1 |
| 335 | Multiple sclerosis | neurological | DR | 0.74 | 0.69 | 0.80 | 4.90E-05 | 128777 | 2 |
| 245.21 | Chronic lymphocytic thyroiditis | endocrine/metabolic | DR | 1.32 | 1.23 | 1.42 | 5.44E-05 | 128777 | 2 |
| 750 | Digestive congenital anomalies | congenital anomalies | DR | 1.18 | 1.14 | 1.24 | 5.54E-05 | 111475 | 1 |
| 733.8 | Malunion and nonunion of fracture | musculoskeletal | DR | 1.22 | 1.16 | 1.28 | 5.60E-05 | 128777 | 2 |
| 291.1 | Transient mental disorders due to conditions classified elsewhere | mental disorders | DR | 0.70 | 0.64 | 0.77 | 5.74E-05 | 128777 | 2 |
| 506 | Empyema and pneumothorax | respiratory | DR | 1.18 | 1.13 | 1.22 | 5.91E-05 | 128777 | 2 |
| 627.4 | Premenopausal menorrhagia | genitourinary | DR | 0.55 | 0.47 | 0.64 | 6.36E-05 | 128777 | 2 |
| 751.2 | Congenital anomalies of urinary system | congenital anomalies | DR | 1.13 | 1.10 | 1.17 | 6.39E-05 | 128777 | 2 |
| 360.3 | Hypotony of eye | sense organs | DR | 1.67 | 1.47 | 1.90 | 6.59E-05 | 128777 | 2 |
| 301.1 | Schizoid personality disorder | mental disorders | DR | 0.75 | 0.69 | 0.80 | 6.74E-05 | 111475 | 1 |
| 562.2 | Diverticulitis | digestive | DR | 0.90 | 0.88 | 0.93 | 7.07E-05 | 128777 | 2 |
| 286.4 | Acquired coagulation factor deficiency | hematopoietic | DR | 1.37 | 1.26 | 1.48 | 7.54E-05 | 128777 | 2 |
| 481 | Influenza | respiratory | DR | 1.11 | 1.08 | 1.13 | 7.58E-05 | 128777 | 2 |
| 530.3 | Stricture and stenosis of esophagus | digestive | DR | 1.16 | 1.12 | 1.20 | 8.07E-05 | 128777 | 2 |
| 300.3 | Obsessive-compulsive disorders | mental disorders | DR | 0.83 | 0.79 | 0.87 | 8.85E-05 | 128777 | 2 |
| 614.3 | Pelvic inflammatory disease (PID) | genitourinary | DR | 0.46 | 0.38 | 0.56 | 0.000100882 | 111475 | 1 |
| 480.13 | MRSA pneumonia | respiratory | DR | 1.53 | 1.37 | 1.71 | 0.000102926 | 128777 | 2 |
| 739 | Contracture of joint | musculoskeletal | DR | 1.18 | 1.13 | 1.24 | 0.000109593 | 128777 | 2 |
| 550.1 | Inguinal hernia | digestive | DR | 0.91 | 0.89 | 0.93 | 0.000111594 | 128777 | 2 |
| 218.1 | Uterine leiomyoma | neoplasms | DR | 0.71 | 0.65 | 0.78 | 0.000112725 | 128777 | 2 |
| 327.72 | Sleep related leg cramps | neurological | DR | 1.22 | 1.16 | 1.28 | 0.000113881 | 128777 | 2 |
| 530.15 | Eosinophilic esophagitis | digestive | DR | 0.56 | 0.48 | 0.65 | 0.000115601 | 128777 | 2 |
| 70.4 | Chronic hepatitis | infectious diseases | DR | 0.80 | 0.76 | 0.85 | 0.000122295 | 128777 | 2 |
| 686.1 | Carbuncle and furuncle | dermatologic | DR | 1.12 | 1.08 | 1.15 | 0.000126675 | 128777 | 2 |
| 200.1 | Polycythemia vera | neoplasms | DR | 0.77 | 0.72 | 0.82 | 0.000127204 | 128777 | 2 |
| 686.4 | Pyogenic granuloma | dermatologic | DR | 1.27 | 1.19 | 1.35 | 0.000128207 | 128777 | 2 |
| 870.1 | Open wound or laceration of eye or eyelid | injuries & poisonings | DR | 1.26 | 1.18 | 1.33 | 0.000128532 | 128777 | 2 |
| 756.5 | Congenital osteodystrophies | congenital anomalies | DR | 1.31 | 1.22 | 1.41 | 0.000133471 | 128777 | 2 |
| 364.41 | Keratoconus | sense organs | DR | 0.60 | 0.52 | 0.68 | 0.00013411 | 128777 | 2 |
| 1090 | Acquired absence of organs | | DR | 1.11 | 1.08 | 1.14 | 0.000141166 | 128777 | 2 |
| 289.3 | Personal history of diseases of blood and blood-forming organs | hematopoietic | DR | 1.37 | 1.26 | 1.49 | 0.000150453 | 128777 | 2 |
| 519.9 | Symptoms involving respiratory system and other chest symptoms | respiratory | DR | 1.11 | 1.08 | 1.14 | 0.000154825 | 128777 | 2 |
| 727.4 | Ganglion and cyst of synovium, tendon, and bursa | musculoskeletal | DR | 0.89 | 0.86 | 0.92 | 0.00017471 | 128777 | 2 |
| 353.2 | Nerve root lesions | neurological | DR | 0.86 | 0.83 | 0.90 | 0.000182359 | 128777 | 2 |
| 217 | Vascular hamartomas and non-neoplastic nevi | neoplasms | DR | 1.08 | 1.06 | 1.11 | 0.000184575 | 111475 | 1 |
| 288.1 | Decreased white blood cell count | hematopoietic | DR | 1.14 | 1.10 | 1.18 | 0.000185175 | 128777 | 2 |
| 180.3 | Cervical intraepithelial neoplasia [CIN] [Cervical dysplasia] | neoplasms | DR | 0.63 | 0.56 | 0.72 | 0.00019426 | 128777 | 2 |
| 253.2 | Pituitary hypofunction | endocrine/metabolic | DR | 0.71 | 0.65 | 0.78 | 0.000198776 | 128777 | 2 |
| 376 | Disorders of the orbit | sense organs | DR | 1.45 | 1.32 | 1.61 | 0.000203304 | 128777 | 2 |
| 519.2 | Respiratory complications | respiratory | DR | 1.24 | 1.17 | 1.32 | 0.000211779 | 128777 | 2 |
| 364.1 | Corneal opacity | sense organs | DR | 1.11 | 1.08 | 1.15 | 0.000215473 | 128777 | 2 |
| 496.21 | Obstructive chronic bronchitis | respiratory | DR | 1.07 | 1.05 | 1.09 | 0.000249143 | 128777 | 2 |
| 214.1 | Lipoma of skin and subcutaneous tissue | neoplasms | DR | 0.89 | 0.86 | 0.92 | 0.000252213 | 128777 | 2 |
| 738.4 | Acquired spondylolisthesis | musculoskeletal | DR | 0.88 | 0.85 | 0.91 | 0.000252684 | 128777 | 2 |
| 365.5 | Pseudoexfoliation glaucoma | sense organs | DR | 1.42 | 1.29 | 1.56 | 0.000263257 | 128777 | 2 |
| 287.31 | Primary thrombocytopenia | hematopoietic | DR | 1.28 | 1.20 | 1.37 | 0.000264064 | 128777 | 2 |
| 793.2 | Nonspecific abnormal findings on radiological and other examination of other intrathoracic organs (echocardiogram, etc) | symptoms | DR | 1.09 | 1.07 | 1.12 | 0.000267371 | 128777 | 2 |
| 287.1 | Spontaneous ecchymoses | hematopoietic | DR | 1.25 | 1.17 | 1.32 | 0.00027104 | 128777 | 2 |
| 555.1 | Regional enteritis | digestive | DR | 0.80 | 0.75 | 0.85 | 0.000278582 | 128777 | 2 |
| 575.1 | Cholangitis | digestive | DR | 1.30 | 1.21 | 1.39 | 0.000281539 | 128777 | 2 |
| 278.1 | Obesity | endocrine/metabolic | DR | 0.95 | 0.94 | 0.96 | 0.000290662 | 128777 | 2 |
| 442.3 | Aneurysm of artery of lower extremity | circulatory system | DR | 1.29 | 1.20 | 1.38 | 0.000292261 | 128777 | 2 |
| 721 | Spondylosis and allied disorders | musculoskeletal | DR | 0.95 | 0.94 | 0.96 | 0.000297265 | 128777 | 2 |
| 729.3 | Panniculitis | musculoskeletal | DR | 1.36 | 1.25 | 1.48 | 0.000300191 | 128777 | 2 |
| 743.22 | | | DR | 1.86 | 1.57 | 2.22 | 0.000314369 | 128777 | 2 |
| 782 | | | DR | 2.07 | 1.69 | 2.53 | 0.000336097 | 17302 | 1 |
| 614.33 | Pelvic inflammatory disease, NOS | genitourinary | DR | 0.23 | 0.15 | 0.34 | 0.000340567 | 111475 | 1 |
| 189.1 | Cancer of kidney and renal pelvis | neoplasms | DR | 1.16 | 1.11 | 1.21 | 0.000343843 | 128777 | 2 |
| 535.9 | Gastritis and duodenitis, NOS | digestive | DR | 1.08 | 1.06 | 1.11 | 0.000357158 | 128777 | 2 |

| | | | | | | | | | |
|---|---|---|---|---|---|---|---|---|---|
| 772 | Symptoms of the muscles | symptoms | DR | 1.17 | 1.12 | 1.22 | 0.000366474 | 128777 | 2 |
| 942 | Infusion and transfusion reaction | injuries & poisonings | DR | 1.59 | 1.40 | 1.82 | 0.000368069 | 128777 | 2 |
| 709 | | | DR | 1.22 | 1.15 | 1.29 | 0.000387835 | 111475 | 1 |
| 366.3 | Traumatic cataract | sense organs | DR | 1.35 | 1.24 | 1.47 | 0.000391799 | 128777 | 2 |
| 753.1 | Congenital cataract and lens anomalies | congenital anomalies | DR | 1.33 | 1.23 | 1.44 | 0.000392722 | 128777 | 2 |
| 625.1 | Dyspareunia | genitourinary | DR | 0.64 | 0.56 | 0.72 | 0.000405986 | 128777 | 2 |
| 81.11 | Acute graft-versus-host disease | infectious diseases | DR | 0.27 | 0.18 | 0.39 | 0.000409646 | 128777 | 2 |
| 705 | Disorders of sweat glands | dermatologic | DR | 0.87 | 0.84 | 0.91 | 0.00041454 | 111475 | 1 |
| 586.3 | Vascular disorders of kidney/hypertrophy | genitourinary | DR | 1.43 | 1.29 | 1.58 | 0.000417145 | 128777 | 2 |
| 626.4 | Premenstrual tension syndromes | genitourinary | DR | 0.35 | 0.26 | 0.47 | 0.000417288 | 111475 | 1 |
| 705.3 | Hidradenitis | dermatologic | DR | 0.78 | 0.73 | 0.84 | 0.000420562 | 128777 | 2 |
| 180 | | | DR | 0.64 | 0.57 | 0.73 | 0.0004223 | 111475 | 1 |
| 794 | Abnormal results of other function studies (bladder, pancreas, placenta, spleen, etc) | symptoms | DR | 1.14 | 1.10 | 1.18 | 0.000479635 | 128777 | 2 |
| 626.15 | Infertility, female, associated with anovulation | genitourinary | DR | 0.15 | 0.09 | 0.26 | 0.000501438 | 111475 | 1 |
| 579.2 | Splenomegaly | digestive | DR | 1.13 | 1.09 | 1.17 | 0.000531794 | 128777 | 2 |
| 371.21 | Allergic conjunctivitis | sense organs | DR | 0.92 | 0.90 | 0.94 | 0.000533551 | 128777 | 2 |
| 333.4 | Torsion dystonia | neurological | DR | 0.82 | 0.77 | 0.87 | 0.00055997 | 128777 | 2 |
| 170 | Cancer of bone and connective tissue | neoplasms | DR | 1.23 | 1.16 | 1.31 | 0.000561994 | 111475 | 1 |
| 389.3 | Degenerative and vascular disorders of ear | sense organs | DR | 1.20 | 1.14 | 1.26 | 0.00056756 | 128777 | 2 |
| 441.1 | Acute vascular insufficiency of intestine | circulatory system | DR | 1.33 | 1.22 | 1.44 | 0.000584249 | 128777 | 2 |
| 686.5 | Pyoderma | dermatologic | DR | 1.40 | 1.27 | 1.54 | 0.000595499 | 128777 | 2 |
| 681.2 | Cellulitis and abscess of face/neck | dermatologic | DR | 1.12 | 1.09 | 1.16 | 0.000624557 | 128777 | 2 |
| 155.1 | Malignant neoplasm of liver, primary | neoplasms | DR | 1.20 | 1.14 | 1.26 | 0.000626587 | 128777 | 2 |
| 430.3 | Subdural hemorrhage | circulatory system | DR | 1.25 | 1.17 | 1.33 | 0.000631822 | 128777 | 2 |
| 388 | Other disorders of ear | sense organs | DR | 1.13 | 1.09 | 1.17 | 0.000666489 | 128777 | 2 |
| 614 | Inflammatory diseases of female pelvic organs | genitourinary | DR | 0.80 | 0.74 | 0.85 | 0.000673284 | 111475 | 1 |
| 741.4 | Joint effusions | musculoskeletal | DR | 1.08 | 1.06 | 1.10 | 0.000694075 | 128777 | 2 |
| 789.1 | | | DR | 1.53 | 1.35 | 1.74 | 0.000701588 | 128777 | 2 |
| 594.8 | Renal colic | genitourinary | DR | 0.85 | 0.81 | 0.89 | 0.000715324 | 128777 | 2 |
| 564.8 | Abnormal findings on exam of gastrointestinal tract/ abdominal area | digestive | DR | 1.09 | 1.06 | 1.12 | 0.000719228 | 128777 | 2 |
| 535.8 | Other specified gastritis | digestive | DR | 1.11 | 1.07 | 1.14 | 0.000745069 | 128777 | 2 |
| 695.3 | Rosacea | dermatologic | DR | 1.09 | 1.07 | 1.12 | 0.000749337 | 128777 | 2 |
| 751.21 | Cystic kidney disease | congenital anomalies | DR | 1.12 | 1.08 | 1.15 | 0.000750694 | 128777 | 2 |
| 420.21 | Acute pericarditis | circulatory system | DR | 1.31 | 1.21 | 1.42 | 0.000773664 | 128777 | 2 |
| 530.12 | Ulcer of esophagus | digestive | DR | 1.17 | 1.11 | 1.22 | 0.000812 | 128777 | 2 |
| 1009 | Injury, NOS | NULL | DR | 1.05 | 1.03 | 1.06 | 0.000822903 | 128777 | 2 |
| 352 | Disorders of other cranial nerves | neurological | DR | 1.11 | 1.08 | 1.15 | 0.000837832 | 128777 | 2 |
| 380 | Disorders of external ear | sense organs | DR | 1.12 | 1.08 | 1.15 | 0.000858331 | 128777 | 2 |
| 577 | Diseases of pancreas | digestive | DR | 1.09 | 1.06 | 1.11 | 0.000868707 | 128777 | 2 |
| 472 | Chronic pharyngitis and nasopharyngitis | respiratory | DR | 0.94 | 0.92 | 0.96 | 0.000869185 | 128777 | 2 |
| 189.11 | Malignant neoplasm of kidney, except pelvis | neoplasms | DR | 1.15 | 1.10 | 1.20 | 0.000870264 | 128777 | 2 |
| 530.2 | Esophageal bleeding (varices/hemorrhage) | digestive | DR | 1.12 | 1.08 | 1.16 | 0.000923236 | 128777 | 2 |
| 227.3 | Benign neoplasm of pituitary gland and craniopharyngeal duct (pouch) | neoplasms | DR | 0.78 | 0.72 | 0.84 | 0.000925551 | 128777 | 2 |
| 732.7 | Osteochondritis dissecans | musculoskeletal | DR | 0.63 | 0.54 | 0.72 | 0.000926393 | 111475 | 1 |
| 530.14 | Reflux esophagitis | digestive | DR | 0.93 | 0.91 | 0.95 | 0.000939073 | 128777 | 2 |
| 578.1 | Hematemesis | digestive | DR | 1.17 | 1.12 | 1.23 | 0.00095734 | 128777 | 2 |
| 742.2 | Pathological, developmental or recurrent dislocation | musculoskeletal | DR | 0.77 | 0.71 | 0.83 | 0.000981474 | 128777 | 2 |
| 720.1 | Spinal stenosis of lumbar region | musculoskeletal | DR | 1.06 | 1.04 | 1.08 | 0.001036894 | 128777 | 2 |
| 601.3 | Orchitis and epididymitis | genitourinary | DR | 0.90 | 0.87 | 0.93 | 0.001039139 | 128777 | 2 |
| 609.2 | Abnormal spermatozoa | genitourinary | DR | 0.81 | 0.76 | 0.86 | 0.001045169 | 111475 | 1 |
| 571.51 | Cirrhosis of liver without mention of alcohol | digestive | DR | 1.09 | 1.06 | 1.11 | 0.001064563 | 128777 | 2 |
| 642.1 | Preeclampsia and eclampsia | pregnancy complications | DR | 2.17 | 1.71 | 2.76 | 0.001071712 | 128777 | 2 |
| 569.1 | Toxic gastroenteritis and colitis | digestive | DR | 1.43 | 1.28 | 1.59 | 0.001079002 | 128777 | 2 |
| 564.9 | Personal history of diseases of digestive system | digestive | DR | 1.13 | 1.09 | 1.17 | 0.001083036 | 128777 | 2 |
| 303.1 | Dissociative disorder | mental disorders | DR | 0.70 | 0.63 | 0.78 | 0.001119505 | 111475 | 1 |
| 766 | Neuralgia, neuritis, and radiculitis NOS | symptoms | DR | 0.93 | 0.91 | 0.95 | 0.001121376 | 128777 | 2 |
| 430.1 | Subarachnoid hemorrhage | circulatory system | DR | 1.32 | 1.21 | 1.43 | 0.001128194 | 128777 | 2 |
| 573.4 | Acute and subacute necrosis of liver | digestive | DR | 1.24 | 1.16 | 1.32 | 0.00112936 | 128777 | 2 |
| 620.1 | Dysplasia of cervix | genitourinary | DR | 0.34 | 0.25 | 0.48 | 0.00114602 | 111475 | 1 |
| 626.14 | Irregular menstrual bleeding | genitourinary | DR | 0.64 | 0.56 | 0.74 | 0.001165064 | 128777 | 2 |
| 263 | Other nutritional deficiency | endocrine/metabolic | DR | 1.19 | 1.13 | 1.25 | 0.001165185 | 128777 | 2 |
| 740.11 | Osteoarthrosis, localized, primary | musculoskeletal | DR | 0.96 | 0.94 | 0.97 | 0.001168365 | 128777 | 2 |
| 732 | Osteochondropathies | musculoskeletal | DR | 0.78 | 0.73 | 0.84 | 0.00119974 | 111475 | 1 |
| 158 | Neoplasm of unspecified nature of digestive system | neoplasms | DR | 1.15 | 1.10 | 1.20 | 0.001203816 | 128777 | 2 |
| 333.8 | Other degenerative diseases of the basal ganglia | neurological | DR | 1.43 | 1.28 | 1.60 | 0.001227279 | 128777 | 2 |
| 368.5 | Color vision deficiencies | sense organs | DR | 1.46 | 1.30 | 1.64 | 0.00123242 | 111475 | 1 |
| 743.13 | | | DR | 1.28 | 1.19 | 1.38 | 0.001248696 | 128777 | 2 |
| 972.1 | | | DR | 1.48 | 1.31 | 1.67 | 0.001259108 | 128777 | 2 |
| 189.4 | Malignant neoplasm of other urinary organs | neoplasms | DR | 1.32 | 1.21 | 1.43 | 0.001287435 | 128777 | 2 |
| 836 | Traumatic arthropathy | injuries & poisonings | DR | 0.90 | 0.87 | 0.93 | 0.001324222 | 128777 | 2 |
| 79.9 | Viremia, NOS | infectious diseases | DR | 0.83 | 0.79 | 0.88 | 0.001350804 | 128777 | 2 |
| 531.2 | Gastric ulcer | digestive | DR | 1.13 | 1.09 | 1.17 | 0.001356488 | 128777 | 2 |
| 751 | Genitourinary congenital anomalies | congenital anomalies | DR | 1.09 | 1.06 | 1.12 | 0.001386121 | 111475 | 1 |
| 990 | Effects radiation NOS | injuries & poisonings | DR | 1.12 | 1.08 | 1.16 | 0.00139289 | 128777 | 2 |
| 217.1 | Nevus, non-neoplastic | neoplasms | DR | 1.07 | 1.05 | 1.09 | 0.001420688 | 128777 | 2 |
| 368.91 | Psychophysical visual disturbances | sense organs | DR | 1.25 | 1.17 | 1.35 | 0.001448325 | 128777 | 2 |
| 516 | Abnormal sputum | respiratory | DR | 1.11 | 1.07 | 1.15 | 0.00145892 | 128777 | 2 |
| 242.2 | Toxic multinodular goiter | endocrine/metabolic | DR | 1.44 | 1.29 | 1.62 | 0.001495497 | 128777 | 2 |
| 442.1 | Aortic aneurysm | circulatory system | DR | 1.08 | 1.05 | 1.11 | 0.001560785 | 128777 | 2 |
| 244.5 | Congenital hypothyroidism | endocrine/metabolic | DR | 1.92 | 1.56 | 2.37 | 0.00156101 | 128777 | 2 |
| 558 | Noninfectious gastroenteritis | digestive | DR | 1.06 | 1.04 | 1.09 | 0.001588817 | 128777 | 2 |
| 790.8 | Elevated C-reactive protein (CRP) | symptoms | DR | 1.30 | 1.19 | 1.41 | 0.001651924 | 128777 | 2 |
| 352.2 | Facial nerve disorders [CN7] | neurological | DR | 1.13 | 1.09 | 1.18 | 0.001718852 | 128777 | 2 |
| 516.1 | Hemoptysis | respiratory | DR | 1.11 | 1.08 | 1.15 | 0.001760021 | 128777 | 2 |
| 327.6 | Circadian rhythm sleep disorder | neurological | DR | 0.82 | 0.77 | 0.88 | 0.001770423 | 128777 | 2 |
| 204.1 | Lymphoid leukemia | neoplasms | DR | 1.22 | 1.14 | 1.29 | 0.001771911 | 128777 | 2 |
| 362.1 | Retinopathy of prematurity | sense organs | DR | 2.44 | 1.83 | 3.24 | 0.001778337 | 111475 | 1 |
| 671 | Venous/cerebrovascular complications embolism in pregnancy and the puerperium | pregnancy complications | DR | 1.70 | 1.43 | 2.02 | 0.001879723 | 128777 | 2 |
| 870.2 | Open wound of ear | injuries & poisonings | DR | 1.33 | 1.21 | 1.46 | 0.001904792 | 128777 | 2 |
| 394.4 | Acute rheumatic heart disease | circulatory system | DR | 1.53 | 1.33 | 1.75 | 0.001915182 | 128777 | 2 |
| 204.12 | Lymphoid leukemia, chronic | neoplasms | DR | 1.22 | 1.15 | 1.30 | 0.001915562 | 128777 | 2 |
| 735.23 | Hallux rigidus | musculoskeletal | DR | 1.11 | 1.07 | 1.15 | 0.001934386 | 128777 | 2 |
| 526.42 | Arthralgia/ankylosis of temporomandibular joint | digestive | DR | 0.81 | 0.76 | 0.87 | 0.001936101 | 128777 | 2 |
| 750.2 | Lower gastrointestinal congenital anomalies | congenital anomalies | DR | 1.20 | 1.13 | 1.27 | 0.0019676 | 111475 | 1 |
| 790.9 | Abnormal arterial blood gases | symptoms | DR | 1.63 | 1.39 | 1.92 | 0.001969863 | 111475 | 1 |

| | | | | | | | | | |
|---|---|---|---|---|---|---|---|---|---|
| 324 | Other CNS infection and poliomyelitis | neurological | DR | 1.27 | 1.17 | 1.37 | 0.001971317 | 128777 | 2 |
| 200 | Myeloproliferative disease | neoplasms | DR | 1.12 | 1.08 | 1.17 | 0.002134599 | 128777 | 2 |
| 441.2 | Chronic vascular insufficiency of intestine | circulatory system | DR | 1.40 | 1.26 | 1.57 | 0.002142091 | 128777 | 2 |
| 90.2 | Gonococcal infections | infectious diseases | DR | 0.52 | 0.42 | 0.64 | 0.002460545 | 111475 | 1 |
| 595 | Hydronephrosis | genitourinary | DR | 1.10 | 1.06 | 1.13 | 0.002501419 | 128777 | 2 |
| 967 | Adverse effects of sedatives or other central nervous system depressants and anesthetics | injuries & poisonings | DR | 1.16 | 1.11 | 1.22 | 0.002508475 | 128777 | 2 |
| 253.5 | | | DR | 0.19 | 0.11 | 0.33 | 0.002528523 | 128777 | 2 |
| 612.1 | Galactorrhea | genitourinary | DR | 0.48 | 0.38 | 0.62 | 0.002564049 | 128777 | 2 |
| 731.1 | Osteitis deformans [Paget's disease of bone] | musculoskeletal | DR | 1.53 | 1.33 | 1.77 | 0.002589084 | 111475 | 1 |
| 259.3 | Delay in sexual development and puberty NEC | endocrine/metabolic | DR | 1.24 | 1.15 | 1.33 | 0.002642334 | 111475 | 1 |
| 369.5 | Conjunctivitis, infectious | sense organs | DR | 1.06 | 1.04 | 1.09 | 0.002672584 | 128777 | 2 |
| 277.4 | Disorders of bilirubin excretion | endocrine/metabolic | DR | 1.17 | 1.11 | 1.23 | 0.002687648 | 128777 | 2 |
| 381.3 | Mastoiditis & related conditions | sense organs | DR | 1.21 | 1.13 | 1.29 | 0.002799148 | 128777 | 2 |
| 153.3 | Malignant neoplasm of rectum, rectosigmoid junction, and anus | neoplasms | DR | 1.18 | 1.12 | 1.25 | 0.00288155 | 128777 | 2 |
| 285.3 | Sideroblastic anemia | hematopoietic | DR | 1.90 | 1.53 | 2.35 | 0.002900939 | 111475 | 1 |
| 642 | Hypertension complicating pregnancy, childbirth, and the puerperium | pregnancy complications | DR | 1.60 | 1.36 | 1.87 | 0.002912837 | 128777 | 2 |
| 569 | Other disorders of intestine | digestive | DR | 1.08 | 1.05 | 1.10 | 0.00291408 | 128777 | 2 |
| 323 | Encephalitis | neurological | DR | 1.34 | 1.22 | 1.49 | 0.003027019 | 128777 | 2 |
| 514.1 | Abnormal results of function study of pulmonary system | respiratory | DR | 1.17 | 1.11 | 1.23 | 0.003090272 | 128777 | 2 |
| 255 | Disorders of adrenal glands | endocrine/metabolic | DR | 1.11 | 1.07 | 1.15 | 0.003329323 | 128777 | 2 |
| 704.2 | Hirsutism | dermatologic | DR | 0.64 | 0.55 | 0.75 | 0.003350672 | 128777 | 2 |
| 218 | | | DR | 0.74 | 0.67 | 0.82 | 0.003404909 | 111475 | 1 |
| 359 | | | DR | 1.18 | 1.11 | 1.24 | 0.003530475 | 111475 | 1 |
| 79 | Viral infection | infectious diseases | DR | 1.07 | 1.05 | 1.10 | 0.003631755 | 128777 | 2 |
| 216 | Benign neoplasm of skin | neoplasms | DR | 1.04 | 1.03 | 1.06 | 0.003763482 | 128777 | 2 |
| 555 | | | DR | 0.89 | 0.86 | 0.93 | 0.00395381 | 111475 | 1 |
| 636 | Early or threatened labor; hemorrhage in early pregnancy | pregnancy complications | DR | 0.42 | 0.31 | 0.57 | 0.003976382 | 128777 | 2 |
| 742.8 | Articular cartilage disorder | musculoskeletal | DR | 0.79 | 0.72 | 0.85 | 0.004116682 | 128777 | 2 |
| 740.2 | Osteoarthrosis, generalized | musculoskeletal | DR | 1.06 | 1.04 | 1.09 | 0.004126508 | 128777 | 2 |
| 798.1 | Chronic fatigue syndrome | symptoms | DR | 0.90 | 0.87 | 0.93 | 0.004132528 | 128777 | 2 |
| 540.1 | Appendicitis | digestive | DR | 0.84 | 0.79 | 0.89 | 0.004157083 | 128777 | 2 |
| 987 | Toxic effect of other gases, fumes, or vapors | injuries & poisonings | DR | 0.54 | 0.43 | 0.67 | 0.004223537 | 111475 | 1 |
| 523.32 | Chronic periodontitis | digestive | DR | 1.06 | 1.04 | 1.08 | 0.0042548 | 128777 | 2 |
| 650 | Normal delivery | pregnancy complications | DR | 0.20 | 0.12 | 0.35 | 0.004292741 | 111475 | 1 |
| 737.1 | Kyphosis (acquired) | musculoskeletal | DR | 1.22 | 1.14 | 1.31 | 0.004369439 | 128777 | 2 |
| 41.8 | H. pylori | infectious diseases | DR | 1.14 | 1.09 | 1.19 | 0.004494823 | 128777 | 2 |
| 523.31 | Acute periodontitis | digestive | DR | 1.06 | 1.04 | 1.08 | 0.004691659 | 128777 | 2 |
| 728 | Disorders of muscle, ligament, and fascia | musculoskeletal | DR | 0.87 | 0.83 | 0.92 | 0.004793871 | 128777 | 2 |
| 818.2 | | | DR | 1.47 | 1.28 | 1.70 | 0.005396231 | 128777 | 2 |
| 965.1 | Opiates and related narcotics causing adverse effects in therapeutic use | injuries & poisonings | DR | 1.13 | 1.08 | 1.18 | 0.005407436 | 128777 | 2 |
| 198 | Secondary malignant neoplasm | neoplasms | DR | 1.08 | 1.05 | 1.11 | 0.005445221 | 128777 | 2 |
| 509.5 | Respiratory arrest | respiratory | DR | 1.41 | 1.25 | 1.60 | 0.005496632 | 128777 | 2 |
| 740.1 | Osteoarthritis; localized | musculoskeletal | DR | 1.04 | 1.02 | 1.05 | 0.005517353 | 128777 | 2 |
| 608 | Other disorders of male genital organs | genitourinary | DR | 0.94 | 0.91 | 0.96 | 0.005651498 | 128777 | 2 |
| 741.5 | Hemarthrosis | musculoskeletal | DR | 1.32 | 1.19 | 1.45 | 0.005682242 | 128777 | 2 |
| 1012 | Late effect | NULL | DR | 0.81 | 0.75 | 0.87 | 0.005860256 | 128777 | 2 |
| 562.1 | Diverticulosis | digestive | DR | 1.04 | 1.02 | 1.05 | 0.005861191 | 128777 | 2 |
| 370.1 | Corneal ulcer | sense organs | DR | 1.21 | 1.13 | 1.29 | 0.005905114 | 128777 | 2 |
| 473.4 | Voice disturbance | respiratory | DR | 1.07 | 1.04 | 1.10 | 0.006004716 | 128777 | 2 |
| 601.8 | Other inflammatory disorders of male genital organs | genitourinary | DR | 1.16 | 1.10 | 1.23 | 0.006361867 | 128777 | 2 |
| 614.51 | Cervicitis and endocervicitis | genitourinary | DR | 0.69 | 0.60 | 0.79 | 0.00636255 | 128777 | 2 |
| 620 | Dysplasia of female genital organs | genitourinary | DR | 0.55 | 0.45 | 0.69 | 0.006577071 | 111475 | 1 |
| 525 | Other diseases of the teeth and supporting structures | digestive | DR | 1.04 | 1.02 | 1.05 | 0.006593131 | 128777 | 2 |
| 750.14 | Congenital anomalies of esophagus | congenital anomalies | DR | 1.51 | 1.30 | 1.76 | 0.006930222 | 111475 | 1 |
| 455 | Hemorrhoids | circulatory system | DR | 0.96 | 0.95 | 0.98 | 0.006954122 | 128777 | 2 |
| 733.4 | Aseptic necrosis of bone | musculoskeletal | DR | 0.86 | 0.81 | 0.91 | 0.006970628 | 128777 | 2 |
| 479 | Other upper respiratory disease | respiratory | DR | 0.95 | 0.93 | 0.97 | 0.007003112 | 128777 | 2 |
| 695.22 | Pemphigus and pemphigoid | dermatologic | DR | 1.39 | 1.23 | 1.57 | 0.007055304 | 128777 | 2 |
| 965 | Poisoning by analgesics, antipyretics, and antirheumatics | injuries & poisonings | DR | 1.16 | 1.09 | 1.22 | 0.007064117 | 128777 | 2 |
| 90.3 | Venereal diseases due to Chlamydia trachomatis | infectious diseases | DR | 0.34 | 0.23 | 0.51 | 0.007097706 | 111475 | 1 |
| 389.5 | Disorders of acoustic nerve | sense organs | DR | 1.22 | 1.14 | 1.32 | 0.007113044 | 111475 | 1 |
| 368.1 | Amblyopia | sense organs | DR | 1.10 | 1.06 | 1.14 | 0.007226021 | 128777 | 2 |
| 870.8 | Open wound of genital organs | injuries & poisonings | DR | 1.30 | 1.18 | 1.44 | 0.007342961 | 111475 | 1 |
| 750.21 | Congenital anomalies of intestine | congenital anomalies | DR | 1.21 | 1.13 | 1.30 | 0.007534971 | 128777 | 2 |
| 189.12 | Malignant neoplasm of renal pelvis | neoplasms | DR | 1.21 | 1.13 | 1.30 | 0.007552878 | 128777 | 2 |
| 523.3 | Periodontitis (acute or chronic) | digestive | DR | 1.05 | 1.03 | 1.06 | 0.007743404 | 128777 | 2 |
| 540 | Appendiceal conditions | digestive | DR | 0.86 | 0.81 | 0.91 | 0.007849789 | 111475 | 1 |
| 255.12 | Hyperaldosteronism | endocrine/metabolic | DR | 1.28 | 1.17 | 1.41 | 0.007896959 | 128777 | 2 |
| 315.1 | Learning disorder | mental disorders | DR | 0.68 | 0.59 | 0.79 | 0.00792917 | 111475 | 1 |
| 282.9 | Other hereditary hemolytic anemias | hematopoietic | DR | 1.43 | 1.25 | 1.64 | 0.00805028 | 128777 | 2 |
| 704.1 | Alopecia | dermatologic | DR | 0.86 | 0.81 | 0.91 | 0.008175274 | 128777 | 2 |
| 722.7 | Intervertebral disc disorder with myelopathy | musculoskeletal | DR | 0.90 | 0.86 | 0.94 | 0.00833702 | 128777 | 2 |
| 443.1 | Raynaud's syndrome | circulatory system | DR | 0.79 | 0.73 | 0.87 | 0.008422487 | 128777 | 2 |
| 965.3 | Salicylates causing adverse effects in therapeutic use | injuries & poisonings | DR | 1.57 | 1.32 | 1.86 | 0.008698468 | 111475 | 1 |
| 803.3 | Fracture of clavicle or scapula | injuries & poisonings | DR | 1.16 | 1.10 | 1.23 | 0.008728876 | 128777 | 2 |
| 386.21 | Central origin vertigo | sense organs | DR | 1.17 | 1.10 | 1.25 | 0.008888487 | 128777 | 2 |
| 521.1 | Dental caries | digestive | DR | 0.96 | 0.95 | 0.98 | 0.009202158 | 128777 | 2 |
| 710.3 | Osteopathy resulting from poliomyelitis | musculoskeletal | DR | 3.11 | 2.01 | 4.81 | 0.009330705 | 111475 | 1 |
| 747.11 | Cardiac shunt/ heart septal defect | congenital anomalies | DR | 1.19 | 1.11 | 1.27 | 0.009414231 | 128777 | 2 |
| 593.2 | Microscopic hematuria | genitourinary | DR | 1.06 | 1.04 | 1.08 | 0.009472377 | 128777 | 2 |
| 818.1 | | | DR | 1.31 | 1.18 | 1.45 | 0.009619777 | 128777 | 2 |
| 624.2 | Atrophy of female genital tract | genitourinary | DR | 1.80 | 1.43 | 2.25 | 0.0097906 | 111475 | 1 |
| 560.4 | Other intestinal obstruction | digestive | DR | 1.10 | 1.06 | 1.13 | 0.009808965 | 128777 | 2 |
| 528.12 | Oral aphthae | digestive | DR | 0.84 | 0.78 | 0.90 | 0.009958183 | 128777 | 2 |
| 602 | Other disorders of prostate | genitourinary | DR | 1.08 | 1.05 | 1.12 | 0.010194704 | 128777 | 2 |
| 656 | Other perinatal conditions of fetus or newborn | pregnancy complications | DR | 1.25 | 1.15 | 1.37 | 0.010428854 | 128777 | 2 |
| 170.2 | Cancer of connective tissue | neoplasms | DR | 1.20 | 1.12 | 1.29 | 0.010479765 | 128777 | 2 |
| 695.42 | Systemic lupus erythematosus | dermatologic | DR | 0.77 | 0.69 | 0.85 | 0.010517662 | 128777 | 2 |
| 727.7 | Contracture of tendon (sheath) | musculoskeletal | DR | 1.21 | 1.12 | 1.30 | 0.010650522 | 128777 | 2 |
| 271.9 | Other disorders of carbohydrate transport and metabolism | endocrine/metabolic | DR | 1.30 | 1.17 | 1.44 | 0.010702929 | 128777 | 2 |
| 609.11 | Azoospermia and oligospermia | genitourinary | DR | 0.43 | 0.31 | 0.60 | 0.010705934 | 111475 | 1 |
| 362.5 | Toxic maculopathy of retina | sense organs | DR | 1.34 | 1.19 | 1.50 | 0.010990665 | 128777 | 2 |
| 580.12 | Non-proliferative glomerulonephritis | genitourinary | DR | 1.33 | 1.19 | 1.49 | 0.011243712 | 128777 | 2 |
| 198.5 | Secondary malignancy of brain/spine | neoplasms | DR | 0.81 | 0.75 | 0.88 | 0.011378539 | 128777 | 2 |
| 610.1 | Cystic mastopathy | genitourinary | DR | 0.83 | 0.77 | 0.89 | 0.011587173 | 128777 | 2 |

| | | | | | | | | | |
|---|---|---|---|---|---|---|---|---|---|
| 527.7 | Disturbance of salivary secretion | digestive | DR | 1.08 | 1.05 | 1.11 | 0.011630245 | 128777 | 2 |
| 313.3 | Autism | mental disorders | DR | 0.63 | 0.52 | 0.76 | 0.011646753 | 111475 | 1 |
| 242 | Thyrotoxicosis with or without goiter | endocrine/metabolic | DR | 1.10 | 1.06 | 1.15 | 0.01211719 | 128777 | 2 |
| 70.2 | Viral hepatitis B | infectious diseases | DR | 0.85 | 0.80 | 0.91 | 0.012154823 | 128777 | 2 |
| 1014 | Effects of heat, cold and air pressure | NULL | DR | 1.13 | 1.08 | 1.19 | 0.012162376 | 128777 | 2 |
| 728.2 | Laxity of ligament or hypermobility syndrome | musculoskeletal | DR | 0.76 | 0.68 | 0.85 | 0.0123381 | 128777 | 2 |
| 720 | Spinal stenosis | musculoskeletal | DR | 1.04 | 1.02 | 1.06 | 0.012640545 | 128777 | 2 |
| 971 | Poisoning by drugs primarily affecting the autonomic nervous system | injuries & poisonings | DR | 1.28 | 1.16 | 1.41 | 0.013053744 | 128777 | 2 |
| 504.1 | Idiopathic fibrosing alveolitis | respiratory | DR | 1.15 | 1.09 | 1.21 | 0.01315247 | 128777 | 2 |
| 800.4 | Fracture of patella | injuries & poisonings | DR | 1.18 | 1.10 | 1.26 | 0.013464317 | 128777 | 2 |
| 627.21 | | | DR | 0.67 | 0.57 | 0.79 | 0.014969961 | 128777 | 2 |
| 907 | Injuries to the nervous system | injuries & poisonings | DR | 0.92 | 0.89 | 0.95 | 0.015259671 | 128777 | 2 |
| 627 | Menopausal and postmenopausal disorders | genitourinary | DR | 1.16 | 1.09 | 1.23 | 0.01530357 | 128777 | 2 |
| 420.22 | Chronic pericarditis | circulatory system | DR | 1.28 | 1.15 | 1.41 | 0.015341983 | 128777 | 2 |
| 204.2 | Myeloid leukemia | neoplasms | DR | 1.23 | 1.13 | 1.35 | 0.016430709 | 128777 | 2 |
| 579 | Other symptoms involving abdomen and pelvis | digestive | DR | 1.06 | 1.04 | 1.09 | 0.016441168 | 128777 | 2 |
| 496.2 | Chronic bronchitis | respiratory | DR | 1.04 | 1.03 | 1.06 | 0.01667393 | 128777 | 2 |
| 313.2 | Tics and stuttering | mental disorders | DR | 0.79 | 0.72 | 0.87 | 0.016678715 | 111475 | 1 |
| 614.5 | Inflammatory disease of cervix, vagina, and vulva | genitourinary | DR | 0.85 | 0.79 | 0.91 | 0.017082775 | 128777 | 2 |
| 580.11 | Proliferative glomerulonephritis | genitourinary | DR | 1.46 | 1.24 | 1.71 | 0.017388113 | 111475 | 1 |
| 753.2 | Congenital anomalies of posterior segment of eye | congenital anomalies | DR | 1.21 | 1.12 | 1.31 | 0.017435287 | 128777 | 2 |
| 529 | Diseases and other conditions of the tongue | digestive | DR | 0.89 | 0.84 | 0.93 | 0.017820808 | 128777 | 2 |
| 755 | Congenital anomalies of limbs | congenital anomalies | DR | 1.06 | 1.03 | 1.09 | 0.018180146 | 111475 | 1 |
| 1007 | Injury to blood vessels | NULL | DR | 1.16 | 1.09 | 1.24 | 0.018201445 | 128777 | 2 |
| 1100 | Family history | NULL | DR | 0.95 | 0.92 | 0.97 | 0.018354146 | 128777 | 2 |
| 860 | Bone marrow or stem cell transplant | neoplasms | DR | 0.75 | 0.66 | 0.84 | 0.018493504 | 128777 | 2 |
| 333.2 | Myoclonus | neurological | DR | 1.14 | 1.08 | 1.21 | 0.018988508 | 128777 | 2 |
| 184.1 | Malignant neoplasm of ovary and other uterine adnexa | neoplasms | DR | 1.47 | 1.25 | 1.74 | 0.019048147 | 128777 | 2 |
| 973 | Poisoning by agents primarily affecting the gastrointestinal system | injuries & poisonings | DR | 1.60 | 1.31 | 1.96 | 0.019246062 | 111475 | 1 |
| 972.2 | | | DR | 1.34 | 1.18 | 1.51 | 0.019436579 | 111475 | 1 |
| 130 | Spirochetal infection | infectious diseases | DR | 1.11 | 1.06 | 1.16 | 0.01943821 | 111475 | 1 |
| 315.2 | Speech and language disorder | mental disorders | DR | 1.26 | 1.14 | 1.39 | 0.019540339 | 128777 | 2 |
| 726.2 | Synoviopathy | musculoskeletal | DR | 0.92 | 0.89 | 0.95 | 0.019611616 | 128777 | 2 |
| 214 | Lipoma | neoplasms | DR | 0.95 | 0.92 | 0.97 | 0.020562242 | 128777 | 2 |
| 654.1 | Abnormality of organs and soft tissues of pelvis complicating pregnancy, childbirth, or the puerperium | pregnancy complications | DR | 1.58 | 1.30 | 1.93 | 0.020863363 | 128777 | 2 |
| 752.11 | Spina bifida | congenital anomalies | DR | 0.55 | 0.42 | 0.71 | 0.021092677 | 111475 | 1 |
| 283.2 | Non-autoimmune hemolytic anemias | hematopoietic | DR | 1.47 | 1.24 | 1.73 | 0.021349687 | 111475 | 1 |
| 530.5 | Disorders of esophageal motility | digestive | DR | 1.12 | 1.07 | 1.18 | 0.021371739 | 128777 | 2 |
| 930 | Allergic reaction to food | injuries & poisonings | DR | 0.87 | 0.81 | 0.92 | 0.021552934 | 128777 | 2 |
| 706.3 | | | DR | 1.14 | 1.08 | 1.21 | 0.021716892 | 111475 | 1 |
| 575.7 | Other disorders of gallbladder | digestive | DR | 1.09 | 1.05 | 1.13 | 0.021745931 | 128777 | 2 |
| 755.1 | Congenital deformities of feet | congenital anomalies | DR | 1.06 | 1.03 | 1.09 | 0.023153351 | 128777 | 2 |
| 752.1 | Neural tube defects | congenital anomalies | DR | 0.67 | 0.56 | 0.80 | 0.023374425 | 111475 | 1 |
| 781.1 | Loss of height | symptoms | DR | 1.47 | 1.24 | 1.74 | 0.023390086 | 128777 | 2 |
| 420.1 | Myocarditis | circulatory system | DR | 1.44 | 1.23 | 1.69 | 0.023463188 | 128777 | 2 |
| 613 | Other nonmalignant breast conditions | genitourinary | DR | 1.10 | 1.05 | 1.14 | 0.023589 | 111475 | 1 |
| 529.6 | Glossodynia | digestive | DR | 0.73 | 0.63 | 0.84 | 0.023917288 | 128777 | 2 |
| 622.1 | Polyp of corpus uteri | genitourinary | DR | 0.72 | 0.62 | 0.83 | 0.024300315 | 128777 | 2 |
| 279 | Disorders involving the immune mechanism | endocrine/metabolic | DR | 1.14 | 1.08 | 1.21 | 0.024361514 | 128777 | 2 |
| 783.1 | Postprocedural fever | symptoms | DR | 1.20 | 1.11 | 1.30 | 0.024405487 | 128777 | 2 |
| 514.2 | Solitary pulmonary nodule | respiratory | DR | 1.05 | 1.02 | 1.07 | 0.02485188 | 128777 | 2 |
| 78 | Viral warts & HPV | infectious diseases | DR | 0.96 | 0.94 | 0.98 | 0.025096583 | 128777 | 2 |
| 286.12 | Congenital deficiency of other clotting factors (including factor VII) | hematopoietic | DR | 0.77 | 0.69 | 0.87 | 0.026126838 | 128777 | 2 |
| 530.6 | Diverticulum of esophagus, acquired | digestive | DR | 1.30 | 1.15 | 1.46 | 0.026789161 | 128777 | 2 |
| 345 | Epilepsy, recurrent seizures, convulsions | neurological | DR | 0.95 | 0.92 | 0.97 | 0.026978246 | 128777 | 2 |
| 729.1 | Rheumatism, unspecified and fibrositis | musculoskeletal | DR | 0.84 | 0.78 | 0.91 | 0.027573601 | 111475 | 1 |
| 755.3 | Congenital anomaly of fingers/toes | congenital anomalies | DR | 1.34 | 1.17 | 1.52 | 0.027608759 | 111475 | 1 |
| 586.11 | Small kidney | genitourinary | DR | 1.53 | 1.26 | 1.86 | 0.027628999 | 111475 | 1 |
| 736.2 | Acquired deformities of finger | musculoskeletal | DR | 0.86 | 0.80 | 0.92 | 0.028044775 | 128777 | 2 |
| 355 | Complex regional/central pain syndrome | neurological | DR | 0.90 | 0.86 | 0.94 | 0.028142025 | 128777 | 2 |
| 180.1 | Cervical cancer | neoplasms | DR | 1.44 | 1.22 | 1.70 | 0.028285842 | 128777 | 2 |
| 381.9 | Otorrhea | sense organs | DR | 1.14 | 1.07 | 1.21 | 0.02874416 | 128777 | 2 |
| 618 | Genital prolapse | genitourinary | DR | 1.22 | 1.11 | 1.34 | 0.02888465 | 128777 | 2 |
| 415.1 | Acute pulmonary heart disease | circulatory system | DR | 1.07 | 1.04 | 1.11 | 0.029012455 | 128777 | 2 |
| 253.7 | Other disorders of neurohypophysis | endocrine/metabolic | DR | 1.19 | 1.10 | 1.29 | 0.029122885 | 128777 | 2 |
| 527 | Diseases of the salivary glands | digestive | DR | 1.06 | 1.03 | 1.09 | 0.029332504 | 128777 | 2 |
| 451.2 | Phlebitis and thrombophlebitis of lower extremities | circulatory system | DR | 1.11 | 1.06 | 1.16 | 0.029444449 | 128777 | 2 |
| 750.1 | Upper gastrointestinal congenital anomalies | congenital anomalies | DR | 1.14 | 1.07 | 1.21 | 0.029563199 | 128777 | 2 |
| 389.2 | Conductive hearing loss | sense organs | DR | 1.07 | 1.04 | 1.11 | 0.029955421 | 128777 | 2 |
| 656.3 | Endocrine and metabolic disturbances of fetus and newborn | pregnancy complications | DR | 1.59 | 1.28 | 1.97 | 0.030361874 | 111475 | 1 |
| 286.6 | Defibrination syndrome | hematopoietic | DR | 1.36 | 1.18 | 1.57 | 0.030647242 | 128777 | 2 |
| 614.32 | Chronic inflammatory pelvic disease | genitourinary | DR | 0.60 | 0.47 | 0.76 | 0.030694667 | 128777 | 2 |
| 471 | Nasal polyps | respiratory | DR | 0.90 | 0.86 | 0.95 | 0.030727465 | 128777 | 2 |
| 574.12 | Cholelithiasis with other cholecystitis | digestive | DR | 1.08 | 1.04 | 1.12 | 0.031316557 | 128777 | 2 |
| 446.9 | Arteritis NOS | circulatory system | DR | 1.19 | 1.10 | 1.29 | 0.031477865 | 128777 | 2 |
| 1010.5 | potential health hazards related to communicable diseases potential health hazards related to communicable diseases | | DR | 1.03 | 1.02 | 1.05 | 0.032747805 | 128777 | 2 |
| 529.1 | Glossitis | digestive | DR | 0.82 | 0.75 | 0.90 | 0.033015966 | 128777 | 2 |
| 187.2 | Malignant neoplasm of testis | neoplasms | DR | 0.80 | 0.73 | 0.89 | 0.03302232 | 111475 | 1 |
| 242.3 | Exophthalmos | endocrine/metabolic | DR | 1.18 | 1.09 | 1.28 | 0.033591927 | 128777 | 2 |
| 567 | Peritonitis and retroperitoneal infections | digestive | DR | 1.09 | 1.05 | 1.13 | 0.033939864 | 128777 | 2 |
| 155 | Cancer of liver and intrahepatic bile duct | neoplasms | DR | 1.11 | 1.06 | 1.17 | 0.034151858 | 128777 | 2 |
| 41.21 | Rheumatic fever / chorea | infectious diseases | DR | 1.46 | 1.22 | 1.74 | 0.034555321 | 111475 | 1 |
| 8.51 | Intestinal e.coli | infectious diseases | DR | 1.48 | 1.23 | 1.78 | 0.034655832 | 111475 | 1 |
| 749.2 | Congenital anomalies of skull and face bones | congenital anomalies | DR | 1.98 | 1.43 | 2.75 | 0.034661996 | 111475 | 1 |
| 473.3 | Paralysis/spasm of vocal cords or larynx | respiratory | DR | 1.16 | 1.08 | 1.25 | 0.035394913 | 128777 | 2 |
| 226 | Benign neoplasm of thyroid glands | neoplasms | DR | 1.17 | 1.09 | 1.26 | 0.036718531 | 128777 | 2 |
| 709.3 | Systemic sclerosis | dermatologic | DR | 1.35 | 1.17 | 1.56 | 0.036788898 | 128777 | 2 |
| 278.4 | Abnormal weight gain | endocrine/metabolic | DR | 1.07 | 1.03 | 1.10 | 0.037122008 | 128777 | 2 |
| 79.1 | Varicella infection | infectious diseases | DR | 1.34 | 1.16 | 1.54 | 0.038424839 | 111475 | 1 |
| 782.6 | Pallor and flushing | symptoms | DR | 0.87 | 0.81 | 0.93 | 0.038582343 | 128777 | 2 |
| 644 | Anemia during pregnancy | pregnancy complications | DR | 1.90 | 1.39 | 2.60 | 0.038846074 | 128777 | 2 |
| 182 | Malignant neoplasm of uterus | neoplasms | DR | 1.31 | 1.15 | 1.49 | 0.039112384 | 128777 | 2 |
| 939.1 | Contact and allergic dermatitis of eyelid | dermatologic | DR | 1.22 | 1.11 | 1.35 | 0.03920546 | 128777 | 2 |
| 150 | Cancer of esophagus | neoplasms | DR | 1.16 | 1.08 | 1.25 | 0.039490738 | 128777 | 2 |

| | | | | | | | | | |
|---|---|---|---|---|---|---|---|---|---|
| 132.1 | Pediculosis and phthirus infestation | infectious diseases | DR | 0.72 | 0.61 | 0.85 | 0.039576357 | 111475 | 1 |
| 555.2 | Ulcerative colitis | digestive | DR | 0.92 | 0.88 | 0.96 | 0.040103349 | 128777 | 2 |
| 610.8 | Other specified benign mammary dysplasias | genitourinary | DR | 0.77 | 0.68 | 0.88 | 0.041145107 | 128777 | 2 |
| 701.3 | Circumscribed scleroderma | dermatologic | DR | 1.22 | 1.11 | 1.34 | 0.041658812 | 128777 | 2 |
| 614.52 | Vaginitis and vulvovaginitis | genitourinary | DR | 0.88 | 0.82 | 0.94 | 0.041719387 | 128777 | 2 |
| 626.21 | Mittelschmerz | genitourinary | DR | 0.12 | 0.04 | 0.34 | 0.041992866 | 111475 | 1 |
| 976 | Poisoning by agents primarily affecting skin & mucous membrane, ophthalmological, otorhinolaryngological, & dental drugs | injuries & poisonings | DR | 1.33 | 1.15 | 1.53 | 0.042304443 | 111475 | 1 |
| 117.4 | Aspergillosis | infectious diseases | DR | 0.76 | 0.66 | 0.87 | 0.042324744 | 128777 | 2 |
| 380.1 | Otitis externa | sense organs | DR | 1.04 | 1.02 | 1.06 | 0.042379987 | 128777 | 2 |
| 245.2 | Chronic thyroiditis | endocrine/metabolic | DR | 0.82 | 0.74 | 0.90 | 0.042509337 | 111475 | 1 |
| 735.1 | Flat foot | musculoskeletal | DR | 1.05 | 1.02 | 1.07 | 0.043024256 | 128777 | 2 |
| 540.11 | Acute appendicitis | digestive | DR | 0.88 | 0.83 | 0.94 | 0.044256729 | 128777 | 2 |
| 656.9 | Neonatal bradycardia or tachycardia | pregnancy complications | DR | 2.34 | 1.53 | 3.57 | 0.044310693 | 111475 | 1 |
| 701.4 | Keloid scar | dermatologic | DR | 1.10 | 1.05 | 1.16 | 0.044366977 | 128777 | 2 |
| 621 | Endometrial hyperplasia | genitourinary | DR | 1.31 | 1.14 | 1.49 | 0.044868087 | 128777 | 2 |
| 292.6 | Hallucinations | mental disorders | DR | 0.89 | 0.84 | 0.94 | 0.045286567 | 128777 | 2 |
| 989 | Toxic effect of other substances, chiefly nonmedicinal as to source | injuries & poisonings | DR | 1.13 | 1.06 | 1.20 | 0.045402203 | 128777 | 2 |
| 442.11 | Abdominal aortic aneurysm | circulatory system | DR | 1.06 | 1.03 | 1.09 | 0.045508743 | 128777 | 2 |
| 597.8 | Urethral hypermobility/ISD | genitourinary | DR | 0.69 | 0.57 | 0.83 | 0.045576998 | 128777 | 2 |
| 174.1 | Breast cancer [female] | neoplasms | DR | 1.19 | 1.09 | 1.29 | 0.046582543 | 128777 | 2 |
| 557 | Intestinal malabsorption (non-celiac) | digestive | DR | 1.17 | 1.08 | 1.27 | 0.047410965 | 128777 | 2 |
| 961 | Poisoning by other anti-infectives | injuries & poisonings | DR | 1.19 | 1.09 | 1.31 | 0.049442503 | 128777 | 2 |
| 444.5 | Atheroembolism | circulatory system | DR | 1.37 | 1.17 | 1.61 | 0.050833383 | 111475 | 1 |
| 303.31 | | | DR | 1.42 | 1.18 | 1.69 | 0.051237098 | 111475 | 1 |
| 724.1 | Disorders of sacrum | musculoskeletal | DR | 0.91 | 0.87 | 0.96 | 0.051736315 | 128777 | 2 |
| 913 | Toxic effect of venom | injuries & poisonings | DR | 0.90 | 0.85 | 0.95 | 0.05187467 | 128777 | 2 |
| 622.2 | Mucous polyp of cervix | genitourinary | DR | 0.70 | 0.58 | 0.84 | 0.052717502 | 128777 | 2 |
| 480.5 | Bronchopneumonia and lung abscess | respiratory | DR | 1.11 | 1.05 | 1.16 | 0.052961086 | 128777 | 2 |
| 286.1 | Congenital coagulation defects | hematopoietic | DR | 0.82 | 0.74 | 0.91 | 0.053041097 | 111475 | 1 |
| 962.1 | | | DR | 0.86 | 0.80 | 0.93 | 0.053601337 | 128777 | 2 |
| 958.2 | Traumatic and surgical subcutaneous emphysema | injuries & poisonings | DR | 0.73 | 0.62 | 0.86 | 0.054483796 | 128777 | 2 |
| 614.1 | Pelvic peritoneal adhesions, female (postoperative) (postinfection) | genitourinary | DR | 0.63 | 0.50 | 0.80 | 0.05549533 | 128777 | 2 |
| 270.11 | Disturbances of sulphur-bearing amino-acid metabolism | endocrine/metabolic | DR | 1.42 | 1.18 | 1.70 | 0.056222625 | 111475 | 1 |
| 694.1 | Vitiligo | dermatologic | DR | 1.19 | 1.08 | 1.30 | 0.056366573 | 128777 | 2 |
| 555.21 | Ulcerative colitis (chronic) | digestive | DR | 0.88 | 0.83 | 0.94 | 0.05766154 | 128777 | 2 |
| 220 | Benign neoplasm of ovary | neoplasms | DR | 0.69 | 0.57 | 0.84 | 0.057882855 | 128777 | 2 |
| 696.2 | Parapsoriasis | dermatologic | DR | 1.34 | 1.15 | 1.56 | 0.058182617 | 111475 | 1 |
| 536.7 | Complications of gastrostomy, colostomy and enterostomy | digestive | DR | 1.18 | 1.08 | 1.29 | 0.058318044 | 128777 | 2 |
| 384.4 | Perforation of tympanic membrane | sense organs | DR | 1.10 | 1.05 | 1.16 | 0.058690719 | 128777 | 2 |
| 292.5 | Transient alteration of awareness | mental disorders | DR | 1.09 | 1.04 | 1.14 | 0.058972574 | 128777 | 2 |
| 261.1 | Vitamin A deficiency | endocrine/metabolic | DR | 1.34 | 1.15 | 1.56 | 0.059783789 | 111475 | 1 |
| 565 | Anal and rectal conditions | digestive | DR | 0.96 | 0.94 | 0.98 | 0.059857986 | 128777 | 2 |
| 199.4 | Neurofibromatosis | neoplasms | DR | 0.75 | 0.65 | 0.87 | 0.059971931 | 111475 | 1 |
| 285.8 | Hemoglobinuria | hematopoietic | DR | 1.40 | 1.17 | 1.67 | 0.059986136 | 111475 | 1 |
| 577.2 | Chronic pancreatitis | digestive | DR | 0.92 | 0.88 | 0.96 | 0.060861596 | 128777 | 2 |
| 611.11 | Mammographic microcalcification | genitourinary | DR | 1.20 | 1.09 | 1.32 | 0.063554343 | 128777 | 2 |
| 686.3 | Pilonidal cyst | dermatologic | DR | 0.87 | 0.81 | 0.94 | 0.063569607 | 128777 | 2 |
| 70.9 | Hepatitis NOS | infectious diseases | DR | 0.92 | 0.88 | 0.96 | 0.06364554 | 128777 | 2 |
| 323.8 | Encephalitis, non-infectious | neurological | DR | 1.26 | 1.11 | 1.42 | 0.063999377 | 128777 | 2 |
| 31 | Diseases due to other mycobacteria | infectious diseases | DR | 1.22 | 1.09 | 1.35 | 0.065047859 | 128777 | 2 |
| 654 | Other and unspecified complications of birth; puerperium affecting management of mother | pregnancy complications | DR | 0.61 | 0.47 | 0.80 | 0.065128034 | 128777 | 2 |
| 557.1 | Celiac disease | digestive | DR | 1.20 | 1.09 | 1.33 | 0.066843258 | 128777 | 2 |
| 347 | Cataplexy and narcolepsy | neurological | DR | 0.85 | 0.77 | 0.93 | 0.067995582 | 128777 | 2 |
| 687.4 | Disturbance of skin sensation | dermatologic | DR | 0.97 | 0.96 | 0.99 | 0.068602322 | 128777 | 2 |
| 603.2 | Spermatocele | genitourinary | DR | 0.90 | 0.85 | 0.95 | 0.06891622 | 128777 | 2 |
| 225.2 | Benign neoplasm of spinal cord, meninges | neoplasms | DR | 0.67 | 0.54 | 0.84 | 0.069022979 | 111475 | 1 |
| 270.31 | Polyclonal hypergammaglobulinemia | endocrine/metabolic | DR | 1.48 | 1.19 | 1.83 | 0.071463362 | 111475 | 1 |
| 159.3 | Malignant neoplasm of gallbladder and extrahepatic bile ducts | neoplasms | DR | 1.24 | 1.10 | 1.40 | 0.071887888 | 128777 | 2 |
| 338.1 | Acute pain | neurological | DR | 1.03 | 1.01 | 1.05 | 0.072169001 | 128777 | 2 |
| 483 | Acute bronchitis and bronchiolitis | respiratory | DR | 1.03 | 1.01 | 1.04 | 0.072412681 | 128777 | 2 |
| 496.1 | Emphysema | respiratory | DR | 0.95 | 0.93 | 0.98 | 0.072419872 | 128777 | 2 |
| 710.2 | | | DR | 1.47 | 1.19 | 1.82 | 0.072443129 | 111475 | 1 |
| 281.13 | Folate-deficiency anemia | hematopoietic | DR | 1.16 | 1.07 | 1.26 | 0.073067127 | 128777 | 2 |
| 277.2 | Other disorders of purine and pyrimidine metabolism | endocrine/metabolic | DR | 2.19 | 1.41 | 3.39 | 0.073596036 | 111475 | 1 |
| 345.3 | Convulsions | neurological | DR | 0.95 | 0.93 | 0.98 | 0.074598588 | 128777 | 2 |
| 601.1 | Prostatitis | genitourinary | DR | 1.05 | 1.02 | 1.08 | 0.074661338 | 128777 | 2 |
| 244.3 | | | DR | 0.58 | 0.43 | 0.79 | 0.074974189 | 111475 | 1 |
| 358.1 | Myasthenia gravis | neurological | DR | 1.18 | 1.08 | 1.30 | 0.075894103 | 128777 | 2 |
| 521 | Diseases of hard tissues of teeth | digestive | DR | 0.98 | 0.96 | 0.99 | 0.075912921 | 111475 | 1 |
| 958.1 | Postoperative shock | injuries & poisonings | DR | 1.24 | 1.10 | 1.40 | 0.077512415 | 128777 | 2 |
| 345.11 | Generalized convulsive epilepsy | neurological | DR | 0.87 | 0.80 | 0.94 | 0.077982549 | 128777 | 2 |
| 348.4 | Cerebral cysts | neurological | DR | 0.80 | 0.70 | 0.91 | 0.078496184 | 128777 | 2 |
| 170.1 | Bone cancer | neoplasms | DR | 1.17 | 1.07 | 1.28 | 0.078907482 | 128777 | 2 |
| 255.22 | | | DR | 1.87 | 1.31 | 2.68 | 0.079484462 | 111475 | 1 |
| 448 | Disease of capillaries | circulatory system | DR | 1.13 | 1.05 | 1.21 | 0.079510698 | 111475 | 1 |
| 259.1 | Nonspecific abnormal results of other endocrine function study | endocrine/metabolic | DR | 0.84 | 0.77 | 0.93 | 0.080348586 | 128777 | 2 |
| 669 | Complications of labor and delivery NEC | pregnancy complications | DR | 1.54 | 1.20 | 1.96 | 0.08040408 | 128777 | 2 |
| 602.3 | Dysplasia of prostate | genitourinary | DR | 1.14 | 1.06 | 1.22 | 0.082347457 | 111475 | 1 |
| 528.41 | Cyst of the salivary gland | digestive | DR | 0.76 | 0.65 | 0.89 | 0.082780547 | 111475 | 1 |
| 722.3 | Schmorl's nodes | musculoskeletal | DR | 0.75 | 0.63 | 0.88 | 0.083726883 | 111475 | 1 |
| 634.3 | Ectopic pregnancy | pregnancy complications | DR | 0.26 | 0.12 | 0.57 | 0.085222095 | 111475 | 1 |
| 985 | Toxic effect of other metals | injuries & poisonings | DR | 1.22 | 1.09 | 1.37 | 0.085524086 | 111475 | 1 |
| 539 | Bariatric surgery | digestive | DR | 0.92 | 0.88 | 0.97 | 0.085796536 | 128777 | 2 |
| 385 | Other disorders of middle ear and mastoid | sense organs | DR | 1.11 | 1.04 | 1.18 | 0.08627521 | 128777 | 2 |
| 305.2 | Eating disorder | mental disorders | DR | 0.89 | 0.83 | 0.95 | 0.086482941 | 128777 | 2 |
| 184.2 | Cancer of other female genital organs (excluding uterus and ovary) | neoplasms | DR | 1.38 | 1.14 | 1.66 | 0.086937392 | 128777 | 2 |
| 751.3 | Obstructive genitourinary defect | congenital anomalies | DR | 1.30 | 1.11 | 1.51 | 0.087356773 | 128777 | 2 |
| 745 | Pain in joint | musculoskeletal | DR | 0.97 | 0.96 | 0.99 | 0.088364998 | 128777 | 2 |
| 381.11 | Suppurative and unspecified otitis media | sense organs | DR | 0.96 | 0.94 | 0.99 | 0.089163344 | 128777 | 2 |
| 415.11 | Pulmonary embolism and infarction, acute | circulatory system | DR | 1.05 | 1.02 | 1.09 | 0.09029344 | 128777 | 2 |
| 215 | Other benign neoplasm of connective and other soft tissue | neoplasms | DR | 0.93 | 0.89 | 0.97 | 0.090820467 | 128777 | 2 |
| 653 | Problems associated with amniotic cavity and membranes | pregnancy complications | DR | 1.46 | 1.17 | 1.83 | 0.090920174 | 128777 | 2 |

| | | | | | | | | | |
|---|---|---|---|---|---|---|---|---|---|
| 614.54 | Abscess or ulceration of vulva | genitourinary | DR | 1.36 | 1.13 | 1.63 | 0.091021691 | 128777 | 2 |
| 594.1 | Calculus of kidney | genitourinary | DR | 0.97 | 0.95 | 0.99 | 0.091125446 | 128777 | 2 |
| 331.1 | Hydrocephalus | neurological | DR | 1.15 | 1.06 | 1.24 | 0.091497732 | 128777 | 2 |
| 386.3 | Labyrinthitis | sense organs | DR | 1.10 | 1.04 | 1.17 | 0.094151359 | 128777 | 2 |
| 711.2 | Reiter's disease | musculoskeletal | DR | 0.76 | 0.64 | 0.90 | 0.094795616 | 111475 | 1 |
| 270.34 | Alpha-1-antitrypsin deficiency | endocrine/metabolic | DR | 0.67 | 0.53 | 0.85 | 0.094829754 | 111475 | 1 |
| 528.7 | Sialolithiasis | digestive | DR | 0.83 | 0.74 | 0.93 | 0.094863777 | 111475 | 1 |
| 250.5 | Glycosuria or Acetonuria | endocrine/metabolic | DR | 1.16 | 1.06 | 1.26 | 0.094917757 | 128777 | 2 |
| 647.3 | Major puerperal infection | pregnancy complications | DR | 1.96 | 1.31 | 2.92 | 0.095810489 | 111475 | 1 |
| 452.8 | Postphlebitic syndrome | circulatory system | DR | 1.26 | 1.10 | 1.44 | 0.096597014 | 128777 | 2 |
| 604.3 | Peyronie's disease | genitourinary | DR | 0.90 | 0.84 | 0.96 | 0.096796257 | 128777 | 2 |
| 665 | Obstetrical/birth trauma | pregnancy complications | DR | 0.77 | 0.66 | 0.90 | 0.100314372 | 128777 | 2 |
| 696.42 | Psoriatic arthropathy | dermatologic | DR | 0.91 | 0.85 | 0.96 | 0.100339148 | 128777 | 2 |
| 736.4 | Genu valgum or varum (acquired) | musculoskeletal | DR | 0.89 | 0.83 | 0.96 | 0.100640015 | 128777 | 2 |
| 663 | Umbilical cord complications during labor and delivery | pregnancy complications | DR | 0.57 | 0.41 | 0.80 | 0.100728514 | 128777 | 2 |
| 255.1 | Adrenal hyperfunction | endocrine/metabolic | DR | 1.14 | 1.05 | 1.23 | 0.100804526 | 111475 | 1 |
| 320 | Meningitis | neurological | DR | 1.13 | 1.05 | 1.23 | 0.100896299 | 128777 | 2 |
| 705.1 | Dyshidrosis | dermatologic | DR | 1.09 | 1.03 | 1.15 | 0.100916974 | 128777 | 2 |
| 535.1 | Acute gastritis | digestive | DR | 1.06 | 1.02 | 1.11 | 0.101167464 | 128777 | 2 |
| 634 | Miscarriage; stillbirth | pregnancy complications | DR | 0.79 | 0.69 | 0.91 | 0.101755 | 128777 | 2 |
| 603.1 | Hydrocele | genitourinary | DR | 1.06 | 1.02 | 1.10 | 0.10201556 | 128777 | 2 |
| 259.8 | Polyglandular activity in multiple endocrine adenomatosis | endocrine/metabolic | DR | 0.49 | 0.32 | 0.76 | 0.102117635 | 111475 | 1 |
| 521.2 | Dental abrasion, erosion and attrition | digestive | DR | 1.04 | 1.02 | 1.07 | 0.104380514 | 111475 | 1 |
| 201 | Hodgkin's disease | neoplasms | DR | 0.86 | 0.78 | 0.94 | 0.104497636 | 128777 | 2 |
| 384.1 | Myringitis | sense organs | DR | 0.81 | 0.71 | 0.92 | 0.10479963 | 128777 | 2 |
| 270 | | | DR | 1.05 | 1.02 | 1.09 | 0.104920256 | 111475 | 1 |
| 535.6 | Duodenitis | digestive | DR | 1.06 | 1.02 | 1.10 | 0.105223971 | 128777 | 2 |
| 855 | | | DR | 0.81 | 0.70 | 0.92 | 0.108998605 | 128777 | 2 |
| 701.6 | Acquired acanthosis nigricans | dermatologic | DR | 1.17 | 1.06 | 1.29 | 0.110395931 | 128777 | 2 |
| 149.9 | Cancer of of nasal cavities | neoplasms | DR | 1.24 | 1.08 | 1.41 | 0.11140177 | 128777 | 2 |
| 260.1 | Cachexia | endocrine/metabolic | DR | 1.14 | 1.05 | 1.24 | 0.111456825 | 128777 | 2 |
| 381.2 | Eustachian tube disorders | sense organs | DR | 0.96 | 0.93 | 0.98 | 0.113047988 | 128777 | 2 |
| 941 | Adverse reaction to serum or vaccine | injuries & poisonings | DR | 1.19 | 1.06 | 1.32 | 0.115048991 | 111475 | 1 |
| 242.1 | Graves' disease | endocrine/metabolic | DR | 0.88 | 0.82 | 0.96 | 0.115784873 | 128777 | 2 |
| 636.1 | Threatened premature labor | pregnancy complications | DR | 0.54 | 0.37 | 0.80 | 0.12060244 | 128777 | 2 |
| 251.8 | Abnormality of secretion of glucagon or gastrin | endocrine/metabolic | DR | 1.61 | 1.18 | 2.20 | 0.121621892 | 111475 | 1 |
| 283.1 | Autoimmune hemolytic anemias | hematopoietic | DR | 1.29 | 1.10 | 1.53 | 0.121653858 | 111475 | 1 |
| 324.1 | Jakob-Creutzfeldt disease | neurological | DR | 2.04 | 1.28 | 3.23 | 0.123322746 | 111475 | 1 |
| 165.1 | Cancer of bronchus; lung | neoplasms | DR | 0.95 | 0.92 | 0.98 | 0.124207675 | 128777 | 2 |
| 873 | Broken tooth | injuries & poisonings | DR | 1.06 | 1.02 | 1.10 | 0.124979234 | 128777 | 2 |
| 619 | Noninflammatory female genital disorders | genitourinary | DR | 0.90 | 0.84 | 0.96 | 0.124983191 | 128777 | 2 |
| 528.6 | Leukoplakia of oral mucosa | digestive | DR | 0.91 | 0.86 | 0.97 | 0.126452506 | 128777 | 2 |
| 520.2 | Disturbances in tooth eruption | digestive | DR | 0.93 | 0.89 | 0.98 | 0.128282978 | 128777 | 2 |
| 253.11 | Acromegaly and gigantism | endocrine/metabolic | DR | 0.67 | 0.51 | 0.87 | 0.129186102 | 111475 | 1 |
| 655 | Known or suspected fetal abnormality affecting management of mother | pregnancy complications | DR | 1.28 | 1.09 | 1.50 | 0.129830092 | 128777 | 2 |
| 10 | Tuberculosis | infectious diseases | DR | 0.91 | 0.85 | 0.97 | 0.131715289 | 128777 | 2 |
| 350.6 | Disturbances of sensation of smell and taste | neurological | DR | 1.10 | 1.03 | 1.18 | 0.13648739 | 128777 | 2 |
| 289.1 | Myelofibrosis | hematopoietic | DR | 1.36 | 1.10 | 1.67 | 0.13721477 | 128777 | 2 |
| 728.1 | Muscular calcification and ossification | musculoskeletal | DR | 1.17 | 1.05 | 1.29 | 0.137931283 | 128777 | 2 |
| 592.2 | Urethritis and urethral syndrome | genitourinary | DR | 0.90 | 0.83 | 0.96 | 0.138170583 | 128777 | 2 |
| 198.2 | Secondary malignancy of respiratory organs | neoplasms | DR | 0.92 | 0.87 | 0.97 | 0.138876268 | 128777 | 2 |
| 758 | Chromosomal anomalies and genetic disorders | congenital anomalies | DR | 0.80 | 0.68 | 0.93 | 0.13912462 | 111475 | 1 |
| 651 | Multiple gestation | pregnancy complications | DR | 0.20 | 0.07 | 0.60 | 0.141925133 | 111475 | 1 |
| 627.5 | Premature menopause and other ovarian failure | genitourinary | DR | 0.82 | 0.72 | 0.94 | 0.142149339 | 128777 | 2 |
| 277.8 | Carnitine deficiencies | endocrine/metabolic | DR | 5.17 | 1.69 | 15.84 | 0.142201423 | 111475 | 1 |
| 498 | Acute bronchospasm | respiratory | DR | 0.93 | 0.88 | 0.98 | 0.142311601 | 128777 | 2 |
| 709.2 | Sicca syndrome | dermatologic | DR | 0.92 | 0.87 | 0.97 | 0.14261345 | 128777 | 2 |
| 145 | Cancer of mouth | neoplasms | DR | 1.07 | 1.02 | 1.13 | 0.143171427 | 128777 | 2 |
| 750.15 | Congenital anomalies of stomach | congenital anomalies | DR | 1.22 | 1.06 | 1.39 | 0.144898304 | 111475 | 1 |
| 8.7 | Intestinal infection due to protozoa | infectious diseases | DR | 1.28 | 1.08 | 1.51 | 0.146928333 | 111475 | 1 |
| 384 | Other disorders of tympanic membrane | sense organs | DR | 1.07 | 1.02 | 1.12 | 0.146980986 | 128777 | 2 |
| 573.2 | Liver replaced by transplant | digestive | DR | 1.13 | 1.04 | 1.24 | 0.147567133 | 128777 | 2 |
| 289.5 | Diseases of spleen | hematopoietic | DR | 1.09 | 1.03 | 1.15 | 0.148294971 | 128777 | 2 |
| 345.1 | Epilepsy | neurological | DR | 0.95 | 0.92 | 0.99 | 0.151580244 | 128777 | 2 |
| 695.4 | Lupus (localized and systemic) | dermatologic | DR | 0.86 | 0.78 | 0.96 | 0.152316904 | 111475 | 1 |
| 272.12 | Hyperglyceridemia | endocrine/metabolic | DR | 0.97 | 0.95 | 0.99 | 0.15282496 | 128777 | 2 |
| 364.9 | Cornea replaced by transplant | sense organs | DR | 1.14 | 1.04 | 1.25 | 0.153036403 | 128777 | 2 |
| 565.1 | Anal and rectal polyp | digestive | DR | 0.96 | 0.93 | 0.99 | 0.155350932 | 128777 | 2 |
| 792.1 | Papanicolaou smear of cervix or vagina with atypical squamous cells | genitourinary | DR | 0.86 | 0.78 | 0.96 | 0.156903077 | 128777 | 2 |
| 647.1 | Infections of genitourinary tract during pregnancy | pregnancy complications | DR | 1.30 | 1.08 | 1.56 | 0.157675408 | 128777 | 2 |
| 694.3 | Vascular disorders of skin | dermatologic | DR | 1.16 | 1.04 | 1.29 | 0.158988866 | 111475 | 1 |
| 198.1 | Secondary malignancy of lymph nodes | neoplasms | DR | 0.95 | 0.91 | 0.98 | 0.160151244 | 128777 | 2 |
| 721.8 | Other allied disorders of spine | musculoskeletal | DR | 1.06 | 1.02 | 1.11 | 0.161061949 | 128777 | 2 |
| 259 | Other endocrine disorders | endocrine/metabolic | DR | 0.93 | 0.89 | 0.98 | 0.163479256 | 128777 | 2 |
| 726 | Peripheral enthesopathies and allied syndromes | musculoskeletal | DR | 1.02 | 1.00 | 1.03 | 0.166857607 | 128777 | 2 |
| 611 | Abnormal findings on mammogram or breast exam | genitourinary | DR | 1.05 | 1.01 | 1.09 | 0.167984903 | 111475 | 1 |
| 736.1 | Acquired deformities of forearm | musculoskeletal | DR | 1.14 | 1.04 | 1.25 | 0.169117973 | 111475 | 1 |
| 614.31 | Acute inflammatory pelvic disease | genitourinary | DR | 0.49 | 0.29 | 0.82 | 0.169655449 | 111475 | 1 |
| 715.3 | Spinal enthesopathy | musculoskeletal | DR | 0.85 | 0.76 | 0.96 | 0.171488591 | 128777 | 2 |
| 378.2 | Nystagmus and other irregular eye movements | sense organs | DR | 1.12 | 1.03 | 1.21 | 0.172827821 | 128777 | 2 |
| 716.1 | | | DR | 1.11 | 1.03 | 1.20 | 0.173709264 | 128777 | 2 |
| 174.3 | Neoplasm of uncertain behavior of breast | neoplasms | DR | 1.20 | 1.05 | 1.38 | 0.175140653 | 128777 | 2 |
| 159 | Malignant neoplasm of other and ill-defined sites within the digestive organs and peritoneum | neoplasms | DR | 1.08 | 1.02 | 1.15 | 0.175313294 | 128777 | 2 |
| 568 | Other disorders of peritoneum | digestive | DR | 1.06 | 1.02 | 1.11 | 0.176416744 | 128777 | 2 |
| 748 | Anomalies of respiratory system, congenital | congenital anomalies | DR | 1.22 | 1.05 | 1.41 | 0.179915271 | 128777 | 2 |
| 983 | Toxic effect of corrosive aromatics, acids, and caustic alkalis | injuries & poisonings | DR | 1.25 | 1.06 | 1.47 | 0.180061446 | 111475 | 1 |
| 767 | | | DR | 0.86 | 0.77 | 0.96 | 0.181351493 | 111475 | 1 |
| 691.1 | Ichthyosis congenita | dermatologic | DR | 1.23 | 1.05 | 1.43 | 0.18330361 | 111475 | 1 |
| 303.3 | Psychogenic disorder | mental disorders | DR | 0.92 | 0.86 | 0.98 | 0.183742319 | 128777 | 2 |
| 614.4 | Inflammatory diseases of uterus, except cervix | genitourinary | DR | 0.66 | 0.49 | 0.90 | 0.185788759 | 128777 | 2 |
| 210 | Benign neoplasm of lip, oral cavity, and pharynx | neoplasms | DR | 0.95 | 0.91 | 0.99 | 0.186284435 | 128777 | 2 |
| 196 | | | DR | 1.07 | 1.02 | 1.12 | 0.187231247 | 128777 | 2 |
| 272.14 | Hyperchylomicronemia | endocrine/metabolic | DR | 1.25 | 1.05 | 1.48 | 0.188797098 | 111475 | 1 |
| 912 | Insect bite | injuries & poisonings | DR | 1.04 | 1.01 | 1.07 | 0.188809842 | 128777 | 2 |
| 575.8 | Other disorders of biliary tract | digestive | DR | 1.07 | 1.02 | 1.12 | 0.18895809 | 128777 | 2 |
| 334.2 | Anterior horn cell disease | neurological | DR | 1.14 | 1.03 | 1.27 | 0.191885579 | 111475 | 1 |

| | | | | | | | | | |
|---|---|---|---|---|---|---|---|---|---|
| 643 | Excessive vomiting in pregnancy | pregnancy complications | DR | 1.54 | 1.11 | 2.14 | 0.192462658 | 128777 | 2 |
| 522 | Diseases of pulp and periapical tissues | digestive | DR | 0.97 | 0.95 | 0.99 | 0.194565241 | 111475 | 1 |
| 258 | Iatrogenic endocrine disorders | endocrine/metabolic | DR | 0.83 | 0.72 | 0.96 | 0.194694523 | 128777 | 2 |
| 159.2 | Malignant neoplasm of small intestine, including duodenum | neoplasms | DR | 0.85 | 0.76 | 0.97 | 0.196310633 | 128777 | 2 |
| 686.2 | Impetigo | dermatologic | DR | 1.08 | 1.02 | 1.14 | 0.19645514 | 128777 | 2 |
| 635.3 | Placenta previa and abruptio placenta | pregnancy complications | DR | 0.22 | 0.07 | 0.71 | 0.196636118 | 111475 | 1 |
| 652 | Malposition and malpresentation of fetus or obstruction | pregnancy complications | DR | 1.25 | 1.05 | 1.48 | 0.197458821 | 128777 | 2 |
| 255.13 | Medulloadrenal hyperfunction | endocrine/metabolic | DR | 0.44 | 0.23 | 0.83 | 0.197607052 | 111475 | 1 |
| 149.5 | | | DR | 0.87 | 0.77 | 0.97 | 0.19929611 | 128777 | 2 |
| 573.9 | Abnormal serum enzyme levels | digestive | DR | 1.04 | 1.01 | 1.06 | 0.203611989 | 111475 | 1 |
| 495.1 | | | DR | 0.95 | 0.92 | 0.99 | 0.204483678 | 128777 | 2 |
| 255.11 | Cushing's syndrome | endocrine/metabolic | DR | 0.85 | 0.75 | 0.97 | 0.205494495 | 128777 | 2 |
| 610.2 | Fibroadenosis of breast | genitourinary | DR | 0.81 | 0.69 | 0.96 | 0.209808862 | 128777 | 2 |
| 447.7 | Aortic ectasia | circulatory system | DR | 1.07 | 1.01 | 1.13 | 0.210126591 | 128777 | 2 |
| 381.1 | Otitis media | sense organs | DR | 0.98 | 0.96 | 1.00 | 0.210228499 | 128777 | 2 |
| 286.13 | Congenital factor VIII disorder | hematopoietic | DR | 0.69 | 0.52 | 0.93 | 0.211281128 | 111475 | 1 |
| 252.2 | Hypoparathyroidism | endocrine/metabolic | DR | 1.15 | 1.03 | 1.29 | 0.212011163 | 128777 | 2 |
| 601.12 | Chronic prostatitis | genitourinary | DR | 1.05 | 1.01 | 1.10 | 0.214101754 | 128777 | 2 |
| 751.22 | Other specified congenital anomalies of kidney | congenital anomalies | DR | 1.13 | 1.02 | 1.25 | 0.214748616 | 128777 | 2 |
| 187.1 | Malignant neoplasm of unspecified male genital organ | neoplasms | DR | 1.14 | 1.02 | 1.26 | 0.215789339 | 111475 | 1 |
| 575.9 | Nonspecific abnormal findings on radiological and other examination of biliary tract | digestive | DR | 0.95 | 0.92 | 0.99 | 0.216498992 | 128777 | 2 |
| 803.21 | | | DR | 1.16 | 1.03 | 1.30 | 0.218325967 | 128777 | 2 |
| 975 | Poisoning by agents primarily acting on the smooth and skeletal muscles and respiratory system | injuries & poisonings | DR | 0.81 | 0.69 | 0.96 | 0.219468194 | 111475 | 1 |
| 145.5 | Cancer of the mouth floor | neoplasms | DR | 1.26 | 1.04 | 1.52 | 0.219709228 | 111475 | 1 |
| 198.6 | Secondary malignancy of bone | neoplasms | DR | 1.06 | 1.01 | 1.12 | 0.219836545 | 128777 | 2 |
| 618.6 | Vaginal enterocele, congenital or acquired | genitourinary | DR | 0.67 | 0.48 | 0.93 | 0.220356821 | 128777 | 2 |
| 966 | Poisoning by anticonvulsants and anti-Parkinsonism drugs | injuries & poisonings | DR | 0.90 | 0.82 | 0.98 | 0.222408818 | 128777 | 2 |
| 646 | Other complications of pregnancy NEC | pregnancy complications | DR | 1.28 | 1.05 | 1.56 | 0.222470661 | 128777 | 2 |
| 1010.4 | Genetic Test | | DR | 0.78 | 0.64 | 0.96 | 0.222679191 | 128777 | 2 |
| 202 | Cancer of other lymphoid, histiocytic tissue | neoplasms | DR | 1.05 | 1.01 | 1.10 | 0.224051918 | 128777 | 2 |
| 198.7 | Secondary malignant neoplasm of skin | neoplasms | DR | 1.19 | 1.03 | 1.38 | 0.224645721 | 128777 | 2 |
| 348.2 | Cerebral edema and compression of brain | neurological | DR | 1.08 | 1.01 | 1.16 | 0.224884302 | 128777 | 2 |
| 360.2 | Progressive myopia | sense organs | DR | 1.10 | 1.02 | 1.18 | 0.225535402 | 128777 | 2 |
| 110.2 | Dermatomycoses | infectious diseases | DR | 1.05 | 1.01 | 1.10 | 0.226189998 | 128777 | 2 |
| 315.3 | Mental retardation | mental disorders | DR | 0.79 | 0.65 | 0.96 | 0.22779772 | 128777 | 2 |
| 526.3 | Anomalies of jaw size/symmetry | digestive | DR | 0.86 | 0.76 | 0.97 | 0.229797254 | 128777 | 2 |
| 604.2 | | | DR | 1.37 | 1.05 | 1.79 | 0.232851788 | 111475 | 1 |
| 528.5 | Diseases of lips | digestive | DR | 1.08 | 1.01 | 1.15 | 0.2339007 | 128777 | 2 |
| 256.1 | Hyperestrogenism | endocrine/metabolic | DR | 0.27 | 0.09 | 0.82 | 0.234788871 | 111475 | 1 |
| 737 | Curvature of spine | musculoskeletal | DR | 1.05 | 1.01 | 1.09 | 0.235704195 | 128777 | 2 |
| 714.2 | Juvenile rheumatoid arthritis | musculoskeletal | DR | 0.76 | 0.60 | 0.96 | 0.236034858 | 111475 | 1 |
| 341 | Other demyelinating diseases of central nervous system | neurological | DR | 0.86 | 0.75 | 0.98 | 0.236723826 | 128777 | 2 |
| 289.9 | Abnormality of red blood cells | hematopoietic | DR | 1.07 | 1.01 | 1.14 | 0.237110273 | 128777 | 2 |
| 656.5 | Hematological disorders of newborn | pregnancy complications | DR | 0.47 | 0.24 | 0.89 | 0.237563811 | 111475 | 1 |
| 184 | Cancer of other female genital organs | neoplasms | DR | 1.16 | 1.02 | 1.31 | 0.237573511 | 128777 | 2 |
| 524.3 | Anomalies of tooth position/malocclusion | digestive | DR | 1.05 | 1.01 | 1.09 | 0.237969785 | 128777 | 2 |
| 575.2 | Obstruction of bile duct | digestive | DR | 0.92 | 0.86 | 0.99 | 0.241871772 | 128777 | 2 |
| 611.1 | Abnormal mammogram | genitourinary | DR | 0.95 | 0.90 | 0.99 | 0.243561283 | 128777 | 2 |
| 594.2 | Calculus of lower urinary tract | genitourinary | DR | 1.06 | 1.01 | 1.11 | 0.244484188 | 128777 | 2 |
| 741.1 | Ankylosis of joint | musculoskeletal | DR | 1.10 | 1.01 | 1.20 | 0.245642795 | 128777 | 2 |
| 495.11 | | | DR | 0.93 | 0.88 | 0.99 | 0.246175115 | 128777 | 2 |
| 736.6 | Unequal leg length (acquired) | musculoskeletal | DR | 1.05 | 1.01 | 1.09 | 0.247421358 | 128777 | 2 |
| 145.1 | Cancer of lip | neoplasms | DR | 1.16 | 1.02 | 1.32 | 0.247438555 | 111475 | 1 |
| 656.7 | Conditions involving the integument and temperature regulation of fetus and newborn | pregnancy complications | DR | 1.31 | 1.04 | 1.66 | 0.247860056 | 111475 | 1 |
| 279.2 | Autoimmune disease NEC | endocrine/metabolic | DR | 0.80 | 0.66 | 0.97 | 0.247948323 | 111475 | 1 |
| 586.1 | Anatomical abnormatilies of kidney and ureters | genitourinary | DR | 1.18 | 1.02 | 1.36 | 0.248498297 | 111475 | 1 |
| 227.1 | Benign neoplasm of adrenal gland | neoplasms | DR | 1.06 | 1.01 | 1.11 | 0.248848341 | 128777 | 2 |
| 300.13 | Phobia | mental disorders | DR | 1.07 | 1.01 | 1.13 | 0.250907355 | 128777 | 2 |
| 668 | Complications of the administration of anesthetic or other sedation in labor and delivery | pregnancy complications | DR | 2.38 | 1.12 | 5.07 | 0.251956098 | 111475 | 1 |
| 446.7 | Takayasu's disease | circulatory system | DR | 1.53 | 1.06 | 2.23 | 0.252935689 | 111475 | 1 |
| 986 | Toxic effect of carbon monoxide | injuries & poisonings | DR | 0.75 | 0.58 | 0.96 | 0.253819462 | 111475 | 1 |
| 165 | Cancer within the respiratory system | neoplasms | DR | 0.96 | 0.93 | 1.00 | 0.257423624 | 128777 | 2 |
| 622 | Polyp of female genital organs | genitourinary | DR | 0.86 | 0.75 | 0.98 | 0.257855489 | 111475 | 1 |
| 988 | Toxic effect of noxious substances eaten as food | injuries & poisonings | DR | 1.34 | 1.03 | 1.74 | 0.258167876 | 111475 | 1 |
| 870.4 | Open wound of nose and sinus | injuries & poisonings | DR | 1.12 | 1.01 | 1.23 | 0.258260801 | 128777 | 2 |
| 709.4 | Polymyositis | dermatologic | DR | 1.28 | 1.03 | 1.59 | 0.26103865 | 111475 | 1 |
| 724.2 | | | DR | 1.09 | 1.01 | 1.18 | 0.262312158 | 128777 | 2 |
| 465 | Acute upper respiratory infections of multiple or unspecified sites | respiratory | DR | 1.01 | 1.00 | 1.03 | 0.263429952 | 128777 | 2 |
| 592.21 | Urethral syndrome | genitourinary | DR | 1.30 | 1.03 | 1.64 | 0.26475714 | 111475 | 1 |
| 218.2 | Other benign neoplasm of uterus | neoplasms | DR | 0.75 | 0.58 | 0.97 | 0.265844366 | 111475 | 1 |
| 444.2 | Embolism and thrombosis of abdominal aorta | circulatory system | DR | 1.12 | 1.01 | 1.24 | 0.267213548 | 128777 | 2 |
| 656.26 | Transitory tachypnea or apnea of newborn | pregnancy complications | DR | 0.65 | 0.44 | 0.96 | 0.267488536 | 111475 | 1 |
| 709.7 | Unspecified diffuse connective tissue disease | dermatologic | DR | 0.86 | 0.76 | 0.99 | 0.267777339 | 128777 | 2 |
| 288.11 | Neutropenia | hematopoietic | DR | 1.05 | 1.00 | 1.10 | 0.267899546 | 128777 | 2 |
| 371.9 | Chronic inflammatory disorders of orbit | sense organs | DR | 1.30 | 1.03 | 1.65 | 0.268762795 | 111475 | 1 |
| 292.12 | Symbolic dysfunction | mental disorders | DR | 1.11 | 1.01 | 1.23 | 0.268923413 | 128777 | 2 |
| 284.2 | Constitutional aplastic anemia | hematopoietic | DR | 1.84 | 1.06 | 3.21 | 0.271077531 | 111475 | 1 |
| 756.21 | Pectus excavatum | congenital anomalies | DR | 2.01 | 1.06 | 3.79 | 0.271909726 | 111475 | 1 |
| 627.1 | Postmenopausal bleeding | genitourinary | DR | 1.09 | 1.01 | 1.19 | 0.271926004 | 128777 | 2 |
| 229 | Benign neoplasm of unspecified sites | neoplasms | DR | 1.06 | 1.01 | 1.12 | 0.272430114 | 128777 | 2 |
| 191.11 | Cancer of brain | neoplasms | DR | 0.91 | 0.83 | 0.99 | 0.273311808 | 128777 | 2 |
| 465.4 | Acute laryngitis and tracheitis | respiratory | DR | 0.94 | 0.89 | 0.99 | 0.27333684 | 128777 | 2 |
| 174 | Breast cancer | neoplasms | DR | 1.08 | 1.01 | 1.15 | 0.278906687 | 128777 | 2 |
| 749 | Congenital anomalies of face and neck | congenital anomalies | DR | 1.09 | 1.01 | 1.18 | 0.279990696 | 128777 | 2 |
| 277.6 | Other deficiencies of circulating enzymes | endocrine/metabolic | DR | 0.73 | 0.54 | 0.98 | 0.281728445 | 111475 | 1 |
| 260.21 | Kwashiorkor | endocrine/metabolic | DR | 1.33 | 1.02 | 1.73 | 0.281931353 | 111475 | 1 |
| 695.1 | Toxic erythema | dermatologic | DR | 1.15 | 1.01 | 1.30 | 0.287013688 | 111475 | 1 |
| 521.4 | Tooth complications likely association with other diseases | digestive | DR | 0.79 | 0.63 | 0.99 | 0.287095797 | 111475 | 1 |
| 704 | Diseases of hair and hair follicles | dermatologic | DR | 0.98 | 0.96 | 1.00 | 0.287926052 | 128777 | 2 |
| 577.3 | Cyst and pseudocyst of pancreas | digestive | DR | 1.06 | 1.00 | 1.11 | 0.28844547 | 128777 | 2 |
| 520.1 | Hereditary disturbances in tooth structure | digestive | DR | 1.05 | 1.00 | 1.10 | 0.288884989 | 111475 | 1 |
| 704.8 | Other specified diseases of hair and hair follicles | dermatologic | DR | 0.97 | 0.95 | 1.00 | 0.289491024 | 128777 | 2 |
| 636.2 | Early onset of delivery | pregnancy complications | DR | 1.29 | 1.01 | 1.65 | 0.292174977 | 128777 | 2 |
| 202.23 | Lymphosarcoma | neoplasms | DR | 1.16 | 1.01 | 1.34 | 0.294166245 | 128777 | 2 |

| | | | | | | | | | |
|---|---|---|---|---|---|---|---|---|---|
| 755.4 | Congenital anomalies of upper limb, including shoulder girdle | congenital anomalies | DR | 0.80 | 0.64 | 0.99 | 0.294470775 | 111475 | 1 |
| 278.3 | Localized adiposity | endocrine/metabolic | DR | 1.09 | 1.00 | 1.17 | 0.295460526 | 128777 | 2 |
| 446.2 | Acute febrile mucocutaneous lymph node syndrome (Kawasaki disease) | circulatory system | DR | 0.64 | 0.41 | 0.98 | 0.297351895 | 111475 | 1 |
| 525.2 | Atrophy of edentulous alveolar ridge | digestive | DR | 1.04 | 1.00 | 1.08 | 0.298675436 | 111475 | 1 |
| 254 | Diseases of thymus gland | endocrine/metabolic | DR | 0.78 | 0.62 | 0.99 | 0.300321478 | 111475 | 1 |
| 527.2 | Sialoadenitis | digestive | DR | 0.95 | 0.90 | 1.00 | 0.304641359 | 128777 | 2 |
| 277.51 | Lipoprotein disorders | endocrine/metabolic | DR | 1.06 | 1.00 | 1.13 | 0.305512005 | 128777 | 2 |
| 732.1 | Juvenile osteochondrosis | musculoskeletal | DR | 0.90 | 0.81 | 1.00 | 0.309338999 | 128777 | 2 |
| 627.22 | | | DR | 1.11 | 1.00 | 1.23 | 0.312886229 | 128777 | 2 |
| 198.4 | Secondary malignant neoplasm of liver | neoplasms | DR | 0.95 | 0.89 | 1.00 | 0.314823789 | 128777 | 2 |
| 613.1 | Inflammatory disease of breast | genitourinary | DR | 1.11 | 1.00 | 1.24 | 0.315026432 | 128777 | 2 |
| 984 | Toxic effect of lead and its compounds (including fumes) | injuries & poisonings | DR | 0.51 | 0.26 | 1.00 | 0.315132512 | 111475 | 1 |
| 704.11 | Alopecia Areata | dermatologic | DR | 0.87 | 0.75 | 1.00 | 0.315821808 | 128777 | 2 |
| 736 | Other acquired deformities of limbs | musculoskeletal | DR | 1.03 | 1.00 | 1.05 | 0.317088707 | 128777 | 2 |
| 711.3 | Behcet's syndrome | musculoskeletal | DR | 0.65 | 0.42 | 1.00 | 0.318178083 | 111475 | 1 |
| 656.22 | Interstitial emphysema and related conditions of newborn | pregnancy complications | DR | 2.70 | 0.99 | 7.37 | 0.322940678 | 111475 | 1 |
| 613.9 | Breast disorder NOS | genitourinary | DR | 1.08 | 1.00 | 1.18 | 0.32599894 | 128777 | 2 |
| 737.3 | Kyphoscoliosis and scoliosis | musculoskeletal | DR | 0.96 | 0.92 | 1.00 | 0.327554905 | 128777 | 2 |
| 325 | Phlebitis and thrombophlebitis of intracranial venous sinuses | neurological | DR | 0.68 | 0.46 | 1.01 | 0.3278708 | 111475 | 1 |
| 427.9 | Palpitations | circulatory system | DR | 1.02 | 1.00 | 1.04 | 0.328001055 | 128777 | 2 |
| 550.5 | Ventral hernia | digestive | DR | 0.97 | 0.95 | 1.00 | 0.329141456 | 128777 | 2 |
| 528.11 | Stomatitis and mucositis (ulcerative) | digestive | DR | 0.91 | 0.83 | 1.00 | 0.330243778 | 128777 | 2 |
| 523 | Gingival and periodontal diseases | digestive | DR | 0.99 | 0.97 | 1.00 | 0.330397045 | 128777 | 2 |
| 709.5 | Dermatomyositis | dermatologic | DR | 1.22 | 0.99 | 1.51 | 0.332635254 | 128777 | 2 |
| 255.3 | Adrenogenital disorders | endocrine/metabolic | DR | 0.70 | 0.48 | 1.01 | 0.332668265 | 111475 | 1 |
| 803.2 | Fracture of radius and ulna | injuries & poisonings | DR | 1.04 | 1.00 | 1.08 | 0.336255856 | 128777 | 2 |
| 131 | Protozoan infection | infectious diseases | DR | 0.87 | 0.75 | 1.01 | 0.33630718 | 111475 | 1 |
| 612 | Breast conditions, congenital or relating to hormones | genitourinary | DR | 0.96 | 0.92 | 1.00 | 0.337389149 | 111475 | 1 |
| 764 | Sciatica | symptoms | DR | 0.98 | 0.96 | 1.00 | 0.337591996 | 128777 | 2 |
| 758.1 | Chromosomal anomalies | congenital anomalies | DR | 0.86 | 0.73 | 1.01 | 0.337981611 | 128777 | 2 |
| 749.1 | Cleft palate | congenital anomalies | DR | 1.41 | 0.98 | 2.02 | 0.339529595 | 111475 | 1 |
| 315 | Develomental delays and disorders | mental disorders | DR | 1.04 | 1.00 | 1.09 | 0.342607871 | 128777 | 2 |
| 202.2 | Non-Hodgkins lymphoma | neoplasms | DR | 1.04 | 1.00 | 1.09 | 0.342658235 | 128777 | 2 |
| 204.3 | Monocytic leukemia | neoplasms | DR | 1.17 | 0.99 | 1.39 | 0.345077118 | 128777 | 2 |
| 870.6 | Open wound of neck | injuries & poisonings | DR | 1.13 | 0.99 | 1.28 | 0.345193556 | 111475 | 1 |
| 676 | Other disorders of the breast associated with childbirth and disorders of lactation | pregnancy complications | DR | 0.35 | 0.11 | 1.07 | 0.345750188 | 111475 | 1 |
| 327.31 | Central/nonobstructive sleep apnea | neurological | DR | 1.03 | 1.00 | 1.06 | 0.349813243 | 128777 | 2 |
| 619.1 | Noninflammatory disorders of ovary, fallopian tube, and broad ligament | genitourinary | DR | 0.86 | 0.74 | 1.01 | 0.34999626 | 128777 | 2 |
| 619.2 | Disorders of uterus, NEC | genitourinary | DR | 0.93 | 0.85 | 1.01 | 0.352073911 | 128777 | 2 |
| 592.3 | Urethral stricture due to infecton | genitourinary | DR | 1.25 | 0.98 | 1.59 | 0.358520317 | 111475 | 1 |
| 446.4 | Wegener's granulomatosis | circulatory system | DR | 0.82 | 0.66 | 1.02 | 0.365529024 | 128777 | 2 |
| 656.8 | Perinatal jaundice | pregnancy complications | DR | 1.82 | 0.93 | 3.56 | 0.372462197 | 111475 | 1 |
| 145.4 | Cancer of the gums | neoplasms | DR | 1.30 | 0.97 | 1.74 | 0.372837803 | 111475 | 1 |
| 795.81 | Elevated carcinoembryonic antigen [CEA] | symptoms | DR | 1.18 | 0.98 | 1.41 | 0.372952542 | 128777 | 2 |
| 624.1 | Dystrophy of female genital tract | genitourinary | DR | 1.23 | 0.97 | 1.55 | 0.375212977 | 128777 | 2 |
| 696.3 | Pityriasis | dermatologic | DR | 0.89 | 0.79 | 1.01 | 0.376298981 | 111475 | 1 |
| 619.4 | Noninflammatory disorders of vagina | genitourinary | DR | 0.92 | 0.85 | 1.01 | 0.376595555 | 128777 | 2 |
| 270.1 | Disturbances of amino-acid transport | endocrine/metabolic | DR | 1.14 | 0.98 | 1.31 | 0.378051701 | 111475 | 1 |
| 519.1 | Tracheostomy complications | respiratory | DR | 1.13 | 0.98 | 1.29 | 0.379120399 | 128777 | 2 |
| 473.1 | Chronic laryngitis | respiratory | DR | 0.94 | 0.88 | 1.01 | 0.381028557 | 128777 | 2 |
| 204.22 | Myeloid leukemia, chronic | neoplasms | DR | 1.11 | 0.98 | 1.25 | 0.386514414 | 128777 | 2 |
| 275.1 | Disorders of iron metabolism | hematopoietic | DR | 0.95 | 0.90 | 1.01 | 0.388897613 | 128777 | 2 |
| 723.1 | Torticollis | musculoskeletal | DR | 0.93 | 0.86 | 1.01 | 0.397081418 | 128777 | 2 |
| 715.2 | Ankylosing spondylitis | musculoskeletal | DR | 0.93 | 0.86 | 1.01 | 0.398371731 | 128777 | 2 |
| 291 | | | DR | 0.98 | 0.95 | 1.00 | 0.400428671 | 111475 | 1 |
| 657 | Infections specific to the perinatal period | pregnancy complications | DR | 0.84 | 0.68 | 1.03 | 0.400831188 | 111475 | 1 |
| 617 | Disorders secondary to childbirth, surgery, trauma | genitourinary | DR | 1.10 | 0.98 | 1.24 | 0.401033001 | 128777 | 2 |
| 526.5 | Inflammatory conditions of jaw | digestive | DR | 0.95 | 0.89 | 1.01 | 0.402099236 | 128777 | 2 |
| 550.3 | Femoral hernia | digestive | DR | 1.16 | 0.97 | 1.38 | 0.402231 | 111475 | 1 |
| 858 | Complication of internal orthopedic device | injuries & poisonings | DR | 1.03 | 0.99 | 1.06 | 0.402677938 | 128777 | 2 |
| 536.8 | Dyspepsia and other specified disorders of function of stomach | digestive | DR | 1.02 | 1.00 | 1.05 | 0.402863995 | 128777 | 2 |
| 204.11 | Lymphoid leukemia, acute | neoplasms | DR | 1.13 | 0.98 | 1.31 | 0.404170429 | 128777 | 2 |
| 795.82 | Elevated cancer antigen 125 [CA 125] | symptoms | DR | 1.40 | 0.94 | 2.09 | 0.404373685 | 111475 | 1 |
| 741.2 | Stiffness of joint | musculoskeletal | DR | 1.03 | 0.99 | 1.06 | 0.410668048 | 128777 | 2 |
| 212 | Benign neoplasm of respiratory and intrathoracic organs | neoplasms | DR | 0.95 | 0.89 | 1.01 | 0.41158586 | 128777 | 2 |
| 575.6 | Cholesterolosis of gallbladder | digestive | DR | 0.92 | 0.84 | 1.02 | 0.412802931 | 128777 | 2 |
| 714.1 | Rheumatoid arthritis | musculoskeletal | DR | 1.03 | 0.99 | 1.06 | 0.415350648 | 128777 | 2 |
| 246 | Other disorders of thyroid | endocrine/metabolic | DR | 1.03 | 0.99 | 1.07 | 0.416041178 | 128777 | 2 |
| 781.2 | Abnormal posture | symptoms | DR | 1.07 | 0.98 | 1.17 | 0.416067355 | 128777 | 2 |
| 255.2 | | | DR | 1.06 | 0.99 | 1.13 | 0.419369147 | 128777 | 2 |
| 246.2 | | | DR | 1.10 | 0.98 | 1.25 | 0.42022443 | 128777 | 2 |
| 202.22 | Reticulosarcoma | neoplasms | DR | 1.07 | 0.98 | 1.17 | 0.421197203 | 128777 | 2 |
| 695.41 | Cutaneous lupus erythematosus | dermatologic | DR | 0.90 | 0.78 | 1.03 | 0.421683622 | 128777 | 2 |
| 184.11 | Malignant neoplasm of ovary | neoplasms | DR | 1.16 | 0.96 | 1.40 | 0.422515183 | 128777 | 2 |
| 611.3 | Lump or mass in breast | genitourinary | DR | 0.97 | 0.94 | 1.01 | 0.42303458 | 128777 | 2 |
| 385.3 | Cholesteatoma | sense organs | DR | 1.06 | 0.98 | 1.15 | 0.42497655 | 128777 | 2 |
| 636.3 | Hemorrhage in early pregnancy | pregnancy complications | DR | 0.82 | 0.65 | 1.05 | 0.426644603 | 128777 | 2 |
| 244.1 | Secondary hypothyroidism | endocrine/metabolic | DR | 1.04 | 0.99 | 1.08 | 0.429436248 | 128777 | 2 |
| 726.3 | Bursitis | musculoskeletal | DR | 1.02 | 1.00 | 1.04 | 0.430195682 | 128777 | 2 |
| 573 | Other disorders of liver | digestive | DR | 0.98 | 0.95 | 1.01 | 0.430352659 | 128777 | 2 |
| 275.2 | Disorders of copper metabolism | endocrine/metabolic | DR | 0.74 | 0.50 | 1.09 | 0.432870243 | 111475 | 1 |
| 569.2 | Gastrointestinal complications | digestive | DR | 1.03 | 0.99 | 1.08 | 0.437994266 | 128777 | 2 |
| 278.11 | Morbid obesity | endocrine/metabolic | DR | 0.99 | 0.98 | 1.00 | 0.439533723 | 128777 | 2 |
| 795 | Other and nonspecific abnormal cytological, histological and immunological findings | symptoms | DR | 0.87 | 0.73 | 1.04 | 0.440294308 | 128777 | 2 |
| 293.1 | Swelling, mass, or lump in head and neck [Space-occupying lesion, intracranial NOS] | mental disorders | DR | 0.98 | 0.96 | 1.01 | 0.442524448 | 128777 | 2 |
| 696.4 | Psoriasis | dermatologic | DR | 1.02 | 0.99 | 1.05 | 0.444292807 | 111475 | 1 |
| 618.1 | Prolapse of vaginal walls | genitourinary | DR | 1.07 | 0.98 | 1.17 | 0.444924964 | 128777 | 2 |
| 522.1 | Pulpitis and necrosis of tooth pulp | digestive | DR | 0.98 | 0.96 | 1.01 | 0.446468113 | 111475 | 1 |
| 526.9 | Jaw disease NOS | digestive | DR | 1.05 | 0.98 | 1.13 | 0.451490234 | 128777 | 2 |
| 643.1 | Hyperemesis gravidarum | pregnancy complications | DR | 0.42 | 0.13 | 1.34 | 0.453355071 | 111475 | 1 |
| 594 | Urinary calculus | genitourinary | DR | 0.99 | 0.97 | 1.00 | 0.454036829 | 128777 | 2 |
| 785 | Abdominal pain | symptoms | DR | 0.99 | 0.98 | 1.00 | 0.454556763 | 128777 | 2 |

| | | | | | | | | | |
|---|---|---|---|---|---|---|---|---|---|
| 385.5 | Tympanosclerosis and middle ear disease related to otitis media | sense organs | DR | 1.09 | 0.97 | 1.23 | 0.456313755 | 128777 | 2 |
| 756.2 | Pectus and other congenital anomalies of ribs/sternum | congenital anomalies | DR | 1.29 | 0.92 | 1.82 | 0.457866148 | 111475 | 1 |
| 446.8 | Thrombotic microangiopathy | circulatory system | DR | 0.75 | 0.51 | 1.11 | 0.47008326 | 111475 | 1 |
| 221 | Benign neoplasm of other female genital organs | neoplasms | DR | 1.17 | 0.94 | 1.44 | 0.471028132 | 128777 | 2 |
| 957 | | | DR | 0.88 | 0.73 | 1.05 | 0.473608387 | 111475 | 1 |
| 442.4 | Arterial dissection | circulatory system | DR | 1.10 | 0.96 | 1.25 | 0.474969584 | 128777 | 2 |
| 603 | Other disorders of testis | genitourinary | DR | 1.02 | 0.99 | 1.06 | 0.47502832 | 111475 | 1 |
| 842 | | | DR | 1.02 | 0.99 | 1.05 | 0.475213619 | 128777 | 2 |
| 709.6 | Other specified diffuse diseases of connective tissue | dermatologic | DR | 0.85 | 0.68 | 1.07 | 0.475470664 | 128777 | 2 |
| 741.6 | Villonodular synovitis | musculoskeletal | DR | 1.12 | 0.96 | 1.31 | 0.475843052 | 111475 | 1 |
| 560.3 | Peritoneal or intestinal adhesions | digestive | DR | 1.07 | 0.97 | 1.17 | 0.476726431 | 128777 | 2 |
| 499 | Cystic fibrosis | respiratory | DR | 1.21 | 0.92 | 1.59 | 0.477380089 | 128777 | 2 |
| 751.11 | Congenital anomalies of female genital organs | congenital anomalies | DR | 0.79 | 0.57 | 1.10 | 0.48107227 | 111475 | 1 |
| 193 | Thyroid cancer | neoplasms | DR | 0.95 | 0.89 | 1.02 | 0.482046035 | 128777 | 2 |
| 282.5 | Sickle cell anemia | hematopoietic | DR | 1.20 | 0.92 | 1.57 | 0.482932141 | 111475 | 1 |
| 612.2 | Hypertrophy of breast (Gynecomastia) | genitourinary | DR | 0.97 | 0.93 | 1.01 | 0.486790944 | 128777 | 2 |
| 750.22 | Congenital anomaly of gallbladder, bile ducts, liver, pancreas | congenital anomalies | DR | 1.06 | 0.97 | 1.16 | 0.487822182 | 128777 | 2 |
| 197 | Chemotherapy | neoplasms | DR | 0.98 | 0.95 | 1.01 | 0.493579791 | 128777 | 2 |
| 635 | Hemorrhage during pregnancy; childbirth and postpartum | pregnancy complications | DR | 0.74 | 0.47 | 1.16 | 0.499067143 | 111475 | 1 |
| 452.1 | | | DR | 1.09 | 0.96 | 1.23 | 0.499174345 | 128777 | 2 |
| 279.11 | Deficiency of humoral immunity | endocrine/metabolic | DR | 0.94 | 0.85 | 1.03 | 0.500086352 | 128777 | 2 |
| 157 | Pancreatic cancer | neoplasms | DR | 1.04 | 0.98 | 1.11 | 0.500788871 | 128777 | 2 |
| 446.3 | Hypersensitivity angiitis | circulatory system | DR | 1.13 | 0.94 | 1.36 | 0.50102089 | 111475 | 1 |
| 571.6 | Primary biliary cirrhosis | digestive | DR | 1.10 | 0.96 | 1.26 | 0.501198743 | 128777 | 2 |
| 559 | Ileostomy status | digestive | DR | 1.05 | 0.97 | 1.14 | 0.501284158 | 128777 | 2 |
| 530.7 | Gastroesophageal laceration-hemorrhage syndrome | digestive | DR | 1.08 | 0.96 | 1.20 | 0.504064316 | 128777 | 2 |
| 327.7 | Sleep related movement disorders | neurological | DR | 0.99 | 0.97 | 1.01 | 0.504509406 | 128777 | 2 |
| 938.1 | Acute dermatitis due to solar radiation | dermatologic | DR | 0.96 | 0.91 | 1.02 | 0.505563755 | 128777 | 2 |
| 736.3 | Acquired deformities of hip | musculoskeletal | DR | 0.89 | 0.75 | 1.06 | 0.512958462 | 111475 | 1 |
| 871.1 | Open wound of hand except finger(s) | injuries & poisonings | DR | 1.02 | 0.99 | 1.06 | 0.513108588 | 128777 | 2 |
| 577.1 | Acute pancreatitis | digestive | DR | 1.02 | 0.99 | 1.05 | 0.514202228 | 128777 | 2 |
| 756.1 | Congenital anomalies of abdominal wall; diaphram | congenital anomalies | DR | 1.10 | 0.95 | 1.27 | 0.515768656 | 128777 | 2 |
| 334.21 | Amyotrophic Lateral Sclerosis | neurological | DR | 1.10 | 0.95 | 1.29 | 0.5173849 | 111475 | 1 |
| 371.33 | Noninfectious dermatoses of eyelid | sense organs | DR | 1.11 | 0.94 | 1.31 | 0.520418174 | 111475 | 1 |
| 381 | | | DR | 0.99 | 0.97 | 1.01 | 0.522445675 | 111475 | 1 |
| 697 | Sarcoidosis | dermatologic | DR | 0.95 | 0.88 | 1.03 | 0.524370438 | 128777 | 2 |
| 277.1 | Disorders of porphyrin metabolism | endocrine/metabolic | DR | 0.90 | 0.77 | 1.06 | 0.524778592 | 111475 | 1 |
| 716.2 | Unspecified monoarthritis | musculoskeletal | DR | 1.03 | 0.98 | 1.09 | 0.52940091 | 111475 | 1 |
| 353.1 | Nerve plexus lesions | neurological | DR | 0.96 | 0.89 | 1.03 | 0.529531421 | 128777 | 2 |
| 599.7 | Urethral discharge | genitourinary | DR | 1.09 | 0.95 | 1.26 | 0.529608882 | 111475 | 1 |
| 202.21 | Nodular lymphoma | neoplasms | DR | 0.94 | 0.86 | 1.04 | 0.529658945 | 128777 | 2 |
| 751.1 | Congenital anomalies of genital organs | congenital anomalies | DR | 0.97 | 0.91 | 1.02 | 0.532658785 | 111475 | 1 |
| 772.4 | Rhabdomyolysis | symptoms | DR | 1.03 | 0.98 | 1.09 | 0.533696573 | 128777 | 2 |
| 696 | | | DR | 1.02 | 0.99 | 1.04 | 0.536027243 | 111475 | 1 |
| 149 | Cancer of larynx, pharynx, nasal cavities | neoplasms | DR | 1.03 | 0.98 | 1.09 | 0.539538734 | 128777 | 2 |
| 804 | Fracture of hand or wrist | injuries & poisonings | DR | 0.98 | 0.95 | 1.01 | 0.541365379 | 128777 | 2 |
| 289.4 | Lymphadenitis | hematopoietic | DR | 1.01 | 0.99 | 1.04 | 0.542439702 | 128777 | 2 |
| 704.12 | Telogen effluvium | dermatologic | DR | 0.88 | 0.71 | 1.09 | 0.543615961 | 128777 | 2 |
| 716.8 | Palindromic rheumatism | musculoskeletal | DR | 0.90 | 0.74 | 1.08 | 0.549191312 | 111475 | 1 |
| 225 | Benign neoplasm of brain and other parts of nervous system | neoplasms | DR | 1.04 | 0.98 | 1.10 | 0.549733589 | 128777 | 2 |
| 528 | Diseases of the oral soft tissues, excluding lesions specific for gingiva and tongue | digestive | DR | 0.99 | 0.97 | 1.01 | 0.551731161 | 128777 | 2 |
| 658 | Maternal complication of pregnancy affecting fetus or newborn | pregnancy complications | DR | 0.90 | 0.74 | 1.08 | 0.557904776 | 128777 | 2 |
| 696.41 | Psoriasis vulgaris | dermatologic | DR | 1.02 | 0.99 | 1.04 | 0.559089549 | 128777 | 2 |
| 751.12 | Congenital anomalies of male genital organs | congenital anomalies | DR | 0.95 | 0.87 | 1.04 | 0.567600577 | 111475 | 1 |
| 530.11 | GERD | digestive | DR | 0.99 | 0.98 | 1.01 | 0.568734305 | 128777 | 2 |
| 510.2 | Lung transplant | respiratory | DR | 1.07 | 0.95 | 1.21 | 0.569376991 | 128777 | 2 |
| 225.1 | Benign neoplasm of brain, cranial nerves, meninges | neoplasms | DR | 1.03 | 0.97 | 1.10 | 0.570380157 | 128777 | 2 |
| 282.8 | Other hemoglobinopathies | hematopoietic | DR | 0.95 | 0.87 | 1.04 | 0.573503405 | 128777 | 2 |
| 752 | Nervous system congenital anomalies | congenital anomalies | DR | 0.95 | 0.87 | 1.04 | 0.574254298 | 111475 | 1 |
| 149.2 | Cancer of nasopharynx | neoplasms | DR | 1.11 | 0.92 | 1.35 | 0.574557719 | 111475 | 1 |
| 359.1 | Muscular dystrophies | neurological | DR | 0.91 | 0.77 | 1.08 | 0.574853598 | 128777 | 2 |
| 752.2 | Other specified congenital anomalies of nervous system | congenital anomalies | DR | 1.06 | 0.95 | 1.19 | 0.57635701 | 128777 | 2 |
| 750.5 | Congenital hypertrophic pyloric stenosis | congenital anomalies | DR | 0.54 | 0.17 | 1.66 | 0.581084812 | 111475 | 1 |
| 716.3 | Kaschin-Beck disease | musculoskeletal | DR | 1.13 | 0.90 | 1.43 | 0.583300266 | 111475 | 1 |
| 564 | Functional digestive disorders | digestive | DR | 0.99 | 0.97 | 1.01 | 0.586350026 | 128777 | 2 |
| 1006 | Crushing injury | NULL | DR | 1.03 | 0.97 | 1.10 | 0.586631985 | 128777 | 2 |
| 149.3 | Cancer of hypopharynx | neoplasms | DR | 0.91 | 0.76 | 1.09 | 0.587033202 | 111475 | 1 |
| 379.1 | Scleritis and episcleritis | sense organs | DR | 1.04 | 0.97 | 1.12 | 0.588015761 | 128777 | 2 |
| 230 | Kaposi's sarcoma | neoplasms | DR | 0.90 | 0.75 | 1.09 | 0.591392477 | 111475 | 1 |
| 382 | Otalgia | sense organs | DR | 0.99 | 0.96 | 1.01 | 0.591417167 | 128777 | 2 |
| 270.21 | Disorders of urea cycle metabolism | endocrine/metabolic | DR | 0.94 | 0.84 | 1.05 | 0.591716653 | 111475 | 1 |
| 524 | Dentofacial anomalies, including malocclusion | digestive | DR | 1.02 | 0.99 | 1.04 | 0.591927697 | 128777 | 2 |
| 759.1 | Anomalies of endocrine glands, congenital | congenital anomalies | DR | 1.16 | 0.88 | 1.53 | 0.59299299 | 111475 | 1 |
| 149.1 | Cancer of oropharynx | neoplasms | DR | 0.96 | 0.88 | 1.04 | 0.593117146 | 128777 | 2 |
| 334 | Degenerative disease of the spinal cord | neurological | DR | 0.98 | 0.94 | 1.02 | 0.59634829 | 128777 | 2 |
| 733.2 | Cyst of bone | musculoskeletal | DR | 0.96 | 0.89 | 1.04 | 0.597499165 | 128777 | 2 |
| 259.4 | Precocious sexual development and puberty NEC | endocrine/metabolic | DR | 2.11 | 0.51 | 8.71 | 0.597780421 | 111475 | 1 |
| 513.31 | Apnea | respiratory | DR | 1.02 | 0.98 | 1.07 | 0.598203527 | 128777 | 2 |
| 623 | Hypertrophy of female genital organs | genitourinary | DR | 0.92 | 0.78 | 1.08 | 0.603417851 | 128777 | 2 |
| 695.21 | Dermatitis herpetiformis | dermatologic | DR | 1.08 | 0.93 | 1.24 | 0.605713434 | 111475 | 1 |
| 853 | Complication of colostomy or enterostomy | injuries & poisonings | DR | 0.95 | 0.86 | 1.05 | 0.606321343 | 128777 | 2 |
| 963 | Poisoning by primarily systemic agents | injuries & poisonings | DR | 0.97 | 0.93 | 1.03 | 0.609946265 | 128777 | 2 |
| 755.6 | Other congenital anomalies of lower limb, including pelvic girdle | congenital anomalies | DR | 1.06 | 0.95 | 1.18 | 0.611814075 | 111475 | 1 |
| 433.32 | Moyamoya disease | circulatory system | DR | 2.06 | 0.49 | 8.62 | 0.613722303 | 111475 | 1 |
| 656.2 | Respiratory conditions of fetus and newborn | pregnancy complications | DR | 1.12 | 0.89 | 1.41 | 0.61506935 | 111475 | 1 |
| 117.2 | Coccidioidomycosis | infectious diseases | DR | 1.09 | 0.92 | 1.30 | 0.616193729 | 111475 | 1 |
| 520 | Disorders of tooth development | digestive | DR | 1.02 | 0.98 | 1.05 | 0.61902285 | 111475 | 1 |
| 386.1 | Meniere's disease | sense organs | DR | 1.03 | 0.97 | 1.11 | 0.620018458 | 128777 | 2 |
| 496.3 | Bronchiectasis | respiratory | DR | 1.03 | 0.97 | 1.10 | 0.621395776 | 128777 | 2 |
| 174.2 | Breast cancer [male] | neoplasms | DR | 1.09 | 0.91 | 1.30 | 0.634370236 | 128777 | 2 |
| 958 | Certain early complications of trauma or procedure | injuries & poisonings | DR | 1.03 | 0.97 | 1.10 | 0.639920396 | 128777 | 2 |
| 151 | Cancer of stomach | neoplasms | DR | 1.04 | 0.96 | 1.13 | 0.640499662 | 128777 | 2 |
| 550.2 | Diaphragmatic hernia | digestive | DR | 0.99 | 0.97 | 1.01 | 0.644353607 | 128777 | 2 |
| 568.1 | Peritoneal adhesions (postoperative) (postinfection) | digestive | DR | 0.98 | 0.93 | 1.03 | 0.644948178 | 128777 | 2 |
| 264.3 | Delayed milestones | endocrine/metabolic | DR | 1.94 | 0.46 | 8.17 | 0.646187832 | 111475 | 1 |

| | | | | | | | | | |
|---|---|---|---|---|---|---|---|---|---|
| 346.3 | Nonspecific abnormal findings in cerebrospinal fluid | neurological | DR | 1.09 | 0.90 | 1.32 | 0.647609671 | 128777 | 2 |
| 624.9 | stress incontinence, female | genitourinary | DR | 1.02 | 0.97 | 1.08 | 0.649990303 | 128777 | 2 |
| 736.5 | Acquired deformities of knee | musculoskeletal | DR | 1.05 | 0.94 | 1.18 | 0.651192767 | 111475 | 1 |
| 531.5 | Gastrojejunal ulcer | digestive | DR | 1.07 | 0.92 | 1.24 | 0.651772691 | 128777 | 2 |
| 227.2 | Benign neoplasm of parathyroid gland | neoplasms | DR | 1.05 | 0.95 | 1.16 | 0.651883111 | 128777 | 2 |
| 130.1 | Lyme disease | infectious diseases | DR | 1.04 | 0.95 | 1.13 | 0.659426668 | 111475 | 1 |
| 426.4 | Anomalous atrioventricular excitation | circulatory system | DR | 0.93 | 0.80 | 1.09 | 0.66301722 | 111475 | 1 |
| 624 | Symptoms involving female genital tract | genitourinary | DR | 0.95 | 0.83 | 1.08 | 0.665246272 | 128777 | 2 |
| 279.8 | Other specified disorders involving the immune mechanism | endocrine/metabolic | DR | 1.07 | 0.91 | 1.26 | 0.667035891 | 128777 | 2 |
| 614.53 | Cyst or abscess of Bartholin's gland | genitourinary | DR | 0.89 | 0.68 | 1.17 | 0.668612369 | 111475 | 1 |
| 287.4 | Qualitative platelet defects | hematopoietic | DR | 0.95 | 0.85 | 1.07 | 0.669875404 | 128777 | 2 |
| 270.35 | Macroglobulinemia | endocrine/metabolic | DR | 1.11 | 0.87 | 1.43 | 0.672367681 | 111475 | 1 |
| 727.5 | Rupture of synovium | musculoskeletal | DR | 1.02 | 0.97 | 1.07 | 0.673121927 | 128777 | 2 |
| 159.4 | Malignant neoplasm of retroperitoneum and peritoneum | neoplasms | DR | 0.91 | 0.74 | 1.13 | 0.674398891 | 111475 | 1 |
| 634.1 | Missed abortion/Hydatidiform mole | pregnancy complications | DR | 0.93 | 0.79 | 1.10 | 0.677101035 | 128777 | 2 |
| 573.5 | Jaundice (not of newborn) | digestive | DR | 1.02 | 0.97 | 1.07 | 0.677921756 | 128777 | 2 |
| 656.1 | Isoimmunization of fetus or newborn | pregnancy complications | DR | 1.44 | 0.59 | 3.54 | 0.683127311 | 111475 | 1 |
| 637 | Short gestation; low birth weight; and fetal growth retardation | pregnancy complications | DR | 1.14 | 0.82 | 1.59 | 0.685806189 | 111475 | 1 |
| 333.3 | Tics and choreas | neurological | DR | 0.94 | 0.82 | 1.09 | 0.685903665 | 128777 | 2 |
| 612.3 | Congenital anomalies of breast | genitourinary | DR | 0.85 | 0.57 | 1.27 | 0.686016567 | 111475 | 1 |
| 618.5 | Prolapse of vaginal vault after hysterectomy | genitourinary | DR | 1.08 | 0.90 | 1.29 | 0.6906432 | 128777 | 2 |
| 528.1 | Stomatitis and mucositis | digestive | DR | 1.02 | 0.97 | 1.07 | 0.69076273 | 128777 | 2 |
| 575 | Other biliary tract disease | digestive | DR | 1.01 | 0.98 | 1.04 | 0.690937777 | 111475 | 1 |
| 164 | Cancer of intrathoracic organs | neoplasms | DR | 1.05 | 0.92 | 1.20 | 0.691935792 | 128777 | 2 |
| 737.2 | Lordosis (acquired) | musculoskeletal | DR | 0.95 | 0.82 | 1.09 | 0.703263757 | 111475 | 1 |
| 343 | Infantile cerebral palsy | neurological | DR | 1.14 | 0.81 | 1.60 | 0.704101108 | 111475 | 1 |
| 70.1 | Viral hepatitis A | infectious diseases | DR | 0.96 | 0.85 | 1.08 | 0.707330681 | 111475 | 1 |
| 287.2 | Allergic purpura | hematopoietic | DR | 0.94 | 0.78 | 1.12 | 0.707897723 | 111475 | 1 |
| 433.5 | Cerebral aneurysm | circulatory system | DR | 1.03 | 0.95 | 1.11 | 0.7098847 | 128777 | 2 |
| 840.1 | | | DR | 1.02 | 0.96 | 1.09 | 0.709948822 | 128777 | 2 |
| 133 | Arthropod-borne diseases | infectious diseases | DR | 1.05 | 0.92 | 1.19 | 0.712193053 | 111475 | 1 |
| 277.5 | Other disorders of lipoid metabolism | endocrine/metabolic | DR | 1.01 | 0.98 | 1.04 | 0.713388766 | 128777 | 2 |
| 260.22 | Nutritional marasmus | endocrine/metabolic | DR | 0.95 | 0.81 | 1.10 | 0.714990724 | 128777 | 2 |
| 175 | Acquired absence of breast | neoplasms | DR | 0.95 | 0.82 | 1.10 | 0.716570172 | 128777 | 2 |
| 627.3 | Postmenopausal atrophic vaginitis | genitourinary | DR | 1.02 | 0.96 | 1.09 | 0.716879031 | 128777 | 2 |
| 750.13 | Congenital anomalies of mouth/tongue | congenital anomalies | DR | 1.05 | 0.91 | 1.22 | 0.7294968 | 128777 | 2 |
| 635.2 | Antepartum hemorrhage, abruptio placentae, and placenta previa | pregnancy complications | DR | 1.11 | 0.82 | 1.50 | 0.732215992 | 128777 | 2 |
| 316.1 | Polyneuropathy due to drugs | mental disorders | DR | 0.96 | 0.85 | 1.08 | 0.734046426 | 128777 | 2 |
| 946 | Anaphylactic shock NOS | injuries & poisonings | DR | 0.98 | 0.93 | 1.04 | 0.734493554 | 128777 | 2 |
| 645 | Late pregnancy and failed induction | pregnancy complications | DR | 0.88 | 0.61 | 1.28 | 0.736008497 | 128777 | 2 |
| 727.6 | Rupture of tendon, nontraumatic | musculoskeletal | DR | 0.99 | 0.97 | 1.02 | 0.736806588 | 128777 | 2 |
| 323.2 | Acute (transverse) myelitis | neurological | DR | 0.90 | 0.67 | 1.22 | 0.738443518 | 111475 | 1 |
| 187 | Cancer of other male genital organs | neoplasms | DR | 0.98 | 0.93 | 1.04 | 0.740910819 | 111475 | 1 |
| 286.8 | Hypercoagulable state | hematopoietic | DR | 0.98 | 0.92 | 1.04 | 0.742489138 | 128777 | 2 |
| 353 | Nerve root and plexus disorders | neurological | DR | 1.01 | 0.98 | 1.04 | 0.745027943 | 128777 | 2 |
| 816 | Cerebral laceration and contusion | injuries & poisonings | DR | 0.95 | 0.83 | 1.10 | 0.747069084 | 128777 | 2 |
| 194 | Cancer of other endocrine glands | neoplasms | DR | 1.02 | 0.95 | 1.10 | 0.749251776 | 128777 | 2 |
| 279.7 | Other immunological findings | endocrine/metabolic | DR | 0.98 | 0.92 | 1.04 | 0.752301651 | 111475 | 1 |
| 149.4 | Cancer of larynx | neoplasms | DR | 0.98 | 0.90 | 1.06 | 0.754378675 | 128777 | 2 |
| 528.3 | Cellulitis and abscess of oral soft tissues | digestive | DR | 0.99 | 0.94 | 1.03 | 0.760414698 | 128777 | 2 |
| 117.3 | Blastomycotic infection | infectious diseases | DR | 1.09 | 0.83 | 1.43 | 0.760592664 | 111475 | 1 |
| 209 | Neuroendocrine tumors | neoplasms | DR | 1.03 | 0.93 | 1.15 | 0.763580352 | 128777 | 2 |
| 601.11 | Acute prostatitis | genitourinary | DR | 0.99 | 0.94 | 1.03 | 0.768586158 | 128777 | 2 |
| 425.8 | Other cardiomyopathy | circulatory system | DR | 0.97 | 0.87 | 1.08 | 0.769754087 | 128777 | 2 |
| 334.1 | Spinocerebellar disease | neurological | DR | 0.97 | 0.87 | 1.08 | 0.770743599 | 128777 | 2 |
| 475.9 | Postnasal drip | respiratory | DR | 1.01 | 0.97 | 1.06 | 0.77543203 | 128777 | 2 |
| 655.1 | Abnormality in fetal heart rate or rhythm | pregnancy complications | DR | 0.92 | 0.70 | 1.22 | 0.775816548 | 128777 | 2 |
| 227 | Benign neoplasm of other endocrine glands and related structures | neoplasms | DR | 0.99 | 0.95 | 1.03 | 0.77610617 | 111475 | 1 |
| 530.13 | Barrett's esophagus | digestive | DR | 0.99 | 0.96 | 1.02 | 0.776331913 | 128777 | 2 |
| 592.13 | Chronic interstitial cystitis | genitourinary | DR | 0.97 | 0.85 | 1.09 | 0.777061082 | 128777 | 2 |
| 823 | | | DR | 0.00 | 0.00 | 6.68E+09 | 0.777659411 | 111475 | 1 |
| 952 | Spinal cord injury without evidence of spinal bone injury | injuries & poisonings | DR | 0.98 | 0.92 | 1.05 | 0.777919326 | 128777 | 2 |
| 442.2 | Aneurysm of iliac artery | circulatory system | DR | 0.98 | 0.91 | 1.05 | 0.778715011 | 128777 | 2 |
| 286.3 | Coagulation defects complicating pregnancy or postpartum | hematopoietic | DR | 0.00 | 0.00 | 5.57E+09 | 0.781366579 | 111475 | 1 |
| 145.2 | Cancer of tongue | neoplasms | DR | 1.02 | 0.94 | 1.11 | 0.782642741 | 128777 | 2 |
| 282 | | | DR | 0.98 | 0.91 | 1.06 | 0.783588839 | 111475 | 1 |
| 526.1 | Cysts of the jaws | digestive | DR | 1.03 | 0.92 | 1.16 | 0.784610981 | 128777 | 2 |
| 446.1 | Thromboangiitis obliterans | circulatory system | DR | 0.95 | 0.78 | 1.16 | 0.786834848 | 111475 | 1 |
| 350.5 | Abnormal reflex | neurological | DR | 0.97 | 0.87 | 1.08 | 0.78855042 | 128777 | 2 |
| 213 | Benign neoplasm of bone and articular cartilage | neoplasms | DR | 1.02 | 0.95 | 1.09 | 0.789160208 | 128777 | 2 |
| 754 | Congenital musculoskeletal deformities of spine | congenital anomalies | DR | 0.99 | 0.94 | 1.04 | 0.789868832 | 128777 | 2 |
| 871.2 | Open wound of finger(s) | injuries & poisonings | DR | 0.99 | 0.96 | 1.02 | 0.791642325 | 128777 | 2 |
| 79.2 | Infectious mononucleosis | infectious diseases | DR | 1.06 | 0.86 | 1.31 | 0.791742781 | 128777 | 2 |
| 523.1 | Gingivitis | digestive | DR | 1.01 | 0.99 | 1.02 | 0.79352277 | 128777 | 2 |
| 346.2 | Nonspecific abnormal results of function study of brain and central nervous system | neurological | DR | 1.02 | 0.95 | 1.09 | 0.796754768 | 128777 | 2 |
| 714 | Rheumatoid arthritis and other inflammatory polyarthropathies | musculoskeletal | DR | 1.01 | 0.98 | 1.04 | 0.799179671 | 128777 | 2 |
| 756.22 | Pectus carinatum | congenital anomalies | DR | 0.00 | 0.00 | 1.14E+10 | 0.801708288 | 111475 | 1 |
| 270.12 | Phenylketonuria [PKU] | endocrine/metabolic | DR | 0.00 | 0.00 | 4.32E+11 | 0.802288702 | 111475 | 1 |
| 286.11 | Von willebrand's disease | hematopoietic | DR | 1.06 | 0.83 | 1.35 | 0.802750759 | 111475 | 1 |
| 425.11 | Hypertrophic obstructive cardiomyopathy | circulatory system | DR | 1.02 | 0.93 | 1.13 | 0.803488731 | 128777 | 2 |
| 743.12 | | | DR | 0.97 | 0.86 | 1.10 | 0.805855937 | 128777 | 2 |
| 963.1 | Antineoplastic and immunosuppressive drugs causing adverse effects | injuries & poisonings | DR | 1.01 | 0.96 | 1.06 | 0.808271919 | 128777 | 2 |
| 246.7 | Abnormal results of function study of thyroid | endocrine/metabolic | DR | 0.99 | 0.96 | 1.03 | 0.809770468 | 128777 | 2 |
| 245.1 | Thyroiditis, acute and subacute | endocrine/metabolic | DR | 0.96 | 0.82 | 1.13 | 0.812118511 | 128777 | 2 |
| 728.7 | Fasciitis | musculoskeletal | DR | 1.00 | 0.98 | 1.01 | 0.812403804 | 128777 | 2 |
| 729.7 | Nontraumatic compartment syndrome | musculoskeletal | DR | 1.04 | 0.88 | 1.22 | 0.812679429 | 111475 | 1 |
| 261.3 | Vitamin C deficiencies | endocrine/metabolic | DR | 0.94 | 0.73 | 1.22 | 0.813817122 | 111475 | 1 |
| 613.7 | Other signs and symptoms in breast | genitourinary | DR | 0.98 | 0.92 | 1.06 | 0.816010303 | 128777 | 2 |
| 721.2 | Spondylosis with myelopathy | musculoskeletal | DR | 0.99 | 0.96 | 1.03 | 0.817395084 | 128777 | 2 |
| 960.3 | | | DR | 1.12 | 0.69 | 1.82 | 0.818859888 | 111475 | 1 |
| 174.11 | Malignant neoplasm of female breast | neoplasms | DR | 1.02 | 0.95 | 1.09 | 0.819994643 | 128777 | 2 |
| 695.81 | Erythema nodosum | dermatologic | DR | 0.94 | 0.72 | 1.23 | 0.825326491 | 111475 | 1 |
| 528.4 | Cysts of oral soft tissues | digestive | DR | 0.98 | 0.88 | 1.09 | 0.825378365 | 111475 | 1 |

| | | | | | | | | | |
|---|---|---|---|---|---|---|---|---|---|
| 610.3 | Fibrosclerosis of breast | genitourinary | DR | 1.04 | 0.88 | 1.22 | 0.830421542 | 128777 | 2 |
| 636.8 | Cervical incompetence | pregnancy complications | DR | 0.00 | 0.00 | 1.82E+16 | 0.831121969 | 111475 | 1 |
| 379.51 | Pigmentary iris degeneration | sense organs | DR | 0.98 | 0.91 | 1.06 | 0.831242237 | 128777 | 2 |
| 1003 | | | DR | 1.09 | 0.73 | 1.63 | 0.831949981 | 111475 | 1 |
| 647 | Infectious and parasitic complications affecting pregnancy | pregnancy complications | DR | 1.05 | 0.82 | 1.35 | 0.835095332 | 128777 | 2 |
| 306.1 | Mental disorders durring/after pregnancy | mental disorders | DR | 1.05 | 0.82 | 1.36 | 0.83633297 | 128777 | 2 |
| 656.6 | Perinatal disorders of digestive system | pregnancy complications | DR | 1.06 | 0.81 | 1.38 | 0.839331699 | 111475 | 1 |
| 425.12 | Other hypertrophic cardiomyopathy | circulatory system | DR | 1.02 | 0.92 | 1.14 | 0.839738531 | 128777 | 2 |
| 145.3 | Cancer of major salivary glands | neoplasms | DR | 1.02 | 0.92 | 1.13 | 0.839847353 | 128777 | 2 |
| 649 | Other conditions or status of the mother complicating pregnancy, childbirth, or the puerperium | pregnancy complications | DR | 1.03 | 0.90 | 1.18 | 0.843902262 | 128777 | 2 |
| 187.8 | Neoplasm of uncertain behavior of male genital organs | neoplasms | DR | 1.02 | 0.93 | 1.12 | 0.844347327 | 111475 | 1 |
| 261.41 | Rickets or osteomalacia | endocrine/metabolic | DR | 0.98 | 0.87 | 1.10 | 0.846649315 | 128777 | 2 |
| 530 | Diseases of esophagus | digestive | DR | 1.00 | 0.99 | 1.02 | 0.851465713 | 128777 | 2 |
| 639 | Complications following abortion or ectopic and molar pregnancies | pregnancy complications | DR | 0.00 | 0.00 | 3.29E+15 | 0.851597645 | 111475 | 1 |
| 754.2 | Spondylolisthesis, congenital | congenital anomalies | DR | 0.99 | 0.93 | 1.05 | 0.85281569 | 128777 | 2 |
| 870.5 | Open wound of lip and mouth | injuries & poisonings | DR | 0.99 | 0.91 | 1.07 | 0.853839323 | 128777 | 2 |
| 264.9 | Lack of normal physiological development, unspecified | endocrine/metabolic | DR | 1.15 | 0.53 | 2.49 | 0.85528806 | 111475 | 1 |
| 530.1 | Esophagitis, GERD and related diseases | digestive | DR | 1.00 | 0.98 | 1.01 | 0.855454788 | 128777 | 2 |
| 691.3 | Congenital pigmentary anomalies of skin | dermatologic | DR | 0.95 | 0.73 | 1.24 | 0.856883154 | 111475 | 1 |
| 638 | Other high-risk pregnancy | pregnancy complications | DR | 1.03 | 0.85 | 1.25 | 0.859879284 | 128777 | 2 |
| 618.2 | Uterine/Uterovaginal prolapse | genitourinary | DR | 1.03 | 0.88 | 1.19 | 0.861111662 | 128777 | 2 |
| 191.1 | Cancer of brain and nervous system | neoplasms | DR | 0.99 | 0.91 | 1.07 | 0.864193682 | 128777 | 2 |
| 795.8 | Abnormal tumor markers | symptoms | DR | 0.97 | 0.83 | 1.14 | 0.865727672 | 111475 | 1 |
| 755.61 | Congenital hip dysplasia and deformity | congenital anomalies | DR | 1.06 | 0.75 | 1.49 | 0.866543686 | 111475 | 1 |
| 229.1 | Benign neoplasm of lymph nodes | neoplasms | DR | 0.98 | 0.86 | 1.11 | 0.866751321 | 111475 | 1 |
| 264.1 | Short stature | endocrine/metabolic | DR | 1.23 | 0.35 | 4.28 | 0.867341439 | 111475 | 1 |
| 654.2 | Rhesus isoimmunization in pregnancy | pregnancy complications | DR | 0.88 | 0.40 | 1.94 | 0.869555059 | 111475 | 1 |
| 610 | | | DR | 1.01 | 0.95 | 1.08 | 0.871344468 | 111475 | 1 |
| 204.21 | Myeloid leukemia, acute | neoplasms | DR | 0.98 | 0.89 | 1.09 | 0.871485359 | 128777 | 2 |
| 346 | Abnormal findings on study of brain and/or nervous system | neurological | DR | 1.01 | 0.95 | 1.07 | 0.87425678 | 128777 | 2 |
| 383 | Otosclerosis | sense organs | DR | 1.02 | 0.90 | 1.15 | 0.87834322 | 111475 | 1 |
| 705.8 | Hyperhidrosis | dermatologic | DR | 0.99 | 0.96 | 1.03 | 0.879723747 | 128777 | 2 |
| 191 | Manlignant and unknown neoplasms of brain and nervous system | neoplasms | DR | 1.01 | 0.95 | 1.07 | 0.88280092 | 128777 | 2 |
| 242.31 | | | DR | 0.95 | 0.69 | 1.32 | 0.882913478 | 111475 | 1 |
| 613.5 | Mastodynia | genitourinary | DR | 0.99 | 0.94 | 1.05 | 0.883218394 | 128777 | 2 |
| 132 | Infestation (lice, mites) | infectious diseases | DR | 1.01 | 0.96 | 1.06 | 0.889957925 | 128777 | 2 |
| 259.2 | Carcinoid syndrome | endocrine/metabolic | DR | 1.02 | 0.85 | 1.23 | 0.891820982 | 111475 | 1 |
| 527.8 | Other specified diseases of the salivary glands | digestive | DR | 0.99 | 0.90 | 1.09 | 0.899219055 | 128777 | 2 |
| 258.1 | Postablative ovarian failure | endocrine/metabolic | DR | 0.95 | 0.62 | 1.45 | 0.901064792 | 111475 | 1 |
| 526.8 | | | DR | 0.99 | 0.92 | 1.07 | 0.902415577 | 111475 | 1 |
| 446.6 | Polyarteritis nodosa | circulatory system | DR | 1.03 | 0.80 | 1.34 | 0.903070791 | 111475 | 1 |
| 293 | Symptoms involving head and neck | mental disorders | DR | 1.01 | 0.95 | 1.07 | 0.906073726 | 128777 | 2 |
| 627.2 | Symptomatic menopause | genitourinary | DR | 1.01 | 0.95 | 1.06 | 0.909833198 | 128777 | 2 |
| 965.2 | | | DR | 1.01 | 0.89 | 1.15 | 0.909950846 | 111475 | 1 |
| 371.2 | Conjunctivitis, noninfectious | sense organs | DR | 1.00 | 0.98 | 1.03 | 0.913299323 | 128777 | 2 |
| 613.8 | Other specified disorders of breast | genitourinary | DR | 0.99 | 0.93 | 1.07 | 0.917638772 | 128777 | 2 |
| 345.12 | Partial epilepsy | neurological | DR | 1.01 | 0.96 | 1.06 | 0.917997082 | 128777 | 2 |
| 286.81 | Primary hypercoagulable state | hematopoietic | DR | 1.01 | 0.95 | 1.07 | 0.919653544 | 128777 | 2 |
| 352.1 | Trigeminal nerve disorders [CN5] | neurological | DR | 1.00 | 0.95 | 1.05 | 0.92113268 | 128777 | 2 |
| 522.5 | Periapical abscess | digestive | DR | 1.00 | 0.97 | 1.02 | 0.921220168 | 128777 | 2 |
| 134 | Helminthiases | infectious diseases | DR | 1.01 | 0.90 | 1.13 | 0.924387764 | 111475 | 1 |
| 619.3 | Noninflammatory disorders of cervix | genitourinary | DR | 1.01 | 0.87 | 1.18 | 0.931208233 | 128777 | 2 |
| 674 | Other complications of the puerperium NEC | pregnancy complications | DR | 1.02 | 0.83 | 1.24 | 0.931517311 | 128777 | 2 |
| 198.3 | Secondary malignant neoplasm of digestive systems | neoplasms | DR | 1.01 | 0.93 | 1.09 | 0.935443192 | 128777 | 2 |
| 512.1 | Wheezing | respiratory | DR | 1.00 | 0.98 | 1.03 | 0.935852601 | 128777 | 2 |
| 480.12 | Pseudomonal pneumonia | respiratory | DR | 1.01 | 0.90 | 1.13 | 0.938371656 | 128777 | 2 |
| 327.71 | Restless legs syndrome | neurological | DR | 1.00 | 0.98 | 1.03 | 0.939254264 | 128777 | 2 |
| 619.5 | Noninflammatory disorders of vulva and perineum | genitourinary | DR | 1.01 | 0.88 | 1.16 | 0.9408539 | 128777 | 2 |
| 255.21 | Glucocorticoid deficiency | endocrine/metabolic | DR | 1.00 | 0.93 | 1.06 | 0.952828123 | 128777 | 2 |
| 754.1 | | | DR | 1.01 | 0.91 | 1.11 | 0.958135129 | 111475 | 1 |
| 260.7 | Polyphagia | endocrine/metabolic | DR | 0.99 | 0.86 | 1.14 | 0.963815123 | 111475 | 1 |
| 500.1 | Extrinsic allergic alveolitis | respiratory | DR | 0.99 | 0.88 | 1.12 | 0.963938527 | 111475 | 1 |
| 202.24 | Large cell lymphoma | neoplasms | DR | 1.01 | 0.86 | 1.18 | 0.965426542 | 128777 | 2 |
| 527.1 | Hypertrophy of salivary gland | digestive | DR | 1.01 | 0.85 | 1.20 | 0.971023837 | 111475 | 1 |
| 750.11 | Esophageal atresia/tracheoesophageal fistula | congenital anomalies | DR | 1.00 | 0.91 | 1.10 | 0.973865241 | 111475 | 1 |
| 305.21 | Anorexia nervosa | mental disorders | DR | 1.01 | 0.77 | 1.33 | 0.974606642 | 111475 | 1 |
| 610.4 | Benign neoplasm of breast | genitourinary | DR | 1.00 | 0.92 | 1.10 | 0.975609893 | 128777 | 2 |
| 283.21 | Hemolytic-uremic syndrome | hematopoietic | DR | 1.01 | 0.63 | 1.62 | 0.980426201 | 111475 | 1 |
| 586.12 | Vesicoureteral reflux | genitourinary | DR | 1.00 | 0.79 | 1.26 | 0.98617403 | 128777 | 2 |
| 31.1 | Leprosy | infectious diseases | DR | 1.01 | 0.42 | 2.45 | 0.989428226 | 111475 | 1 |
| 134.1 | Intestinal helminthiases | infectious diseases | DR | 1.00 | 0.85 | 1.17 | 0.990846462 | 111475 | 1 |
| 759 | Other and unspecified congenital anomalies | congenital anomalies | DR | 1.00 | 0.92 | 1.10 | 0.990848199 | 128777 | 2 |
| 245 | Thyroiditis | endocrine/metabolic | DR | 1.00 | 0.93 | 1.07 | 0.991254071 | 128777 | 2 |
| 733.6 | | | DR | 1.00 | 0.94 | 1.06 | 0.991470452 | 128777 | 2 |
| 656.4 | Hemorrhage of fetus or newborn | pregnancy complications | DR | 1.00 | 0.65 | 1.56 | 0.992532349 | 111475 | 1 |
