## Supplemental Table 9 for "Development of Portable Electronic Health Record Based Algorithms to Identify Individuals with Diabetic Retinopathy"

| Supplemental Table 9. Hispanic-Ethnicity Phenome-Wide Association Study Results for Diabetic Retinopathy Algorithm Designation adjusted for age, sex, and duration of diabetes | | | | | | | | |
|---|---|---|---|---|---|---|---|---|
| PheCode | Phenotype | Phenotype Goup | Predictor | OR | LCI | UCI | p | n_total |
| 250.7 | Diabetic retinopathy | endocrine/metabolic | DR | 635.20 | 579.65 | 696.08 | <1E-300 | 15621 |
| 250.23 | Type 2 diabetes with ophthalmic manifestations | endocrine/metabolic | DR | 100.14 | 94.38 | 106.24 | <1E-300 | 15621 |
| 366 | Cataract | sense organs | DR | 8.70 | 8.22 | 9.20 | <1E-300 | 15621 |
| 250.24 | Type 2 diabetes with neurological manifestations | endocrine/metabolic | DR | 3.80 | 3.67 | 3.94 | <1E-300 | 15621 |
| 250.22 | Type 2 diabetes with renal manifestations | endocrine/metabolic | DR | 4.24 | 4.08 | 4.41 | 3.06E-292 | 15621 |
| 362 | Other retinal disorders | sense organs | DR | 3.50 | 3.38 | 3.63 | 1.46E-261 | 15621 |
| 366.2 | Senile cataract | sense organs | DR | 4.89 | 4.67 | 5.12 | 5.49E-251 | 15621 |
| 585.3 | Chronic renal failure [CKD] | genitourinary | DR | 3.71 | 3.56 | 3.86 | 4.66E-238 | 15621 |
| 585 | | | DR | 3.26 | 3.14 | 3.38 | 5.34E-233 | 15621 |
| 250.1 | Type 1 diabetes | endocrine/metabolic | DR | 3.56 | 3.42 | 3.71 | 3.60E-217 | 15621 |
| 250.3 | Insulin pump user | endocrine/metabolic | DR | 4.40 | 4.19 | 4.61 | 4.42E-212 | 15621 |
| 401.22 | Hypertensive chronic kidney disease | circulatory system | DR | 3.86 | 3.70 | 4.04 | 5.70E-205 | 15621 |
| 250.6 | Polyneuropathy in diabetes | endocrine/metabolic | DR | 3.96 | 3.78 | 4.14 | 7.09E-204 | 15621 |
| 401.2 | Hypertensive heart and/or renal disease | circulatory system | DR | 3.05 | 2.93 | 3.16 | 1.69E-194 | 15621 |
| 585.33 | Chronic Kidney Disease, Stage III | genitourinary | DR | 3.65 | 3.49 | 3.81 | 5.21E-185 | 15621 |
| 285 | Other anemias | hematopoietic | DR | 2.80 | 2.70 | 2.90 | 3.34E-178 | 15621 |
| 707 | Chronic ulcer of skin | dermatologic | DR | 3.89 | 3.69 | 4.09 | 5.12E-155 | 15621 |
| 285.2 | Anemia of chronic disease | hematopoietic | DR | 4.34 | 4.11 | 4.59 | 5.94E-155 | 15621 |
| 443 | | | DR | 3.04 | 2.91 | 3.17 | 9.26E-154 | 15621 |
| 443.9 | Peripheral vascular disease, unspecified | circulatory system | DR | 3.06 | 2.93 | 3.19 | 6.43E-152 | 15621 |
| 585.1 | Acute renal failure | genitourinary | DR | 2.79 | 2.69 | 2.91 | 2.48E-149 | 15621 |
| 428 | | | DR | 2.94 | 2.82 | 3.07 | 3.06E-148 | 15621 |
| 362.2 | | | DR | 3.04 | 2.91 | 3.17 | 1.75E-146 | 15621 |
| 707.2 | Chronic ulcer of leg or foot | dermatologic | DR | 4.70 | 4.42 | 4.99 | 3.34E-144 | 15621 |
| 285.21 | Anemia in chronic kidney disease | hematopoietic | DR | 5.98 | 5.56 | 6.42 | 3.20E-136 | 15621 |
| 411.4 | Coronary atherosclerosis | circulatory system | DR | 2.43 | 2.34 | 2.52 | 8.47E-134 | 15621 |
| 585.34 | Chronic Kidney Disease, Stage IV | genitourinary | DR | 6.67 | 6.18 | 7.21 | 1.12E-133 | 15621 |
| 250.41 | Impaired fasting glucose | endocrine/metabolic | DR | 0.25 | 0.23 | 0.26 | 1.97E-132 | 15621 |
| 276.13 | Hyperpotassemia | endocrine/metabolic | DR | 3.34 | 3.18 | 3.52 | 1.67E-129 | 15621 |
| 250.25 | Diabetes type 2 with peripheral circulatory disorders | endocrine/metabolic | DR | 3.56 | 3.38 | 3.75 | 1.14E-128 | 15621 |
| 249 | Secondary diabetes mellitus | endocrine/metabolic | DR | 3.36 | 3.19 | 3.54 | 4.61E-123 | 15621 |
| 428.1 | Congestive heart failure (CHF) NOS | circulatory system | DR | 2.93 | 2.80 | 3.07 | 6.17E-123 | 15621 |
| 362.9 | Retinal edema | sense organs | DR | 11.17 | 10.08 | 12.38 | 3.26E-121 | 15621 |
| 379 | Other disorders of eye | sense organs | DR | 2.39 | 2.30 | 2.48 | 7.59E-119 | 15621 |
| 110.11 | Dermatophytosis of nail | infectious diseases | DR | 2.24 | 2.16 | 2.32 | 1.26E-118 | 15621 |
| 411 | | | DR | 2.25 | 2.17 | 2.33 | 3.57E-117 | 15621 |
| 276 | | | DR | 2.19 | 2.12 | 2.27 | 1.47E-112 | 15621 |
| 585.32 | End stage renal disease | genitourinary | DR | 7.66 | 6.99 | 8.40 | 5.75E-109 | 15621 |
| 440 | Atherosclerosis | circulatory system | DR | 3.13 | 2.97 | 3.29 | 5.16E-108 | 15621 |
| 269 | Proteinuria | endocrine/metabolic | DR | 3.03 | 2.88 | 3.19 | 1.34E-107 | 15621 |
| 362.3 | Other nondiabetic retinopathy | sense organs | DR | 3.15 | 2.99 | 3.32 | 1.88E-107 | 15621 |
| 411.8 | Other chronic ischemic heart disease, unspecified | circulatory system | DR | 2.35 | 2.26 | 2.44 | 4.18E-107 | 15621 |
| 782.3 | Edema | symptoms | DR | 2.23 | 2.15 | 2.31 | 1.13E-105 | 15621 |
| 401.1 | Essential hypertension | circulatory system | DR | 6.46 | 5.92 | 7.04 | 1.84E-103 | 15621 |
| 401 | Hypertension | circulatory system | DR | 6.70 | 6.13 | 7.32 | 1.23E-102 | 15621 |
| 428.3 | Heart failure with reduced EF [Systolic or combined heart failure] | circulatory system | DR | 3.01 | 2.86 | 3.17 | 2.15E-101 | 15621 |
| 276.1 | Electrolyte imbalance | endocrine/metabolic | DR | 2.16 | 2.08 | 2.24 | 4.44E-99 | 15621 |
| 433 | Cerebrovascular disease | circulatory system | DR | 2.24 | 2.16 | 2.33 | 1.04E-97 | 15621 |
| 580 | | | DR | 4.20 | 3.92 | 4.50 | 1.34E-95 | 15621 |
| 588 | Disorders resulting from impaired renal function | genitourinary | DR | 5.25 | 4.85 | 5.69 | 1.65E-94 | 15621 |
| 703 | Diseases of nail, NOS | dermatologic | DR | 2.22 | 2.14 | 2.31 | 1.09E-91 | 15621 |
| 440.2 | Atherosclerosis of the extremities | circulatory system | DR | 3.56 | 3.34 | 3.79 | 1.53E-90 | 15621 |
| 280 | | | DR | 2.40 | 2.30 | 2.50 | 1.59E-90 | 15621 |
| 251.1 | Hypoglycemia | endocrine/metabolic | DR | 3.33 | 3.14 | 3.54 | 3.28E-89 | 15621 |
| 588.2 | Secondary hyperparathyroidism (of renal origin) | genitourinary | DR | 6.84 | 6.21 | 7.53 | 4.76E-88 | 15621 |
| 280.1 | Iron deficiency anemias, unspecified or not due to blood loss | hematopoietic | DR | 2.42 | 2.32 | 2.53 | 7.10E-88 | 15621 |
| 580.3 | Nephritis and nephropathy without mention of glomerulonephritis | genitourinary | DR | 4.64 | 4.29 | 5.02 | 1.60E-86 | 15621 |
| 367.4 | Presbyopia | sense organs | DR | 2.48 | 2.37 | 2.60 | 2.29E-85 | 15621 |
| 428.4 | Heart failure with preserved EF [Diastolic heart failure] | circulatory system | DR | 3.12 | 2.94 | 3.31 | 5.18E-82 | 15621 |
| 379.2 | Disorders of vitreous body | sense organs | DR | 2.18 | 2.09 | 2.27 | 1.92E-81 | 15621 |
| 710 | Osteomyelitis, periostitis, and other infections involving bone | musculoskeletal | DR | 4.26 | 3.94 | 4.59 | 5.37E-81 | 15621 |
| 710.1 | Osteomyelitis | musculoskeletal | DR | 4.39 | 4.06 | 4.74 | 9.12E-81 | 15621 |
| 580.31 | Nephritis and nephropathy in diseases classified elsewhere | genitourinary | DR | 5.86 | 5.34 | 6.43 | 1.34E-80 | 15621 |
| 250.13 | Type 1 diabetes with ophthalmic manifestations | endocrine/metabolic | DR | 24.94 | 21.03 | 29.58 | 2.47E-79 | 15621 |
| 681 | Superficial cellulitis and abscess | dermatologic | DR | 1.91 | 1.85 | 1.98 | 4.59E-79 | 15621 |
| 362.26 | Macular puckering of retina | sense organs | DR | 2.96 | 2.80 | 3.14 | 8.80E-79 | 15621 |
| 458 | Hypotension | circulatory system | DR | 2.18 | 2.09 | 2.28 | 1.22E-78 | 15621 |
| 585.2 | Renal failure NOS | genitourinary | DR | 4.25 | 3.94 | 4.59 | 1.75E-78 | 15621 |
| 367 | Disorders of refraction and accommodation; blindness and low vision | sense organs | DR | 3.51 | 3.28 | 3.75 | 2.91E-77 | 15621 |
| 362.4 | Retinal vascular changes and abnomalities | sense organs | DR | 3.76 | 3.50 | 4.04 | 5.12E-77 | 15621 |
| 411.2 | Myocardial infarction | circulatory system | DR | 2.31 | 2.21 | 2.42 | 1.25E-74 | 15621 |
| 585.31 | Renal dialysis | genitourinary | DR | 9.52 | 8.42 | 10.77 | 2.10E-74 | 15621 |
| 110.1 | Dermatophytosis | infectious diseases | DR | 1.88 | 1.81 | 1.94 | 3.37E-74 | 15621 |
| 110 | | | DR | 1.86 | 1.80 | 1.93 | 3.35E-72 | 15621 |
| 700 | Corns and callosities | dermatologic | DR | 2.22 | 2.12 | 2.32 | 3.39E-72 | 15621 |
| 350.2 | Abnormality of gait | neurological | DR | 1.94 | 1.87 | 2.01 | 6.45E-71 | 15621 |
| 365 | Glaucoma | sense organs | DR | 1.84 | 1.78 | 1.91 | 4.67E-69 | 15621 |
| 710.19 | Unspecified osteomyelitis | musculoskeletal | DR | 4.68 | 4.28 | 5.12 | 5.08E-67 | 15621 |
| 276.6 | Fluid overload | endocrine/metabolic | DR | 3.88 | 3.59 | 4.20 | 1.02E-65 | 15621 |
| 350 | | | DR | 1.83 | 1.76 | 1.89 | 1.33E-64 | 15621 |
| 250.4 | Abnormal glucose | endocrine/metabolic | DR | 0.54 | 0.52 | 0.56 | 1.67E-64 | 15621 |
| 362.23 | Cystoid macular degeneration of retina | sense organs | DR | 4.77 | 4.35 | 5.23 | 9.63E-64 | 15621 |
| 710.11 | Acute osteomyelitis | musculoskeletal | DR | 6.22 | 5.57 | 6.95 | 4.91E-62 | 15621 |
| 433.1 | Occlusion and stenosis of precerebral arteries | circulatory system | DR | 2.42 | 2.29 | 2.55 | 6.84E-62 | 15621 |
| 276.4 | Acid-base balance disorder | endocrine/metabolic | DR | 2.46 | 2.33 | 2.59 | 1.41E-61 | 15621 |
| 356 | Hereditary and idiopathic peripheral neuropathy | neurological | DR | 2.07 | 1.98 | 2.16 | 1.96E-61 | 15621 |
| 276.41 | Acidosis | endocrine/metabolic | DR | 2.57 | 2.43 | 2.72 | 4.52E-61 | 15621 |
| 41 | Bacterial infection NOS | infectious diseases | DR | 1.95 | 1.88 | 2.03 | 4.61E-61 | 15621 |
| 854 | Complications of cardiac/vascular device, implant, and graft | injuries & poisonings | DR | 3.77 | 3.48 | 4.09 | 1.30E-60 | 15621 |
| 277 | Other disorders of metabolism | endocrine/metabolic | DR | 3.54 | 3.27 | 3.82 | 5.20E-60 | 15621 |
| 871 | Open wounds of extremities | injuries & poisonings | DR | 2.05 | 1.96 | 2.14 | 2.08E-59 | 15621 |
| 458.9 | Hypotension NOS | circulatory system | DR | 2.15 | 2.05 | 2.26 | 8.97E-59 | 15621 |
| 38 | Septicemia | infectious diseases | DR | 2.20 | 2.09 | 2.31 | 1.07E-58 | 15621 |

| | | | | | | | | |
|---|---|---|---|---|---|---|---|---|
| 710.12 | Chronic osteomyelitis | musculoskeletal | DR | 5.37 | 4.84 | 5.96 | 2.59E-58 | 15621 |
| 433.3 | Cerebral ischemia | circulatory system | DR | 2.17 | 2.07 | 2.28 | 7.73E-58 | 15621 |
| 707.1 | Decubitus ulcer | dermatologic | DR | 3.91 | 3.58 | 4.26 | 4.20E-56 | 15621 |
| 1089 | Acquired absence of limb | | DR | 6.10 | 5.43 | 6.85 | 1.12E-54 | 15621 |
| 271.3 | Intestinal disaccharidase deficiencies and disaccharide malabsorption | endocrine/metabolic | DR | 0.23 | 0.21 | 0.25 | 2.60E-54 | 15621 |
| 433.31 | Transient cerebral ischemia | circulatory system | DR | 2.17 | 2.06 | 2.28 | 2.71E-54 | 15621 |
| 275 | Disorders of mineral metabolism | endocrine/metabolic | DR | 2.04 | 1.95 | 2.14 | 3.52E-54 | 15621 |
| 871.3 | Open wound of foot except toe(s) alone | injuries & poisonings | DR | 3.56 | 3.28 | 3.87 | 6.64E-54 | 15621 |
| 271 | | | DR | 0.24 | 0.22 | 0.27 | 8.85E-54 | 15621 |
| 791 | Gangrene | symptoms | DR | 9.94 | 8.56 | 11.54 | 2.14E-53 | 15621 |
| 507 | Pleurisy; pleural effusion | respiratory | DR | 2.57 | 2.42 | 2.73 | 2.24E-53 | 15621 |
| 681.5 | Cellulitis and abscess of leg, except foot | dermatologic | DR | 2.17 | 2.06 | 2.28 | 4.62E-53 | 15621 |
| 433.2 | Occlusion of cerebral arteries | circulatory system | DR | 2.34 | 2.21 | 2.47 | 9.65E-53 | 15621 |
| 440.21 | Atherosclerosis of native arteries of the extremities with ulceration or gangrene | circulatory system | DR | 9.00 | 7.79 | 10.40 | 3.56E-52 | 15621 |
| 772.3 | Muscle weakness | symptoms | DR | 1.95 | 1.87 | 2.04 | 4.79E-52 | 15621 |
| 427.2 | Atrial fibrillation and flutter | circulatory system | DR | 2.08 | 1.98 | 2.19 | 8.70E-51 | 15621 |
| 426 | Cardiac conduction disorders | circulatory system | DR | 1.76 | 1.69 | 1.82 | 1.29E-50 | 15621 |
| 362.8 | Retinal hemorrhage/ischemia | sense organs | DR | 4.10 | 3.74 | 4.51 | 1.38E-50 | 15621 |
| 600 | Hyperplasia of prostate | genitourinary | DR | 1.73 | 1.67 | 1.79 | 2.42E-50 | 15621 |
| 276.5 | Hypovolemia | endocrine/metabolic | DR | 1.91 | 1.82 | 1.99 | 3.56E-50 | 15621 |
| 433.21 | Cerebral artery occlusion, with cerebral infarction | circulatory system | DR | 2.31 | 2.18 | 2.44 | 4.95E-50 | 15621 |
| 379.3 | Aphakia and other disorders of lens | sense organs | DR | 2.91 | 2.70 | 3.12 | 1.45E-49 | 15621 |
| 250.14 | Type 1 diabetes with neurological manifestations | endocrine/metabolic | DR | 3.88 | 3.53 | 4.25 | 1.10E-48 | 15621 |
| 427.21 | Atrial fibrillation | circulatory system | DR | 2.07 | 1.97 | 2.18 | 1.20E-48 | 15621 |
| 411.3 | Angina pectoris | circulatory system | DR | 1.82 | 1.75 | 1.90 | 2.04E-48 | 15621 |
| 798 | Malaise and fatigue | symptoms | DR | 1.69 | 1.63 | 1.75 | 5.68E-48 | 15621 |
| 681.6 | Cellulitis and abscess of foot, toe | dermatologic | DR | 4.30 | 3.89 | 4.75 | 8.05E-48 | 15621 |
| 367.9 | Blindness and low vision | sense organs | DR | 2.09 | 1.98 | 2.20 | 1.46E-47 | 15621 |
| 994.2 | Sepsis | injuries & poisonings | DR | 2.17 | 2.05 | 2.29 | 8.23E-47 | 15621 |
| 365.1 | Open-angle glaucoma | sense organs | DR | 1.68 | 1.62 | 1.75 | 1.34E-45 | 15621 |
| 591 | Urinary tract infection | genitourinary | DR | 1.74 | 1.67 | 1.81 | 1.41E-45 | 15621 |
| 994 | | | DR | 2.10 | 1.99 | 2.21 | 1.76E-45 | 15621 |
| 707.3 | Chronic ulcer of unspecified site | dermatologic | DR | 3.40 | 3.12 | 3.71 | 2.38E-45 | 15621 |
| 290 | Delirium dementia and amnestic and other cognitive disorders | mental disorders | DR | 1.98 | 1.89 | 2.08 | 3.13E-45 | 15621 |
| 459 | Other disorders of circulatory system | circulatory system | DR | 2.14 | 2.03 | 2.26 | 6.30E-45 | 15621 |
| 509 | Respiratory failure, insufficiency, arrest | respiratory | DR | 1.94 | 1.85 | 2.04 | 7.49E-44 | 15621 |
| 480 | Pneumonia | respiratory | DR | 1.80 | 1.72 | 1.88 | 1.63E-43 | 15621 |
| 411.1 | Unstable angina (intermediate coronary syndrome) | circulatory system | DR | 2.10 | 1.99 | 2.22 | 1.65E-43 | 15621 |
| 585.4 | Chronic kidney disease, Stage I or II | genitourinary | DR | 2.44 | 2.29 | 2.61 | 5.19E-43 | 15621 |
| 365.11 | Primary open angle glaucoma | sense organs | DR | 2.04 | 1.94 | 2.15 | 5.64E-43 | 15621 |
| 532 | Dysphagia | digestive | DR | 1.78 | 1.70 | 1.85 | 1.37E-42 | 15621 |
| 586 | Other disorders of the kidney and ureters | genitourinary | DR | 1.85 | 1.77 | 1.93 | 2.75E-42 | 15621 |
| 509.1 | Respiratory failure | respiratory | DR | 2.10 | 1.99 | 2.22 | 3.69E-42 | 15621 |
| 426.2 | Atrioventricular [AV] block | circulatory system | DR | 2.73 | 2.53 | 2.94 | 1.89E-41 | 15621 |
| 276.12 | Hyposmolality and/or hyponatremia | endocrine/metabolic | DR | 1.98 | 1.88 | 2.08 | 1.98E-41 | 15621 |
| 440.22 | Atherosclerosis of native arteries of the extremities with intermittent claudication | circulatory system | DR | 2.72 | 2.53 | 2.93 | 5.81E-41 | 15621 |
| 275.5 | Disorders of calcium/phosphorus metabolism | endocrine/metabolic | DR | 2.48 | 2.32 | 2.66 | 1.92E-40 | 15621 |
| 375.1 | Dry eyes | sense organs | DR | 1.57 | 1.51 | 1.62 | 4.53E-39 | 15621 |
| 401.21 | Hypertensive heart disease | circulatory system | DR | 1.83 | 1.75 | 1.92 | 5.10E-39 | 15621 |
| 872 | Traumatic amputation | injuries & poisonings | DR | 4.37 | 3.91 | 4.90 | 8.42E-39 | 15621 |
| 401.3 | Other hypertensive complications | circulatory system | DR | 2.48 | 2.31 | 2.66 | 1.32E-38 | 15621 |
| 275.53 | Disorders of phosphorus metabolism | endocrine/metabolic | DR | 3.03 | 2.78 | 3.30 | 2.11E-38 | 15621 |
| 41.1 | Staphylococcus infections | infectious diseases | DR | 2.33 | 2.18 | 2.49 | 2.89E-38 | 15621 |
| 427 | | | DR | 1.55 | 1.50 | 1.60 | 1.15E-37 | 15621 |
| 681.1 | Cellulitis and abscess of fingers/toes | dermatologic | DR | 2.02 | 1.91 | 2.14 | 7.54E-37 | 15621 |
| 395 | Heart valve disorders | circulatory system | DR | 1.98 | 1.87 | 2.09 | 1.17E-36 | 15621 |
| 290.1 | Dementias | mental disorders | DR | 2.24 | 2.10 | 2.39 | 1.97E-36 | 15621 |
| 456 | Chronic venous insufficiency [CVI] | circulatory system | DR | 1.93 | 1.83 | 2.04 | 2.69E-36 | 15621 |
| 741.3 | Difficulty in walking | musculoskeletal | DR | 1.94 | 1.84 | 2.04 | 2.90E-36 | 15621 |
| 433.8 | Late effects of cerebrovascular disease | circulatory system | DR | 2.14 | 2.02 | 2.28 | 3.29E-36 | 15621 |
| 292.4 | Altered mental status | mental disorders | DR | 2.02 | 1.91 | 2.14 | 5.46E-35 | 15621 |
| 512.7 | Shortness of breath | respiratory | DR | 1.54 | 1.48 | 1.59 | 6.20E-35 | 15621 |
| 38.3 | Bacteremia | infectious diseases | DR | 2.46 | 2.29 | 2.65 | 8.77E-35 | 15621 |
| 425 | Cardiomyopathy | circulatory system | DR | 2.08 | 1.96 | 2.21 | 1.08E-34 | 15621 |
| 286.2 | | | DR | 2.00 | 1.89 | 2.11 | 1.14E-34 | 15621 |
| 433.6 | Acute, but ill-defined cerebrovascular disease | circulatory system | DR | 2.30 | 2.15 | 2.46 | 1.46E-34 | 15621 |
| 458.1 | Orthostatic hypotension | circulatory system | DR | 2.09 | 1.97 | 2.22 | 3.18E-34 | 15621 |
| 447 | Other disorders of arteries and arterioles | circulatory system | DR | 3.05 | 2.78 | 3.34 | 5.43E-34 | 15621 |
| 426.9 | Cardiac pacemaker/device in situ | circulatory system | DR | 2.37 | 2.21 | 2.54 | 5.86E-34 | 15621 |
| 605 | Erectile dysfunction [ED] | genitourinary | DR | 1.52 | 1.47 | 1.57 | 7.88E-34 | 15621 |
| 871.4 | Open wound of toe(s) | injuries & poisonings | DR | 4.11 | 3.66 | 4.63 | 2.67E-33 | 15621 |
| 443.8 | Other specified peripheral vascular diseases | circulatory system | DR | 3.90 | 3.48 | 4.36 | 3.37E-33 | 15621 |
| 425.1 | Primary/intrinsic cardiomyopathies | circulatory system | DR | 2.11 | 1.98 | 2.24 | 3.64E-33 | 15621 |
| 427.3 | Other specified cardiac dysrhythmias | circulatory system | DR | 1.69 | 1.62 | 1.77 | 2.09E-32 | 15621 |
| 735.2 | Acquired toe deformities | musculoskeletal | DR | 1.70 | 1.63 | 1.78 | 2.18E-32 | 15621 |
| 389.1 | Sensorineural hearing loss | sense organs | DR | 1.52 | 1.46 | 1.57 | 3.87E-32 | 15621 |
| 599.2 | Retention of urine | genitourinary | DR | 1.80 | 1.71 | 1.89 | 1.77E-31 | 15621 |
| 459.9 | Circulatory disease NEC | circulatory system | DR | 1.98 | 1.86 | 2.10 | 6.09E-31 | 15621 |
| 367.8 | Hypermetropia | sense organs | DR | 1.51 | 1.46 | 1.56 | 8.70E-31 | 15621 |
| 41.2 | Streptococcus infection | infectious diseases | DR | 2.61 | 2.40 | 2.84 | 8.72E-31 | 15621 |
| 916 | Contusion | injuries & poisonings | DR | 1.60 | 1.54 | 1.67 | 1.17E-30 | 15621 |
| 706.8 | Other specified diseases of sebaceous glands | dermatologic | DR | 1.73 | 1.65 | 1.81 | 1.89E-30 | 15621 |
| 272 | | | DR | 2.73 | 2.50 | 2.98 | 2.45E-30 | 15621 |
| 563 | Constipation | digestive | DR | 1.55 | 1.49 | 1.61 | 3.38E-30 | 15621 |
| 272.1 | Hyperlipidemia | endocrine/metabolic | DR | 2.71 | 2.49 | 2.96 | 3.62E-30 | 15621 |
| 337 | Disorders of the autonomic nervous system | neurological | DR | 2.04 | 1.92 | 2.18 | 8.39E-30 | 15621 |
| 525.1 | Loss of teeth or edentulism | digestive | DR | 1.53 | 1.48 | 1.59 | 9.79E-30 | 15621 |
| 367.2 | Astigmatism | sense organs | DR | 1.47 | 1.42 | 1.52 | 1.43E-29 | 15621 |
| 503 | Pulmonary congestion and hypostasis | respiratory | DR | 3.17 | 2.86 | 3.52 | 2.89E-29 | 15621 |
| 771.1 | Swelling of limb | symptoms | DR | 1.82 | 1.72 | 1.92 | 4.83E-29 | 15621 |
| 457 | | | DR | 2.14 | 2.00 | 2.29 | 7.17E-29 | 15621 |
| 337.1 | Peripheral autonomic neuropathy | neurological | DR | 2.06 | 1.93 | 2.20 | 8.28E-29 | 15621 |
| 426.91 | Cardiac pacemaker in situ | circulatory system | DR | 2.54 | 2.33 | 2.76 | 8.70E-29 | 15621 |
| 252.1 | Hyperparathyroidism | endocrine/metabolic | DR | 3.35 | 3.00 | 3.74 | 1.40E-28 | 15621 |

| | | | | | | | | |
|---|---|---|---|---|---|---|---|---|
| 252 | Disorders of parathyroid gland | endocrine/metabolic | DR | 3.14 | 2.83 | 3.48 | 2.54E-28 | 15621 |
| 275.3 | Disorders of magnesium metabolism | endocrine/metabolic | DR | 1.96 | 1.84 | 2.09 | 1.61E-27 | 15621 |
| 915 | Superficial injury without mention of infection | injuries & poisonings | DR | 1.64 | 1.57 | 1.72 | 4.72E-27 | 15621 |
| 851 | Complications of transplants and reattached limbs | injuries & poisonings | DR | 2.48 | 2.28 | 2.69 | 6.21E-27 | 15621 |
| 418 | Nonspecific chest pain | circulatory system | DR | 1.44 | 1.39 | 1.49 | 6.33E-27 | 15621 |
| 911 | Blister | injuries & poisonings | DR | 2.74 | 2.50 | 3.01 | 9.88E-27 | 15621 |
| 208 | Benign neoplasm of colon | neoplasms | DR | 1.44 | 1.39 | 1.49 | 3.05E-26 | 15621 |
| 368.4 | Visual field defects | sense organs | DR | 1.99 | 1.86 | 2.12 | 3.32E-26 | 15621 |
| 429 | | | DR | 1.78 | 1.69 | 1.88 | 4.35E-26 | 15621 |
| 362.21 | Macular degeneration, dry | sense organs | DR | 2.07 | 1.93 | 2.22 | 7.77E-26 | 15621 |
| 457.3 | | | DR | 2.10 | 1.96 | 2.26 | 1.27E-25 | 15621 |
| 340 | Migraine | neurological | DR | 0.53 | 0.50 | 0.57 | 2.28E-25 | 15621 |
| 743 | | | DR | 1.74 | 1.65 | 1.83 | 2.99E-25 | 15621 |
| 395.2 | Nonrheumatic aortic valve disorders | circulatory system | DR | 2.05 | 1.91 | 2.19 | 1.04E-24 | 15621 |
| 1015 | Effects of other external causes | NULL | DR | 1.55 | 1.49 | 1.62 | 1.29E-24 | 15621 |
| 588.1 | Renal osteodystrophy | genitourinary | DR | 4.18 | 3.63 | 4.81 | 1.66E-24 | 15621 |
| 41.11 | Methicillin sensitive Staphylococcus aureus | infectious diseases | DR | 2.49 | 2.28 | 2.73 | 2.13E-24 | 15621 |
| 427.22 | Atrial flutter | circulatory system | DR | 2.13 | 1.97 | 2.29 | 2.26E-24 | 15621 |
| 788 | Syncope and collapse | symptoms | DR | 1.59 | 1.52 | 1.67 | 2.30E-24 | 15621 |
| 1011 | Complications of surgical and medical procedures | NULL | DR | 1.81 | 1.71 | 1.92 | 2.85E-24 | 15621 |
| 395.1 | Nonrheumatic mitral valve disorders | circulatory system | DR | 2.13 | 1.98 | 2.30 | 5.32E-24 | 15621 |
| 426.21 | First degree AV block | circulatory system | DR | 2.82 | 2.55 | 3.13 | 6.30E-24 | 15621 |
| 260 | Protein-calorie malnutrition | endocrine/metabolic | DR | 2.01 | 1.88 | 2.16 | 9.75E-24 | 15621 |
| 525 | Other diseases of the teeth and supporting structures | digestive | DR | 1.41 | 1.36 | 1.45 | 1.18E-23 | 15621 |
| 374 | Other disorders of eyelids | sense organs | DR | 1.61 | 1.54 | 1.69 | 1.22E-23 | 15621 |
| 580.2 | Nephrotic syndrome without mention of glomerulonephritis | genitourinary | DR | 4.88 | 4.17 | 5.72 | 1.36E-23 | 15621 |
| 416 | Cardiomegaly | circulatory system | DR | 1.86 | 1.75 | 1.98 | 1.37E-23 | 15621 |
| 874 | Complication of amputation stump | injuries & poisonings | DR | 4.81 | 4.11 | 5.62 | 1.56E-23 | 15621 |
| 415.2 | Chronic pulmonary heart disease | circulatory system | DR | 2.03 | 1.89 | 2.18 | 2.80E-23 | 15621 |
| 735.21 | Hammer toe (acquired) | musculoskeletal | DR | 1.80 | 1.70 | 1.91 | 3.58E-23 | 15621 |
| 351 | Other peripheral nerve disorders | neurological | DR | 1.41 | 1.36 | 1.46 | 3.75E-23 | 15621 |
| 701.1 | Keratoderma, acquired | dermatologic | DR | 1.83 | 1.72 | 1.95 | 7.55E-23 | 15621 |
| 380.4 | Impacted cerumen | sense organs | DR | 1.50 | 1.44 | 1.56 | 7.56E-23 | 15621 |
| 348.8 | Encephalopathy, not elsewhere classified | neurological | DR | 2.09 | 1.94 | 2.25 | 8.20E-23 | 15621 |
| 428.2 | Heart failure NOS | circulatory system | DR | 2.47 | 2.25 | 2.71 | 1.04E-22 | 15621 |
| 280.2 | Iron deficiency anemia secondary to blood loss (chronic) | hematopoietic | DR | 2.17 | 2.01 | 2.35 | 1.10E-22 | 15621 |
| 573.7 | Abnormal results of function study of liver | digestive | DR | 0.67 | 0.64 | 0.69 | 2.19E-22 | 15621 |
| 366.1 | Nonsenile Cataract | sense organs | DR | 2.41 | 2.20 | 2.64 | 2.59E-22 | 15621 |
| 372 | Disorders of conjunctiva | sense organs | DR | 1.58 | 1.51 | 1.66 | 2.81E-22 | 15621 |
| 292 | Neurological disorders | mental disorders | DR | 1.45 | 1.39 | 1.50 | 3.03E-22 | 15621 |
| 348 | Other conditions of brain | neurological | DR | 1.95 | 1.82 | 2.09 | 3.45E-22 | 15621 |
| 368 | Visual disturbances | sense organs | DR | 1.42 | 1.37 | 1.48 | 4.91E-22 | 15621 |
| 300.1 | Anxiety disorder | mental disorders | DR | 0.72 | 0.69 | 0.74 | 1.07E-21 | 15621 |
| 285.1 | Acute posthemorrhagic anemia | hematopoietic | DR | 2.03 | 1.88 | 2.18 | 2.50E-21 | 15621 |
| 1013 | Asphyxia and hypoxemia | NULL | DR | 1.84 | 1.72 | 1.96 | 2.57E-21 | 15621 |
| 362.29 | Macular degeneration (senile) of retina NOS | sense organs | DR | 2.45 | 2.23 | 2.69 | 3.06E-21 | 15621 |
| 370 | Keratitis | sense organs | DR | 1.69 | 1.60 | 1.78 | 3.48E-21 | 15621 |
| 480.1 | Bacterial pneumonia | respiratory | DR | 1.88 | 1.76 | 2.02 | 4.96E-21 | 15621 |
| 426.3 | Bundle branch block | circulatory system | DR | 1.86 | 1.74 | 1.98 | 1.01E-20 | 15621 |
| 789 | Nausea and vomiting | symptoms | DR | 1.47 | 1.41 | 1.53 | 1.88E-20 | 15621 |
| 41.12 | Methicillin resistant Staphylococcus aureus | infectious diseases | DR | 2.52 | 2.28 | 2.78 | 2.08E-20 | 15621 |
| 731 | Osteitis deformans and osteopathies associated with other disorders classified elsewhere | musculoskeletal | DR | 5.12 | 4.29 | 6.11 | 2.95E-20 | 15621 |
| 415 | Pulmonary heart disease | circulatory system | DR | 1.75 | 1.65 | 1.86 | 5.10E-20 | 15621 |
| 735 | Acquired foot deformities | musculoskeletal | DR | 1.40 | 1.35 | 1.46 | 6.03E-20 | 15621 |
| 458.2 | Iatrogenic hypotension | circulatory system | DR | 2.35 | 2.14 | 2.58 | 6.27E-20 | 15621 |
| 327.32 | Obstructive sleep apnea | neurological | DR | 0.73 | 0.71 | 0.76 | 6.64E-20 | 15621 |
| 379.5 | Disorders of iris and ciliary body | sense organs | DR | 2.45 | 2.22 | 2.71 | 9.08E-20 | 15621 |
| 799 | | | DR | 1.76 | 1.65 | 1.87 | 1.02E-19 | 15621 |
| 713.5 | Arthropathy associated with neurological disorders | musculoskeletal | DR | 6.69 | 5.43 | 8.25 | 1.07E-19 | 15621 |
| 587 | Kidney replaced by transpant | genitourinary | DR | 5.82 | 4.79 | 7.06 | 1.21E-19 | 15621 |
| 505 | Other pulmonary inflamation or edema | respiratory | DR | 3.21 | 2.82 | 3.66 | 1.40E-19 | 15621 |
| 290.16 | Vascular dementia | mental disorders | DR | 2.46 | 2.23 | 2.72 | 1.48E-19 | 15621 |
| 357 | Inflammatory and toxic neuropathy | neurological | DR | 1.82 | 1.70 | 1.94 | 2.20E-19 | 15621 |
| 686 | Other local infections of skin and subcutaneous tissue | dermatologic | DR | 1.83 | 1.71 | 1.95 | 2.82E-19 | 15621 |
| 250.12 | Type 1 diabetes with renal manifestations | endocrine/metabolic | DR | 6.41 | 5.21 | 7.89 | 3.85E-19 | 15621 |
| 420 | Carditis | circulatory system | DR | 2.00 | 1.85 | 2.16 | 4.86E-19 | 15621 |
| 327.3 | Sleep apnea | neurological | DR | 0.74 | 0.71 | 0.76 | 5.23E-19 | 15621 |
| 378.5 | Paralytic strabismus | sense organs | DR | 2.73 | 2.44 | 3.05 | 6.65E-19 | 15621 |
| 452 | Other venous embolism and thrombosis | circulatory system | DR | 1.69 | 1.59 | 1.79 | 7.05E-19 | 15621 |
| 819 | Skull and face fracture and other intercranial injury | injuries & poisonings | DR | 0.56 | 0.52 | 0.60 | 1.39E-18 | 15621 |
| 703.1 | Ingrowing nail | dermatologic | DR | 1.50 | 1.43 | 1.57 | 1.76E-18 | 15621 |
| 426.7 | Abnormal electrocardiogram [ECG] [EKG] | circulatory system | DR | 1.45 | 1.39 | 1.52 | 1.92E-18 | 15621 |
| 536.3 | Gastroparesis | digestive | DR | 2.39 | 2.16 | 2.64 | 3.06E-18 | 15621 |
| 512 | Other symptoms of respiratory system | respiratory | DR | 1.36 | 1.31 | 1.41 | 4.63E-18 | 15621 |
| 365.2 | Primary angle-closure glaucoma | sense organs | DR | 1.94 | 1.80 | 2.10 | 5.45E-18 | 15621 |
| 411.9 | Other acute and subacute forms of ischemic heart disease | circulatory system | DR | 2.64 | 2.36 | 2.96 | 6.30E-18 | 15621 |
| 394 | Rheumatic disease of the heart valves | circulatory system | DR | 2.11 | 1.93 | 2.31 | 1.71E-17 | 15621 |
| 41.4 | E. coli | infectious diseases | DR | 2.14 | 1.95 | 2.34 | 1.80E-17 | 15621 |
| 414 | Other forms of chronic heart disease | circulatory system | DR | 1.81 | 1.69 | 1.95 | 2.39E-17 | 15621 |
| 370.31 | Keratoconjunctivitis sicca | sense organs | DR | 1.78 | 1.66 | 1.90 | 2.41E-17 | 15621 |
| 713 | Arthropathy associated with other disorders classified elsewhere | musculoskeletal | DR | 3.64 | 3.13 | 4.25 | 3.02E-17 | 15621 |
| 270.2 | Disorders of amino-acid metabolism | endocrine/metabolic | DR | 0.30 | 0.26 | 0.35 | 4.18E-17 | 15621 |
| 508 | Pulmonary collapse; interstitial and compensatory emphysema | respiratory | DR | 1.76 | 1.64 | 1.88 | 5.32E-17 | 15621 |
| 361 | Retinal detachments and defects | sense organs | DR | 1.75 | 1.63 | 1.87 | 8.00E-17 | 15621 |
| 426.92 | Cardiac defibrillator in situ | circulatory system | DR | 2.18 | 1.98 | 2.39 | 8.47E-17 | 15621 |
| 370.3 | Keratoconjunctivitis | sense organs | DR | 1.71 | 1.61 | 1.83 | 9.14E-17 | 15621 |
| 361.1 | Retinal detachment with retinal defect | sense organs | DR | 2.88 | 2.54 | 3.27 | 1.03E-16 | 15621 |
| 272.11 | Hypercholesterolemia | endocrine/metabolic | DR | 1.36 | 1.31 | 1.41 | 1.31E-16 | 15621 |
| 292.1 | Aphasia/speech disturbance | mental disorders | DR | 1.72 | 1.61 | 1.83 | 1.47E-16 | 15621 |
| 396 | Abnormal heart sounds | circulatory system | DR | 1.72 | 1.61 | 1.84 | 1.62E-16 | 15621 |
| 571.5 | Other chronic nonalcoholic liver disease | digestive | DR | 0.71 | 0.69 | 0.74 | 3.45E-16 | 15621 |
| 514 | Abnormal findings examination of lungs | respiratory | DR | 1.47 | 1.40 | 1.54 | 3.50E-16 | 15621 |
| 1002 | Symptoms concerning nutrition, metabolism, and development | NULL | DR | 1.55 | 1.47 | 1.64 | 5.57E-16 | 15621 |
| 374.6 | Dermatochalasis | sense organs | DR | 1.70 | 1.60 | 1.82 | 6.49E-16 | 15621 |
| 278 | Overweight, obesity and other hyperalimentation | endocrine/metabolic | DR | 0.73 | 0.70 | 0.76 | 7.97E-16 | 15621 |
| 278.1 | Obesity | endocrine/metabolic | DR | 0.74 | 0.72 | 0.77 | 1.07E-15 | 15621 |

| | | | | | | | | |
|---|---|---|---|---|---|---|---|---|
| 426.24 | Atrioventricular block, complete | circulatory system | DR | 2.69 | 2.37 | 3.04 | 1.53E-15 | 15621 |
| 599.1 | Urinary obstruction | genitourinary | DR | 2.18 | 1.98 | 2.41 | 1.70E-15 | 15621 |
| 727.1 | Synovitis and tenosynovitis | musculoskeletal | DR | 1.43 | 1.37 | 1.50 | 2.23E-15 | 15621 |
| 444 | Arterial embolism and thrombosis | circulatory system | DR | 2.46 | 2.20 | 2.76 | 2.24E-15 | 15621 |
| 427.4 | | | DR | 2.59 | 2.30 | 2.92 | 2.48E-15 | 15621 |
| 371 | Inflammation of the eye | sense organs | DR | 1.35 | 1.30 | 1.40 | 2.88E-15 | 15621 |
| 362.22 | Macular degeneration, wet | sense organs | DR | 2.88 | 2.52 | 3.29 | 3.20E-15 | 15621 |
| 444.1 | Arterial embolism and thrombosis of lower extremity artery | circulatory system | DR | 3.65 | 3.10 | 4.30 | 3.75E-15 | 15621 |
| 980 | | | DR | 2.69 | 2.37 | 3.06 | 3.94E-15 | 15621 |
| 496 | Chronic airway obstruction | respiratory | DR | 1.36 | 1.31 | 1.42 | 4.09E-15 | 15621 |
| 561.1 | Diarrhea | digestive | DR | 1.35 | 1.30 | 1.40 | 6.37E-15 | 15621 |
| 270.3 | | | DR | 2.26 | 2.04 | 2.51 | 7.50E-15 | 15621 |
| 80 | Postoperative infection | infectious diseases | DR | 1.95 | 1.79 | 2.12 | 9.28E-15 | 15621 |
| 589 | Abnormal results of function study of kidney | genitourinary | DR | 2.24 | 2.02 | 2.49 | 9.41E-15 | 15621 |
| 743.1 | | | DR | 1.87 | 1.72 | 2.02 | 1.01E-14 | 15621 |
| 287 | Purpura and other hemorrhagic conditions | hematopoietic | DR | 1.52 | 1.44 | 1.61 | 1.11E-14 | 15621 |
| 509.8 | Dependence on respirator [Ventilator] or supplemental oxygen | respiratory | DR | 1.80 | 1.67 | 1.94 | 1.17E-14 | 15621 |
| 743.11 | Osteoporosis NOS | musculoskeletal | DR | 1.87 | 1.72 | 2.03 | 1.23E-14 | 15621 |
| 261.2 | Vitamin B-complex deficiencies | endocrine/metabolic | DR | 1.58 | 1.49 | 1.67 | 1.25E-14 | 15621 |
| 427.42 | Cardiac arrest | circulatory system | DR | 2.84 | 2.48 | 3.26 | 2.20E-14 | 15621 |
| 38.1 | Gram negative septicemia | infectious diseases | DR | 2.12 | 1.92 | 2.34 | 2.33E-14 | 15621 |
| 571 | | | DR | 0.74 | 0.71 | 0.77 | 4.30E-14 | 15621 |
| 599 | Other symptoms/disorders or the urinary system | genitourinary | DR | 1.31 | 1.26 | 1.35 | 4.41E-14 | 15621 |
| 740.9 | Osteoarthrosis NOS | musculoskeletal | DR | 1.29 | 1.25 | 1.34 | 4.47E-14 | 15621 |
| 427.5 | Arrhythmia (cardiac) NOS | circulatory system | DR | 1.40 | 1.34 | 1.46 | 5.13E-14 | 15621 |
| 512.8 | Cough | respiratory | DR | 1.29 | 1.25 | 1.34 | 5.21E-14 | 15621 |
| 433.11 | Occlusion of cerebral arteries, with cerebral infarction | circulatory system | DR | 2.87 | 2.49 | 3.30 | 5.40E-14 | 15621 |
| 561 | Symptoms involving digestive system | digestive | DR | 1.31 | 1.26 | 1.36 | 6.22E-14 | 15621 |
| 281 | Other deficiency anemia | hematopoietic | DR | 1.58 | 1.48 | 1.67 | 6.79E-14 | 15621 |
| 371.3 | Inflammation of eyelids | sense organs | DR | 1.37 | 1.31 | 1.43 | 7.51E-14 | 15621 |
| 580.32 | Nephritis and nephropathy with pathological lesion | genitourinary | DR | 2.69 | 2.35 | 3.08 | 1.32E-13 | 15621 |
| 706 | | | DR | 1.32 | 1.27 | 1.37 | 1.41E-13 | 15621 |
| 426.23 | Second degree AV block | circulatory system | DR | 3.18 | 2.71 | 3.71 | 1.62E-13 | 15621 |
| 440.9 | Atherosclerosis of aorta | circulatory system | DR | 2.17 | 1.96 | 2.41 | 1.76E-13 | 15621 |
| 38.2 | Gram positive septicemia | infectious diseases | DR | 3.06 | 2.63 | 3.56 | 1.77E-13 | 15621 |
| 368.2 | Diplopia and disorders of binocular vision | sense organs | DR | 1.71 | 1.59 | 1.84 | 2.08E-13 | 15621 |
| 386.9 | Dizziness and giddiness (Light-headedness and vertigo) | sense organs | DR | 1.30 | 1.26 | 1.35 | 2.43E-13 | 15621 |
| 578.9 | Hemorrhage of gastrointestinal tract | digestive | DR | 1.50 | 1.42 | 1.59 | 2.51E-13 | 15621 |
| 729 | Other disorders of soft tissues | musculoskeletal | DR | 1.60 | 1.50 | 1.71 | 5.56E-13 | 15621 |
| 454.11 | Varicose veins of lower extremity, symptomtic | circulatory system | DR | 1.73 | 1.61 | 1.87 | 6.69E-13 | 15621 |
| 743.9 | Osteopenia or other disorder of bone and cartilage | musculoskeletal | DR | 1.63 | 1.52 | 1.74 | 6.72E-13 | 15621 |
| 443.7 | Peripheral angiopathy in diseases classified elsewhere | circulatory system | DR | 6.21 | 4.81 | 8.00 | 6.74E-13 | 15621 |
| 281.9 | Deficiency anemias | hematopoietic | DR | 2.36 | 2.09 | 2.66 | 6.77E-13 | 15621 |
| 783 | Fever of unknown origin | symptoms | DR | 1.41 | 1.34 | 1.48 | 6.95E-13 | 15621 |
| 797 | Shock | symptoms | DR | 2.55 | 2.23 | 2.90 | 7.39E-13 | 15621 |
| 741 | Symptoms and disorders of the joints | musculoskeletal | DR | 1.32 | 1.27 | 1.37 | 9.20E-13 | 15621 |
| 513 | Respiratory abnormalities | respiratory | DR | 1.49 | 1.41 | 1.57 | 1.10E-12 | 15621 |
| 459.7 | Blood vessel replaced | circulatory system | DR | 2.78 | 2.40 | 3.21 | 1.37E-12 | 15621 |
| 523.31 | Acute periodontitis | digestive | DR | 1.34 | 1.29 | 1.40 | 1.54E-12 | 15621 |
| 8.52 | Intestinal infection due to C. difficile | infectious diseases | DR | 2.12 | 1.90 | 2.36 | 1.86E-12 | 15621 |
| 875 | | | DR | 2.24 | 2.00 | 2.51 | 2.37E-12 | 15621 |
| 452.2 | Deep vein thrombosis [DVT] | circulatory system | DR | 1.71 | 1.58 | 1.85 | 2.76E-12 | 15621 |
| 110.12 | Althete's foot | infectious diseases | DR | 1.32 | 1.27 | 1.38 | 3.96E-12 | 15621 |
| 287.3 | Thrombocytopenia | hematopoietic | DR | 1.49 | 1.40 | 1.57 | 4.13E-12 | 15621 |
| 427.8 | Sinoatrial node dysfunction (Bradycardia) | circulatory system | DR | 2.28 | 2.03 | 2.57 | 4.49E-12 | 15621 |
| 300 | Anxiety disorders | mental disorders | DR | 0.79 | 0.76 | 0.81 | 4.90E-12 | 15621 |
| 286 | | | DR | 1.64 | 1.53 | 1.77 | 4.98E-12 | 15621 |
| 420.3 | Endocarditis | circulatory system | DR | 2.02 | 1.82 | 2.23 | 6.05E-12 | 15621 |
| 523.3 | Periodontitis (acute or chronic) | digestive | DR | 1.30 | 1.25 | 1.35 | 6.28E-12 | 15621 |
| 454.1 | Varicose veins of lower extremity | circulatory system | DR | 1.54 | 1.45 | 1.64 | 7.36E-12 | 15621 |
| 727 | Other disorders of synovium, tendon, and bursa | musculoskeletal | DR | 1.29 | 1.24 | 1.34 | 7.79E-12 | 15621 |
| 433.12 | Cerebral atherosclerosis | circulatory system | DR | 2.63 | 2.28 | 3.03 | 8.04E-12 | 15621 |
| 800 | Fracture of lower limb | injuries & poisonings | DR | 1.69 | 1.57 | 1.83 | 1.06E-11 | 15621 |
| 850 | Hemorrhage or hematoma complicating a procedure | injuries & poisonings | DR | 1.88 | 1.71 | 2.06 | 1.39E-11 | 15621 |
| 288 | Diseases of white blood cells | hematopoietic | DR | 1.41 | 1.34 | 1.48 | 1.50E-11 | 15621 |
| 374.3 | Ptosis of eyelid | sense organs | DR | 1.62 | 1.51 | 1.74 | 1.50E-11 | 15621 |
| 426.32 | Left bundle branch block | circulatory system | DR | 1.99 | 1.80 | 2.20 | 1.56E-11 | 15621 |
| 521.1 | Dental caries | digestive | DR | 1.26 | 1.21 | 1.30 | 1.72E-11 | 15621 |
| 521 | Diseases of hard tissues of teeth | digestive | DR | 1.26 | 1.21 | 1.30 | 1.85E-11 | 15621 |
| 429.9 | Cardiac complications, not elsewhere classified | circulatory system | DR | 2.52 | 2.20 | 2.90 | 2.16E-11 | 15621 |
| 313.1 | Attention deficit hyperactivity disorder | mental disorders | DR | 0.34 | 0.29 | 0.40 | 2.21E-11 | 15621 |
| 429.3 | Symptoms involving cardiovascular system | circulatory system | DR | 1.70 | 1.57 | 1.84 | 2.26E-11 | 15621 |
| 275.51 | Hypocalcemia | endocrine/metabolic | DR | 2.31 | 2.04 | 2.62 | 2.59E-11 | 15621 |
| 317 | Alcohol-related disorders | mental disorders | DR | 0.78 | 0.75 | 0.81 | 2.93E-11 | 15621 |
| 362.27 | Drusen (degenerative) of retina | sense organs | DR | 1.69 | 1.56 | 1.83 | 3.28E-11 | 15621 |
| 740 | | | DR | 1.26 | 1.22 | 1.30 | 3.53E-11 | 15621 |
| 426.31 | Right bundle branch block | circulatory system | DR | 1.74 | 1.60 | 1.90 | 5.01E-11 | 15621 |
| 807 | Fracture of ribs | injuries & poisonings | DR | 1.68 | 1.55 | 1.82 | 5.16E-11 | 15621 |
| 501 | Pneumonitis due to inhalation of food or vomitus | respiratory | DR | 2.21 | 1.96 | 2.49 | 5.24E-11 | 15621 |
| 994.21 | Septic shock | injuries & poisonings | DR | 2.06 | 1.84 | 2.30 | 5.36E-11 | 15621 |
| 250.2 | Type 2 diabetes | endocrine/metabolic | DR | 46.43 | 25.84 | 83.45 | 5.87E-11 | 15621 |
| 270.32 | Paraproteinemia | endocrine/metabolic | DR | 2.48 | 2.16 | 2.85 | 6.09E-11 | 15621 |
| 427.1 | Paroxysmal tachycardia, unspecified | circulatory system | DR | 1.62 | 1.51 | 1.75 | 6.34E-11 | 15621 |
| 260.3 | Adult failure to thrive | endocrine/metabolic | DR | 2.26 | 1.99 | 2.56 | 7.56E-11 | 15621 |
| 870 | Open wounds of head; neck; and trunk | injuries & poisonings | DR | 1.41 | 1.34 | 1.49 | 7.87E-11 | 15621 |
| 427.12 | Paroxysmal ventricular tachycardia | circulatory system | DR | 1.89 | 1.71 | 2.08 | 9.27E-11 | 15621 |
| 728.71 | Contracture of palmar fascia [Dupuytren's disease] | musculoskeletal | DR | 2.22 | 1.96 | 2.51 | 9.29E-11 | 15621 |
| 497 | Bronchitis | respiratory | DR | 1.32 | 1.27 | 1.38 | 9.45E-11 | 15621 |
| 574 | | | DR | 1.40 | 1.33 | 1.47 | 1.01E-10 | 15621 |
| 250.15 | Diabetes type 1 with peripheral circulatory disorders | endocrine/metabolic | DR | 5.72 | 4.36 | 7.51 | 1.38E-10 | 15621 |
| 429.2 | Abnormal function study of cardiovascular system | circulatory system | DR | 1.62 | 1.51 | 1.75 | 1.47E-10 | 15621 |
| 781 | Symptoms involving nervous and musculoskeletal systems | symptoms | DR | 1.53 | 1.43 | 1.64 | 1.52E-10 | 15621 |
| 747 | Cardiac and circulatory congenital anomalies | congenital anomalies | DR | 1.85 | 1.68 | 2.04 | 1.59E-10 | 15621 |
| 286.9 | Abnormal coagulation profile | hematopoietic | DR | 1.95 | 1.76 | 2.17 | 1.80E-10 | 15621 |
| 604.1 | Redundant prepuce and phimosis/BXO | genitourinary | DR | 1.55 | 1.45 | 1.66 | 1.98E-10 | 15621 |
| 681.3 | Cellulitis and abscess of arm/hand | dermatologic | DR | 1.60 | 1.48 | 1.72 | 2.34E-10 | 15621 |

| | | | | | | | | |
|---|---|---|---|---|---|---|---|---|
| 342 | Hemiplegia | neurological | DR | 2.02 | 1.81 | 2.26 | 2.36E-10 | 15621 |
| 453 | Chronic venous hypertension | circulatory system | DR | 2.47 | 2.14 | 2.86 | 3.51E-10 | 15621 |
| 480.11 | Pneumococcal pneumonia | respiratory | DR | 2.24 | 1.97 | 2.54 | 3.94E-10 | 15621 |
| 296.1 | Bipolar | mental disorders | DR | 0.68 | 0.64 | 0.72 | 4.03E-10 | 15621 |
| 317.1 | Alcoholism | mental disorders | DR | 0.78 | 0.75 | 0.81 | 4.21E-10 | 15621 |
| 797.1 | Cardiogenic shock | symptoms | DR | 3.38 | 2.78 | 4.11 | 4.33E-10 | 15621 |
| 288.2 | Elevated white blood cell count | hematopoietic | DR | 1.41 | 1.33 | 1.49 | 6.23E-10 | 15621 |
| 599.4 | Urinary incontinence | genitourinary | DR | 1.37 | 1.30 | 1.44 | 7.80E-10 | 15621 |
| 772.6 | Facial weakness | symptoms | DR | 2.18 | 1.92 | 2.47 | 7.84E-10 | 15621 |
| 512.9 | Other dyspnea | respiratory | DR | 1.24 | 1.20 | 1.29 | 8.94E-10 | 15621 |
| 363 | Chorioretinal inflammations, scars, and other disorders of choroid | sense organs | DR | 1.58 | 1.46 | 1.70 | 9.71E-10 | 15621 |
| 801 | Fracture of ankle and foot | injuries & poisonings | DR | 1.46 | 1.38 | 1.56 | 9.84E-10 | 15621 |
| 8 | Intestinal infection | infectious diseases | DR | 1.43 | 1.35 | 1.51 | 9.90E-10 | 15621 |
| 274 | | | DR | 1.35 | 1.28 | 1.42 | 1.00E-09 | 15621 |
| 743.2 | Pathologic fracture | musculoskeletal | DR | 1.93 | 1.73 | 2.16 | 1.45E-09 | 15621 |
| 377 | Disorders of optic nerve and visual pathways | sense organs | DR | 1.52 | 1.41 | 1.62 | 1.65E-09 | 15621 |
| 817 | Concussion | injuries & poisonings | DR | 0.55 | 0.50 | 0.61 | 1.99E-09 | 15621 |
| 773 | Pain in limb | symptoms | DR | 1.23 | 1.18 | 1.27 | 2.05E-09 | 15621 |
| 374.1 | Ectropion or entropion | sense organs | DR | 1.89 | 1.70 | 2.11 | 2.23E-09 | 15621 |
| 859 | Complication due to other implant and internal device | injuries & poisonings | DR | 2.00 | 1.78 | 2.25 | 2.52E-09 | 15621 |
| 244 | | | DR | 1.32 | 1.26 | 1.38 | 2.64E-09 | 15621 |
| 572 | Ascites (non malignant) | digestive | DR | 1.72 | 1.57 | 1.89 | 2.70E-09 | 15621 |
| 244.4 | Hypothyroidism NOS | endocrine/metabolic | DR | 1.32 | 1.26 | 1.38 | 2.77E-09 | 15621 |
| 297.1 | Suicidal ideation | mental disorders | DR | 0.69 | 0.65 | 0.74 | 3.04E-09 | 15621 |
| 702.1 | Actinic keratosis | dermatologic | DR | 1.44 | 1.36 | 1.54 | 3.26E-09 | 15621 |
| 250 | Diabetes mellitus | endocrine/metabolic | DR | 68.18 | 33.38 | 139.27 | 3.39E-09 | 15621 |
| 300.9 | Posttraumatic stress disorder | mental disorders | DR | 0.82 | 0.79 | 0.85 | 4.05E-09 | 15621 |
| 803 | Fracture of upper limb | injuries & poisonings | DR | 1.58 | 1.46 | 1.71 | 4.16E-09 | 15621 |
| 53 | Herpes zoster | infectious diseases | DR | 1.43 | 1.34 | 1.52 | 4.35E-09 | 15621 |
| 440.1 | Atherosclerosis of renal artery | circulatory system | DR | 3.49 | 2.82 | 4.32 | 4.49E-09 | 15621 |
| 523.32 | Chronic periodontitis | digestive | DR | 1.29 | 1.24 | 1.35 | 4.82E-09 | 15621 |
| 352 | Disorders of other cranial nerves | neurological | DR | 1.48 | 1.38 | 1.58 | 5.06E-09 | 15621 |
| 250.42 | Other abnormal glucose | endocrine/metabolic | DR | 0.80 | 0.77 | 0.83 | 5.31E-09 | 15621 |
| 261 | Vitamin deficiency | endocrine/metabolic | DR | 1.22 | 1.18 | 1.26 | 5.38E-09 | 15621 |
| 523 | Gingival and periodontal diseases | digestive | DR | 1.22 | 1.18 | 1.26 | 6.36E-09 | 15621 |
| 702 | Degenerative skin conditions and other dermatoses | dermatologic | DR | 1.27 | 1.22 | 1.32 | 6.63E-09 | 15621 |
| 260.2 | severe protein-calorie malnutrition | endocrine/metabolic | DR | 2.39 | 2.05 | 2.77 | 7.22E-09 | 15621 |
| 681.7 | Cellulitis and abscess of trunk | dermatologic | DR | 1.46 | 1.37 | 1.56 | 7.62E-09 | 15621 |
| 447.1 | Stricture of artery | circulatory system | DR | 2.52 | 2.15 | 2.96 | 8.06E-09 | 15621 |
| 796 | Elevated prostate specific antigen [PSA] | genitourinary | DR | 1.31 | 1.25 | 1.37 | 8.40E-09 | 15621 |
| 394.7 | Disease of tricuspid valve | circulatory system | DR | 2.11 | 1.85 | 2.40 | 8.83E-09 | 15621 |
| 442 | Other aneurysm | circulatory system | DR | 1.55 | 1.44 | 1.68 | 1.04E-08 | 15621 |
| 367.1 | Myopia | sense organs | DR | 1.22 | 1.18 | 1.26 | 1.10E-08 | 15621 |
| 420.2 | Pericarditis | circulatory system | DR | 1.90 | 1.70 | 2.13 | 1.10E-08 | 15621 |
| 590 | Pyelonephritis | genitourinary | DR | 1.75 | 1.58 | 1.92 | 1.35E-08 | 15621 |
| 562.1 | Diverticulosis | digestive | DR | 1.24 | 1.20 | 1.29 | 1.46E-08 | 15621 |
| 577 | Diseases of pancreas | digestive | DR | 1.44 | 1.35 | 1.54 | 1.73E-08 | 15621 |
| 426.25 | Other heart block | circulatory system | DR | 2.44 | 2.08 | 2.85 | 1.81E-08 | 15621 |
| 224.1 | Benign neoplasm of eye, uveal | neoplasms | DR | 1.57 | 1.45 | 1.70 | 1.86E-08 | 15621 |
| 483 | Acute bronchitis and bronchiolitis | respiratory | DR | 1.24 | 1.19 | 1.29 | 2.14E-08 | 15621 |
| 609.1 | Infertility, male | genitourinary | DR | 0.32 | 0.26 | 0.39 | 2.18E-08 | 15621 |
| 1005 | Other symptoms | NULL | DR | 1.21 | 1.17 | 1.25 | 2.31E-08 | 15621 |
| 250.21 | Type 2 diabetes with ketoacidosis | endocrine/metabolic | DR | 1.77 | 1.59 | 1.96 | 3.14E-08 | 15621 |
| 389 | Hearing loss | sense organs | DR | 1.21 | 1.17 | 1.26 | 3.38E-08 | 15621 |
| 510 | Other diseases of lung | respiratory | DR | 1.39 | 1.31 | 1.47 | 3.55E-08 | 15621 |
| 457.2 | | | DR | 2.37 | 2.02 | 2.77 | 3.96E-08 | 15621 |
| 596 | Other disorders of bladder | genitourinary | DR | 1.33 | 1.26 | 1.40 | 4.73E-08 | 15621 |
| 363.3 | Chorioretinal scars | sense organs | DR | 1.54 | 1.42 | 1.67 | 4.77E-08 | 15621 |
| 378 | Strabismus and other disorders of binocular eye movements | sense organs | DR | 1.43 | 1.34 | 1.52 | 5.50E-08 | 15621 |
| 686.1 | Carbuncle and furuncle | dermatologic | DR | 1.47 | 1.37 | 1.58 | 5.71E-08 | 15621 |
| 579.8 | Nonspecific abnormal findings in stool contents | digestive | DR | 1.41 | 1.32 | 1.50 | 6.00E-08 | 15621 |
| 8.5 | Bacterial enteritis | infectious diseases | DR | 1.60 | 1.46 | 1.74 | 6.64E-08 | 15621 |
| 81 | Infection/inflammation of internal prosthetic device; implant; and graft | infectious diseases | DR | 1.90 | 1.68 | 2.14 | 8.27E-08 | 15621 |
| 172 | | | DR | 1.46 | 1.36 | 1.56 | 8.31E-08 | 15621 |
| 402 | Elevated blood pressure reading without diagnosis of hypertension | circulatory system | DR | 0.77 | 0.73 | 0.81 | 8.81E-08 | 15621 |
| 519 | Other diseases of respiratory system, not elsewhere classified | respiratory | DR | 1.34 | 1.27 | 1.41 | 9.57E-08 | 15621 |
| 599.6 | Oliguria and anuria | genitourinary | DR | 3.03 | 2.46 | 3.74 | 1.03E-07 | 15621 |
| 805 | Fracture of vertebral column without mention of spinal cord injury | injuries & poisonings | DR | 1.77 | 1.59 | 1.97 | 1.03E-07 | 15621 |
| 1010.6 | Persons encountering health services in circumstances related to reproduction | | DR | 0.40 | 0.34 | 0.48 | 1.07E-07 | 15621 |
| 604 | Disorders of penis | genitourinary | DR | 1.36 | 1.28 | 1.44 | 1.12E-07 | 15621 |
| 474 | Acute and chronic tonsillitis | respiratory | DR | 0.58 | 0.52 | 0.64 | 1.13E-07 | 15621 |
| 300.11 | Generalized anxiety disorder | mental disorders | DR | 0.76 | 0.72 | 0.80 | 1.13E-07 | 15621 |
| 513.3 | Hypoventilation | respiratory | DR | 1.57 | 1.44 | 1.71 | 1.19E-07 | 15621 |
| 364 | Corneal opacity and other disorders of cornea | sense organs | DR | 1.48 | 1.38 | 1.60 | 1.21E-07 | 15621 |
| 513.32 | Orthopnea | respiratory | DR | 2.27 | 1.94 | 2.65 | 1.22E-07 | 15621 |
| 349 | Other and unspecified disorders of the nervous system | neurological | DR | 1.71 | 1.55 | 1.90 | 1.22E-07 | 15621 |
| 331.9 | Cerebral degeneration, unspecified | neurological | DR | 2.15 | 1.86 | 2.48 | 1.25E-07 | 15621 |
| 290.11 | Alzheimer's disease | mental disorders | DR | 1.94 | 1.71 | 2.20 | 1.26E-07 | 15621 |
| 292.2 | Mild cognitive impairment | mental disorders | DR | 1.37 | 1.29 | 1.46 | 1.29E-07 | 15621 |
| 565.1 | Anal and rectal polyp | digestive | DR | 1.48 | 1.37 | 1.59 | 1.37E-07 | 15621 |
| 771.2 | | | DR | 1.54 | 1.42 | 1.67 | 1.38E-07 | 15621 |
| 712 | Infective connective tissue disorders | musculoskeletal | DR | 2.91 | 2.38 | 3.58 | 1.71E-07 | 15621 |
| 41.9 | Infection with drug-resistant microorganisms | infectious diseases | DR | 2.03 | 1.77 | 2.32 | 1.71E-07 | 15621 |
| 790 | Nonspecific findings on examination of blood | symptoms | DR | 1.80 | 1.61 | 2.02 | 2.22E-07 | 15621 |
| 224 | Benign neoplasm of eye | neoplasms | DR | 1.46 | 1.36 | 1.58 | 2.31E-07 | 15621 |
| 316 | Substance addiction and disorders | mental disorders | DR | 0.80 | 0.77 | 0.84 | 2.31E-07 | 15621 |
| 454 | Varicose veins | circulatory system | DR | 1.34 | 1.26 | 1.41 | 2.59E-07 | 15621 |
| 339 | Other headache syndromes | neurological | DR | 0.83 | 0.80 | 0.86 | 3.18E-07 | 15621 |
| 772.1 | Muscular wasting and disuse atrophy | symptoms | DR | 2.38 | 2.01 | 2.82 | 3.42E-07 | 15621 |
| 429.1 | Heart transplant/surgery | circulatory system | DR | 1.90 | 1.67 | 2.15 | 3.86E-07 | 15621 |
| 377.1 | Optic atrophy | sense organs | DR | 1.58 | 1.45 | 1.74 | 3.95E-07 | 15621 |
| 835 | Internal derangement of knee | injuries & poisonings | DR | 0.78 | 0.74 | 0.82 | 4.11E-07 | 15621 |
| 531 | Peptic ulcer (excl. esophageal) | digestive | DR | 1.36 | 1.28 | 1.45 | 4.19E-07 | 15621 |

| | | | | | | | | |
|---|---|---|---|---|---|---|---|---|
| 962.2 | | | DR | 3.38 | 2.65 | 4.29 | 4.32E-07 | 15621 |
| 687 | Symptoms affecting skin | dermatologic | DR | 1.29 | 1.23 | 1.36 | 4.45E-07 | 15621 |
| 593 | Hematuria | genitourinary | DR | 1.24 | 1.19 | 1.29 | 5.23E-07 | 15621 |
| 473.4 | Voice disturbance | respiratory | DR | 1.43 | 1.33 | 1.53 | 5.40E-07 | 15621 |
| 262 | Mineral deficiency NEC | endocrine/metabolic | DR | 1.69 | 1.52 | 1.88 | 5.47E-07 | 15621 |
| 509.2 | Respiratory insufficiency | respiratory | DR | 1.74 | 1.56 | 1.94 | 6.28E-07 | 15621 |
| 726.1 | Enthesopathy | musculoskeletal | DR | 1.21 | 1.16 | 1.26 | 6.70E-07 | 15621 |
| 962 | Poisoning by hormones and synthetic substitutes | injuries & poisonings | DR | 1.74 | 1.56 | 1.95 | 7.14E-07 | 15621 |
| 371.1 | Uveitis, noninfectious or NOS | sense organs | DR | 1.60 | 1.45 | 1.76 | 7.57E-07 | 15621 |
| 574.1 | Cholelithiasis | digestive | DR | 1.31 | 1.24 | 1.39 | 7.87E-07 | 15621 |
| 276.42 | Alkalosis | endocrine/metabolic | DR | 1.89 | 1.66 | 2.15 | 8.05E-07 | 15621 |
| 274.1 | Gout | endocrine/metabolic | DR | 1.29 | 1.22 | 1.35 | 8.58E-07 | 15621 |
| 522 | Diseases of pulp and periapical tissues | digestive | DR | 1.25 | 1.19 | 1.31 | 8.90E-07 | 15621 |
| 327 | Sleep disorders | neurological | DR | 0.85 | 0.82 | 0.88 | 1.02E-06 | 15621 |
| 609 | | | DR | 0.52 | 0.46 | 0.60 | 1.04E-06 | 15621 |
| 270.38 | Other specified disorders of plasma protein metabolism | endocrine/metabolic | DR | 2.29 | 1.93 | 2.71 | 1.04E-06 | 15621 |
| 495 | Asthma | respiratory | DR | 0.79 | 0.75 | 0.83 | 1.04E-06 | 15621 |
| 689 | Disorder of skin and subcutaneous tissue NOS | dermatologic | DR | 1.24 | 1.19 | 1.30 | 1.06E-06 | 15621 |
| 960 | Poisoning by antibiotics | injuries & poisonings | DR | 1.67 | 1.50 | 1.85 | 1.07E-06 | 15621 |
| 592.1 | Cystitis | genitourinary | DR | 1.47 | 1.36 | 1.59 | 1.11E-06 | 15621 |
| 747.2 | Congenital anomalies of peripheral vascular system | congenital anomalies | DR | 2.12 | 1.82 | 2.48 | 1.11E-06 | 15621 |
| 379.4 | Anomalies of pupillary function | sense organs | DR | 1.87 | 1.64 | 2.13 | 1.12E-06 | 15621 |
| 414.2 | | | DR | 2.06 | 1.77 | 2.39 | 1.12E-06 | 15621 |
| 857 | Mechanical complication of unspecified genitourinary device, implant, and graft | injuries & poisonings | DR | 1.72 | 1.54 | 1.92 | 1.25E-06 | 15621 |
| 276.14 | Hypopotassemia | endocrine/metabolic | DR | 1.26 | 1.20 | 1.33 | 1.27E-06 | 15621 |
| 470 | Septal Deviations/Turbinate Hypertrophy | respiratory | DR | 0.71 | 0.66 | 0.76 | 1.28E-06 | 15621 |
| 284 | Aplastic anemia | hematopoietic | DR | 1.63 | 1.47 | 1.80 | 1.28E-06 | 15621 |
| 1010 | Other tests | | DR | 2.11 | 1.81 | 2.46 | 1.29E-06 | 15621 |
| 972 | Poisoning by agents primarily affecting the cardiovascular system | injuries & poisonings | DR | 1.81 | 1.60 | 2.05 | 1.37E-06 | 15621 |
| 153 | Colorectal cancer | neoplasms | DR | 1.54 | 1.41 | 1.68 | 1.42E-06 | 15621 |
| 350.3 | Lack of coordination | neurological | DR | 1.57 | 1.43 | 1.72 | 1.50E-06 | 15621 |
| 331 | Other cerebral degenerations | neurological | DR | 1.75 | 1.56 | 1.97 | 1.54E-06 | 15621 |
| 281.1 | Megaloblastic anemia | hematopoietic | DR | 1.38 | 1.29 | 1.48 | 1.56E-06 | 15621 |
| 172.2 | Other non-epithelial cancer of skin | neoplasms | DR | 1.44 | 1.33 | 1.55 | 1.68E-06 | 15621 |
| 599.9 | Other abnormality of urination | genitourinary | DR | 1.30 | 1.23 | 1.37 | 1.80E-06 | 15621 |
| 274.21 | Chondrocalcinosis | endocrine/metabolic | DR | 1.85 | 1.62 | 2.10 | 1.83E-06 | 15621 |
| 580.1 | | | DR | 2.17 | 1.85 | 2.56 | 1.89E-06 | 15621 |
| 313 | Pervasive developmental disorders | mental disorders | DR | 0.56 | 0.49 | 0.63 | 2.14E-06 | 15621 |
| 274.2 | Crystal arthropathies | endocrine/metabolic | DR | 1.71 | 1.53 | 1.92 | 2.31E-06 | 15621 |
| 562 | | | DR | 1.19 | 1.15 | 1.24 | 2.35E-06 | 15621 |
| 574.3 | Cholecystitis without cholelithiasis | digestive | DR | 1.46 | 1.35 | 1.58 | 2.49E-06 | 15621 |
| 300.12 | Agorophobia, social phobia, and panic disorder | mental disorders | DR | 0.72 | 0.68 | 0.78 | 2.58E-06 | 15621 |
| 601.4 | Balanoposthitis | genitourinary | DR | 1.37 | 1.28 | 1.47 | 2.61E-06 | 15621 |
| 512.2 | Painful respiration | respiratory | DR | 1.29 | 1.22 | 1.36 | 2.72E-06 | 15621 |
| 153.2 | Colon cancer | neoplasms | DR | 1.55 | 1.41 | 1.70 | 2.76E-06 | 15621 |
| 284.1 | Pancytopenia | hematopoietic | DR | 1.65 | 1.48 | 1.83 | 2.81E-06 | 15621 |
| 263 | Other nutritional deficiency | endocrine/metabolic | DR | 1.81 | 1.59 | 2.06 | 3.00E-06 | 15621 |
| 599.5 | Frequency of urination and polyuria | genitourinary | DR | 1.22 | 1.17 | 1.28 | 3.32E-06 | 15621 |
| 601 | | | DR | 1.23 | 1.17 | 1.28 | 3.65E-06 | 15621 |
| 979 | Adverse drug events and drug allergies | injuries & poisonings | DR | 1.30 | 1.23 | 1.38 | 3.74E-06 | 15621 |
| 536 | Disorders of function of stomach | digestive | DR | 1.28 | 1.21 | 1.35 | 3.79E-06 | 15621 |
| 112 | Candidiasis | infectious diseases | DR | 1.34 | 1.26 | 1.43 | 3.79E-06 | 15621 |
| 297 | Suicidal ideation or attempt | mental disorders | DR | 0.78 | 0.74 | 0.82 | 3.80E-06 | 15621 |
| 394.1 | Mitral valve stenosis and aortic valve stenosis | circulatory system | DR | 2.10 | 1.78 | 2.47 | 4.59E-06 | 15621 |
| 514.2 | Solitary pulmonary nodule | respiratory | DR | 1.32 | 1.24 | 1.40 | 7.87E-06 | 15621 |
| 601.1 | Prostatitis | genitourinary | DR | 1.33 | 1.25 | 1.42 | 8.14E-06 | 15621 |
| 379.9 | Pain, swelling or discharge of eye | sense organs | DR | 1.33 | 1.25 | 1.42 | 9.06E-06 | 15621 |
| 964 | Poisoning by agents primarily affecting blood constituents | injuries & poisonings | DR | 2.16 | 1.82 | 2.58 | 9.33E-06 | 15621 |
| 427.6 | Premature beats | circulatory system | DR | 1.43 | 1.32 | 1.56 | 1.00E-05 | 15621 |
| 711 | Arthropathy associated with infections | musculoskeletal | DR | 1.61 | 1.45 | 1.79 | 1.03E-05 | 15621 |
| 297.2 | Suicide or self-inflicted injury | mental disorders | DR | 0.64 | 0.58 | 0.71 | 1.08E-05 | 15621 |
| 364.2 | Corneal edema | sense organs | DR | 2.01 | 1.71 | 2.35 | 1.14E-05 | 15621 |
| 474.2 | Chronic tonsillitis and adenoiditis | respiratory | DR | 0.50 | 0.43 | 0.59 | 1.24E-05 | 15621 |
| 564.1 | Irritable Bowel Syndrome | digestive | DR | 0.69 | 0.63 | 0.75 | 1.25E-05 | 15621 |
| 702.2 | Seborrheic keratosis | dermatologic | DR | 1.21 | 1.15 | 1.26 | 1.26E-05 | 15621 |
| 441 | Vascular insufficiency of intestine | circulatory system | DR | 2.22 | 1.85 | 2.66 | 1.35E-05 | 15621 |
| 701 | Other hypertrophic and atrophic conditions of skin | dermatologic | DR | 1.19 | 1.14 | 1.24 | 1.38E-05 | 15621 |
| 785 | Abdominal pain | symptoms | DR | 1.16 | 1.12 | 1.20 | 1.68E-05 | 15621 |
| 136 | Other infectious and parasitic diseases | infectious diseases | DR | 1.38 | 1.28 | 1.49 | 1.97E-05 | 15621 |
| 281.12 | Other vitamin B12 deficiency anemia | hematopoietic | DR | 1.35 | 1.26 | 1.45 | 2.07E-05 | 15621 |
| 352.2 | Facial nerve disorders [CN7] | neurological | DR | 1.42 | 1.31 | 1.54 | 2.08E-05 | 15621 |
| 522.5 | Periapical abscess | digestive | DR | 1.30 | 1.22 | 1.38 | 2.30E-05 | 15621 |
| 415.21 | Primary pulmonary hypertension | circulatory system | DR | 1.88 | 1.62 | 2.18 | 2.33E-05 | 15621 |
| 876 | | | DR | 1.89 | 1.62 | 2.19 | 2.35E-05 | 15621 |
| 395.6 | Heart valve replaced | circulatory system | DR | 1.65 | 1.46 | 1.86 | 2.57E-05 | 15621 |
| 963 | Poisoning by primarily systemic agents | injuries & poisonings | DR | 1.91 | 1.64 | 2.23 | 2.59E-05 | 15621 |
| 275.6 | Hypercalcemia | endocrine/metabolic | DR | 1.56 | 1.40 | 1.73 | 2.63E-05 | 15621 |
| 473 | Diseases of the larynx and vocal cords | respiratory | DR | 1.31 | 1.23 | 1.40 | 2.68E-05 | 15621 |
| 386 | Vertiginous syndromes and other disorders of vestibular system | sense organs | DR | 1.25 | 1.19 | 1.32 | 2.80E-05 | 15621 |
| 578.2 | Blood in stool | digestive | DR | 1.21 | 1.16 | 1.27 | 2.83E-05 | 15621 |
| 301.2 | Antisocial/borderline personality disorder | mental disorders | DR | 0.64 | 0.57 | 0.71 | 2.83E-05 | 15621 |
| 560.1 | Paralytic ileus | digestive | DR | 1.62 | 1.44 | 1.81 | 2.88E-05 | 15621 |
| 189.11 | Malignant neoplasm of kidney, except pelvis | neoplasms | DR | 1.57 | 1.41 | 1.75 | 3.02E-05 | 15621 |
| 425.2 | Secondary/extrinsic cardiomyopathies | circulatory system | DR | 1.71 | 1.51 | 1.95 | 3.20E-05 | 15621 |
| 360 | Disorders of the globe | sense organs | DR | 1.82 | 1.58 | 2.11 | 3.41E-05 | 15621 |
| 537.1 | Lesions of stomach and duodenum | digestive | DR | 2.24 | 1.84 | 2.72 | 3.47E-05 | 15621 |
| 286.7 | Other and unspecified coagulation defects | hematopoietic | DR | 1.52 | 1.37 | 1.68 | 3.71E-05 | 15621 |
| 364.1 | Corneal opacity | sense organs | DR | 1.36 | 1.26 | 1.47 | 3.74E-05 | 15621 |
| 747.1 | Cardiac congenital anomalies | congenital anomalies | DR | 1.66 | 1.47 | 1.88 | 3.83E-05 | 15621 |
| 596.5 | Functional disorders of bladder | genitourinary | DR | 1.30 | 1.22 | 1.39 | 3.84E-05 | 15621 |
| 359.2 | Myopathy | neurological | DR | 1.86 | 1.60 | 2.16 | 4.17E-05 | 15621 |
| 698 | Pruritus and related conditions | dermatologic | DR | 1.25 | 1.18 | 1.32 | 4.33E-05 | 15621 |
| 560 | | | DR | 1.35 | 1.25 | 1.45 | 4.38E-05 | 15621 |
| 1019 | Other ill-defined and unknown causes of morbidity and mortality | NULL | DR | 1.18 | 1.14 | 1.23 | 4.38E-05 | 15621 |
| 272.13 | Mixed hyperlipidemia | endocrine/metabolic | DR | 1.15 | 1.11 | 1.19 | 4.49E-05 | 15621 |
| 340.1 | Migrain with aura | neurological | DR | 0.59 | 0.51 | 0.67 | 4.60E-05 | 15621 |

| | | | | | | | | |
|---|---|---|---|---|---|---|---|---|
| 596.1 | Bladder neck obstruction | genitourinary | DR | 1.50 | 1.35 | 1.65 | 4.76E-05 | 15621 |
| 304 | Adjustment reaction | mental disorders | DR | 0.86 | 0.83 | 0.89 | 4.78E-05 | 15621 |
| 695.9 | Unspecified erythematous condition | dermatologic | DR | 1.75 | 1.53 | 2.01 | 4.84E-05 | 15621 |
| 513.8 | Disorders of diaphragm | respiratory | DR | 1.73 | 1.51 | 1.99 | 4.84E-05 | 15621 |
| 296.22 | Major depressive disorder | mental disorders | DR | 0.87 | 0.84 | 0.90 | 4.86E-05 | 15621 |
| 577.1 | Acute pancreatitis | digestive | DR | 1.36 | 1.26 | 1.47 | 4.88E-05 | 15621 |
| 359 | | | DR | 1.78 | 1.54 | 2.05 | 4.92E-05 | 15621 |
| 450 | Noninfectious disorders of lymphatic channels | circulatory system | DR | 1.62 | 1.44 | 1.82 | 5.27E-05 | 15621 |
| 242 | Thyrotoxicosis with or without goiter | endocrine/metabolic | DR | 1.54 | 1.39 | 1.72 | 5.34E-05 | 15621 |
| 711.1 | Pyogenic arthritis | musculoskeletal | DR | 1.72 | 1.51 | 1.97 | 5.69E-05 | 15621 |
| 426.8 | Other cardiac conduction disorders | circulatory system | DR | 1.87 | 1.60 | 2.18 | 5.70E-05 | 15621 |
| 790.6 | Other abnormal blood chemistry | symptoms | DR | 1.17 | 1.12 | 1.21 | 5.74E-05 | 15621 |
| 301 | Personality disorders | mental disorders | DR | 0.76 | 0.71 | 0.82 | 5.80E-05 | 15621 |
| 327.4 | Insomnia | neurological | DR | 0.86 | 0.83 | 0.90 | 5.88E-05 | 15621 |
| 598 | Abnormal findings on examination of urine | genitourinary | DR | 1.34 | 1.24 | 1.44 | 6.34E-05 | 15621 |
| 531.1 | Hemorrhage from gastrointestinal ulcer | digestive | DR | 1.67 | 1.47 | 1.91 | 6.40E-05 | 15621 |
| 964.1 | Anticoagulants causing adverse effects | injuries & poisonings | DR | 2.06 | 1.72 | 2.47 | 6.43E-05 | 15621 |
| 295.1 | Schizophrenia | mental disorders | DR | 0.74 | 0.69 | 0.80 | 6.64E-05 | 15621 |
| 803.2 | Fracture of radius and ulna | injuries & poisonings | DR | 1.51 | 1.36 | 1.67 | 6.71E-05 | 15621 |
| 598.9 | Other nonspecific findings on examination of urine | genitourinary | DR | 1.34 | 1.25 | 1.45 | 6.82E-05 | 15621 |
| 701.6 | Acquired acanthosis nigricans | dermatologic | DR | 0.49 | 0.41 | 0.59 | 7.43E-05 | 15621 |
| 395.3 | Nonrheumatic tricuspid valve disorders | circulatory system | DR | 1.64 | 1.45 | 1.86 | 7.57E-05 | 15621 |
| 733 | Other disorders of bone and cartilage | musculoskeletal | DR | 1.28 | 1.20 | 1.36 | 8.00E-05 | 15621 |
| 535 | Gastritis and duodenitis | digestive | DR | 1.19 | 1.14 | 1.24 | 8.02E-05 | 15621 |
| 418.1 | Precordial pain | circulatory system | DR | 1.43 | 1.31 | 1.57 | 8.27E-05 | 15621 |
| 756 | Other congenital musculoskeletal anomalies | congenital anomalies | DR | 1.74 | 1.51 | 2.01 | 8.61E-05 | 15621 |
| 1001 | Foreign body injury | NULL | DR | 1.30 | 1.22 | 1.39 | 8.92E-05 | 15621 |
| 189.1 | Cancer of kidney and renal pelvis | neoplasms | DR | 1.52 | 1.36 | 1.69 | 9.01E-05 | 15621 |
| 521.2 | Dental abrasion, erosion and attrition | digestive | DR | 1.24 | 1.17 | 1.30 | 9.03E-05 | 15621 |
| 592.11 | Acute cystitis | genitourinary | DR | 1.53 | 1.37 | 1.70 | 9.10E-05 | 15621 |
| 910 | | | DR | 1.81 | 1.56 | 2.11 | 9.41E-05 | 15621 |
| 939 | Atopic/contact dermatitis due to other or unspecified | dermatologic | DR | 1.15 | 1.11 | 1.19 | 9.50E-05 | 15621 |
| 370.2 | Superficial keratitis | sense organs | DR | 1.58 | 1.41 | 1.78 | 9.54E-05 | 15621 |
| 523.1 | Gingivitis | digestive | DR | 1.18 | 1.13 | 1.23 | 9.99E-05 | 15621 |
| 716.9 | Arthropathy NOS | musculoskeletal | DR | 1.19 | 1.13 | 1.24 | 0.000102 | 15621 |
| 592 | | | DR | 1.32 | 1.23 | 1.42 | 0.000104 | 15621 |
| 496.21 | Obstructive chronic bronchitis | respiratory | DR | 1.31 | 1.22 | 1.41 | 0.000105 | 15621 |
| 71.1 | HIV infection, symptomatic | infectious diseases | DR | 0.49 | 0.41 | 0.59 | 0.000107 | 15621 |
| 274.11 | Gouty arthropathy | endocrine/metabolic | DR | 1.37 | 1.26 | 1.48 | 0.000111 | 15621 |
| 537 | Other disorders of stomach and duodenum | digestive | DR | 1.31 | 1.22 | 1.40 | 0.000111 | 15621 |
| 189 | Cancer of urinary organs (incl. kidney and bladder) | neoplasms | DR | 1.37 | 1.26 | 1.48 | 0.000117 | 15621 |
| 172.1 | Melanomas of skin, dx or hx | neoplasms | DR | 1.47 | 1.33 | 1.63 | 0.000125 | 15621 |
| 963.1 | Antineoplastic and immunosuppressive drugs causing adverse effects | injuries & poisonings | DR | 1.87 | 1.59 | 2.20 | 0.000127 | 15621 |
| 200 | Myeloproliferative disease | neoplasms | DR | 1.54 | 1.38 | 1.73 | 0.000132 | 15621 |
| 726 | Peripheral enthesopathies and allied syndromes | musculoskeletal | DR | 1.14 | 1.10 | 1.18 | 0.000142 | 15621 |
| 290.12 | Dementia with cerebral degenerations | mental disorders | DR | 2.03 | 1.69 | 2.45 | 0.000142 | 15621 |
| 531.4 | Peptic ulcer, site unspecified | digestive | DR | 1.35 | 1.25 | 1.46 | 0.000143 | 15621 |
| 480.2 | Viral pneumonia | respiratory | DR | 1.70 | 1.48 | 1.95 | 0.00015 | 15621 |
| 173 | Neoplasm of uncertain behavior of skin | neoplasms | DR | 1.21 | 1.15 | 1.27 | 0.000155 | 15621 |
| 727.2 | Bursitis disorders | musculoskeletal | DR | 1.33 | 1.23 | 1.43 | 0.000157 | 15621 |
| 71 | Human immunodeficiency virus [HIV] disease | infectious diseases | DR | 0.52 | 0.44 | 0.62 | 0.000163 | 15621 |
| 278.11 | Morbid obesity | endocrine/metabolic | DR | 0.86 | 0.83 | 0.90 | 0.00017 | 15621 |
| 8.6 | Viral Enteritis | infectious diseases | DR | 1.44 | 1.31 | 1.59 | 0.00017 | 15621 |
| 276.11 | Hyperosmolality and/or hypernatremia | endocrine/metabolic | DR | 1.65 | 1.44 | 1.89 | 0.000174 | 15621 |
| 536.7 | Complications of gastrostomy, colostomy and enterostomy | digestive | DR | 2.70 | 2.07 | 3.52 | 0.000177 | 15621 |
| 117 | Mycoses | infectious diseases | DR | 1.49 | 1.34 | 1.65 | 0.000179 | 15621 |
| 614.54 | Abscess or ulceration of vulva | genitourinary | DR | 6.95 | 4.14 | 11.67 | 0.00018 | 15621 |
| 427.41 | Ventricular fibrillation and flutter | circulatory system | DR | 2.13 | 1.74 | 2.60 | 0.000184 | 15621 |
| 535.6 | Duodenitis | digestive | DR | 1.49 | 1.34 | 1.66 | 0.000188 | 15621 |
| 442.1 | Aortic aneurysm | circulatory system | DR | 1.40 | 1.28 | 1.53 | 0.000194 | 15621 |
| 427.11 | Paroxysmal supraventricular tachycardia | circulatory system | DR | 1.48 | 1.33 | 1.64 | 0.000195 | 15621 |
| 972.6 | Antihypertensive agents causing adverse effects | injuries & poisonings | DR | 1.72 | 1.49 | 1.99 | 0.000202 | 15621 |
| 575 | Other biliary tract disease | digestive | DR | 1.25 | 1.18 | 1.33 | 0.000208 | 15621 |
| 368.7 | Disorders of accommodation | sense organs | DR | 0.26 | 0.18 | 0.38 | 0.000231 | 15621 |
| 1000 | Burns | NULL | DR | 1.39 | 1.27 | 1.52 | 0.000234 | 15621 |
| 706.1 | Acne | dermatologic | DR | 0.72 | 0.66 | 0.79 | 0.000251 | 15621 |
| 430.2 | Intracerebral hemorrhage | circulatory system | DR | 1.76 | 1.51 | 2.05 | 0.000254 | 15621 |
| 292.11 | Aphasia | mental disorders | DR | 1.62 | 1.42 | 1.85 | 0.000255 | 15621 |
| 803.1 | Fracture of humerus | injuries & poisonings | DR | 1.68 | 1.46 | 1.94 | 0.000265 | 15621 |
| 283 | Acquired hemolytic anemias | hematopoietic | DR | 2.31 | 1.84 | 2.91 | 0.00028 | 15621 |
| 261.4 | Vitamin D deficiency | endocrine/metabolic | DR | 1.13 | 1.10 | 1.17 | 0.00028 | 15621 |
| 296 | Mood disorders | mental disorders | DR | 0.88 | 0.85 | 0.91 | 0.000296 | 15621 |
| 296.2 | | | DR | 0.88 | 0.85 | 0.91 | 0.000318 | 15621 |
| 302.1 | Decreased libido | mental disorders | DR | 0.63 | 0.56 | 0.72 | 0.00034 | 15621 |
| 375.2 | Epiphora | sense organs | DR | 1.71 | 1.47 | 1.98 | 0.00035 | 15621 |
| 430 | Intracranial hemorrhage | circulatory system | DR | 1.45 | 1.31 | 1.61 | 0.000366 | 15621 |
| 571.8 | Liver abscess and sequelae of chronic liver disease | digestive | DR | 1.34 | 1.24 | 1.46 | 0.000372 | 15621 |
| 386.2 | Peripheral or central vertigo | sense organs | DR | 1.22 | 1.15 | 1.29 | 0.000387 | 15621 |
| 368.3 | Anisometropia | sense organs | DR | 1.40 | 1.27 | 1.54 | 0.000398 | 15621 |
| 800.2 | Fracture of unspecified part of femur | injuries & poisonings | DR | 1.90 | 1.59 | 2.28 | 0.000407 | 15621 |
| 575.7 | Other disorders of gallbladder | digestive | DR | 1.40 | 1.27 | 1.55 | 0.000501 | 15621 |
| 516 | Abnormal sputum | respiratory | DR | 1.37 | 1.25 | 1.50 | 0.000508 | 15621 |
| 626 | Disorders of menstruation and other abnormal bleeding from female genital tract | genitourinary | DR | 0.57 | 0.48 | 0.67 | 0.000511 | 15621 |
| 724.8 | Other symptoms referable to back | musculoskeletal | DR | 0.76 | 0.70 | 0.82 | 0.000551 | 15621 |
| 597.1 | Urethral stricture (not specified as infectious) | genitourinary | DR | 1.50 | 1.34 | 1.69 | 0.000562 | 15621 |
| 687.2 | Localized superficial swelling, mass, or lump | dermatologic | DR | 1.21 | 1.15 | 1.28 | 0.000566 | 15621 |
| 442.11 | Abdominal aortic aneurysm | circulatory system | DR | 1.41 | 1.28 | 1.56 | 0.000582 | 15621 |
| 800.4 | Fracture of patella | injuries & poisonings | DR | 1.81 | 1.52 | 2.16 | 0.000602 | 15621 |
| 295.2 | Paranoid disorders | mental disorders | DR | 0.60 | 0.52 | 0.70 | 0.000629 | 15621 |
| 155 | Cancer of liver and intrahepatic bile duct | neoplasms | DR | 1.44 | 1.30 | 1.61 | 0.000639 | 15621 |
| 738 | Other acquired musculoskeletal deformity | musculoskeletal | DR | 0.78 | 0.73 | 0.84 | 0.000644 | 15621 |
| 348.7 | Coma | neurological | DR | 3.27 | 2.31 | 4.63 | 0.000651 | 15621 |
| 333.1 | Essential tremor | neurological | DR | 1.34 | 1.23 | 1.47 | 0.000656 | 15621 |
| 112.3 | Candidiasis of skin and nails | infectious diseases | DR | 1.81 | 1.52 | 2.15 | 0.000657 | 15621 |
| 386.3 | Labyrinthitis | sense organs | DR | 1.69 | 1.45 | 1.97 | 0.000675 | 15621 |

| | | | | | | | | |
|---|---|---|---|---|---|---|---|---|
| 556.11 | Angiodysplasia of intestine (without mention of hemorrhage) | digestive | DR | 1.89 | 1.57 | 2.28 | 0.00071 | 15621 |
| 380.1 | Otitis externa | sense organs | DR | 1.19 | 1.13 | 1.26 | 0.000719 | 15621 |
| 377.3 | Optic neuritis/neuropathy | sense organs | DR | 1.44 | 1.29 | 1.61 | 0.000733 | 15621 |
| 735.3 | Hallux valgus (Bunion) | musculoskeletal | DR | 1.23 | 1.16 | 1.31 | 0.000738 | 15621 |
| 578 | | | DR | 1.14 | 1.09 | 1.18 | 0.000762 | 15621 |
| 375 | Disorders of lacrimal system | sense organs | DR | 1.57 | 1.37 | 1.80 | 0.000786 | 15621 |
| 496.2 | Chronic bronchitis | respiratory | DR | 1.23 | 1.16 | 1.31 | 0.000793 | 15621 |
| 374.2 | Lagophthalmos | sense organs | DR | 1.88 | 1.56 | 2.27 | 0.000798 | 15621 |
| 686.2 | Impetigo | dermatologic | DR | 1.83 | 1.53 | 2.19 | 0.000809 | 15621 |
| 474.1 | Acute tonsillitis | respiratory | DR | 0.65 | 0.58 | 0.74 | 0.000814 | 15621 |
| 516.1 | Hemoptysis | respiratory | DR | 1.39 | 1.26 | 1.54 | 0.000838 | 15621 |
| 800.3 | Fracture of tibia and fibula | injuries & poisonings | DR | 1.45 | 1.30 | 1.63 | 0.000859 | 15621 |
| 530.5 | Disorders of esophageal motility | digestive | DR | 1.73 | 1.47 | 2.05 | 0.000885 | 15621 |
| 53.1 | Herpes zoster with nervous system complications | infectious diseases | DR | 1.62 | 1.40 | 1.87 | 0.000912 | 15621 |
| 442.8 | Aneurysm of other specified artery | circulatory system | DR | 1.95 | 1.59 | 2.38 | 0.000915 | 15621 |
| 344 | Other paralytic syndromes | neurological | DR | 1.39 | 1.26 | 1.53 | 0.000919 | 15621 |
| 574.2 | Calculus of bile duct | digestive | DR | 1.43 | 1.29 | 1.60 | 0.000923 | 15621 |
| 960.1 | Adverse effects of antibacterials (not penicillins) | injuries & poisonings | DR | 1.87 | 1.55 | 2.26 | 0.00094 | 15621 |
| 455 | Hemorrhoids | circulatory system | DR | 1.12 | 1.08 | 1.16 | 0.000958 | 15621 |
| 599.8 | Other symptoms involving urinary system | genitourinary | DR | 1.24 | 1.16 | 1.32 | 0.001013 | 15621 |
| 333 | Extrapyramidal disease and abnormal movement disorders | neurological | DR | 1.23 | 1.16 | 1.32 | 0.001052 | 15621 |
| 204 | Leukemia | neoplasms | DR | 1.58 | 1.37 | 1.82 | 0.001089 | 15621 |
| 172.21 | Basal cell carcinoma | neoplasms | DR | 1.47 | 1.30 | 1.65 | 0.001091 | 15621 |
| 290.3 | Other persistent mental disorders due to conditions classified elsewhere | mental disorders | DR | 1.22 | 1.15 | 1.29 | 0.001099 | 15621 |
| 480.5 | Bronchopneumonia and lung abscess | respiratory | DR | 1.64 | 1.41 | 1.91 | 0.001128 | 15621 |
| 172.22 | Squamous cell carcinoma | neoplasms | DR | 1.61 | 1.39 | 1.86 | 0.001133 | 15621 |
| 626.1 | Irregular menstrual cycle/bleeding | genitourinary | DR | 0.57 | 0.48 | 0.68 | 0.001146 | 15621 |
| 765 | Cervical radiculitis | symptoms | DR | 0.84 | 0.79 | 0.89 | 0.001165 | 15621 |
| 756.3 | Congenital anomalies of muscle, tendon, fascia, and connective tissue | congenital anomalies | DR | 2.47 | 1.87 | 3.26 | 0.001169 | 15621 |
| 525.2 | Atrophy of edentulous alveolar ridge | digestive | DR | 1.29 | 1.19 | 1.39 | 0.001185 | 15621 |
| 565 | Anal and rectal conditions | digestive | DR | 1.18 | 1.12 | 1.24 | 0.00122 | 15621 |
| 257.1 | Testicular hypofunction | endocrine/metabolic | DR | 0.82 | 0.77 | 0.87 | 0.001273 | 15621 |
| 527 | Diseases of the salivary glands | digestive | DR | 1.22 | 1.15 | 1.30 | 0.00134 | 15621 |
| 522.1 | Pulpitis and necrosis of tooth pulp | digestive | DR | 1.18 | 1.12 | 1.25 | 0.001365 | 15621 |
| 809 | Fracture of unspecified bones | injuries & poisonings | DR | 1.30 | 1.20 | 1.41 | 0.001365 | 15621 |
| 446 | Polyarteritis nodosa and allied conditions | circulatory system | DR | 1.68 | 1.43 | 1.97 | 0.001383 | 15621 |
| 539 | Bariatric surgery | digestive | DR | 0.59 | 0.50 | 0.70 | 0.001409 | 15621 |
| 260.6 | Anorexia | endocrine/metabolic | DR | 1.49 | 1.31 | 1.68 | 0.001412 | 15621 |
| 1010.7 | Persons with potential health hazards related to socioeconomic, psychosocial, and other circumstances | | DR | 0.78 | 0.72 | 0.85 | 0.001487 | 15621 |
| 627 | Menopausal and postmenopausal disorders | genitourinary | DR | 1.70 | 1.44 | 2.01 | 0.001488 | 15621 |
| 476 | Allergic rhinitis | respiratory | DR | 0.90 | 0.86 | 0.93 | 0.001507 | 15621 |
| 204.4 | Multiple myeloma | neoplasms | DR | 2.10 | 1.66 | 2.65 | 0.001537 | 15621 |
| 597.2 | Urinary complications NEC | genitourinary | DR | 1.61 | 1.39 | 1.87 | 0.001545 | 15621 |
| 350.1 | Abnormal involuntary movements | neurological | DR | 1.20 | 1.13 | 1.27 | 0.001578 | 15621 |
| 312.3 | Impulse control disorder | mental disorders | DR | 0.70 | 0.63 | 0.79 | 0.001582 | 15621 |
| 198 | Secondary malignant neoplasm | neoplasms | DR | 1.30 | 1.20 | 1.42 | 0.001588 | 15621 |
| 363.4 | Choroidal degenerations | sense organs | DR | 2.76 | 2.00 | 3.81 | 0.001647 | 15621 |
| 717 | Polymyalgia Rheumatica | musculoskeletal | DR | 2.14 | 1.68 | 2.72 | 0.001729 | 15621 |
| 155.1 | Malignant neoplasm of liver, primary | neoplasms | DR | 1.42 | 1.27 | 1.58 | 0.001738 | 15621 |
| 556 | Ulceration of the lower GI tract | digestive | DR | 1.53 | 1.33 | 1.75 | 0.001794 | 15621 |
| 447.7 | Aortic ectasia | circulatory system | DR | 1.78 | 1.48 | 2.14 | 0.001796 | 15621 |
| 535.9 | Gastritis and duodenitis, NOS | digestive | DR | 1.19 | 1.13 | 1.26 | 0.001803 | 15621 |
| 994.1 | Systemic inflammatory response syndrome (SIRS) | injuries & poisonings | DR | 1.49 | 1.31 | 1.69 | 0.001803 | 15621 |
| 661 | Fetal distress and abnormal forces of labor | pregnancy complications | DR | 2.54 | 1.89 | 3.43 | 0.001806 | 15621 |
| 368.9 | Subjective visual disturbances | sense organs | DR | 1.19 | 1.13 | 1.26 | 0.001807 | 15621 |
| 345 | Epilepsy, recurrent seizures, convulsions | neurological | DR | 1.22 | 1.15 | 1.30 | 0.001824 | 15621 |
| 977 | | | DR | 2.29 | 1.75 | 2.98 | 0.001825 | 15621 |
| 556.1 | Ulceration of intestine | digestive | DR | 1.57 | 1.36 | 1.81 | 0.001831 | 15621 |
| 279.8 | Other specified disorders involving the immune mechanism | endocrine/metabolic | DR | 3.63 | 2.40 | 5.49 | 0.001849 | 15621 |
| 389.4 | Tinnitus | sense organs | DR | 0.90 | 0.86 | 0.93 | 0.001863 | 15621 |
| 524.3 | Anomalies of tooth position/malocclusion | digestive | DR | 1.27 | 1.18 | 1.37 | 0.001919 | 15621 |
| 722.6 | Degeneration of intervertebral disc | musculoskeletal | DR | 0.88 | 0.85 | 0.92 | 0.001923 | 15621 |
| 840.2 | | | DR | 1.23 | 1.15 | 1.32 | 0.00194 | 15621 |
| 256.4 | Polycystic ovaries | endocrine/metabolic | DR | 0.31 | 0.21 | 0.45 | 0.001943 | 15621 |
| 257 | Testicular dysfunction | endocrine/metabolic | DR | 0.84 | 0.79 | 0.89 | 0.001967 | 15621 |
| 781.2 | Abnormal posture | symptoms | DR | 0.37 | 0.27 | 0.51 | 0.002055 | 15621 |
| 459.1 | Hemorrhage NOS | circulatory system | DR | 1.68 | 1.42 | 1.98 | 0.002083 | 15621 |
| 427.61 | Supraventricular premature beats | circulatory system | DR | 1.48 | 1.30 | 1.68 | 0.0022 | 15621 |
| 803.21 | | | DR | 2.60 | 1.90 | 3.55 | 0.00223 | 15621 |
| 1014 | Effects of heat, cold and air pressure | NULL | DR | 1.55 | 1.34 | 1.80 | 0.002284 | 15621 |
| 530 | Diseases of esophagus | digestive | DR | 1.11 | 1.07 | 1.15 | 0.002344 | 15621 |
| 575.1 | Cholangitis | digestive | DR | 1.78 | 1.47 | 2.15 | 0.002365 | 15621 |
| 256 | Ovarian dysfunction | endocrine/metabolic | DR | 0.33 | 0.23 | 0.48 | 0.002408 | 15621 |
| 451 | Phlebitis and thrombophlebitis | circulatory system | DR | 1.39 | 1.24 | 1.54 | 0.002409 | 15621 |
| 840.3 | Joint/ligament sprain | injuries & poisonings | DR | 0.84 | 0.79 | 0.89 | 0.00251 | 15621 |
| 279 | Disorders involving the immune mechanism | endocrine/metabolic | DR | 1.58 | 1.36 | 1.84 | 0.002526 | 15621 |
| 558 | Noninfectious gastroenteritis | digestive | DR | 1.16 | 1.10 | 1.22 | 0.002552 | 15621 |
| 716 | Other arthropathies | musculoskeletal | DR | 1.13 | 1.08 | 1.17 | 0.002615 | 15621 |
| 527.7 | Disturbance of salivary secretion | digestive | DR | 1.24 | 1.15 | 1.33 | 0.002638 | 15621 |
| 1009 | Injury, NOS | NULL | DR | 1.12 | 1.08 | 1.17 | 0.002653 | 15621 |
| 593.2 | Microscopic hematuria | genitourinary | DR | 1.17 | 1.11 | 1.23 | 0.002655 | 15621 |
| 145 | Cancer of mouth | neoplasms | DR | 1.64 | 1.39 | 1.93 | 0.002705 | 15621 |
| 281.11 | Pernicious anemia | hematopoietic | DR | 1.78 | 1.47 | 2.16 | 0.002712 | 15621 |
| 394.3 | Aortic valve disease | circulatory system | DR | 2.80 | 1.98 | 3.94 | 0.00272 | 15621 |
| 332 | Parkinson's disease | neurological | DR | 1.41 | 1.26 | 1.59 | 0.002865 | 15621 |
| 519.8 | Other diseases of respiratory system, NEC | respiratory | DR | 1.31 | 1.19 | 1.43 | 0.00289 | 15621 |
| 756.5 | Congenital osteodystrophies | congenital anomalies | DR | 1.84 | 1.50 | 2.26 | 0.002902 | 15621 |
| 580.13 | Acute glomerulonephritis, NOS | genitourinary | DR | 2.53 | 1.85 | 3.46 | 0.003012 | 15621 |
| 276.8 | Polydipsia | endocrine/metabolic | DR | 0.51 | 0.41 | 0.64 | 0.003018 | 15621 |
| 369 | Infection of the eye | sense organs | DR | 1.15 | 1.10 | 1.21 | 0.003049 | 15621 |
| 574.11 | Cholelithiasis with acute cholecystitis | digestive | DR | 1.43 | 1.27 | 1.61 | 0.003056 | 15621 |
| 364.4 | Corneal degenerations | sense organs | DR | 1.37 | 1.23 | 1.53 | 0.003216 | 15621 |
| 426.22 | | | DR | 2.67 | 1.91 | 3.72 | 0.003238 | 15621 |
| 312 | Conduct disorders | mental disorders | DR | 0.76 | 0.69 | 0.83 | 0.00324 | 15621 |

| | | | | | | | | |
|---|---|---|---|---|---|---|---|---|
| 578.8 | Hemorrhage of rectum and anus | digestive | DR | 0.86 | 0.81 | 0.90 | 0.003247 | 15621 |
| 990 | Effects radiation NOS | injuries & poisonings | DR | 1.40 | 1.25 | 1.56 | 0.003249 | 15621 |
| 285.22 | Anemia in neoplastic disease | hematopoietic | DR | 1.65 | 1.39 | 1.95 | 0.003351 | 15621 |
| 757 | Congenital anomalies of the integument | congenital anomalies | DR | 1.77 | 1.45 | 2.14 | 0.003366 | 15621 |
| 681.2 | Cellulitis and abscess of face/neck | dermatologic | DR | 1.29 | 1.18 | 1.41 | 0.003422 | 15621 |
| 199 | Neoplasm of uncertain behavior | neoplasms | DR | 1.19 | 1.12 | 1.27 | 0.003471 | 15621 |
| 571.6 | Primary biliary cirrhosis | digestive | DR | 2.90 | 2.01 | 4.19 | 0.003605 | 15621 |
| 960.2 | Allergy/adverse effect of penicillin | injuries & poisonings | DR | 1.49 | 1.30 | 1.70 | 0.003675 | 15621 |
| 380 | Disorders of external ear | sense organs | DR | 1.27 | 1.17 | 1.38 | 0.003713 | 15621 |
| 597 | Other disorders of urethra and urinary tract | genitourinary | DR | 1.37 | 1.23 | 1.53 | 0.003733 | 15621 |
| 465 | Acute upper respiratory infections of multiple or unspecified sites | respiratory | DR | 1.10 | 1.07 | 1.14 | 0.00376 | 15621 |
| 289.3 | Personal history of diseases of blood and blood-forming organs | hematopoietic | DR | 2.28 | 1.72 | 3.04 | 0.0038 | 15621 |
| 389.3 | Degenerative and vascular disorders of ear | sense organs | DR | 1.52 | 1.32 | 1.76 | 0.003904 | 15621 |
| 519.9 | Symptoms involving respiratory system and other chest symptoms | respiratory | DR | 1.27 | 1.17 | 1.38 | 0.003997 | 15621 |
| 535.8 | Other specified gastritis | digestive | DR | 1.25 | 1.16 | 1.36 | 0.004039 | 15621 |
| 446.5 | Giant cell arteritis | circulatory system | DR | 2.04 | 1.59 | 2.62 | 0.004368 | 15621 |
| 531.2 | Gastric ulcer | digestive | DR | 1.33 | 1.20 | 1.47 | 0.004413 | 15621 |
| 575.8 | Other disorders of biliary tract | digestive | DR | 1.41 | 1.25 | 1.60 | 0.004509 | 15621 |
| 626.11 | Absent or infrequent menstruation | genitourinary | DR | 0.42 | 0.31 | 0.57 | 0.004554 | 15621 |
| 352.1 | Trigeminal nerve disorders [CN5] | neurological | DR | 1.38 | 1.23 | 1.54 | 0.004561 | 15621 |
| 571.51 | Cirrhosis of liver without mention of alcohol | digestive | DR | 1.17 | 1.11 | 1.24 | 0.004622 | 15621 |
| 526.8 | | | DR | 1.61 | 1.36 | 1.90 | 0.004656 | 15621 |
| 441.1 | Acute vascular insufficiency of intestine | circulatory system | DR | 2.26 | 1.70 | 3.03 | 0.00477 | 15621 |
| 442.3 | Aneurysm of artery of lower extremity | circulatory system | DR | 1.85 | 1.49 | 2.31 | 0.004967 | 15621 |
| 569.2 | Gastrointestinal complications | digestive | DR | 1.49 | 1.29 | 1.72 | 0.005115 | 15621 |
| 873 | Broken tooth | injuries & poisonings | DR | 1.27 | 1.17 | 1.38 | 0.00515 | 15621 |
| 656 | Other perinatal conditions of fetus or newborn | pregnancy complications | DR | 1.79 | 1.45 | 2.21 | 0.005199 | 15621 |
| 727.6 | Rupture of tendon, nontraumatic | musculoskeletal | DR | 1.19 | 1.12 | 1.27 | 0.005242 | 15621 |
| 741.5 | Hemarthrosis | musculoskeletal | DR | 2.00 | 1.56 | 2.57 | 0.005724 | 15621 |
| 972.2 | | | DR | 2.75 | 1.91 | 3.96 | 0.005745 | 15621 |
| 345.3 | Convulsions | neurological | DR | 1.23 | 1.14 | 1.32 | 0.005851 | 15621 |
| 573.1 | Chronic passive congestion of liver | digestive | DR | 2.37 | 1.73 | 3.25 | 0.0059 | 15621 |
| 751.21 | Cystic kidney disease | congenital anomalies | DR | 1.30 | 1.18 | 1.44 | 0.006075 | 15621 |
| 577.3 | Cyst and pseudocyst of pancreas | digestive | DR | 1.49 | 1.29 | 1.72 | 0.006121 | 15621 |
| 803.3 | Fracture of clavicle or scapula | injuries & poisonings | DR | 1.59 | 1.34 | 1.89 | 0.006208 | 15621 |
| 569 | Other disorders of intestine | digestive | DR | 1.23 | 1.14 | 1.33 | 0.006298 | 15621 |
| 481 | Influenza | respiratory | DR | 1.18 | 1.11 | 1.25 | 0.006379 | 15621 |
| 197 | Chemotherapy | neoplasms | DR | 1.31 | 1.19 | 1.45 | 0.006649 | 15621 |
| 578.1 | Hematemesis | digestive | DR | 1.36 | 1.22 | 1.53 | 0.006728 | 15621 |
| 802 | Fracture of pelvis | injuries & poisonings | DR | 2.03 | 1.56 | 2.63 | 0.006886 | 15621 |
| 425.8 | Other cardiomyopathy | circulatory system | DR | 2.17 | 1.63 | 2.90 | 0.006961 | 15621 |
| 291.4 | Specific nonpsychotic mental disorders due to brain damage | mental disorders | DR | 0.74 | 0.66 | 0.83 | 0.007066 | 15621 |
| 724 | | | DR | 0.85 | 0.80 | 0.90 | 0.007654 | 15621 |
| 531.3 | Duodenal ulcer | digestive | DR | 1.43 | 1.25 | 1.64 | 0.007667 | 15621 |
| 365.5 | Pseudoexfoliation glaucoma | sense organs | DR | 2.02 | 1.55 | 2.63 | 0.007697 | 15621 |
| 524 | Dentofacial anomalies, including malocclusion | digestive | DR | 1.17 | 1.10 | 1.25 | 0.007848 | 15621 |
| 172.3 | Carcinoma in situ of skin | neoplasms | DR | 1.82 | 1.46 | 2.29 | 0.00785 | 15621 |
| 575.2 | Obstruction of bile duct | digestive | DR | 1.60 | 1.34 | 1.91 | 0.008037 | 15621 |
| 627.2 | Symptomatic menopause | genitourinary | DR | 1.58 | 1.33 | 1.88 | 0.008087 | 15621 |
| 527.2 | Sialoadenitis | digestive | DR | 1.43 | 1.25 | 1.63 | 0.008339 | 15621 |
| 444.5 | Atheroembolism | circulatory system | DR | 15.90 | 5.56 | 45.48 | 0.008484 | 15621 |
| 687.3 | Changes in skin texture | dermatologic | DR | 1.64 | 1.36 | 1.98 | 0.008541 | 15621 |
| 729.3 | Panniculitis | musculoskeletal | DR | 1.97 | 1.52 | 2.54 | 0.008569 | 15621 |
| 381.1 | Otitis media | sense organs | DR | 1.13 | 1.08 | 1.19 | 0.008668 | 15621 |
| 753 | Congenital anomalies of the eye | congenital anomalies | DR | 1.42 | 1.24 | 1.63 | 0.008669 | 15621 |
| 287.31 | Primary thrombocytopenia | hematopoietic | DR | 1.65 | 1.36 | 2.00 | 0.008675 | 15621 |
| 287.32 | Secondary thrombocytopenia | hematopoietic | DR | 1.36 | 1.21 | 1.53 | 0.00879 | 15621 |
| 364.5 | Corneal dystrophy | sense organs | DR | 1.51 | 1.29 | 1.76 | 0.009097 | 15621 |
| 520 | Disorders of tooth development | digestive | DR | 1.20 | 1.12 | 1.28 | 0.009147 | 15621 |
| 800.1 | Fracture of neck of femur | injuries & poisonings | DR | 1.60 | 1.34 | 1.92 | 0.009287 | 15621 |
| 260.1 | Cachexia | endocrine/metabolic | DR | 1.74 | 1.40 | 2.15 | 0.009608 | 15621 |
| 362.7 | Hereditary retinal dystrophies | sense organs | DR | 1.44 | 1.25 | 1.66 | 0.009651 | 15621 |
| 514.1 | Abnormal results of function study of pulmonary system | respiratory | DR | 1.48 | 1.27 | 1.72 | 0.00971 | 15621 |
| 856 | Vascular complications of surgery and medical procedures | injuries & poisonings | DR | 1.87 | 1.47 | 2.39 | 0.010148 | 15621 |
| 772 | Symptoms of the muscles | symptoms | DR | 1.34 | 1.20 | 1.50 | 0.010209 | 15621 |
| 362.5 | Toxic maculopathy of retina | sense organs | DR | 2.07 | 1.56 | 2.76 | 0.010371 | 15621 |
| 971 | Poisoning by drugs primarily affecting the autonomic nervous system | injuries & poisonings | DR | 2.05 | 1.55 | 2.72 | 0.01045 | 15621 |
| 285.3 | Sideroblastic anemia | hematopoietic | DR | 3.83 | 2.27 | 6.48 | 0.010626 | 15621 |
| 368.91 | Psychophysical visual disturbances | sense organs | DR | 1.57 | 1.32 | 1.88 | 0.010904 | 15621 |
| 306.9 | Tension headache | mental disorders | DR | 0.75 | 0.67 | 0.84 | 0.011174 | 15621 |
| 253.3 | Diabetes insipidus | endocrine/metabolic | DR | 1.70 | 1.38 | 2.09 | 0.0112 | 15621 |
| 728.2 | Laxity of ligament or hypermobility syndrome | musculoskeletal | DR | 0.52 | 0.40 | 0.67 | 0.011209 | 15621 |
| 601.12 | Chronic prostatitis | genitourinary | DR | 1.30 | 1.17 | 1.44 | 0.011257 | 15621 |
| 90.3 | Venereal diseases due to Chlamydia trachomatis | infectious diseases | DR | 0.23 | 0.13 | 0.42 | 0.011281 | 15621 |
| 753.1 | Congenital cataract and lens anomalies | congenital anomalies | DR | 1.73 | 1.39 | 2.15 | 0.011583 | 15621 |
| 745 | Pain in joint | musculoskeletal | DR | 1.12 | 1.07 | 1.17 | 0.012103 | 15621 |
| 290.2 | Delirium due to conditions classified elsewhere | mental disorders | DR | 1.27 | 1.15 | 1.39 | 0.012625 | 15621 |
| 430.1 | Subarachnoid hemorrhage | circulatory system | DR | 1.72 | 1.38 | 2.13 | 0.012887 | 15621 |
| 560.2 | Impaction of intestine | digestive | DR | 1.45 | 1.25 | 1.68 | 0.012938 | 15621 |
| 695 | Erythematous conditions | dermatologic | DR | 1.13 | 1.07 | 1.18 | 0.013391 | 15621 |
| 571.81 | Portal hypertension | digestive | DR | 1.28 | 1.16 | 1.41 | 0.013495 | 15621 |
| 204.1 | Lymphoid leukemia | neoplasms | DR | 1.78 | 1.41 | 2.25 | 0.013714 | 15621 |
| 54 | Herpes simplex | infectious diseases | DR | 0.82 | 0.75 | 0.89 | 0.013785 | 15621 |
| 735.1 | Flat foot | musculoskeletal | DR | 0.87 | 0.82 | 0.92 | 0.014748 | 15621 |
| 300.13 | Phobia | mental disorders | DR | 1.42 | 1.23 | 1.63 | 0.015019 | 15621 |
| 580.14 | Chronic glomerulonephritis, NOS | genitourinary | DR | 1.94 | 1.48 | 2.56 | 0.015362 | 15621 |
| 191.1 | Cancer of brain and nervous system | neoplasms | DR | 1.76 | 1.39 | 2.22 | 0.015397 | 15621 |
| 506 | Empyema and pneumothorax | respiratory | DR | 1.42 | 1.23 | 1.64 | 0.015611 | 15621 |
| 251 | Other disorders of pancreatic internal secretion | endocrine/metabolic | DR | 1.97 | 1.49 | 2.61 | 0.015617 | 15621 |
| 577.2 | Chronic pancreatitis | digestive | DR | 1.34 | 1.19 | 1.52 | 0.015675 | 15621 |
| 520.1 | Hereditary disturbances in tooth structure | digestive | DR | 1.27 | 1.15 | 1.41 | 0.015934 | 15621 |
| 723 | Other disorders of cervical region | musculoskeletal | DR | 0.77 | 0.69 | 0.86 | 0.015954 | 15621 |
| 763 | Thoracic or lumbosacral neuritis or radiculitis, unspecified | symptoms | DR | 0.90 | 0.86 | 0.94 | 0.016052 | 15621 |
| 381 | | | DR | 1.11 | 1.07 | 1.17 | 0.016145 | 15621 |
| 360.3 | Hypotony of eye | sense organs | DR | 2.27 | 1.61 | 3.18 | 0.016155 | 15621 |
| 382 | Otalgia | sense organs | DR | 1.15 | 1.09 | 1.22 | 0.016409 | 15621 |

| | | | | | | | | |
|---|---|---|---|---|---|---|---|---|
| 196 | | | DR | 1.44 | 1.24 | 1.68 | 0.016427 | 15621 |
| 615 | Endometriosis | genitourinary | DR | 0.39 | 0.26 | 0.57 | 0.016716 | 15621 |
| 327.72 | Sleep related leg cramps | neurological | DR | 1.41 | 1.22 | 1.62 | 0.016752 | 15621 |
| 153.3 | Malignant neoplasm of rectum, rectosigmoid junction, and anus | neoplasms | DR | 1.46 | 1.25 | 1.72 | 0.017078 | 15621 |
| 871.2 | Open wound of finger(s) | injuries & poisonings | DR | 1.22 | 1.12 | 1.32 | 0.017098 | 15621 |
| 739 | Contracture of joint | musculoskeletal | DR | 1.33 | 1.18 | 1.51 | 0.017172 | 15621 |
| 818 | Intracranial hemorrhage (injury) | injuries & poisonings | DR | 1.38 | 1.21 | 1.58 | 0.017346 | 15621 |
| 656.3 | Endocrine and metabolic disturbances of fetus and newborn | pregnancy complications | DR | 5.27 | 2.61 | 10.62 | 0.017787 | 15621 |
| 338.1 | Acute pain | neurological | DR | 1.11 | 1.06 | 1.16 | 0.017855 | 15621 |
| 912 | Insect bite | injuries & poisonings | DR | 1.23 | 1.13 | 1.34 | 0.017903 | 15621 |
| 567 | Peritonitis and retroperitoneal infections | digestive | DR | 1.29 | 1.16 | 1.44 | 0.017986 | 15621 |
| 191 | Manlignant and unknown neoplasms of brain and nervous system | neoplasms | DR | 1.54 | 1.28 | 1.84 | 0.018036 | 15621 |
| 364.9 | Cornea replaced by transplant | sense organs | DR | 2.01 | 1.50 | 2.70 | 0.018099 | 15621 |
| 1090 | Acquired absence of organs | | DR | 1.21 | 1.11 | 1.31 | 0.018393 | 15621 |
| 771 | Musculoskeletal symptoms referable to limbs | symptoms | DR | 1.33 | 1.18 | 1.49 | 0.018996 | 15621 |
| 145.2 | Cancer of tongue | neoplasms | DR | 1.89 | 1.44 | 2.49 | 0.020095 | 15621 |
| 187.2 | Malignant neoplasm of testis | neoplasms | DR | 0.53 | 0.40 | 0.70 | 0.020159 | 15621 |
| 793.2 | Nonspecific abnormal findings on radiological and other examination of other intrathoracic organs (echocardiogram, etc) | symptoms | DR | 1.19 | 1.11 | 1.29 | 0.020213 | 15621 |
| 743.21 | Pathologic fracture of vertebrae | musculoskeletal | DR | 1.68 | 1.34 | 2.10 | 0.020433 | 15621 |
| 288.3 | Eosinophilia | hematopoietic | DR | 1.55 | 1.28 | 1.88 | 0.020567 | 15621 |
| 690 | Erythematosquamous dermatosis | dermatologic | DR | 1.14 | 1.07 | 1.20 | 0.021676 | 15621 |
| 477 | Epistaxis or throat hemorrhage | respiratory | DR | 1.17 | 1.09 | 1.26 | 0.021725 | 15621 |
| 286.8 | Hypercoagulable state | hematopoietic | DR | 1.56 | 1.28 | 1.89 | 0.021939 | 15621 |
| 586.3 | Vascular disorders of kidney/hypertrophy | genitourinary | DR | 2.08 | 1.51 | 2.87 | 0.02221 | 15621 |
| 646 | Other complications of pregnancy NEC | pregnancy complications | DR | 0.10 | 0.03 | 0.27 | 0.022614 | 15621 |
| 362.31 | Separation of retinal layers | sense organs | DR | 1.37 | 1.19 | 1.57 | 0.023167 | 15621 |
| 792.1 | Papanicolaou smear of cervix or vagina with atypical squamous cells | genitourinary | DR | 0.47 | 0.34 | 0.65 | 0.023211 | 15621 |
| 480.3 | Pneumonia due to fungus (mycoses) | respiratory | DR | 1.39 | 1.20 | 1.60 | 0.023242 | 15621 |
| 198.2 | Secondary malignancy of respiratory organs | neoplasms | DR | 1.50 | 1.25 | 1.79 | 0.023287 | 15621 |
| 870.3 | Other open wound of head and face | injuries & poisonings | DR | 1.27 | 1.14 | 1.42 | 0.023653 | 15621 |
| 690.1 | Seborrheic dermatitis | dermatologic | DR | 1.14 | 1.07 | 1.20 | 0.023678 | 15621 |
| 348.9 | Other conditions of brain, NOS | neurological | DR | 1.28 | 1.15 | 1.42 | 0.023732 | 15621 |
| 586.4 | Stricture/obstruction of ureter | genitourinary | DR | 1.27 | 1.14 | 1.42 | 0.023756 | 15621 |
| 223 | Benign neoplasm of kidney and other urinary organs | neoplasms | DR | 1.40 | 1.21 | 1.63 | 0.023942 | 15621 |
| 562.2 | Diverticulitis | digestive | DR | 0.85 | 0.79 | 0.91 | 0.02401 | 15621 |
| 480.13 | MRSA pneumonia | respiratory | DR | 2.39 | 1.62 | 3.51 | 0.024035 | 15621 |
| 627.1 | Postmenopausal bleeding | genitourinary | DR | 1.85 | 1.41 | 2.42 | 0.02405 | 15621 |
| 473.3 | Paralysis/spasm of vocal cords or larynx | respiratory | DR | 1.53 | 1.26 | 1.84 | 0.024469 | 15621 |
| 595 | Hydronephrosis | genitourinary | DR | 1.22 | 1.12 | 1.33 | 0.024568 | 15621 |
| 1004 | Other signs and symptoms involving emotional state | NULL | DR | 0.83 | 0.76 | 0.90 | 0.024792 | 15621 |
| 722 | | | DR | 0.92 | 0.89 | 0.95 | 0.024844 | 15621 |
| 369.5 | Conjunctivitis, infectious | sense organs | DR | 1.12 | 1.07 | 1.18 | 0.025211 | 15621 |
| 580.11 | Proliferative glomerulonephritis | genitourinary | DR | 2.03 | 1.48 | 2.78 | 0.025889 | 15621 |
| 726.4 | Calcaneal spur; Exostosis NOS | musculoskeletal | DR | 1.14 | 1.07 | 1.20 | 0.026219 | 15621 |
| 502 | Postinflammatory pulmonary fibrosis | respiratory | DR | 1.28 | 1.15 | 1.44 | 0.026596 | 15621 |
| 149 | Cancer of larynx, pharynx, nasal cavities | neoplasms | DR | 1.45 | 1.23 | 1.72 | 0.027351 | 15621 |
| 411.41 | Aneurysm and dissection of heart | circulatory system | DR | 1.71 | 1.34 | 2.19 | 0.02754 | 15621 |
| 755.3 | Congenital anomaly of fingers/toes | congenital anomalies | DR | 2.42 | 1.62 | 3.62 | 0.027686 | 15621 |
| 709 | | | DR | 1.34 | 1.17 | 1.53 | 0.027757 | 15621 |
| 530.1 | Esophagitis, GERD and related diseases | digestive | DR | 1.08 | 1.04 | 1.11 | 0.028218 | 15621 |
| 760 | Back pain | symptoms | DR | 0.92 | 0.89 | 0.96 | 0.028754 | 15621 |
| 974 | Poisoning by water, mineral, and uric acid metabolism drugs | injuries & poisonings | DR | 1.96 | 1.44 | 2.67 | 0.028821 | 15621 |
| 601.11 | Acute prostatitis | genitourinary | DR | 1.25 | 1.13 | 1.39 | 0.029311 | 15621 |
| 586.2 | Cyst of kidney, acquired | genitourinary | DR | 1.17 | 1.09 | 1.26 | 0.029359 | 15621 |
| 722.1 | Displacement of intervertebral disc | musculoskeletal | DR | 0.88 | 0.83 | 0.93 | 0.029686 | 15621 |
| 530.3 | Stricture and stenosis of esophagus | digestive | DR | 1.36 | 1.18 | 1.56 | 0.029932 | 15621 |
| 614.53 | Cyst or abscess of Bartholin's gland | genitourinary | DR | 3.16 | 1.86 | 5.38 | 0.030312 | 15621 |
| 495.2 | Asthma with exacerbation | respiratory | DR | 0.82 | 0.74 | 0.90 | 0.030351 | 15621 |
| 972.1 | | | DR | 3.79 | 2.05 | 7.03 | 0.03072 | 15621 |
| 907 | Injuries to the nervous system | injuries & poisonings | DR | 1.20 | 1.10 | 1.31 | 0.031375 | 15621 |
| 801.1 | Fracture of foot | injuries & poisonings | DR | 1.25 | 1.13 | 1.39 | 0.031429 | 15621 |
| 364.51 | Fuchs' dystrophy | sense organs | DR | 1.52 | 1.25 | 1.84 | 0.031508 | 15621 |
| 550.4 | Umbilical hernia | digestive | DR | 0.86 | 0.80 | 0.92 | 0.03187 | 15621 |
| 195 | Cancer, suspected or other | neoplasms | DR | 1.20 | 1.10 | 1.30 | 0.032132 | 15621 |
| 149.1 | Cancer of oropharynx | neoplasms | DR | 1.85 | 1.39 | 2.46 | 0.032274 | 15621 |
| 704.11 | Alopecia Areata | dermatologic | DR | 0.63 | 0.50 | 0.78 | 0.032914 | 15621 |
| 509.3 | Pulmonary insufficiency or respiratory failure following trauma and surgery | respiratory | DR | 1.34 | 1.17 | 1.53 | 0.033637 | 15621 |
| 855 | | | DR | 0.30 | 0.17 | 0.53 | 0.033765 | 15621 |
| 288.1 | Decreased white blood cell count | hematopoietic | DR | 1.25 | 1.13 | 1.39 | 0.033858 | 15621 |
| 530.6 | Diverticulum of esophagus, acquired | digestive | DR | 2.24 | 1.53 | 3.28 | 0.034104 | 15621 |
| 1008 | Crushing or internal injury to organs | NULL | DR | 1.29 | 1.14 | 1.45 | 0.034202 | 15621 |
| 530.11 | GERD | digestive | DR | 1.07 | 1.04 | 1.11 | 0.034618 | 15621 |
| 159.3 | Malignant neoplasm of gallbladder and extrahepatic bile ducts | neoplasms | DR | 2.11 | 1.48 | 3.00 | 0.035614 | 15621 |
| 324 | Other CNS infection and poliomyelitis | neurological | DR | 1.55 | 1.26 | 1.91 | 0.036297 | 15621 |
| 704.1 | Alopecia | dermatologic | DR | 0.78 | 0.70 | 0.88 | 0.036466 | 15621 |
| 211 | Benign neoplasm of other parts of digestive system | neoplasms | DR | 1.20 | 1.10 | 1.30 | 0.036532 | 15621 |
| 751.2 | Congenital anomalies of urinary system | congenital anomalies | DR | 1.21 | 1.10 | 1.32 | 0.037551 | 15621 |
| 204.12 | Lymphoid leukemia, chronic | neoplasms | DR | 1.77 | 1.35 | 2.34 | 0.038167 | 15621 |
| 389.2 | Conductive hearing loss | sense organs | DR | 1.20 | 1.10 | 1.31 | 0.038355 | 15621 |
| 611.3 | Lump or mass in breast | genitourinary | DR | 1.27 | 1.13 | 1.43 | 0.038968 | 15621 |
| 724.2 | | | DR | 1.56 | 1.26 | 1.94 | 0.039108 | 15621 |
| 368.1 | Amblyopia | sense organs | DR | 1.25 | 1.12 | 1.40 | 0.039256 | 15621 |
| 250.11 | Type 1 diabetes with ketoacidosis | endocrine/metabolic | DR | 1.50 | 1.23 | 1.82 | 0.039377 | 15621 |
| 241.1 | Nontoxic uninodular goiter | endocrine/metabolic | DR | 1.20 | 1.10 | 1.31 | 0.03979 | 15621 |
| 738.4 | Acquired spondylolisthesis | musculoskeletal | DR | 0.83 | 0.75 | 0.91 | 0.040407 | 15621 |
| 327.5 | Parasomnia | neurological | DR | 0.87 | 0.81 | 0.93 | 0.040485 | 15621 |
| 962.3 | Hormones and synthetic substitutes causing adverse effects in therapeutic use | injuries & poisonings | DR | 1.37 | 1.17 | 1.59 | 0.04059 | 15621 |
| 695.8 | Other specified erythematous conditions | dermatologic | DR | 1.19 | 1.09 | 1.30 | 0.04116 | 15621 |
| 526 | Diseases of the jaws | digestive | DR | 1.13 | 1.06 | 1.19 | 0.041577 | 15621 |
| 395.4 | Nonrheumatic pulmonary valve disorders | circulatory system | DR | 1.59 | 1.27 | 1.99 | 0.041611 | 15621 |
| 283.2 | Non-autoimmune hemolytic anemias | hematopoietic | DR | 2.49 | 1.59 | 3.90 | 0.042363 | 15621 |
| 649.1 | Diabetes or abnormal glucose tolerance complicating pregnancy | pregnancy complications | DR | 0.43 | 0.29 | 0.65 | 0.042899 | 15621 |
| 320 | Meningitis | neurological | DR | 1.50 | 1.23 | 1.84 | 0.042998 | 15621 |

| | | | | | | | | |
|---|---|---|---|---|---|---|---|---|
| 165 | Cancer within the respiratory system | neoplasms | DR | 1.27 | 1.13 | 1.43 | 0.043487 | 15621 |
| 626.13 | Irregular menstrual cycle | genitourinary | DR | 0.62 | 0.48 | 0.78 | 0.043495 | 15621 |
| 965 | Poisoning by analgesics, antipyretics, and antirheumatics | injuries & poisonings | DR | 1.37 | 1.17 | 1.60 | 0.043767 | 15621 |
| 574.12 | Cholelithiasis with other cholecystitis | digestive | DR | 1.21 | 1.10 | 1.33 | 0.044358 | 15621 |
| 185 | Cancer of prostate | neoplasms | DR | 1.13 | 1.06 | 1.20 | 0.044569 | 15621 |
| 721.1 | Spondylosis without myelopathy | musculoskeletal | DR | 0.93 | 0.89 | 0.96 | 0.044849 | 15621 |
| 720 | Spinal stenosis | musculoskeletal | DR | 1.09 | 1.05 | 1.14 | 0.045019 | 15621 |
| 289 | Other diseases of blood and blood-forming organs | hematopoietic | DR | 1.24 | 1.12 | 1.39 | 0.045561 | 15621 |
| 702.4 | Degenerative skin disorders | dermatologic | DR | 2.35 | 1.53 | 3.59 | 0.045649 | 15621 |
| 580.12 | Non-proliferative glomerulonephritis | genitourinary | DR | 1.66 | 1.29 | 2.15 | 0.046235 | 15621 |
| 216 | Benign neoplasm of skin | neoplasms | DR | 1.10 | 1.05 | 1.15 | 0.046355 | 15621 |
| 792 | Abnormal Papanicolaou smear of cervix and cervical HPV | genitourinary | DR | 0.64 | 0.51 | 0.80 | 0.046405 | 15621 |
| 649 | Other conditions or status of the mother complicating pregnancy, childbirth, or the puerperium | pregnancy complications | DR | 0.37 | 0.22 | 0.61 | 0.047169 | 15621 |
| 513.31 | Apnea | respiratory | DR | 1.32 | 1.15 | 1.52 | 0.047308 | 15621 |
| 626.2 | Dysmenorrhea | genitourinary | DR | 0.59 | 0.45 | 0.77 | 0.047441 | 15621 |
| 479 | Other upper respiratory disease | respiratory | DR | 0.90 | 0.86 | 0.95 | 0.04745 | 15621 |
| 695.3 | Rosacea | dermatologic | DR | 0.82 | 0.74 | 0.91 | 0.04759 | 15621 |
| 782.6 | Pallor and flushing | symptoms | DR | 1.55 | 1.24 | 1.93 | 0.048133 | 15621 |
| 528.4 | Cysts of oral soft tissues | digestive | DR | 0.53 | 0.38 | 0.73 | 0.048434 | 15621 |
| 292.3 | Memory loss | mental disorders | DR | 1.13 | 1.06 | 1.20 | 0.048501 | 15621 |
| 222 | Benign neoplasm of male genital organs | neoplasms | DR | 1.19 | 1.09 | 1.29 | 0.048603 | 15621 |
| 580.4 | Renal sclerosis, NOS | genitourinary | DR | 1.78 | 1.33 | 2.38 | 0.048637 | 15621 |
| 159 | Malignant neoplasm of other and ill-defined sites within the digestive organs and peritoneum | neoplasms | DR | 1.42 | 1.19 | 1.69 | 0.049059 | 15621 |
| 180 | | | DR | 0.52 | 0.37 | 0.72 | 0.049339 | 15621 |
| 255.2 | | | DR | 1.45 | 1.20 | 1.74 | 0.049386 | 15621 |
| 345.1 | Epilepsy | neurological | DR | 1.19 | 1.09 | 1.30 | 0.049607 | 15621 |
| 564.8 | Abnormal findings on exam of gastrointestinal tract/ abdominal area | digestive | DR | 1.15 | 1.07 | 1.24 | 0.0501 | 15621 |
| 593.1 | Gross hematuria | genitourinary | DR | 1.14 | 1.07 | 1.22 | 0.050367 | 15621 |
| 430.3 | Subdural hemorrhage | circulatory system | DR | 1.35 | 1.16 | 1.58 | 0.051438 | 15621 |
| 568 | Other disorders of peritoneum | digestive | DR | 1.26 | 1.12 | 1.42 | 0.051724 | 15621 |
| 1007 | Injury to blood vessels | NULL | DR | 1.45 | 1.20 | 1.75 | 0.052158 | 15621 |
| 496.3 | Bronchiectasis | respiratory | DR | 1.42 | 1.18 | 1.71 | 0.053985 | 15621 |
| 290.13 | Senile dementia | mental disorders | DR | 1.68 | 1.28 | 2.19 | 0.055104 | 15621 |
| 241 | Nontoxic nodular goiter | endocrine/metabolic | DR | 1.18 | 1.08 | 1.28 | 0.055375 | 15621 |
| 283.1 | Autoimmune hemolytic anemias | hematopoietic | DR | 2.55 | 1.56 | 4.16 | 0.056386 | 15621 |
| 451.2 | Phlebitis and thrombophlebitis of lower extremities | circulatory system | DR | 1.31 | 1.14 | 1.52 | 0.056573 | 15621 |
| 656.2 | Respiratory conditions of fetus and newborn | pregnancy complications | DR | 3.81 | 1.89 | 7.68 | 0.056869 | 15621 |
| 289.8 | Polycythemia, secondary | hematopoietic | DR | 0.76 | 0.66 | 0.88 | 0.056985 | 15621 |
| 609.11 | Azoospermia and oligospermia | genitourinary | DR | 0.35 | 0.20 | 0.61 | 0.057117 | 15621 |
| 117.2 | Coccidioidomycosis | infectious diseases | DR | 1.77 | 1.31 | 2.40 | 0.05723 | 15621 |
| 736 | Other acquired deformities of limbs | musculoskeletal | DR | 1.14 | 1.06 | 1.22 | 0.057843 | 15621 |
| 958.2 | Traumatic and surgical subcutaneous emphysema | injuries & poisonings | DR | 2.47 | 1.53 | 3.98 | 0.057902 | 15621 |
| 761 | Cervicalgia | symptoms | DR | 0.93 | 0.90 | 0.97 | 0.058379 | 15621 |
| 755.1 | Congenital deformities of feet | congenital anomalies | DR | 0.88 | 0.82 | 0.94 | 0.05883 | 15621 |
| 985 | Toxic effect of other metals | injuries & poisonings | DR | 1.80 | 1.32 | 2.46 | 0.059226 | 15621 |
| 573.3 | Hepatomegaly | digestive | DR | 0.86 | 0.79 | 0.93 | 0.059451 | 15621 |
| 191.11 | Cancer of brain | neoplasms | DR | 1.63 | 1.25 | 2.11 | 0.060453 | 15621 |
| 601.8 | Other inflammatory disorders of male genital organs | genitourinary | DR | 1.28 | 1.12 | 1.47 | 0.060814 | 15621 |
| 789.1 | | | DR | 1.95 | 1.37 | 2.79 | 0.061018 | 15621 |
| 341 | Other demyelinating diseases of central nervous system | neurological | DR | 0.45 | 0.30 | 0.69 | 0.061246 | 15621 |
| 958 | Certain early complications of trauma or procedure | injuries & poisonings | DR | 1.40 | 1.17 | 1.67 | 0.061257 | 15621 |
| 1012 | Late effect | NULL | DR | 0.71 | 0.59 | 0.85 | 0.061532 | 15621 |
| 740.1 | Osteoarthritis; localized | musculoskeletal | DR | 1.07 | 1.03 | 1.10 | 0.061578 | 15621 |
| 709.2 | Sicca syndrome | dermatologic | DR | 1.35 | 1.15 | 1.59 | 0.062018 | 15621 |
| 333.2 | Myoclonus | neurological | DR | 1.38 | 1.16 | 1.64 | 0.062413 | 15621 |
| 530.2 | Esophageal bleeding (varices/hemorrhage) | digestive | DR | 1.17 | 1.08 | 1.27 | 0.06305 | 15621 |
| 289.5 | Diseases of spleen | hematopoietic | DR | 1.38 | 1.16 | 1.65 | 0.063373 | 15621 |
| 41.8 | H. pylori | infectious diseases | DR | 1.15 | 1.07 | 1.24 | 0.063465 | 15621 |
| 446.1 | Thromboangiitis obliterans | circulatory system | DR | 4.55 | 2.01 | 10.30 | 0.063857 | 15621 |
| 145.5 | Cancer of the mouth floor | neoplasms | DR | 2.47 | 1.51 | 4.02 | 0.063981 | 15621 |
| 710.3 | Osteopathy resulting from poliomyelitis | musculoskeletal | DR | 4.55 | 2.00 | 10.36 | 0.065582 | 15621 |
| 327.41 | Organic or persistent insomnia | neurological | DR | 0.83 | 0.75 | 0.92 | 0.065611 | 15621 |
| 427.9 | Palpitations | circulatory system | DR | 1.10 | 1.05 | 1.17 | 0.066138 | 15621 |
| 794 | Abnormal results of other function studies (bladder, pancreas, placenta, spleen, etc) | symptoms | DR | 1.22 | 1.10 | 1.37 | 0.066147 | 15621 |
| 172.11 | Melanomas of skin | neoplasms | DR | 1.45 | 1.18 | 1.77 | 0.06711 | 15621 |
| 366.3 | Traumatic cataract | sense organs | DR | 1.59 | 1.23 | 2.06 | 0.068829 | 15621 |
| 747.12 | Valvular heart disease/ heart chambers | congenital anomalies | DR | 1.57 | 1.23 | 2.02 | 0.069226 | 15621 |
| 701.4 | Keloid scar | dermatologic | DR | 0.78 | 0.68 | 0.89 | 0.069497 | 15621 |
| 388 | Other disorders of ear | sense organs | DR | 1.19 | 1.08 | 1.31 | 0.069712 | 15621 |
| 250.5 | Glycosuria or Acetonuria | endocrine/metabolic | DR | 1.53 | 1.21 | 1.93 | 0.069993 | 15621 |
| 180.3 | Cervical intraepithelial neoplasia [CIN] [Cervical dysplasia] | neoplasms | DR | 0.52 | 0.36 | 0.74 | 0.070128 | 15621 |
| 394.2 | Mitral valve disease | circulatory system | DR | 1.61 | 1.24 | 2.09 | 0.070408 | 15621 |
| 303.4 | Somatoform disorder | mental disorders | DR | 0.87 | 0.80 | 0.94 | 0.07074 | 15621 |
| 818.2 | | | DR | 2.24 | 1.43 | 3.50 | 0.070945 | 15621 |
| 942 | Infusion and transfusion reaction | injuries & poisonings | DR | 2.03 | 1.37 | 3.00 | 0.072167 | 15621 |
| 362.6 | Peripheral retinal degenerations | sense organs | DR | 1.14 | 1.06 | 1.23 | 0.072255 | 15621 |
| 573.4 | Acute and subacute necrosis of liver | digestive | DR | 1.32 | 1.13 | 1.55 | 0.072438 | 15621 |
| 804 | Fracture of hand or wrist | injuries & poisonings | DR | 1.15 | 1.06 | 1.24 | 0.073404 | 15621 |
| 277.1 | Disorders of porphyrin metabolism | endocrine/metabolic | DR | 2.78 | 1.57 | 4.92 | 0.07396 | 15621 |
| 184 | Cancer of other female genital organs | neoplasms | DR | 2.02 | 1.36 | 2.99 | 0.074387 | 15621 |
| 433.5 | Cerebral aneurysm | circulatory system | DR | 1.40 | 1.16 | 1.70 | 0.075465 | 15621 |
| 303 | Psychogenic and somatoform disorders | mental disorders | DR | 0.88 | 0.82 | 0.95 | 0.075504 | 15621 |
| 381.3 | Mastoiditis & related conditions | sense organs | DR | 1.35 | 1.14 | 1.60 | 0.076058 | 15621 |
| 620.1 | Dysplasia of cervix | genitourinary | DR | 0.26 | 0.12 | 0.56 | 0.077373 | 15621 |
| 259.2 | Carcinoid syndrome | endocrine/metabolic | DR | 0.37 | 0.21 | 0.65 | 0.077607 | 15621 |
| 715.2 | Ankylosing spondylitis | musculoskeletal | DR | 0.67 | 0.53 | 0.84 | 0.077979 | 15621 |
| 599.3 | Dysuria | genitourinary | DR | 1.11 | 1.05 | 1.18 | 0.078252 | 15621 |
| 624 | Symptoms involving female genital tract | genitourinary | DR | 0.49 | 0.33 | 0.74 | 0.078944 | 15621 |
| 277.5 | Other disorders of lipoid metabolism | endocrine/metabolic | DR | 0.89 | 0.83 | 0.95 | 0.079547 | 15621 |
| 378.1 | Strabismus (not specified as paralytic) | sense organs | DR | 1.16 | 1.07 | 1.26 | 0.080477 | 15621 |
| 318 | Tobacco use disorder | mental disorders | DR | 1.06 | 1.03 | 1.10 | 0.081007 | 15621 |
| 626.12 | Excessive or frequent menstruation | genitourinary | DR | 0.67 | 0.53 | 0.84 | 0.081176 | 15621 |
| 259 | Other endocrine disorders | endocrine/metabolic | DR | 1.27 | 1.11 | 1.46 | 0.081421 | 15621 |

| | | | | | | | | |
|---|---|---|---|---|---|---|---|---|
| 361.2 | Retinoschisis and retinal cysts | sense organs | DR | 1.35 | 1.13 | 1.60 | 0.082278 | 15621 |
| 287.1 | Spontaneous ecchymoses | hematopoietic | DR | 1.36 | 1.14 | 1.63 | 0.083205 | 15621 |
| 446.9 | Arteritis NOS | circulatory system | DR | 1.46 | 1.17 | 1.82 | 0.083288 | 15621 |
| 691 | Congenital anomalies of skin | dermatologic | DR | 1.25 | 1.10 | 1.42 | 0.083616 | 15621 |
| 519.2 | Respiratory complications | respiratory | DR | 1.36 | 1.14 | 1.63 | 0.083929 | 15621 |
| 281.13 | Folate-deficiency anemia | hematopoietic | DR | 1.58 | 1.21 | 2.07 | 0.085574 | 15621 |
| 793 | Nonspecific abnormal findings on radiological and other examination of musculoskeletal system | symptoms | DR | 1.18 | 1.07 | 1.30 | 0.086002 | 15621 |
| 938.1 | Acute dermatitis due to solar radiation | dermatologic | DR | 0.71 | 0.58 | 0.87 | 0.086255 | 15621 |
| 149.5 | | | DR | 2.25 | 1.40 | 3.62 | 0.087785 | 15621 |
| 202.2 | Non-Hodgkins lymphoma | neoplasms | DR | 1.25 | 1.10 | 1.42 | 0.089022 | 15621 |
| 270.31 | Polyclonal hypergammaglobulinemia | endocrine/metabolic | DR | 2.14 | 1.37 | 3.36 | 0.089126 | 15621 |
| 420.22 | Chronic pericarditis | circulatory system | DR | 1.88 | 1.30 | 2.73 | 0.089262 | 15621 |
| 611 | Abnormal findings on mammogram or breast exam | genitourinary | DR | 1.21 | 1.08 | 1.35 | 0.090442 | 15621 |
| 726.2 | Synoviopathy | musculoskeletal | DR | 1.14 | 1.05 | 1.23 | 0.090717 | 15621 |
| 293.1 | Swelling, mass, or lump in head and neck [Space-occupying lesion, intracranial NOS] | mental disorders | DR | 1.12 | 1.05 | 1.20 | 0.091909 | 15621 |
| 586.12 | Vesicoureteral reflux | genitourinary | DR | 6.81 | 2.18 | 21.27 | 0.092225 | 15621 |
| 656.6 | Perinatal disorders of digestive system | pregnancy complications | DR | 2.34 | 1.41 | 3.88 | 0.092454 | 15621 |
| 427.7 | Tachycardia NOS | circulatory system | DR | 1.09 | 1.04 | 1.15 | 0.092544 | 15621 |
| 723.1 | Torticollis | musculoskeletal | DR | 0.71 | 0.58 | 0.87 | 0.092809 | 15621 |
| 259.1 | Nonspecific abnormal results of other endocrine function study | endocrine/metabolic | DR | 0.54 | 0.38 | 0.78 | 0.093718 | 15621 |
| 747.11 | Cardiac shunt/ heart septal defect | congenital anomalies | DR | 1.47 | 1.17 | 1.85 | 0.096025 | 15621 |
| 79 | Viral infection | infectious diseases | DR | 1.10 | 1.04 | 1.16 | 0.096818 | 15621 |
| 255.21 | Glucocorticoid deficiency | endocrine/metabolic | DR | 1.41 | 1.15 | 1.74 | 0.097275 | 15621 |
| 528 | Diseases of the oral soft tissues, excluding lesions specific for gingiva and tongue | digestive | DR | 1.10 | 1.04 | 1.17 | 0.098409 | 15621 |
| 195.1 | Malignant neoplasm, other | neoplasms | DR | 1.21 | 1.08 | 1.36 | 0.100166 | 15621 |
| 446.3 | Hypersensitivity angiitis | circulatory system | DR | 3.07 | 1.55 | 6.08 | 0.100261 | 15621 |
| 614.1 | Pelvic peritoneal adhesions, female (postoperative) (postinfection) | genitourinary | DR | 0.18 | 0.06 | 0.51 | 0.101958 | 15621 |
| 70.9 | Hepatitis NOS | infectious diseases | DR | 1.16 | 1.06 | 1.27 | 0.103172 | 15621 |
| 288.11 | Neutropenia | hematopoietic | DR | 1.26 | 1.09 | 1.45 | 0.103815 | 15621 |
| 472 | Chronic pharyngitis and nasopharyngitis | respiratory | DR | 0.92 | 0.87 | 0.97 | 0.104398 | 15621 |
| 573.5 | Jaundice (not of newborn) | digestive | DR | 1.24 | 1.09 | 1.42 | 0.104961 | 15621 |
| 755 | Congenital anomalies of limbs | congenital anomalies | DR | 0.90 | 0.84 | 0.96 | 0.105576 | 15621 |
| 420.1 | Myocarditis | circulatory system | DR | 2.49 | 1.42 | 4.38 | 0.105649 | 15621 |
| 736.2 | Acquired deformities of finger | musculoskeletal | DR | 1.32 | 1.11 | 1.57 | 0.105755 | 15621 |
| 165.1 | Cancer of bronchus; lung | neoplasms | DR | 1.22 | 1.08 | 1.39 | 0.107642 | 15621 |
| 741.2 | Stiffness of joint | musculoskeletal | DR | 1.16 | 1.06 | 1.28 | 0.10821 | 15621 |
| 189.21 | Malignant neoplasm of bladder | neoplasms | DR | 1.22 | 1.08 | 1.38 | 0.108723 | 15621 |
| 627.3 | Postmenopausal atrophic vaginitis | genitourinary | DR | 1.41 | 1.14 | 1.74 | 0.109068 | 15621 |
| 614.5 | Inflammatory disease of cervix, vagina, and vulva | genitourinary | DR | 1.32 | 1.11 | 1.57 | 0.109987 | 15621 |
| 696.4 | Psoriasis | dermatologic | DR | 1.12 | 1.04 | 1.20 | 0.11081 | 15621 |
| 510.2 | Lung transplant | respiratory | DR | 2.38 | 1.38 | 4.11 | 0.111151 | 15621 |
| 614.52 | Vaginitis and vulvovaginitis | genitourinary | DR | 1.33 | 1.11 | 1.59 | 0.111717 | 15621 |
| 626.15 | Infertility, female, associated with anovulation | genitourinary | DR | 0.36 | 0.19 | 0.68 | 0.112097 | 15621 |
| 704.8 | Other specified diseases of hair and hair follicles | dermatologic | DR | 1.10 | 1.04 | 1.17 | 0.113187 | 15621 |
| 286.81 | Primary hypercoagulable state | hematopoietic | DR | 1.40 | 1.13 | 1.72 | 0.115488 | 15621 |
| 1010.3 | screening for other diseases and disorders | | DR | 1.06 | 1.02 | 1.10 | 0.115726 | 15621 |
| 79.2 | Infectious mononucleosis | infectious diseases | DR | 0.18 | 0.06 | 0.54 | 0.115845 | 15621 |
| 790.1 | Elevated sedimentation rate | symptoms | DR | 1.42 | 1.14 | 1.78 | 0.116626 | 15621 |
| 198.6 | Secondary malignancy of bone | neoplasms | DR | 1.27 | 1.09 | 1.49 | 0.117263 | 15621 |
| 742.2 | Pathological, developmental or recurrent dislocation | musculoskeletal | DR | 0.74 | 0.61 | 0.90 | 0.117766 | 15621 |
| 947 | Urticaria | dermatologic | DR | 0.89 | 0.83 | 0.96 | 0.118339 | 15621 |
| 602 | Other disorders of prostate | genitourinary | DR | 1.14 | 1.05 | 1.24 | 0.118652 | 15621 |
| 671 | Venous/cerebrovascular complications embolism in pregnancy and the puerperium | pregnancy complications | DR | 0.30 | 0.14 | 0.65 | 0.119671 | 15621 |
| 870.2 | Open wound of ear | injuries & poisonings | DR | 1.57 | 1.17 | 2.11 | 0.122098 | 15621 |
| 750.22 | Congenital anomaly of gallbladder, bile ducts, liver, pancreas | congenital anomalies | DR | 1.44 | 1.14 | 1.82 | 0.122627 | 15621 |
| 870.8 | Open wound of genital organs | injuries & poisonings | DR | 1.42 | 1.13 | 1.77 | 0.122916 | 15621 |
| 560.4 | Other intestinal obstruction | digestive | DR | 1.17 | 1.06 | 1.29 | 0.123433 | 15621 |
| 720.1 | Spinal stenosis of lumbar region | musculoskeletal | DR | 1.08 | 1.03 | 1.14 | 0.124055 | 15621 |
| 227.1 | Benign neoplasm of adrenal gland | neoplasms | DR | 1.26 | 1.08 | 1.47 | 0.124115 | 15621 |
| 292.5 | Transient alteration of awareness | mental disorders | DR | 1.22 | 1.07 | 1.38 | 0.124853 | 15621 |
| 117.4 | Aspergillosis | infectious diseases | DR | 1.73 | 1.21 | 2.48 | 0.126508 | 15621 |
| 240 | Simple and unspecified goiter | endocrine/metabolic | DR | 1.36 | 1.11 | 1.67 | 0.126664 | 15621 |
| 694.3 | Vascular disorders of skin | dermatologic | DR | 1.53 | 1.16 | 2.02 | 0.127022 | 15621 |
| 818.1 | | | DR | 1.46 | 1.14 | 1.87 | 0.127953 | 15621 |
| 189.12 | Malignant neoplasm of renal pelvis | neoplasms | DR | 1.34 | 1.10 | 1.61 | 0.12841 | 15621 |
| 70 | Viral hepatitis | infectious diseases | DR | 1.09 | 1.03 | 1.15 | 0.129883 | 15621 |
| 568.1 | Peritoneal adhesions (postoperative) (postinfection) | digestive | DR | 1.22 | 1.07 | 1.39 | 0.129919 | 15621 |
| 327.31 | Central/nonobstroctive sleep apnea | neurological | DR | 1.16 | 1.05 | 1.28 | 0.130145 | 15621 |
| 609.2 | Abnormal spermatozoa | genitourinary | DR | 0.77 | 0.65 | 0.92 | 0.130425 | 15621 |
| 315.2 | Speech and language disorder | mental disorders | DR | 1.48 | 1.14 | 1.93 | 0.130633 | 15621 |
| 610.3 | Fibrosclerosis of breast | genitourinary | DR | 2.42 | 1.35 | 4.34 | 0.131929 | 15621 |
| 750.2 | Lower gastrointestinal congenital anomalies | congenital anomalies | DR | 1.32 | 1.10 | 1.58 | 0.132266 | 15621 |
| 736.5 | Acquired deformities of knee | musculoskeletal | DR | 0.63 | 0.46 | 0.86 | 0.134215 | 15621 |
| 663 | Umbilical cord complications during labor and delivery | pregnancy complications | DR | 5.43 | 1.75 | 16.82 | 0.134658 | 15621 |
| 696 | | | DR | 1.11 | 1.04 | 1.19 | 0.134803 | 15621 |
| 737.2 | Lordosis (acquired) | musculoskeletal | DR | 0.62 | 0.45 | 0.85 | 0.136554 | 15621 |
| 613.8 | Other specified disorders of breast | genitourinary | DR | 0.67 | 0.51 | 0.88 | 0.137707 | 15621 |
| 70.3 | Viral hepatitis C | infectious diseases | DR | 1.09 | 1.03 | 1.15 | 0.138026 | 15621 |
| 149.4 | Cancer of larynx | neoplasms | DR | 1.52 | 1.15 | 2.02 | 0.138045 | 15621 |
| 721 | Spondylosis and allied disorders | musculoskeletal | DR | 0.95 | 0.91 | 0.98 | 0.140077 | 15621 |
| 194 | Cancer of other endocrine glands | neoplasms | DR | 1.35 | 1.10 | 1.65 | 0.140164 | 15621 |
| 379.1 | Scleritis and episcleritis | sense organs | DR | 1.32 | 1.09 | 1.60 | 0.140615 | 15621 |
| 696.41 | Psoriasis vulgaris | dermatologic | DR | 1.11 | 1.03 | 1.20 | 0.141423 | 15621 |
| 278.4 | Abnormal weight gain | endocrine/metabolic | DR | 1.16 | 1.05 | 1.28 | 0.14143 | 15621 |
| 242.2 | Toxic multinodular goiter | endocrine/metabolic | DR | 0.47 | 0.28 | 0.79 | 0.142301 | 15621 |
| 317.11 | Alcoholic liver damage | mental disorders | DR | 0.90 | 0.85 | 0.97 | 0.143197 | 15621 |
| 772.4 | Rhabdomyolysis | symptoms | DR | 1.21 | 1.06 | 1.39 | 0.144674 | 15621 |
| 1010.4 | Genetic Test | | DR | 0.32 | 0.15 | 0.70 | 0.144993 | 15621 |
| 255 | Disorders of adrenal glands | endocrine/metabolic | DR | 1.16 | 1.05 | 1.29 | 0.145459 | 15621 |
| 701.5 | Abnormal granulation tissue | dermatologic | DR | 1.28 | 1.08 | 1.53 | 0.14688 | 15621 |
| 733.8 | Malunion and nonunion of fracture | musculoskeletal | DR | 1.26 | 1.07 | 1.47 | 0.147206 | 15621 |
| 527.8 | Other specified diseases of the salivary glands | digestive | DR | 1.43 | 1.12 | 1.83 | 0.148093 | 15621 |
| 272.14 | Hyperchylomicronemia | endocrine/metabolic | DR | 2.20 | 1.27 | 3.80 | 0.148619 | 15621 |

| | | | | | | | | |
|---|---|---|---|---|---|---|---|---|
| 150 | Cancer of esophagus | neoplasms | DR | 1.42 | 1.11 | 1.82 | 0.149115 | 15621 |
| 272.9 | Unspecified disorder of lipoid metabolism | endocrine/metabolic | DR | 1.16 | 1.05 | 1.28 | 0.150694 | 15621 |
| 315 | Develomental delays and disorders | mental disorders | DR | 1.17 | 1.05 | 1.30 | 0.151009 | 15621 |
| 504 | Other alveolar and parietoalveolar pneumonopathy | respiratory | DR | 1.20 | 1.06 | 1.36 | 0.151298 | 15621 |
| 1010.2 | screening for malignant neoplasms | | DR | 0.95 | 0.92 | 0.98 | 0.152031 | 15621 |
| 965.1 | Opiates and related narcotics causing adverse effects in therapeutic use | injuries & poisonings | DR | 1.23 | 1.06 | 1.42 | 0.152056 | 15621 |
| 751.3 | Obstructive genitourinary defect | congenital anomalies | DR | 0.45 | 0.25 | 0.78 | 0.153049 | 15621 |
| 755.6 | Other congenital anomalies of lower limb, including pelvic girdle | congenital anomalies | DR | 0.64 | 0.47 | 0.88 | 0.15352 | 15621 |
| 740.2 | Osteoarthrosis, generalized | musculoskeletal | DR | 1.09 | 1.03 | 1.16 | 0.153809 | 15621 |
| 701.3 | Circumscribed scleroderma | dermatologic | DR | 1.43 | 1.11 | 1.84 | 0.154055 | 15621 |
| 714.2 | Juvenile rheumatoid arthritis | musculoskeletal | DR | 0.39 | 0.20 | 0.76 | 0.155867 | 15621 |
| 369.2 | Eye infection, viral | sense organs | DR | 1.17 | 1.05 | 1.30 | 0.159723 | 15621 |
| 359.1 | Muscular dystrophies | neurological | DR | 1.63 | 1.15 | 2.30 | 0.159997 | 15621 |
| 350.5 | Abnormal reflex | neurological | DR | 0.59 | 0.40 | 0.86 | 0.161977 | 15621 |
| 766 | Neuralgia, neuritis, and radiculitis NOS | symptoms | DR | 1.08 | 1.02 | 1.15 | 0.163464 | 15621 |
| 278.3 | Localized adiposity | endocrine/metabolic | DR | 0.69 | 0.52 | 0.90 | 0.163578 | 15621 |
| 530.15 | Eosinophilic esophagitis | digestive | DR | 0.54 | 0.35 | 0.84 | 0.167661 | 15621 |
| 687.1 | Rash and other nonspecific skin eruption | dermatologic | DR | 1.05 | 1.01 | 1.09 | 0.167981 | 15621 |
| 743.12 | | | DR | 2.08 | 1.22 | 3.55 | 0.168038 | 15621 |
| 695.7 | Prurigo and Lichen | dermatologic | DR | 1.10 | 1.03 | 1.18 | 0.168175 | 15621 |
| 749 | Congenital anomalies of face and neck | congenital anomalies | DR | 1.32 | 1.08 | 1.61 | 0.16875 | 15621 |
| 170 | Cancer of bone and connective tissue | neoplasms | DR | 1.26 | 1.07 | 1.50 | 0.169296 | 15621 |
| 130.1 | Lyme disease | infectious diseases | DR | 0.51 | 0.31 | 0.83 | 0.170273 | 15621 |
| 345.11 | Generalized convulsive epilepsy | neurological | DR | 0.76 | 0.62 | 0.93 | 0.17099 | 15621 |
| 189.2 | Cancer of bladder | neoplasms | DR | 1.17 | 1.04 | 1.32 | 0.171204 | 15621 |
| 750 | Digestive congenital anomalies | congenital anomalies | DR | 1.21 | 1.05 | 1.38 | 0.171365 | 15621 |
| 170.1 | Bone cancer | neoplasms | DR | 1.41 | 1.09 | 1.81 | 0.173209 | 15621 |
| 530.14 | Reflux esophagitis | digestive | DR | 1.09 | 1.02 | 1.16 | 0.173273 | 15621 |
| 225 | Benign neoplasm of brain and other parts of nervous system | neoplasms | DR | 1.24 | 1.06 | 1.46 | 0.175261 | 15621 |
| 597.8 | Urethral hypermobility/ISD | genitourinary | DR | 0.42 | 0.22 | 0.80 | 0.17601 | 15621 |
| 930 | Allergic reaction to food | injuries & poisonings | DR | 0.81 | 0.69 | 0.95 | 0.178476 | 15621 |
| 706.2 | Sebaceous cyst | dermatologic | DR | 1.07 | 1.02 | 1.12 | 0.17952 | 15621 |
| 70.2 | Viral hepatitis B | infectious diseases | DR | 1.17 | 1.04 | 1.32 | 0.180299 | 15621 |
| 559 | Ileostomy status | digestive | DR | 1.37 | 1.08 | 1.74 | 0.181401 | 15621 |
| 279.1 | Immunity deficiency | endocrine/metabolic | DR | 1.34 | 1.08 | 1.68 | 0.181772 | 15621 |
| 302 | Sexual and gender identity disorders | mental disorders | DR | 0.90 | 0.83 | 0.97 | 0.182126 | 15621 |
| 327.1 | Hypersomnia | neurological | DR | 0.89 | 0.82 | 0.97 | 0.183282 | 15621 |
| 981 | Toxic effect of (non-ethyl) alcohol and petroleum and other solvents | injuries & poisonings | DR | 0.54 | 0.34 | 0.86 | 0.184066 | 15621 |
| 212 | Benign neoplasm of respiratory and intrathoracic organs | neoplasms | DR | 1.30 | 1.07 | 1.58 | 0.185623 | 15621 |
| 870.1 | Open wound or laceration of eye or eyelid | injuries & poisonings | DR | 1.21 | 1.05 | 1.39 | 0.187218 | 15621 |
| 285.8 | Hemoglobinuria | hematopoietic | DR | 2.01 | 1.18 | 3.41 | 0.18733 | 15621 |
| 841 | | | DR | 0.92 | 0.87 | 0.98 | 0.187633 | 15621 |
| 241.2 | Nontoxic multinodular goiter | endocrine/metabolic | DR | 1.21 | 1.05 | 1.39 | 0.188682 | 15621 |
| 132 | Infestation (lice, mites) | infectious diseases | DR | 1.15 | 1.03 | 1.29 | 0.18892 | 15621 |
| 214.1 | Lipoma of skin and subcutaneous tissue | neoplasms | DR | 0.90 | 0.83 | 0.97 | 0.189893 | 15621 |
| 614 | Inflammatory diseases of female pelvic organs | genitourinary | DR | 1.24 | 1.05 | 1.47 | 0.19076 | 15621 |
| 555.1 | Regional enteritis | digestive | DR | 0.76 | 0.61 | 0.94 | 0.19105 | 15621 |
| 736.1 | Acquired deformities of forearm | musculoskeletal | DR | 1.46 | 1.09 | 1.94 | 0.191724 | 15621 |
| 376 | Disorders of the orbit | sense organs | DR | 1.41 | 1.08 | 1.83 | 0.192349 | 15621 |
| 949 | Allergies, other | injuries & poisonings | DR | 0.93 | 0.87 | 0.98 | 0.193498 | 15621 |
| 701.2 | Scar conditions and fibrosis of skin | dermatologic | DR | 1.12 | 1.03 | 1.23 | 0.195644 | 15621 |
| 187.1 | Malignant neoplasm of unspecified male genital organ | neoplasms | DR | 0.64 | 0.45 | 0.90 | 0.196453 | 15621 |
| 110.2 | Dermatomycoses | infectious diseases | DR | 1.13 | 1.03 | 1.24 | 0.196793 | 15621 |
| 300.8 | Acute reaction to stress | mental disorders | DR | 0.89 | 0.82 | 0.97 | 0.197207 | 15621 |
| 594.3 | Calculus of ureter | genitourinary | DR | 0.90 | 0.82 | 0.98 | 0.198016 | 15621 |
| 619.3 | Noninflammatory disorders of cervix | genitourinary | DR | 0.55 | 0.35 | 0.88 | 0.198411 | 15621 |
| 331.1 | Hydrocephalus | neurological | DR | 1.37 | 1.07 | 1.76 | 0.19923 | 15621 |
| 478 | Throat pain | respiratory | DR | 0.89 | 0.81 | 0.97 | 0.199591 | 15621 |
| 244.2 | Acquired hypothyroidism | endocrine/metabolic | DR | 1.15 | 1.03 | 1.29 | 0.200401 | 15621 |
| 386.21 | Central origin vertigo | sense organs | DR | 1.24 | 1.05 | 1.47 | 0.200406 | 15621 |
| 259.3 | Delay in sexual development and puberty NEC | endocrine/metabolic | DR | 1.34 | 1.07 | 1.69 | 0.201599 | 15621 |
| 190 | Cancer of eye | neoplasms | DR | 1.46 | 1.08 | 1.96 | 0.202437 | 15621 |
| 444.2 | Embolism and thrombosis of abdominal aorta | circulatory system | DR | 1.70 | 1.12 | 2.59 | 0.202952 | 15621 |
| 836 | Traumatic arthropathy | injuries & poisonings | DR | 1.10 | 1.02 | 1.19 | 0.203293 | 15621 |
| 348.2 | Cerebral edema and compression of brain | neurological | DR | 1.30 | 1.06 | 1.60 | 0.205592 | 15621 |
| 962.1 | | | DR | 1.40 | 1.07 | 1.82 | 0.207373 | 15621 |
| 728.1 | Muscular calcification and ossification | musculoskeletal | DR | 1.51 | 1.09 | 2.10 | 0.210443 | 15621 |
| 316.1 | Polyneuropathy due to drugs | mental disorders | DR | 1.69 | 1.11 | 2.57 | 0.211526 | 15621 |
| 747.13 | Congenital anomalies of great vessels | congenital anomalies | DR | 1.47 | 1.08 | 2.00 | 0.212241 | 15621 |
| 333.8 | Other degenerative diseases of the basal ganglia | neurological | DR | 1.57 | 1.09 | 2.27 | 0.213557 | 15621 |
| 527.1 | Hypertrophy of salivary gland | digestive | DR | 1.64 | 1.10 | 2.45 | 0.21379 | 15621 |
| 465.2 | Acute pharyngitis | respiratory | DR | 0.94 | 0.90 | 0.99 | 0.213927 | 15621 |
| 8.7 | Intestinal infection due to protozoa | infectious diseases | DR | 1.60 | 1.10 | 2.32 | 0.214121 | 15621 |
| 213 | Benign neoplasm of bone and articular cartilage | neoplasms | DR | 1.29 | 1.05 | 1.57 | 0.214506 | 15621 |
| 313.2 | Tics and stuttering | mental disorders | DR | 1.34 | 1.06 | 1.69 | 0.215639 | 15621 |
| 592.2 | Urethritis and urethral syndrome | genitourinary | DR | 0.81 | 0.69 | 0.96 | 0.216629 | 15621 |
| 214 | Lipoma | neoplasms | DR | 0.93 | 0.87 | 0.99 | 0.216989 | 15621 |
| 769 | Nonallopathic lesions NEC | symptoms | DR | 0.89 | 0.81 | 0.98 | 0.217176 | 15621 |
| 217 | Vascular hamartomas and non-neoplastic nevi | neoplasms | DR | 1.09 | 1.02 | 1.17 | 0.218121 | 15621 |
| 327.6 | Circadian rhythm sleep disorder | neurological | DR | 0.81 | 0.69 | 0.96 | 0.218738 | 15621 |
| 286.13 | Congenital factor VIII disorder | hematopoietic | DR | 0.26 | 0.09 | 0.79 | 0.222433 | 15621 |
| 695.1 | Toxic erythema | dermatologic | DR | 1.55 | 1.08 | 2.23 | 0.223106 | 15621 |
| 202 | Cancer of other lymphoid, histiocytic tissue | neoplasms | DR | 1.17 | 1.03 | 1.33 | 0.224158 | 15621 |
| 696.3 | Pityriasis | dermatologic | DR | 0.68 | 0.50 | 0.94 | 0.225033 | 15621 |
| 604.3 | Peyronie's disease | genitourinary | DR | 0.82 | 0.69 | 0.97 | 0.227979 | 15621 |
| 696.42 | Psoriatic arthropathy | dermatologic | DR | 1.21 | 1.03 | 1.41 | 0.228193 | 15621 |
| 260.22 | Nutritional marasmus | endocrine/metabolic | DR | 1.83 | 1.11 | 3.03 | 0.22858 | 15621 |
| 614.33 | Pelvic inflammatory disease, NOS | genitourinary | DR | 1.94 | 1.12 | 3.37 | 0.228682 | 15621 |
| 842 | | | DR | 1.09 | 1.01 | 1.17 | 0.230806 | 15621 |
| 732.7 | Osteochondritis dissecans | musculoskeletal | DR | 0.72 | 0.54 | 0.95 | 0.231449 | 15621 |
| 535.2 | Atrophic gastritis | digestive | DR | 1.11 | 1.02 | 1.21 | 0.232356 | 15621 |
| 592.13 | Chronic interstitial cystitis | genitourinary | DR | 0.63 | 0.42 | 0.93 | 0.23696 | 15621 |
| 145.4 | Cancer of the gums | neoplasms | DR | 2.40 | 1.14 | 5.03 | 0.23755 | 15621 |
| 303.3 | Psychogenic disorder | mental disorders | DR | 0.84 | 0.73 | 0.97 | 0.238328 | 15621 |
| 627.4 | Premenopausal menorrhagia | genitourinary | DR | 0.55 | 0.34 | 0.91 | 0.238678 | 15621 |

| | | | | | | | | |
|---|---|---|---|---|---|---|---|---|
| 961.1 | Poisoning/allergy of sulfonamides | injuries & poisonings | DR | 1.39 | 1.05 | 1.84 | 0.240454 | 15621 |
| 618.5 | Prolapse of vaginal vault after hysterectomy | genitourinary | DR | 2.10 | 1.11 | 3.98 | 0.24232 | 15621 |
| 637 | Short gestation; low birth weight; and fetal growth retardation | pregnancy complications | DR | 0.28 | 0.10 | 0.83 | 0.242467 | 15621 |
| 721.8 | Other allied disorders of spine | musculoskeletal | DR | 1.17 | 1.02 | 1.33 | 0.24378 | 15621 |
| 528.41 | Cyst of the salivary gland | digestive | DR | 0.60 | 0.39 | 0.93 | 0.243826 | 15621 |
| 790.8 | Elevated C-reactive protein (CRP) | symptoms | DR | 1.42 | 1.05 | 1.93 | 0.250945 | 15621 |
| 426.4 | Anomalous atrioventricular excitation | circulatory system | DR | 0.57 | 0.35 | 0.93 | 0.252455 | 15621 |
| 622.2 | Mucous polyp of cervix | genitourinary | DR | 1.74 | 1.07 | 2.82 | 0.252591 | 15621 |
| 742.8 | Articular cartilage disorder | musculoskeletal | DR | 0.78 | 0.62 | 0.97 | 0.252643 | 15621 |
| 264.2 | Failure to thrive (childhood) | endocrine/metabolic | DR | 1.63 | 1.06 | 2.49 | 0.25396 | 15621 |
| 260.7 | Polyphagia | endocrine/metabolic | DR | 1.78 | 1.07 | 2.95 | 0.254581 | 15621 |
| 635 | Hemorrhage during pregnancy; childbirth and postpartum | pregnancy complications | DR | 4.11 | 1.19 | 14.20 | 0.255206 | 15621 |
| 635.2 | Antepartum hemorrhage, abruptio placentae, and placenta previa | pregnancy complications | DR | 4.11 | 1.19 | 14.20 | 0.255206 | 15621 |
| 617 | Disorders secondary to childbirth, surgery, trauma | genitourinary | DR | 0.63 | 0.42 | 0.95 | 0.255298 | 15621 |
| 913 | Toxic effect of venom | injuries & poisonings | DR | 0.81 | 0.67 | 0.98 | 0.257386 | 15621 |
| 353 | Nerve root and plexus disorders | neurological | DR | 1.09 | 1.01 | 1.19 | 0.259257 | 15621 |
| 586.1 | Anatomical abnormatilies of kidney and ureters | genitourinary | DR | 1.75 | 1.06 | 2.88 | 0.259977 | 15621 |
| 626.8 | Infertility, female | genitourinary | DR | 0.68 | 0.48 | 0.96 | 0.261282 | 15621 |
| 530.7 | Gastroesophageal laceration-hemorrhage syndrome | digestive | DR | 0.73 | 0.55 | 0.97 | 0.262425 | 15621 |
| 733.6 | | | DR | 1.15 | 1.02 | 1.31 | 0.263535 | 15621 |
| 338.2 | Chronic pain | neurological | DR | 0.96 | 0.92 | 1.00 | 0.263614 | 15621 |
| 519.1 | Tracheostomy complications | respiratory | DR | 1.54 | 1.05 | 2.27 | 0.264302 | 15621 |
| 742.1 | Loose body in joint | musculoskeletal | DR | 1.38 | 1.03 | 1.84 | 0.265221 | 15621 |
| 338 | | | DR | 1.04 | 1.00 | 1.08 | 0.267647 | 15621 |
| 749.1 | Cleft palate | congenital anomalies | DR | 0.29 | 0.09 | 0.89 | 0.268134 | 15621 |
| 742 | Derangement of joint, non-traumatic | musculoskeletal | DR | 0.93 | 0.88 | 0.99 | 0.268321 | 15621 |
| 676 | Other disorders of the breast associated with childbirth and disorders of lactation | pregnancy complications | DR | 0.30 | 0.10 | 0.89 | 0.268814 | 15621 |
| 619 | Noninflammatory female genital disorders | genitourinary | DR | 1.20 | 1.02 | 1.43 | 0.276857 | 15621 |
| 500.2 | Pneumoconiosis | respiratory | DR | 0.71 | 0.52 | 0.97 | 0.27689 | 15621 |
| 180.1 | Cervical cancer | neoplasms | DR | 0.43 | 0.20 | 0.94 | 0.279 | 15621 |
| 647.3 | Major puerperal infection | pregnancy complications | DR | 3.78 | 1.10 | 12.91 | 0.279699 | 15621 |
| 722.8 | Postlaminectomy syndrome | musculoskeletal | DR | 0.88 | 0.78 | 0.99 | 0.28108 | 15621 |
| 529.6 | Glossodynia | digestive | DR | 0.67 | 0.47 | 0.97 | 0.281124 | 15621 |
| 504.1 | Idiopathic fibrosing alveolitis | respiratory | DR | 1.20 | 1.01 | 1.42 | 0.282099 | 15621 |
| 452.8 | Postphlebitic syndrome | circulatory system | DR | 0.53 | 0.29 | 0.96 | 0.283159 | 15621 |
| 735.23 | Hallux rigidus | musculoskeletal | DR | 1.11 | 1.01 | 1.22 | 0.284817 | 15621 |
| 627.21 | | | DR | 1.65 | 1.03 | 2.64 | 0.284998 | 15621 |
| 743.13 | | | DR | 1.45 | 1.02 | 2.07 | 0.285233 | 15621 |
| 277.7 | Dysmetabolic syndrome X | endocrine/metabolic | DR | 0.91 | 0.84 | 0.99 | 0.285468 | 15621 |
| 724.1 | Disorders of sacrum | musculoskeletal | DR | 0.88 | 0.78 | 0.99 | 0.286149 | 15621 |
| 751.22 | Other specified congenital anomalies of kidney | congenital anomalies | DR | 0.74 | 0.55 | 0.98 | 0.288358 | 15621 |
| 612.2 | Hypertrophy of breast (Gynecomastia) | genitourinary | DR | 0.87 | 0.76 | 0.99 | 0.289597 | 15621 |
| 1010.5 | potential health hazards related to communicable diseases potential health hazards related to communicable diseases | | DR | 1.05 | 1.00 | 1.09 | 0.291327 | 15621 |
| 70.1 | Viral hepatitis A | infectious diseases | DR | 0.78 | 0.62 | 0.99 | 0.291415 | 15621 |
| 261.3 | Vitamin C deficiencies | endocrine/metabolic | DR | 1.90 | 1.03 | 3.49 | 0.291904 | 15621 |
| 727.5 | Rupture of synovium | musculoskeletal | DR | 0.86 | 0.75 | 0.99 | 0.293697 | 15621 |
| 728 | Disorders of muscle, ligament, and fascia | musculoskeletal | DR | 1.15 | 1.01 | 1.30 | 0.293828 | 15621 |
| 81.1 | Graft-versus-host disease | infectious diseases | DR | 3.38 | 1.06 | 10.76 | 0.293989 | 15621 |
| 78 | Viral warts & HPV | infectious diseases | DR | 0.94 | 0.89 | 1.00 | 0.29427 | 15621 |
| 145.1 | Cancer of lip | neoplasms | DR | 1.91 | 1.03 | 3.54 | 0.29454 | 15621 |
| 709.6 | Other specified diffuse diseases of connective tissue | dermatologic | DR | 1.96 | 1.03 | 3.72 | 0.294906 | 15621 |
| 535.1 | Acute gastritis | digestive | DR | 1.10 | 1.00 | 1.20 | 0.295614 | 15621 |
| 573.6 | Nonspecific elevation of levels of transaminase or lactic acid dehydrogenase [LDH] | digestive | DR | 0.93 | 0.88 | 1.00 | 0.29762 | 15621 |
| 225.1 | Benign neoplasm of brain, cranial nerves, meninges | neoplasms | DR | 1.19 | 1.01 | 1.40 | 0.298562 | 15621 |
| 780 | Hypothermia/Chills | symptoms | DR | 1.17 | 1.01 | 1.36 | 0.29983 | 15621 |
| 695.81 | Erythema nodosum | dermatologic | DR | 0.50 | 0.25 | 0.98 | 0.300671 | 15621 |
| 149.2 | Cancer of nasopharynx | neoplasms | DR | 0.43 | 0.19 | 0.97 | 0.300987 | 15621 |
| 526.4 | Temporomandibular joint disorders | digestive | DR | 1.08 | 1.00 | 1.17 | 0.302483 | 15621 |
| 452.1 | | | DR | 0.60 | 0.37 | 0.99 | 0.304189 | 15621 |
| 751 | Genitourinary congenital anomalies | congenital anomalies | DR | 1.08 | 1.00 | 1.16 | 0.304429 | 15621 |
| 669 | Complications of labor and delivery NEC | pregnancy complications | DR | 0.32 | 0.11 | 0.97 | 0.304723 | 15621 |
| 79.9 | Viremia, NOS | infectious diseases | DR | 0.88 | 0.78 | 1.00 | 0.305371 | 15621 |
| 772.2 | Spasm of muscle | symptoms | DR | 1.05 | 1.00 | 1.11 | 0.306003 | 15621 |
| 244.3 | | | DR | 1.93 | 1.02 | 3.68 | 0.306144 | 15621 |
| 509.5 | Respiratory arrest | respiratory | DR | 1.42 | 1.01 | 2.00 | 0.306339 | 15621 |
| 260.21 | Kwashiorkor | endocrine/metabolic | DR | 0.32 | 0.11 | 0.98 | 0.307306 | 15621 |
| 751.12 | Congenital anomalies of male genital organs | congenital anomalies | DR | 1.22 | 1.00 | 1.48 | 0.307354 | 15621 |
| 764 | Sciatica | symptoms | DR | 1.05 | 1.00 | 1.10 | 0.308843 | 15621 |
| 291.8 | Alteration of consciousness | mental disorders | DR | 1.10 | 1.00 | 1.20 | 0.309383 | 15621 |
| 603.2 | Spermatocele | genitourinary | DR | 0.86 | 0.74 | 1.00 | 0.310282 | 15621 |
| 228.1 | Hemangioma of skin and subcutaneous tissue | neoplasms | DR | 1.12 | 1.00 | 1.24 | 0.311015 | 15621 |
| 253.1 | Pituitary hyperfunction | endocrine/metabolic | DR | 0.80 | 0.64 | 1.00 | 0.311191 | 15621 |
| 705.3 | Hidradenitis | dermatologic | DR | 1.17 | 1.00 | 1.37 | 0.311686 | 15621 |
| 364.41 | Keratoconus | sense organs | DR | 0.75 | 0.56 | 1.00 | 0.312596 | 15621 |
| 599.7 | Urethral discharge | genitourinary | DR | 1.31 | 1.00 | 1.72 | 0.313155 | 15621 |
| 151 | Cancer of stomach | neoplasms | DR | 1.22 | 1.00 | 1.49 | 0.316455 | 15621 |
| 187.8 | Neoplasm of uncertain behavior of male genital organs | neoplasms | DR | 1.28 | 1.00 | 1.63 | 0.318573 | 15621 |
| 965.2 | | | DR | 1.45 | 1.00 | 2.10 | 0.318774 | 15621 |
| 695.2 | Bullous dermatoses | dermatologic | DR | 1.25 | 1.00 | 1.58 | 0.320683 | 15621 |
| 628 | Ovarian cyst | genitourinary | DR | 0.80 | 0.65 | 1.00 | 0.322244 | 15621 |
| 714 | Rheumatoid arthritis and other inflammatory polyarthropathies | musculoskeletal | DR | 1.08 | 1.00 | 1.16 | 0.322627 | 15621 |
| 938.2 | Chronic dermatitis due to solar radiation | dermatologic | DR | 1.14 | 1.00 | 1.30 | 0.322697 | 15621 |
| 381.11 | Suppurative and unspecified otitis media | sense organs | DR | 1.05 | 1.00 | 1.11 | 0.323916 | 15621 |
| 253.7 | Other disorders of neurohypophysis | endocrine/metabolic | DR | 1.25 | 1.00 | 1.57 | 0.32422 | 15621 |
| 220 | Benign neoplasm of ovary | neoplasms | DR | 1.78 | 0.99 | 3.20 | 0.328033 | 15621 |
| 315.1 | Learning disorder | mental disorders | DR | 0.68 | 0.46 | 1.01 | 0.328063 | 15621 |
| 528.11 | Stomatitis and mucositis (ulcerative) | digestive | DR | 1.32 | 0.99 | 1.76 | 0.328775 | 15621 |
| 686.4 | Pyogenic granuloma | dermatologic | DR | 1.20 | 1.00 | 1.45 | 0.330037 | 15621 |
| 598.4 | | | DR | 1.33 | 0.99 | 1.78 | 0.330553 | 15621 |
| 622 | Polyp of female genital organs | genitourinary | DR | 1.38 | 0.99 | 1.91 | 0.332293 | 15621 |
| 323.8 | Encephalitis, non-infectious | neurological | DR | 1.35 | 0.99 | 1.85 | 0.334222 | 15621 |
| 270.34 | Alpha-1-antitrypsin deficiency | endocrine/metabolic | DR | 0.55 | 0.29 | 1.02 | 0.334848 | 15621 |
| 253.2 | Pituitary hypofunction | endocrine/metabolic | DR | 0.78 | 0.60 | 1.01 | 0.335187 | 15621 |
| 305.2 | Eating disorder | mental disorders | DR | 0.82 | 0.67 | 1.01 | 0.335926 | 15621 |

| | | | | | | | | |
|---|---|---|---|---|---|---|---|---|
| 295 | Schizophrenia and other psychotic disorders | mental disorders | DR | 0.95 | 0.90 | 1.00 | 0.336201 | 15621 |
| 656.4 | Hemorrhage of fetus or newborn | pregnancy complications | DR | 2.09 | 0.97 | 4.52 | 0.336514 | 15621 |
| 264 | | | DR | 1.47 | 0.98 | 2.20 | 0.337279 | 15621 |
| 695.22 | Pemphigus and pemphigoid | dermatologic | DR | 1.39 | 0.98 | 1.95 | 0.33999 | 15621 |
| 41.21 | Rheumatic fever / chorea | infectious diseases | DR | 0.56 | 0.30 | 1.03 | 0.340509 | 15621 |
| 957 | | | DR | 0.56 | 0.30 | 1.03 | 0.341658 | 15621 |
| 425.12 | Other hypertrophic cardiomyopathy | circulatory system | DR | 0.73 | 0.52 | 1.02 | 0.341884 | 15621 |
| 579.2 | Splenomegaly | digestive | DR | 1.11 | 0.99 | 1.23 | 0.34251 | 15621 |
| 334 | Degenerative disease of the spinal cord | neurological | DR | 1.11 | 0.99 | 1.25 | 0.343348 | 15621 |
| 716.1 | | | DR | 1.20 | 0.99 | 1.47 | 0.343819 | 15621 |
| 716.3 | Kaschin-Beck disease | musculoskeletal | DR | 2.28 | 0.95 | 5.45 | 0.344248 | 15621 |
| 346.2 | Nonspecific abnormal results of function study of brain and central nervous system | neurological | DR | 1.17 | 0.99 | 1.39 | 0.34558 | 15621 |
| 132.1 | Pediculosis and phthirus infestation | infectious diseases | DR | 1.49 | 0.98 | 2.27 | 0.346838 | 15621 |
| 709.3 | Systemic sclerosis | dermatologic | DR | 1.57 | 0.97 | 2.56 | 0.35043 | 15621 |
| 10 | Tuberculosis | infectious diseases | DR | 1.12 | 0.99 | 1.27 | 0.350846 | 15621 |
| 225.2 | Benign neoplasm of spinal cord, meninges | neoplasms | DR | 1.78 | 0.96 | 3.29 | 0.351292 | 15621 |
| 415.1 | Acute pulmonary heart disease | circulatory system | DR | 1.10 | 0.99 | 1.22 | 0.351729 | 15621 |
| 634 | Miscarriage; stillbirth | pregnancy complications | DR | 0.72 | 0.50 | 1.03 | 0.352845 | 15621 |
| 560.3 | Peritoneal or intestinal adhesions | digestive | DR | 1.26 | 0.98 | 1.63 | 0.354597 | 15621 |
| 198.1 | Secondary malignancy of lymph nodes | neoplasms | DR | 1.13 | 0.99 | 1.30 | 0.355377 | 15621 |
| 665 | Obstetrical/birth trauma | pregnancy complications | DR | 0.65 | 0.41 | 1.04 | 0.355805 | 15621 |
| 512.3 | | | DR | 1.32 | 0.98 | 1.78 | 0.356344 | 15621 |
| 371.21 | Allergic conjunctivitis | sense organs | DR | 1.06 | 1.00 | 1.13 | 0.356686 | 15621 |
| 634.3 | Ectopic pregnancy | pregnancy complications | DR | 0.36 | 0.12 | 1.09 | 0.358113 | 15621 |
| 626.4 | Premenstrual tension syndromes | genitourinary | DR | 0.48 | 0.21 | 1.07 | 0.358545 | 15621 |
| 989 | Toxic effect of other substances, chiefly nonmedicinal as to source | injuries & poisonings | DR | 0.82 | 0.67 | 1.02 | 0.358607 | 15621 |
| 749.2 | Congenital anomalies of skull and face bones | congenital anomalies | DR | 2.04 | 0.94 | 4.45 | 0.360082 | 15621 |
| 735.22 | Claw toe (acquired) | musculoskeletal | DR | 1.36 | 0.97 | 1.90 | 0.363032 | 15621 |
| 272.12 | Hyperglyceridemia | endocrine/metabolic | DR | 0.96 | 0.91 | 1.00 | 0.365253 | 15621 |
| 217.1 | Nevus, non-neoplastic | neoplasms | DR | 1.07 | 0.99 | 1.14 | 0.366754 | 15621 |
| 383 | Otosclerosis | sense organs | DR | 0.76 | 0.56 | 1.03 | 0.366966 | 15621 |
| 420.21 | Acute pericarditis | circulatory system | DR | 1.25 | 0.98 | 1.60 | 0.368219 | 15621 |
| 530.13 | Barrett's esophagus | digestive | DR | 1.09 | 0.99 | 1.20 | 0.369339 | 15621 |
| 767 | | | DR | 1.32 | 0.97 | 1.80 | 0.369735 | 15621 |
| 282 | | | DR | 1.16 | 0.98 | 1.36 | 0.369828 | 15621 |
| 204.21 | Myeloid leukemia, acute | neoplasms | DR | 1.38 | 0.96 | 1.99 | 0.370021 | 15621 |
| 277.51 | Lipoprotein disorders | endocrine/metabolic | DR | 1.14 | 0.98 | 1.32 | 0.370427 | 15621 |
| 353.2 | Nerve root lesions | neurological | DR | 0.91 | 0.82 | 1.01 | 0.373042 | 15621 |
| 949.1 | Diaper or napkin rash | injuries & poisonings | DR | 1.34 | 0.96 | 1.87 | 0.374261 | 15621 |
| 174.3 | Neoplasm of uncertain behavior of breast | neoplasms | DR | 0.61 | 0.35 | 1.06 | 0.374354 | 15621 |
| 315.3 | Mental retardation | mental disorders | DR | 0.67 | 0.42 | 1.05 | 0.374702 | 15621 |
| 721.2 | Spondylosis with myelopathy | musculoskeletal | DR | 1.09 | 0.99 | 1.20 | 0.375051 | 15621 |
| 946 | Anaphylactic shock NOS | injuries & poisonings | DR | 1.13 | 0.98 | 1.29 | 0.375695 | 15621 |
| 624.2 | Atrophy of female genital tract | genitourinary | DR | 2.12 | 0.91 | 4.98 | 0.377124 | 15621 |
| 358 | Myoneural disorders | neurological | DR | 1.24 | 0.97 | 1.59 | 0.377876 | 15621 |
| 425.11 | Hypertrophic obstructive cardiomyopathy | circulatory system | DR | 1.31 | 0.96 | 1.79 | 0.377934 | 15621 |
| 251.8 | Abnormality of secretion of glucagon or gastrin | endocrine/metabolic | DR | 2.28 | 0.89 | 5.83 | 0.381866 | 15621 |
| 795.82 | Elevated cancer antigen 125 [CA 125] | symptoms | DR | 2.49 | 0.88 | 7.06 | 0.382396 | 15621 |
| 279.2 | Autoimmune disease NEC | endocrine/metabolic | DR | 1.50 | 0.94 | 2.38 | 0.382823 | 15621 |
| 184.1 | Malignant neoplasm of ovary and other uterine adnexa | neoplasms | DR | 1.72 | 0.92 | 3.19 | 0.382988 | 15621 |
| 1006 | Crushing injury | NULL | DR | 1.15 | 0.98 | 1.35 | 0.383163 | 15621 |
| 612 | Breast conditions, congenital or relating to hormones | genitourinary | DR | 0.90 | 0.79 | 1.02 | 0.385046 | 15621 |
| 355 | Complex regional/central pain syndrome | neurological | DR | 1.13 | 0.98 | 1.29 | 0.385138 | 15621 |
| 528.7 | Sialolithiasis | digestive | DR | 1.28 | 0.96 | 1.69 | 0.385483 | 15621 |
| 300.3 | Obsessive-compulsive disorders | mental disorders | DR | 1.13 | 0.98 | 1.29 | 0.387105 | 15621 |
| 31 | Diseases due to other mycobacteria | infectious diseases | DR | 1.27 | 0.96 | 1.69 | 0.388992 | 15621 |
| 199.4 | Neurofibromatosis | neoplasms | DR | 0.66 | 0.40 | 1.07 | 0.389474 | 15621 |
| 550.1 | Inguinal hernia | digestive | DR | 0.95 | 0.89 | 1.01 | 0.390063 | 15621 |
| 715.3 | Spinal enthesopathy | musculoskeletal | DR | 1.29 | 0.96 | 1.75 | 0.391542 | 15621 |
| 976 | Poisoning by agents primarily affecting skin & mucous membrane, ophthalmological, otorhinolaryngological, & dental drugs | injuries & poisonings | DR | 1.47 | 0.94 | 2.31 | 0.392517 | 15621 |
| 740.11 | Osteoarthrosis, localized, primary | musculoskeletal | DR | 1.03 | 0.99 | 1.07 | 0.392814 | 15621 |
| 193 | Thyroid cancer | neoplasms | DR | 0.84 | 0.68 | 1.03 | 0.39688 | 15621 |
| 614.4 | Inflammatory diseases of uterus, except cervix | genitourinary | DR | 2.19 | 0.87 | 5.52 | 0.397316 | 15621 |
| 255.11 | Cushing's syndrome | endocrine/metabolic | DR | 0.74 | 0.52 | 1.06 | 0.403476 | 15621 |
| 752.11 | Spina bifida | congenital anomalies | DR | 0.57 | 0.29 | 1.12 | 0.40363 | 15621 |
| 198.3 | Secondary malignant neoplasm of digestive systems | neoplasms | DR | 1.22 | 0.96 | 1.54 | 0.406382 | 15621 |
| 732.1 | Juvenile osteochondrosis | musculoskeletal | DR | 0.78 | 0.58 | 1.05 | 0.406674 | 15621 |
| 759.1 | Anomalies of endocrine glands, congenital | congenital anomalies | DR | 1.76 | 0.89 | 3.47 | 0.408989 | 15621 |
| 528.12 | Oral aphthae | digestive | DR | 0.86 | 0.72 | 1.03 | 0.408992 | 15621 |
| 696.2 | Parapsoriasis | dermatologic | DR | 1.33 | 0.94 | 1.88 | 0.41046 | 15621 |
| 529 | Diseases and other conditions of the tongue | digestive | DR | 1.12 | 0.98 | 1.28 | 0.410653 | 15621 |
| 722.9 | Other and unspecified disc disorder | musculoskeletal | DR | 0.94 | 0.87 | 1.01 | 0.411668 | 15621 |
| 626.14 | Irregular menstrual bleeding | genitourinary | DR | 0.76 | 0.54 | 1.06 | 0.411671 | 15621 |
| 620 | Dysplasia of female genital organs | genitourinary | DR | 0.65 | 0.39 | 1.10 | 0.411771 | 15621 |
| 175 | Acquired absence of breast | neoplasms | DR | 0.63 | 0.36 | 1.10 | 0.411986 | 15621 |
| 528.6 | Leukoplakia of oral mucosa | digestive | DR | 1.16 | 0.97 | 1.39 | 0.412302 | 15621 |
| 592.12 | Chronic cystitis | genitourinary | DR | 1.17 | 0.97 | 1.41 | 0.413873 | 15621 |
| 755.61 | Congenital hip dysplasia and deformity | congenital anomalies | DR | 0.36 | 0.10 | 1.26 | 0.414928 | 15621 |
| 714.1 | Rheumatoid arthritis | musculoskeletal | DR | 1.08 | 0.98 | 1.18 | 0.417145 | 15621 |
| 621 | Endometrial hyperplasia | genitourinary | DR | 0.70 | 0.45 | 1.09 | 0.419866 | 15621 |
| 709.7 | Unspecified diffuse connective tissue disease | dermatologic | DR | 0.75 | 0.52 | 1.07 | 0.424021 | 15621 |
| 289.9 | Abnormality of red blood cells | hematopoietic | DR | 1.14 | 0.97 | 1.34 | 0.424794 | 15621 |
| 748 | Anomalies of respiratory system, congenital | congenital anomalies | DR | 0.65 | 0.38 | 1.11 | 0.4251 | 15621 |
| 705.8 | Hyperhidrosis | dermatologic | DR | 1.09 | 0.98 | 1.22 | 0.425873 | 15621 |
| 983 | Toxic effect of corrosive aromatics, acids, and caustic alkalis | injuries & poisonings | DR | 0.74 | 0.50 | 1.08 | 0.426059 | 15621 |
| 371.33 | Noninfectious dermatoses of eyelid | sense organs | DR | 0.67 | 0.41 | 1.11 | 0.426561 | 15621 |
| 638 | Other high-risk pregnancy | pregnancy complications | DR | 0.69 | 0.44 | 1.10 | 0.427921 | 15621 |
| 227 | Benign neoplasm of other endocrine glands and related structures | neoplasms | DR | 1.09 | 0.98 | 1.23 | 0.430317 | 15621 |
| 655.1 | Abnormality in fetal heart rate or rhythm | pregnancy complications | DR | 0.41 | 0.13 | 1.27 | 0.430642 | 15621 |
| 608 | Other disorders of male genital organs | genitourinary | DR | 0.95 | 0.90 | 1.01 | 0.431974 | 15621 |
| 277.2 | Other disorders of purine and pyrimidine metabolism | endocrine/metabolic | DR | 1.86 | 0.84 | 4.09 | 0.432475 | 15621 |
| 716.2 | Unspecified monoarthritis | musculoskeletal | DR | 1.12 | 0.97 | 1.29 | 0.433582 | 15621 |
| 134 | Helminthiases | infectious diseases | DR | 1.21 | 0.95 | 1.55 | 0.436098 | 15621 |
| 275.1 | Disorders of iron metabolism | hematopoietic | DR | 1.12 | 0.97 | 1.30 | 0.436184 | 15621 |
| 385.5 | Tympanosclerosis and middle ear disease related to otitis media | sense organs | DR | 1.27 | 0.93 | 1.73 | 0.436523 | 15621 |

| | | | | | | | | |
|---|---|---|---|---|---|---|---|---|
| 384 | Other disorders of tympanic membrane | sense organs | DR | 1.09 | 0.98 | 1.22 | 0.437194 | 15621 |
| 526.5 | Inflammatory conditions of jaw | digestive | DR | 0.88 | 0.74 | 1.04 | 0.437933 | 15621 |
| 625.1 | Dyspareunia | genitourinary | DR | 1.27 | 0.93 | 1.73 | 0.43902 | 15621 |
| 246.2 | | | DR | 0.74 | 0.51 | 1.09 | 0.44075 | 15621 |
| 500 | Lung disease due to external agents | respiratory | DR | 0.87 | 0.73 | 1.04 | 0.440967 | 15621 |
| 164 | Cancer of intrathoracic organs | neoplasms | DR | 0.66 | 0.38 | 1.13 | 0.441342 | 15621 |
| 202.23 | Lymphosarcoma | neoplasms | DR | 0.63 | 0.35 | 1.15 | 0.442671 | 15621 |
| 741.6 | Villonodular synovitis | musculoskeletal | DR | 0.70 | 0.44 | 1.11 | 0.442918 | 15621 |
| 594.2 | Calculus of lower urinary tract | genitourinary | DR | 0.89 | 0.77 | 1.04 | 0.445374 | 15621 |
| 642.1 | Preeclampsia and eclampsia | pregnancy complications | DR | 2.57 | 0.74 | 8.88 | 0.446682 | 15621 |
| 371.2 | Conjunctivitis, noninfectious | sense organs | DR | 1.05 | 0.99 | 1.11 | 0.448287 | 15621 |
| 362.1 | Retinopathy of prematurity | sense organs | DR | 1.63 | 0.85 | 3.13 | 0.448922 | 15621 |
| 526.1 | Cysts of the jaws | digestive | DR | 0.76 | 0.54 | 1.09 | 0.449763 | 15621 |
| 526.3 | Anomalies of jaw size/symmetry | digestive | DR | 0.77 | 0.55 | 1.09 | 0.449921 | 15621 |
| 686.3 | Pilonidal cyst | dermatologic | DR | 0.86 | 0.70 | 1.05 | 0.450018 | 15621 |
| 740.3 | Osteoarthrosis involving more than one site, but not specified as generalized | musculoskeletal | DR | 1.09 | 0.97 | 1.22 | 0.452013 | 15621 |
| 853 | Complication of colostomy or enterostomy | injuries & poisonings | DR | 1.24 | 0.93 | 1.64 | 0.452451 | 15621 |
| 204.2 | Myeloid leukemia | neoplasms | DR | 1.21 | 0.94 | 1.57 | 0.455384 | 15621 |
| 270.33 | Amyloidosis | endocrine/metabolic | DR | 1.23 | 0.93 | 1.61 | 0.455906 | 15621 |
| 614.31 | Acute inflammatory pelvic disease | genitourinary | DR | 0.43 | 0.14 | 1.34 | 0.45777 | 15621 |
| 654 | Other and unspecified complications of birth; puerperium affecting management of mother | pregnancy complications | DR | 0.54 | 0.24 | 1.24 | 0.458081 | 15621 |
| 759 | Other and unspecified congenital anomalies | congenital anomalies | DR | 1.22 | 0.93 | 1.59 | 0.458236 | 15621 |
| 623 | Hypertrophy of female genital organs | genitourinary | DR | 1.36 | 0.90 | 2.07 | 0.458955 | 15621 |
| 594 | Urinary calculus | genitourinary | DR | 0.96 | 0.92 | 1.01 | 0.459503 | 15621 |
| 475.9 | Postnasal drip | respiratory | DR | 1.11 | 0.97 | 1.26 | 0.459629 | 15621 |
| 227.3 | Benign neoplasm of pituitary gland and craniopharyngeal duct (pouch) | neoplasms | DR | 0.87 | 0.72 | 1.05 | 0.460101 | 15621 |
| 346.1 | Nonspecific abnormal findings on radiological and other examination of skull and head | neurological | DR | 1.10 | 0.97 | 1.25 | 0.464684 | 15621 |
| 499 | Cystic fibrosis | respiratory | DR | 2.53 | 0.71 | 9.00 | 0.465031 | 15621 |
| 737 | Curvature of spine | musculoskeletal | DR | 0.92 | 0.82 | 1.03 | 0.466724 | 15621 |
| 594.1 | Calculus of kidney | genitourinary | DR | 0.96 | 0.91 | 1.01 | 0.467101 | 15621 |
| 795.81 | Elevated carcinoembryonic antigen [CEA] | symptoms | DR | 1.49 | 0.86 | 2.59 | 0.467726 | 15621 |
| 202.21 | Nodular lymphoma | neoplasms | DR | 1.25 | 0.92 | 1.69 | 0.469985 | 15621 |
| 379.51 | Pigmentary iris degeneration | sense organs | DR | 1.18 | 0.94 | 1.49 | 0.470914 | 15621 |
| 528.5 | Diseases of lips | digestive | DR | 1.14 | 0.95 | 1.37 | 0.471323 | 15621 |
| 705.1 | Dyshidrosis | dermatologic | DR | 0.90 | 0.79 | 1.04 | 0.471685 | 15621 |
| 253.4 | | | DR | 0.90 | 0.78 | 1.04 | 0.473217 | 15621 |
| 743.22 | | | DR | 1.63 | 0.82 | 3.22 | 0.475051 | 15621 |
| 346.3 | Nonspecific abnormal findings in cerebrospinal fluid | neurological | DR | 0.60 | 0.29 | 1.23 | 0.475268 | 15621 |
| 816 | Cerebral laceration and contusion | injuries & poisonings | DR | 1.23 | 0.92 | 1.64 | 0.4765 | 15621 |
| 586.11 | Small kidney | genitourinary | DR | 1.74 | 0.80 | 3.80 | 0.477359 | 15621 |
| 742.9 | Other derangement of joint | musculoskeletal | DR | 0.95 | 0.89 | 1.02 | 0.478808 | 15621 |
| 958.1 | Postoperative shock | injuries & poisonings | DR | 1.38 | 0.88 | 2.16 | 0.478921 | 15621 |
| 202.22 | Reticulosarcoma | neoplasms | DR | 1.20 | 0.93 | 1.54 | 0.479493 | 15621 |
| 187 | Cancer of other male genital organs | neoplasms | DR | 0.89 | 0.76 | 1.05 | 0.484573 | 15621 |
| 711.3 | Behcet's syndrome | musculoskeletal | DR | 0.55 | 0.24 | 1.29 | 0.484864 | 15621 |
| 442.4 | Arterial dissection | circulatory system | DR | 1.34 | 0.88 | 2.03 | 0.486478 | 15621 |
| 840.1 | | | DR | 1.12 | 0.95 | 1.32 | 0.487046 | 15621 |
| 252.2 | Hypoparathyroidism | endocrine/metabolic | DR | 1.30 | 0.89 | 1.91 | 0.488694 | 15621 |
| 656.8 | Perinatal jaundice | pregnancy complications | DR | 2.37 | 0.68 | 8.27 | 0.490079 | 15621 |
| 579 | Other symptoms involving abdomen and pelvis | digestive | DR | 1.05 | 0.98 | 1.13 | 0.491227 | 15621 |
| 360.2 | Progressive myopia | sense organs | DR | 1.16 | 0.93 | 1.45 | 0.491256 | 15621 |
| 170.2 | Cancer of connective tissue | neoplasms | DR | 1.17 | 0.93 | 1.46 | 0.491344 | 15621 |
| 555.21 | Ulcerative colitis (chronic) | digestive | DR | 1.13 | 0.95 | 1.35 | 0.491395 | 15621 |
| 741.1 | Ankylosis of joint | musculoskeletal | DR | 0.85 | 0.66 | 1.08 | 0.491858 | 15621 |
| 333.3 | Tics and choreas | neurological | DR | 0.76 | 0.50 | 1.14 | 0.492791 | 15621 |
| 242.1 | Graves' disease | endocrine/metabolic | DR | 1.15 | 0.94 | 1.40 | 0.494306 | 15621 |
| 612.3 | Congenital anomalies of breast | genitourinary | DR | 1.91 | 0.74 | 4.96 | 0.494807 | 15621 |
| 770 | Myalgia and myositis unspecified | symptoms | DR | 1.03 | 0.99 | 1.08 | 0.4964 | 15621 |
| 441.2 | Chronic vascular insufficiency of intestine | circulatory system | DR | 1.31 | 0.88 | 1.96 | 0.497165 | 15621 |
| 575.9 | Nonspecific abnormal findings on radiological and other examination of biliary tract | digestive | DR | 1.07 | 0.97 | 1.17 | 0.49753 | 15621 |
| 287.4 | Qualitative platelet defects | hematopoietic | DR | 1.25 | 0.90 | 1.75 | 0.49992 | 15621 |
| 473.1 | Chronic laryngitis | respiratory | DR | 1.12 | 0.94 | 1.34 | 0.501668 | 15621 |
| 564 | Functional digestive disorders | digestive | DR | 1.03 | 0.98 | 1.09 | 0.501943 | 15621 |
| 370.1 | Corneal ulcer | sense organs | DR | 1.13 | 0.94 | 1.37 | 0.505502 | 15621 |
| 741.4 | Joint effusions | musculoskeletal | DR | 1.04 | 0.98 | 1.11 | 0.511574 | 15621 |
| 614.32 | Chronic inflammatory pelvic disease | genitourinary | DR | 0.64 | 0.32 | 1.26 | 0.512462 | 15621 |
| 750.14 | Congenital anomalies of esophagus | congenital anomalies | DR | 0.64 | 0.32 | 1.27 | 0.512785 | 15621 |
| 271.9 | Other disorders of carbohydrate transport and metabolism | endocrine/metabolic | DR | 0.77 | 0.51 | 1.15 | 0.513436 | 15621 |
| 636.2 | Early onset of delivery | pregnancy complications | DR | 0.47 | 0.15 | 1.50 | 0.515896 | 15621 |
| 90 | Sexually transmitted infections (not HIV or hepatitis) | infectious diseases | DR | 0.94 | 0.86 | 1.03 | 0.52232 | 15621 |
| 939.1 | Contact and allergic dermatitis of eyelid | dermatologic | DR | 1.22 | 0.89 | 1.67 | 0.522642 | 15621 |
| 157 | Pancreatic cancer | neoplasms | DR | 1.13 | 0.93 | 1.37 | 0.522912 | 15621 |
| 752 | Nervous system congenital anomalies | congenital anomalies | DR | 0.85 | 0.66 | 1.10 | 0.524987 | 15621 |
| 228 | Hemangioma and lymphangioma, any site | neoplasms | DR | 1.06 | 0.97 | 1.15 | 0.52587 | 15621 |
| 496.1 | Emphysema | respiratory | DR | 1.07 | 0.96 | 1.20 | 0.525909 | 15621 |
| 301.1 | Schizoid personality disorder | mental disorders | DR | 0.87 | 0.69 | 1.09 | 0.526545 | 15621 |
| 195.3 | Malignant neoplasm of head, face, and neck | neoplasms | DR | 1.14 | 0.92 | 1.42 | 0.527546 | 15621 |
| 334.21 | Amyotrophic Lateral Sclerosis | neurological | DR | 1.30 | 0.86 | 1.96 | 0.527931 | 15621 |
| 724.9 | Other unspecified back disorders | musculoskeletal | DR | 0.93 | 0.82 | 1.05 | 0.530907 | 15621 |
| 602.3 | Dysplasia of prostate | genitourinary | DR | 0.88 | 0.72 | 1.08 | 0.531845 | 15621 |
| 754.2 | Spondylolisthesis, congenital | congenital anomalies | DR | 1.11 | 0.94 | 1.32 | 0.531991 | 15621 |
| 729.7 | Nontraumatic compartment syndrome | musculoskeletal | DR | 1.27 | 0.87 | 1.86 | 0.532229 | 15621 |
| 182 | Malignant neoplasm of uterus | neoplasms | DR | 1.42 | 0.81 | 2.49 | 0.53393 | 15621 |
| 613.1 | Inflammatory disease of breast | genitourinary | DR | 1.23 | 0.88 | 1.72 | 0.535341 | 15621 |
| 184.11 | Malignant neoplasm of ovary | neoplasms | DR | 1.73 | 0.71 | 4.19 | 0.53636 | 15621 |
| 727.4 | Ganglion and cyst of synovium, tendon, and bursa | musculoskeletal | DR | 0.95 | 0.87 | 1.03 | 0.541332 | 15621 |
| 347 | Cataplexy and narcolepsy | neurological | DR | 0.84 | 0.62 | 1.12 | 0.543661 | 15621 |
| 704.12 | Telogen effluvium | dermatologic | DR | 1.34 | 0.83 | 2.16 | 0.544689 | 15621 |
| 90.2 | Gonococcal infections | infectious diseases | DR | 1.22 | 0.88 | 1.69 | 0.544903 | 15621 |
| 291 | | | DR | 0.96 | 0.89 | 1.03 | 0.545001 | 15621 |
| 261.1 | Vitamin A deficiency | endocrine/metabolic | DR | 1.22 | 0.88 | 1.69 | 0.549671 | 15621 |
| 31.1 | Leprosy | infectious diseases | DR | 0.60 | 0.26 | 1.41 | 0.551833 | 15621 |

| | | | | | | | | |
|---|---|---|---|---|---|---|---|---|
| 531.5 | Gastrojejunal ulcer | digestive | DR | 0.66 | 0.33 | 1.33 | 0.552176 | 15621 |
| 619.4 | Noninflammatory disorders of vagina | genitourinary | DR | 1.17 | 0.90 | 1.52 | 0.554525 | 15621 |
| 385.3 | Cholesteatoma | sense organs | DR | 1.13 | 0.92 | 1.40 | 0.555063 | 15621 |
| 415.11 | Pulmonary embolism and infarction, acute | circulatory system | DR | 1.06 | 0.96 | 1.18 | 0.557288 | 15621 |
| 275.11 | Hereditary hemochromatosis | hematopoietic | DR | 0.80 | 0.55 | 1.17 | 0.558971 | 15621 |
| 216.1 | | | DR | 1.05 | 0.97 | 1.14 | 0.559645 | 15621 |
| 227.2 | Benign neoplasm of parathyroid gland | neoplasms | DR | 0.80 | 0.54 | 1.18 | 0.560876 | 15621 |
| 656.5 | Hematological disorders of newborn | pregnancy complications | DR | 0.51 | 0.16 | 1.63 | 0.561652 | 15621 |
| 528.3 | Cellulitis and abscess of oral soft tissues | digestive | DR | 1.07 | 0.95 | 1.20 | 0.561944 | 15621 |
| 159.2 | Malignant neoplasm of small intestine, including duodenum | neoplasms | DR | 1.22 | 0.86 | 1.74 | 0.565135 | 15621 |
| 327.7 | Sleep related movement disorders | neurological | DR | 0.97 | 0.91 | 1.03 | 0.567749 | 15621 |
| 550 | Abdominal hernia | digestive | DR | 0.98 | 0.94 | 1.02 | 0.567859 | 15621 |
| 446.6 | Polyarteritis nodosa | circulatory system | DR | 0.61 | 0.26 | 1.44 | 0.568038 | 15621 |
| 255.12 | Hyperaldosteronism | endocrine/metabolic | DR | 1.16 | 0.89 | 1.50 | 0.568529 | 15621 |
| 550.2 | Diaphragmatic hernia | digestive | DR | 0.96 | 0.90 | 1.03 | 0.569621 | 15621 |
| 386.1 | Meniere's disease | sense organs | DR | 1.11 | 0.92 | 1.33 | 0.575131 | 15621 |
| 261.41 | Rickets or osteomalacia | endocrine/metabolic | DR | 1.20 | 0.87 | 1.65 | 0.577327 | 15621 |
| 550.3 | Femoral hernia | digestive | DR | 1.24 | 0.84 | 1.83 | 0.579865 | 15621 |
| 381.9 | Otorrhea | sense organs | DR | 1.10 | 0.92 | 1.32 | 0.580874 | 15621 |
| 783.1 | Postprocedural fever | symptoms | DR | 1.13 | 0.91 | 1.41 | 0.583385 | 15621 |
| 495.11 | | | DR | 1.11 | 0.92 | 1.33 | 0.58362 | 15621 |
| 722.7 | Intervertebral disc disorder with myelopathy | musculoskeletal | DR | 1.06 | 0.95 | 1.18 | 0.583703 | 15621 |
| 529.1 | Glossitis | digestive | DR | 0.87 | 0.67 | 1.13 | 0.585957 | 15621 |
| 530.12 | Ulcer of esophagus | digestive | DR | 1.09 | 0.93 | 1.27 | 0.586695 | 15621 |
| 740.12 | Osteoarthrosis, localized, secondary | musculoskeletal | DR | 0.96 | 0.90 | 1.03 | 0.587652 | 15621 |
| 334.2 | Anterior horn cell disease | neurological | DR | 1.17 | 0.88 | 1.56 | 0.588749 | 15621 |
| 500.1 | Extrinsic allergic alveolitis | respiratory | DR | 0.82 | 0.56 | 1.19 | 0.590626 | 15621 |
| 733.2 | Cyst of bone | musculoskeletal | DR | 1.12 | 0.91 | 1.37 | 0.590787 | 15621 |
| 242.3 | Exophthalmos | endocrine/metabolic | DR | 1.11 | 0.91 | 1.35 | 0.592388 | 15621 |
| 619.1 | Noninflammatory disorders of ovary, fallopian tube, and broad ligament | genitourinary | DR | 1.26 | 0.82 | 1.92 | 0.592856 | 15621 |
| 245.21 | Chronic lymphocytic thyroiditis | endocrine/metabolic | DR | 1.15 | 0.88 | 1.51 | 0.597221 | 15621 |
| 159.4 | Malignant neoplasm of retroperitoneum and peritoneum | neoplasms | DR | 1.37 | 0.75 | 2.48 | 0.597968 | 15621 |
| 245.1 | Thyroiditis, acute and subacute | endocrine/metabolic | DR | 1.27 | 0.81 | 1.98 | 0.598392 | 15621 |
| 658 | Maternal complication of pregnancy affecting fetus or newborn | pregnancy complications | DR | 0.72 | 0.38 | 1.35 | 0.599164 | 15621 |
| 259.8 | Polyglandular activity in multiple endocrine adenomatosis | endocrine/metabolic | DR | 0.64 | 0.27 | 1.51 | 0.601162 | 15621 |
| 750.11 | Esophageal atresia/tracheoesophageal fistula | congenital anomalies | DR | 1.18 | 0.86 | 1.64 | 0.602467 | 15621 |
| 189.4 | Malignant neoplasm of other urinary organs | neoplasms | DR | 1.17 | 0.86 | 1.59 | 0.602564 | 15621 |
| 530.9 | Heartburn | digestive | DR | 0.96 | 0.89 | 1.04 | 0.60386 | 15621 |
| 561.2 | Flatulence | digestive | DR | 1.04 | 0.97 | 1.11 | 0.603951 | 15621 |
| 130 | Spirochetal infection | infectious diseases | DR | 1.09 | 0.92 | 1.28 | 0.604667 | 15621 |
| 303.31 | | | DR | 1.36 | 0.75 | 2.49 | 0.605612 | 15621 |
| 705 | Disorders of sweat glands | dermatologic | DR | 0.95 | 0.86 | 1.05 | 0.607081 | 15621 |
| 750.21 | Congenital anomalies of intestine | congenital anomalies | DR | 1.17 | 0.86 | 1.58 | 0.610237 | 15621 |
| 938 | Dermatitis due to solar radiation | dermatologic | DR | 0.95 | 0.87 | 1.05 | 0.610422 | 15621 |
| 755.4 | Congenital anomalies of upper limb, including shoulder girdle | congenital anomalies | DR | 0.75 | 0.43 | 1.32 | 0.61243 | 15621 |
| 697 | Sarcoidosis | dermatologic | DR | 0.88 | 0.68 | 1.14 | 0.61329 | 15621 |
| 174.2 | Breast cancer [male] | neoplasms | DR | 1.35 | 0.74 | 2.49 | 0.618562 | 15621 |
| 465.4 | Acute laryngitis and tracheitis | respiratory | DR | 0.93 | 0.80 | 1.08 | 0.62167 | 15621 |
| 695.41 | Cutaneous lupus erythematosus | dermatologic | DR | 1.18 | 0.84 | 1.67 | 0.622353 | 15621 |
| 289.4 | Lymphadenitis | hematopoietic | DR | 1.04 | 0.96 | 1.11 | 0.623809 | 15621 |
| 686.5 | Pyoderma | dermatologic | DR | 1.14 | 0.88 | 1.47 | 0.625503 | 15621 |
| 611.1 | Abnormal mammogram | genitourinary | DR | 1.09 | 0.91 | 1.31 | 0.627234 | 15621 |
| 480.12 | Pseudomonal pneumonia | respiratory | DR | 1.24 | 0.80 | 1.92 | 0.627751 | 15621 |
| 752.1 | Neural tube defects | congenital anomalies | DR | 0.81 | 0.51 | 1.26 | 0.629287 | 15621 |
| 625 | Pain and other symptoms associated with female genital organs | genitourinary | DR | 0.91 | 0.75 | 1.11 | 0.63259 | 15621 |
| 303.1 | Dissociative disorder | mental disorders | DR | 1.15 | 0.86 | 1.54 | 0.632773 | 15621 |
| 8.51 | Intestinal e.coli | infectious diseases | DR | 0.81 | 0.52 | 1.26 | 0.632934 | 15621 |
| 668 | Complications of the administration of anesthetic or other sedation in labor and delivery | pregnancy complications | DR | 0.56 | 0.16 | 1.91 | 0.634744 | 15621 |
| 245.2 | Chronic thyroiditis | endocrine/metabolic | DR | 1.13 | 0.88 | 1.45 | 0.63504 | 15621 |
| 282.8 | Other hemoglobinopathies | hematopoietic | DR | 1.11 | 0.89 | 1.40 | 0.635333 | 15621 |
| 201 | Hodgkin's disease | neoplasms | DR | 0.86 | 0.62 | 1.19 | 0.635354 | 15621 |
| 246.7 | Abnormal results of function study of thyroid | endocrine/metabolic | DR | 1.05 | 0.95 | 1.17 | 0.637561 | 15621 |
| 295.3 | Psychosis | mental disorders | DR | 0.97 | 0.91 | 1.03 | 0.637704 | 15621 |
| 966 | Poisoning by anticonvulsants and anti-Parkinsonism drugs | injuries & poisonings | DR | 1.14 | 0.86 | 1.49 | 0.639467 | 15621 |
| 555 | | | DR | 0.95 | 0.84 | 1.06 | 0.639836 | 15621 |
| 592.3 | Urethral stricture due to infecton | genitourinary | DR | 0.75 | 0.40 | 1.40 | 0.639958 | 15621 |
| 647.1 | Infections of genitourinary tract during pregnancy | pregnancy complications | DR | 0.79 | 0.47 | 1.32 | 0.643196 | 15621 |
| 758.1 | Chromosomal anomalies | congenital anomalies | DR | 1.24 | 0.78 | 1.97 | 0.643291 | 15621 |
| 870.5 | Open wound of lip and mouth | injuries & poisonings | DR | 1.11 | 0.89 | 1.38 | 0.643971 | 15621 |
| 647 | Infectious and parasitic complications affecting pregnancy | pregnancy complications | DR | 0.57 | 0.17 | 1.93 | 0.644232 | 15621 |
| 727.8 | Plica syndrome | musculoskeletal | DR | 0.86 | 0.63 | 1.19 | 0.64426 | 15621 |
| 752.2 | Other specified congenital anomalies of nervous system | congenital anomalies | DR | 1.16 | 0.84 | 1.61 | 0.645431 | 15621 |
| 117.3 | Blastomycotic infection | infectious diseases | DR | 1.40 | 0.68 | 2.89 | 0.645756 | 15621 |
| 270.1 | Disturbances of amino-acid transport | endocrine/metabolic | DR | 0.79 | 0.48 | 1.32 | 0.647289 | 15621 |
| 674 | Other complications of the puerperium NEC | pregnancy complications | DR | 0.66 | 0.26 | 1.65 | 0.647399 | 15621 |
| 292.6 | Hallucinations | mental disorders | DR | 1.06 | 0.93 | 1.21 | 0.647456 | 15621 |
| 969 | Poisoning by psychotropic agents | injuries & poisonings | DR | 0.94 | 0.83 | 1.07 | 0.648162 | 15621 |
| 634.1 | Missed abortion/Hydatidiform mole | pregnancy complications | DR | 0.82 | 0.53 | 1.27 | 0.648242 | 15621 |
| 333.4 | Torsion dystonia | neurological | DR | 0.93 | 0.80 | 1.09 | 0.649016 | 15621 |
| 687.4 | Disturbance of skin sensation | dermatologic | DR | 0.98 | 0.94 | 1.02 | 0.650234 | 15621 |
| 293 | Symptoms involving head and neck | mental disorders | DR | 1.08 | 0.91 | 1.27 | 0.653746 | 15621 |
| 286.5 | Hemorrhagic disorder due to intrinsic circulating anticoagulants | hematopoietic | DR | 1.12 | 0.87 | 1.44 | 0.654529 | 15621 |
| 381.2 | Eustachian tube disorders | sense organs | DR | 1.04 | 0.96 | 1.13 | 0.655095 | 15621 |
| 149.3 | Cancer of hypopharynx | neoplasms | DR | 1.31 | 0.71 | 2.43 | 0.65574 | 15621 |
| 790.9 | Abnormal arterial blood gases | symptoms | DR | 1.32 | 0.71 | 2.45 | 0.656475 | 15621 |
| 210 | Benign neoplasm of lip, oral cavity, and pharynx | neoplasms | DR | 1.05 | 0.94 | 1.19 | 0.657353 | 15621 |
| 622.1 | Polyp of corpus uteri | genitourinary | DR | 1.23 | 0.77 | 1.97 | 0.658193 | 15621 |
| 1010.1 | screening for infectious and parasitic diseases | | DR | 0.98 | 0.94 | 1.02 | 0.658562 | 15621 |
| 323 | Encephalitis | neurological | DR | 1.12 | 0.87 | 1.45 | 0.65975 | 15621 |
| 736.3 | Acquired deformities of hip | musculoskeletal | DR | 0.77 | 0.42 | 1.40 | 0.660924 | 15621 |
| 244.1 | Secondary hypothyroidism | endocrine/metabolic | DR | 1.07 | 0.92 | 1.24 | 0.661014 | 15621 |
| 230 | Kaposi's sarcoma | neoplasms | DR | 1.21 | 0.78 | 1.89 | 0.662436 | 15621 |
| 975 | Poisoning by agents primarily acting on the smooth and skeletal muscles and respiratory system | injuries & poisonings | DR | 1.23 | 0.77 | 1.98 | 0.663349 | 15621 |
| 313.3 | Autism | mental disorders | DR | 0.67 | 0.27 | 1.69 | 0.664814 | 15621 |

| | | | | | | | | |
|---|---|---|---|---|---|---|---|---|
| 279.11 | Deficiency of humoral immunity | endocrine/metabolic | DR | 0.86 | 0.61 | 1.22 | 0.664951 | 15621 |
| 729.1 | Rheumatism, unspecified and fibrositis | musculoskeletal | DR | 1.09 | 0.89 | 1.33 | 0.665417 | 15621 |
| 306 | Other mental disorder | mental disorders | DR | 0.98 | 0.95 | 1.02 | 0.666038 | 15621 |
| 158 | Neoplasm of unspecified nature of digestive system | neoplasms | DR | 1.06 | 0.93 | 1.21 | 0.669107 | 15621 |
| 1100 | Family history | NULL | DR | 0.97 | 0.91 | 1.04 | 0.670691 | 15621 |
| 737.3 | Kyphoscoliosis and scoliosis | musculoskeletal | DR | 0.94 | 0.82 | 1.08 | 0.671737 | 15621 |
| 286.11 | Von willebrand's disease | hematopoietic | DR | 1.30 | 0.70 | 2.42 | 0.672279 | 15621 |
| 540.11 | Acute appendicitis | digestive | DR | 1.06 | 0.92 | 1.24 | 0.672552 | 15621 |
| 656.26 | Transitory tachypnea or apnea of newborn | pregnancy complications | DR | 1.53 | 0.56 | 4.15 | 0.672887 | 15621 |
| 733.9 | | | DR | 0.95 | 0.83 | 1.08 | 0.674141 | 15621 |
| 244.5 | Congenital hypothyroidism | endocrine/metabolic | DR | 0.69 | 0.28 | 1.67 | 0.674321 | 15621 |
| 569.1 | Toxic gastroenteritis and colitis | digestive | DR | 1.16 | 0.82 | 1.64 | 0.674435 | 15621 |
| 575.6 | Cholesterolosis of gallbladder | digestive | DR | 0.91 | 0.72 | 1.14 | 0.677519 | 15621 |
| 286.6 | Defibrination syndrome | hematopoietic | DR | 1.16 | 0.81 | 1.65 | 0.677733 | 15621 |
| 327.71 | Restless legs syndrome | neurological | DR | 1.04 | 0.95 | 1.13 | 0.678283 | 15621 |
| 830 | Dislocation | injuries & poisonings | DR | 0.98 | 0.92 | 1.03 | 0.679757 | 15621 |
| 325 | Phlebitis and thrombophlebitis of intracranial venous sinuses | neurological | DR | 0.62 | 0.20 | 1.98 | 0.68073 | 15621 |
| 743.4 | Stress fracture | musculoskeletal | DR | 1.07 | 0.90 | 1.28 | 0.681032 | 15621 |
| 733.4 | Aseptic necrosis of bone | musculoskeletal | DR | 1.07 | 0.91 | 1.25 | 0.686024 | 15621 |
| 245 | Thyroiditis | endocrine/metabolic | DR | 1.09 | 0.88 | 1.33 | 0.688715 | 15621 |
| 642 | Hypertension complicating pregnancy, childbirth, and the puerperium | pregnancy complications | DR | 1.20 | 0.76 | 1.88 | 0.689352 | 15621 |
| 204.3 | Monocytic leukemia | neoplasms | DR | 0.78 | 0.41 | 1.47 | 0.693235 | 15621 |
| 961 | Poisoning by other anti-infectives | injuries & poisonings | DR | 0.86 | 0.59 | 1.26 | 0.694197 | 15621 |
| 987 | Toxic effect of other gases, fumes, or vapors | injuries & poisonings | DR | 1.23 | 0.73 | 2.06 | 0.694233 | 15621 |
| 564.9 | Personal history of diseases of digestive system | digestive | DR | 1.04 | 0.94 | 1.15 | 0.694315 | 15621 |
| 270 | | | DR | 0.97 | 0.90 | 1.05 | 0.694963 | 15621 |
| 131 | Protozoan infection | infectious diseases | DR | 0.88 | 0.63 | 1.22 | 0.695476 | 15621 |
| 282.5 | Sickle cell anemia | hematopoietic | DR | 1.14 | 0.82 | 1.59 | 0.695862 | 15621 |
| 603 | Other disorders of testis | genitourinary | DR | 0.97 | 0.89 | 1.05 | 0.697847 | 15621 |
| 218.2 | Other benign neoplasm of uterus | neoplasms | DR | 0.73 | 0.32 | 1.65 | 0.699426 | 15621 |
| 282.9 | Other hereditary hemolytic anemias | hematopoietic | DR | 1.12 | 0.83 | 1.51 | 0.70243 | 15621 |
| 345.12 | Partial epilepsy | neurological | DR | 1.05 | 0.92 | 1.21 | 0.703826 | 15621 |
| 750.15 | Congenital anomalies of stomach | congenital anomalies | DR | 0.86 | 0.58 | 1.27 | 0.704355 | 15621 |
| 1003 | | | DR | 0.63 | 0.18 | 2.15 | 0.707654 | 15621 |
| 871.1 | Open wound of hand except finger(s) | injuries & poisonings | DR | 1.04 | 0.93 | 1.16 | 0.709217 | 15621 |
| 781.1 | Loss of height | symptoms | DR | 1.24 | 0.70 | 2.20 | 0.709665 | 15621 |
| 612.1 | Galactorrhea | genitourinary | DR | 0.82 | 0.48 | 1.40 | 0.711079 | 15621 |
| 528.1 | Stomatitis and mucositis | digestive | DR | 0.95 | 0.84 | 1.08 | 0.711239 | 15621 |
| 253.11 | Acromegaly and gigantism | endocrine/metabolic | DR | 0.79 | 0.41 | 1.50 | 0.712031 | 15621 |
| 149.9 | Cancer of of nasal cavities | neoplasms | DR | 1.15 | 0.78 | 1.71 | 0.712674 | 15621 |
| 513.4 | Hyperventilation | respiratory | DR | 1.08 | 0.87 | 1.34 | 0.71598 | 15621 |
| 174.1 | Breast cancer [female] | neoplasms | DR | 1.10 | 0.85 | 1.43 | 0.719553 | 15621 |
| 550.6 | Incisional hernia | digestive | DR | 0.96 | 0.86 | 1.08 | 0.723877 | 15621 |
| 722.3 | Schmorl's nodes | musculoskeletal | DR | 0.87 | 0.59 | 1.29 | 0.724778 | 15621 |
| 200.1 | Polycythemia vera | neoplasms | DR | 1.07 | 0.89 | 1.29 | 0.725289 | 15621 |
| 446.2 | Acute febrile mucocutaneous lymph node syndrome (Kawasaki disease) | circulatory system | DR | 1.64 | 0.40 | 6.75 | 0.727156 | 15621 |
| 573.2 | Liver replaced by transplant | digestive | DR | 1.07 | 0.88 | 1.32 | 0.728276 | 15621 |
| 636.3 | Hemorrhage in early pregnancy | pregnancy complications | DR | 1.21 | 0.70 | 2.07 | 0.72854 | 15621 |
| 198.7 | Secondary malignant neoplasm of skin | neoplasms | DR | 0.82 | 0.47 | 1.44 | 0.729244 | 15621 |
| 557 | Intestinal malabsorption (non-celiac) | digestive | DR | 1.10 | 0.84 | 1.43 | 0.729649 | 15621 |
| 627.5 | Premature menopause and other ovarian failure | genitourinary | DR | 1.14 | 0.78 | 1.67 | 0.729937 | 15621 |
| 221 | Benign neoplasm of other female genital organs | neoplasms | DR | 0.79 | 0.40 | 1.56 | 0.733947 | 15621 |
| 870.4 | Open wound of nose and sinus | injuries & poisonings | DR | 0.91 | 0.68 | 1.21 | 0.738562 | 15621 |
| 283.21 | Hemolytic-uremic syndrome | hematopoietic | DR | 1.61 | 0.38 | 6.75 | 0.741506 | 15621 |
| 610.4 | Benign neoplasm of breast | genitourinary | DR | 0.90 | 0.67 | 1.23 | 0.742089 | 15621 |
| 540 | Appendiceal conditions | digestive | DR | 1.04 | 0.91 | 1.19 | 0.743338 | 15621 |
| 737.1 | Kyphosis (acquired) | musculoskeletal | DR | 1.08 | 0.86 | 1.35 | 0.745898 | 15621 |
| 726.3 | Bursitis | musculoskeletal | DR | 0.98 | 0.92 | 1.04 | 0.749609 | 15621 |
| 603.1 | Hydrocele | genitourinary | DR | 1.03 | 0.94 | 1.13 | 0.749624 | 15621 |
| 656.7 | Conditions involving the integument and temperature regulation of fetus and newborn | pregnancy complications | DR | 1.26 | 0.61 | 2.58 | 0.74966 | 15621 |
| 443.1 | Raynaud's syndrome | circulatory system | DR | 0.89 | 0.63 | 1.27 | 0.750611 | 15621 |
| 652 | Malposition and malpresentation of fetus or obstruction | pregnancy complications | DR | 0.80 | 0.39 | 1.64 | 0.753773 | 15621 |
| 305.21 | Anorexia nervosa | mental disorders | DR | 1.27 | 0.59 | 2.75 | 0.756559 | 15621 |
| 732 | Osteochondropathies | musculoskeletal | DR | 0.95 | 0.80 | 1.12 | 0.757195 | 15621 |
| 619.2 | Disorders of uterus, NEC | genitourinary | DR | 1.08 | 0.85 | 1.37 | 0.759672 | 15621 |
| 614.3 | Pelvic inflammatory disease (PID) | genitourinary | DR | 1.12 | 0.77 | 1.63 | 0.761833 | 15621 |
| 592.21 | Urethral syndrome | genitourinary | DR | 0.81 | 0.41 | 1.60 | 0.762051 | 15621 |
| 988 | Toxic effect of noxious substances eaten as food | injuries & poisonings | DR | 1.19 | 0.66 | 2.16 | 0.764512 | 15621 |
| 526.42 | Arthralgia/ankylosis of temporomandibular joint | digestive | DR | 1.04 | 0.91 | 1.20 | 0.766233 | 15621 |
| 694.2 | Other dyschromia | dermatologic | DR | 0.98 | 0.92 | 1.05 | 0.767702 | 15621 |
| 117.1 | Histoplasmosis | infectious diseases | DR | 1.14 | 0.73 | 1.77 | 0.767998 | 15621 |
| 750.13 | Congenital anomalies of mouth/tongue | congenital anomalies | DR | 1.15 | 0.72 | 1.84 | 0.768761 | 15621 |
| 254 | Diseases of thymus gland | endocrine/metabolic | DR | 1.19 | 0.65 | 2.18 | 0.77126 | 15621 |
| 526.41 | Temporomandibular joint disorder, unspecified | digestive | DR | 1.03 | 0.94 | 1.12 | 0.771282 | 15621 |
| 795.8 | Abnormal tumor markers | symptoms | DR | 0.89 | 0.59 | 1.34 | 0.772484 | 15621 |
| 350.6 | Disturbances of sensation of smell and taste | neurological | DR | 1.06 | 0.87 | 1.28 | 0.773303 | 15621 |
| 300.4 | Dysthymic disorder | mental disorders | DR | 0.99 | 0.94 | 1.04 | 0.775525 | 15621 |
| 270.35 | Macroglobulinemia | endocrine/metabolic | DR | 1.20 | 0.63 | 2.31 | 0.77781 | 15621 |
| 754 | Congenital musculoskeletal deformities of spine | congenital anomalies | DR | 1.04 | 0.91 | 1.19 | 0.77874 | 15621 |
| 695.42 | Systemic lupus erythematosus | dermatologic | DR | 0.92 | 0.69 | 1.23 | 0.782462 | 15621 |
| 860 | Bone marrow or stem cell transplant | neoplasms | DR | 0.87 | 0.53 | 1.43 | 0.782839 | 15621 |
| 218 | | | DR | 1.06 | 0.86 | 1.31 | 0.782912 | 15621 |
| 624.9 | stress incontinence, female | genitourinary | DR | 0.96 | 0.84 | 1.11 | 0.783163 | 15621 |
| 618.1 | Prolapse of vaginal walls | genitourinary | DR | 1.09 | 0.80 | 1.48 | 0.784171 | 15621 |
| 385 | Other disorders of middle ear and mastoid | sense organs | DR | 1.05 | 0.89 | 1.23 | 0.787491 | 15621 |
| 751.1 | Congenital anomalies of genital organs | congenital anomalies | DR | 0.97 | 0.85 | 1.10 | 0.790928 | 15621 |
| 348.4 | Cerebral cysts | neurological | DR | 1.08 | 0.80 | 1.47 | 0.791351 | 15621 |
| 355.1 | Chronic pain syndrome | neurological | DR | 1.01 | 0.96 | 1.07 | 0.791844 | 15621 |
| 695.21 | Dermatitis herpetiformis | dermatologic | DR | 0.89 | 0.58 | 1.37 | 0.791912 | 15621 |
| 870.6 | Open wound of neck | injuries & poisonings | DR | 0.90 | 0.62 | 1.33 | 0.792192 | 15621 |
| 204.11 | Lymphoid leukemia, acute | neoplasms | DR | 1.13 | 0.70 | 1.83 | 0.792393 | 15621 |
| 495.1 | | | DR | 0.97 | 0.86 | 1.09 | 0.793263 | 15621 |
| 695.4 | Lupus (localized and systemic) | dermatologic | DR | 1.07 | 0.84 | 1.36 | 0.79403 | 15621 |
| 258 | Iatrogenic endocrine disorders | endocrine/metabolic | DR | 0.89 | 0.58 | 1.37 | 0.794143 | 15621 |

| | | | | | | | | |
|---|---|---|---|---|---|---|---|---|
| 624.1 | Dystrophy of female genital tract | genitourinary | DR | 0.75 | 0.24 | 2.28 | 0.794343 | 15621 |
| 229.1 | Benign neoplasm of lymph nodes | neoplasms | DR | 0.91 | 0.64 | 1.30 | 0.794985 | 15621 |
| 716.8 | Palindromic rheumatism | musculoskeletal | DR | 0.90 | 0.59 | 1.37 | 0.79574 | 15621 |
| 704 | Diseases of hair and hair follicles | dermatologic | DR | 1.01 | 0.96 | 1.07 | 0.796378 | 15621 |
| 134.1 | Intestinal helminthiases | infectious diseases | DR | 1.09 | 0.78 | 1.52 | 0.796525 | 15621 |
| 218.1 | Uterine leiomyoma | neoplasms | DR | 1.06 | 0.85 | 1.31 | 0.796741 | 15621 |
| 613.5 | Mastodynia | genitourinary | DR | 1.04 | 0.89 | 1.23 | 0.797863 | 15621 |
| 291.1 | Transient mental disorders due to conditions classified elsewhere | mental disorders | DR | 0.95 | 0.78 | 1.16 | 0.798804 | 15621 |
| 573 | Other disorders of liver | digestive | DR | 1.02 | 0.95 | 1.09 | 0.801008 | 15621 |
| 209 | Neuroendocrine tumors | neoplasms | DR | 1.08 | 0.79 | 1.47 | 0.80145 | 15621 |
| 198.5 | Secondary malignancy of brain/spine | neoplasms | DR | 1.07 | 0.82 | 1.41 | 0.80162 | 15621 |
| 540.1 | Appendicitis | digestive | DR | 1.04 | 0.90 | 1.19 | 0.803425 | 15621 |
| 270.11 | Disturbances of sulphur-bearing amino-acid metabolism | endocrine/metabolic | DR | 0.81 | 0.34 | 1.93 | 0.80521 | 15621 |
| 736.6 | Unequal leg length (acquired) | musculoskeletal | DR | 1.03 | 0.91 | 1.16 | 0.811407 | 15621 |
| 471 | Nasal polyps | respiratory | DR | 1.03 | 0.90 | 1.18 | 0.811442 | 15621 |
| 323.2 | Acute (transverse) myelitis | neurological | DR | 0.83 | 0.38 | 1.82 | 0.812746 | 15621 |
| 753.2 | Congenital anomalies of posterior segment of eye | congenital anomalies | DR | 0.95 | 0.76 | 1.19 | 0.814271 | 15621 |
| 557.1 | Celiac disease | digestive | DR | 1.09 | 0.76 | 1.56 | 0.817369 | 15621 |
| 611.11 | Mammographic microcalcification | genitourinary | DR | 1.10 | 0.72 | 1.69 | 0.818447 | 15621 |
| 378.2 | Nystagmus and other irregular eye movements | sense organs | DR | 0.95 | 0.76 | 1.19 | 0.821242 | 15621 |
| 715.1 | Sacroiliitis NEC | musculoskeletal | DR | 1.02 | 0.93 | 1.13 | 0.821609 | 15621 |
| 498 | Acute bronchospasm | respiratory | DR | 1.03 | 0.91 | 1.17 | 0.822004 | 15621 |
| 618 | Genital prolapse | genitourinary | DR | 1.06 | 0.81 | 1.40 | 0.823204 | 15621 |
| 174.11 | Malignant neoplasm of female breast | neoplasms | DR | 1.06 | 0.81 | 1.39 | 0.824513 | 15621 |
| 536.8 | Dyspepsia and other specified disorders of function of stomach | digestive | DR | 0.99 | 0.93 | 1.05 | 0.825831 | 15621 |
| 358.1 | Myasthenia gravis | neurological | DR | 1.07 | 0.77 | 1.49 | 0.826087 | 15621 |
| 550.5 | Ventral hernia | digestive | DR | 0.98 | 0.91 | 1.06 | 0.826632 | 15621 |
| 286.4 | Acquired coagulation factor deficiency | hematopoietic | DR | 1.06 | 0.82 | 1.36 | 0.826912 | 15621 |
| 79.1 | Varicella infection | infectious diseases | DR | 1.09 | 0.72 | 1.65 | 0.829611 | 15621 |
| 384.1 | Myringitis | sense organs | DR | 1.06 | 0.81 | 1.39 | 0.830409 | 15621 |
| 795 | Other and nonspecific abnormal cytological, histological and immunological findings | symptoms | DR | 0.88 | 0.48 | 1.60 | 0.830974 | 15621 |
| 618.2 | Uterine/Uterovaginal prolapse | genitourinary | DR | 1.14 | 0.62 | 2.09 | 0.831351 | 15621 |
| 253 | Disorders of the pituitary gland and its hypothalamic control | endocrine/metabolic | DR | 1.02 | 0.93 | 1.12 | 0.831809 | 15621 |
| 941 | Adverse reaction to serum or vaccine | injuries & poisonings | DR | 0.93 | 0.68 | 1.29 | 0.833765 | 15621 |
| 657 | Infections specific to the perinatal period | pregnancy complications | DR | 0.90 | 0.56 | 1.46 | 0.834948 | 15621 |
| 464 | Acute sinusitis | respiratory | DR | 0.99 | 0.95 | 1.04 | 0.840844 | 15621 |
| 613.7 | Other signs and symptoms in breast | genitourinary | DR | 0.95 | 0.75 | 1.21 | 0.841752 | 15621 |
| 967 | Adverse effects of sedatives or other central nervous system depressants and anesthetics | injuries & poisonings | DR | 1.04 | 0.86 | 1.25 | 0.842697 | 15621 |
| 610.2 | Fibroadenosis of breast | genitourinary | DR | 1.08 | 0.72 | 1.62 | 0.843949 | 15621 |
| 715 | Other inflammatory spondylopathies | musculoskeletal | DR | 1.02 | 0.94 | 1.10 | 0.844129 | 15621 |
| 242.31 | | | DR | 0.86 | 0.41 | 1.83 | 0.845616 | 15621 |
| 286.1 | Congenital coagulation defects | hematopoietic | DR | 1.08 | 0.72 | 1.64 | 0.846487 | 15621 |
| 706.3 | | | DR | 1.04 | 0.86 | 1.26 | 0.84671 | 15621 |
| 279.7 | Other immunological findings | endocrine/metabolic | DR | 1.03 | 0.90 | 1.17 | 0.849855 | 15621 |
| 246 | Other disorders of thyroid | endocrine/metabolic | DR | 0.98 | 0.87 | 1.10 | 0.851573 | 15621 |
| 270.21 | Disorders of urea cycle metabolism | endocrine/metabolic | DR | 1.04 | 0.84 | 1.29 | 0.851804 | 15621 |
| 751.11 | Congenital anomalies of female genital organs | congenital anomalies | DR | 0.89 | 0.49 | 1.64 | 0.853554 | 15621 |
| 446.8 | Thrombotic microangiopathy | circulatory system | DR | 1.20 | 0.44 | 3.32 | 0.855239 | 15621 |
| 526.9 | Jaw disease NOS | digestive | DR | 1.04 | 0.85 | 1.26 | 0.857984 | 15621 |
| 594.8 | Renal colic | genitourinary | DR | 0.98 | 0.86 | 1.11 | 0.858094 | 15621 |
| 986 | Toxic effect of carbon monoxide | injuries & poisonings | DR | 0.80 | 0.23 | 2.78 | 0.859762 | 15621 |
| 334.1 | Spinocerebellar disease | neurological | DR | 1.06 | 0.77 | 1.44 | 0.86093 | 15621 |
| 731.1 | Osteitis deformans [Paget's disease of bone] | musculoskeletal | DR | 0.92 | 0.57 | 1.49 | 0.861567 | 15621 |
| 389.5 | Disorders of acoustic nerve | sense organs | DR | 0.97 | 0.80 | 1.17 | 0.863763 | 15621 |
| 654.1 | Abnormality of organs and soft tissues of pelvis complicating pregnancy, childbirth, or the puerperium | pregnancy complications | DR | 0.89 | 0.43 | 1.82 | 0.867236 | 15621 |
| 202.24 | Large cell lymphoma | neoplasms | DR | 1.09 | 0.63 | 1.90 | 0.87016 | 15621 |
| 711.2 | Reiter's disease | musculoskeletal | DR | 1.07 | 0.71 | 1.60 | 0.870819 | 15621 |
| 655 | Known or suspected fetal abnormality affecting management of mother | pregnancy complications | DR | 0.92 | 0.55 | 1.55 | 0.878511 | 15621 |
| 756.2 | Pectus and other congenital anomalies of ribs/sternum | congenital anomalies | DR | 1.11 | 0.53 | 2.34 | 0.886451 | 15621 |
| 555.2 | Ulcerative colitis | digestive | DR | 1.02 | 0.90 | 1.16 | 0.886956 | 15621 |
| 653 | Problems associated with amniotic cavity and membranes | pregnancy complications | DR | 0.00 | 0.00 | 5.13E+29 | 0.890038 | 15621 |
| 442.2 | Aneurysm of iliac artery | circulatory system | DR | 0.96 | 0.74 | 1.25 | 0.890299 | 15621 |
| 710.2 | | | DR | 0.90 | 0.41 | 1.97 | 0.892659 | 15621 |
| 798.1 | Chronic fatigue syndrome | symptoms | DR | 1.02 | 0.90 | 1.14 | 0.895214 | 15621 |
| 619.5 | Noninflammatory disorders of vulva and perineum | genitourinary | DR | 0.95 | 0.62 | 1.44 | 0.895578 | 15621 |
| 650 | Normal delivery | pregnancy complications | DR | 0.89 | 0.35 | 2.23 | 0.895924 | 15621 |
| 756.1 | Congenital anomalies of abdominal wall; diaphram | congenital anomalies | DR | 1.06 | 0.67 | 1.68 | 0.896072 | 15621 |
| 614.51 | Cervicitis and endocervicitis | genitourinary | DR | 1.04 | 0.76 | 1.43 | 0.896857 | 15621 |
| 384.4 | Perforation of tympanic membrane | sense organs | DR | 1.02 | 0.90 | 1.15 | 0.897484 | 15621 |
| 353.1 | Nerve plexus lesions | neurological | DR | 1.03 | 0.84 | 1.25 | 0.898058 | 15621 |
| 259.4 | Precocious sexual development and puberty NEC | endocrine/metabolic | DR | 0.00 | 0.00 | 7.72E+30 | 0.898174 | 15621 |
| 736.4 | Genu valgum or varum (acquired) | musculoskeletal | DR | 1.02 | 0.86 | 1.21 | 0.898618 | 15621 |
| 81.12 | Chronic graft-versus-host disease | infectious diseases | DR | 3.76E+04 | 0.00 | 1.17E+41 | 0.900241 | 15621 |
| 858 | Complication of internal orthopedic device | injuries & poisonings | DR | 1.01 | 0.92 | 1.12 | 0.900823 | 15621 |
| 255.22 | | | DR | 1.20E+05 | 0.00 | 2.89E+46 | 0.90228 | 15621 |
| 750.5 | Congenital hypertrophic pyloric stenosis | congenital anomalies | DR | 3.00E+04 | 0.00 | 1.45E+41 | 0.902877 | 15621 |
| 184.2 | Cancer of other female genital organs (excluding uterus and ovary) | neoplasms | DR | 0.91 | 0.40 | 2.05 | 0.903709 | 15621 |
| 750.1 | Upper gastrointestinal congenital anomalies | congenital anomalies | DR | 1.03 | 0.83 | 1.26 | 0.903949 | 15621 |
| 965.3 | Salicylates causing adverse effects in therapeutic use | injuries & poisonings | DR | 1.06 | 0.67 | 1.66 | 0.904263 | 15621 |
| 645 | Late pregnancy and failed induction | pregnancy complications | DR | 0.00 | 0.00 | 4.88E+31 | 0.904276 | 15621 |
| 306.1 | Mental disorders durring/after pregnancy | mental disorders | DR | 0.00 | 0.00 | 1.90E+32 | 0.904871 | 15621 |
| 643 | Excessive vomiting in pregnancy | pregnancy complications | DR | 0.00 | 0.00 | 2.00E+32 | 0.906276 | 15621 |
| 694 | | | DR | 1.01 | 0.95 | 1.07 | 0.907301 | 15621 |
| 446.4 | Wegener's granulomatosis | circulatory system | DR | 0.00 | 0.00 | 2.64E+32 | 0.908899 | 15621 |
| 931 | Contact dermatitis and other eczema due to plants [except food] | dermatologic | DR | 1.02 | 0.87 | 1.20 | 0.909157 | 15621 |
| 70.4 | Chronic hepatitis | infectious diseases | DR | 0.99 | 0.88 | 1.11 | 0.910534 | 15621 |
| 284.2 | Constitutional aplastic anemia | hematopoietic | DR | 0.87 | 0.25 | 3.01 | 0.911596 | 15621 |
| 277.4 | Disorders of bilirubin excretion | endocrine/metabolic | DR | 0.99 | 0.87 | 1.12 | 0.912621 | 15621 |
| 613 | Other nonmalignant breast conditions | genitourinary | DR | 1.01 | 0.90 | 1.15 | 0.913228 | 15621 |
| 145.3 | Cancer of major salivary glands | neoplasms | DR | 0.97 | 0.71 | 1.31 | 0.914127 | 15621 |
| 289.1 | Myelofibrosis | hematopoietic | DR | 4.63E+05 | 0.00 | 1.76E+58 | 0.914188 | 15621 |
| 198.4 | Secondary malignant neoplasm of liver | neoplasms | DR | 1.02 | 0.86 | 1.21 | 0.914787 | 15621 |
| 229 | Benign neoplasm of unspecified sites | neoplasms | DR | 0.98 | 0.84 | 1.15 | 0.915469 | 15621 |

| | | | | | | | | |
|---|---|---|---|---|---|---|---|---|
| 286.12 | Congenital deficiency of other clotting factors (including factor VII) | hematopoietic | DR | 1.08 | 0.51 | 2.28 | 0.915998 | 15621 |
| 324.1 | Jakob-Creutzfeldt disease | neurological | DR | 0.88 | 0.27 | 2.93 | 0.916073 | 15621 |
| 287.2 | Allergic purpura | hematopoietic | DR | 1.05 | 0.65 | 1.70 | 0.916697 | 15621 |
| 226 | Benign neoplasm of thyroid glands | neoplasms | DR | 1.03 | 0.80 | 1.33 | 0.917843 | 15621 |
| 277.6 | Other deficiencies of circulating enzymes | endocrine/metabolic | DR | 1.06 | 0.57 | 1.97 | 0.919238 | 15621 |
| 253.5 | | | DR | 0.00 | 0.00 | 6.09E+43 | 0.920162 | 15621 |
| 275.2 | Disorders of copper metabolism | endocrine/metabolic | DR | 1.09 | 0.48 | 2.48 | 0.920289 | 15621 |
| 694.1 | Vitiligo | dermatologic | DR | 0.98 | 0.84 | 1.16 | 0.921358 | 15621 |
| 255.3 | Adrenogenital disorders | endocrine/metabolic | DR | 0.00 | 0.00 | 1.07E+44 | 0.921586 | 15621 |
| 960.3 | | | DR | 1.14E+05 | 0.00 | 8.75E+56 | 0.922342 | 15621 |
| 823 | | | DR | 0.00 | 0.00 | 2.06E+44 | 0.922515 | 15621 |
| 255.13 | Medulloadrenal hyperfunction | endocrine/metabolic | DR | 0.00 | 0.00 | 7.77E+46 | 0.923322 | 15621 |
| 756.22 | Pectus carinatum | congenital anomalies | DR | 7.18E+04 | 0.00 | 5.50E+56 | 0.925427 | 15621 |
| 286.3 | Coagulation defects complicating pregnancy or postpartum | hematopoietic | DR | 6.10E+04 | 0.00 | 4.68E+56 | 0.92651 | 15621 |
| 258.1 | Postablative ovarian failure | endocrine/metabolic | DR | 0.00 | 0.00 | 2.57E+43 | 0.926538 | 15621 |
| 264.9 | Lack of normal physiological development, unspecified | endocrine/metabolic | DR | 0.00 | 0.00 | 1.62E+47 | 0.928205 | 15621 |
| 446.7 | Takayasu's disease | circulatory system | DR | 1.14 | 0.27 | 4.76 | 0.929105 | 15621 |
| 618.6 | Vaginal enterocele, congenital or acquired | genitourinary | DR | 0.00 | 0.00 | 3.66E+44 | 0.929331 | 15621 |
| 475 | Chronic sinusitis | respiratory | DR | 1.00 | 0.95 | 1.04 | 0.930721 | 15621 |
| 255.1 | Adrenal hyperfunction | endocrine/metabolic | DR | 0.98 | 0.80 | 1.21 | 0.9314 | 15621 |
| 840 | Sprains and strains | injuries & poisonings | DR | 1.00 | 0.96 | 1.03 | 0.931509 | 15621 |
| 656.9 | Neonatal bradycardia or tachycardia | pregnancy complications | DR | 1.09 | 0.40 | 2.99 | 0.93166 | 15621 |
| 636.8 | Cervical incompetence | pregnancy complications | DR | 2.71E+04 | 0.00 | 2.07E+56 | 0.931924 | 15621 |
| 651 | Multiple gestation | pregnancy complications | DR | 0.00 | 0.00 | 2.89E+47 | 0.932048 | 15621 |
| 704.2 | Hirsutism | dermatologic | DR | 0.97 | 0.70 | 1.35 | 0.932389 | 15621 |
| 601.3 | Orchitis and epididymitis | genitourinary | DR | 1.01 | 0.93 | 1.09 | 0.932665 | 15621 |
| 756.21 | Pectus excavatum | congenital anomalies | DR | 0.00 | 0.00 | 3.24E+47 | 0.93281 | 15621 |
| 654.2 | Rhesus isoimmunization in pregnancy | pregnancy complications | DR | 0.00 | 0.00 | 3.69E+47 | 0.933678 | 15621 |
| 520.2 | Disturbances in tooth eruption | digestive | DR | 0.99 | 0.90 | 1.10 | 0.935532 | 15621 |
| 639 | Complications following abortion or ectopic and molar pregnancies | pregnancy complications | DR | 0.00 | 0.00 | 5.11E+47 | 0.935849 | 15621 |
| 346 | Abnormal findings on study of brain and/or nervous system | neurological | DR | 0.99 | 0.86 | 1.14 | 0.935922 | 15621 |
| 644 | Anemia during pregnancy | pregnancy complications | DR | 0.00 | 0.00 | 6.01E+47 | 0.936935 | 15621 |
| 691.1 | Ichthyosis congenita | dermatologic | DR | 0.96 | 0.57 | 1.63 | 0.939775 | 15621 |
| 512.1 | Wheezing | respiratory | DR | 1.01 | 0.93 | 1.09 | 0.939823 | 15621 |
| 394.4 | Acute rheumatic heart disease | circulatory system | DR | 1.03 | 0.67 | 1.60 | 0.941725 | 15621 |
| 110.13 | Dermatophytosis of the body | infectious diseases | DR | 1.00 | 0.94 | 1.06 | 0.944928 | 15621 |
| 448 | Disease of capillaries | circulatory system | DR | 0.98 | 0.77 | 1.25 | 0.945843 | 15621 |
| 610 | | | DR | 0.99 | 0.81 | 1.20 | 0.946164 | 15621 |
| 754.1 | | | DR | 1.02 | 0.79 | 1.30 | 0.947785 | 15621 |
| 292.12 | Symbolic dysfunction | mental disorders | DR | 1.02 | 0.78 | 1.32 | 0.947881 | 15621 |
| 973 | Poisoning by agents primarily affecting the gastrointestinal system | injuries & poisonings | DR | 0.96 | 0.45 | 2.01 | 0.951537 | 15621 |
| 610.8 | Other specified benign mammary dysplasias | genitourinary | DR | 1.02 | 0.71 | 1.48 | 0.953603 | 15621 |
| 952 | Spinal cord injury without evidence of spinal bone injury | injuries & poisonings | DR | 0.99 | 0.83 | 1.18 | 0.95426 | 15621 |
| 727.7 | Contracture of tendon (sheath) | musculoskeletal | DR | 1.01 | 0.82 | 1.25 | 0.954847 | 15621 |
| 215 | Other benign neoplasm of connective and other soft tissue | neoplasms | DR | 0.99 | 0.88 | 1.12 | 0.95664 | 15621 |
| 604.2 | | | DR | 1.03 | 0.57 | 1.89 | 0.957341 | 15621 |
| 709.4 | Polymyositis | dermatologic | DR | 0.97 | 0.57 | 1.67 | 0.95796 | 15621 |
| 691.3 | Congenital pigmentary anomalies of skin | dermatologic | DR | 0.97 | 0.54 | 1.75 | 0.958821 | 15621 |
| 728.7 | Fasciitis | musculoskeletal | DR | 1.00 | 0.96 | 1.04 | 0.95885 | 15621 |
| 709.5 | Dermatomyositis | dermatologic | DR | 1.03 | 0.61 | 1.73 | 0.961354 | 15621 |
| 573.9 | Abnormal serum enzyme levels | digestive | DR | 1.00 | 0.94 | 1.07 | 0.961995 | 15621 |
| 610.1 | Cystic mastopathy | genitourinary | DR | 1.01 | 0.79 | 1.29 | 0.963338 | 15621 |
| 613.9 | Breast disorder NOS | genitourinary | DR | 0.99 | 0.78 | 1.26 | 0.963667 | 15621 |
| 133 | Arthropod-borne diseases | infectious diseases | DR | 0.98 | 0.70 | 1.38 | 0.964224 | 15621 |
| 174 | Breast cancer | neoplasms | DR | 0.99 | 0.78 | 1.25 | 0.964711 | 15621 |
| 343 | Infantile cerebral palsy | neurological | DR | 0.98 | 0.55 | 1.74 | 0.96631 | 15621 |
| 636 | Early or threatened labor; hemorrhage in early pregnancy | pregnancy complications | DR | 1.02 | 0.66 | 1.57 | 0.96928 | 15621 |
| 627.22 | | | DR | 1.01 | 0.70 | 1.47 | 0.976102 | 15621 |
| 371.9 | Chronic inflammatory disorders of orbit | sense organs | DR | 1.02 | 0.55 | 1.90 | 0.977607 | 15621 |
| 758 | Chromosomal anomalies and genetic disorders | congenital anomalies | DR | 1.01 | 0.67 | 1.54 | 0.978257 | 15621 |
| 204.22 | Myeloid leukemia, chronic | neoplasms | DR | 0.99 | 0.67 | 1.47 | 0.984163 | 15621 |
| 368.5 | Color vision deficiencies | sense organs | DR | 0.99 | 0.69 | 1.43 | 0.984818 | 15621 |
| 636.1 | Threatened premature labor | pregnancy complications | DR | 0.99 | 0.41 | 2.37 | 0.987296 | 15621 |
| 335 | Multiple sclerosis | neurological | DR | 1.00 | 0.78 | 1.29 | 0.988796 | 15621 |
| 984 | Toxic effect of lead and its compounds (including fumes) | injuries & poisonings | DR | 0.99 | 0.22 | 4.35 | 0.991954 | 15621 |
| 626.21 | Mittelschmerz | genitourinary | DR | 1.01 | 0.30 | 3.46 | 0.992101 | 15621 |
| 521.4 | Tooth complications likely association with other diseases | digestive | DR | 1.00 | 0.61 | 1.64 | 0.995625 | 15621 |
