## Supplemental Table 10 for "Development of Portable Electronic Health Record Based Algorithms to Identify Individuals with Diabetic Retinopathy"

**Supplemental Table 10. Details of imaging-based DR diagnosis and confirmation of staging validation for algorithmically-determined DR cases with fundus imaging available**

| **Patient** | **Algorithm-derived DR category** | **cpt_fundus** | **cpt_oct** | **cpt_tri** | **Is there available imaging? Y/N** | **Confirmation of DR via image reveiw? Y/N** | **Confirmation of staging via image review? Y/N** |
| --- | --- | --- | --- | --- | --- | --- | --- |
| **Patient 1** | MNPDR | 0 | 0 | 1 | Y | Y | Y MNPDR |
|  | NSDR | 0 | 0 | 1 | N | NA- no photos available | NA |
| **Patient 2** | MNPDR | 0 | 0 | 1 | Y | Y | Y MNPDR |
| **Patient 3** | snpdr_hf | 1 | 0 | 1 | Y | Y | Y SNPDR |
|  | snpdr_hf | 0 | 0 | 1 | Y | Y | Y SNPDR |
|  | MNPDR | 0 | 0 | 1 | Y | Y | N looks more like SNPDR |
|  | MNPDR | 0 | 0 | 1 | Y | Y | N SNPDR OD; MNDPR OS |
| **Patient 4** | NSDR | 1 | 0 | 1 | Y | Y | Y has mild NPDR OD |
|  | NSDR | 0 | 0 | 1 | Y | Y | Y has mild NPDR OD |
|  | MNPDR | 1 | 0 | 1 | Y | Y | Y has mild NPDR OD |
|  | MNPDR | 0 | 0 | 1 | Y | Y | Y has mild NPDR OD |
| **Patient 5** | NSDR | 0 | 0 | 1 | N | NA- no photos available | NA |
|  | MNPDR | 0 | 0 | 1 | N | NA- no photos available | NA |
|  | NSDR | 0 | 0 | 1 | N | NA- no photos available | NA |
|  | NSDR | 0 | 0 | 1 | N | NA- no photos available | NA |
|  | NSDR | 0 | 0 | 1 | N | NA- no photos available | NA |
|  | MNPDR | 0 | 1 | 1 | N | NA- no photos available | NA |
|  | MNPDR | 0 | 0 | 1 | N | NA- no photos available | NA |
| **Patient 6** | NSDR | 0 | 0 | 1 | N | NA- no photos available | NA |
|  | NSDR | 0 | 0 | 1 | N | NA- no photos available | NA |
|  | NSDR | 0 | 0 | 1 | N | NA- no photos available | NA |
|  | NSDR | 0 | 0 | 1 | N | NA- no photos available | NA |
|  | NSDR | 0 | 0 | 1 | N | NA- no photos available | NA |
|  | NSDR | 0 | 0 | 1 | N | NA- no photos available | NA |
|  | NSDR | 0 | 0 | 1 | N | NA- no photos available | NA |
|  | MNPDR | 0 | 0 | 1 | N | NA- no photos available | NA |
|  | MNPDR | 0 | 0 | 1 | N | NA- no photos available | NA |
|  | MNPDR | 0 | 0 | 1 | N | NA- no photos available | NA |
|  | MNPDR | 0 | 0 | 1 | N | NA- no photos available | NA |
|  | MNPDR | 0 | 0 | 1 | N | NA- no photos available | NA |
|  | MNPDR | 0 | 0 | 1 | N | NA- no photos available | NA |
| **Patient 7** | NSDR | 0 | 0 | 1 | N | NA-no photos available | NA |
| **Patient 8** | MNPDR | 0 | 1 | 1 | Y, only OCT | NA-only OCT available, cannot see DR | NA |
|  | MNPDR | 0 | 0 | 1 | Y, only OCT | NA-only OCT available, cannot see DR | NA |
| **Patient 9** | NSDR | 0 | 0 | 1 | N | NA-no photos available | NA |
|  | MNPDR | 0 | 0 | 1 | N | NA-no photos available | NA |
|  | NSDR | 0 | 0 | 1 | N | NA-no photos available | NA |
|  | NSDR | 0 | 0 | 1 | N | NA-no photos available | NA |
|  | MNPDR | 0 | 0 | 1 | N | NA-no photos available | NA |
|  | NSDR | 0 | 1 | 1 | Y, only OCT | NA-only OCT available, cannot see DR | NA |
|  | NSDR | 0 | 1 | 1 | Y | Y | Y-mild NPDR |
|  | NSDR | 1 | 0 | 1 | Y | Y | Y-mild NPDR |
|  | NSDR | 0 | 0 | 1 | Y | Y | Y-mild NPDR |
|  | NSDR | 0 | 1 | 1 | Y, only OCT | NA-only OCT available, cannot see DR | NA |
|  | NSDR | 0 | 1 | 1 | Y, only OCT | NA-only OCT available, cannot see DR | NA |
|  | MNPDR | 0 | 1 | 1 | Y, only OCT | NA-only OCT available, cannot see DR | NA |
|  | NSDR | 0 | 1 | 1 | Y, only OCT | NA-only OCT available, cannot see DR | NA |
|  | NSDR | 0 | 1 | 1 | Y, only OCT | NA-only OCT available, cannot see DR | NA |
|  | MNPDR | 0 | 1 | 1 | Y, only OCT | NA-only OCT available, cannot see DR | NA |
|  | NSDR | 0 | 1 | 1 | Y, only OCT | NA-only OCT available, cannot see DR | NA |
|  | MNPDR | 0 | 1 | 1 | Y, only OCT | NA-only OCT available, cannot see DR | NA |
|  | NSDR | 0 | 1 | 1 | Y, only OCT | NA-only OCT available, cannot see DR | NA |
|  | MNPDR | 0 | 0 | 1 | Y, only OCT | NA-only OCT available, cannot see DR | NA |
|  | MNPDR | 0 | 1 | 1 | Y, only OCT | NA-only OCT available, cannot see DR | NA |
|  | NSDR | 0 | 1 | 1 | Y, only OCT | NA-only OCT available, cannot see DR | NA |
|  | MNPDR | 0 | 1 | 1 | Y, only OCT | NA-only OCT available, cannot see DR | NA |
|  | NSDR | 0 | 0 | 1 | Y, only OCT | NA-only OCT available, cannot see DR | NA |
|  | NSDR | 0 | 1 | 1 | Y, only OCT | NA-only OCT available, cannot see DR | NA |
|  | MNPDR | 0 | 0 | 1 | Y, only OCT | NA-only OCT available, cannot see DR | NA |
|  | MNPDR | 0 | 1 | 1 | Y, only OCT | NA-only OCT available, cannot see DR | NA |
|  | NSDR | 0 | 0 | 1 | Y, only OCT | NA-only OCT available, cannot see DR | NA |
|  | NSDR | 0 | 1 | 1 | Y, only OCT | NA-only OCT available, cannot see DR | NA |
|  | MNPDR | 0 | 0 | 1 | Y, only OCT | NA-only OCT available, cannot see DR | NA |
|  | MNPDR | 0 | 1 | 1 | Y, only OCT | NA-only OCT available, cannot see DR | NA |
|  | NSDR | 0 | 0 | 1 | Y, only OCT | NA-only OCT available, cannot see DR | NA |
|  | NSDR | 0 | 1 | 1 | Y, only OCT | NA-only OCT available, cannot see DR | NA |
|  | NSDR | 0 | 0 | 1 | Y, only OCT | NA-only OCT available, cannot see DR | NA |
|  | NSDR | 0 | 1 | 1 | Y, OCT and FANG | Y | N - should be MNPDR |
|  | MNPDR | 0 | 1 | 1 | Y, OCT only | NA-only OCT available, cannot see DR | NA |
|  | MNPDR | 0 | 0 | 1 | Y, OCT only | NA-only OCT available, cannot see DR | NA |
|  | MNPDR | 0 | 1 | 1 | Y, OCT only | NA-only OCT available, cannot see DR | NA |
|  | MNPDR | 0 | 0 | 1 | Y, OCT only | NA-only OCT available, cannot see DR | NA |
|  | MNPDR | 0 | 1 | 1 | Y, OCT only | NA-only OCT available, cannot see DR | NA |
|  | MNPDR | 0 | 0 | 1 | Y, OCT only | NA-only OCT available, cannot see DR | NA |
|  | MNPDR | 0 | 1 | 1 | Y, OCT only | NA-only OCT available, cannot see DR | NA |
|  | MNPDR | 0 | 0 | 1 | Y, OCT only | NA-only OCT available, cannot see DR | NA |
|  | MNPDR | 0 | 1 | 1 | Y, OCT only | NA-only OCT available, cannot see DR | NA |
|  | MNPDR | 0 | 0 | 1 | Y, OCT only | NA-only OCT available, cannot see DR | NA |
|  | MNPDR | 0 | 1 | 1 | Y, OCT only | NA-only OCT available, cannot see DR | NA |
|  | MNPDR | 0 | 0 | 1 | Y, OCT only | NA-only OCT available, cannot see DR | NA |
|  | MNPDR | 0 | 1 | 1 | Y, OCT only | NA-only OCT available, cannot see DR | NA |
|  | MNPDR | 0 | 0 | 1 | Y, OCT only | NA-only OCT available, cannot see DR | NA |
|  | MNPDR | 0 | 0 | 1 | Y, OCT only | NA-only OCT available, cannot see DR | NA |
|  | MNPDR | 0 | 1 | 1 | Y, OCT only | NA-only OCT available, cannot see DR | NA |
|  | MNPDR | 0 | 0 | 1 | Y, OCT only | NA-only OCT available, cannot see DR | NA |
|  | MNPDR | 0 | 1 | 1 | Y, OCT only | NA-only OCT available, cannot see DR | NA |
|  | MNPDR | 0 | 0 | 1 | Y, OCT only | NA-only OCT available, cannot see DR | NA |
|  | MNPDR | 0 | 1 | 1 | Y, OCT only | NA-only OCT available, cannot see DR | NA |
|  | MNPDR | 0 | 0 | 1 | Y, OCT only | NA-only OCT available, cannot see DR | NA |
|  | MNPDR | 0 | 1 | 1 | Y, OCT only | NA-only OCT available, cannot see DR | NA |
| **Patient 10** | MNPDR | 1 | 0 | 1 | Y | Y | Y |
|  | MNPDR | 0 | 0 | 1 | Y | Y | Y |
| **Patient 11** | MNPDR | 1 | 0 | 1 | Y | Y | Y |
|  | MNPDR | 0 | 0 | 1 | Y | Y | Y |
|  | MNPDR | 0 | 0 | 1 | Y - OCT only | NA-only OCT available, cannot see DR | NA |
|  | MNPDR | 0 | 0 | 1 | Y | Y | Y |
|  | MNPDR | 0 | 0 | 1 | Y | Y | Y |
|  | snpdr_hf | 0 | 0 | 1 | Y | Y | Y |
| **Patient 12** | MNPDR | 0 | 0 | 1 | N | NA | NA |
|  | NSDR | 0 | 0 | 1 | N | NA | NA |
|  | MNPDR | 0 | 0 | 1 | N | NA | NA |
|  | MNPDR | 0 | 1 | 1 | Y - OCT only | NA-only OCT available, cannot see DR | NA |
|  | MNPDR | 0 | 0 | 1 | Y - OCT only | NA-only OCT available, cannot see DR | NA |
|  | MNPDR | 0 | 0 | 1 | N | NA | NA |
| **Patient 13** | NSDR | 0 | 1 | 1 | N | NA | NA |
|  | NSDR | 0 | 1 | 1 | N OCT coded but not visible | NA | NA |
|  | NSDR | 0 | 0 | 1 | N OCT coded but not visible | NA | NA |
|  | MNPDR | 0 | 1 | 1 | N OCT coded but not visible | NA | NA |
|  | MNPDR | 0 | 0 | 1 | N OCT coded but not visible | NA | NA |
|  | NSDR | 0 | 0 | 1 | Y - OCT only | NA | NA |
|  | MNPDR | 0 | 1 | 1 | Y - OCT and photo | Y | N - lists NS but definitely MNDR |
|  | MNPDR | 0 | 0 | 1 | Y - OCT and photo | Y | N - lists NS but definitely MNDR |
|  | NSDR | 0 | 1 | 1 | Y | Y | N - lists NS but definitely MNDR |
|  | NSDR | 1 | 0 | 1 | Y | Y | Y |
|  | NSDR | 0 | 0 | 1 | Y | Y | Y |
|  | MNPDR | 0 | 0 | 1 | Y - OCT and photo | Y | Y |
|  | MNPDR | 0 | 0 | 1 | Y - OCT and photo | Y | NA |
| **Patient 14** | pdr_hr | 0 | 0 | 1 | Y | Y | Y |
| **Patient 15** | NSDR | 0 | 0 | 1 | N | NA | NA |
| **Patient 16** | NSDR | 1 | 0 | 1 | Y | Y | Y |
|  | NSDR | 0 | 0 | 1 | Y | Y | Y |
|  | MNPDR | 1 | 0 | 1 | Y | Y | Y - mild NPDR |
|  | MNPDR | 0 | 0 | 1 | Y | Y | Y |
|  | MNPDR | 0 | 0 | 1 | Y | Y | Y |
|  | MNPDR | 0 | 0 | 1 | Y | Y | Y |
| **Patient 17** | NSDR | 0 | 0 | 1 | N - only RNFL OCT | NA | NA |
| **Patient 18** | MNDR | 0 | 0 | 1 | Y | Y | Y |
| **Patient 19** | NSDR | 1 | 0 | 1 | Y | Y | Y |
|  | NSDR | 0 | 0 | 1 | Y | Y | Y |
|  | MNPDR | 1 | 0 | 1 | Y | Y | Y |
|  | MNPDR | 0 | 0 | 1 | Y | Y | Y |
| **Patient 20** | MNPDR | 0 | 1 | 1 | N - OCT only | NA-only OCT available, cannot see DR | NA |
|  | MNPDR | 0 | 0 | 1 | N - OCT only | NA-only OCT available, cannot see DR | NA |
|  | MNPDR | 1 | 0 | 1 | Y | N - 1 poor quality photo | N - lists MNDR but PDR also coded |
|  | MNPDR | 0 | 0 | 1 | Y | N - 1 poor quality photo | N - lists MNDR but PDR also coded |
|  | MNPDR | 0 | 1 | 1 | Y | Y | N - Should be PDR (has NVD) |
|  | MNPDR | 1 | 0 | 1 | Y | Y | N - Should be PDR (has NVD) |
|  | MNPDR | 0 | 0 | 1 | Y | Y | N - Should be PDR (has NVD) |
| **Patient 21** | NSDR | 0 | 0 | 1 | N | NA | NA |
| **Patient 22** | MNPDR | 1 | 0 | 1 | Y | Y | Y - MNDR |
|  | MNPDR | 0 | 1 | 1 | Y | Y | Y - MNDR |
|  | MNPDR | 0 | 0 | 1 | Y | Y | Y - MNDR |
|  | MNPDR | 0 | 1 | 1 | Y OCT only | NA-only OCT available, cannot see DR | NA |
|  | MNPDR | 0 | 0 | 1 | Y OCT only | NA-only OCT available, cannot see DR | NA |
|  | MNPDR | 0 | 1 | 1 | Y | Y | Y - MNDR |
|  | MNPDR | 1 | 0 | 1 | Y | Y | Y - MNDR |
|  | MNPDR | 0 | 0 | 1 | Y | Y | Y - MNDR |
|  | MNPDR | 0 | 1 | 1 | Y | Y | Y - MNDR |
|  | MNPDR | 1 | 0 | 1 | Y | Y | Y - MNDR |
|  | MNPDR | 0 | 0 | 1 | Y | Y | Y - MNDR |
|  | MNPDR | 1 | 0 | 1 | Y | Y | Y - MNDR |
|  | MNPDR | 0 | 1 | 1 | Y | Y | Y - MNDR |
|  | MNPDR | 0 | 0 | 1 | Y | Y | Y - MNDR |
| **Patient 23** | MNPDR | 1 | 0 | 1 | Y | Y | Y -MNPDR |
|  | MNPDR | 0 | 0 | 1 | Y | Y | Y -MNPDR |
|  | MNPDR | 0 | 1 | 1 | Y OCT only | NA-only OCT available, cannot see DR | NA |
|  | MNPDR | 0 | 0 | 1 | Y OCT only | NA-only OCT available, cannot see DR | NA |
|  | MNPDR | 0 | 1 | 1 | Y OCT only | NA-only OCT available, cannot see DR | NA |
|  | MNPDR | 0 | 0 | 1 | Y OCT only | NA-only OCT available, cannot see DR | NA |
|  | MNPDR | 0 | 1 | 1 | Y OCT only | NA-only OCT available, cannot see DR | NA |
|  | MNPDR | 0 | 0 | 1 | Y OCT only | NA-only OCT available, cannot see DR | NA |
|  | MNPDR | 0 | 1 | 1 | Y OCT only | NA-only OCT available, cannot see DR | NA |
|  | MNPDR | 0 | 0 | 1 | Y OCT only | NA-only OCT available, cannot see DR | NA |
|  | MNPDR | 0 | 1 | 1 | Y OCT only | NA-only OCT available, cannot see DR | NA |
|  | MNPDR | 0 | 0 | 1 | Y OCT only | NA-only OCT available, cannot see DR | NA |
|  | MNPDR | 0 | 1 | 1 | Y OCT only | NA-only OCT available, cannot see DR | NA |
|  | MNPDR | 0 | 0 | 1 | Y OCT only | NA-only OCT available, cannot see DR | NA |
|  | MNPDR | 0 | 1 | 1 | Y OCT only | NA-only OCT available, cannot see DR | NA |
|  | MNPDR | 0 | 0 | 1 | Y OCT only | NA-only OCT available, cannot see DR | NA |
|  | MNPDR | 0 | 1 | 1 | Y OCT only | NA-only OCT available, cannot see DR | NA |
|  | MNPDR | 0 | 0 | 1 | Y OCT only | NA-only OCT available, cannot see DR | NA |
|  | MNPDR | 0 | 1 | 1 | N | N - OCT was coded as photos | NA |
|  | MNPDR | 0 | 0 | 1 | N | N - OCT was coded as photos | NA |
|  | MNPDR | 0 | 0 | 1 | N | N - OCT was coded as photos | NA |
| **Patient 24** | MNPDR | 0 | 0 | 1 | Y | Y | Y |
|  | MNPDR | 0 | 1 | 1 | Y OCT only | NA-only OCT available, cannot see DR | NA |
|  | MNPDR | 0 | 0 | 1 | Y OCT only | NA-only OCT available, cannot see DR | NA |
|  | MNPDR | 0 | 0 | 1 | Y OCT only | NA-only OCT available, cannot see DR | NA |
|  | MNPDR | 0 | 0 | 1 | Y OCT only | NA-only OCT available, cannot see DR | NA |
|  | MNPDR | 0 | 1 | 1 | Y OCT only | NA-only OCT available, cannot see DR | NA |
|  | MNPDR | 0 | 0 | 1 | Y OCT only | NA-only OCT available, cannot see DR | NA |
|  | MNPDR | 0 | 1 | 1 | Y OCT only | NA-only OCT available, cannot see DR | NA |
|  | MNPDR | 0 | 0 | 1 | Y OCT only | NA-only OCT available, cannot see DR | NA |
|  | MNPDR | 0 | 1 | 1 | Y OCT only | NA-only OCT available, cannot see DR | NA |
|  | MNPDR | 0 | 0 | 1 | Y OCT only | NA-only OCT available, cannot see DR | NA |
|  | MNPDR | 0 | 1 | 1 | Y OCT only | NA-only OCT available, cannot see DR | NA |
|  | MNPDR | 0 | 0 | 1 | Y OCT only | NA-only OCT available, cannot see DR | NA |
|  | MNPDR | 0 | 1 | 1 | Y OCT only | NA-only OCT available, cannot see DR | NA |
|  | MNPDR | 0 | 0 | 1 | Y OCT only | NA-only OCT available, cannot see DR | NA |
|  | MNPDR | 0 | 0 | 1 | Y OCT only | NA-only OCT available, cannot see DR | NA |
|  | MNPDR | 0 | 1 | 1 | Y OCT only | NA-only OCT available, cannot see DR | NA |
|  | MNPDR | 0 | 0 | 1 | Y OCT only | NA-only OCT available, cannot see DR | NA |
|  | MNPDR | 0 | 1 | 1 | Y OCT only | NA-only OCT available, cannot see DR | NA |
|  | MNPDR | 0 | 0 | 1 | Y OCT only | NA-only OCT available, cannot see DR | NA |
|  | MNPDR | 0 | 0 | 1 | N | NA | NA |
|  | MNPDR | 0 | 0 | 1 | Y OCT only | NA-only OCT available, cannot see DR | NA |
|  | MNPDR | 0 | 1 | 1 | Y OCT only | NA-only OCT available, cannot see DR | NA |
|  | MNPDR | 0 | 0 | 1 | Y OCT only | NA-only OCT available, cannot see DR | NA |
|  | MNPDR | 0 | 1 | 1 | Y OCT only | NA-only OCT available, cannot see DR | NA |
|  | MNPDR | 0 | 0 | 1 | Y OCT only | NA-only OCT available, cannot see DR | NA |
|  | MNPDR | 0 | 1 | 1 | Y OCT only | NA-only OCT available, cannot see DR | NA |
|  | MNPDR | 0 | 0 | 1 | Y OCT only | NA-only OCT available, cannot see DR | NA |
|  | MNPDR | 0 | 1 | 1 | Y OCT only | NA-only OCT available, cannot see DR | NA |
|  | MNPDR | 0 | 1 | 1 | Y OCT only | NA-only OCT available, cannot see DR | NA |
|  | MNPDR | 0 | 0 | 1 | Y OCT only | NA-only OCT available, cannot see DR | NA |
| **Patient 25** | NSDR | 0 | 0 | 1 | No imaging available | NA | NA |
| **Patient 26** | MNPDR | 0 | 0 | 1 | Y OCT only | NA-only OCT available, cannot see DR | NA |
|  | MNPDR | 0 | 1 | 1 | Y OCT only | NA-only OCT available, cannot see DR | NA |
|  | MNPDR | 0 | 0 | 1 | Y OCT only | NA-only OCT available, cannot see DR | NA |
| **Patient 27** | NSDR | 0 | 0 | 1 | No imaging available | NA | NA |
|  | MNPDR | 0 | 0 | 1 | No imaging available | NA | NA |
| **Patient 28** | NSDR | 1 | 0 | 1 | Y | Y | N - MNDR (exudates OS) |
|  | NSDR | 0 | 0 | 1 | Y | Y | N - MNDR (exudates OS) |
|  | MNPDR | 0 | 1 | 1 | Y | Y | Y |
|  | MNPDR | 0 | 0 | 1 | Y OCT only | NA-only OCT available, cannot see DR | NA |
|  | MNPDR | 0 | 1 | 1 | Y OCT only | NA-only OCT available, cannot see DR | NA |
|  | MNPDR | 0 | 0 | 1 | Y OCT only | NA-only OCT available, cannot see DR | NA |
|  | MNPDR | 0 | 1 | 1 | Y OCT only | NA-only OCT available, cannot see DR | NA |
|  | MNPDR | 0 | 0 | 1 | Y OCT only | NA-only OCT available, cannot see DR | NA |
|  | MNPDR | 0 | 1 | 1 | Y OCT only | NA-only OCT available, cannot see DR | NA |
|  | MNPDR | 0 | 0 | 1 | Y OCT only | NA-only OCT available, cannot see DR | NA |
|  | MNPDR | 0 | 1 | 1 | Y OCT only | NA-only OCT available, cannot see DR | NA |
|  | MNPDR | 0 | 0 | 1 | Y OCT only | NA-only OCT available, cannot see DR | NA |
|  | MNPDR | 0 | 1 | 1 | Y OCT only | NA-only OCT available, cannot see DR | NA |
|  | MNPDR | 0 | 0 | 1 | Y OCT only | NA-only OCT available, cannot see DR | NA |
|  | MNPDR | 0 | 1 | 1 | Y OCT only | NA-only OCT available, cannot see DR | NA |
|  | MNPDR | 0 | 0 | 1 | Y OCT only | NA-only OCT available, cannot see DR | NA |
|  | MNPDR | 0 | 1 | 1 | Y OCT only | NA-only OCT available, cannot see DR | NA |
|  | MNPDR | 0 | 0 | 1 | Y OCT only | NA-only OCT available, cannot see DR | NA |
|  | MNPDR | 0 | 1 | 1 | Y OCT only | NA-only OCT available, cannot see DR | NA |
|  | MNPDR | 0 | 0 | 1 | Y OCT only | NA-only OCT available, cannot see DR | NA |
|  | MNPDR | 0 | 1 | 1 | No imaging available | NA | NA |
|  | MNPDR | 0 | 0 | 1 | No imaging available | NA | NA |
|  | MNPDR | 0 | 1 | 1 | Y OCT only | NA-only OCT available, cannot see DR | NA |
|  | MNPDR | 0 | 0 | 1 | Y OCT only | NA-only OCT available, cannot see DR | NA |
|  | MNPDR | 0 | 1 | 1 | Y OCT only | NA-only OCT available, cannot see DR | NA |
|  | MNPDR | 0 | 0 | 1 | Y OCT only | NA-only OCT available, cannot see DR | NA |
|  | MNPDR | 0 | 1 | 1 | N | NA | NA |
|  | MNPDR | 1 | 0 | 1 | N | N - OCT was coded as photos | NA |
|  | MNPDR | 0 | 0 | 1 | N | NA | NA |
|  | MNPDR | 0 | 1 | 1 | Y OCT only | NA-only OCT available, cannot see DR | NA |
|  | MNPDR | 0 | 0 | 1 | Y OCT only | NA-only OCT available, cannot see DR | NA |
|  | MNPDR | 0 | 1 | 1 | Y OCT only | NA-only OCT available, cannot see DR | NA |
|  | MNPDR | 1 | 0 | 1 | N | N - OCT was coded as photos | NA |
|  | MNPDR | 0 | 0 | 1 | Y OCT only | NA-only OCT available, cannot see DR | NA |
|  | MNPDR | 0 | 1 | 1 | Y OCT only | NA-only OCT available, cannot see DR | NA |
|  | MNPDR | 0 | 0 | 1 | Y OCT only | NA-only OCT available, cannot see DR | NA |
|  | MNPDR | 0 | 1 | 1 | Y OCT only | NA-only OCT available, cannot see DR | NA |
|  | MNPDR | 0 | 0 | 1 | Y OCT only | NA-only OCT available, cannot see DR | NA |
|  | MNPDR | 0 | 1 | 1 | Y OCT only | NA-only OCT available, cannot see DR | NA |
|  | MNPDR | 0 | 0 | 1 | Y OCT only | NA-only OCT available, cannot see DR | NA |
|  | MNPDR | 0 | 1 | 1 | No imaging available | NA | NA |
|  | MNPDR | 0 | 0 | 1 | No imaging available | NA | NA |
|  | MNPDR | 0 | 1 | 1 | Y OCT only | NA-only OCT available, cannot see DR | NA |
|  | MNPDR | 0 | 0 | 1 | Y OCT only | NA-only OCT available, cannot see DR | NA |
| **Patient 29** | NSDR | 0 | 0 | 1 | No imaging available | NA | NA |
|  | NSDR | 0 | 1 | 1 | Y OCT only | NA-only OCT available, cannot see DR | NA |
| **Patient 30** | NSDR | 0 | 0 | 1 | No imaging available | NA | NA |
| **Patient 31** | MNPDR | 1 | 0 | 1 | No imaging available | NA | NA |
|  | MNPDR | 0 | 1 | 1 | Y OCT only | NA-only OCT available, cannot see DR | NA |
|  | MNPDR | 0 | 0 | 1 | Y OCT only | NA-only OCT available, cannot see DR | NA |
|  | MNPDR | 0 | 1 | 1 | Y OCT only | NA-only OCT available, cannot see DR | NA |
|  | MNPDR | 0 | 0 | 1 | Y OCT only | NA-only OCT available, cannot see DR | NA |
|  | MNPDR | 0 | 0 | 1 | Y OCT only | NA-only OCT available, cannot see DR | NA |
|  | NSDR | 0 | 0 | 1 | No imaging available | NA | NA |
|  | MNPDR | 0 | 1 | 1 | Y OCT only | NA-only OCT available, cannot see DR | NA |
|  | MNPDR | 0 | 0 | 1 | Y OCT only | NA-only OCT available, cannot see DR | NA |
|  | MNPDR | 0 | 1 | 1 | Y OCT only | NA-only OCT available, cannot see DR | NA |
|  | MNPDR | 0 | 0 | 1 | Y OCT only | NA-only OCT available, cannot see DR | NA |
|  | MNPDR | 0 | 1 | 1 | Y OCT only | NA-only OCT available, cannot see DR | NA |
|  | MNPDR | 0 | 0 | 1 | Y OCT only | NA-only OCT available, cannot see DR | NA |
|  | MNPDR | 0 | 1 | 1 | Y OCT only | NA-only OCT available, cannot see DR | NA |
|  | MNPDR | 0 | 0 | 1 | Y OCT only | NA-only OCT available, cannot see DR | NA |
|  | MNPDR | 0 | 1 | 1 | Y OCT only | NA-only OCT available, cannot see DR | NA |
|  | MNPDR | 0 | 0 | 1 | Y OCT only | NA-only OCT available, cannot see DR | NA |
|  | MNPDR | 0 | 0 | 1 | No imaging available | NA | NA |
|  | MNPDR | 0 | 1 | 1 | Y | Y | Y - MNPDR |
|  | MNPDR | 1 | 0 | 1 | Y | Y | Y - MNPDR |
|  | MNPDR | 0 | 0 | 1 | Y | Y | Y - MNPDR |
|  | MNPDR | 1 | 0 | 1 | Y | Y | Y - MNPDR |
|  | MNPDR | 0 | 1 | 1 | Y | Y | Y - MNPDR |
|  | MNPDR | 0 | 0 | 1 | Y | Y | Y - MNPDR |
|  | MNPDR | 0 | 1 | 1 | Y | Y | Y - MNPDR |
|  | MNPDR | 1 | 0 | 1 | Y | Y | Y - MNPDR |
|  | MNPDR | 0 | 0 | 1 | Y | Y | Y - MNPDR |
|  | MNPDR | 1 | 0 | 1 | N | NA - no photos available | NA |
|  | MNPDR | 0 | 1 | 1 | N | NA - no photos available | NA |
|  | MNPDR | 0 | 0 | 1 | N | NA - no photos available | NA |
|  | MNPDR | 0 | 1 | 1 | Y OCT only | NA-only OCT available, cannot see DR | NA |
|  | MNPDR | 0 | 0 | 1 | Y OCT only | NA-only OCT available, cannot see DR | NA |
|  | MNPDR | 1 | 0 | 1 | Y | Y | Y - MNPDR |
|  | MNPDR | 0 | 1 | 1 | Y | Y | Y - MNPDR |
|  | MNPDR | 0 | 0 | 1 | Y | Y | Y - MNPDR |
|  | MNPDR | 0 | 1 | 1 | Y | Y | Y - MNPDR |
|  | MNPDR | 1 | 0 | 1 | Y | Y | Y - MNPDR |
|  | MNPDR | 0 | 0 | 1 | Y | Y | Y - MNPDR |
| **Patient 32** | NSDR | 0 | 0 | 1 | No imaging available | NA | NA |
|  | NSDR | 0 | 0 | 1 | Y OCT only | NA-only OCT available, cannot see DR | NA |
|  | NSDR | 0 | 1 | 1 | Y OCT only | NA-only OCT available, cannot see DR | NA |
|  | NSDR | 0 | 0 | 1 | Y OCT only | NA-only OCT available, cannot see DR | NA |
|  | MNPDR | 0 | 1 | 1 | Y OCT only | NA-only OCT available, cannot see DR | NA |
|  | MNPDR | 0 | 0 | 1 | Y OCT only | NA-only OCT available, cannot see DR | NA |
| **Patient 33** | MNPDR | 0 | 0 | 1 | No imaging available | NA | NA |
|  | MNPDR | 1 | 0 | 1 | Y | Y | Y- MNPDR |
|  | MNPDR | 0 | 0 | 1 | Y | Y | Y- MNPDR |
|  | NSDR | 1 | 0 | 1 | Y | Y | Y- MNPDR |
|  | NSDR | 0 | 0 | 1 | Y | Y | Y- MNPDR |
|  | MNPDR | 1 | 0 | 1 | Y | Y | Y- MNPDR |
|  | MNPDR | 0 | 0 | 1 | Y | Y | Y- MNPDR |
|  | NSDR | 1 | 0 | 1 | Y | Y | Y- MNPDR |
|  | NSDR | 0 | 0 | 1 | Y | Y | Y- MNPDR |
|  | NSDR | 1 | 0 | 1 | Y | Y | Y- MNPDR |
|  | NSDR | 0 | 0 | 1 | Y | Y | Y- MNPDR |
|  | MNPDR | 1 | 0 | 1 | Y | Y | Y- MNPDR |
|  | MNPDR | 0 | 0 | 1 | Y | Y | Y- MNPDR |
|  | NSDR | 0 | 0 | 1 | No imaging available | NA | NA |
| **Patient 34** | snpdr_hf | 1 | 0 | 1 | Y | Y | Y- SNPDR |
|  | snpdr_hf | 0 | 0 | 1 | Y | Y | Y- SNPDR |
| **Patient 35** | NSDR | 0 | 0 | 1 | No imaging available | NA | NA |
|  | NSDR | 0 | 0 | 1 | No imaging available | NA | NA |
|  | NSDR | 0 | 0 | 1 | No imaging available | NA | NA |
| **Patient 36** | NSDR | 0 | 0 | 1 | No imaging available | NA | NA |
| **Patient 37** | NSDR | 0 | 0 | 1 | Y | Y | Y-NSDR |
|  | NSDR | 0 | 0 | 1 | No imaging available | NA | NA |
|  | NSDR | 0 | 0 | 1 | No imaging available | NA | NA |
|  | NSDR | 0 | 0 | 1 | No imaging available | NA | NA |
|  | NSDR | 0 | 1 | 1 | Y OCT only | NA-only OCT available, cannot see DR | NA |
|  | NSDR | 0 | 0 | 1 | Y OCT only | NA-only OCT available, cannot see DR | NA |
|  | MNPDR | 0 | 0 | 1 | No imaging available | NA | NA |
|  | NSDR | 0 | 1 | 1 | Y OCT only | NA-only OCT available, cannot see DR | NA |
|  | NSDR | 0 | 0 | 1 | Y OCT only | NA-only OCT available, cannot see DR | NA |
| **Patient 38** | NSDR | 0 | 0 | 1 | No imaging available | NA | NA |
|  | NSDR | 0 | 0 | 1 | No imaging available | NA | NA |
| **Patient 39** | NSDR | 0 | 0 | 1 | No imaging available | NA | NA |
|  | NSDR | 0 | 0 | 1 | No imaging available | NA | NA |
|  | NSDR | 0 | 0 | 1 | No imaging available | NA | NA |
|  | NSDR | 0 | 0 | 1 | No imaging available | NA | NA |
|  | NSDR | 0 | 0 |  | No imaging available | NA | NA |
|  | NSDR | 0 | 0 | 1 | No imaging available | NA | NA |
| **Patient 40** | NSDR | 0 | 0 | 1 | Y | Y | Y MNPDR |
|  | NSDR | 0 | 1 | 1 | Y OCT only | NA-only OCT available, cannot see DR | NA |
|  | NSDR | 0 | 0 | 1 | Y OCT only | NA-only OCT available, cannot see DR | NA |
|  | NSDR | 0 | 1 | 1 | Y OCT only | NA-only OCT available, cannot see DR | NA |
|  | NSDR | 0 | 0 | 1 | Y OCT only | NA-only OCT available, cannot see DR | NA |
| **Patient 41** | MNPDR | 0 | 1 | 1 | Y OCT only | NA-only OCT available, cannot see DR | NA |
|  | MNPDR | 0 | 0 | 1 | Y OCT only | NA-only OCT available, cannot see DR | NA |
|  | NSDR | 0 | 1 | 1 | Y OCT only | NA-only OCT available, cannot see DR | NA |
|  | NSDR | 0 | 0 | 1 | Y OCT only | NA-only OCT available, cannot see DR | NA |
|  | MNPDR | 0 | 1 | 1 | Y OCT only | NA-only OCT available, cannot see DR | NA |
|  | MNPDR | 0 | 0 | 1 | Y OCT only | NA-only OCT available, cannot see DR | NA |
|  | MNPDR | 0 | 1 | 1 | Y OCT only | NA-only OCT available, cannot see DR | NA |
|  | MNPDR | 0 | 0 | 1 | Y OCT only | NA-only OCT available, cannot see DR | NA |
|  | MNPDR | 0 | 1 | 1 | Y | Y | Y- MNPDR |
|  | MNPDR | 0 | 0 | 1 | Y | Y | Y- MNPDR |
|  | MNPDR | 0 | 1 | 1 | Y | Y | N- IRMA listed so should be SNPDR |
|  | MNPDR | 1 | 0 | 1 | Y | Y | N- IRMA listed so should be SNPDR |
|  | MNPDR | 0 | 0 | 1 | Y | Y | N- IRMA listed so should be SNPDR |
|  | MNPDR | 0 | 1 | 1 | Y | Y | Y- MNPDR |
|  | MNPDR | 0 | 0 | 1 | Y | Y | Y- MNPDR |
|  | MNPDR | 0 | 1 | 1 | Y OCT only | NA-only OCT available, cannot see DR | NA |
|  | MNPDR | 0 | 0 | 1 | Y OCT only | NA-only OCT available, cannot see DR | NA |
| **Patient 42** | NSDR | 0 | 0 | 1 | No imaging available | NA | NA |
| **Patient 43** | MNPDR | 1 | 0 | 1 | Y | N - image too poor | Unable |
|  | MNPDR | 0 | 0 | 1 | Y | N - image too poor | Unable |
|  | snpdr_hf | 1 | 0 | 1 | Y | N - image too poor | Unable |
|  | snpdr_hf | 0 | 0 | 1 | Y | N - image too poor | Unable |
| **Patient 44** | MNPDR | 0 | 0 | 1 | No imaging available | NA | NA |
|  |  |  |  | N-total visits with imaging | 105 | 99 | 84 |
|  |  |  |  | % Accuracy |  | 94.29% | 84.85% |
|  |  |  |  | N-unique individuals with imaging | 24 | 23 | 21 |
|  |  |  |  | % Accuracy |  | 95.83% | 91.30% |
